# Molecular Stratification of Chronic Kidney Disease

**DOI:** 10.1101/2021.09.09.21263234

**Authors:** Anna Reznichenko, Viji Nair, Sean Eddy, Mark Tomilo, Timothy Slidel, Wenjun Ju, James P. Conway, Shawn S. Badal, Johnna Wesley, John T. Liles, Sven Moosmang, Julie M. Williams, Carol Moreno Quinn, Markus Bitzer, Anil Karihaloo, Matthew D. Breyer, Kevin L. Duffin, Uptal D. Patel, Maria Chiara Magnone, Ratan Bhat, Matthias Kretzler

## Abstract

Current classification of chronic kidney disease (CKD) into stages based on the indirect measures of kidney functional state, estimated glomerular filtration rate and albuminuria, is agnostic to the heterogeneity of underlying etiologies, histopathology, and molecular processes. We used genome-wide transcriptomics from patients’ kidney biopsies, directly reflecting kidney biological processes, to stratify patients from three independent CKD cohorts. Unsupervised Self-Organizing Maps (SOM), an artificial neural network algorithm, assembled CKD patients into four novel subgroups, molecular categories, based on the similarity of their kidney transcriptomics profiles. The unbiased, molecular categories were present across CKD stages and histopathological diagnoses, highlighting heterogeneity of conventional clinical subgroups at the molecular level. CKD molecular categories were distinct in terms of biological pathways, transcriptional regulation and associated kidney cell types, indicating that the molecular categorization is founded on biologically meaningful mechanisms. Importantly, our results revealed that not all biological pathways are equally activated in all patients; instead, different pathways could be more dominant in different subgroups and thereby differentially influencing disease progression and outcomes. This first kidney-centric unbiased categorization of CKD paves the way to an integrated clinical, morphological and molecular diagnosis. This is a key step towards enabling precision medicine for this heterogeneous condition with the potential to advance biological understanding, clinical management, and drug development, as well as establish a roadmap for molecular reclassification of CKD and other complex diseases.

**One sentence summary:** Unbiased grouping of patients based on kidney biopsy transcriptomics profiles generated a novel molecular categorization of chronic kidney disease.

## Introduction

Chronic kidney disease (CKD) is a global health burden affecting nearly 700 million people worldwide, with a high cost of care to individuals and health systems. It is an independent risk factor for cardiovascular disease and is associated with increased morbidity and mortality (*1, 2*).

Biologically, CKD is a highly heterogeneous group of disorders characterized by sustained alterations in kidney structure and function arising from a wide range of etiologies (the most common are diabetic and hypertensive nephropathies) and associated with a multitude of underlying molecular processes in the kidney (*3*). Even within the same etiological group, individual disease presentation, histopathological features, progression rates and treatment responses are variable, reflecting the underlying biological heterogeneity.

According to the KDIGO (Kidney Disease Improving Global Outcomes) guidelines, CKD is classified into five stages (CKD 1-5) based on gradations of glomerular filtration rate (GFR), estimated from serum creatinine, and/or albuminuria (*3*). This, now widely accepted classification system, was a major achievement in the field as it provided a common vocabulary and standardized approach to disease management (*4-7*). However, the classification relies on systemic, indirect measures and is therefore agnostic to the intra-renal biology. As a result, biologically diverse cases presenting with the same GFR or albuminuria values are classified under the same CKD stage category, thereby precluding personalized prognosis and treatment options (*8*).

Histopathological diagnosis based on kidney biopsy provides more direct insights on the intra-renal state (*9*). However, several tissue morphological features may overlap between various kidney diseases. Furthermore, tissue molecular signatures also overlap, as previously revealed by a transcriptomics study of CKD biopsies, where patterns of molecular similarity spanned histopathological diagnoses (*10*).

Thus, an unbiased re-categorization of CKD by using kidney molecular profiles may circumvent the shortcomings of GFR staging or histopathological grouping. In a field where specific therapies are still lacking, such a mechanistic classification based on underlying disease biology is urgently needed to fuel diagnostic, prognostic, and therapeutic development that will enable the transition to precision medicine (*11-13*).

The overarching goal of this study was to determine whether a mechanistic disease classification based on intra-renal molecular characteristics can be achieved across multiple cohorts, as a critical step towards development of novel personalized treatments for CKD. We used individual transcriptomics profiles from kidney biopsies to group patients in an unbiased, data-driven manner. We analyzed an extensive collection of kidney transcriptomics data across three CKD patient cohorts from North America and Europe that encompass a broad range of kidney function and wide spectrum of diseases. Focusing on the tubulointerstitial compartment, the most abundantly available tissue fraction in kidney biopsies and closely linked to long-term outcomes of CKD (40), we employed an unsupervised artificial neural network algorithm, Self-Organizing Maps (SOM) (*14, 15*), to reveal inherent patient sub-groups. We then substantiated the clinical relevance of these sub-groups by assessing their association with biological pathways and clinical parameters.

## Results

### CKD patient population map construction and segmentation

A total of 314 CKD patients from European Renal cDNA Bank (ERCB)(*16*), Clinical Phenotyping Resource and Biobank Core (C-PROBE)(*17*), and Nephrotic Syndrome Study Network (NEPTUNE)(*18*) cohorts with available genome-wide transcriptomics data from kidney biopsy tubulointerstitial fraction were included in this study. Patient clinical characteristics are summarized in *Table 1*.

**Table 1.**
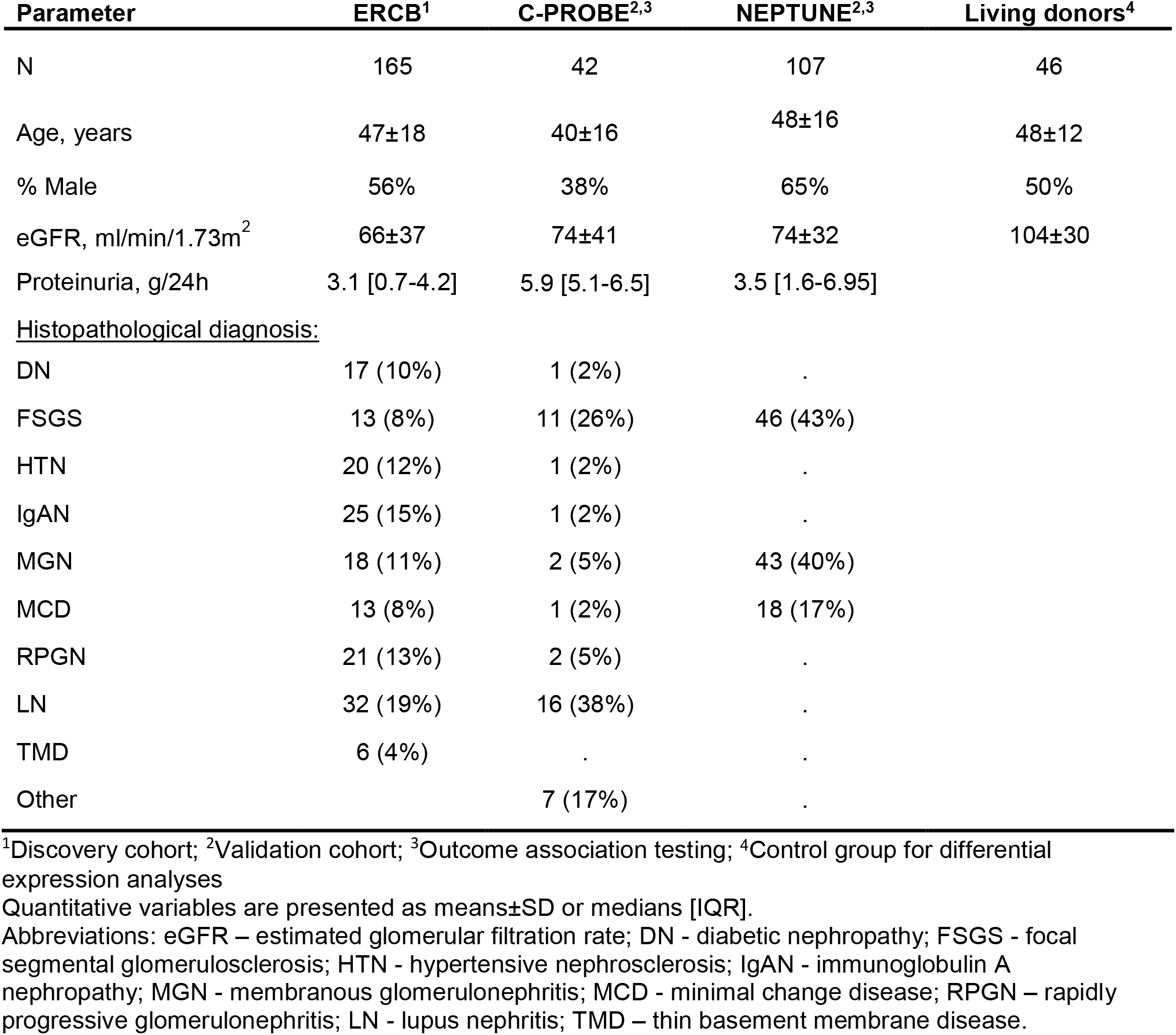
Patient clinical characteristics for discovery and validation cohorts

| Parameter | ERCB<sup>1</sup> | C-PROBE<sup>2,3</sup> | NEPTUNE<sup>2,3</sup> | Living donors<sup>4</sup> |
| --- | --- | --- | --- | --- |
| N | 165 | 42 | 107 | 46 |
| Age, years | 47±18 | 40±16 | 48±16 | 48±12 |
| % Male | 56% | 38% | 65% | 50% |
| eGFR, ml/min/1.73m<sup>2</sup> | 66±37 | 74±41 | 74±32 | 104±30 |
| Proteinuria, g/24h | 3.1 [0.7-4.2] | 5.9 [5.1-6.5] | 3.5 [1.6-6.95] |  |
| <u>Histopathological diagnosis:</u> |  |  |  |  |
| DN | 17 (10%) | 1 (2%) | . |  |
| FSGS | 13 (8%) | 11 (26%) | 46 (43%) |  |
| HTN | 20 (12%) | 1 (2%) | . |  |
| IgAN | 25 (15%) | 1 (2%) | . |  |
| MGN | 18 (11%) | 2 (5%) | 43 (40%) |  |
| MCD | 13 (8%) | 1 (2%) | 18 (17%) |  |
| RPGN | 21 (13%) | 2 (5%) | . |  |
| LN | 32 (19%) | 16 (38%) | . |  |
| TMD | 6 (4%) | . | . |  |
| Other |  | 7 (17%) | . |  |

<sup>1</sup>Discovery cohort; <sup>2</sup>Validation cohort; <sup>3</sup>Outcome association testing; <sup>4</sup>Control group for differential expression analyses
Quantitative variables are presented as means±SD or medians [IQR].
Abbreviations: eGFR – estimated glomerular filtration rate; DN - diabetic nephropathy; FSGS - focal segmental glomerulosclerosis; HTN - hypertensive nephrosclerosis; IgAN - immunoglobulin A nephropathy; MGN - membranous glomerulonephritis; MCD - minimal change disease; RPGN – rapidly progressive glomerulonephritis; LN - lupus nephritis; TMD – thin basement membrane disease.

Based on the large sample size as well as broadest range of kidney function and spectrum of histopathological diagnoses, the ERCB dataset was selected as the discovery dataset to maximize the pattern detection potential in the data. The independent datasets from C-PROBE and NEPTUNE, contributing complementary GFR ranges and etiologies, were then used for validation.

An unsupervised SOM of the ERCB CKD population was constructed based solely on patient kidney transcriptomics profiles, comprised of expression values for 8,454 genes. The algorithm mapped patients onto the 8×8 SOM grid, resulting in 1 to 8 individuals (median of 2) per each of the 64 SOM units (*Fig 1A*), placed in a topological order based on the relative similarities of their transcriptional profiles. Hierarchical clustering of SOM units was then performed to delineate subgroups with similar gene expression profiles. Four inherent patient clusters were thus identified within the discovery CKD population, named “molecular categories” and assigned arbitrary colors – Blue, Gold, Olive, Plum – in line with the unbiased nature of their discovery (*Fig 1B*). The larger Blue and Plum molecular categories accounted for 35% and 27% of the cases while 20% and 18% of the patients mapped to the Gold and Olive categories, respectively. Global gene expression levels were not different between the four molecular categories (*Fig S1*), indicating absence of technical bias but rather, qualitative differences in patterns underlying the subgrouping. This was further evident from inspecting the transcriptomics profiles visualization, showing distinct patterns between the molecular categories (*Fig 1C*).

**Fig 1.**
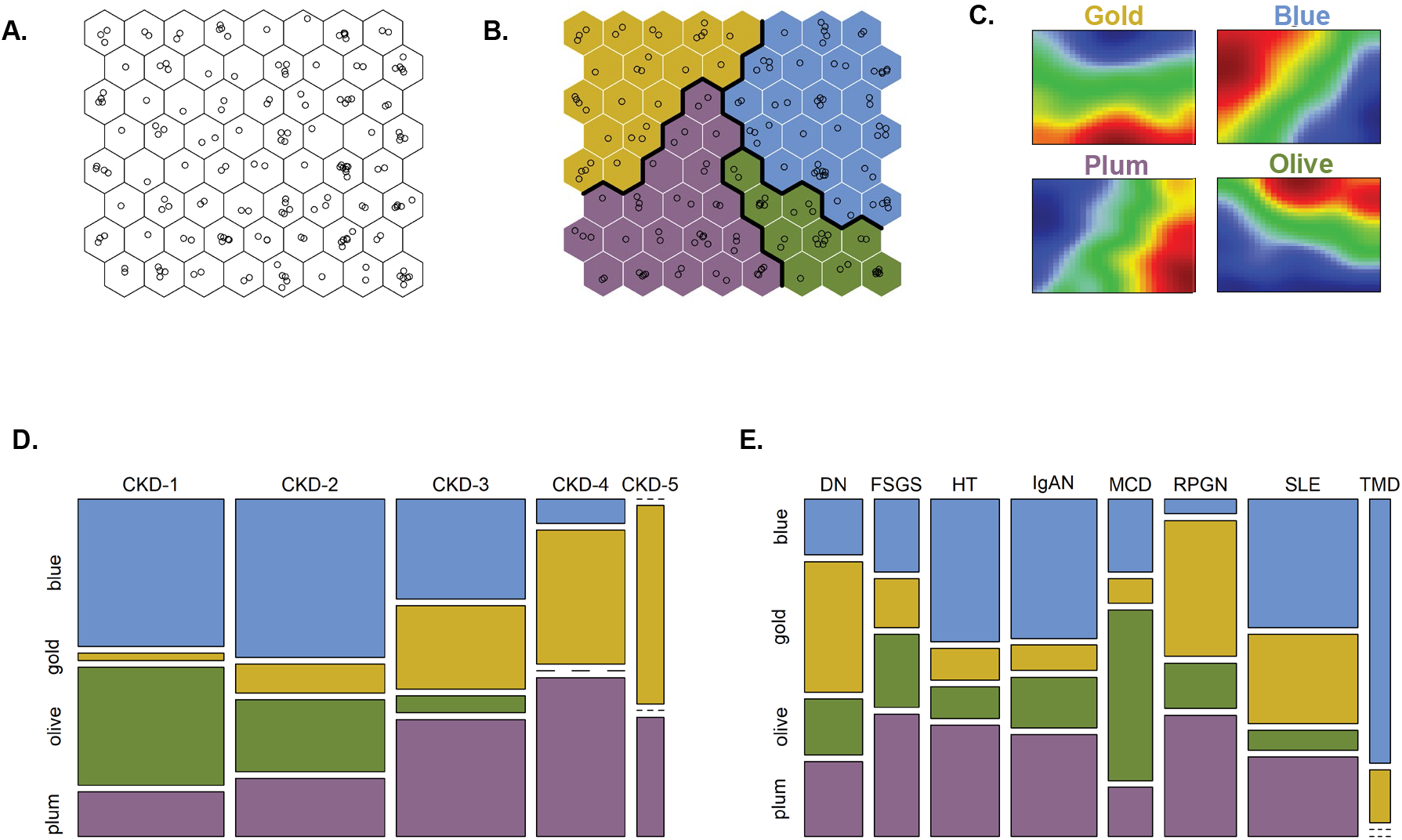
Unbiased kidney transcriptomics stratification identified four inherent subgroups of patients (“molecular categories”) within a CKD cohort. Molecular categories were present at different CKD stages and histopathological diagnoses, providing orthogonal biomechanistic information regardless of the disease etiology or severity. **(A)** Self-Organizing Map of a CKD population based on kidney gene expression profiling. Individual patients (shown as open circles) were arranged in a topological order by similarities of their multivariable transcriptomics profiles. **(B)** Clustering of similar SOM units identified subgroups of similar patients. Thick lines indicate cluster boundaries. Clusters were assigned arbitrary colors. **(C)** Group-level summarized transcriptomics profiles (“expression portraits”) show distinct patterns between the molecular categories. Color scale reflects relative gene expression levels (red – high, blue – low, green – average) of 8,454 transcripts mapped. **(D-E)** Mosaic plots show the correspondence between molecular categorization and CKD stages and histopathological diagnoses, respectively. Width of the bands reflects the relative proportions of cases, the numbers of patients per category are also indicated by numbers.

### CKD stages consist of molecularly heterogeneous categories

Having thus identified different molecular categories of patients with CKD based on kidney transcriptomics features, we next wanted to compare them with clinical CKD classifications.

*Figs 1D-E* visualize the correspondence between novel molecular categories and conventional clinical classification. The molecular categories were present across all CKD stages and histopathological diagnoses, highlighting heterogeneity of clinical subgroups at the molecular level. Gold and Plum categories were relatively more frequent in advanced disease stages (CKD 3-5), while CKD 1-3 patients were predominantly mapped to Blue and Olive categories (*Fig 1D*). However, the fact that the same molecular categories were observed at different CKD stages indicates independence from GFR as a potential confounder (*Fig S2* shows similar estimated GFR levels across molecular categories within same CKD stages strata). Likewise, each histopathologically defined patient group (*Fig 1E*) contained multiple molecular categories, highlighting within-group heterogeneity as well as between-group similarities. *Fig 2* illustrates the kidney molecular heterogeneity in a sample of 9 patients (three patients per each of three CKD histopathological groups: focal segmental glomerulosclerosis, diabetic nephropathy, and immunoglobulin A nephropathy) and highlights differences in grouping in the two classification approaches.

**Fig 2.**
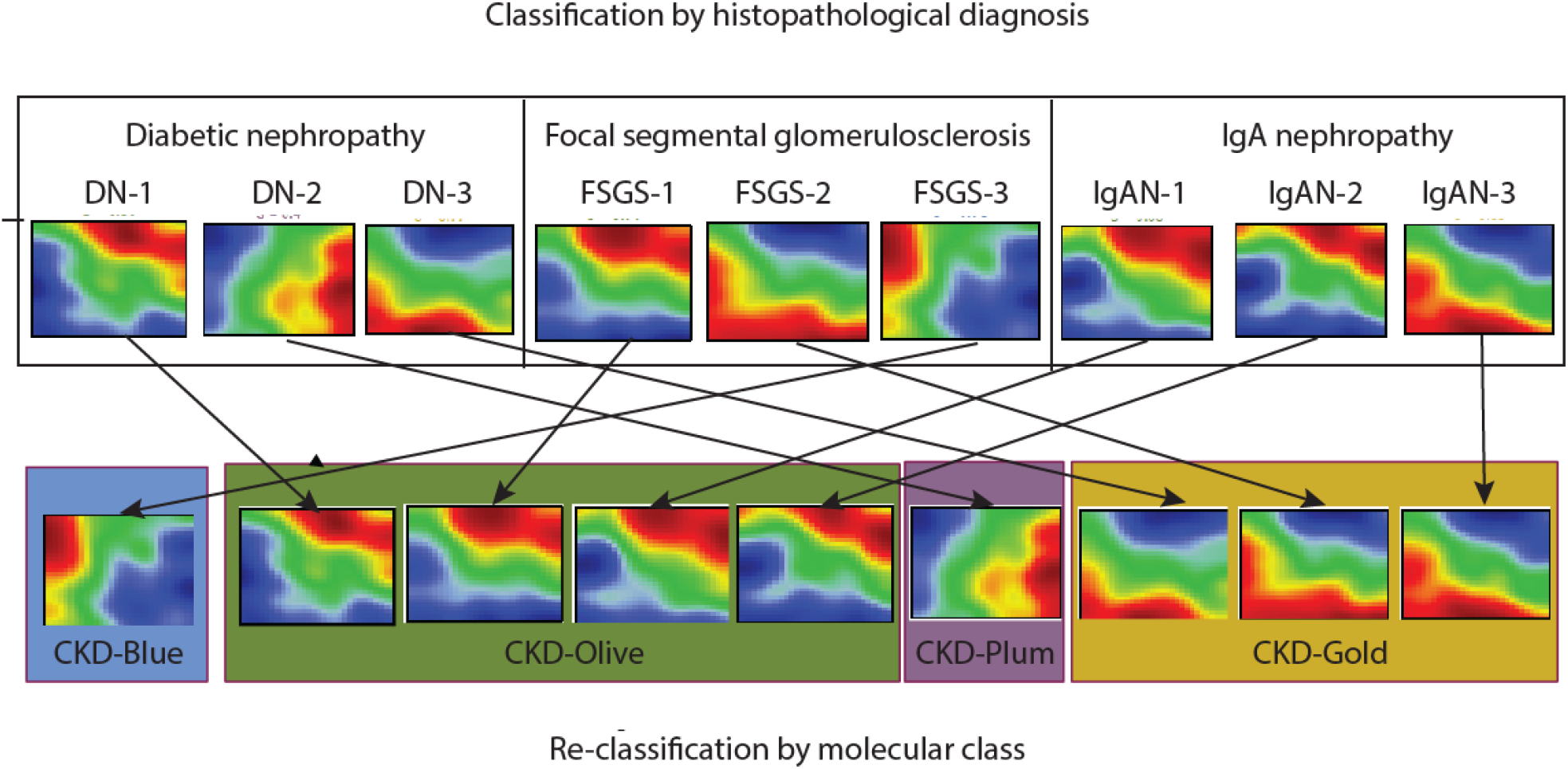
Heterogeneity of individual CKD transcriptomics profiles and re-classification based on molecular similarity independently from the conventional categories. Sample of 9 patients’ kidney gene expression profiles demonstrate marked heterogeneity, even within same diagnostic groups. Genes are represented in a fixed order on a grid to enable visual comparisons between samples. Color scale reflects gene expression levels relative to the population average (red – high, blue – low, green – average).

Therefore, CKD molecular categories represent a novel characterization, not captured by existing classifications, and provide orthogonal biomechanistic information, independent of clinical manifestations or disease severity.

### CKD molecular categories are biologically distinct

In order to obtain functional insights into biological processes underlying the molecular categorization, we compared differences in kidney gene expression between each molecular category to those of healthy controls (living kidney donors). The volcano plots (*Fig 3 A*), highlight substantial transcriptomics changes in each CKD molecular categories. Interestingly, the numbers of significantly differentially expressed genes (DEGs) in each comparison were not driven by the differences in sample size. Blue, the largest CKD molecular category, yielded the fewest number of differentially expressed genes (216), predominantly down-regulated. In contrast, Plum and Gold categories had many more DEGs (3,125 and 3,472, respectively), evenly proportioned between up- and down-regulation (*Table 2*). Finally, when the CKD samples were pooled together and contrasted to the healthy controls in a conventional case-control analysis, the differential expression was less pronounced (1,341 DEGs), likely dampened by the heterogeneity of cases. The list of all significant DEGs (at FDR q<0.0001) per CKD molecular category is provided in *Table S1*.

**Table 2.**
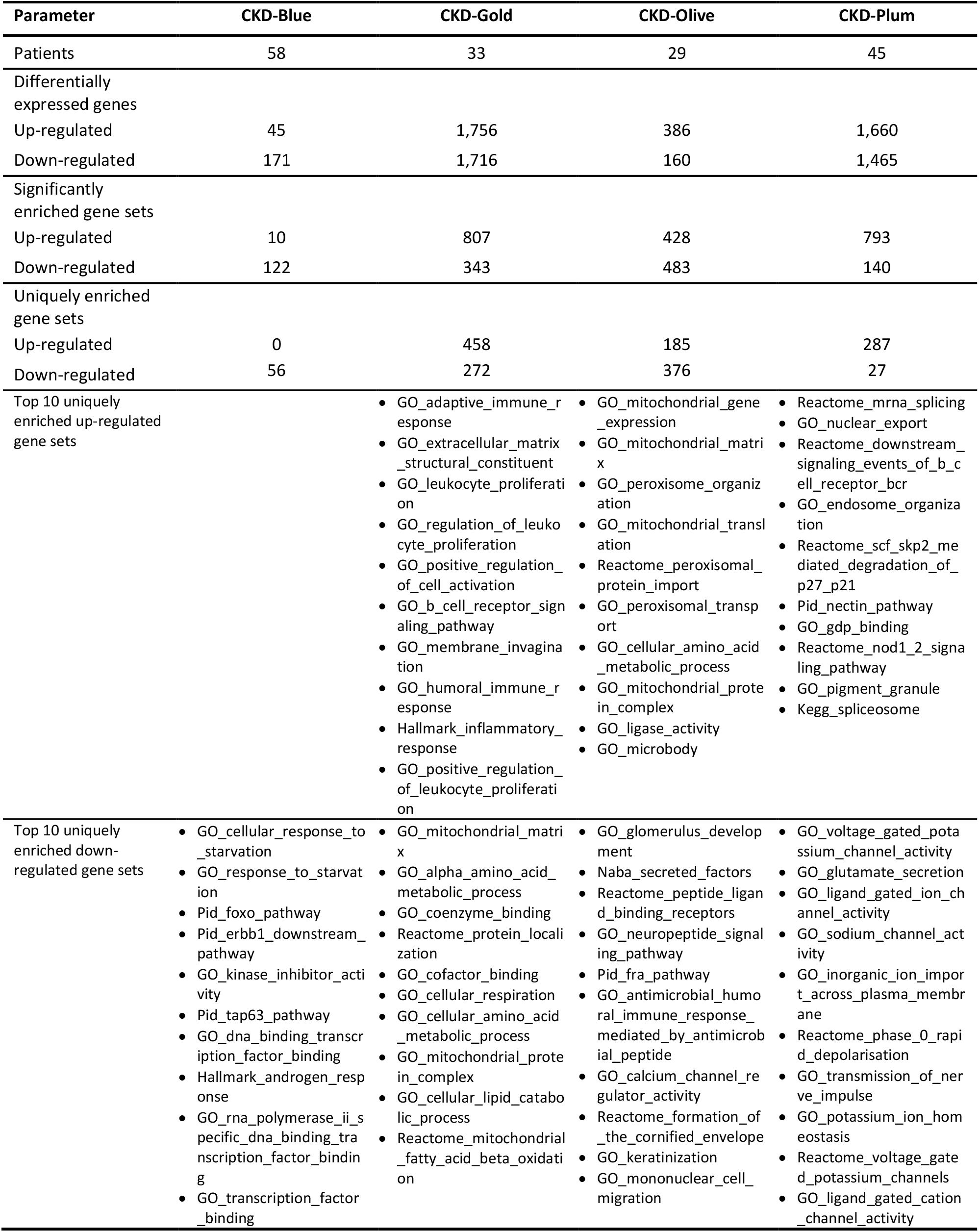
Summary of differential expression and enrichment gene sets by molecular CKD category

**Fig. 3.**
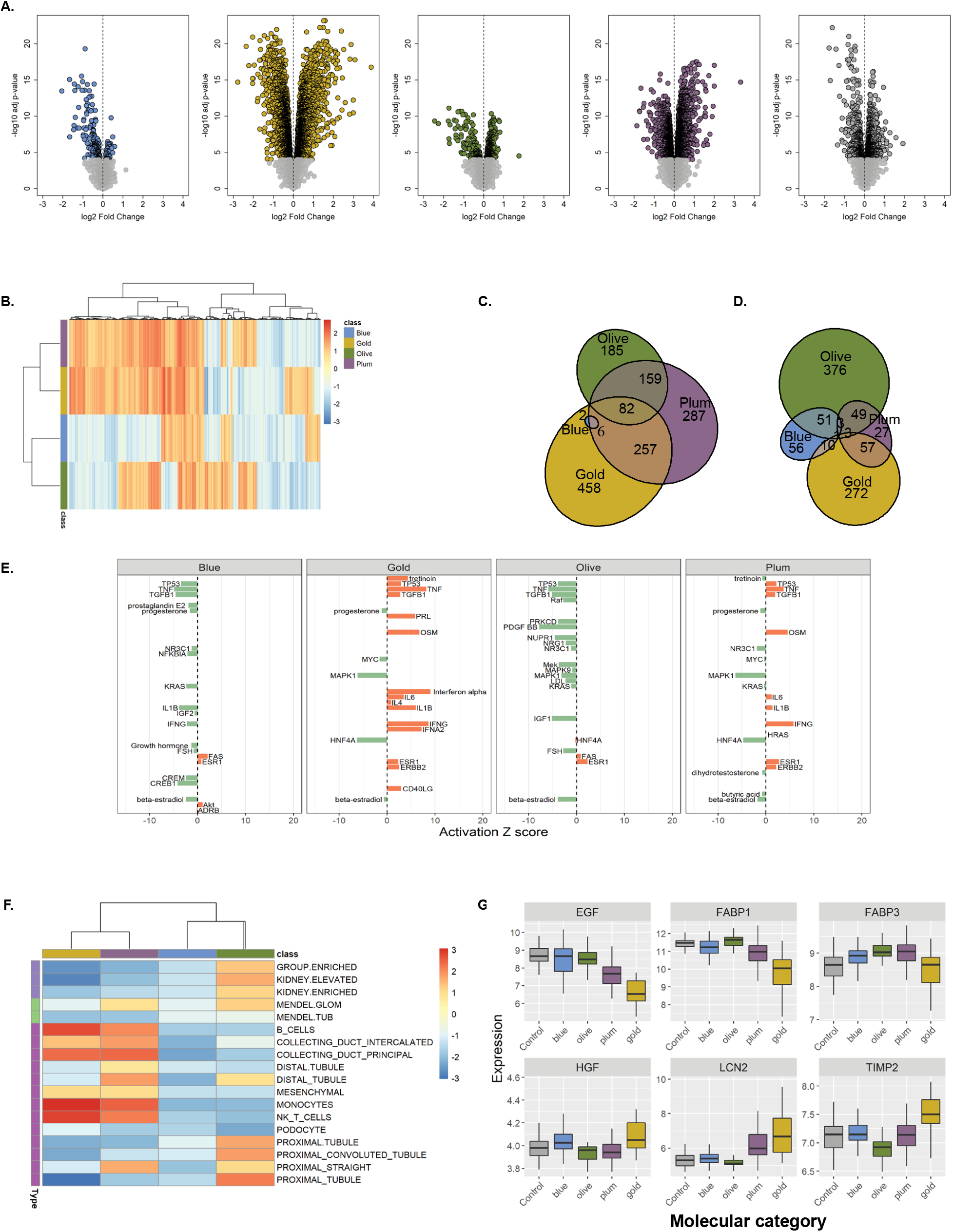
CKD molecular categories differ from healthy kidney transcriptomics profile and are biologically distinct. **(A)** Volcano plots visualize differential gene expression analysis results (log fold changes by statistical significance) per each molecular category contrasted with healthy control (living kidney donors, N=46). Each point represents a gene. Significantly modulated genes (q<0.0001) are highlighted in category-specific colors. Vertical dotted line indicates zero-fold change. The table shows numbers of differentially expressed genes, stratified by the direction of regulation, per each comparison. **(B)** Heatmap of gene set enrichment analysis results. The 5,205 gene sets (columns) were tested in each CKD molecular category (rows). The colors reflect normalized enrichment score (NES) values (maroon – positive, blue – negative) per gene set. The color bar indicates respective molecular category (Olive, Gold, Plum, Blue). Hierarchical clustering groups similar gene sets. **(C-D)** Venn diagrams visualize overlaps in significantly enriched (FDR q<0.05) up- and down-regulated gene sets, respectively, demonstrating presence of shared and unique pathways between CKD molecular categories. **(E)** Transcription regulator analysis. Activation Z-score for top 20 endogenous upstream regulators per CKD molecular category (orange – activated, green – inhibited). **(F)** Heatmap of hypothesis-driven enrichment analysis visualizing enrichments scores for CKD-relevant gene sets (kidney cell-type specific signatures, Mendelian genes known to cause renal phenotypes, genes with kidney-enriched expression). **(G)** Gene expression levels of known kidney injury markers in healthy controls and CKD molecular categories. Boxplots visualize median values per group, whiskers indicate IQR.

Specific biological themes or pathways in each CKD molecular category were then identified using gene set enrichment analysis (GSEA). In the first instance, a hypothesis-free GSEA using canonical, CKD-agnostic gene sets (Hallmark, Curated Canonical Pathways, Gene Ontology) was performed. The resulting heatmap of gene set enrichment score values (*Fig 3 B*) shows distinct patterns across the molecular categories and presence of both unique and shared pathways, as corroborated by Venn diagrams of the significantly enriched (FDR q<0.05) up- and down-regulated gene sets (*Fig 3 C and D*, respectively). The top 10 uniquely enriched gene sets (*Table 2*, while a complete list of significantly enriched gene sets per CKD molecular category is provided in *Table S2*), indicate pathways related to transcription, signaling, response to stimuli, apoptosis were selectively down-regulated in the Blue category patients. Gold category revealed a strong pro-inflammatory and pro-fibrotic drive, as the top activated processes including immune cell proliferation and migration, cytokine production, and extracellular matrix organization, while several metabolic pathways were down-regulated. In the Olive category, mitochondrial and peroxisomal processes were up-regulated while immune and membrane transport pathways were inhibited. The Plum category showed enrichment for signaling, protein synthesis, and vesicle transport processes; at the same time, ion transport pathways were down-regulated.

In line with the vast transcriptomics differences, upstream transcriptional regulator analysis revealed a diverse array of activated regulatory molecules in each of the CKD molecular category (top 20 upstream regulators are shown in *Fig 3 E*, while a complete list per CKD molecular category is provided in *Table S3*).

As the next step, hypothesis-driven gene sets of CKD relevance, including kidney-specific genes, Mendelian kidney disease genes, kidney cell type-specific markers, and kidney injury markers, were tested for enrichment across CKD molecular categories (*Fig 3 F*). All three groups of kidney-specific transcriptome were positively enriched in the Olive category.Mendelian genes known to cause glomerulopathies but not tubulopathies were enriched in the Olive and Plum categories. The Gold and Plum molecular categories showed enrichment in markers of immune cells and collecting duct, while the Olive category was specifically enriched for the proximal and distal tubular signatures. Furthermore, a set of genes encoding established urinary biomarkers of kidney damage showed diverse trends across the molecular categories (*Fig 3 G*).

Taken together, these results highlight that patients from different CKD molecular categories are distinct in terms of the biological pathways affected in the kidney, indicating that the molecular categorization has a biomechanistic basis.

### CKD molecular categories validation in independent cohorts

We validated our principal findings and determined whether these new molecular categories could be identified in other CKD populations using data from two independent cohorts.

CKD patients from the C-PROBE and NEPTUNE cohorts were mapped onto the discovery SOM based on their kidney biopsy transcriptomics profiles (*Fig 4 A-B*). As a negative control, permuted data produced nonsensical mapping. All four molecular categories were present among the C-PROBE and NEPTUNE patients. The proportions of patients per molecular categories differed between the studied CKD cohorts (chi squared p=0.0019; *Fig 4 C*).

**Fig. 4.**
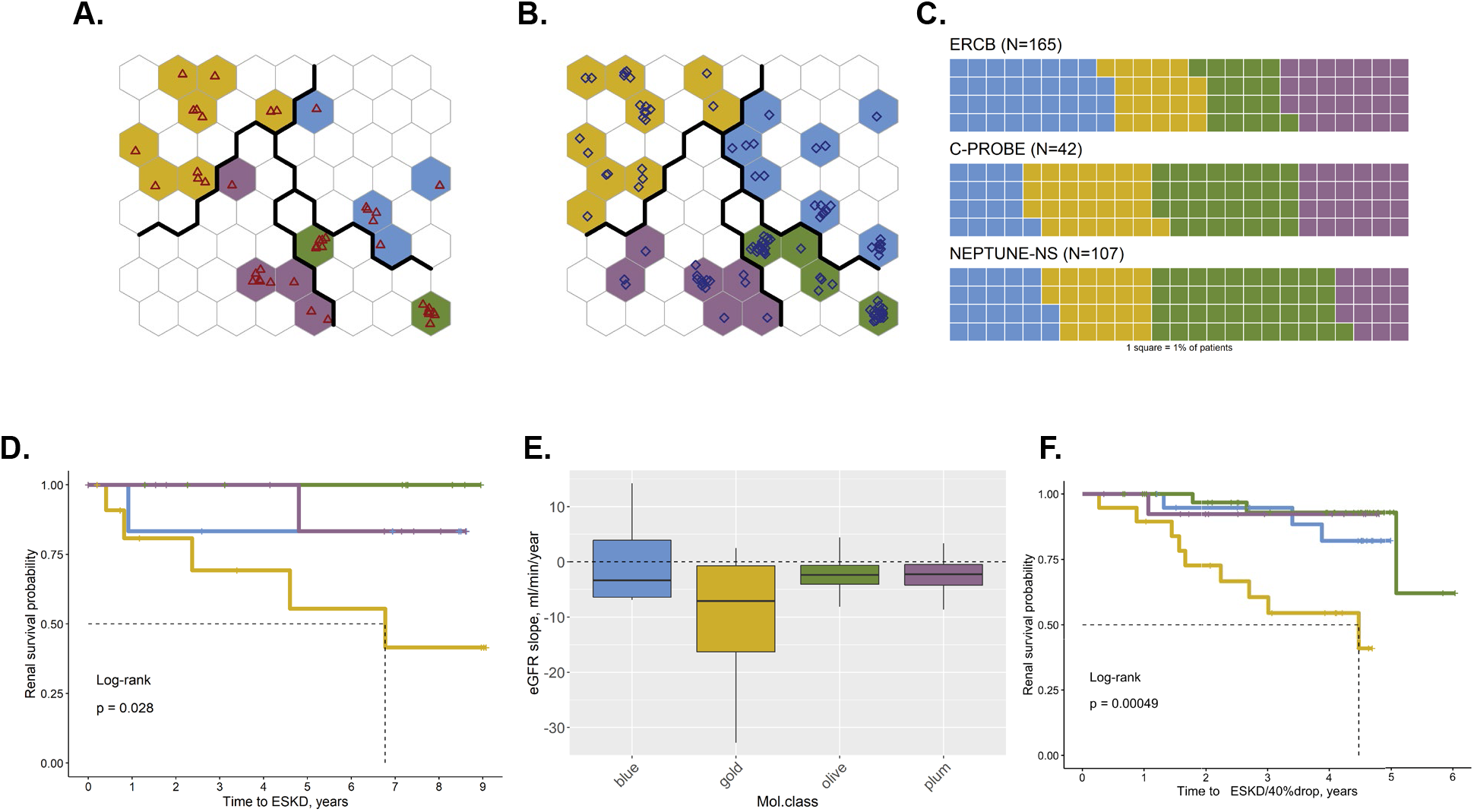
CKD molecular categorization in independent patient cohorts and disease progression comparison across CKD molecular categories. **(A)** C-PROBE cohort patients (N=42), mapped onto the trained discovery SOM; individual patients are shown as red triangles. **(B)** NEPTUNE-NS cohort patients (N=107) mapped onto the trained discovery SOM; individual patients are shown as blue diamonds. **(C)** Waffle charts visualize relative proportions of patients assigned to the different molecular categories in each CKD cohort (1 square=1%). **(D)** Kaplan-Meier curves of ESKD incidence by CKD molecular category in C-PROBE patients. Statistical significance of differences was tested using log-rank test. **(E)** Slope of eGFR, ml/min/year, by CKD molecular category in C-PROBE patients. Boxplots visualize median values per molecular class, whiskers indicate IQR. Dotted horizontal line denotes zero (no change in eGFR over time). **(F)** Kaplan-Meier curves of ESKD incidence by CKD molecular category in NEPTUNE patients. Statistical significance of differences was tested using log-rank test.

To determine whether the underlying differences in biological mechanisms between the CKD molecular categories would manifest in disparate disease progression rates, we analyzed kidney outcomes in patients with available long-term clinical follow-up data (42 C-PROBE and 90 NEPTUNE patients, respectively).

In the C-PROBE cohort, during the follow-up period (median duration 4.7 years, maximal 9.1 years), 7 out of 42 patients progressed to end-stage kidney disease (ESKD), defined as initiation of renal replacement therapy (dialysis or kidney transplantation). Even in this limited analysis, the rate of reaching the end-point varied significantly depending on the molecular category (log-rank p=0.028). Kaplan-Meier survival curves stratified by molecular category (*Fig 4D*) demonstrate the significantly higher probability of ESKD incidence in the Gold category, with 5 out of 7 events occurring in this subgroup of patients. The rate of disease progression was thus faster for patients in the Gold category with a median survival probability of 6.8 years, while no other molecular category reached median survival during the follow-up period. Cox regression hazard ratios (HR) for ESKD in Gold compared to other molecular categories was 7.9 [95%CI 1.5-41.1] (p=0.014) in the crude model and 5.6 [95%CI 1.0-31.3] in the model adjusted for baseline eGFR (p=0.048). For 37 C-PROBE patients, repeated kidney function values were available over time from which eGFR slopes were calculated. Consistent with the hard end-point (ESKD) prediction, the eGFR slopes also showed a trend for greater annual loss of kidney function in the Gold category (*Fig 4E*).

In NEPTUNE, where the outcome was defined as a composite of ESKD or loss of >40% eGFR, there were 16 events among 90 patients during the follow-up period (median duration 4.1 years, maximal 6.0 years). Likewise, the rate of disease progression was significantly different between the molecular categories (log-rank p=0.00049, *Fig 4F*), with the Gold category patients showing higher event incidence (9 out of 16) and median renal survival probability of 4.5 years. Cox regression HR for Gold compared to other molecular categories was 6.7 [95%CI 2.4-19.0] (p=0.00032) in the crude model and 4.3 [95%CI 1.3-14.2] (p=0.017) in the model adjusted for baseline eGFR (4 observations were excluded from the adjusted analysis due to missing values).

Interestingly, in contrast to the molecular categories, if stratified by CKD stage at baseline in the C-PROBE cohort (*Fig S3A*) or histopathological diagnosis in NEPTUNE (*Fig S3B*), the event rates were not significantly different.

The presence of the molecular categories in two independent cohorts thus strengthens the generalizability of our findings. The differences in disease progression rates between CKD molecular classes were consistent across both cohorts.

## Discussion

Our findings represent the first kidney transcriptomics-driven approach for molecular categorization of CKD, a significant step towards precision medicine for this complex and heterogeneous condition. This has the potential to transform the CKD field with respect to biological understanding, clinical management, and drug development, as well as pave the way towards molecular reclassification of other complex diseases.

This study was motivated by the growing realization in the field that mechanistic categorization of the various CKD etiologies is lacking (*8, 10, 19, 20*). The call to redefine CKD to boost therapeutic development is becoming increasingly urgent as its global prevalence is reaching epidemic proportions (*21*). Integration of molecular features into clinico-morphological diagnosis promises a more mechanistic framework for classification of CKD patients, as a foundation for development of personalized treatment options based on individual disease biology (*22-24*).

Additive value of molecular characterization on top of clinical parameters has been shown, previously, in studies evaluating the use of circulating or urinary biomarkers to improve disease stratification and prognosis prediction (*25, 26*). However, a fundamental limitation of such systemic phenotyping is that due to the indirect assessment, the underlying molecular changes in the kidney remain unknown. We pursued a different approach, leveraging available molecular profiles from kidney biopsies, directly reflecting intra-renal biology, as the basis of our categorization strategy.

Transcriptomics profiling provides a fingerprint of genome-wide gene expression patterns reflecting tissue structural and functional states (*27-30*). These include cell type composition, activity of biological pathways causing, perpetuating or counteracting the disease, interplay of co-morbidities, as well as the effects of treatments (*29-32*). Our findings highlight the value of transcriptomics in grouping patients based on similar profiles. The fact that permuted data, where the expression values of each gene were randomized across the samples, failed to map onto SOM signifies the importance of the individual specificity of each patient’s transcriptomics profile as well as gene covariance modular structure.

We identified four novel patient subgroups within a CKD population in an unbiased data-driven manner, based on kidney transcriptomics data and without imposing any patient clinical information or preconceived, domain biological knowledge. The emergent molecular categories did not overlap with the conventional clinical classification and were detected orthogonally at all the different CKD stages and histopathological diagnoses, highlighting molecular heterogeneity among CKD patients and underscoring the biological complexity of this multifaceted condition. We propose that molecular information can, therefore, add a new dimension to the existing framework of CKD classification.

Our results highlighted that CKD molecular categories were distinct from the healthy controls as well as one another in terms of biological pathways affected in the kidney. This shows that molecular categorization represents a mechanistic classification of disease that is based on underlying biological processes. Differential expression analysis of the molecular categories vs healthy control produced a multitude of modulated genes. Living kidney donor biopsies were chosen to best represent the “healthy” kidney state, as these individuals were specifically screened for absence of disease and selected for superior kidney function to qualify for donation. Using such a control group, as opposed to commonly used tumor nephrectomy or autopsy tissues, provides a robust baseline and enhances our ability to detect biologically meaningful differences by maximizing the contrast with the disease state. The case-control contrast was further boosted by CKD molecular categories forming intrinsically more uniform subgroups as compared to the heterogeneous all-comer CKD population. This highlights that the molecular categorization is also able to capture elements of the biological basis of the disease.

Through both hypothesis-free exploration and hypothesis-driven analyses, we found that distinct biological pathways, transcriptional regulators, injury markers, and cell type context were perturbed in each of the CKD molecular categories – an important step in unraveling the biological basis of disease heterogeneity. Many of the enriched pathways, such as inflammation, apoptosis, metabolism, have been previously implicated in CKD (*33-36*). However, the novelty of our results is in that not all pathways were found equally activated in all patients; instead, different pathways could be more dominant in different subgroups and thereby differentially influencing disease progression and outcomes. For example, inflammation and fibrosis, generally accepted as universal pathophysiological mechanisms in CKD, showed uneven distribution within the patient population with specific enrichment in one of the molecular categories, CKD-Gold. This challenges existing dogma with implications for disease understanding as well as anti-inflammatory drug development, further emphasizing the importance of precision medicine and mechanistic enrichment approaches to patient selection.

The identified molecular categories were replicated in independent cohorts indicating that the molecular categorization can be extended to other CKD populations. The ability to predict the molecular category of previously unseen patient data strengthens generalizability and robustness of our findings as it demonstrates robustness across CKD etiology and transcriptomics platforms between research centers in the US and Europe. The outcome association was replicated in two independent cohorts. The observation of different ESKD risk across molecular categories conserved between both cohorts is encouraging and advocates for further investigation as the molecular categorization might help identify patients at greater risk for rapid disease progression.

Cross-sectional study design does not allow to observe the anticipated changes over time in the transcriptomics profile following the disease progression processes. However, as repeated kidney biopsies are rarely performed in clinical practice, the temporal trends of molecular changes will need to be ascertained through non-invasive means or through time-course studies in animal models. Finally, to enable implementation in clinical practice, non-invasive biomarkers, e.g. urinary proteins reflecting the intra-renal molecular characteristics of each category, will be needed to allow comprehensive capture in large scale cohorts. Our findings showing differential expression patterns of genes encoding known markers of kidney injury across molecular categories offer a starting point to develop a non-invasive biomarker panel to recognize these categories.

There are several limitations to this study. We analyzed the largest available kidney transcriptomics datasets from CKD patients nevertheless were limited in sample size. Future collaborative effort will be needed to generate more data that may help further refine the classification as well as expand the repertoire of ‘omics’ beyond transcriptomics. Studied cohorts were subject to an inherent selection bias as patients undergoing clinically indicated kidney biopsy, therefore may differ from the general CKD population in composition. Our analysis focused on the more readily available tubulointerstitial tissue component; we plan to extend similar analysis to the glomerular compartment in the future. However, tubulointerstitial biology is highly relevant for CKD. Tubulointerstitial lesions are known to be a strong predictor of CKD progression. Even in the cases of primary or secondary glomerular disease, studies have suggested that it is the extent of accompanying tubulointerstitial histologic injury that correlates best with renal function decline and progression to ESKD (*38-41*). Transcriptomics profiling of renal tubulointerstitium has proved informative in deciphering molecular basis of disease (*42-44*). In the subsequent studies, it would be highly relevant to include also genetics and comorbidities information for further exploration and interpretation of the characteristics of each molecular class, which for ethical concerns were not available for the discovery cohort.

Our results have the potential to take the CKD disease classification to the next, mechanistic level. A transition to an integrated clinical, morphological and molecular diagnosis is warranted for development of individualized treatments in CKD. Elucidation of the molecular drivers of different patient subgroups can lead to new biological hypotheses, therapeutic targets, and biomarkers. Establishing a first framework of molecular categories in CKD is an important milestone in CKD research, but replication across further disease cohorts and development of non-invasive means to capture the intra-renal categories are urgently needed to accelerate progress towards precision medicine for CKD.

## Materials & Methods

### Renal transcriptomics data

We analyzed publicly available renal tubulointerstitial microarray transcriptomics data from three CKD cohorts. Consent was obtained from individual patients at enrollment, and the studies were approved by Institutional Review Boards of participating institutions.

ERCB (European Renal cDNA Bank) is a European multicenter CKD study established to collect renal biopsy tissue for gene expression analysis at the time of a clinically indicated biopsy (*45*). Affymetrix U133 array transcriptomics data from 165 patients were used for the discovery analysis (GEO accession ID: GSE104954).

C-PROBE (Clinical Phenotyping Resource and Biobank Core) is a multicenter longitudinal observational CKD cohort (*17*). Tubulointerstitial transcriptomics data generated on Affymetrix Human Genome U133 Plus 2.0 Array were available for 42 patients (GEO accession ID: GSE69438).

NEPTUNE (Nephrotic Syndrome Study Network) is a multi-center, prospective study of children and adults with nephrotic range proteinuria (>500mg/day), recruited at the time of first clinically indicated baseline renal biopsy (*46*). Affymetrix Human Gene 2.1 ST Array tubulointerstitial transcriptomics data from 107 NEPTUNE-NS adult CKD cases were included in the analysis (GEO accession ID: GSE108112).

Tubulointerstitial transcriptomics data from 46 living kidney donors (GSE104954) were used as controls for differential expression analysis.

### Self-Organizing Maps algorithm

All analyses and visualizations were run using R version 3.6.0, unless indicated otherwise. SOM functions implemented in R packages *kohonen* v3.0.8, *popsom* v4.2.1, and *oposSOM* v2.2.0 (*49*) were used. Inherent data clustering tendency in ERCB dataset was assessed with Hopkins statistics and VAT (Visual Assessment of cluster Tendency) function using *factoextra* R package (*Fig. S4*). Genes with low variance across samples (median absolute deviation <0.15, 27% of genes) were excluded from the analysis as non-informative. The retained 8,454 gene expression variables were centered and scaled to give them equal importance during the SOM training process and prevent bias from highly abundant genes. Different map grid sizes (4×4 through 12×12) and number of iterations (100 through 50,000) were tested, and mapping quality indices (quantization error, topographic accuracy, embedding accuracy) assessed (*Fig. S5*). Considering the random nature of SOM initialization, behaviors of SOMs from 10 independent runs were compared (*Fig. S5 B*). Final model parameters were set to achieve the highest convergence. The optimal map size was determined to be 8×8 (64 SOM units), consistent with the commonly used empirical rule of thumb 5x√N samples (*50*). Hexagonal topology and a bubble neighborhood function with sum-of-squares as distance measure were used. In order to isolate groups of samples with similar profiles, Ward’s hierarchical clustering was run on the codebook vectors. The optimal number of clusters was guided by the ‘elbow method’ using *NbClust* R package based on the simplest model with the lowest within-cluster sum-of-squares. Diagnostic plots are presented in *Fig. S5-S6*. Validation mapping of the independent CKD cohorts was performed using ‘map.kohonen’ function of *kohonen* R package (*14*). New data for the 8,454 genes were extracted from C-PROBE and NEPTUNE transcriptomics, expression values for missing genes (378 and 427 for C-PROBE and NEPTUNE, respectively) were set to “NA”. The data were scaled using the ERCB scaling parameters and then presented to the trained SOM, and distances from the new data points to the closest units were calculated.

### Downstream analyses

Individual and group-level transcriptomics profiles were visualized using *oposSOM* R package with default parameters (*49*). Differential gene expression analyses comparing patients’ and control samples were performed with *limma* R package(*52*). The output was ranked by *sign(logFC)*-log10(adj*.*p-value*), and functional enrichment analyses were run with Gene Set Enrichment Analysis application using 5,205 gene sets belonging to Hallmark, Curated Canonical Pathways, and Gene Ontology from MSigDB database of molecular signatures (*53, 54*). Upstream regulator analysis was performed using IPA (QIAGEN Inc., https://www.qiagenbioinformatics.com/products/ingenuitypathway-analysis, (*55*)), focusing on endogenous regulators only. A list of 413 genes with preferential renal expression (53 kidney-enriched, 229 kidney-elevated, and 131 group-enriched) was extracted from Human Protein Atlas v19.3 (www.proteinatlas.org (*56*)). Mendelian genes responsible for glomerulopathies (172) and tubulopathies (55) were extracted from Parsa et al. (*57*). A list of renal cell-type specific markers was curated from literature (*58*) as well as derived from single-cell RNA-seq data(*59*). Outcome association testing was performed using *survival* R package.

## Data Availability

ERCB (European Renal cDNA Bank) data were accessed from GEO accession ID: GSE104954; C-PROBE (Clinical Phenotyping Resource and Biobank Core) tubulointerstitial transcriptomics data generated on Affymetrix Human Genome U133 Plus 2.0 Array were available for 42 patients (GEO accession ID: GSE69438). Affymetrix Human Gene 2.1 ST Array tubulointerstitial transcriptomics data from 107 NEPTUNE (Nephrotic Syndrome Study Network) adult CKD were accessed from GEO accession ID: GSE108112. Tubulointerstitial transcriptomics data from 46 living kidney donors were accessed from GEO accession ID:GSE104954.

## Acknowledgements

We thank Dr. Lalita Subramanian for the excellent writing assistance and diligent proofreading of this manuscript. See Supplemental Acknowledgments for NEPTUNE consortium details. ERCB members at the time of the study: Clemens David Cohen, Holger Schmid, Michael Fischereder, Lutz Weber, Matthias Kretzler, Detlef Schlöndorff, Munich/Zurich/AnnArbor/New York; Jean Daniel. Sraer, Pierre Ronco, Paris; Maria Pia Rastaldi, Giuseppe D’Amico, Milano; Peter Doran, Hugh Brady, Dublin; Detlev Mönks, Christoph Wanner, Würzburg; Andrew Rees, Aberdeen and Vienna; Frank Strutz, Gerhard Anton Müller, Göttingen; Peter Mertens, Jürgen Floege, Aachen; Norbert Braun, Teut Risler, Tübingen; Loreto Gesualdo, Francesco Paolo Schena, Bari; Gunter Wolf, Jena; Rainer Oberbauer, Dontscho Kerjaschki, Vienna; Bernhard Banas, Bernhard Krämer, Regensburg; Moin Saleem, Bristol; Rudolf Wüthrich, Zurich; Walter Samtleben, Munich; Harm Peters, Hans-Hellmut Neumayer, Berlin; Mohamed Daha, Leiden; Katrin Ivens, Bernd Grabensee, Düsseldorf; Francisco Mampaso(†), Madrid; Jun Oh, Franz Schaefer, Martin Zeier, Hermann-Joseph Gröne, Heidelberg; Peter Gross, Dresden; Giancarlo Tonolo; Sassari; Vladimir Tesar, Prague; Harald Rupprecht, Bayreuth; Hermann Pavenstädt, Münster; Hans-Peter Marti, Bern; Peter Mertens, Magdeburg, Jens Gerth, Zwickau.

## Author contributions

All co-authors have contributed to the manuscript. Specifically, A.R. conceived the project. A.R., M.C.M., M.K., V.N., R.B. designed and directed the project. A.R., V.N., S.E., M.T., T.S. analyzed the data and produced the figures. All authors interpreted the results, and provided critical feedback that helped shape the research, analysis and manuscript. A.R. drafted the manuscript. All authors approved the final version and accept accountability for the overall work by ensuring that questions pertaining to the accuracy or integrity of any portion of the work were appropriately investigated and resolved.

### Data and materials availability

All source data used in this study are publicly available and referenced in the Materials and Methods section in the main text. All data resulting from this study are available in the main text or the supplementary materials.

## Supplementary Materials

**Supplemental Acknowledgement**. NEPTUNE Consortia members

**Fig S1**. Gene expression levels across CKD molecular categories

**Fig S2**. eGFR by molecular category stratified by CKD stage

**Fig S3**. Kaplan-Meier survival curves for prediction of ESKD outcome stratified by CKD stage (A) and histopathological diagnosis (B)

**Fig S4**. Data clustering tendency assessment: pairwise Spearman correlation-based distances between samples (A); VAT, Visual Assessment of cluster Tendency (B)

**Fig S5**. Building Self-Organizing Map of CKD population: quantization error vs map size (A); convergence -embedding accuracy & topographic accuracy (B); learning progress across 10 independent models (C)

**Fig S6**. Final model SOM diagnostic plots: counts (A); mapping quality (B); neighbor distances (C); number of clusters (D)

**Table S1**. Differentially expressed genes per CKD molecular category vs healthy control

**Table S2**. Enriched pathways per CKD molecular category

**Table S3**. Upstream transcriptional regulators per CKD molecular category

