## Supplementary materials for "Molecular Stratification of Chronic Kidney Disease"

#### Supplemental Acknowledgement

##### Members of the Nephrotic Syndrome Study Network (NEPTUNE)

NEPTUNE Enrolling Centers

*Cleveland Clinic, Cleveland, OH*: K Dell[*], J Sedor[**], M Schachere[#], J Negrey[#]
*Children's Hospital, Los Angeles, CA*: K Lemley[*], E Lim[#]
*Children's Mercy Hospital, Kansas City, MO*: T Srivastava[*], A Garrett[#]
*Cohen Children's Hospital, New Hyde Park, NY:* C Sethna[*], K Laurent [#]
*Columbia University, New York, NY:* P Canetta[*], A Pradhan[#]
*Emory University, Atlanta, GA:* L Greenbaum[*], C Wang**, C Kang[#]
*Harbor-University of California Los Angeles Medical Center:* S Adler[*], J LaPage[#]
*John H. Stroger Jr. Hospital of Cook County, Chicago, IL:* A Athavale[*], M Itteera
*Johns Hopkins Medicine, Baltimore, MD:* M Atkinson[*], S Boynton[#]
*Mayo Clinic, Rochester, MN:* F Fervenza[*], M Hogan**, J Lieske[*], V Chernitskiy[#]
*Montefiore Medical Center, Bronx, NY:* F Kaskel[*], M Ross[*], P Flynn[#]
*NIDDK Intramural, Bethesda MD:* J Kopp[*], J Blake[#]
*New York University Medical Center, New York, NY:* H Trachtman[*], O Zhdanova**, F Modersitzki[#], S Vento[#]
*Stanford University, Stanford, CA:* R Lafayette[*], K Mehta[#]
*Temple University, Philadelphia, PA:* C Gadegbeku[*], S Quinn-Boyle[#]
*University Health Network Toronto:* M Hladunewich**, H Reich**, P Ling[#], M Romano[#]
*University of Miami, Miami, FL:* A Fornoni[*], C Bidot[#]
*University of Michigan, Ann Arbor, MI:* M Kretzler[*], D Gipson*, A Williams[#], J LaVigne[#]
*University of North Carolina, Chapel Hill, NC:* V Derebail[*], K Gibson[*], E Cole[#], J Ormond-Foster[#]
*University of Pennsylvania, Philadelphia, PA:* L Holzman[*], K Meyers**, K Kallem[#], A Swenson[#]
*University of Texas Southwestern, Dallas, TX:* K Sambandam[*], Z Wang[#], M Rogers[#]
*University of Washington, Seattle, WA:* A Jefferson[*], S Hingorani**, K Tuttle**[§], M Bray [#], M Kelton[#], A Cooper[#§]
*Wake Forest University Baptist Health, Winston-Salem, NC:* JJ Lin*, Stefanie Baker[#]

*Data Analysis and Coordinating Center*: M Kretzler, L Barisoni, J Bixler, H Desmond, S Eddy, D Fermin, C Gadegbeku, B Gillespie, D Gipson, L Holzman, V Kurtz, M Larkina, J Lavigne, S Li, S Li, CC Lienczewski, J Liu, T Mainieri, L Mariani, M Sampson, J Sedor, A Smith, A Williams, J Zee.

*Digital Pathology Committee*: Carmen Avila-Casado (University Health Network, Toronto), Serena Bagnasco (Johns Hopkins University), Joseph Gaut (Washington University in St Louis), Stephen Hewitt (National Cancer Institute), Jeff Hodgin (University of Michigan), Kevin Lemley (Children's Hospital of Los Angeles), Laura Mariani (University of Michigan), Matthew Palmer (University of Pennsylvania), Avi Rosenberg (Johns Hopkins University), Virginie Royal (University of Montreal), David Thomas (University of Miami), Jarcy Zee (University of Pennsylvania) Co-Chairs: Laura Barisoni (Duke University) and Cynthia Nast (Cedar Sinai).

*Principal Investigator; **Co-investigator; [#]Study Coordinator
[§]Providence Medical Research Center, Spokane, WA

#### Fig S1. Gene expression levels across CKD molecular categories and healthy controls

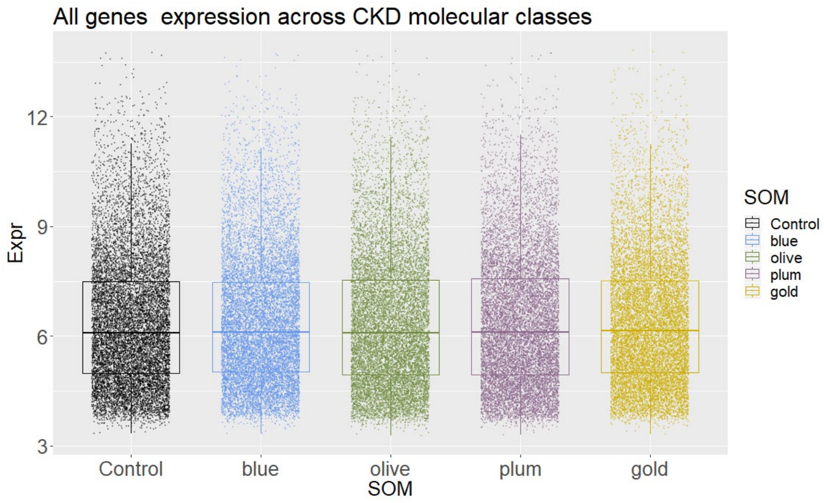

**Fig S2. eGFR by molecular category stratified by CKD stage**

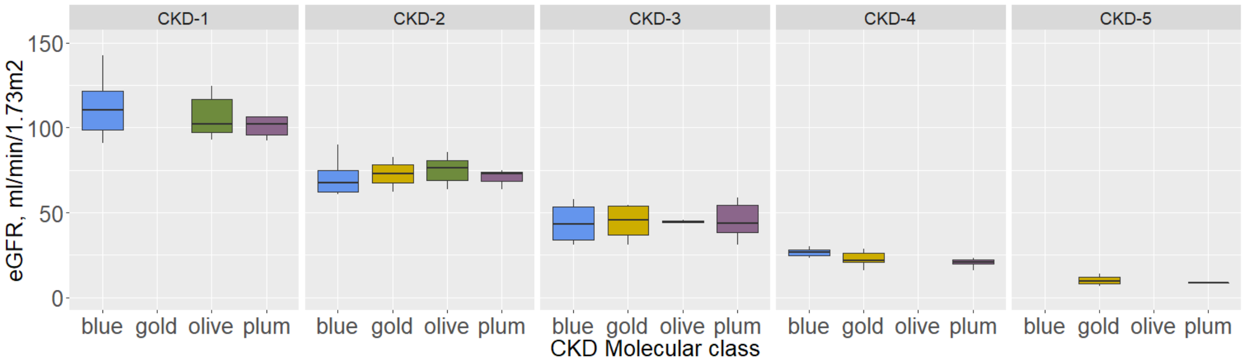

**Fig S3. Kaplan-Meier survival curves for prediction of ESKD outcome stratified by (A) CKD stage (C-PROBE) and (B) histopathological diagnosis (NEPTUNE)**

A.

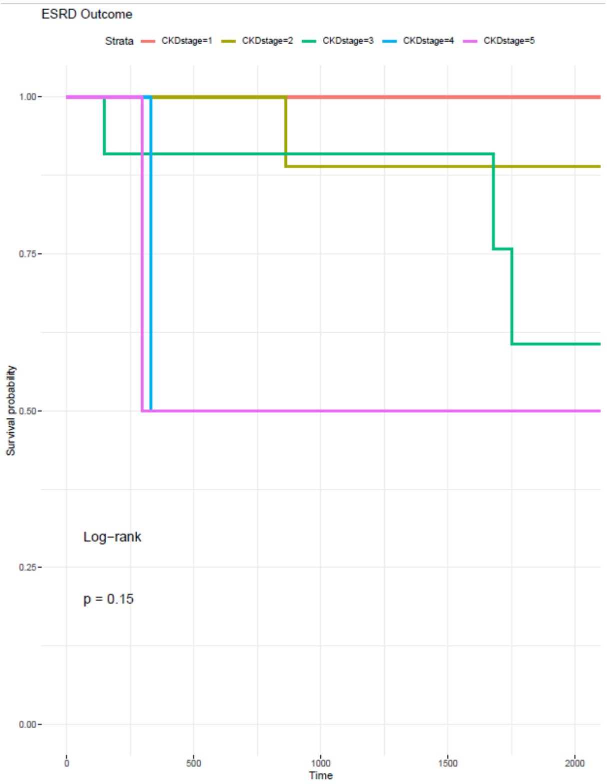


B.

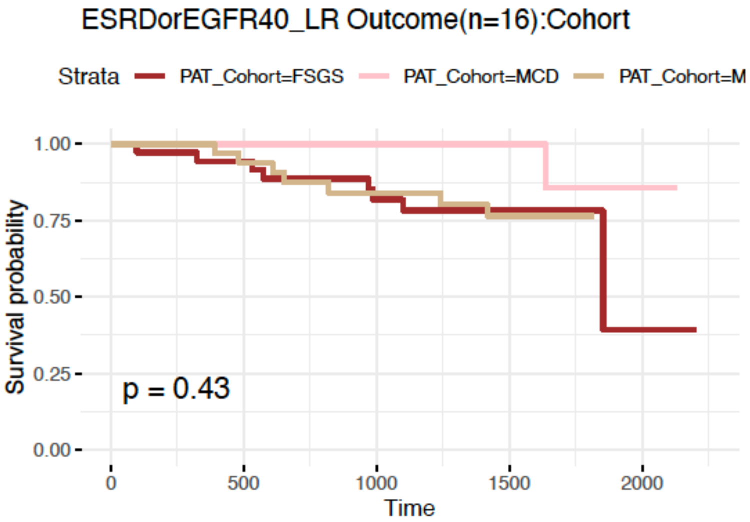

**Fig S4. Data clustering tendency assessment**

**(A)** Pairwise Spearman correlation-based distances between samples to assess similarity between observations. **(B)** VAT (Visual Assessment of cluster Tendency). The value of Hopkins statistic is below 0.5, implying that the dataset is significantly clusterable, that is, contains inherent grouping structure.

**A.**

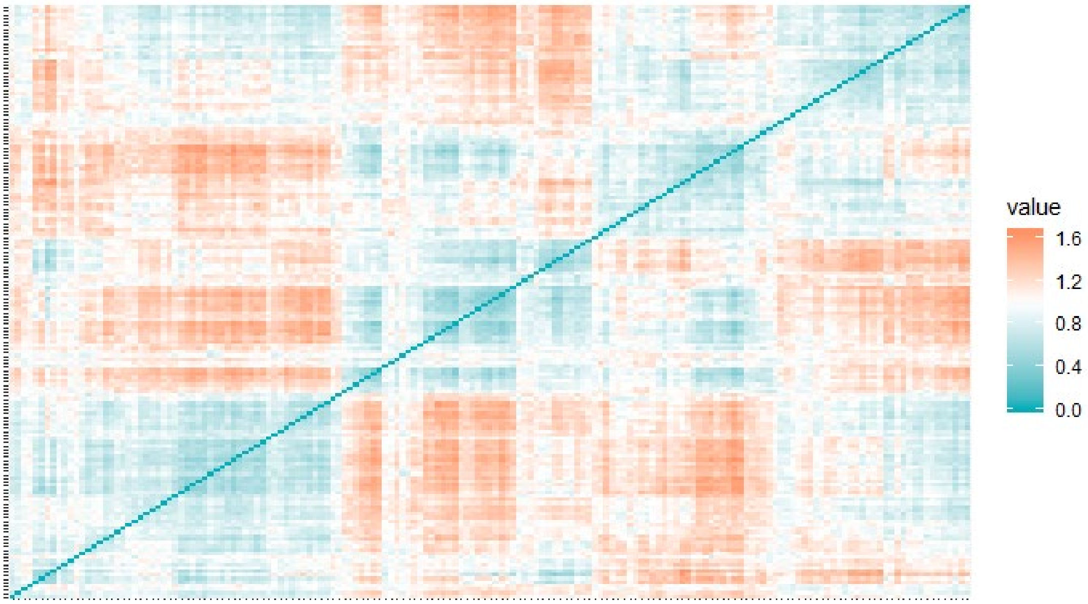


**B.**

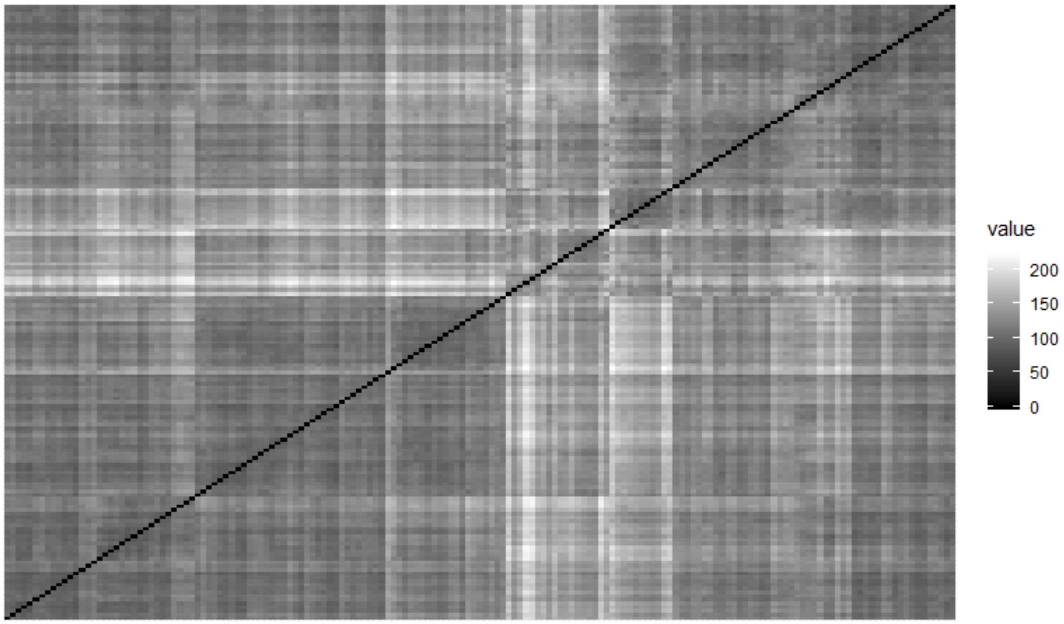


```
$hopkins_stat
[1] 0.3526008
```

**Fig S5. Building Self-Organizing Map of CKD population**

(A) SOM mapping quality, estimated by the average distance of each data point to its corresponding codebook vector (BMU), plotted as a function of map grid size. Is a measure of map resolution. 8x8 grid is the simplest model achieving the best mapping.
(B) Convergence as a linear combination of the two SOM quality indices, embedding accuracy and estimated topographic accuracy, was evaluated across various N iterations and reached 0.8 at 50,000. The embedding accuracy (black line) is the variance of the training data captured by the map. Evaluated by Kolmogorov-Smirnov test. Grows with the number of iterations, acceptable at 50,000. Topographic accuracy (blue line) is the proportion of data points for which first and second BMUs are adjacent units. Measures topology preservation. Is perfect already at 1,000 iterations.
(C) Learning progress is similar across 10 independent models. Considering the random nature of SOM initialization, behaviors of SOMs from 10 independent runs were compared. The results and conclusions were not materially different.

**A. Quantization error vs map size**

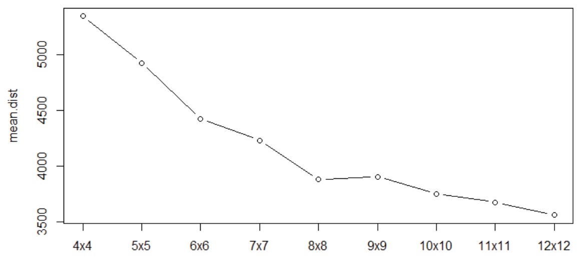


**B. Convergence vs N iterations**

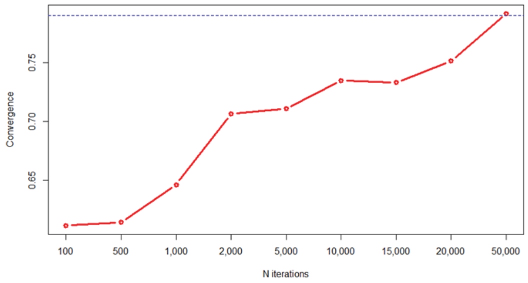


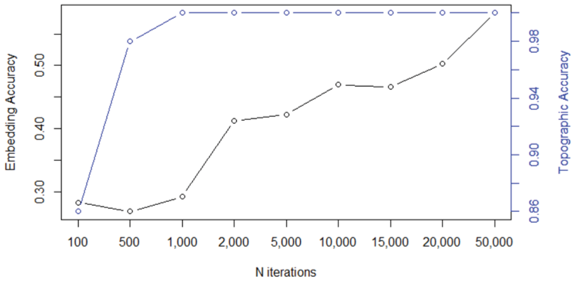


**C. Learning progress across 10 independent models**

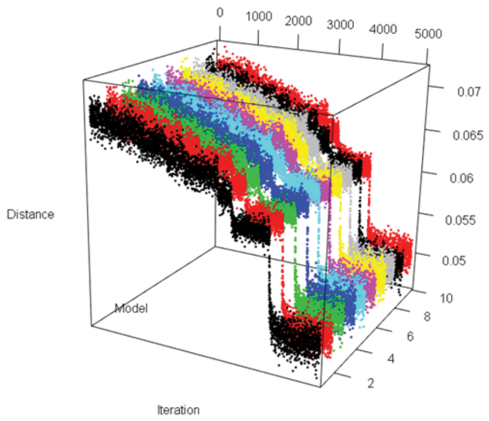

**Fig S6. Final model SOM properties**

**(A)** Diagnostic counts plots; **(B)** mapping quality; **(C)** neighbor distances; **(D)** number of clusters. **(E)** The simplest model with the lowest within-cluster sum-of-squares is selected.

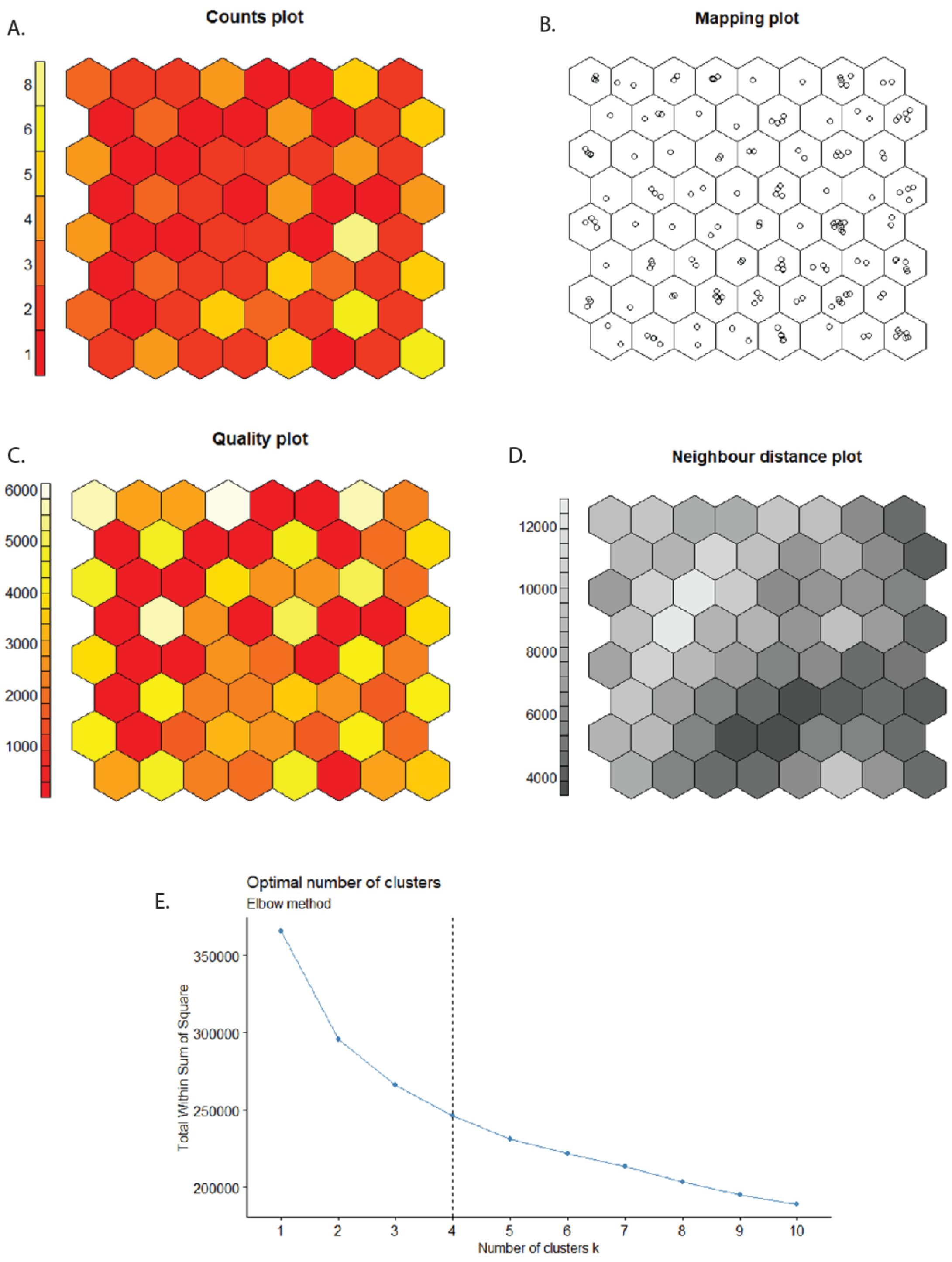

Table S1 Blue vs. Control

| Gene | logFC | UnlogFC | AveExpr | t | P.Value | adj.P.Val | B |
|---|---|---|---|---|---|---|---|
| ZFAND5 | -0.88879 | 0.540069 | 9.558462 | -13.0917 | 6.13E-24 | 5.18E-20 | 43.5909 |
| RHOB | -1.06268 | 0.478741 | 9.158654 | -11.2635 | 7.22E-20 | 3.05E-16 | 34.53365 |
| PDK4 | -1.66283 | 0.315818 | 7.451346 | -10.986 | 3.05E-19 | 8.60E-16 | 33.13811 |
| CEBPB | -1.01112 | 0.496159 | 8.251538 | -10.7939 | 8.29E-19 | 1.75E-15 | 32.17016 |
| PTP4A1 | -0.4373 | 0.738515 | 7.598077 | -10.5894 | 2.41E-18 | 3.68E-15 | 31.13808 |
| GADD45B | -1.42193 | 0.373214 | 7.985481 | -10.5737 | 2.61E-18 | 3.68E-15 | 31.05908 |
| NFKBIA | -0.77826 | 0.583069 | 9.409231 | -10.5106 | 3.63E-18 | 4.38E-15 | 30.74022 |
| KLF6 | -1.2353 | 0.424754 | 8.020865 | -10.3494 | 8.40E-18 | 8.88E-15 | 29.92552 |
| SGK1 | -0.9093 | 0.532442 | 10.47702 | -10.1156 | 2.84E-17 | 2.65E-14 | 28.74337 |
| ZFP36 | -1.03391 | 0.488386 | 8.256442 | -10.0968 | 3.14E-17 | 2.65E-14 | 28.64841 |
| DUSP1 | -1.39842 | 0.379345 | 7.426635 | -10.0527 | 3.95E-17 | 3.03E-14 | 28.42551 |
| FAM134B | -0.68334 | 0.622724 | 9.235865 | -10.0278 | 4.49E-17 | 3.17E-14 | 28.29956 |
| MAFF | -2.05172 | 0.241196 | 6.295769 | -9.99219 | 5.41E-17 | 3.52E-14 | 28.11949 |
| JUND | -0.55109 | 0.682502 | 5.964615 | -9.662 | 3.02E-16 | 1.83E-13 | 26.45018 |
| NEDD9 | -0.65863 | 0.63348 | 7.288558 | -9.62829 | 3.61E-16 | 2.03E-13 | 26.27983 |
| SLC19A2 | -0.70598 | 0.613025 | 6.815192 | -9.55572 | 5.26E-16 | 2.78E-13 | 25.91319 |
| APOLD1 | -0.8281 | 0.563269 | 5.593173 | -9.42557 | 1.04E-15 | 5.15E-13 | 25.25613 |
| CISH | -0.76475 | 0.588557 | 8.148942 | -9.1562 | 4.20E-15 | 1.97E-12 | 23.89821 |
| MCL1 | -0.52537 | 0.694779 | 7.715481 | -9.08681 | 6.01E-15 | 2.68E-12 | 23.54895 |
| CEBPD | -0.82184 | 0.565718 | 9.885577 | -9.02129 | 8.45E-15 | 3.57E-12 | 23.21945 |
| RGS2 | -1.21088 | 0.432006 | 7.761442 | -8.90275 | 1.56E-14 | 6.28E-12 | 22.62399 |
| GADD45A | -0.72603 | 0.604563 | 10.4401 | -8.75949 | 3.27E-14 | 1.26E-11 | 21.9057 |
| CCNL1 | -0.48427 | 0.714857 | 7.405577 | -8.71458 | 4.12E-14 | 1.46E-11 | 21.68088 |
| CDKN1A | -0.67052 | 0.628281 | 8.146058 | -8.71334 | 4.15E-14 | 1.46E-11 | 21.67468 |
| ZBTB10 | -0.37768 | 0.769672 | 4.810673 | -8.68999 | 4.68E-14 | 1.58E-11 | 21.55785 |
| KLF9 | -0.59654 | 0.66134 | 6.925577 | -8.64234 | 5.98E-14 | 1.95E-11 | 21.31958 |
| DUSP6 | -0.81092 | 0.570017 | 7.654712 | -8.52846 | 1.07E-13 | 3.37E-11 | 20.75096 |
| SOCS2 | -0.81054 | 0.570169 | 6.630577 | -8.4349 | 1.74E-13 | 5.25E-11 | 20.28476 |
| GDF15 | -1.37634 | 0.385194 | 7.708077 | -8.22149 | 5.18E-13 | 1.51E-10 | 19.22495 |
| MTERF1 | 0.410082 | 1.328762 | 5.079135 | 8.1929 | 6.00E-13 | 1.69E-10 | 19.08337 |
| TIPARP | -0.95013 | 0.517587 | 8.469904 | -8.08508 | 1.04E-12 | 2.76E-10 | 18.5504 |
| JOSD1 | -0.37407 | 0.771602 | 7.318558 | -8.08399 | 1.04E-12 | 2.76E-10 | 18.54505 |
| SERTAD3 | -0.37379 | 0.771755 | 6.675673 | -7.99731 | 1.62E-12 | 4.16E-10 | 18.11773 |
| NR4A1 | -0.95534 | 0.515718 | 6.167212 | -7.98524 | 1.72E-12 | 4.29E-10 | 18.05833 |
| FILIP1L | -0.56673 | 0.675145 | 5.604808 | -7.97713 | 1.80E-12 | 4.34E-10 | 18.01837 |
| FERMT2 | -0.34543 | 0.787075 | 7.726923 | -7.9571 | 1.99E-12 | 4.67E-10 | 17.91984 |
| RGS3 | -0.46291 | 0.725522 | 6.334231 | -7.69973 | 7.30E-12 | 1.67E-09 | 16.65927 |
| FOSB | -1.44188 | 0.368087 | 5.944135 | -7.66529 | 8.68E-12 | 1.93E-09 | 16.49141 |
| BHLHE40 | -0.60767 | 0.656256 | 7.635673 | -7.65232 | 9.27E-12 | 2.01E-09 | 16.42824 |
| RND3 | -0.61581 | 0.652564 | 7.415481 | -7.63721 | 1.00E-11 | 2.11E-09 | 16.35467 |
| TRIB1 | -0.68239 | 0.623132 | 7.171827 | -7.5131 | 1.86E-11 | 3.84E-09 | 15.75214 |
| KLF10 | -0.83191 | 0.561784 | 7.267788 | -7.50131 | 1.97E-11 | 3.97E-09 | 15.69502 |
| EGR1 | -1.26379 | 0.41645 | 6.954327 | -7.48013 | 2.19E-11 | 4.31E-09 | 15.59252 |
| IER3 | -0.94246 | 0.520345 | 8.499615 | -7.4731 | 2.27E-11 | 4.36E-09 | 15.55854 |
| CYR61 | -1.18675 | 0.439292 | 7.185769 | -7.46903 | 2.32E-11 | 4.36E-09 | 15.53885 |

| | | | | | | | |
|---|---|---|---|---|---|---|---|
| JUNB | -0.84981 | 0.55486 | 6.403462 | -7.37635 | 3.68E-11 | 6.76E-09 | 15.09157 |
| ATF3 | -1.46087 | 0.363274 | 6.086154 | -7.35118 | 4.17E-11 | 7.50E-09 | 14.9704 |
| EIF2AK3 | -0.47436 | 0.719785 | 5.776538 | -7.33244 | 4.58E-11 | 8.06E-09 | 14.88025 |
| IL6R | -0.48115 | 0.716404 | 6.200577 | -7.31558 | 4.98E-11 | 8.59E-09 | 14.79923 |
| CHSY1 | -0.4784 | 0.717772 | 6.752981 | -7.27611 | 6.05E-11 | 1.02E-08 | 14.60973 |
| ETS2 | -0.53504 | 0.690141 | 6.735962 | -7.08621 | 1.54E-10 | 2.56E-08 | 13.70287 |
| RAB20 | -0.52031 | 0.697223 | 8.735481 | -7.02618 | 2.07E-10 | 3.37E-08 | 13.41793 |
| SLC38A2 | -0.43419 | 0.740109 | 9.988942 | -7.01007 | 2.24E-10 | 3.58E-08 | 13.34158 |
| CYP27B1 | -0.97914 | 0.507283 | 8.498942 | -6.99558 | 2.41E-10 | 3.77E-08 | 13.273 |
| MIR22HG | -0.54181 | 0.686907 | 8.478269 | -6.97589 | 2.65E-10 | 4.07E-08 | 13.17983 |
| VPS37B | -0.43485 | 0.739771 | 6.345096 | -6.96576 | 2.78E-10 | 4.20E-08 | 13.13198 |
| GEM | -0.85882 | 0.551405 | 6.338654 | -6.93276 | 3.27E-10 | 4.85E-08 | 12.97619 |
| IFRD1 | -0.42274 | 0.746008 | 5.811635 | -6.92087 | 3.47E-10 | 5.05E-08 | 12.92009 |
| JUN | -0.98238 | 0.506143 | 6.563654 | -6.88161 | 4.20E-10 | 6.01E-08 | 12.73522 |
| JADE1 | -0.39541 | 0.760272 | 8.197308 | -6.844 | 5.04E-10 | 7.10E-08 | 12.55847 |
| IFIT2 | 0.492616 | 1.406994 | 6.43625 | 6.829115 | 5.42E-10 | 7.41E-08 | 12.48861 |
| IER2 | -0.64073 | 0.641386 | 9.377885 | -6.82729 | 5.46E-10 | 7.41E-08 | 12.48006 |
| FOS | -1.65891 | 0.316678 | 6.64875 | -6.82501 | 5.52E-10 | 7.41E-08 | 12.46937 |
| CYP51A1 | -0.45387 | 0.730083 | 6.635577 | -6.75465 | 7.77E-10 | 1.03E-07 | 12.14004 |
| PER1 | -0.40331 | 0.75612 | 7.489423 | -6.73567 | 8.51E-10 | 1.11E-07 | 12.05144 |
| ERF | -0.27409 | 0.826974 | 6.721058 | -6.70145 | 1.00E-09 | 1.29E-07 | 11.89191 |
| TFRC | -0.66217 | 0.631929 | 8.032019 | -6.68937 | 1.06E-09 | 1.34E-07 | 11.83568 |
| CTGF | -0.91728 | 0.529507 | 8.577788 | -6.65207 | 1.27E-09 | 1.58E-07 | 11.6623 |
| PQLC1 | -0.47813 | 0.717906 | 9.066827 | -6.60736 | 1.58E-09 | 1.93E-07 | 11.45499 |
| YRDC | -0.22533 | 0.855399 | 7.067596 | -6.55147 | 2.06E-09 | 2.49E-07 | 11.19665 |
| COQ10B | -0.21993 | 0.85861 | 8.403654 | -6.54865 | 2.09E-09 | 2.49E-07 | 11.18363 |
| ZSCAN26 | 0.249243 | 1.188583 | 6.116827 | 6.501955 | 2.61E-09 | 3.06E-07 | 10.96853 |
| SLC20A1 | -0.45942 | 0.727281 | 7.150962 | -6.47976 | 2.90E-09 | 3.36E-07 | 10.86653 |
| SMAD7 | -0.35203 | 0.78348 | 6.659327 | -6.40705 | 4.10E-09 | 4.68E-07 | 10.53336 |
| SPRY2 | -0.3183 | 0.802015 | 6.585096 | -6.39588 | 4.32E-09 | 4.87E-07 | 10.48234 |
| HNRNPA0 | -0.18567 | 0.879238 | 8.039712 | -6.37452 | 4.78E-09 | 5.31E-07 | 10.38481 |
| HES1 | -0.45954 | 0.727217 | 7.110673 | -6.36186 | 5.07E-09 | 5.56E-07 | 10.32711 |
| SLCO4A1 | -0.78788 | 0.579195 | 6.654519 | -6.35944 | 5.13E-09 | 5.56E-07 | 10.3161 |
| MYLIP | 0.380442 | 1.301741 | 7.250865 | 6.344829 | 5.49E-09 | 5.87E-07 | 10.24954 |
| HBEGF | -0.51239 | 0.701059 | 5.691635 | -6.34256 | 5.55E-09 | 5.87E-07 | 10.2392 |
| ARF6 | -0.25196 | 0.839757 | 7.821442 | -6.31616 | 6.29E-09 | 6.56E-07 | 10.11921 |
| SPATA2L | -0.43485 | 0.739771 | 7.355096 | -6.31305 | 6.38E-09 | 6.58E-07 | 10.10505 |
| UPF2 | 0.223306 | 1.167406 | 6.441058 | 6.303881 | 6.66E-09 | 6.79E-07 | 10.06344 |
| RNF138 | -0.25929 | 0.8355 | 7.242788 | -6.29964 | 6.80E-09 | 6.83E-07 | 10.04421 |
| ZBED8 | 0.324978 | 1.252645 | 4.538846 | 6.297611 | 6.86E-09 | 6.83E-07 | 10.03499 |
| PPID | 0.307781 | 1.237802 | 7.874038 | 6.287398 | 7.20E-09 | 7.08E-07 | 9.988676 |
| PRKRA | -0.23085 | 0.852134 | 7.719519 | -6.26419 | 8.03E-09 | 7.70E-07 | 9.883563 |
| RBBP8 | -0.42292 | 0.745912 | 9.003269 | -6.26222 | 8.10E-09 | 7.70E-07 | 9.874636 |
| PPP1R15A | -0.34663 | 0.786417 | 5.91625 | -6.26208 | 8.11E-09 | 7.70E-07 | 9.874018 |
| PIM1 | -0.30756 | 0.808009 | 6.455 | -6.23519 | 9.20E-09 | 8.58E-07 | 9.752468 |
| SLC29A3 | 0.272451 | 1.207858 | 6.867596 | 6.234403 | 9.23E-09 | 8.58E-07 | 9.748897 |
| BTG3 | -0.44905 | 0.732526 | 7.564135 | -6.17096 | 1.24E-08 | 1.14E-06 | 9.463083 |

| | | | | | | | |
|---|---|---|---|---|---|---|---|
| SDPR | -0.31475 | 0.803989 | 5.305769 | -6.1696 | 1.25E-08 | 1.14E-06 | 9.456939 |
| KLF5 | -0.3075 | 0.808039 | 4.943942 | -6.1352 | 1.47E-08 | 1.32E-06 | 9.302553 |
| RNASEL | 0.247886 | 1.187466 | 6.523462 | 6.129737 | 1.50E-08 | 1.34E-06 | 9.278072 |
| LIPG | -0.5307 | 0.69222 | 5.274904 | -6.12425 | 1.54E-08 | 1.36E-06 | 9.253488 |
| LDLR | -0.45283 | 0.730606 | 5.256154 | -6.12233 | 1.56E-08 | 1.36E-06 | 9.244896 |
| ELL2 | -0.58286 | 0.667637 | 6.212115 | -6.10425 | 1.69E-08 | 1.46E-06 | 9.163982 |
| ETF1 | -0.19114 | 0.875914 | 8.845577 | -6.10102 | 1.72E-08 | 1.47E-06 | 9.149523 |
| EPHA4 | 0.301454 | 1.232386 | 6.179423 | 6.089738 | 1.81E-08 | 1.53E-06 | 9.099124 |
| FAM102A | -0.27332 | 0.827413 | 7.217788 | -6.08252 | 1.87E-08 | 1.56E-06 | 9.066896 |
| CD83 | -0.46983 | 0.722051 | 7.967981 | -6.08003 | 1.89E-08 | 1.56E-06 | 9.055787 |
| IFIT3 | 0.587504 | 1.502645 | 7.347212 | 6.079135 | 1.90E-08 | 1.56E-06 | 9.05178 |
| TUBB2A | -0.44806 | 0.733029 | 9.634904 | -6.05178 | 2.16E-08 | 1.75E-06 | 8.929817 |
| MAP2K3 | -0.25314 | 0.839068 | 7.853173 | -6.03994 | 2.28E-08 | 1.84E-06 | 8.877095 |
| ZNF165 | -0.4743 | 0.719818 | 4.61875 | -5.98866 | 2.89E-08 | 2.30E-06 | 8.649433 |
| GABARAPL | -0.30177 | 0.811257 | 8.261923 | -5.98422 | 2.95E-08 | 2.31E-06 | 8.629749 |
| FANCC | 0.240375 | 1.1813 | 6.530577 | 5.981296 | 2.99E-08 | 2.31E-06 | 8.616797 |
| VMP1 | -0.25153 | 0.840006 | 8.682115 | -5.97993 | 3.01E-08 | 2.31E-06 | 8.610752 |
| FOSL2 | -0.27972 | 0.823749 | 5.716827 | -5.97917 | 3.02E-08 | 2.31E-06 | 8.607372 |
| SERPINE1 | -0.61939 | 0.650945 | 5.514135 | -5.97787 | 3.04E-08 | 2.31E-06 | 8.601611 |
| AMOT | 0.272174 | 1.207626 | 10.10462 | 5.961194 | 3.28E-08 | 2.47E-06 | 8.527852 |
| TSC22D3 | -0.70821 | 0.61208 | 8.827212 | -5.95765 | 3.33E-08 | 2.49E-06 | 8.512177 |
| RCAN1 | -0.45361 | 0.730216 | 7.745288 | -5.93481 | 3.70E-08 | 2.72E-06 | 8.411335 |
| SLC39A14 | -0.33961 | 0.790255 | 8.055385 | -5.93377 | 3.72E-08 | 2.72E-06 | 8.406748 |
| AP3M2 | 0.244955 | 1.185056 | 7.056827 | 5.931913 | 3.75E-08 | 2.72E-06 | 8.39855 |
| CITED2 | 0.438448 | 1.355146 | 7.159519 | 5.9311 | 3.76E-08 | 2.72E-06 | 8.394964 |
| DDX5 | -0.23587 | 0.849173 | 9.934327 | -5.92282 | 3.91E-08 | 2.78E-06 | 8.358463 |
| SRSF2 | -0.19749 | 0.872067 | 8.043558 | -5.92073 | 3.94E-08 | 2.78E-06 | 8.34924 |
| ARG2 | -0.40979 | 0.752733 | 8.337115 | -5.92071 | 3.94E-08 | 2.78E-06 | 8.349163 |
| HMGCR | -0.35941 | 0.779484 | 6.610865 | -5.91849 | 3.98E-08 | 2.78E-06 | 8.339388 |
| TXNL4B | 0.231162 | 1.17378 | 6.654135 | 5.91511 | 4.05E-08 | 2.80E-06 | 8.324493 |
| MLF2 | -0.23588 | 0.849169 | 8.420192 | -5.88564 | 4.63E-08 | 3.18E-06 | 8.194854 |
| BTG2 | -0.61126 | 0.654625 | 7.130192 | -5.87731 | 4.81E-08 | 3.28E-06 | 8.158258 |
| ZNF331 | -0.48635 | 0.713829 | 6.745288 | -5.84186 | 5.65E-08 | 3.82E-06 | 8.002874 |
| EIF5 | -0.18867 | 0.877417 | 8.745 | -5.83423 | 5.85E-08 | 3.93E-06 | 7.969475 |
| YWHAZ | -0.30143 | 0.811447 | 9.832981 | -5.82439 | 6.12E-08 | 4.07E-06 | 7.926485 |
| PNRC1 | -0.26131 | 0.834329 | 10.05644 | -5.80756 | 6.61E-08 | 4.36E-06 | 7.852977 |
| TACC1 | -0.3452 | 0.787198 | 8.345962 | -5.79254 | 7.07E-08 | 4.63E-06 | 7.787469 |
| CSNK1G1 | 0.155 | 1.113422 | 5.116442 | 5.771802 | 7.77E-08 | 5.05E-06 | 7.697182 |
| IDI1 | -0.23786 | 0.848 | 8.769519 | -5.7662 | 7.97E-08 | 5.14E-06 | 7.672813 |
| PCDH17 | 0.242961 | 1.183419 | 5.267019 | 5.746586 | 8.70E-08 | 5.57E-06 | 7.587618 |
| GCLM | -0.34918 | 0.785029 | 6.820481 | -5.74309 | 8.84E-08 | 5.62E-06 | 7.572456 |
| ZNF227 | 0.224453 | 1.168334 | 4.748654 | 5.728319 | 9.45E-08 | 5.96E-06 | 7.508402 |
| CDKN1C | -0.41066 | 0.752279 | 8.3275 | -5.70943 | 1.03E-07 | 6.44E-06 | 7.426622 |
| WDR13 | -0.20425 | 0.86799 | 8.849135 | -5.70468 | 1.05E-07 | 6.53E-06 | 7.40607 |
| NFIL3 | -0.52274 | 0.696047 | 8.016731 | -5.69362 | 1.10E-07 | 6.82E-06 | 7.358285 |
| LINC00472 | -0.30299 | 0.81057 | 4.962981 | -5.68783 | 1.13E-07 | 6.95E-06 | 7.333279 |
| FAN1 | 0.194243 | 1.144124 | 6.346154 | 5.678917 | 1.18E-07 | 7.18E-06 | 7.294826 |

| | | | | | | | |
|---|---|---|---|---|---|---|---|
| FAM131A | -0.22352 | 0.856471 | 5.939038 | -5.66369 | 1.26E-07 | 7.60E-06 | 7.229209 |
| ZNF184 | -0.29417 | 0.815543 | 5.839423 | -5.66294 | 1.27E-07 | 7.60E-06 | 7.225961 |
| IARS | -0.25405 | 0.83854 | 9.657885 | -5.6548 | 1.32E-07 | 7.83E-06 | 7.190923 |
| ZNF816 | 0.299895 | 1.231055 | 6.789423 | 5.649337 | 1.35E-07 | 7.97E-06 | 7.16741 |
| CHD1 | -0.24009 | 0.846693 | 6.166538 | -5.62994 | 1.47E-07 | 8.63E-06 | 7.084062 |
| BAG3 | -0.42401 | 0.74535 | 9.32875 | -5.60493 | 1.64E-07 | 9.58E-06 | 6.976784 |
| KCTD15 | 0.179648 | 1.132607 | 5.751058 | 5.574465 | 1.88E-07 | 1.09E-05 | 6.846486 |
| BAIAP2 | -0.26845 | 0.830212 | 5.870288 | -5.57218 | 1.90E-07 | 1.09E-05 | 6.836723 |
| PPP1R3C | 0.483703 | 1.398328 | 7.175192 | 5.568161 | 1.94E-07 | 1.11E-05 | 6.819571 |
| ACKR3 | -0.49968 | 0.707265 | 6.273942 | -5.56645 | 1.95E-07 | 1.11E-05 | 6.81229 |
| SOWAHC | -0.35501 | 0.781862 | 7.34375 | -5.53543 | 2.24E-07 | 1.26E-05 | 6.680082 |
| NR4A3 | -0.38197 | 0.767388 | 4.595673 | -5.53304 | 2.26E-07 | 1.27E-05 | 6.66991 |
| LIFR | -0.22244 | 0.857112 | 5.249423 | -5.52284 | 2.37E-07 | 1.32E-05 | 6.626539 |
| GATA6 | -0.31064 | 0.806281 | 4.151538 | -5.51522 | 2.45E-07 | 1.35E-05 | 6.594183 |
| AVPI1 | -0.2752 | 0.826334 | 7.145 | -5.49834 | 2.64E-07 | 1.44E-05 | 6.522567 |
| CHIC2 | -0.22979 | 0.852759 | 7.0525 | -5.49824 | 2.64E-07 | 1.44E-05 | 6.52213 |
| MFAP1 | 0.191274 | 1.141772 | 8.278846 | 5.487861 | 2.76E-07 | 1.50E-05 | 6.478178 |
| CCDC51 | 0.246492 | 1.186319 | 8.199423 | 5.442568 | 3.37E-07 | 1.82E-05 | 6.286818 |
| LMAN2L | 0.146432 | 1.106829 | 8.215577 | 5.434021 | 3.50E-07 | 1.87E-05 | 6.250808 |
| HECA | -0.22323 | 0.856645 | 7.165288 | -5.42967 | 3.57E-07 | 1.90E-05 | 6.232509 |
| HNRNPDL | -0.17711 | 0.884475 | 8.590577 | -5.41769 | 3.76E-07 | 1.99E-05 | 6.182086 |
| F3 | -0.54786 | 0.684032 | 6.121635 | -5.41213 | 3.85E-07 | 2.02E-05 | 6.158702 |
| CHI3L1 | -0.74519 | 0.59659 | 5.746154 | -5.40716 | 3.94E-07 | 2.06E-05 | 6.137825 |
| ADM | -0.63734 | 0.642898 | 9.290865 | -5.37993 | 4.44E-07 | 2.30E-05 | 6.023636 |
| GMNN | -0.28824 | 0.818901 | 7.749904 | -5.37831 | 4.47E-07 | 2.30E-05 | 6.016848 |
| SPTSSA | -0.191 | 0.876 | 7.619135 | -5.37291 | 4.57E-07 | 2.34E-05 | 5.99424 |
| AKIRIN1 | -0.23972 | 0.846913 | 7.318269 | -5.34947 | 5.07E-07 | 2.58E-05 | 5.896298 |
| ATG101 | -0.2016 | 0.869583 | 7.403654 | -5.3341 | 5.42E-07 | 2.74E-05 | 5.832188 |
| KDELC1 | 0.314303 | 1.243411 | 5.031154 | 5.321353 | 5.73E-07 | 2.88E-05 | 5.779089 |
| DDX3X | -0.33513 | 0.79271 | 7.75875 | -5.31915 | 5.78E-07 | 2.89E-05 | 5.769929 |
| DLK2 | 0.162444 | 1.119181 | 5.592115 | 5.309685 | 6.02E-07 | 3.00E-05 | 5.730557 |
| RYBP | -0.20893 | 0.86518 | 7.604135 | -5.30399 | 6.17E-07 | 3.05E-05 | 5.706906 |
| HNF1B | 0.249445 | 1.18875 | 9.488462 | 5.30151 | 6.24E-07 | 3.07E-05 | 5.696592 |
| RRAD | -0.55349 | 0.681368 | 5.146538 | -5.30009 | 6.28E-07 | 3.07E-05 | 5.690676 |
| DUSP5 | -0.50522 | 0.704554 | 5.208462 | -5.29874 | 6.32E-07 | 3.07E-05 | 5.685073 |
| STEAP1 | 0.329888 | 1.256915 | 6.892019 | 5.288434 | 6.60E-07 | 3.19E-05 | 5.642328 |
| VEGFA | -0.26213 | 0.833857 | 7.330769 | -5.27831 | 6.90E-07 | 3.30E-05 | 5.60036 |
| ADRM1 | -0.17004 | 0.888815 | 7.720385 | -5.27821 | 6.90E-07 | 3.30E-05 | 5.599963 |
| KNOP1 | 0.195067 | 1.144778 | 6.055962 | 5.240478 | 8.12E-07 | 3.86E-05 | 5.443974 |
| ATF7IP | 0.227324 | 1.170661 | 7.062212 | 5.236065 | 8.28E-07 | 3.91E-05 | 5.425775 |
| NOL8 | 0.212796 | 1.158932 | 6.799327 | 5.232567 | 8.41E-07 | 3.94E-05 | 5.411355 |
| HOMER1 | -0.38428 | 0.766161 | 5.517212 | -5.23174 | 8.44E-07 | 3.94E-05 | 5.407929 |
| NAMPT | -0.28504 | 0.82072 | 7.525385 | -5.2304 | 8.48E-07 | 3.94E-05 | 5.402442 |
| UAP1 | -0.42223 | 0.746272 | 7.977788 | -5.22845 | 8.56E-07 | 3.95E-05 | 5.394391 |
| MCM2 | 0.198396 | 1.147422 | 6.256731 | 5.224708 | 8.70E-07 | 4.00E-05 | 5.378978 |
| METTL21B | -0.36088 | 0.778691 | 7.780481 | -5.21524 | 9.06E-07 | 4.14E-05 | 5.34002 |
| NDUFAF4 | -0.25039 | 0.840669 | 7.595577 | -5.20519 | 9.46E-07 | 4.30E-05 | 5.298684 |

| | | | | | | | |
|---|---|---|---|---|---|---|---|
| CD55 | -0.24268 | 0.845176 | 5.760096 | -5.18671 | 1.02E-06 | 4.63E-05 | 5.222848 |
| TMEM140 | 0.347391 | 1.272258 | 8.131346 | 5.183801 | 1.04E-06 | 4.66E-05 | 5.210912 |
| CHGB | -0.49055 | 0.711755 | 5.564904 | -5.17713 | 1.07E-06 | 4.77E-05 | 5.18358 |
| CUTC | -0.22964 | 0.852848 | 6.650192 | -5.17128 | 1.09E-06 | 4.87E-05 | 5.159613 |
| DNAAF2 | -0.17149 | 0.887924 | 5.832404 | -5.16289 | 1.13E-06 | 5.02E-05 | 5.125313 |
| NGRN | -0.14722 | 0.902989 | 9.445288 | -5.1448 | 1.22E-06 | 5.39E-05 | 5.051401 |
| SLC25A32 | -0.2893 | 0.818302 | 7.031923 | -5.13788 | 1.26E-06 | 5.51E-05 | 5.023184 |
| ALX1 | 0.176259 | 1.12995 | 4.917212 | 5.137338 | 1.26E-06 | 5.51E-05 | 5.020967 |
| FKBP5 | -0.9089 | 0.532592 | 7.815288 | -5.10792 | 1.43E-06 | 6.21E-05 | 4.901229 |
| ZNF606 | 0.200037 | 1.148728 | 6.102212 | 5.099481 | 1.49E-06 | 6.40E-05 | 4.866956 |
| C2orf44 | 0.156957 | 1.114933 | 5.065577 | 5.098424 | 1.49E-06 | 6.40E-05 | 4.862669 |
| JAG1 | -0.18949 | 0.876916 | 6.100192 | -5.08241 | 1.60E-06 | 6.82E-05 | 4.797734 |
| TSPYL2 | -0.26645 | 0.83136 | 6.890096 | -5.07726 | 1.63E-06 | 6.93E-05 | 4.776885 |
| WEE1 | -0.21304 | 0.862715 | 6.754231 | -5.07088 | 1.68E-06 | 7.09E-05 | 4.751072 |
| ZNF638 | 0.196304 | 1.14576 | 8.018173 | 5.06393 | 1.73E-06 | 7.26E-05 | 4.722955 |
| KIAA0753 | 0.177436 | 1.130872 | 6.851346 | 5.057181 | 1.78E-06 | 7.43E-05 | 4.695684 |
| GUCY1A3 | 0.30512 | 1.235521 | 7.19125 | 5.051284 | 1.82E-06 | 7.58E-05 | 4.671877 |
| TNFRSF1A | -0.3085 | 0.80748 | 7.754038 | -5.04473 | 1.87E-06 | 7.76E-05 | 4.645418 |
| BHLHB9 | 0.289303 | 1.22205 | 5.542212 | 5.029833 | 1.99E-06 | 8.22E-05 | 4.585416 |
| CRY1 | -0.29166 | 0.816959 | 5.865385 | -5.02193 | 2.06E-06 | 8.44E-05 | 4.553598 |
| SLBP | -0.19403 | 0.874163 | 8.06375 | -5.02096 | 2.07E-06 | 8.44E-05 | 4.549712 |
| ZC3H7A | 0.22976 | 1.17264 | 6.365962 | 5.020274 | 2.08E-06 | 8.44E-05 | 4.546956 |
| DLG3 | 0.131664 | 1.095557 | 6.120385 | 5.017646 | 2.10E-06 | 8.49E-05 | 4.536393 |
| UTP18 | -0.1884 | 0.877581 | 8.668846 | -5.00733 | 2.19E-06 | 8.82E-05 | 4.494946 |
| EFNA1 | -0.37932 | 0.768801 | 8.159327 | -4.99384 | 2.32E-06 | 9.29E-05 | 4.440835 |
| RAE1 | 0.167759 | 1.123312 | 7.463558 | 4.989496 | 2.36E-06 | 9.42E-05 | 4.423438 |
| FZD5 | -0.2082 | 0.865616 | 6.980192 | -4.98332 | 2.42E-06 | 9.60E-05 | 4.398702 |
| UBE2M | -0.23996 | 0.846767 | 7.611827 | -4.98278 | 2.43E-06 | 9.60E-05 | 4.39656 |
| FOXO1 | -0.18831 | 0.877636 | 7.423462 | -4.97402 | 2.52E-06 | 9.91E-05 | 4.361508 |
| GNA13 | -0.26088 | 0.834576 | 5.852115 | -4.97272 | 2.53E-06 | 9.92E-05 | 4.3563 |

Table S1 Gold vs. Control

| Gene | logFC | UnlogFC | AveExpr | t | P.Value | adj.P.Val | B |
|---|---|---|---|---|---|---|---|
| TMSB10 | 1.591107 | 3.012804 | 11.67899 | 16.45152 | 1.35E-27 | 6.95E-24 | 52.34374 |
| HLA-B | 1.511812 | 2.851679 | 11.53304 | 16.3997 | 1.64E-27 | 6.95E-24 | 52.14914 |
| ANXA2 | 1.508531 | 2.845202 | 10.68949 | 15.72669 | 2.21E-26 | 6.03E-23 | 49.58955 |
| PLP2 | 1.320692 | 2.497858 | 7.491899 | 15.66198 | 2.85E-26 | 6.03E-23 | 49.34028 |
| PCBP2 | -0.83478 | 0.56067 | 9.264557 | -15.4461 | 6.66E-26 | 1.13E-22 | 48.50481 |
| TIMP1 | 2.401943 | 5.285146 | 8.73443 | 15.26821 | 1.35E-25 | 1.90E-22 | 47.81162 |
| HLA-DRA | 1.722352 | 3.299739 | 10.1962 | 15.20572 | 1.72E-25 | 2.08E-22 | 47.56713 |
| B2M | 0.664025 | 1.584497 | 12.68759 | 15.12228 | 2.40E-25 | 2.54E-22 | 47.23987 |
| SUGCT | -1.10625 | 0.464501 | 8.530506 | -14.9709 | 4.40E-25 | 4.13E-22 | 46.64404 |
| CYP4A11 | -1.44953 | 0.36614 | 8.146456 | -14.7802 | 9.46E-25 | 8.00E-22 | 45.88873 |
| HPD | -2.03084 | 0.244713 | 10.95494 | -14.6378 | 1.68E-24 | 1.26E-21 | 45.32182 |
| HAGH | -0.93465 | 0.523169 | 9.123924 | -14.6217 | 1.79E-24 | 1.26E-21 | 45.25725 |
| CRYL1 | -0.87579 | 0.544955 | 10.96177 | -14.5759 | 2.16E-24 | 1.40E-21 | 45.07436 |
| TKFC | -1.27369 | 0.413601 | 6.800127 | -14.4798 | 3.19E-24 | 1.93E-21 | 44.68937 |
| UPB1 | -1.6622 | 0.315957 | 8.193924 | -14.2933 | 6.82E-24 | 3.84E-21 | 43.93903 |
| HDAC6 | -0.76596 | 0.588064 | 7.975696 | -14.239 | 8.52E-24 | 4.50E-21 | 43.71975 |
| HLA-DMA | 1.503129 | 2.834568 | 8.475063 | 14.22155 | 9.15E-24 | 4.52E-21 | 43.64909 |
| ECHDC2 | -0.66067 | 0.632586 | 10.33772 | -14.2094 | 9.62E-24 | 4.52E-21 | 43.59991 |
| TNFAIP8 | 1.667536 | 3.176716 | 6.985696 | 14.17056 | 1.13E-23 | 5.02E-21 | 43.44268 |
| CTSS | 1.627918 | 3.090667 | 5.470886 | 14.11582 | 1.41E-23 | 5.97E-21 | 43.22067 |
| CYB5A | -0.66244 | 0.63181 | 11.77633 | -14.0122 | 2.16E-23 | 8.71E-21 | 42.79936 |
| PLSCR1 | 1.271179 | 2.413588 | 8.918608 | 13.97835 | 2.49E-23 | 9.56E-21 | 42.66147 |
| SLC25A10 | -0.88918 | 0.539922 | 6.918354 | -13.9639 | 2.64E-23 | 9.58E-21 | 42.60249 |
| ACOT13 | -1.05295 | 0.481981 | 9.721899 | -13.9569 | 2.72E-23 | 9.58E-21 | 42.57382 |
| AMT | -0.853 | 0.553631 | 8.767595 | -13.9055 | 3.36E-23 | 1.14E-20 | 42.36429 |
| S100A11 | 1.23556 | 2.354727 | 9.261772 | 13.84791 | 4.27E-23 | 1.37E-20 | 42.12863 |
| MMP7 | 2.883379 | 7.378765 | 9.753797 | 13.83865 | 4.43E-23 | 1.37E-20 | 42.0907 |
| HLA-DMB | 1.39726 | 2.634008 | 8.066709 | 13.83251 | 4.55E-23 | 1.37E-20 | 42.06556 |
| MAPT | -0.92167 | 0.527899 | 7.37 | -13.8231 | 4.73E-23 | 1.38E-20 | 42.0269 |
| VIM | 1.590435 | 3.011401 | 10.81392 | 13.79688 | 5.27E-23 | 1.49E-20 | 41.91958 |
| CLU | 1.733248 | 3.324754 | 9.093797 | 13.69871 | 7.93E-23 | 2.16E-20 | 41.51648 |
| ALDH1L1 | -1.18194 | 0.440759 | 9.583671 | -13.6615 | 9.26E-23 | 2.45E-20 | 41.3632 |
| APOH | -2.35253 | 0.195802 | 8.031646 | -13.653 | 9.59E-23 | 2.46E-20 | 41.32848 |
| LPCAT1 | 1.118419 | 2.171089 | 6.722405 | 13.6114 | 1.14E-22 | 2.79E-20 | 41.15694 |
| GHR | -1.22721 | 0.427142 | 7.323671 | -13.6087 | 1.15E-22 | 2.79E-20 | 41.14588 |
| COL4A1 | 1.275323 | 2.42053 | 8.433165 | 13.48913 | 1.90E-22 | 4.47E-20 | 40.65185 |
| SERPING1 | 1.652049 | 3.142796 | 8.436835 | 13.42688 | 2.47E-22 | 5.64E-20 | 40.39398 |
| CD53 | 1.874209 | 3.666007 | 8.110506 | 13.41279 | 2.62E-22 | 5.81E-20 | 40.33554 |
| ACAA1 | -1.07428 | 0.474907 | 10.00038 | -13.4073 | 2.68E-22 | 5.81E-20 | 40.31262 |
| CD47 | 0.686943 | 1.609869 | 7.213038 | 13.3747 | 3.07E-22 | 6.27E-20 | 40.17741 |
| CDHR5 | -1.19142 | 0.437873 | 6.859494 | -13.3685 | 3.16E-22 | 6.27E-20 | 40.15171 |
| XPNPEP2 | -1.63392 | 0.322212 | 9.142911 | -13.3673 | 3.17E-22 | 6.27E-20 | 40.14651 |
| ELK3 | 0.642991 | 1.561563 | 5.187722 | 13.36126 | 3.25E-22 | 6.27E-20 | 40.1216 |
| WFDC2 | 2.51834 | 5.729225 | 9.735443 | 13.36057 | 3.26E-22 | 6.27E-20 | 40.11876 |
| CRADD | -0.51827 | 0.698207 | 7.92481 | -13.3262 | 3.77E-22 | 7.08E-20 | 39.9757 |

| | | | | | | | |
|---|---|---|---|---|---|---|---|
| ANXA5 | 0.704519 | 1.629601 | 9.91038 | 13.31267 | 3.99E-22 | 7.33E-20 | 39.91959 |
| GPD1 | -1.12871 | 0.457325 | 8.580253 | -13.2976 | 4.25E-22 | 7.65E-20 | 39.85682 |
| AHCY | -0.94557 | 0.519223 | 9.942405 | -13.2596 | 4.99E-22 | 8.79E-20 | 39.69881 |
| BPHL | -1.22259 | 0.428513 | 9.137342 | -13.2459 | 5.29E-22 | 9.12E-20 | 39.64166 |
| HRSP12 | -1.13209 | 0.456253 | 11.56101 | -13.2405 | 5.41E-22 | 9.14E-20 | 39.61916 |
| PDK2 | -0.84066 | 0.558389 | 7.721013 | -13.2294 | 5.67E-22 | 9.34E-20 | 39.57277 |
| S100A10 | 0.97139 | 1.960729 | 11.05468 | 13.22611 | 5.75E-22 | 9.34E-20 | 39.55897 |
| PSMB9 | 1.853235 | 3.613093 | 7.156962 | 13.20842 | 6.19E-22 | 9.88E-20 | 39.48514 |
| MARCKS | 0.998063 | 1.997317 | 5.534304 | 13.18794 | 6.75E-22 | 1.04E-19 | 39.39965 |
| TSPAN13 | 1.210323 | 2.313894 | 7.521013 | 13.18716 | 6.77E-22 | 1.04E-19 | 39.39641 |
| HLA-DRB1 | 1.418906 | 2.673828 | 10.83532 | 13.17825 | 7.03E-22 | 1.06E-19 | 39.3592 |
| PSMB8 | 1.376515 | 2.596404 | 7.55 | 13.16931 | 7.31E-22 | 1.08E-19 | 39.32185 |
| LAPTM5 | 2.106805 | 4.307363 | 7.598101 | 13.16584 | 7.41E-22 | 1.08E-19 | 39.30735 |
| ARPC1B | 1.358267 | 2.563771 | 8.477595 | 13.15772 | 7.67E-22 | 1.10E-19 | 39.27339 |
| SLC12A6 | -0.90384 | 0.534462 | 6.72962 | -13.1048 | 9.60E-22 | 1.35E-19 | 39.05216 |
| LHPP | -0.67868 | 0.624736 | 7.492152 | -13.0977 | 9.89E-22 | 1.37E-19 | 39.02227 |
| CNNM2 | -0.56078 | 0.677934 | 6.586835 | -13.0945 | 1.00E-21 | 1.37E-19 | 39.0089 |
| BHMT2 | -0.9604 | 0.513914 | 11.20165 | -13.0371 | 1.28E-21 | 1.72E-19 | 38.76842 |
| C1S | 2.113123 | 4.326266 | 7.721392 | 13.03314 | 1.30E-21 | 1.72E-19 | 38.75174 |
| IFI16 | 1.553966 | 2.936232 | 6.433038 | 13.00475 | 1.47E-21 | 1.91E-19 | 38.6326 |
| TPBG | 1.119466 | 2.172666 | 7.100886 | 12.98714 | 1.58E-21 | 2.01E-19 | 38.55867 |
| NMI | 1.129908 | 2.188447 | 7.163291 | 12.9858 | 1.59E-21 | 2.01E-19 | 38.55302 |
| TGFBI | 1.73137 | 3.32043 | 7.796709 | 12.97229 | 1.68E-21 | 2.09E-19 | 38.49626 |
| ECI2 | -0.8524 | 0.553861 | 10.39937 | -12.9542 | 1.82E-21 | 2.23E-19 | 38.42033 |
| HLA-E | 0.869407 | 1.826912 | 9.950127 | 12.93866 | 1.94E-21 | 2.35E-19 | 38.35489 |
| PNP | -0.89639 | 0.537229 | 9.791646 | -12.9276 | 2.04E-21 | 2.41E-19 | 38.30854 |
| FBXO7 | -0.48937 | 0.712337 | 9.979494 | -12.9206 | 2.10E-21 | 2.41E-19 | 38.27875 |
| MYOF | 1.178281 | 2.263069 | 7.419367 | 12.91898 | 2.11E-21 | 2.41E-19 | 38.27208 |
| EAF2 | -1.10256 | 0.465689 | 8.721392 | -12.9188 | 2.11E-21 | 2.41E-19 | 38.27151 |
| LY96 | 2.215211 | 4.643494 | 5.481646 | 12.89128 | 2.38E-21 | 2.62E-19 | 38.15545 |
| CD74 | 1.227029 | 2.340844 | 9.888861 | 12.89047 | 2.39E-21 | 2.62E-19 | 38.15204 |
| CTH | -0.98705 | 0.504509 | 6.765949 | -12.8898 | 2.39E-21 | 2.62E-19 | 38.14926 |
| TRIM22 | 1.680837 | 3.206138 | 7.818861 | 12.88712 | 2.42E-21 | 2.62E-19 | 38.1379 |
| GADD45A | -1.27803 | 0.412358 | 10.31114 | -12.8803 | 2.49E-21 | 2.66E-19 | 38.10939 |
| ITM2C | 1.082101 | 2.117118 | 8.986582 | 12.87556 | 2.54E-21 | 2.69E-19 | 38.08923 |
| ACTN1 | 1.219974 | 2.329425 | 7.155696 | 12.81306 | 3.32E-21 | 3.46E-19 | 37.82561 |
| PARP12 | 1.04556 | 2.064167 | 6.712405 | 12.80115 | 3.49E-21 | 3.60E-19 | 37.77531 |
| ADH6 | -1.019 | 0.493459 | 8.303038 | -12.7201 | 4.94E-21 | 4.98E-19 | 37.43265 |
| YWHAH | 1.086324 | 2.123323 | 7.542911 | 12.71938 | 4.95E-21 | 4.98E-19 | 37.42962 |
| ARPC2 | 0.502437 | 1.416605 | 9.916835 | 12.70532 | 5.26E-21 | 5.23E-19 | 37.37009 |
| PCK2 | -1.40736 | 0.377 | 9.973418 | -12.7008 | 5.36E-21 | 5.25E-19 | 37.35103 |
| RGS10 | 1.525883 | 2.879629 | 6.498481 | 12.6992 | 5.40E-21 | 5.25E-19 | 37.34418 |
| LIPA | 0.888959 | 1.85184 | 9.401772 | 12.68213 | 5.81E-21 | 5.58E-19 | 37.27189 |
| DPYS | -1.32312 | 0.399669 | 10.36861 | -12.6629 | 6.31E-21 | 5.99E-19 | 37.19054 |
| PC | -0.94918 | 0.517928 | 8.213291 | -12.6397 | 6.97E-21 | 6.54E-19 | 37.09195 |
| COL6A3 | 2.098511 | 4.282672 | 6.570506 | 12.59753 | 8.35E-21 | 7.76E-19 | 36.91301 |
| ARPC5 | 0.700751 | 1.625351 | 9.319241 | 12.57252 | 9.30E-21 | 8.54E-19 | 36.8068 |

| | | | | | | | |
|---|---|---|---|---|---|---|---|
| IFITM1 | 1.819111 | 3.528636 | 9.125316 | 12.56539 | 9.58E-21 | 8.71E-19 | 36.77649 |
| NECAP2 | 0.631238 | 1.548894 | 7.467595 | 12.55374 | 1.01E-20 | 9.06E-19 | 36.72694 |
| ARHGDIB | 1.510138 | 2.848374 | 7.898861 | 12.54858 | 1.03E-20 | 9.17E-19 | 36.705 |
| DCK | 0.8833 | 1.84459 | 5.466582 | 12.52952 | 1.12E-20 | 9.85E-19 | 36.62395 |
| C2orf47 | -0.5624 | 0.677173 | 9.340506 | -12.5054 | 1.24E-20 | 1.08E-18 | 36.52146 |
| DCXR | -1.41625 | 0.374686 | 11.21101 | -12.4862 | 1.35E-20 | 1.16E-18 | 36.43936 |
| HPGD | -1.48743 | 0.356647 | 7.90519 | -12.4758 | 1.41E-20 | 1.20E-18 | 36.39522 |
| DNPH1 | -0.63682 | 0.64313 | 9.758987 | -12.4715 | 1.44E-20 | 1.21E-18 | 36.37702 |
| ACOX2 | -1.15797 | 0.448142 | 9.007595 | -12.4541 | 1.55E-20 | 1.30E-18 | 36.30257 |
| NNMT | 2.824177 | 7.082097 | 7.614937 | 12.43686 | 1.67E-20 | 1.38E-18 | 36.22921 |
| GCHFR | -0.78602 | 0.579941 | 8.562532 | -12.4332 | 1.69E-20 | 1.39E-18 | 36.21372 |
| MSRA | -1.0094 | 0.496753 | 10.65987 | -12.4299 | 1.72E-20 | 1.40E-18 | 36.19961 |
| ALDH6A1 | -1.26454 | 0.416232 | 10.82025 | -12.419 | 1.80E-20 | 1.45E-18 | 36.15309 |
| COL4A2 | 1.151937 | 2.22212 | 7.753797 | 12.39345 | 2.01E-20 | 1.60E-18 | 36.04397 |
| HLA-DPA1 | 1.767292 | 3.404145 | 9.983671 | 12.38042 | 2.13E-20 | 1.67E-18 | 35.98833 |
| CASP1 | 1.446014 | 2.724543 | 5.314684 | 12.37937 | 2.14E-20 | 1.67E-18 | 35.98385 |
| LYZ | 3.083228 | 8.475086 | 6.762278 | 12.3747 | 2.18E-20 | 1.67E-18 | 35.96389 |
| VEGFA | -0.80362 | 0.572909 | 7.141266 | -12.3734 | 2.19E-20 | 1.67E-18 | 35.95814 |
| ALDH2 | -0.58393 | 0.667146 | 11.39304 | -12.3733 | 2.19E-20 | 1.67E-18 | 35.95782 |
| CHPT1 | -0.69771 | 0.616551 | 9.438987 | -12.3688 | 2.24E-20 | 1.69E-18 | 35.93861 |
| TES | 0.857404 | 1.811776 | 6.837722 | 12.35086 | 2.42E-20 | 1.81E-18 | 35.862 |
| AZGP1 | -1.64496 | 0.319755 | 9.768734 | -12.3437 | 2.49E-20 | 1.85E-18 | 35.8312 |
| C1QA | 2.051436 | 4.145184 | 6.79519 | 12.33012 | 2.64E-20 | 1.94E-18 | 35.7733 |
| GSTA1 | -1.68038 | 0.312 | 11.71633 | -12.327 | 2.68E-20 | 1.95E-18 | 35.75987 |
| SLC13A3 | -1.17769 | 0.442057 | 10.08544 | -12.3123 | 2.85E-20 | 2.06E-18 | 35.69721 |
| STUB1 | -0.37486 | 0.771179 | 9.396456 | -12.3074 | 2.91E-20 | 2.09E-18 | 35.67626 |
| TUBA1A | 1.423979 | 2.683245 | 9.935696 | 12.30516 | 2.94E-20 | 2.09E-18 | 35.66649 |
| PEPD | -1.00931 | 0.496784 | 11.29861 | -12.2909 | 3.13E-20 | 2.21E-18 | 35.60557 |
| PROM1 | 2.21002 | 4.626816 | 8.178608 | 12.26874 | 3.45E-20 | 2.41E-18 | 35.51055 |
| ALPL | -0.99348 | 0.502265 | 5.828481 | -12.2654 | 3.50E-20 | 2.42E-18 | 35.49615 |
| GRPEL1 | -0.66313 | 0.631507 | 8.625823 | -12.2636 | 3.52E-20 | 2.42E-18 | 35.4887 |
| CPVL | 1.032088 | 2.044982 | 8.428734 | 12.24752 | 3.78E-20 | 2.58E-18 | 35.41963 |
| GALK1 | -0.72653 | 0.604354 | 6.676076 | -12.2423 | 3.86E-20 | 2.61E-18 | 35.39734 |
| DHRS3 | -0.70831 | 0.612038 | 8.773038 | -12.2157 | 4.34E-20 | 2.91E-18 | 35.28331 |
| HYAL1 | -0.83424 | 0.560877 | 9.331519 | -12.2065 | 4.51E-20 | 3.00E-18 | 35.2436 |
| CKLF | 1.371469 | 2.587339 | 6.803544 | 12.19399 | 4.77E-20 | 3.15E-18 | 35.19001 |
| SLC37A4 | -0.94013 | 0.521185 | 9.627722 | -12.19 | 4.85E-20 | 3.18E-18 | 35.17274 |
| VCAN | 2.007227 | 4.020087 | 5.249114 | 12.18832 | 4.88E-20 | 3.18E-18 | 35.16563 |
| C11orf71 | -0.86689 | 0.548327 | 7.23962 | -12.1277 | 6.35E-20 | 4.10E-18 | 34.90522 |
| TAP1 | 1.045244 | 2.063715 | 7.305316 | 12.11406 | 6.74E-20 | 4.32E-18 | 34.84652 |
| SHMT1 | -0.97426 | 0.509002 | 9.906076 | -12.1048 | 7.02E-20 | 4.46E-18 | 34.8066 |
| DNMT1 | 0.767451 | 1.702259 | 6.419494 | 12.09019 | 7.48E-20 | 4.72E-18 | 34.74382 |
| RAB27A | 0.783827 | 1.721692 | 6.183291 | 12.07855 | 7.87E-20 | 4.93E-18 | 34.69373 |
| HADH | -0.92892 | 0.525252 | 9.942405 | -12.0619 | 8.46E-20 | 5.26E-18 | 34.6221 |
| PXDN | 1.31413 | 2.486524 | 6.28481 | 12.05129 | 8.86E-20 | 5.47E-18 | 34.57632 |
| ITGB2 | 1.631798 | 3.098991 | 6.256203 | 12.02319 | 1.00E-19 | 6.13E-18 | 34.45521 |
| TYROBP | 2.11726 | 4.33869 | 7.247468 | 12.01509 | 1.04E-19 | 6.31E-18 | 34.42031 |

| | | | | | | | |
|---|---|---|---|---|---|---|---|
| PRODH2 | -1.37536 | 0.385456 | 10.80418 | -12.0117 | 1.05E-19 | 6.36E-18 | 34.40571 |
| TPM1 | 1.016179 | 2.022555 | 8.917089 | 12.00738 | 1.07E-19 | 6.42E-18 | 34.38706 |
| FTCD | -1.39675 | 0.379783 | 8.706329 | -12.0063 | 1.08E-19 | 6.42E-18 | 34.38237 |
| RAB11FIP3 | -1.0764 | 0.47421 | 7.999494 | -11.9919 | 1.15E-19 | 6.79E-18 | 34.3201 |
| HLA-C | 0.894862 | 1.859432 | 11.79076 | 11.98091 | 1.20E-19 | 7.04E-18 | 34.27285 |
| PKLR | -0.76795 | 0.587251 | 6.455949 | -11.9803 | 1.21E-19 | 7.04E-18 | 34.27035 |
| EMP3 | 1.208794 | 2.311444 | 7.053418 | 11.97149 | 1.25E-19 | 7.26E-18 | 34.23221 |
| TRIOBP | 0.496561 | 1.410847 | 7.911772 | 11.95257 | 1.36E-19 | 7.84E-18 | 34.1505 |
| S100A4 | 1.412069 | 2.661184 | 8.002025 | 11.95048 | 1.38E-19 | 7.85E-18 | 34.1415 |
| HCLS1 | 1.693636 | 3.23471 | 6.467468 | 11.94748 | 1.39E-19 | 7.88E-18 | 34.12854 |
| AUH | -0.50535 | 0.70449 | 10.08456 | -11.9467 | 1.40E-19 | 7.88E-18 | 34.12524 |
| IDH3B | -0.39673 | 0.75958 | 9.167975 | -11.926 | 1.53E-19 | 8.57E-18 | 34.0359 |
| PHYH | -1.01472 | 0.494925 | 10.8357 | -11.9098 | 1.64E-19 | 9.13E-18 | 33.96576 |
| HLA-F | 1.182451 | 2.26962 | 9.857848 | 11.90677 | 1.66E-19 | 9.20E-18 | 33.95262 |
| HDHD3 | -0.80088 | 0.574 | 7.847848 | -11.901 | 1.71E-19 | 9.37E-18 | 33.92752 |
| HLA-J | 1.008847 | 2.012302 | 8.207722 | 11.89819 | 1.73E-19 | 9.42E-18 | 33.91552 |
| AFM | -1.94978 | 0.258855 | 7.835316 | -11.8936 | 1.76E-19 | 9.55E-18 | 33.89586 |
| SDHB | -0.67379 | 0.626859 | 9.313544 | -11.8758 | 1.91E-19 | 1.03E-17 | 33.81882 |
| G6PC | -2.21359 | 0.215597 | 7.564684 | -11.859 | 2.05E-19 | 1.10E-17 | 33.74588 |
| MICU1 | -0.72629 | 0.604456 | 8.769873 | -11.8557 | 2.08E-19 | 1.11E-17 | 33.7316 |
| MNS1 | 1.114796 | 2.165644 | 5.587848 | 11.83108 | 2.32E-19 | 1.22E-17 | 33.62505 |
| NIT1 | -0.52138 | 0.696707 | 8.200253 | -11.8287 | 2.34E-19 | 1.23E-17 | 33.61463 |
| LRRC23 | -0.56036 | 0.678135 | 6.288101 | -11.8146 | 2.49E-19 | 1.30E-17 | 33.55344 |
| TM4SF5 | -0.91919 | 0.528806 | 8.108861 | -11.8125 | 2.51E-19 | 1.30E-17 | 33.54459 |
| QPCT | 1.640264 | 3.117228 | 5.364304 | 11.80871 | 2.56E-19 | 1.32E-17 | 33.52811 |
| MS4A6A | 1.719605 | 3.293462 | 7.12962 | 11.79269 | 2.74E-19 | 1.40E-17 | 33.45867 |
| HSD17B8 | -0.50185 | 0.7062 | 8.977975 | -11.7659 | 3.08E-19 | 1.57E-17 | 33.34256 |
| OCEL1 | -0.5396 | 0.687959 | 8.163291 | -11.7609 | 3.15E-19 | 1.60E-17 | 33.32078 |
| ESYT1 | 0.729552 | 1.658124 | 7.968228 | 11.75121 | 3.29E-19 | 1.65E-17 | 33.27877 |
| ASB9 | -1.20418 | 0.434015 | 6.893291 | -11.7323 | 3.57E-19 | 1.79E-17 | 33.19646 |
| LTF | 3.889394 | 14.81918 | 8.414684 | 11.72708 | 3.66E-19 | 1.82E-17 | 33.17401 |
| LAPTM4B | 0.668445 | 1.589359 | 9.778354 | 11.69174 | 4.27E-19 | 2.11E-17 | 33.02048 |
| CAMK2G | -0.51746 | 0.698599 | 6.947975 | -11.6884 | 4.33E-19 | 2.13E-17 | 33.0061 |
| COL3A1 | 2.30971 | 4.957835 | 7.541772 | 11.6754 | 4.59E-19 | 2.24E-17 | 32.94945 |
| C1R | 1.692253 | 3.23161 | 7.776456 | 11.67411 | 4.61E-19 | 2.24E-17 | 32.94385 |
| SUCLG1 | -0.69067 | 0.619565 | 12.05671 | -11.6587 | 4.93E-19 | 2.38E-17 | 32.87693 |
| CLIC1 | 0.689598 | 1.612834 | 10.12089 | 11.65488 | 5.02E-19 | 2.40E-17 | 32.8602 |
| PXMP2 | -0.88623 | 0.541025 | 9.179367 | -11.6546 | 5.02E-19 | 2.40E-17 | 32.85912 |
| TRPM3 | -0.79634 | 0.575807 | 6.992785 | -11.6373 | 5.42E-19 | 2.57E-17 | 32.78369 |
| SH2B3 | 0.873564 | 1.832183 | 7.231646 | 11.63621 | 5.45E-19 | 2.57E-17 | 32.779 |
| ETFB | -0.59335 | 0.662804 | 9.92519 | -11.633 | 5.53E-19 | 2.60E-17 | 32.76497 |
| CTSV | -1.59865 | 0.330186 | 8.270253 | -11.6212 | 5.82E-19 | 2.72E-17 | 32.71348 |
| SLC25A1 | -0.67941 | 0.624419 | 8.150759 | -11.6147 | 5.99E-19 | 2.78E-17 | 32.68537 |
| CD48 | 2.175428 | 4.517198 | 6.72481 | 11.61204 | 6.06E-19 | 2.80E-17 | 32.67377 |
| RBKS | -0.57814 | 0.669829 | 8.977848 | -11.6109 | 6.09E-19 | 2.80E-17 | 32.66888 |
| ANXA1 | 1.715949 | 3.285126 | 7.532658 | 11.60785 | 6.17E-19 | 2.82E-17 | 32.65553 |
| MME | -1.46833 | 0.3614 | 10.19165 | -11.5914 | 6.64E-19 | 3.02E-17 | 32.58378 |

| | | | | | | | |
|---|---|---|---|---|---|---|---|
| UCP2 | 1.058906 | 2.083352 | 7.074937 | 11.56592 | 7.42E-19 | 3.36E-17 | 32.47287 |
| DSE | 1.101476 | 2.145741 | 7.039241 | 11.5595 | 7.64E-19 | 3.43E-17 | 32.44492 |
| KIAA1551 | 0.81643 | 1.761042 | 6.245823 | 11.5517 | 7.90E-19 | 3.53E-17 | 32.4109 |
| ECSIT | -0.54187 | 0.68688 | 6.795823 | -11.5477 | 8.04E-19 | 3.58E-17 | 32.39339 |
| GLYAT | -1.21067 | 0.432069 | 10.15797 | -11.532 | 8.62E-19 | 3.81E-17 | 32.32519 |
| ETNK2 | -1.13213 | 0.456242 | 7.148608 | -11.5237 | 8.94E-19 | 3.94E-17 | 32.28872 |
| METTL21B | -1.03022 | 0.489634 | 7.551392 | -11.5037 | 9.76E-19 | 4.28E-17 | 32.20155 |
| ACSF2 | -1.03094 | 0.48939 | 9.376962 | -11.4975 | 1.00E-18 | 4.37E-17 | 32.17434 |
| SLC22A8 | -1.48776 | 0.356566 | 9.931139 | -11.4965 | 1.01E-18 | 4.37E-17 | 32.16999 |
| CRYM | -1.02539 | 0.491278 | 10.66646 | -11.4894 | 1.04E-18 | 4.49E-17 | 32.13896 |
| ST13 | -0.50416 | 0.705069 | 10.53114 | -11.4882 | 1.05E-18 | 4.49E-17 | 32.13383 |
| ACTR3 | 0.561324 | 1.475623 | 8.548608 | 11.48407 | 1.06E-18 | 4.55E-17 | 32.11583 |
| EPHX1 | -1.00823 | 0.497157 | 8.879494 | -11.4738 | 1.11E-18 | 4.73E-17 | 32.07114 |
| C2CD2 | 0.683603 | 1.606146 | 6.693165 | 11.45941 | 1.19E-18 | 5.02E-17 | 32.00808 |
| PSMB10 | 1.264756 | 2.402866 | 6.75962 | 11.45836 | 1.19E-18 | 5.02E-17 | 32.00352 |
| GLRX | -0.49159 | 0.711242 | 10.21139 | -11.4534 | 1.22E-18 | 5.10E-17 | 31.98184 |
| KMO | -1.14689 | 0.451597 | 8.862658 | -11.4451 | 1.26E-18 | 5.24E-17 | 31.94555 |
| PHF11 | 0.625356 | 1.542591 | 8.544051 | 11.44495 | 1.27E-18 | 5.24E-17 | 31.94493 |
| DDC | -1.15312 | 0.449651 | 10.10962 | -11.435 | 1.32E-18 | 5.45E-17 | 31.90151 |
| ANKRD46 | -0.74474 | 0.596777 | 8.453038 | -11.4217 | 1.40E-18 | 5.75E-17 | 31.84333 |
| SLC22A6 | -1.35191 | 0.391773 | 9.426582 | -11.4102 | 1.48E-18 | 6.02E-17 | 31.79319 |
| TNC | 1.586785 | 3.003793 | 6.005443 | 11.39901 | 1.55E-18 | 6.30E-17 | 31.74405 |
| RABGGTB | -0.56632 | 0.675338 | 7.982785 | -11.3904 | 1.61E-18 | 6.51E-17 | 31.70643 |
| MRPS15 | -0.43895 | 0.737673 | 7.523165 | -11.3865 | 1.64E-18 | 6.59E-17 | 31.68946 |
| PECAM1 | 1.05832 | 2.082505 | 7.130127 | 11.38322 | 1.66E-18 | 6.64E-17 | 31.67495 |
| RTP4 | 1.017345 | 2.024191 | 5.688228 | 11.38286 | 1.67E-18 | 6.64E-17 | 31.67336 |
| PYCARD | 1.455165 | 2.741879 | 5.47481 | 11.38037 | 1.68E-18 | 6.68E-17 | 31.66248 |
| ANXA2P2 | 0.78334 | 1.721111 | 6.255696 | 11.36195 | 1.83E-18 | 7.21E-17 | 31.58185 |
| GBP2 | 1.35695 | 2.561431 | 5.521392 | 11.35554 | 1.88E-18 | 7.39E-17 | 31.55376 |
| GALNT3 | 0.786798 | 1.725242 | 6.078228 | 11.32998 | 2.10E-18 | 8.22E-17 | 31.4418 |
| SPAG5 | -0.89533 | 0.537624 | 7.707089 | -11.3294 | 2.11E-18 | 8.22E-17 | 31.43947 |
| TLR2 | 0.811574 | 1.755126 | 5.805316 | 11.30176 | 2.38E-18 | 9.24E-17 | 31.31813 |
| HSD17B14 | -0.942 | 0.52051 | 7.729114 | -11.2928 | 2.48E-18 | 9.57E-17 | 31.27887 |
| STAT1 | 1.185567 | 2.274527 | 7.279367 | 11.2849 | 2.57E-18 | 9.87E-17 | 31.24421 |
| SFXN1 | -0.68989 | 0.619899 | 7.877468 | -11.2835 | 2.58E-18 | 9.89E-17 | 31.23805 |
| METTL1 | -0.77661 | 0.583735 | 7.983418 | -11.2796 | 2.63E-18 | 1.00E-16 | 31.22077 |
| UBE2L6 | 1.024651 | 2.034467 | 8.023671 | 11.26602 | 2.79E-18 | 1.06E-16 | 31.16141 |
| ABCC6 | -0.9417 | 0.520619 | 6.899241 | -11.2641 | 2.82E-18 | 1.06E-16 | 31.15288 |
| IQGAP1 | 0.623412 | 1.540515 | 7.862152 | 11.26274 | 2.83E-18 | 1.06E-16 | 31.14701 |
| XRCC5 | 0.442437 | 1.358898 | 9.581772 | 11.25367 | 2.95E-18 | 1.10E-16 | 31.10721 |
| CES2 | -0.79082 | 0.578017 | 9.578354 | -11.2492 | 3.01E-18 | 1.12E-16 | 31.08742 |
| FAH | -0.84911 | 0.555127 | 8.559873 | -11.249 | 3.01E-18 | 1.12E-16 | 31.0869 |
| ALDH7A1 | -0.59052 | 0.664103 | 6.213544 | -11.2323 | 3.24E-18 | 1.20E-16 | 31.01346 |
| HRG | -1.39023 | 0.381504 | 8.232532 | -11.2297 | 3.28E-18 | 1.21E-16 | 31.00178 |
| CKAP4 | 0.586153 | 1.501238 | 7.693544 | 11.22652 | 3.33E-18 | 1.22E-16 | 30.98802 |
| BDH2 | -0.64628 | 0.638927 | 10.74025 | -11.2216 | 3.40E-18 | 1.24E-16 | 30.9663 |
| SERPINA6 | -1.43318 | 0.370315 | 8.43481 | -11.1778 | 4.13E-18 | 1.50E-16 | 30.77403 |

| | | | | | | | |
|---|---|---|---|---|---|---|---|
| GPS1 | -0.53125 | 0.691957 | 7.735696 | -11.1685 | 4.30E-18 | 1.56E-16 | 30.73326 |
| RUNDC3B | -0.73428 | 0.60112 | 5.660886 | -11.1645 | 4.38E-18 | 1.58E-16 | 30.71548 |
| PDLIM1 | 0.884697 | 1.846377 | 9.434557 | 11.15281 | 4.62E-18 | 1.65E-16 | 30.66412 |
| SCRN1 | 0.804717 | 1.746803 | 7.536582 | 11.152 | 4.63E-18 | 1.65E-16 | 30.66056 |
| TREH | -1.76458 | 0.294311 | 6.732025 | -11.148 | 4.72E-18 | 1.68E-16 | 30.6428 |
| IFNGR1 | 0.719717 | 1.646859 | 8.066076 | 11.14449 | 4.79E-18 | 1.69E-16 | 30.62752 |
| EVI2A | 1.91643 | 3.774877 | 5.635316 | 11.13797 | 4.93E-18 | 1.74E-16 | 30.59887 |
| PLOD1 | -0.62974 | 0.646294 | 7.746076 | -11.129 | 5.13E-18 | 1.80E-16 | 30.55961 |
| CCDC109B | 1.03529 | 2.049525 | 5.650506 | 11.12474 | 5.23E-18 | 1.83E-16 | 30.54064 |
| TLE1 | 0.531542 | 1.445473 | 6.095949 | 11.11905 | 5.36E-18 | 1.87E-16 | 30.51563 |
| FBP1 | -1.2606 | 0.41737 | 10.9319 | -11.1176 | 5.40E-18 | 1.87E-16 | 30.50927 |
| CAT | -0.63227 | 0.645162 | 10.97937 | -11.1141 | 5.48E-18 | 1.88E-16 | 30.49385 |
| HOPX | 1.849321 | 3.603307 | 6.359241 | 11.11402 | 5.48E-18 | 1.88E-16 | 30.49349 |
| C4orf27 | 0.49083 | 1.405253 | 7.873291 | 11.09102 | 6.07E-18 | 2.08E-16 | 30.39225 |
| LYRM1 | -0.43214 | 0.741161 | 9.634051 | -11.0901 | 6.10E-18 | 2.08E-16 | 30.388 |
| KHK | -0.93795 | 0.521974 | 9.644937 | -11.0833 | 6.29E-18 | 2.13E-16 | 30.35807 |
| LYN | 1.019789 | 2.027623 | 5.844684 | 11.07147 | 6.63E-18 | 2.24E-16 | 30.30613 |
| ITGB6 | 1.953775 | 3.873868 | 5.604177 | 11.05415 | 7.16E-18 | 2.41E-16 | 30.22985 |
| CFDP1 | -0.70036 | 0.61542 | 9.38962 | -11.0488 | 7.33E-18 | 2.46E-16 | 30.20613 |
| ECHS1 | -0.6002 | 0.659664 | 11.96494 | -11.048 | 7.36E-18 | 2.46E-16 | 30.20288 |
| FSTL1 | 1.160586 | 2.235483 | 8.959367 | 11.03869 | 7.67E-18 | 2.55E-16 | 30.16171 |
| SEMA3B | -0.63843 | 0.642411 | 7.141139 | -11.037 | 7.73E-18 | 2.56E-16 | 30.1543 |
| MAOB | -0.66926 | 0.628831 | 10.70848 | -11.0325 | 7.88E-18 | 2.60E-16 | 30.13428 |
| MPZL1 | 0.450929 | 1.36692 | 6.943797 | 11.02285 | 8.23E-18 | 2.71E-16 | 30.09186 |
| SP110 | 0.626686 | 1.544015 | 6.497215 | 11.01919 | 8.36E-18 | 2.74E-16 | 30.07572 |
| TBC1D13 | -0.50595 | 0.704197 | 7.612785 | -10.9837 | 9.80E-18 | 3.20E-16 | 29.91905 |
| IFITM2 | 1.293775 | 2.451687 | 9.328481 | 10.97974 | 9.97E-18 | 3.24E-16 | 29.90172 |
| CD44 | 0.962424 | 1.948581 | 5.972025 | 10.9785 | 1.00E-17 | 3.24E-16 | 29.89625 |
| SLC25A44 | -0.5516 | 0.682263 | 7.889367 | -10.9785 | 1.00E-17 | 3.24E-16 | 29.89611 |
| AKR1B1 | 0.764519 | 1.698804 | 9.625443 | 10.96765 | 1.05E-17 | 3.38E-16 | 29.84837 |
| S100A6 | 0.965797 | 1.953142 | 10.0043 | 10.96699 | 1.06E-17 | 3.38E-16 | 29.84546 |
| PDE10A | -0.33292 | 0.793925 | 4.746582 | -10.9407 | 1.19E-17 | 3.79E-16 | 29.72936 |
| PARP8 | 0.631232 | 1.548887 | 6.099114 | 10.92476 | 1.27E-17 | 4.05E-16 | 29.659 |
| CCNB1IP1 | -0.64073 | 0.641388 | 9.191266 | -10.9216 | 1.29E-17 | 4.09E-16 | 29.64513 |
| CALM2 | 0.401607 | 1.320979 | 12.69646 | 10.91393 | 1.34E-17 | 4.22E-16 | 29.61114 |
| SPON2 | 1.480316 | 2.790099 | 7.465316 | 10.91299 | 1.34E-17 | 4.22E-16 | 29.60698 |
| NFS1 | -0.64706 | 0.638582 | 8.519494 | -10.9119 | 1.35E-17 | 4.22E-16 | 29.60233 |
| CFB | 1.762378 | 3.392569 | 7.616835 | 10.90725 | 1.38E-17 | 4.30E-16 | 29.58165 |
| HLA-A | 0.881028 | 1.841687 | 12.29063 | 10.90584 | 1.39E-17 | 4.31E-16 | 29.57538 |
| FDX1 | -0.65244 | 0.636205 | 9.020506 | -10.9023 | 1.41E-17 | 4.36E-16 | 29.55992 |
| ECH1 | -0.43603 | 0.739167 | 11.30025 | -10.8899 | 1.49E-17 | 4.59E-16 | 29.50505 |
| RAB11FIP5 | -0.54626 | 0.684791 | 8.018987 | -10.8885 | 1.50E-17 | 4.60E-16 | 29.49895 |
| DIP2C | -0.8691 | 0.547489 | 8.298481 | -10.8781 | 1.57E-17 | 4.80E-16 | 29.45288 |
| BLNK | 0.914236 | 1.884571 | 7.543418 | 10.87684 | 1.58E-17 | 4.80E-16 | 29.44724 |
| FABP1 | -1.63947 | 0.320974 | 10.74342 | -10.8767 | 1.58E-17 | 4.80E-16 | 29.44681 |
| CMTM6 | 0.639835 | 1.558151 | 9.125316 | 10.85806 | 1.72E-17 | 5.20E-16 | 29.36416 |
| ALOX5AP | 1.400922 | 2.640703 | 6.732152 | 10.84686 | 1.80E-17 | 5.45E-16 | 29.31463 |

| | | | | | | | |
|---|---|---|---|---|---|---|---|
| NCF2 | 1.148617 | 2.217012 | 5.109367 | 10.84059 | 1.86E-17 | 5.57E-16 | 29.28687 |
| PTPRC | 1.35087 | 2.550658 | 5.153418 | 10.84018 | 1.86E-17 | 5.57E-16 | 29.28508 |
| C3 | 2.445296 | 5.446375 | 8.657975 | 10.83893 | 1.87E-17 | 5.58E-16 | 29.27955 |
| CASP4 | 0.905059 | 1.872621 | 6.389367 | 10.83766 | 1.88E-17 | 5.60E-16 | 29.27394 |
| SELENBP1 | -1.02545 | 0.491255 | 8.821646 | -10.8286 | 1.96E-17 | 5.81E-16 | 29.23381 |
| S100A13 | 0.979947 | 1.972393 | 9.131519 | 10.82788 | 1.96E-17 | 5.81E-16 | 29.23065 |
| GPR137B | -0.87329 | 0.545902 | 9.454557 | -10.8258 | 1.98E-17 | 5.84E-16 | 29.22164 |
| HAAO | -0.65167 | 0.636545 | 7.702785 | -10.822 | 2.02E-17 | 5.92E-16 | 29.20485 |
| H2AFY | 0.413505 | 1.331917 | 8.513165 | 10.8175 | 2.06E-17 | 6.02E-16 | 29.18474 |
| POLD3 | -0.84302 | 0.557474 | 7.476329 | -10.8109 | 2.12E-17 | 6.18E-16 | 29.15558 |
| C1QB | 2.112727 | 4.325081 | 6.422532 | 10.80798 | 2.15E-17 | 6.24E-16 | 29.14259 |
| ITGAV | 0.782477 | 1.720082 | 9.354684 | 10.80728 | 2.15E-17 | 6.24E-16 | 29.13947 |
| GRHPR | -0.71552 | 0.608985 | 10.36633 | -10.8023 | 2.20E-17 | 6.33E-16 | 29.11759 |
| KLK1 | -2.0001 | 0.249983 | 8.187342 | -10.8023 | 2.20E-17 | 6.33E-16 | 29.11742 |
| PLG | -2.02401 | 0.245875 | 10.02975 | -10.7981 | 2.24E-17 | 6.43E-16 | 29.09892 |
| AKR1A1 | -0.70559 | 0.61319 | 11.74722 | -10.7917 | 2.31E-17 | 6.59E-16 | 29.07052 |
| PAFAH1B3 | 0.541601 | 1.455587 | 7.096456 | 10.78929 | 2.33E-17 | 6.64E-16 | 29.05982 |
| PIPOX | -1.21756 | 0.430009 | 10.09835 | -10.7797 | 2.44E-17 | 6.91E-16 | 29.01744 |
| RAC2 | 1.366785 | 2.578953 | 6.853544 | 10.76909 | 2.55E-17 | 7.22E-16 | 28.97034 |
| LGALS1 | 1.374644 | 2.59304 | 8.661392 | 10.76066 | 2.65E-17 | 7.48E-16 | 28.933 |
| CLPP | -0.38717 | 0.764626 | 8.365443 | -10.7537 | 2.74E-17 | 7.69E-16 | 28.90198 |
| IL10RA | 1.301924 | 2.465574 | 5.749494 | 10.75126 | 2.77E-17 | 7.75E-16 | 28.89134 |
| TUBG1 | -0.73746 | 0.599793 | 7.966076 | -10.7478 | 2.81E-17 | 7.84E-16 | 28.87592 |
| ALAD | -0.82747 | 0.563516 | 7.43 | -10.7463 | 2.83E-17 | 7.87E-16 | 28.86957 |
| ASPA | -1.20985 | 0.432314 | 9.05962 | -10.7431 | 2.87E-17 | 7.96E-16 | 28.85497 |
| PPP1R16B | -1.19053 | 0.438143 | 8.28443 | -10.742 | 2.88E-17 | 7.97E-16 | 28.85016 |
| NET1 | -0.83293 | 0.561387 | 8.319241 | -10.7345 | 2.98E-17 | 8.21E-16 | 28.81728 |
| TRAC | 1.558874 | 2.946237 | 6.561392 | 10.69616 | 3.54E-17 | 9.72E-16 | 28.64709 |
| GALNT1 | 0.738636 | 1.668598 | 7.513544 | 10.69038 | 3.63E-17 | 9.92E-16 | 28.62142 |
| CIDEB | -0.71502 | 0.609197 | 8.480886 | -10.6902 | 3.64E-17 | 9.92E-16 | 28.62065 |
| HLA-G | 0.523577 | 1.437515 | 8.412405 | 10.68694 | 3.69E-17 | 1.00E-15 | 28.60618 |
| CYBB | 1.242003 | 2.365266 | 5.576203 | 10.68633 | 3.70E-17 | 1.00E-15 | 28.60346 |
| DEK | 0.512688 | 1.426706 | 9.453291 | 10.68621 | 3.70E-17 | 1.00E-15 | 28.60291 |
| PEX5 | -0.42196 | 0.746412 | 6.915696 | -10.6772 | 3.85E-17 | 1.04E-15 | 28.56315 |
| PLAC8 | 1.759901 | 3.386749 | 5.147975 | 10.67347 | 3.92E-17 | 1.05E-15 | 28.54639 |
| APMAP | 0.462095 | 1.377541 | 8.709114 | 10.67184 | 3.95E-17 | 1.06E-15 | 28.53918 |
| CALML3 | -1.44808 | 0.366508 | 6.132278 | -10.6699 | 3.98E-17 | 1.06E-15 | 28.53062 |
| ASS1 | -1.22063 | 0.429095 | 12.1662 | -10.6682 | 4.01E-17 | 1.07E-15 | 28.5229 |
| LXN | 0.702213 | 1.626999 | 7.682025 | 10.65367 | 4.28E-17 | 1.14E-15 | 28.45853 |
| FAM129A | 0.897332 | 1.862618 | 4.971139 | 10.64788 | 4.40E-17 | 1.16E-15 | 28.43282 |
| SS18L1 | -0.60659 | 0.656745 | 6.679873 | -10.6364 | 4.63E-17 | 1.22E-15 | 28.38194 |
| CRIP2 | 0.910007 | 1.879054 | 7.108608 | 10.63293 | 4.70E-17 | 1.23E-15 | 28.36644 |
| RRAS | 0.798004 | 1.738694 | 6.43443 | 10.61935 | 5.00E-17 | 1.31E-15 | 28.30614 |
| GIMAP4 | 0.968992 | 1.957473 | 6.947595 | 10.60518 | 5.33E-17 | 1.39E-15 | 28.24321 |
| TGFBR2 | 0.493123 | 1.407488 | 7.054684 | 10.59928 | 5.47E-17 | 1.42E-15 | 28.21697 |
| FUZ | -0.66447 | 0.630922 | 6.644177 | -10.5835 | 5.87E-17 | 1.52E-15 | 28.1469 |
| SNX5 | -0.464 | 0.724974 | 9.849873 | -10.5816 | 5.92E-17 | 1.53E-15 | 28.1384 |

| | | | | | | | |
|---|---|---|---|---|---|---|---|
| RNASE6 | 1.739407 | 3.338979 | 6.293544 | 10.58076 | 5.94E-17 | 1.53E-15 | 28.13471 |
| TGFBR3 | -0.94233 | 0.520391 | 8.415063 | -10.5792 | 5.99E-17 | 1.54E-15 | 28.12765 |
| GRAMD1B | -0.94408 | 0.519759 | 7.609114 | -10.5687 | 6.27E-17 | 1.60E-15 | 28.08124 |
| RPS19 | 0.486693 | 1.401229 | 12.90722 | 10.56839 | 6.28E-17 | 1.60E-15 | 28.07972 |
| ACO1 | -0.77219 | 0.585529 | 10.03266 | -10.5659 | 6.35E-17 | 1.62E-15 | 28.06875 |
| AMMECR1 | 0.506482 | 1.420582 | 5.764177 | 10.56553 | 6.36E-17 | 1.62E-15 | 28.06698 |
| ISOC2 | -0.75603 | 0.592124 | 8.746582 | -10.5586 | 6.57E-17 | 1.66E-15 | 28.03602 |
| PSME1 | 0.433735 | 1.350726 | 10.13835 | 10.55722 | 6.61E-17 | 1.67E-15 | 28.03007 |
| IRF7 | 0.848452 | 1.800568 | 6.032025 | 10.55449 | 6.69E-17 | 1.68E-15 | 28.01792 |
| QDPR | -0.80044 | 0.574173 | 10.63759 | -10.5541 | 6.70E-17 | 1.68E-15 | 28.0163 |
| CD63 | 0.408986 | 1.327752 | 11.66519 | 10.54681 | 6.92E-17 | 1.73E-15 | 27.98376 |
| IVD | -0.65657 | 0.634383 | 7.96443 | -10.5438 | 7.02E-17 | 1.75E-15 | 27.97036 |
| WDR13 | -0.40244 | 0.756579 | 8.794937 | -10.5331 | 7.36E-17 | 1.83E-15 | 27.92278 |
| RHOA | 0.495105 | 1.409424 | 10.92747 | 10.52875 | 7.51E-17 | 1.86E-15 | 27.90344 |
| RAI2 | 0.697444 | 1.621629 | 6.211772 | 10.52015 | 7.80E-17 | 1.93E-15 | 27.8652 |
| MACROD1 | -0.9239 | 0.527082 | 7.544937 | -10.5178 | 7.89E-17 | 1.94E-15 | 27.85454 |
| ACBD4 | -0.46893 | 0.722499 | 6.667595 | -10.5159 | 7.95E-17 | 1.95E-15 | 27.84624 |
| SYT11 | 0.520132 | 1.434086 | 4.446835 | 10.51427 | 8.01E-17 | 1.96E-15 | 27.83904 |
| ACKR1 | 1.286772 | 2.439816 | 5.833165 | 10.51151 | 8.11E-17 | 1.98E-15 | 27.82675 |
| JAK1 | 0.628465 | 1.545919 | 8.657089 | 10.51146 | 8.11E-17 | 1.98E-15 | 27.82651 |
| DLEU1 | -0.55028 | 0.682886 | 7.47557 | -10.505 | 8.35E-17 | 2.03E-15 | 27.79784 |
| SNTA1 | -0.56477 | 0.676063 | 7.085823 | -10.5002 | 8.53E-17 | 2.07E-15 | 27.7766 |
| ALB | -2.76862 | 0.146744 | 9.295443 | -10.498 | 8.62E-17 | 2.08E-15 | 27.76655 |
| DNAJB9 | -0.5918 | 0.663515 | 9.118228 | -10.4961 | 8.70E-17 | 2.09E-15 | 27.75796 |
| CBX6 | 0.482345 | 1.397013 | 8.159747 | 10.49432 | 8.76E-17 | 2.10E-15 | 27.75027 |
| SH3GL2 | -0.89326 | 0.538396 | 7.080127 | -10.489 | 8.98E-17 | 2.15E-15 | 27.72656 |
| NQO2 | -1.13941 | 0.453944 | 11.00861 | -10.4819 | 9.27E-17 | 2.21E-15 | 27.69478 |
| RPA3 | -0.49194 | 0.711067 | 9.103418 | -10.4806 | 9.32E-17 | 2.22E-15 | 27.68897 |
| CSF1R | 0.956107 | 1.940067 | 6.708734 | 10.4614 | 1.02E-16 | 2.41E-15 | 27.6037 |
| CD24 | 0.614361 | 1.53088 | 10.64924 | 10.45615 | 1.04E-16 | 2.46E-15 | 27.58032 |
| CD52 | 2.075955 | 4.216235 | 6.051519 | 10.45534 | 1.04E-16 | 2.47E-15 | 27.57672 |
| SAMSN1 | 1.232767 | 2.350173 | 4.525823 | 10.44181 | 1.11E-16 | 2.61E-15 | 27.51648 |
| UBE2I | 0.338551 | 1.264486 | 7.646203 | 10.4101 | 1.28E-16 | 3.01E-15 | 27.3752 |
| HAO2 | -1.51744 | 0.349306 | 10.03418 | -10.4084 | 1.29E-16 | 3.02E-15 | 27.36752 |
| RNASET2 | 0.799262 | 1.740211 | 8.284304 | 10.40764 | 1.29E-16 | 3.02E-15 | 27.36423 |
| CYP27B1 | -2.03045 | 0.244778 | 8.196835 | -10.4041 | 1.31E-16 | 3.06E-15 | 27.3486 |
| MLF2 | -0.51447 | 0.700052 | 8.336835 | -10.4035 | 1.32E-16 | 3.06E-15 | 27.34587 |
| TRAM1 | 0.609736 | 1.52598 | 9.98557 | 10.40219 | 1.33E-16 | 3.07E-15 | 27.33992 |
| UBE2E1 | 0.567174 | 1.481618 | 9.429747 | 10.39652 | 1.36E-16 | 3.14E-15 | 27.31467 |
| DAO | -1.33914 | 0.395255 | 9.022785 | -10.391 | 1.40E-16 | 3.21E-15 | 27.28992 |
| PJA1 | 0.520731 | 1.434682 | 8.098608 | 10.37862 | 1.47E-16 | 3.39E-15 | 27.23488 |
| ANXA3 | 1.162418 | 2.238322 | 6.707089 | 10.37823 | 1.48E-16 | 3.39E-15 | 27.23313 |
| TPD52L1 | 0.720652 | 1.647927 | 8.10038 | 10.37366 | 1.51E-16 | 3.45E-15 | 27.21275 |
| MARCKSL1 | 0.915599 | 1.886353 | 7.598987 | 10.36542 | 1.57E-16 | 3.57E-15 | 27.176 |
| LAMC1 | 0.586831 | 1.501944 | 7.967089 | 10.35785 | 1.62E-16 | 3.67E-15 | 27.14225 |
| RAB31 | 1.012708 | 2.017694 | 6.907595 | 10.35777 | 1.62E-16 | 3.67E-15 | 27.1419 |
| ROGDI | -0.59425 | 0.662389 | 7.613291 | -10.3392 | 1.76E-16 | 3.98E-15 | 27.05897 |

| | | | | | | | |
|---|---|---|---|---|---|---|---|
| SLC25A11 | -0.51616 | 0.699231 | 8.537215 | -10.3382 | 1.77E-16 | 3.99E-15 | 27.05455 |
| CD81 | 0.592207 | 1.507551 | 10.26759 | 10.3375 | 1.78E-16 | 3.99E-15 | 27.05147 |
| BAIAP2 | -0.63182 | 0.645363 | 5.756076 | -10.3285 | 1.85E-16 | 4.13E-15 | 27.01153 |
| MOXD1 | 1.435277 | 2.70434 | 6.430633 | 10.32852 | 1.85E-16 | 4.13E-15 | 27.01142 |
| HN1 | 0.841206 | 1.791547 | 7.532911 | 10.3201 | 1.92E-16 | 4.28E-15 | 26.97383 |
| HEXB | 0.667042 | 1.587814 | 10.1619 | 10.31923 | 1.93E-16 | 4.29E-15 | 26.96998 |
| SDCBP | 0.630257 | 1.547841 | 10.48392 | 10.31833 | 1.94E-16 | 4.29E-15 | 26.96597 |
| ETFA | -0.48827 | 0.712881 | 9.895823 | -10.3169 | 1.95E-16 | 4.31E-15 | 26.95958 |
| COL1A2 | 1.63054 | 3.096289 | 6.916329 | 10.30578 | 2.05E-16 | 4.52E-15 | 26.90994 |
| EBP | -0.59877 | 0.660318 | 8.535316 | -10.3048 | 2.06E-16 | 4.53E-15 | 26.90553 |
| HLA-DPB1 | 1.521515 | 2.870924 | 8.33557 | 10.30298 | 2.07E-16 | 4.55E-15 | 26.89743 |
| TACO1 | -0.55392 | 0.681167 | 8.054051 | -10.3015 | 2.09E-16 | 4.57E-15 | 26.89072 |
| ECHDC3 | -0.74582 | 0.596327 | 10.16367 | -10.2976 | 2.12E-16 | 4.64E-15 | 26.87345 |
| ENTPD5 | -0.92969 | 0.524971 | 8.250127 | -10.2954 | 2.15E-16 | 4.68E-15 | 26.86341 |
| PPIC | 0.5861 | 1.501183 | 8.475696 | 10.28322 | 2.27E-16 | 4.93E-15 | 26.80922 |
| SLC2A5 | -1.1961 | 0.436454 | 8.049494 | -10.2828 | 2.27E-16 | 4.93E-15 | 26.80719 |
| KDELR3 | 0.535224 | 1.449167 | 5.766835 | 10.28144 | 2.29E-16 | 4.94E-15 | 26.80126 |
| PDIA6 | 0.490632 | 1.405061 | 9.828861 | 10.27943 | 2.31E-16 | 4.97E-15 | 26.79231 |
| HERC6 | 1.144993 | 2.211451 | 6.48481 | 10.2725 | 2.38E-16 | 5.12E-15 | 26.76136 |
| DIO1 | -1.39682 | 0.379764 | 10.17304 | -10.2686 | 2.42E-16 | 5.18E-15 | 26.74398 |
| NUP85 | 0.513327 | 1.427338 | 6.185949 | 10.2686 | 2.42E-16 | 5.18E-15 | 26.74397 |
| GCH1 | -1.07243 | 0.475517 | 8.993544 | -10.2645 | 2.47E-16 | 5.26E-15 | 26.72572 |
| SAR1B | -0.49974 | 0.707233 | 8.261899 | -10.2644 | 2.47E-16 | 5.26E-15 | 26.72517 |
| ADI1 | -0.83086 | 0.562193 | 11.29228 | -10.2636 | 2.48E-16 | 5.26E-15 | 26.72173 |
| SLCO3A1 | 0.488399 | 1.402887 | 5.103797 | 10.25706 | 2.55E-16 | 5.41E-15 | 26.6924 |
| FCER1G | 1.394466 | 2.628913 | 7.520759 | 10.25021 | 2.63E-16 | 5.56E-15 | 26.66182 |
| SLC22A4 | -1.03062 | 0.4895 | 7.552532 | -10.2459 | 2.68E-16 | 5.66E-15 | 26.64235 |
| BTN2A2 | 0.45004 | 1.366078 | 5.578861 | 10.23557 | 2.81E-16 | 5.90E-15 | 26.59644 |
| LUM | 1.786014 | 3.448609 | 8.636709 | 10.23547 | 2.81E-16 | 5.90E-15 | 26.59596 |
| TXNDC9 | 0.472345 | 1.387363 | 8.79557 | 10.23313 | 2.84E-16 | 5.93E-15 | 26.58553 |
| GJB1 | -0.65092 | 0.636876 | 7.88962 | -10.2331 | 2.84E-16 | 5.93E-15 | 26.5853 |
| MCUR1 | -0.69873 | 0.616115 | 8.56443 | -10.2325 | 2.85E-16 | 5.94E-15 | 26.5825 |
| ABAT | -0.9784 | 0.507543 | 9.641519 | -10.2221 | 2.99E-16 | 6.20E-15 | 26.53638 |
| FXYD5 | 1.019328 | 2.026975 | 6.131013 | 10.2218 | 2.99E-16 | 6.20E-15 | 26.53487 |
| A2M | 1.213755 | 2.319405 | 10.02962 | 10.21302 | 3.11E-16 | 6.43E-15 | 26.49565 |
| LGALS3BP | 0.677484 | 1.599348 | 8.694304 | 10.20961 | 3.16E-16 | 6.52E-15 | 26.48043 |
| BNIP3 | -0.54572 | 0.685047 | 10.04443 | -10.1904 | 3.45E-16 | 7.09E-15 | 26.39469 |
| APOBEC3G | 1.169144 | 2.248782 | 5.906203 | 10.18977 | 3.46E-16 | 7.09E-15 | 26.39173 |
| DHRS11 | -0.60903 | 0.65564 | 6.16038 | -10.1884 | 3.48E-16 | 7.12E-15 | 26.38558 |
| AKR7A3 | -1.09564 | 0.467929 | 9.894937 | -10.1871 | 3.50E-16 | 7.14E-15 | 26.37982 |
| GZMA | 1.722095 | 3.299151 | 5.415443 | 10.17943 | 3.62E-16 | 7.38E-15 | 26.34552 |
| NXN | 0.703946 | 1.628954 | 6.422532 | 10.17624 | 3.67E-16 | 7.45E-15 | 26.33125 |
| RNF34 | 0.44556 | 1.361843 | 7.561772 | 10.17616 | 3.68E-16 | 7.45E-15 | 26.33088 |
| APOM | -1.34974 | 0.392362 | 10.79684 | -10.1756 | 3.69E-16 | 7.45E-15 | 26.32827 |
| CSDC2 | -0.63436 | 0.644226 | 6.932405 | -10.163 | 3.90E-16 | 7.87E-15 | 26.27214 |
| SAMHD1 | 0.770968 | 1.706415 | 6.188354 | 10.15897 | 3.97E-16 | 7.99E-15 | 26.25403 |
| CMC2 | -0.43206 | 0.741205 | 9.324304 | -10.1586 | 3.98E-16 | 7.99E-15 | 26.25243 |

| | | | | | | | |
|---|---|---|---|---|---|---|---|
| MFSD10 | 0.556212 | 1.470404 | 5.817342 | 10.15422 | 4.06E-16 | 8.13E-15 | 26.23276 |
| RENBP | -0.67361 | 0.626936 | 7.372532 | -10.1314 | 4.50E-16 | 8.99E-15 | 26.13086 |
| RXRA | -0.42163 | 0.746582 | 6.739747 | -10.1287 | 4.55E-16 | 9.08E-15 | 26.11881 |
| CYP3A4 | -0.55161 | 0.682256 | 6.612405 | -10.1238 | 4.66E-16 | 9.27E-15 | 26.09658 |
| BHMT | -1.18668 | 0.439313 | 12.15734 | -10.1224 | 4.69E-16 | 9.30E-15 | 26.0904 |
| KLHDC2 | -0.48621 | 0.7139 | 10.02038 | -10.1215 | 4.71E-16 | 9.32E-15 | 26.08644 |
| ALOX5 | 1.030257 | 2.042388 | 4.761013 | 10.12071 | 4.72E-16 | 9.33E-15 | 26.08286 |
| STK10 | 0.459216 | 1.374795 | 5.532911 | 10.11084 | 4.94E-16 | 9.73E-15 | 26.03868 |
| ST5 | 0.578682 | 1.493485 | 6.086076 | 10.08518 | 5.55E-16 | 1.09E-14 | 25.92386 |
| INTS8 | 0.484526 | 1.399126 | 7.566962 | 10.08218 | 5.62E-16 | 1.10E-14 | 25.91043 |
| LHFP | 1.284341 | 2.435708 | 7.093671 | 10.07475 | 5.81E-16 | 1.14E-14 | 25.87715 |
| ISG20 | 1.249822 | 2.378121 | 5.573165 | 10.07305 | 5.86E-16 | 1.14E-14 | 25.86957 |
| CLEC2B | 1.111726 | 2.16104 | 6.615696 | 10.07213 | 5.88E-16 | 1.15E-14 | 25.86543 |
| ALDOB | -1.11611 | 0.461337 | 12.64443 | -10.0661 | 6.05E-16 | 1.18E-14 | 25.83822 |
| BASP1 | 1.105837 | 2.152237 | 7.123671 | 10.05007 | 6.50E-16 | 1.26E-14 | 25.76664 |
| TST | -0.71983 | 0.60717 | 11.03975 | -10.0439 | 6.69E-16 | 1.29E-14 | 25.73892 |
| TRAPPC6A | -0.50791 | 0.70324 | 8.035443 | -10.041 | 6.77E-16 | 1.31E-14 | 25.72602 |
| GLUD1 | -0.71321 | 0.609962 | 10.08316 | -10.0264 | 7.24E-16 | 1.39E-14 | 25.66044 |
| ALDH3A2 | -0.91175 | 0.531539 | 9.813924 | -10.0148 | 7.63E-16 | 1.47E-14 | 25.6086 |
| PTTG1IP | 0.427503 | 1.344904 | 11.01532 | 10.00424 | 8.00E-16 | 1.53E-14 | 25.56134 |
| VWA5A | 0.48444 | 1.399043 | 7.096709 | 10.00408 | 8.01E-16 | 1.53E-14 | 25.56062 |
| EPM2A | -0.42003 | 0.747408 | 5.992152 | -10.003 | 8.04E-16 | 1.53E-14 | 25.55595 |
| ARSF | -0.82026 | 0.566341 | 7.876709 | -10.0016 | 8.10E-16 | 1.54E-14 | 25.54939 |
| CYFIP1 | 0.470593 | 1.385679 | 9.95962 | 10.00103 | 8.12E-16 | 1.54E-14 | 25.54699 |
| SHMT2 | -0.34887 | 0.785201 | 7.362532 | -10.0004 | 8.14E-16 | 1.54E-14 | 25.54402 |
| ZNF207 | 0.365053 | 1.287929 | 8.945316 | 9.998185 | 8.22E-16 | 1.55E-14 | 25.53422 |
| CHST15 | 0.806614 | 1.749101 | 5.589114 | 9.991818 | 8.46E-16 | 1.60E-14 | 25.50568 |
| CD2 | 1.233715 | 2.351719 | 5.757089 | 9.986985 | 8.65E-16 | 1.62E-14 | 25.48402 |
| IQGAP2 | -0.7604 | 0.590335 | 8.463671 | -9.98681 | 8.66E-16 | 1.62E-14 | 25.48326 |
| GIMAP6 | 1.048162 | 2.067894 | 5.472405 | 9.986628 | 8.66E-16 | 1.62E-14 | 25.48243 |
| FOLR1 | -0.80527 | 0.572255 | 10.39101 | -9.98159 | 8.86E-16 | 1.66E-14 | 25.45985 |
| FBN1 | 0.546706 | 1.460747 | 5.019241 | 9.978473 | 8.99E-16 | 1.68E-14 | 25.44587 |
| ALDH4A1 | -0.82845 | 0.563136 | 9.39481 | -9.97603 | 9.09E-16 | 1.69E-14 | 25.43492 |
| RPA1 | 0.466186 | 1.381452 | 7.435823 | 9.976011 | 9.09E-16 | 1.69E-14 | 25.43484 |
| C1RL | 0.476034 | 1.390915 | 4.874937 | 9.975354 | 9.12E-16 | 1.69E-14 | 25.43189 |
| COLEC11 | -1.00328 | 0.498864 | 9.008734 | -9.97311 | 9.21E-16 | 1.70E-14 | 25.42185 |
| CAP1 | 0.405547 | 1.324591 | 10.3181 | 9.968116 | 9.42E-16 | 1.74E-14 | 25.39945 |
| TP53BP1 | 0.365679 | 1.288487 | 6.471013 | 9.960565 | 9.75E-16 | 1.80E-14 | 25.3656 |
| CD3D | 1.46305 | 2.756906 | 5.766582 | 9.952467 | 1.01E-15 | 1.86E-14 | 25.32929 |
| CXADR | 1.048458 | 2.068319 | 7.354051 | 9.948004 | 1.03E-15 | 1.89E-14 | 25.30928 |
| ABCC2 | -0.83543 | 0.560417 | 6.864937 | -9.94659 | 1.04E-15 | 1.90E-14 | 25.30295 |
| ARL2 | -0.3996 | 0.758066 | 9.031772 | -9.94577 | 1.04E-15 | 1.90E-14 | 25.29928 |
| CCND2 | 0.52888 | 1.442809 | 5.13962 | 9.941831 | 1.06E-15 | 1.93E-14 | 25.2816 |
| GNAI3 | 0.41496 | 1.333262 | 7.607468 | 9.940967 | 1.07E-15 | 1.94E-14 | 25.27773 |
| HCP5 | 1.029394 | 2.041167 | 6.11 | 9.922748 | 1.16E-15 | 2.10E-14 | 25.19602 |
| GMFG | 1.234585 | 2.353136 | 6.816582 | 9.917346 | 1.19E-15 | 2.15E-14 | 25.1718 |
| LTB | 1.280804 | 2.429743 | 6.349367 | 9.916132 | 1.19E-15 | 2.15E-14 | 25.16635 |

| | | | | | | | |
|---|---|---|---|---|---|---|---|
| A1CF | -0.91327 | 0.530981 | 8.303291 | -9.91327 | 1.21E-15 | 2.18E-14 | 25.15352 |
| PPP6R2 | -0.3556 | 0.781545 | 6.074937 | -9.91183 | 1.22E-15 | 2.19E-14 | 25.14707 |
| EPHX2 | -0.94447 | 0.519622 | 9.867215 | -9.9041 | 1.26E-15 | 2.26E-14 | 25.11237 |
| ETNPPL | -1.32429 | 0.399346 | 5.537468 | -9.90006 | 1.28E-15 | 2.30E-14 | 25.09427 |
| RPLP2 | 0.647016 | 1.565926 | 11.57962 | 9.888726 | 1.35E-15 | 2.41E-14 | 25.0434 |
| PGM1 | -0.43292 | 0.740762 | 9.833291 | -9.88812 | 1.35E-15 | 2.41E-14 | 25.04069 |
| PRPF38B | 0.425343 | 1.342891 | 6.253544 | 9.88078 | 1.40E-15 | 2.49E-14 | 25.00775 |
| IRF8 | 1.413755 | 2.664297 | 7.163165 | 9.866996 | 1.49E-15 | 2.65E-14 | 24.9459 |
| TLN2 | -0.73818 | 0.599494 | 8.181646 | -9.86599 | 1.50E-15 | 2.65E-14 | 24.94137 |
| PCYT2 | -0.59813 | 0.66061 | 7.207975 | -9.86254 | 1.52E-15 | 2.69E-14 | 24.9259 |
| TDRD7 | 0.493939 | 1.408285 | 6.626329 | 9.859627 | 1.54E-15 | 2.72E-14 | 24.91283 |
| GYPC | 0.775323 | 1.711573 | 7.804304 | 9.855254 | 1.57E-15 | 2.77E-14 | 24.8932 |
| VSIG4 | 1.487352 | 2.803738 | 6.583038 | 9.852869 | 1.59E-15 | 2.79E-14 | 24.8825 |
| FAM3A | -0.34681 | 0.786324 | 6.957089 | -9.84517 | 1.64E-15 | 2.88E-14 | 24.84794 |
| DUSP22 | 0.484038 | 1.398653 | 7.789367 | 9.842493 | 1.66E-15 | 2.91E-14 | 24.83592 |
| TXN2 | -0.32355 | 0.799101 | 8.815063 | -9.83307 | 1.74E-15 | 3.04E-14 | 24.7936 |
| GK | -0.8693 | 0.547414 | 8.764051 | -9.83255 | 1.74E-15 | 3.04E-14 | 24.79128 |
| KIAA0930 | 0.43583 | 1.352689 | 5.925316 | 9.828723 | 1.77E-15 | 3.08E-14 | 24.77411 |
| SLIT2 | -0.83393 | 0.560998 | 8.120127 | -9.82273 | 1.82E-15 | 3.16E-14 | 24.74721 |
| ERF | -0.50331 | 0.705488 | 6.663671 | -9.81494 | 1.89E-15 | 3.27E-14 | 24.71223 |
| ACOX1 | -0.58589 | 0.666239 | 7.945696 | -9.7985 | 2.03E-15 | 3.51E-14 | 24.6384 |
| HNF4A | -0.43831 | 0.737997 | 6.267342 | -9.79638 | 2.05E-15 | 3.54E-14 | 24.62889 |
| ALDH18A1 | 0.593636 | 1.509046 | 7.887975 | 9.794184 | 2.07E-15 | 3.57E-14 | 24.61902 |
| TLR7 | 0.469453 | 1.384585 | 3.932405 | 9.792645 | 2.09E-15 | 3.59E-14 | 24.61211 |
| ABCC1 | 0.417299 | 1.335425 | 5.848228 | 9.789785 | 2.11E-15 | 3.63E-14 | 24.59926 |
| ARHGEF40 | 0.436963 | 1.353752 | 6.464051 | 9.784244 | 2.17E-15 | 3.71E-14 | 24.57437 |
| SRGN | 1.35253 | 2.553595 | 8.420633 | 9.780437 | 2.21E-15 | 3.77E-14 | 24.55727 |
| NDUFS1 | -0.54009 | 0.68773 | 8.564177 | -9.77767 | 2.23E-15 | 3.81E-14 | 24.54485 |
| TSKU | -0.97026 | 0.510415 | 8.164051 | -9.77423 | 2.27E-15 | 3.86E-14 | 24.5294 |
| MANSC1 | 0.646581 | 1.565454 | 5.609873 | 9.773518 | 2.28E-15 | 3.86E-14 | 24.52619 |
| FCHSD2 | 0.737727 | 1.667547 | 6.363165 | 9.769896 | 2.31E-15 | 3.92E-14 | 24.50992 |
| CDH3 | 0.803874 | 1.745782 | 6.701013 | 9.769474 | 2.32E-15 | 3.92E-14 | 24.50803 |
| ZCCHC6 | 0.436028 | 1.352874 | 6.589747 | 9.767087 | 2.34E-15 | 3.96E-14 | 24.49731 |
| MRPL40 | -0.37458 | 0.771327 | 8.912658 | -9.76603 | 2.36E-15 | 3.97E-14 | 24.49254 |
| FGL2 | 1.109644 | 2.157924 | 8.345696 | 9.749488 | 2.54E-15 | 4.27E-14 | 24.41824 |
| RNF186 | -1.21945 | 0.429447 | 8.982785 | -9.73719 | 2.68E-15 | 4.50E-14 | 24.36299 |
| NCK1 | 0.519111 | 1.433072 | 6.332278 | 9.732213 | 2.75E-15 | 4.60E-14 | 24.34062 |
| ARFIP2 | -0.39903 | 0.758371 | 8.208101 | -9.72871 | 2.79E-15 | 4.66E-14 | 24.32488 |
| EIF4G3 | 0.393544 | 1.313616 | 5.595696 | 9.728141 | 2.80E-15 | 4.66E-14 | 24.32232 |
| KDM5B | 0.407339 | 1.326237 | 6.694937 | 9.727495 | 2.81E-15 | 4.67E-14 | 24.31942 |
| TSPO | 0.577148 | 1.491897 | 8.34 | 9.723032 | 2.86E-15 | 4.76E-14 | 24.29936 |
| TPCN1 | -0.50923 | 0.702598 | 8.399241 | -9.71855 | 2.92E-15 | 4.84E-14 | 24.27921 |
| MRPL46 | -0.38675 | 0.764849 | 7.938228 | -9.71724 | 2.94E-15 | 4.86E-14 | 24.27335 |
| CLPTM1 | -0.53977 | 0.687881 | 8.031266 | -9.71534 | 2.97E-15 | 4.90E-14 | 24.26478 |
| SMPDL3A | -0.98624 | 0.504792 | 9.169114 | -9.71337 | 2.99E-15 | 4.93E-14 | 24.25593 |
| CDK18 | -0.51932 | 0.6977 | 6.176329 | -9.70821 | 3.06E-15 | 5.04E-14 | 24.23273 |
| SAE1 | 0.417793 | 1.335883 | 7.954304 | 9.689626 | 3.33E-15 | 5.47E-14 | 24.14921 |

| | | | | | | | |
|---|---|---|---|---|---|---|---|
| LY86 | 1.028175 | 2.039443 | 6.741013 | 9.682249 | 3.45E-15 | 5.65E-14 | 24.11604 |
| TM6SF1 | 0.636522 | 1.554577 | 4.969367 | 9.681554 | 3.46E-15 | 5.65E-14 | 24.11292 |
| MTTP | -1.02714 | 0.490682 | 6.220506 | -9.66826 | 3.67E-15 | 5.99E-14 | 24.05313 |
| CX3CR1 | 1.615072 | 3.06327 | 6.092911 | 9.663109 | 3.76E-15 | 6.12E-14 | 24.02999 |
| ENOSF1 | -0.47646 | 0.718738 | 7.943797 | -9.65603 | 3.88E-15 | 6.31E-14 | 23.99817 |
| IFI27 | 1.711897 | 3.275913 | 8.156835 | 9.654425 | 3.91E-15 | 6.34E-14 | 23.99094 |
| SLC22A7 | -0.84186 | 0.557925 | 7.297468 | -9.65404 | 3.92E-15 | 6.34E-14 | 23.98922 |
| SEL1L3 | 0.939289 | 1.917582 | 6.986709 | 9.645912 | 4.06E-15 | 6.57E-14 | 23.95266 |
| KCTD12 | 0.781686 | 1.719139 | 8.516962 | 9.633763 | 4.30E-15 | 6.93E-14 | 23.89802 |
| PAOX | -0.38675 | 0.764849 | 6.408228 | -9.62513 | 4.47E-15 | 7.19E-14 | 23.85917 |
| ANGPTL3 | -1.42738 | 0.371806 | 7.968101 | -9.62079 | 4.56E-15 | 7.31E-14 | 23.83968 |
| DHX15 | 0.353847 | 1.277964 | 9.449114 | 9.620729 | 4.56E-15 | 7.31E-14 | 23.83939 |
| ADAM10 | 0.610856 | 1.527165 | 7.277342 | 9.617544 | 4.62E-15 | 7.40E-14 | 23.82507 |
| CSTA | 1.603762 | 3.039347 | 5.061013 | 9.608247 | 4.82E-15 | 7.71E-14 | 23.78325 |
| LRRC49 | 0.454163 | 1.369988 | 5.497975 | 9.605436 | 4.89E-15 | 7.79E-14 | 23.7706 |
| KYNU | -0.47572 | 0.719109 | 7.092152 | -9.60379 | 4.92E-15 | 7.84E-14 | 23.76319 |
| GRINA | -0.49926 | 0.707468 | 6.931013 | -9.60168 | 4.97E-15 | 7.89E-14 | 23.75372 |
| PEBP1 | -0.47616 | 0.718889 | 11.54392 | -9.60159 | 4.97E-15 | 7.89E-14 | 23.75329 |
| FUT3 | -0.71637 | 0.608627 | 6.662278 | -9.59736 | 5.07E-15 | 8.02E-14 | 23.73427 |
| ACADSB | -0.85903 | 0.551322 | 8.137468 | -9.59649 | 5.09E-15 | 8.04E-14 | 23.73035 |
| SLC13A2 | -0.84453 | 0.556891 | 7.274177 | -9.59462 | 5.13E-15 | 8.09E-14 | 23.72196 |
| DHRS12 | -0.30517 | 0.809346 | 6.237089 | -9.58948 | 5.25E-15 | 8.27E-14 | 23.69882 |
| ARHGAP15 | 1.198643 | 2.295237 | 6.694177 | 9.588709 | 5.27E-15 | 8.28E-14 | 23.69535 |
| EVI2B | 1.446509 | 2.725477 | 5.010759 | 9.586641 | 5.32E-15 | 8.34E-14 | 23.68605 |
| SLC2A4RG | -0.67929 | 0.624473 | 7.441899 | -9.58651 | 5.32E-15 | 8.34E-14 | 23.68546 |
| GYPA | -0.37476 | 0.771232 | 4.503671 | -9.57774 | 5.54E-15 | 8.66E-14 | 23.64598 |
| PIGR | 0.93137 | 1.907086 | 8.202532 | 9.567899 | 5.79E-15 | 9.04E-14 | 23.60172 |
| IFRD2 | -0.42623 | 0.744203 | 7.981519 | -9.56746 | 5.81E-15 | 9.04E-14 | 23.59975 |
| SLC26A6 | -0.58383 | 0.667188 | 5.781772 | -9.56368 | 5.91E-15 | 9.18E-14 | 23.58271 |
| UQCRC1 | -0.44219 | 0.736015 | 10.57203 | -9.56215 | 5.95E-15 | 9.23E-14 | 23.57585 |
| STRA6 | -0.56478 | 0.67606 | 6.367342 | -9.55742 | 6.08E-15 | 9.41E-14 | 23.55454 |
| PBLD | -0.83089 | 0.562183 | 10.78835 | -9.55551 | 6.13E-15 | 9.48E-14 | 23.54597 |
| RALYL | -1.04416 | 0.484926 | 7.32557 | -9.55009 | 6.28E-15 | 9.69E-14 | 23.52158 |
| TPST1 | 0.528518 | 1.442446 | 6.283165 | 9.548124 | 6.34E-15 | 9.76E-14 | 23.51272 |
| BTD | -0.47109 | 0.721421 | 7.024304 | -9.53938 | 6.60E-15 | 1.01E-13 | 23.47339 |
| ENO3 | -0.51289 | 0.700816 | 5.013797 | -9.53764 | 6.65E-15 | 1.02E-13 | 23.46554 |
| CRIP1 | 0.956383 | 1.940439 | 6.784937 | 9.533489 | 6.78E-15 | 1.04E-13 | 23.44685 |
| ZDHHC13 | 0.566943 | 1.481382 | 6.092911 | 9.53196 | 6.82E-15 | 1.04E-13 | 23.43997 |
| SLC6A13 | -0.83011 | 0.562488 | 8.817595 | -9.53111 | 6.85E-15 | 1.05E-13 | 23.43613 |
| CYP3A7 | -1.34526 | 0.393582 | 6.863924 | -9.52998 | 6.89E-15 | 1.05E-13 | 23.43107 |
| TTC38 | -0.71121 | 0.61081 | 9.311392 | -9.52728 | 6.97E-15 | 1.06E-13 | 23.41889 |
| HMGCL | -0.47874 | 0.717603 | 9.769367 | -9.5244 | 7.06E-15 | 1.07E-13 | 23.40594 |
| BCL2L2 | -0.3705 | 0.773514 | 7.210886 | -9.5182 | 7.26E-15 | 1.10E-13 | 23.37804 |
| CALM1 | -0.51269 | 0.700912 | 10.11823 | -9.51651 | 7.32E-15 | 1.11E-13 | 23.37044 |
| MX2 | 0.905758 | 1.873528 | 5.928354 | 9.510742 | 7.52E-15 | 1.13E-13 | 23.34446 |
| CRAT | -0.43172 | 0.741378 | 7.646835 | -9.51043 | 7.53E-15 | 1.13E-13 | 23.34306 |
| NMRK1 | -0.66119 | 0.632355 | 8.189241 | -9.50259 | 7.80E-15 | 1.17E-13 | 23.30777 |

| | | | | | | | |
|---|---|---|---|---|---|---|---|
| MACF1 | 0.530823 | 1.444754 | 7.871519 | 9.49779 | 7.97E-15 | 1.20E-13 | 23.28615 |
| MAP4K4 | 0.498841 | 1.413077 | 5.416203 | 9.487893 | 8.34E-15 | 1.25E-13 | 23.24159 |
| HK1 | 0.696126 | 1.620149 | 7.94557 | 9.486394 | 8.40E-15 | 1.26E-13 | 23.23484 |
| SH3BGRL3 | 0.628808 | 1.546286 | 8.278101 | 9.482363 | 8.55E-15 | 1.28E-13 | 23.21669 |
| ACAT1 | -0.62003 | 0.650659 | 11.08709 | -9.48088 | 8.61E-15 | 1.28E-13 | 23.21002 |
| USH1C | -0.66742 | 0.629633 | 8.814684 | -9.47755 | 8.74E-15 | 1.30E-13 | 23.19504 |
| PTPRE | 0.699585 | 1.624038 | 5.278101 | 9.471186 | 9.00E-15 | 1.34E-13 | 23.16637 |
| CSF2RB | 1.218953 | 2.327777 | 5.171139 | 9.467608 | 9.15E-15 | 1.36E-13 | 23.15025 |
| FAAH | -0.55217 | 0.681992 | 6.391519 | -9.46538 | 9.24E-15 | 1.37E-13 | 23.14023 |
| ANXA4 | 0.657839 | 1.577718 | 10.71392 | 9.458082 | 9.55E-15 | 1.41E-13 | 23.10736 |
| EPB41L2 | 0.602075 | 1.517898 | 6.232152 | 9.457664 | 9.57E-15 | 1.41E-13 | 23.10547 |
| DPEP1 | -1.52493 | 0.347495 | 10.11278 | -9.45248 | 9.80E-15 | 1.44E-13 | 23.08215 |
| CCL5 | 1.439256 | 2.711809 | 5.453165 | 9.449268 | 9.94E-15 | 1.46E-13 | 23.06766 |
| ILVBL | -0.50997 | 0.702238 | 8.179367 | -9.44663 | 1.01E-14 | 1.48E-13 | 23.05579 |
| PRKCH | 0.442925 | 1.359357 | 5.819367 | 9.444969 | 1.01E-14 | 1.49E-13 | 23.0483 |
| SAYSD1 | 0.355929 | 1.279809 | 6.019114 | 9.437442 | 1.05E-14 | 1.53E-13 | 23.0144 |
| ACADS | -0.37496 | 0.771127 | 7.039241 | -9.42899 | 1.09E-14 | 1.59E-13 | 22.97632 |
| GRB10 | -0.49493 | 0.709597 | 7.821519 | -9.41993 | 1.14E-14 | 1.66E-13 | 22.93551 |
| MYL6 | 0.411001 | 1.329608 | 12.19038 | 9.412776 | 1.17E-14 | 1.71E-13 | 22.9033 |
| SMC4 | 0.500889 | 1.415086 | 5.633797 | 9.408398 | 1.20E-14 | 1.74E-13 | 22.88358 |
| C1orf115 | -0.58544 | 0.666445 | 9.012405 | -9.40403 | 1.22E-14 | 1.77E-13 | 22.86389 |
| FHL1 | 1.070837 | 2.100651 | 6.524051 | 9.397163 | 1.26E-14 | 1.82E-13 | 22.83297 |
| CSNK1A1 | 0.304427 | 1.234928 | 9.034557 | 9.388434 | 1.31E-14 | 1.90E-13 | 22.79364 |
| SLC19A2 | -0.92861 | 0.525364 | 6.821013 | -9.38522 | 1.33E-14 | 1.92E-13 | 22.77916 |
| NSMAF | 0.452233 | 1.368156 | 5.943038 | 9.383656 | 1.34E-14 | 1.93E-13 | 22.77212 |
| NAGLU | -0.54832 | 0.683816 | 7.432911 | -9.38213 | 1.35E-14 | 1.94E-13 | 22.76523 |
| IRF9 | 0.83386 | 1.782448 | 9.016582 | 9.38121 | 1.36E-14 | 1.95E-13 | 22.76109 |
| FPR3 | 0.999552 | 1.999379 | 5.161013 | 9.377365 | 1.38E-14 | 1.98E-13 | 22.74377 |
| SLC25A28 | -0.28939 | 0.818246 | 6.919114 | -9.37567 | 1.39E-14 | 1.99E-13 | 22.73615 |
| GSS | -0.376 | 0.77057 | 8.759241 | -9.37547 | 1.39E-14 | 1.99E-13 | 22.73524 |
| SLC22A11 | -0.82415 | 0.564815 | 8.53443 | -9.3716 | 1.42E-14 | 2.02E-13 | 22.7178 |
| TNFRSF1B | 0.596733 | 1.512288 | 5.164051 | 9.367945 | 1.44E-14 | 2.05E-13 | 22.70133 |
| DPP4 | -0.96677 | 0.51165 | 9.110506 | -9.36305 | 1.47E-14 | 2.09E-13 | 22.67928 |
| PMM1 | -0.45066 | 0.731709 | 7.983924 | -9.35753 | 1.51E-14 | 2.14E-13 | 22.65439 |
| MS4A4A | 1.56973 | 2.968491 | 5.488101 | 9.354155 | 1.53E-14 | 2.17E-13 | 22.63919 |
| HSPA9 | -0.42574 | 0.744458 | 8.567595 | -9.35156 | 1.55E-14 | 2.19E-13 | 22.6275 |
| CD163 | 1.284486 | 2.435953 | 5.530253 | 9.346301 | 1.59E-14 | 2.24E-13 | 22.6038 |
| AP1S2 | 0.751765 | 1.683852 | 6.716203 | 9.342795 | 1.61E-14 | 2.27E-13 | 22.588 |
| CLCN6 | -0.40665 | 0.754371 | 6.677089 | -9.33414 | 1.68E-14 | 2.36E-13 | 22.54899 |
| LST1 | 1.064631 | 2.091635 | 6.624937 | 9.331374 | 1.70E-14 | 2.39E-13 | 22.53653 |
| BCKDHA | -0.34181 | 0.78905 | 8.745696 | -9.32758 | 1.73E-14 | 2.43E-13 | 22.51944 |
| BZW2 | 0.417332 | 1.335456 | 8.550633 | 9.327006 | 1.73E-14 | 2.43E-13 | 22.51685 |
| RNF219 | 0.481094 | 1.395801 | 5.488354 | 9.32269 | 1.77E-14 | 2.47E-13 | 22.4974 |
| KIAA0355 | 0.402951 | 1.32221 | 7.016582 | 9.317687 | 1.81E-14 | 2.53E-13 | 22.47485 |
| CARD8 | 0.574816 | 1.489487 | 6.257722 | 9.315516 | 1.83E-14 | 2.54E-13 | 22.46507 |
| PPT1 | 0.443347 | 1.359755 | 9.892152 | 9.315516 | 1.83E-14 | 2.54E-13 | 22.46506 |
| PAAF1 | -0.53482 | 0.690247 | 8.078987 | -9.30891 | 1.88E-14 | 2.62E-13 | 22.43527 |

| | | | | | | | |
|---|---|---|---|---|---|---|---|
| ATP6V1B2 | -0.55208 | 0.682038 | 10.31873 | -9.29296 | 2.03E-14 | 2.81E-13 | 22.36342 |
| SMARCE1 | 0.474578 | 1.389512 | 8.580633 | 9.282937 | 2.12E-14 | 2.93E-13 | 22.31822 |
| SLC27A3 | 0.572688 | 1.487292 | 5.838354 | 9.279307 | 2.16E-14 | 2.98E-13 | 22.30186 |
| CCL19 | 2.225191 | 4.675728 | 5.82038 | 9.277805 | 2.17E-14 | 2.99E-13 | 22.29509 |
| DECR2 | -0.78038 | 0.582215 | 8.830759 | -9.2676 | 2.27E-14 | 3.13E-13 | 22.24908 |
| TSFM | -0.36995 | 0.773811 | 6.153291 | -9.25891 | 2.37E-14 | 3.25E-13 | 22.20991 |
| SUOX | -0.50248 | 0.705894 | 8.582278 | -9.25877 | 2.37E-14 | 3.25E-13 | 22.20927 |
| GPD1L | -0.45811 | 0.72794 | 8.771899 | -9.25259 | 2.44E-14 | 3.34E-13 | 22.18144 |
| KCNAB2 | -0.34574 | 0.786905 | 6.121013 | -9.24856 | 2.48E-14 | 3.39E-13 | 22.16327 |
| MFAP1 | 0.397523 | 1.317244 | 8.338228 | 9.246972 | 2.50E-14 | 3.41E-13 | 22.15609 |
| LRP5 | -0.42999 | 0.742265 | 5.348861 | -9.24455 | 2.53E-14 | 3.44E-13 | 22.14515 |
| CD320 | -0.34363 | 0.788056 | 7.574937 | -9.24444 | 2.53E-14 | 3.44E-13 | 22.14469 |
| PDZD2 | -0.93439 | 0.523265 | 6.313165 | -9.24062 | 2.57E-14 | 3.50E-13 | 22.12746 |
| GALNT7 | 0.938123 | 1.916033 | 6.97557 | 9.236987 | 2.61E-14 | 3.55E-13 | 22.11107 |
| APOC3 | -1.76806 | 0.293604 | 6.482532 | -9.22679 | 2.74E-14 | 3.71E-13 | 22.06511 |
| IFI44 | 1.48054 | 2.790532 | 5.823671 | 9.224011 | 2.77E-14 | 3.75E-13 | 22.05257 |
| PNPO | -0.40644 | 0.754485 | 8.206962 | -9.21695 | 2.86E-14 | 3.87E-13 | 22.02073 |
| APITD1 | -0.71111 | 0.610851 | 8.328608 | -9.21668 | 2.87E-14 | 3.87E-13 | 22.01952 |
| TRIM15 | -0.36417 | 0.776916 | 6.071139 | -9.21621 | 2.87E-14 | 3.87E-13 | 22.01742 |
| LCP1 | 0.967306 | 1.955186 | 7.426456 | 9.214927 | 2.89E-14 | 3.89E-13 | 22.01161 |
| ASL | -0.62599 | 0.647973 | 9.123291 | -9.2137 | 2.91E-14 | 3.90E-13 | 22.00609 |
| CXCR4 | 1.329928 | 2.5139 | 6.382278 | 9.213359 | 2.91E-14 | 3.90E-13 | 22.00454 |
| TMEM87A | 0.422563 | 1.340306 | 7.784557 | 9.209472 | 2.96E-14 | 3.96E-13 | 21.98702 |
| ARL6IP5 | 0.70448 | 1.629557 | 9.699494 | 9.207485 | 2.99E-14 | 3.99E-13 | 21.97806 |
| AGMAT | -1.0891 | 0.470055 | 11.14658 | -9.20508 | 3.02E-14 | 4.03E-13 | 21.96723 |
| SLC7A7 | -1.03462 | 0.488145 | 11.65456 | -9.20012 | 3.09E-14 | 4.12E-13 | 21.94485 |
| MTHFD2 | 0.964335 | 1.951163 | 5.921519 | 9.198417 | 3.12E-14 | 4.14E-13 | 21.93717 |
| DNAJC16 | -0.51448 | 0.700045 | 7.589873 | -9.19692 | 3.14E-14 | 4.17E-13 | 21.93041 |
| PAK2 | 0.384242 | 1.305174 | 6.960506 | 9.195589 | 3.16E-14 | 4.18E-13 | 21.92442 |
| PROSC | -0.39317 | 0.761455 | 8.937722 | -9.19422 | 3.18E-14 | 4.20E-13 | 21.91823 |
| TUSC3 | 0.719321 | 1.646408 | 6.567215 | 9.189812 | 3.24E-14 | 4.28E-13 | 21.89837 |
| YEATS2 | 0.448663 | 1.364775 | 5.266329 | 9.162814 | 3.67E-14 | 4.84E-13 | 21.77662 |
| MTFR1 | -0.49733 | 0.708419 | 8.39443 | -9.16193 | 3.68E-14 | 4.84E-13 | 21.77262 |
| IFI30 | 0.769005 | 1.704094 | 9.631013 | 9.161862 | 3.68E-14 | 4.84E-13 | 21.77233 |
| DHTKD1 | -0.66225 | 0.631894 | 8.542278 | -9.15839 | 3.74E-14 | 4.91E-13 | 21.75668 |
| CDHR2 | -0.55207 | 0.682042 | 7.687215 | -9.15569 | 3.79E-14 | 4.96E-13 | 21.74449 |
| XPO1 | 0.432213 | 1.349302 | 8.689241 | 9.155462 | 3.79E-14 | 4.96E-13 | 21.74346 |
| ATP9A | 0.70695 | 1.632349 | 6.819873 | 9.146485 | 3.95E-14 | 5.16E-13 | 21.70298 |
| DCP2 | 0.49191 | 1.406306 | 6.284177 | 9.143666 | 4.00E-14 | 5.22E-13 | 21.69027 |
| FCGR2B | 1.203076 | 2.302301 | 5.221899 | 9.142606 | 4.02E-14 | 5.23E-13 | 21.68549 |
| DHRS1 | -0.56713 | 0.674956 | 8.811139 | -9.14248 | 4.02E-14 | 5.23E-13 | 21.68492 |
| ZGPAT | -1.27847 | 0.412232 | 8.292911 | -9.13923 | 4.08E-14 | 5.30E-13 | 21.67026 |
| HCK | 1.042372 | 2.059611 | 4.977595 | 9.129434 | 4.27E-14 | 5.54E-13 | 21.62608 |
| LPXN | 0.678287 | 1.600239 | 6.748987 | 9.124811 | 4.36E-14 | 5.64E-13 | 21.60523 |
| DYRK2 | 0.535856 | 1.449803 | 5.191013 | 9.119828 | 4.46E-14 | 5.77E-13 | 21.58275 |
| OGG1 | -0.45964 | 0.727166 | 6.905823 | -9.11632 | 4.53E-14 | 5.85E-13 | 21.56695 |
| NIT2 | -0.53322 | 0.69101 | 10.66139 | -9.11498 | 4.56E-14 | 5.88E-13 | 21.56091 |

| | | | | | | | |
|---|---|---|---|---|---|---|---|
| GPX7 | 0.688228 | 1.611303 | 5.956835 | 9.112481 | 4.61E-14 | 5.93E-13 | 21.54962 |
| SPRY1 | 0.509578 | 1.423634 | 7.030253 | 9.10992 | 4.67E-14 | 6.00E-13 | 21.53807 |
| IGKC | 2.990586 | 7.947969 | 10.9338 | 9.109574 | 4.67E-14 | 6.00E-13 | 21.53651 |
| ICAM2 | 0.738248 | 1.668148 | 6.573165 | 9.107045 | 4.73E-14 | 6.06E-13 | 21.5251 |
| PTMS | -0.62406 | 0.64884 | 6.238228 | -9.10388 | 4.80E-14 | 6.13E-13 | 21.51082 |
| PMPCB | -0.28619 | 0.820067 | 9.099367 | -9.10059 | 4.87E-14 | 6.22E-13 | 21.49601 |
| YTHDC1 | 0.374638 | 1.296514 | 7.79519 | 9.099388 | 4.90E-14 | 6.24E-13 | 21.49056 |
| PLD1 | -0.53634 | 0.689519 | 6.349873 | -9.08727 | 5.17E-14 | 6.59E-13 | 21.43592 |
| FHIT | -0.56406 | 0.676397 | 7.601772 | -9.08668 | 5.19E-14 | 6.60E-13 | 21.43322 |
| ADIPOR1 | -0.42623 | 0.744203 | 8.251519 | -9.0801 | 5.35E-14 | 6.79E-13 | 21.40359 |
| STAU1 | -0.61202 | 0.654282 | 10.63 | -9.07451 | 5.48E-14 | 6.95E-13 | 21.37835 |
| CRLF3 | 0.807563 | 1.750252 | 6.90038 | 9.073103 | 5.52E-14 | 6.99E-13 | 21.37201 |
| PTEN | 0.445158 | 1.361463 | 6.98443 | 9.06935 | 5.61E-14 | 7.10E-13 | 21.35508 |
| LANCL2 | -0.44482 | 0.734675 | 7.298101 | -9.06659 | 5.69E-14 | 7.17E-13 | 21.34262 |
| EIF2D | -0.27919 | 0.824054 | 8.926203 | -9.06365 | 5.76E-14 | 7.26E-13 | 21.32935 |
| USP2 | -1.15202 | 0.449996 | 6.91443 | -9.06181 | 5.81E-14 | 7.30E-13 | 21.32107 |
| EMX1 | -0.37252 | 0.77243 | 6.227215 | -9.06177 | 5.81E-14 | 7.30E-13 | 21.32087 |
| OSMR | 0.860955 | 1.81624 | 5.268987 | 9.061504 | 5.82E-14 | 7.30E-13 | 21.31968 |
| TMEM50B | 0.396891 | 1.316667 | 8.684051 | 9.053115 | 6.05E-14 | 7.57E-13 | 21.28184 |
| TUBAL3 | -0.7636 | 0.589023 | 7.093418 | -9.0481 | 6.19E-14 | 7.74E-13 | 21.25921 |
| NAGK | 0.428748 | 1.346065 | 8.768228 | 9.042515 | 6.35E-14 | 7.92E-13 | 21.23403 |
| TMEM43 | 0.405 | 1.324089 | 8.284177 | 9.042274 | 6.35E-14 | 7.92E-13 | 21.23294 |
| MNDA | 1.475 | 2.779836 | 4.821139 | 9.040772 | 6.40E-14 | 7.96E-13 | 21.22617 |
| FHL2 | 1.167773 | 2.246647 | 7.452152 | 9.039387 | 6.44E-14 | 8.00E-13 | 21.21992 |
| LAIR1 | 0.6033 | 1.519188 | 6.02962 | 9.037044 | 6.51E-14 | 8.08E-13 | 21.20935 |
| RARS | 0.467727 | 1.382929 | 8.85038 | 9.034957 | 6.57E-14 | 8.14E-13 | 21.19993 |
| PINK1 | -0.32883 | 0.79618 | 9.053291 | -9.02685 | 6.82E-14 | 8.43E-13 | 21.16338 |
| TCIRG1 | 0.662688 | 1.583029 | 5.545949 | 9.026723 | 6.82E-14 | 8.43E-13 | 21.16279 |
| PDZD3 | -0.5515 | 0.682309 | 7.116582 | -9.02602 | 6.84E-14 | 8.44E-13 | 21.1596 |
| ACAD8 | -0.70618 | 0.612941 | 7.692405 | -9.02557 | 6.86E-14 | 8.45E-13 | 21.15759 |
| ST3GAL6 | -0.35831 | 0.780076 | 5.140759 | -9.02398 | 6.91E-14 | 8.50E-13 | 21.15042 |
| COQ6 | -0.39986 | 0.757931 | 7.161013 | -9.02091 | 7.00E-14 | 8.61E-13 | 21.13655 |
| PRKACB | 0.501206 | 1.415396 | 7.610886 | 9.016421 | 7.15E-14 | 8.77E-13 | 21.11632 |
| EPB41L3 | -0.76874 | 0.586932 | 7.836709 | -9.01506 | 7.19E-14 | 8.81E-13 | 21.1102 |
| HIF1A | 0.653221 | 1.572676 | 10.61873 | 9.014728 | 7.20E-14 | 8.81E-13 | 21.10868 |
| MEF2C | 0.501311 | 1.415499 | 5.776582 | 9.013922 | 7.23E-14 | 8.83E-13 | 21.10505 |
| CFD | 0.875362 | 1.834469 | 5.766962 | 9.008983 | 7.39E-14 | 9.02E-13 | 21.08276 |
| UBD | 1.580586 | 2.990914 | 7.61481 | 9.005372 | 7.52E-14 | 9.15E-13 | 21.06648 |
| ITGB3 | 0.559453 | 1.473711 | 5.52 | 9.005275 | 7.52E-14 | 9.15E-13 | 21.06604 |
| SOWAHC | -0.70931 | 0.611611 | 7.245443 | -9.00362 | 7.58E-14 | 9.20E-13 | 21.05859 |
| GUCY1A3 | 0.752852 | 1.685121 | 7.33557 | 8.995615 | 7.86E-14 | 9.53E-13 | 21.02246 |
| EYA2 | -0.59127 | 0.663758 | 5.656709 | -8.99447 | 7.90E-14 | 9.57E-13 | 21.01729 |
| HIST1H2BK | 1.018729 | 2.026133 | 7.199241 | 8.991746 | 8.00E-14 | 9.68E-13 | 21.005 |
| SLC4A4 | -0.81792 | 0.567257 | 9.278987 | -8.98963 | 8.08E-14 | 9.75E-13 | 20.99545 |
| SLC5A12 | -0.98674 | 0.504617 | 7.484557 | -8.98787 | 8.14E-14 | 9.82E-13 | 20.98751 |
| NDUFA2 | -0.27374 | 0.827171 | 9.68 | -8.98669 | 8.19E-14 | 9.86E-13 | 20.9822 |
| THTPA | -0.36478 | 0.776589 | 7.700886 | -8.98528 | 8.24E-14 | 9.91E-13 | 20.97586 |

| | | | | | | | |
|---|---|---|---|---|---|---|---|
| DNAJC10 | 0.477984 | 1.392796 | 5.965316 | 8.982083 | 8.36E-14 | 1.00E-12 | 20.96141 |
| GNPTAB | 0.56303 | 1.477369 | 6.62519 | 8.981876 | 8.37E-14 | 1.00E-12 | 20.96048 |
| SETD3 | -0.32526 | 0.798156 | 9.218481 | -8.97809 | 8.51E-14 | 1.02E-12 | 20.9434 |
| EGF | -1.78625 | 0.289924 | 7.907975 | -8.97764 | 8.53E-14 | 1.02E-12 | 20.94136 |
| LIME1 | -0.78254 | 0.581341 | 7.710506 | -8.97676 | 8.57E-14 | 1.02E-12 | 20.9374 |
| UTP6 | 0.445474 | 1.361762 | 7.641519 | 8.972393 | 8.74E-14 | 1.04E-12 | 20.9177 |
| RNASE1 | 1.132378 | 2.192198 | 9.513671 | 8.968087 | 8.91E-14 | 1.06E-12 | 20.89827 |
| KLF9 | -0.89584 | 0.537435 | 6.884051 | -8.96507 | 9.03E-14 | 1.07E-12 | 20.88465 |
| SLC25A20 | -0.48835 | 0.712838 | 7.89557 | -8.96404 | 9.08E-14 | 1.08E-12 | 20.88003 |
| CSPP1 | -0.44084 | 0.736704 | 6.420633 | -8.96386 | 9.08E-14 | 1.08E-12 | 20.87921 |
| POLR2G | 0.390362 | 1.310723 | 9.429367 | 8.963019 | 9.12E-14 | 1.08E-12 | 20.87541 |
| MAP7 | -0.51163 | 0.701428 | 7.953671 | -8.9629 | 9.12E-14 | 1.08E-12 | 20.87488 |
| SNX4 | -0.45282 | 0.730614 | 8.512152 | -8.96262 | 9.14E-14 | 1.08E-12 | 20.87363 |
| FBXO21 | -0.79121 | 0.577861 | 9.627975 | -8.96222 | 9.15E-14 | 1.08E-12 | 20.8718 |
| PIK3IP1 | 0.455046 | 1.370827 | 7.49443 | 8.956339 | 9.40E-14 | 1.11E-12 | 20.84527 |
| CYC1 | -0.43973 | 0.737273 | 9.893924 | -8.95325 | 9.54E-14 | 1.12E-12 | 20.83136 |
| TUBA1B | 0.701989 | 1.626747 | 11.20367 | 8.952965 | 9.55E-14 | 1.12E-12 | 20.83005 |
| CELF2 | 0.510962 | 1.425 | 5.111266 | 8.951622 | 9.61E-14 | 1.13E-12 | 20.824 |
| YWHAB | 0.30143 | 1.232365 | 10.0157 | 8.939402 | 1.02E-13 | 1.19E-12 | 20.76886 |
| ZNF22 | 0.355316 | 1.279266 | 7.69038 | 8.936465 | 1.03E-13 | 1.20E-12 | 20.75562 |
| HIBCH | -0.81088 | 0.570036 | 9.763671 | -8.93445 | 1.04E-13 | 1.21E-12 | 20.74654 |
| KIAA0101 | 0.606785 | 1.522862 | 5.196076 | 8.933089 | 1.05E-13 | 1.22E-12 | 20.74038 |
| MYO7A | -0.31891 | 0.801677 | 5.464177 | -8.93289 | 1.05E-13 | 1.22E-12 | 20.7395 |
| MMD | 0.798933 | 1.739814 | 7.440253 | 8.932697 | 1.05E-13 | 1.22E-12 | 20.73862 |
| SLC9A3R1 | -0.5204 | 0.697181 | 9.843924 | -8.91358 | 1.14E-13 | 1.33E-12 | 20.65239 |
| ALDH1A3 | 0.738281 | 1.668187 | 6.07557 | 8.908578 | 1.17E-13 | 1.36E-12 | 20.6298 |
| CXCL6 | 2.588775 | 6.015875 | 5.10443 | 8.907646 | 1.17E-13 | 1.36E-12 | 20.6256 |
| MCM2 | 0.429065 | 1.34636 | 6.325316 | 8.905566 | 1.19E-13 | 1.37E-12 | 20.61622 |
| HSBP1 | 0.33359 | 1.260145 | 9.57 | 8.905182 | 1.19E-13 | 1.37E-12 | 20.61448 |
| LRRFIP1 | 0.505072 | 1.419195 | 7.759241 | 8.901006 | 1.21E-13 | 1.39E-12 | 20.59564 |
| SERPINF2 | -1.05325 | 0.481882 | 7.130253 | -8.90098 | 1.21E-13 | 1.39E-12 | 20.59554 |
| SLC34A1 | -1.37003 | 0.386882 | 8.735316 | -8.90037 | 1.21E-13 | 1.40E-12 | 20.59277 |
| DENND5B | -0.30669 | 0.808493 | 4.307975 | -8.89392 | 1.25E-13 | 1.44E-12 | 20.5637 |
| HGD | -0.62042 | 0.650484 | 9.406709 | -8.88758 | 1.29E-13 | 1.48E-12 | 20.53507 |
| PRLR | -0.5165 | 0.699065 | 5.326203 | -8.88473 | 1.30E-13 | 1.49E-12 | 20.52221 |
| PHB | -0.42489 | 0.744893 | 9.673165 | -8.88247 | 1.32E-13 | 1.51E-12 | 20.51203 |
| MCCC2 | -0.49213 | 0.710972 | 8.777468 | -8.87847 | 1.34E-13 | 1.53E-12 | 20.49397 |
| GMPR2 | -0.34053 | 0.789753 | 8.809494 | -8.87532 | 1.36E-13 | 1.55E-12 | 20.47978 |
| BTN3A2 | 1.029532 | 2.041362 | 5.838101 | 8.875121 | 1.36E-13 | 1.55E-12 | 20.47887 |
| VASH1 | 0.40919 | 1.32794 | 4.768101 | 8.874707 | 1.36E-13 | 1.55E-12 | 20.477 |
| SLPI | 1.665086 | 3.171325 | 8.060759 | 8.873359 | 1.37E-13 | 1.56E-12 | 20.47092 |
| ASB8 | -0.31143 | 0.805843 | 6.840127 | -8.87232 | 1.38E-13 | 1.57E-12 | 20.46626 |
| MPC1 | -0.57038 | 0.673438 | 10.53 | -8.8678 | 1.41E-13 | 1.60E-12 | 20.44586 |
| FZD5 | -0.49449 | 0.709814 | 6.889747 | -8.86136 | 1.45E-13 | 1.64E-12 | 20.41681 |
| RFTN1 | 0.950007 | 1.931881 | 7.535316 | 8.854364 | 1.50E-13 | 1.69E-12 | 20.38523 |
| SLC12A3 | -1.01163 | 0.495987 | 7.683291 | -8.85123 | 1.52E-13 | 1.71E-12 | 20.37111 |
| SLA | 0.789967 | 1.729035 | 6.047595 | 8.850394 | 1.52E-13 | 1.72E-12 | 20.36732 |

| | | | | | | | |
|---|---|---|---|---|---|---|---|
| VCAM1 | 1.222345 | 2.333257 | 10.56886 | 8.842 | 1.58E-13 | 1.78E-12 | 20.32945 |
| F11 | -0.6451 | 0.639449 | 6.768354 | -8.83926 | 1.60E-13 | 1.80E-12 | 20.3171 |
| AVPI1 | -0.57696 | 0.670373 | 7.057468 | -8.83673 | 1.62E-13 | 1.82E-12 | 20.3057 |
| DDX50 | 0.547477 | 1.461527 | 8.031519 | 8.83439 | 1.64E-13 | 1.84E-12 | 20.29512 |
| C1GALT1 | -0.41654 | 0.749219 | 6.587089 | -8.83076 | 1.67E-13 | 1.87E-12 | 20.27873 |
| APOL1 | 0.470916 | 1.385989 | 6.51519 | 8.830228 | 1.67E-13 | 1.87E-12 | 20.27634 |
| SPHK2 | -0.44291 | 0.735652 | 7.056076 | -8.82524 | 1.71E-13 | 1.91E-12 | 20.25384 |
| HSDL2 | -0.52223 | 0.696296 | 7.726203 | -8.81957 | 1.75E-13 | 1.96E-12 | 20.22827 |
| BNIP3L | -0.3972 | 0.75933 | 9.357342 | -8.81562 | 1.79E-13 | 1.99E-12 | 20.21046 |
| CYP27A1 | -0.53174 | 0.69172 | 9.07962 | -8.81476 | 1.79E-13 | 2.00E-12 | 20.20657 |
| GGH | -0.94592 | 0.519098 | 10.12291 | -8.81383 | 1.80E-13 | 2.00E-12 | 20.20238 |
| RGS19 | 0.546667 | 1.460707 | 6.328354 | 8.808623 | 1.84E-13 | 2.05E-12 | 20.17888 |
| P3H1 | 0.345277 | 1.270395 | 6.515316 | 8.808174 | 1.85E-13 | 2.05E-12 | 20.17686 |
| SLC25A36 | -0.4585 | 0.727744 | 8.371519 | -8.80657 | 1.86E-13 | 2.06E-12 | 20.1696 |
| DCTPP1 | 0.313123 | 1.242394 | 8.579494 | 8.802541 | 1.90E-13 | 2.10E-12 | 20.15144 |
| CD99 | 0.596785 | 1.512343 | 8.561899 | 8.800409 | 1.92E-13 | 2.11E-12 | 20.14183 |
| NDRG1 | -0.51283 | 0.700845 | 11.12013 | -8.79565 | 1.96E-13 | 2.16E-12 | 20.12036 |
| PRKCZ | -0.63367 | 0.644535 | 8.262911 | -8.79184 | 1.99E-13 | 2.19E-12 | 20.10318 |
| MLX | -0.3085 | 0.807482 | 8.454177 | -8.79077 | 2.00E-13 | 2.20E-12 | 20.09833 |
| SOD1 | -0.2879 | 0.819094 | 12.1143 | -8.79007 | 2.01E-13 | 2.20E-12 | 20.09518 |
| SELP | 0.608314 | 1.524476 | 6.523671 | 8.787233 | 2.03E-13 | 2.23E-12 | 20.08239 |
| CBFB | 0.45836 | 1.373979 | 7.18038 | 8.77772 | 2.12E-13 | 2.33E-12 | 20.03948 |
| PDZK1IP1 | -0.71069 | 0.61103 | 12.61139 | -8.77386 | 2.16E-13 | 2.36E-12 | 20.02208 |
| RBP1 | 0.831113 | 1.779058 | 6.12 | 8.772977 | 2.17E-13 | 2.37E-12 | 20.01809 |
| FOXO4 | -0.39329 | 0.761393 | 5.695063 | -8.77053 | 2.19E-13 | 2.39E-12 | 20.00706 |
| RORA | -0.58922 | 0.664701 | 6.284304 | -8.76841 | 2.22E-13 | 2.41E-12 | 19.99749 |
| CCR2 | 0.955217 | 1.938872 | 4.683797 | 8.768399 | 2.22E-13 | 2.41E-12 | 19.99743 |
| REV3L | 0.503491 | 1.41764 | 5.863797 | 8.758677 | 2.32E-13 | 2.52E-12 | 19.95358 |
| IFT46 | -0.36539 | 0.77626 | 7.632152 | -8.75671 | 2.34E-13 | 2.54E-12 | 19.94469 |
| SLC25A16 | -0.43525 | 0.739565 | 5.471013 | -8.7561 | 2.34E-13 | 2.54E-12 | 19.94197 |
| CLEC4A | 0.663294 | 1.583694 | 4.376203 | 8.755422 | 2.35E-13 | 2.55E-12 | 19.9389 |
| RAB11A | 0.345231 | 1.270354 | 8.755949 | 8.754015 | 2.37E-13 | 2.56E-12 | 19.93255 |
| TESC | 1.053452 | 2.07549 | 6.252658 | 8.751384 | 2.40E-13 | 2.59E-12 | 19.92069 |
| TLR3 | 0.625744 | 1.543007 | 6.30443 | 8.74958 | 2.41E-13 | 2.60E-12 | 19.91255 |
| REEP4 | 0.36691 | 1.289588 | 6.766962 | 8.746797 | 2.45E-13 | 2.63E-12 | 19.89999 |
| PEX7 | -0.44667 | 0.733736 | 7.173418 | -8.74469 | 2.47E-13 | 2.66E-12 | 19.89048 |
| RORC | -0.75657 | 0.591903 | 7.071139 | -8.74354 | 2.48E-13 | 2.67E-12 | 19.88531 |
| RAE1 | 0.360909 | 1.284235 | 7.520759 | 8.740536 | 2.52E-13 | 2.70E-12 | 19.87176 |
| ALDH3B1 | -0.40875 | 0.753277 | 6.620127 | -8.73868 | 2.54E-13 | 2.72E-12 | 19.86339 |
| ANXA11 | 0.412997 | 1.331449 | 7.050127 | 8.734839 | 2.58E-13 | 2.76E-12 | 19.84606 |
| DOCK2 | 0.974223 | 1.964582 | 4.771519 | 8.73338 | 2.60E-13 | 2.78E-12 | 19.83948 |
| CALML4 | -0.66146 | 0.63224 | 8.51 | -8.73081 | 2.63E-13 | 2.81E-12 | 19.82788 |
| HHEX | 0.548419 | 1.462482 | 4.214304 | 8.730025 | 2.64E-13 | 2.81E-12 | 19.82435 |
| ZNF816 | 0.559341 | 1.473596 | 6.855823 | 8.729848 | 2.64E-13 | 2.81E-12 | 19.82355 |
| MGAM | -1.22207 | 0.428668 | 8.527342 | -8.72972 | 2.64E-13 | 2.81E-12 | 19.82298 |
| DDR1 | 0.491528 | 1.405933 | 8.992278 | 8.728496 | 2.66E-13 | 2.82E-12 | 19.81745 |
| AXL | 0.37946 | 1.300855 | 6.503291 | 8.727206 | 2.67E-13 | 2.84E-12 | 19.81163 |

| | | | | | | | |
|---|---|---|---|---|---|---|---|
| C3orf18 | -0.37231 | 0.772543 | 6.828608 | -8.72371 | 2.72E-13 | 2.88E-12 | 19.79588 |
| HSPA12A | -0.49204 | 0.711018 | 7.726203 | -8.72288 | 2.73E-13 | 2.89E-12 | 19.79212 |
| PTH1R | -0.8111 | 0.569947 | 10.20532 | -8.72236 | 2.73E-13 | 2.89E-12 | 19.78976 |
| AGXT | -1.46547 | 0.362116 | 7.082405 | -8.7213 | 2.75E-13 | 2.90E-12 | 19.78497 |
| GATM | -0.79567 | 0.576075 | 12.12785 | -8.72021 | 2.76E-13 | 2.91E-12 | 19.7801 |
| MYL5 | -0.45469 | 0.729667 | 6.213544 | -8.71267 | 2.86E-13 | 3.01E-12 | 19.74609 |
| HNRNPM | 0.439578 | 1.356208 | 9.071013 | 8.710118 | 2.89E-13 | 3.04E-12 | 19.73456 |
| TCF4 | 0.64886 | 1.567929 | 5.574304 | 8.70977 | 2.90E-13 | 3.04E-12 | 19.73299 |
| STK26 | 0.441831 | 1.358327 | 4.281519 | 8.706819 | 2.94E-13 | 3.08E-12 | 19.71968 |
| IFNGR2 | 0.422767 | 1.340496 | 8.377468 | 8.702671 | 2.99E-13 | 3.13E-12 | 19.70097 |
| SLC39A4 | -0.67278 | 0.627297 | 8.321139 | -8.70104 | 3.01E-13 | 3.15E-12 | 19.69361 |
| SLC7A9 | -1.13547 | 0.455187 | 10.67873 | -8.68452 | 3.25E-13 | 3.40E-12 | 19.61912 |
| DERA | -0.42957 | 0.742482 | 9.741646 | -8.68252 | 3.28E-13 | 3.42E-12 | 19.6101 |
| CYP4F12 | -0.54983 | 0.683101 | 6.550759 | -8.67316 | 3.42E-13 | 3.57E-12 | 19.5679 |
| PEX19 | -0.29918 | 0.812712 | 7.806329 | -8.6715 | 3.45E-13 | 3.59E-12 | 19.5604 |
| CCDC106 | -0.33026 | 0.795391 | 5.382911 | -8.66768 | 3.51E-13 | 3.65E-12 | 19.54318 |
| THEMIS2 | 0.769124 | 1.704234 | 5.133671 | 8.666869 | 3.52E-13 | 3.66E-12 | 19.53951 |
| PSTPIP2 | 0.51166 | 1.42569 | 4.750253 | 8.664679 | 3.56E-13 | 3.69E-12 | 19.52964 |
| GPR65 | 0.866647 | 1.82342 | 4.406582 | 8.662622 | 3.59E-13 | 3.72E-12 | 19.52036 |
| LAMTOR5 | -0.45871 | 0.727637 | 11.37013 | -8.65765 | 3.67E-13 | 3.80E-12 | 19.49796 |
| TUBG2 | -0.42416 | 0.745274 | 5.353038 | -8.65192 | 3.77E-13 | 3.90E-12 | 19.47208 |
| SLC25A46 | -0.41091 | 0.752149 | 8.388354 | -8.64838 | 3.83E-13 | 3.96E-12 | 19.45612 |
| COQ4 | -0.4053 | 0.755078 | 7.965696 | -8.64667 | 3.86E-13 | 3.98E-12 | 19.44841 |
| NOTCH2 | 0.315797 | 1.244699 | 6.832785 | 8.646636 | 3.86E-13 | 3.98E-12 | 19.44828 |
| BBS1 | -0.36404 | 0.776987 | 8.040759 | -8.6444 | 3.90E-13 | 4.01E-12 | 19.43817 |
| IL10RB | 0.510455 | 1.424499 | 5.298228 | 8.639957 | 3.98E-13 | 4.09E-12 | 19.41816 |
| GPNMB | 1.098564 | 2.141414 | 7.850633 | 8.634441 | 4.08E-13 | 4.19E-12 | 19.39329 |
| TLR1 | 0.776173 | 1.712581 | 4.528354 | 8.627852 | 4.21E-13 | 4.31E-12 | 19.36358 |
| SACS | 0.660573 | 1.58071 | 4.333544 | 8.626239 | 4.24E-13 | 4.34E-12 | 19.3563 |
| CD93 | 0.639848 | 1.558166 | 7.362278 | 8.620368 | 4.35E-13 | 4.45E-12 | 19.32983 |
| COASY | -0.36099 | 0.778631 | 8.627468 | -8.61933 | 4.38E-13 | 4.47E-12 | 19.32517 |
| FGGY | -0.4447 | 0.734739 | 7.589241 | -8.61767 | 4.41E-13 | 4.50E-12 | 19.31765 |
| HMOX2 | -0.36016 | 0.779079 | 7.706076 | -8.61384 | 4.49E-13 | 4.57E-12 | 19.30041 |
| TRAM2 | 0.384769 | 1.305651 | 6.188987 | 8.609282 | 4.58E-13 | 4.66E-12 | 19.27985 |
| GZMK | 1.381258 | 2.604955 | 6.316329 | 8.607958 | 4.61E-13 | 4.68E-12 | 19.27388 |
| SUCLG2 | -0.54564 | 0.685088 | 9.414684 | -8.60433 | 4.68E-13 | 4.75E-12 | 19.25752 |
| YRDC | -0.39235 | 0.761887 | 7.029367 | -8.60118 | 4.75E-13 | 4.82E-12 | 19.2433 |
| GSTK1 | -0.35925 | 0.77957 | 10.35646 | -8.5951 | 4.89E-13 | 4.95E-12 | 19.21592 |
| ARAP2 | 0.488564 | 1.403048 | 5.335823 | 8.591847 | 4.96E-13 | 5.02E-12 | 19.20124 |
| DUS1L | -0.39221 | 0.76196 | 7.797468 | -8.58981 | 5.01E-13 | 5.06E-12 | 19.19206 |
| ACSL5 | 0.504644 | 1.418773 | 5.707975 | 8.588628 | 5.03E-13 | 5.08E-12 | 19.18673 |
| VTI1B | -0.31658 | 0.802971 | 9.967975 | -8.58819 | 5.04E-13 | 5.08E-12 | 19.18476 |
| SLC39A6 | 0.670125 | 1.591211 | 7.641013 | 8.588074 | 5.05E-13 | 5.08E-12 | 19.18424 |
| PCK1 | -1.23353 | 0.425275 | 11.88038 | -8.58726 | 5.06E-13 | 5.09E-12 | 19.18056 |
| C3AR1 | 0.883544 | 1.844902 | 6.28038 | 8.58721 | 5.07E-13 | 5.09E-12 | 19.18034 |
| ALS2CL | -0.45099 | 0.731542 | 6.719873 | -8.5863 | 5.09E-13 | 5.10E-12 | 19.17623 |
| SYNGR2 | 0.412358 | 1.33086 | 8.867468 | 8.583901 | 5.14E-13 | 5.15E-12 | 19.16542 |

| | | | | | | | |
|---|---|---|---|---|---|---|---|
| HSP90B1 | 0.640145 | 1.558486 | 9.803924 | 8.575287 | 5.35E-13 | 5.35E-12 | 19.12659 |
| UPF2 | 0.394993 | 1.314937 | 6.481519 | 8.570202 | 5.47E-13 | 5.47E-12 | 19.10367 |
| DDX58 | 0.729901 | 1.658525 | 5.387722 | 8.565713 | 5.59E-13 | 5.58E-12 | 19.08343 |
| CRY2 | -0.46134 | 0.726309 | 7.067722 | -8.55645 | 5.83E-13 | 5.81E-12 | 19.04166 |
| FAM13B | 0.557767 | 1.471989 | 7.348861 | 8.553222 | 5.91E-13 | 5.89E-12 | 19.02713 |
| ACO2 | -0.5401 | 0.687724 | 9.997215 | -8.5527 | 5.93E-13 | 5.90E-12 | 19.02479 |
| ACTG1 | 0.501733 | 1.415913 | 12.30937 | 8.54708 | 6.08E-13 | 6.04E-12 | 18.99945 |
| SIRT2 | -0.32405 | 0.798823 | 8.270506 | -8.54647 | 6.10E-13 | 6.05E-12 | 18.9967 |
| COL15A1 | 1.019625 | 2.027391 | 5.502658 | 8.545186 | 6.14E-13 | 6.08E-12 | 18.99091 |
| DBN1 | 0.402227 | 1.321546 | 6.823671 | 8.544973 | 6.14E-13 | 6.08E-12 | 18.98995 |
| TRIM38 | 0.549967 | 1.464052 | 7.727342 | 8.541802 | 6.23E-13 | 6.16E-12 | 18.97566 |
| GNAS | 0.32388 | 1.251692 | 11.21899 | 8.541438 | 6.24E-13 | 6.16E-12 | 18.97401 |
| AGRN | 0.377549 | 1.299133 | 6.853797 | 8.54088 | 6.26E-13 | 6.17E-12 | 18.9715 |
| ISG15 | 1.558235 | 2.944932 | 7.328734 | 8.539402 | 6.30E-13 | 6.21E-12 | 18.96484 |
| PERP | 0.68083 | 1.603062 | 6.422658 | 8.537023 | 6.37E-13 | 6.27E-12 | 18.95412 |
| PIGV | -0.32835 | 0.796449 | 7.540886 | -8.5356 | 6.41E-13 | 6.30E-12 | 18.94771 |
| P4HA2 | -0.85936 | 0.551197 | 9.133418 | -8.52054 | 6.87E-13 | 6.74E-12 | 18.87983 |
| TDP2 | -0.45377 | 0.730133 | 10.27089 | -8.51987 | 6.89E-13 | 6.75E-12 | 18.87682 |
| SLC15A3 | 0.464045 | 1.379404 | 5.899494 | 8.518138 | 6.94E-13 | 6.80E-12 | 18.869 |
| VGLL4 | 0.313426 | 1.242655 | 7.70962 | 8.507777 | 7.28E-13 | 7.12E-12 | 18.82231 |
| SARDH | -0.39999 | 0.757865 | 5.359873 | -8.50749 | 7.29E-13 | 7.12E-12 | 18.821 |
| MUT | -0.4916 | 0.711235 | 8.27443 | -8.50627 | 7.33E-13 | 7.15E-12 | 18.81553 |
| ACOT11 | -0.51708 | 0.698784 | 7.289873 | -8.50164 | 7.48E-13 | 7.30E-12 | 18.79464 |
| GSTM4 | -0.66005 | 0.632858 | 6.484937 | -8.49359 | 7.76E-13 | 7.56E-12 | 18.75837 |
| PTPRN2 | 0.384967 | 1.30583 | 6.173418 | 8.489825 | 7.90E-13 | 7.68E-12 | 18.74142 |
| RSPH14 | -0.46509 | 0.724428 | 6.350506 | -8.48758 | 7.98E-13 | 7.75E-12 | 18.73132 |
| HPRT1 | 0.485059 | 1.399643 | 8.473924 | 8.486377 | 8.02E-13 | 7.79E-12 | 18.72589 |
| EHHADH | -0.82379 | 0.564957 | 8.610886 | -8.48019 | 8.25E-13 | 8.00E-12 | 18.69802 |
| LRPPRC | -0.40724 | 0.754065 | 8.952278 | -8.47911 | 8.29E-13 | 8.02E-12 | 18.69312 |
| ARHGAP29 | 0.651897 | 1.571233 | 9.124051 | 8.478818 | 8.30E-13 | 8.02E-12 | 18.69183 |
| VSIG10 | 0.339756 | 1.265543 | 5.228228 | 8.478647 | 8.31E-13 | 8.02E-12 | 18.69105 |
| FAM57A | 0.456087 | 1.371816 | 5.90443 | 8.47854 | 8.31E-13 | 8.02E-12 | 18.69057 |
| FKBP8 | -0.36675 | 0.775526 | 6.206582 | -8.4766 | 8.39E-13 | 8.09E-12 | 18.68184 |
| ADGRE5 | 0.672569 | 1.593909 | 5.747468 | 8.474309 | 8.48E-13 | 8.16E-12 | 18.67151 |
| NEK11 | 0.368966 | 1.291427 | 6.103038 | 8.472606 | 8.54E-13 | 8.22E-12 | 18.66384 |
| EED | 0.365441 | 1.288276 | 4.645696 | 8.471243 | 8.60E-13 | 8.26E-12 | 18.6577 |
| SLC1A1 | -0.66848 | 0.629167 | 8.730759 | -8.46514 | 8.84E-13 | 8.48E-12 | 18.6302 |
| SULT1C2 | -0.52841 | 0.693317 | 8.682532 | -8.46282 | 8.93E-13 | 8.56E-12 | 18.61973 |
| SLC22A13 | -0.74986 | 0.594661 | 5.79481 | -8.45553 | 9.23E-13 | 8.84E-12 | 18.58693 |
| IFIT3 | 0.957101 | 1.941405 | 7.419367 | 8.455338 | 9.24E-13 | 8.84E-12 | 18.58604 |
| KLRB1 | 1.258393 | 2.392291 | 5.966962 | 8.454947 | 9.26E-13 | 8.84E-12 | 18.58428 |
| MRPL49 | -0.26358 | 0.833016 | 9.977722 | -8.45471 | 9.27E-13 | 8.84E-12 | 18.5832 |
| TLR5 | 0.568037 | 1.482505 | 5.460759 | 8.453783 | 9.31E-13 | 8.87E-12 | 18.57904 |
| SUN2 | -0.37291 | 0.772226 | 8.135316 | -8.45178 | 9.39E-13 | 8.94E-12 | 18.57003 |
| DHPS | -0.28749 | 0.819326 | 7.170127 | -8.45093 | 9.43E-13 | 8.97E-12 | 18.56617 |
| ABHD6 | -0.70325 | 0.614188 | 7.906456 | -8.45007 | 9.47E-13 | 8.99E-12 | 18.56233 |
| TXNRD2 | -0.38723 | 0.764598 | 6.807595 | -8.44639 | 9.63E-13 | 9.13E-12 | 18.54572 |

| | | | | | | | |
|---|---|---|---|---|---|---|---|
| RAB32 | 0.461482 | 1.376956 | 8.07038 | 8.444476 | 9.71E-13 | 9.20E-12 | 18.53711 |
| FBXL5 | -0.43774 | 0.73829 | 9.09519 | -8.44441 | 9.71E-13 | 9.20E-12 | 18.5368 |
| DPY19L1 | 0.430896 | 1.34807 | 5.433038 | 8.444193 | 9.72E-13 | 9.20E-12 | 18.53584 |
| PROZ | -1.04502 | 0.484638 | 7.273038 | -8.44315 | 9.77E-13 | 9.23E-12 | 18.53115 |
| DNAJA3 | -0.2931 | 0.816145 | 9.264304 | -8.44254 | 9.80E-13 | 9.24E-12 | 18.52841 |
| CRTC3 | 0.388248 | 1.308803 | 7.856962 | 8.442459 | 9.80E-13 | 9.24E-12 | 18.52803 |
| PKP2 | -0.45242 | 0.730817 | 5.189494 | -8.43154 | 1.03E-12 | 9.70E-12 | 18.47882 |
| GATB | -0.36974 | 0.773924 | 6.904684 | -8.42886 | 1.04E-12 | 9.80E-12 | 18.46677 |
| HSD17B10 | -0.34482 | 0.787405 | 10.09987 | -8.42867 | 1.04E-12 | 9.80E-12 | 18.46591 |
| POLDIP2 | -0.3358 | 0.792346 | 8.768861 | -8.42694 | 1.05E-12 | 9.87E-12 | 18.45811 |
| CCS | -0.40366 | 0.75594 | 7.635949 | -8.42608 | 1.06E-12 | 9.90E-12 | 18.45426 |
| CALHM2 | 0.393379 | 1.313467 | 5.523671 | 8.423403 | 1.07E-12 | 1.00E-11 | 18.44219 |
| CD83 | -0.71818 | 0.607863 | 7.93 | -8.42213 | 1.08E-12 | 1.01E-11 | 18.43646 |
| CCT6B | -0.56931 | 0.67394 | 5.402405 | -8.42005 | 1.09E-12 | 1.01E-11 | 18.42709 |
| TNFSF10 | 0.955988 | 1.939908 | 9.786076 | 8.412758 | 1.12E-12 | 1.05E-11 | 18.39425 |
| PLOD2 | 0.623762 | 1.540887 | 7.151646 | 8.412292 | 1.12E-12 | 1.05E-11 | 18.39215 |
| PMVK | -0.39034 | 0.762952 | 8.474557 | -8.41128 | 1.13E-12 | 1.05E-11 | 18.38759 |
| ARG2 | -0.75565 | 0.592279 | 8.25 | -8.40916 | 1.14E-12 | 1.06E-11 | 18.37804 |
| PFKP | 0.869987 | 1.827646 | 7.916456 | 8.404903 | 1.16E-12 | 1.08E-11 | 18.35887 |
| RARRES1 | 1.573287 | 2.97582 | 5.957848 | 8.404853 | 1.16E-12 | 1.08E-11 | 18.35865 |
| GLRX5 | -0.35728 | 0.780635 | 10.60228 | -8.40284 | 1.17E-12 | 1.09E-11 | 18.34959 |
| PALM | -0.48866 | 0.712685 | 7.416962 | -8.39638 | 1.21E-12 | 1.12E-11 | 18.32049 |
| MRC1 | 1.368379 | 2.581804 | 6.825949 | 8.389293 | 1.25E-12 | 1.16E-11 | 18.28858 |
| ARL4C | 1.27139 | 2.41394 | 6.35 | 8.38565 | 1.27E-12 | 1.17E-11 | 18.27218 |
| MAPKAP1 | -0.48317 | 0.715405 | 7.440127 | -8.37991 | 1.30E-12 | 1.20E-11 | 18.24635 |
| SUPV3L1 | -0.33497 | 0.792799 | 7.108987 | -8.3794 | 1.31E-12 | 1.20E-11 | 18.24405 |
| MUC1 | 0.773274 | 1.709144 | 8.126709 | 8.376896 | 1.32E-12 | 1.22E-11 | 18.23276 |
| ACAA2 | -0.34991 | 0.784631 | 7.044051 | -8.37346 | 1.34E-12 | 1.23E-11 | 18.2173 |
| LCP2 | 0.786331 | 1.724682 | 4.576076 | 8.369642 | 1.37E-12 | 1.25E-11 | 18.2001 |
| ZNF358 | -0.38738 | 0.764518 | 6.152532 | -8.36954 | 1.37E-12 | 1.25E-11 | 18.19964 |
| IMPDH1 | 0.674717 | 1.596283 | 7.532278 | 8.364418 | 1.40E-12 | 1.28E-11 | 18.17658 |
| ZNF200 | 0.330428 | 1.257387 | 5.219114 | 8.361634 | 1.42E-12 | 1.30E-11 | 18.16405 |
| IL7R | 1.26639 | 2.405589 | 5.572911 | 8.360002 | 1.43E-12 | 1.31E-11 | 18.1567 |
| ADM | -1.1957 | 0.436575 | 9.146835 | -8.35962 | 1.43E-12 | 1.31E-11 | 18.15498 |
| TRIM32 | 0.339071 | 1.264942 | 6.116203 | 8.355571 | 1.46E-12 | 1.33E-11 | 18.13675 |
| AOAH | 0.795949 | 1.736219 | 5.088354 | 8.350233 | 1.49E-12 | 1.36E-11 | 18.11272 |
| GCDH | -0.75402 | 0.59295 | 7.603291 | -8.34672 | 1.52E-12 | 1.38E-11 | 18.09692 |
| ATXN7L1 | -0.37419 | 0.771539 | 4.661519 | -8.34667 | 1.52E-12 | 1.38E-11 | 18.09669 |
| IDH3G | -0.31547 | 0.803587 | 8.862785 | -8.34368 | 1.54E-12 | 1.40E-11 | 18.08323 |
| SURF1 | -0.3947 | 0.760649 | 9.280127 | -8.34227 | 1.55E-12 | 1.41E-11 | 18.07688 |
| TSC22D1 | 0.391482 | 1.31174 | 8.661139 | 8.337664 | 1.58E-12 | 1.43E-11 | 18.05614 |
| CSTB | 0.686726 | 1.609626 | 10.92316 | 8.336113 | 1.59E-12 | 1.44E-11 | 18.04916 |
| IL7 | 0.492497 | 1.406877 | 3.888987 | 8.335048 | 1.60E-12 | 1.45E-11 | 18.04437 |
| STOML2 | -0.31538 | 0.803642 | 8.83 | -8.33396 | 1.61E-12 | 1.45E-11 | 18.03949 |
| P4HA1 | -0.81328 | 0.569086 | 8.058101 | -8.33309 | 1.61E-12 | 1.46E-11 | 18.03557 |
| FRAT2 | 0.48865 | 1.403131 | 5.516076 | 8.329944 | 1.64E-12 | 1.48E-11 | 18.0214 |
| IGHM | 2.435507 | 5.409545 | 6.80519 | 8.328614 | 1.65E-12 | 1.48E-11 | 18.01541 |

| | | | | | | | |
|---|---|---|---|---|---|---|---|
| PLXND1 | 0.423162 | 1.340863 | 5.796329 | 8.328037 | 1.65E-12 | 1.49E-11 | 18.01282 |
| OAT | -0.69288 | 0.618618 | 10.58557 | -8.32106 | 1.70E-12 | 1.53E-11 | 17.98142 |
| AURKAIP1 | -0.28479 | 0.820862 | 9.567342 | -8.31801 | 1.73E-12 | 1.55E-11 | 17.96769 |
| HYI | -0.34615 | 0.786683 | 7.46519 | -8.29963 | 1.88E-12 | 1.69E-11 | 17.88499 |
| MYO1F | 0.666304 | 1.587002 | 6.122025 | 8.298233 | 1.89E-12 | 1.70E-11 | 17.87869 |
| PANX1 | 0.343155 | 1.268528 | 5.30443 | 8.294434 | 1.92E-12 | 1.72E-11 | 17.8616 |
| SWAP70 | 0.497444 | 1.41171 | 6.088228 | 8.29432 | 1.93E-12 | 1.72E-11 | 17.86109 |
| NDUFAF3 | -0.34853 | 0.785383 | 8.975063 | -8.28851 | 1.98E-12 | 1.77E-11 | 17.83497 |
| DEXI | -0.43871 | 0.737795 | 7.948481 | -8.28717 | 1.99E-12 | 1.78E-11 | 17.82891 |
| APOC1 | 1.53691 | 2.901724 | 8.375696 | 8.286707 | 1.99E-12 | 1.78E-11 | 17.82683 |
| SLC35D1 | -0.45614 | 0.728934 | 6.907722 | -8.28594 | 2.00E-12 | 1.78E-11 | 17.82337 |
| IGSF6 | 0.896653 | 1.861742 | 4.127595 | 8.284079 | 2.02E-12 | 1.80E-11 | 17.81501 |
| PPAP2C | 0.672207 | 1.593509 | 4.871013 | 8.278482 | 2.07E-12 | 1.84E-11 | 17.78983 |
| MIR22HG | -0.76074 | 0.590194 | 8.462658 | -8.27562 | 2.10E-12 | 1.86E-11 | 17.77697 |
| MID1 | 0.587563 | 1.502706 | 6.438481 | 8.274914 | 2.10E-12 | 1.87E-11 | 17.77378 |
| ELF2 | 0.385744 | 1.306534 | 6.844177 | 8.274753 | 2.10E-12 | 1.87E-11 | 17.77305 |
| FN1 | 0.616779 | 1.533447 | 6.171772 | 8.274445 | 2.11E-12 | 1.87E-11 | 17.77167 |
| ARHGAP25 | 0.568327 | 1.482803 | 5.603924 | 8.273213 | 2.12E-12 | 1.87E-11 | 17.76613 |
| ACSM3 | -0.8264 | 0.563933 | 8.173924 | -8.26054 | 2.25E-12 | 1.98E-11 | 17.70913 |
| TUBB | 0.515178 | 1.42917 | 9.199114 | 8.259179 | 2.26E-12 | 1.99E-11 | 17.703 |
| TMED3 | 0.449914 | 1.365959 | 7.827722 | 8.254869 | 2.30E-12 | 2.03E-11 | 17.68361 |
| DDX60 | 0.862253 | 1.817875 | 6.679747 | 8.253815 | 2.32E-12 | 2.04E-11 | 17.67887 |
| NSMF | -0.44108 | 0.736583 | 6.735316 | -8.25035 | 2.35E-12 | 2.07E-11 | 17.66329 |
| GABARAPL | -0.58173 | 0.668161 | 8.187215 | -8.24745 | 2.38E-12 | 2.09E-11 | 17.65026 |
| AGBL5 | -0.26945 | 0.829634 | 7.081139 | -8.24709 | 2.39E-12 | 2.10E-11 | 17.64863 |
| CCL21 | 1.029506 | 2.041325 | 6.174177 | 8.246814 | 2.39E-12 | 2.10E-11 | 17.64738 |
| TOLLIP | -0.36346 | 0.777299 | 7.347089 | -8.24591 | 2.40E-12 | 2.10E-11 | 17.64334 |
| LCK | 0.687681 | 1.610693 | 4.902911 | 8.242767 | 2.43E-12 | 2.13E-11 | 17.62919 |
| KRT19 | 1.199453 | 2.296526 | 8.017342 | 8.238942 | 2.48E-12 | 2.17E-11 | 17.61198 |
| KIAA0226L | 0.520863 | 1.434813 | 4.058228 | 8.234942 | 2.52E-12 | 2.20E-11 | 17.59399 |
| COX6A2 | -0.50188 | 0.706187 | 5.904051 | -8.23238 | 2.55E-12 | 2.23E-11 | 17.58247 |
| G0S2 | -1.20002 | 0.435269 | 9.013291 | -8.23038 | 2.58E-12 | 2.25E-11 | 17.57348 |
| PPL | 0.614895 | 1.531446 | 6.666203 | 8.224997 | 2.64E-12 | 2.30E-11 | 17.54927 |
| ITGAM | 0.775547 | 1.711839 | 4.922658 | 8.220919 | 2.69E-12 | 2.34E-11 | 17.53094 |
| ENPP2 | -0.53154 | 0.691815 | 9.464051 | -8.22075 | 2.69E-12 | 2.34E-11 | 17.53018 |
| SLC2A10 | 0.605184 | 1.521173 | 4.89519 | 8.215114 | 2.76E-12 | 2.40E-11 | 17.50484 |
| PARP1 | 0.294592 | 1.226538 | 7.862405 | 8.212808 | 2.79E-12 | 2.42E-11 | 17.49447 |
| PDIA4 | 0.471008 | 1.386077 | 7.533924 | 8.212162 | 2.80E-12 | 2.42E-11 | 17.49157 |
| GHITM | -0.30102 | 0.811678 | 11.33013 | -8.21053 | 2.82E-12 | 2.44E-11 | 17.48421 |
| PPIB | 0.448215 | 1.364351 | 10.22962 | 8.208991 | 2.84E-12 | 2.45E-11 | 17.47731 |
| CYB5R1 | -0.41072 | 0.752249 | 9.334304 | -8.20874 | 2.84E-12 | 2.45E-11 | 17.4762 |
| GSTA3 | -0.56591 | 0.67553 | 7.178608 | -8.20749 | 2.86E-12 | 2.47E-11 | 17.47055 |
| GNB5 | -0.49196 | 0.711057 | 7.767975 | -8.20693 | 2.87E-12 | 2.47E-11 | 17.46805 |
| NIPSNAP1 | -0.53141 | 0.691878 | 9.233671 | -8.20377 | 2.91E-12 | 2.50E-11 | 17.45382 |
| BAZ1A | 0.48749 | 1.402004 | 4.873418 | 8.200454 | 2.95E-12 | 2.54E-11 | 17.43893 |
| CECR1 | 1.103742 | 2.149114 | 7.526709 | 8.197694 | 2.99E-12 | 2.57E-11 | 17.42652 |
| KLHL3 | -0.65321 | 0.635862 | 7.619747 | -8.19515 | 3.02E-12 | 2.60E-11 | 17.41508 |

| | | | | | | | |
|---|---|---|---|---|---|---|---|
| NECAB3 | -0.31992 | 0.801114 | 6.368101 | -8.19392 | 3.04E-12 | 2.61E-11 | 17.40958 |
| SMAD1 | 0.506107 | 1.420212 | 7.240759 | 8.193854 | 3.04E-12 | 2.61E-11 | 17.40926 |
| CUBN | -1.0715 | 0.475826 | 11.17785 | -8.19337 | 3.05E-12 | 2.61E-11 | 17.40709 |
| FAM110B | 0.55419 | 1.468344 | 6.053671 | 8.190842 | 3.08E-12 | 2.64E-11 | 17.39573 |
| MXI1 | -0.3936 | 0.761229 | 9.186456 | -8.18873 | 3.11E-12 | 2.66E-11 | 17.38625 |
| PEX10 | -0.20341 | 0.868498 | 6.281772 | -8.17988 | 3.24E-12 | 2.77E-11 | 17.34644 |
| PEA15 | 0.511166 | 1.425202 | 7.534177 | 8.176588 | 3.29E-12 | 2.80E-11 | 17.33166 |
| ACADM | -0.6484 | 0.637988 | 10.29937 | -8.17606 | 3.30E-12 | 2.81E-11 | 17.32927 |
| TOP2B | 0.417437 | 1.335553 | 7.916329 | 8.174273 | 3.33E-12 | 2.83E-11 | 17.32125 |
| CCND3 | 0.430237 | 1.347455 | 8.334937 | 8.170917 | 3.38E-12 | 2.87E-11 | 17.30617 |
| MX1 | 1.21805 | 2.326321 | 7.239241 | 8.170705 | 3.38E-12 | 2.87E-11 | 17.30522 |
| IGFBP7 | 0.337457 | 1.263528 | 12.04835 | 8.16945 | 3.40E-12 | 2.88E-11 | 17.29958 |
| NFKB1 | 0.369657 | 1.292046 | 7.623544 | 8.168967 | 3.41E-12 | 2.89E-11 | 17.29741 |
| BCKDHB | -0.42779 | 0.743401 | 6.97 | -8.16821 | 3.42E-12 | 2.89E-11 | 17.29403 |
| COL1A1 | 0.743439 | 1.674162 | 5.266203 | 8.165519 | 3.46E-12 | 2.93E-11 | 17.28192 |
| CLPB | -0.43137 | 0.741557 | 5.996329 | -8.16393 | 3.49E-12 | 2.94E-11 | 17.27478 |
| SLC38A10 | -0.34102 | 0.789482 | 7.093418 | -8.16372 | 3.49E-12 | 2.94E-11 | 17.27384 |
| VPS13C | 0.459084 | 1.374669 | 7.633291 | 8.161273 | 3.53E-12 | 2.97E-11 | 17.26283 |
| MSH3 | -0.45257 | 0.73074 | 6.09443 | -8.16105 | 3.53E-12 | 2.97E-11 | 17.26185 |
| TOR4A | 0.430369 | 1.347578 | 6.154557 | 8.15836 | 3.58E-12 | 3.01E-11 | 17.24975 |
| CRELD2 | 0.465474 | 1.380771 | 8.209873 | 8.152894 | 3.67E-12 | 3.08E-11 | 17.22519 |
| CLTB | -0.37245 | 0.772469 | 7.770506 | -8.1499 | 3.72E-12 | 3.12E-11 | 17.21175 |
| MCCC1 | -0.46775 | 0.72309 | 8.665696 | -8.14769 | 3.75E-12 | 3.15E-11 | 17.20182 |
| KL | -1.01449 | 0.495005 | 10.10253 | -8.14618 | 3.78E-12 | 3.17E-11 | 17.19503 |
| NOX4 | -0.96573 | 0.512019 | 10.40051 | -8.14235 | 3.85E-12 | 3.22E-11 | 17.17781 |
| KIF2A | 0.300356 | 1.231448 | 5.413291 | 8.140726 | 3.87E-12 | 3.24E-11 | 17.17052 |
| VAMP2 | -0.28879 | 0.81859 | 6.199367 | -8.13877 | 3.91E-12 | 3.27E-11 | 17.16171 |
| GLS | -0.46094 | 0.726512 | 10.22506 | -8.13658 | 3.95E-12 | 3.29E-11 | 17.15189 |
| NME1 | 0.495184 | 1.409501 | 9.209241 | 8.132767 | 4.02E-12 | 3.35E-11 | 17.13477 |
| IPO13 | -0.33486 | 0.792864 | 7.211646 | -8.13148 | 4.04E-12 | 3.36E-11 | 17.129 |
| HLA-DQB1 | 1.726061 | 3.308232 | 6.771013 | 8.131467 | 4.04E-12 | 3.36E-11 | 17.12892 |
| NECAB2 | -0.808 | 0.571174 | 7.479873 | -8.1308 | 4.05E-12 | 3.37E-11 | 17.12595 |
| FUBP1 | 0.34836 | 1.273112 | 7.62443 | 8.126441 | 4.13E-12 | 3.43E-11 | 17.10635 |
| THEM6 | -0.38956 | 0.763363 | 6.775316 | -8.12463 | 4.17E-12 | 3.46E-11 | 17.09823 |
| CERS2 | -0.3551 | 0.781816 | 10.24949 | -8.12429 | 4.17E-12 | 3.46E-11 | 17.09669 |
| RHOF | 0.407971 | 1.326818 | 6.459114 | 8.123454 | 4.19E-12 | 3.47E-11 | 17.09293 |
| RPS3 | 0.424816 | 1.342401 | 12.22506 | 8.1213 | 4.23E-12 | 3.50E-11 | 17.08326 |
| IGLL3P | 2.995264 | 7.973778 | 6.225316 | 8.116548 | 4.32E-12 | 3.57E-11 | 17.06192 |
| TMEM123 | 0.595481 | 1.510976 | 8.712658 | 8.113379 | 4.39E-12 | 3.62E-11 | 17.04769 |
| BNIP2 | 0.358491 | 1.282085 | 5.668228 | 8.11104 | 4.43E-12 | 3.66E-11 | 17.03718 |
| CUTC | -0.37129 | 0.77309 | 6.623165 | -8.10704 | 4.52E-12 | 3.72E-11 | 17.0192 |
| BTN3A3 | 0.617668 | 1.534393 | 4.776709 | 8.098656 | 4.69E-12 | 3.86E-11 | 16.98157 |
| SNX27 | -0.30689 | 0.808382 | 7.063544 | -8.09842 | 4.70E-12 | 3.86E-11 | 16.98052 |
| MAPK13 | 0.410296 | 1.328959 | 6.102911 | 8.09731 | 4.72E-12 | 3.88E-11 | 16.97553 |
| HNRNPR | 0.37081 | 1.293079 | 8.387722 | 8.097107 | 4.72E-12 | 3.88E-11 | 16.97461 |
| GPHN | -0.54181 | 0.686911 | 7.420633 | -8.09597 | 4.75E-12 | 3.89E-11 | 16.96953 |
| SLC39A14 | -0.58115 | 0.668432 | 8.002025 | -8.08241 | 5.05E-12 | 4.14E-11 | 16.90862 |

| | | | | | | | |
|---|---|---|---|---|---|---|---|
| ST7 | -0.34866 | 0.785315 | 6.763924 | -8.08067 | 5.09E-12 | 4.17E-11 | 16.9008 |
| AGPAT3 | -0.39819 | 0.758811 | 7.21519 | -8.07968 | 5.11E-12 | 4.18E-11 | 16.89637 |
| ABHD14A | -0.78701 | 0.579544 | 8.21038 | -8.07866 | 5.14E-12 | 4.20E-11 | 16.89178 |
| VPS35 | 0.350771 | 1.275242 | 10.01848 | 8.078025 | 5.15E-12 | 4.20E-11 | 16.88894 |
| TYRO3 | -0.38094 | 0.767936 | 5.738481 | -8.07615 | 5.20E-12 | 4.24E-11 | 16.88053 |
| XPO7 | -0.37599 | 0.770577 | 7.676203 | -8.07482 | 5.23E-12 | 4.26E-11 | 16.87455 |
| SELL | 1.080784 | 2.115185 | 5.35038 | 8.074181 | 5.24E-12 | 4.27E-11 | 16.87169 |
| BTF3 | 0.353742 | 1.277871 | 10.09342 | 8.073681 | 5.25E-12 | 4.27E-11 | 16.86944 |
| NAP1L1 | 0.507154 | 1.421244 | 8.329241 | 8.067268 | 5.41E-12 | 4.39E-11 | 16.84066 |
| PDE8A | -0.4214 | 0.746698 | 8.098101 | -8.06652 | 5.43E-12 | 4.40E-11 | 16.83729 |
| MDFIC | 0.279381 | 1.213674 | 4.589747 | 8.066153 | 5.44E-12 | 4.41E-11 | 16.83565 |
| CDK14 | -0.36321 | 0.777434 | 6.929367 | -8.06526 | 5.46E-12 | 4.42E-11 | 16.83165 |
| WDR45 | -0.37281 | 0.772279 | 7.412532 | -8.05898 | 5.62E-12 | 4.55E-11 | 16.80346 |
| ESM1 | -0.90796 | 0.532937 | 5.510506 | -8.04935 | 5.87E-12 | 4.74E-11 | 16.76023 |
| LGALSL | -0.36214 | 0.778009 | 6.523291 | -8.04798 | 5.91E-12 | 4.77E-11 | 16.75409 |
| CD247 | 0.653577 | 1.573064 | 4.716709 | 8.047244 | 5.93E-12 | 4.78E-11 | 16.75079 |
| SPECC1L | 0.314816 | 1.243853 | 6.899114 | 8.046471 | 5.95E-12 | 4.79E-11 | 16.74732 |
| MYL12A | 0.535988 | 1.449935 | 10.73063 | 8.045548 | 5.97E-12 | 4.81E-11 | 16.74318 |
| MAN2B1 | 0.63865 | 1.556871 | 7.188734 | 8.043086 | 6.04E-12 | 4.86E-11 | 16.73213 |
| KCNH6 | -0.5031 | 0.705591 | 5.025063 | -8.04149 | 6.08E-12 | 4.89E-11 | 16.72499 |
| GALT | -0.3603 | 0.779001 | 7.159494 | -8.04005 | 6.12E-12 | 4.92E-11 | 16.71851 |
| PVALB | -1.45486 | 0.364792 | 7.643797 | -8.03963 | 6.13E-12 | 4.92E-11 | 16.71661 |
| TEX264 | -0.40623 | 0.754592 | 7.859873 | -8.03893 | 6.15E-12 | 4.93E-11 | 16.71348 |
| FLNA | 0.520573 | 1.434525 | 6.745063 | 8.038318 | 6.17E-12 | 4.94E-11 | 16.71074 |
| AK2 | -0.38414 | 0.766234 | 7.006709 | -8.03468 | 6.27E-12 | 5.02E-11 | 16.69441 |
| GNAI1 | 0.83753 | 1.786988 | 6.210506 | 8.033972 | 6.29E-12 | 5.03E-11 | 16.69124 |
| GSTP1 | 0.615303 | 1.53188 | 9.102025 | 8.029848 | 6.41E-12 | 5.12E-11 | 16.67274 |
| MFNG | 0.500119 | 1.41433 | 5.151519 | 8.029006 | 6.44E-12 | 5.13E-11 | 16.66896 |
| LRRC19 | -0.70122 | 0.615052 | 8.608608 | -8.02699 | 6.50E-12 | 5.18E-11 | 16.65992 |
| PON2 | 0.78193 | 1.71943 | 8.490759 | 8.024684 | 6.57E-12 | 5.23E-11 | 16.64957 |
| TMEM50A | 0.34809 | 1.272874 | 8.906709 | 8.018279 | 6.76E-12 | 5.38E-11 | 16.62084 |
| ZCCHC24 | 0.595975 | 1.511494 | 6.338734 | 8.015637 | 6.84E-12 | 5.44E-11 | 16.60899 |
| GUSBP11 | 2.808663 | 7.006348 | 8.072152 | 8.014207 | 6.89E-12 | 5.46E-11 | 16.60257 |
| AKAP12 | 1.012602 | 2.017547 | 8.301899 | 8.014176 | 6.89E-12 | 5.46E-11 | 16.60243 |
| SMARCD3 | 0.616818 | 1.533489 | 5.742658 | 8.013541 | 6.91E-12 | 5.47E-11 | 16.59959 |
| ADCY7 | 0.370237 | 1.292565 | 4.319873 | 8.01173 | 6.96E-12 | 5.51E-11 | 16.59146 |
| APOE | -0.90149 | 0.535334 | 11.24734 | -8.00938 | 7.04E-12 | 5.57E-11 | 16.58093 |
| AGAP1 | -0.40621 | 0.754602 | 6.515316 | -8.00771 | 7.09E-12 | 5.60E-11 | 16.57343 |
| CLIC2 | 0.706166 | 1.631463 | 4.530633 | 8.007448 | 7.10E-12 | 5.60E-11 | 16.57226 |
| FAM89B | 0.261673 | 1.198868 | 7.002785 | 8.005636 | 7.16E-12 | 5.65E-11 | 16.56413 |
| SMARCA5 | 0.333142 | 1.259754 | 6.333291 | 7.995917 | 7.48E-12 | 5.90E-11 | 16.52054 |
| PMP22 | 0.716285 | 1.642945 | 7.207468 | 7.992393 | 7.60E-12 | 5.98E-11 | 16.50474 |
| FANCL | 0.468452 | 1.383624 | 5.583291 | 7.992246 | 7.61E-12 | 5.98E-11 | 16.50408 |
| CST6 | 0.640909 | 1.559311 | 4.577722 | 7.988542 | 7.74E-12 | 6.08E-11 | 16.48748 |
| VSTM4 | -0.49061 | 0.711723 | 5.836582 | -7.98778 | 7.76E-12 | 6.09E-11 | 16.48405 |
| POSTN | 1.798241 | 3.477959 | 6.317468 | 7.987461 | 7.78E-12 | 6.10E-11 | 16.48263 |
| C9orf16 | 0.317358 | 1.246047 | 7.772785 | 7.985799 | 7.83E-12 | 6.14E-11 | 16.47518 |

| | | | | | | | |
|---|---|---|---|---|---|---|---|
| MAP3K12 | -0.24993 | 0.840935 | 6.52038 | -7.98179 | 7.98E-12 | 6.25E-11 | 16.4572 |
| RARRES3 | 0.975922 | 1.966898 | 9.19962 | 7.981259 | 8.00E-12 | 6.25E-11 | 16.45482 |
| IFNAR2 | 0.379855 | 1.301211 | 5.652152 | 7.975918 | 8.19E-12 | 6.40E-11 | 16.43088 |
| ERGIC2 | 0.43222 | 1.349308 | 7.797722 | 7.973175 | 8.30E-12 | 6.48E-11 | 16.41858 |
| ATP10D | 0.556416 | 1.470612 | 7.920253 | 7.971488 | 8.36E-12 | 6.52E-11 | 16.41102 |
| TMED9 | 0.299914 | 1.231071 | 9.435063 | 7.96902 | 8.45E-12 | 6.59E-11 | 16.39996 |
| DLG3 | 0.272437 | 1.207847 | 6.160759 | 7.967985 | 8.49E-12 | 6.61E-11 | 16.39532 |
| SRPK2 | 0.285059 | 1.21846 | 6.68038 | 7.966071 | 8.57E-12 | 6.66E-11 | 16.38674 |
| HPN | -0.5164 | 0.699113 | 9.103418 | -7.96573 | 8.58E-12 | 6.67E-11 | 16.38522 |
| TXNIP | 0.647602 | 1.566562 | 10.74443 | 7.963563 | 8.67E-12 | 6.73E-11 | 16.3755 |
| GAMT | -0.48456 | 0.714716 | 7.390633 | -7.96186 | 8.73E-12 | 6.77E-11 | 16.36785 |
| CDK10 | -0.26426 | 0.832628 | 6.742658 | -7.96052 | 8.79E-12 | 6.81E-11 | 16.36187 |
| RASSF2 | 1.007925 | 2.011016 | 5.17038 | 7.960315 | 8.80E-12 | 6.81E-11 | 16.36094 |
| MPI | -0.37282 | 0.772272 | 6.70557 | -7.95633 | 8.96E-12 | 6.93E-11 | 16.3431 |
| STC2 | -0.39436 | 0.760826 | 5.712658 | -7.95348 | 9.07E-12 | 7.01E-11 | 16.3303 |
| COA4 | -0.27504 | 0.826428 | 8.629241 | -7.94546 | 9.41E-12 | 7.26E-11 | 16.29437 |
| AP1AR | -0.5521 | 0.682026 | 5.51481 | -7.94435 | 9.46E-12 | 7.29E-11 | 16.28939 |
| BIN1 | -0.46839 | 0.722769 | 8.483038 | -7.94197 | 9.56E-12 | 7.37E-11 | 16.27874 |
| MECR | -0.33528 | 0.792628 | 6.85038 | -7.94079 | 9.61E-12 | 7.40E-11 | 16.27344 |
| TP53 | 0.367813 | 1.290395 | 5.168861 | 7.937321 | 9.76E-12 | 7.51E-11 | 16.2579 |
| ESRRA | -0.33165 | 0.794625 | 6.343418 | -7.93441 | 9.89E-12 | 7.60E-11 | 16.24483 |
| CEL | -0.63275 | 0.644944 | 5.091772 | -7.93335 | 9.94E-12 | 7.63E-11 | 16.2401 |
| NDUFA10 | -0.34377 | 0.787977 | 7.958354 | -7.93222 | 9.99E-12 | 7.67E-11 | 16.23506 |
| UBE2C | 0.692009 | 1.615532 | 5.734937 | 7.925234 | 1.03E-11 | 7.91E-11 | 16.20374 |
| MCM3 | 0.382181 | 1.30331 | 6.985949 | 7.924615 | 1.03E-11 | 7.92E-11 | 16.20097 |
| MYDGF | 0.321087 | 1.249271 | 8.213038 | 7.923581 | 1.04E-11 | 7.95E-11 | 16.19634 |
| PSMA6 | 0.444328 | 1.36068 | 10.41582 | 7.921141 | 1.05E-11 | 8.03E-11 | 16.18541 |
| ZNF91 | -0.58325 | 0.66746 | 7.706582 | -7.92025 | 1.06E-11 | 8.06E-11 | 16.18142 |
| NSUN5 | 0.410777 | 1.329402 | 5.742025 | 7.92 | 1.06E-11 | 8.06E-11 | 16.1803 |
| NETO2 | 0.361271 | 1.284557 | 4.307215 | 7.91667 | 1.07E-11 | 8.17E-11 | 16.16538 |
| PNMAL1 | 0.775514 | 1.7118 | 6.430253 | 7.91197 | 1.10E-11 | 8.34E-11 | 16.14433 |
| ACOT7 | -0.88891 | 0.540023 | 8.886076 | -7.90891 | 1.11E-11 | 8.45E-11 | 16.13061 |
| UQCRC2 | -0.28239 | 0.822227 | 9.98443 | -7.90704 | 1.12E-11 | 8.52E-11 | 16.12225 |
| CA2 | -0.56293 | 0.676925 | 11.26203 | -7.90319 | 1.14E-11 | 8.66E-11 | 16.10502 |
| PPP4R1 | 0.398979 | 1.318574 | 8.012532 | 7.89994 | 1.16E-11 | 8.78E-11 | 16.09046 |
| ALDH9A1 | -0.41649 | 0.749246 | 11.01494 | -7.89935 | 1.16E-11 | 8.80E-11 | 16.0878 |
| CCNA2 | 0.335599 | 1.261902 | 4.416709 | 7.888772 | 1.22E-11 | 9.22E-11 | 16.04045 |
| LOXL1 | 1.070145 | 2.099644 | 5.823544 | 7.888609 | 1.22E-11 | 9.22E-11 | 16.03972 |
| DCAF8 | -0.25767 | 0.836439 | 6.653671 | -7.88636 | 1.23E-11 | 9.30E-11 | 16.02967 |
| EGOT | -0.78952 | 0.578537 | 6.509114 | -7.88263 | 1.25E-11 | 9.45E-11 | 16.01297 |
| PHF21A | 0.331667 | 1.258466 | 6.023544 | 7.882252 | 1.25E-11 | 9.46E-11 | 16.01126 |
| CLDN3 | 0.779552 | 1.716598 | 6.919114 | 7.880843 | 1.26E-11 | 9.51E-11 | 16.00496 |
| OGDHL | -0.53551 | 0.689916 | 9.938481 | -7.87683 | 1.28E-11 | 9.68E-11 | 15.987 |
| FH | -0.31635 | 0.803099 | 7.54481 | -7.87558 | 1.29E-11 | 9.73E-11 | 15.9814 |
| MCAM | 0.445343 | 1.361637 | 6.081899 | 7.87198 | 1.31E-11 | 9.88E-11 | 15.96528 |
| RAPGEF6 | 0.328353 | 1.255579 | 5.277595 | 7.871725 | 1.31E-11 | 9.88E-11 | 15.96414 |
| DNAJC12 | -0.62395 | 0.648894 | 8.470886 | -7.86651 | 1.35E-11 | 1.01E-10 | 15.94078 |

| | | | | | | | |
|---|---|---|---|---|---|---|---|
| GPX3 | -0.4597 | 0.727139 | 13.53797 | -7.86557 | 1.35E-11 | 1.01E-10 | 15.9366 |
| TPI1 | -0.30119 | 0.811581 | 10.67962 | -7.86495 | 1.36E-11 | 1.02E-10 | 15.93383 |
| SLC27A2 | -0.90702 | 0.533287 | 10.73177 | -7.86479 | 1.36E-11 | 1.02E-10 | 15.9331 |
| SET | 0.303162 | 1.233846 | 10.4062 | 7.864561 | 1.36E-11 | 1.02E-10 | 15.93208 |
| DYNLL1 | 0.340553 | 1.266242 | 11.68443 | 7.863226 | 1.37E-11 | 1.02E-10 | 15.9261 |
| NDUFS8 | -0.36411 | 0.776948 | 8.697468 | -7.86239 | 1.37E-11 | 1.02E-10 | 15.92237 |
| ARHGEF6 | 0.589572 | 1.5048 | 6.61519 | 7.862366 | 1.37E-11 | 1.02E-10 | 15.92225 |
| SGK1 | -0.87777 | 0.544209 | 10.61747 | -7.86216 | 1.37E-11 | 1.02E-10 | 15.92133 |
| EXT1 | 0.419144 | 1.337134 | 7.202911 | 7.860238 | 1.39E-11 | 1.03E-10 | 15.91273 |
| CYFIP2 | -0.71895 | 0.607541 | 9.006203 | -7.85211 | 1.44E-11 | 1.07E-10 | 15.87638 |
| STK16 | -0.40942 | 0.752926 | 6.745063 | -7.85017 | 1.45E-11 | 1.08E-10 | 15.86768 |
| GLIPR1 | 0.720619 | 1.647889 | 6.407975 | 7.848926 | 1.46E-11 | 1.08E-10 | 15.86211 |
| CAMK2N1 | -0.37793 | 0.76954 | 9.684304 | -7.84765 | 1.47E-11 | 1.09E-10 | 15.85639 |
| CD27 | 0.799262 | 1.740211 | 5.224304 | 7.843997 | 1.49E-11 | 1.11E-10 | 15.84006 |
| SNN | 0.326719 | 1.254158 | 5.404304 | 7.842001 | 1.50E-11 | 1.11E-10 | 15.83113 |
| NPLOC4 | -0.2852 | 0.820625 | 8.703038 | -7.83463 | 1.56E-11 | 1.15E-10 | 15.79817 |
| MYOM2 | -0.62146 | 0.650015 | 7.396709 | -7.83443 | 1.56E-11 | 1.15E-10 | 15.79728 |
| ECI1 | -0.36296 | 0.777565 | 9.183165 | -7.82245 | 1.64E-11 | 1.21E-10 | 15.74366 |
| HAX1 | -0.29829 | 0.813217 | 9.049747 | -7.81957 | 1.67E-11 | 1.23E-10 | 15.73082 |
| FNTA | 0.348491 | 1.273229 | 8.114051 | 7.817958 | 1.68E-11 | 1.24E-10 | 15.72359 |
| ISOC1 | -0.47629 | 0.718823 | 8.104304 | -7.8173 | 1.68E-11 | 1.24E-10 | 15.72064 |
| CAPN2 | 0.451667 | 1.367619 | 10.78367 | 7.817161 | 1.68E-11 | 1.24E-10 | 15.72002 |
| PRNP | -0.48493 | 0.71453 | 9.057215 | -7.81352 | 1.71E-11 | 1.26E-10 | 15.70376 |
| MOCS1 | -0.38608 | 0.765206 | 6.423291 | -7.81339 | 1.71E-11 | 1.26E-10 | 15.70316 |
| PHKA2 | -0.27561 | 0.826103 | 6.129873 | -7.80829 | 1.75E-11 | 1.29E-10 | 15.68035 |
| SH3BGR | -0.37976 | 0.768564 | 6.106582 | -7.80639 | 1.77E-11 | 1.30E-10 | 15.67187 |
| HRH1 | 0.352563 | 1.276827 | 4.935316 | 7.802669 | 1.80E-11 | 1.32E-10 | 15.65522 |
| PRKCDBP | 0.529038 | 1.442967 | 5.873165 | 7.800377 | 1.82E-11 | 1.33E-10 | 15.64498 |
| PLTP | 1.364025 | 2.574023 | 7.45 | 7.798922 | 1.83E-11 | 1.34E-10 | 15.63847 |
| SPP1 | 0.481153 | 1.395859 | 12.72468 | 7.797837 | 1.84E-11 | 1.34E-10 | 15.63362 |
| SMAGP | 0.559967 | 1.474236 | 6.741519 | 7.797645 | 1.84E-11 | 1.34E-10 | 15.63276 |
| ACADL | -0.48682 | 0.713597 | 7.151646 | -7.79512 | 1.86E-11 | 1.36E-10 | 15.62149 |
| MGAT4B | 0.439934 | 1.356542 | 7.728987 | 7.793773 | 1.87E-11 | 1.37E-10 | 15.61545 |
| QPRT | -0.71165 | 0.61062 | 9.584684 | -7.79327 | 1.88E-11 | 1.37E-10 | 15.6132 |
| BAG3 | -0.74582 | 0.596327 | 9.253671 | -7.79006 | 1.90E-11 | 1.39E-10 | 15.59886 |
| SLC26A2 | 0.577635 | 1.492401 | 5.357595 | 7.786773 | 1.93E-11 | 1.41E-10 | 15.58417 |
| ENAH | 0.395356 | 1.315267 | 6.177975 | 7.78603 | 1.94E-11 | 1.41E-10 | 15.58084 |
| TFPI2 | 1.341627 | 2.53437 | 5.614557 | 7.781649 | 1.98E-11 | 1.44E-10 | 15.56127 |
| AASS | -0.54627 | 0.684788 | 6.100506 | -7.78022 | 1.99E-11 | 1.44E-10 | 15.55487 |
| MRPS2 | -0.37016 | 0.773698 | 7.271899 | -7.77974 | 2.00E-11 | 1.45E-10 | 15.55275 |
| PDLIM2 | -0.52682 | 0.694084 | 6.834937 | -7.77397 | 2.05E-11 | 1.48E-10 | 15.52695 |
| TP53TG1 | -0.29057 | 0.817577 | 6.701013 | -7.77309 | 2.06E-11 | 1.49E-10 | 15.52303 |
| MAOA | -0.7287 | 0.603446 | 9.665823 | -7.77253 | 2.06E-11 | 1.49E-10 | 15.5205 |
| GLB1L2 | -0.67092 | 0.628105 | 8.652785 | -7.76884 | 2.10E-11 | 1.51E-10 | 15.50401 |
| CDH6 | 1.001298 | 2.0018 | 7.008481 | 7.767263 | 2.11E-11 | 1.52E-10 | 15.49698 |
| JCHAIN | 2.301126 | 4.928424 | 7.421013 | 7.766416 | 2.12E-11 | 1.53E-10 | 15.49319 |
| PIGH | -0.47101 | 0.72146 | 8.266076 | -7.76364 | 2.15E-11 | 1.55E-10 | 15.4808 |

| | | | | | | | |
|---|---|---|---|---|---|---|---|
| C1orf216 | 0.421054 | 1.338905 | 5.282405 | 7.761848 | 2.16E-11 | 1.56E-10 | 15.47278 |
| TPMT | -0.39837 | 0.758714 | 7.837722 | -7.76123 | 2.17E-11 | 1.56E-10 | 15.47003 |
| RMND1 | -0.61201 | 0.654285 | 7.168481 | -7.76008 | 2.18E-11 | 1.57E-10 | 15.46491 |
| MRPS34 | -0.29362 | 0.815854 | 7.772785 | -7.7503 | 2.28E-11 | 1.64E-10 | 15.42122 |
| GAL3ST1 | -0.64429 | 0.639808 | 6.861519 | -7.74756 | 2.31E-11 | 1.66E-10 | 15.40896 |
| SDC3 | 0.273676 | 1.208884 | 5.10519 | 7.746354 | 2.32E-11 | 1.66E-10 | 15.40357 |
| SLC31A1 | -0.39449 | 0.76076 | 8.101519 | -7.73562 | 2.44E-11 | 1.75E-10 | 15.35565 |
| OAS3 | 0.647444 | 1.566391 | 5.490886 | 7.732691 | 2.47E-11 | 1.77E-10 | 15.34256 |
| RITA1 | -0.34194 | 0.788978 | 7.026076 | -7.72847 | 2.52E-11 | 1.80E-10 | 15.3237 |
| GTPBP6 | -0.37551 | 0.770834 | 7.765316 | -7.72545 | 2.55E-11 | 1.82E-10 | 15.31021 |
| IFIT2 | 0.732418 | 1.661421 | 6.467468 | 7.721364 | 2.60E-11 | 1.85E-10 | 15.29199 |
| FMO5 | -0.83647 | 0.560013 | 6.894937 | -7.72133 | 2.60E-11 | 1.85E-10 | 15.29186 |
| WDR82 | 0.327661 | 1.254977 | 8.577089 | 7.714389 | 2.68E-11 | 1.91E-10 | 15.26085 |
| KCNK5 | -0.87073 | 0.54687 | 8.26519 | -7.71334 | 2.69E-11 | 1.92E-10 | 15.25617 |
| MTHFD1 | -0.50949 | 0.702473 | 9.678481 | -7.71153 | 2.72E-11 | 1.93E-10 | 15.24809 |
| NKG7 | 0.633972 | 1.551832 | 4.577215 | 7.711163 | 2.72E-11 | 1.93E-10 | 15.24645 |
| XAF1 | 0.907879 | 1.876285 | 5.419241 | 7.709786 | 2.74E-11 | 1.95E-10 | 15.24031 |
| CYB5R4 | 0.438939 | 1.355607 | 6.748354 | 7.696806 | 2.90E-11 | 2.06E-10 | 15.18238 |
| SLC25A13 | -0.48158 | 0.716192 | 7.184051 | -7.69558 | 2.92E-11 | 2.07E-10 | 15.17689 |
| SLAMF8 | 0.613485 | 1.52995 | 4.821266 | 7.6955 | 2.92E-11 | 2.07E-10 | 15.17655 |
| GLUD2 | -0.35622 | 0.781209 | 7.347722 | -7.69435 | 2.94E-11 | 2.08E-10 | 15.17144 |
| CDC25B | 0.494592 | 1.408922 | 6.645949 | 7.694327 | 2.94E-11 | 2.08E-10 | 15.17132 |
| TRPV1 | -0.32769 | 0.796809 | 5.390506 | -7.68884 | 3.01E-11 | 2.13E-10 | 15.14683 |
| TRAF5 | 0.489407 | 1.403868 | 5.491392 | 7.686367 | 3.04E-11 | 2.15E-10 | 15.1358 |
| ECT2 | 0.448867 | 1.364968 | 4.239241 | 7.683901 | 3.08E-11 | 2.17E-10 | 15.1248 |
| CLEC10A | 0.952866 | 1.935714 | 5.076076 | 7.681898 | 3.11E-11 | 2.19E-10 | 15.11587 |
| TBC1D2B | 0.706891 | 1.632282 | 7.163544 | 7.680435 | 3.13E-11 | 2.20E-10 | 15.10934 |
| CD38 | 0.912213 | 1.881931 | 5.859747 | 7.675495 | 3.20E-11 | 2.25E-10 | 15.0873 |
| PTTG1 | 0.644868 | 1.563596 | 7.57481 | 7.672738 | 3.24E-11 | 2.28E-10 | 15.07501 |
| BTN2A1 | 0.255599 | 1.193832 | 7.003291 | 7.67026 | 3.27E-11 | 2.30E-10 | 15.06396 |
| RAC1 | 0.256028 | 1.194186 | 11.14456 | 7.668965 | 3.29E-11 | 2.31E-10 | 15.05818 |
| MAP2K3 | -0.41859 | 0.748155 | 7.819494 | -7.65943 | 3.44E-11 | 2.41E-10 | 15.01565 |
| SLC28A1 | -0.5041 | 0.705098 | 7.017468 | -7.6587 | 3.45E-11 | 2.42E-10 | 15.01242 |
| SLC17A5 | -0.53843 | 0.688519 | 8.262911 | -7.65762 | 3.47E-11 | 2.43E-10 | 15.00758 |
| IGHV4-28 | 2.193498 | 4.574132 | 4.798228 | 7.654012 | 3.52E-11 | 2.47E-10 | 14.99151 |
| BIRC3 | 0.938103 | 1.916007 | 5.020127 | 7.652669 | 3.55E-11 | 2.48E-10 | 14.98552 |
| PRKCA | -0.23666 | 0.848708 | 5.79962 | -7.64646 | 3.65E-11 | 2.55E-10 | 14.95782 |
| SIGIRR | -0.67877 | 0.624696 | 8.973418 | -7.64423 | 3.68E-11 | 2.57E-10 | 14.94792 |
| FNBP1 | 0.409308 | 1.328049 | 6.420759 | 7.641905 | 3.72E-11 | 2.60E-10 | 14.93754 |
| GCLC | -0.55044 | 0.682811 | 7.912025 | -7.64184 | 3.72E-11 | 2.60E-10 | 14.93725 |
| HTR2B | 1.044097 | 2.062076 | 4.69962 | 7.640329 | 3.75E-11 | 2.61E-10 | 14.93051 |
| GALK2 | -0.40842 | 0.753449 | 6.564177 | -7.63643 | 3.82E-11 | 2.65E-10 | 14.91315 |
| SMTN | -0.33366 | 0.793519 | 6.056709 | -7.63428 | 3.85E-11 | 2.68E-10 | 14.90357 |
| QSOX1 | 0.347516 | 1.272368 | 6.928861 | 7.633876 | 3.86E-11 | 2.68E-10 | 14.90175 |
| CLCN5 | -0.58802 | 0.665254 | 7.917848 | -7.63268 | 3.88E-11 | 2.69E-10 | 14.89644 |
| PRDX3 | -0.28775 | 0.819177 | 11.39089 | -7.63008 | 3.93E-11 | 2.72E-10 | 14.88483 |
| AKR7A2 | -0.49843 | 0.707879 | 9.988101 | -7.62493 | 4.02E-11 | 2.79E-10 | 14.86188 |

| | | | | | | | |
|---|---|---|---|---|---|---|---|
| FYN | -0.39057 | 0.76283 | 6.757722 | -7.62385 | 4.04E-11 | 2.80E-10 | 14.85706 |
| ERAL1 | -0.30343 | 0.810322 | 6.506076 | -7.62269 | 4.06E-11 | 2.81E-10 | 14.85189 |
| DDX10 | -0.65301 | 0.635952 | 8.802658 | -7.61978 | 4.11E-11 | 2.84E-10 | 14.83894 |
| ADCY9 | -0.31303 | 0.804949 | 7.069241 | -7.61882 | 4.13E-11 | 2.85E-10 | 14.83464 |
| MRPL34 | -0.40632 | 0.754543 | 8.911139 | -7.61751 | 4.16E-11 | 2.87E-10 | 14.82885 |
| PSMB7 | -0.22947 | 0.85295 | 10.48089 | -7.61678 | 4.17E-11 | 2.88E-10 | 14.82559 |
| LCN2 | 1.469532 | 2.769321 | 5.941899 | 7.615942 | 4.19E-11 | 2.88E-10 | 14.82184 |
| DNASE1L1 | -0.33327 | 0.793733 | 7.617089 | -7.61002 | 4.30E-11 | 2.96E-10 | 14.79547 |
| CASP6 | 0.38668 | 1.307381 | 6.858481 | 7.609774 | 4.30E-11 | 2.96E-10 | 14.79436 |
| FAM208B | 0.34307 | 1.268453 | 5.994177 | 7.60901 | 4.32E-11 | 2.97E-10 | 14.79096 |
| MPST | -0.39356 | 0.761247 | 8.348861 | -7.60863 | 4.33E-11 | 2.97E-10 | 14.78928 |
| CD14 | 1.030876 | 2.043265 | 7.658228 | 7.60667 | 4.36E-11 | 3.00E-10 | 14.78054 |
| MCM5 | 0.373123 | 1.295153 | 5.244557 | 7.603104 | 4.44E-11 | 3.04E-10 | 14.76466 |
| THBS2 | 1.177437 | 2.261747 | 7.073797 | 7.596353 | 4.57E-11 | 3.13E-10 | 14.73459 |
| MSMO1 | -0.49493 | 0.709594 | 9.023038 | -7.59207 | 4.66E-11 | 3.19E-10 | 14.71553 |
| PRKCD | -0.44213 | 0.736048 | 6.256835 | -7.59126 | 4.68E-11 | 3.20E-10 | 14.7119 |
| MLLT3 | 0.37749 | 1.29908 | 5.607468 | 7.589191 | 4.72E-11 | 3.23E-10 | 14.7027 |
| BOLA1 | -0.31588 | 0.803363 | 6.525443 | -7.58298 | 4.86E-11 | 3.32E-10 | 14.67506 |
| NRP1 | 0.762075 | 1.695928 | 6.988987 | 7.582286 | 4.87E-11 | 3.32E-10 | 14.67196 |
| DTL | 0.326482 | 1.253952 | 3.858987 | 7.579928 | 4.92E-11 | 3.36E-10 | 14.66146 |
| ACSM5 | -0.62055 | 0.650424 | 7.297089 | -7.57955 | 4.93E-11 | 3.36E-10 | 14.6598 |
| SPINT2 | 0.315889 | 1.244779 | 9.901519 | 7.577016 | 4.99E-11 | 3.40E-10 | 14.6485 |
| CDC7 | 0.478788 | 1.393572 | 4.5 | 7.576368 | 5.00E-11 | 3.40E-10 | 14.64561 |
| EXTL1 | -0.2781 | 0.824675 | 4.75557 | -7.57517 | 5.03E-11 | 3.42E-10 | 14.64028 |
| SERPINE2 | 0.96197 | 1.947968 | 9.656835 | 7.571646 | 5.11E-11 | 3.47E-10 | 14.6246 |
| NPR3 | -0.82385 | 0.564933 | 9.104557 | -7.56931 | 5.17E-11 | 3.51E-10 | 14.6142 |
| NUDT6 | -0.53191 | 0.691638 | 6.759114 | -7.56695 | 5.22E-11 | 3.54E-10 | 14.60371 |
| PLK2 | 0.968399 | 1.956668 | 4.934304 | 7.565181 | 5.26E-11 | 3.57E-10 | 14.59582 |
| ACSS3 | -0.42653 | 0.74405 | 7.639873 | -7.56344 | 5.31E-11 | 3.59E-10 | 14.58808 |
| IFT52 | 0.305665 | 1.235989 | 7.888987 | 7.560166 | 5.38E-11 | 3.64E-10 | 14.57351 |
| NCALD | -0.50163 | 0.706306 | 6.797848 | -7.55769 | 5.44E-11 | 3.68E-10 | 14.56251 |
| AHR | 0.504763 | 1.41889 | 6.610633 | 7.556565 | 5.47E-11 | 3.70E-10 | 14.55749 |
| TRIM21 | 0.337022 | 1.263147 | 6.598608 | 7.555901 | 5.49E-11 | 3.70E-10 | 14.55453 |
| RUNX3 | 0.469065 | 1.384212 | 4.702025 | 7.552431 | 5.58E-11 | 3.76E-10 | 14.5391 |
| MAGI2 | -0.35127 | 0.783893 | 5.216962 | -7.55067 | 5.62E-11 | 3.79E-10 | 14.53127 |
| FNDC3B | 0.55139 | 1.465497 | 7.689241 | 7.550391 | 5.63E-11 | 3.79E-10 | 14.53002 |
| DOK5 | 0.512681 | 1.426699 | 6.69481 | 7.55034 | 5.63E-11 | 3.79E-10 | 14.52979 |
| C1GALT1C | 0.41166 | 1.330216 | 7.638481 | 7.548002 | 5.69E-11 | 3.82E-10 | 14.51939 |
| KPTN | -0.31895 | 0.801655 | 6.313291 | -7.54623 | 5.73E-11 | 3.85E-10 | 14.51152 |
| HNF1A | -0.4074 | 0.753982 | 6.408734 | -7.54546 | 5.75E-11 | 3.86E-10 | 14.50807 |
| TMEM254 | -0.26604 | 0.831599 | 5.114304 | -7.54235 | 5.84E-11 | 3.91E-10 | 14.49425 |
| NDUFAB1 | -0.27057 | 0.828994 | 11.18785 | -7.54078 | 5.88E-11 | 3.94E-10 | 14.48729 |
| VIL1 | -0.89566 | 0.537502 | 7.293038 | -7.53939 | 5.91E-11 | 3.96E-10 | 14.48107 |
| SEC61G | 0.369177 | 1.291615 | 9.60443 | 7.538669 | 5.93E-11 | 3.97E-10 | 14.47788 |
| MAP4K2 | -0.34097 | 0.789511 | 5.021266 | -7.53779 | 5.96E-11 | 3.98E-10 | 14.47397 |
| TRMT61B | -0.47899 | 0.717479 | 7.567089 | -7.53608 | 6.00E-11 | 4.01E-10 | 14.46636 |
| GDPD3 | -0.99838 | 0.500562 | 7.678608 | -7.53444 | 6.05E-11 | 4.03E-10 | 14.45905 |

| | | | | | | | |
|---|---|---|---|---|---|---|---|
| CYP2B6 | -0.59178 | 0.663524 | 5.723671 | -7.53442 | 6.05E-11 | 4.03E-10 | 14.45899 |
| INSR | -0.42607 | 0.744288 | 7.543544 | -7.53372 | 6.07E-11 | 4.04E-10 | 14.45585 |
| ATP5SL | -0.23103 | 0.852024 | 8.752405 | -7.53163 | 6.12E-11 | 4.08E-10 | 14.44658 |
| ZNF331 | -0.79228 | 0.577431 | 6.68557 | -7.52788 | 6.23E-11 | 4.14E-10 | 14.42991 |
| WARS | 0.403505 | 1.322717 | 8.468987 | 7.525319 | 6.30E-11 | 4.19E-10 | 14.41852 |
| BCL2L13 | -0.32081 | 0.80062 | 7.623165 | -7.52504 | 6.31E-11 | 4.19E-10 | 14.41729 |
| NPY1R | 1.292227 | 2.449057 | 8.175443 | 7.524554 | 6.32E-11 | 4.20E-10 | 14.41511 |
| DDB2 | 0.43386 | 1.350843 | 5.759494 | 7.52347 | 6.35E-11 | 4.21E-10 | 14.41029 |
| BLVRA | 0.246397 | 1.186241 | 8.815316 | 7.518198 | 6.51E-11 | 4.31E-10 | 14.38686 |
| PNMA2 | 0.503202 | 1.417355 | 5.370633 | 7.515189 | 6.60E-11 | 4.37E-10 | 14.37348 |
| FAM102A | -0.41143 | 0.751878 | 7.198354 | -7.51447 | 6.62E-11 | 4.38E-10 | 14.37029 |
| ASNS | 0.648735 | 1.567793 | 5.423165 | 7.513881 | 6.63E-11 | 4.39E-10 | 14.36767 |
| SP140L | 0.616568 | 1.533223 | 5.40038 | 7.51324 | 6.65E-11 | 4.39E-10 | 14.36481 |
| REG1A | 1.951706 | 3.868317 | 5.821139 | 7.510929 | 6.72E-11 | 4.44E-10 | 14.35454 |
| BST1 | -0.55935 | 0.678606 | 5.977215 | -7.51082 | 6.73E-11 | 4.44E-10 | 14.35404 |
| GUCY1B3 | 0.696034 | 1.620045 | 6.176835 | 7.510686 | 6.73E-11 | 4.44E-10 | 14.35346 |
| CST7 | 0.665211 | 1.5858 | 6.094177 | 7.506966 | 6.84E-11 | 4.51E-10 | 14.33693 |
| RBM7 | 0.413768 | 1.332161 | 7.362405 | 7.505699 | 6.88E-11 | 4.53E-10 | 14.3313 |
| FRYL | 0.329881 | 1.25691 | 6.46519 | 7.505088 | 6.90E-11 | 4.54E-10 | 14.32859 |
| EOGT | 0.381568 | 1.302757 | 4.297215 | 7.504863 | 6.91E-11 | 4.54E-10 | 14.32758 |
| CAPN1 | 0.340823 | 1.266479 | 6.582152 | 7.496741 | 7.17E-11 | 4.70E-10 | 14.29149 |
| CD300A | 0.521126 | 1.435075 | 4.927468 | 7.495417 | 7.21E-11 | 4.73E-10 | 14.28561 |
| UBE2M | -0.45565 | 0.72918 | 7.555316 | -7.49173 | 7.33E-11 | 4.80E-10 | 14.26921 |
| MAN1B1 | 0.267161 | 1.203437 | 6.927468 | 7.487391 | 7.48E-11 | 4.89E-10 | 14.24995 |
| CDC14B | -0.23692 | 0.848553 | 6.59038 | -7.48725 | 7.48E-11 | 4.89E-10 | 14.24933 |
| CTNNBIP1 | -0.31629 | 0.803132 | 7.781139 | -7.48388 | 7.60E-11 | 4.97E-10 | 14.23435 |
| SEC22A | 0.279875 | 1.21409 | 4.945823 | 7.483701 | 7.60E-11 | 4.97E-10 | 14.23357 |
| CHST8 | -0.30178 | 0.811252 | 5.51481 | -7.48337 | 7.61E-11 | 4.97E-10 | 14.23209 |
| RRM2 | 0.913702 | 1.883874 | 4.746456 | 7.482141 | 7.66E-11 | 4.99E-10 | 14.22663 |
| DENND5A | 0.325935 | 1.253477 | 7.815063 | 7.479357 | 7.75E-11 | 5.05E-10 | 14.21427 |
| LBR | 0.49747 | 1.411736 | 8.452152 | 7.479052 | 7.76E-11 | 5.06E-10 | 14.21291 |
| PLEKHO1 | 0.716258 | 1.642915 | 5.333544 | 7.478336 | 7.79E-11 | 5.07E-10 | 14.20974 |
| TUFM | -0.25171 | 0.839899 | 9.140506 | -7.47704 | 7.83E-11 | 5.09E-10 | 14.20397 |
| PNPLA3 | -0.64956 | 0.637475 | 6.086709 | -7.47034 | 8.07E-11 | 5.25E-10 | 14.17422 |
| C2 | 0.841588 | 1.792021 | 6.64481 | 7.468935 | 8.12E-11 | 5.28E-10 | 14.16799 |
| FKBP11 | 1.015626 | 2.02178 | 7.504684 | 7.46311 | 8.34E-11 | 5.41E-10 | 14.14212 |
| VDR | -0.53483 | 0.690241 | 7.142025 | -7.45829 | 8.52E-11 | 5.53E-10 | 14.12074 |
| TMEM258 | 0.351924 | 1.276261 | 10.53266 | 7.457764 | 8.54E-11 | 5.53E-10 | 14.11839 |
| MET | 0.430784 | 1.347966 | 7.528861 | 7.457695 | 8.55E-11 | 5.53E-10 | 14.11809 |
| FZD6 | 0.528597 | 1.442526 | 7.054937 | 7.45764 | 8.55E-11 | 5.53E-10 | 14.11784 |
| MDK | 0.460935 | 1.376434 | 5.646456 | 7.447291 | 8.96E-11 | 5.79E-10 | 14.07191 |
| SLC3A2 | -0.63039 | 0.646002 | 9.426456 | -7.44599 | 9.01E-11 | 5.82E-10 | 14.06614 |
| PRKD2 | 0.282404 | 1.21622 | 6.912532 | 7.443724 | 9.10E-11 | 5.87E-10 | 14.05607 |
| TUBB6 | 0.813412 | 1.757363 | 7.291519 | 7.438127 | 9.33E-11 | 6.02E-10 | 14.03124 |
| SUB1 | 0.276831 | 1.211531 | 9.597595 | 7.432864 | 9.56E-11 | 6.16E-10 | 14.00789 |
| KLC1 | 0.325066 | 1.252722 | 5.49557 | 7.432333 | 9.58E-11 | 6.17E-10 | 14.00553 |
| DLST | -0.5775 | 0.670122 | 8.982025 | -7.4308 | 9.65E-11 | 6.21E-10 | 13.99874 |

| | | | | | | | |
|---|---|---|---|---|---|---|---|
| RAP2B | 0.397497 | 1.31722 | 4.874304 | 7.429426 | 9.71E-11 | 6.24E-10 | 13.99264 |
| MALT1 | 0.541779 | 1.455766 | 6.225443 | 7.427568 | 9.79E-11 | 6.28E-10 | 13.98439 |
| PCCB | -0.45426 | 0.729883 | 9.16481 | -7.42754 | 9.79E-11 | 6.28E-10 | 13.98426 |
| CRABP1 | -0.47647 | 0.718735 | 5.405316 | -7.42678 | 9.82E-11 | 6.30E-10 | 13.98091 |
| POLR2I | -0.29383 | 0.815731 | 9.262911 | -7.42382 | 9.95E-11 | 6.38E-10 | 13.96776 |
| ELF1 | 0.35525 | 1.279208 | 7.47557 | 7.423175 | 9.98E-11 | 6.39E-10 | 13.96491 |
| FBXO9 | -0.25688 | 0.836893 | 8.122911 | -7.4197 | 1.01E-10 | 6.49E-10 | 13.94952 |
| ST3GAL1 | -0.60147 | 0.659082 | 6.908101 | -7.41563 | 1.03E-10 | 6.60E-10 | 13.93144 |
| MAST2 | -0.26008 | 0.835042 | 6.30962 | -7.41344 | 1.04E-10 | 6.67E-10 | 13.92174 |
| TTF1 | 0.290303 | 1.222897 | 5.951266 | 7.413048 | 1.04E-10 | 6.67E-10 | 13.92 |
| NR0B2 | -0.56829 | 0.674414 | 6.498481 | -7.41036 | 1.06E-10 | 6.75E-10 | 13.90809 |
| TRIB3 | 0.292971 | 1.225161 | 5.786076 | 7.406276 | 1.08E-10 | 6.87E-10 | 13.88997 |
| NPDC1 | 0.575804 | 1.490508 | 5.879873 | 7.405447 | 1.08E-10 | 6.89E-10 | 13.8863 |
| CHTOP | 0.272484 | 1.207885 | 7.710127 | 7.4042 | 1.09E-10 | 6.92E-10 | 13.88077 |
| NR4A1 | -1.10848 | 0.463781 | 6.236962 | -7.40264 | 1.09E-10 | 6.97E-10 | 13.87385 |
| SLC38A3 | -0.43121 | 0.741642 | 5.538354 | -7.40047 | 1.11E-10 | 7.03E-10 | 13.86422 |
| KIF20A | 0.40697 | 1.325898 | 4.34 | 7.398934 | 1.11E-10 | 7.07E-10 | 13.85743 |
| CD151 | 0.542958 | 1.456957 | 7.803544 | 7.397571 | 1.12E-10 | 7.11E-10 | 13.85138 |
| BST2 | 0.947879 | 1.929034 | 7.475949 | 7.395353 | 1.13E-10 | 7.18E-10 | 13.84155 |
| NDUFAF1 | -0.3429 | 0.788456 | 9.286329 | -7.38946 | 1.16E-10 | 7.36E-10 | 13.81544 |
| CD86 | 0.431047 | 1.348212 | 4.558101 | 7.38638 | 1.18E-10 | 7.46E-10 | 13.80179 |
| EXT2 | 0.260942 | 1.198261 | 7.651392 | 7.38557 | 1.18E-10 | 7.48E-10 | 13.7982 |
| TWF2 | 0.387372 | 1.308008 | 6.718987 | 7.385369 | 1.18E-10 | 7.48E-10 | 13.79731 |
| PFKL | -0.27738 | 0.825085 | 8.05 | -7.38518 | 1.18E-10 | 7.48E-10 | 13.79645 |
| TMEM45A | 0.601377 | 1.517164 | 4.533165 | 7.384725 | 1.19E-10 | 7.49E-10 | 13.79445 |
| PGPEP1 | -0.48181 | 0.716081 | 7.835696 | -7.38119 | 1.21E-10 | 7.61E-10 | 13.7788 |
| NDUFC1 | -0.29158 | 0.817006 | 10.58342 | -7.38095 | 1.21E-10 | 7.61E-10 | 13.77771 |
| CTBP2 | 0.336186 | 1.262415 | 7.341519 | 7.380557 | 1.21E-10 | 7.62E-10 | 13.77599 |
| ALDH5A1 | -0.46347 | 0.725239 | 6.828354 | -7.37789 | 1.22E-10 | 7.70E-10 | 13.76415 |
| MTCH2 | -0.33165 | 0.794629 | 9.881899 | -7.37754 | 1.23E-10 | 7.71E-10 | 13.76261 |
| TRIM37 | 0.323709 | 1.251544 | 7.578481 | 7.375211 | 1.24E-10 | 7.79E-10 | 13.7523 |
| IL13RA1 | 0.384137 | 1.305079 | 7.11481 | 7.369446 | 1.27E-10 | 7.98E-10 | 13.72676 |
| VTCN1 | 0.887661 | 1.850175 | 6.561013 | 7.368287 | 1.28E-10 | 8.02E-10 | 13.72163 |
| CYP1B1 | 0.78193 | 1.71943 | 6.620759 | 7.368186 | 1.28E-10 | 8.02E-10 | 13.72118 |
| PVRIG | 0.661653 | 1.581895 | 5.59443 | 7.364951 | 1.30E-10 | 8.13E-10 | 13.70685 |
| FAM49A | 0.419137 | 1.337127 | 4.13443 | 7.360903 | 1.32E-10 | 8.27E-10 | 13.68893 |
| RPL3 | 0.447879 | 1.364033 | 11.56709 | 7.359449 | 1.33E-10 | 8.32E-10 | 13.68249 |
| TXN | -0.33337 | 0.793679 | 11.49987 | -7.35878 | 1.33E-10 | 8.34E-10 | 13.67955 |
| CISH | -0.70604 | 0.613 | 8.280506 | -7.35847 | 1.34E-10 | 8.34E-10 | 13.67816 |
| SGCE | 0.431818 | 1.348933 | 8.17038 | 7.353097 | 1.37E-10 | 8.54E-10 | 13.65436 |
| DEFB1 | -0.69132 | 0.619288 | 12.03557 | -7.35263 | 1.37E-10 | 8.55E-10 | 13.65228 |
| GADD45B | -1.25272 | 0.419656 | 8.25519 | -7.35061 | 1.38E-10 | 8.63E-10 | 13.64338 |
| ARL6IP4 | -0.29065 | 0.817536 | 8.807722 | -7.34841 | 1.40E-10 | 8.70E-10 | 13.63364 |
| NDUFS7 | -0.32661 | 0.797406 | 9.271392 | -7.34821 | 1.40E-10 | 8.71E-10 | 13.63273 |
| ZNF227 | 0.374401 | 1.296301 | 4.779873 | 7.348058 | 1.40E-10 | 8.71E-10 | 13.63206 |
| IGKV4-1 | 1.74164 | 3.344152 | 6.528608 | 7.347549 | 1.40E-10 | 8.72E-10 | 13.6298 |
| PER1 | -0.55495 | 0.680679 | 7.482532 | -7.34612 | 1.41E-10 | 8.77E-10 | 13.62349 |

| | | | | | | | |
|---|---|---|---|---|---|---|---|
| FLI1 | 0.396456 | 1.31627 | 4.484304 | 7.344996 | 1.42E-10 | 8.81E-10 | 13.6185 |
| PRC1 | 0.517128 | 1.431103 | 4.519494 | 7.336573 | 1.47E-10 | 9.14E-10 | 13.58122 |
| IGHV3-21 | 2.020613 | 4.057561 | 4.742532 | 7.33463 | 1.49E-10 | 9.21E-10 | 13.57262 |
| CAPN6 | 0.562121 | 1.476438 | 4.39481 | 7.333373 | 1.49E-10 | 9.26E-10 | 13.56706 |
| TIMM17B | -0.3007 | 0.811859 | 7.795696 | -7.33145 | 1.51E-10 | 9.33E-10 | 13.55854 |
| KRT81 | -0.33146 | 0.794734 | 5.697848 | -7.33051 | 1.51E-10 | 9.36E-10 | 13.55441 |
| ARHGAP32 | -0.26553 | 0.831895 | 5.685823 | -7.32908 | 1.52E-10 | 9.42E-10 | 13.54805 |
| DDIT3 | -0.47832 | 0.717813 | 7.642152 | -7.32545 | 1.55E-10 | 9.57E-10 | 13.53199 |
| EFHD1 | -0.45319 | 0.730423 | 10.34873 | -7.32393 | 1.56E-10 | 9.62E-10 | 13.52526 |
| ERI3 | -0.42567 | 0.744495 | 6.780886 | -7.32379 | 1.56E-10 | 9.62E-10 | 13.52467 |
| SNRPB | 0.381976 | 1.303126 | 8.713038 | 7.323417 | 1.56E-10 | 9.63E-10 | 13.52301 |
| CHN1 | 0.357253 | 1.280984 | 6.243797 | 7.321273 | 1.58E-10 | 9.72E-10 | 13.51353 |
| MYLIP | 0.521001 | 1.434951 | 7.256329 | 7.320117 | 1.59E-10 | 9.76E-10 | 13.50841 |
| AFP | -0.41597 | 0.749516 | 5.284937 | -7.31981 | 1.59E-10 | 9.77E-10 | 13.50706 |
| FCMR | 0.426153 | 1.343646 | 5.176709 | 7.313684 | 1.63E-10 | 1.00E-09 | 13.47996 |
| DOK1 | 0.315283 | 1.244256 | 5.521266 | 7.308765 | 1.67E-10 | 1.03E-09 | 13.45821 |
| PIK3CB | -0.30329 | 0.810404 | 6.342658 | -7.30431 | 1.70E-10 | 1.04E-09 | 13.43849 |
| OGFOD2 | -0.22166 | 0.857578 | 6.780886 | -7.30259 | 1.72E-10 | 1.05E-09 | 13.43091 |
| PTPN12 | 0.347852 | 1.272665 | 5.481392 | 7.297049 | 1.76E-10 | 1.08E-09 | 13.4064 |
| PLCB4 | 0.584868 | 1.499902 | 4.579747 | 7.296936 | 1.76E-10 | 1.08E-09 | 13.4059 |
| CXCL9 | 1.672701 | 3.188109 | 6.16481 | 7.294885 | 1.78E-10 | 1.09E-09 | 13.39683 |
| L2HGDH | -0.46663 | 0.723654 | 5.495949 | -7.29354 | 1.79E-10 | 1.09E-09 | 13.3909 |
| CPT2 | -0.36231 | 0.777917 | 7.752785 | -7.29306 | 1.79E-10 | 1.09E-09 | 13.38877 |
| ICT1 | -0.36937 | 0.774122 | 7.76962 | -7.29245 | 1.80E-10 | 1.10E-09 | 13.38608 |
| CDH11 | 0.834196 | 1.782864 | 5.929114 | 7.291164 | 1.81E-10 | 1.10E-09 | 13.38039 |
| CD84 | 0.371693 | 1.29387 | 5.094177 | 7.285193 | 1.86E-10 | 1.13E-09 | 13.354 |
| BAG1 | -0.29397 | 0.815657 | 8.453291 | -7.28388 | 1.87E-10 | 1.14E-09 | 13.34822 |
| TPGS2 | 0.345329 | 1.270441 | 6.853165 | 7.281577 | 1.89E-10 | 1.15E-09 | 13.33802 |
| ASB13 | -0.42292 | 0.745911 | 9.358987 | -7.2768 | 1.93E-10 | 1.17E-09 | 13.31692 |
| ADIRF | -0.93171 | 0.524238 | 9.894937 | -7.27397 | 1.95E-10 | 1.19E-09 | 13.30441 |
| UBA5 | -0.28447 | 0.821045 | 7.662911 | -7.27309 | 1.96E-10 | 1.19E-09 | 13.30053 |
| ITPR3 | 0.331014 | 1.257898 | 5.873924 | 7.271332 | 1.97E-10 | 1.20E-09 | 13.29275 |
| UROS | -0.40196 | 0.756828 | 6.85557 | -7.26959 | 1.99E-10 | 1.21E-09 | 13.28505 |
| PTP4A2 | 0.274038 | 1.209188 | 8.941646 | 7.268848 | 2.00E-10 | 1.21E-09 | 13.28178 |
| ZMYM6NB | 0.384796 | 1.305675 | 8.192911 | 7.268157 | 2.00E-10 | 1.21E-09 | 13.27873 |
| ADAMDEC | 1.134256 | 2.195053 | 4.350759 | 7.2666 | 2.02E-10 | 1.22E-09 | 13.27185 |
| NUAK1 | 0.595784 | 1.511294 | 6.602785 | 7.266237 | 2.02E-10 | 1.22E-09 | 13.27025 |
| NDUFS3 | -0.29013 | 0.817827 | 9.919241 | -7.26517 | 2.03E-10 | 1.23E-09 | 13.26553 |
| DENND2D | 0.502833 | 1.416993 | 7.345696 | 7.264986 | 2.03E-10 | 1.23E-09 | 13.26472 |
| SSR4 | 0.452648 | 1.36855 | 10.14734 | 7.26272 | 2.05E-10 | 1.24E-09 | 13.25471 |
| GAD1 | -0.30445 | 0.809749 | 4.501519 | -7.26096 | 2.07E-10 | 1.25E-09 | 13.24693 |
| B4GAT1 | -0.38292 | 0.766885 | 8.614177 | -7.26078 | 2.07E-10 | 1.25E-09 | 13.24615 |
| LETMD1 | -0.34008 | 0.789998 | 8.236203 | -7.25889 | 2.09E-10 | 1.26E-09 | 13.23778 |
| CTSZ | 0.637984 | 1.556153 | 6.032152 | 7.252158 | 2.15E-10 | 1.30E-09 | 13.20807 |
| FUT6 | -0.53757 | 0.688931 | 8.183924 | -7.24582 | 2.21E-10 | 1.33E-09 | 13.1801 |
| SFN | 0.732391 | 1.661391 | 7.233544 | 7.243408 | 2.24E-10 | 1.34E-09 | 13.16945 |
| TNFRSF17 | 1.087793 | 2.125487 | 4.204177 | 7.243286 | 2.24E-10 | 1.34E-09 | 13.16891 |

| | | | | | | | |
|---|---|---|---|---|---|---|---|
| EHD4 | 0.406509 | 1.325474 | 7.626329 | 7.242782 | 2.24E-10 | 1.35E-09 | 13.16668 |
| CD40 | 0.351001 | 1.275446 | 5.885316 | 7.24141 | 2.26E-10 | 1.35E-09 | 13.16063 |
| GOT1 | -0.46593 | 0.724005 | 9.924937 | -7.23907 | 2.28E-10 | 1.37E-09 | 13.15031 |
| SAMD9 | 0.365356 | 1.288199 | 4.415443 | 7.23829 | 2.29E-10 | 1.37E-09 | 13.14686 |
| GLUL | 0.513676 | 1.427683 | 8.395443 | 7.23754 | 2.30E-10 | 1.37E-09 | 13.14355 |
| WBP2 | -0.2904 | 0.817678 | 7.05 | -7.23225 | 2.35E-10 | 1.41E-09 | 13.12018 |
| PHTF1 | 0.286957 | 1.220064 | 4.972911 | 7.23183 | 2.36E-10 | 1.41E-09 | 13.11835 |
| TRPV5 | -0.32601 | 0.797737 | 5.233165 | -7.23135 | 2.36E-10 | 1.41E-09 | 13.11623 |
| GLI3 | -0.39569 | 0.760125 | 7.109494 | -7.22798 | 2.40E-10 | 1.43E-09 | 13.10136 |
| FGF7 | 0.643709 | 1.56234 | 4.482152 | 7.225193 | 2.43E-10 | 1.45E-09 | 13.08906 |
| IQSEC2 | -0.2779 | 0.824792 | 5.758481 | -7.21878 | 2.50E-10 | 1.49E-09 | 13.06078 |
| CASP3 | 0.498393 | 1.412639 | 6.369494 | 7.21741 | 2.51E-10 | 1.50E-09 | 13.05473 |
| BHLHE41 | 0.345856 | 1.270905 | 5.041646 | 7.214905 | 2.54E-10 | 1.51E-09 | 13.04368 |
| BHLHE40 | -0.7085 | 0.611954 | 7.678608 | -7.21312 | 2.56E-10 | 1.52E-09 | 13.0358 |
| APOBEC3B | 0.867997 | 1.825128 | 4.48519 | 7.211387 | 2.58E-10 | 1.53E-09 | 13.02817 |
| DMPK | -0.38999 | 0.763137 | 5.804051 | -7.21002 | 2.60E-10 | 1.54E-09 | 13.02212 |
| SENP2 | -0.30784 | 0.807851 | 6.952278 | -7.20993 | 2.60E-10 | 1.54E-09 | 13.02172 |
| CAPRIN1 | 0.315323 | 1.24429 | 8.002152 | 7.209476 | 2.61E-10 | 1.54E-09 | 13.01974 |
| PHYHIP | -0.35994 | 0.779197 | 5.175949 | -7.20819 | 2.62E-10 | 1.55E-09 | 13.01408 |
| COX8A | -0.2593 | 0.835492 | 11.66038 | -7.20778 | 2.63E-10 | 1.55E-09 | 13.01225 |
| SH3YL1 | -0.59507 | 0.662011 | 10.11316 | -7.20521 | 2.66E-10 | 1.57E-09 | 13.00091 |
| OPTN | -0.34495 | 0.787337 | 8.818734 | -7.20251 | 2.69E-10 | 1.59E-09 | 12.98902 |
| CDK5 | -0.35854 | 0.779951 | 7.233924 | -7.20109 | 2.71E-10 | 1.60E-09 | 12.98275 |
| BLVRB | -0.33071 | 0.795144 | 9.066203 | -7.2 | 2.72E-10 | 1.61E-09 | 12.97796 |
| RBBP7 | 0.280995 | 1.215032 | 9.167595 | 7.199808 | 2.72E-10 | 1.61E-09 | 12.97712 |
| PDCD4 | 0.392563 | 1.312723 | 6.472025 | 7.199251 | 2.73E-10 | 1.61E-09 | 12.97466 |
| LY75 | 0.971522 | 1.960908 | 5.184304 | 7.193422 | 2.80E-10 | 1.65E-09 | 12.94896 |
| ASCC3 | 0.337319 | 1.263406 | 5.370253 | 7.190893 | 2.83E-10 | 1.67E-09 | 12.93782 |
| SORBS3 | -0.27475 | 0.826594 | 5.972405 | -7.18515 | 2.91E-10 | 1.71E-09 | 12.91249 |
| NUP37 | 0.362925 | 1.286031 | 7.615949 | 7.173247 | 3.06E-10 | 1.80E-09 | 12.86007 |
| RBFOX2 | 0.332286 | 1.259007 | 7.320759 | 7.172489 | 3.07E-10 | 1.81E-09 | 12.85673 |
| AGFG1 | 0.237391 | 1.178859 | 6.611772 | 7.168843 | 3.13E-10 | 1.83E-09 | 12.84067 |
| UROD | -0.23038 | 0.852413 | 9.200506 | -7.16783 | 3.14E-10 | 1.84E-09 | 12.83621 |
| CD1D | 0.424414 | 1.342027 | 3.927722 | 7.157788 | 3.28E-10 | 1.93E-09 | 12.79199 |
| PQLC1 | -0.62469 | 0.648559 | 9.072532 | -7.15525 | 3.32E-10 | 1.95E-09 | 12.7808 |
| NUP93 | 0.302411 | 1.233204 | 7.259367 | 7.155091 | 3.32E-10 | 1.95E-09 | 12.78012 |
| C11orf63 | -0.31972 | 0.801224 | 5.472532 | -7.15444 | 3.33E-10 | 1.95E-09 | 12.77723 |
| VRK2 | 0.309163 | 1.238989 | 7.052405 | 7.154356 | 3.33E-10 | 1.95E-09 | 12.77688 |
| OXA1L | -0.26638 | 0.831401 | 9.763291 | -7.15241 | 3.36E-10 | 1.97E-09 | 12.76831 |
| OSBPL1A | -0.39022 | 0.763011 | 8.298734 | -7.14885 | 3.42E-10 | 2.00E-09 | 12.75263 |
| SMYD2 | 0.399776 | 1.319303 | 8.558734 | 7.148331 | 3.43E-10 | 2.00E-09 | 12.75036 |
| PBX3 | 0.41614 | 1.334352 | 5.87557 | 7.146779 | 3.45E-10 | 2.01E-09 | 12.74352 |
| MLST8 | -0.27992 | 0.823636 | 6.61481 | -7.14398 | 3.49E-10 | 2.04E-09 | 12.7312 |
| BLOC1S1 | -0.30406 | 0.809967 | 8.971899 | -7.14336 | 3.50E-10 | 2.04E-09 | 12.72848 |
| CD69 | 0.758445 | 1.691667 | 4.245949 | 7.142368 | 3.52E-10 | 2.05E-09 | 12.72411 |
| ABCC10 | 0.285112 | 1.218505 | 6.588228 | 7.137632 | 3.59E-10 | 2.09E-09 | 12.70327 |
| SLC5A2 | -0.74327 | 0.597385 | 9.134304 | -7.13271 | 3.67E-10 | 2.13E-09 | 12.6816 |

| | | | | | | | |
|---|---|---|---|---|---|---|---|
| SORBS2 | 0.4533 | 1.369169 | 7.696962 | 7.131819 | 3.69E-10 | 2.14E-09 | 12.67769 |
| GPC4 | 0.589269 | 1.504484 | 7.495063 | 7.130003 | 3.72E-10 | 2.16E-09 | 12.66971 |
| ROBO1 | 0.474295 | 1.389239 | 5.645949 | 7.128408 | 3.74E-10 | 2.17E-09 | 12.66269 |
| LRRC16A | 0.449368 | 1.365442 | 7.623797 | 7.127653 | 3.76E-10 | 2.18E-09 | 12.65937 |
| PSMG1 | -0.39427 | 0.760875 | 8.431392 | -7.122 | 3.85E-10 | 2.23E-09 | 12.63451 |
| TRPV4 | -0.46126 | 0.726353 | 5.637975 | -7.12169 | 3.86E-10 | 2.23E-09 | 12.63315 |
| PFN1 | 0.54191 | 1.455899 | 8.675063 | 7.117959 | 3.92E-10 | 2.27E-09 | 12.61673 |
| TIAM1 | 0.326805 | 1.254233 | 5.004557 | 7.112088 | 4.03E-10 | 2.33E-09 | 12.59092 |
| FANCG | 0.261713 | 1.198901 | 6.043671 | 7.112038 | 4.03E-10 | 2.33E-09 | 12.5907 |
| NUCB1 | -0.39181 | 0.762175 | 8.153291 | -7.11173 | 4.03E-10 | 2.33E-09 | 12.58935 |
| MRPS22 | -0.20474 | 0.867693 | 8.830127 | -7.10953 | 4.07E-10 | 2.35E-09 | 12.57969 |
| ZNF688 | -0.24457 | 0.844066 | 5.803924 | -7.10731 | 4.11E-10 | 2.37E-09 | 12.56992 |
| TAGLN2 | 0.476219 | 1.391093 | 8.502405 | 7.107042 | 4.12E-10 | 2.37E-09 | 12.56873 |
| IMPA2 | -0.37896 | 0.768992 | 10.76127 | -7.10098 | 4.23E-10 | 2.44E-09 | 12.5421 |
| GNPDA1 | -0.53663 | 0.689381 | 9.056709 | -7.09984 | 4.25E-10 | 2.45E-09 | 12.53709 |
| PRKD3 | 0.302286 | 1.233097 | 5.968228 | 7.09799 | 4.29E-10 | 2.47E-09 | 12.52895 |
| MRPL2 | -0.36984 | 0.773867 | 7.378987 | -7.09165 | 4.41E-10 | 2.53E-09 | 12.5011 |
| IL2RB | 0.60641 | 1.522466 | 6.282658 | 7.089014 | 4.47E-10 | 2.56E-09 | 12.48951 |
| ST6GAL1 | 0.662095 | 1.582379 | 7.252658 | 7.087756 | 4.49E-10 | 2.58E-09 | 12.48398 |
| RFNG | -0.32871 | 0.796249 | 6.70443 | -7.08743 | 4.50E-10 | 2.58E-09 | 12.48256 |
| FKBP3 | 0.362233 | 1.285414 | 7.795443 | 7.086164 | 4.52E-10 | 2.59E-09 | 12.47699 |
| COPZ2 | -0.38404 | 0.766286 | 7.593924 | -7.08394 | 4.57E-10 | 2.61E-09 | 12.46722 |
| CHP2 | -0.94218 | 0.520446 | 6.370127 | -7.0832 | 4.58E-10 | 2.62E-09 | 12.46395 |
| FAM96B | -0.26685 | 0.831132 | 9.621139 | -7.08107 | 4.63E-10 | 2.64E-09 | 12.4546 |
| HPCAL1 | -0.34462 | 0.787517 | 7.992785 | -7.07822 | 4.69E-10 | 2.68E-09 | 12.44208 |
| PSMB6 | -0.24457 | 0.844066 | 9.323924 | -7.07791 | 4.69E-10 | 2.68E-09 | 12.44075 |
| NUP214 | 0.285665 | 1.218972 | 6.140633 | 7.077701 | 4.70E-10 | 2.68E-09 | 12.43982 |
| TCEB3-AS1 | -0.37988 | 0.768501 | 6.463924 | -7.07439 | 4.77E-10 | 2.72E-09 | 12.42528 |
| GM2A | 0.292174 | 1.224484 | 6.839873 | 7.072834 | 4.80E-10 | 2.73E-09 | 12.41844 |
| POMGNT1 | -0.30632 | 0.8087 | 6.102911 | -7.07126 | 4.83E-10 | 2.75E-09 | 12.41155 |
| HACD1 | 0.484427 | 1.39903 | 5.169747 | 7.069772 | 4.87E-10 | 2.77E-09 | 12.405 |
| SRI | 0.335343 | 1.261677 | 8.485949 | 7.068567 | 4.89E-10 | 2.78E-09 | 12.39971 |
| PCYOX1 | -0.68566 | 0.621722 | 8.050759 | -7.06564 | 4.96E-10 | 2.82E-09 | 12.38687 |
| ROCK1 | 0.407846 | 1.326703 | 6.417975 | 7.061465 | 5.05E-10 | 2.87E-09 | 12.36853 |
| ABCB7 | -0.29699 | 0.813949 | 7.560506 | -7.05958 | 5.09E-10 | 2.89E-09 | 12.36025 |
| FAM214B | -0.28558 | 0.820412 | 6.70962 | -7.05653 | 5.16E-10 | 2.93E-09 | 12.34687 |
| P3H4 | 0.287721 | 1.22071 | 5.326709 | 7.054142 | 5.22E-10 | 2.95E-09 | 12.33639 |
| NFKBIE | 0.628551 | 1.546011 | 6.137342 | 7.054094 | 5.22E-10 | 2.95E-09 | 12.33618 |
| NDUFA3 | -0.26885 | 0.829979 | 10.75291 | -7.05233 | 5.26E-10 | 2.98E-09 | 12.32846 |
| MMP2 | 0.453498 | 1.369356 | 6.601392 | 7.050521 | 5.30E-10 | 3.00E-09 | 12.3205 |
| ESR1 | -0.19738 | 0.872134 | 5.251899 | -7.04843 | 5.35E-10 | 3.02E-09 | 12.31134 |
| IGLV3-19 | 1.657536 | 3.154773 | 5.521519 | 7.047605 | 5.37E-10 | 3.03E-09 | 12.30771 |
| RPAP3 | 0.322312 | 1.250333 | 5.220506 | 7.046797 | 5.39E-10 | 3.04E-09 | 12.30416 |
| MAST3 | -0.30801 | 0.807755 | 6.321772 | -7.04488 | 5.44E-10 | 3.07E-09 | 12.29577 |
| UPF3B | 0.308696 | 1.238587 | 5.580253 | 7.044621 | 5.44E-10 | 3.07E-09 | 12.29461 |
| CEP68 | 0.286318 | 1.219523 | 6.010253 | 7.040727 | 5.54E-10 | 3.12E-09 | 12.27753 |
| DDRGK1 | 0.296647 | 1.228286 | 8.028481 | 7.040564 | 5.54E-10 | 3.12E-09 | 12.27682 |

| | | | | | | | |
|---|---|---|---|---|---|---|---|
| DCTD | 0.295889 | 1.227642 | 7.943165 | 7.038951 | 5.58E-10 | 3.14E-09 | 12.26974 |
| SDHD | -0.45133 | 0.731368 | 10.73886 | -7.03791 | 5.61E-10 | 3.15E-09 | 12.26517 |
| NRL | -0.25648 | 0.837127 | 5.400253 | -7.03226 | 5.75E-10 | 3.23E-09 | 12.24041 |
| CLEC7A | 0.662984 | 1.583354 | 4.707595 | 7.029984 | 5.81E-10 | 3.26E-09 | 12.23042 |
| CLIC4 | -0.53435 | 0.690471 | 9.741139 | -7.02988 | 5.81E-10 | 3.26E-09 | 12.22997 |
| LGALS2 | -0.76375 | 0.588964 | 10.10684 | -7.02901 | 5.83E-10 | 3.27E-09 | 12.22615 |
| DDN | -0.48522 | 0.714386 | 7.194051 | -7.02774 | 5.87E-10 | 3.29E-09 | 12.22058 |
| NDUFB7 | -0.35229 | 0.783342 | 10.00089 | -7.0253 | 5.93E-10 | 3.32E-09 | 12.20988 |
| GOSR2 | -0.27866 | 0.824355 | 6.784684 | -7.02527 | 5.93E-10 | 3.32E-09 | 12.20975 |
| RNF19B | -0.32645 | 0.797497 | 6.813418 | -7.02423 | 5.96E-10 | 3.33E-09 | 12.20519 |
| AFTPH | -0.30738 | 0.808105 | 9.837468 | -7.01936 | 6.09E-10 | 3.40E-09 | 12.18386 |
| LUC7L3 | 0.39892 | 1.31852 | 7.806203 | 7.018804 | 6.11E-10 | 3.41E-09 | 12.18141 |
| RB1 | 0.275435 | 1.210359 | 5.53962 | 7.016351 | 6.17E-10 | 3.44E-09 | 12.17066 |
| RNF123 | -0.30269 | 0.810737 | 6.745949 | -7.01245 | 6.28E-10 | 3.50E-09 | 12.15356 |
| MGP | 1.064644 | 2.091654 | 10.3519 | 7.011445 | 6.31E-10 | 3.52E-09 | 12.14916 |
| DROSHA | 0.287358 | 1.220404 | 7.130253 | 7.009144 | 6.37E-10 | 3.55E-09 | 12.13907 |
| PIBF1 | 0.385382 | 1.306206 | 5.137722 | 7.00819 | 6.40E-10 | 3.56E-09 | 12.13489 |
| CD96 | 0.350283 | 1.274811 | 3.910886 | 7.006227 | 6.46E-10 | 3.59E-09 | 12.12629 |
| TMX1 | 0.467859 | 1.383055 | 7.87 | 7.005788 | 6.47E-10 | 3.59E-09 | 12.12437 |
| HIPK3 | -0.46877 | 0.722581 | 5.41962 | -7.00578 | 6.47E-10 | 3.59E-09 | 12.12432 |
| CPN2 | -0.75939 | 0.590744 | 5.432785 | -7.00164 | 6.59E-10 | 3.66E-09 | 12.10619 |
| CYP2A13 | -0.33517 | 0.79269 | 5.604557 | -6.99932 | 6.66E-10 | 3.69E-09 | 12.09603 |
| THAP7 | -0.28758 | 0.819274 | 6.401392 | -6.99774 | 6.71E-10 | 3.72E-09 | 12.0891 |
| RNF167 | -0.23636 | 0.848882 | 8.091266 | -6.99734 | 6.72E-10 | 3.72E-09 | 12.08735 |
| PITX3 | -0.29646 | 0.81425 | 5.947468 | -6.99672 | 6.74E-10 | 3.73E-09 | 12.08465 |
| MDH2 | -0.26499 | 0.832206 | 9.281266 | -6.99461 | 6.80E-10 | 3.76E-09 | 12.07541 |
| ARHGEF16 | -0.30079 | 0.811807 | 6.666962 | -6.9909 | 6.91E-10 | 3.82E-09 | 12.05917 |
| TMBIM6 | -0.32879 | 0.796202 | 12.09418 | -6.98945 | 6.96E-10 | 3.85E-09 | 12.05279 |
| TRANK1 | 0.400132 | 1.319628 | 5.066709 | 6.98453 | 7.11E-10 | 3.93E-09 | 12.03127 |
| SLC12A9 | -0.26763 | 0.830684 | 6.37038 | -6.98325 | 7.15E-10 | 3.95E-09 | 12.02566 |
| PRPSAP2 | -0.3133 | 0.804799 | 7.531519 | -6.98296 | 7.16E-10 | 3.95E-09 | 12.02438 |
| APOL3 | 0.32247 | 1.25047 | 5.894051 | 6.980588 | 7.24E-10 | 3.99E-09 | 12.01401 |
| IGHV3-23 | 1.699888 | 3.248757 | 5.185949 | 6.979324 | 7.28E-10 | 4.01E-09 | 12.00848 |
| SERPINA4 | -0.56775 | 0.674666 | 5.883924 | -6.97652 | 7.37E-10 | 4.06E-09 | 11.99619 |
| GTF2E1 | 0.358597 | 1.282178 | 7.033924 | 6.976269 | 7.38E-10 | 4.06E-09 | 11.9951 |
| USP3 | 0.349664 | 1.274264 | 7.753671 | 6.975712 | 7.40E-10 | 4.07E-09 | 11.99267 |
| TLR4 | 0.333702 | 1.260243 | 4.594177 | 6.975611 | 7.40E-10 | 4.07E-09 | 11.99223 |
| TP53TG5 | -0.39183 | 0.762162 | 5.739367 | -6.97449 | 7.44E-10 | 4.08E-09 | 11.98732 |
| ADCK3 | -0.38791 | 0.764239 | 7.674051 | -6.9732 | 7.48E-10 | 4.10E-09 | 11.98167 |
| RYK | 0.246884 | 1.186641 | 6.712911 | 6.971861 | 7.53E-10 | 4.13E-09 | 11.97581 |
| DYNLT1 | 0.273999 | 1.209155 | 10.85076 | 6.971626 | 7.53E-10 | 4.13E-09 | 11.97479 |
| GPR183 | 0.509954 | 1.424005 | 4.643671 | 6.968735 | 7.63E-10 | 4.18E-09 | 11.96214 |
| TMPRSS4 | 0.611845 | 1.528212 | 6.639494 | 6.96846 | 7.64E-10 | 4.18E-09 | 11.96093 |
| PTRF | 0.310494 | 1.240132 | 6.33557 | 6.96712 | 7.69E-10 | 4.20E-09 | 11.95507 |
| MYB | 0.332885 | 1.25953 | 4.172532 | 6.96669 | 7.70E-10 | 4.21E-09 | 11.95319 |
| CANT1 | 0.228643 | 1.171732 | 6.758987 | 6.966519 | 7.71E-10 | 4.21E-09 | 11.95244 |
| SERPINB1 | 0.427016 | 1.34445 | 7.357722 | 6.964893 | 7.76E-10 | 4.24E-09 | 11.94533 |

| | | | | | | | |
|---|---|---|---|---|---|---|---|
| PDK4 | -1.42021 | 0.373658 | 7.785443 | -6.96096 | 7.90E-10 | 4.31E-09 | 11.92812 |
| PCCA | -0.42248 | 0.746142 | 9.805696 | -6.96052 | 7.91E-10 | 4.31E-09 | 11.92621 |
| CORO1A | 0.877708 | 1.837453 | 5.736203 | 6.960139 | 7.93E-10 | 4.32E-09 | 11.92453 |
| SAMM50 | -0.39493 | 0.760524 | 9.06481 | -6.95992 | 7.94E-10 | 4.32E-09 | 11.92359 |
| AEBP1 | 0.830626 | 1.778457 | 6.842405 | 6.959715 | 7.94E-10 | 4.32E-09 | 11.92268 |
| CTSO | 0.410744 | 1.329372 | 7.88962 | 6.959599 | 7.95E-10 | 4.32E-09 | 11.92217 |
| INVS | -0.39576 | 0.760087 | 6.316203 | -6.95944 | 7.95E-10 | 4.32E-09 | 11.92145 |
| DDX25 | -0.38007 | 0.768403 | 5.506456 | -6.95791 | 8.01E-10 | 4.35E-09 | 11.91476 |
| DDO | -0.40236 | 0.756617 | 6.128228 | -6.9559 | 8.08E-10 | 4.38E-09 | 11.90597 |
| IK | 0.271383 | 1.206965 | 9.063797 | 6.955832 | 8.08E-10 | 4.38E-09 | 11.90569 |
| PAPSS1 | 0.390343 | 1.310705 | 8.843924 | 6.951803 | 8.23E-10 | 4.46E-09 | 11.88807 |
| CFH | 1.00168 | 2.00233 | 6.30038 | 6.951734 | 8.23E-10 | 4.46E-09 | 11.88777 |
| TM7SF3 | -0.45605 | 0.728981 | 6.066456 | -6.95167 | 8.23E-10 | 4.46E-09 | 11.88749 |
| SRPX | 0.815125 | 1.759451 | 5.066582 | 6.950841 | 8.26E-10 | 4.47E-09 | 11.88386 |
| HDAC1 | 0.38052 | 1.301811 | 7.248734 | 6.948644 | 8.34E-10 | 4.51E-09 | 11.87426 |
| GNG11 | 0.67496 | 1.596553 | 7.936076 | 6.944542 | 8.50E-10 | 4.59E-09 | 11.85632 |
| CDC123 | 0.28247 | 1.216276 | 8.587342 | 6.941833 | 8.60E-10 | 4.64E-09 | 11.84448 |
| LPPR1 | -0.72455 | 0.605188 | 8.747342 | -6.93983 | 8.68E-10 | 4.68E-09 | 11.83574 |
| IFIH1 | 0.714025 | 1.640374 | 5.848481 | 6.938731 | 8.72E-10 | 4.70E-09 | 11.83092 |
| ABHD10 | -0.49409 | 0.710009 | 8.858608 | -6.93799 | 8.75E-10 | 4.71E-09 | 11.82769 |
| NUP62 | 0.330099 | 1.257099 | 5.845063 | 6.937038 | 8.79E-10 | 4.73E-09 | 11.82352 |
| SPATA2L | -0.60549 | 0.657249 | 7.344684 | -6.93699 | 8.79E-10 | 4.73E-09 | 11.8233 |
| MR1 | 0.264236 | 1.201 | 5.831899 | 6.936873 | 8.79E-10 | 4.73E-09 | 11.8228 |
| RLN2 | 0.343386 | 1.268731 | 4.161266 | 6.930694 | 9.04E-10 | 4.86E-09 | 11.7958 |
| SHARPIN | -0.23724 | 0.848367 | 6.973291 | -6.93003 | 9.06E-10 | 4.87E-09 | 11.79289 |
| PIK3R3 | 0.294308 | 1.226297 | 5.637722 | 6.928183 | 9.14E-10 | 4.90E-09 | 11.78483 |
| DARS2 | -0.33458 | 0.793016 | 6.947848 | -6.92744 | 9.17E-10 | 4.92E-09 | 11.78158 |
| SEMA4G | -0.3019 | 0.811185 | 5.772152 | -6.9264 | 9.21E-10 | 4.94E-09 | 11.77702 |
| SHANK2 | -0.3701 | 0.77373 | 6.148228 | -6.92515 | 9.26E-10 | 4.96E-09 | 11.77159 |
| IGLV1-40 | 1.433393 | 2.700811 | 7.325063 | 6.921911 | 9.40E-10 | 5.03E-09 | 11.75743 |
| MANF | 0.442899 | 1.359333 | 9.285443 | 6.919652 | 9.49E-10 | 5.08E-09 | 11.74756 |
| SHBG | -0.40362 | 0.755957 | 6.158354 | -6.91871 | 9.53E-10 | 5.10E-09 | 11.74344 |
| TM7SF2 | -0.40211 | 0.756748 | 7.700506 | -6.91588 | 9.65E-10 | 5.16E-09 | 11.73108 |
| CDK1 | 0.403333 | 1.32256 | 4.658481 | 6.915676 | 9.66E-10 | 5.16E-09 | 11.73019 |
| PARK2 | -0.28621 | 0.820052 | 5.105443 | -6.91437 | 9.72E-10 | 5.19E-09 | 11.72449 |
| TNNC1 | -0.56381 | 0.676514 | 4.964051 | -6.91394 | 9.73E-10 | 5.19E-09 | 11.7226 |
| PFKFB2 | -0.49236 | 0.710859 | 6.660633 | -6.90999 | 9.91E-10 | 5.28E-09 | 11.70536 |
| XBP1 | 0.527971 | 1.4419 | 7.889241 | 6.905817 | 1.01E-09 | 5.38E-09 | 11.68715 |
| SAFB2 | -0.2316 | 0.851689 | 7.623038 | -6.90488 | 1.01E-09 | 5.40E-09 | 11.68303 |
| KCNK10 | -0.42657 | 0.744026 | 5.300506 | -6.90462 | 1.01E-09 | 5.40E-09 | 11.68192 |
| ZNHIT1 | -0.29789 | 0.81344 | 9.688608 | -6.89804 | 1.04E-09 | 5.55E-09 | 11.6532 |
| TRHDE | -0.59372 | 0.662631 | 6.801772 | -6.89799 | 1.04E-09 | 5.55E-09 | 11.65296 |
| BCAS3 | -0.27166 | 0.828366 | 6.88 | -6.8942 | 1.06E-09 | 5.64E-09 | 11.63643 |
| CALU | 0.244519 | 1.184698 | 7.088228 | 6.893733 | 1.06E-09 | 5.65E-09 | 11.6344 |
| SOD2 | 0.980613 | 1.973303 | 7.998101 | 6.892667 | 1.07E-09 | 5.67E-09 | 11.62975 |
| COL5A1 | 0.313702 | 1.242893 | 4.645823 | 6.889858 | 1.08E-09 | 5.74E-09 | 11.61749 |
| POU2AF1 | 1.328874 | 2.512065 | 4.925316 | 6.888542 | 1.09E-09 | 5.77E-09 | 11.61175 |

| | | | | | | | |
|---|---|---|---|---|---|---|---|
| OGDH | -0.40105 | 0.757308 | 7.38443 | -6.88799 | 1.09E-09 | 5.78E-09 | 11.60932 |
| RPL36 | 0.443603 | 1.359997 | 10.04291 | 6.887933 | 1.09E-09 | 5.78E-09 | 11.60909 |
| MPDZ | -0.46623 | 0.723853 | 7.85481 | -6.88229 | 1.12E-09 | 5.92E-09 | 11.58448 |
| CCDC90B | 0.319361 | 1.247778 | 7.401013 | 6.8814 | 1.12E-09 | 5.94E-09 | 11.5806 |
| TNNT2 | -0.56242 | 0.677163 | 5.715063 | -6.88109 | 1.13E-09 | 5.95E-09 | 11.57926 |
| MGAT3 | -0.232 | 0.851452 | 5.215696 | -6.87961 | 1.13E-09 | 5.98E-09 | 11.57277 |
| COPG2IT1 | -0.34451 | 0.787578 | 4.846962 | -6.87803 | 1.14E-09 | 6.02E-09 | 11.56592 |
| MICAL2 | 0.25917 | 1.19679 | 5.2 | 6.87563 | 1.15E-09 | 6.08E-09 | 11.55543 |
| EMD | -0.29024 | 0.817768 | 7.263544 | -6.87522 | 1.16E-09 | 6.09E-09 | 11.55365 |
| FOSB | -1.66735 | 0.314831 | 6.051772 | -6.87364 | 1.16E-09 | 6.13E-09 | 11.54673 |
| MS4A1 | 0.82334 | 1.769498 | 4.892405 | 6.872904 | 1.17E-09 | 6.14E-09 | 11.54354 |
| FCN1 | 0.770547 | 1.705916 | 4.95557 | 6.87256 | 1.17E-09 | 6.15E-09 | 11.54205 |
| RHOG | 0.334776 | 1.261182 | 7.586582 | 6.872478 | 1.17E-09 | 6.15E-09 | 11.54169 |
| TFB2M | -0.38968 | 0.763297 | 7.914177 | -6.8717 | 1.17E-09 | 6.17E-09 | 11.53828 |
| ADA | 0.421673 | 1.33948 | 5.29962 | 6.868622 | 1.19E-09 | 6.25E-09 | 11.52487 |
| SLC1A3 | 0.449618 | 1.365679 | 5.506076 | 6.864305 | 1.21E-09 | 6.36E-09 | 11.50606 |
| TAPBP | 0.298037 | 1.22947 | 6.927975 | 6.862795 | 1.22E-09 | 6.40E-09 | 11.49947 |
| PCTP | -0.51843 | 0.698133 | 8.189747 | -6.86277 | 1.22E-09 | 6.40E-09 | 11.49938 |
| PTPN6 | 0.412055 | 1.33058 | 5.777342 | 6.860764 | 1.23E-09 | 6.45E-09 | 11.49062 |
| LPGAT1 | 0.509447 | 1.423504 | 5.500633 | 6.860395 | 1.23E-09 | 6.46E-09 | 11.48901 |
| COQ9 | -0.30621 | 0.808762 | 9.657089 | -6.85975 | 1.24E-09 | 6.47E-09 | 11.48619 |
| GLB1 | 0.330962 | 1.257852 | 8.896076 | 6.859526 | 1.24E-09 | 6.47E-09 | 11.48522 |
| CSK | 0.34946 | 1.274083 | 7.070759 | 6.858095 | 1.25E-09 | 6.51E-09 | 11.47899 |
| TEX30 | -0.33632 | 0.79206 | 6.998861 | -6.85608 | 1.26E-09 | 6.57E-09 | 11.4702 |
| MTRR | -0.3915 | 0.762336 | 8.093418 | -6.85511 | 1.26E-09 | 6.59E-09 | 11.46599 |
| SLC30A9 | -0.38053 | 0.768153 | 9.254304 | -6.85169 | 1.28E-09 | 6.69E-09 | 11.45108 |
| NELFCD | -0.29487 | 0.815147 | 8.841392 | -6.85129 | 1.29E-09 | 6.69E-09 | 11.44935 |
| HTR6 | -0.23371 | 0.850446 | 4.749114 | -6.84928 | 1.30E-09 | 6.75E-09 | 11.44058 |
| CCBL1 | -0.31588 | 0.803359 | 6.906962 | -6.84767 | 1.31E-09 | 6.79E-09 | 11.43358 |
| FLRT3 | 0.76307 | 1.697098 | 8.34962 | 6.847052 | 1.31E-09 | 6.81E-09 | 11.43087 |
| TNFAIP2 | 0.450422 | 1.36644 | 6.550759 | 6.845614 | 1.32E-09 | 6.85E-09 | 11.42461 |
| MPP1 | -0.33756 | 0.791377 | 8.575949 | -6.8385 | 1.36E-09 | 7.06E-09 | 11.39362 |
| PGRMC2 | -0.3491 | 0.785071 | 8.317215 | -6.83764 | 1.37E-09 | 7.09E-09 | 11.38987 |
| WDR1 | 0.275863 | 1.210718 | 9.020886 | 6.835459 | 1.38E-09 | 7.15E-09 | 11.38038 |
| UBE2D2 | 0.194302 | 1.14417 | 7.467468 | 6.834947 | 1.38E-09 | 7.16E-09 | 11.37816 |
| PKIG | -0.25491 | 0.838041 | 8.997215 | -6.83359 | 1.39E-09 | 7.20E-09 | 11.37225 |
| LOC10013( | 0.879895 | 1.840241 | 6.411899 | 6.833202 | 1.39E-09 | 7.21E-09 | 11.37056 |
| LSM14B | -0.28128 | 0.822858 | 5.369241 | -6.83253 | 1.40E-09 | 7.23E-09 | 11.36764 |
| R3HCC1 | -0.22502 | 0.855583 | 8.16557 | -6.83218 | 1.40E-09 | 7.23E-09 | 11.36613 |
| ZNF468 | 0.32892 | 1.256072 | 5.436962 | 6.827267 | 1.43E-09 | 7.39E-09 | 11.34472 |
| DENR | 0.339104 | 1.264971 | 8.668608 | 6.827001 | 1.43E-09 | 7.39E-09 | 11.34356 |
| ENDOG | -0.44044 | 0.736909 | 7.537975 | -6.82675 | 1.43E-09 | 7.40E-09 | 11.34248 |
| ESRRG | -0.49831 | 0.707934 | 7.992278 | -6.8254 | 1.44E-09 | 7.43E-09 | 11.33659 |
| LAMA5 | 0.351028 | 1.275469 | 7.679241 | 6.824652 | 1.45E-09 | 7.46E-09 | 11.33334 |
| ATP6V1G2 | -0.35736 | 0.780593 | 4.940506 | -6.82273 | 1.46E-09 | 7.51E-09 | 11.32496 |
| CAV1 | 0.675955 | 1.597654 | 6.416709 | 6.815351 | 1.51E-09 | 7.76E-09 | 11.29287 |
| RAB17 | -0.4735 | 0.720213 | 8.711772 | -6.81498 | 1.51E-09 | 7.77E-09 | 11.29124 |

| | | | | | | | |
|---|---|---|---|---|---|---|---|
| CYP46A1 | -0.44026 | 0.737003 | 6.015443 | -6.81457 | 1.51E-09 | 7.78E-09 | 11.28947 |
| ETFDH | -0.45068 | 0.731699 | 6.778481 | -6.81195 | 1.53E-09 | 7.86E-09 | 11.27807 |
| APOBEC3F | 0.348426 | 1.27317 | 4.279241 | 6.811929 | 1.53E-09 | 7.86E-09 | 11.27798 |
| MAF | -0.61319 | 0.65375 | 7.60038 | -6.8085 | 1.55E-09 | 7.97E-09 | 11.26308 |
| ITGA3 | 0.554328 | 1.468485 | 6.671772 | 6.806699 | 1.57E-09 | 8.03E-09 | 11.25523 |
| PLXNA1 | 0.204493 | 1.152281 | 5.657595 | 6.804517 | 1.58E-09 | 8.11E-09 | 11.24574 |
| DDX56 | -0.22298 | 0.856791 | 7.376203 | -6.80007 | 1.61E-09 | 8.26E-09 | 11.22639 |
| RMDN1 | -0.35715 | 0.780707 | 7.491899 | -6.79959 | 1.62E-09 | 8.27E-09 | 11.22431 |
| ATF7IP | 0.338505 | 1.264445 | 7.076835 | 6.797174 | 1.63E-09 | 8.36E-09 | 11.21381 |
| RNF44 | 0.273333 | 1.208597 | 7.604177 | 6.796888 | 1.64E-09 | 8.36E-09 | 11.21257 |
| MAP3K1 | 0.359236 | 1.282746 | 5.256582 | 6.795645 | 1.64E-09 | 8.40E-09 | 11.20717 |
| GINS1 | 0.466542 | 1.381793 | 5.443797 | 6.793171 | 1.66E-09 | 8.49E-09 | 11.19641 |
| PLEKHO2 | 0.460165 | 1.375699 | 7.324177 | 6.792942 | 1.66E-09 | 8.49E-09 | 11.19542 |
| ZBED5 | 0.366278 | 1.289023 | 8.362785 | 6.792216 | 1.67E-09 | 8.52E-09 | 11.19226 |
| HSD11B2 | -0.6761 | 0.625855 | 9.526709 | -6.78759 | 1.70E-09 | 8.69E-09 | 11.17214 |
| CENPM | 0.431291 | 1.34844 | 5.071899 | 6.786917 | 1.71E-09 | 8.71E-09 | 11.16923 |
| ZNHIT6 | 0.325797 | 1.253357 | 5.716962 | 6.778095 | 1.78E-09 | 9.05E-09 | 11.1309 |
| COMT | 0.240066 | 1.181047 | 8.155063 | 6.777346 | 1.78E-09 | 9.07E-09 | 11.12764 |
| CCNG1 | -0.2949 | 0.815128 | 10.96899 | -6.77524 | 1.80E-09 | 9.15E-09 | 11.11848 |
| PTOV1 | -0.34718 | 0.786119 | 8.263671 | -6.77523 | 1.80E-09 | 9.15E-09 | 11.11846 |
| TCEA2 | -0.2998 | 0.812367 | 6.527595 | -6.7751 | 1.80E-09 | 9.15E-09 | 11.1179 |
| CPA3 | 1.398524 | 2.636318 | 5.835063 | 6.774464 | 1.81E-09 | 9.17E-09 | 11.11512 |
| PDXDC1 | -0.25676 | 0.836966 | 8.954051 | -6.77422 | 1.81E-09 | 9.17E-09 | 11.11408 |
| MICB | 0.531976 | 1.445909 | 4.775696 | 6.773653 | 1.81E-09 | 9.19E-09 | 11.1116 |
| FBRS | -0.24378 | 0.844529 | 5.621646 | -6.77129 | 1.83E-09 | 9.28E-09 | 11.10132 |
| SGPL1 | 0.230198 | 1.172996 | 6.450506 | 6.770162 | 1.84E-09 | 9.32E-09 | 11.09644 |
| RGN | -0.78323 | 0.581063 | 8.49 | -6.76915 | 1.85E-09 | 9.36E-09 | 11.09205 |
| FAM49B | 0.310132 | 1.239821 | 7.099114 | 6.760991 | 1.92E-09 | 9.70E-09 | 11.05661 |
| TXLNA | 0.312286 | 1.241674 | 6.132405 | 6.756315 | 1.96E-09 | 9.89E-09 | 11.03632 |
| VPS51 | -0.25912 | 0.835599 | 9.127848 | -6.75382 | 1.98E-09 | 1.00E-08 | 11.0255 |
| FIP1L1 | 0.295395 | 1.227221 | 5.617089 | 6.753238 | 1.98E-09 | 1.00E-08 | 11.02296 |
| PRF1 | 0.347115 | 1.272014 | 5.591519 | 6.751387 | 2.00E-09 | 1.01E-08 | 11.01493 |
| CHN2 | -0.42858 | 0.742991 | 6.243797 | -6.74871 | 2.02E-09 | 1.02E-08 | 11.00331 |
| PDCD5 | 0.43307 | 1.350103 | 6.171772 | 6.748702 | 2.02E-09 | 1.02E-08 | 11.00328 |
| ADH5 | -0.36653 | 0.775643 | 10.15646 | -6.74302 | 2.07E-09 | 1.05E-08 | 10.97864 |
| MLXIPL | -0.42671 | 0.743955 | 6.802405 | -6.74222 | 2.08E-09 | 1.05E-08 | 10.97517 |
| NELL1 | -0.66718 | 0.629736 | 6.25 | -6.73813 | 2.12E-09 | 1.07E-08 | 10.95742 |
| ST6GALNA | 0.336561 | 1.262743 | 5.234937 | 6.736737 | 2.13E-09 | 1.07E-08 | 10.95137 |
| SERPINC1 | -0.54919 | 0.683404 | 4.793418 | -6.73296 | 2.17E-09 | 1.09E-08 | 10.935 |
| FOLR2 | 0.58166 | 1.49657 | 7.389494 | 6.73229 | 2.18E-09 | 1.09E-08 | 10.93209 |
| ORC6 | 0.361733 | 1.284968 | 4.240886 | 6.730304 | 2.19E-09 | 1.10E-08 | 10.92348 |
| TDO2 | 0.768498 | 1.703495 | 4.577975 | 6.729802 | 2.20E-09 | 1.10E-08 | 10.9213 |
| IL32 | 1.226054 | 2.339263 | 7.733671 | 6.728271 | 2.21E-09 | 1.11E-08 | 10.91466 |
| MSH2 | 0.404855 | 1.323956 | 5.447595 | 6.719259 | 2.30E-09 | 1.15E-08 | 10.8756 |
| SMPD1 | -0.39883 | 0.758471 | 7.334051 | -6.71749 | 2.32E-09 | 1.16E-08 | 10.86792 |
| SYK | 0.363874 | 1.286876 | 5.417215 | 6.715384 | 2.34E-09 | 1.17E-08 | 10.85881 |
| ENTPD7 | 0.34251 | 1.267961 | 5.563291 | 6.714426 | 2.35E-09 | 1.18E-08 | 10.85465 |

| | | | | | | | |
|---|---|---|---|---|---|---|---|
| PPARA | -0.27648 | 0.825602 | 5.521899 | -6.71293 | 2.37E-09 | 1.18E-08 | 10.84815 |
| PTP4A1 | -0.36499 | 0.776476 | 7.689494 | -6.71256 | 2.37E-09 | 1.19E-08 | 10.84656 |
| PBX2 | -0.26276 | 0.833492 | 5.577848 | -6.71006 | 2.40E-09 | 1.20E-08 | 10.83573 |
| MYH9 | 0.310487 | 1.240127 | 7.987089 | 6.703655 | 2.47E-09 | 1.23E-08 | 10.80799 |
| SLC27A5 | -0.25811 | 0.836183 | 6.105443 | -6.70334 | 2.47E-09 | 1.23E-08 | 10.80663 |
| HECA | 0.401126 | 1.320539 | 7.457342 | 6.702645 | 2.48E-09 | 1.24E-08 | 10.80362 |
| TPK1 | 0.420389 | 1.338288 | 5.245823 | 6.701774 | 2.49E-09 | 1.24E-08 | 10.79985 |
| ENTPD4 | 0.230586 | 1.173312 | 5.950886 | 6.701168 | 2.50E-09 | 1.24E-08 | 10.79722 |
| HNMT | -0.29355 | 0.815892 | 7.647595 | -6.70089 | 2.50E-09 | 1.24E-08 | 10.79603 |
| C17orf62 | 0.406831 | 1.325771 | 5.901899 | 6.699615 | 2.51E-09 | 1.25E-08 | 10.7905 |
| SSBP2 | 0.349756 | 1.274345 | 5.632405 | 6.699538 | 2.51E-09 | 1.25E-08 | 10.79016 |
| METTL7A | -0.43322 | 0.740606 | 9.353165 | -6.69806 | 2.53E-09 | 1.26E-08 | 10.78374 |
| ZBED8 | 0.429664 | 1.34692 | 4.537089 | 6.696493 | 2.55E-09 | 1.26E-08 | 10.77698 |
| TBKBP1 | -0.25191 | 0.839784 | 5.686076 | -6.68609 | 2.67E-09 | 1.32E-08 | 10.73193 |
| COL5A2 | 0.358682 | 1.282254 | 4.744177 | 6.685189 | 2.68E-09 | 1.33E-08 | 10.72805 |
| SIGLEC1 | 0.335369 | 1.2617 | 5.279873 | 6.683185 | 2.70E-09 | 1.34E-08 | 10.71937 |
| MIOS | 0.27415 | 1.209282 | 6.855823 | 6.679556 | 2.74E-09 | 1.36E-08 | 10.70367 |
| ANP32B | 0.317938 | 1.246548 | 9.259114 | 6.679129 | 2.75E-09 | 1.36E-08 | 10.70182 |
| RBX1 | 0.290718 | 1.223249 | 9.73557 | 6.677437 | 2.77E-09 | 1.37E-08 | 10.6945 |
| SIK3 | -0.22014 | 0.858483 | 6.31 | -6.67609 | 2.79E-09 | 1.38E-08 | 10.68867 |
| ADGRL4 | 0.568762 | 1.48325 | 5.323671 | 6.675158 | 2.80E-09 | 1.38E-08 | 10.68465 |
| CXCL12 | 0.813617 | 1.757612 | 8.71443 | 6.674218 | 2.81E-09 | 1.39E-08 | 10.68058 |
| KIAA0753 | 0.303669 | 1.23428 | 6.879241 | 6.673778 | 2.82E-09 | 1.39E-08 | 10.67868 |
| LHFPL2 | 0.416482 | 1.334669 | 6.626582 | 6.671987 | 2.84E-09 | 1.40E-08 | 10.67093 |
| ZFP36 | -0.89244 | 0.538703 | 8.460253 | -6.67192 | 2.84E-09 | 1.40E-08 | 10.67064 |
| AGPAT1 | -0.27521 | 0.82633 | 8.008734 | -6.66456 | 2.93E-09 | 1.44E-08 | 10.63879 |
| DNASE2 | 0.322859 | 1.250807 | 6.52443 | 6.664466 | 2.93E-09 | 1.44E-08 | 10.6384 |
| KDELC1 | 0.663827 | 1.58428 | 5.133165 | 6.658935 | 3.01E-09 | 1.48E-08 | 10.61449 |
| PLIN3 | 0.430652 | 1.347843 | 7.999241 | 6.655741 | 3.05E-09 | 1.50E-08 | 10.60068 |
| LAMP3 | 0.849381 | 1.801727 | 4.727848 | 6.653282 | 3.08E-09 | 1.51E-08 | 10.59006 |
| COPB1 | 0.236238 | 1.177917 | 9.887595 | 6.651846 | 3.10E-09 | 1.52E-08 | 10.58385 |
| ARFIP1 | 0.314993 | 1.244006 | 6.708101 | 6.651372 | 3.11E-09 | 1.53E-08 | 10.5818 |
| AOC1 | -1.44853 | 0.366394 | 10.43557 | -6.65045 | 3.12E-09 | 1.53E-08 | 10.57783 |
| ASAP1 | 0.265224 | 1.201823 | 5.284051 | 6.646596 | 3.17E-09 | 1.56E-08 | 10.56117 |
| NAE1 | 0.384348 | 1.30527 | 8.916203 | 6.64616 | 3.18E-09 | 1.56E-08 | 10.55928 |
| MAFB | 0.772095 | 1.707748 | 7.758608 | 6.645756 | 3.18E-09 | 1.56E-08 | 10.55754 |
| ZDHHC4 | -0.25377 | 0.838699 | 8.125949 | -6.64495 | 3.20E-09 | 1.56E-08 | 10.55404 |
| SHC1 | 0.25747 | 1.195381 | 7.311899 | 6.644163 | 3.21E-09 | 1.57E-08 | 10.55066 |
| CXCL1 | 0.796897 | 1.737361 | 4.97962 | 6.641651 | 3.24E-09 | 1.59E-08 | 10.5398 |
| GRAMD1C | -0.48111 | 0.716425 | 6.896203 | -6.64073 | 3.26E-09 | 1.59E-08 | 10.53581 |
| SLC25A32 | -0.49872 | 0.707737 | 6.984937 | -6.6401 | 3.26E-09 | 1.59E-08 | 10.53309 |
| C12orf10 | -0.30897 | 0.80722 | 7.922025 | -6.63902 | 3.28E-09 | 1.60E-08 | 10.52846 |
| CTSK | 0.641871 | 1.560351 | 6.355949 | 6.636297 | 3.32E-09 | 1.62E-08 | 10.51668 |
| MBIP | -0.23366 | 0.850477 | 5.786962 | -6.63536 | 3.33E-09 | 1.63E-08 | 10.51263 |
| SIPA1L1 | -0.20292 | 0.868787 | 6.490886 | -6.63474 | 3.34E-09 | 1.63E-08 | 10.50994 |
| PTGS1 | 0.394776 | 1.314739 | 5.071646 | 6.634183 | 3.35E-09 | 1.63E-08 | 10.50755 |
| NHEJ1 | -0.26951 | 0.829604 | 7.553291 | -6.63413 | 3.35E-09 | 1.63E-08 | 10.5073 |

| | | | | | | | |
|---|---|---|---|---|---|---|---|
| LIN7A | -0.34036 | 0.789847 | 5.16 | -6.63306 | 3.37E-09 | 1.64E-08 | 10.50268 |
| TIA1 | 0.298142 | 1.22956 | 6.123671 | 6.632772 | 3.37E-09 | 1.64E-08 | 10.50145 |
| ACSBG2 | -0.42704 | 0.743789 | 5.336835 | -6.63198 | 3.38E-09 | 1.64E-08 | 10.49806 |
| NCF4 | 0.412701 | 1.331176 | 4.898481 | 6.63067 | 3.40E-09 | 1.65E-08 | 10.49238 |
| TYK2 | -0.2686 | 0.830123 | 7.67519 | -6.62973 | 3.42E-09 | 1.66E-08 | 10.48831 |
| PLEK2 | 0.310152 | 1.239838 | 6.884557 | 6.628386 | 3.44E-09 | 1.67E-08 | 10.48252 |
| TM4SF1 | 0.708682 | 1.634311 | 6.69038 | 6.627661 | 3.45E-09 | 1.67E-08 | 10.47939 |
| DNAJC6 | -0.2253 | 0.855415 | 6.270886 | -6.6253 | 3.48E-09 | 1.69E-08 | 10.46918 |
| PARM1 | -0.48189 | 0.716039 | 8.155443 | -6.62472 | 3.49E-09 | 1.69E-08 | 10.46671 |
| SERPINA3 | 1.932128 | 3.816176 | 8.18557 | 6.616814 | 3.62E-09 | 1.75E-08 | 10.43258 |
| BCLAF1 | 0.285889 | 1.219162 | 7.198987 | 6.613513 | 3.67E-09 | 1.77E-08 | 10.41833 |
| STAG1 | 0.303669 | 1.23428 | 6.429241 | 6.612732 | 3.68E-09 | 1.78E-08 | 10.41496 |
| SLC10A2 | -0.90055 | 0.535681 | 7.951646 | -6.61176 | 3.70E-09 | 1.79E-08 | 10.41078 |
| MAPK10 | -0.24238 | 0.845347 | 6.42962 | -6.61048 | 3.72E-09 | 1.80E-08 | 10.40523 |
| FTO | -0.23778 | 0.848049 | 8.857848 | -6.60854 | 3.75E-09 | 1.81E-08 | 10.39689 |
| GCC2 | 0.380204 | 1.301526 | 7.871646 | 6.604471 | 3.82E-09 | 1.84E-08 | 10.37933 |
| TAPBPL | 0.526113 | 1.440044 | 6.487595 | 6.60065 | 3.88E-09 | 1.87E-08 | 10.36286 |
| ALDH1B1 | -0.60599 | 0.657018 | 7.551646 | -6.59996 | 3.89E-09 | 1.88E-08 | 10.3599 |
| ANXA9 | -0.65063 | 0.637004 | 6.922785 | -6.59666 | 3.95E-09 | 1.90E-08 | 10.34565 |
| CNOT8 | 0.278478 | 1.212915 | 6.657848 | 6.596404 | 3.96E-09 | 1.90E-08 | 10.34455 |
| DOCK10 | 0.442233 | 1.358706 | 4.138861 | 6.595175 | 3.98E-09 | 1.91E-08 | 10.33925 |
| ST3GAL5 | 0.354756 | 1.27877 | 6.119494 | 6.593214 | 4.01E-09 | 1.93E-08 | 10.3308 |
| NOP16 | -0.32443 | 0.798616 | 6.327089 | -6.59215 | 4.03E-09 | 1.94E-08 | 10.3262 |
| PLA2G4C | 0.720211 | 1.647423 | 6.622152 | 6.589659 | 4.07E-09 | 1.96E-08 | 10.31548 |
| PRDX4 | 0.483037 | 1.397683 | 9.230253 | 6.587629 | 4.11E-09 | 1.97E-08 | 10.30673 |
| ACACB | -0.37933 | 0.768796 | 6.886329 | -6.58687 | 4.12E-09 | 1.98E-08 | 10.30348 |
| RPS24 | 0.254829 | 1.193194 | 13.04101 | 6.584268 | 4.17E-09 | 2.00E-08 | 10.29225 |
| BRD3 | 0.342431 | 1.267891 | 7.661519 | 6.583845 | 4.18E-09 | 2.00E-08 | 10.29042 |
| AHNAK | 0.361206 | 1.284499 | 7.342405 | 6.578931 | 4.27E-09 | 2.04E-08 | 10.26926 |
| PLEKHB2 | -0.34502 | 0.787297 | 9.265443 | -6.57818 | 4.29E-09 | 2.05E-08 | 10.26602 |
| AGR2 | 0.998511 | 1.997937 | 5.511013 | 6.578096 | 4.29E-09 | 2.05E-08 | 10.26566 |
| KCNN2 | -1.01779 | 0.493871 | 5.245063 | -6.57588 | 4.33E-09 | 2.07E-08 | 10.2561 |
| RHCG | -0.97589 | 0.508426 | 8.242785 | -6.57359 | 4.37E-09 | 2.09E-08 | 10.24624 |
| SSTR2 | -0.38015 | 0.768357 | 5.116203 | -6.57118 | 4.42E-09 | 2.11E-08 | 10.23588 |
| GNA11 | -0.23762 | 0.848142 | 7.541392 | -6.57036 | 4.43E-09 | 2.12E-08 | 10.23234 |
| PAH | -1.05329 | 0.481869 | 11.75937 | -6.5698 | 4.45E-09 | 2.12E-08 | 10.22995 |
| FAM198B | 0.403636 | 1.322838 | 4.308608 | 6.569413 | 4.45E-09 | 2.12E-08 | 10.22826 |
| LPAR6 | 0.457365 | 1.373032 | 7.479747 | 6.568967 | 4.46E-09 | 2.13E-08 | 10.22635 |
| CPNE6 | -1.0669 | 0.477342 | 5.739114 | -6.568 | 4.48E-09 | 2.13E-08 | 10.22219 |
| LYPD1 | 0.593478 | 1.50888 | 5.23443 | 6.566979 | 4.50E-09 | 2.14E-08 | 10.21778 |
| CTSB | -0.42505 | 0.744812 | 10.83962 | -6.56509 | 4.54E-09 | 2.16E-08 | 10.20963 |
| NDUFS4 | -0.27074 | 0.828896 | 9.387342 | -6.56315 | 4.58E-09 | 2.18E-08 | 10.20129 |
| LETM1 | -0.27305 | 0.827568 | 7.220506 | -6.56088 | 4.62E-09 | 2.20E-08 | 10.19153 |
| TOX | 0.283742 | 1.217348 | 4.234177 | 6.557873 | 4.68E-09 | 2.22E-08 | 10.17859 |
| OFD1 | 0.384018 | 1.304972 | 7.702152 | 6.556835 | 4.71E-09 | 2.23E-08 | 10.17413 |
| OPA1 | -0.25684 | 0.836916 | 6.623797 | -6.55435 | 4.76E-09 | 2.26E-08 | 10.16345 |
| TCF12 | 0.349736 | 1.274328 | 8.166962 | 6.554014 | 4.76E-09 | 2.26E-08 | 10.16198 |

| | | | | | | | |
|---|---|---|---|---|---|---|---|
| NDUFV1 | -0.23628 | 0.848933 | 10.69152 | -6.55165 | 4.81E-09 | 2.28E-08 | 10.15181 |
| PMS1 | 0.369578 | 1.291975 | 6.681772 | 6.54657 | 4.92E-09 | 2.33E-08 | 10.12996 |
| GGTLC1 | -0.44821 | 0.732953 | 9.318861 | -6.54473 | 4.96E-09 | 2.35E-08 | 10.12205 |
| DECR1 | -0.27163 | 0.828385 | 10.54241 | -6.54248 | 5.01E-09 | 2.37E-08 | 10.11236 |
| CAPG | 0.635679 | 1.553668 | 8.163797 | 6.536424 | 5.15E-09 | 2.43E-08 | 10.08633 |
| NPHS1 | -0.61253 | 0.654049 | 5.448481 | -6.53319 | 5.22E-09 | 2.46E-08 | 10.07241 |
| FAS | 0.651113 | 1.57038 | 6.34481 | 6.53066 | 5.28E-09 | 2.49E-08 | 10.06156 |
| PSIP1 | 0.35365 | 1.277789 | 7.204684 | 6.52674 | 5.37E-09 | 2.53E-08 | 10.04471 |
| RAC3 | -0.31375 | 0.804545 | 4.436329 | -6.52634 | 5.38E-09 | 2.54E-08 | 10.04299 |
| CDK2AP1 | 0.262391 | 1.199465 | 9.917215 | 6.524146 | 5.43E-09 | 2.56E-08 | 10.03356 |
| BPI | -1.21841 | 0.429755 | 5.824304 | -6.5239 | 5.44E-09 | 2.56E-08 | 10.0325 |
| TRAP1 | -0.28996 | 0.817924 | 9.449747 | -6.52156 | 5.49E-09 | 2.58E-08 | 10.02247 |
| ALDH3A1 | -0.27411 | 0.82696 | 4.925063 | -6.52109 | 5.50E-09 | 2.59E-08 | 10.02044 |
| HDDC2 | -0.33941 | 0.790366 | 7.721266 | -6.52041 | 5.52E-09 | 2.60E-08 | 10.01751 |
| TEX10 | 0.231542 | 1.174089 | 6.690633 | 6.519519 | 5.54E-09 | 2.60E-08 | 10.01368 |
| DENND1A | -0.35742 | 0.780557 | 5.355696 | -6.51608 | 5.62E-09 | 2.64E-08 | 9.998893 |
| HMGN4 | 0.267951 | 1.204097 | 8.60519 | 6.512923 | 5.70E-09 | 2.68E-08 | 9.985353 |
| KCNJ8 | 0.661258 | 1.581461 | 5.89557 | 6.511012 | 5.75E-09 | 2.70E-08 | 9.97715 |
| EFHC1 | 0.409157 | 1.327909 | 5.375696 | 6.508836 | 5.81E-09 | 2.72E-08 | 9.967805 |
| NR2F2 | 0.354381 | 1.278437 | 7.656076 | 6.506866 | 5.86E-09 | 2.74E-08 | 9.959349 |
| PCSK1N | -0.5986 | 0.660396 | 8.325823 | -6.50423 | 5.92E-09 | 2.77E-08 | 9.948047 |
| USE1 | -0.24951 | 0.841184 | 7.011646 | -6.50403 | 5.93E-09 | 2.78E-08 | 9.947185 |
| PRR5 | -0.32254 | 0.799659 | 6.742658 | -6.50291 | 5.96E-09 | 2.79E-08 | 9.942364 |
| SETX | 0.260086 | 1.19755 | 6.608861 | 6.500657 | 6.02E-09 | 2.81E-08 | 9.932696 |
| IMPDH2 | 0.407833 | 1.326691 | 9.051013 | 6.49959 | 6.05E-09 | 2.83E-08 | 9.928116 |
| FOLH1 | -0.98631 | 0.504767 | 7.635823 | -6.49816 | 6.08E-09 | 2.84E-08 | 9.921996 |
| TBP | 0.239631 | 1.180691 | 6.685316 | 6.494108 | 6.19E-09 | 2.89E-08 | 9.904592 |
| PLIN2 | -0.67543 | 0.626143 | 10.59329 | -6.4908 | 6.28E-09 | 2.93E-08 | 9.890386 |
| ATP6V0E2 | -0.58521 | 0.666552 | 8.629241 | -6.48632 | 6.41E-09 | 2.99E-08 | 9.871185 |
| PKM | 0.525896 | 1.439827 | 8.487722 | 6.485804 | 6.42E-09 | 2.99E-08 | 9.868972 |
| C9orf91 | 0.309783 | 1.239521 | 4.65962 | 6.484927 | 6.45E-09 | 3.00E-08 | 9.865209 |
| TNFRSF10I | -0.34992 | 0.784627 | 5.28557 | -6.48457 | 6.46E-09 | 3.01E-08 | 9.863661 |
| CSGALNAC | 0.629032 | 1.546527 | 6.076456 | 6.482068 | 6.53E-09 | 3.04E-08 | 9.852949 |
| GADD45GI | -0.33889 | 0.790651 | 6.591266 | -6.4814 | 6.55E-09 | 3.04E-08 | 9.850088 |
| ZKSCAN7 | 0.259209 | 1.196823 | 4.810886 | 6.480335 | 6.58E-09 | 3.06E-08 | 9.845519 |
| TIPARP | -0.99766 | 0.500811 | 8.583038 | -6.47896 | 6.62E-09 | 3.07E-08 | 9.839625 |
| STAB1 | 0.386752 | 1.307447 | 6.081772 | 6.477278 | 6.66E-09 | 3.09E-08 | 9.832412 |
| GYS1 | -0.28704 | 0.819581 | 6.976835 | -6.47159 | 6.83E-09 | 3.17E-08 | 9.808021 |
| SLCO2A1 | 0.465646 | 1.380935 | 7.11038 | 6.470444 | 6.87E-09 | 3.18E-08 | 9.803121 |
| EVX1 | -0.36308 | 0.777501 | 5.030506 | -6.46939 | 6.90E-09 | 3.20E-08 | 9.798585 |
| OXSM | -0.29493 | 0.815113 | 7.585063 | -6.4693 | 6.90E-09 | 3.20E-08 | 9.798227 |
| CHFR | 0.333748 | 1.260284 | 6.153544 | 6.469162 | 6.91E-09 | 3.20E-08 | 9.797625 |
| ENG | 0.335949 | 1.262207 | 6.156203 | 6.467835 | 6.95E-09 | 3.21E-08 | 9.791938 |
| RPA2 | 0.359974 | 1.283402 | 7.796456 | 6.466075 | 7.00E-09 | 3.24E-08 | 9.784397 |
| ITK | 0.42365 | 1.341316 | 4.193924 | 6.464208 | 7.06E-09 | 3.26E-08 | 9.776397 |
| RHBDF2 | 0.3511 | 1.275533 | 5.312532 | 6.462209 | 7.12E-09 | 3.29E-08 | 9.767835 |
| TMEM184 | 0.319091 | 1.247544 | 6.973291 | 6.461991 | 7.12E-09 | 3.29E-08 | 9.7669 |

| | | | | | | | |
|---|---|---|---|---|---|---|---|
| ESRP2 | -0.34555 | 0.787006 | 6.848481 | -6.46152 | 7.14E-09 | 3.29E-08 | 9.764905 |
| SPARC | 0.70444 | 1.629512 | 8.428608 | 6.460345 | 7.18E-09 | 3.31E-08 | 9.75985 |
| CALCOCO1 | -0.27644 | 0.825624 | 6.962785 | -6.45863 | 7.23E-09 | 3.33E-08 | 9.752498 |
| PPT2 | -0.24643 | 0.84298 | 7.712278 | -6.45788 | 7.25E-09 | 3.34E-08 | 9.74931 |
| ARSA | -0.27177 | 0.828302 | 6.175823 | -6.45625 | 7.31E-09 | 3.36E-08 | 9.742318 |
| MCAT | -0.4191 | 0.747892 | 7.686456 | -6.45484 | 7.35E-09 | 3.38E-08 | 9.736276 |
| NLGN4X | 0.359025 | 1.282559 | 3.89519 | 6.450587 | 7.49E-09 | 3.44E-08 | 9.718061 |
| HMGN3 | 0.349888 | 1.274462 | 10.86203 | 6.449058 | 7.54E-09 | 3.47E-08 | 9.711517 |
| DUSP1 | -1.14864 | 0.451049 | 7.726709 | -6.44763 | 7.59E-09 | 3.49E-08 | 9.705416 |
| CCRN4L | -0.34161 | 0.789158 | 5.430127 | -6.44708 | 7.60E-09 | 3.49E-08 | 9.70303 |
| NT5M | -0.2531 | 0.83909 | 5.091013 | -6.44261 | 7.75E-09 | 3.56E-08 | 9.683925 |
| GIPC2 | -0.71606 | 0.608757 | 9.330886 | -6.44208 | 7.77E-09 | 3.56E-08 | 9.681665 |
| TRIM26 | -0.27715 | 0.825221 | 7.545316 | -6.43667 | 7.96E-09 | 3.65E-08 | 9.658478 |
| CCR5 | 0.448709 | 1.364818 | 6.655696 | 6.434097 | 8.05E-09 | 3.69E-08 | 9.647488 |
| TIMM10 | -0.30457 | 0.809682 | 8.268861 | -6.42741 | 8.28E-09 | 3.79E-08 | 9.618892 |
| CENPU | 0.382833 | 1.303899 | 4.40557 | 6.426788 | 8.31E-09 | 3.80E-08 | 9.616227 |
| GBA3 | -0.64498 | 0.639502 | 10.21101 | -6.4266 | 8.31E-09 | 3.80E-08 | 9.615442 |
| TEX2 | -0.30964 | 0.806844 | 7.676962 | -6.42634 | 8.32E-09 | 3.81E-08 | 9.614307 |
| RPL35A | 0.382668 | 1.303751 | 11.88354 | 6.423977 | 8.41E-09 | 3.84E-08 | 9.604205 |
| IFIT5 | 0.43193 | 1.349037 | 7.204557 | 6.422977 | 8.45E-09 | 3.86E-08 | 9.599932 |
| OLFM4 | 1.720982 | 3.296606 | 5.622152 | 6.420524 | 8.54E-09 | 3.90E-08 | 9.589445 |
| SLC31A2 | -0.61318 | 0.653753 | 8.108861 | -6.42009 | 8.55E-09 | 3.90E-08 | 9.587598 |
| DNPEP | -0.20419 | 0.868026 | 8.342532 | -6.41912 | 8.59E-09 | 3.92E-08 | 9.583423 |
| YBX3 | 0.479914 | 1.394661 | 9.590253 | 6.418322 | 8.62E-09 | 3.93E-08 | 9.580032 |
| IGFBP6 | 1.063347 | 2.089773 | 7.461139 | 6.416832 | 8.68E-09 | 3.95E-08 | 9.57366 |
| ARNTL2 | 0.277148 | 1.211797 | 4.344684 | 6.416319 | 8.69E-09 | 3.96E-08 | 9.571467 |
| SAP18 | -0.21903 | 0.859146 | 8.853291 | -6.41498 | 8.75E-09 | 3.98E-08 | 9.565759 |
| KRT10 | 0.24832 | 1.187823 | 8.841772 | 6.41068 | 8.91E-09 | 4.05E-08 | 9.54737 |
| INPP5D | 0.310613 | 1.240234 | 5.288228 | 6.410392 | 8.92E-09 | 4.06E-08 | 9.546138 |
| SHTN1 | 0.497582 | 1.411846 | 6.986329 | 6.40987 | 8.94E-09 | 4.06E-08 | 9.543909 |
| GOLGA3 | 0.26139 | 1.198633 | 6.728101 | 6.403298 | 9.20E-09 | 4.18E-08 | 9.515831 |
| LMNB1 | 0.295797 | 1.227563 | 4.54443 | 6.403064 | 9.21E-09 | 4.18E-08 | 9.514832 |
| C2orf72 | -0.27132 | 0.828559 | 5.382532 | -6.40063 | 9.31E-09 | 4.22E-08 | 9.504436 |
| IL1R1 | 0.313169 | 1.242434 | 5.628861 | 6.400452 | 9.32E-09 | 4.22E-08 | 9.503678 |
| PRKCQ | 0.466825 | 1.382064 | 6.438481 | 6.398247 | 9.41E-09 | 4.26E-08 | 9.494262 |
| MYL3 | -0.31043 | 0.806402 | 7.409241 | -6.39161 | 9.68E-09 | 4.38E-08 | 9.465942 |
| PYGB | 0.280777 | 1.214849 | 7.907722 | 6.386234 | 9.91E-09 | 4.49E-08 | 9.442976 |
| SKIV2L2 | 0.261574 | 1.198786 | 7.88557 | 6.385681 | 9.94E-09 | 4.49E-08 | 9.440618 |
| SLC22A5 | -0.48736 | 0.713327 | 9.067722 | -6.38447 | 9.99E-09 | 4.52E-08 | 9.435447 |
| SLFN12 | 0.37087 | 1.293132 | 4.464051 | 6.384059 | 1.00E-08 | 4.52E-08 | 9.433696 |
| GZMH | 0.614592 | 1.531124 | 5.296076 | 6.383809 | 1.00E-08 | 4.52E-08 | 9.432628 |
| EFEMP1 | 0.559374 | 1.47363 | 8.228228 | 6.383214 | 1.00E-08 | 4.53E-08 | 9.430088 |
| SLC16A4 | -0.42416 | 0.745274 | 10.04304 | -6.38319 | 1.00E-08 | 4.53E-08 | 9.429981 |
| SEC11A | 0.230527 | 1.173263 | 10.71456 | 6.381002 | 1.01E-08 | 4.57E-08 | 9.420651 |
| TESK1 | -0.29194 | 0.816801 | 7.196962 | -6.37899 | 1.02E-08 | 4.61E-08 | 9.412082 |
| HMGB2 | 0.537556 | 1.451511 | 7.949114 | 6.378187 | 1.03E-08 | 4.62E-08 | 9.408643 |
| NAB2 | -0.33037 | 0.795333 | 5.947215 | -6.37786 | 1.03E-08 | 4.63E-08 | 9.407254 |

| | | | | | | | |
|---|---|---|---|---|---|---|---|
| SRPX2 | 0.336812 | 1.262962 | 4.367215 | 6.376614 | 1.03E-08 | 4.65E-08 | 9.401932 |
| NUP155 | 0.263801 | 1.200638 | 5.422152 | 6.376022 | 1.04E-08 | 4.66E-08 | 9.399405 |
| GNL3 | 0.353636 | 1.277777 | 7.167722 | 6.375535 | 1.04E-08 | 4.67E-08 | 9.397329 |
| RNF128 | -0.43352 | 0.740451 | 9.183038 | -6.37473 | 1.04E-08 | 4.68E-08 | 9.393897 |
| FGF1 | -0.82715 | 0.56364 | 6.677089 | -6.37283 | 1.05E-08 | 4.72E-08 | 9.385808 |
| B4GALT5 | 0.621067 | 1.538012 | 8.182911 | 6.367162 | 1.08E-08 | 4.83E-08 | 9.361622 |
| RBM22 | 0.232918 | 1.17521 | 7.323165 | 6.366751 | 1.08E-08 | 4.84E-08 | 9.35987 |
| LEPROT | 0.375217 | 1.297035 | 10.08152 | 6.366724 | 1.08E-08 | 4.84E-08 | 9.359755 |
| AP1M2 | -0.42088 | 0.746967 | 8.358101 | -6.35992 | 1.11E-08 | 4.98E-08 | 9.330747 |
| TCN2 | -0.43931 | 0.737488 | 9.106709 | -6.35942 | 1.11E-08 | 4.99E-08 | 9.328618 |
| MPZL2 | 0.345718 | 1.270783 | 4.893544 | 6.356827 | 1.13E-08 | 5.04E-08 | 9.317572 |
| EXOSC10 | 0.396476 | 1.316288 | 5.679747 | 6.356005 | 1.13E-08 | 5.06E-08 | 9.31407 |
| ADAR | 0.333702 | 1.260243 | 9.784177 | 6.355184 | 1.13E-08 | 5.07E-08 | 9.310573 |
| NTAN1 | 0.270949 | 1.206601 | 6.864051 | 6.354621 | 1.14E-08 | 5.08E-08 | 9.308171 |
| KAT5 | -0.20664 | 0.866553 | 7.737595 | -6.35155 | 1.15E-08 | 5.15E-08 | 9.295085 |
| FAM193A | 0.312312 | 1.241696 | 5.966329 | 6.349485 | 1.16E-08 | 5.19E-08 | 9.286294 |
| FOCAD | 0.454974 | 1.370758 | 6.401139 | 6.349402 | 1.16E-08 | 5.19E-08 | 9.285937 |
| CUL1 | 0.293103 | 1.225273 | 7.135696 | 6.347905 | 1.17E-08 | 5.22E-08 | 9.279562 |
| DDX39A | 0.290909 | 1.223411 | 6.871519 | 6.342137 | 1.20E-08 | 5.35E-08 | 9.254999 |
| SH3BP2 | -0.30188 | 0.811192 | 6.579114 | -6.34187 | 1.20E-08 | 5.35E-08 | 9.253857 |
| SLC17A1 | -0.70917 | 0.611672 | 9.542025 | -6.34016 | 1.21E-08 | 5.39E-08 | 9.246564 |
| C1orf116 | 0.299196 | 1.230459 | 5.290633 | 6.338742 | 1.22E-08 | 5.42E-08 | 9.240543 |
| FERMT2 | -0.37714 | 0.769962 | 7.762025 | -6.33865 | 1.22E-08 | 5.42E-08 | 9.240155 |
| TG | -0.44302 | 0.735591 | 6.023418 | -6.33847 | 1.22E-08 | 5.42E-08 | 9.2394 |
| IRF1 | 0.757628 | 1.690709 | 6.544304 | 6.337172 | 1.23E-08 | 5.45E-08 | 9.23386 |
| SEPHS2 | -0.33569 | 0.792408 | 10.34304 | -6.33451 | 1.24E-08 | 5.51E-08 | 9.222533 |
| SDC1 | -0.5125 | 0.701005 | 9.544177 | -6.33281 | 1.25E-08 | 5.55E-08 | 9.215275 |
| TNS1 | -0.29202 | 0.81676 | 7.503671 | -6.33224 | 1.25E-08 | 5.56E-08 | 9.212868 |
| ACACA | 0.309941 | 1.239657 | 6.613165 | 6.331468 | 1.26E-08 | 5.57E-08 | 9.209585 |
| INPPL1 | -0.21684 | 0.860445 | 7.150506 | -6.33117 | 1.26E-08 | 5.58E-08 | 9.208299 |
| TBCB | 0.240481 | 1.181386 | 8.829367 | 6.330815 | 1.26E-08 | 5.58E-08 | 9.206804 |
| DHODH | -0.26291 | 0.833408 | 5.091266 | -6.32885 | 1.27E-08 | 5.63E-08 | 9.19846 |
| TMEM248 | 0.236871 | 1.178434 | 8.561772 | 6.327385 | 1.28E-08 | 5.66E-08 | 9.192209 |
| GLYR1 | -0.23974 | 0.8469 | 5.878987 | -6.32707 | 1.28E-08 | 5.67E-08 | 9.190859 |
| NCKAP1L | 0.549084 | 1.463157 | 6.310886 | 6.325799 | 1.29E-08 | 5.70E-08 | 9.185462 |
| TREM2 | 0.446937 | 1.363143 | 6.304304 | 6.321967 | 1.31E-08 | 5.79E-08 | 9.169159 |
| ADD3 | -0.35208 | 0.783453 | 10.0838 | -6.31803 | 1.33E-08 | 5.88E-08 | 9.152429 |
| HSPA4L | -0.44593 | 0.734112 | 6.873291 | -6.31779 | 1.33E-08 | 5.89E-08 | 9.1514 |
| SH2D1A | 0.337385 | 1.263464 | 4.515063 | 6.315884 | 1.35E-08 | 5.93E-08 | 9.143292 |
| RSRP1 | 0.485474 | 1.400046 | 7.288228 | 6.315768 | 1.35E-08 | 5.93E-08 | 9.142797 |
| C11orf73 | 0.296212 | 1.227916 | 4.468734 | 6.314876 | 1.35E-08 | 5.95E-08 | 9.139004 |
| APOA1 | -0.30918 | 0.807102 | 5.120633 | -6.31127 | 1.37E-08 | 6.04E-08 | 9.123658 |
| HOXA10 | 0.337971 | 1.263978 | 6.979873 | 6.309536 | 1.38E-08 | 6.09E-08 | 9.116303 |
| HIP1R | -0.30501 | 0.809435 | 6.130633 | -6.30329 | 1.42E-08 | 6.25E-08 | 9.08978 |
| SUCLA2 | -0.4304 | 0.742055 | 8.903038 | -6.30329 | 1.42E-08 | 6.25E-08 | 9.089768 |
| TMED10 | 0.247108 | 1.186826 | 9.551266 | 6.29999 | 1.44E-08 | 6.33E-08 | 9.075737 |
| PTPRK | 0.381008 | 1.302251 | 9.126329 | 6.299808 | 1.44E-08 | 6.33E-08 | 9.074966 |

| | | | | | | | |
|---|---|---|---|---|---|---|---|
| RNF139 | 0.260817 | 1.198157 | 8.240253 | 6.299722 | 1.44E-08 | 6.33E-08 | 9.074602 |
| FAR2 | 0.415929 | 1.334157 | 5.284177 | 6.29844 | 1.45E-08 | 6.37E-08 | 9.069153 |
| MPHOSPH | 0.246206 | 1.186083 | 8.149367 | 6.297251 | 1.46E-08 | 6.39E-08 | 9.064104 |
| GCNT3 | 0.746001 | 1.677138 | 6.035316 | 6.297201 | 1.46E-08 | 6.39E-08 | 9.063891 |
| CXCL10 | 1.234868 | 2.353599 | 6.351266 | 6.296555 | 1.46E-08 | 6.41E-08 | 9.061145 |
| ZBTB7B | -0.28512 | 0.820674 | 5.383291 | -6.29312 | 1.48E-08 | 6.50E-08 | 9.046564 |
| GPN3 | -0.23372 | 0.850438 | 7.902152 | -6.29253 | 1.49E-08 | 6.51E-08 | 9.044034 |
| FAM212B | -0.27385 | 0.827111 | 5.794304 | -6.29056 | 1.50E-08 | 6.57E-08 | 9.035702 |
| JADE3 | 0.312095 | 1.241509 | 5.266456 | 6.288931 | 1.51E-08 | 6.61E-08 | 9.028769 |
| ACADVL | -0.28094 | 0.823057 | 10.52873 | -6.28884 | 1.51E-08 | 6.61E-08 | 9.028376 |
| LAX1 | 0.374071 | 1.296005 | 4.315823 | 6.288672 | 1.51E-08 | 6.61E-08 | 9.027668 |
| S100A8 | 2.071601 | 4.203528 | 6.85557 | 6.286271 | 1.53E-08 | 6.68E-08 | 9.017475 |
| IGFBP4 | -0.56012 | 0.678246 | 9.483418 | -6.28598 | 1.53E-08 | 6.68E-08 | 9.016229 |
| WIF1 | -0.35969 | 0.779332 | 4.138228 | -6.28593 | 1.53E-08 | 6.68E-08 | 9.016017 |
| PIK3C2B | 0.332503 | 1.259196 | 7.400633 | 6.282026 | 1.56E-08 | 6.79E-08 | 8.999455 |
| ABCG1 | 0.439058 | 1.355719 | 5.841013 | 6.280928 | 1.57E-08 | 6.82E-08 | 8.994797 |
| GDAP2 | -0.23823 | 0.847786 | 5.441139 | -6.27812 | 1.58E-08 | 6.90E-08 | 8.982869 |
| BCL11A | 0.271535 | 1.207091 | 4.668861 | 6.277336 | 1.59E-08 | 6.92E-08 | 8.97955 |
| RAB8B | 0.329407 | 1.256497 | 6.664557 | 6.27516 | 1.60E-08 | 6.98E-08 | 8.970318 |
| IRF2BP1 | -0.2481 | 0.842007 | 7.346582 | -6.27513 | 1.61E-08 | 6.98E-08 | 8.970176 |
| CDADC1 | -0.3027 | 0.810733 | 5.427468 | -6.27507 | 1.61E-08 | 6.98E-08 | 8.969949 |
| MAGED1 | 0.253939 | 1.192459 | 9.886076 | 6.274948 | 1.61E-08 | 6.98E-08 | 8.969419 |
| SERPINA5 | -0.62212 | 0.649715 | 10.07013 | -6.27348 | 1.62E-08 | 7.02E-08 | 8.963197 |
| ALDH8A1 | -0.59697 | 0.661141 | 10.17063 | -6.27287 | 1.62E-08 | 7.03E-08 | 8.960597 |
| GPC3 | -0.91132 | 0.531697 | 7.59519 | -6.27186 | 1.63E-08 | 7.06E-08 | 8.956319 |
| ARF3 | -0.21802 | 0.859742 | 8.732405 | -6.26884 | 1.65E-08 | 7.15E-08 | 8.943526 |
| STS | 0.323603 | 1.251452 | 5.222785 | 6.268359 | 1.65E-08 | 7.16E-08 | 8.941469 |
| FEV | -0.29877 | 0.812946 | 5.080633 | -6.2681 | 1.65E-08 | 7.16E-08 | 8.940372 |
| VAMP5 | 0.479967 | 1.394712 | 6.558101 | 6.266006 | 1.67E-08 | 7.22E-08 | 8.931492 |
| NEU1 | 0.405711 | 1.324742 | 8.640127 | 6.265511 | 1.67E-08 | 7.24E-08 | 8.929394 |
| XYLB | -0.1981 | 0.871696 | 3.848987 | -6.26056 | 1.71E-08 | 7.39E-08 | 8.9084 |
| IER5 | 0.641159 | 1.559582 | 5.9 | 6.259649 | 1.72E-08 | 7.41E-08 | 8.904537 |
| ADGRG1 | 0.483893 | 1.398513 | 8.572785 | 6.258264 | 1.73E-08 | 7.46E-08 | 8.898669 |
| PCDH1 | -0.23831 | 0.84774 | 5.219367 | -6.25557 | 1.75E-08 | 7.54E-08 | 8.887269 |
| SSPN | 0.36222 | 1.285402 | 5.118481 | 6.254083 | 1.76E-08 | 7.58E-08 | 8.880945 |
| STK17B | 0.322062 | 1.250116 | 4.288228 | 6.252563 | 1.77E-08 | 7.63E-08 | 8.874505 |
| ZNF14 | 0.274137 | 1.209271 | 6.778861 | 6.250315 | 1.79E-08 | 7.70E-08 | 8.86498 |
| NDUFB11 | -0.21617 | 0.860846 | 9.42557 | -6.24952 | 1.79E-08 | 7.72E-08 | 8.861625 |
| FSCN1 | 0.256271 | 1.194388 | 5.768354 | 6.249425 | 1.79E-08 | 7.72E-08 | 8.861211 |
| NEDD4L | -0.33812 | 0.791074 | 7.583544 | -6.24911 | 1.80E-08 | 7.73E-08 | 8.859892 |
| EFNB2 | 0.440777 | 1.357335 | 6.134557 | 6.245131 | 1.83E-08 | 7.86E-08 | 8.84302 |
| SOX4 | 0.823373 | 1.769538 | 7.46481 | 6.244596 | 1.83E-08 | 7.87E-08 | 8.840751 |
| ATOX1 | -0.29558 | 0.814745 | 10.35544 | -6.24454 | 1.83E-08 | 7.87E-08 | 8.840529 |
| RPL13A | 0.196146 | 1.145634 | 13.67215 | 6.243516 | 1.84E-08 | 7.90E-08 | 8.836177 |
| SLC6A20 | 0.506462 | 1.420563 | 4.568734 | 6.24219 | 1.85E-08 | 7.94E-08 | 8.830561 |
| SLC35G2 | 0.292826 | 1.225038 | 4.739494 | 6.240846 | 1.86E-08 | 7.99E-08 | 8.824871 |
| PPP1R11 | -0.23689 | 0.848572 | 9.362785 | -6.23678 | 1.89E-08 | 8.12E-08 | 8.80766 |

| | | | | | | | |
|---|---|---|---|---|---|---|---|
| TUBB4B | -0.34559 | 0.786988 | 10.61608 | -6.23621 | 1.90E-08 | 8.14E-08 | 8.805247 |
| APEH | -0.28216 | 0.822358 | 8.661266 | -6.2302 | 1.95E-08 | 8.35E-08 | 8.779786 |
| FYB | 0.353274 | 1.277456 | 5.251266 | 6.227206 | 1.97E-08 | 8.45E-08 | 8.767129 |
| TBC1D16 | 0.432661 | 1.349721 | 6.455949 | 6.225682 | 1.99E-08 | 8.50E-08 | 8.760682 |
| CLMN | -0.37327 | 0.772028 | 7.44038 | -6.21849 | 2.05E-08 | 8.77E-08 | 8.730263 |
| MRPL4 | -0.25931 | 0.835488 | 7.131899 | -6.21532 | 2.08E-08 | 8.88E-08 | 8.716832 |
| VAC14 | -0.21756 | 0.860017 | 6.336076 | -6.21445 | 2.09E-08 | 8.91E-08 | 8.713178 |
| LZTS3 | -0.28783 | 0.819132 | 8.559114 | -6.21427 | 2.09E-08 | 8.92E-08 | 8.712429 |
| ACTL8 | -0.31559 | 0.803521 | 6.730127 | -6.21361 | 2.09E-08 | 8.94E-08 | 8.709621 |
| ADRA2A | 0.430231 | 1.347449 | 4.336456 | 6.212888 | 2.10E-08 | 8.96E-08 | 8.706569 |
| LONP1 | -0.27612 | 0.825809 | 8.218354 | -6.21224 | 2.11E-08 | 8.98E-08 | 8.703843 |
| PHACTR2 | 0.345046 | 1.270192 | 5.778481 | 6.212042 | 2.11E-08 | 8.98E-08 | 8.702992 |
| CLDN1 | 0.539163 | 1.45313 | 5.288481 | 6.211916 | 2.11E-08 | 8.98E-08 | 8.702459 |
| KITLG | 0.405843 | 1.324863 | 4.709747 | 6.210807 | 2.12E-08 | 9.02E-08 | 8.697768 |
| SETD1B | 0.278768 | 1.213159 | 8.911013 | 6.209179 | 2.13E-08 | 9.08E-08 | 8.690888 |
| TRA2A | 0.365296 | 1.288146 | 8.199114 | 6.208468 | 2.14E-08 | 9.10E-08 | 8.687883 |
| SPON1 | 0.503208 | 1.417362 | 5.029114 | 6.207605 | 2.15E-08 | 9.13E-08 | 8.684235 |
| ATAD2B | 0.263788 | 1.200627 | 4.87519 | 6.206545 | 2.16E-08 | 9.17E-08 | 8.679753 |
| IGLV2-14 | 1.535652 | 2.899195 | 8.055823 | 6.203392 | 2.19E-08 | 9.29E-08 | 8.66643 |
| SGMS1 | 0.468287 | 1.383466 | 6.261266 | 6.202503 | 2.20E-08 | 9.32E-08 | 8.662674 |
| NCK2 | 0.238557 | 1.179812 | 7.882911 | 6.202432 | 2.20E-08 | 9.32E-08 | 8.662374 |
| CD37 | 0.655 | 1.574616 | 4.808608 | 6.200574 | 2.22E-08 | 9.39E-08 | 8.654519 |
| ANG | -0.50698 | 0.703696 | 7.289747 | -6.19908 | 2.23E-08 | 9.45E-08 | 8.648212 |
| CTCF | 0.277668 | 1.212234 | 7.654684 | 6.196284 | 2.26E-08 | 9.56E-08 | 8.636397 |
| PECR | -0.50321 | 0.705533 | 7.722405 | -6.19464 | 2.27E-08 | 9.62E-08 | 8.62947 |
| PLEK | 0.61193 | 1.528303 | 5.859747 | 6.193826 | 2.28E-08 | 9.65E-08 | 8.626016 |
| RBM12 | 0.311871 | 1.241316 | 6.258101 | 6.192329 | 2.30E-08 | 9.71E-08 | 8.619695 |
| MRPS35 | -0.21252 | 0.863031 | 9.782532 | -6.18791 | 2.34E-08 | 9.89E-08 | 8.601017 |
| IDI1 | -0.3243 | 0.798689 | 8.766709 | -6.18713 | 2.35E-08 | 9.92E-08 | 8.597729 |
| CYTIP | 0.757273 | 1.690292 | 5.106329 | 6.186544 | 2.35E-08 | 9.94E-08 | 8.595265 |
| MTNR1A | -0.60461 | 0.657649 | 5.072658 | -6.18518 | 2.37E-08 | 9.99E-08 | 8.589517 |
| ISCA1 | -0.27781 | 0.824844 | 9.437215 | -6.18486 | 2.37E-08 | 1.00E-07 | 8.588139 |
| EIF4ENIF1 | 0.251087 | 1.190103 | 6.763797 | 6.182925 | 2.39E-08 | 1.01E-07 | 8.579986 |
| IL4R | 0.372905 | 1.294958 | 6.574684 | 6.182454 | 2.40E-08 | 1.01E-07 | 8.578 |
| AKAP1 | -0.1963 | 0.872787 | 6.972785 | -6.18192 | 2.40E-08 | 1.01E-07 | 8.575764 |
| PLA2G12A | -0.25353 | 0.838841 | 6.619747 | -6.17912 | 2.43E-08 | 1.02E-07 | 8.563946 |
| IHH | -0.29328 | 0.816044 | 6.465316 | -6.17692 | 2.45E-08 | 1.03E-07 | 8.554666 |
| MOCS2 | -0.27773 | 0.824886 | 7.760506 | -6.17544 | 2.47E-08 | 1.04E-07 | 8.548421 |
| SIRT3 | -0.22019 | 0.858452 | 6.522152 | -6.17399 | 2.48E-08 | 1.04E-07 | 8.54229 |
| CLDN15 | -0.2691 | 0.829835 | 8.090633 | -6.17326 | 2.49E-08 | 1.05E-07 | 8.539204 |
| ZNF76 | -0.219 | 0.859162 | 6.997215 | -6.17183 | 2.51E-08 | 1.05E-07 | 8.533155 |
| ATP2B4 | 0.218043 | 1.163155 | 5.973038 | 6.170832 | 2.52E-08 | 1.06E-07 | 8.528963 |
| PTPRCAP | 0.47809 | 1.392898 | 5.681013 | 6.169872 | 2.53E-08 | 1.06E-07 | 8.524915 |
| HLA-DQB2 | 0.400935 | 1.320364 | 5.681392 | 6.166356 | 2.57E-08 | 1.08E-07 | 8.510085 |
| RPIA | 0.298327 | 1.229717 | 6.871139 | 6.165919 | 2.57E-08 | 1.08E-07 | 8.508242 |
| DZIP1 | -0.30132 | 0.811507 | 6.59 | -6.1636 | 2.60E-08 | 1.09E-07 | 8.498484 |
| DLGAP5 | 0.347299 | 1.272177 | 3.728987 | 6.16266 | 2.61E-08 | 1.09E-07 | 8.494503 |

| | | | | | | | |
|---|---|---|---|---|---|---|---|
| PNPLA4 | -0.47187 | 0.721029 | 7.945063 | -6.16171 | 2.62E-08 | 1.10E-07 | 8.490494 |
| GARS | 0.268011 | 1.204146 | 8.981519 | 6.161225 | 2.62E-08 | 1.10E-07 | 8.488451 |
| MAP3K13 | -0.22122 | 0.85784 | 5.079114 | -6.16081 | 2.63E-08 | 1.10E-07 | 8.486689 |
| HN1L | 0.245659 | 1.185634 | 7.255443 | 6.160594 | 2.63E-08 | 1.10E-07 | 8.485791 |
| MAML1 | 0.224717 | 1.168548 | 6.814304 | 6.160341 | 2.63E-08 | 1.10E-07 | 8.484723 |
| NEIL1 | -0.21715 | 0.860261 | 5.921899 | -6.1596 | 2.64E-08 | 1.10E-07 | 8.481612 |
| UBA6 | 0.297187 | 1.228746 | 5.473924 | 6.159289 | 2.65E-08 | 1.10E-07 | 8.480291 |
| LSP1 | 0.332708 | 1.259375 | 5.303544 | 6.159185 | 2.65E-08 | 1.10E-07 | 8.479853 |
| PIGP | -0.26538 | 0.831982 | 9.240886 | -6.15754 | 2.67E-08 | 1.11E-07 | 8.4729 |
| ZMAT3 | 0.261653 | 1.198852 | 6.537342 | 6.155541 | 2.69E-08 | 1.12E-07 | 8.464495 |
| CDCP1 | 0.265758 | 1.202267 | 4.891013 | 6.155401 | 2.69E-08 | 1.12E-07 | 8.463903 |
| PIP4K2A | 0.271976 | 1.207461 | 6.837089 | 6.155034 | 2.70E-08 | 1.12E-07 | 8.462359 |
| GZMB | 0.763188 | 1.697237 | 4.402278 | 6.154408 | 2.70E-08 | 1.12E-07 | 8.459721 |
| ZNF292 | 0.305408 | 1.235768 | 5.948228 | 6.154289 | 2.70E-08 | 1.12E-07 | 8.45922 |
| VWF | 0.478366 | 1.393165 | 5.777215 | 6.15113 | 2.74E-08 | 1.14E-07 | 8.445906 |
| CAST | 0.284282 | 1.217804 | 8.70962 | 6.149842 | 2.76E-08 | 1.14E-07 | 8.440479 |
| CELF3 | -0.23326 | 0.85071 | 6.205823 | -6.14568 | 2.81E-08 | 1.17E-07 | 8.422962 |
| RSU1 | 0.35888 | 1.28243 | 7.298608 | 6.144343 | 2.82E-08 | 1.17E-07 | 8.41732 |
| VEZF1 | 0.27392 | 1.209088 | 7.758987 | 6.143181 | 2.84E-08 | 1.18E-07 | 8.412423 |
| BAZ2B | 0.308142 | 1.238112 | 6.717848 | 6.142727 | 2.84E-08 | 1.18E-07 | 8.410512 |
| MILR1 | 0.332628 | 1.259306 | 4.121772 | 6.139598 | 2.88E-08 | 1.19E-07 | 8.397338 |
| PLCL1 | -0.69916 | 0.615932 | 7.203165 | -6.13957 | 2.88E-08 | 1.19E-07 | 8.397233 |
| INADL | -0.23425 | 0.850127 | 4.673671 | -6.13956 | 2.88E-08 | 1.19E-07 | 8.397191 |
| SEC31A | -0.24625 | 0.843084 | 9.581266 | -6.1374 | 2.91E-08 | 1.20E-07 | 8.38808 |
| EFNA4 | 0.372286 | 1.294402 | 5.527468 | 6.136773 | 2.92E-08 | 1.21E-07 | 8.385445 |
| ATP5D | -0.28089 | 0.823083 | 7.798101 | -6.13613 | 2.92E-08 | 1.21E-07 | 8.382753 |
| TBXAS1 | 0.389934 | 1.310334 | 5.288101 | 6.136105 | 2.92E-08 | 1.21E-07 | 8.382631 |
| INPP4B | 0.35614 | 1.279996 | 4.270506 | 6.135014 | 2.94E-08 | 1.21E-07 | 8.378041 |
| DNAJB5 | -0.37343 | 0.771947 | 5.385316 | -6.13365 | 2.95E-08 | 1.22E-07 | 8.372314 |
| GNAI2 | 0.294545 | 1.226498 | 7.533038 | 6.133253 | 2.96E-08 | 1.22E-07 | 8.370629 |
| FAM65B | 0.354183 | 1.278262 | 4.511646 | 6.126373 | 3.05E-08 | 1.26E-07 | 8.341677 |
| PILRA | 0.296713 | 1.228342 | 4.833291 | 6.126366 | 3.05E-08 | 1.26E-07 | 8.341647 |
| CTDP1 | -0.24537 | 0.8436 | 6.277722 | -6.12239 | 3.10E-08 | 1.28E-07 | 8.32494 |
| SUV420H1 | 0.38581 | 1.306593 | 7.088987 | 6.121296 | 3.12E-08 | 1.28E-07 | 8.320323 |
| IL2RG | 0.576746 | 1.491481 | 6.132658 | 6.120564 | 3.13E-08 | 1.29E-07 | 8.317245 |
| SPAST | 0.237134 | 1.178649 | 4.861013 | 6.120458 | 3.13E-08 | 1.29E-07 | 8.3168 |
| BACH2 | 0.22025 | 1.164936 | 4.244177 | 6.120033 | 3.13E-08 | 1.29E-07 | 8.315011 |
| EIF4EBP2 | -0.24371 | 0.844571 | 7.444937 | -6.11964 | 3.14E-08 | 1.29E-07 | 8.313365 |
| CAP2 | 0.437945 | 1.354673 | 6.917722 | 6.119474 | 3.14E-08 | 1.29E-07 | 8.31266 |
| FCGRT | -0.3584 | 0.78003 | 9.490506 | -6.11683 | 3.18E-08 | 1.30E-07 | 8.301523 |
| LONRF3 | -0.40729 | 0.754041 | 5.642911 | -6.11597 | 3.19E-08 | 1.31E-07 | 8.29793 |
| ZNF638 | 0.306456 | 1.236666 | 8.036709 | 6.11321 | 3.23E-08 | 1.32E-07 | 8.286322 |
| GBP1 | 0.77307 | 1.708902 | 6.393797 | 6.109566 | 3.28E-08 | 1.34E-07 | 8.271008 |
| TARS | 0.274578 | 1.209641 | 8.097089 | 6.108628 | 3.29E-08 | 1.35E-07 | 8.267066 |
| C9orf78 | -0.22265 | 0.856987 | 7.760253 | -6.10413 | 3.35E-08 | 1.37E-07 | 8.248178 |
| EZH2 | 0.3639 | 1.2869 | 4.141139 | 6.102954 | 3.37E-08 | 1.38E-07 | 8.243221 |
| MAPK8 | -0.26536 | 0.831994 | 6.156329 | -6.10161 | 3.39E-08 | 1.39E-07 | 8.237585 |

| | | | | | | | |
|---|---|---|---|---|---|---|---|
| SEMA4C | 0.236864 | 1.178429 | 7.023291 | 6.101421 | 3.39E-08 | 1.39E-07 | 8.236784 |
| COL16A1 | 0.526805 | 1.440735 | 5.198101 | 6.101357 | 3.39E-08 | 1.39E-07 | 8.236511 |
| TFDP2 | -0.24194 | 0.845609 | 7.366329 | -6.09882 | 3.43E-08 | 1.40E-07 | 8.225867 |
| BBOX1 | -0.57518 | 0.671203 | 11.60582 | -6.09408 | 3.50E-08 | 1.43E-07 | 8.205934 |
| CD59 | 0.29585 | 1.227608 | 10.48228 | 6.094038 | 3.50E-08 | 1.43E-07 | 8.205777 |
| COPS7A | -0.23655 | 0.84877 | 8.135316 | -6.09391 | 3.50E-08 | 1.43E-07 | 8.205249 |
| FZD2 | 0.319137 | 1.247584 | 4.682658 | 6.093425 | 3.51E-08 | 1.43E-07 | 8.203202 |
| RPL39L | 0.301186 | 1.232157 | 4.471899 | 6.093253 | 3.51E-08 | 1.43E-07 | 8.202477 |
| BACE1 | -0.22352 | 0.856471 | 6.730759 | -6.09304 | 3.52E-08 | 1.43E-07 | 8.201585 |
| ADNP | 0.329743 | 1.25679 | 7.857089 | 6.091921 | 3.53E-08 | 1.44E-07 | 8.196885 |
| S100A9 | 0.97552 | 1.96635 | 5.372278 | 6.091099 | 3.55E-08 | 1.44E-07 | 8.193438 |
| PAQR5 | -0.39138 | 0.762398 | 6.926076 | -6.09042 | 3.56E-08 | 1.45E-07 | 8.19057 |
| CLIC5 | -0.71069 | 0.611027 | 6.492911 | -6.08674 | 3.61E-08 | 1.47E-07 | 8.175157 |
| GCA | 0.346713 | 1.27166 | 6.134177 | 6.086605 | 3.62E-08 | 1.47E-07 | 8.17457 |
| SEC24D | 0.329256 | 1.256365 | 4.179494 | 6.085094 | 3.64E-08 | 1.48E-07 | 8.168228 |
| RECK | 0.390395 | 1.310752 | 5.121772 | 6.084613 | 3.65E-08 | 1.48E-07 | 8.166214 |
| CDH9 | -0.26129 | 0.834341 | 5.329114 | -6.07373 | 3.82E-08 | 1.55E-07 | 8.120555 |
| PLCH2 | -0.362 | 0.778084 | 5.521392 | -6.07014 | 3.88E-08 | 1.57E-07 | 8.105523 |
| INHA | -0.25496 | 0.83801 | 4.589367 | -6.06905 | 3.90E-08 | 1.58E-07 | 8.100936 |
| RPARP-AS1 | -0.34248 | 0.788682 | 5.590633 | -6.0627 | 4.01E-08 | 1.62E-07 | 8.074328 |
| DGAT1 | -0.24043 | 0.846494 | 6.658481 | -6.05837 | 4.08E-08 | 1.65E-07 | 8.056187 |
| HIRIP3 | -0.21383 | 0.862247 | 7.37481 | -6.05759 | 4.09E-08 | 1.66E-07 | 8.052903 |
| CLCN7 | -0.19773 | 0.871923 | 7.102405 | -6.05628 | 4.12E-08 | 1.67E-07 | 8.04743 |
| MIF | -0.25006 | 0.840862 | 11.73924 | -6.05396 | 4.16E-08 | 1.68E-07 | 8.0377 |
| NDNF | -1.1043 | 0.46513 | 6.630886 | -6.04868 | 4.25E-08 | 1.72E-07 | 8.015576 |
| CXorf21 | 0.349335 | 1.273973 | 3.82962 | 6.047985 | 4.27E-08 | 1.72E-07 | 8.012675 |
| SRD5A2 | -0.24966 | 0.841096 | 5.386582 | -6.04686 | 4.29E-08 | 1.73E-07 | 8.007975 |
| MT1X | -1.02463 | 0.491536 | 11.62177 | -6.04606 | 4.30E-08 | 1.74E-07 | 8.004604 |
| WIPI1 | -0.43675 | 0.738799 | 8.065823 | -6.04554 | 4.31E-08 | 1.74E-07 | 8.002444 |
| THOC1 | 0.238327 | 1.179624 | 7.356076 | 6.044858 | 4.32E-08 | 1.74E-07 | 7.999584 |
| FUS | 0.277339 | 1.211957 | 7.280633 | 6.042219 | 4.37E-08 | 1.76E-07 | 7.988534 |
| DMXL1 | -0.5793 | 0.669291 | 8.17519 | -6.03924 | 4.43E-08 | 1.78E-07 | 7.976063 |
| ERAP1 | 0.270059 | 1.205857 | 5.179114 | 6.038872 | 4.44E-08 | 1.79E-07 | 7.974528 |
| CD1C | 0.594526 | 1.509976 | 6.472911 | 6.038384 | 4.44E-08 | 1.79E-07 | 7.972486 |
| DDAH1 | -0.24235 | 0.845366 | 11.00203 | -6.03682 | 4.47E-08 | 1.80E-07 | 7.96595 |
| ZWILCH | 0.281581 | 1.215526 | 4.762405 | 6.036229 | 4.49E-08 | 1.80E-07 | 7.963467 |
| NUP107 | 0.310547 | 1.240178 | 7.573418 | 6.033228 | 4.54E-08 | 1.83E-07 | 7.950914 |
| ING2 | -0.30795 | 0.807788 | 6.798101 | -6.0319 | 4.57E-08 | 1.84E-07 | 7.945369 |
| ZZZ3 | 0.39193 | 1.312148 | 7.087848 | 6.031608 | 4.58E-08 | 1.84E-07 | 7.944139 |
| NR1I3 | -0.71414 | 0.60957 | 5.127342 | -6.03136 | 4.58E-08 | 1.84E-07 | 7.943118 |
| NPPA | -0.30691 | 0.808371 | 6.088101 | -6.03117 | 4.58E-08 | 1.84E-07 | 7.942317 |
| OAS2 | 0.480968 | 1.39568 | 5.107215 | 6.029294 | 4.62E-08 | 1.85E-07 | 7.934458 |
| NR2F6 | -0.21582 | 0.861058 | 6.573544 | -6.02815 | 4.64E-08 | 1.86E-07 | 7.929679 |
| GALNT10 | 0.253689 | 1.192252 | 6.143797 | 6.027192 | 4.66E-08 | 1.87E-07 | 7.925667 |
| CETN3 | 0.422866 | 1.340588 | 7.424684 | 6.025647 | 4.69E-08 | 1.88E-07 | 7.919208 |
| PCBD1 | -0.42728 | 0.743663 | 9.173038 | -6.02467 | 4.71E-08 | 1.89E-07 | 7.915137 |
| RRM1 | 0.264809 | 1.201477 | 6.539747 | 6.023025 | 4.75E-08 | 1.90E-07 | 7.908247 |

| | | | | | | | |
|---|---|---|---|---|---|---|---|
| ELOVL6 | 0.323202 | 1.251104 | 5.245443 | 6.021663 | 4.77E-08 | 1.91E-07 | 7.90255 |
| PIGN | 0.265092 | 1.201713 | 5.93443 | 6.019242 | 4.82E-08 | 1.93E-07 | 7.892434 |
| COX7B | -0.27716 | 0.825213 | 11.31835 | -6.01798 | 4.85E-08 | 1.94E-07 | 7.887163 |
| CSHL1 | -0.33302 | 0.793871 | 5.429367 | -6.01592 | 4.89E-08 | 1.95E-07 | 7.878532 |
| ERMP1 | 0.420711 | 1.338588 | 8.451392 | 6.0137 | 4.94E-08 | 1.97E-07 | 7.869272 |
| USP1 | 0.283913 | 1.217493 | 7.234684 | 6.009309 | 5.03E-08 | 2.01E-07 | 7.850928 |
| CEBPA-AS1 | -0.29616 | 0.814418 | 6.489114 | -6.00644 | 5.09E-08 | 2.03E-07 | 7.838948 |
| HMX1 | -0.31927 | 0.801476 | 6.437722 | -6.00348 | 5.16E-08 | 2.06E-07 | 7.826566 |
| ECHDC1 | -0.34834 | 0.785487 | 8.951013 | -6.00321 | 5.17E-08 | 2.06E-07 | 7.825441 |
| NMT2 | 0.439631 | 1.356257 | 5.968861 | 6.001327 | 5.21E-08 | 2.07E-07 | 7.817595 |
| SERPINB9 | 0.491805 | 1.406203 | 5.658481 | 6.000899 | 5.22E-08 | 2.08E-07 | 7.81581 |
| PHF1 | -0.19834 | 0.871553 | 8.313671 | -6.00069 | 5.22E-08 | 2.08E-07 | 7.81493 |
| POGLUT1 | 0.266542 | 1.202921 | 6.580253 | 5.994235 | 5.37E-08 | 2.13E-07 | 7.787995 |
| MAT2A | -0.56788 | 0.674608 | 7.632785 | -5.98922 | 5.48E-08 | 2.18E-07 | 7.767088 |
| IFI44L | 1.324275 | 2.504071 | 5.99557 | 5.989014 | 5.49E-08 | 2.18E-07 | 7.766213 |
| C18orf8 | -0.3275 | 0.796914 | 7.106456 | -5.98739 | 5.53E-08 | 2.19E-07 | 7.759455 |
| SKAP2 | 0.284539 | 1.218021 | 7.26038 | 5.98722 | 5.53E-08 | 2.19E-07 | 7.758727 |
| HADHA | -0.18715 | 0.878337 | 9.39443 | -5.98561 | 5.57E-08 | 2.21E-07 | 7.752003 |
| TRIM24 | 0.331212 | 1.25807 | 6.118354 | 5.984853 | 5.59E-08 | 2.22E-07 | 7.748857 |
| APBB1 | -0.25187 | 0.839807 | 5.746962 | -5.9837 | 5.61E-08 | 2.22E-07 | 7.744048 |
| NPHS2 | -1.26985 | 0.414703 | 8.474557 | -5.98218 | 5.65E-08 | 2.24E-07 | 7.737695 |
| HMGXB4 | 0.224756 | 1.16858 | 6.47519 | 5.981578 | 5.66E-08 | 2.24E-07 | 7.735201 |
| RALGDS | 0.23442 | 1.176434 | 6.006835 | 5.980461 | 5.69E-08 | 2.25E-07 | 7.730541 |
| HOXB1 | -0.32958 | 0.795765 | 5.816456 | -5.98026 | 5.70E-08 | 2.25E-07 | 7.729703 |
| IFI6 | 1.239545 | 2.361241 | 8.112785 | 5.980195 | 5.70E-08 | 2.25E-07 | 7.729431 |
| LRRC1 | 0.395968 | 1.315826 | 7.526709 | 5.978731 | 5.73E-08 | 2.27E-07 | 7.723328 |
| ID3 | 0.580982 | 1.495867 | 6.665949 | 5.977402 | 5.77E-08 | 2.28E-07 | 7.71779 |
| PSMD1 | 0.199592 | 1.148373 | 8.667722 | 5.975551 | 5.81E-08 | 2.29E-07 | 7.710074 |
| PCOLCE2 | -1.008 | 0.497236 | 5.676329 | -5.97523 | 5.82E-08 | 2.30E-07 | 7.708737 |
| FOXN1 | -0.21466 | 0.861751 | 4.896203 | -5.97278 | 5.88E-08 | 2.32E-07 | 7.698526 |
| PRKAG2 | -0.34682 | 0.786316 | 7.220127 | -5.97261 | 5.89E-08 | 2.32E-07 | 7.697806 |
| TLR8 | 0.392009 | 1.31222 | 4.29962 | 5.970005 | 5.95E-08 | 2.35E-07 | 7.68696 |
| TRA2B | 0.216621 | 1.162008 | 6.541139 | 5.968152 | 6.00E-08 | 2.36E-07 | 7.679241 |
| POLE2 | 0.354045 | 1.278139 | 4.963544 | 5.965899 | 6.06E-08 | 2.38E-07 | 7.669856 |
| IRF2 | 0.259987 | 1.197468 | 6.331646 | 5.963197 | 6.13E-08 | 2.41E-07 | 7.6586 |
| HECW1 | -0.28411 | 0.821248 | 4.870886 | -5.963 | 6.13E-08 | 2.41E-07 | 7.657768 |
| VWA8 | -0.3554 | 0.781655 | 7.277848 | -5.95642 | 6.31E-08 | 2.48E-07 | 7.630384 |
| ANKRD6 | 0.26807 | 1.204196 | 5.407848 | 5.956129 | 6.31E-08 | 2.48E-07 | 7.629171 |
| ZCCHC14 | -0.21063 | 0.864162 | 5.926582 | -5.95601 | 6.32E-08 | 2.48E-07 | 7.628695 |
| POLR1E | 0.301601 | 1.232511 | 6.336203 | 5.95254 | 6.41E-08 | 2.52E-07 | 7.614236 |
| UBE2J1 | 0.312451 | 1.241815 | 6.45443 | 5.952194 | 6.42E-08 | 2.52E-07 | 7.612796 |
| RPP40 | 0.294901 | 1.226801 | 6.591013 | 5.951715 | 6.43E-08 | 2.52E-07 | 7.610803 |
| PDGFRA | 0.591225 | 1.506526 | 7.013924 | 5.950182 | 6.47E-08 | 2.54E-07 | 7.60442 |
| PCDH17 | 0.364539 | 1.28747 | 5.283797 | 5.949738 | 6.49E-08 | 2.54E-07 | 7.602574 |
| PGRMC1 | -0.31347 | 0.804707 | 10.93949 | -5.946 | 6.59E-08 | 2.58E-07 | 7.587041 |
| CASP8 | 0.246897 | 1.186652 | 5.839873 | 5.940374 | 6.75E-08 | 2.64E-07 | 7.563622 |
| DMTN | -0.22946 | 0.852954 | 7.169367 | -5.93968 | 6.77E-08 | 2.65E-07 | 7.560727 |

| | | | | | | | |
|---|---|---|---|---|---|---|---|
| PBK | 0.484104 | 1.398717 | 3.824177 | 5.93308 | 6.96E-08 | 2.72E-07 | 7.533298 |
| ELF4 | 0.335296 | 1.261637 | 5.486582 | 5.932674 | 6.98E-08 | 2.73E-07 | 7.531609 |
| GFPT1 | 0.340336 | 1.266051 | 7.374557 | 5.93255 | 6.98E-08 | 2.73E-07 | 7.531093 |
| SLC2A11 | -0.23207 | 0.851413 | 5.170886 | -5.93232 | 6.99E-08 | 2.73E-07 | 7.530159 |
| NOL8 | 0.327833 | 1.255126 | 6.817595 | 5.931632 | 7.01E-08 | 2.74E-07 | 7.527279 |
| PLEKHA6 | -0.27866 | 0.824355 | 6.414684 | -5.92855 | 7.10E-08 | 2.77E-07 | 7.514492 |
| SEMA4D | 0.383241 | 1.304269 | 6.981392 | 5.928234 | 7.11E-08 | 2.77E-07 | 7.513157 |
| PFKFB3 | 0.774137 | 1.710167 | 8.147722 | 5.928048 | 7.11E-08 | 2.77E-07 | 7.512386 |
| GGA2 | -0.20295 | 0.868772 | 7.176962 | -5.92721 | 7.14E-08 | 2.78E-07 | 7.508894 |
| GMDS | 0.282022 | 1.215898 | 6.050633 | 5.926233 | 7.17E-08 | 2.79E-07 | 7.504846 |
| CRISP2 | -0.36556 | 0.776168 | 4.181646 | -5.92418 | 7.23E-08 | 2.82E-07 | 7.49632 |
| COL18A1 | 0.396225 | 1.31606 | 7.657468 | 5.923592 | 7.25E-08 | 2.82E-07 | 7.493873 |
| TGM5 | -0.26539 | 0.831975 | 6.193924 | -5.92354 | 7.25E-08 | 2.82E-07 | 7.493657 |
| XYLT2 | 0.275145 | 1.210116 | 6.076456 | 5.922684 | 7.28E-08 | 2.83E-07 | 7.490101 |
| PSAT1 | -0.6467 | 0.63874 | 9.567468 | -5.92179 | 7.31E-08 | 2.84E-07 | 7.486407 |
| SLC38A6 | -0.48579 | 0.714106 | 5.944684 | -5.92105 | 7.33E-08 | 2.85E-07 | 7.483299 |
| AGO1 | -0.20489 | 0.867602 | 6.735063 | -5.91925 | 7.38E-08 | 2.87E-07 | 7.475819 |
| HSF4 | -0.29924 | 0.812683 | 6.578481 | -5.9166 | 7.47E-08 | 2.90E-07 | 7.464826 |
| MCM7 | 0.193689 | 1.143684 | 6.388734 | 5.913095 | 7.58E-08 | 2.94E-07 | 7.450285 |
| KRT7 | 0.70946 | 1.635192 | 6.391139 | 5.912811 | 7.59E-08 | 2.94E-07 | 7.449106 |
| ZNF646 | -0.27609 | 0.825828 | 5.340759 | -5.91256 | 7.60E-08 | 2.94E-07 | 7.44805 |
| QARS | 0.243676 | 1.184006 | 9.372658 | 5.911909 | 7.62E-08 | 2.95E-07 | 7.445362 |
| EIF5 | -0.2281 | 0.853761 | 8.754937 | -5.90806 | 7.74E-08 | 3.00E-07 | 7.429389 |
| PSD3 | 0.340204 | 1.265936 | 4.504937 | 5.906671 | 7.79E-08 | 3.02E-07 | 7.423626 |
| DVL1 | -0.21697 | 0.860371 | 6.349367 | -5.90539 | 7.83E-08 | 3.03E-07 | 7.418328 |
| NBEA | 0.35058 | 1.275073 | 5.492532 | 5.905148 | 7.84E-08 | 3.03E-07 | 7.417307 |
| PRKD1 | 0.408472 | 1.327279 | 6.713671 | 5.90455 | 7.86E-08 | 3.04E-07 | 7.414825 |
| PIGG | 0.290744 | 1.223271 | 6.999494 | 5.904232 | 7.87E-08 | 3.04E-07 | 7.413506 |
| PRX | -0.27692 | 0.825353 | 6.272152 | -5.90374 | 7.89E-08 | 3.05E-07 | 7.41146 |
| SLC38A1 | 0.523702 | 1.43764 | 7.613544 | 5.902941 | 7.91E-08 | 3.05E-07 | 7.408151 |
| ZFAND1 | -0.32984 | 0.795627 | 7.744177 | -5.89882 | 8.05E-08 | 3.11E-07 | 7.391077 |
| ADAM17 | 0.239216 | 1.180351 | 5.021013 | 5.89668 | 8.13E-08 | 3.13E-07 | 7.382182 |
| TFEC | -0.4859 | 0.714054 | 7.258987 | -5.89599 | 8.15E-08 | 3.14E-07 | 7.379318 |
| DIMT1 | 0.215046 | 1.160741 | 7.084177 | 5.889706 | 8.37E-08 | 3.23E-07 | 7.353275 |
| TRIL | 0.381686 | 1.302864 | 5.799873 | 5.888602 | 8.41E-08 | 3.24E-07 | 7.348699 |
| ZNF787 | -0.267 | 0.831044 | 5.466076 | -5.88856 | 8.41E-08 | 3.24E-07 | 7.34851 |
| NUCB2 | 0.384097 | 1.305043 | 6.823924 | 5.886338 | 8.49E-08 | 3.27E-07 | 7.339315 |
| PRKX | 0.493037 | 1.407404 | 4.92443 | 5.881814 | 8.66E-08 | 3.33E-07 | 7.320573 |
| CCR7 | 0.453123 | 1.369 | 4.497975 | 5.880002 | 8.72E-08 | 3.35E-07 | 7.31307 |
| QTRT1 | -0.25292 | 0.839193 | 6.96 | -5.87923 | 8.75E-08 | 3.36E-07 | 7.309861 |
| CRYBG3 | 0.222497 | 1.166751 | 7.156203 | 5.875664 | 8.88E-08 | 3.41E-07 | 7.295104 |
| CACNB3 | 0.228841 | 1.171893 | 5.473418 | 5.869819 | 9.11E-08 | 3.50E-07 | 7.270907 |
| SCAND1 | -0.22471 | 0.855767 | 8.704177 | -5.86868 | 9.15E-08 | 3.51E-07 | 7.266186 |
| MFSD1 | 0.264091 | 1.200879 | 9.515316 | 5.865388 | 9.28E-08 | 3.56E-07 | 7.252571 |
| NKX2-5 | -0.23091 | 0.852098 | 5.203544 | -5.86348 | 9.35E-08 | 3.59E-07 | 7.244663 |
| PLS3 | 0.31587 | 1.244762 | 9.936076 | 5.863369 | 9.36E-08 | 3.59E-07 | 7.244215 |
| TRBC1 | 0.976331 | 1.967455 | 6.875443 | 5.861251 | 9.44E-08 | 3.62E-07 | 7.235455 |

| | | | | | | | |
|---|---|---|---|---|---|---|---|
| PEMT | -0.51422 | 0.700173 | 7.709114 | -5.86017 | 9.49E-08 | 3.63E-07 | 7.230989 |
| FASTKD3 | -0.40895 | 0.75317 | 7.287215 | -5.85969 | 9.51E-08 | 3.64E-07 | 7.229017 |
| MLLT11 | 0.339868 | 1.265641 | 5.772405 | 5.85935 | 9.52E-08 | 3.64E-07 | 7.22759 |
| ZXDC | -0.20783 | 0.865837 | 6.442532 | -5.8588 | 9.54E-08 | 3.65E-07 | 7.225335 |
| PDLIM4 | 0.239447 | 1.18054 | 4.597848 | 5.855979 | 9.66E-08 | 3.69E-07 | 7.213652 |
| TTLL7 | 0.375211 | 1.297029 | 4.713038 | 5.855741 | 9.67E-08 | 3.69E-07 | 7.212666 |
| MT1F | -0.66308 | 0.63153 | 12.14367 | -5.85424 | 9.73E-08 | 3.71E-07 | 7.206449 |
| P2RY13 | 0.522899 | 1.436839 | 4.528861 | 5.85193 | 9.82E-08 | 3.75E-07 | 7.196912 |
| TTC39A | 0.857009 | 1.81128 | 6.258861 | 5.846131 | 1.01E-07 | 3.84E-07 | 7.172946 |
| IL33 | 0.471989 | 1.387021 | 3.977595 | 5.846111 | 1.01E-07 | 3.84E-07 | 7.172861 |
| VLDLR | 0.401364 | 1.320756 | 6.522658 | 5.844195 | 1.01E-07 | 3.87E-07 | 7.164945 |
| ZNF423 | 0.335415 | 1.26174 | 4.669241 | 5.841608 | 1.03E-07 | 3.91E-07 | 7.154256 |
| MRPS33 | -0.23285 | 0.850951 | 9.011646 | -5.84099 | 1.03E-07 | 3.92E-07 | 7.151723 |
| CACNA2D3 | 0.425264 | 1.342818 | 4.531772 | 5.835858 | 1.05E-07 | 4.00E-07 | 7.130509 |
| ACTR1B | -0.21341 | 0.862495 | 7.809114 | -5.83549 | 1.05E-07 | 4.01E-07 | 7.12899 |
| TNFRSF11B | 0.660033 | 1.580119 | 7.428101 | 5.834567 | 1.06E-07 | 4.02E-07 | 7.12518 |
| LSM4 | -0.26195 | 0.83396 | 8.821013 | -5.83426 | 1.06E-07 | 4.02E-07 | 7.123916 |
| C10orf10 | 0.41915 | 1.33714 | 6.541392 | 5.833904 | 1.06E-07 | 4.03E-07 | 7.12244 |
| SCML1 | -0.31553 | 0.803557 | 5.314937 | -5.83374 | 1.06E-07 | 4.03E-07 | 7.121758 |
| SDHAF3 | -0.27094 | 0.828782 | 5.912911 | -5.8326 | 1.07E-07 | 4.05E-07 | 7.117044 |
| FAM149A | -0.55646 | 0.679971 | 7.438861 | -5.83126 | 1.07E-07 | 4.07E-07 | 7.111523 |
| UHRF1BP1 | -0.23944 | 0.847074 | 5.310633 | -5.82778 | 1.09E-07 | 4.12E-07 | 7.097165 |
| TBX2 | -0.24655 | 0.842907 | 6.491139 | -5.82775 | 1.09E-07 | 4.12E-07 | 7.097042 |
| C19orf24 | -0.22043 | 0.858311 | 7.336835 | -5.82696 | 1.09E-07 | 4.14E-07 | 7.093758 |
| CDH15 | -0.23361 | 0.850504 | 4.806329 | -5.82612 | 1.10E-07 | 4.15E-07 | 7.090317 |
| SHFM1 | 0.223755 | 1.167769 | 7.056076 | 5.823689 | 1.11E-07 | 4.19E-07 | 7.080282 |
| TNFRSF21 | -0.50026 | 0.706981 | 8.83038 | -5.8231 | 1.11E-07 | 4.20E-07 | 7.077852 |
| UBE2Z | 0.188656 | 1.139702 | 8.389241 | 5.819363 | 1.13E-07 | 4.26E-07 | 7.062437 |
| MAPK12 | -0.33495 | 0.792809 | 5.63443 | -5.81737 | 1.14E-07 | 4.30E-07 | 7.054228 |
| HOOK2 | -0.26516 | 0.832108 | 8.100759 | -5.81079 | 1.17E-07 | 4.42E-07 | 7.027092 |
| DNAJB1 | -0.77923 | 0.582678 | 9.106456 | -5.80969 | 1.17E-07 | 4.44E-07 | 7.02255 |
| IQSEC3 | -0.25523 | 0.837853 | 5.471646 | -5.80882 | 1.18E-07 | 4.45E-07 | 7.018958 |
| PTPRD | -0.44946 | 0.732317 | 7.327468 | -5.80776 | 1.18E-07 | 4.47E-07 | 7.01458 |
| BID | 0.332951 | 1.259587 | 5.987342 | 5.805788 | 1.19E-07 | 4.50E-07 | 7.00647 |
| DTYMK | 0.269005 | 1.204977 | 6.542152 | 5.804897 | 1.20E-07 | 4.52E-07 | 7.002801 |
| CRYBB3 | -0.35322 | 0.782834 | 5.356582 | -5.80454 | 1.20E-07 | 4.52E-07 | 7.001336 |
| ZC3H7B | -0.20269 | 0.868926 | 6.647722 | -5.80345 | 1.21E-07 | 4.54E-07 | 6.996844 |
| ORMDL2 | 0.265942 | 1.202421 | 8.468481 | 5.801935 | 1.21E-07 | 4.57E-07 | 6.990597 |
| IFRD1 | -0.43588 | 0.739245 | 5.865316 | -5.80097 | 1.22E-07 | 4.59E-07 | 6.986613 |
| CD58 | 0.31448 | 1.243563 | 7.056582 | 5.800251 | 1.22E-07 | 4.60E-07 | 6.983661 |
| LEPR | 0.31473 | 1.243779 | 5.138861 | 5.799067 | 1.23E-07 | 4.62E-07 | 6.978784 |
| METRN | -0.36136 | 0.778432 | 6.432532 | -5.79821 | 1.23E-07 | 4.63E-07 | 6.975271 |
| NARS2 | -0.29337 | 0.815996 | 7.995063 | -5.79779 | 1.23E-07 | 4.64E-07 | 6.97353 |
| CPNE7 | -0.25115 | 0.840225 | 5.701392 | -5.79658 | 1.24E-07 | 4.66E-07 | 6.968526 |
| MSRB2 | -0.20336 | 0.868526 | 8.551139 | -5.79578 | 1.25E-07 | 4.67E-07 | 6.965237 |
| WT1 | -0.45787 | 0.728063 | 5.695696 | -5.79464 | 1.25E-07 | 4.69E-07 | 6.960539 |
| ZNF146 | 0.269183 | 1.205125 | 7.151139 | 5.794226 | 1.25E-07 | 4.70E-07 | 6.958851 |

| | | | | | | | |
|---|---|---|---|---|---|---|---|
| PLA2G7 | 0.414552 | 1.332885 | 3.711646 | 5.79337 | 1.26E-07 | 4.72E-07 | 6.955323 |
| CDKL1 | -0.30232 | 0.810948 | 5.699367 | -5.7921 | 1.26E-07 | 4.74E-07 | 6.950103 |
| AQP1 | -0.45176 | 0.731151 | 11.59759 | -5.78839 | 1.28E-07 | 4.81E-07 | 6.934823 |
| SERTAD3 | -0.31655 | 0.802985 | 6.751899 | -5.78643 | 1.30E-07 | 4.85E-07 | 6.926739 |
| PLA2G4A | 0.617701 | 1.534428 | 4.589114 | 5.786395 | 1.30E-07 | 4.85E-07 | 6.926614 |
| MRPS7 | -0.2476 | 0.842299 | 8.601139 | -5.78599 | 1.30E-07 | 4.85E-07 | 6.924948 |
| ZC3H4 | 0.325856 | 1.253408 | 8.503291 | 5.785563 | 1.30E-07 | 4.86E-07 | 6.92319 |
| FAM50B | -0.25987 | 0.83516 | 7.682532 | -5.78366 | 1.31E-07 | 4.90E-07 | 6.915342 |
| WBP1L | 0.31865 | 1.247163 | 8.465063 | 5.780512 | 1.33E-07 | 4.96E-07 | 6.902411 |
| MYO5C | 0.480455 | 1.395183 | 7.725696 | 5.775182 | 1.36E-07 | 5.07E-07 | 6.880492 |
| ATP6V1B1 | -0.44854 | 0.732782 | 7.136329 | -5.77481 | 1.36E-07 | 5.08E-07 | 6.878972 |
| GPR18 | 0.683808 | 1.606374 | 4.296076 | 5.774179 | 1.36E-07 | 5.09E-07 | 6.87637 |
| UBQLN2 | 0.431304 | 1.348452 | 7.198861 | 5.773863 | 1.37E-07 | 5.09E-07 | 6.875069 |
| FMO4 | -0.69292 | 0.618601 | 9.144684 | -5.77381 | 1.37E-07 | 5.09E-07 | 6.874837 |
| TM9SF4 | -0.2326 | 0.851098 | 7.253924 | -5.77042 | 1.39E-07 | 5.16E-07 | 6.860934 |
| SLC26A1 | -0.222 | 0.857378 | 4.568354 | -5.77033 | 1.39E-07 | 5.16E-07 | 6.860527 |
| DLAT | -0.34034 | 0.789857 | 7.145443 | -5.76999 | 1.39E-07 | 5.17E-07 | 6.859163 |
| PCGF2 | -0.21609 | 0.860897 | 6.985823 | -5.76895 | 1.39E-07 | 5.19E-07 | 6.854871 |
| NFIL3 | -0.61675 | 0.65214 | 8.050633 | -5.76853 | 1.40E-07 | 5.19E-07 | 6.853146 |
| C7 | 0.791094 | 1.730386 | 9.697848 | 5.768171 | 1.40E-07 | 5.20E-07 | 6.851675 |
| TTC13 | 0.207754 | 1.154889 | 5.465696 | 5.766254 | 1.41E-07 | 5.24E-07 | 6.843797 |
| BAG5 | 0.237879 | 1.179258 | 7.379367 | 5.76522 | 1.42E-07 | 5.26E-07 | 6.839546 |
| TRDC | 0.258788 | 1.196473 | 4.398101 | 5.764992 | 1.42E-07 | 5.26E-07 | 6.838609 |
| ANAPC15 | -0.20183 | 0.869446 | 7.708734 | -5.76305 | 1.43E-07 | 5.30E-07 | 6.830646 |
| PLEKHF2 | -0.35834 | 0.780062 | 7.986835 | -5.7619 | 1.44E-07 | 5.33E-07 | 6.825919 |
| GAS2 | -0.63684 | 0.643118 | 6.025063 | -5.76182 | 1.44E-07 | 5.33E-07 | 6.825579 |
| MAPK1IP1 | -0.19882 | 0.871262 | 8.554557 | -5.76026 | 1.45E-07 | 5.36E-07 | 6.819175 |
| PRR36 | -0.29379 | 0.815754 | 5.763797 | -5.75961 | 1.45E-07 | 5.37E-07 | 6.816497 |
| ALKBH4 | -0.2348 | 0.849801 | 5.451266 | -5.75822 | 1.46E-07 | 5.40E-07 | 6.810806 |
| SASH3 | 0.438801 | 1.355477 | 5.500253 | 5.755409 | 1.48E-07 | 5.46E-07 | 6.799252 |
| IGHV1-2 | 1.040679 | 2.057195 | 4.547975 | 5.754107 | 1.48E-07 | 5.49E-07 | 6.793905 |
| RAB40B | -0.37075 | 0.77338 | 8.288608 | -5.75039 | 1.51E-07 | 5.57E-07 | 6.778651 |
| GPATCH3 | -0.24271 | 0.845154 | 6.48557 | -5.74926 | 1.51E-07 | 5.60E-07 | 6.774011 |
| COX11 | -0.22158 | 0.857625 | 7.922658 | -5.74318 | 1.55E-07 | 5.74E-07 | 6.749055 |
| METTL9 | -0.20735 | 0.866126 | 10.22165 | -5.74244 | 1.56E-07 | 5.76E-07 | 6.746019 |
| WNT10B | -0.37497 | 0.771123 | 6.650759 | -5.74182 | 1.56E-07 | 5.77E-07 | 6.74349 |
| CTSF | -0.24059 | 0.846397 | 8.126456 | -5.74128 | 1.57E-07 | 5.78E-07 | 6.741251 |
| CLCN3 | 0.307273 | 1.237366 | 7.648354 | 5.740148 | 1.57E-07 | 5.80E-07 | 6.736629 |
| CCNB1 | 0.415375 | 1.333646 | 4.931772 | 5.739694 | 1.58E-07 | 5.81E-07 | 6.734765 |
| ITGA2 | 0.431838 | 1.348951 | 7.655823 | 5.737902 | 1.59E-07 | 5.85E-07 | 6.727418 |
| SLC2A9 | -0.6387 | 0.64229 | 8.753418 | -5.73651 | 1.60E-07 | 5.88E-07 | 6.721726 |
| EPN1 | -0.30111 | 0.811626 | 6.341392 | -5.73351 | 1.62E-07 | 5.95E-07 | 6.70943 |
| PPFIBP1 | 0.447661 | 1.363828 | 5.437215 | 5.733491 | 1.62E-07 | 5.95E-07 | 6.709331 |
| STAT3 | 0.301686 | 1.232584 | 8.816456 | 5.732453 | 1.63E-07 | 5.98E-07 | 6.705077 |
| KIR3DL2 | -0.26016 | 0.834993 | 4.999367 | -5.73182 | 1.63E-07 | 5.99E-07 | 6.702482 |
| PDIA5 | 0.388281 | 1.308833 | 7.219367 | 5.730402 | 1.64E-07 | 6.02E-07 | 6.696672 |
| CACNA1F | -0.24475 | 0.843962 | 5.734937 | -5.72981 | 1.64E-07 | 6.04E-07 | 6.69424 |

| | | | | | | | |
|---|---|---|---|---|---|---|---|
| ATP1A3 | -0.17358 | 0.886642 | 4.273797 | -5.72927 | 1.65E-07 | 6.05E-07 | 6.69203 |
| HOXB6 | 0.411838 | 1.33038 | 6.447468 | 5.727832 | 1.66E-07 | 6.08E-07 | 6.686137 |
| SOCS7 | -0.19484 | 0.873669 | 5.097089 | -5.72748 | 1.66E-07 | 6.09E-07 | 6.684679 |
| NR1H2 | -0.22316 | 0.85669 | 6.695696 | -5.72673 | 1.66E-07 | 6.11E-07 | 6.681604 |
| POR | -0.44301 | 0.735598 | 7.53038 | -5.72577 | 1.67E-07 | 6.13E-07 | 6.677686 |
| EVL | 0.357134 | 1.280879 | 6.241139 | 5.722354 | 1.70E-07 | 6.21E-07 | 6.663697 |
| SLC22A18/ | -0.44385 | 0.735168 | 7.79481 | -5.7202 | 1.71E-07 | 6.27E-07 | 6.654891 |
| PTPRM | -0.27182 | 0.828271 | 8.497975 | -5.71944 | 1.72E-07 | 6.28E-07 | 6.651768 |
| NABP2 | -0.19082 | 0.876106 | 5.390506 | -5.71736 | 1.73E-07 | 6.34E-07 | 6.643256 |
| BIRC2 | 0.232411 | 1.174797 | 9.330127 | 5.716666 | 1.74E-07 | 6.35E-07 | 6.640406 |
| NYX | -0.20929 | 0.864964 | 5.048228 | -5.71506 | 1.75E-07 | 6.39E-07 | 6.633845 |
| GTPBP4 | 0.199769 | 1.148515 | 6.606709 | 5.71495 | 1.75E-07 | 6.39E-07 | 6.633381 |
| SLCO4C1 | -0.44772 | 0.7332 | 6.986456 | -5.71431 | 1.75E-07 | 6.41E-07 | 6.630745 |
| NOS2 | -0.35556 | 0.781566 | 5.835823 | -5.71175 | 1.77E-07 | 6.47E-07 | 6.62027 |
| TUG1 | 0.268669 | 1.204696 | 8.03962 | 5.706345 | 1.81E-07 | 6.62E-07 | 6.598166 |
| ASPSCR1 | -0.40159 | 0.757025 | 5.638987 | -5.70577 | 1.82E-07 | 6.63E-07 | 6.595815 |
| VPS13A | -0.26521 | 0.832077 | 5.422911 | -5.70464 | 1.83E-07 | 6.66E-07 | 6.591202 |
| ARNT2 | 0.399671 | 1.319207 | 6.543038 | 5.701392 | 1.85E-07 | 6.75E-07 | 6.577903 |
| VAV1 | 0.294578 | 1.226526 | 4.535443 | 5.70124 | 1.85E-07 | 6.75E-07 | 6.577282 |
| BCAP29 | 0.226825 | 1.170256 | 5.778228 | 5.70024 | 1.86E-07 | 6.78E-07 | 6.573196 |
| ZNF574 | -0.22659 | 0.85465 | 6.728608 | -5.69744 | 1.88E-07 | 6.85E-07 | 6.561731 |
| THBS4 | -0.2952 | 0.814961 | 5.397342 | -5.69743 | 1.88E-07 | 6.85E-07 | 6.561686 |
| DRAM1 | 0.348966 | 1.273647 | 8.424684 | 5.697111 | 1.88E-07 | 6.85E-07 | 6.560402 |
| EHBP1 | -0.23344 | 0.850605 | 7.376835 | -5.6971 | 1.88E-07 | 6.85E-07 | 6.560365 |
| SOX13 | -0.23587 | 0.849173 | 6.397342 | -5.69625 | 1.89E-07 | 6.87E-07 | 6.556899 |
| MT1G | -0.6861 | 0.621532 | 13.48253 | -5.69615 | 1.89E-07 | 6.87E-07 | 6.556463 |
| UGCG | 0.677062 | 1.59888 | 7.951519 | 5.695228 | 1.90E-07 | 6.90E-07 | 6.5527 |
| COMMD3 | 0.320237 | 1.248536 | 8.988987 | 5.694684 | 1.90E-07 | 6.91E-07 | 6.550479 |
| PEX1 | -0.22895 | 0.853258 | 6.220886 | -5.69344 | 1.91E-07 | 6.94E-07 | 6.545404 |
| F2RL1 | 0.339513 | 1.265329 | 7.92443 | 5.691712 | 1.93E-07 | 6.99E-07 | 6.538334 |
| ITGB8 | 0.328603 | 1.255797 | 4.629873 | 5.68903 | 1.95E-07 | 7.07E-07 | 6.527373 |
| AOX1 | -0.71959 | 0.607269 | 9.415063 | -5.68871 | 1.95E-07 | 7.07E-07 | 6.52605 |
| ZBTB10 | -0.31433 | 0.804222 | 4.89 | -5.68829 | 1.96E-07 | 7.08E-07 | 6.524356 |
| ANKMY2 | 0.348755 | 1.273461 | 8.263291 | 5.684367 | 1.99E-07 | 7.20E-07 | 6.508322 |
| APOO | -0.29472 | 0.815229 | 9.677975 | -5.68346 | 2.00E-07 | 7.22E-07 | 6.504602 |
| THG1L | 0.258906 | 1.196571 | 6.200759 | 5.683447 | 2.00E-07 | 7.22E-07 | 6.504565 |
| KNG1 | -0.84228 | 0.557762 | 7.814684 | -5.68009 | 2.02E-07 | 7.32E-07 | 6.490859 |
| WIPF1 | 0.310995 | 1.240563 | 5.450127 | 5.677919 | 2.04E-07 | 7.38E-07 | 6.481994 |
| MTMR14 | -0.22531 | 0.855411 | 7.482405 | -5.67681 | 2.05E-07 | 7.41E-07 | 6.477452 |
| TOM1L2 | -0.21588 | 0.861023 | 6.587215 | -5.67644 | 2.05E-07 | 7.42E-07 | 6.475952 |
| INPP5J | -0.34123 | 0.789371 | 6.780506 | -5.67607 | 2.06E-07 | 7.43E-07 | 6.474444 |
| SPTLC2 | 0.285804 | 1.219089 | 6.648734 | 5.674879 | 2.07E-07 | 7.46E-07 | 6.469587 |
| GPM6B | 0.342134 | 1.267631 | 4.639873 | 5.674742 | 2.07E-07 | 7.46E-07 | 6.469027 |
| ATP1B2 | -0.29845 | 0.813128 | 5.876203 | -5.67164 | 2.10E-07 | 7.56E-07 | 6.456349 |
| ELMO1 | 0.284038 | 1.217598 | 4.945823 | 5.670978 | 2.10E-07 | 7.58E-07 | 6.453666 |
| RPP14 | -0.25179 | 0.839853 | 7.478734 | -5.67063 | 2.11E-07 | 7.58E-07 | 6.452239 |
| MICALL2 | 0.428129 | 1.345488 | 5.451013 | 5.669727 | 2.11E-07 | 7.61E-07 | 6.448562 |

| | | | | | | | |
|---|---|---|---|---|---|---|---|
| HS3ST1 | 0.621831 | 1.538827 | 5.036709 | 5.667884 | 2.13E-07 | 7.66E-07 | 6.441044 |
| PTGER3 | -0.43982 | 0.737229 | 6.943671 | -5.66779 | 2.13E-07 | 7.66E-07 | 6.440661 |
| TIMM23 | -0.22041 | 0.858322 | 7.792278 | -5.66667 | 2.14E-07 | 7.70E-07 | 6.436089 |
| PDSS2 | -0.34774 | 0.785814 | 8.092785 | -5.66561 | 2.15E-07 | 7.73E-07 | 6.431753 |
| IZUMO4 | -0.25891 | 0.835721 | 5.489241 | -5.66339 | 2.17E-07 | 7.80E-07 | 6.422733 |
| NDN | 0.404157 | 1.323315 | 7.498608 | 5.660494 | 2.20E-07 | 7.89E-07 | 6.410904 |
| MRPL48 | -0.25024 | 0.840754 | 8.551772 | -5.6595 | 2.21E-07 | 7.92E-07 | 6.406865 |
| MAP7D1 | 0.298241 | 1.229644 | 6.500886 | 5.65892 | 2.21E-07 | 7.93E-07 | 6.404488 |
| CORO1C | 0.320329 | 1.248616 | 8.807722 | 5.654739 | 2.25E-07 | 8.07E-07 | 6.387448 |
| ANK2 | -0.70735 | 0.612443 | 9.032785 | -5.65449 | 2.25E-07 | 8.08E-07 | 6.386432 |
| TRIM5 | 0.27247 | 1.207874 | 4.333165 | 5.654201 | 2.26E-07 | 8.08E-07 | 6.385256 |
| HSD3B2 | -0.25204 | 0.839711 | 4.924937 | -5.65364 | 2.26E-07 | 8.10E-07 | 6.382973 |
| ADAMTS1 | 1.104065 | 2.149595 | 6.942278 | 5.651365 | 2.28E-07 | 8.17E-07 | 6.373699 |
| TXNL4B | 0.252661 | 1.191403 | 6.630759 | 5.64869 | 2.31E-07 | 8.26E-07 | 6.362804 |
| YARS2 | 0.255758 | 1.193963 | 6.846835 | 5.646727 | 2.33E-07 | 8.32E-07 | 6.354808 |
| PXMP4 | -0.21449 | 0.861853 | 7.046709 | -5.64482 | 2.35E-07 | 8.38E-07 | 6.347054 |
| CALCRL | 0.289506 | 1.222222 | 6.005063 | 5.644806 | 2.35E-07 | 8.38E-07 | 6.346985 |
| RBM4B | 0.3 | 1.231144 | 6.895316 | 5.644047 | 2.35E-07 | 8.41E-07 | 6.343891 |
| CAPN3 | -1.03221 | 0.488959 | 8.950127 | -5.63965 | 2.40E-07 | 8.56E-07 | 6.326001 |
| ITM2A | 0.540402 | 1.454378 | 6.722911 | 5.638862 | 2.40E-07 | 8.58E-07 | 6.322783 |
| GRN | 0.296601 | 1.228247 | 8.899114 | 5.637711 | 2.42E-07 | 8.62E-07 | 6.3181 |
| ABHD8 | -0.20617 | 0.866838 | 5.118228 | -5.63743 | 2.42E-07 | 8.62E-07 | 6.316969 |
| LIMA1 | 0.282549 | 1.216342 | 8.399114 | 5.637425 | 2.42E-07 | 8.62E-07 | 6.316936 |
| JAM2 | 0.397141 | 1.316896 | 5.486329 | 5.635257 | 2.44E-07 | 8.70E-07 | 6.308115 |
| YOD1 | -0.26113 | 0.834436 | 4.271139 | -5.63394 | 2.45E-07 | 8.74E-07 | 6.302748 |
| ACOT8 | -0.28303 | 0.821863 | 7.381772 | -5.63334 | 2.46E-07 | 8.76E-07 | 6.300304 |
| ZCCHC10 | 0.285501 | 1.218833 | 5.698608 | 5.632873 | 2.46E-07 | 8.77E-07 | 6.298415 |
| ST8SIA4 | 0.310672 | 1.240285 | 3.974557 | 5.631492 | 2.48E-07 | 8.82E-07 | 6.292795 |
| WDR18 | -0.38976 | 0.763255 | 7.152405 | -5.62893 | 2.51E-07 | 8.91E-07 | 6.282373 |
| MT1HL1 | -0.30685 | 0.808404 | 7.65443 | -5.62775 | 2.52E-07 | 8.95E-07 | 6.277589 |
| MEIS2 | 0.67973 | 1.60184 | 6.306329 | 5.62733 | 2.52E-07 | 8.96E-07 | 6.275866 |
| GEMIN6 | -0.21553 | 0.861231 | 7.746709 | -5.62675 | 2.53E-07 | 8.98E-07 | 6.273515 |
| TUT1 | -0.25219 | 0.839623 | 5.789873 | -5.62627 | 2.53E-07 | 9.00E-07 | 6.27155 |
| WWOX | -0.22978 | 0.852767 | 7.202278 | -5.62567 | 2.54E-07 | 9.02E-07 | 6.269112 |
| CACNB2 | -0.25791 | 0.836301 | 4.978354 | -5.62543 | 2.54E-07 | 9.02E-07 | 6.26813 |
| EXOC1 | 0.31496 | 1.243978 | 8.035696 | 5.625371 | 2.54E-07 | 9.02E-07 | 6.2679 |
| SPAG9 | -0.23146 | 0.851775 | 7.18962 | -5.62358 | 2.56E-07 | 9.08E-07 | 6.260635 |
| IDH1 | -0.49589 | 0.709124 | 11.24329 | -5.6213 | 2.59E-07 | 9.17E-07 | 6.251338 |
| P4HB | 0.305949 | 1.236231 | 9.793671 | 5.619487 | 2.61E-07 | 9.23E-07 | 6.243981 |
| TMEM127 | 0.210534 | 1.157116 | 7.804684 | 5.614839 | 2.66E-07 | 9.41E-07 | 6.225097 |
| ERCC2 | -0.25967 | 0.835279 | 6.315443 | -5.61363 | 2.67E-07 | 9.45E-07 | 6.220203 |
| TAC1 | 1.140705 | 2.204887 | 4.573671 | 5.609535 | 2.72E-07 | 9.61E-07 | 6.203551 |
| NDUFA7 | -0.22299 | 0.856787 | 8.697722 | -5.60868 | 2.73E-07 | 9.64E-07 | 6.200078 |
| RECQL | 0.247905 | 1.187482 | 6.067468 | 5.607047 | 2.74E-07 | 9.70E-07 | 6.193448 |
| ZDHHC14 | 0.268906 | 1.204894 | 6.374937 | 5.604129 | 2.78E-07 | 9.82E-07 | 6.181602 |
| NPL | -0.65974 | 0.632991 | 8.845063 | -5.60291 | 2.79E-07 | 9.86E-07 | 6.176637 |
| NUDT13 | -0.35923 | 0.779581 | 5.031899 | -5.60236 | 2.80E-07 | 9.88E-07 | 6.174418 |

| | | | | | | | |
|---|---|---|---|---|---|---|---|
| IGSF3 | 0.283874 | 1.217459 | 7.153797 | 5.601666 | 2.81E-07 | 9.90E-07 | 6.171606 |
| FBXW4 | -0.24534 | 0.843615 | 7.721646 | -5.60134 | 2.81E-07 | 9.91E-07 | 6.170296 |
| VNN2 | 0.561765 | 1.476074 | 5.066835 | 5.599806 | 2.83E-07 | 9.97E-07 | 6.164056 |
| TACSTD2 | 0.600514 | 1.516257 | 9.752152 | 5.599597 | 2.83E-07 | 9.98E-07 | 6.163207 |
| NINL | -0.33264 | 0.794085 | 7.459747 | -5.59899 | 2.84E-07 | 1.00E-06 | 6.160728 |
| COTL1 | -0.6504 | 0.637103 | 9.381139 | -5.59782 | 2.85E-07 | 1.00E-06 | 6.155996 |
| UIMC1 | 0.25332 | 1.191947 | 6.478861 | 5.594719 | 2.89E-07 | 1.02E-06 | 6.143415 |
| BTNL3 | -0.25874 | 0.83582 | 6.469747 | -5.59429 | 2.89E-07 | 1.02E-06 | 6.141668 |
| AIFM1 | -0.36638 | 0.775728 | 9.78 | -5.59326 | 2.91E-07 | 1.02E-06 | 6.137495 |
| GRK5 | 0.233379 | 1.175585 | 5.376835 | 5.59284 | 2.91E-07 | 1.02E-06 | 6.135794 |
| TMEM115 | -0.25858 | 0.835912 | 6.653291 | -5.5911 | 2.93E-07 | 1.03E-06 | 6.128751 |
| CTSD | 0.378024 | 1.29956 | 8.44443 | 5.590452 | 2.94E-07 | 1.03E-06 | 6.12611 |
| TSTA3 | 0.287095 | 1.220181 | 7.391013 | 5.589872 | 2.95E-07 | 1.03E-06 | 6.123761 |
| PNMA1 | 0.270975 | 1.206623 | 8.437975 | 5.589849 | 2.95E-07 | 1.03E-06 | 6.123666 |
| FOXO1 | -0.27848 | 0.82446 | 7.412152 | -5.58963 | 2.95E-07 | 1.04E-06 | 6.122798 |
| BAX | 0.426285 | 1.343768 | 6.216329 | 5.589057 | 2.96E-07 | 1.04E-06 | 6.120453 |
| PPA2 | -0.20886 | 0.86522 | 10.06949 | -5.58793 | 2.97E-07 | 1.04E-06 | 6.115871 |
| CA4 | -0.48328 | 0.715349 | 7.525949 | -5.58222 | 3.04E-07 | 1.07E-06 | 6.092758 |
| BCL2A1 | 0.613729 | 1.530209 | 4.045063 | 5.5801 | 3.07E-07 | 1.08E-06 | 6.084151 |
| HBB | 1.936515 | 3.827799 | 8.793924 | 5.579947 | 3.07E-07 | 1.08E-06 | 6.083531 |
| SLC7A8 | -0.49939 | 0.707404 | 9.291392 | -5.57975 | 3.07E-07 | 1.08E-06 | 6.082737 |
| PPP1R10 | -0.30628 | 0.808725 | 7.092278 | -5.57954 | 3.08E-07 | 1.08E-06 | 6.081863 |
| ADAM9 | 0.465072 | 1.380387 | 8.622532 | 5.578888 | 3.09E-07 | 1.08E-06 | 6.079242 |
| CCNB2 | 0.432016 | 1.349117 | 4.81481 | 5.575223 | 3.13E-07 | 1.10E-06 | 6.064396 |
| PHLDA3 | 0.287253 | 1.220314 | 6.624557 | 5.57479 | 3.14E-07 | 1.10E-06 | 6.06264 |
| WDR3 | 0.244559 | 1.18473 | 6.979114 | 5.574775 | 3.14E-07 | 1.10E-06 | 6.062582 |
| DHRS9 | 0.231653 | 1.17418 | 5.37481 | 5.573147 | 3.16E-07 | 1.10E-06 | 6.055987 |
| KIAA0391 | -0.21445 | 0.861877 | 8.197595 | -5.57112 | 3.19E-07 | 1.11E-06 | 6.047784 |
| HLA-DQA1 | 1.869921 | 3.655126 | 6.609367 | 5.568173 | 3.23E-07 | 1.13E-06 | 6.035851 |
| MTG1 | -0.22702 | 0.8544 | 7.745823 | -5.56561 | 3.26E-07 | 1.14E-06 | 6.025487 |
| SH3BGRL | 0.269618 | 1.205489 | 9.120886 | 5.562265 | 3.31E-07 | 1.15E-06 | 6.011941 |
| IL18 | 0.439631 | 1.356257 | 5.158861 | 5.55876 | 3.35E-07 | 1.17E-06 | 5.997761 |
| STK39 | 0.387154 | 1.307811 | 5.469114 | 5.558502 | 3.36E-07 | 1.17E-06 | 5.996719 |
| CYP4F2 | -1.10434 | 0.465115 | 8.741519 | -5.55607 | 3.39E-07 | 1.18E-06 | 5.986897 |
| EIF5B | 0.191686 | 1.142098 | 7.010506 | 5.556019 | 3.39E-07 | 1.18E-06 | 5.986677 |
| PCOLCE | 0.387701 | 1.308307 | 5.803038 | 5.554532 | 3.41E-07 | 1.19E-06 | 5.980665 |
| TBC1D17 | -0.24481 | 0.843928 | 6.038608 | -5.5496 | 3.48E-07 | 1.21E-06 | 5.96073 |
| DUSP26 | -0.28643 | 0.819929 | 5.18557 | -5.54847 | 3.50E-07 | 1.22E-06 | 5.956175 |
| IGKV1D-8 | 0.573399 | 1.488025 | 4.344304 | 5.547847 | 3.51E-07 | 1.22E-06 | 5.953641 |
| TACR2 | -0.2543 | 0.838393 | 5.367468 | -5.54778 | 3.51E-07 | 1.22E-06 | 5.953375 |
| CCL8 | 0.758887 | 1.692184 | 5.184177 | 5.546338 | 3.53E-07 | 1.23E-06 | 5.947541 |
| CCDC25 | -0.22065 | 0.858177 | 7.648481 | -5.54519 | 3.55E-07 | 1.23E-06 | 5.942891 |
| KIAA0895 | -0.26311 | 0.83329 | 4.958354 | -5.54188 | 3.60E-07 | 1.25E-06 | 5.929526 |
| BAAT | -0.21455 | 0.861818 | 5.23038 | -5.53552 | 3.69E-07 | 1.28E-06 | 5.903861 |
| REXO2 | -0.26955 | 0.829577 | 10.29392 | -5.53285 | 3.74E-07 | 1.29E-06 | 5.893058 |
| MMP25 | -0.22457 | 0.855853 | 6.020759 | -5.53188 | 3.75E-07 | 1.30E-06 | 5.88916 |
| MTERF4 | -0.1896 | 0.876846 | 5.889494 | -5.53171 | 3.75E-07 | 1.30E-06 | 5.888485 |

| | | | | | | | |
|---|---|---|---|---|---|---|---|
| SCP2 | -0.2453 | 0.843639 | 9.892532 | -5.53042 | 3.77E-07 | 1.31E-06 | 5.883269 |
| ARAF | -0.18861 | 0.877451 | 7.050127 | -5.52995 | 3.78E-07 | 1.31E-06 | 5.881376 |
| RPL23 | 0.212885 | 1.159004 | 12.51241 | 5.529343 | 3.79E-07 | 1.31E-06 | 5.878916 |
| SNRNP70 | 0.298393 | 1.229773 | 7.555949 | 5.528898 | 3.80E-07 | 1.31E-06 | 5.877121 |
| LPO | -0.20928 | 0.864972 | 4.83519 | -5.52627 | 3.84E-07 | 1.33E-06 | 5.866501 |
| TSPAN8 | 1.01361 | 2.018957 | 8.039494 | 5.525955 | 3.84E-07 | 1.33E-06 | 5.865248 |
| CPPED1 | -0.20378 | 0.868272 | 5.738354 | -5.52572 | 3.85E-07 | 1.33E-06 | 5.864305 |
| RAB5C | -0.24807 | 0.842022 | 8.510506 | -5.52469 | 3.86E-07 | 1.33E-06 | 5.860145 |
| PPCDC | 0.25195 | 1.190816 | 5.61481 | 5.524179 | 3.87E-07 | 1.34E-06 | 5.858085 |
| RAB6B | 0.21448 | 1.160285 | 6.11481 | 5.521103 | 3.92E-07 | 1.35E-06 | 5.84568 |
| GOLIM4 | -0.21089 | 0.864004 | 5.877342 | -5.51991 | 3.94E-07 | 1.36E-06 | 5.840847 |
| CDA | -0.39242 | 0.761848 | 5.716076 | -5.51988 | 3.94E-07 | 1.36E-06 | 5.840755 |
| ID2 | -0.56026 | 0.678181 | 8.765316 | -5.51691 | 3.99E-07 | 1.37E-06 | 5.82877 |
| FAM134B | -0.45514 | 0.72944 | 9.426835 | -5.5166 | 4.00E-07 | 1.38E-06 | 5.827512 |
| TONSL | -0.25382 | 0.838672 | 5.086582 | -5.51268 | 4.06E-07 | 1.40E-06 | 5.811715 |
| TMEM59L | -0.29785 | 0.813466 | 6.007975 | -5.51028 | 4.10E-07 | 1.41E-06 | 5.802035 |
| POLM | -0.25729 | 0.83666 | 5.36557 | -5.50888 | 4.12E-07 | 1.42E-06 | 5.796426 |
| LILRB2 | 0.309091 | 1.238927 | 3.989114 | 5.507013 | 4.16E-07 | 1.43E-06 | 5.788893 |
| GABARAPL | -0.24011 | 0.84668 | 4.58557 | -5.50581 | 4.18E-07 | 1.44E-06 | 5.784036 |
| RAB21 | -0.27378 | 0.827149 | 8.969114 | -5.50534 | 4.19E-07 | 1.44E-06 | 5.78216 |
| CD200 | 0.226502 | 1.169995 | 5.042658 | 5.504005 | 4.21E-07 | 1.44E-06 | 5.77678 |
| ACTR2 | 0.358333 | 1.281944 | 8.634684 | 5.502165 | 4.24E-07 | 1.45E-06 | 5.769369 |
| CLUH | -0.24208 | 0.845524 | 9.129747 | -5.50194 | 4.24E-07 | 1.46E-06 | 5.768475 |
| RBM15 | 0.352134 | 1.276448 | 7.354051 | 5.496905 | 4.33E-07 | 1.49E-06 | 5.748194 |
| HABP4 | -0.20302 | 0.868728 | 6.893671 | -5.49424 | 4.38E-07 | 1.50E-06 | 5.737464 |
| SLC12A2 | -0.27547 | 0.826182 | 6.997975 | -5.49335 | 4.40E-07 | 1.51E-06 | 5.733903 |
| DKK3 | 0.658063 | 1.577963 | 4.932278 | 5.493093 | 4.40E-07 | 1.51E-06 | 5.732856 |
| SLC47A1 | -0.54739 | 0.684256 | 10.69873 | -5.49295 | 4.41E-07 | 1.51E-06 | 5.732295 |
| HAUS4 | -0.39436 | 0.760826 | 7.822658 | -5.49157 | 4.43E-07 | 1.52E-06 | 5.72673 |
| BORA | 0.246509 | 1.186333 | 4.739494 | 5.491282 | 4.44E-07 | 1.52E-06 | 5.72557 |
| CTPS1 | -0.5431 | 0.686296 | 6.498354 | -5.49034 | 4.45E-07 | 1.52E-06 | 5.721781 |
| WDR41 | 0.370744 | 1.29302 | 6.552911 | 5.490332 | 4.45E-07 | 1.52E-06 | 5.721751 |
| EIF4G2 | 0.203155 | 1.151214 | 10.26595 | 5.489781 | 4.46E-07 | 1.52E-06 | 5.719533 |
| ACTB | 0.410929 | 1.329542 | 11.45709 | 5.489591 | 4.47E-07 | 1.53E-06 | 5.718768 |
| HNF1B | 0.329137 | 1.256262 | 9.486835 | 5.489279 | 4.47E-07 | 1.53E-06 | 5.717513 |
| URGCP | -0.21086 | 0.864024 | 7.809747 | -5.48818 | 4.49E-07 | 1.53E-06 | 5.7131 |
| ALG5 | 0.215942 | 1.161462 | 9.507595 | 5.487783 | 4.50E-07 | 1.53E-06 | 5.711496 |
| SQRDL | 0.298801 | 1.230122 | 9.561772 | 5.487154 | 4.51E-07 | 1.54E-06 | 5.708968 |
| ABCG2 | -0.52861 | 0.693222 | 5.978101 | -5.48411 | 4.57E-07 | 1.56E-06 | 5.696739 |
| MPDU1 | -0.24929 | 0.841311 | 8.861519 | -5.48367 | 4.58E-07 | 1.56E-06 | 5.694979 |
| DCLK1 | 0.223623 | 1.167662 | 3.966456 | 5.483004 | 4.59E-07 | 1.56E-06 | 5.692283 |
| MAP3K10 | -0.21663 | 0.860571 | 6.821899 | -5.48294 | 4.59E-07 | 1.56E-06 | 5.692038 |
| ETV2 | -0.20729 | 0.866165 | 4.346456 | -5.48088 | 4.63E-07 | 1.58E-06 | 5.683741 |
| OXR1 | 0.338524 | 1.264463 | 5.852278 | 5.480124 | 4.64E-07 | 1.58E-06 | 5.680706 |
| PTDSS1 | -0.24952 | 0.841177 | 9.474684 | -5.47677 | 4.71E-07 | 1.60E-06 | 5.667221 |
| GBAS | 0.330949 | 1.25784 | 8.569114 | 5.475437 | 4.74E-07 | 1.61E-06 | 5.661877 |
| KRAS | 0.28886 | 1.221675 | 7.603924 | 5.474954 | 4.74E-07 | 1.61E-06 | 5.659935 |

| | | | | | | | |
|---|---|---|---|---|---|---|---|
| SOAT1 | 0.259875 | 1.197375 | 5.267468 | 5.474596 | 4.75E-07 | 1.61E-06 | 5.658494 |
| PPP1R3C | 0.540323 | 1.454298 | 7.131139 | 5.473979 | 4.76E-07 | 1.62E-06 | 5.65602 |
| SSRP1 | 0.185593 | 1.137284 | 7.56557 | 5.473287 | 4.78E-07 | 1.62E-06 | 5.653239 |
| PLAG1 | 0.241476 | 1.182201 | 4.23 | 5.472598 | 4.79E-07 | 1.62E-06 | 5.650469 |
| MAD2L1 | 0.286337 | 1.21954 | 4.205696 | 5.472533 | 4.79E-07 | 1.62E-06 | 5.650209 |
| ZEB2 | 0.287167 | 1.220242 | 4.124304 | 5.471376 | 4.82E-07 | 1.63E-06 | 5.645564 |
| NAT8B | -0.80256 | 0.573333 | 8.18519 | -5.47107 | 4.82E-07 | 1.63E-06 | 5.64435 |
| ALG6 | 0.252734 | 1.191463 | 7.004051 | 5.469532 | 4.85E-07 | 1.64E-06 | 5.638157 |
| PCNA | 0.295072 | 1.226947 | 8.701519 | 5.468355 | 4.88E-07 | 1.65E-06 | 5.633433 |
| NT5DC2 | 0.31612 | 1.244978 | 5.108354 | 5.46691 | 4.90E-07 | 1.66E-06 | 5.627633 |
| ARID4B | 0.267747 | 1.203926 | 7.062278 | 5.464396 | 4.96E-07 | 1.68E-06 | 5.61754 |
| IL13 | -0.26395 | 0.832803 | 5.632785 | -5.46071 | 5.03E-07 | 1.70E-06 | 5.60273 |
| EXOC2 | 0.265659 | 1.202185 | 5.583797 | 5.459993 | 5.05E-07 | 1.71E-06 | 5.599871 |
| KLF15 | -0.31282 | 0.805067 | 6.860633 | -5.45871 | 5.07E-07 | 1.71E-06 | 5.594726 |
| CIRBP | -0.27117 | 0.828646 | 9.517595 | -5.45435 | 5.17E-07 | 1.74E-06 | 5.577241 |
| C1orf54 | 0.589295 | 1.504511 | 8.008987 | 5.453882 | 5.18E-07 | 1.75E-06 | 5.575357 |
| SHPK | -0.22155 | 0.857645 | 5.715063 | -5.45345 | 5.18E-07 | 1.75E-06 | 5.573613 |
| CDKN1C | -0.4541 | 0.729967 | 8.366835 | -5.45109 | 5.23E-07 | 1.77E-06 | 5.564159 |
| RNF126 | -0.21914 | 0.859075 | 5.940633 | -5.4419 | 5.44E-07 | 1.83E-06 | 5.527316 |
| FKBP6 | -0.20462 | 0.867768 | 5.281266 | -5.44038 | 5.47E-07 | 1.84E-06 | 5.521256 |
| ZNF839 | -0.18709 | 0.878373 | 5.650759 | -5.43905 | 5.50E-07 | 1.85E-06 | 5.515926 |
| CHUK | -0.29157 | 0.817014 | 5.61038 | -5.43651 | 5.56E-07 | 1.87E-06 | 5.505728 |
| TNFAIP6 | 0.379223 | 1.300641 | 4.197975 | 5.431872 | 5.67E-07 | 1.91E-06 | 5.487179 |
| BMP8B | -0.29562 | 0.814723 | 4.914557 | -5.43133 | 5.68E-07 | 1.91E-06 | 5.485019 |
| SMCO4 | 0.364295 | 1.287253 | 7.9 | 5.429977 | 5.71E-07 | 1.92E-06 | 5.479596 |
| SPATA7 | -0.25899 | 0.835675 | 6.037468 | -5.42706 | 5.78E-07 | 1.94E-06 | 5.467919 |
| MT4 | -0.54405 | 0.685842 | 9.608608 | -5.42557 | 5.81E-07 | 1.95E-06 | 5.461956 |
| FBXL15 | -0.23584 | 0.849189 | 6.641266 | -5.42478 | 5.83E-07 | 1.96E-06 | 5.458792 |
| CXCL11 | 0.587727 | 1.502877 | 3.790506 | 5.422553 | 5.89E-07 | 1.98E-06 | 5.449896 |
| DPF1 | -0.24206 | 0.84554 | 6.023671 | -5.42103 | 5.92E-07 | 1.99E-06 | 5.443806 |
| HHAT | 0.34722 | 1.272107 | 5.617215 | 5.420307 | 5.94E-07 | 1.99E-06 | 5.440912 |
| TOB1 | -0.42432 | 0.745189 | 9.891013 | -5.41481 | 6.08E-07 | 2.04E-06 | 5.41895 |
| ARSB | -0.36254 | 0.777792 | 5.895949 | -5.41334 | 6.11E-07 | 2.05E-06 | 5.413077 |
| ST14 | 0.321621 | 1.249734 | 6.52 | 5.411785 | 6.15E-07 | 2.06E-06 | 5.406846 |
| GPT | -0.40018 | 0.757765 | 6.843924 | -5.4112 | 6.17E-07 | 2.07E-06 | 5.404492 |
| APRT | -0.23752 | 0.848204 | 8.097089 | -5.40775 | 6.26E-07 | 2.09E-06 | 5.390745 |
| LARP4 | -0.34554 | 0.787013 | 5.615443 | -5.40559 | 6.31E-07 | 2.11E-06 | 5.382103 |
| TMCO3 | 0.348347 | 1.273101 | 6.467468 | 5.405258 | 6.32E-07 | 2.11E-06 | 5.380774 |
| ADAMTSL3 | 0.353169 | 1.277363 | 5.07557 | 5.405112 | 6.32E-07 | 2.11E-06 | 5.380194 |
| ZMYND8 | 0.316173 | 1.245023 | 6.336203 | 5.4046 | 6.34E-07 | 2.12E-06 | 5.378147 |
| PRPF19 | 0.21558 | 1.16117 | 7.751139 | 5.404077 | 6.35E-07 | 2.12E-06 | 5.376059 |
| ASF1A | 0.304631 | 1.235103 | 6.877468 | 5.401905 | 6.41E-07 | 2.14E-06 | 5.367387 |
| RAB38 | 0.323689 | 1.251527 | 6.503038 | 5.401833 | 6.41E-07 | 2.14E-06 | 5.367099 |
| ORAI3 | -0.23752 | 0.848204 | 8.187089 | -5.39988 | 6.46E-07 | 2.16E-06 | 5.359291 |
| PYGL | 0.331074 | 1.257949 | 5.190253 | 5.396172 | 6.56E-07 | 2.19E-06 | 5.344503 |
| ATP8B2 | 0.269974 | 1.205786 | 5.908861 | 5.39513 | 6.59E-07 | 2.20E-06 | 5.340345 |
| RSAD2 | 0.733162 | 1.662278 | 4.685823 | 5.394643 | 6.60E-07 | 2.20E-06 | 5.338405 |

| | | | | | | | |
|---|---|---|---|---|---|---|---|
| DBR1 | 0.268702 | 1.204724 | 5.032025 | 5.394433 | 6.61E-07 | 2.20E-06 | 5.337568 |
| NUP133 | 0.229196 | 1.172182 | 7.231392 | 5.393943 | 6.62E-07 | 2.21E-06 | 5.33561 |
| TSPYL2 | -0.33082 | 0.795086 | 6.900506 | -5.39343 | 6.63E-07 | 2.21E-06 | 5.333555 |
| FZD4 | -0.25855 | 0.835927 | 8.007215 | -5.39268 | 6.65E-07 | 2.21E-06 | 5.330584 |
| NR4A3 | -0.48567 | 0.714168 | 4.605823 | -5.39251 | 6.66E-07 | 2.22E-06 | 5.329885 |
| CYP2C8 | -0.60586 | 0.657081 | 4.179747 | -5.39162 | 6.68E-07 | 2.22E-06 | 5.326331 |
| SPTBN4 | -0.19493 | 0.873617 | 5.586835 | -5.39114 | 6.70E-07 | 2.23E-06 | 5.324421 |
| ST3GAL4 | -0.35165 | 0.783685 | 5.825063 | -5.38849 | 6.77E-07 | 2.25E-06 | 5.313874 |
| CFHR2 | 0.347273 | 1.272153 | 4.355063 | 5.387254 | 6.80E-07 | 2.26E-06 | 5.308932 |
| UCKL1 | -0.19379 | 0.874303 | 6.39557 | -5.38636 | 6.83E-07 | 2.27E-06 | 5.305382 |
| FARP1 | 0.268399 | 1.204471 | 8.681899 | 5.38622 | 6.83E-07 | 2.27E-06 | 5.30481 |
| MELK | 0.32753 | 1.254863 | 4.857468 | 5.385985 | 6.84E-07 | 2.27E-06 | 5.303875 |
| MOK | -0.26123 | 0.834379 | 6.373924 | -5.38542 | 6.86E-07 | 2.27E-06 | 5.301617 |
| CALB1 | -0.80086 | 0.574006 | 10.93481 | -5.3825 | 6.94E-07 | 2.30E-06 | 5.28999 |
| PHIP | 0.278155 | 1.212643 | 6.872278 | 5.38006 | 7.01E-07 | 2.32E-06 | 5.280259 |
| UBE2D1 | 0.223129 | 1.167263 | 4.99038 | 5.374632 | 7.16E-07 | 2.37E-06 | 5.258636 |
| FAM117A | -0.29284 | 0.816294 | 7.463544 | -5.37371 | 7.19E-07 | 2.38E-06 | 5.254966 |
| FGFR3 | -0.27171 | 0.828339 | 7.540633 | -5.37316 | 7.21E-07 | 2.39E-06 | 5.25279 |
| HECTD3 | -0.29211 | 0.816708 | 6.954937 | -5.37294 | 7.21E-07 | 2.39E-06 | 5.251889 |
| PIKFYVE | -0.26772 | 0.830631 | 5.961646 | -5.36883 | 7.34E-07 | 2.43E-06 | 5.235515 |
| LTBP3 | 0.343913 | 1.269194 | 5.659747 | 5.368547 | 7.35E-07 | 2.43E-06 | 5.234404 |
| ATP13A1 | 0.245264 | 1.185309 | 6.836582 | 5.366286 | 7.41E-07 | 2.45E-06 | 5.225406 |
| PRPF4 | -0.27905 | 0.824133 | 7.614304 | -5.36235 | 7.53E-07 | 2.49E-06 | 5.209747 |
| FRZB | 0.62809 | 1.545517 | 5.603671 | 5.361737 | 7.55E-07 | 2.49E-06 | 5.207303 |
| KIF22 | -0.20402 | 0.868129 | 5.563038 | -5.35664 | 7.71E-07 | 2.55E-06 | 5.187032 |
| OXT | -0.30616 | 0.808792 | 5.994937 | -5.35402 | 7.79E-07 | 2.57E-06 | 5.176633 |
| MPP5 | -0.3676 | 0.77507 | 6.992532 | -5.35376 | 7.80E-07 | 2.57E-06 | 5.175577 |
| TOP2A | 0.393202 | 1.313305 | 3.814684 | 5.352486 | 7.84E-07 | 2.59E-06 | 5.170517 |
| CHST12 | 0.218874 | 1.163824 | 5.941646 | 5.351881 | 7.86E-07 | 2.59E-06 | 5.16811 |
| RCAN1 | -0.48856 | 0.712734 | 7.794177 | -5.35152 | 7.87E-07 | 2.59E-06 | 5.166664 |
| PPP6R1 | -0.18596 | 0.879063 | 7.069494 | -5.34977 | 7.93E-07 | 2.61E-06 | 5.159705 |
| SMARCA1 | 0.258057 | 1.195867 | 6.726709 | 5.348632 | 7.97E-07 | 2.62E-06 | 5.155199 |
| CPE | 0.774354 | 1.710424 | 6.467595 | 5.346374 | 8.04E-07 | 2.65E-06 | 5.146227 |
| CUL9 | -0.15781 | 0.896387 | 5.887342 | -5.34559 | 8.07E-07 | 2.65E-06 | 5.14313 |
| ABCC8 | -0.26116 | 0.834417 | 5.058734 | -5.34534 | 8.08E-07 | 2.66E-06 | 5.142116 |
| AGO4 | 0.202642 | 1.150804 | 4.82443 | 5.342652 | 8.16E-07 | 2.68E-06 | 5.131443 |
| EXOSC8 | 0.274776 | 1.209806 | 6.271519 | 5.342312 | 8.18E-07 | 2.69E-06 | 5.130094 |
| TCF20 | 0.212101 | 1.158374 | 7.183165 | 5.340462 | 8.24E-07 | 2.71E-06 | 5.122744 |
| LSM7 | 0.324374 | 1.252121 | 8.015063 | 5.3389 | 8.29E-07 | 2.72E-06 | 5.116542 |
| ELMO3 | -0.31024 | 0.806505 | 6.016709 | -5.33717 | 8.35E-07 | 2.74E-06 | 5.109695 |
| TMEM160 | -0.32437 | 0.798648 | 6.913418 | -5.33709 | 8.35E-07 | 2.74E-06 | 5.109345 |
| ANKH | -0.1679 | 0.890138 | 4.62443 | -5.33466 | 8.44E-07 | 2.77E-06 | 5.099732 |
| PLA2G16 | -0.37564 | 0.770764 | 9.805696 | -5.33456 | 8.44E-07 | 2.77E-06 | 5.099316 |
| TNNI1 | -0.28336 | 0.821675 | 4.527722 | -5.33392 | 8.46E-07 | 2.77E-06 | 5.096757 |
| NCOA6 | 0.233366 | 1.175575 | 7.309873 | 5.333457 | 8.48E-07 | 2.78E-06 | 5.094938 |
| SUPT16H | 0.255619 | 1.193848 | 8.208734 | 5.327648 | 8.68E-07 | 2.84E-06 | 5.071893 |
| CTF1 | -0.30245 | 0.810874 | 6.209747 | -5.32705 | 8.70E-07 | 2.85E-06 | 5.069522 |

| | | | | | | | |
|---|---|---|---|---|---|---|---|
| MLLT4-AS1 | -0.29739 | 0.813722 | 5.103165 | -5.32688 | 8.71E-07 | 2.85E-06 | 5.068843 |
| EVPL | -0.213 | 0.862743 | 6.063418 | -5.32539 | 8.76E-07 | 2.87E-06 | 5.06294 |
| STT3A | 0.351634 | 1.276005 | 8.229494 | 5.325229 | 8.77E-07 | 2.87E-06 | 5.062301 |
| NAPA | -0.23465 | 0.849891 | 7.756329 | -5.32382 | 8.82E-07 | 2.88E-06 | 5.056695 |
| PARP6 | -0.18755 | 0.878096 | 7.37557 | -5.32321 | 8.84E-07 | 2.89E-06 | 5.05431 |
| ME1 | 0.420949 | 1.338808 | 4.926709 | 5.323003 | 8.85E-07 | 2.89E-06 | 5.053474 |
| DKFZP586I | 0.361324 | 1.284604 | 7.185063 | 5.322196 | 8.88E-07 | 2.90E-06 | 5.050274 |
| ESPL1 | -0.55943 | 0.678572 | 6.443924 | -5.31701 | 9.07E-07 | 2.96E-06 | 5.029714 |
| SAMD4B | -0.22336 | 0.856568 | 5.952785 | -5.31629 | 9.09E-07 | 2.97E-06 | 5.026856 |
| C16orf95 | -0.20963 | 0.864758 | 5.457215 | -5.3156 | 9.12E-07 | 2.97E-06 | 5.024141 |
| LITAF | 0.381443 | 1.302644 | 9.366076 | 5.315468 | 9.12E-07 | 2.97E-06 | 5.023611 |
| SLC43A3 | 0.259842 | 1.197347 | 6.075063 | 5.31375 | 9.19E-07 | 2.99E-06 | 5.016805 |
| CCDC28A | -0.22773 | 0.853979 | 9.339873 | -5.31005 | 9.33E-07 | 3.04E-06 | 5.002132 |
| RHOB | -0.6113 | 0.654605 | 9.495949 | -5.30709 | 9.44E-07 | 3.07E-06 | 4.990417 |
| CCL18 | 0.568623 | 1.483108 | 4.13557 | 5.307007 | 9.44E-07 | 3.07E-06 | 4.990101 |
| CAV2 | 0.60504 | 1.52102 | 6.268608 | 5.306803 | 9.45E-07 | 3.07E-06 | 4.989294 |
| CNOT6 | 0.264499 | 1.201219 | 7.051139 | 5.304871 | 9.52E-07 | 3.10E-06 | 4.981645 |
| CIB2 | 0.232484 | 1.174856 | 6.483418 | 5.304264 | 9.55E-07 | 3.10E-06 | 4.979242 |
| CGRRF1 | -0.2069 | 0.866399 | 7.106835 | -5.304 | 9.56E-07 | 3.11E-06 | 4.978192 |
| TBRG4 | -0.21304 | 0.862715 | 6.604051 | -5.30205 | 9.63E-07 | 3.13E-06 | 4.970468 |
| IL15RA | 0.276311 | 1.211094 | 5.967595 | 5.301957 | 9.64E-07 | 3.13E-06 | 4.970112 |
| NAT1 | -0.3092 | 0.807088 | 7.566709 | -5.30173 | 9.65E-07 | 3.13E-06 | 4.969212 |
| PANK2 | 0.240889 | 1.181721 | 7.80519 | 5.301653 | 9.65E-07 | 3.13E-06 | 4.968911 |
| FABP2 | -0.20449 | 0.867844 | 4.672405 | -5.30026 | 9.70E-07 | 3.15E-06 | 4.9634 |
| BZRAP1 | -0.22214 | 0.857292 | 5.481772 | -5.29709 | 9.83E-07 | 3.19E-06 | 4.950849 |
| SNX10 | 0.506166 | 1.420271 | 8.037089 | 5.296952 | 9.84E-07 | 3.19E-06 | 4.950311 |
| HNRNPH3 | 0.211495 | 1.157888 | 7.702911 | 5.296848 | 9.84E-07 | 3.19E-06 | 4.949901 |
| MZB1 | 0.800975 | 1.742278 | 6.279367 | 5.29522 | 9.91E-07 | 3.21E-06 | 4.943464 |
| CTNS | -0.22067 | 0.858166 | 7.273038 | -5.29373 | 9.97E-07 | 3.23E-06 | 4.937573 |
| STK25 | -0.25975 | 0.835233 | 6.773671 | -5.28815 | 1.02E-06 | 3.30E-06 | 4.915514 |
| LPAR1 | 0.30751 | 1.23757 | 5.413671 | 5.287669 | 1.02E-06 | 3.30E-06 | 4.913613 |
| AVP | -0.316 | 0.803293 | 5.514304 | -5.28364 | 1.04E-06 | 3.36E-06 | 4.897701 |
| PSMG2 | 0.183228 | 1.135421 | 9.700886 | 5.283092 | 1.04E-06 | 3.36E-06 | 4.895527 |
| CYP17A1 | -1.31575 | 0.401716 | 8.358861 | -5.28289 | 1.04E-06 | 3.36E-06 | 4.894725 |
| NSF | 0.23722 | 1.178719 | 8.301266 | 5.28233 | 1.04E-06 | 3.37E-06 | 4.892518 |
| CDH5 | 0.306765 | 1.236931 | 5.245316 | 5.280509 | 1.05E-06 | 3.39E-06 | 4.885326 |
| MSRB1 | -0.48438 | 0.714804 | 10.34962 | -5.27593 | 1.07E-06 | 3.46E-06 | 4.86723 |
| SPG21 | -0.21321 | 0.862613 | 9.803544 | -5.27163 | 1.09E-06 | 3.52E-06 | 4.850261 |
| ATP6V0D1 | -0.23125 | 0.851899 | 10.05101 | -5.27024 | 1.10E-06 | 3.54E-06 | 4.844772 |
| ENPEP | -0.64572 | 0.639175 | 9.921139 | -5.26954 | 1.10E-06 | 3.54E-06 | 4.842008 |
| OSBPL8 | -0.33036 | 0.79534 | 8.884177 | -5.26932 | 1.10E-06 | 3.55E-06 | 4.841148 |
| DUSP3 | -0.21688 | 0.860426 | 7.708101 | -5.26747 | 1.11E-06 | 3.57E-06 | 4.833869 |
| PROCR | 0.341482 | 1.267058 | 5.020253 | 5.264903 | 1.12E-06 | 3.61E-06 | 4.823737 |
| GLB1L | -0.27173 | 0.828328 | 7.96519 | -5.25953 | 1.15E-06 | 3.68E-06 | 4.802565 |
| HMG20B | -0.21802 | 0.859742 | 7.712405 | -5.25905 | 1.15E-06 | 3.69E-06 | 4.800666 |
| AZIN1 | 0.259018 | 1.196664 | 8.124937 | 5.258616 | 1.15E-06 | 3.70E-06 | 4.798952 |
| MFSD5 | 0.23247 | 1.174845 | 7.126456 | 5.25538 | 1.16E-06 | 3.74E-06 | 4.7862 |

| | | | | | | | |
|---|---|---|---|---|---|---|---|
| RND3 | 0.555329 | 1.469504 | 7.990886 | 5.255216 | 1.17E-06 | 3.74E-06 | 4.785551 |
| PRODH | -0.46481 | 0.724567 | 5.746709 | -5.25371 | 1.17E-06 | 3.77E-06 | 4.779637 |
| LRP10 | 0.22502 | 1.168793 | 8.53443 | 5.252395 | 1.18E-06 | 3.78E-06 | 4.774438 |
| POMT1 | -0.23315 | 0.850776 | 6.41 | -5.24988 | 1.19E-06 | 3.82E-06 | 4.764523 |
| MGST2 | -0.23885 | 0.847418 | 9.465443 | -5.24974 | 1.19E-06 | 3.82E-06 | 4.763964 |
| DCN | 0.741265 | 1.671641 | 9.597468 | 5.246499 | 1.21E-06 | 3.87E-06 | 4.751222 |
| GSTZ1 | -0.27287 | 0.827674 | 7.377975 | -5.24637 | 1.21E-06 | 3.87E-06 | 4.750723 |
| PNPLA2 | -0.19929 | 0.87098 | 7.352405 | -5.24563 | 1.21E-06 | 3.88E-06 | 4.747804 |
| CD36 | 0.438775 | 1.355453 | 4.416329 | 5.244414 | 1.22E-06 | 3.90E-06 | 4.743011 |
| DLGAP2 | -0.22106 | 0.857934 | 4.962658 | -5.24279 | 1.23E-06 | 3.92E-06 | 4.736625 |
| NUDT1 | 0.270665 | 1.206364 | 5.259367 | 5.241424 | 1.23E-06 | 3.94E-06 | 4.731246 |
| GABRR2 | -0.25358 | 0.838814 | 6.34038 | -5.24141 | 1.23E-06 | 3.94E-06 | 4.731172 |
| SLC1A5 | -0.29379 | 0.815757 | 6.682278 | -5.24063 | 1.24E-06 | 3.95E-06 | 4.728132 |
| SPATS2 | 0.237793 | 1.179188 | 5.159114 | 5.240384 | 1.24E-06 | 3.96E-06 | 4.727152 |
| TSPAN1 | 0.426884 | 1.344327 | 11.5781 | 5.238476 | 1.25E-06 | 3.99E-06 | 4.719645 |
| IGBP1 | -0.20924 | 0.864991 | 8.857595 | -5.2343 | 1.27E-06 | 4.05E-06 | 4.703239 |
| ITGAL | 0.255791 | 1.19399 | 4.509241 | 5.233197 | 1.27E-06 | 4.07E-06 | 4.698884 |
| SSR1 | 0.233215 | 1.175451 | 7.92481 | 5.232224 | 1.28E-06 | 4.08E-06 | 4.69506 |
| NAT10 | 0.184361 | 1.136314 | 6.91962 | 5.23201 | 1.28E-06 | 4.09E-06 | 4.694217 |
| HSD17B11 | -0.37852 | 0.769224 | 9.591013 | -5.23166 | 1.28E-06 | 4.09E-06 | 4.692825 |
| ARHGEF17 | -0.24596 | 0.843253 | 7.10443 | -5.23161 | 1.28E-06 | 4.09E-06 | 4.692626 |
| FOXRED2 | -0.23049 | 0.852343 | 5.537848 | -5.23015 | 1.29E-06 | 4.11E-06 | 4.686908 |
| FKBP4 | -0.20881 | 0.865252 | 6.747342 | -5.22616 | 1.31E-06 | 4.18E-06 | 4.671221 |
| EIF4A3 | 0.199203 | 1.148064 | 9.087342 | 5.226077 | 1.31E-06 | 4.18E-06 | 4.670899 |
| SMARCA4 | 0.193597 | 1.143611 | 6.85 | 5.225906 | 1.31E-06 | 4.18E-06 | 4.670228 |
| TRMT1 | 0.196937 | 1.146262 | 4.679873 | 5.223535 | 1.32E-06 | 4.22E-06 | 4.660911 |
| C19orf57 | -0.2107 | 0.864115 | 6.50481 | -5.22225 | 1.33E-06 | 4.24E-06 | 4.655845 |
| XPC | -0.24841 | 0.841826 | 8.407975 | -5.22055 | 1.34E-06 | 4.26E-06 | 4.649173 |
| ATP6V1D | -0.23827 | 0.847763 | 10.18025 | -5.21768 | 1.36E-06 | 4.31E-06 | 4.63791 |
| PPP1R26 | -0.22669 | 0.854595 | 6.459873 | -5.21609 | 1.37E-06 | 4.34E-06 | 4.63169 |
| CD55 | 0.340626 | 1.266306 | 6.037722 | 5.215709 | 1.37E-06 | 4.34E-06 | 4.630181 |
| PHGDH | -0.57571 | 0.670955 | 7.948861 | -5.21386 | 1.38E-06 | 4.37E-06 | 4.622923 |
| RFC2 | 0.220962 | 1.16551 | 5.890127 | 5.213307 | 1.38E-06 | 4.38E-06 | 4.620753 |
| GAK | -0.18659 | 0.878678 | 8.125316 | -5.21279 | 1.38E-06 | 4.39E-06 | 4.618737 |
| LSM8 | 0.211884 | 1.1582 | 6.613291 | 5.211871 | 1.39E-06 | 4.41E-06 | 4.615115 |
| STAT4 | 0.416133 | 1.334346 | 5.447089 | 5.210441 | 1.40E-06 | 4.43E-06 | 4.609504 |
| USP33 | 0.203814 | 1.151739 | 8.154051 | 5.209073 | 1.40E-06 | 4.45E-06 | 4.604138 |
| TFEB | -0.19108 | 0.87595 | 6.589747 | -5.20752 | 1.41E-06 | 4.48E-06 | 4.598048 |
| GPER1 | -0.32061 | 0.80073 | 5.017595 | -5.20726 | 1.41E-06 | 4.48E-06 | 4.597032 |
| PPP1R12A | 0.244796 | 1.184925 | 6.30443 | 5.206958 | 1.42E-06 | 4.48E-06 | 4.595843 |
| SFRP1 | 0.575283 | 1.48997 | 7.729873 | 5.206134 | 1.42E-06 | 4.50E-06 | 4.592611 |
| MAN2A2 | -0.18911 | 0.877146 | 6.30557 | -5.20564 | 1.42E-06 | 4.51E-06 | 4.590675 |
| HPGDS | 0.397194 | 1.316944 | 4.834177 | 5.204151 | 1.43E-06 | 4.53E-06 | 4.584835 |
| SGSH | 0.204881 | 1.152592 | 5.867975 | 5.20361 | 1.44E-06 | 4.54E-06 | 4.582711 |
| TRIM13 | -0.31402 | 0.804398 | 7.437089 | -5.20098 | 1.45E-06 | 4.59E-06 | 4.572405 |
| CHMP6 | -0.19929 | 0.87098 | 6.412405 | -5.19994 | 1.46E-06 | 4.60E-06 | 4.56834 |
| IFI35 | 0.454466 | 1.370276 | 7.378101 | 5.197701 | 1.47E-06 | 4.64E-06 | 4.559548 |

| | | | | | | | |
|---|---|---|---|---|---|---|---|
| NDUFAF4 | -0.31704 | 0.802718 | 7.602785 | -5.19624 | 1.48E-06 | 4.67E-06 | 4.553819 |
| ALLC | -0.28904 | 0.818444 | 4.898608 | -5.19507 | 1.49E-06 | 4.69E-06 | 4.549231 |
| TSR2 | -0.17425 | 0.886229 | 7.808734 | -5.19492 | 1.49E-06 | 4.69E-06 | 4.548651 |
| OR1D2 | -0.21316 | 0.862644 | 5.731392 | -5.19232 | 1.50E-06 | 4.74E-06 | 4.538458 |
| ASIC4 | -0.20891 | 0.865193 | 5.490127 | -5.19175 | 1.51E-06 | 4.75E-06 | 4.536228 |
| IL36A | -0.2187 | 0.859338 | 5.008861 | -5.19155 | 1.51E-06 | 4.75E-06 | 4.535446 |
| AIM2 | 0.415224 | 1.333506 | 5.136709 | 5.187868 | 1.53E-06 | 4.82E-06 | 4.521035 |
| RIN1 | -0.22729 | 0.854237 | 6.10962 | -5.18758 | 1.53E-06 | 4.82E-06 | 4.519903 |
| STARD8 | -0.3309 | 0.795043 | 6.948734 | -5.18729 | 1.53E-06 | 4.82E-06 | 4.518779 |
| PRKACG | -0.21503 | 0.861526 | 5.162785 | -5.18727 | 1.53E-06 | 4.82E-06 | 4.518675 |
| PRSS16 | 0.284835 | 1.218271 | 4.582025 | 5.185474 | 1.54E-06 | 4.86E-06 | 4.511662 |
| KANK2 | 0.264374 | 1.201115 | 7.63 | 5.184198 | 1.55E-06 | 4.88E-06 | 4.50667 |
| F12 | 0.36031 | 1.283701 | 5.778987 | 5.183203 | 1.56E-06 | 4.90E-06 | 4.502776 |
| PRIM1 | 0.320152 | 1.248462 | 5.958734 | 5.182863 | 1.56E-06 | 4.90E-06 | 4.501446 |
| ZDHHC24 | -0.1737 | 0.886569 | 7.181139 | -5.17927 | 1.58E-06 | 4.97E-06 | 4.487391 |
| SOCS2 | -0.70109 | 0.615106 | 6.789747 | -5.17656 | 1.60E-06 | 5.03E-06 | 4.476768 |
| UBE2B | -0.16039 | 0.894784 | 8.392785 | -5.17626 | 1.60E-06 | 5.03E-06 | 4.475605 |
| ABHD11 | 0.244084 | 1.184341 | 7.058481 | 5.175997 | 1.60E-06 | 5.03E-06 | 4.474586 |
| KCNG2 | -0.21791 | 0.859809 | 4.726582 | -5.17595 | 1.61E-06 | 5.03E-06 | 4.474388 |
| APOLD1 | -0.64894 | 0.637749 | 5.783924 | -5.17543 | 1.61E-06 | 5.04E-06 | 4.472377 |
| CRMP1 | -0.2435 | 0.844695 | 6.616329 | -5.17451 | 1.61E-06 | 5.06E-06 | 4.468789 |
| TPPP3 | -0.37511 | 0.771046 | 5.824177 | -5.17217 | 1.63E-06 | 5.10E-06 | 4.459623 |
| DPAGT1 | 0.224638 | 1.168484 | 7.242532 | 5.170913 | 1.64E-06 | 5.13E-06 | 4.454708 |
| S1PR1 | 0.414618 | 1.332946 | 5.206456 | 5.169171 | 1.65E-06 | 5.16E-06 | 4.4479 |
| TMBIM1 | 0.233162 | 1.175408 | 8.676962 | 5.167366 | 1.66E-06 | 5.20E-06 | 4.440847 |
| ADGRE2 | 0.308248 | 1.238203 | 4.083544 | 5.166738 | 1.67E-06 | 5.21E-06 | 4.438394 |
| FEZ2 | 0.210547 | 1.157127 | 8.831646 | 5.165992 | 1.67E-06 | 5.22E-06 | 4.435481 |
| DPYSL2 | 0.326285 | 1.25378 | 9.464557 | 5.164832 | 1.68E-06 | 5.25E-06 | 4.430949 |
| SNAPC2 | -0.2116 | 0.863578 | 6.031392 | -5.16455 | 1.68E-06 | 5.25E-06 | 4.429855 |
| NDC80 | 0.273933 | 1.209099 | 3.825949 | 5.1644 | 1.68E-06 | 5.25E-06 | 4.429259 |
| KBTBD2 | 0.253057 | 1.191729 | 7.06962 | 5.162718 | 1.69E-06 | 5.28E-06 | 4.42269 |
| SLC16A5 | 0.387437 | 1.308068 | 5.843797 | 5.160405 | 1.71E-06 | 5.33E-06 | 4.413659 |
| CSE1L | 0.221087 | 1.165611 | 7.811266 | 5.159124 | 1.72E-06 | 5.36E-06 | 4.408656 |
| SNRPF | 0.234071 | 1.176149 | 8.527342 | 5.15896 | 1.72E-06 | 5.36E-06 | 4.408017 |
| OPLAH | -0.3058 | 0.808995 | 5.781392 | -5.15877 | 1.72E-06 | 5.36E-06 | 4.407268 |
| MISP | -0.27374 | 0.827171 | 7.81 | -5.1578 | 1.73E-06 | 5.38E-06 | 4.403499 |
| CARHSP1 | 0.331377 | 1.258214 | 8.27038 | 5.157768 | 1.73E-06 | 5.38E-06 | 4.403362 |
| NPRL2 | -0.167 | 0.890695 | 7.876329 | -5.15515 | 1.75E-06 | 5.43E-06 | 4.393132 |
| TP53BP2 | 0.326001 | 1.253534 | 6.669873 | 5.153869 | 1.75E-06 | 5.46E-06 | 4.388143 |
| MYD88 | 0.259671 | 1.197205 | 8.704557 | 5.153386 | 1.76E-06 | 5.47E-06 | 4.386262 |
| PPP1R8 | 0.184163 | 1.136158 | 8.42519 | 5.153228 | 1.76E-06 | 5.47E-06 | 4.385645 |
| MFAP2 | 0.365646 | 1.288458 | 5.518608 | 5.152179 | 1.77E-06 | 5.49E-06 | 4.381551 |
| UAP1L1 | -0.29107 | 0.817297 | 4.864937 | -5.15137 | 1.77E-06 | 5.51E-06 | 4.378378 |
| GRAMD4 | 0.216574 | 1.161971 | 5.591772 | 5.15094 | 1.77E-06 | 5.51E-06 | 4.376715 |
| PPIH | 0.358966 | 1.282506 | 7.028861 | 5.150907 | 1.78E-06 | 5.51E-06 | 4.376588 |
| AMZ2 | -0.23525 | 0.849538 | 8.324557 | -5.15017 | 1.78E-06 | 5.53E-06 | 4.373722 |
| ABCC3 | 0.480883 | 1.395597 | 6.596962 | 5.147154 | 1.80E-06 | 5.59E-06 | 4.361949 |

| | | | | | | | |
|---|---|---|---|---|---|---|---|
| TSPYL5 | 0.411225 | 1.329815 | 6.958734 | 5.144381 | 1.82E-06 | 5.65E-06 | 4.351137 |
| WASF2 | 0.303228 | 1.233902 | 7.131013 | 5.143973 | 1.83E-06 | 5.66E-06 | 4.349548 |
| CEND1 | -0.21748 | 0.860068 | 6.096329 | -5.14237 | 1.84E-06 | 5.70E-06 | 4.343304 |
| ASCC2 | -0.19759 | 0.872007 | 5.650506 | -5.14206 | 1.84E-06 | 5.70E-06 | 4.342094 |
| GCAT | -0.32174 | 0.800105 | 7.647342 | -5.14161 | 1.84E-06 | 5.71E-06 | 4.340318 |
| KRT4 | -0.23903 | 0.847314 | 5.396456 | -5.1406 | 1.85E-06 | 5.73E-06 | 4.336387 |
| MYL10 | -0.2044 | 0.867899 | 5.961139 | -5.13873 | 1.86E-06 | 5.77E-06 | 4.329102 |
| SH3GLB1 | 0.187431 | 1.138734 | 8.671772 | 5.138068 | 1.87E-06 | 5.78E-06 | 4.326533 |
| BMP7 | -0.18671 | 0.878605 | 5.202658 | -5.13775 | 1.87E-06 | 5.79E-06 | 4.325313 |
| CD8A | 0.477563 | 1.392389 | 5.952532 | 5.137683 | 1.87E-06 | 5.79E-06 | 4.325034 |
| ANOS1 | 0.31195 | 1.241384 | 4.759873 | 5.137154 | 1.88E-06 | 5.80E-06 | 4.322973 |
| SEZ6L2 | 0.312984 | 1.242275 | 5.791392 | 5.137082 | 1.88E-06 | 5.80E-06 | 4.322691 |
| DDX47 | 0.192167 | 1.142479 | 9.04962 | 5.1348 | 1.89E-06 | 5.85E-06 | 4.313805 |
| SEMA6C | -0.26495 | 0.832229 | 6.482152 | -5.13407 | 1.90E-06 | 5.86E-06 | 4.310979 |
| HOXB5 | 0.224578 | 1.168436 | 6.556203 | 5.13391 | 1.90E-06 | 5.87E-06 | 4.310337 |
| GAR1 | 0.219677 | 1.164473 | 7.406329 | 5.133477 | 1.90E-06 | 5.87E-06 | 4.30865 |
| BTBD1 | 0.268623 | 1.204658 | 8.920253 | 5.130767 | 1.92E-06 | 5.94E-06 | 4.298097 |
| INTS9 | 0.218788 | 1.163755 | 6.741392 | 5.130474 | 1.93E-06 | 5.94E-06 | 4.296959 |
| AURKA | 0.233511 | 1.175693 | 5.086456 | 5.129664 | 1.93E-06 | 5.96E-06 | 4.293804 |
| CMAHP | -0.40267 | 0.756458 | 5.278101 | -5.1283 | 1.94E-06 | 5.99E-06 | 4.288476 |
| BCCIP | -0.29707 | 0.813905 | 7.428734 | -5.12819 | 1.94E-06 | 5.99E-06 | 4.28807 |
| ULK1 | -0.26041 | 0.834852 | 6.26557 | -5.12467 | 1.97E-06 | 6.07E-06 | 4.274351 |
| SDR39U1 | -0.1952 | 0.873449 | 8.650633 | -5.12452 | 1.97E-06 | 6.07E-06 | 4.273801 |
| HEPH | 0.459018 | 1.374606 | 4.958481 | 5.121023 | 2.00E-06 | 6.16E-06 | 4.260179 |
| DYNC1LI1 | -0.21417 | 0.862042 | 7.203797 | -5.11995 | 2.01E-06 | 6.18E-06 | 4.255994 |
| TRPM6 | -0.25571 | 0.837574 | 5.122532 | -5.11826 | 2.02E-06 | 6.22E-06 | 4.249431 |
| SLC25A3 | -0.14457 | 0.904648 | 12.1557 | -5.11637 | 2.04E-06 | 6.27E-06 | 4.242072 |
| RIOK3 | -0.18415 | 0.880167 | 7.431772 | -5.1127 | 2.07E-06 | 6.36E-06 | 4.22781 |
| CDX2 | -0.2445 | 0.844109 | 5.337215 | -5.11156 | 2.08E-06 | 6.38E-06 | 4.223394 |
| PSMA4 | 0.192819 | 1.142995 | 9.129241 | 5.109855 | 2.09E-06 | 6.42E-06 | 4.216765 |
| CA10 | -0.27513 | 0.826375 | 4.600506 | -5.10879 | 2.10E-06 | 6.45E-06 | 4.212624 |
| CXCL14 | -0.43872 | 0.737788 | 12.12152 | -5.10784 | 2.11E-06 | 6.47E-06 | 4.20895 |
| CSNK1G3 | 0.24386 | 1.184157 | 6.840127 | 5.10613 | 2.12E-06 | 6.51E-06 | 4.202297 |
| MORN1 | -0.21175 | 0.863488 | 5.316329 | -5.10342 | 2.15E-06 | 6.58E-06 | 4.191781 |
| FAHD2A | -0.26062 | 0.83473 | 6.024177 | -5.10329 | 2.15E-06 | 6.58E-06 | 4.191253 |
| GPX5 | -0.16979 | 0.888973 | 4.23038 | -5.10144 | 2.16E-06 | 6.63E-06 | 4.184079 |
| FLII | 0.219598 | 1.164409 | 7.244557 | 5.099478 | 2.18E-06 | 6.68E-06 | 4.176469 |
| WNT8B | -0.27158 | 0.828411 | 5.581772 | -5.09825 | 2.19E-06 | 6.71E-06 | 4.17169 |
| HIST1H2AI | -0.29536 | 0.814871 | 4.763797 | -5.0978 | 2.20E-06 | 6.72E-06 | 4.169975 |
| BARD1 | 0.216008 | 1.161515 | 4.832405 | 5.096482 | 2.21E-06 | 6.75E-06 | 4.164843 |
| CKAP2 | 0.264104 | 1.20089 | 4.572278 | 5.095721 | 2.21E-06 | 6.77E-06 | 4.161892 |
| ENO1 | 0.260909 | 1.198234 | 10.60899 | 5.095168 | 2.22E-06 | 6.78E-06 | 4.159745 |
| DCAF11 | -0.1874 | 0.878188 | 7.780633 | -5.0938 | 2.23E-06 | 6.82E-06 | 4.15442 |
| RAB3A | -0.20049 | 0.870252 | 5.18038 | -5.09345 | 2.23E-06 | 6.83E-06 | 4.153098 |
| MRPL44 | -0.29942 | 0.812579 | 6.291013 | -5.08928 | 2.27E-06 | 6.94E-06 | 4.13692 |
| MTOR | -0.24096 | 0.846181 | 7.401519 | -5.08924 | 2.27E-06 | 6.94E-06 | 4.136757 |
| SMYD5 | -0.22859 | 0.853468 | 6.638861 | -5.08729 | 2.29E-06 | 6.99E-06 | 4.129184 |

| | | | | | | | |
|---|---|---|---|---|---|---|---|
| ADRB2 | 0.305553 | 1.235893 | 4.62481 | 5.08687 | 2.29E-06 | 7.00E-06 | 4.127565 |
| FAM111A | 0.242095 | 1.182709 | 7.497215 | 5.086626 | 2.30E-06 | 7.00E-06 | 4.12662 |
| ACOX3 | -0.20327 | 0.868577 | 5.921392 | -5.08316 | 2.33E-06 | 7.10E-06 | 4.113178 |
| CCT2 | 0.215823 | 1.161367 | 9.424937 | 5.081764 | 2.34E-06 | 7.13E-06 | 4.107777 |
| SCNN1B | -0.49297 | 0.71056 | 6.42038 | -5.08056 | 2.35E-06 | 7.17E-06 | 4.10311 |
| USP22 | 0.211337 | 1.157761 | 8.449367 | 5.07927 | 2.37E-06 | 7.20E-06 | 4.098119 |
| STX18 | 0.204895 | 1.152602 | 7.964937 | 5.07684 | 2.39E-06 | 7.27E-06 | 4.088707 |
| SNW1 | 0.195883 | 1.145425 | 7.542911 | 5.076413 | 2.39E-06 | 7.28E-06 | 4.087053 |
| SMARCAL1 | 0.241311 | 1.182066 | 6.407975 | 5.075054 | 2.41E-06 | 7.31E-06 | 4.081792 |
| PGAM2 | -0.24607 | 0.843188 | 4.620253 | -5.07471 | 2.41E-06 | 7.32E-06 | 4.080452 |
| CPD | 0.323076 | 1.250995 | 6.834304 | 5.074567 | 2.41E-06 | 7.32E-06 | 4.079905 |
| OTUD3 | -0.27219 | 0.828059 | 5.793038 | -5.07376 | 2.42E-06 | 7.34E-06 | 4.076776 |
| FRAT1 | 0.31108 | 1.240636 | 5.40038 | 5.07289 | 2.43E-06 | 7.37E-06 | 4.073412 |
| ALPPL2 | -0.24271 | 0.845158 | 5.484051 | -5.07233 | 2.43E-06 | 7.38E-06 | 4.071259 |
| CCT5 | 0.2517 | 1.190609 | 9.002532 | 5.071415 | 2.44E-06 | 7.41E-06 | 4.067704 |
| NID2 | 0.5122 | 1.426224 | 6.455696 | 5.070813 | 2.45E-06 | 7.42E-06 | 4.065374 |
| PARD3 | 0.181133 | 1.133774 | 7.283924 | 5.069729 | 2.46E-06 | 7.45E-06 | 4.061179 |
| NEDD9 | -0.45102 | 0.731525 | 7.467468 | -5.06886 | 2.47E-06 | 7.47E-06 | 4.057817 |
| SLC48A1 | 0.254631 | 1.193031 | 6.536582 | 5.068845 | 2.47E-06 | 7.47E-06 | 4.057759 |
| CA11 | -0.22843 | 0.853562 | 5.452405 | -5.06563 | 2.50E-06 | 7.56E-06 | 4.045314 |
| ARHGEF28 | -0.28428 | 0.82115 | 7.97038 | -5.06463 | 2.51E-06 | 7.59E-06 | 4.041447 |
| CA8 | -0.40441 | 0.755543 | 5.350633 | -5.06304 | 2.52E-06 | 7.64E-06 | 4.035293 |
| JUND | -0.38408 | 0.766269 | 6.111519 | -5.06221 | 2.53E-06 | 7.66E-06 | 4.032077 |
| TTK | 0.214025 | 1.15992 | 3.78962 | 5.059595 | 2.56E-06 | 7.74E-06 | 4.021988 |
| MSN | 0.2911 | 1.223573 | 8.497468 | 5.058718 | 2.57E-06 | 7.76E-06 | 4.018594 |
| RNF208 | -0.23063 | 0.852265 | 5.608228 | -5.05857 | 2.57E-06 | 7.76E-06 | 4.018019 |
| KDM7A | 0.220112 | 1.164824 | 5.786076 | 5.058452 | 2.57E-06 | 7.76E-06 | 4.017567 |
| SGPP1 | -0.47993 | 0.71701 | 7.694304 | -5.05842 | 2.57E-06 | 7.76E-06 | 4.017452 |
| TMC6 | 0.254848 | 1.19321 | 5.336456 | 5.058247 | 2.57E-06 | 7.77E-06 | 4.016774 |
| FN3K | -0.23357 | 0.850527 | 4.217215 | -5.05651 | 2.59E-06 | 7.82E-06 | 4.010065 |
| ABCC5 | 0.242352 | 1.182919 | 7.177975 | 5.05172 | 2.64E-06 | 7.96E-06 | 3.991559 |
| MAP1LC3C | 0.283768 | 1.21737 | 4.458101 | 5.051711 | 2.64E-06 | 7.96E-06 | 3.991524 |
| RIN2 | 0.309895 | 1.239617 | 9.513797 | 5.043296 | 2.73E-06 | 8.23E-06 | 3.959035 |
| RBM10 | 0.207793 | 1.15492 | 6.856582 | 5.042941 | 2.73E-06 | 8.24E-06 | 3.957662 |
| AREL1 | 0.294183 | 1.226191 | 7.096582 | 5.041076 | 2.75E-06 | 8.30E-06 | 3.950468 |
| TRAPPC13 | -0.24256 | 0.845243 | 4.750633 | -5.04084 | 2.76E-06 | 8.30E-06 | 3.949548 |
| TRAF3IP3 | 0.480876 | 1.395591 | 4.938481 | 5.039359 | 2.77E-06 | 8.35E-06 | 3.943845 |
| OIP5 | 0.184137 | 1.136137 | 4.121266 | 5.038527 | 2.78E-06 | 8.37E-06 | 3.940634 |
| PYROXD1 | -0.34472 | 0.787459 | 9.317089 | -5.03655 | 2.80E-06 | 8.44E-06 | 3.932999 |
| ICE2 | 0.235237 | 1.1771 | 5.718481 | 5.035947 | 2.81E-06 | 8.45E-06 | 3.930682 |
| COX15 | -0.19673 | 0.872524 | 6.983038 | -5.03453 | 2.83E-06 | 8.50E-06 | 3.925233 |
| STK38 | 0.307121 | 1.237236 | 7.093291 | 5.033801 | 2.83E-06 | 8.52E-06 | 3.922409 |
| SSBP3 | -0.35593 | 0.781366 | 6.910886 | -5.03063 | 2.87E-06 | 8.62E-06 | 3.910169 |
| WTAP | 0.234315 | 1.176348 | 8.291139 | 5.029572 | 2.88E-06 | 8.66E-06 | 3.906108 |
| ENC1 | 0.256324 | 1.194431 | 5.236203 | 5.029063 | 2.89E-06 | 8.67E-06 | 3.904148 |
| NRXN2 | -0.28735 | 0.819409 | 5.296709 | -5.02868 | 2.89E-06 | 8.68E-06 | 3.902685 |
| CELSR2 | 0.207885 | 1.154994 | 6.305316 | 5.028663 | 2.89E-06 | 8.68E-06 | 3.902606 |

| | | | | | | | |
|---|---|---|---|---|---|---|---|
| ARHGEF9 | -0.27637 | 0.825666 | 6.156076 | -5.02865 | 2.89E-06 | 8.68E-06 | 3.902556 |
| ARF4 | 0.206186 | 1.153634 | 9.907215 | 5.028448 | 2.90E-06 | 8.68E-06 | 3.901778 |
| GNLY | 0.314223 | 1.243342 | 4.715823 | 5.027641 | 2.91E-06 | 8.71E-06 | 3.898669 |
| C1QL1 | -0.22606 | 0.854966 | 6.23557 | -5.02569 | 2.93E-06 | 8.77E-06 | 3.891169 |
| LINC00472 | -0.34984 | 0.784674 | 4.985823 | -5.0253 | 2.93E-06 | 8.78E-06 | 3.889647 |
| TMEM53 | -0.26873 | 0.830051 | 7.314051 | -5.02436 | 2.94E-06 | 8.81E-06 | 3.886031 |
| NSDHL | -0.21479 | 0.861672 | 6.406582 | -5.02351 | 2.95E-06 | 8.84E-06 | 3.882768 |
| SIDT1 | 0.22776 | 1.171016 | 3.962532 | 5.021931 | 2.97E-06 | 8.89E-06 | 3.876674 |
| PCDHGB5 | -0.2124 | 0.863098 | 5.756709 | -5.02134 | 2.98E-06 | 8.91E-06 | 3.874415 |
| MIA3 | -0.18182 | 0.881587 | 5.94557 | -5.02028 | 2.99E-06 | 8.94E-06 | 3.870337 |
| TEF | -0.22353 | 0.856467 | 5.362278 | -5.01302 | 3.08E-06 | 9.20E-06 | 3.842389 |
| ASMTL | -0.21898 | 0.859173 | 7.472658 | -5.0111 | 3.10E-06 | 9.27E-06 | 3.834988 |
| BCL7A | -0.20841 | 0.865493 | 6.314684 | -5.01091 | 3.10E-06 | 9.27E-06 | 3.834249 |
| RASAL1 | 0.225 | 1.168777 | 6.638987 | 5.010871 | 3.11E-06 | 9.27E-06 | 3.834114 |
| CLN6 | -0.19002 | 0.876594 | 5.92519 | -5.01004 | 3.12E-06 | 9.30E-06 | 3.830905 |
| GPRASP1 | -0.35773 | 0.780389 | 6.247089 | -5.00858 | 3.13E-06 | 9.35E-06 | 3.825289 |
| RPS20 | 0.344427 | 1.269647 | 11.15127 | 4.999007 | 3.25E-06 | 9.70E-06 | 3.788509 |
| THY1 | -0.8066 | 0.571727 | 9.447848 | -4.99899 | 3.25E-06 | 9.70E-06 | 3.788451 |
| SLC29A1 | 0.249381 | 1.188697 | 5.907215 | 4.997099 | 3.28E-06 | 9.77E-06 | 3.781182 |
| ARMCX1 | 0.30025 | 1.231358 | 7.717595 | 4.994245 | 3.32E-06 | 9.88E-06 | 3.770221 |
| FAM63A | 0.208452 | 1.155448 | 7.934684 | 4.992811 | 3.34E-06 | 9.93E-06 | 3.764715 |
| BLCAP | -0.17194 | 0.887646 | 9.277089 | -4.99263 | 3.34E-06 | 9.94E-06 | 3.76404 |
| GPR32 | -0.22202 | 0.857367 | 4.072911 | -4.99078 | 3.36E-06 | 1.00E-05 | 3.756903 |
| MAN2C1 | -0.19088 | 0.876074 | 6.922658 | -4.9894 | 3.38E-06 | 1.01E-05 | 3.751631 |
| KCNK15 | 0.30807 | 1.23805 | 4.204557 | 4.988896 | 3.39E-06 | 1.01E-05 | 3.74969 |
| CCL20 | 0.838386 | 1.788049 | 4.963038 | 4.987265 | 3.41E-06 | 1.01E-05 | 3.743434 |
| NAP1L3 | 0.297424 | 1.228948 | 4.799241 | 4.986525 | 3.42E-06 | 1.02E-05 | 3.740594 |
| HMGCS2 | -0.85443 | 0.553085 | 7.565696 | -4.98444 | 3.45E-06 | 1.02E-05 | 3.732593 |
| ICAM1 | 0.342148 | 1.267642 | 6.066835 | 4.983592 | 3.46E-06 | 1.03E-05 | 3.729346 |
| BEGAIN | -0.26674 | 0.831196 | 5.315316 | -4.98274 | 3.47E-06 | 1.03E-05 | 3.726091 |
| PTPRF | 0.245145 | 1.185212 | 7.063924 | 4.98267 | 3.47E-06 | 1.03E-05 | 3.725808 |
| CDC37L1 | -0.28586 | 0.820251 | 7.194937 | -4.98143 | 3.49E-06 | 1.04E-05 | 3.721038 |
| TBC1D15 | -0.24857 | 0.84173 | 7.135949 | -4.98056 | 3.50E-06 | 1.04E-05 | 3.717727 |
| PDE6A | -0.27417 | 0.826926 | 4.858734 | -4.97867 | 3.53E-06 | 1.05E-05 | 3.710487 |
| R3HDM2 | -0.34848 | 0.785412 | 8.302911 | -4.97849 | 3.53E-06 | 1.05E-05 | 3.709778 |
| VPS4B | 0.266028 | 1.202492 | 7.708734 | 4.977693 | 3.54E-06 | 1.05E-05 | 3.706728 |
| F3 | -0.57354 | 0.671967 | 6.187595 | -4.97669 | 3.56E-06 | 1.05E-05 | 3.7029 |
| SNRPD1 | 0.175962 | 1.129717 | 7.326329 | 4.976362 | 3.56E-06 | 1.05E-05 | 3.701627 |
| CYP2J2 | -0.37496 | 0.771127 | 6.279241 | -4.97559 | 3.57E-06 | 1.06E-05 | 3.69867 |
| MGAT2 | 0.217951 | 1.163081 | 6.344304 | 4.97474 | 3.58E-06 | 1.06E-05 | 3.695412 |
| NOD2 | 0.201989 | 1.150283 | 4.03481 | 4.974736 | 3.58E-06 | 1.06E-05 | 3.695395 |
| HTR1B | -0.19301 | 0.874778 | 4.92481 | -4.97383 | 3.60E-06 | 1.06E-05 | 3.691925 |
| OAS1 | 0.658083 | 1.577984 | 6.227722 | 4.973079 | 3.61E-06 | 1.07E-05 | 3.689047 |
| RBM38 | -0.22981 | 0.852748 | 6.169873 | -4.97284 | 3.61E-06 | 1.07E-05 | 3.688126 |
| HMHA1 | 0.255758 | 1.193963 | 5.866835 | 4.972445 | 3.62E-06 | 1.07E-05 | 3.686619 |
| AP3M2 | 0.218267 | 1.163336 | 7.011392 | 4.969908 | 3.65E-06 | 1.08E-05 | 3.676901 |
| FOSL1 | -0.4148 | 0.750122 | 4.086076 | -4.96953 | 3.66E-06 | 1.08E-05 | 3.675436 |

| | | | | | | | |
|---|---|---|---|---|---|---|---|
| TIMP2 | 0.365896 | 1.288682 | 7.290886 | 4.968439 | 3.67E-06 | 1.08E-05 | 3.671278 |
| UBA7 | 0.228142 | 1.171326 | 6.64443 | 4.967902 | 3.68E-06 | 1.09E-05 | 3.66922 |
| TAOK3 | 0.246298 | 1.186159 | 7.178101 | 4.967215 | 3.69E-06 | 1.09E-05 | 3.666593 |
| ARL5A | -0.25933 | 0.835477 | 8.466456 | -4.96629 | 3.70E-06 | 1.09E-05 | 3.663033 |
| TMEM140 | 0.415119 | 1.333408 | 8.111013 | 4.965703 | 3.71E-06 | 1.09E-05 | 3.660801 |
| EBAG9 | -0.18742 | 0.878176 | 8.03519 | -4.96393 | 3.74E-06 | 1.10E-05 | 3.654017 |
| GCLM | -0.37794 | 0.769533 | 6.857342 | -4.96256 | 3.76E-06 | 1.11E-05 | 3.648757 |
| VEGFC | 0.339901 | 1.26567 | 5.52481 | 4.96102 | 3.78E-06 | 1.11E-05 | 3.642884 |
| NFE2L1 | -0.26257 | 0.833602 | 9.553797 | -4.96095 | 3.78E-06 | 1.11E-05 | 3.642616 |
| EHBP1L1 | 0.215072 | 1.160762 | 5.028101 | 4.960888 | 3.78E-06 | 1.11E-05 | 3.642378 |
| ELL2 | -0.5599 | 0.678349 | 6.303291 | -4.96033 | 3.79E-06 | 1.11E-05 | 3.640249 |
| MCTP1 | 0.234262 | 1.176305 | 4.093291 | 4.960288 | 3.79E-06 | 1.11E-05 | 3.640082 |
| TLE2 | 0.24448 | 1.184665 | 5.527342 | 4.959113 | 3.81E-06 | 1.12E-05 | 3.635588 |
| C2orf44 | 0.20529 | 1.152918 | 5.063797 | 4.956513 | 3.85E-06 | 1.13E-05 | 3.625642 |
| P2RY14 | 0.339605 | 1.26541 | 4.273165 | 4.95561 | 3.86E-06 | 1.13E-05 | 3.622191 |
| ABCA5 | -0.21404 | 0.862117 | 6.354937 | -4.95473 | 3.88E-06 | 1.14E-05 | 3.618843 |
| B3GNT4 | -0.27818 | 0.82463 | 6.933797 | -4.95459 | 3.88E-06 | 1.14E-05 | 3.618297 |
| PNMA3 | -0.24221 | 0.845451 | 5.528608 | -4.95332 | 3.90E-06 | 1.14E-05 | 3.613418 |
| HSPA8 | -0.2099 | 0.864596 | 12.15949 | -4.9523 | 3.92E-06 | 1.15E-05 | 3.609536 |
| SRSF6 | -0.23995 | 0.846776 | 9.007595 | -4.95156 | 3.93E-06 | 1.15E-05 | 3.606695 |
| FGR | 0.35836 | 1.281967 | 5.248608 | 4.949282 | 3.96E-06 | 1.16E-05 | 3.598002 |
| PEX3 | -0.2502 | 0.840777 | 8.132658 | -4.94813 | 3.98E-06 | 1.17E-05 | 3.593601 |
| ACSL1 | -0.37294 | 0.772205 | 10.01443 | -4.94757 | 3.99E-06 | 1.17E-05 | 3.591457 |
| HR | -0.20281 | 0.868855 | 5.295063 | -4.94742 | 3.99E-06 | 1.17E-05 | 3.590889 |
| MPC2 | -0.22107 | 0.857931 | 10.46418 | -4.94697 | 4.00E-06 | 1.17E-05 | 3.589161 |
| COA7 | -0.2184 | 0.859519 | 7.248987 | -4.94633 | 4.01E-06 | 1.17E-05 | 3.586738 |
| KCNJ2 | 0.498287 | 1.412536 | 5.713797 | 4.944116 | 4.04E-06 | 1.18E-05 | 3.578268 |
| HOXA1 | 0.193406 | 1.14346 | 4.734051 | 4.944043 | 4.04E-06 | 1.18E-05 | 3.57799 |
| MAFF | -1.30303 | 0.405274 | 6.895696 | -4.94327 | 4.06E-06 | 1.19E-05 | 3.575037 |
| MSH6 | 0.263808 | 1.200643 | 6.640633 | 4.941473 | 4.09E-06 | 1.19E-05 | 3.568178 |
| DDOST | 0.213992 | 1.159893 | 10.12722 | 4.939267 | 4.12E-06 | 1.20E-05 | 3.559754 |
| UBXN6 | -0.21503 | 0.861526 | 7.482785 | -4.93925 | 4.12E-06 | 1.20E-05 | 3.559685 |
| PIGL | -0.2791 | 0.824106 | 5.724937 | -4.93906 | 4.12E-06 | 1.20E-05 | 3.558963 |
| ITPKC | -0.26024 | 0.834951 | 6.176076 | -4.93857 | 4.13E-06 | 1.21E-05 | 3.557111 |
| ABTB2 | -0.22518 | 0.85549 | 6.542025 | -4.93828 | 4.14E-06 | 1.21E-05 | 3.555998 |
| STX4 | 0.246126 | 1.186018 | 7.927595 | 4.938214 | 4.14E-06 | 1.21E-05 | 3.555735 |
| THNSL2 | -0.6228 | 0.649409 | 7.036582 | -4.93297 | 4.22E-06 | 1.23E-05 | 3.535746 |
| MUC13 | -0.78797 | 0.579158 | 6.222152 | -4.92986 | 4.28E-06 | 1.25E-05 | 3.523884 |
| PSORS1C2 | -0.23947 | 0.847055 | 5.278228 | -4.92964 | 4.28E-06 | 1.25E-05 | 3.523023 |
| TAF7 | 0.23581 | 1.177568 | 8.626329 | 4.926983 | 4.33E-06 | 1.26E-05 | 3.5129 |
| SMAD2 | 0.173689 | 1.127939 | 7.70038 | 4.925567 | 4.35E-06 | 1.27E-05 | 3.507502 |
| GJA3 | -0.19225 | 0.875238 | 4.790127 | -4.92536 | 4.35E-06 | 1.27E-05 | 3.506717 |
| KLRD1 | 0.212391 | 1.158607 | 4.616329 | 4.924734 | 4.36E-06 | 1.27E-05 | 3.504329 |
| RTEL1 | -0.24642 | 0.842984 | 7.370759 | -4.92384 | 4.38E-06 | 1.27E-05 | 3.500913 |
| FLRT1 | -0.25825 | 0.836099 | 5.388861 | -4.92377 | 4.38E-06 | 1.27E-05 | 3.500658 |
| PROC | -0.54725 | 0.684325 | 7.515316 | -4.92282 | 4.40E-06 | 1.28E-05 | 3.497039 |
| GALE | -0.2487 | 0.841657 | 8.03481 | -4.92243 | 4.40E-06 | 1.28E-05 | 3.495551 |

| | | | | | | | |
|---|---|---|---|---|---|---|---|
| GOT2 | -0.21301 | 0.862735 | 10.36646 | -4.92007 | 4.45E-06 | 1.29E-05 | 3.486571 |
| BCL6 | 0.400784 | 1.320225 | 6.346329 | 4.917358 | 4.49E-06 | 1.30E-05 | 3.476229 |
| TAC3 | -0.25669 | 0.837004 | 6.908861 | -4.91691 | 4.50E-06 | 1.30E-05 | 3.474521 |
| CHRND | -0.18628 | 0.878866 | 4.363924 | -4.9169 | 4.50E-06 | 1.30E-05 | 3.474476 |
| PPM1D | 0.258913 | 1.196577 | 5.769241 | 4.916462 | 4.51E-06 | 1.31E-05 | 3.472818 |
| SPRY2 | -0.32291 | 0.799455 | 6.627722 | -4.91591 | 4.52E-06 | 1.31E-05 | 3.470709 |
| GDPD2 | -0.28518 | 0.820637 | 6.328481 | -4.91382 | 4.56E-06 | 1.32E-05 | 3.462742 |
| CDC42SE1 | 0.297095 | 1.228668 | 8.08519 | 4.911605 | 4.60E-06 | 1.33E-05 | 3.454329 |
| GUCA2A | -0.2425 | 0.845281 | 5.655443 | -4.91141 | 4.60E-06 | 1.33E-05 | 3.453603 |
| MAPK8IP2 | -0.19377 | 0.874315 | 4.951013 | -4.91106 | 4.61E-06 | 1.33E-05 | 3.452274 |
| DACT1 | 0.451871 | 1.367813 | 5.066582 | 4.91092 | 4.61E-06 | 1.33E-05 | 3.451725 |
| TCTN2 | 0.285804 | 1.219089 | 8.708734 | 4.909181 | 4.64E-06 | 1.34E-05 | 3.445108 |
| FAM182B | -0.40016 | 0.757775 | 5.589367 | -4.90843 | 4.65E-06 | 1.34E-05 | 3.442268 |
| ACKR3 | -0.56625 | 0.675372 | 6.316076 | -4.90843 | 4.65E-06 | 1.34E-05 | 3.44226 |
| LMAN2 | 0.219361 | 1.164218 | 8.139241 | 4.908259 | 4.66E-06 | 1.34E-05 | 3.441598 |
| ITGA7 | -0.22287 | 0.856862 | 6.168861 | -4.90723 | 4.68E-06 | 1.35E-05 | 3.437683 |
| HOXC10 | -0.27696 | 0.825327 | 7.892785 | -4.90718 | 4.68E-06 | 1.35E-05 | 3.437512 |
| GNMT | -0.25504 | 0.837964 | 4.907595 | -4.90677 | 4.68E-06 | 1.35E-05 | 3.435944 |
| STMN1 | 0.183412 | 1.135567 | 6.338354 | 4.904474 | 4.73E-06 | 1.36E-05 | 3.427203 |
| AHCYL2 | -0.22963 | 0.852853 | 6.868861 | -4.90268 | 4.76E-06 | 1.37E-05 | 3.42038 |
| CEP135 | -0.23507 | 0.849646 | 5.402025 | -4.90132 | 4.79E-06 | 1.38E-05 | 3.415222 |
| NSUN5P1 | 0.352951 | 1.277171 | 5.595696 | 4.897024 | 4.87E-06 | 1.40E-05 | 3.398887 |
| CLTCL1 | -0.19528 | 0.873401 | 5.928861 | -4.89683 | 4.87E-06 | 1.40E-05 | 3.398161 |
| FKBP5 | -1.03581 | 0.487742 | 7.889494 | -4.89621 | 4.88E-06 | 1.41E-05 | 3.395803 |
| SLC25A24 | 0.26475 | 1.201428 | 7.883418 | 4.893116 | 4.94E-06 | 1.42E-05 | 3.384043 |
| RPN2 | 0.160652 | 1.117792 | 10.36646 | 4.891589 | 4.97E-06 | 1.43E-05 | 3.378244 |
| DHX9 | 0.200329 | 1.148961 | 6.217595 | 4.891397 | 4.98E-06 | 1.43E-05 | 3.377516 |
| ABHD17B | -0.24003 | 0.846726 | 6.807342 | -4.89138 | 4.98E-06 | 1.43E-05 | 3.377462 |
| LEMD3 | 0.189078 | 1.140035 | 7.662025 | 4.890205 | 5.00E-06 | 1.44E-05 | 3.372991 |
| C10orf2 | 0.230198 | 1.172996 | 5.970506 | 4.889888 | 5.00E-06 | 1.44E-05 | 3.371788 |
| COQ2 | 0.27552 | 1.210431 | 6.099873 | 4.889278 | 5.02E-06 | 1.44E-05 | 3.369473 |
| FUT4 | 0.297497 | 1.22901 | 5.462532 | 4.888185 | 5.04E-06 | 1.45E-05 | 3.365323 |
| NOV | 0.438109 | 1.354828 | 5.369747 | 4.885841 | 5.08E-06 | 1.46E-05 | 3.356425 |
| GALNT6 | 0.23527 | 1.177127 | 4.560886 | 4.885558 | 5.09E-06 | 1.46E-05 | 3.355352 |
| ARHGAP28 | -0.21492 | 0.861593 | 4.516962 | -4.88449 | 5.11E-06 | 1.47E-05 | 3.351317 |
| SMAD7 | -0.36292 | 0.777587 | 6.704051 | -4.88361 | 5.13E-06 | 1.47E-05 | 3.347966 |
| PDCD10 | 0.195086 | 1.144792 | 9.806709 | 4.88223 | 5.16E-06 | 1.48E-05 | 3.342726 |
| TSPAN15 | 0.237615 | 1.179042 | 5.220127 | 4.881888 | 5.16E-06 | 1.48E-05 | 3.341429 |
| KIAA1467 | -0.2599 | 0.835145 | 6.478608 | -4.8817 | 5.17E-06 | 1.48E-05 | 3.340712 |
| GJB4 | -0.25845 | 0.835984 | 5.68443 | -4.88009 | 5.20E-06 | 1.49E-05 | 3.334608 |
| MTERF1 | 0.25108 | 1.190098 | 4.955316 | 4.880088 | 5.20E-06 | 1.49E-05 | 3.334604 |
| CACNA2D1 | -0.20628 | 0.866767 | 6.04557 | -4.8789 | 5.23E-06 | 1.50E-05 | 3.3301 |
| TMSB15A | 0.366713 | 1.289412 | 5.042532 | 4.878621 | 5.23E-06 | 1.50E-05 | 3.329042 |
| ATR | 0.232312 | 1.174716 | 5.842911 | 4.878315 | 5.24E-06 | 1.50E-05 | 3.327881 |
| MRPL24 | -0.19129 | 0.875822 | 10.11835 | -4.87633 | 5.28E-06 | 1.51E-05 | 3.320344 |
| RHBDL1 | -0.22385 | 0.856279 | 4.68519 | -4.87606 | 5.28E-06 | 1.51E-05 | 3.319316 |
| ZNF267 | 0.320698 | 1.248935 | 5.112658 | 4.874507 | 5.32E-06 | 1.52E-05 | 3.313447 |

| | | | | | | | |
|---|---|---|---|---|---|---|---|
| FCGR3B | 0.919466 | 1.891416 | 4.907342 | 4.874038 | 5.33E-06 | 1.52E-05 | 3.311666 |
| CDV3 | 0.243933 | 1.184216 | 6.803418 | 4.873655 | 5.33E-06 | 1.52E-05 | 3.310218 |
| KIAA0907 | 0.321304 | 1.24946 | 7.302911 | 4.873349 | 5.34E-06 | 1.52E-05 | 3.309057 |
| HAUS3 | 0.217003 | 1.162316 | 5.563038 | 4.871995 | 5.37E-06 | 1.53E-05 | 3.303927 |
| RDX | -0.25301 | 0.839144 | 8.229747 | -4.8718 | 5.37E-06 | 1.53E-05 | 3.303191 |
| CASP8AP2 | 0.210395 | 1.157005 | 5.306582 | 4.870783 | 5.39E-06 | 1.54E-05 | 3.299336 |
| NFKBIL1 | -0.20355 | 0.868411 | 6.33519 | -4.86898 | 5.43E-06 | 1.55E-05 | 3.292507 |
| SST | -0.42111 | 0.746851 | 4.759747 | -4.86891 | 5.43E-06 | 1.55E-05 | 3.292236 |
| GNB2 | 0.195764 | 1.145331 | 7.760253 | 4.867745 | 5.46E-06 | 1.55E-05 | 3.287829 |
| ADAP2 | 0.385909 | 1.306683 | 6.336203 | 4.8663 | 5.49E-06 | 1.56E-05 | 3.282358 |
| RMI1 | 0.234868 | 1.176799 | 6.163544 | 4.866079 | 5.49E-06 | 1.56E-05 | 3.281519 |
| RRAS2 | -0.37792 | 0.769547 | 7.371266 | -4.86543 | 5.51E-06 | 1.57E-05 | 3.279049 |
| NASP | 0.165184 | 1.121309 | 6.491392 | 4.864991 | 5.52E-06 | 1.57E-05 | 3.2774 |
| CEP83 | 0.219466 | 1.164303 | 4.434937 | 4.862604 | 5.57E-06 | 1.58E-05 | 3.268363 |
| OLFML2B | 0.310652 | 1.240268 | 4.879114 | 4.857305 | 5.69E-06 | 1.62E-05 | 3.248316 |
| MECOM | 0.51751 | 1.431482 | 8.171392 | 4.856523 | 5.70E-06 | 1.62E-05 | 3.245358 |
| SURF2 | -0.2144 | 0.861904 | 5.766962 | -4.85545 | 5.73E-06 | 1.63E-05 | 3.241288 |
| PRDX2 | -0.20435 | 0.867931 | 10.50899 | -4.85483 | 5.74E-06 | 1.63E-05 | 3.238936 |
| RPS16 | 0.175 | 1.128964 | 12.9381 | 4.854571 | 5.75E-06 | 1.63E-05 | 3.237977 |
| PRSS8 | -0.35417 | 0.78232 | 7.395316 | -4.85435 | 5.75E-06 | 1.63E-05 | 3.237127 |
| LIMD1 | 0.190237 | 1.140951 | 5.804684 | 4.851468 | 5.82E-06 | 1.65E-05 | 3.226246 |
| GPR21 | -0.18026 | 0.882542 | 6.14557 | -4.85071 | 5.83E-06 | 1.65E-05 | 3.223362 |
| FCER1A | 0.973689 | 1.963856 | 4.744557 | 4.848605 | 5.88E-06 | 1.67E-05 | 3.215423 |
| MINA | 0.207918 | 1.15502 | 6.497722 | 4.848054 | 5.90E-06 | 1.67E-05 | 3.213342 |
| HARS2 | 0.219921 | 1.16467 | 7.410127 | 4.84709 | 5.92E-06 | 1.68E-05 | 3.2097 |
| ITGB1BP1 | 0.235731 | 1.177503 | 6.194557 | 4.84698 | 5.92E-06 | 1.68E-05 | 3.209284 |
| PRRG2 | -0.22618 | 0.854896 | 5.932911 | -4.84693 | 5.92E-06 | 1.68E-05 | 3.209078 |
| FRMD1 | -0.19092 | 0.87605 | 5.691772 | -4.84322 | 6.01E-06 | 1.70E-05 | 3.195081 |
| NR1H3 | -0.24211 | 0.845509 | 6.765823 | -4.84117 | 6.06E-06 | 1.71E-05 | 3.187322 |
| FGF13 | 0.377213 | 1.298831 | 6.841266 | 4.840299 | 6.08E-06 | 1.72E-05 | 3.184051 |
| IQCA1 | 0.274875 | 1.209889 | 5.308734 | 4.838004 | 6.13E-06 | 1.73E-05 | 3.17539 |
| DLD | -0.24013 | 0.846672 | 9.028608 | -4.83743 | 6.14E-06 | 1.74E-05 | 3.173212 |
| NCS1 | -0.27046 | 0.829055 | 5.123544 | -4.83719 | 6.15E-06 | 1.74E-05 | 3.172308 |
| ZP2 | -0.28796 | 0.819057 | 5.059494 | -4.8354 | 6.19E-06 | 1.75E-05 | 3.165552 |
| PMPCA | -0.20906 | 0.865098 | 7.886582 | -4.8349 | 6.21E-06 | 1.75E-05 | 3.163675 |
| TMEM47 | 0.284895 | 1.218321 | 7.988354 | 4.831742 | 6.28E-06 | 1.77E-05 | 3.151762 |
| ADGRL2 | 0.242378 | 1.182941 | 6.901899 | 4.830535 | 6.31E-06 | 1.78E-05 | 3.14721 |
| UCHL1 | 0.598307 | 1.513939 | 8.691013 | 4.829452 | 6.34E-06 | 1.79E-05 | 3.143128 |
| TOR3A | 0.251594 | 1.190522 | 6.716835 | 4.828373 | 6.37E-06 | 1.80E-05 | 3.139057 |
| CD3G | 0.377233 | 1.298849 | 4.896709 | 4.827904 | 6.38E-06 | 1.80E-05 | 3.13729 |
| HSPB8 | 0.568366 | 1.482843 | 8.60481 | 4.826712 | 6.41E-06 | 1.81E-05 | 3.132795 |
| SULT2B1 | -0.23135 | 0.851837 | 6.035316 | -4.82628 | 6.42E-06 | 1.81E-05 | 3.131159 |
| RAB29 | -0.32549 | 0.798029 | 8.191646 | -4.8249 | 6.45E-06 | 1.82E-05 | 3.125958 |
| CRYZ | -0.20405 | 0.868109 | 11.44063 | -4.82459 | 6.46E-06 | 1.82E-05 | 3.124802 |
| SREK1 | 0.220758 | 1.165345 | 6.697215 | 4.823862 | 6.48E-06 | 1.82E-05 | 3.122053 |
| MTO1 | -0.24608 | 0.843184 | 7.771772 | -4.8237 | 6.48E-06 | 1.82E-05 | 3.121432 |
| PRDM14 | -0.23094 | 0.852078 | 5.701139 | -4.82336 | 6.49E-06 | 1.83E-05 | 3.120179 |

| | | | | | | | |
|---|---|---|---|---|---|---|---|
| OR7E12P | -0.36817 | 0.774765 | 5.983165 | -4.82331 | 6.49E-06 | 1.83E-05 | 3.119992 |
| NAA60 | -0.17111 | 0.888157 | 8.415696 | -4.81926 | 6.60E-06 | 1.85E-05 | 3.104702 |
| MRAS | 0.312352 | 1.24173 | 6.307215 | 4.819159 | 6.60E-06 | 1.85E-05 | 3.10434 |
| OVOL1 | -0.24967 | 0.841088 | 6.38962 | -4.81897 | 6.60E-06 | 1.85E-05 | 3.103608 |
| DAB2 | -0.2603 | 0.834913 | 10.63127 | -4.8176 | 6.64E-06 | 1.86E-05 | 3.098449 |
| GLDC | -0.38438 | 0.766108 | 9.611392 | -4.81748 | 6.64E-06 | 1.86E-05 | 3.098009 |
| UBFD1 | 0.313887 | 1.243052 | 6.143291 | 4.817407 | 6.64E-06 | 1.86E-05 | 3.097741 |
| TACC3 | 0.288155 | 1.221078 | 4.926456 | 4.817045 | 6.65E-06 | 1.87E-05 | 3.096379 |
| GTF2IRD1 | 0.226719 | 1.170171 | 8.022532 | 4.814854 | 6.71E-06 | 1.88E-05 | 3.088129 |
| IFT122 | 0.266304 | 1.202723 | 7.364937 | 4.814656 | 6.72E-06 | 1.88E-05 | 3.087384 |
| SALL2 | 0.225053 | 1.16882 | 7.276835 | 4.813847 | 6.74E-06 | 1.89E-05 | 3.08434 |
| GDF2 | -0.20383 | 0.86824 | 5.880506 | -4.81273 | 6.77E-06 | 1.90E-05 | 3.080121 |
| GSTO1 | -0.2574 | 0.836595 | 10.72139 | -4.81252 | 6.77E-06 | 1.90E-05 | 3.079333 |
| SNCG | -0.23731 | 0.848328 | 6.618481 | -4.81227 | 6.78E-06 | 1.90E-05 | 3.078409 |
| COLEC12 | 0.358808 | 1.282366 | 4.955316 | 4.810799 | 6.82E-06 | 1.91E-05 | 3.072868 |
| UTRN | 0.187332 | 1.138656 | 5.584557 | 4.810059 | 6.84E-06 | 1.91E-05 | 3.070085 |
| ACE2 | -0.61893 | 0.651155 | 9.193418 | -4.80958 | 6.85E-06 | 1.92E-05 | 3.06827 |
| TRMT11 | -0.22628 | 0.854837 | 6.545696 | -4.80875 | 6.87E-06 | 1.92E-05 | 3.065176 |
| ITPA | 0.193979 | 1.143914 | 8.331899 | 4.808166 | 6.89E-06 | 1.92E-05 | 3.062963 |
| TOM1L1 | -0.22783 | 0.853921 | 9.222658 | -4.80766 | 6.90E-06 | 1.93E-05 | 3.061053 |
| HLA-DOA | 0.212207 | 1.158459 | 5.838861 | 4.806944 | 6.92E-06 | 1.93E-05 | 3.058367 |
| CCDC176 | 0.256897 | 1.194906 | 6.824051 | 4.806543 | 6.93E-06 | 1.94E-05 | 3.056857 |
| RPL8 | 0.184592 | 1.136495 | 12.44646 | 4.806447 | 6.93E-06 | 1.94E-05 | 3.056497 |
| DUSP9 | -0.52734 | 0.693833 | 6.364937 | -4.79895 | 7.14E-06 | 1.99E-05 | 3.028328 |
| C1orf50 | -0.16308 | 0.893119 | 6.992532 | -4.79859 | 7.15E-06 | 1.99E-05 | 3.02695 |
| SIRPG | 0.252266 | 1.191077 | 4.191899 | 4.797697 | 7.17E-06 | 2.00E-05 | 3.023609 |
| CCND1 | -0.31326 | 0.804821 | 6.772405 | -4.79767 | 7.17E-06 | 2.00E-05 | 3.023515 |
| CPSF6 | 0.165204 | 1.121325 | 6.446835 | 4.795742 | 7.23E-06 | 2.01E-05 | 3.016263 |
| IL6R | -0.42225 | 0.746262 | 6.292532 | -4.79542 | 7.24E-06 | 2.02E-05 | 3.01507 |
| RBFA | -0.17976 | 0.882852 | 6.888608 | -4.79468 | 7.26E-06 | 2.02E-05 | 3.012285 |
| PTGES2 | -0.16807 | 0.890033 | 7.893924 | -4.79406 | 7.28E-06 | 2.02E-05 | 3.009948 |
| CD2AP | 0.238129 | 1.179462 | 7.541646 | 4.794053 | 7.28E-06 | 2.02E-05 | 3.00992 |
| LPIN1 | 0.214177 | 1.160042 | 6.344684 | 4.793022 | 7.30E-06 | 2.03E-05 | 3.006049 |
| CHPF2 | 0.178775 | 1.131922 | 6.547722 | 4.791901 | 7.34E-06 | 2.04E-05 | 3.001841 |
| ERLIN2 | -0.25265 | 0.83935 | 7.107722 | -4.78926 | 7.41E-06 | 2.06E-05 | 2.99192 |
| PIGT | 0.194592 | 1.1444 | 8.410633 | 4.788196 | 7.44E-06 | 2.07E-05 | 2.987932 |
| UPK2 | -0.16987 | 0.888924 | 4.488608 | -4.78593 | 7.51E-06 | 2.09E-05 | 2.979431 |
| SCN4A | -0.18559 | 0.879288 | 4.50443 | -4.78498 | 7.54E-06 | 2.09E-05 | 2.975858 |
| HERC1 | -0.25929 | 0.8355 | 6.487342 | -4.78333 | 7.59E-06 | 2.11E-05 | 2.969661 |
| FNDC8 | -0.20258 | 0.868998 | 4.77038 | -4.78256 | 7.61E-06 | 2.11E-05 | 2.966798 |
| SAP30L | -0.19843 | 0.871501 | 5.963418 | -4.78154 | 7.64E-06 | 2.12E-05 | 2.962979 |
| IDO1 | 0.537668 | 1.451624 | 5.573291 | 4.780696 | 7.66E-06 | 2.13E-05 | 2.959802 |
| RPAP1 | -0.16325 | 0.893013 | 6.172025 | -4.78049 | 7.67E-06 | 2.13E-05 | 2.959029 |
| TMPRSS2 | -0.38421 | 0.766199 | 7.141899 | -4.78009 | 7.68E-06 | 2.13E-05 | 2.957534 |
| AKIP1 | 0.190138 | 1.140873 | 6.047468 | 4.779763 | 7.69E-06 | 2.13E-05 | 2.956302 |
| TMEM259 | -0.19247 | 0.875106 | 4.950253 | -4.77831 | 7.73E-06 | 2.14E-05 | 2.950864 |
| SRSF7 | 0.285231 | 1.218605 | 7.600886 | 4.775392 | 7.82E-06 | 2.17E-05 | 2.93992 |

| | | | | | | | |
|---|---|---|---|---|---|---|---|
| CRYZL1 | -0.23822 | 0.84779 | 6.98962 | -4.77333 | 7.89E-06 | 2.18E-05 | 2.932187 |
| ERVMER34 | -0.30455 | 0.809694 | 5.234304 | -4.77291 | 7.90E-06 | 2.19E-05 | 2.93063 |
| RUFY1 | 0.17695 | 1.130491 | 8.188481 | 4.772155 | 7.92E-06 | 2.19E-05 | 2.927792 |
| UBXN7 | 0.193333 | 1.143402 | 5.920759 | 4.771992 | 7.93E-06 | 2.19E-05 | 2.927183 |
| SLC34A2 | 0.567062 | 1.481503 | 5.42557 | 4.771283 | 7.95E-06 | 2.20E-05 | 2.924525 |
| DSCR3 | -0.21849 | 0.859464 | 6.330253 | -4.77104 | 7.96E-06 | 2.20E-05 | 2.923611 |
| ZNF771 | -0.23907 | 0.847291 | 6.89557 | -4.76965 | 8.00E-06 | 2.21E-05 | 2.918394 |
| CSTF2T | 0.205744 | 1.153281 | 6.458987 | 4.766768 | 8.09E-06 | 2.23E-05 | 2.907622 |
| ATG4A | -0.18449 | 0.879962 | 7.399241 | -4.7653 | 8.13E-06 | 2.25E-05 | 2.902113 |
| ARHGAP33 | -0.17992 | 0.882751 | 5.856582 | -4.76375 | 8.18E-06 | 2.26E-05 | 2.896314 |
| ERH | 0.172365 | 1.126904 | 10.9057 | 4.763548 | 8.19E-06 | 2.26E-05 | 2.895569 |
| STK19 | 0.510995 | 1.425032 | 7.463671 | 4.762894 | 8.21E-06 | 2.26E-05 | 2.893123 |
| LIN7B | -0.24877 | 0.841611 | 6.083038 | -4.76122 | 8.26E-06 | 2.28E-05 | 2.886868 |
| RAB25 | 0.359308 | 1.282811 | 6.929873 | 4.760658 | 8.28E-06 | 2.28E-05 | 2.884759 |
| AGPAT2 | 0.196311 | 1.145765 | 6.904177 | 4.757734 | 8.38E-06 | 2.31E-05 | 2.87382 |
| CAPZA2 | 0.233004 | 1.17528 | 9.123418 | 4.757306 | 8.39E-06 | 2.31E-05 | 2.872222 |
| MCTS1 | 0.203491 | 1.151482 | 7.988481 | 4.755914 | 8.44E-06 | 2.32E-05 | 2.867016 |
| TRIM45 | 0.215007 | 1.160709 | 5.743291 | 4.754386 | 8.49E-06 | 2.34E-05 | 2.861304 |
| GINS2 | 0.263439 | 1.200336 | 4.825696 | 4.754335 | 8.49E-06 | 2.34E-05 | 2.861113 |
| TBC1D8B | -0.59829 | 0.660538 | 6.08443 | -4.75082 | 8.60E-06 | 2.37E-05 | 2.847988 |
| ACP5 | -0.54096 | 0.687313 | 9.436203 | -4.74963 | 8.64E-06 | 2.38E-05 | 2.84352 |
| NDUFS2 | -0.17635 | 0.884939 | 10.43329 | -4.74848 | 8.68E-06 | 2.39E-05 | 2.839244 |
| ARHGEF10 | 0.202134 | 1.150399 | 6.891392 | 4.748073 | 8.70E-06 | 2.39E-05 | 2.837717 |
| HSPA5 | 0.380217 | 1.301538 | 9.698608 | 4.747536 | 8.71E-06 | 2.39E-05 | 2.835709 |
| HIST1H2BL | -0.18285 | 0.880963 | 3.761013 | -4.74574 | 8.77E-06 | 2.41E-05 | 2.828996 |
| C12orf29 | -0.23059 | 0.852288 | 7.159114 | -4.74262 | 8.88E-06 | 2.44E-05 | 2.817349 |
| SLC16A3 | 0.388538 | 1.309066 | 5.770127 | 4.741677 | 8.91E-06 | 2.45E-05 | 2.813835 |
| CLCN1 | -0.19106 | 0.875962 | 5.09519 | -4.74129 | 8.93E-06 | 2.45E-05 | 2.812383 |
| RNPS1 | 0.230527 | 1.173263 | 8.274557 | 4.741241 | 8.93E-06 | 2.45E-05 | 2.812206 |
| CEP55 | 0.262978 | 1.199953 | 3.822025 | 4.73664 | 9.09E-06 | 2.49E-05 | 2.79504 |
| FTL | -0.20516 | 0.86744 | 13.48734 | -4.7358 | 9.12E-06 | 2.50E-05 | 2.7919 |
| COX5B | -0.20717 | 0.866232 | 11.27063 | -4.73572 | 9.12E-06 | 2.50E-05 | 2.791625 |
| NCL | 0.20975 | 1.156487 | 10.55544 | 4.73569 | 9.12E-06 | 2.50E-05 | 2.791499 |
| PDIA3 | 0.283656 | 1.217276 | 9.223924 | 4.73535 | 9.13E-06 | 2.50E-05 | 2.790232 |
| CHP1 | -0.26886 | 0.829975 | 8.92443 | -4.73434 | 9.17E-06 | 2.51E-05 | 2.786479 |
| COQ3 | -0.24442 | 0.844155 | 7.388987 | -4.73421 | 9.17E-06 | 2.51E-05 | 2.785965 |
| CLPX | -0.27889 | 0.824223 | 6.167848 | -4.73325 | 9.21E-06 | 2.52E-05 | 2.782386 |
| AGPAT4 | 0.21863 | 1.163628 | 4.967848 | 4.726285 | 9.46E-06 | 2.59E-05 | 2.75644 |
| ZNF264 | 0.245856 | 1.185796 | 6.449873 | 4.724926 | 9.51E-06 | 2.60E-05 | 2.751377 |
| C3orf14 | 0.23724 | 1.178735 | 6.576709 | 4.724837 | 9.51E-06 | 2.60E-05 | 2.751046 |
| PRRG1 | 0.354578 | 1.278612 | 6.830506 | 4.723973 | 9.54E-06 | 2.61E-05 | 2.747829 |
| RGR | -0.19313 | 0.874706 | 4.532152 | -4.72287 | 9.58E-06 | 2.62E-05 | 2.743718 |
| PAPPA | 0.351594 | 1.27597 | 5.868608 | 4.722773 | 9.59E-06 | 2.62E-05 | 2.743359 |
| GOLGA4 | -0.18951 | 0.876902 | 9.038228 | -4.72142 | 9.64E-06 | 2.63E-05 | 2.738335 |
| TMEM110 | -0.25227 | 0.839577 | 6.708101 | -4.72084 | 9.66E-06 | 2.64E-05 | 2.736148 |
| PARD6A | -0.2121 | 0.863279 | 6.536835 | -4.71873 | 9.74E-06 | 2.66E-05 | 2.728305 |
| LY6G6D | -0.17442 | 0.886124 | 4.568228 | -4.71844 | 9.75E-06 | 2.66E-05 | 2.72722 |

| | | | | | | | |
|---|---|---|---|---|---|---|---|
| CACNA1I | -0.17599 | 0.885161 | 5.009747 | -4.71743 | 9.79E-06 | 2.67E-05 | 2.723455 |
| OSGIN1 | -0.25208 | 0.839688 | 6.054051 | -4.71592 | 9.84E-06 | 2.68E-05 | 2.717846 |
| SECTM1 | 0.348762 | 1.273467 | 6.181772 | 4.715007 | 9.88E-06 | 2.69E-05 | 2.714452 |
| AAMDC | -0.19099 | 0.876002 | 6.86 | -4.71233 | 9.98E-06 | 2.72E-05 | 2.704479 |
| GADD45G | -0.47619 | 0.718872 | 5.341519 | -4.7097 | 1.01E-05 | 2.75E-05 | 2.69473 |
| SLC22A1 | -0.25118 | 0.840209 | 4.787468 | -4.70927 | 1.01E-05 | 2.75E-05 | 2.69311 |
| OGFRL1 | 0.237819 | 1.179209 | 5.233038 | 4.709182 | 1.01E-05 | 2.75E-05 | 2.69279 |
| PALLD | 0.376357 | 1.29806 | 8.498734 | 4.708353 | 1.01E-05 | 2.76E-05 | 2.689707 |
| SLC12A8 | 0.2883 | 1.221201 | 4.883038 | 4.707445 | 1.02E-05 | 2.77E-05 | 2.686332 |
| MMP9 | 0.518972 | 1.432934 | 5.134177 | 4.707081 | 1.02E-05 | 2.77E-05 | 2.68498 |
| MPPE1 | 0.198821 | 1.14776 | 8.495443 | 4.706664 | 1.02E-05 | 2.77E-05 | 2.683429 |
| LGSN | -0.28112 | 0.822952 | 3.851266 | -4.70616 | 1.02E-05 | 2.78E-05 | 2.68156 |
| UBAC1 | -0.18204 | 0.881454 | 7.405696 | -4.70599 | 1.02E-05 | 2.78E-05 | 2.680934 |
| HOMER3 | 0.306726 | 1.236898 | 6.93443 | 4.705026 | 1.03E-05 | 2.79E-05 | 2.677342 |
| LIM2 | -0.21168 | 0.863531 | 5.22962 | -4.70442 | 1.03E-05 | 2.79E-05 | 2.675108 |
| APBB3 | -0.21744 | 0.860088 | 6.578734 | -4.70426 | 1.03E-05 | 2.79E-05 | 2.674489 |
| SLC37A1 | 0.221403 | 1.165867 | 5.338354 | 4.703876 | 1.03E-05 | 2.80E-05 | 2.673069 |
| TEAD4 | 0.302484 | 1.233266 | 6.092658 | 4.703547 | 1.03E-05 | 2.80E-05 | 2.671846 |
| CNN2 | 0.257655 | 1.195534 | 7.419367 | 4.703475 | 1.03E-05 | 2.80E-05 | 2.671581 |
| GLRB | 0.208966 | 1.155859 | 4.116203 | 4.703276 | 1.03E-05 | 2.80E-05 | 2.67084 |
| CAMK2A | -0.23013 | 0.852561 | 5.862785 | -4.70291 | 1.04E-05 | 2.80E-05 | 2.669463 |
| SHC3 | -0.16663 | 0.890923 | 5.531266 | -4.70217 | 1.04E-05 | 2.81E-05 | 2.666716 |
| SASH1 | 0.257536 | 1.195435 | 7.276709 | 4.700784 | 1.04E-05 | 2.82E-05 | 2.661585 |
| HDGFRP3 | 0.226041 | 1.169621 | 6.158987 | 4.699817 | 1.05E-05 | 2.83E-05 | 2.657994 |
| NAMPT | 0.362622 | 1.28576 | 7.835823 | 4.699784 | 1.05E-05 | 2.83E-05 | 2.657869 |
| GRPR | -0.21914 | 0.859075 | 6.880633 | -4.69944 | 1.05E-05 | 2.84E-05 | 2.656608 |
| C1QBP | -0.21798 | 0.859766 | 9.333291 | -4.6987 | 1.05E-05 | 2.84E-05 | 2.653834 |
| PRRX1 | 0.323149 | 1.251058 | 4.497595 | 4.69832 | 1.05E-05 | 2.85E-05 | 2.652436 |
| PRSS3 | -0.43511 | 0.73964 | 6.827595 | -4.69799 | 1.05E-05 | 2.85E-05 | 2.65119 |
| GUK1 | -0.17864 | 0.883534 | 9.181899 | -4.69648 | 1.06E-05 | 2.87E-05 | 2.645594 |
| SPRYD7 | -0.21621 | 0.860823 | 4.914684 | -4.69596 | 1.06E-05 | 2.87E-05 | 2.643673 |
| ARMCX3 | 0.226318 | 1.169845 | 7.69519 | 4.695157 | 1.07E-05 | 2.88E-05 | 2.640693 |
| RETSAT | -0.3751 | 0.771053 | 9.401139 | -4.69499 | 1.07E-05 | 2.88E-05 | 2.64007 |
| CD180 | 0.228426 | 1.171556 | 4.809114 | 4.692427 | 1.08E-05 | 2.91E-05 | 2.63056 |
| FAU | 0.195494 | 1.145116 | 11.95253 | 4.689422 | 1.09E-05 | 2.94E-05 | 2.619414 |
| CD9 | 0.388814 | 1.309317 | 9.976329 | 4.688368 | 1.09E-05 | 2.95E-05 | 2.615505 |
| SLC16A7 | -0.35993 | 0.779204 | 5.812911 | -4.68787 | 1.10E-05 | 2.96E-05 | 2.613672 |
| RDH11 | -0.23431 | 0.850092 | 9.417342 | -4.68351 | 1.12E-05 | 3.00E-05 | 2.597502 |
| CFAP45 | -0.22108 | 0.857923 | 6.317215 | -4.68106 | 1.13E-05 | 3.03E-05 | 2.588402 |
| GAB2 | 0.187101 | 1.138474 | 6.087722 | 4.680279 | 1.13E-05 | 3.04E-05 | 2.585522 |
| IRAK1 | 0.257062 | 1.195042 | 9.166076 | 4.678663 | 1.14E-05 | 3.06E-05 | 2.579536 |
| EDRF1 | 0.183162 | 1.13537 | 5.266076 | 4.678561 | 1.14E-05 | 3.06E-05 | 2.579161 |
| CDKN3 | 0.275132 | 1.210105 | 4.729494 | 4.676444 | 1.15E-05 | 3.08E-05 | 2.571318 |
| FMO1 | -0.67854 | 0.624798 | 11.05873 | -4.67607 | 1.15E-05 | 3.09E-05 | 2.569947 |
| MMP1 | 0.338452 | 1.264399 | 3.758987 | 4.6759 | 1.15E-05 | 3.09E-05 | 2.569305 |
| TRAPPC2L | -0.17254 | 0.887277 | 7.945316 | -4.67495 | 1.15E-05 | 3.10E-05 | 2.565803 |
| ZC2HC1C | -0.21167 | 0.863535 | 5.878101 | -4.6706 | 1.17E-05 | 3.15E-05 | 2.549693 |

| | | | | | | | |
|---|---|---|---|---|---|---|---|
| TSPAN7 | -0.32804 | 0.79662 | 7.819494 | -4.66999 | 1.17E-05 | 3.15E-05 | 2.547432 |
| DLG5 | 0.274941 | 1.209944 | 6.623544 | 4.668258 | 1.18E-05 | 3.17E-05 | 2.541023 |
| TMEM246 | 0.29081 | 1.223327 | 5.624304 | 4.668206 | 1.18E-05 | 3.17E-05 | 2.540829 |
| MIA2 | -0.20831 | 0.865548 | 4.413418 | -4.66705 | 1.19E-05 | 3.19E-05 | 2.536548 |
| RIC8A | 0.15303 | 1.111903 | 8.143924 | 4.666976 | 1.19E-05 | 3.19E-05 | 2.536279 |
| IFNA10 | 0.26475 | 1.201428 | 4.253418 | 4.666233 | 1.19E-05 | 3.19E-05 | 2.53353 |
| NTRK2 | 0.237661 | 1.17908 | 4.919494 | 4.664358 | 1.20E-05 | 3.22E-05 | 2.526598 |
| SGK2 | -0.42696 | 0.743829 | 7.498608 | -4.6641 | 1.20E-05 | 3.22E-05 | 2.525654 |
| SCAF8 | 0.210613 | 1.157179 | 7.626456 | 4.662627 | 1.21E-05 | 3.24E-05 | 2.5202 |
| ZFC3H1 | 0.223709 | 1.167732 | 6.896709 | 4.661674 | 1.21E-05 | 3.25E-05 | 2.516677 |
| BAIAP3 | -0.18655 | 0.878706 | 6.574684 | -4.66145 | 1.21E-05 | 3.25E-05 | 2.515829 |
| NOX5 | -0.17119 | 0.888108 | 5.793924 | -4.66049 | 1.22E-05 | 3.26E-05 | 2.512316 |
| GTF3C1 | -0.18922 | 0.877082 | 7.489873 | -4.66019 | 1.22E-05 | 3.26E-05 | 2.511203 |
| DNM1L | 0.199717 | 1.148473 | 7.038861 | 4.659372 | 1.22E-05 | 3.27E-05 | 2.508169 |
| CBR3 | 0.278972 | 1.21333 | 5.393924 | 4.65864 | 1.23E-05 | 3.28E-05 | 2.505463 |
| GMFB | 0.23747 | 1.178924 | 8.083544 | 4.658214 | 1.23E-05 | 3.28E-05 | 2.503888 |
| RASGRP1 | 0.49226 | 1.406646 | 6.313671 | 4.657467 | 1.23E-05 | 3.29E-05 | 2.50113 |
| KLHL20 | -0.22159 | 0.857617 | 5.925696 | -4.65746 | 1.23E-05 | 3.29E-05 | 2.501116 |
| CGA | -0.49035 | 0.711853 | 5.495823 | -4.65691 | 1.23E-05 | 3.30E-05 | 2.499063 |
| ATP2C2 | -0.33868 | 0.790763 | 5.094177 | -4.65639 | 1.24E-05 | 3.30E-05 | 2.497136 |
| GCNT2 | -0.22293 | 0.856823 | 5.344051 | -4.65491 | 1.24E-05 | 3.32E-05 | 2.491684 |
| SLC9A3 | -0.18628 | 0.878866 | 4.473924 | -4.65188 | 1.26E-05 | 3.36E-05 | 2.480507 |
| AKAP3 | -0.20828 | 0.865568 | 5.245823 | -4.65183 | 1.26E-05 | 3.36E-05 | 2.480315 |
| UBP1 | 0.26359 | 1.200462 | 7.000759 | 4.650355 | 1.27E-05 | 3.38E-05 | 2.474865 |
| MAP6D1 | -0.19353 | 0.874463 | 5.59481 | -4.64957 | 1.27E-05 | 3.39E-05 | 2.471966 |
| C6orf15 | -0.28497 | 0.82076 | 6.468354 | -4.64876 | 1.27E-05 | 3.39E-05 | 2.468986 |
| C8B | -0.21575 | 0.861098 | 5.528354 | -4.64866 | 1.27E-05 | 3.39E-05 | 2.468598 |
| RNASEL | 0.225389 | 1.169092 | 6.479367 | 4.648084 | 1.28E-05 | 3.40E-05 | 2.466485 |
| DDX1 | -0.15304 | 0.899355 | 9.677595 | -4.64741 | 1.28E-05 | 3.41E-05 | 2.463995 |
| TRAT1 | 0.284638 | 1.218104 | 3.697595 | 4.647363 | 1.28E-05 | 3.41E-05 | 2.463825 |
| CBR4 | -0.26808 | 0.830426 | 8.033671 | -4.6469 | 1.28E-05 | 3.41E-05 | 2.462099 |
| TTLL1 | 0.226891 | 1.17031 | 5.943038 | 4.645699 | 1.29E-05 | 3.43E-05 | 2.457684 |
| PPP1R15A | -0.33381 | 0.79344 | 5.970127 | -4.64564 | 1.29E-05 | 3.43E-05 | 2.457457 |
| ZNF184 | -0.28499 | 0.820745 | 5.88443 | -4.64525 | 1.29E-05 | 3.43E-05 | 2.456018 |
| CDKN1B | 0.336548 | 1.262732 | 8.087975 | 4.644989 | 1.29E-05 | 3.43E-05 | 2.455068 |
| PTGES | 0.226772 | 1.170214 | 5.58038 | 4.644984 | 1.29E-05 | 3.43E-05 | 2.455048 |
| HBQ1 | -0.21661 | 0.860583 | 5.127342 | -4.64489 | 1.29E-05 | 3.43E-05 | 2.454687 |
| STAM | 0.261258 | 1.198524 | 7.228481 | 4.643294 | 1.30E-05 | 3.45E-05 | 2.448816 |
| ZNHIT2 | -0.16852 | 0.889756 | 5.697215 | -4.64131 | 1.31E-05 | 3.48E-05 | 2.441514 |
| DSG2 | 0.337115 | 1.263228 | 8.277342 | 4.639441 | 1.32E-05 | 3.50E-05 | 2.434608 |
| ZNF112 | 0.242721 | 1.183222 | 4.862911 | 4.637679 | 1.33E-05 | 3.52E-05 | 2.428113 |
| UQCR11 | -0.15918 | 0.895536 | 9.913291 | -4.63754 | 1.33E-05 | 3.53E-05 | 2.427621 |
| PSENEN | 0.245283 | 1.185325 | 7.932025 | 4.63748 | 1.33E-05 | 3.53E-05 | 2.427381 |
| HYAL2 | 0.212055 | 1.158337 | 7.133797 | 4.63594 | 1.34E-05 | 3.55E-05 | 2.421707 |
| MCM6 | 0.29143 | 1.223852 | 5.901519 | 4.63397 | 1.35E-05 | 3.57E-05 | 2.41445 |
| LAMB1 | 0.307787 | 1.237807 | 7.819873 | 4.63383 | 1.35E-05 | 3.57E-05 | 2.413934 |
| SLC4A1 | -0.51524 | 0.699675 | 8.326076 | -4.63145 | 1.36E-05 | 3.60E-05 | 2.405171 |

| | | | | | | | |
|---|---|---|---|---|---|---|---|
| H1FX | 0.308959 | 1.238814 | 6.179494 | 4.627251 | 1.38E-05 | 3.66E-05 | 2.389711 |
| TBC1D9 | 0.30643 | 1.236643 | 7.502785 | 4.626943 | 1.38E-05 | 3.66E-05 | 2.388577 |
| EHD1 | -0.18534 | 0.87944 | 7.036709 | -4.62615 | 1.39E-05 | 3.67E-05 | 2.385676 |
| THAP1 | 0.247569 | 1.187205 | 5.894937 | 4.624616 | 1.40E-05 | 3.69E-05 | 2.380014 |
| SND1 | 0.197899 | 1.147026 | 8.388101 | 4.62186 | 1.41E-05 | 3.73E-05 | 2.369876 |
| STEAP4 | 0.324137 | 1.251915 | 4.019747 | 4.620743 | 1.42E-05 | 3.75E-05 | 2.365767 |
| BIN2 | 0.364592 | 1.287517 | 5.391646 | 4.620197 | 1.42E-05 | 3.75E-05 | 2.363758 |
| GPR171 | 0.411542 | 1.330106 | 4.345823 | 4.616183 | 1.44E-05 | 3.81E-05 | 2.349002 |
| ACE | -0.27329 | 0.827432 | 4.95519 | -4.61581 | 1.44E-05 | 3.81E-05 | 2.347635 |
| SDF2L1 | 0.329506 | 1.256583 | 7.871772 | 4.615365 | 1.45E-05 | 3.82E-05 | 2.345994 |
| ARHGAP17 | 0.194783 | 1.144552 | 7.286582 | 4.615076 | 1.45E-05 | 3.82E-05 | 2.344933 |
| RPF1 | 0.174453 | 1.128537 | 9.024177 | 4.614038 | 1.45E-05 | 3.84E-05 | 2.341118 |
| PLEKHG6 | -0.20326 | 0.868585 | 4.698354 | -4.61377 | 1.46E-05 | 3.84E-05 | 2.340146 |
| UNC13B | -0.20225 | 0.869192 | 8.475949 | -4.6085 | 1.49E-05 | 3.92E-05 | 2.320792 |
| BTN3A1 | 0.219776 | 1.164553 | 5.973544 | 4.608388 | 1.49E-05 | 3.92E-05 | 2.320365 |
| TTF2 | 0.210072 | 1.156746 | 4.041013 | 4.607739 | 1.49E-05 | 3.92E-05 | 2.317982 |
| TACC2 | -0.33339 | 0.793668 | 7.03443 | -4.60737 | 1.49E-05 | 3.93E-05 | 2.316629 |
| HMGB3 | 0.245639 | 1.185618 | 7.02 | 4.605503 | 1.50E-05 | 3.96E-05 | 2.309776 |
| NXF1 | 0.204526 | 1.152307 | 7.41 | 4.604823 | 1.51E-05 | 3.96E-05 | 2.307278 |
| STXBP1 | 0.24054 | 1.181435 | 7.235696 | 4.604548 | 1.51E-05 | 3.97E-05 | 2.30627 |
| CLPS | -0.19233 | 0.875194 | 4.596835 | -4.60222 | 1.52E-05 | 4.00E-05 | 2.297717 |
| P4HTM | -0.20852 | 0.865426 | 8.070506 | -4.60109 | 1.53E-05 | 4.02E-05 | 2.293579 |
| GPSM2 | 0.199335 | 1.148169 | 5.066962 | 4.600405 | 1.53E-05 | 4.03E-05 | 2.29107 |
| SLC2A8 | -0.17876 | 0.883461 | 6.219241 | -4.60022 | 1.53E-05 | 4.03E-05 | 2.290375 |
| NGF | -0.21551 | 0.861239 | 5.133671 | -4.59967 | 1.54E-05 | 4.03E-05 | 2.288367 |
| SEC24C | 0.193636 | 1.143643 | 7.480886 | 4.599654 | 1.54E-05 | 4.03E-05 | 2.288314 |
| SC5D | -0.16812 | 0.89 | 6.266076 | -4.59613 | 1.56E-05 | 4.09E-05 | 2.275399 |
| SULT1B1 | -0.21848 | 0.859472 | 5.447215 | -4.596 | 1.56E-05 | 4.09E-05 | 2.274929 |
| THOC6 | 0.230145 | 1.172953 | 5.902658 | 4.595267 | 1.56E-05 | 4.10E-05 | 2.272228 |
| MCF2L | -0.17781 | 0.884046 | 6.898987 | -4.59465 | 1.57E-05 | 4.11E-05 | 2.269978 |
| HLA-DOB | 0.271113 | 1.206739 | 5.716076 | 4.594027 | 1.57E-05 | 4.11E-05 | 2.267684 |
| GNB3 | -0.21716 | 0.860257 | 6.273418 | -4.59398 | 1.57E-05 | 4.11E-05 | 2.267515 |
| CNPY3 | 0.227141 | 1.170513 | 6.155316 | 4.593789 | 1.57E-05 | 4.12E-05 | 2.266812 |
| STK32B | -0.31993 | 0.80111 | 7.37962 | -4.59323 | 1.57E-05 | 4.12E-05 | 2.26478 |
| ZBTB48 | -0.17949 | 0.883013 | 5.627848 | -4.59269 | 1.58E-05 | 4.13E-05 | 2.262771 |
| RTCA | -0.31182 | 0.805626 | 8.989747 | -4.59147 | 1.59E-05 | 4.15E-05 | 2.258322 |
| MYL7 | -0.21529 | 0.861373 | 5.782025 | -4.58454 | 1.63E-05 | 4.26E-05 | 2.23295 |
| HPSE | 0.329328 | 1.256428 | 5.152785 | 4.58319 | 1.64E-05 | 4.28E-05 | 2.227991 |
| ARR3 | -0.26377 | 0.832906 | 6.181772 | -4.58297 | 1.64E-05 | 4.28E-05 | 2.227192 |
| NFATC4 | -0.16328 | 0.892992 | 5.74962 | -4.58254 | 1.64E-05 | 4.29E-05 | 2.225603 |
| SLC22A18 | -0.60528 | 0.657345 | 9.976076 | -4.58236 | 1.64E-05 | 4.29E-05 | 2.224945 |
| GGCX | -0.15918 | 0.895532 | 6.81481 | -4.58208 | 1.64E-05 | 4.29E-05 | 2.223927 |
| NFE2L2 | 0.254717 | 1.193101 | 9.296835 | 4.581095 | 1.65E-05 | 4.31E-05 | 2.220328 |
| FIS1 | -0.16455 | 0.89221 | 9.881266 | -4.58007 | 1.66E-05 | 4.32E-05 | 2.216587 |
| PRKCI | 0.256225 | 1.19435 | 7.968987 | 4.579682 | 1.66E-05 | 4.33E-05 | 2.215158 |
| TAP2 | 0.169137 | 1.124386 | 5.21 | 4.578168 | 1.67E-05 | 4.35E-05 | 2.209618 |
| POLR2H | -0.15664 | 0.897112 | 8.358481 | -4.57802 | 1.67E-05 | 4.35E-05 | 2.209081 |

| | | | | | | | |
|---|---|---|---|---|---|---|---|
| HNRNPA3 | 0.189144 | 1.140087 | 7.816835 | 4.57726 | 1.67E-05 | 4.36E-05 | 2.206296 |
| SIRT4 | -0.18099 | 0.882099 | 4.762658 | -4.57713 | 1.67E-05 | 4.36E-05 | 2.205823 |
| SPCS3 | 0.258702 | 1.196402 | 7.807848 | 4.576152 | 1.68E-05 | 4.38E-05 | 2.202245 |
| CCDC91 | 0.23444 | 1.17645 | 6.532278 | 4.574355 | 1.69E-05 | 4.41E-05 | 2.195677 |
| TOPBP1 | 0.20807 | 1.155142 | 5.902785 | 4.572156 | 1.71E-05 | 4.44E-05 | 2.187637 |
| BUD31 | 0.225422 | 1.169119 | 8.081772 | 4.570195 | 1.72E-05 | 4.47E-05 | 2.180469 |
| CCNH | -0.17738 | 0.884305 | 8.461772 | -4.56903 | 1.73E-05 | 4.49E-05 | 2.176209 |
| TERT | -0.18764 | 0.878044 | 5.055316 | -4.56825 | 1.73E-05 | 4.51E-05 | 2.173365 |
| IL15 | 0.169842 | 1.124935 | 4.307468 | 4.567463 | 1.74E-05 | 4.52E-05 | 2.170489 |
| UGT2A3 | -0.43534 | 0.739522 | 7.690759 | -4.56715 | 1.74E-05 | 4.52E-05 | 2.169345 |
| ZNF165 | -0.47841 | 0.717767 | 4.683418 | -4.56634 | 1.74E-05 | 4.53E-05 | 2.166395 |
| CRTAP | -0.19733 | 0.872166 | 7.539747 | -4.56538 | 1.75E-05 | 4.55E-05 | 2.162885 |
| PPID | 0.263366 | 1.200276 | 7.812405 | 4.563096 | 1.77E-05 | 4.59E-05 | 2.154544 |
| KLHDC4 | -0.19252 | 0.875078 | 6.000886 | -4.56276 | 1.77E-05 | 4.59E-05 | 2.153303 |
| KIN | 0.207516 | 1.154699 | 4.71038 | 4.56245 | 1.77E-05 | 4.60E-05 | 2.152187 |
| PLA2R1 | -0.32439 | 0.798637 | 4.207975 | -4.56197 | 1.77E-05 | 4.60E-05 | 2.150432 |
| PARG | -0.20939 | 0.864904 | 7.651013 | -4.561 | 1.78E-05 | 4.62E-05 | 2.146875 |
| HAO1 | -0.31497 | 0.803869 | 4.475823 | -4.56083 | 1.78E-05 | 4.62E-05 | 2.146262 |
| ABCD4 | -0.1658 | 0.891432 | 6.101392 | -4.56037 | 1.78E-05 | 4.63E-05 | 2.144604 |
| PLAGL2 | -0.17014 | 0.888757 | 5.650886 | -4.56025 | 1.78E-05 | 4.63E-05 | 2.144169 |
| CDKN2C | -0.18421 | 0.880131 | 6.355443 | -4.55997 | 1.79E-05 | 4.63E-05 | 2.143145 |
| IGHV4-34 | 0.782721 | 1.720372 | 5.038481 | 4.558776 | 1.79E-05 | 4.65E-05 | 2.138778 |
| CEBPE | -0.19257 | 0.875046 | 4.843038 | -4.55856 | 1.80E-05 | 4.65E-05 | 2.138007 |
| GLT8D2 | 0.348781 | 1.273484 | 5.487215 | 4.555989 | 1.81E-05 | 4.70E-05 | 2.128611 |
| GJA1 | 0.694829 | 1.618692 | 7.28481 | 4.554696 | 1.82E-05 | 4.72E-05 | 2.123894 |
| C2orf54 | -0.47023 | 0.721849 | 5.586835 | -4.55456 | 1.82E-05 | 4.72E-05 | 2.123393 |
| IQCC | -0.2065 | 0.866636 | 5.255696 | -4.5529 | 1.84E-05 | 4.75E-05 | 2.117341 |
| DSTN | 0.148524 | 1.108435 | 11.51291 | 4.55285 | 1.84E-05 | 4.75E-05 | 2.117165 |
| SMUG1 | -0.22825 | 0.853667 | 6.371392 | -4.55261 | 1.84E-05 | 4.75E-05 | 2.116275 |
| TUBB2A | -0.40266 | 0.756462 | 9.716582 | -4.55131 | 1.85E-05 | 4.77E-05 | 2.111553 |
| SLC33A1 | -0.23981 | 0.846857 | 6.925696 | -4.55073 | 1.85E-05 | 4.78E-05 | 2.109427 |
| CBX3 | 0.273485 | 1.208724 | 7.869241 | 4.550495 | 1.85E-05 | 4.78E-05 | 2.10858 |
| EIF3D | 0.212747 | 1.158893 | 9.424304 | 4.550266 | 1.85E-05 | 4.79E-05 | 2.107744 |
| RGS7 | -0.57964 | 0.669129 | 4.925696 | -4.54965 | 1.86E-05 | 4.80E-05 | 2.105494 |
| USP14 | 0.144921 | 1.10567 | 7.993797 | 4.549332 | 1.86E-05 | 4.80E-05 | 2.104342 |
| RHOH | 0.337721 | 1.263758 | 5.917595 | 4.54887 | 1.86E-05 | 4.81E-05 | 2.102659 |
| WNK1 | -0.24159 | 0.84581 | 8.377342 | -4.54802 | 1.87E-05 | 4.82E-05 | 2.099545 |
| HAUS7 | -0.49825 | 0.707966 | 6.307089 | -4.54791 | 1.87E-05 | 4.82E-05 | 2.099168 |
| TFF3 | 0.376957 | 1.298599 | 5.710506 | 4.546778 | 1.88E-05 | 4.84E-05 | 2.095035 |
| PDK3 | -0.16864 | 0.889683 | 6.324557 | -4.5461 | 1.88E-05 | 4.85E-05 | 2.092556 |
| LRRC3 | -0.14289 | 0.905706 | 4.196835 | -4.54354 | 1.90E-05 | 4.90E-05 | 2.083247 |
| ZNF84 | 0.223379 | 1.167465 | 5.532658 | 4.543518 | 1.90E-05 | 4.90E-05 | 2.083162 |
| GPS2 | -0.23366 | 0.850473 | 7.448481 | -4.54074 | 1.92E-05 | 4.95E-05 | 2.073037 |
| RWDD2B | -0.16514 | 0.891839 | 7.979494 | -4.53937 | 1.93E-05 | 4.97E-05 | 2.068074 |
| PTPRT | -0.22875 | 0.853375 | 5.425316 | -4.53844 | 1.94E-05 | 4.99E-05 | 2.064681 |
| HNRNPF | 0.195349 | 1.145001 | 8.195949 | 4.538181 | 1.94E-05 | 4.99E-05 | 2.063737 |
| PAPD7 | 0.204216 | 1.15206 | 7.211392 | 4.53489 | 1.96E-05 | 5.05E-05 | 2.051763 |

| | | | | | | | |
|---|---|---|---|---|---|---|---|
| MYO1B | 0.268999 | 1.204971 | 7.703671 | 4.534383 | 1.97E-05 | 5.06E-05 | 2.049919 |
| GPR3 | -0.21173 | 0.8635 | 5.141772 | -4.53382 | 1.97E-05 | 5.07E-05 | 2.047875 |
| CYBRD1 | 0.370191 | 1.292524 | 5.710506 | 4.52959 | 2.00E-05 | 5.15E-05 | 2.032494 |
| TJP1 | 0.240026 | 1.181014 | 7.994177 | 4.526058 | 2.03E-05 | 5.22E-05 | 2.019658 |
| CCT6A | 0.232121 | 1.174561 | 7.926962 | 4.524114 | 2.05E-05 | 5.25E-05 | 2.012597 |
| CARKD | -0.19821 | 0.871628 | 8.93481 | -4.52406 | 2.05E-05 | 5.25E-05 | 2.012387 |
| LCN1 | -0.22655 | 0.854677 | 4.587975 | -4.52385 | 2.05E-05 | 5.25E-05 | 2.011653 |
| MGRN1 | 0.17774 | 1.131111 | 7.796203 | 4.523318 | 2.05E-05 | 5.26E-05 | 2.009707 |
| SERINC3 | 0.207872 | 1.154983 | 8.628354 | 4.523107 | 2.05E-05 | 5.27E-05 | 2.008939 |
| NOP10 | 0.163221 | 1.119785 | 10.32405 | 4.522886 | 2.06E-05 | 5.27E-05 | 2.008138 |
| CD164 | -0.26377 | 0.832906 | 10.11177 | -4.52179 | 2.06E-05 | 5.29E-05 | 2.004159 |
| CKM | -0.18268 | 0.881064 | 4.633038 | -4.51976 | 2.08E-05 | 5.33E-05 | 1.996788 |
| ZNF266 | 0.308814 | 1.238689 | 6.892911 | 4.519397 | 2.08E-05 | 5.33E-05 | 1.995469 |
| METTL4 | 0.173636 | 1.127898 | 5.162532 | 4.519334 | 2.08E-05 | 5.33E-05 | 1.995241 |
| WDR61 | -0.17637 | 0.884927 | 8.867848 | -4.51643 | 2.11E-05 | 5.39E-05 | 1.984713 |
| LHB | -0.22003 | 0.85855 | 5.644177 | -4.51607 | 2.11E-05 | 5.40E-05 | 1.983394 |
| RPL28 | 0.167207 | 1.122882 | 7.855063 | 4.515296 | 2.12E-05 | 5.41E-05 | 1.980587 |
| TSC22D4 | -0.20949 | 0.864841 | 5.845316 | -4.51528 | 2.12E-05 | 5.41E-05 | 1.980523 |
| VRK1 | 0.210863 | 1.15738 | 5.528734 | 4.514947 | 2.12E-05 | 5.41E-05 | 1.979323 |
| FAM76A | -0.18322 | 0.880734 | 4.847595 | -4.51374 | 2.13E-05 | 5.44E-05 | 1.974931 |
| ME3 | -0.27412 | 0.826956 | 6.526582 | -4.51165 | 2.14E-05 | 5.48E-05 | 1.967375 |
| ZMAT5 | -0.16601 | 0.891302 | 6.46 | -4.51155 | 2.15E-05 | 5.48E-05 | 1.967005 |
| FAM98A | 0.260968 | 1.198283 | 8.535316 | 4.511125 | 2.15E-05 | 5.49E-05 | 1.96546 |
| S100A2 | 0.844018 | 1.795043 | 9.474304 | 4.511013 | 2.15E-05 | 5.49E-05 | 1.965052 |
| PSMB5 | -0.19947 | 0.870873 | 9.723418 | -4.50567 | 2.19E-05 | 5.60E-05 | 1.945673 |
| DAPK2 | -0.15359 | 0.89901 | 4.99519 | -4.50549 | 2.19E-05 | 5.60E-05 | 1.945041 |
| SP3 | 0.291324 | 1.223763 | 7.415823 | 4.505022 | 2.20E-05 | 5.61E-05 | 1.943339 |
| RARG | -0.18278 | 0.881004 | 7.015823 | -4.5027 | 2.22E-05 | 5.65E-05 | 1.934942 |
| TOPORS | -0.19233 | 0.875194 | 5.806835 | -4.4995 | 2.24E-05 | 5.72E-05 | 1.923335 |
| LRRC8D | -0.21871 | 0.859334 | 8.70038 | -4.49906 | 2.25E-05 | 5.73E-05 | 1.921757 |
| NDST2 | 0.189177 | 1.140113 | 6.969241 | 4.498981 | 2.25E-05 | 5.73E-05 | 1.921461 |
| GSR | -0.27526 | 0.826303 | 7.819367 | -4.49867 | 2.25E-05 | 5.73E-05 | 1.920354 |
| USP34 | 0.184723 | 1.136599 | 7.036076 | 4.495186 | 2.28E-05 | 5.81E-05 | 1.907729 |
| LAT | 0.226667 | 1.170128 | 5.244684 | 4.494752 | 2.29E-05 | 5.82E-05 | 1.906159 |
| MMADHC | -0.18136 | 0.881873 | 9.877722 | -4.49177 | 2.31E-05 | 5.88E-05 | 1.895374 |
| LARP7 | 0.241561 | 1.182271 | 7.550253 | 4.488747 | 2.34E-05 | 5.94E-05 | 1.88444 |
| MFHAS1 | 0.296515 | 1.228174 | 6.898861 | 4.488233 | 2.34E-05 | 5.95E-05 | 1.882584 |
| SP140 | 0.238755 | 1.179974 | 4.187342 | 4.487038 | 2.35E-05 | 5.98E-05 | 1.878263 |
| AMBN | -0.18559 | 0.879292 | 4.482911 | -4.48568 | 2.36E-05 | 6.01E-05 | 1.873361 |
| ZFP30 | 0.200501 | 1.149097 | 6.028101 | 4.484084 | 2.38E-05 | 6.04E-05 | 1.867588 |
| ERBB3 | -0.31511 | 0.803792 | 8.047722 | -4.48188 | 2.40E-05 | 6.09E-05 | 1.859629 |
| HOXB7 | 0.275336 | 1.210276 | 7.732405 | 4.481715 | 2.40E-05 | 6.09E-05 | 1.859031 |
| CLCNKB | -0.41101 | 0.752094 | 8.132658 | -4.48026 | 2.41E-05 | 6.12E-05 | 1.853783 |
| SLC35B1 | -0.19236 | 0.875174 | 8.42443 | -4.47991 | 2.42E-05 | 6.13E-05 | 1.852531 |
| RAB8A | 0.211166 | 1.157623 | 9.058861 | 4.479912 | 2.42E-05 | 6.13E-05 | 1.852521 |
| C1orf95 | -0.21019 | 0.864423 | 5.556329 | -4.47984 | 2.42E-05 | 6.13E-05 | 1.852256 |
| BANK1 | 0.262009 | 1.199148 | 4.925316 | 4.478801 | 2.43E-05 | 6.15E-05 | 1.84851 |

| | | | | | | | |
|---|---|---|---|---|---|---|---|
| EIF6 | -0.16797 | 0.890094 | 9.601139 | -4.4785 | 2.43E-05 | 6.16E-05 | 1.847427 |
| PLEKHF1 | 0.230896 | 1.173564 | 6.279494 | 4.478286 | 2.43E-05 | 6.16E-05 | 1.846651 |
| MFN1 | 0.205777 | 1.153308 | 6.091392 | 4.477313 | 2.44E-05 | 6.18E-05 | 1.843139 |
| PLS1 | 0.244506 | 1.184687 | 8.771266 | 4.475545 | 2.46E-05 | 6.22E-05 | 1.836755 |
| PROP1 | -0.2208 | 0.858091 | 5.261899 | -4.47531 | 2.46E-05 | 6.22E-05 | 1.83591 |
| HHLA3 | -0.26848 | 0.830191 | 5.247848 | -4.47442 | 2.47E-05 | 6.24E-05 | 1.832704 |
| GTF3C4 | 0.194664 | 1.144458 | 6.533924 | 4.471948 | 2.49E-05 | 6.30E-05 | 1.823778 |
| KPNB1 | 0.20722 | 1.154461 | 8.738734 | 4.471812 | 2.49E-05 | 6.30E-05 | 1.823287 |
| TOR1B | 0.182536 | 1.134877 | 6.91038 | 4.47169 | 2.49E-05 | 6.30E-05 | 1.822848 |
| DPEP2 | 0.267892 | 1.204047 | 4.428861 | 4.470994 | 2.50E-05 | 6.31E-05 | 1.820338 |
| ATP5G2 | -0.18524 | 0.879504 | 10.25241 | -4.47098 | 2.50E-05 | 6.31E-05 | 1.820279 |
| WWC1 | 0.259677 | 1.197211 | 7.893038 | 4.470622 | 2.50E-05 | 6.32E-05 | 1.818996 |
| CEP192 | 0.247464 | 1.187118 | 5.429241 | 4.469925 | 2.51E-05 | 6.33E-05 | 1.816485 |
| HOXB2 | 0.308551 | 1.238463 | 8.783671 | 4.46852 | 2.52E-05 | 6.37E-05 | 1.811417 |
| PPWD1 | 0.264361 | 1.201104 | 5.643038 | 4.468347 | 2.52E-05 | 6.37E-05 | 1.810792 |
| USP5 | -0.21599 | 0.860952 | 6.184557 | -4.46814 | 2.53E-05 | 6.37E-05 | 1.810063 |
| CLCC1 | 0.162319 | 1.119084 | 5.032152 | 4.467164 | 2.53E-05 | 6.39E-05 | 1.806528 |
| C7orf43 | -0.18376 | 0.880405 | 6.192152 | -4.46497 | 2.56E-05 | 6.44E-05 | 1.798607 |
| PTDSS2 | -0.20014 | 0.870467 | 6.848354 | -4.46296 | 2.58E-05 | 6.49E-05 | 1.791363 |
| KCNA6 | -0.21478 | 0.86168 | 5.253544 | -4.46282 | 2.58E-05 | 6.49E-05 | 1.790872 |
| MORF4L2 | 0.172233 | 1.126801 | 11.47608 | 4.461282 | 2.59E-05 | 6.53E-05 | 1.785335 |
| RHCE | -0.22662 | 0.854634 | 4.864684 | -4.4604 | 2.60E-05 | 6.55E-05 | 1.782147 |
| C8orf33 | 0.283986 | 1.217554 | 5.977975 | 4.460027 | 2.60E-05 | 6.55E-05 | 1.780817 |
| TPP2 | 0.17749 | 1.130915 | 6.863924 | 4.458979 | 2.61E-05 | 6.58E-05 | 1.777042 |
| UBL3 | -0.22711 | 0.854342 | 8.528608 | -4.45772 | 2.63E-05 | 6.61E-05 | 1.772506 |
| HABP2 | 0.787233 | 1.725762 | 7.217975 | 4.455133 | 2.65E-05 | 6.67E-05 | 1.763199 |
| PUS7L | 0.17195 | 1.12658 | 5.651392 | 4.454388 | 2.66E-05 | 6.69E-05 | 1.760515 |
| ETV7 | -0.21044 | 0.864273 | 5.654051 | -4.45149 | 2.69E-05 | 6.76E-05 | 1.7501 |
| ECM2 | -0.4537 | 0.730167 | 7.505696 | -4.451 | 2.69E-05 | 6.77E-05 | 1.748319 |
| APOA4 | -0.20182 | 0.86945 | 5.347215 | -4.44981 | 2.70E-05 | 6.80E-05 | 1.744056 |
| EIF2B2 | -0.19419 | 0.874064 | 8.096709 | -4.44785 | 2.72E-05 | 6.84E-05 | 1.736988 |
| SLC22A17 | -0.23046 | 0.852362 | 7.120253 | -4.44707 | 2.73E-05 | 6.86E-05 | 1.73419 |
| POLL | -0.16739 | 0.890451 | 6.007468 | -4.44706 | 2.73E-05 | 6.86E-05 | 1.734147 |
| OSTF1 | 0.327187 | 1.254565 | 8.156456 | 4.446261 | 2.74E-05 | 6.88E-05 | 1.731289 |
| GNG7 | -0.17937 | 0.883086 | 6.380506 | -4.44551 | 2.75E-05 | 6.90E-05 | 1.728592 |
| FANCF | 0.210507 | 1.157095 | 6.410759 | 4.443278 | 2.77E-05 | 6.95E-05 | 1.720567 |
| SPSB1 | -0.35289 | 0.783013 | 5.360633 | -4.44265 | 2.78E-05 | 6.97E-05 | 1.718324 |
| PRMT1 | 0.158491 | 1.116119 | 7.334684 | 4.439083 | 2.82E-05 | 7.06E-05 | 1.705501 |
| ARL15 | -0.22487 | 0.855669 | 4.892152 | -4.43854 | 2.82E-05 | 7.07E-05 | 1.703547 |
| FPR1 | 0.544209 | 1.458221 | 5.964937 | 4.437602 | 2.83E-05 | 7.09E-05 | 1.700181 |
| TPX2 | 0.288307 | 1.221206 | 4.621519 | 4.437596 | 2.83E-05 | 7.09E-05 | 1.70016 |
| TKT | 0.385863 | 1.306641 | 8.546835 | 4.437219 | 2.84E-05 | 7.10E-05 | 1.698806 |
| SMC3 | 0.244974 | 1.185071 | 5.363418 | 4.437015 | 2.84E-05 | 7.10E-05 | 1.698077 |
| TMEM14A | -0.24316 | 0.844891 | 9.668861 | -4.43607 | 2.85E-05 | 7.12E-05 | 1.694699 |
| MGA | 0.250514 | 1.189631 | 5.815949 | 4.434911 | 2.86E-05 | 7.15E-05 | 1.690523 |
| C21orf2 | -0.18773 | 0.877984 | 6.338101 | -4.43451 | 2.86E-05 | 7.16E-05 | 1.68907 |
| SFXN3 | -0.21546 | 0.861271 | 7.281519 | -4.43296 | 2.88E-05 | 7.20E-05 | 1.683523 |

| | | | | | | | |
|---|---|---|---|---|---|---|---|
| RBM28 | 0.184111 | 1.136116 | 5.977342 | 4.432322 | 2.89E-05 | 7.22E-05 | 1.681233 |
| ME2 | 0.20002 | 1.148714 | 7.408987 | 4.430277 | 2.91E-05 | 7.27E-05 | 1.673898 |
| NUP54 | 0.202615 | 1.150783 | 7.210506 | 4.429469 | 2.92E-05 | 7.29E-05 | 1.671001 |
| PLCE1 | -0.32085 | 0.800598 | 5.172278 | -4.42589 | 2.96E-05 | 7.38E-05 | 1.658176 |
| PPARGC1A | -0.3741 | 0.771588 | 7.770253 | -4.42544 | 2.96E-05 | 7.40E-05 | 1.656541 |
| CREB5 | 0.421831 | 1.339627 | 4.943165 | 4.425037 | 2.97E-05 | 7.40E-05 | 1.655112 |
| BEST1 | -0.20838 | 0.865509 | 5.468608 | -4.42436 | 2.97E-05 | 7.42E-05 | 1.652671 |
| CECR5 | -0.18438 | 0.880027 | 7.564937 | -4.42333 | 2.99E-05 | 7.45E-05 | 1.648989 |
| DICER1 | 0.228544 | 1.171652 | 7.371772 | 4.422922 | 2.99E-05 | 7.46E-05 | 1.64753 |
| C22orf46 | -0.1302 | 0.913706 | 4.431266 | -4.42221 | 3.00E-05 | 7.47E-05 | 1.644964 |
| ZNF573 | 0.271772 | 1.20729 | 4.984177 | 4.421551 | 3.01E-05 | 7.49E-05 | 1.642621 |
| AQP3 | -0.43692 | 0.738708 | 9.286835 | -4.42016 | 3.02E-05 | 7.53E-05 | 1.637638 |
| CIDEC | -0.22021 | 0.85844 | 5.336709 | -4.41977 | 3.03E-05 | 7.54E-05 | 1.636247 |
| RBPMS | 0.23724 | 1.178735 | 7.756709 | 4.41748 | 3.05E-05 | 7.60E-05 | 1.628042 |
| SCN3A | 0.221528 | 1.165968 | 4.159494 | 4.416789 | 3.06E-05 | 7.61E-05 | 1.625566 |
| GPR162 | -0.18758 | 0.878076 | 5.353165 | -4.41582 | 3.07E-05 | 7.64E-05 | 1.622115 |
| NAT8 | -0.49043 | 0.711811 | 12.67557 | -4.41445 | 3.09E-05 | 7.68E-05 | 1.61718 |
| ZWINT | 0.382747 | 1.303822 | 6.635316 | 4.414244 | 3.09E-05 | 7.68E-05 | 1.616456 |
| FOS | -1.30179 | 0.405622 | 7.030127 | -4.41423 | 3.09E-05 | 7.68E-05 | 1.61642 |
| UAP1 | -0.43569 | 0.739343 | 8.031266 | -4.4136 | 3.10E-05 | 7.69E-05 | 1.614157 |
| EFCC1 | -0.24298 | 0.845 | 5.196329 | -4.41359 | 3.10E-05 | 7.69E-05 | 1.614115 |
| AUP1 | -0.18273 | 0.881036 | 7.733671 | -4.41209 | 3.11E-05 | 7.73E-05 | 1.608764 |
| SERPINF1 | 0.832984 | 1.781366 | 7.408608 | 4.411097 | 3.13E-05 | 7.76E-05 | 1.605199 |
| TMEM134 | -0.18607 | 0.878999 | 6.893797 | -4.4109 | 3.13E-05 | 7.76E-05 | 1.604511 |
| SLC2A2 | -0.55937 | 0.6786 | 8.100253 | -4.40835 | 3.16E-05 | 7.83E-05 | 1.595359 |
| RNMTL1 | -0.15505 | 0.898104 | 7.120886 | -4.40834 | 3.16E-05 | 7.83E-05 | 1.595342 |
| CUL5 | -0.17298 | 0.887006 | 7.037089 | -4.40792 | 3.16E-05 | 7.84E-05 | 1.593848 |
| GTF2A2 | 0.195171 | 1.14486 | 8.086962 | 4.407879 | 3.16E-05 | 7.84E-05 | 1.593691 |
| PPEF1 | -0.25445 | 0.838309 | 4.130886 | -4.40705 | 3.17E-05 | 7.86E-05 | 1.59074 |
| KLHL2 | 0.197062 | 1.146361 | 7.831013 | 4.405505 | 3.19E-05 | 7.91E-05 | 1.585204 |
| SNRNP25 | -0.2154 | 0.86131 | 7.896329 | -4.40474 | 3.20E-05 | 7.93E-05 | 1.582462 |
| RRP1 | -0.16366 | 0.89276 | 6.216203 | -4.40457 | 3.20E-05 | 7.93E-05 | 1.58187 |
| RHBDF1 | 0.167872 | 1.1234 | 5.721646 | 4.403213 | 3.22E-05 | 7.97E-05 | 1.577013 |
| MTMR6 | 0.189592 | 1.140441 | 6.223544 | 4.40071 | 3.25E-05 | 8.04E-05 | 1.568071 |
| DRP2 | -0.17076 | 0.888372 | 5.00519 | -4.40064 | 3.25E-05 | 8.04E-05 | 1.567813 |
| DERL1 | 0.186094 | 1.137679 | 7.180127 | 4.399008 | 3.27E-05 | 8.09E-05 | 1.561991 |
| GNB1 | 0.149545 | 1.10922 | 9.147468 | 4.398871 | 3.27E-05 | 8.09E-05 | 1.561501 |
| NFYB | 0.179025 | 1.132119 | 5.83 | 4.397085 | 3.29E-05 | 8.14E-05 | 1.555125 |
| CYBA | 0.367813 | 1.290395 | 9.778861 | 4.396213 | 3.30E-05 | 8.16E-05 | 1.552012 |
| SNRPD2 | 0.242628 | 1.183146 | 10.55418 | 4.395408 | 3.31E-05 | 8.19E-05 | 1.549137 |
| EXPH5 | -0.25351 | 0.838852 | 5.70519 | -4.3953 | 3.31E-05 | 8.19E-05 | 1.548752 |
| MRPS12 | -0.14986 | 0.901337 | 5.635443 | -4.39475 | 3.32E-05 | 8.20E-05 | 1.546789 |
| SCD | -0.24092 | 0.846204 | 7.112405 | -4.39461 | 3.32E-05 | 8.20E-05 | 1.546306 |
| SUGP1 | 0.204605 | 1.152371 | 6.291772 | 4.394094 | 3.33E-05 | 8.22E-05 | 1.544447 |
| ELP4 | -0.17798 | 0.883937 | 7.4 | -4.38992 | 3.38E-05 | 8.34E-05 | 1.529543 |
| HTATIP2 | 0.276423 | 1.211188 | 7.071772 | 4.389222 | 3.39E-05 | 8.36E-05 | 1.527067 |
| SOBP | -0.26192 | 0.833975 | 6.414937 | -4.38873 | 3.40E-05 | 8.37E-05 | 1.52533 |

| | | | | | | | |
|---|---|---|---|---|---|---|---|
| RGS6 | -0.14057 | 0.907163 | 5.032152 | -4.38786 | 3.41E-05 | 8.40E-05 | 1.522196 |
| AGPS | 0.294657 | 1.226594 | 6.887215 | 4.387404 | 3.41E-05 | 8.41E-05 | 1.520585 |
| CALR | 0.188893 | 1.139889 | 6.914557 | 4.386295 | 3.43E-05 | 8.44E-05 | 1.516631 |
| MLYCD | -0.21372 | 0.86231 | 7.780506 | -4.38584 | 3.43E-05 | 8.45E-05 | 1.51502 |
| ID1 | 0.619348 | 1.536181 | 8.229367 | 4.38513 | 3.44E-05 | 8.47E-05 | 1.51248 |
| RPS21 | 0.270909 | 1.206568 | 11.64316 | 4.381832 | 3.48E-05 | 8.58E-05 | 1.500727 |
| NINJ2 | 0.23056 | 1.17329 | 4.606962 | 4.380402 | 3.50E-05 | 8.62E-05 | 1.495633 |
| APOBEC3A | 0.328808 | 1.255975 | 3.882785 | 4.380184 | 3.51E-05 | 8.62E-05 | 1.494856 |
| RTN3 | 0.237194 | 1.178698 | 9.467342 | 4.379833 | 3.51E-05 | 8.63E-05 | 1.493608 |
| PRLH | -0.27315 | 0.827511 | 5.463291 | -4.37929 | 3.52E-05 | 8.65E-05 | 1.491657 |
| MRPL16 | -0.21592 | 0.860996 | 8.647848 | -4.37683 | 3.55E-05 | 8.72E-05 | 1.482924 |
| PITPNC1 | -0.2091 | 0.865078 | 6.894177 | -4.37592 | 3.56E-05 | 8.75E-05 | 1.479657 |
| VWA7 | -0.2014 | 0.869704 | 5.83 | -4.37533 | 3.57E-05 | 8.77E-05 | 1.477584 |
| ATF3 | -1.11208 | 0.462626 | 6.436329 | -4.3746 | 3.58E-05 | 8.79E-05 | 1.474991 |
| ADRBK1 | 0.161627 | 1.118548 | 5.811646 | 4.372415 | 3.61E-05 | 8.86E-05 | 1.467202 |
| EHF | 0.351469 | 1.275859 | 4.447468 | 4.371477 | 3.62E-05 | 8.89E-05 | 1.463865 |
| HPSE2 | -0.2164 | 0.860713 | 5.127215 | -4.36952 | 3.65E-05 | 8.95E-05 | 1.456913 |
| DYNLT3 | -0.22608 | 0.854955 | 10.52013 | -4.36872 | 3.66E-05 | 8.97E-05 | 1.454047 |
| PCSK5 | 0.197958 | 1.147074 | 4.35443 | 4.366819 | 3.68E-05 | 9.03E-05 | 1.4473 |
| SLC25A5 | -0.18049 | 0.882401 | 12.38873 | -4.36631 | 3.69E-05 | 9.05E-05 | 1.445475 |
| TM9SF1 | 0.211482 | 1.157877 | 7.935949 | 4.365593 | 3.70E-05 | 9.07E-05 | 1.442943 |
| KIF13B | -0.22748 | 0.854123 | 7.663671 | -4.36418 | 3.72E-05 | 9.11E-05 | 1.437925 |
| ADH1B | 0.942859 | 1.922334 | 8.003418 | 4.364085 | 3.72E-05 | 9.11E-05 | 1.437581 |
| CCT3 | 0.160474 | 1.117655 | 9.787468 | 4.361147 | 3.76E-05 | 9.21E-05 | 1.427145 |
| IGLV3-25 | 0.856976 | 1.811238 | 4.396456 | 4.359212 | 3.79E-05 | 9.27E-05 | 1.420272 |
| C19orf73 | -0.14158 | 0.906525 | 4.836076 | -4.35819 | 3.80E-05 | 9.31E-05 | 1.416635 |
| MAL | 0.30664 | 1.236824 | 10.49418 | 4.356555 | 3.83E-05 | 9.36E-05 | 1.410837 |
| TRIP12 | 0.181733 | 1.134245 | 8.805696 | 4.356275 | 3.83E-05 | 9.37E-05 | 1.409843 |
| ZDHHC6 | -0.21484 | 0.861641 | 8.618734 | -4.35626 | 3.83E-05 | 9.37E-05 | 1.409796 |
| DHCR7 | -0.23027 | 0.852475 | 6.336203 | -4.35614 | 3.83E-05 | 9.37E-05 | 1.409355 |
| MED8 | -0.1329 | 0.911997 | 6.844051 | -4.35482 | 3.85E-05 | 9.41E-05 | 1.404694 |
| HTRA1 | -0.42244 | 0.74616 | 8.898101 | -4.35426 | 3.86E-05 | 9.43E-05 | 1.402682 |
| ZBTB22 | -0.18217 | 0.881374 | 6.286076 | -4.35211 | 3.89E-05 | 9.50E-05 | 1.395049 |
| GLTSCR2 | 0.225237 | 1.168969 | 11.4143 | 4.350473 | 3.91E-05 | 9.55E-05 | 1.389258 |
| NEMF | 0.279137 | 1.213469 | 6.975949 | 4.34884 | 3.94E-05 | 9.61E-05 | 1.383466 |
| YWHAE | -0.25428 | 0.838408 | 9.411392 | -4.34802 | 3.95E-05 | 9.64E-05 | 1.380558 |
| ATP6V1E1 | -0.16466 | 0.89214 | 10.66709 | -4.34471 | 4.00E-05 | 9.75E-05 | 1.368842 |
| GNB2L1 | 0.180929 | 1.133614 | 12.31101 | 4.343578 | 4.01E-05 | 9.79E-05 | 1.364813 |
| RPL14 | 0.183188 | 1.13539 | 11.96 | 4.341126 | 4.05E-05 | 9.88E-05 | 1.356129 |
| SGCA | -0.1752 | 0.885642 | 4.218987 | -4.34016 | 4.07E-05 | 9.91E-05 | 1.352707 |
| ARC | -0.23328 | 0.850698 | 4.53038 | -4.33975 | 4.07E-05 | 9.92E-05 | 1.351239 |
| LYRM9 | -0.21675 | 0.8605 | 6.239241 | -4.3396 | 4.07E-05 | 9.92E-05 | 1.35073 |
| THRB | -0.21543 | 0.86129 | 6.493924 | -4.33859 | 4.09E-05 | 9.96E-05 | 1.347137 |

Table S1 Olive vs. Control

| Gene | logFC | UnlogFC | AveExpr | t | P.Value | adj.P.Val | B |
|---|---|---|---|---|---|---|---|
| DUSP1 | -1.73342 | 0.300739 | 7.536267 | -10.0838 | 9.26E-16 | 7.83E-12 | 25.41705 |
| CEBPB | -1.15716 | 0.448395 | 8.368 | -9.62969 | 6.89E-15 | 2.15E-11 | 23.47789 |
| RHOB | -1.0913 | 0.469337 | 9.329333 | -9.6068 | 7.62E-15 | 2.15E-11 | 23.37979 |
| ZFP36 | -1.28408 | 0.410633 | 8.336533 | -9.50602 | 1.19E-14 | 2.52E-11 | 22.94751 |
| TDP2 | 0.383703 | 1.304686 | 10.6088 | 9.314425 | 2.79E-14 | 4.72E-11 | 22.12436 |
| NFKBIA | -0.92085 | 0.528199 | 9.4872 | -9.21982 | 4.25E-14 | 5.99E-11 | 21.71729 |
| FOSL2 | -0.51765 | 0.698507 | 5.672667 | -9.14681 | 5.88E-14 | 6.51E-11 | 21.40292 |
| SGK1 | -1.03516 | 0.48796 | 10.58387 | -9.13633 | 6.16E-14 | 6.51E-11 | 21.3578 |
| FILIP1L | -0.78501 | 0.580349 | 5.617333 | -8.89322 | 1.82E-13 | 1.71E-10 | 20.30971 |
| MTERF1 | 0.4906 | 1.405029 | 5.040133 | 8.866635 | 2.05E-13 | 1.73E-10 | 20.19502 |
| ZZZ3 | 0.524835 | 1.438769 | 7.127067 | 8.666232 | 5.00E-13 | 3.61E-10 | 19.33003 |
| CCDC51 | 0.447354 | 1.363537 | 8.234933 | 8.646126 | 5.47E-13 | 3.61E-10 | 19.24323 |
| ABCG1 | 0.742046 | 1.672547 | 5.944533 | 8.64266 | 5.56E-13 | 3.61E-10 | 19.22826 |
| RGS2 | -1.46122 | 0.363185 | 7.871733 | -8.6266 | 5.97E-13 | 3.61E-10 | 19.15891 |
| KLF6 | -1.37289 | 0.386118 | 8.178933 | -8.52847 | 9.25E-13 | 4.82E-10 | 18.73522 |
| MAFF | -2.44483 | 0.183668 | 6.494667 | -8.50376 | 1.03E-12 | 4.82E-10 | 18.62853 |
| ZSCAN26 | 0.397001 | 1.316768 | 6.131333 | 8.502699 | 1.04E-12 | 4.82E-10 | 18.62394 |
| PDK4 | -1.76525 | 0.294176 | 7.696133 | -8.49614 | 1.07E-12 | 4.82E-10 | 18.59563 |
| ZBED8 | 0.531012 | 1.444942 | 4.562933 | 8.493184 | 1.08E-12 | 4.82E-10 | 18.58285 |
| ZFAND5 | -0.78275 | 0.581257 | 9.751467 | -8.46295 | 1.24E-12 | 5.24E-10 | 18.45231 |
| CITED2 | 0.785345 | 1.723504 | 7.218667 | 8.434438 | 1.41E-12 | 5.45E-10 | 18.32921 |
| SYNJ1 | 0.410802 | 1.329425 | 5.5308 | 8.432451 | 1.42E-12 | 5.45E-10 | 18.32063 |
| CDKN1A | -0.88862 | 0.54013 | 8.1764 | -8.38359 | 1.76E-12 | 6.48E-10 | 18.10967 |
| PER1 | -0.60814 | 0.656042 | 7.4792 | -8.33809 | 2.16E-12 | 7.61E-10 | 17.91326 |
| USP8 | 0.335067 | 1.261436 | 7.301733 | 8.317989 | 2.36E-12 | 7.94E-10 | 17.8265 |
| MGAT2 | 0.339843 | 1.265618 | 6.384667 | 8.310749 | 2.44E-12 | 7.94E-10 | 17.79525 |
| CRNKL1 | 0.436027 | 1.352874 | 7.199467 | 8.2705 | 2.92E-12 | 9.02E-10 | 17.62154 |
| FBXO28 | 0.456619 | 1.372322 | 7.853733 | 8.265278 | 2.99E-12 | 9.02E-10 | 17.59901 |
| FOS | -2.26564 | 0.207958 | 6.697867 | -8.22734 | 3.54E-12 | 1.03E-09 | 17.4353 |
| B4GALT4 | 0.364033 | 1.287019 | 6.812933 | 8.133298 | 5.38E-12 | 1.52E-09 | 17.02964 |
| CDC23 | 0.401634 | 1.321003 | 7.355733 | 8.113522 | 5.88E-12 | 1.57E-09 | 16.94437 |
| GOLPH3L | 0.342091 | 1.267593 | 8.744667 | 8.104148 | 6.13E-12 | 1.57E-09 | 16.90395 |
| EGR1 | -1.68223 | 0.3116 | 7.008667 | -8.10358 | 6.14E-12 | 1.57E-09 | 16.90151 |
| CNOT6 | 0.339348 | 1.265185 | 7.071867 | 8.096794 | 6.33E-12 | 1.57E-09 | 16.87224 |
| ZC3H7A | 0.446312 | 1.362553 | 6.4104 | 8.075952 | 6.95E-12 | 1.65E-09 | 16.78239 |
| SPRY4 | -0.52864 | 0.693206 | 6.3532 | -8.07333 | 7.03E-12 | 1.65E-09 | 16.77111 |
| PPID | 0.470022 | 1.385131 | 7.884133 | 8.038422 | 8.21E-12 | 1.88E-09 | 16.62063 |
| APOLD1 | -0.96534 | 0.512156 | 5.681733 | -7.99261 | 1.01E-11 | 2.24E-09 | 16.42323 |
| IER3 | -1.28487 | 0.410407 | 8.5284 | -7.95711 | 1.18E-11 | 2.48E-09 | 16.27034 |
| DUSP6 | -0.88558 | 0.541271 | 7.764533 | -7.95591 | 1.18E-11 | 2.48E-09 | 16.26519 |
| TOMM70A | 0.344475 | 1.269689 | 8.299067 | 7.952934 | 1.20E-11 | 2.48E-09 | 16.25236 |
| ARMC1 | 0.382474 | 1.303575 | 7.820933 | 7.886222 | 1.61E-11 | 3.25E-09 | 15.96517 |
| RTCA | 0.447586 | 1.363757 | 9.293067 | 7.823605 | 2.13E-11 | 4.13E-09 | 15.6958 |
| KMO | 0.562054 | 1.47637 | 9.559067 | 7.822111 | 2.15E-11 | 4.13E-09 | 15.68938 |
| CEBPD | -0.8615 | 0.55038 | 10.0108 | -7.80242 | 2.34E-11 | 4.40E-09 | 15.60469 |

| | | | | | | | |
|---|---|---|---|---|---|---|---|
| NOL11 | 0.375712 | 1.29748 | 8.262667 | 7.780669 | 2.58E-11 | 4.74E-09 | 15.51121 |
| JUNB | -1.15153 | 0.450148 | 6.432133 | -7.74963 | 2.96E-11 | 5.33E-09 | 15.37784 |
| GADD45B | -1.4271 | 0.371878 | 8.226667 | -7.74018 | 3.09E-11 | 5.44E-09 | 15.33722 |
| CISH | -0.8913 | 0.539129 | 8.2308 | -7.69013 | 3.86E-11 | 6.65E-09 | 15.12231 |
| RGS3 | -0.58136 | 0.668335 | 6.3676 | -7.65644 | 4.48E-11 | 7.57E-09 | 14.97768 |
| ARMT1 | 0.579063 | 1.493879 | 8.5276 | 7.637015 | 4.88E-11 | 8.09E-09 | 14.89436 |
| MYLIP | 0.525097 | 1.439031 | 7.241733 | 7.54733 | 7.26E-11 | 1.18E-08 | 14.50993 |
| PLCL2 | 0.528276 | 1.442205 | 7.364267 | 7.532895 | 7.74E-11 | 1.23E-08 | 14.44811 |
| AIF1 | -0.41814 | 0.748388 | 7.662667 | -7.49056 | 9.33E-11 | 1.46E-08 | 14.26688 |
| PRKACB | 0.366754 | 1.289448 | 7.543333 | 7.486892 | 9.48E-11 | 1.46E-08 | 14.2512 |
| BHLHB9 | 0.497751 | 1.412011 | 5.573333 | 7.477307 | 9.89E-11 | 1.49E-08 | 14.21019 |
| ANGPTL3 | 0.804273 | 1.746265 | 8.875333 | 7.45947 | 1.07E-10 | 1.59E-08 | 14.1339 |
| CCT6A | 0.364138 | 1.287112 | 7.9708 | 7.430684 | 1.22E-10 | 1.77E-08 | 14.01083 |
| ETS2 | -0.65055 | 0.637035 | 6.7828 | -7.40756 | 1.35E-10 | 1.93E-08 | 13.91202 |
| MTMR4 | 0.408201 | 1.32703 | 7.916533 | 7.377191 | 1.54E-10 | 2.17E-08 | 13.78231 |
| RASA1 | 0.28404 | 1.2176 | 7.296133 | 7.349151 | 1.74E-10 | 2.41E-08 | 13.66263 |
| ANK3 | 0.274775 | 1.209805 | 8.917333 | 7.336559 | 1.84E-10 | 2.51E-08 | 13.60891 |
| MTIF2 | 0.290517 | 1.223079 | 8.087333 | 7.324977 | 1.94E-10 | 2.60E-08 | 13.55951 |
| OARD1 | 0.4203 | 1.338206 | 8.227733 | 7.307481 | 2.09E-10 | 2.76E-08 | 13.4849 |
| POLR3B | 0.355982 | 1.279856 | 7.293733 | 7.273405 | 2.43E-10 | 3.16E-08 | 13.33969 |
| DHX40 | 0.360682 | 1.284033 | 7.955333 | 7.265738 | 2.51E-10 | 3.22E-08 | 13.30703 |
| CTGF | -1.12693 | 0.457888 | 8.6536 | -7.23744 | 2.85E-10 | 3.59E-08 | 13.18652 |
| JUND | -0.5754 | 0.671098 | 6.049467 | -7.22707 | 2.98E-10 | 3.71E-08 | 13.14239 |
| DRAM1 | 0.385915 | 1.306688 | 8.428133 | 7.198059 | 3.39E-10 | 4.15E-08 | 13.019 |
| RCBTB1 | 0.621732 | 1.538721 | 6.444533 | 7.193375 | 3.46E-10 | 4.17E-08 | 12.99909 |
| CYR61 | -1.38416 | 0.383112 | 7.3124 | -7.18618 | 3.57E-10 | 4.22E-08 | 12.96852 |
| STX3 | 0.499385 | 1.413611 | 7.2844 | 7.184382 | 3.60E-10 | 4.22E-08 | 12.96085 |
| FAM134B | -0.58765 | 0.665428 | 9.389733 | -7.11138 | 4.95E-10 | 5.74E-08 | 12.65084 |
| ALDH8A1 | 0.375862 | 1.297615 | 10.56533 | 7.104775 | 5.10E-10 | 5.80E-08 | 12.6228 |
| TRMT1L | 0.408051 | 1.326892 | 6.053867 | 7.102511 | 5.15E-10 | 5.80E-08 | 12.6132 |
| ZBED5 | 0.3847 | 1.305588 | 8.358533 | 7.098275 | 5.25E-10 | 5.84E-08 | 12.59523 |
| TMEM135 | 0.466207 | 1.381473 | 7.730267 | 7.079374 | 5.70E-10 | 6.26E-08 | 12.51509 |
| BBS10 | 0.486057 | 1.400612 | 5.895333 | 7.045026 | 6.62E-10 | 7.18E-08 | 12.36955 |
| NR4A1 | -1.12069 | 0.459874 | 6.266667 | -7.02655 | 7.18E-10 | 7.69E-08 | 12.29133 |
| TUBAL3 | 0.478298 | 1.3931 | 7.597333 | 7.012436 | 7.64E-10 | 8.07E-08 | 12.23159 |
| ABCA1 | 0.613171 | 1.529617 | 8.554267 | 6.984247 | 8.64E-10 | 9.02E-08 | 12.11237 |
| MOSPD1 | 0.290397 | 1.222977 | 7.0912 | 6.978916 | 8.84E-10 | 9.12E-08 | 12.08983 |
| NUP107 | 0.311132 | 1.240681 | 7.564 | 6.972686 | 9.09E-10 | 9.26E-08 | 12.0635 |
| CCNL1 | -0.49289 | 0.710598 | 7.485067 | -6.96485 | 9.41E-10 | 9.47E-08 | 12.0304 |
| ZNF268 | 0.419048 | 1.337045 | 6.167467 | 6.936414 | 1.06E-09 | 1.06E-07 | 11.91029 |
| RND3 | -0.74926 | 0.59491 | 7.4692 | -6.92875 | 1.10E-09 | 1.08E-07 | 11.87795 |
| C2orf42 | 0.33599 | 1.262243 | 6.620133 | 6.91362 | 1.18E-09 | 1.14E-07 | 11.8141 |
| GDF15 | -1.38082 | 0.383999 | 7.941733 | -6.90462 | 1.22E-09 | 1.16E-07 | 11.77613 |
| GLT8D1 | 0.235022 | 1.176925 | 7.728267 | 6.904176 | 1.23E-09 | 1.16E-07 | 11.77426 |
| FOSB | -1.73757 | 0.299874 | 6.0764 | -6.87274 | 1.41E-09 | 1.32E-07 | 11.64175 |
| TSPYL2 | -0.44111 | 0.736568 | 6.868133 | -6.86112 | 1.48E-09 | 1.37E-07 | 11.59281 |
| RAB40B | 0.378591 | 1.300071 | 8.589867 | 6.8541 | 1.52E-09 | 1.39E-07 | 11.56325 |

| | | | | | | | |
|---|---|---|---|---|---|---|---|
| MTTP | 0.546987 | 1.461031 | 6.861067 | 6.853511 | 1.53E-09 | 1.39E-07 | 11.56077 |
| ASF1A | 0.378058 | 1.299592 | 6.8964 | 6.84382 | 1.59E-09 | 1.43E-07 | 11.51997 |
| FAM98A | 0.358868 | 1.282419 | 8.565067 | 6.806242 | 1.88E-09 | 1.67E-07 | 11.3619 |
| ZNF302 | 0.415735 | 1.333978 | 5.880533 | 6.796281 | 1.96E-09 | 1.73E-07 | 11.32003 |
| METTL9 | 0.206567 | 1.153939 | 10.38813 | 6.793351 | 1.99E-09 | 1.73E-07 | 11.30772 |
| LIPG | -0.78535 | 0.58021 | 5.2672 | -6.78832 | 2.03E-09 | 1.75E-07 | 11.28658 |
| PIK3R3 | 0.291079 | 1.223555 | 5.627333 | 6.778257 | 2.12E-09 | 1.81E-07 | 11.24431 |
| NUP37 | 0.313583 | 1.242791 | 7.5856 | 6.771382 | 2.18E-09 | 1.85E-07 | 11.21545 |
| CSTF2T | 0.284198 | 1.217733 | 6.482933 | 6.7691 | 2.21E-09 | 1.85E-07 | 11.20586 |
| PPP1R15A | -0.49681 | 0.708674 | 5.917467 | -6.727 | 2.65E-09 | 2.18E-07 | 11.02925 |
| COMMD8 | 0.314423 | 1.243514 | 9.698533 | 6.726027 | 2.66E-09 | 2.18E-07 | 11.02517 |
| ATF7IP | 0.339393 | 1.265224 | 7.066667 | 6.702292 | 2.95E-09 | 2.40E-07 | 10.92572 |
| JUN | -1.28773 | 0.409595 | 6.6136 | -6.69869 | 2.99E-09 | 2.41E-07 | 10.91064 |
| EXOC1 | 0.352766 | 1.277007 | 8.040533 | 6.694232 | 3.05E-09 | 2.43E-07 | 10.89196 |
| PHKB | 0.329145 | 1.256269 | 7.7664 | 6.67218 | 3.36E-09 | 2.65E-07 | 10.79967 |
| DNAJC13 | 0.259153 | 1.196776 | 6.023467 | 6.62411 | 4.13E-09 | 3.24E-07 | 10.59875 |
| PPCS | 0.224783 | 1.168601 | 9.552133 | 6.608056 | 4.43E-09 | 3.43E-07 | 10.53174 |
| TERF1 | 0.247594 | 1.187225 | 7.659867 | 6.606066 | 4.47E-09 | 3.43E-07 | 10.52343 |
| RAB29 | 0.396874 | 1.316652 | 8.481067 | 6.585288 | 4.89E-09 | 3.72E-07 | 10.43676 |
| SUOX | 0.278171 | 1.212656 | 8.899733 | 6.582209 | 4.95E-09 | 3.74E-07 | 10.42393 |
| BAIAP2 | -0.41103 | 0.752084 | 5.861067 | -6.56808 | 5.27E-09 | 3.94E-07 | 10.36506 |
| FPGT | 0.367429 | 1.290052 | 5.945333 | 6.542059 | 5.89E-09 | 4.37E-07 | 10.25668 |
| PTGDS | -0.97798 | 0.507691 | 7.947067 | -6.53067 | 6.19E-09 | 4.55E-07 | 10.20929 |
| VPS45 | 0.268598 | 1.204637 | 7.381467 | 6.522995 | 6.39E-09 | 4.64E-07 | 10.17737 |
| DCP2 | 0.320615 | 1.248863 | 6.202667 | 6.521896 | 6.42E-09 | 4.64E-07 | 10.1728 |
| KRIT1 | 0.291919 | 1.224268 | 5.420267 | 6.514565 | 6.63E-09 | 4.75E-07 | 10.14232 |
| TNS2 | -0.36036 | 0.77897 | 6.5924 | -6.5033 | 6.96E-09 | 4.94E-07 | 10.09552 |
| CHI3L1 | -1.01484 | 0.494882 | 5.769333 | -6.50157 | 7.01E-09 | 4.94E-07 | 10.08832 |
| MCL1 | -0.49469 | 0.709717 | 7.8172 | -6.49907 | 7.09E-09 | 4.95E-07 | 10.07793 |
| RCBTB2 | 0.287811 | 1.220787 | 8.1452 | 6.49476 | 7.22E-09 | 5.00E-07 | 10.06002 |
| FAM69A | 0.483906 | 1.398525 | 7.129067 | 6.487514 | 7.45E-09 | 5.12E-07 | 10.02992 |
| RARS | 0.321207 | 1.249375 | 8.7792 | 6.469711 | 8.04E-09 | 5.48E-07 | 9.956027 |
| NR4A3 | -0.60214 | 0.658774 | 4.575867 | -6.46395 | 8.24E-09 | 5.57E-07 | 9.932111 |
| GPATCH1 | 0.274858 | 1.209875 | 5.5228 | 6.462096 | 8.31E-09 | 5.57E-07 | 9.924436 |
| ATF3 | -1.71259 | 0.305111 | 6.238667 | -6.45483 | 8.57E-09 | 5.71E-07 | 9.894298 |
| TSC22D3 | -0.93804 | 0.521943 | 8.859467 | -6.42049 | 9.93E-09 | 6.54E-07 | 9.752022 |
| FNTA | 0.252211 | 1.191031 | 8.066 | 6.419318 | 9.98E-09 | 6.54E-07 | 9.747166 |
| KIAA0391 | 0.227309 | 1.170649 | 8.375067 | 6.400797 | 1.08E-08 | 7.03E-07 | 9.670518 |
| NR0B2 | -0.50346 | 0.705415 | 6.5412 | -6.38038 | 1.18E-08 | 7.57E-07 | 9.586116 |
| LDLR | -0.65249 | 0.636182 | 5.2564 | -6.37994 | 1.18E-08 | 7.57E-07 | 9.584292 |
| BTG2 | -0.84591 | 0.556358 | 7.144 | -6.36792 | 1.24E-08 | 7.91E-07 | 9.53461 |
| BET1 | 0.278951 | 1.213312 | 7.9496 | 6.353078 | 1.33E-08 | 8.36E-07 | 9.473341 |
| FASTKD3 | 0.431957 | 1.349062 | 7.625067 | 6.34523 | 1.37E-08 | 8.58E-07 | 9.440957 |
| STX7 | 0.276499 | 1.211252 | 8.958 | 6.337978 | 1.41E-08 | 8.79E-07 | 9.411039 |
| RBM47 | 0.281094 | 1.215116 | 9.771733 | 6.327599 | 1.48E-08 | 9.12E-07 | 9.368241 |
| RWDD3 | 0.378823 | 1.300281 | 6.218 | 6.312195 | 1.58E-08 | 9.63E-07 | 9.304765 |
| TAOK3 | 0.298576 | 1.22993 | 7.190667 | 6.31137 | 1.58E-08 | 9.63E-07 | 9.301366 |

| | | | | | | | |
|---|---|---|---|---|---|---|---|
| SEPHS2 | 0.261222 | 1.198493 | 10.58427 | 6.307814 | 1.61E-08 | 9.71E-07 | 9.286717 |
| DENR | 0.286147 | 1.219379 | 8.6376 | 6.302435 | 1.65E-08 | 9.85E-07 | 9.264568 |
| CCNG2 | 0.357774 | 1.281447 | 8.3216 | 6.301106 | 1.65E-08 | 9.85E-07 | 9.259096 |
| STEAP1 | 0.518853 | 1.432816 | 6.908667 | 6.287775 | 1.75E-08 | 1.04E-06 | 9.204228 |
| TRIP4 | 0.31593 | 1.244813 | 8.319333 | 6.276594 | 1.84E-08 | 1.08E-06 | 9.158233 |
| SDPR | -0.42372 | 0.745501 | 5.317467 | -6.27052 | 1.89E-08 | 1.09E-06 | 9.13325 |
| FAM179B | 0.35994 | 1.283373 | 7.166133 | 6.270195 | 1.89E-08 | 1.09E-06 | 9.131925 |
| MINPP1 | 0.385862 | 1.30664 | 7.7592 | 6.266021 | 1.92E-08 | 1.11E-06 | 9.114766 |
| NEDD9 | -0.54104 | 0.687274 | 7.446667 | -6.2643 | 1.94E-08 | 1.11E-06 | 9.107679 |
| NR3C1 | 0.306004 | 1.236279 | 7.4768 | 6.261997 | 1.96E-08 | 1.11E-06 | 9.098228 |
| NUDT9 | 0.258793 | 1.196477 | 8.315067 | 6.257761 | 1.99E-08 | 1.12E-06 | 9.080825 |
| NOC3L | 0.332339 | 1.259053 | 6.6172 | 6.247308 | 2.08E-08 | 1.17E-06 | 9.037889 |
| RAB20 | -0.59565 | 0.661745 | 8.795333 | -6.23886 | 2.16E-08 | 1.20E-06 | 9.003223 |
| LAMTOR3 | 0.28063 | 1.214725 | 8.435467 | 6.23586 | 2.19E-08 | 1.21E-06 | 8.990895 |
| CTCF | 0.232684 | 1.175019 | 7.628667 | 6.230252 | 2.24E-08 | 1.23E-06 | 8.967881 |
| CHSY1 | -0.47702 | 0.718458 | 6.835333 | -6.22359 | 2.30E-08 | 1.26E-06 | 8.940548 |
| CDK7 | 0.375262 | 1.297075 | 7.584667 | 6.211235 | 2.43E-08 | 1.31E-06 | 8.889897 |
| NAT1 | 0.380682 | 1.301957 | 7.843067 | 6.209225 | 2.45E-08 | 1.32E-06 | 8.881659 |
| TYRO3 | -0.30968 | 0.806822 | 5.777867 | -6.20665 | 2.47E-08 | 1.32E-06 | 8.871092 |
| ZBTB11 | 0.367361 | 1.289991 | 6.788133 | 6.19697 | 2.58E-08 | 1.37E-06 | 8.831451 |
| MGEA5 | 0.344393 | 1.269617 | 8.1336 | 6.191873 | 2.63E-08 | 1.39E-06 | 8.810579 |
| SLCO4A1 | -1.00253 | 0.499123 | 6.706267 | -6.18024 | 2.77E-08 | 1.45E-06 | 8.762949 |
| VAV3 | 0.415885 | 1.334116 | 8.6032 | 6.178868 | 2.78E-08 | 1.45E-06 | 8.757345 |
| KANSL1L | 0.300517 | 1.231586 | 5.9212 | 6.173973 | 2.84E-08 | 1.47E-06 | 8.737316 |
| TBL1XR1 | 0.378966 | 1.300409 | 6.616533 | 6.16774 | 2.92E-08 | 1.49E-06 | 8.711824 |
| HBEGF | -0.69636 | 0.617129 | 5.708133 | -6.16692 | 2.93E-08 | 1.49E-06 | 8.708479 |
| CDK8 | 0.25521 | 1.193509 | 5.429333 | 6.166478 | 2.93E-08 | 1.49E-06 | 8.706665 |
| RGS16 | -0.23771 | 0.848088 | 5.177867 | -6.16097 | 3.00E-08 | 1.52E-06 | 8.684159 |
| DAB2 | 0.23931 | 1.180428 | 10.83253 | 6.15664 | 3.06E-08 | 1.54E-06 | 8.666444 |
| SLC35A5 | 0.29967 | 1.230863 | 7.134133 | 6.152741 | 3.11E-08 | 1.56E-06 | 8.650508 |
| TFEC | 0.478388 | 1.393186 | 7.646933 | 6.128928 | 3.44E-08 | 1.71E-06 | 8.553268 |
| TMEM251 | 0.340315 | 1.266033 | 7.685067 | 6.128176 | 3.45E-08 | 1.71E-06 | 8.550198 |
| RIOK3 | 0.193718 | 1.143708 | 7.5836 | 6.122689 | 3.53E-08 | 1.73E-06 | 8.52781 |
| ZNF112 | 0.293651 | 1.225738 | 4.875067 | 6.122491 | 3.53E-08 | 1.73E-06 | 8.527 |
| HES1 | -0.56868 | 0.674233 | 7.147067 | -6.09663 | 3.94E-08 | 1.92E-06 | 8.421561 |
| GNPDA1 | 0.378441 | 1.299936 | 9.4272 | 6.089625 | 4.06E-08 | 1.96E-06 | 8.393045 |
| GEM | -0.97451 | 0.508914 | 6.4408 | -6.07298 | 4.36E-08 | 2.09E-06 | 8.325311 |
| NAA35 | 0.237159 | 1.178669 | 5.766267 | 6.071695 | 4.38E-08 | 2.09E-06 | 8.320065 |
| DHX29 | 0.435862 | 1.352719 | 7.828533 | 6.063531 | 4.53E-08 | 2.15E-06 | 8.28686 |
| NPL | 0.466934 | 1.382169 | 9.3012 | 6.051487 | 4.77E-08 | 2.25E-06 | 8.237903 |
| YARS2 | 0.224483 | 1.168358 | 6.8268 | 6.047154 | 4.86E-08 | 2.28E-06 | 8.220295 |
| UBXN8 | 0.248418 | 1.187904 | 7.204533 | 6.046514 | 4.87E-08 | 2.28E-06 | 8.217694 |
| KIAA0430 | 0.307309 | 1.237397 | 6.776 | 6.029468 | 5.23E-08 | 2.43E-06 | 8.148484 |
| DMXL1 | 0.535585 | 1.44953 | 8.624267 | 6.025803 | 5.32E-08 | 2.46E-06 | 8.133611 |
| IER2 | -0.71142 | 0.610717 | 9.460133 | -6.01429 | 5.58E-08 | 2.56E-06 | 8.0869 |
| ZNF638 | 0.30096 | 1.231964 | 8.025067 | 6.013291 | 5.60E-08 | 2.56E-06 | 8.082858 |
| IFIH1 | 0.697024 | 1.621157 | 5.819733 | 6.007975 | 5.73E-08 | 2.60E-06 | 8.06131 |

| | | | | | | | |
|---|---|---|---|---|---|---|---|
| NUP54 | 0.24413 | 1.184379 | 7.220267 | 6.006858 | 5.76E-08 | 2.60E-06 | 8.05678 |
| FMR1 | 0.283478 | 1.217126 | 9.116133 | 5.959308 | 7.03E-08 | 3.15E-06 | 7.864323 |
| LPCAT3 | 0.323576 | 1.251428 | 6.765333 | 5.957981 | 7.07E-08 | 3.15E-06 | 7.858959 |
| COPB2 | 0.184888 | 1.136728 | 9.324533 | 5.957721 | 7.08E-08 | 3.15E-06 | 7.857909 |
| TIPARP | -0.89047 | 0.539438 | 8.655467 | -5.94987 | 7.31E-08 | 3.24E-06 | 7.826175 |
| ACVR1 | 0.343411 | 1.268753 | 8.372133 | 5.941959 | 7.56E-08 | 3.33E-06 | 7.794239 |
| SOCS2 | -0.77295 | 0.585218 | 6.783733 | -5.93857 | 7.67E-08 | 3.36E-06 | 7.780551 |
| NFASC | -0.38217 | 0.767281 | 5.0544 | -5.93666 | 7.73E-08 | 3.37E-06 | 7.772836 |
| DUSP5 | -0.70953 | 0.61152 | 5.215867 | -5.93452 | 7.80E-08 | 3.38E-06 | 7.764194 |
| UNC119B | 0.207661 | 1.154815 | 9.2416 | 5.932121 | 7.88E-08 | 3.40E-06 | 7.754533 |
| GMFB | 0.297721 | 1.229201 | 8.099467 | 5.926513 | 8.06E-08 | 3.46E-06 | 7.731906 |
| FYCO1 | 0.278036 | 1.212543 | 6.525333 | 5.924539 | 8.13E-08 | 3.47E-06 | 7.723945 |
| SKAP2 | 0.242961 | 1.183419 | 7.235467 | 5.919665 | 8.30E-08 | 3.53E-06 | 7.704288 |
| WDFY3 | 0.266469 | 1.20286 | 7.3676 | 5.903578 | 8.88E-08 | 3.75E-06 | 7.639458 |
| NAT8B | 0.717496 | 1.644326 | 8.797867 | 5.866231 | 1.04E-07 | 4.36E-06 | 7.489195 |
| NLGN1 | 0.354048 | 1.278142 | 5.167333 | 5.855222 | 1.09E-07 | 4.53E-06 | 7.44497 |
| ZNF33B | 0.30003 | 1.23117 | 6.082533 | 5.854681 | 1.09E-07 | 4.53E-06 | 7.442797 |
| PDSS2 | 0.316784 | 1.245551 | 8.360533 | 5.842651 | 1.14E-07 | 4.74E-06 | 7.394507 |
| EIF2B1 | 0.172016 | 1.126632 | 8.1376 | 5.836776 | 1.17E-07 | 4.84E-06 | 7.370935 |
| SLC25A38 | 0.20422 | 1.152064 | 7.9844 | 5.830991 | 1.20E-07 | 4.93E-06 | 7.347735 |
| GPRC5A | -0.34169 | 0.789118 | 6.288533 | -5.81862 | 1.27E-07 | 5.17E-06 | 7.298141 |
| COX15 | 0.236852 | 1.178418 | 7.1568 | 5.808282 | 1.32E-07 | 5.37E-06 | 7.256748 |
| CKAP2 | 0.272871 | 1.20821 | 4.567467 | 5.807069 | 1.33E-07 | 5.37E-06 | 7.251892 |
| AP3M2 | 0.264265 | 1.201024 | 7.0224 | 5.797478 | 1.38E-07 | 5.56E-06 | 7.213505 |
| NARS | 0.176612 | 1.130226 | 11.10133 | 5.78045 | 1.48E-07 | 5.94E-06 | 7.145416 |
| STAMBP | 0.207114 | 1.154377 | 7.709867 | 5.770322 | 1.55E-07 | 6.17E-06 | 7.104954 |
| SPOCK2 | -0.36736 | 0.775199 | 6.741867 | -5.76218 | 1.60E-07 | 6.30E-06 | 7.072443 |
| WDR12 | 0.296649 | 1.228288 | 8.7084 | 5.761838 | 1.60E-07 | 6.30E-06 | 7.071083 |
| THAP10 | 0.261882 | 1.199041 | 6.748 | 5.761748 | 1.60E-07 | 6.30E-06 | 7.070724 |
| FGFR1OP | 0.306754 | 1.236922 | 6.760133 | 5.756924 | 1.63E-07 | 6.40E-06 | 7.051473 |
| ADD3 | 0.292579 | 1.224828 | 10.344 | 5.753826 | 1.66E-07 | 6.42E-06 | 7.039111 |
| JADE1 | -0.38162 | 0.767576 | 8.270267 | -5.7538 | 1.66E-07 | 6.42E-06 | 7.039014 |
| LIAS | 0.431019 | 1.348186 | 6.3884 | 5.750015 | 1.68E-07 | 6.49E-06 | 7.023911 |
| DPP6 | -0.56463 | 0.676131 | 5.5832 | -5.7365 | 1.78E-07 | 6.84E-06 | 6.97005 |
| POT1 | 0.231694 | 1.174213 | 6.363067 | 5.72598 | 1.86E-07 | 7.11E-06 | 6.928131 |
| AVPI1 | -0.38606 | 0.765214 | 7.1492 | -5.72381 | 1.88E-07 | 7.14E-06 | 6.919478 |
| ZNF573 | 0.300382 | 1.231471 | 4.9868 | 5.715704 | 1.94E-07 | 7.35E-06 | 6.887228 |
| PCTP | 0.381972 | 1.303121 | 8.554 | 5.705021 | 2.03E-07 | 7.65E-06 | 6.844738 |
| RFC4 | 0.28404 | 1.2176 | 6.566133 | 5.701412 | 2.06E-07 | 7.71E-06 | 6.830389 |
| ATF1 | 0.272256 | 1.207695 | 6.788533 | 5.701085 | 2.06E-07 | 7.71E-06 | 6.829089 |
| RNF14 | 0.270742 | 1.206428 | 7.4236 | 5.696041 | 2.10E-07 | 7.83E-06 | 6.809044 |
| RAB22A | 0.201484 | 1.149881 | 7.545733 | 5.694063 | 2.12E-07 | 7.86E-06 | 6.801185 |
| MYO1C | -0.21195 | 0.86337 | 7.724133 | -5.69087 | 2.15E-07 | 7.93E-06 | 6.788483 |
| DTX4 | 0.427811 | 1.345191 | 8.059333 | 5.6853 | 2.20E-07 | 8.08E-06 | 6.766379 |
| PRPF18 | 0.277931 | 1.212455 | 7.207467 | 5.682194 | 2.23E-07 | 8.15E-06 | 6.754049 |
| MFAP1 | 0.232654 | 1.174994 | 8.262133 | 5.676847 | 2.28E-07 | 8.27E-06 | 6.732826 |
| RNF146 | 0.277369 | 1.211982 | 7.067467 | 5.676419 | 2.28E-07 | 8.27E-06 | 6.731128 |

| | | | | | | | |
|---|---|---|---|---|---|---|---|
| MSH3 | 0.336867 | 1.26301 | 6.413733 | 5.672743 | 2.32E-07 | 8.37E-06 | 6.716545 |
| MAP7 | 0.297436 | 1.228959 | 8.2824 | 5.668534 | 2.36E-07 | 8.47E-06 | 6.69985 |
| MYO6 | 0.250397 | 1.189535 | 8.275733 | 5.667531 | 2.37E-07 | 8.47E-06 | 6.695872 |
| MKL2 | 0.289078 | 1.221859 | 7.363733 | 5.664868 | 2.39E-07 | 8.53E-06 | 6.685312 |
| ALS2CL | -0.29723 | 0.813815 | 6.793333 | -5.66027 | 2.44E-07 | 8.66E-06 | 6.667102 |
| OSGEPL1 | 0.27533 | 1.210271 | 6.8232 | 5.653726 | 2.50E-07 | 8.86E-06 | 6.641154 |
| NDNF | -1.12597 | 0.458195 | 6.6568 | -5.63619 | 2.69E-07 | 9.48E-06 | 6.571711 |
| PEX3 | 0.269378 | 1.205288 | 8.341333 | 5.628601 | 2.78E-07 | 9.74E-06 | 6.541705 |
| SMIM10L1 | 0.26946 | 1.205357 | 7.6368 | 5.627319 | 2.79E-07 | 9.75E-06 | 6.536637 |
| STAM | 0.290997 | 1.223485 | 7.231867 | 5.624887 | 2.82E-07 | 9.81E-06 | 6.52702 |
| VPS4B | 0.272736 | 1.208097 | 7.703067 | 5.621082 | 2.86E-07 | 9.90E-06 | 6.511977 |
| PPP2R3C | 0.365817 | 1.288611 | 6.846667 | 5.620757 | 2.87E-07 | 9.90E-06 | 6.510694 |
| MAP3K4 | 0.259048 | 1.196689 | 7.4656 | 5.617315 | 2.91E-07 | 9.98E-06 | 6.497092 |
| MED8 | 0.168366 | 1.123785 | 6.964667 | 5.615782 | 2.93E-07 | 9.98E-06 | 6.491036 |
| NOL8 | 0.274865 | 1.209881 | 6.786933 | 5.615666 | 2.93E-07 | 9.98E-06 | 6.490576 |
| SHTN1 | 0.390832 | 1.311149 | 6.9296 | 5.608119 | 3.02E-07 | 1.03E-05 | 6.460766 |
| HARS2 | 0.223808 | 1.167812 | 7.4048 | 5.606854 | 3.04E-07 | 1.03E-05 | 6.455773 |
| RBM22 | 0.187579 | 1.138851 | 7.2984 | 5.604573 | 3.07E-07 | 1.03E-05 | 6.446766 |
| NSL1 | 0.237541 | 1.178982 | 7.027067 | 5.603724 | 3.08E-07 | 1.03E-05 | 6.443416 |
| VAMP2 | -0.19828 | 0.871592 | 6.243333 | -5.59781 | 3.15E-07 | 1.05E-05 | 6.420084 |
| PIM1 | -0.33652 | 0.791948 | 6.4964 | -5.59141 | 3.24E-07 | 1.08E-05 | 6.394836 |
| ECD | 0.187076 | 1.138454 | 7.391467 | 5.59015 | 3.25E-07 | 1.08E-05 | 6.389858 |
| HTR1E | -0.21795 | 0.859784 | 4.463333 | -5.58268 | 3.35E-07 | 1.11E-05 | 6.360427 |
| MFSD1 | 0.205 | 1.152686 | 9.484267 | 5.578909 | 3.41E-07 | 1.12E-05 | 6.34555 |
| GRAMD1C | 0.421792 | 1.33959 | 7.260267 | 5.572552 | 3.50E-07 | 1.15E-05 | 6.320507 |
| SORBS2 | 0.328253 | 1.255492 | 7.634533 | 5.570112 | 3.53E-07 | 1.15E-05 | 6.310899 |
| NAT8 | 0.379565 | 1.30095 | 13.0272 | 5.568982 | 3.55E-07 | 1.15E-05 | 6.306448 |
| BTG3 | -0.51594 | 0.699335 | 7.615067 | -5.56364 | 3.63E-07 | 1.17E-05 | 6.285424 |
| ITPKC | -0.27616 | 0.825785 | 6.178 | -5.55976 | 3.68E-07 | 1.19E-05 | 6.270135 |
| STK38 | 0.319483 | 1.247883 | 7.088533 | 5.55744 | 3.72E-07 | 1.19E-05 | 6.261025 |
| SPSB1 | -0.47908 | 0.717436 | 5.3228 | -5.55726 | 3.72E-07 | 1.19E-05 | 6.260298 |
| FLRT3 | 0.618096 | 1.534848 | 8.269867 | 5.556325 | 3.74E-07 | 1.19E-05 | 6.256638 |
| POP5 | 0.255352 | 1.193627 | 8.937867 | 5.556125 | 3.74E-07 | 1.19E-05 | 6.255853 |
| CPPED1 | 0.201694 | 1.150048 | 5.901467 | 5.551257 | 3.81E-07 | 1.21E-05 | 6.236707 |
| YTHDF3 | 0.30482 | 1.235265 | 7.873733 | 5.550291 | 3.83E-07 | 1.21E-05 | 6.232907 |
| GTF3C3 | 0.226199 | 1.169749 | 6.443333 | 5.548309 | 3.86E-07 | 1.21E-05 | 6.225117 |
| NXT1 | -0.30599 | 0.808887 | 7.301467 | -5.54441 | 3.92E-07 | 1.23E-05 | 6.209806 |
| KPNA4 | 0.24006 | 1.181042 | 6.015867 | 5.542735 | 3.95E-07 | 1.23E-05 | 6.203207 |
| PDHX | 0.28928 | 1.222031 | 7.805333 | 5.541468 | 3.97E-07 | 1.23E-05 | 6.198229 |
| PROS1 | 0.314033 | 1.243178 | 7.5236 | 5.537109 | 4.04E-07 | 1.25E-05 | 6.181105 |
| SLC25A13 | 0.327541 | 1.254873 | 7.511867 | 5.534748 | 4.08E-07 | 1.26E-05 | 6.171832 |
| COL4A3 | -0.31754 | 0.802436 | 5.112 | -5.53408 | 4.09E-07 | 1.26E-05 | 6.169209 |
| BBIP1 | 0.198996 | 1.147899 | 7.893467 | 5.532565 | 4.12E-07 | 1.26E-05 | 6.16326 |
| MTUS1 | 0.231927 | 1.174402 | 7.2212 | 5.531946 | 4.13E-07 | 1.26E-05 | 6.160831 |
| CAND1 | 0.244655 | 1.18481 | 7.3396 | 5.528317 | 4.19E-07 | 1.27E-05 | 6.146582 |
| FANCF | 0.250622 | 1.18972 | 6.419733 | 5.525447 | 4.24E-07 | 1.28E-05 | 6.135318 |
| CFDP1 | 0.291964 | 1.224306 | 9.795067 | 5.522329 | 4.29E-07 | 1.29E-05 | 6.123084 |

| | | | | | | | |
|---|---|---|---|---|---|---|---|
| ATG14 | 0.259108 | 1.196739 | 6.768667 | 5.521357 | 4.31E-07 | 1.29E-05 | 6.119272 |
| GADD45A | -0.56466 | 0.676117 | 10.62667 | -5.52107 | 4.31E-07 | 1.29E-05 | 6.118151 |
| ZC3H15 | 0.233778 | 1.17591 | 8.752133 | 5.514671 | 4.43E-07 | 1.32E-05 | 6.093047 |
| TLR3 | 0.36937 | 1.291789 | 6.185867 | 5.514397 | 4.43E-07 | 1.32E-05 | 6.091975 |
| AMOT | 0.322001 | 1.250064 | 10.07733 | 5.513903 | 4.44E-07 | 1.32E-05 | 6.090037 |
| ZNF271P | 0.282249 | 1.216089 | 6.098267 | 5.510741 | 4.50E-07 | 1.33E-05 | 6.07764 |
| ZNF331 | -0.61618 | 0.652397 | 6.778267 | -5.50893 | 4.53E-07 | 1.33E-05 | 6.070556 |
| SNX2 | 0.226439 | 1.169944 | 8.6656 | 5.507964 | 4.55E-07 | 1.33E-05 | 6.066757 |
| GTF2H1 | 0.288148 | 1.221072 | 7.0412 | 5.507528 | 4.56E-07 | 1.33E-05 | 6.065045 |
| AKAP9 | 0.281664 | 1.215596 | 7.745867 | 5.505102 | 4.61E-07 | 1.34E-05 | 6.055541 |
| ANKRD17 | 0.218711 | 1.163693 | 7.474133 | 5.504165 | 4.62E-07 | 1.34E-05 | 6.051869 |
| SYNPO | -0.3295 | 0.795813 | 6.429333 | -5.50256 | 4.65E-07 | 1.34E-05 | 6.045564 |
| SAP18 | 0.171079 | 1.125901 | 9.010933 | 5.502428 | 4.66E-07 | 1.34E-05 | 6.045064 |
| ANGEL2 | 0.272699 | 1.208065 | 6.1024 | 5.501863 | 4.67E-07 | 1.34E-05 | 6.042852 |
| CDC123 | 0.213066 | 1.159149 | 8.551733 | 5.501791 | 4.67E-07 | 1.34E-05 | 6.04257 |
| SCAF8 | 0.242556 | 1.183087 | 7.632267 | 5.500246 | 4.70E-07 | 1.34E-05 | 6.036517 |
| TNFAIP3 | -0.41765 | 0.748645 | 6.355467 | -5.49746 | 4.75E-07 | 1.35E-05 | 6.025587 |
| IARS2 | 0.253703 | 1.192264 | 9.398533 | 5.497261 | 4.76E-07 | 1.35E-05 | 6.024826 |
| CD58 | 0.298921 | 1.230224 | 7.0408 | 5.494245 | 4.81E-07 | 1.36E-05 | 6.013018 |
| SACM1L | 0.238516 | 1.179778 | 9.0544 | 5.494171 | 4.82E-07 | 1.36E-05 | 6.012728 |
| PPWD1 | 0.259805 | 1.197317 | 5.633067 | 5.492194 | 4.85E-07 | 1.36E-05 | 6.004989 |
| NBR1 | 0.262354 | 1.199434 | 8.7384 | 5.489684 | 4.90E-07 | 1.37E-05 | 5.995165 |
| PSPH | 0.408351 | 1.327168 | 5.5092 | 5.488046 | 4.94E-07 | 1.38E-05 | 5.988756 |
| CXCL2 | -1.07333 | 0.475221 | 4.971067 | -5.48102 | 5.08E-07 | 1.41E-05 | 5.961253 |
| DBR1 | 0.295735 | 1.22751 | 5.034133 | 5.479226 | 5.12E-07 | 1.42E-05 | 5.954254 |
| AIFM1 | 0.274543 | 1.209611 | 10.0392 | 5.478749 | 5.13E-07 | 1.42E-05 | 5.952388 |
| TACC1 | -0.40606 | 0.754679 | 8.381467 | -5.47444 | 5.22E-07 | 1.44E-05 | 5.935537 |
| PCOLCE2 | -0.98325 | 0.505838 | 5.7172 | -5.46929 | 5.33E-07 | 1.46E-05 | 5.915414 |
| CASP6 | 0.245802 | 1.185752 | 6.792 | 5.461715 | 5.50E-07 | 1.50E-05 | 5.885827 |
| KLK7 | -0.42133 | 0.746738 | 5.526 | -5.46036 | 5.53E-07 | 1.50E-05 | 5.880539 |
| LRRC40 | 0.311912 | 1.241351 | 7.713867 | 5.459937 | 5.54E-07 | 1.50E-05 | 5.878883 |
| TACSTD2 | -0.54406 | 0.685837 | 9.290933 | -5.45957 | 5.54E-07 | 1.50E-05 | 5.877467 |
| SUV420H1 | 0.304933 | 1.235361 | 7.045733 | 5.459278 | 5.55E-07 | 1.50E-05 | 5.876311 |
| LIPT1 | 0.345817 | 1.270871 | 6.918933 | 5.455836 | 5.63E-07 | 1.52E-05 | 5.862875 |
| UROD | 0.169813 | 1.124912 | 9.3624 | 5.44778 | 5.82E-07 | 1.56E-05 | 5.831439 |
| DDN | -0.41019 | 0.752526 | 7.238133 | -5.43824 | 6.05E-07 | 1.62E-05 | 5.794258 |
| CUL5 | 0.19479 | 1.144558 | 7.184667 | 5.435749 | 6.11E-07 | 1.63E-05 | 5.784533 |
| ZNF587B | 0.410742 | 1.329369 | 5.447733 | 5.435401 | 6.12E-07 | 1.63E-05 | 5.783178 |
| SMARCE1 | 0.221402 | 1.165866 | 8.468 | 5.434273 | 6.14E-07 | 1.63E-05 | 5.77878 |
| MYL9 | -0.43475 | 0.739824 | 6.717333 | -5.4276 | 6.31E-07 | 1.67E-05 | 5.752803 |
| OXSM | 0.198981 | 1.147887 | 7.7852 | 5.418746 | 6.54E-07 | 1.72E-05 | 5.718318 |
| THAP1 | 0.28572 | 1.219018 | 5.902 | 5.414983 | 6.64E-07 | 1.74E-05 | 5.703678 |
| ZNF226 | 0.233156 | 1.175403 | 5.409067 | 5.412134 | 6.72E-07 | 1.76E-05 | 5.692595 |
| EI24 | 0.188546 | 1.139614 | 7.6716 | 5.411879 | 6.73E-07 | 1.76E-05 | 5.691604 |
| ATP6V1C1 | 0.240217 | 1.181171 | 7.402667 | 5.404039 | 6.94E-07 | 1.80E-05 | 5.661118 |
| NES | -0.34257 | 0.788635 | 7.0108 | -5.40376 | 6.95E-07 | 1.80E-05 | 5.660033 |
| COPS4 | 0.171454 | 1.126193 | 8.8176 | 5.400958 | 7.03E-07 | 1.81E-05 | 5.649147 |

| | | | | | | | |
|---|---|---|---|---|---|---|---|
| JOSD1 | -0.33062 | 0.795193 | 7.399333 | -5.4002 | 7.05E-07 | 1.81E-05 | 5.646208 |
| CNN2 | -0.27415 | 0.826936 | 7.205733 | -5.40012 | 7.06E-07 | 1.81E-05 | 5.645905 |
| TPRKB | 0.290405 | 1.222983 | 8.185333 | 5.395526 | 7.19E-07 | 1.84E-05 | 5.628043 |
| KATNA1 | 0.259588 | 1.197137 | 7.2632 | 5.39283 | 7.27E-07 | 1.86E-05 | 5.617573 |
| KRBOX4 | 0.310525 | 1.240159 | 6.7592 | 5.391457 | 7.31E-07 | 1.86E-05 | 5.612239 |
| TRIM14 | 0.269865 | 1.205695 | 6.62 | 5.39074 | 7.33E-07 | 1.86E-05 | 5.609457 |
| VPS37B | -0.43588 | 0.73924 | 6.419067 | -5.37616 | 7.77E-07 | 1.97E-05 | 5.552898 |
| PTGER4 | -0.3742 | 0.771534 | 5.872267 | -5.37418 | 7.84E-07 | 1.98E-05 | 5.545191 |
| TNFRSF11I | 0.532436 | 1.44637 | 7.358267 | 5.371036 | 7.94E-07 | 2.00E-05 | 5.533013 |
| SMARCAL1 | 0.262481 | 1.19954 | 6.408667 | 5.36393 | 8.17E-07 | 2.05E-05 | 5.505475 |
| ABHD6 | 0.3553 | 1.279251 | 8.3376 | 5.361506 | 8.25E-07 | 2.06E-05 | 5.496085 |
| DCHS2 | -0.17839 | 0.88369 | 4.899067 | -5.35945 | 8.32E-07 | 2.07E-05 | 5.488131 |
| FAM206A | 0.21958 | 1.164395 | 7.3736 | 5.351583 | 8.58E-07 | 2.13E-05 | 5.457666 |
| SULT1C2 | 0.248118 | 1.187657 | 8.9992 | 5.349679 | 8.65E-07 | 2.14E-05 | 5.450297 |
| TRIB1 | -0.60308 | 0.658347 | 7.3192 | -5.34887 | 8.68E-07 | 2.15E-05 | 5.447155 |
| ERBB3 | 0.304445 | 1.234944 | 8.297067 | 5.348027 | 8.71E-07 | 2.15E-05 | 5.443906 |
| MGA | 0.330075 | 1.257079 | 5.838933 | 5.337003 | 9.10E-07 | 2.24E-05 | 5.401274 |
| AGA | 0.298298 | 1.229693 | 6.987733 | 5.325885 | 9.52E-07 | 2.33E-05 | 5.358319 |
| KIAA1033 | 0.245705 | 1.185672 | 7.858267 | 5.322151 | 9.67E-07 | 2.36E-05 | 5.343902 |
| UTP14C | 0.254063 | 1.192561 | 8.186933 | 5.321996 | 9.67E-07 | 2.36E-05 | 5.343306 |
| UBE2L3 | 0.213711 | 1.159667 | 7.9572 | 5.320331 | 9.74E-07 | 2.36E-05 | 5.336876 |
| DDAH1 | 0.176739 | 1.130326 | 11.1716 | 5.320026 | 9.75E-07 | 2.36E-05 | 5.335698 |
| DET1 | 0.218036 | 1.163149 | 5.872133 | 5.319045 | 9.79E-07 | 2.36E-05 | 5.331913 |
| TMEM184 | 0.210367 | 1.156983 | 7.853733 | 5.314962 | 9.95E-07 | 2.40E-05 | 5.316157 |
| SERPINI1 | 0.518568 | 1.432533 | 8.7416 | 5.31302 | 1.00E-06 | 2.41E-05 | 5.308666 |
| PREPL | 0.209625 | 1.156388 | 8.104533 | 5.312402 | 1.01E-06 | 2.41E-05 | 5.30628 |
| PLA1A | 0.45587 | 1.371609 | 6.9604 | 5.302666 | 1.05E-06 | 2.50E-05 | 5.268744 |
| RB1CC1 | 0.264123 | 1.200906 | 7.343867 | 5.295745 | 1.07E-06 | 2.56E-05 | 5.242077 |
| ATG5 | 0.202489 | 1.150682 | 6.8096 | 5.293813 | 1.08E-06 | 2.57E-05 | 5.234634 |
| NAB2 | -0.28384 | 0.821403 | 5.975467 | -5.29325 | 1.09E-06 | 2.57E-05 | 5.232472 |
| GNAI3 | 0.191732 | 1.142134 | 7.508267 | 5.292993 | 1.09E-06 | 2.57E-05 | 5.231478 |
| SERPINE1 | -0.7537 | 0.593079 | 5.568133 | -5.28475 | 1.12E-06 | 2.65E-05 | 5.199754 |
| F3 | -0.66269 | 0.631699 | 6.170933 | -5.28185 | 1.14E-06 | 2.67E-05 | 5.18859 |
| IL6R | -0.47857 | 0.71769 | 6.283867 | -5.27792 | 1.15E-06 | 2.70E-05 | 5.173456 |
| DROSHA | 0.202886 | 1.150999 | 7.088667 | 5.274861 | 1.17E-06 | 2.73E-05 | 5.161707 |
| TEFM | 0.334483 | 1.260925 | 7.339333 | 5.272216 | 1.18E-06 | 2.75E-05 | 5.151542 |
| PRDM14 | -0.23761 | 0.84815 | 5.705733 | -5.26865 | 1.20E-06 | 2.78E-05 | 5.137849 |
| SMC4 | 0.223021 | 1.167175 | 5.5108 | 5.259545 | 1.24E-06 | 2.88E-05 | 5.10286 |
| GIN1 | 0.245675 | 1.185647 | 6.291733 | 5.258336 | 1.25E-06 | 2.88E-05 | 5.098218 |
| GLCE | 0.281897 | 1.215792 | 5.554 | 5.254741 | 1.27E-06 | 2.92E-05 | 5.084418 |
| CSE1L | 0.208673 | 1.155625 | 7.7996 | 5.25328 | 1.27E-06 | 2.93E-05 | 5.078814 |
| B4GALT5 | 0.456522 | 1.372229 | 8.1 | 5.252276 | 1.28E-06 | 2.93E-05 | 5.074959 |
| YRDC | -0.2405 | 0.846451 | 7.100267 | -5.24987 | 1.29E-06 | 2.95E-05 | 5.065743 |
| BDKRB2 | -0.43798 | 0.738166 | 6.191733 | -5.24478 | 1.32E-06 | 3.00E-05 | 5.046188 |
| TRIP12 | 0.177804 | 1.13116 | 8.798533 | 5.243966 | 1.32E-06 | 3.01E-05 | 5.043085 |
| ZNF544 | 0.238793 | 1.180005 | 7.147333 | 5.242857 | 1.33E-06 | 3.01E-05 | 5.038833 |
| SNX13 | 0.174483 | 1.12856 | 6.987467 | 5.241263 | 1.34E-06 | 3.02E-05 | 5.032721 |

| | | | | | | | |
|---|---|---|---|---|---|---|---|
| HBB | 1.761552 | 3.390626 | 8.666133 | 5.240176 | 1.34E-06 | 3.03E-05 | 5.028554 |
| FAM46C | 0.272519 | 1.207915 | 6.3132 | 5.239331 | 1.35E-06 | 3.03E-05 | 5.025314 |
| CHGB | -0.60848 | 0.655888 | 5.6032 | -5.23888 | 1.35E-06 | 3.03E-05 | 5.023573 |
| PLK3 | -0.3213 | 0.800346 | 5.157067 | -5.22666 | 1.42E-06 | 3.17E-05 | 4.976769 |
| MIS18A | 0.188576 | 1.139638 | 5.998133 | 5.226393 | 1.42E-06 | 3.17E-05 | 4.975755 |
| UPF2 | 0.26003 | 1.197504 | 6.417067 | 5.225469 | 1.42E-06 | 3.17E-05 | 4.972219 |
| FRK | 0.309925 | 1.239643 | 5.078533 | 5.223607 | 1.43E-06 | 3.18E-05 | 4.965089 |
| APOE | 0.341259 | 1.266862 | 11.75587 | 5.221758 | 1.45E-06 | 3.19E-05 | 4.958012 |
| LARS2 | 0.267631 | 1.20383 | 7.178267 | 5.221756 | 1.45E-06 | 3.19E-05 | 4.958006 |
| C3orf14 | 0.306529 | 1.236729 | 6.596133 | 5.220911 | 1.45E-06 | 3.19E-05 | 4.954771 |
| PSMD1 | 0.147031 | 1.107289 | 8.6412 | 5.21736 | 1.47E-06 | 3.23E-05 | 4.941188 |
| ELL2 | -0.56476 | 0.676068 | 6.3188 | -5.21445 | 1.49E-06 | 3.26E-05 | 4.93007 |
| DPP8 | 0.235787 | 1.177549 | 8.339867 | 5.207146 | 1.53E-06 | 3.35E-05 | 4.902134 |
| TUG1 | 0.234333 | 1.176363 | 8.018 | 5.200593 | 1.57E-06 | 3.43E-05 | 4.877096 |
| ZNF106 | 0.276649 | 1.211378 | 7.920667 | 5.197407 | 1.59E-06 | 3.46E-05 | 4.864931 |
| USP11 | -0.21514 | 0.861461 | 7.893333 | -5.19571 | 1.60E-06 | 3.48E-05 | 4.858451 |
| SREBF2 | -0.19195 | 0.875422 | 6.341867 | -5.18814 | 1.65E-06 | 3.57E-05 | 4.829554 |
| ZNF468 | 0.269745 | 1.205595 | 5.403867 | 5.181497 | 1.70E-06 | 3.66E-05 | 4.804225 |
| CBR4 | 0.314693 | 1.243747 | 8.267333 | 5.180226 | 1.71E-06 | 3.67E-05 | 4.799379 |
| TRIM8 | 0.184865 | 1.136711 | 8.362133 | 5.177797 | 1.72E-06 | 3.70E-05 | 4.79012 |
| ATR | 0.213786 | 1.159727 | 5.828533 | 5.165976 | 1.81E-06 | 3.86E-05 | 4.745085 |
| CEP57 | 0.20982 | 1.156544 | 6.482 | 5.16306 | 1.83E-06 | 3.90E-05 | 4.733985 |
| SERPINE2 | 0.613621 | 1.530094 | 9.492267 | 5.162169 | 1.83E-06 | 3.90E-05 | 4.730594 |
| SPATA7 | 0.235382 | 1.177219 | 6.236667 | 5.158456 | 1.86E-06 | 3.94E-05 | 4.716465 |
| FRY | -0.34551 | 0.78703 | 4.990533 | -5.1567 | 1.87E-06 | 3.94E-05 | 4.709775 |
| NUP43 | 0.30422 | 1.234751 | 6.793067 | 5.156114 | 1.88E-06 | 3.94E-05 | 4.707556 |
| PMS1 | 0.238471 | 1.179741 | 6.6196 | 5.155902 | 1.88E-06 | 3.94E-05 | 4.706748 |
| NIF3L1 | 0.199768 | 1.148513 | 8.5392 | 5.155753 | 1.88E-06 | 3.94E-05 | 4.70618 |
| WDR37 | 0.218928 | 1.163868 | 7.004 | 5.155727 | 1.88E-06 | 3.94E-05 | 4.706083 |
| SPRY2 | -0.3033 | 0.810398 | 6.645333 | -5.15498 | 1.89E-06 | 3.95E-05 | 4.703241 |
| GATA6 | -0.39685 | 0.759514 | 4.171333 | -5.15181 | 1.91E-06 | 3.99E-05 | 4.691194 |
| NSF | 0.201274 | 1.149713 | 8.28 | 5.147019 | 1.95E-06 | 4.05E-05 | 4.672972 |
| GNS | 0.310735 | 1.240339 | 9.474933 | 5.136996 | 2.03E-06 | 4.21E-05 | 4.634897 |
| DYNC1LI1 | 0.210877 | 1.157392 | 7.3748 | 5.136178 | 2.03E-06 | 4.21E-05 | 4.63179 |
| TMEM140 | 0.456874 | 1.372565 | 8.114267 | 5.134813 | 2.04E-06 | 4.22E-05 | 4.626607 |
| UBE2B | 0.141597 | 1.103125 | 8.514533 | 5.132536 | 2.06E-06 | 4.25E-05 | 4.617963 |
| DBT | 0.308178 | 1.238143 | 6.905467 | 5.129651 | 2.09E-06 | 4.29E-05 | 4.607016 |
| MRS2 | 0.269048 | 1.205012 | 6.369467 | 5.128694 | 2.09E-06 | 4.30E-05 | 4.603383 |
| SPATA5L1 | 0.228718 | 1.171793 | 6.062133 | 5.125728 | 2.12E-06 | 4.34E-05 | 4.592133 |
| THSD4 | -0.22915 | 0.85314 | 5.072267 | -5.12452 | 2.13E-06 | 4.34E-05 | 4.587537 |
| CALML4 | 0.308868 | 1.238735 | 8.905733 | 5.124072 | 2.13E-06 | 4.34E-05 | 4.585851 |
| GRSF1 | 0.18012 | 1.132978 | 7.615733 | 5.12241 | 2.15E-06 | 4.36E-05 | 4.579548 |
| PEX12 | 0.193471 | 1.143511 | 7.3072 | 5.118334 | 2.18E-06 | 4.42E-05 | 4.564093 |
| AK2 | 0.239723 | 1.180766 | 7.259867 | 5.11347 | 2.22E-06 | 4.50E-05 | 4.545662 |
| CETN3 | 0.340577 | 1.266263 | 7.379733 | 5.110464 | 2.25E-06 | 4.54E-05 | 4.534276 |
| CETN2 | 0.168321 | 1.12375 | 9.839867 | 5.108816 | 2.26E-06 | 4.56E-05 | 4.528034 |
| EPHA2 | -0.4475 | 0.733314 | 5.876533 | -5.10791 | 2.27E-06 | 4.56E-05 | 4.524615 |

| | | | | | | | |
|---|---|---|---|---|---|---|---|
| ERBB2IP | 0.342504 | 1.267955 | 8.272 | 5.107544 | 2.28E-06 | 4.56E-05 | 4.523215 |
| XPNPEP1 | 0.191552 | 1.141991 | 8.239067 | 5.104913 | 2.30E-06 | 4.60E-05 | 4.513255 |
| PHF1 | -0.17756 | 0.884199 | 8.327867 | -5.10161 | 2.33E-06 | 4.65E-05 | 4.50075 |
| PDE6G | -0.18753 | 0.878106 | 6.6764 | -5.09532 | 2.39E-06 | 4.75E-05 | 4.476968 |
| ZNF83 | 0.238103 | 1.179441 | 5.607067 | 5.094737 | 2.39E-06 | 4.75E-05 | 4.474751 |
| FRYL | 0.208816 | 1.155739 | 6.408133 | 5.088753 | 2.45E-06 | 4.85E-05 | 4.452126 |
| DUSP2 | -0.30486 | 0.809522 | 5.2256 | -5.08747 | 2.46E-06 | 4.87E-05 | 4.447281 |
| DRG1 | 0.140735 | 1.102466 | 9.4892 | 5.083352 | 2.50E-06 | 4.94E-05 | 4.431713 |
| CDKN1C | -0.38169 | 0.767536 | 8.408933 | -5.08003 | 2.54E-06 | 4.99E-05 | 4.419186 |
| LSM5 | 0.198426 | 1.147446 | 5.819333 | 5.079574 | 2.54E-06 | 4.99E-05 | 4.417444 |
| PIK3C2A | 0.201027 | 1.149516 | 7.5836 | 5.078836 | 2.55E-06 | 4.99E-05 | 4.41466 |
| NEDD4 | 0.268853 | 1.20485 | 6.562 | 5.074381 | 2.59E-06 | 5.07E-05 | 4.397838 |
| KIDINS220 | 0.23066 | 1.173371 | 7.272667 | 5.072875 | 2.61E-06 | 5.08E-05 | 4.392153 |
| TNFAIP8 | 0.55949 | 1.473748 | 6.505467 | 5.070791 | 2.63E-06 | 5.11E-05 | 4.384289 |
| ERGIC2 | 0.257309 | 1.195247 | 7.716667 | 5.069009 | 2.65E-06 | 5.14E-05 | 4.377568 |
| FBXL4 | 0.223081 | 1.167224 | 5.743867 | 5.064704 | 2.70E-06 | 5.21E-05 | 4.361328 |
| BRCC3 | 0.315322 | 1.24429 | 6.054533 | 5.058789 | 2.76E-06 | 5.32E-05 | 4.339029 |
| NUP133 | 0.202624 | 1.150789 | 7.214 | 5.058672 | 2.76E-06 | 5.32E-05 | 4.338586 |
| GAS6 | -0.25191 | 0.839783 | 7.649333 | -5.05668 | 2.78E-06 | 5.35E-05 | 4.331063 |
| PFAS | 0.211439 | 1.157843 | 7.1948 | 5.055763 | 2.79E-06 | 5.35E-05 | 4.327625 |
| PDZRN3 | 0.348786 | 1.273488 | 5.905733 | 5.054259 | 2.81E-06 | 5.37E-05 | 4.321959 |
| RBBP8 | -0.39499 | 0.760493 | 9.0864 | -5.05063 | 2.85E-06 | 5.44E-05 | 4.308301 |
| ZNF146 | 0.216822 | 1.16217 | 7.122533 | 5.04965 | 2.86E-06 | 5.45E-05 | 4.304599 |
| RNF128 | 0.240697 | 1.181563 | 9.4572 | 5.048815 | 2.87E-06 | 5.45E-05 | 4.301455 |
| RIN2 | 0.313583 | 1.242791 | 9.5056 | 5.044628 | 2.92E-06 | 5.53E-05 | 4.285694 |
| DNAJC22 | 0.278688 | 1.213091 | 7.324933 | 5.043093 | 2.94E-06 | 5.54E-05 | 4.279919 |
| GGA2 | 0.166882 | 1.122629 | 7.326267 | 5.042892 | 2.94E-06 | 5.54E-05 | 4.27916 |
| KIFAP3 | 0.202789 | 1.150921 | 8.164933 | 5.042681 | 2.94E-06 | 5.54E-05 | 4.278369 |
| GEMIN6 | 0.206709 | 1.154053 | 7.916667 | 5.040137 | 2.97E-06 | 5.58E-05 | 4.268796 |
| CTR9 | 0.296042 | 1.227771 | 7.6336 | 5.036387 | 3.01E-06 | 5.65E-05 | 4.254692 |
| RNF34 | 0.214003 | 1.159902 | 7.4584 | 5.03334 | 3.05E-06 | 5.70E-05 | 4.243233 |
| E2F4 | -0.18374 | 0.880417 | 8.237867 | -5.03094 | 3.08E-06 | 5.74E-05 | 4.234211 |
| RAD23B | 0.164393 | 1.120694 | 8.684 | 5.030156 | 3.09E-06 | 5.74E-05 | 4.231265 |
| LINC00963 | -0.20445 | 0.867868 | 4.687467 | -5.03014 | 3.09E-06 | 5.74E-05 | 4.231198 |
| IMPACT | 0.212339 | 1.158565 | 5.3408 | 5.029043 | 3.10E-06 | 5.74E-05 | 4.227083 |
| SNAP23 | 0.257496 | 1.195402 | 6.76 | 5.028742 | 3.11E-06 | 5.74E-05 | 4.225949 |
| KLF4 | -0.46905 | 0.722441 | 5.4332 | -5.02681 | 3.13E-06 | 5.78E-05 | 4.218691 |
| SSX2IP | 0.23997 | 1.180968 | 6.996267 | 5.025028 | 3.15E-06 | 5.80E-05 | 4.211996 |
| ZNF84 | 0.228238 | 1.171404 | 5.5276 | 5.024149 | 3.16E-06 | 5.81E-05 | 4.208694 |
| GDI1 | -0.17196 | 0.887638 | 7.415467 | -5.02345 | 3.17E-06 | 5.81E-05 | 4.206075 |
| RHOBTB3 | -0.34648 | 0.786502 | 6.467333 | -5.02081 | 3.20E-06 | 5.86E-05 | 4.196146 |
| CINP | 0.250892 | 1.189943 | 7.108533 | 5.019823 | 3.22E-06 | 5.87E-05 | 4.192451 |
| AMPH | -0.22598 | 0.855013 | 4.866533 | -5.01932 | 3.22E-06 | 5.87E-05 | 4.190566 |
| EGR3 | -0.59053 | 0.664098 | 4.4284 | -5.01281 | 3.31E-06 | 6.01E-05 | 4.166114 |
| ATP10A | -0.27999 | 0.823595 | 5.965867 | -5.01146 | 3.32E-06 | 6.03E-05 | 4.161054 |
| TNNT2 | -0.41759 | 0.748676 | 5.788533 | -5.0101 | 3.34E-06 | 6.05E-05 | 4.155949 |
| MAEA | 0.188741 | 1.139768 | 8.699067 | 5.009643 | 3.35E-06 | 6.05E-05 | 4.154249 |

| | | | | | | | |
|---|---|---|---|---|---|---|---|
| R3HDM1 | 0.185052 | 1.136858 | 7.595467 | 5.004961 | 3.41E-06 | 6.13E-05 | 4.136692 |
| CCNC | 0.194295 | 1.144165 | 9.781867 | 5.004933 | 3.41E-06 | 6.13E-05 | 4.136587 |
| PTP4A1 | -0.25506 | 0.837952 | 7.743333 | -5.00343 | 3.43E-06 | 6.16E-05 | 4.130953 |
| PRELP | -0.18778 | 0.877955 | 6.51 | -5.00159 | 3.45E-06 | 6.19E-05 | 4.124041 |
| RPE | 0.181042 | 1.133702 | 7.084133 | 4.998692 | 3.49E-06 | 6.25E-05 | 4.113197 |
| ATOX1 | 0.212466 | 1.158667 | 10.56107 | 4.997834 | 3.51E-06 | 6.25E-05 | 4.109983 |
| CFAP45 | -0.21715 | 0.860262 | 6.3256 | -4.99537 | 3.54E-06 | 6.30E-05 | 4.10076 |
| CCL4 | -0.65236 | 0.636238 | 6.646667 | -4.9897 | 3.62E-06 | 6.43E-05 | 4.07954 |
| PLCE1 | -0.36251 | 0.777809 | 5.166133 | -4.98517 | 3.68E-06 | 6.53E-05 | 4.062555 |
| GLS | 0.192736 | 1.142929 | 10.49213 | 4.984402 | 3.70E-06 | 6.54E-05 | 4.059699 |
| ACTR6 | 0.28066 | 1.21475 | 6.962 | 4.983214 | 3.71E-06 | 6.55E-05 | 4.055255 |
| IL1RAPL2 | -0.21006 | 0.864501 | 5.085733 | -4.98054 | 3.75E-06 | 6.61E-05 | 4.045243 |
| MXRA8 | -0.25551 | 0.837691 | 7.305333 | -4.97909 | 3.77E-06 | 6.63E-05 | 4.039822 |
| PDCD10 | 0.193403 | 1.143458 | 9.8 | 4.97119 | 3.89E-06 | 6.82E-05 | 4.010303 |
| NME5 | 0.265142 | 1.201755 | 8.726 | 4.970799 | 3.90E-06 | 6.82E-05 | 4.008843 |
| KIAA0040 | -0.19782 | 0.871868 | 5.715467 | -4.96697 | 3.96E-06 | 6.91E-05 | 3.994548 |
| PLSCR4 | 0.292609 | 1.224853 | 8.310533 | 4.965287 | 3.98E-06 | 6.94E-05 | 3.988256 |
| SMAP1 | 0.248186 | 1.187713 | 8.0864 | 4.960778 | 4.05E-06 | 7.05E-05 | 3.971424 |
| KLF9 | -0.44481 | 0.73468 | 7.086267 | -4.95656 | 4.12E-06 | 7.15E-05 | 3.955674 |
| RPA1 | 0.21512 | 1.1608 | 7.324267 | 4.952244 | 4.19E-06 | 7.26E-05 | 3.939587 |
| SEMA3G | -0.43278 | 0.740832 | 5.650267 | -4.94287 | 4.35E-06 | 7.51E-05 | 3.904643 |
| FERMT2 | -0.25508 | 0.837939 | 7.820933 | -4.94143 | 4.37E-06 | 7.54E-05 | 3.899299 |
| ARHGEF17 | -0.21442 | 0.861895 | 7.124267 | -4.93965 | 4.40E-06 | 7.58E-05 | 3.892636 |
| METTL18 | 0.263411 | 1.200313 | 5.6912 | 4.93741 | 4.44E-06 | 7.63E-05 | 3.884313 |
| KHK | 0.27533 | 1.210271 | 10.1432 | 4.936224 | 4.46E-06 | 7.65E-05 | 3.879897 |
| USP47 | 0.202271 | 1.150508 | 7.739733 | 4.934473 | 4.49E-06 | 7.69E-05 | 3.873382 |
| SLC20A1 | -0.45855 | 0.727716 | 7.229867 | -4.93322 | 4.51E-06 | 7.71E-05 | 3.868724 |
| FAM8A1 | 0.232151 | 1.174585 | 9.0552 | 4.92836 | 4.60E-06 | 7.84E-05 | 3.850633 |
| RIT1 | 0.439475 | 1.356111 | 6.7208 | 4.926629 | 4.63E-06 | 7.88E-05 | 3.844194 |
| CSRP1 | -0.32164 | 0.800159 | 8.911067 | -4.92133 | 4.73E-06 | 8.02E-05 | 3.824481 |
| UBR2 | 0.173921 | 1.12812 | 7.227467 | 4.918183 | 4.78E-06 | 8.08E-05 | 3.812799 |
| XPOT | 0.235742 | 1.177512 | 7.995067 | 4.918165 | 4.79E-06 | 8.08E-05 | 3.812732 |
| RRAGC | 0.226034 | 1.169616 | 8.6224 | 4.917924 | 4.79E-06 | 8.08E-05 | 3.811837 |
| GGCT | 0.222241 | 1.166545 | 9.160933 | 4.915575 | 4.83E-06 | 8.11E-05 | 3.803108 |
| ZNF277 | 0.283711 | 1.217322 | 6.994267 | 4.915334 | 4.84E-06 | 8.11E-05 | 3.802215 |
| DNAJC16 | 0.267976 | 1.204117 | 7.9084 | 4.915089 | 4.84E-06 | 8.11E-05 | 3.801305 |
| MPP3 | -0.24049 | 0.846455 | 5.6644 | -4.91472 | 4.85E-06 | 8.11E-05 | 3.799917 |
| TIMP3 | -0.34644 | 0.786523 | 7.478 | -4.91443 | 4.86E-06 | 8.11E-05 | 3.798853 |
| SKIV2L2 | 0.195075 | 1.144784 | 7.851733 | 4.912485 | 4.89E-06 | 8.16E-05 | 3.791633 |
| EDN1 | -0.35689 | 0.780847 | 5.906133 | -4.91097 | 4.92E-06 | 8.19E-05 | 3.786002 |
| CAPN7 | 0.253951 | 1.192468 | 7.024933 | 4.909441 | 4.95E-06 | 8.22E-05 | 3.780331 |
| FABP3 | 0.442556 | 1.35901 | 8.7696 | 4.907456 | 4.99E-06 | 8.26E-05 | 3.772961 |
| OSBPL9 | 0.237091 | 1.178614 | 9.559067 | 4.907289 | 4.99E-06 | 8.26E-05 | 3.772344 |
| PSMD14 | 0.200172 | 1.148836 | 9.7924 | 4.906397 | 5.01E-06 | 8.27E-05 | 3.769033 |
| PPFIBP2 | 0.294798 | 1.226713 | 7.737467 | 4.90512 | 5.03E-06 | 8.30E-05 | 3.764293 |
| UBA6 | 0.197804 | 1.146951 | 5.426267 | 4.901614 | 5.10E-06 | 8.38E-05 | 3.751286 |
| TPMT | 0.217249 | 1.162515 | 8.088133 | 4.901589 | 5.10E-06 | 8.38E-05 | 3.751193 |

| | | | | | | | |
|---|---|---|---|---|---|---|---|
| ZNF330 | 0.181222 | 1.133844 | 6.963333 | 4.900366 | 5.13E-06 | 8.40E-05 | 3.746658 |
| USP16 | 0.177729 | 1.131102 | 7.6172 | 4.899583 | 5.14E-06 | 8.41E-05 | 3.743751 |
| CDKN1B | 0.353988 | 1.278089 | 8.084267 | 4.898037 | 5.17E-06 | 8.44E-05 | 3.738019 |
| CRISPLD2 | -0.5466 | 0.68463 | 7.609733 | -4.89714 | 5.19E-06 | 8.46E-05 | 3.734698 |
| GPR183 | -0.24169 | 0.845756 | 4.3372 | -4.89464 | 5.24E-06 | 8.52E-05 | 3.725434 |
| GPR137B | 0.277549 | 1.212134 | 9.926667 | 4.890814 | 5.32E-06 | 8.64E-05 | 3.711249 |
| C1orf21 | -0.22028 | 0.858396 | 5.407867 | -4.88913 | 5.36E-06 | 8.68E-05 | 3.704996 |
| ACP2 | 0.208546 | 1.155523 | 8.699333 | 4.887501 | 5.39E-06 | 8.71E-05 | 3.698974 |
| ITPR2 | 0.173021 | 1.127417 | 5.741467 | 4.884723 | 5.45E-06 | 8.79E-05 | 3.688687 |
| ZNF185 | -0.2145 | 0.861846 | 6.5788 | -4.87876 | 5.58E-06 | 8.98E-05 | 3.666617 |
| CTSA | 0.233201 | 1.17544 | 10.99387 | 4.877243 | 5.61E-06 | 9.02E-05 | 3.661003 |
| LDAH | 0.217496 | 1.162714 | 6.454533 | 4.876134 | 5.63E-06 | 9.03E-05 | 3.6569 |
| IL4R | -0.25926 | 0.835518 | 6.318667 | -4.87594 | 5.64E-06 | 9.03E-05 | 3.656165 |
| CPT2 | 0.23449 | 1.176491 | 7.9948 | 4.873426 | 5.69E-06 | 9.08E-05 | 3.646884 |
| TFB2M | 0.295112 | 1.226981 | 8.191067 | 4.873338 | 5.69E-06 | 9.08E-05 | 3.646556 |
| PQLC1 | -0.44762 | 0.733253 | 9.1604 | -4.87183 | 5.73E-06 | 9.12E-05 | 3.64097 |
| TRPM6 | -0.26107 | 0.834468 | 5.1284 | -4.87041 | 5.76E-06 | 9.15E-05 | 3.635723 |
| WDR11 | 0.301177 | 1.232149 | 6.984933 | 4.869368 | 5.78E-06 | 9.17E-05 | 3.631881 |
| ICE1 | 0.240142 | 1.181109 | 7.521333 | 4.866995 | 5.84E-06 | 9.24E-05 | 3.623109 |
| VDAC3 | 0.186394 | 1.137916 | 10.40533 | 4.863531 | 5.92E-06 | 9.33E-05 | 3.610308 |
| RBM7 | 0.245607 | 1.185592 | 7.284533 | 4.863406 | 5.92E-06 | 9.33E-05 | 3.609849 |
| SMARCA5 | 0.190352 | 1.141042 | 6.267733 | 4.85577 | 6.10E-06 | 9.57E-05 | 3.58165 |
| HIST1H2BI | 0.364708 | 1.287621 | 9.644933 | 4.855619 | 6.10E-06 | 9.57E-05 | 3.581094 |
| UEVLD | 0.195367 | 1.145016 | 5.942933 | 4.855488 | 6.10E-06 | 9.57E-05 | 3.580607 |
| PPP1R3C | 0.482841 | 1.397493 | 7.092133 | 4.854294 | 6.13E-06 | 9.60E-05 | 3.5762 |
| DENND4B | -0.18374 | 0.880417 | 6.837867 | -4.85326 | 6.15E-06 | 9.62E-05 | 3.572373 |
| BACE2 | -0.41673 | 0.74912 | 7.079733 | -4.85271 | 6.17E-06 | 9.62E-05 | 3.570363 |
| SYNRG | 0.186792 | 1.13823 | 7.6644 | 4.849612 | 6.24E-06 | 9.72E-05 | 3.558926 |
| TNFSF8 | -0.17633 | 0.884953 | 5.065733 | -4.84608 | 6.33E-06 | 9.83E-05 | 3.545887 |
| RUFY1 | 0.173021 | 1.127417 | 8.181467 | 4.843206 | 6.40E-06 | 9.93E-05 | 3.535305 |
| CCDC90B | 0.19446 | 1.144296 | 7.3428 | 4.842662 | 6.41E-06 | 9.93E-05 | 3.533297 |

Table S1 Plum vs. Control

| Gene | logFC | UnlogFC | AveExpr | t | P.Value | adj.P.Val | B |
|---|---|---|---|---|---|---|---|
| COL4A2 | 1.08828 | 2.126204 | 7.810769 | 12.60573 | 7.54E-22 | 3.22E-18 | 39.1825 |
| COL4A1 | 1.136454 | 2.1984 | 8.462418 | 12.60372 | 7.61E-22 | 3.22E-18 | 39.17328 |
| LPCAT1 | 0.908783 | 1.877461 | 6.704615 | 12.42445 | 1.77E-21 | 4.98E-18 | 38.34874 |
| TMSB10 | 1.18343 | 2.271161 | 11.59956 | 12.26851 | 3.68E-21 | 7.78E-18 | 37.62882 |
| TNFAIP8 | 1.249314 | 2.377284 | 6.906923 | 12.21801 | 4.67E-21 | 7.90E-18 | 37.39518 |
| WFDC2 | 2.002077 | 4.005764 | 9.673516 | 12.13636 | 6.88E-21 | 9.69E-18 | 37.01683 |
| HEXB | 0.793406 | 1.733161 | 10.2756 | 12.04328 | 1.07E-20 | 1.29E-17 | 36.58472 |
| LAPTM4B | 0.582203 | 1.497134 | 9.787033 | 11.87544 | 2.37E-20 | 2.51E-17 | 35.80352 |
| CASP6 | 0.480821 | 1.395538 | 6.934725 | 11.81967 | 3.09E-20 | 2.91E-17 | 35.54335 |
| MYOF | 0.853048 | 1.806314 | 7.349011 | 11.79576 | 3.47E-20 | 2.93E-17 | 35.43175 |
| S100A11 | 1.026126 | 2.036548 | 9.253077 | 11.75861 | 4.14E-20 | 3.18E-17 | 35.2582 |
| PLOD2 | 0.659135 | 1.579136 | 7.217033 | 11.71166 | 5.18E-20 | 3.65E-17 | 35.03874 |
| TXNDC9 | 0.487739 | 1.402246 | 8.839451 | 11.60129 | 8.77E-20 | 5.70E-17 | 34.52201 |
| YWHAH | 0.844647 | 1.795826 | 7.506813 | 11.43275 | 1.96E-19 | 1.19E-16 | 33.73102 |
| GALNT1 | 0.582778 | 1.49773 | 7.493187 | 11.35318 | 2.88E-19 | 1.61E-16 | 33.35677 |
| MAPT | -0.68789 | 0.620762 | 7.414835 | -11.341 | 3.05E-19 | 1.61E-16 | 33.29965 |
| SERPINE2 | 1.155222 | 2.227186 | 9.826264 | 11.32055 | 3.37E-19 | 1.67E-16 | 33.20315 |
| PDIA6 | 0.515198 | 1.42919 | 9.878681 | 11.25638 | 4.58E-19 | 2.11E-16 | 32.90083 |
| PPIC | 0.582019 | 1.496943 | 8.518681 | 11.24936 | 4.74E-19 | 2.11E-16 | 32.8677 |
| PROM1 | 1.731454 | 3.320623 | 8.111648 | 11.19072 | 6.28E-19 | 2.65E-16 | 32.59112 |
| ANXA2 | 1.074874 | 2.106539 | 10.59088 | 11.16691 | 7.04E-19 | 2.84E-16 | 32.47878 |
| ARPC2 | 0.394599 | 1.314577 | 9.902088 | 11.06853 | 1.13E-18 | 4.35E-16 | 32.01405 |
| GALNT7 | 0.930527 | 1.905972 | 7.043846 | 10.93094 | 2.20E-18 | 8.07E-16 | 31.36299 |
| NR0B2 | -0.72054 | 0.606872 | 6.37956 | -10.8591 | 3.11E-18 | 1.09E-15 | 31.02231 |
| RAB11A | 0.32915 | 1.256273 | 8.774505 | 10.78483 | 4.45E-18 | 1.50E-15 | 30.67026 |
| CMTM6 | 0.55129 | 1.465395 | 9.130659 | 10.77642 | 4.63E-18 | 1.51E-15 | 30.63033 |
| PPIB | 0.516275 | 1.430258 | 10.29769 | 10.76234 | 4.96E-18 | 1.52E-15 | 30.56352 |
| YWHAB | 0.319773 | 1.248134 | 10.04791 | 10.75963 | 5.03E-18 | 1.52E-15 | 30.55067 |
| NMI | 0.866473 | 1.823201 | 7.11978 | 10.75246 | 5.20E-18 | 1.52E-15 | 30.51664 |
| VIM | 1.181101 | 2.267498 | 10.73363 | 10.72857 | 5.84E-18 | 1.65E-15 | 30.40319 |
| QPCT | 1.423758 | 2.682835 | 5.383187 | 10.69301 | 6.94E-18 | 1.89E-15 | 30.23427 |
| LTF | 3.318444 | 9.975882 | 8.430989 | 10.67257 | 7.66E-18 | 2.02E-15 | 30.13719 |
| PLSCR1 | 0.876614 | 1.83606 | 8.821099 | 10.62418 | 9.69E-18 | 2.48E-15 | 29.90718 |
| PHF1 | -0.30941 | 0.806971 | 8.243516 | -10.5846 | 1.17E-17 | 2.92E-15 | 29.71906 |
| TPM1 | 0.806502 | 1.748966 | 8.891429 | 10.52197 | 1.59E-17 | 3.84E-15 | 29.42094 |
| PDIA4 | 0.517493 | 1.431465 | 7.593077 | 10.51067 | 1.68E-17 | 3.94E-15 | 29.36715 |
| PSMB8 | 0.957667 | 1.942166 | 7.448571 | 10.50406 | 1.73E-17 | 3.96E-15 | 29.33567 |
| LAMC1 | 0.450932 | 1.366923 | 7.944945 | 10.44027 | 2.36E-17 | 5.26E-15 | 29.03191 |
| CFB | 1.547126 | 2.922343 | 7.645714 | 10.41763 | 2.64E-17 | 5.72E-15 | 28.92408 |
| HLA-B | 0.898256 | 1.863812 | 11.34571 | 10.4022 | 2.84E-17 | 6.01E-15 | 28.85054 |
| MMP7 | 2.002652 | 4.00736 | 9.53967 | 10.35906 | 3.51E-17 | 7.23E-15 | 28.64493 |
| ARFIP1 | 0.376145 | 1.297869 | 6.762527 | 10.31619 | 4.32E-17 | 8.69E-15 | 28.44054 |
| FARP1 | 0.450662 | 1.366667 | 8.792637 | 10.28645 | 4.99E-17 | 9.81E-15 | 28.29869 |
| S100A10 | 0.761976 | 1.695812 | 11.02571 | 10.23226 | 6.49E-17 | 1.25E-14 | 28.04014 |
| APOH | -1.65368 | 0.317828 | 8.196593 | -10.2054 | 7.40E-17 | 1.39E-14 | 27.91218 |

| | | | | | | | |
|---|---|---|---|---|---|---|---|
| NEU1 | 0.527126 | 1.441055 | 8.731319 | 10.19252 | 7.88E-17 | 1.45E-14 | 27.85048 |
| ACTN1 | 0.939246 | 1.917526 | 7.110549 | 10.17285 | 8.67E-17 | 1.56E-14 | 27.75657 |
| ALDH18A1 | 0.565333 | 1.479729 | 7.91956 | 10.11667 | 1.14E-16 | 2.01E-14 | 27.4883 |
| IL32 | 1.5747 | 2.978736 | 8.00022 | 10.09868 | 1.24E-16 | 2.14E-14 | 27.40236 |
| PLP2 | 0.878894 | 1.838965 | 7.374835 | 10.09427 | 1.27E-16 | 2.15E-14 | 27.38131 |
| HRG | -1.13659 | 0.454832 | 8.251209 | -10.0774 | 1.38E-16 | 2.29E-14 | 27.30092 |
| FKBP3 | 0.47587 | 1.390756 | 7.879451 | 10.07326 | 1.41E-16 | 2.29E-14 | 27.28094 |
| VWA5A | 0.411874 | 1.330413 | 7.098022 | 10.04402 | 1.62E-16 | 2.59E-14 | 27.14124 |
| SHTN1 | 0.667522 | 1.588342 | 7.108571 | 10.01838 | 1.84E-16 | 2.88E-14 | 27.0187 |
| WWC1 | 0.464546 | 1.379883 | 8.014286 | 10.01235 | 1.89E-16 | 2.91E-14 | 26.98988 |
| ESYT1 | 0.608744 | 1.524931 | 7.964505 | 9.99276 | 2.08E-16 | 3.14E-14 | 26.89624 |
| CLIC1 | 0.599396 | 1.515082 | 10.12923 | 9.963284 | 2.40E-16 | 3.56E-14 | 26.75534 |
| P4HB | 0.44813 | 1.364271 | 9.887473 | 9.956426 | 2.49E-16 | 3.62E-14 | 26.72256 |
| TIMP1 | 1.592691 | 3.016114 | 8.518681 | 9.949645 | 2.57E-16 | 3.68E-14 | 26.69014 |
| S100A13 | 0.848048 | 1.800064 | 9.141538 | 9.94205 | 2.67E-16 | 3.76E-14 | 26.65383 |
| GNAI3 | 0.373425 | 1.295425 | 7.618791 | 9.936932 | 2.73E-16 | 3.79E-14 | 26.62936 |
| SMARCE1 | 0.424053 | 1.341692 | 8.592088 | 9.926727 | 2.87E-16 | 3.92E-14 | 26.58056 |
| JAK1 | 0.54499 | 1.459011 | 8.664066 | 9.906654 | 3.17E-16 | 4.25E-14 | 26.48459 |
| CD63 | 0.334986 | 1.261365 | 11.66 | 9.891579 | 3.41E-16 | 4.50E-14 | 26.4125 |
| DCK | 0.580836 | 1.495715 | 5.384835 | 9.887118 | 3.48E-16 | 4.53E-14 | 26.39116 |
| FTCD | -1.03689 | 0.487376 | 8.777033 | -9.87895 | 3.62E-16 | 4.64E-14 | 26.35212 |
| CBX6 | 0.405295 | 1.324359 | 8.158681 | 9.872352 | 3.74E-16 | 4.72E-14 | 26.32055 |
| PON2 | 0.784758 | 1.722804 | 8.552198 | 9.857227 | 4.03E-16 | 5.01E-14 | 26.24821 |
| SERPING1 | 1.126372 | 2.183091 | 8.303736 | 9.83807 | 4.42E-16 | 5.42E-14 | 26.15658 |
| CLU | 1.172217 | 2.253578 | 8.949451 | 9.824115 | 4.73E-16 | 5.72E-14 | 26.08983 |
| TUBA1A | 1.124686 | 2.180541 | 9.897033 | 9.807418 | 5.14E-16 | 6.12E-14 | 26.00995 |
| NAGK | 0.387536 | 1.308157 | 8.780769 | 9.774912 | 6.02E-16 | 7.07E-14 | 25.85444 |
| RPA1 | 0.382913 | 1.303972 | 7.43044 | 9.765957 | 6.29E-16 | 7.28E-14 | 25.8116 |
| MANSC1 | 0.619995 | 1.53687 | 5.646374 | 9.733756 | 7.36E-16 | 8.40E-14 | 25.65753 |
| RARS | 0.493444 | 1.407802 | 8.899011 | 9.727985 | 7.56E-16 | 8.53E-14 | 25.62991 |
| HN1 | 0.863812 | 1.81984 | 7.608681 | 9.678824 | 9.61E-16 | 1.07E-13 | 25.39466 |
| PIGG | 0.40529 | 1.324355 | 7.078462 | 9.654627 | 1.08E-15 | 1.19E-13 | 25.27886 |
| HOMER3 | 0.601473 | 1.517265 | 7.103736 | 9.643305 | 1.14E-15 | 1.24E-13 | 25.22467 |
| DNAJC10 | 0.41057 | 1.329211 | 5.968681 | 9.636397 | 1.18E-15 | 1.27E-13 | 25.19161 |
| ZNF22 | 0.347377 | 1.272245 | 7.713736 | 9.609606 | 1.35E-15 | 1.42E-13 | 25.06339 |
| LIPA | 0.634232 | 1.552111 | 9.344066 | 9.607265 | 1.36E-15 | 1.42E-13 | 25.05218 |
| NME1 | 0.530275 | 1.444205 | 9.264615 | 9.595744 | 1.44E-15 | 1.49E-13 | 24.99704 |
| DHX15 | 0.321362 | 1.24951 | 9.46022 | 9.586042 | 1.51E-15 | 1.54E-13 | 24.95061 |
| ANKRD6 | 0.36013 | 1.283542 | 5.473956 | 9.574126 | 1.60E-15 | 1.61E-13 | 24.89357 |
| CDH3 | 0.687894 | 1.61093 | 6.705385 | 9.555727 | 1.75E-15 | 1.74E-13 | 24.80551 |
| BAIAP2 | -0.52622 | 0.694371 | 5.75978 | -9.5477 | 1.82E-15 | 1.79E-13 | 24.7671 |
| ALS2CL | -0.42893 | 0.742814 | 6.696154 | -9.54207 | 1.87E-15 | 1.82E-13 | 24.74015 |
| C2CD2 | 0.485502 | 1.400073 | 6.647692 | 9.539921 | 1.89E-15 | 1.82E-13 | 24.72985 |
| ZZZ3 | 0.534314 | 1.448253 | 7.188352 | 9.532539 | 1.96E-15 | 1.86E-13 | 24.69451 |
| ADAM9 | 0.657739 | 1.577608 | 8.753516 | 9.52569 | 2.03E-15 | 1.91E-13 | 24.66173 |
| UBE2E1 | 0.459618 | 1.375178 | 9.42011 | 9.522085 | 2.06E-15 | 1.92E-13 | 24.64447 |
| PTTG1IP | 0.356372 | 1.280202 | 11.01297 | 9.513022 | 2.16E-15 | 1.98E-13 | 24.60109 |

| | | | | | | | |
|---|---|---|---|---|---|---|---|
| ATP10D | 0.631285 | 1.548944 | 8 | 9.497839 | 2.32E-15 | 2.08E-13 | 24.52842 |
| C1GALT1C | 0.427478 | 1.344881 | 7.677912 | 9.497219 | 2.33E-15 | 2.08E-13 | 24.52545 |
| SDCBP | 0.480459 | 1.395187 | 10.45824 | 9.496585 | 2.34E-15 | 2.08E-13 | 24.52241 |
| AMMECR1 | 0.459614 | 1.375173 | 5.77989 | 9.481649 | 2.51E-15 | 2.21E-13 | 24.45092 |
| B2M | 0.414449 | 1.33279 | 12.61516 | 9.476061 | 2.58E-15 | 2.25E-13 | 24.42417 |
| APMAP | 0.339024 | 1.264901 | 8.683736 | 9.46434 | 2.74E-15 | 2.36E-13 | 24.36807 |
| CD47 | 0.418802 | 1.336817 | 7.133187 | 9.460532 | 2.79E-15 | 2.37E-13 | 24.34984 |
| GCNT3 | 0.987415 | 1.98263 | 6.211978 | 9.459412 | 2.80E-15 | 2.37E-13 | 24.34448 |
| TSPAN13 | 0.649676 | 1.568816 | 7.336703 | 9.453689 | 2.88E-15 | 2.41E-13 | 24.31709 |
| XRCC5 | 0.347266 | 1.272147 | 9.568681 | 9.444788 | 3.01E-15 | 2.49E-13 | 24.27448 |
| CKLF | 1.03557 | 2.049923 | 6.742747 | 9.439864 | 3.08E-15 | 2.53E-13 | 24.25091 |
| PJA1 | 0.39558 | 1.315471 | 8.076703 | 9.401791 | 3.71E-15 | 3.02E-13 | 24.06867 |
| PTPRK | 0.398604 | 1.318232 | 9.164286 | 9.384317 | 4.04E-15 | 3.25E-13 | 23.98503 |
| DEK | 0.381314 | 1.302528 | 9.427692 | 9.378898 | 4.15E-15 | 3.29E-13 | 23.95909 |
| GMDS | 0.352063 | 1.276384 | 6.106923 | 9.378334 | 4.16E-15 | 3.29E-13 | 23.95639 |
| PRKACB | 0.464478 | 1.379818 | 7.631209 | 9.344103 | 4.92E-15 | 3.85E-13 | 23.79255 |
| XPNPEP1 | 0.354556 | 1.278592 | 8.34033 | 9.327602 | 5.33E-15 | 4.13E-13 | 23.71358 |
| SPON2 | 1.20171 | 2.300122 | 7.441209 | 9.323716 | 5.43E-15 | 4.17E-13 | 23.69498 |
| PIGR | 0.760077 | 1.693581 | 8.189341 | 9.315448 | 5.66E-15 | 4.31E-13 | 23.65541 |
| VAMP2 | -0.31311 | 0.804904 | 6.165165 | -9.3111 | 5.78E-15 | 4.34E-13 | 23.63462 |
| YARS2 | 0.368444 | 1.29096 | 6.922198 | 9.310104 | 5.81E-15 | 4.34E-13 | 23.62983 |
| CDH6 | 1.062227 | 2.088152 | 7.115495 | 9.290199 | 6.40E-15 | 4.71E-13 | 23.53457 |
| ARHGEF40 | 0.411145 | 1.329741 | 6.484835 | 9.289741 | 6.41E-15 | 4.71E-13 | 23.53238 |
| TGFBR3 | -0.7647 | 0.588578 | 8.430549 | -9.26449 | 7.25E-15 | 5.27E-13 | 23.41153 |
| CSTB | 0.727696 | 1.655992 | 10.99615 | 9.262994 | 7.30E-15 | 5.27E-13 | 23.40438 |
| C1RL | 0.383469 | 1.304474 | 4.865714 | 9.261703 | 7.35E-15 | 5.27E-13 | 23.3982 |
| ADAM10 | 0.478715 | 1.393502 | 7.258901 | 9.240096 | 8.17E-15 | 5.78E-13 | 23.29481 |
| H2AFY | 0.344676 | 1.269866 | 8.510879 | 9.239198 | 8.20E-15 | 5.78E-13 | 23.29051 |
| B4GALT5 | 0.755633 | 1.688372 | 8.297143 | 9.226796 | 8.71E-15 | 6.09E-13 | 23.23117 |
| HSP90B1 | 0.600367 | 1.516102 | 9.833407 | 9.221922 | 8.92E-15 | 6.18E-13 | 23.20785 |
| MGAT4B | 0.589449 | 1.504672 | 7.836703 | 9.209175 | 9.50E-15 | 6.53E-13 | 23.14685 |
| ANXA4 | 0.509092 | 1.423154 | 10.69088 | 9.2075 | 9.57E-15 | 6.53E-13 | 23.13884 |
| RAB32 | 0.429947 | 1.347184 | 8.09022 | 9.193815 | 1.02E-14 | 6.92E-13 | 23.07336 |
| GYPA | -0.31688 | 0.802802 | 4.503516 | -9.18507 | 1.07E-14 | 7.17E-13 | 23.03153 |
| RUFY1 | 0.281657 | 1.21559 | 8.253846 | 9.157594 | 1.22E-14 | 8.13E-13 | 22.90008 |
| IL10RB | 0.479 | 1.393777 | 5.321868 | 9.144338 | 1.30E-14 | 8.59E-13 | 22.83667 |
| MTMR2 | 0.325324 | 1.252946 | 6.57044 | 9.143105 | 1.31E-14 | 8.59E-13 | 22.83077 |
| HSBP1 | 0.282237 | 1.216079 | 9.57022 | 9.137769 | 1.34E-14 | 8.75E-13 | 22.80525 |
| TTC39A | 1.285797 | 2.438167 | 6.536703 | 9.125103 | 1.43E-14 | 9.23E-13 | 22.74466 |
| TNFSF10 | 0.914372 | 1.884748 | 9.838901 | 9.122541 | 1.45E-14 | 9.28E-13 | 22.73241 |
| MFAP1 | 0.336048 | 1.262294 | 8.338352 | 9.116553 | 1.49E-14 | 9.40E-13 | 22.70377 |
| TPD52L1 | 0.603986 | 1.51991 | 8.098022 | 9.115519 | 1.50E-14 | 9.40E-13 | 22.69882 |
| SPP1 | 0.491193 | 1.405607 | 12.76659 | 9.115217 | 1.50E-14 | 9.40E-13 | 22.69738 |
| CD83 | -0.76089 | 0.590133 | 7.853736 | -9.11105 | 1.53E-14 | 9.52E-13 | 22.67744 |
| COQ2 | 0.434551 | 1.35149 | 6.19967 | 9.109391 | 1.54E-14 | 9.53E-13 | 22.66951 |
| SEL1L3 | 0.80143 | 1.742828 | 6.990659 | 9.085824 | 1.73E-14 | 1.06E-12 | 22.5568 |
| PRDX4 | 0.499966 | 1.41418 | 9.275714 | 9.083086 | 1.76E-14 | 1.07E-12 | 22.54371 |

| | | | | | | | |
|---|---|---|---|---|---|---|---|
| NUP37 | 0.410986 | 1.329594 | 7.667582 | 9.080684 | 1.78E-14 | 1.07E-12 | 22.53222 |
| MYO6 | 0.347976 | 1.272774 | 8.350989 | 9.070374 | 1.87E-14 | 1.12E-12 | 22.48292 |
| GMFB | 0.477874 | 1.39269 | 8.220659 | 9.049327 | 2.07E-14 | 1.23E-12 | 22.38228 |
| CXADR | 0.773246 | 1.709111 | 7.298462 | 9.045514 | 2.11E-14 | 1.25E-12 | 22.36405 |
| ANXA5 | 0.48658 | 1.401119 | 9.856703 | 9.040936 | 2.16E-14 | 1.27E-12 | 22.34216 |
| NECAP2 | 0.423865 | 1.341516 | 7.413516 | 9.03863 | 2.18E-14 | 1.27E-12 | 22.33114 |
| ZFP36 | -1.04082 | 0.486051 | 8.318352 | -9.03014 | 2.27E-14 | 1.31E-12 | 22.29054 |
| HAGH | -0.50079 | 0.706719 | 9.266703 | -9.02961 | 2.28E-14 | 1.31E-12 | 22.28799 |
| PAFAH1B3 | 0.440449 | 1.357027 | 7.088022 | 9.007853 | 2.53E-14 | 1.45E-12 | 22.184 |
| ACBD4 | -0.35437 | 0.782213 | 6.688242 | -8.98119 | 2.88E-14 | 1.64E-12 | 22.05657 |
| MNS1 | 0.840271 | 1.790386 | 5.537692 | 8.966041 | 3.11E-14 | 1.75E-12 | 21.98415 |
| TPGS2 | 0.395087 | 1.315022 | 6.904286 | 8.96277 | 3.16E-14 | 1.77E-12 | 21.96852 |
| DPYS | -0.66575 | 0.630361 | 10.59209 | -8.96005 | 3.20E-14 | 1.78E-12 | 21.95554 |
| STRA6 | -0.50015 | 0.707033 | 6.355934 | -8.95762 | 3.24E-14 | 1.78E-12 | 21.9439 |
| G6PC | -1.50601 | 0.352083 | 7.744615 | -8.95736 | 3.24E-14 | 1.78E-12 | 21.94264 |
| SMARCA5 | 0.318314 | 1.246873 | 6.351538 | 8.950388 | 3.35E-14 | 1.83E-12 | 21.90935 |
| PSMB10 | 0.876473 | 1.835882 | 6.664725 | 8.94136 | 3.50E-14 | 1.90E-12 | 21.86621 |
| IQGAP1 | 0.418928 | 1.336933 | 7.808901 | 8.924862 | 3.79E-14 | 2.04E-12 | 21.78738 |
| CDC123 | 0.313763 | 1.242946 | 8.624505 | 8.913849 | 4.00E-14 | 2.14E-12 | 21.73476 |
| LHPP | -0.46232 | 0.725819 | 7.547033 | -8.91298 | 4.02E-14 | 2.14E-12 | 21.73064 |
| NR4A1 | -1.16156 | 0.44703 | 6.125604 | -8.89654 | 4.36E-14 | 2.30E-12 | 21.65209 |
| ACTR3 | 0.410758 | 1.329385 | 8.517253 | 8.891219 | 4.47E-14 | 2.35E-12 | 21.62666 |
| TRIM37 | 0.364295 | 1.287252 | 7.623407 | 8.870738 | 4.94E-14 | 2.57E-12 | 21.52884 |
| SRP54 | 0.31258 | 1.241926 | 8.580659 | 8.869808 | 4.96E-14 | 2.57E-12 | 21.5244 |
| CLCN3 | 0.353111 | 1.277312 | 7.694615 | 8.859914 | 5.21E-14 | 2.68E-12 | 21.47715 |
| HDAC6 | -0.42076 | 0.747029 | 8.087582 | -8.85363 | 5.37E-14 | 2.74E-12 | 21.44715 |
| PARP12 | 0.66257 | 1.5829 | 6.603297 | 8.852834 | 5.39E-14 | 2.74E-12 | 21.44334 |
| TFEB | -0.26734 | 0.830848 | 6.537363 | -8.84673 | 5.55E-14 | 2.81E-12 | 21.41421 |
| GADD45A | -0.76478 | 0.588544 | 10.46681 | -8.83025 | 6.01E-14 | 3.03E-12 | 21.33551 |
| FOXO4 | -0.30268 | 0.810744 | 5.70967 | -8.82394 | 6.20E-14 | 3.10E-12 | 21.30537 |
| ZNF331 | -0.8163 | 0.567897 | 6.612857 | -8.81378 | 6.52E-14 | 3.24E-12 | 21.25686 |
| CAP1 | 0.371749 | 1.29392 | 10.33253 | 8.807429 | 6.72E-14 | 3.32E-12 | 21.22656 |
| VPS35 | 0.351377 | 1.275778 | 10.04571 | 8.796875 | 7.08E-14 | 3.48E-12 | 21.17618 |
| AMPH | -0.35725 | 0.780653 | 4.777253 | -8.79152 | 7.26E-14 | 3.53E-12 | 21.15062 |
| MME | -0.90322 | 0.534691 | 10.35835 | -8.79115 | 7.27E-14 | 3.53E-12 | 21.14887 |
| COPB2 | 0.243401 | 1.18378 | 9.373407 | 8.790018 | 7.32E-14 | 3.53E-12 | 21.14346 |
| MFSD5 | 0.337319 | 1.263406 | 7.196154 | 8.777587 | 7.77E-14 | 3.73E-12 | 21.08413 |
| CAP2 | 0.524773 | 1.438707 | 6.994286 | 8.771948 | 7.99E-14 | 3.81E-12 | 21.05722 |
| CNNM2 | -0.34598 | 0.786776 | 6.65 | -8.75135 | 8.83E-14 | 4.19E-12 | 20.95893 |
| LPCAT3 | 0.444894 | 1.361214 | 6.86022 | 8.749615 | 8.90E-14 | 4.21E-12 | 20.95066 |
| NNMT | 1.951005 | 3.866437 | 7.4 | 8.742414 | 9.22E-14 | 4.29E-12 | 20.91631 |
| CYFIP1 | 0.375179 | 1.297 | 9.948571 | 8.742291 | 9.23E-14 | 4.29E-12 | 20.91572 |
| CCT6A | 0.408 | 1.326845 | 8.031758 | 8.742021 | 9.24E-14 | 4.29E-12 | 20.91443 |
| DYRK2 | 0.410159 | 1.328833 | 5.17 | 8.739495 | 9.35E-14 | 4.32E-12 | 20.90239 |
| SGCE | 0.418889 | 1.336898 | 8.197143 | 8.736432 | 9.49E-14 | 4.36E-12 | 20.88777 |
| MGAT2 | 0.30585 | 1.236147 | 6.404505 | 8.728471 | 9.87E-14 | 4.51E-12 | 20.8498 |
| RNF34 | 0.369014 | 1.29147 | 7.558132 | 8.726327 | 9.97E-14 | 4.53E-12 | 20.83957 |

| | | | | | | | |
|---|---|---|---|---|---|---|---|
| EXOC1 | 0.397647 | 1.317358 | 8.100769 | 8.718439 | 1.04E-13 | 4.68E-12 | 20.80195 |
| RBM7 | 0.41999 | 1.337919 | 7.397253 | 8.711141 | 1.07E-13 | 4.83E-12 | 20.76714 |
| HLA-A | 0.632502 | 1.550252 | 12.23538 | 8.698369 | 1.14E-13 | 5.11E-12 | 20.70623 |
| EAF2 | -0.67551 | 0.62611 | 8.847912 | -8.69622 | 1.15E-13 | 5.14E-12 | 20.69598 |
| TMED10 | 0.268179 | 1.204287 | 9.580659 | 8.692138 | 1.18E-13 | 5.20E-12 | 20.67652 |
| RPN2 | 0.237319 | 1.1788 | 10.4167 | 8.69135 | 1.18E-13 | 5.20E-12 | 20.67276 |
| PRODH2 | -0.8707 | 0.546883 | 10.94813 | -8.67954 | 1.25E-13 | 5.48E-12 | 20.61645 |
| ANXA11 | 0.35928 | 1.282786 | 7.055275 | 8.676641 | 1.27E-13 | 5.52E-12 | 20.60263 |
| C1S | 1.189527 | 2.280779 | 7.426923 | 8.676158 | 1.27E-13 | 5.52E-12 | 20.60033 |
| VCAM1 | 1.022628 | 2.031616 | 10.56396 | 8.665029 | 1.34E-13 | 5.77E-12 | 20.54728 |
| APOC3 | -1.53331 | 0.345484 | 6.462857 | -8.66477 | 1.34E-13 | 5.77E-12 | 20.54603 |
| CD99 | 0.518947 | 1.432909 | 8.569231 | 8.656469 | 1.40E-13 | 5.98E-12 | 20.50647 |
| SMAP1 | 0.370454 | 1.29276 | 8.173626 | 8.65337 | 1.42E-13 | 6.04E-12 | 20.4917 |
| SLC39A6 | 0.569135 | 1.483634 | 7.642527 | 8.643663 | 1.49E-13 | 6.30E-12 | 20.44544 |
| ITGB6 | 1.55529 | 2.938928 | 5.557143 | 8.632611 | 1.57E-13 | 6.60E-12 | 20.39277 |
| LEPROT | 0.430995 | 1.348163 | 10.13791 | 8.631393 | 1.58E-13 | 6.60E-12 | 20.38697 |
| CYP3A4 | -0.47905 | 0.717451 | 6.605934 | -8.63104 | 1.58E-13 | 6.60E-12 | 20.38531 |
| SLC19A2 | -0.7018 | 0.614804 | 6.861868 | -8.62563 | 1.63E-13 | 6.74E-12 | 20.35949 |
| FAM13B | 0.467686 | 1.38289 | 7.347143 | 8.623432 | 1.64E-13 | 6.78E-12 | 20.34903 |
| FAM98A | 0.429696 | 1.346949 | 8.638791 | 8.620466 | 1.67E-13 | 6.84E-12 | 20.3349 |
| GFPT1 | 0.394498 | 1.314485 | 7.427473 | 8.610769 | 1.75E-13 | 7.12E-12 | 20.2887 |
| TMEM50A | 0.327585 | 1.254911 | 8.923297 | 8.610428 | 1.75E-13 | 7.12E-12 | 20.28708 |
| BLNK | 0.642478 | 1.561008 | 7.479231 | 8.593983 | 1.90E-13 | 7.67E-12 | 20.20874 |
| SH3BGRL3 | 0.499454 | 1.413679 | 8.262418 | 8.586266 | 1.97E-13 | 7.93E-12 | 20.17199 |
| VEGFA | -0.45896 | 0.727512 | 7.25 | -8.58215 | 2.01E-13 | 8.05E-12 | 20.1524 |
| DENR | 0.370377 | 1.29269 | 8.71011 | 8.580833 | 2.02E-13 | 8.06E-12 | 20.14611 |
| DCTD | 0.318879 | 1.247361 | 7.977253 | 8.574092 | 2.09E-13 | 8.29E-12 | 20.11401 |
| ECT2 | 0.500039 | 1.414251 | 4.299011 | 8.5609 | 2.23E-13 | 8.80E-12 | 20.0512 |
| MAPK13 | 0.425589 | 1.343121 | 6.141978 | 8.555843 | 2.28E-13 | 8.98E-12 | 20.02712 |
| PFN1 | 0.600193 | 1.51592 | 8.745495 | 8.547905 | 2.37E-13 | 9.28E-12 | 19.98933 |
| RSU1 | 0.399527 | 1.319075 | 7.346264 | 8.54709 | 2.38E-13 | 9.28E-12 | 19.98545 |
| ITM2C | 0.629213 | 1.546721 | 8.845714 | 8.536315 | 2.51E-13 | 9.73E-12 | 19.93416 |
| HLA-DMA | 0.928826 | 1.903726 | 8.306484 | 8.532162 | 2.56E-13 | 9.88E-12 | 19.91439 |
| METTL21B | -0.66796 | 0.629395 | 7.651429 | -8.52666 | 2.63E-13 | 1.01E-11 | 19.88819 |
| BAX | 0.560406 | 1.474684 | 6.315385 | 8.524252 | 2.66E-13 | 1.02E-11 | 19.87675 |
| TNS2 | -0.41707 | 0.748943 | 6.525495 | -8.51398 | 2.80E-13 | 1.07E-11 | 19.82786 |
| UBE2D2 | 0.215918 | 1.161443 | 7.493077 | 8.510733 | 2.84E-13 | 1.08E-11 | 19.81241 |
| RASA1 | 0.292807 | 1.225021 | 7.331099 | 8.505846 | 2.91E-13 | 1.10E-11 | 19.78916 |
| GUCA2A | -0.32563 | 0.797951 | 5.595714 | -8.50499 | 2.92E-13 | 1.10E-11 | 19.78507 |
| TRAM1 | 0.440686 | 1.35725 | 9.948791 | 8.486538 | 3.20E-13 | 1.20E-11 | 19.69731 |
| UBE2K | 0.292005 | 1.22434 | 7.774615 | 8.467326 | 3.51E-13 | 1.31E-11 | 19.60593 |
| EIF4G3 | 0.326029 | 1.253558 | 5.592527 | 8.464464 | 3.56E-13 | 1.32E-11 | 19.59232 |
| IFI30 | 0.605106 | 1.521091 | 9.609011 | 8.463578 | 3.57E-13 | 1.32E-11 | 19.58811 |
| USP3 | 0.307502 | 1.237563 | 7.75967 | 8.462118 | 3.60E-13 | 1.32E-11 | 19.58117 |
| PCBP2 | -0.42393 | 0.745393 | 9.403626 | -8.45739 | 3.68E-13 | 1.35E-11 | 19.55868 |
| KLHL3 | -0.66883 | 0.629016 | 7.561868 | -8.44775 | 3.86E-13 | 1.41E-11 | 19.51286 |
| ANXA3 | 0.875812 | 1.83504 | 6.654615 | 8.443368 | 3.94E-13 | 1.43E-11 | 19.49202 |

| | | | | | | | |
|---|---|---|---|---|---|---|---|
| NARS | 0.249179 | 1.18853 | 11.15626 | 8.439796 | 4.01E-13 | 1.45E-11 | 19.47504 |
| CISH | -0.69743 | 0.616668 | 8.230549 | -8.43494 | 4.10E-13 | 1.47E-11 | 19.45195 |
| STAT1 | 0.761203 | 1.694903 | 7.160549 | 8.434631 | 4.11E-13 | 1.47E-11 | 19.45049 |
| C4orf27 | 0.330406 | 1.257367 | 7.831648 | 8.433713 | 4.13E-13 | 1.47E-11 | 19.44613 |
| CAPRIN1 | 0.309121 | 1.238952 | 8.023297 | 8.428382 | 4.24E-13 | 1.50E-11 | 19.42079 |
| PER1 | -0.56124 | 0.677721 | 7.436813 | -8.42393 | 4.33E-13 | 1.53E-11 | 19.39962 |
| HNRNPR | 0.270952 | 1.206604 | 8.366813 | 8.408128 | 4.67E-13 | 1.65E-11 | 19.32454 |
| PSMB9 | 1.014729 | 2.020524 | 6.884615 | 8.405057 | 4.74E-13 | 1.66E-11 | 19.30994 |
| ZNF207 | 0.292729 | 1.224956 | 8.937582 | 8.39792 | 4.91E-13 | 1.71E-11 | 19.27604 |
| GALNT3 | 0.613768 | 1.530251 | 6.053077 | 8.397683 | 4.92E-13 | 1.71E-11 | 19.27491 |
| MICU1 | -0.51882 | 0.697944 | 8.816703 | -8.39041 | 5.09E-13 | 1.76E-11 | 19.24034 |
| FLRT3 | 0.841797 | 1.792281 | 8.447143 | 8.389614 | 5.11E-13 | 1.76E-11 | 19.23658 |
| CTCF | 0.273527 | 1.208759 | 7.673956 | 8.388924 | 5.13E-13 | 1.76E-11 | 19.2333 |
| DPAGT1 | 0.304193 | 1.234728 | 7.299121 | 8.384118 | 5.25E-13 | 1.80E-11 | 19.21047 |
| DNMT1 | 0.499754 | 1.413972 | 6.346044 | 8.373946 | 5.51E-13 | 1.88E-11 | 19.16216 |
| CD58 | 0.495005 | 1.409325 | 7.17 | 8.371543 | 5.58E-13 | 1.89E-11 | 19.15075 |
| GNAI1 | 0.805126 | 1.747298 | 6.258791 | 8.369982 | 5.62E-13 | 1.90E-11 | 19.14334 |
| AVPI1 | -0.45848 | 0.727753 | 7.071758 | -8.36526 | 5.75E-13 | 1.93E-11 | 19.1209 |
| FKBP8 | -0.30067 | 0.811874 | 6.211099 | -8.36474 | 5.77E-13 | 1.93E-11 | 19.11846 |
| KLK1 | -1.72705 | 0.302069 | 8.168791 | -8.36325 | 5.81E-13 | 1.94E-11 | 19.11138 |
| PSENEN | 0.363768 | 1.286782 | 8.009451 | 8.362392 | 5.83E-13 | 1.94E-11 | 19.1073 |
| VPS4B | 0.345058 | 1.270202 | 7.768242 | 8.36 | 5.90E-13 | 1.96E-11 | 19.09594 |
| BLVRA | 0.261164 | 1.198445 | 8.841538 | 8.347105 | 6.28E-13 | 2.07E-11 | 19.03472 |
| AGPS | 0.567203 | 1.481648 | 7.044615 | 8.347024 | 6.28E-13 | 2.07E-11 | 19.03434 |
| DTYMK | 0.387551 | 1.308171 | 6.621429 | 8.34294 | 6.41E-13 | 2.10E-11 | 19.01495 |
| PNMA1 | 0.357884 | 1.281545 | 8.501758 | 8.333111 | 6.72E-13 | 2.19E-11 | 18.9683 |
| PMS1 | 0.359498 | 1.282979 | 6.705165 | 8.331531 | 6.77E-13 | 2.20E-11 | 18.9608 |
| FZD6 | 0.529425 | 1.443354 | 7.095934 | 8.327462 | 6.91E-13 | 2.24E-11 | 18.94149 |
| ADGRA3 | 0.280155 | 1.214325 | 5.995495 | 8.322847 | 7.06E-13 | 2.28E-11 | 18.91959 |
| FNTA | 0.336855 | 1.263 | 8.135055 | 8.321371 | 7.11E-13 | 2.29E-11 | 18.91259 |
| UBQLN2 | 0.48686 | 1.401391 | 7.259451 | 8.314879 | 7.34E-13 | 2.35E-11 | 18.88178 |
| GLB1 | 0.363729 | 1.286748 | 8.937692 | 8.313157 | 7.40E-13 | 2.36E-11 | 18.87361 |
| BCAP29 | 0.316966 | 1.245708 | 5.84022 | 8.311751 | 7.45E-13 | 2.36E-11 | 18.86694 |
| DSE | 0.763092 | 1.697124 | 6.956484 | 8.311604 | 7.46E-13 | 2.36E-11 | 18.86624 |
| AKR1B1 | 0.624802 | 1.541999 | 9.615055 | 8.306507 | 7.64E-13 | 2.41E-11 | 18.84206 |
| FOSB | -1.71604 | 0.304383 | 5.89967 | -8.30468 | 7.71E-13 | 2.42E-11 | 18.83341 |
| ASCC3 | 0.331541 | 1.258357 | 5.393297 | 8.302094 | 7.81E-13 | 2.44E-11 | 18.82113 |
| PHF11 | 0.410063 | 1.328744 | 8.485604 | 8.294843 | 8.09E-13 | 2.52E-11 | 18.78673 |
| SKAP2 | 0.351812 | 1.276162 | 7.315495 | 8.289234 | 8.31E-13 | 2.58E-11 | 18.76013 |
| ORMDL2 | 0.392609 | 1.312765 | 8.551538 | 8.280057 | 8.69E-13 | 2.69E-11 | 18.71661 |
| CORO1C | 0.470976 | 1.386047 | 8.906813 | 8.278814 | 8.74E-13 | 2.70E-11 | 18.71071 |
| DUSP1 | -1.24386 | 0.422243 | 7.591429 | -8.2708 | 9.08E-13 | 2.79E-11 | 18.67272 |
| CRY2 | -0.44843 | 0.732837 | 7.038681 | -8.2661 | 9.29E-13 | 2.85E-11 | 18.65046 |
| EOGT | 0.381507 | 1.302702 | 4.326484 | 8.252741 | 9.91E-13 | 3.03E-11 | 18.58711 |
| HLA-DRA | 0.925928 | 1.899905 | 9.934615 | 8.25005 | 1.00E-12 | 3.05E-11 | 18.57435 |
| MFN1 | 0.306787 | 1.23695 | 6.157143 | 8.243151 | 1.04E-12 | 3.15E-11 | 18.54166 |
| CPVL | 0.706169 | 1.631466 | 8.346813 | 8.239235 | 1.06E-12 | 3.19E-11 | 18.5231 |

| | | | | | | | |
|---|---|---|---|---|---|---|---|
| TSPYL2 | -0.4507 | 0.73169 | 6.815824 | -8.23345 | 1.09E-12 | 3.27E-11 | 18.49568 |
| INHA | -0.26654 | 0.831313 | 4.564066 | -8.23192 | 1.10E-12 | 3.29E-11 | 18.48843 |
| ZBED5 | 0.367329 | 1.289962 | 8.391429 | 8.225989 | 1.13E-12 | 3.37E-11 | 18.46034 |
| GTPBP4 | 0.294517 | 1.226474 | 6.668901 | 8.224627 | 1.14E-12 | 3.38E-11 | 18.45388 |
| NOP10 | 0.294575 | 1.226524 | 10.40154 | 8.221229 | 1.15E-12 | 3.42E-11 | 18.43779 |
| WDR1 | 0.305459 | 1.235812 | 9.056703 | 8.214096 | 1.19E-12 | 3.53E-11 | 18.404 |
| FAM208B | 0.347353 | 1.272224 | 6.022637 | 8.212372 | 1.20E-12 | 3.55E-11 | 18.39583 |
| LSM1 | 0.250179 | 1.189354 | 9.171758 | 8.211117 | 1.21E-12 | 3.56E-11 | 18.38989 |
| IL7 | 0.480517 | 1.395243 | 3.920879 | 8.210146 | 1.22E-12 | 3.56E-11 | 18.38529 |
| RARRES3 | 0.973377 | 1.963431 | 9.273297 | 8.207379 | 1.23E-12 | 3.58E-11 | 18.37218 |
| COL3A1 | 1.553043 | 2.934355 | 7.344945 | 8.207366 | 1.23E-12 | 3.58E-11 | 18.37213 |
| CLCN6 | -0.35896 | 0.779728 | 6.669451 | -8.20678 | 1.24E-12 | 3.58E-11 | 18.36935 |
| TES | 0.548657 | 1.462723 | 6.750879 | 8.204664 | 1.25E-12 | 3.61E-11 | 18.35933 |
| MVP | 0.515754 | 1.429741 | 8.663956 | 8.19989 | 1.28E-12 | 3.68E-11 | 18.33672 |
| HPGD | -0.84986 | 0.55484 | 8.106264 | -8.19475 | 1.31E-12 | 3.76E-11 | 18.3124 |
| ARMCX3 | 0.340681 | 1.266354 | 7.769121 | 8.192603 | 1.33E-12 | 3.79E-11 | 18.30222 |
| CCDC90B | 0.300614 | 1.231668 | 7.416264 | 8.185362 | 1.37E-12 | 3.91E-11 | 18.26793 |
| NSF | 0.278493 | 1.212927 | 8.33989 | 8.183053 | 1.39E-12 | 3.94E-11 | 18.257 |
| DYNLL1 | 0.330271 | 1.257249 | 11.70549 | 8.181477 | 1.40E-12 | 3.95E-11 | 18.24954 |
| TARS | 0.336053 | 1.262299 | 8.148571 | 8.167764 | 1.49E-12 | 4.21E-11 | 18.18464 |
| NUDT9 | 0.320111 | 1.248427 | 8.373297 | 8.166052 | 1.51E-12 | 4.23E-11 | 18.17653 |
| CD2AP | 0.355159 | 1.279127 | 7.617802 | 8.162182 | 1.54E-12 | 4.30E-11 | 18.15822 |
| ERGIC2 | 0.390604 | 1.310942 | 7.81033 | 8.15718 | 1.57E-12 | 4.39E-11 | 18.13455 |
| ARPC1B | 0.841338 | 1.791711 | 8.326264 | 8.155417 | 1.59E-12 | 4.41E-11 | 18.12621 |
| NRP1 | 0.743126 | 1.673798 | 7.038132 | 8.153454 | 1.60E-12 | 4.44E-11 | 18.11692 |
| CSNK1A1 | 0.258831 | 1.196509 | 9.035385 | 8.15148 | 1.62E-12 | 4.46E-11 | 18.10758 |
| TLR3 | 0.545845 | 1.459876 | 6.312967 | 8.151201 | 1.62E-12 | 4.46E-11 | 18.10626 |
| KLF9 | -0.67093 | 0.628103 | 6.926484 | -8.14911 | 1.64E-12 | 4.49E-11 | 18.09636 |
| XPNPEP2 | -0.8441 | 0.557058 | 9.408022 | -8.14099 | 1.70E-12 | 4.65E-11 | 18.05794 |
| PAK2 | 0.329111 | 1.256239 | 6.962747 | 8.137748 | 1.73E-12 | 4.71E-11 | 18.04262 |
| PYCARD | 0.837488 | 1.786936 | 5.281099 | 8.136659 | 1.74E-12 | 4.72E-11 | 18.03748 |
| ATF7IP | 0.376343 | 1.298047 | 7.121538 | 8.133367 | 1.76E-12 | 4.78E-11 | 18.0219 |
| GINS1 | 0.545309 | 1.459333 | 5.518571 | 8.130233 | 1.79E-12 | 4.84E-11 | 18.00708 |
| HLA-F | 0.71542 | 1.641961 | 9.717692 | 8.127222 | 1.82E-12 | 4.88E-11 | 17.99284 |
| GALK1 | -0.49734 | 0.70841 | 6.733626 | -8.12698 | 1.82E-12 | 4.88E-11 | 17.99172 |
| TRIOBP | 0.324763 | 1.252459 | 7.864945 | 8.125343 | 1.83E-12 | 4.91E-11 | 17.98396 |
| HMGN3 | 0.35013 | 1.274676 | 10.88901 | 8.11697 | 1.91E-12 | 5.08E-11 | 17.94436 |
| DROSHA | 0.274227 | 1.209346 | 7.145824 | 8.116875 | 1.91E-12 | 5.08E-11 | 17.94392 |
| AFM | -1.16512 | 0.445928 | 8.073626 | -8.11627 | 1.92E-12 | 5.08E-11 | 17.94106 |
| FANCF | 0.319396 | 1.247808 | 6.480769 | 8.111732 | 1.96E-12 | 5.17E-11 | 17.9196 |
| TNC | 1.059169 | 2.083731 | 5.866374 | 8.107596 | 2.00E-12 | 5.26E-11 | 17.90005 |
| GUCY1B3 | 0.731913 | 1.66084 | 6.248022 | 8.105062 | 2.02E-12 | 5.31E-11 | 17.88807 |
| ARPC5 | 0.4497 | 1.365757 | 9.248901 | 8.102587 | 2.05E-12 | 5.35E-11 | 17.87637 |
| C17orf62 | 0.389821 | 1.310231 | 5.924725 | 8.102189 | 2.05E-12 | 5.35E-11 | 17.87449 |
| RBPMS | 0.43128 | 1.34843 | 7.870879 | 8.100128 | 2.07E-12 | 5.39E-11 | 17.86475 |
| CALM2 | 0.255527 | 1.193771 | 12.65505 | 8.099368 | 2.08E-12 | 5.39E-11 | 17.86116 |
| CD24 | 0.499614 | 1.413835 | 10.63967 | 8.097186 | 2.10E-12 | 5.43E-11 | 17.85085 |

| | | | | | | | |
|---|---|---|---|---|---|---|---|
| SLC26A6 | -0.45587 | 0.729068 | 5.80022 | -8.09111 | 2.16E-12 | 5.58E-11 | 17.82214 |
| CLCC1 | 0.231208 | 1.173817 | 5.078681 | 8.090381 | 2.17E-12 | 5.58E-11 | 17.81869 |
| MCTS1 | 0.291633 | 1.224025 | 8.047692 | 8.086009 | 2.22E-12 | 5.68E-11 | 17.79803 |
| KDELC1 | 0.62813 | 1.545561 | 5.166484 | 8.083078 | 2.25E-12 | 5.74E-11 | 17.78418 |
| CDHR5 | -0.6634 | 0.63139 | 7.029121 | -8.08009 | 2.28E-12 | 5.81E-11 | 17.77006 |
| TRIM38 | 0.526169 | 1.4401 | 7.757802 | 8.079019 | 2.29E-12 | 5.82E-11 | 17.765 |
| TM9SF1 | 0.302614 | 1.233377 | 7.997253 | 8.077501 | 2.31E-12 | 5.85E-11 | 17.75783 |
| HNF4A | -0.29843 | 0.813134 | 6.302857 | -8.06919 | 2.40E-12 | 6.07E-11 | 17.71858 |
| ARL6IP5 | 0.56256 | 1.476888 | 9.683407 | 8.067977 | 2.42E-12 | 6.08E-11 | 17.71284 |
| RGS16 | -0.28089 | 0.823081 | 5.130879 | -8.06198 | 2.49E-12 | 6.24E-11 | 17.68449 |
| CNOT6 | 0.35557 | 1.279491 | 7.116484 | 8.056909 | 2.55E-12 | 6.38E-11 | 17.66057 |
| NOL11 | 0.33172 | 1.258513 | 8.281429 | 8.054786 | 2.58E-12 | 6.43E-11 | 17.65054 |
| TNNT2 | -0.58044 | 0.668758 | 5.662967 | -8.05298 | 2.60E-12 | 6.46E-11 | 17.642 |
| GSPT2 | 0.498372 | 1.412619 | 6.753187 | 8.049146 | 2.65E-12 | 6.56E-11 | 17.62391 |
| DRP2 | -0.2483 | 0.841888 | 4.953736 | -8.04598 | 2.69E-12 | 6.65E-11 | 17.60898 |
| DDN | -0.52518 | 0.694871 | 7.137033 | -8.03467 | 2.84E-12 | 7.00E-11 | 17.55555 |
| UBE2I | 0.214106 | 1.159985 | 7.610659 | 8.032322 | 2.87E-12 | 7.06E-11 | 17.54449 |
| C1R | 0.915101 | 1.885702 | 7.522088 | 8.031467 | 2.88E-12 | 7.07E-11 | 17.54046 |
| ALDH2 | -0.33251 | 0.794152 | 11.47253 | -8.02303 | 3.00E-12 | 7.34E-11 | 17.50066 |
| DRAM1 | 0.405754 | 1.324781 | 8.47956 | 8.015672 | 3.11E-12 | 7.58E-11 | 17.46591 |
| TPBG | 0.595406 | 1.510898 | 6.927692 | 8.012549 | 3.16E-12 | 7.67E-11 | 17.45118 |
| PANX1 | 0.317135 | 1.245854 | 5.317912 | 8.01104 | 3.18E-12 | 7.71E-11 | 17.44406 |
| TALDO1 | 0.337179 | 1.263284 | 10.21978 | 8.007979 | 3.23E-12 | 7.80E-11 | 17.42962 |
| HMGN4 | 0.259184 | 1.196801 | 8.621429 | 8.00583 | 3.26E-12 | 7.86E-11 | 17.41948 |
| TSTA3 | 0.378691 | 1.300161 | 7.458352 | 8.002792 | 3.31E-12 | 7.95E-11 | 17.40515 |
| STAM | 0.387541 | 1.308162 | 7.310989 | 8.001589 | 3.33E-12 | 7.97E-11 | 17.39948 |
| GATM | -0.47555 | 0.719192 | 12.22505 | -7.99852 | 3.38E-12 | 8.07E-11 | 17.38501 |
| SYDE1 | -0.22073 | 0.858131 | 6.198022 | -7.998 | 3.39E-12 | 8.07E-11 | 17.38254 |
| EIF5B | 0.231787 | 1.174289 | 7.045055 | 7.989883 | 3.52E-12 | 8.36E-11 | 17.34427 |
| SIRT2 | -0.26876 | 0.830034 | 8.272967 | -7.98406 | 3.62E-12 | 8.58E-11 | 17.31679 |
| KDELR2 | 0.269246 | 1.205178 | 9.249231 | 7.981657 | 3.66E-12 | 8.65E-11 | 17.30549 |
| GSTA1 | -1.18604 | 0.439508 | 11.83176 | -7.98105 | 3.68E-12 | 8.65E-11 | 17.30263 |
| CKAP4 | 0.349971 | 1.274535 | 7.621758 | 7.977234 | 3.74E-12 | 8.78E-11 | 17.28463 |
| NAE1 | 0.414792 | 1.333107 | 8.960769 | 7.976893 | 3.75E-12 | 8.78E-11 | 17.28302 |
| NUP85 | 0.396034 | 1.315885 | 6.167363 | 7.970411 | 3.87E-12 | 9.03E-11 | 17.25247 |
| DNAH3 | -0.23692 | 0.848556 | 5.791538 | -7.96518 | 3.97E-12 | 9.24E-11 | 17.22779 |
| TRIM32 | 0.304768 | 1.23522 | 6.125275 | 7.964042 | 3.99E-12 | 9.26E-11 | 17.22245 |
| RBM22 | 0.22413 | 1.168073 | 7.336703 | 7.95977 | 4.07E-12 | 9.43E-11 | 17.20232 |
| DCP2 | 0.327527 | 1.25486 | 6.240659 | 7.955259 | 4.16E-12 | 9.61E-11 | 17.18106 |
| SLPI | 1.502116 | 2.832579 | 8.108022 | 7.952103 | 4.22E-12 | 9.73E-11 | 17.16619 |
| PDK2 | -0.4824 | 0.715788 | 7.833626 | -7.94316 | 4.41E-12 | 1.01E-10 | 17.12406 |
| CDH15 | -0.27236 | 0.827965 | 4.769231 | -7.93546 | 4.58E-12 | 1.05E-10 | 17.08779 |
| IRF9 | 0.652184 | 1.571545 | 8.990769 | 7.935066 | 4.58E-12 | 1.05E-10 | 17.08593 |
| PSMD1 | 0.228541 | 1.17165 | 8.697363 | 7.934936 | 4.59E-12 | 1.05E-10 | 17.08532 |
| ANXA2P2 | 0.531966 | 1.445898 | 6.191538 | 7.927954 | 4.74E-12 | 1.08E-10 | 17.05244 |
| PRKD1 | 0.439179 | 1.355832 | 6.76022 | 7.92537 | 4.80E-12 | 1.09E-10 | 17.04027 |
| GNPTAB | 0.355333 | 1.279281 | 6.565714 | 7.915618 | 5.03E-12 | 1.14E-10 | 16.99435 |

| | | | | | | | |
|---|---|---|---|---|---|---|---|
| TGFBI | 0.942077 | 1.921293 | 7.539341 | 7.91488 | 5.05E-12 | 1.14E-10 | 16.99088 |
| F2RL1 | 0.400058 | 1.319561 | 7.98044 | 7.911315 | 5.14E-12 | 1.16E-10 | 16.9741 |
| LYZ | 1.904097 | 3.742745 | 6.415934 | 7.909615 | 5.18E-12 | 1.16E-10 | 16.96609 |
| C3 | 1.566812 | 2.962493 | 8.411319 | 7.90756 | 5.23E-12 | 1.17E-10 | 16.95642 |
| MIOS | 0.280918 | 1.214968 | 6.88022 | 7.903963 | 5.32E-12 | 1.19E-10 | 16.93949 |
| TP53TG5 | -0.36571 | 0.776087 | 5.722198 | -7.8905 | 5.68E-12 | 1.26E-10 | 16.87612 |
| TAP1 | 0.587304 | 1.502437 | 7.159121 | 7.88788 | 5.75E-12 | 1.28E-10 | 16.86381 |
| KIDINS220 | 0.288077 | 1.221012 | 7.325934 | 7.884192 | 5.85E-12 | 1.30E-10 | 16.84646 |
| PLS1 | 0.363536 | 1.286576 | 8.848901 | 7.883353 | 5.88E-12 | 1.30E-10 | 16.84251 |
| GPC4 | 0.569531 | 1.484041 | 7.530549 | 7.878478 | 6.02E-12 | 1.32E-10 | 16.81958 |
| PXDN | 0.72213 | 1.649616 | 6.092967 | 7.87686 | 6.06E-12 | 1.33E-10 | 16.81197 |
| STK38 | 0.411222 | 1.329812 | 7.168352 | 7.876764 | 6.07E-12 | 1.33E-10 | 16.81152 |
| NOL8 | 0.34557 | 1.270653 | 6.851538 | 7.876632 | 6.07E-12 | 1.33E-10 | 16.8109 |
| DYNLT1 | 0.325696 | 1.253269 | 10.89736 | 7.874907 | 6.12E-12 | 1.33E-10 | 16.80278 |
| CASP8 | 0.29815 | 1.229566 | 5.884176 | 7.872022 | 6.21E-12 | 1.35E-10 | 16.78922 |
| GAD1 | -0.28803 | 0.81902 | 4.486264 | -7.87152 | 6.22E-12 | 1.35E-10 | 16.78685 |
| NUP107 | 0.328527 | 1.25573 | 7.606154 | 7.871319 | 6.23E-12 | 1.35E-10 | 16.78591 |
| SMAD1 | 0.403541 | 1.322751 | 7.228901 | 7.864208 | 6.44E-12 | 1.39E-10 | 16.75247 |
| HAAO | -0.46478 | 0.724583 | 7.745165 | -7.85695 | 6.67E-12 | 1.43E-10 | 16.71834 |
| RFC4 | 0.401696 | 1.32106 | 6.654945 | 7.856745 | 6.68E-12 | 1.43E-10 | 16.71738 |
| NR4A3 | -0.59647 | 0.661369 | 4.513736 | -7.83422 | 7.44E-12 | 1.59E-10 | 16.61154 |
| NCK1 | 0.31301 | 1.242297 | 6.27022 | 7.825226 | 7.77E-12 | 1.66E-10 | 16.56926 |
| RARG | -0.26217 | 0.833831 | 6.962527 | -7.82512 | 7.77E-12 | 1.66E-10 | 16.56877 |
| TKFC | -0.70373 | 0.613983 | 6.984176 | -7.82309 | 7.85E-12 | 1.67E-10 | 16.55921 |
| AMT | -0.43769 | 0.738315 | 8.907473 | -7.82115 | 7.92E-12 | 1.68E-10 | 16.5501 |
| ZNF112 | 0.345367 | 1.270474 | 4.932308 | 7.820209 | 7.96E-12 | 1.68E-10 | 16.5457 |
| TMEM258 | 0.31257 | 1.241918 | 10.54022 | 7.816477 | 8.10E-12 | 1.71E-10 | 16.52817 |
| SMARCA1 | 0.325754 | 1.253319 | 6.78 | 7.813439 | 8.22E-12 | 1.73E-10 | 16.5139 |
| MTERF1 | 0.384676 | 1.305567 | 5.040659 | 7.813018 | 8.24E-12 | 1.73E-10 | 16.51192 |
| SLIT2 | -0.62559 | 0.648155 | 8.159121 | -7.81047 | 8.34E-12 | 1.74E-10 | 16.49993 |
| PTEN | 0.318633 | 1.247148 | 6.956044 | 7.809846 | 8.36E-12 | 1.75E-10 | 16.49703 |
| MYL12A | 0.409261 | 1.328005 | 10.70912 | 7.807441 | 8.46E-12 | 1.76E-10 | 16.48573 |
| RNF219 | 0.31572 | 1.244632 | 5.443516 | 7.802705 | 8.65E-12 | 1.80E-10 | 16.4635 |
| TYRO3 | -0.34316 | 0.78831 | 5.727912 | -7.8004 | 8.75E-12 | 1.81E-10 | 16.45268 |
| SRSF10 | 0.35815 | 1.281781 | 7.443846 | 7.799069 | 8.81E-12 | 1.82E-10 | 16.44643 |
| STAG1 | 0.307609 | 1.237655 | 6.454505 | 7.798935 | 8.81E-12 | 1.82E-10 | 16.4458 |
| C19orf57 | -0.26216 | 0.833839 | 6.463187 | -7.78774 | 9.30E-12 | 1.91E-10 | 16.39323 |
| FANCL | 0.416836 | 1.334996 | 5.593736 | 7.7873 | 9.32E-12 | 1.91E-10 | 16.39119 |
| SGPL1 | 0.216097 | 1.161587 | 6.461209 | 7.785505 | 9.40E-12 | 1.92E-10 | 16.38276 |
| NCBP2 | 0.257159 | 1.195123 | 7.379341 | 7.779346 | 9.68E-12 | 1.98E-10 | 16.35386 |
| ARHGEF17 | -0.3054 | 0.80922 | 7.056154 | -7.77622 | 9.82E-12 | 2.00E-10 | 16.3392 |
| CLDN3 | 0.681411 | 1.603707 | 6.93044 | 7.772982 | 9.98E-12 | 2.03E-10 | 16.324 |
| COPB1 | 0.244643 | 1.184799 | 9.90989 | 7.763479 | 1.04E-11 | 2.12E-10 | 16.27943 |
| GPX7 | 0.585763 | 1.500833 | 5.959011 | 7.762573 | 1.05E-11 | 2.12E-10 | 16.27518 |
| SLC12A6 | -0.47717 | 0.718383 | 6.871209 | -7.75687 | 1.08E-11 | 2.17E-10 | 16.24845 |
| UBD | 1.308324 | 2.476536 | 7.601538 | 7.75503 | 1.09E-11 | 2.19E-10 | 16.2398 |
| SAE1 | 0.382884 | 1.303946 | 7.969121 | 7.749315 | 1.12E-11 | 2.24E-10 | 16.21301 |

| | | | | | | | |
|---|---|---|---|---|---|---|---|
| FBXO7 | -0.28925 | 0.818329 | 10.04088 | -7.74223 | 1.16E-11 | 2.32E-10 | 16.17978 |
| TMX1 | 0.406546 | 1.325508 | 7.875604 | 7.737007 | 1.19E-11 | 2.37E-10 | 16.15531 |
| MYL6 | 0.302638 | 1.233397 | 12.16835 | 7.729129 | 1.23E-11 | 2.45E-10 | 16.11839 |
| NEIL1 | -0.23794 | 0.847954 | 5.894945 | -7.72702 | 1.24E-11 | 2.47E-10 | 16.10849 |
| TMEM50B | 0.39085 | 1.311166 | 8.711538 | 7.722635 | 1.27E-11 | 2.52E-10 | 16.08796 |
| HSF4 | -0.29992 | 0.812296 | 6.555165 | -7.71503 | 1.32E-11 | 2.61E-10 | 16.05236 |
| BTN2A2 | 0.27113 | 1.206753 | 5.524945 | 7.705842 | 1.38E-11 | 2.72E-10 | 16.0093 |
| MYL10 | -0.24474 | 0.843966 | 5.925495 | -7.69819 | 1.43E-11 | 2.81E-10 | 15.97347 |
| CALML3 | -1.05406 | 0.48161 | 6.215934 | -7.6955 | 1.45E-11 | 2.84E-10 | 15.9609 |
| USP8 | 0.268271 | 1.204363 | 7.304835 | 7.692157 | 1.47E-11 | 2.88E-10 | 15.94523 |
| KLHL2 | 0.265971 | 1.202445 | 7.88022 | 7.682959 | 1.54E-11 | 3.00E-10 | 15.90217 |
| HLA-DMB | 0.656957 | 1.576753 | 7.807912 | 7.678949 | 1.56E-11 | 3.05E-10 | 15.88341 |
| CTSV | -0.8636 | 0.54958 | 8.510989 | -7.67598 | 1.59E-11 | 3.09E-10 | 15.86954 |
| ALDOB | -0.47243 | 0.72075 | 12.87703 | -7.67382 | 1.60E-11 | 3.12E-10 | 15.85942 |
| HTR6 | -0.22274 | 0.856937 | 4.736593 | -7.67256 | 1.61E-11 | 3.13E-10 | 15.85351 |
| NRL | -0.22784 | 0.853915 | 5.394725 | -7.66864 | 1.64E-11 | 3.17E-10 | 15.83516 |
| LONRF3 | -0.41082 | 0.752195 | 5.60989 | -7.6685 | 1.64E-11 | 3.17E-10 | 15.83451 |
| NAA35 | 0.293435 | 1.225555 | 5.81967 | 7.661667 | 1.70E-11 | 3.27E-10 | 15.80255 |
| DDX58 | 0.589174 | 1.504385 | 5.374176 | 7.661607 | 1.70E-11 | 3.27E-10 | 15.80227 |
| MACROD1 | -0.59043 | 0.664147 | 7.638901 | -7.66051 | 1.71E-11 | 3.28E-10 | 15.79713 |
| SERPINB1 | 0.380652 | 1.30193 | 7.367582 | 7.649703 | 1.80E-11 | 3.44E-10 | 15.74661 |
| RPF1 | 0.209362 | 1.156177 | 9.054835 | 7.648831 | 1.81E-11 | 3.45E-10 | 15.74253 |
| NECAB3 | -0.27685 | 0.825391 | 6.364835 | -7.64732 | 1.82E-11 | 3.47E-10 | 15.73545 |
| TNFRSF11I | 0.766053 | 1.700611 | 7.531209 | 7.644947 | 1.84E-11 | 3.50E-10 | 15.72437 |
| PNMA2 | 0.499565 | 1.413787 | 5.407473 | 7.639872 | 1.89E-11 | 3.57E-10 | 15.70065 |
| ACAA1 | -0.51046 | 0.701997 | 10.1967 | -7.63584 | 1.92E-11 | 3.64E-10 | 15.6818 |
| STEAP1 | 0.581068 | 1.495956 | 6.995385 | 7.624812 | 2.03E-11 | 3.82E-10 | 15.63026 |
| FEV | -0.29877 | 0.812946 | 5.057692 | -7.62339 | 2.04E-11 | 3.84E-10 | 15.6236 |
| SMYD2 | 0.438928 | 1.355596 | 8.608791 | 7.620881 | 2.06E-11 | 3.88E-10 | 15.6119 |
| UCHL3 | 0.311362 | 1.240879 | 8.595275 | 7.620542 | 2.07E-11 | 3.88E-10 | 15.61031 |
| SLC5A5 | -0.2362 | 0.848977 | 7.144066 | -7.61975 | 2.08E-11 | 3.88E-10 | 15.60663 |
| RPAP3 | 0.32213 | 1.250175 | 5.245165 | 7.616237 | 2.11E-11 | 3.94E-10 | 15.5902 |
| BECN1 | 0.19643 | 1.145859 | 8.036484 | 7.612205 | 2.15E-11 | 4.01E-10 | 15.57137 |
| PPP1R16B | -0.76729 | 0.587518 | 8.402308 | -7.61096 | 2.16E-11 | 4.01E-10 | 15.56554 |
| CD59 | 0.31086 | 1.240447 | 10.51242 | 7.610923 | 2.17E-11 | 4.01E-10 | 15.56538 |
| COL6A3 | 1.224976 | 2.337515 | 6.29967 | 7.60962 | 2.18E-11 | 4.03E-10 | 15.55929 |
| UBXN8 | 0.298633 | 1.229978 | 7.256154 | 7.608517 | 2.19E-11 | 4.04E-10 | 15.55414 |
| SNX10 | 0.688792 | 1.611934 | 8.166264 | 7.607762 | 2.20E-11 | 4.05E-10 | 15.55062 |
| C6orf15 | -0.35561 | 0.781537 | 6.411538 | -7.6044 | 2.23E-11 | 4.10E-10 | 15.53492 |
| PLA2G6 | -0.22759 | 0.854061 | 6.745934 | -7.60029 | 2.28E-11 | 4.18E-10 | 15.51573 |
| TSPAN1 | 0.519773 | 1.43373 | 11.65681 | 7.599788 | 2.28E-11 | 4.18E-10 | 15.51338 |
| ZNF200 | 0.257357 | 1.195287 | 5.208352 | 7.592752 | 2.36E-11 | 4.31E-10 | 15.48053 |
| PDK4 | -1.3887 | 0.38191 | 7.691978 | -7.59005 | 2.39E-11 | 4.36E-10 | 15.46794 |
| DNAJB6 | 0.317783 | 1.246413 | 6.477363 | 7.58298 | 2.47E-11 | 4.50E-10 | 15.43493 |
| ERF | -0.34769 | 0.785841 | 6.701978 | -7.57775 | 2.54E-11 | 4.60E-10 | 15.4105 |
| SCN4A | -0.23485 | 0.849776 | 4.465824 | -7.57715 | 2.54E-11 | 4.60E-10 | 15.40771 |
| KDELR3 | 0.334295 | 1.260761 | 5.708571 | 7.572531 | 2.60E-11 | 4.70E-10 | 15.38617 |

| | | | | | | | |
|---|---|---|---|---|---|---|---|
| CHTOP | 0.243918 | 1.184204 | 7.716923 | 7.571319 | 2.62E-11 | 4.71E-10 | 15.38052 |
| DCTPP1 | 0.294193 | 1.226199 | 8.594176 | 7.569814 | 2.63E-11 | 4.74E-10 | 15.37349 |
| CDK18 | -0.37259 | 0.772392 | 6.209011 | -7.56587 | 2.68E-11 | 4.82E-10 | 15.35511 |
| GCC2 | 0.363174 | 1.286253 | 7.892418 | 7.558801 | 2.78E-11 | 4.97E-10 | 15.32213 |
| SLC12A3 | -0.9192 | 0.528801 | 7.651319 | -7.55685 | 2.80E-11 | 5.01E-10 | 15.31305 |
| TBXA2R | -0.21314 | 0.862658 | 5.553297 | -7.55482 | 2.83E-11 | 5.05E-10 | 15.30358 |
| ELK3 | 0.293536 | 1.225641 | 5.064286 | 7.554326 | 2.84E-11 | 5.05E-10 | 15.30126 |
| UBAP2 | 0.258778 | 1.196465 | 7.282967 | 7.552636 | 2.86E-11 | 5.08E-10 | 15.29338 |
| CASP4 | 0.549585 | 1.463664 | 6.283077 | 7.548003 | 2.92E-11 | 5.18E-10 | 15.27178 |
| CPNE7 | -0.2843 | 0.821137 | 5.665714 | -7.54621 | 2.95E-11 | 5.21E-10 | 15.26343 |
| CAMK2G | -0.32369 | 0.799026 | 7.004066 | -7.53984 | 3.04E-11 | 5.36E-10 | 15.23374 |
| RAB27A | 0.450575 | 1.366585 | 6.078681 | 7.538135 | 3.06E-11 | 5.39E-10 | 15.22578 |
| SEC61G | 0.334227 | 1.260702 | 9.615495 | 7.53644 | 3.09E-11 | 5.43E-10 | 15.21789 |
| SPINT2 | 0.268435 | 1.2045 | 9.902308 | 7.532488 | 3.15E-11 | 5.52E-10 | 15.19947 |
| ATR | 0.287908 | 1.220869 | 5.888242 | 7.530323 | 3.18E-11 | 5.56E-10 | 15.18938 |
| BPHL | -0.59982 | 0.659836 | 9.351429 | -7.50681 | 3.56E-11 | 6.21E-10 | 15.07987 |
| CLDN15 | -0.29527 | 0.814922 | 8.057033 | -7.50593 | 3.57E-11 | 6.22E-10 | 15.07576 |
| FAM102A | -0.38377 | 0.766431 | 7.18044 | -7.49974 | 3.68E-11 | 6.40E-10 | 15.04697 |
| MET | 0.374865 | 1.296718 | 7.534286 | 7.498879 | 3.69E-11 | 6.41E-10 | 15.04295 |
| TXLNA | 0.327821 | 1.255116 | 6.164066 | 7.48965 | 3.86E-11 | 6.68E-10 | 15 |
| FOXN1 | -0.22831 | 0.853632 | 4.872967 | -7.48862 | 3.88E-11 | 6.70E-10 | 14.99518 |
| FLII | 0.294507 | 1.226466 | 7.298462 | 7.48749 | 3.90E-11 | 6.72E-10 | 14.98995 |
| DBR1 | 0.334662 | 1.261082 | 5.085275 | 7.484726 | 3.95E-11 | 6.80E-10 | 14.97709 |
| TGM5 | -0.26612 | 0.831555 | 6.173187 | -7.48418 | 3.96E-11 | 6.80E-10 | 14.97456 |
| EXT2 | 0.215831 | 1.161373 | 7.649121 | 7.481226 | 4.01E-11 | 6.88E-10 | 14.9608 |
| PLK2 | 0.782662 | 1.720302 | 4.916813 | 7.478761 | 4.06E-11 | 6.95E-10 | 14.94933 |
| AGA | 0.392275 | 1.312462 | 7.066374 | 7.474517 | 4.15E-11 | 7.08E-10 | 14.92959 |
| DAPK2 | -0.22557 | 0.855257 | 4.947802 | -7.47374 | 4.16E-11 | 7.09E-10 | 14.926 |
| DERL2 | 0.285343 | 1.2187 | 8.501538 | 7.473227 | 4.17E-11 | 7.09E-10 | 14.9236 |
| NAB2 | -0.34966 | 0.784768 | 5.912308 | -7.47236 | 4.19E-11 | 7.11E-10 | 14.91958 |
| AGFG1 | 0.278502 | 1.212935 | 6.65033 | 7.469411 | 4.25E-11 | 7.19E-10 | 14.90585 |
| G0S2 | -0.96012 | 0.514014 | 9.03978 | -7.4688 | 4.26E-11 | 7.20E-10 | 14.903 |
| IFNGR2 | 0.338464 | 1.264409 | 8.368242 | 7.465613 | 4.32E-11 | 7.30E-10 | 14.88819 |
| TP53 | 0.330116 | 1.257114 | 5.178462 | 7.465028 | 4.34E-11 | 7.30E-10 | 14.88547 |
| MAP9 | 0.3547 | 1.27872 | 5.366923 | 7.462306 | 4.39E-11 | 7.38E-10 | 14.87281 |
| HN1L | 0.282507 | 1.216307 | 7.292527 | 7.45838 | 4.48E-11 | 7.51E-10 | 14.85456 |
| SERPINA4 | -0.56753 | 0.67477 | 5.84044 | -7.45647 | 4.52E-11 | 7.56E-10 | 14.84566 |
| HLA-C | 0.532155 | 1.446087 | 11.68011 | 7.455428 | 4.54E-11 | 7.58E-10 | 14.84083 |
| ALAS1 | 0.415826 | 1.334062 | 7.787802 | 7.449463 | 4.67E-11 | 7.79E-10 | 14.81311 |
| SRPK2 | 0.222251 | 1.166552 | 6.671209 | 7.443608 | 4.80E-11 | 7.99E-10 | 14.7859 |
| ZDHHC13 | 0.374357 | 1.296262 | 6.041209 | 7.439046 | 4.91E-11 | 8.15E-10 | 14.76471 |
| HHAT | 0.331604 | 1.258412 | 5.636154 | 7.43793 | 4.93E-11 | 8.17E-10 | 14.75952 |
| SORBS3 | -0.2374 | 0.848275 | 5.96978 | -7.43739 | 4.94E-11 | 8.18E-10 | 14.75704 |
| PLA2G4C | 0.730251 | 1.658928 | 6.682418 | 7.434497 | 5.01E-11 | 8.28E-10 | 14.74358 |
| FBRS | -0.23192 | 0.851499 | 5.608791 | -7.4341 | 5.02E-11 | 8.28E-10 | 14.74174 |
| ARF4 | 0.282691 | 1.216462 | 9.960879 | 7.433157 | 5.04E-11 | 8.30E-10 | 14.73735 |
| TRPM6 | -0.32468 | 0.798475 | 5.068791 | -7.43204 | 5.07E-11 | 8.33E-10 | 14.73215 |

| | | | | | | | |
|---|---|---|---|---|---|---|---|
| PPID | 0.420942 | 1.338801 | 7.910549 | 7.429265 | 5.14E-11 | 8.42E-10 | 14.71927 |
| RBBP7 | 0.246894 | 1.186649 | 9.172308 | 7.427604 | 5.18E-11 | 8.47E-10 | 14.71156 |
| ZNF816 | 0.414493 | 1.33283 | 6.827143 | 7.425644 | 5.23E-11 | 8.53E-10 | 14.70246 |
| GSTA3 | -0.48878 | 0.712629 | 7.173297 | -7.4233 | 5.29E-11 | 8.61E-10 | 14.69157 |
| SCARB2 | 0.401928 | 1.321272 | 7.575495 | 7.421198 | 5.34E-11 | 8.68E-10 | 14.68182 |
| MAP4K4 | 0.367285 | 1.289923 | 5.389451 | 7.42004 | 5.37E-11 | 8.71E-10 | 14.67644 |
| IQSEC2 | -0.24323 | 0.844851 | 5.754286 | -7.41849 | 5.41E-11 | 8.76E-10 | 14.66926 |
| LRRFIP1 | 0.381739 | 1.302912 | 7.737033 | 7.41238 | 5.57E-11 | 9.00E-10 | 14.64088 |
| XPO1 | 0.327082 | 1.254474 | 8.67044 | 7.411863 | 5.58E-11 | 9.00E-10 | 14.63848 |
| MAP3K10 | -0.22972 | 0.852798 | 6.798791 | -7.40962 | 5.64E-11 | 9.08E-10 | 14.62807 |
| HIF1A | 0.521908 | 1.435853 | 10.60396 | 7.408862 | 5.66E-11 | 9.10E-10 | 14.62455 |
| KDM5B | 0.292773 | 1.224993 | 6.66956 | 7.405141 | 5.76E-11 | 9.24E-10 | 14.60728 |
| PRX | -0.28894 | 0.818505 | 6.244945 | -7.39474 | 6.05E-11 | 9.69E-10 | 14.55901 |
| KIAA0930 | 0.306072 | 1.236337 | 5.894615 | 7.394225 | 6.07E-11 | 9.70E-10 | 14.55663 |
| ZP2 | -0.39178 | 0.762187 | 4.986044 | -7.39287 | 6.11E-11 | 9.74E-10 | 14.55036 |
| C3orf18 | -0.30457 | 0.809681 | 6.833516 | -7.39087 | 6.17E-11 | 9.82E-10 | 14.54108 |
| MAPK1 | 0.300043 | 1.231182 | 7.00033 | 7.389564 | 6.20E-11 | 9.84E-10 | 14.53501 |
| TSPO | 0.417976 | 1.336052 | 8.305604 | 7.389538 | 6.20E-11 | 9.84E-10 | 14.53489 |
| TUBG2 | -0.35955 | 0.779407 | 5.352418 | -7.38843 | 6.24E-11 | 9.87E-10 | 14.52977 |
| CD74 | 0.755918 | 1.688706 | 9.75011 | 7.385465 | 6.33E-11 | 9.98E-10 | 14.516 |
| LPGAT1 | 0.536618 | 1.450568 | 5.553187 | 7.385385 | 6.33E-11 | 9.98E-10 | 14.51563 |
| EPHX1 | -0.61621 | 0.652384 | 8.995934 | -7.3835 | 6.38E-11 | 1.01E-09 | 14.50687 |
| DBI | 0.333845 | 1.260368 | 11.86813 | 7.38165 | 6.44E-11 | 1.01E-09 | 14.49831 |
| TRIM24 | 0.412444 | 1.330939 | 6.183956 | 7.378092 | 6.55E-11 | 1.03E-09 | 14.48181 |
| CCDC51 | 0.372932 | 1.294982 | 8.246374 | 7.375792 | 6.62E-11 | 1.04E-09 | 14.47115 |
| RXRA | -0.31631 | 0.803119 | 6.759451 | -7.37284 | 6.71E-11 | 1.05E-09 | 14.45747 |
| BZRAP1 | -0.28301 | 0.821875 | 5.434615 | -7.37138 | 6.76E-11 | 1.05E-09 | 14.4507 |
| TUG1 | 0.30572 | 1.236035 | 8.078571 | 7.369932 | 6.81E-11 | 1.06E-09 | 14.44399 |
| C2orf72 | -0.2492 | 0.841361 | 5.372637 | -7.36507 | 6.97E-11 | 1.08E-09 | 14.42144 |
| GAR1 | 0.309435 | 1.239222 | 7.467582 | 7.363825 | 7.01E-11 | 1.09E-09 | 14.41568 |
| KIAA0101 | 0.588058 | 1.503222 | 5.233407 | 7.362697 | 7.05E-11 | 1.09E-09 | 14.41045 |
| SQRDL | 0.318377 | 1.246927 | 9.594396 | 7.362283 | 7.06E-11 | 1.09E-09 | 14.40854 |
| FAM3A | -0.23707 | 0.848468 | 6.984725 | -7.35354 | 7.36E-11 | 1.14E-09 | 14.36803 |
| ARHGAP29 | 0.529816 | 1.443745 | 9.113736 | 7.350621 | 7.46E-11 | 1.15E-09 | 14.35451 |
| HNRNPM | 0.339942 | 1.265706 | 9.055495 | 7.3506 | 7.46E-11 | 1.15E-09 | 14.35441 |
| ADNP | 0.332208 | 1.258938 | 7.883626 | 7.349396 | 7.50E-11 | 1.15E-09 | 14.34883 |
| RB1 | 0.238101 | 1.17944 | 5.542308 | 7.346733 | 7.60E-11 | 1.16E-09 | 14.3365 |
| TMEM87A | 0.258179 | 1.195968 | 7.735714 | 7.344886 | 7.67E-11 | 1.17E-09 | 14.32795 |
| C3orf14 | 0.364169 | 1.28714 | 6.657692 | 7.343817 | 7.70E-11 | 1.17E-09 | 14.323 |
| UPB1 | -0.82448 | 0.564684 | 8.480549 | -7.34361 | 7.71E-11 | 1.17E-09 | 14.32206 |
| FOCAD | 0.457802 | 1.373448 | 6.437473 | 7.343037 | 7.73E-11 | 1.18E-09 | 14.31938 |
| MACF1 | 0.383773 | 1.30475 | 7.83956 | 7.33771 | 7.93E-11 | 1.20E-09 | 14.29472 |
| PRR36 | -0.29941 | 0.812584 | 5.738462 | -7.33595 | 8.00E-11 | 1.21E-09 | 14.28655 |
| SUV420H1 | 0.366618 | 1.289327 | 7.109121 | 7.334754 | 8.04E-11 | 1.22E-09 | 14.28103 |
| RAB38 | 0.319952 | 1.248289 | 6.526044 | 7.331067 | 8.18E-11 | 1.24E-09 | 14.26397 |
| BUB3 | 0.237512 | 1.178958 | 7.835495 | 7.325892 | 8.39E-11 | 1.26E-09 | 14.24001 |
| DDRGK1 | 0.303435 | 1.234079 | 8.054615 | 7.317246 | 8.74E-11 | 1.31E-09 | 14.20001 |

| | | | | | | | |
|---|---|---|---|---|---|---|---|
| TRA2B | 0.231126 | 1.17375 | 6.564945 | 7.316026 | 8.79E-11 | 1.32E-09 | 14.19436 |
| TMEM140 | 0.535725 | 1.44967 | 8.202527 | 7.31476 | 8.84E-11 | 1.32E-09 | 14.18851 |
| CDK14 | -0.3042 | 0.809892 | 6.930659 | -7.31426 | 8.86E-11 | 1.33E-09 | 14.1862 |
| IRAK1 | 0.372638 | 1.294718 | 9.242967 | 7.308183 | 9.12E-11 | 1.36E-09 | 14.15808 |
| CYP27B1 | -1.28678 | 0.409865 | 8.408681 | -7.30322 | 9.34E-11 | 1.39E-09 | 14.13513 |
| CCNB1 | 0.617295 | 1.533996 | 5.063516 | 7.301675 | 9.40E-11 | 1.40E-09 | 14.12799 |
| CRELD2 | 0.338343 | 1.264304 | 8.182747 | 7.297827 | 9.58E-11 | 1.42E-09 | 14.1102 |
| SEC22A | 0.255976 | 1.194143 | 4.955495 | 7.293661 | 9.77E-11 | 1.45E-09 | 14.09094 |
| PIK3R3 | 0.271662 | 1.207198 | 5.649121 | 7.292574 | 9.82E-11 | 1.45E-09 | 14.08591 |
| SEMA3G | -0.53472 | 0.690293 | 5.553187 | -7.28749 | 1.01E-10 | 1.49E-09 | 14.0624 |
| RAB22A | 0.249952 | 1.189167 | 7.591429 | 7.28195 | 1.03E-10 | 1.52E-09 | 14.03681 |
| ZNF146 | 0.279749 | 1.213983 | 7.177033 | 7.27925 | 1.05E-10 | 1.54E-09 | 14.02433 |
| HTATIP2 | 0.462362 | 1.377796 | 7.184945 | 7.277099 | 1.06E-10 | 1.55E-09 | 14.01439 |
| FAM212B | -0.26514 | 0.832118 | 5.777582 | -7.27431 | 1.07E-10 | 1.57E-09 | 14.00149 |
| OR1D2 | -0.24688 | 0.842717 | 5.698352 | -7.27408 | 1.07E-10 | 1.57E-09 | 14.00046 |
| RAC1 | 0.222614 | 1.166845 | 11.14769 | 7.271824 | 1.08E-10 | 1.58E-09 | 13.99003 |
| WNT10B | -0.36828 | 0.774705 | 6.625275 | -7.27118 | 1.09E-10 | 1.59E-09 | 13.98704 |
| S100A6 | 0.585797 | 1.500868 | 9.890549 | 7.27004 | 1.09E-10 | 1.59E-09 | 13.98179 |
| IL13RA1 | 0.365652 | 1.288464 | 7.135165 | 7.266976 | 1.11E-10 | 1.61E-09 | 13.96764 |
| GTF2H1 | 0.320662 | 1.248903 | 7.088352 | 7.264906 | 1.12E-10 | 1.63E-09 | 13.95808 |
| AFP | -0.3287 | 0.796256 | 5.296154 | -7.26445 | 1.12E-10 | 1.63E-09 | 13.95596 |
| DPF1 | -0.25323 | 0.839018 | 5.99956 | -7.26255 | 1.13E-10 | 1.64E-09 | 13.94721 |
| CAMK2A | -0.2958 | 0.814619 | 5.812637 | -7.26107 | 1.14E-10 | 1.65E-09 | 13.94037 |
| FAIM | 0.529841 | 1.44377 | 5.994835 | 7.260532 | 1.14E-10 | 1.65E-09 | 13.93788 |
| ADGRG1 | 0.530014 | 1.443944 | 8.632747 | 7.259203 | 1.15E-10 | 1.65E-09 | 13.93174 |
| SMAD2 | 0.202841 | 1.150962 | 7.728132 | 7.258867 | 1.15E-10 | 1.65E-09 | 13.93019 |
| GRN | 0.319227 | 1.247662 | 8.933077 | 7.257252 | 1.16E-10 | 1.66E-09 | 13.92274 |
| ESR1 | -0.19613 | 0.872892 | 5.237363 | -7.2568 | 1.16E-10 | 1.67E-09 | 13.92064 |
| FUZ | -0.42707 | 0.74377 | 6.710549 | -7.25508 | 1.17E-10 | 1.68E-09 | 13.91272 |
| HPD | -0.89415 | 0.538064 | 11.3611 | -7.25456 | 1.17E-10 | 1.68E-09 | 13.91033 |
| TRIM15 | -0.24615 | 0.843144 | 6.101538 | -7.25064 | 1.20E-10 | 1.71E-09 | 13.89222 |
| LBR | 0.423652 | 1.341319 | 8.453846 | 7.249453 | 1.20E-10 | 1.71E-09 | 13.88674 |
| CCNB1IP1 | -0.39714 | 0.759365 | 9.262527 | -7.24663 | 1.22E-10 | 1.73E-09 | 13.87371 |
| RWDD3 | 0.350478 | 1.274983 | 6.244835 | 7.245634 | 1.23E-10 | 1.74E-09 | 13.86912 |
| PITX3 | -0.29042 | 0.817667 | 5.927692 | -7.23995 | 1.26E-10 | 1.78E-09 | 13.8429 |
| EMP3 | 0.719744 | 1.64689 | 6.904396 | 7.239178 | 1.26E-10 | 1.79E-09 | 13.83933 |
| SWAP70 | 0.374899 | 1.296748 | 6.065824 | 7.236267 | 1.28E-10 | 1.81E-09 | 13.8259 |
| TUBB | 0.439865 | 1.356477 | 9.201429 | 7.231156 | 1.31E-10 | 1.85E-09 | 13.80232 |
| RRAGC | 0.307222 | 1.237323 | 8.686923 | 7.230183 | 1.32E-10 | 1.85E-09 | 13.79784 |
| DAD1 | 0.231498 | 1.174053 | 10.61187 | 7.224049 | 1.36E-10 | 1.91E-09 | 13.76955 |
| PRDM14 | -0.3185 | 0.801905 | 5.64011 | -7.22348 | 1.36E-10 | 1.91E-09 | 13.76691 |
| PSME1 | 0.282382 | 1.216201 | 10.09681 | 7.222391 | 1.37E-10 | 1.91E-09 | 13.76191 |
| EIF4G2 | 0.212024 | 1.158312 | 10.28593 | 7.220824 | 1.38E-10 | 1.92E-09 | 13.75468 |
| ATP10B | -0.29168 | 0.81695 | 5.54011 | -7.22053 | 1.38E-10 | 1.92E-09 | 13.75331 |
| SIRT3 | -0.2168 | 0.860474 | 6.506923 | -7.22009 | 1.38E-10 | 1.92E-09 | 13.75128 |
| TMED9 | 0.267773 | 1.203948 | 9.442198 | 7.219731 | 1.38E-10 | 1.93E-09 | 13.74964 |
| SPA17 | 0.435406 | 1.352291 | 6.698571 | 7.218784 | 1.39E-10 | 1.93E-09 | 13.74527 |

| | | | | | | | |
|---|---|---|---|---|---|---|---|
| MFSD10 | 0.440556 | 1.357127 | 5.802857 | 7.216833 | 1.40E-10 | 1.94E-09 | 13.73628 |
| LAPTM5 | 1.079512 | 2.113321 | 7.251868 | 7.216567 | 1.41E-10 | 1.94E-09 | 13.73505 |
| ABCC10 | 0.302425 | 1.233216 | 6.618681 | 7.209994 | 1.45E-10 | 2.00E-09 | 13.70476 |
| HLA-DRB1 | 0.81628 | 1.76086 | 10.64626 | 7.204696 | 1.49E-10 | 2.05E-09 | 13.68035 |
| RNF139 | 0.259807 | 1.197318 | 8.25978 | 7.200446 | 1.52E-10 | 2.09E-09 | 13.66077 |
| CELF3 | -0.24193 | 0.845615 | 6.183626 | -7.19795 | 1.53E-10 | 2.11E-09 | 13.64925 |
| DUSP26 | -0.31077 | 0.80621 | 5.151538 | -7.19738 | 1.54E-10 | 2.11E-09 | 13.64666 |
| ALB | -1.85773 | 0.275909 | 9.533297 | -7.19682 | 1.54E-10 | 2.11E-09 | 13.64405 |
| FADD | 0.320271 | 1.248565 | 7.710549 | 7.194341 | 1.56E-10 | 2.13E-09 | 13.63265 |
| LRP10 | 0.278454 | 1.212895 | 8.578132 | 7.19231 | 1.58E-10 | 2.15E-09 | 13.62329 |
| VANGL1 | 0.322372 | 1.250385 | 5.436154 | 7.18467 | 1.63E-10 | 2.23E-09 | 13.58811 |
| HDAC1 | 0.351106 | 1.275538 | 7.263407 | 7.18272 | 1.65E-10 | 2.24E-09 | 13.57914 |
| ALDH6A1 | -0.51448 | 0.700046 | 11.09407 | -7.18257 | 1.65E-10 | 2.24E-09 | 13.57846 |
| KRT83 | -0.25453 | 0.838262 | 5.28044 | -7.18206 | 1.65E-10 | 2.24E-09 | 13.57609 |
| GPD1 | -0.50885 | 0.702782 | 8.80011 | -7.18145 | 1.66E-10 | 2.25E-09 | 13.57331 |
| MCM3 | 0.324807 | 1.252497 | 6.986923 | 7.178338 | 1.68E-10 | 2.28E-09 | 13.55896 |
| NDNF | -1.28862 | 0.409343 | 6.454945 | -7.17548 | 1.71E-10 | 2.30E-09 | 13.54579 |
| CA7 | -0.31189 | 0.805586 | 5.800769 | -7.17289 | 1.73E-10 | 2.33E-09 | 13.53389 |
| BZW2 | 0.310362 | 1.240019 | 8.52978 | 7.169921 | 1.75E-10 | 2.36E-09 | 13.52023 |
| GBP2 | 0.79699 | 1.737473 | 5.348681 | 7.169617 | 1.75E-10 | 2.36E-09 | 13.51883 |
| ARHGAP33 | -0.22129 | 0.857795 | 5.822308 | -7.16816 | 1.77E-10 | 2.37E-09 | 13.51211 |
| GABRR2 | -0.27475 | 0.826594 | 6.31044 | -7.16709 | 1.77E-10 | 2.38E-09 | 13.50718 |
| PIM1 | -0.36808 | 0.774814 | 6.444505 | -7.16678 | 1.78E-10 | 2.38E-09 | 13.50578 |
| MARCKSL1 | 0.587478 | 1.502618 | 7.507033 | 7.161704 | 1.82E-10 | 2.43E-09 | 13.48242 |
| SPRY1 | 0.339942 | 1.265706 | 6.985495 | 7.158756 | 1.84E-10 | 2.46E-09 | 13.46887 |
| ARHGEF16 | -0.28439 | 0.821091 | 6.651978 | -7.15686 | 1.86E-10 | 2.48E-09 | 13.46013 |
| SRI | 0.285464 | 1.218802 | 8.487033 | 7.154851 | 1.88E-10 | 2.50E-09 | 13.45091 |
| VCAN | 0.866459 | 1.823182 | 4.839121 | 7.152556 | 1.90E-10 | 2.52E-09 | 13.44035 |
| PDLIM1 | 0.577222 | 1.491974 | 9.35044 | 7.148012 | 1.94E-10 | 2.57E-09 | 13.41946 |
| UBE2D1 | 0.251937 | 1.190805 | 5.021758 | 7.146454 | 1.95E-10 | 2.59E-09 | 13.4123 |
| NFATC4 | -0.20338 | 0.868512 | 5.717253 | -7.14471 | 1.97E-10 | 2.60E-09 | 13.40429 |
| RAB8A | 0.281348 | 1.21533 | 9.10978 | 7.141624 | 2.00E-10 | 2.64E-09 | 13.3901 |
| NPHS1 | -0.57524 | 0.671176 | 5.41989 | -7.14024 | 2.01E-10 | 2.65E-09 | 13.38375 |
| CTSZ | 0.605459 | 1.521463 | 6.065055 | 7.140119 | 2.01E-10 | 2.65E-09 | 13.38318 |
| SHBG | -0.38873 | 0.763799 | 6.134725 | -7.13747 | 2.04E-10 | 2.68E-09 | 13.371 |
| TP53BP1 | 0.246628 | 1.186431 | 6.44022 | 7.132717 | 2.09E-10 | 2.73E-09 | 13.34917 |
| MYOZ1 | -0.257 | 0.836826 | 5.887912 | -7.13158 | 2.10E-10 | 2.74E-09 | 13.34392 |
| ELK1 | -0.20351 | 0.868434 | 6.251319 | -7.13059 | 2.11E-10 | 2.75E-09 | 13.33939 |
| TRIP12 | 0.263329 | 1.200245 | 8.86 | 7.123477 | 2.18E-10 | 2.84E-09 | 13.30672 |
| PPP1R12B | -0.22088 | 0.858039 | 5.600989 | -7.12157 | 2.20E-10 | 2.86E-09 | 13.29796 |
| SLC10A1 | -0.24368 | 0.844588 | 5.303846 | -7.11832 | 2.23E-10 | 2.90E-09 | 13.28304 |
| DNAJB5 | -0.34197 | 0.788963 | 5.372198 | -7.11431 | 2.27E-10 | 2.95E-09 | 13.26463 |
| ALOX5 | 0.469792 | 1.38491 | 4.562967 | 7.106758 | 2.36E-10 | 3.05E-09 | 13.22995 |
| PTGDS | -0.92344 | 0.52725 | 7.868571 | -7.10389 | 2.39E-10 | 3.09E-09 | 13.21678 |
| FSTL1 | 0.657213 | 1.577033 | 8.79956 | 7.103495 | 2.39E-10 | 3.09E-09 | 13.21498 |
| RAE1 | 0.267778 | 1.203952 | 7.502418 | 7.099313 | 2.44E-10 | 3.15E-09 | 13.19579 |
| CEL | -0.52942 | 0.692833 | 5.094286 | -7.09517 | 2.49E-10 | 3.21E-09 | 13.17678 |

| | | | | | | | |
|---|---|---|---|---|---|---|---|
| SOAT1 | 0.360643 | 1.283998 | 5.337253 | 7.0946 | 2.49E-10 | 3.21E-09 | 13.17417 |
| MAGED1 | 0.230444 | 1.173196 | 9.893956 | 7.093651 | 2.50E-10 | 3.22E-09 | 13.16981 |
| CALCOCO1 | -0.26915 | 0.829808 | 6.945165 | -7.09271 | 2.52E-10 | 3.23E-09 | 13.16552 |
| MTPAP | 0.235787 | 1.177549 | 6.837033 | 7.088598 | 2.57E-10 | 3.29E-09 | 13.14663 |
| STT3A | 0.38628 | 1.307019 | 8.273626 | 7.086381 | 2.59E-10 | 3.31E-09 | 13.13647 |
| HSPB8 | 0.828164 | 1.775425 | 8.776923 | 7.086031 | 2.60E-10 | 3.32E-09 | 13.13486 |
| LRRC49 | 0.338628 | 1.264553 | 5.475714 | 7.08406 | 2.62E-10 | 3.34E-09 | 13.12582 |
| SUCO | 0.281647 | 1.215582 | 7.053407 | 7.083407 | 2.63E-10 | 3.35E-09 | 13.12283 |
| CA5B | -0.22547 | 0.855314 | 5.342198 | -7.08144 | 2.65E-10 | 3.37E-09 | 13.11379 |
| GPRASP1 | -0.49052 | 0.711768 | 6.153956 | -7.07296 | 2.76E-10 | 3.50E-09 | 13.07495 |
| MMS19 | 0.256217 | 1.194343 | 6.716484 | 7.067986 | 2.83E-10 | 3.58E-09 | 13.05214 |
| GTF2E1 | 0.30787 | 1.237878 | 7.036374 | 7.063268 | 2.89E-10 | 3.66E-09 | 13.03052 |
| ZNF227 | 0.301633 | 1.232539 | 4.772637 | 7.062889 | 2.89E-10 | 3.66E-09 | 13.02878 |
| DDR1 | 0.352821 | 1.277056 | 8.961429 | 7.060871 | 2.92E-10 | 3.68E-09 | 13.01954 |
| GRPR | -0.26462 | 0.832419 | 6.841319 | -7.06061 | 2.92E-10 | 3.68E-09 | 13.01835 |
| ZWINT | 0.599899 | 1.51561 | 6.772088 | 7.059994 | 2.93E-10 | 3.69E-09 | 13.01552 |
| IGSF3 | 0.336338 | 1.262548 | 7.201538 | 7.055477 | 3.00E-10 | 3.76E-09 | 12.99483 |
| ZNF544 | 0.274111 | 1.209249 | 7.190549 | 7.050148 | 3.07E-10 | 3.85E-09 | 12.97042 |
| CPD | 0.373319 | 1.295329 | 6.883956 | 7.049594 | 3.08E-10 | 3.85E-09 | 12.96789 |
| PPFIBP1 | 0.572671 | 1.487275 | 5.533407 | 7.049525 | 3.08E-10 | 3.85E-09 | 12.96757 |
| PPP4R1 | 0.345242 | 1.270364 | 8.016593 | 7.046138 | 3.13E-10 | 3.91E-09 | 12.95206 |
| IHH | -0.27183 | 0.828271 | 6.453407 | -7.04463 | 3.15E-10 | 3.93E-09 | 12.94515 |
| POLM | -0.27171 | 0.828337 | 5.338681 | -7.0436 | 3.17E-10 | 3.94E-09 | 12.94043 |
| C2 | 0.848295 | 1.800372 | 6.712747 | 7.043243 | 3.17E-10 | 3.94E-09 | 12.93881 |
| EFCC1 | -0.31894 | 0.80166 | 5.14011 | -7.0388 | 3.24E-10 | 4.02E-09 | 12.91848 |
| MID1 | 0.404512 | 1.323641 | 6.393077 | 7.03764 | 3.26E-10 | 4.04E-09 | 12.91316 |
| PHYHIP | -0.31208 | 0.805478 | 5.171978 | -7.03629 | 3.28E-10 | 4.06E-09 | 12.90697 |
| LRRC59 | 0.404802 | 1.323907 | 8.926264 | 7.034923 | 3.30E-10 | 4.08E-09 | 12.90073 |
| UBE2L3 | 0.236546 | 1.178169 | 7.991538 | 7.034223 | 3.31E-10 | 4.08E-09 | 12.89753 |
| DKFZP586I | 0.401425 | 1.320812 | 7.232637 | 7.032705 | 3.33E-10 | 4.11E-09 | 12.89058 |
| CENPU | 0.480126 | 1.394865 | 4.483077 | 7.030544 | 3.37E-10 | 4.14E-09 | 12.88069 |
| SAYSD1 | 0.293343 | 1.225477 | 6.015495 | 7.029482 | 3.38E-10 | 4.16E-09 | 12.87583 |
| MAGI2 | -0.30592 | 0.808927 | 5.212418 | -7.02879 | 3.39E-10 | 4.17E-09 | 12.87264 |
| FBXO21 | -0.53959 | 0.687967 | 9.691648 | -7.02478 | 3.46E-10 | 4.24E-09 | 12.85431 |
| SLC13A2 | -0.56518 | 0.675872 | 7.347473 | -7.01888 | 3.56E-10 | 4.35E-09 | 12.82735 |
| CAPN2 | 0.353667 | 1.277804 | 10.76989 | 7.015753 | 3.61E-10 | 4.41E-09 | 12.81304 |
| PTCRA | -0.21656 | 0.860615 | 6.897692 | -7.01558 | 3.61E-10 | 4.41E-09 | 12.81224 |
| RPP40 | 0.351285 | 1.275696 | 6.641538 | 7.014453 | 3.63E-10 | 4.42E-09 | 12.80709 |
| DNM1L | 0.26701 | 1.203311 | 7.087473 | 7.01275 | 3.66E-10 | 4.45E-09 | 12.79931 |
| HR | -0.23134 | 0.851844 | 5.265385 | -7.01274 | 3.66E-10 | 4.45E-09 | 12.79924 |
| MISP | -0.2859 | 0.820228 | 7.782967 | -7.00981 | 3.71E-10 | 4.50E-09 | 12.78588 |
| MYLIP | 0.446193 | 1.362441 | 7.259341 | 7.006722 | 3.76E-10 | 4.56E-09 | 12.77175 |
| AKAP12 | 0.925309 | 1.899091 | 8.336484 | 7.006173 | 3.77E-10 | 4.56E-09 | 12.76924 |
| ASNS | 0.554048 | 1.4682 | 5.426154 | 7.00597 | 3.78E-10 | 4.56E-09 | 12.76831 |
| KCNN3 | -0.19975 | 0.870699 | 4.482308 | -7.00326 | 3.83E-10 | 4.61E-09 | 12.75594 |
| TOP2B | 0.295155 | 1.227016 | 7.887912 | 7.002851 | 3.83E-10 | 4.62E-09 | 12.75405 |
| SMARCA4 | 0.237758 | 1.179159 | 6.886703 | 7.001752 | 3.85E-10 | 4.63E-09 | 12.74903 |

| | | | | | | | |
|---|---|---|---|---|---|---|---|
| IRF2BP1 | -0.23022 | 0.852506 | 7.336374 | -7.00094 | 3.87E-10 | 4.64E-09 | 12.74533 |
| TPST1 | 0.367164 | 1.289815 | 6.243956 | 7.000319 | 3.88E-10 | 4.65E-09 | 12.74248 |
| FBP1 | -0.60626 | 0.656899 | 11.15868 | -6.99305 | 4.01E-10 | 4.81E-09 | 12.70926 |
| TERT | -0.23058 | 0.852289 | 5.01967 | -6.99209 | 4.03E-10 | 4.81E-09 | 12.7049 |
| GADD45B | -1.11448 | 0.461858 | 8.227363 | -6.992 | 4.03E-10 | 4.81E-09 | 12.70448 |
| MLXIPL | -0.39287 | 0.761611 | 6.786374 | -6.98927 | 4.08E-10 | 4.87E-09 | 12.69199 |
| FRYL | 0.26572 | 1.202236 | 6.458791 | 6.988571 | 4.10E-10 | 4.88E-09 | 12.68882 |
| SEC31B | -0.27902 | 0.824148 | 6.095934 | -6.98767 | 4.11E-10 | 4.89E-09 | 12.68469 |
| USP16 | 0.235522 | 1.177332 | 7.664945 | 6.986752 | 4.13E-10 | 4.91E-09 | 12.68051 |
| NMT2 | 0.434338 | 1.351291 | 6 | 6.984917 | 4.17E-10 | 4.94E-09 | 12.67213 |
| ETNK2 | -0.60797 | 0.656121 | 7.320879 | -6.98484 | 4.17E-10 | 4.94E-09 | 12.67178 |
| NES | -0.40348 | 0.756031 | 6.943736 | -6.98278 | 4.21E-10 | 4.98E-09 | 12.66239 |
| ATG5 | 0.232473 | 1.174847 | 6.846264 | 6.981334 | 4.24E-10 | 5.00E-09 | 12.65577 |
| NEMF | 0.373986 | 1.295928 | 7.044286 | 6.980953 | 4.25E-10 | 5.01E-09 | 12.65403 |
| ALDOC | -0.27977 | 0.823721 | 6.121868 | -6.97894 | 4.29E-10 | 5.05E-09 | 12.64482 |
| NOX5 | -0.22343 | 0.856524 | 5.754945 | -6.97833 | 4.30E-10 | 5.05E-09 | 12.64205 |
| ASPA | -0.63322 | 0.644735 | 9.251868 | -6.97819 | 4.30E-10 | 5.05E-09 | 12.64139 |
| FIP1L1 | 0.251193 | 1.190191 | 5.617912 | 6.977384 | 4.32E-10 | 5.06E-09 | 12.63773 |
| DBN1 | 0.331681 | 1.258479 | 6.81967 | 6.96874 | 4.50E-10 | 5.26E-09 | 12.59828 |
| ENO3 | -0.34404 | 0.78783 | 5.057912 | -6.9671 | 4.53E-10 | 5.30E-09 | 12.59078 |
| AMBN | -0.22577 | 0.85514 | 4.448791 | -6.96198 | 4.64E-10 | 5.42E-09 | 12.56742 |
| GHR | -0.56364 | 0.676594 | 7.557582 | -6.95232 | 4.85E-10 | 5.66E-09 | 12.52338 |
| ESM1 | -0.72445 | 0.605228 | 5.531538 | -6.94877 | 4.93E-10 | 5.75E-09 | 12.50719 |
| COL1A2 | 0.966783 | 1.954477 | 6.713297 | 6.94517 | 5.02E-10 | 5.83E-09 | 12.49078 |
| COMT | 0.214329 | 1.160164 | 8.160769 | 6.945157 | 5.02E-10 | 5.83E-09 | 12.49072 |
| ITGB3 | 0.492362 | 1.406746 | 5.52978 | 6.943343 | 5.06E-10 | 5.87E-09 | 12.48245 |
| EIF4ENIF1 | 0.276865 | 1.211559 | 6.795824 | 6.943126 | 5.07E-10 | 5.87E-09 | 12.48146 |
| SNX2 | 0.271512 | 1.207072 | 8.712308 | 6.938613 | 5.17E-10 | 5.98E-09 | 12.46089 |
| SOD2 | 0.995744 | 1.994109 | 8.080879 | 6.936775 | 5.22E-10 | 6.03E-09 | 12.45252 |
| ACACA | 0.293415 | 1.225538 | 6.628791 | 6.936258 | 5.23E-10 | 6.03E-09 | 12.45016 |
| RGS6 | -0.1862 | 0.878916 | 4.998791 | -6.93346 | 5.30E-10 | 6.10E-09 | 12.43743 |
| COPG2IT1 | -0.30954 | 0.806901 | 4.837802 | -6.93337 | 5.30E-10 | 6.10E-09 | 12.43702 |
| PARP6 | -0.2048 | 0.867658 | 7.352637 | -6.93053 | 5.37E-10 | 6.17E-09 | 12.42407 |
| HECW1 | -0.29934 | 0.812622 | 4.841538 | -6.92686 | 5.47E-10 | 6.27E-09 | 12.40735 |
| PHKA2 | -0.20656 | 0.866604 | 6.142857 | -6.92529 | 5.51E-10 | 6.31E-09 | 12.40018 |
| FAM179B | 0.329488 | 1.256567 | 7.18989 | 6.918773 | 5.68E-10 | 6.49E-09 | 12.37052 |
| ANKRD46 | -0.41813 | 0.748394 | 8.557363 | -6.91577 | 5.76E-10 | 6.58E-09 | 12.35682 |
| SORBS2 | 0.408836 | 1.327614 | 7.70978 | 6.910973 | 5.89E-10 | 6.71E-09 | 12.33501 |
| HLA-J | 0.497473 | 1.411739 | 8.032308 | 6.906579 | 6.01E-10 | 6.84E-09 | 12.31501 |
| CPNE6 | -0.99234 | 0.502662 | 5.694066 | -6.90293 | 6.11E-10 | 6.95E-09 | 12.29842 |
| UBFD1 | 0.398271 | 1.317927 | 6.209121 | 6.901815 | 6.14E-10 | 6.98E-09 | 12.29334 |
| PRL | -0.27317 | 0.827497 | 4.842088 | -6.89898 | 6.22E-10 | 7.06E-09 | 12.28044 |
| NUP62 | 0.315271 | 1.244245 | 5.863077 | 6.897951 | 6.25E-10 | 7.08E-09 | 12.27576 |
| TMEM184 | 0.244 | 1.184272 | 6.960659 | 6.8978 | 6.26E-10 | 7.08E-09 | 12.27507 |
| BET1 | 0.268039 | 1.20417 | 7.974286 | 6.895854 | 6.31E-10 | 7.14E-09 | 12.26622 |
| CTNNA1 | 0.232159 | 1.174592 | 9.651978 | 6.895405 | 6.33E-10 | 7.14E-09 | 12.26417 |
| TCF7L1 | -0.31211 | 0.805462 | 5.490659 | -6.89239 | 6.42E-10 | 7.23E-09 | 12.25048 |

| | | | | | | | |
|---|---|---|---|---|---|---|---|
| TUSC3 | 0.533705 | 1.447642 | 6.530659 | 6.891487 | 6.44E-10 | 7.24E-09 | 12.24636 |
| TCL6 | -0.22739 | 0.854181 | 4.605165 | -6.89138 | 6.45E-10 | 7.24E-09 | 12.24588 |
| WDR82 | 0.250227 | 1.189394 | 8.563956 | 6.891227 | 6.45E-10 | 7.24E-09 | 12.24517 |
| SHC3 | -0.20443 | 0.867884 | 5.49978 | -6.8904 | 6.48E-10 | 7.26E-09 | 12.24142 |
| C1QL1 | -0.28667 | 0.819794 | 6.188242 | -6.88767 | 6.56E-10 | 7.35E-09 | 12.22899 |
| INPP5J | -0.38015 | 0.768355 | 6.735055 | -6.8841 | 6.67E-10 | 7.46E-09 | 12.21276 |
| STAU1 | -0.44343 | 0.735384 | 10.66637 | -6.88195 | 6.74E-10 | 7.52E-09 | 12.203 |
| WNT8B | -0.30744 | 0.808075 | 5.543187 | -6.88182 | 6.74E-10 | 7.52E-09 | 12.20239 |
| CELSR3-AS | -0.243 | 0.844989 | 5.185055 | -6.88102 | 6.77E-10 | 7.54E-09 | 12.19877 |
| GUCY1A3 | 0.500691 | 1.414891 | 7.268681 | 6.879246 | 6.82E-10 | 7.59E-09 | 12.19071 |
| CITED2 | 0.546556 | 1.460594 | 7.185275 | 6.87817 | 6.86E-10 | 7.62E-09 | 12.18581 |
| COMMD8 | 0.287488 | 1.220513 | 9.719121 | 6.877412 | 6.88E-10 | 7.63E-09 | 12.18237 |
| LXN | 0.409082 | 1.327841 | 7.590989 | 6.873455 | 7.01E-10 | 7.77E-09 | 12.16439 |
| MTMR6 | 0.301874 | 1.232745 | 6.293626 | 6.872714 | 7.03E-10 | 7.78E-09 | 12.16102 |
| SLC28A1 | -0.34982 | 0.784681 | 7.055055 | -6.86835 | 7.18E-10 | 7.92E-09 | 12.1412 |
| FYCO1 | 0.271729 | 1.207254 | 6.552198 | 6.868332 | 7.18E-10 | 7.92E-09 | 12.14111 |
| ZWILCH | 0.280106 | 1.214284 | 4.783297 | 6.867727 | 7.20E-10 | 7.93E-09 | 12.13837 |
| XDH | -0.24984 | 0.840989 | 5.373626 | -6.86351 | 7.34E-10 | 8.08E-09 | 12.11919 |
| PPAP2C | 0.454005 | 1.369838 | 4.814725 | 6.861002 | 7.43E-10 | 8.16E-09 | 12.10782 |
| ABI1 | 0.309942 | 1.239658 | 7.500659 | 6.859908 | 7.46E-10 | 8.20E-09 | 12.10285 |
| CBX3 | 0.352556 | 1.27682 | 7.929341 | 6.859073 | 7.49E-10 | 8.22E-09 | 12.09906 |
| CCDC47 | 0.271589 | 1.207137 | 8.455824 | 6.858194 | 7.52E-10 | 8.24E-09 | 12.09507 |
| CCT2 | 0.26944 | 1.20534 | 9.468022 | 6.854533 | 7.65E-10 | 8.36E-09 | 12.07845 |
| PSMD14 | 0.287444 | 1.220476 | 9.857143 | 6.854477 | 7.66E-10 | 8.36E-09 | 12.0782 |
| SUGCT | -0.47216 | 0.720882 | 8.759121 | -6.85388 | 7.68E-10 | 8.37E-09 | 12.0755 |
| MPP2 | -0.23021 | 0.852509 | 5.576593 | -6.85232 | 7.73E-10 | 8.42E-09 | 12.0684 |
| RAB11FIP3 | -0.53713 | 0.68914 | 8.183516 | -6.85195 | 7.75E-10 | 8.43E-09 | 12.06672 |
| TBCB | 0.250198 | 1.18937 | 8.852637 | 6.851164 | 7.77E-10 | 8.45E-09 | 12.06316 |
| CEND1 | -0.23695 | 0.848536 | 6.07 | -6.8501 | 7.81E-10 | 8.48E-09 | 12.05835 |
| ATP1B2 | -0.31087 | 0.806156 | 5.847143 | -6.84901 | 7.85E-10 | 8.51E-09 | 12.05337 |
| THAP1 | 0.357589 | 1.281283 | 5.968352 | 6.848771 | 7.86E-10 | 8.51E-09 | 12.05229 |
| TRPM3 | -0.47432 | 0.719804 | 7.090879 | -6.84845 | 7.87E-10 | 8.51E-09 | 12.05086 |
| MYDGF | 0.265087 | 1.201708 | 8.21 | 6.848025 | 7.89E-10 | 8.52E-09 | 12.04891 |
| ZMYM6NB | 0.296937 | 1.228534 | 8.179011 | 6.847308 | 7.91E-10 | 8.53E-09 | 12.04566 |
| SPTLC2 | 0.262208 | 1.199313 | 6.659011 | 6.846969 | 7.93E-10 | 8.54E-09 | 12.04412 |
| NXPE4 | -0.23939 | 0.847105 | 4.659231 | -6.84629 | 7.95E-10 | 8.55E-09 | 12.04105 |
| CA2 | -0.39384 | 0.761101 | 11.30242 | -6.84595 | 7.96E-10 | 8.56E-09 | 12.03951 |
| RGS2 | -0.93674 | 0.522412 | 7.973516 | -6.84238 | 8.10E-10 | 8.69E-09 | 12.02329 |
| WBP5 | 0.320126 | 1.248439 | 9.693956 | 6.841928 | 8.12E-10 | 8.70E-09 | 12.02124 |
| C1orf95 | -0.26502 | 0.832188 | 5.513077 | -6.84094 | 8.15E-10 | 8.72E-09 | 12.01678 |
| SLC38A3 | -0.37648 | 0.770316 | 5.532308 | -6.8402 | 8.18E-10 | 8.73E-09 | 12.01341 |
| CCRN4L | -0.33216 | 0.794347 | 5.408571 | -6.84014 | 8.18E-10 | 8.73E-09 | 12.01314 |
| PLCB2 | -0.2167 | 0.860534 | 5.801538 | -6.83619 | 8.33E-10 | 8.89E-09 | 11.9952 |
| TAC3 | -0.31342 | 0.804732 | 6.861099 | -6.83526 | 8.37E-10 | 8.91E-09 | 11.99101 |
| ZBED8 | 0.437058 | 1.353841 | 4.573736 | 6.828512 | 8.64E-10 | 9.17E-09 | 11.96039 |
| ADD2 | -0.22576 | 0.855145 | 5.499231 | -6.82851 | 8.64E-10 | 9.17E-09 | 11.96038 |
| KRT4 | -0.26364 | 0.832985 | 5.365934 | -6.8254 | 8.76E-10 | 9.29E-09 | 11.9463 |

| | | | | | | | |
|---|---|---|---|---|---|---|---|
| BSN | -0.25605 | 0.837376 | 4.780989 | -6.82472 | 8.79E-10 | 9.31E-09 | 11.94322 |
| NUP43 | 0.355232 | 1.279191 | 6.851099 | 6.823095 | 8.86E-10 | 9.37E-09 | 11.93583 |
| STX3 | 0.436029 | 1.352875 | 7.306923 | 6.821696 | 8.91E-10 | 9.42E-09 | 11.92949 |
| YIPF3 | 0.238309 | 1.179609 | 7.191758 | 6.821043 | 8.94E-10 | 9.44E-09 | 11.92653 |
| MAD2L1 | 0.38458 | 1.305479 | 4.276264 | 6.819176 | 9.02E-10 | 9.51E-09 | 11.91807 |
| BHMT2 | -0.37571 | 0.770723 | 11.41703 | -6.81777 | 9.08E-10 | 9.56E-09 | 11.91169 |
| RRAS | 0.503135 | 1.41729 | 6.34989 | 6.816005 | 9.15E-10 | 9.62E-09 | 11.9037 |
| DDOST | 0.241729 | 1.182409 | 10.15736 | 6.812716 | 9.29E-10 | 9.76E-09 | 11.8888 |
| GARS | 0.321768 | 1.249861 | 9.028681 | 6.811372 | 9.35E-10 | 9.81E-09 | 11.8827 |
| NBN | 0.279652 | 1.213902 | 6.772637 | 6.810447 | 9.39E-10 | 9.84E-09 | 11.87852 |
| VAMP8 | 0.293923 | 1.225969 | 10.55187 | 6.809918 | 9.42E-10 | 9.85E-09 | 11.87612 |
| NOS1AP | -0.26388 | 0.832845 | 5.834945 | -6.80932 | 9.44E-10 | 9.87E-09 | 11.8734 |
| GRAMD1B | -0.58703 | 0.66571 | 7.713187 | -6.80881 | 9.46E-10 | 9.87E-09 | 11.87108 |
| CYB5A | -0.26971 | 0.829486 | 11.91967 | -6.80868 | 9.47E-10 | 9.87E-09 | 11.87049 |
| SSR1 | 0.251275 | 1.190259 | 7.951648 | 6.807599 | 9.52E-10 | 9.91E-09 | 11.86562 |
| PLTP | 1.242227 | 2.365634 | 7.494505 | 6.807115 | 9.54E-10 | 9.92E-09 | 11.86342 |
| UBE2C | 0.680797 | 1.603025 | 5.782527 | 6.805695 | 9.60E-10 | 9.97E-09 | 11.85699 |
| MAP1A | -0.23629 | 0.848926 | 5.58011 | -6.80363 | 9.69E-10 | 1.01E-08 | 11.84763 |
| CDK8 | 0.261348 | 1.198598 | 5.45989 | 6.798395 | 9.93E-10 | 1.03E-08 | 11.82393 |
| EIF4E2 | 0.343019 | 1.268408 | 8.135495 | 6.797545 | 9.97E-10 | 1.03E-08 | 11.82008 |
| ABCG1 | 0.526614 | 1.440544 | 5.918022 | 6.796297 | 1.00E-09 | 1.04E-08 | 11.81443 |
| MGAT3 | -0.21194 | 0.863374 | 5.207802 | -6.79606 | 1.00E-09 | 1.04E-08 | 11.81334 |
| PTPN12 | 0.318802 | 1.247294 | 5.493736 | 6.795215 | 1.01E-09 | 1.04E-08 | 11.80953 |
| TUBA4B | 0.375024 | 1.296861 | 6.581538 | 6.794745 | 1.01E-09 | 1.04E-08 | 11.80741 |
| AREL1 | 0.300971 | 1.231973 | 7.122527 | 6.794723 | 1.01E-09 | 1.04E-08 | 11.80731 |
| GLT8D1 | 0.215498 | 1.161104 | 7.743956 | 6.794611 | 1.01E-09 | 1.04E-08 | 11.8068 |
| EIF3J | 0.240923 | 1.181748 | 6.750659 | 6.793989 | 1.01E-09 | 1.04E-08 | 11.80399 |
| CTBP2 | 0.292024 | 1.224357 | 7.345495 | 6.793886 | 1.01E-09 | 1.04E-08 | 11.80352 |
| LARS2 | 0.348106 | 1.272889 | 7.246923 | 6.793356 | 1.02E-09 | 1.04E-08 | 11.80112 |
| EGR1 | -1.17424 | 0.443117 | 7.078462 | -6.7893 | 1.04E-09 | 1.06E-08 | 11.78275 |
| BIRC2 | 0.249401 | 1.188713 | 9.356374 | 6.788634 | 1.04E-09 | 1.06E-08 | 11.77975 |
| GJB4 | -0.31839 | 0.801964 | 5.634945 | -6.78857 | 1.04E-09 | 1.06E-08 | 11.77948 |
| HERC6 | 0.708812 | 1.634457 | 6.357033 | 6.786745 | 1.05E-09 | 1.07E-08 | 11.7712 |
| PCLO | 0.296556 | 1.228209 | 5.191648 | 6.785935 | 1.05E-09 | 1.07E-08 | 11.76753 |
| CASP1 | 0.55757 | 1.471788 | 4.986374 | 6.785123 | 1.06E-09 | 1.07E-08 | 11.76386 |
| SPINT1 | 0.335865 | 1.262134 | 7.74 | 6.783912 | 1.06E-09 | 1.08E-08 | 11.75838 |
| MFSD1 | 0.234333 | 1.176363 | 9.520879 | 6.783512 | 1.06E-09 | 1.08E-08 | 11.75657 |
| COPZ2 | -0.28524 | 0.820607 | 7.613297 | -6.78048 | 1.08E-09 | 1.09E-08 | 11.74283 |
| SAC3D1 | 0.353179 | 1.277372 | 5.597692 | 6.779524 | 1.08E-09 | 1.10E-08 | 11.73853 |
| ROCK1 | 0.356169 | 1.280022 | 6.423736 | 6.778335 | 1.09E-09 | 1.10E-08 | 11.73315 |
| STX4 | 0.284551 | 1.218031 | 7.965495 | 6.777071 | 1.10E-09 | 1.11E-08 | 11.72743 |
| KIAA0355 | 0.243517 | 1.183875 | 6.968681 | 6.776848 | 1.10E-09 | 1.11E-08 | 11.72642 |
| SLC25A28 | -0.20111 | 0.86988 | 6.940549 | -6.77317 | 1.12E-09 | 1.12E-08 | 11.70977 |
| SOCS2 | -0.75105 | 0.59417 | 6.711209 | -6.77163 | 1.12E-09 | 1.13E-08 | 11.70281 |
| ADSL | 0.220647 | 1.165256 | 8.948242 | 6.771107 | 1.13E-09 | 1.13E-08 | 11.70046 |
| BUD31 | 0.287947 | 1.220902 | 8.13 | 6.771055 | 1.13E-09 | 1.13E-08 | 11.70022 |
| SLC43A1 | -0.39221 | 0.761963 | 5.516703 | -6.76885 | 1.14E-09 | 1.14E-08 | 11.69025 |

| | | | | | | | |
|---|---|---|---|---|---|---|---|
| TEX40 | -0.27613 | 0.825803 | 5.407582 | -6.76817 | 1.14E-09 | 1.14E-08 | 11.68718 |
| AHCY | -0.43361 | 0.740405 | 10.12297 | -6.76668 | 1.15E-09 | 1.15E-08 | 11.68044 |
| DENND2D | 0.438792 | 1.355469 | 7.352637 | 6.758578 | 1.19E-09 | 1.19E-08 | 11.64382 |
| RFXANK | 0.268039 | 1.20417 | 7.194286 | 6.758168 | 1.20E-09 | 1.19E-08 | 11.64197 |
| HARS2 | 0.241072 | 1.181871 | 7.437473 | 6.757617 | 1.20E-09 | 1.19E-08 | 11.63948 |
| PART1 | -0.24864 | 0.841691 | 4.858352 | -6.757 | 1.20E-09 | 1.20E-08 | 11.63671 |
| EEF1E1 | 0.329908 | 1.256933 | 9.139011 | 6.755235 | 1.21E-09 | 1.21E-08 | 11.62872 |
| PROP1 | -0.27635 | 0.825676 | 5.217473 | -6.75463 | 1.22E-09 | 1.21E-08 | 11.626 |
| TRPV1 | -0.2765 | 0.82559 | 5.390659 | -6.75338 | 1.22E-09 | 1.21E-08 | 11.62036 |
| SYN1 | -0.24432 | 0.844214 | 5.154835 | -6.75082 | 1.24E-09 | 1.23E-08 | 11.60879 |
| CCT5 | 0.327275 | 1.254642 | 9.059231 | 6.749299 | 1.25E-09 | 1.23E-08 | 11.6019 |
| YARS | 0.228628 | 1.17172 | 8.491319 | 6.748496 | 1.25E-09 | 1.24E-08 | 11.59828 |
| DAP3 | 0.219643 | 1.164445 | 9.852527 | 6.746173 | 1.26E-09 | 1.25E-08 | 11.58778 |
| SCRN1 | 0.452454 | 1.368366 | 7.424176 | 6.746112 | 1.27E-09 | 1.25E-08 | 11.58751 |
| DDIT3 | -0.3904 | 0.762918 | 7.648901 | -6.74581 | 1.27E-09 | 1.25E-08 | 11.58612 |
| CAPN1 | 0.272662 | 1.208035 | 6.574615 | 6.744912 | 1.27E-09 | 1.25E-08 | 11.58209 |
| IFT46 | -0.249 | 0.841477 | 7.661648 | -6.74137 | 1.29E-09 | 1.27E-08 | 11.56611 |
| LMAN2 | 0.260169 | 1.197619 | 8.176264 | 6.741295 | 1.29E-09 | 1.27E-08 | 11.56576 |
| HKDC1 | 0.482246 | 1.396917 | 5.06956 | 6.741094 | 1.30E-09 | 1.27E-08 | 11.56485 |
| AMHR2 | -0.25748 | 0.836549 | 5.756154 | -6.741 | 1.30E-09 | 1.27E-08 | 11.56444 |
| UBP1 | 0.275792 | 1.210659 | 7.027033 | 6.740728 | 1.30E-09 | 1.27E-08 | 11.5632 |
| CANT1 | 0.194522 | 1.144345 | 6.75967 | 6.739792 | 1.30E-09 | 1.27E-08 | 11.55897 |
| MPZ | -0.24685 | 0.842737 | 5.29989 | -6.73694 | 1.32E-09 | 1.29E-08 | 11.54612 |
| TSKU | -0.66468 | 0.630828 | 8.240659 | -6.73066 | 1.36E-09 | 1.32E-08 | 11.51773 |
| PSMG2 | 0.219874 | 1.164632 | 9.733077 | 6.729224 | 1.37E-09 | 1.33E-08 | 11.51127 |
| ST6GALNA | 0.325652 | 1.253231 | 5.255385 | 6.728039 | 1.38E-09 | 1.34E-08 | 11.50592 |
| MYO7A | -0.25228 | 0.839568 | 5.472637 | -6.72756 | 1.38E-09 | 1.34E-08 | 11.50378 |
| AACS | 0.253454 | 1.192058 | 7.260769 | 6.726251 | 1.39E-09 | 1.34E-08 | 11.49786 |
| ATP13A1 | 0.28587 | 1.219145 | 6.875495 | 6.725395 | 1.39E-09 | 1.35E-08 | 11.494 |
| DHRS12 | -0.18901 | 0.877208 | 6.271099 | -6.72343 | 1.41E-09 | 1.36E-08 | 11.48512 |
| HRSP12 | -0.42791 | 0.743336 | 11.82231 | -6.72274 | 1.41E-09 | 1.36E-08 | 11.48202 |
| CTNNA3 | -0.22486 | 0.855675 | 4.78989 | -6.72213 | 1.41E-09 | 1.36E-08 | 11.47925 |
| LRRC23 | -0.31751 | 0.802455 | 6.365165 | -6.72061 | 1.42E-09 | 1.37E-08 | 11.47242 |
| DLGAP2 | -0.27544 | 0.826196 | 4.918791 | -6.7199 | 1.43E-09 | 1.38E-08 | 11.4692 |
| AOC4P | -0.20733 | 0.86614 | 4.667692 | -6.71814 | 1.44E-09 | 1.38E-08 | 11.46129 |
| CDIPT | 0.265942 | 1.202421 | 10.8089 | 6.717883 | 1.44E-09 | 1.39E-08 | 11.46011 |
| HLA-E | 0.430155 | 1.347378 | 9.79967 | 6.715652 | 1.46E-09 | 1.40E-08 | 11.45005 |
| POLR2G | 0.282362 | 1.216185 | 9.405934 | 6.712598 | 1.48E-09 | 1.42E-08 | 11.43628 |
| F11 | -0.42983 | 0.742351 | 6.825275 | -6.71214 | 1.48E-09 | 1.42E-08 | 11.43423 |
| STK24 | 0.172546 | 1.127046 | 9.97989 | 6.711198 | 1.49E-09 | 1.42E-08 | 11.42996 |
| ZCWPW1 | -0.20371 | 0.868318 | 6.182527 | -6.71072 | 1.49E-09 | 1.42E-08 | 11.42783 |
| SGK1 | -0.70613 | 0.612962 | 10.63495 | -6.7104 | 1.49E-09 | 1.42E-08 | 11.42636 |
| SGMS1 | 0.443459 | 1.359861 | 6.284945 | 6.705209 | 1.53E-09 | 1.46E-08 | 11.40297 |
| SLFN12 | 0.347092 | 1.271994 | 4.480769 | 6.702895 | 1.55E-09 | 1.47E-08 | 11.39254 |
| NDC1 | 0.259454 | 1.197026 | 5.243736 | 6.701941 | 1.55E-09 | 1.48E-08 | 11.38824 |
| NET1 | -0.46073 | 0.726619 | 8.439341 | -6.70147 | 1.56E-09 | 1.48E-08 | 11.38614 |
| ALDH3B1 | -0.26754 | 0.830737 | 6.658571 | -6.70062 | 1.56E-09 | 1.48E-08 | 11.3823 |

| | | | | | | | |
|---|---|---|---|---|---|---|---|
| CACNA1F | -0.28051 | 0.823301 | 5.698462 | -6.70061 | 1.56E-09 | 1.48E-08 | 11.38225 |
| RRM2 | 1.041217 | 2.057963 | 4.87967 | 6.700323 | 1.56E-09 | 1.48E-08 | 11.38095 |
| RNF7 | 0.282372 | 1.216193 | 7.076374 | 6.697323 | 1.59E-09 | 1.50E-08 | 11.36743 |
| ASB9 | -0.67697 | 0.625477 | 7.061538 | -6.6884 | 1.65E-09 | 1.56E-08 | 11.32725 |
| ARHGAP28 | -0.25207 | 0.839689 | 4.482088 | -6.68831 | 1.65E-09 | 1.56E-08 | 11.32685 |
| ZC3H7A | 0.301507 | 1.232431 | 6.386923 | 6.685366 | 1.68E-09 | 1.58E-08 | 11.31359 |
| TRIM22 | 0.820372 | 1.765861 | 7.522418 | 6.68476 | 1.68E-09 | 1.58E-08 | 11.31085 |
| TRIM5 | 0.285986 | 1.219243 | 4.360769 | 6.681621 | 1.70E-09 | 1.60E-08 | 11.29672 |
| STK19 | 0.706227 | 1.631532 | 7.599451 | 6.680489 | 1.71E-09 | 1.61E-08 | 11.29163 |
| MANF | 0.416232 | 1.334438 | 9.306264 | 6.679791 | 1.72E-09 | 1.61E-08 | 11.28849 |
| GORASP2 | 0.218952 | 1.163888 | 8.291099 | 6.679232 | 1.72E-09 | 1.62E-08 | 11.28597 |
| MPHOSPH | 0.220812 | 1.165389 | 8.155714 | 6.675376 | 1.75E-09 | 1.64E-08 | 11.26862 |
| ALG9 | 0.24814 | 1.187675 | 6.709011 | 6.67432 | 1.76E-09 | 1.65E-08 | 11.26387 |
| PARD3 | 0.246184 | 1.186065 | 7.33 | 6.674317 | 1.76E-09 | 1.65E-08 | 11.26385 |
| OPN3 | 0.28628 | 1.219492 | 7.964176 | 6.671746 | 1.78E-09 | 1.66E-08 | 11.25229 |
| STK39 | 0.402609 | 1.321896 | 5.506484 | 6.671671 | 1.78E-09 | 1.66E-08 | 11.25195 |
| ZNHIT6 | 0.273575 | 1.208799 | 5.716154 | 6.669293 | 1.80E-09 | 1.68E-08 | 11.24125 |
| GNG7 | -0.24477 | 0.843951 | 6.334396 | -6.66904 | 1.81E-09 | 1.68E-08 | 11.24012 |
| GBAS | 0.360242 | 1.283641 | 8.609011 | 6.668543 | 1.81E-09 | 1.68E-08 | 11.23787 |
| MED7 | 0.223802 | 1.167807 | 7.011758 | 6.667527 | 1.82E-09 | 1.69E-08 | 11.2333 |
| S100A1 | 0.674966 | 1.596559 | 7.687253 | 6.666156 | 1.83E-09 | 1.70E-08 | 11.22714 |
| PRKCD | -0.38241 | 0.767155 | 6.252418 | -6.66601 | 1.83E-09 | 1.70E-08 | 11.22648 |
| NPFFR1 | -0.23 | 0.852635 | 5.586264 | -6.66048 | 1.88E-09 | 1.74E-08 | 11.20161 |
| BAG5 | 0.227333 | 1.170669 | 7.392418 | 6.658669 | 1.89E-09 | 1.75E-08 | 11.19347 |
| MAST2 | -0.20159 | 0.869589 | 6.318571 | -6.65849 | 1.90E-09 | 1.75E-08 | 11.19265 |
| SDF2L1 | 0.432536 | 1.349604 | 7.948022 | 6.655638 | 1.92E-09 | 1.77E-08 | 11.17984 |
| HCCS | 0.219164 | 1.164059 | 7.730769 | 6.655049 | 1.93E-09 | 1.77E-08 | 11.17719 |
| PIGN | 0.268749 | 1.204763 | 5.956593 | 6.65352 | 1.94E-09 | 1.78E-08 | 11.17032 |
| OSMR | 0.615097 | 1.531661 | 5.213516 | 6.652908 | 1.95E-09 | 1.79E-08 | 11.16757 |
| SSTR2 | -0.37389 | 0.7717 | 5.09011 | -6.64966 | 1.97E-09 | 1.81E-08 | 11.15296 |
| BTNL3 | -0.28005 | 0.823563 | 6.439341 | -6.64946 | 1.98E-09 | 1.81E-08 | 11.15209 |
| SMC4 | 0.363657 | 1.286683 | 5.604396 | 6.648868 | 1.98E-09 | 1.82E-08 | 11.14941 |
| DEFB1 | -0.64568 | 0.639191 | 12.00505 | -6.64784 | 1.99E-09 | 1.82E-08 | 11.14477 |
| TMEM251 | 0.297633 | 1.229126 | 7.700659 | 6.646321 | 2.01E-09 | 1.83E-08 | 11.13797 |
| CLPS | -0.24273 | 0.845145 | 4.557143 | -6.6452 | 2.02E-09 | 1.84E-08 | 11.13294 |
| CNPY2 | 0.182304 | 1.134695 | 8.923846 | 6.642437 | 2.04E-09 | 1.86E-08 | 11.12052 |
| EMC8 | 0.24185 | 1.182508 | 7.842857 | 6.64078 | 2.06E-09 | 1.87E-08 | 11.11307 |
| INSR | -0.32797 | 0.796659 | 7.559341 | -6.64022 | 2.06E-09 | 1.88E-08 | 11.11057 |
| RASL10A | -0.23328 | 0.850701 | 5.337253 | -6.63821 | 2.08E-09 | 1.89E-08 | 11.10152 |
| NAT10 | 0.232725 | 1.175052 | 6.957692 | 6.636191 | 2.10E-09 | 1.91E-08 | 11.09246 |
| OCA2 | -0.24594 | 0.843265 | 6.010989 | -6.6314 | 2.15E-09 | 1.95E-08 | 11.07094 |
| KRT81 | -0.2623 | 0.833755 | 5.706593 | -6.62952 | 2.17E-09 | 1.96E-08 | 11.06251 |
| RPGRIP1L | 0.256585 | 1.194647 | 4.093187 | 6.627056 | 2.19E-09 | 1.98E-08 | 11.05144 |
| SENP2 | -0.23665 | 0.848715 | 6.963846 | -6.62704 | 2.19E-09 | 1.98E-08 | 11.05137 |
| TMCO3 | 0.383155 | 1.30419 | 6.511429 | 6.626135 | 2.20E-09 | 1.99E-08 | 11.04731 |
| PIBF1 | 0.329483 | 1.256563 | 5.13967 | 6.625569 | 2.21E-09 | 1.99E-08 | 11.04477 |
| PLA1A | 0.548314 | 1.462376 | 7.055275 | 6.625293 | 2.21E-09 | 1.99E-08 | 11.04353 |

| | | | | | | | |
|---|---|---|---|---|---|---|---|
| LARP7 | 0.297763 | 1.229237 | 7.596593 | 6.624407 | 2.22E-09 | 2.00E-08 | 11.03956 |
| APOBEC3B | 0.936058 | 1.913293 | 4.585495 | 6.623425 | 2.23E-09 | 2.00E-08 | 11.03515 |
| S100A4 | 0.675159 | 1.596773 | 7.746044 | 6.62325 | 2.23E-09 | 2.00E-08 | 11.03437 |
| NUP133 | 0.239903 | 1.180914 | 7.254286 | 6.623107 | 2.23E-09 | 2.00E-08 | 11.03372 |
| QPCTL | -0.18785 | 0.877913 | 4.263846 | -6.62235 | 2.24E-09 | 2.01E-08 | 11.03033 |
| CD81 | 0.349338 | 1.273976 | 10.19297 | 6.619777 | 2.27E-09 | 2.03E-08 | 11.01878 |
| CDKN1C | -0.47563 | 0.719151 | 8.321319 | -6.61888 | 2.28E-09 | 2.04E-08 | 11.01478 |
| MSH2 | 0.355522 | 1.279448 | 5.454286 | 6.617726 | 2.29E-09 | 2.04E-08 | 11.00958 |
| SLC35A5 | 0.261517 | 1.198738 | 7.147582 | 6.616996 | 2.30E-09 | 2.05E-08 | 11.0063 |
| BEST1 | -0.26299 | 0.833362 | 5.425604 | -6.61565 | 2.31E-09 | 2.06E-08 | 11.00024 |
| VRK2 | 0.25985 | 1.197354 | 7.051758 | 6.615097 | 2.32E-09 | 2.06E-08 | 10.99778 |
| ALOXE3 | -0.17763 | 0.884153 | 4.558681 | -6.61477 | 2.32E-09 | 2.06E-08 | 10.99634 |
| WIF1 | -0.33781 | 0.791241 | 4.121429 | -6.61209 | 2.35E-09 | 2.09E-08 | 10.98428 |
| SEC61B | 0.219903 | 1.164656 | 10.9944 | 6.611078 | 2.36E-09 | 2.09E-08 | 10.97976 |
| ANXA1 | 0.815242 | 1.759593 | 7.219011 | 6.610988 | 2.36E-09 | 2.09E-08 | 10.97935 |
| FRMPD4 | -0.19983 | 0.870656 | 4.539011 | -6.61023 | 2.37E-09 | 2.10E-08 | 10.97597 |
| RC3H2 | 0.193295 | 1.143372 | 6.233846 | 6.609708 | 2.37E-09 | 2.10E-08 | 10.97361 |
| EDF1 | 0.216527 | 1.161933 | 9.510769 | 6.609056 | 2.38E-09 | 2.11E-08 | 10.97069 |
| CSTF2T | 0.262068 | 1.199196 | 6.502637 | 6.606489 | 2.41E-09 | 2.13E-08 | 10.95918 |
| DHX40 | 0.304353 | 1.234864 | 7.966374 | 6.605982 | 2.41E-09 | 2.13E-08 | 10.9569 |
| REV3L | 0.307188 | 1.237294 | 5.805385 | 6.605197 | 2.42E-09 | 2.14E-08 | 10.95338 |
| OR10H3 | -0.2921 | 0.816714 | 4.511209 | -6.60393 | 2.44E-09 | 2.15E-08 | 10.94773 |
| LOXL1 | 0.606367 | 1.522421 | 5.676374 | 6.603038 | 2.45E-09 | 2.15E-08 | 10.9437 |
| ZNF468 | 0.346879 | 1.271807 | 5.471099 | 6.601353 | 2.47E-09 | 2.17E-08 | 10.93615 |
| ASS1 | -0.48231 | 0.715831 | 12.43758 | -6.59847 | 2.50E-09 | 2.19E-08 | 10.92321 |
| PRSS3 | -0.52646 | 0.694257 | 6.749011 | -6.59569 | 2.53E-09 | 2.22E-08 | 10.91078 |
| RBM12 | 0.281507 | 1.215464 | 6.267033 | 6.594287 | 2.55E-09 | 2.23E-08 | 10.90448 |
| BAIAP3 | -0.22061 | 0.858203 | 6.543516 | -6.59225 | 2.57E-09 | 2.25E-08 | 10.89536 |
| ACTL8 | -0.30285 | 0.810652 | 6.712198 | -6.59126 | 2.58E-09 | 2.26E-08 | 10.89091 |
| PPT1 | 0.266599 | 1.202969 | 9.838791 | 6.585172 | 2.66E-09 | 2.32E-08 | 10.86365 |
| SPOCK2 | -0.39502 | 0.760477 | 6.688571 | -6.5824 | 2.69E-09 | 2.35E-08 | 10.85122 |
| LY75 | 0.820633 | 1.766181 | 5.184286 | 6.581058 | 2.71E-09 | 2.36E-08 | 10.84522 |
| HPSE2 | -0.28094 | 0.823053 | 5.078681 | -6.57851 | 2.74E-09 | 2.38E-08 | 10.83384 |
| PNMA3 | -0.25845 | 0.835986 | 5.501978 | -6.57778 | 2.75E-09 | 2.39E-08 | 10.83055 |
| UBE2J1 | 0.230976 | 1.173629 | 6.438132 | 6.577733 | 2.75E-09 | 2.39E-08 | 10.83034 |
| GK | -0.53117 | 0.691991 | 8.864505 | -6.57414 | 2.79E-09 | 2.43E-08 | 10.81425 |
| CTSD | 0.421256 | 1.339093 | 8.494835 | 6.573279 | 2.81E-09 | 2.43E-08 | 10.8104 |
| UCHL1 | 0.74358 | 1.674325 | 8.808791 | 6.571891 | 2.82E-09 | 2.45E-08 | 10.80419 |
| FGFBP1 | -0.26503 | 0.832179 | 5.972418 | -6.56579 | 2.90E-09 | 2.51E-08 | 10.77689 |
| PDCD10 | 0.253894 | 1.192421 | 9.850769 | 6.565294 | 2.91E-09 | 2.52E-08 | 10.77467 |
| MTIF2 | 0.252778 | 1.191499 | 8.1 | 6.56465 | 2.92E-09 | 2.52E-08 | 10.77179 |
| CLIC5 | -0.74978 | 0.594693 | 6.419011 | -6.56371 | 2.93E-09 | 2.53E-08 | 10.7676 |
| TNS4 | -0.24897 | 0.841496 | 5.573187 | -6.56362 | 2.93E-09 | 2.53E-08 | 10.76717 |
| RAB3A | -0.20769 | 0.865925 | 5.161429 | -6.56321 | 2.94E-09 | 2.53E-08 | 10.76533 |
| EED | 0.237401 | 1.178867 | 4.61044 | 6.561195 | 2.97E-09 | 2.55E-08 | 10.75633 |
| ERMP1 | 0.322792 | 1.250749 | 8.435275 | 6.560069 | 2.98E-09 | 2.56E-08 | 10.7513 |
| ZNF358 | -0.2719 | 0.828226 | 6.17989 | -6.55985 | 2.98E-09 | 2.56E-08 | 10.7503 |

| | | | | | | | |
|---|---|---|---|---|---|---|---|
| MAVS | 0.299068 | 1.230349 | 6.055934 | 6.557315 | 3.02E-09 | 2.59E-08 | 10.73898 |
| RNF208 | -0.26234 | 0.833733 | 5.574835 | -6.55452 | 3.06E-09 | 2.62E-08 | 10.72647 |
| PLA2G3 | -0.22835 | 0.853609 | 5.241209 | -6.55313 | 3.08E-09 | 2.63E-08 | 10.72028 |
| LRRC15 | -0.19576 | 0.873114 | 5.374066 | -6.55088 | 3.11E-09 | 2.66E-08 | 10.71021 |
| GSTP1 | 0.480111 | 1.394851 | 9.082418 | 6.546255 | 3.18E-09 | 2.71E-08 | 10.68954 |
| ICE1 | 0.304188 | 1.234724 | 7.578901 | 6.546012 | 3.18E-09 | 2.71E-08 | 10.68845 |
| LY96 | 0.951029 | 1.933251 | 5.026593 | 6.545037 | 3.19E-09 | 2.72E-08 | 10.68409 |
| XPOT | 0.263643 | 1.200506 | 8.034286 | 6.541093 | 3.25E-09 | 2.77E-08 | 10.66647 |
| CECR1 | 0.763459 | 1.697556 | 7.443187 | 6.532449 | 3.38E-09 | 2.88E-08 | 10.62786 |
| CSNK2A1 | 0.181459 | 1.13403 | 7.665385 | 6.532117 | 3.39E-09 | 2.88E-08 | 10.62638 |
| HAP1 | -0.1706 | 0.888471 | 5.878462 | -6.53104 | 3.41E-09 | 2.89E-08 | 10.62155 |
| EFTUD1 | 0.231705 | 1.174222 | 7.361319 | 6.530169 | 3.42E-09 | 2.90E-08 | 10.61767 |
| PAK3 | -0.26063 | 0.834724 | 5.172857 | -6.52689 | 3.47E-09 | 2.94E-08 | 10.60302 |
| GLI1 | -0.24892 | 0.841527 | 5.535604 | -6.52608 | 3.48E-09 | 2.95E-08 | 10.59941 |
| DUSP2 | -0.30726 | 0.808177 | 5.191538 | -6.52546 | 3.49E-09 | 2.95E-08 | 10.59666 |
| YEATS2 | 0.272198 | 1.207646 | 5.213516 | 6.52345 | 3.53E-09 | 2.98E-08 | 10.58768 |
| DIP2C | -0.4853 | 0.714349 | 8.421538 | -6.5231 | 3.53E-09 | 2.98E-08 | 10.5861 |
| MLEC | 0.292159 | 1.224472 | 8.761648 | 6.519401 | 3.59E-09 | 3.03E-08 | 10.56961 |
| CDC23 | 0.296676 | 1.228311 | 7.347143 | 6.517858 | 3.62E-09 | 3.05E-08 | 10.56272 |
| DCAF6 | 0.252353 | 1.191148 | 7.710659 | 6.517368 | 3.62E-09 | 3.05E-08 | 10.56054 |
| EXTL1 | -0.20463 | 0.867762 | 4.770549 | -6.51676 | 3.64E-09 | 3.05E-08 | 10.55781 |
| SYNGR2 | 0.28056 | 1.214667 | 8.833956 | 6.516346 | 3.64E-09 | 3.06E-08 | 10.55598 |
| POLR2L | 0.206097 | 1.153563 | 7.406264 | 6.515168 | 3.66E-09 | 3.07E-08 | 10.55072 |
| RPS19 | 0.284087 | 1.217639 | 12.8444 | 6.514903 | 3.67E-09 | 3.07E-08 | 10.54954 |
| CTSS | 0.780242 | 1.717418 | 5.176703 | 6.511397 | 3.73E-09 | 3.12E-08 | 10.5339 |
| COMMD3 | 0.286783 | 1.219917 | 8.997033 | 6.511248 | 3.73E-09 | 3.12E-08 | 10.53324 |
| SERPINA3 | 1.787966 | 3.453277 | 8.262637 | 6.510949 | 3.73E-09 | 3.12E-08 | 10.5319 |
| SMARCAL1 | 0.279271 | 1.213581 | 6.445275 | 6.510696 | 3.74E-09 | 3.12E-08 | 10.53077 |
| ITGB8 | 0.379836 | 1.301194 | 4.68044 | 6.508463 | 3.78E-09 | 3.14E-08 | 10.52081 |
| PCNA | 0.357517 | 1.281219 | 8.755055 | 6.508458 | 3.78E-09 | 3.14E-08 | 10.52079 |
| ADIRF | -0.8808 | 0.543067 | 9.848571 | -6.5052 | 3.83E-09 | 3.19E-08 | 10.50625 |
| RHOA | 0.279348 | 1.213646 | 10.85879 | 6.504066 | 3.85E-09 | 3.20E-08 | 10.50121 |
| M6PR | 0.317217 | 1.245925 | 8.111648 | 6.503065 | 3.87E-09 | 3.21E-08 | 10.49674 |
| TRIM44 | 0.230435 | 1.173188 | 8.553516 | 6.500223 | 3.92E-09 | 3.25E-08 | 10.48408 |
| POLD3 | -0.54137 | 0.687119 | 7.560769 | -6.49919 | 3.94E-09 | 3.26E-08 | 10.47946 |
| CDX1 | -0.23068 | 0.852232 | 6.605275 | -6.4979 | 3.96E-09 | 3.28E-08 | 10.47371 |
| SLC30A5 | 0.223271 | 1.167377 | 7.067582 | 6.497402 | 3.97E-09 | 3.29E-08 | 10.4715 |
| CKAP2 | 0.282488 | 1.216291 | 4.601648 | 6.496622 | 3.99E-09 | 3.29E-08 | 10.46802 |
| ITIH4 | -0.23216 | 0.85136 | 5.318022 | -6.49541 | 4.01E-09 | 3.31E-08 | 10.46264 |
| FRK | 0.33286 | 1.259508 | 5.123297 | 6.494396 | 4.03E-09 | 3.32E-08 | 10.4581 |
| ZNF573 | 0.335126 | 1.261487 | 5.036374 | 6.493466 | 4.04E-09 | 3.33E-08 | 10.45396 |
| MIR22HG | -0.57666 | 0.670516 | 8.495275 | -6.49046 | 4.10E-09 | 3.37E-08 | 10.44058 |
| IFT52 | 0.238029 | 1.17938 | 7.879011 | 6.490223 | 4.10E-09 | 3.37E-08 | 10.43951 |
| SYT2 | -0.25651 | 0.837109 | 6.62011 | -6.49016 | 4.11E-09 | 3.37E-08 | 10.43923 |
| EPCAM | 0.289435 | 1.222161 | 10.32769 | 6.490083 | 4.11E-09 | 3.37E-08 | 10.43888 |
| IFI35 | 0.466628 | 1.381876 | 7.419011 | 6.48878 | 4.13E-09 | 3.39E-08 | 10.43308 |
| LGALS1 | 0.852159 | 1.805201 | 8.508571 | 6.484886 | 4.21E-09 | 3.44E-08 | 10.41574 |

| | | | | | | | |
|---|---|---|---|---|---|---|---|
| ENO1 | 0.293333 | 1.225468 | 10.64505 | 6.483898 | 4.22E-09 | 3.46E-08 | 10.41133 |
| EIF3A | 0.226237 | 1.16978 | 8.632527 | 6.4818 | 4.26E-09 | 3.49E-08 | 10.40199 |
| RDH16 | -0.25588 | 0.837477 | 5.888901 | -6.47653 | 4.37E-09 | 3.57E-08 | 10.37852 |
| KIF21B | -0.23748 | 0.848227 | 5.496044 | -6.47643 | 4.37E-09 | 3.57E-08 | 10.37809 |
| CCBL1 | -0.26891 | 0.829945 | 6.905934 | -6.4759 | 4.38E-09 | 3.57E-08 | 10.37572 |
| PCK1 | -0.82076 | 0.566142 | 11.98978 | -6.47347 | 4.43E-09 | 3.61E-08 | 10.36491 |
| CRNN | -0.21705 | 0.860324 | 4.825495 | -6.47302 | 4.44E-09 | 3.61E-08 | 10.36291 |
| C5orf28 | 0.285271 | 1.218639 | 5.258242 | 6.472435 | 4.45E-09 | 3.62E-08 | 10.3603 |
| WAPAL | 0.279319 | 1.213622 | 7.067473 | 6.471588 | 4.47E-09 | 3.63E-08 | 10.35653 |
| FADS3 | 0.394126 | 1.314146 | 6.230549 | 6.468565 | 4.53E-09 | 3.68E-08 | 10.34308 |
| SLC22A8 | -0.89372 | 0.538225 | 10.11066 | -6.46703 | 4.56E-09 | 3.70E-08 | 10.33625 |
| VRK3 | 0.245556 | 1.185549 | 7.951429 | 6.464299 | 4.62E-09 | 3.74E-08 | 10.3241 |
| CSE1L | 0.257976 | 1.1958 | 7.846484 | 6.463238 | 4.64E-09 | 3.76E-08 | 10.31938 |
| LYN | 0.456638 | 1.37234 | 5.644505 | 6.462315 | 4.66E-09 | 3.77E-08 | 10.31527 |
| IMPDH1 | 0.39401 | 1.31404 | 7.445275 | 6.459083 | 4.73E-09 | 3.82E-08 | 10.3009 |
| DIO3 | -0.21116 | 0.86384 | 5.823187 | -6.45717 | 4.77E-09 | 3.85E-08 | 10.29241 |
| TEAD4 | 0.389696 | 1.310117 | 6.159011 | 6.456992 | 4.78E-09 | 3.85E-08 | 10.2916 |
| MED17 | 0.308386 | 1.238322 | 6.28989 | 6.454904 | 4.82E-09 | 3.88E-08 | 10.28231 |
| NEU3 | -0.18946 | 0.876935 | 5.210659 | -6.45402 | 4.84E-09 | 3.89E-08 | 10.27837 |
| RPA3 | -0.27269 | 0.827774 | 9.174066 | -6.45339 | 4.86E-09 | 3.90E-08 | 10.2756 |
| RORC | -0.53629 | 0.689544 | 7.121978 | -6.45316 | 4.86E-09 | 3.90E-08 | 10.27457 |
| SEC23B | 0.231271 | 1.173868 | 7.991538 | 6.451142 | 4.91E-09 | 3.93E-08 | 10.26559 |
| SMAGP | 0.404391 | 1.32353 | 6.707582 | 6.450088 | 4.93E-09 | 3.95E-08 | 10.2609 |
| EPM2A | -0.25761 | 0.836473 | 6.04022 | -6.44924 | 4.95E-09 | 3.96E-08 | 10.25714 |
| PPP6R2 | -0.21503 | 0.861526 | 6.117143 | -6.44767 | 4.98E-09 | 3.99E-08 | 10.25014 |
| PALM | -0.38109 | 0.767859 | 7.432637 | -6.44609 | 5.02E-09 | 4.01E-08 | 10.24314 |
| IFITM1 | 0.893454 | 1.857618 | 8.807253 | 6.446053 | 5.02E-09 | 4.01E-08 | 10.24297 |
| CFAP45 | -0.24979 | 0.84102 | 6.286044 | -6.44492 | 5.05E-09 | 4.02E-08 | 10.23793 |
| SMCP | -0.22562 | 0.855226 | 5.635385 | -6.44472 | 5.05E-09 | 4.02E-08 | 10.23705 |
| UTP6 | 0.291232 | 1.223685 | 7.599451 | 6.443882 | 5.07E-09 | 4.04E-08 | 10.23332 |
| CASP10 | -0.17963 | 0.882931 | 5.877912 | -6.44324 | 5.09E-09 | 4.04E-08 | 10.23048 |
| YRDC | -0.26237 | 0.833716 | 7.063516 | -6.44214 | 5.11E-09 | 4.06E-08 | 10.2256 |
| CHST8 | -0.24465 | 0.844022 | 5.51989 | -6.43817 | 5.20E-09 | 4.13E-08 | 10.20794 |
| DERL1 | 0.210498 | 1.157087 | 7.206484 | 6.435804 | 5.26E-09 | 4.17E-08 | 10.19744 |
| HGSNAT | 0.215087 | 1.160774 | 7.335275 | 6.43414 | 5.30E-09 | 4.20E-08 | 10.19005 |
| UGT8 | 0.52313 | 1.43707 | 6.29956 | 6.43099 | 5.38E-09 | 4.26E-08 | 10.17606 |
| P4HA1 | -0.61471 | 0.653059 | 8.093846 | -6.4303 | 5.39E-09 | 4.27E-08 | 10.17301 |
| CAPZA2 | 0.265024 | 1.201656 | 9.157143 | 6.428774 | 5.43E-09 | 4.29E-08 | 10.16623 |
| PTPRT | -0.27865 | 0.824364 | 5.383077 | -6.42797 | 5.45E-09 | 4.30E-08 | 10.16266 |
| MPZL1 | 0.25901 | 1.196657 | 6.883516 | 6.421185 | 5.62E-09 | 4.43E-08 | 10.13254 |
| PBK | 0.692932 | 1.616566 | 3.964615 | 6.421069 | 5.63E-09 | 4.43E-08 | 10.13202 |
| ITGAV | 0.474174 | 1.389123 | 9.262308 | 6.420118 | 5.65E-09 | 4.45E-08 | 10.12781 |
| MINA | 0.238019 | 1.179372 | 6.528571 | 6.419403 | 5.67E-09 | 4.46E-08 | 10.12464 |
| CSHL1 | -0.31492 | 0.803894 | 5.412747 | -6.41673 | 5.74E-09 | 4.51E-08 | 10.11276 |
| PRRG1 | 0.463609 | 1.378987 | 6.911648 | 6.415574 | 5.77E-09 | 4.53E-08 | 10.10765 |
| UBA6 | 0.31644 | 1.245254 | 5.506264 | 6.415459 | 5.77E-09 | 4.53E-08 | 10.10714 |
| EPX | -0.24327 | 0.844831 | 5.492747 | -6.41266 | 5.85E-09 | 4.58E-08 | 10.09473 |

| | | | | | | | |
|---|---|---|---|---|---|---|---|
| CYP3A7 | -0.93387 | 0.523452 | 6.964066 | -6.41071 | 5.90E-09 | 4.62E-08 | 10.08609 |
| ST13 | -0.25329 | 0.838978 | 10.61648 | -6.41041 | 5.91E-09 | 4.62E-08 | 10.08476 |
| PSMA3 | 0.261193 | 1.19847 | 9.242857 | 6.409902 | 5.92E-09 | 4.62E-08 | 10.08249 |
| AP1G1 | 0.287232 | 1.220297 | 7.617473 | 6.408385 | 5.96E-09 | 4.65E-08 | 10.07576 |
| CADM3 | -0.15313 | 0.899297 | 5.003407 | -6.40263 | 6.12E-09 | 4.77E-08 | 10.05026 |
| AATK | -0.23787 | 0.847997 | 6.948242 | -6.3991 | 6.22E-09 | 4.84E-08 | 10.03463 |
| MGLL | 0.491725 | 1.406125 | 8.910769 | 6.398498 | 6.23E-09 | 4.85E-08 | 10.03194 |
| RAB8B | 0.302599 | 1.233364 | 6.676593 | 6.395824 | 6.31E-09 | 4.90E-08 | 10.02009 |
| HOXC13 | -0.23049 | 0.852347 | 4.604066 | -6.39578 | 6.31E-09 | 4.90E-08 | 10.01991 |
| PPP1R15A | -0.38157 | 0.767604 | 5.920879 | -6.39418 | 6.36E-09 | 4.94E-08 | 10.0128 |
| PRKCDBP | 0.384715 | 1.305602 | 5.842418 | 6.391962 | 6.42E-09 | 4.98E-08 | 10.00298 |
| LILRB3 | -0.24211 | 0.845507 | 8.915275 | -6.38872 | 6.52E-09 | 5.05E-08 | 9.988623 |
| IFRD1 | -0.40406 | 0.75573 | 5.847582 | -6.38868 | 6.52E-09 | 5.05E-08 | 9.98843 |
| PARP8 | 0.275232 | 1.210189 | 5.971538 | 6.38708 | 6.57E-09 | 5.08E-08 | 9.981354 |
| DMWD | -0.20195 | 0.869374 | 6.552308 | -6.38631 | 6.59E-09 | 5.09E-08 | 9.977937 |
| RIC8A | 0.196444 | 1.145871 | 8.177143 | 6.385092 | 6.63E-09 | 5.12E-08 | 9.972553 |
| HSPA5 | 0.463106 | 1.378507 | 9.768791 | 6.382119 | 6.72E-09 | 5.18E-08 | 9.959389 |
| MC1R | -0.28397 | 0.82133 | 5.581099 | -6.38186 | 6.72E-09 | 5.18E-08 | 9.958225 |
| PHTF1 | 0.228512 | 1.171626 | 4.966044 | 6.380007 | 6.78E-09 | 5.22E-08 | 9.950039 |
| KIR3DX1 | -0.35109 | 0.783991 | 5.587253 | -6.37958 | 6.79E-09 | 5.23E-08 | 9.948144 |
| TEF | -0.25899 | 0.835675 | 5.327582 | -6.37793 | 6.85E-09 | 5.26E-08 | 9.940853 |
| TMEM43 | 0.246111 | 1.186006 | 8.236703 | 6.377865 | 6.85E-09 | 5.26E-08 | 9.940558 |
| TLR2 | 0.461473 | 1.376947 | 5.694505 | 6.377234 | 6.87E-09 | 5.27E-08 | 9.937767 |
| PLGRKT | 0.260942 | 1.198261 | 8.651429 | 6.374423 | 6.95E-09 | 5.33E-08 | 9.925324 |
| FHL2 | 0.669874 | 1.590934 | 7.295604 | 6.373492 | 6.98E-09 | 5.35E-08 | 9.921207 |
| PCK2 | -0.65886 | 0.633379 | 10.23549 | -6.37252 | 7.02E-09 | 5.37E-08 | 9.916912 |
| ELAVL3 | -0.22142 | 0.857721 | 5.286593 | -6.37229 | 7.02E-09 | 5.37E-08 | 9.915883 |
| VPS13C | 0.306256 | 1.236495 | 7.592967 | 6.370281 | 7.09E-09 | 5.41E-08 | 9.907001 |
| NPPA | -0.28297 | 0.821897 | 6.076374 | -6.37009 | 7.09E-09 | 5.41E-08 | 9.906156 |
| POLR2E | 0.252227 | 1.191044 | 8.934945 | 6.369283 | 7.12E-09 | 5.43E-08 | 9.902586 |
| EXOSC10 | 0.326314 | 1.253806 | 5.675495 | 6.368285 | 7.15E-09 | 5.45E-08 | 9.89817 |
| MARCKS | 0.442609 | 1.35906 | 5.336264 | 6.367752 | 7.17E-09 | 5.46E-08 | 9.895814 |
| SPDEF | -0.24261 | 0.845216 | 6.922637 | -6.36617 | 7.22E-09 | 5.49E-08 | 9.888824 |
| IFITM2 | 0.70329 | 1.628213 | 9.135824 | 6.363729 | 7.30E-09 | 5.55E-08 | 9.878021 |
| GPX5 | -0.17553 | 0.885444 | 4.214505 | -6.36078 | 7.40E-09 | 5.62E-08 | 9.864966 |
| ALDH1L1 | -0.47561 | 0.719161 | 9.842198 | -6.35978 | 7.43E-09 | 5.64E-08 | 9.860572 |
| PARP1 | 0.223319 | 1.167416 | 7.84978 | 6.359262 | 7.45E-09 | 5.64E-08 | 9.858269 |
| ALLC | -0.31913 | 0.801556 | 4.861538 | -6.35675 | 7.54E-09 | 5.70E-08 | 9.847154 |
| NUP93 | 0.242734 | 1.183233 | 7.253077 | 6.356281 | 7.55E-09 | 5.71E-08 | 9.845089 |
| FUBP1 | 0.23842 | 1.1797 | 7.596813 | 6.355494 | 7.58E-09 | 5.72E-08 | 9.841613 |
| SSTR3 | -0.24366 | 0.844602 | 5.994945 | -6.35548 | 7.58E-09 | 5.72E-08 | 9.841548 |
| CRABP1 | -0.37879 | 0.769081 | 5.417033 | -6.35532 | 7.58E-09 | 5.72E-08 | 9.84086 |
| UBE2L6 | 0.525459 | 1.439391 | 7.855495 | 6.355173 | 7.59E-09 | 5.72E-08 | 9.840193 |
| MRPS17 | 0.20872 | 1.155662 | 7.965604 | 6.354711 | 7.61E-09 | 5.73E-08 | 9.838151 |
| IFI16 | 0.666531 | 1.587252 | 6.113516 | 6.352295 | 7.69E-09 | 5.78E-08 | 9.827474 |
| TNNI1 | -0.29542 | 0.814835 | 4.5 | -6.35171 | 7.71E-09 | 5.79E-08 | 9.82487 |
| SULT2B1 | -0.27107 | 0.828706 | 5.997912 | -6.3512 | 7.73E-09 | 5.80E-08 | 9.822627 |

| | | | | | | | |
|---|---|---|---|---|---|---|---|
| KIN | 0.251193 | 1.190191 | 4.747912 | 6.350761 | 7.74E-09 | 5.80E-08 | 9.820693 |
| TMEM59L | -0.28906 | 0.818436 | 5.989451 | -6.35054 | 7.75E-09 | 5.80E-08 | 9.819728 |
| KYNU | -0.27865 | 0.824364 | 7.153077 | -6.35047 | 7.75E-09 | 5.80E-08 | 9.819409 |
| BHLHE40 | -0.56034 | 0.678141 | 7.697473 | -6.35034 | 7.76E-09 | 5.80E-08 | 9.818848 |
| KRT10 | 0.249957 | 1.189171 | 8.861648 | 6.348619 | 7.82E-09 | 5.84E-08 | 9.811229 |
| TCEB3-AS1 | -0.27639 | 0.825656 | 6.485934 | -6.34846 | 7.82E-09 | 5.84E-08 | 9.81051 |
| TESC | 0.594947 | 1.510417 | 6.106813 | 6.343445 | 8.00E-09 | 5.97E-08 | 9.788374 |
| DDX41 | 0.220493 | 1.165131 | 7.941209 | 6.341821 | 8.06E-09 | 6.01E-08 | 9.7812 |
| MAPK8 | -0.22073 | 0.858131 | 6.158022 | -6.33812 | 8.20E-09 | 6.11E-08 | 9.764867 |
| PTRH2 | 0.245295 | 1.185335 | 7.63956 | 6.337361 | 8.23E-09 | 6.12E-08 | 9.761507 |
| SEC11A | 0.203961 | 1.151857 | 10.71912 | 6.336427 | 8.26E-09 | 6.14E-08 | 9.757384 |
| KIAA1551 | 0.352551 | 1.276816 | 6.079121 | 6.336305 | 8.27E-09 | 6.14E-08 | 9.756847 |
| DSTN | 0.186019 | 1.13762 | 11.54286 | 6.334634 | 8.33E-09 | 6.18E-08 | 9.749467 |
| IVL | -0.24281 | 0.845097 | 5.743407 | -6.33452 | 8.33E-09 | 6.18E-08 | 9.748973 |
| IGFALS | -0.24357 | 0.844653 | 5.628901 | -6.33354 | 8.37E-09 | 6.20E-08 | 9.744644 |
| EXT1 | 0.284396 | 1.2179 | 7.168462 | 6.332088 | 8.43E-09 | 6.24E-08 | 9.738229 |
| TP53I3 | 0.336198 | 1.262425 | 8.385165 | 6.330852 | 8.47E-09 | 6.27E-08 | 9.732777 |
| DNAJC13 | 0.222961 | 1.167127 | 6.033516 | 6.328404 | 8.57E-09 | 6.33E-08 | 9.721972 |
| FBXL4 | 0.252169 | 1.190996 | 5.782308 | 6.327768 | 8.59E-09 | 6.35E-08 | 9.719167 |
| HPX | -0.22003 | 0.858548 | 6.45989 | -6.32629 | 8.65E-09 | 6.38E-08 | 9.712624 |
| SOX10 | -0.20219 | 0.869231 | 5.411538 | -6.32582 | 8.67E-09 | 6.39E-08 | 9.710557 |
| SNX5 | -0.23525 | 0.849537 | 9.927363 | -6.32561 | 8.68E-09 | 6.39E-08 | 9.709658 |
| HMX1 | -0.27398 | 0.827037 | 6.435604 | -6.32257 | 8.80E-09 | 6.47E-08 | 9.696219 |
| MMP25 | -0.2299 | 0.852695 | 6.000879 | -6.32192 | 8.82E-09 | 6.49E-08 | 9.693359 |
| NSMAF | 0.286536 | 1.219708 | 5.895824 | 6.319395 | 8.93E-09 | 6.56E-08 | 9.682231 |
| GNB1 | 0.195 | 1.144724 | 9.181429 | 6.319085 | 8.94E-09 | 6.56E-08 | 9.680861 |
| SEMA4A | -0.23594 | 0.84913 | 6.775934 | -6.31835 | 8.97E-09 | 6.58E-08 | 9.67762 |
| MSRA | -0.35752 | 0.780504 | 10.90473 | -6.31504 | 9.10E-09 | 6.67E-08 | 9.66303 |
| GPR31 | -0.23221 | 0.851328 | 4.875604 | -6.31498 | 9.11E-09 | 6.67E-08 | 9.662779 |
| IL17RB | 0.527406 | 1.441335 | 10.54407 | 6.306845 | 9.45E-09 | 6.91E-08 | 9.6269 |
| CSF3 | -0.2095 | 0.864835 | 6.038791 | -6.30441 | 9.55E-09 | 6.98E-08 | 9.616187 |
| CRIP2 | 0.510188 | 1.424236 | 6.980769 | 6.299561 | 9.76E-09 | 7.13E-08 | 9.594812 |
| KRT76 | -0.22777 | 0.853955 | 4.997802 | -6.29916 | 9.78E-09 | 7.13E-08 | 9.593046 |
| RRM1 | 0.279092 | 1.213431 | 6.567143 | 6.298937 | 9.79E-09 | 7.13E-08 | 9.592061 |
| CDK2AP2 | 0.548488 | 1.462552 | 7.203187 | 6.298936 | 9.79E-09 | 7.13E-08 | 9.592058 |
| INTS8 | 0.287879 | 1.220844 | 7.506923 | 6.296931 | 9.88E-09 | 7.19E-08 | 9.583229 |
| UPF3B | 0.275807 | 1.210671 | 5.587692 | 6.295535 | 9.94E-09 | 7.23E-08 | 9.577078 |
| GPC3 | -0.85209 | 0.553981 | 7.554505 | -6.29479 | 9.98E-09 | 7.25E-08 | 9.573802 |
| NNAT | -0.21971 | 0.858738 | 5.204396 | -6.29441 | 9.99E-09 | 7.25E-08 | 9.572121 |
| CRISP2 | -0.32857 | 0.796325 | 4.171868 | -6.29435 | 1.00E-08 | 7.25E-08 | 9.571874 |
| NBEA | 0.373913 | 1.295863 | 5.530989 | 6.293617 | 1.00E-08 | 7.27E-08 | 9.568631 |
| YTHDC1 | 0.240415 | 1.181333 | 7.757582 | 6.291475 | 1.01E-08 | 7.33E-08 | 9.559201 |
| BTN3A2 | 0.505957 | 1.420065 | 5.658242 | 6.290149 | 1.02E-08 | 7.37E-08 | 9.553366 |
| SNTA1 | -0.32685 | 0.797275 | 7.16011 | -6.2874 | 1.03E-08 | 7.45E-08 | 9.541244 |
| SNRPB | 0.3303 | 1.257274 | 8.716813 | 6.284606 | 1.04E-08 | 7.54E-08 | 9.528966 |
| BTNL8 | -0.25162 | 0.839954 | 4.902747 | -6.2846 | 1.04E-08 | 7.54E-08 | 9.528944 |
| DKKL1 | -0.22788 | 0.853889 | 5.652747 | -6.2822 | 1.06E-08 | 7.61E-08 | 9.518378 |

| | | | | | | | |
|---|---|---|---|---|---|---|---|
| SEMA4D | 0.331807 | 1.258589 | 6.985385 | 6.279487 | 1.07E-08 | 7.70E-08 | 9.506445 |
| ALDH3A2 | -0.54545 | 0.685178 | 9.925055 | -6.27931 | 1.07E-08 | 7.70E-08 | 9.505687 |
| ASCL3 | -0.2675 | 0.830757 | 5.03011 | -6.27835 | 1.07E-08 | 7.73E-08 | 9.501432 |
| VPS45 | 0.253947 | 1.192465 | 7.403187 | 6.273899 | 1.10E-08 | 7.88E-08 | 9.481866 |
| NRAS | 0.369773 | 1.292149 | 7.032637 | 6.267994 | 1.13E-08 | 8.08E-08 | 9.455903 |
| ACOT13 | -0.42841 | 0.743082 | 9.94989 | -6.26629 | 1.13E-08 | 8.14E-08 | 9.448399 |
| PAXIP1 | 0.23513 | 1.177013 | 6.207143 | 6.262216 | 1.16E-08 | 8.28E-08 | 9.430506 |
| PSMD7 | 0.221459 | 1.165912 | 8.655165 | 6.259657 | 1.17E-08 | 8.37E-08 | 9.419264 |
| PNLIPRP1 | -0.26696 | 0.831068 | 5.344725 | -6.25497 | 1.19E-08 | 8.54E-08 | 9.398676 |
| TMEM127 | 0.18215 | 1.134573 | 7.806813 | 6.252102 | 1.21E-08 | 8.65E-08 | 9.386076 |
| CTH | -0.42515 | 0.744761 | 6.968022 | -6.25164 | 1.21E-08 | 8.66E-08 | 9.384046 |
| ARC | -0.2916 | 0.816993 | 4.483626 | -6.25001 | 1.22E-08 | 8.71E-08 | 9.376875 |
| HTR1B | -0.20632 | 0.866743 | 4.903407 | -6.24928 | 1.23E-08 | 8.72E-08 | 9.373697 |
| IFNGR1 | 0.350787 | 1.275256 | 7.938901 | 6.249249 | 1.23E-08 | 8.72E-08 | 9.373548 |
| SPATA5L1 | 0.248749 | 1.188176 | 6.096703 | 6.249217 | 1.23E-08 | 8.72E-08 | 9.373408 |
| CDSN | -0.21575 | 0.861096 | 5.334396 | -6.24846 | 1.23E-08 | 8.74E-08 | 9.370092 |
| AZGP1 | -0.59298 | 0.662972 | 10.16264 | -6.24695 | 1.24E-08 | 8.80E-08 | 9.363469 |
| HIP1R | -0.25671 | 0.836994 | 6.131099 | -6.24507 | 1.25E-08 | 8.86E-08 | 9.35519 |
| PERP | 0.489739 | 1.404191 | 6.38044 | 6.244547 | 1.25E-08 | 8.88E-08 | 9.352908 |
| SLC2A4 | -0.18569 | 0.879231 | 4.502308 | -6.24188 | 1.27E-08 | 8.98E-08 | 9.341219 |
| PEPD | -0.34755 | 0.785917 | 11.54835 | -6.23896 | 1.28E-08 | 9.08E-08 | 9.328378 |
| MBD4 | 0.292512 | 1.224771 | 7.247692 | 6.238954 | 1.28E-08 | 9.08E-08 | 9.328362 |
| METTL5 | 0.271729 | 1.207254 | 8.852198 | 6.238825 | 1.28E-08 | 9.08E-08 | 9.327794 |
| ERCC2 | -0.2568 | 0.836941 | 6.296923 | -6.23775 | 1.29E-08 | 9.12E-08 | 9.323069 |
| RYK | 0.198884 | 1.14781 | 6.708132 | 6.237514 | 1.29E-08 | 9.12E-08 | 9.322042 |
| UPF2 | 0.265034 | 1.201664 | 6.447582 | 6.236417 | 1.30E-08 | 9.16E-08 | 9.317232 |
| DPP4 | -0.57857 | 0.669627 | 9.228242 | -6.23584 | 1.30E-08 | 9.17E-08 | 9.314689 |
| CD1A | -0.24131 | 0.845974 | 4.951538 | -6.23563 | 1.30E-08 | 9.17E-08 | 9.31379 |
| IL13 | -0.2606 | 0.834741 | 5.614176 | -6.23423 | 1.31E-08 | 9.22E-08 | 9.307643 |
| CMPK1 | 0.244266 | 1.18449 | 8.472747 | 6.23266 | 1.32E-08 | 9.28E-08 | 9.300749 |
| ALDH1A1 | 0.365783 | 1.28858 | 10.6411 | 6.23012 | 1.34E-08 | 9.38E-08 | 9.289608 |
| PLAGL2 | -0.21796 | 0.859782 | 5.614176 | -6.2292 | 1.34E-08 | 9.41E-08 | 9.285555 |
| BCAP31 | 0.225333 | 1.169047 | 10.79143 | 6.228399 | 1.35E-08 | 9.44E-08 | 9.282061 |
| SSUH2 | -0.28736 | 0.819402 | 5.386813 | -6.22768 | 1.35E-08 | 9.46E-08 | 9.278903 |
| C21orf2 | -0.23363 | 0.850491 | 6.300989 | -6.227 | 1.35E-08 | 9.48E-08 | 9.27592 |
| MUC1 | 0.510749 | 1.42479 | 8.056264 | 6.225964 | 1.36E-08 | 9.52E-08 | 9.271389 |
| GNL2 | 0.275976 | 1.210813 | 5.615385 | 6.224613 | 1.37E-08 | 9.57E-08 | 9.265465 |
| VPS26A | 0.144536 | 1.105375 | 10.2156 | 6.224101 | 1.37E-08 | 9.58E-08 | 9.26322 |
| EGF | -1.0048 | 0.49834 | 8.157253 | -6.22154 | 1.39E-08 | 9.68E-08 | 9.252001 |
| ST5 | 0.336986 | 1.263115 | 6.010989 | 6.219847 | 1.40E-08 | 9.75E-08 | 9.244574 |
| IFIH1 | 0.577116 | 1.491864 | 5.835604 | 6.216972 | 1.42E-08 | 9.86E-08 | 9.231974 |
| MCAM | 0.319019 | 1.247482 | 6.053626 | 6.216917 | 1.42E-08 | 9.86E-08 | 9.231738 |
| CA8 | -0.38623 | 0.765125 | 5.328571 | -6.21642 | 1.42E-08 | 9.88E-08 | 9.229576 |
| ABLIM1 | 0.229275 | 1.172246 | 8.890769 | 6.215576 | 1.43E-08 | 9.90E-08 | 9.225862 |
| ULK4 | -0.29156 | 0.817018 | 5.235604 | -6.21554 | 1.43E-08 | 9.90E-08 | 9.225706 |
| PGLYRP4 | -0.25643 | 0.837154 | 5.763626 | -6.21186 | 1.45E-08 | 1.01E-07 | 9.209587 |
| SH3GLB1 | 0.1983 | 1.147345 | 8.691538 | 6.21042 | 1.46E-08 | 1.01E-07 | 9.203275 |

| | | | | | | | |
|---|---|---|---|---|---|---|---|
| ZSWIM8-A | -0.24358 | 0.844644 | 6.298242 | -6.20855 | 1.47E-08 | 1.02E-07 | 9.195069 |
| SLC26A2 | 0.50214 | 1.416313 | 5.364615 | 6.206181 | 1.49E-08 | 1.03E-07 | 9.184713 |
| PSMA1 | 0.161319 | 1.118309 | 10.26912 | 6.20559 | 1.49E-08 | 1.03E-07 | 9.182125 |
| TREH | -1.00091 | 0.499685 | 6.974176 | -6.2027 | 1.51E-08 | 1.04E-07 | 9.169483 |
| CTNNBIP1 | -0.25993 | 0.83513 | 7.784725 | -6.20144 | 1.52E-08 | 1.05E-07 | 9.163953 |
| TRIM62 | -0.19153 | 0.875676 | 6.006374 | -6.20041 | 1.53E-08 | 1.05E-07 | 9.15944 |
| SULT1B1 | -0.25314 | 0.839065 | 5.413297 | -6.20032 | 1.53E-08 | 1.05E-07 | 9.15904 |
| NT5DC2 | 0.38014 | 1.301468 | 5.164286 | 6.199958 | 1.53E-08 | 1.05E-07 | 9.157471 |
| EXOC2 | 0.254063 | 1.192561 | 5.598462 | 6.199691 | 1.53E-08 | 1.05E-07 | 9.156301 |
| DNAI1 | -0.26462 | 0.832416 | 4.971099 | -6.19915 | 1.54E-08 | 1.06E-07 | 9.15392 |
| LAMB1 | 0.297362 | 1.228896 | 7.838352 | 6.198925 | 1.54E-08 | 1.06E-07 | 9.152949 |
| ARHGDIB | 0.625734 | 1.542996 | 7.577473 | 6.197331 | 1.55E-08 | 1.06E-07 | 9.145971 |
| TLR1 | 0.456314 | 1.372032 | 4.42978 | 6.19445 | 1.57E-08 | 1.08E-07 | 9.133368 |
| DAAM2 | -0.23735 | 0.848303 | 5.661978 | -6.19429 | 1.57E-08 | 1.08E-07 | 9.13268 |
| ICE2 | 0.196227 | 1.145698 | 5.717253 | 6.19382 | 1.57E-08 | 1.08E-07 | 9.13061 |
| HSD17B8 | -0.27872 | 0.824322 | 9.04978 | -6.1936 | 1.57E-08 | 1.08E-07 | 9.129666 |
| PLOD1 | -0.3218 | 0.800073 | 7.85 | -6.1934 | 1.58E-08 | 1.08E-07 | 9.128791 |
| TBC1D29 | -0.2078 | 0.865855 | 4.676154 | -6.19329 | 1.58E-08 | 1.08E-07 | 9.128302 |
| GDF2 | -0.22365 | 0.856395 | 5.855055 | -6.19273 | 1.58E-08 | 1.08E-07 | 9.125854 |
| BCLAF1 | 0.223768 | 1.16778 | 7.19022 | 6.19019 | 1.60E-08 | 1.09E-07 | 9.11473 |
| DSCAM | -0.19548 | 0.873283 | 5.126813 | -6.18937 | 1.60E-08 | 1.09E-07 | 9.111135 |
| TPSD1 | -0.18454 | 0.879929 | 5.624396 | -6.18911 | 1.61E-08 | 1.09E-07 | 9.110013 |
| LUC7L3 | 0.290213 | 1.22282 | 7.783077 | 6.188312 | 1.61E-08 | 1.10E-07 | 9.106517 |
| ZNF638 | 0.258638 | 1.196348 | 8.036593 | 6.186478 | 1.63E-08 | 1.10E-07 | 9.098499 |
| PNMAL1 | 0.576362 | 1.491085 | 6.391319 | 6.181905 | 1.66E-08 | 1.13E-07 | 9.078502 |
| TMED3 | 0.335106 | 1.26147 | 7.805495 | 6.179119 | 1.68E-08 | 1.14E-07 | 9.066328 |
| ULK1 | -0.27724 | 0.82517 | 6.237253 | -6.17753 | 1.69E-08 | 1.15E-07 | 9.05938 |
| SLC25A24 | 0.294285 | 1.226277 | 7.918352 | 6.177345 | 1.69E-08 | 1.15E-07 | 9.058572 |
| GFRA4 | -0.19477 | 0.87371 | 5.508901 | -6.17701 | 1.70E-08 | 1.15E-07 | 9.057127 |
| CACNA1H | -0.23795 | 0.847948 | 5.974505 | -6.17259 | 1.73E-08 | 1.17E-07 | 9.037818 |
| PAPSS1 | 0.303353 | 1.234009 | 8.830879 | 6.171775 | 1.74E-08 | 1.17E-07 | 9.034234 |
| CDA | -0.40911 | 0.753087 | 5.677692 | -6.17167 | 1.74E-08 | 1.17E-07 | 9.033792 |
| GPR21 | -0.20931 | 0.864948 | 6.117363 | -6.17088 | 1.74E-08 | 1.18E-07 | 9.030322 |
| SCML1 | -0.29074 | 0.817483 | 5.302967 | -6.16904 | 1.76E-08 | 1.18E-07 | 9.022294 |
| PTPRF | 0.262034 | 1.199168 | 7.091099 | 6.168896 | 1.76E-08 | 1.18E-07 | 9.021662 |
| SLC27A3 | 0.403758 | 1.32295 | 5.798791 | 6.168335 | 1.76E-08 | 1.19E-07 | 9.019209 |
| SLC13A3 | -0.49095 | 0.711558 | 10.33462 | -6.16768 | 1.77E-08 | 1.19E-07 | 9.016348 |
| CDK1 | 0.484 | 1.398616 | 4.729341 | 6.166617 | 1.78E-08 | 1.19E-07 | 9.011706 |
| CETN3 | 0.359957 | 1.283387 | 7.426044 | 6.166584 | 1.78E-08 | 1.19E-07 | 9.011562 |
| FBN1 | 0.263575 | 1.20045 | 4.921209 | 6.166552 | 1.78E-08 | 1.19E-07 | 9.011425 |
| ZFAND5 | -0.49457 | 0.709771 | 9.80956 | -6.16619 | 1.78E-08 | 1.19E-07 | 9.009843 |
| TMEM106 | 0.224551 | 1.168413 | 9.455824 | 6.16391 | 1.80E-08 | 1.21E-07 | 8.999887 |
| DNAAF5 | 0.221324 | 1.165803 | 7.419011 | 6.163696 | 1.80E-08 | 1.21E-07 | 8.998951 |
| C11orf71 | -0.39418 | 0.76092 | 7.406813 | -6.16308 | 1.81E-08 | 1.21E-07 | 8.996248 |
| FAS | 0.557174 | 1.471384 | 6.348352 | 6.161641 | 1.82E-08 | 1.21E-07 | 8.989982 |
| LEFTY2 | -0.19622 | 0.872836 | 4.963187 | -6.16138 | 1.82E-08 | 1.21E-07 | 8.988827 |
| RNF44 | 0.235333 | 1.177179 | 7.606374 | 6.161234 | 1.82E-08 | 1.21E-07 | 8.988202 |

| | | | | | | | |
|---|---|---|---|---|---|---|---|
| CRYBB3 | -0.34124 | 0.789362 | 5.335385 | -6.16014 | 1.83E-08 | 1.22E-07 | 8.983438 |
| PPP1R8 | 0.181072 | 1.133726 | 8.437802 | 6.158411 | 1.84E-08 | 1.23E-07 | 8.97588 |
| NSUN5 | 0.296676 | 1.228311 | 5.717143 | 6.156616 | 1.86E-08 | 1.24E-07 | 8.968049 |
| SPAM1 | -0.19333 | 0.874583 | 4.484396 | -6.15586 | 1.86E-08 | 1.24E-07 | 8.964735 |
| CDK7 | 0.364213 | 1.287179 | 7.61967 | 6.153766 | 1.88E-08 | 1.25E-07 | 8.955611 |
| PCDHB1 | -0.19215 | 0.8753 | 4.728242 | -6.15262 | 1.89E-08 | 1.26E-07 | 8.950618 |
| PCDHA9 | -0.21029 | 0.864366 | 4.203187 | -6.15183 | 1.90E-08 | 1.26E-07 | 8.947166 |
| TYROBP | 1.000512 | 2.00071 | 6.857802 | 6.149544 | 1.92E-08 | 1.27E-07 | 8.937193 |
| IL15 | 0.2097 | 1.156448 | 4.34022 | 6.148894 | 1.92E-08 | 1.27E-07 | 8.934358 |
| RUSC2 | -0.19636 | 0.872748 | 6.886593 | -6.14854 | 1.93E-08 | 1.28E-07 | 8.932813 |
| SETX | 0.206671 | 1.154023 | 6.602418 | 6.146769 | 1.94E-08 | 1.28E-07 | 8.925088 |
| CCDC109B | 0.507068 | 1.421159 | 5.468791 | 6.146654 | 1.94E-08 | 1.28E-07 | 8.924588 |
| TBKBP1 | -0.19775 | 0.87191 | 5.693516 | -6.14522 | 1.96E-08 | 1.29E-07 | 8.918342 |
| VGLL4 | 0.220638 | 1.165249 | 7.687802 | 6.14409 | 1.97E-08 | 1.30E-07 | 8.913406 |
| TSPAN6 | 0.311372 | 1.240887 | 9.205714 | 6.141747 | 1.99E-08 | 1.31E-07 | 8.903189 |
| APOBEC3G | 0.468174 | 1.383357 | 5.649341 | 6.140975 | 1.99E-08 | 1.31E-07 | 8.899822 |
| CRYL1 | -0.31428 | 0.804255 | 11.1722 | -6.14034 | 2.00E-08 | 1.32E-07 | 8.897035 |
| MAEA | 0.233024 | 1.175296 | 8.741319 | 6.13948 | 2.01E-08 | 1.32E-07 | 8.893306 |
| SOX13 | -0.22654 | 0.854684 | 6.383846 | -6.13886 | 2.01E-08 | 1.32E-07 | 8.890618 |
| ENTPD4 | 0.188324 | 1.139439 | 5.947692 | 6.138554 | 2.02E-08 | 1.32E-07 | 8.88927 |
| C1orf21 | -0.26749 | 0.830765 | 5.360769 | -6.13693 | 2.03E-08 | 1.33E-07 | 8.882193 |
| ANKMY2 | 0.310836 | 1.240426 | 8.271319 | 6.136578 | 2.03E-08 | 1.33E-07 | 8.880658 |
| CACNG4 | -0.176 | 0.885154 | 6.662967 | -6.13585 | 2.04E-08 | 1.34E-07 | 8.877478 |
| INTS9 | 0.216889 | 1.162225 | 6.757253 | 6.135214 | 2.05E-08 | 1.34E-07 | 8.874715 |
| APBB3 | -0.25801 | 0.836241 | 6.541978 | -6.13351 | 2.06E-08 | 1.35E-07 | 8.867273 |
| TOM1L2 | -0.19339 | 0.874548 | 6.581758 | -6.13257 | 2.07E-08 | 1.35E-07 | 8.86321 |
| POLR3C | 0.196821 | 1.14617 | 6.554286 | 6.132516 | 2.07E-08 | 1.35E-07 | 8.862959 |
| GCH1 | -0.51397 | 0.700295 | 9.187363 | -6.13213 | 2.07E-08 | 1.35E-07 | 8.861293 |
| ARAP2 | 0.268039 | 1.20417 | 5.264286 | 6.131566 | 2.08E-08 | 1.36E-07 | 8.858819 |
| TMPRSS6 | -0.19809 | 0.871703 | 6.627912 | -6.12893 | 2.10E-08 | 1.37E-07 | 8.847355 |
| FAM214B | -0.21936 | 0.858948 | 6.72044 | -6.12771 | 2.12E-08 | 1.38E-07 | 8.842037 |
| KIAA1033 | 0.256295 | 1.194407 | 7.89 | 6.126962 | 2.12E-08 | 1.38E-07 | 8.838765 |
| CA11 | -0.24094 | 0.846195 | 5.428681 | -6.12635 | 2.13E-08 | 1.38E-07 | 8.836078 |
| SLC6A13 | -0.47057 | 0.721679 | 8.931648 | -6.12551 | 2.14E-08 | 1.39E-07 | 8.832441 |
| CDX2 | -0.25446 | 0.838301 | 5.313516 | -6.12534 | 2.14E-08 | 1.39E-07 | 8.831697 |
| CYP4A11 | -0.63018 | 0.646096 | 8.44033 | -6.1235 | 2.16E-08 | 1.40E-07 | 8.823707 |
| MPP3 | -0.26961 | 0.829542 | 5.624066 | -6.12278 | 2.16E-08 | 1.40E-07 | 8.820535 |
| WEE1 | -0.28304 | 0.821855 | 6.733077 | -6.1227 | 2.16E-08 | 1.40E-07 | 8.820221 |
| LYPD3 | -0.28403 | 0.821291 | 6.228022 | -6.12218 | 2.17E-08 | 1.40E-07 | 8.817939 |
| TAF9 | 0.27871 | 1.21311 | 8.54978 | 6.121692 | 2.17E-08 | 1.41E-07 | 8.815816 |
| C1QA | 0.894628 | 1.85913 | 6.380659 | 6.120347 | 2.19E-08 | 1.41E-07 | 8.809957 |
| FOXC2 | -0.20827 | 0.865577 | 4.295055 | -6.1186 | 2.20E-08 | 1.42E-07 | 8.802354 |
| ABCC1 | 0.22342 | 1.167498 | 5.784396 | 6.118379 | 2.21E-08 | 1.42E-07 | 8.801394 |
| PPIF | 0.333565 | 1.260124 | 8.095385 | 6.11779 | 2.21E-08 | 1.43E-07 | 8.79883 |
| GATA6 | -0.37856 | 0.769205 | 4.137582 | -6.11541 | 2.24E-08 | 1.44E-07 | 8.788477 |
| STAP2 | 0.373082 | 1.295117 | 7.563187 | 6.114511 | 2.24E-08 | 1.45E-07 | 8.784558 |
| LRRC1 | 0.38314 | 1.304177 | 7.550769 | 6.113302 | 2.26E-08 | 1.45E-07 | 8.779299 |

| | | | | | | | |
|---|---|---|---|---|---|---|---|
| PPIH | 0.314865 | 1.243895 | 7.034615 | 6.11296 | 2.26E-08 | 1.45E-07 | 8.777809 |
| ASUN | 0.298338 | 1.229727 | 6.952747 | 6.112211 | 2.27E-08 | 1.46E-07 | 8.774548 |
| NEUROD6 | -0.25619 | 0.837297 | 5.804835 | -6.11018 | 2.29E-08 | 1.47E-07 | 8.7657 |
| SH3D21 | -0.23452 | 0.84997 | 5.470769 | -6.10748 | 2.32E-08 | 1.49E-07 | 8.753981 |
| SLC37A4 | -0.35888 | 0.77977 | 9.842967 | -6.10719 | 2.32E-08 | 1.49E-07 | 8.752705 |
| ARHGEF18 | 0.216097 | 1.161587 | 8.141209 | 6.107091 | 2.32E-08 | 1.49E-07 | 8.752277 |
| CAST | 0.218242 | 1.163315 | 8.698791 | 6.104758 | 2.34E-08 | 1.50E-07 | 8.74213 |
| MCM5 | 0.311527 | 1.24102 | 5.242747 | 6.104588 | 2.35E-08 | 1.50E-07 | 8.74139 |
| TTPA | -0.26334 | 0.833155 | 4.239341 | -6.09862 | 2.41E-08 | 1.54E-07 | 8.715458 |
| ENAH | 0.292507 | 1.224767 | 6.157473 | 6.098208 | 2.41E-08 | 1.54E-07 | 8.713653 |
| NRCAM | 0.430155 | 1.347378 | 5.84967 | 6.097661 | 2.42E-08 | 1.54E-07 | 8.711277 |
| SLC25A10 | -0.37423 | 0.771519 | 7.104725 | -6.09759 | 2.42E-08 | 1.54E-07 | 8.710958 |
| QARS | 0.263797 | 1.200635 | 9.401319 | 6.09755 | 2.42E-08 | 1.54E-07 | 8.710793 |
| FKBP6 | -0.20185 | 0.869435 | 5.266923 | -6.09741 | 2.42E-08 | 1.54E-07 | 8.710171 |
| GNRH2 | -0.23 | 0.852635 | 5.086264 | -6.09611 | 2.44E-08 | 1.55E-07 | 8.704537 |
| PIP5K1B | -0.3786 | 0.769182 | 5.765604 | -6.09453 | 2.45E-08 | 1.56E-07 | 8.697669 |
| SEC61A1 | 0.237691 | 1.179104 | 8.513626 | 6.094387 | 2.46E-08 | 1.56E-07 | 8.697046 |
| ABCC6 | -0.44239 | 0.735916 | 7.073846 | -6.09397 | 2.46E-08 | 1.56E-07 | 8.695219 |
| NOTCH4 | -0.18604 | 0.879013 | 5.076044 | -6.09304 | 2.47E-08 | 1.57E-07 | 8.691209 |
| TIPARP | -0.80689 | 0.571611 | 8.600769 | -6.09257 | 2.48E-08 | 1.57E-07 | 8.68917 |
| ATF3 | -1.3342 | 0.396611 | 6.241099 | -6.092 | 2.48E-08 | 1.57E-07 | 8.686664 |
| YTHDF1 | 0.178077 | 1.131375 | 8.801538 | 6.091059 | 2.49E-08 | 1.58E-07 | 8.682587 |
| LOC44160 | -0.24553 | 0.843508 | 6.11989 | -6.09084 | 2.49E-08 | 1.58E-07 | 8.681627 |
| ALPL | -0.47503 | 0.71945 | 6.008571 | -6.08826 | 2.52E-08 | 1.59E-07 | 8.670439 |
| TBX4 | -0.19582 | 0.873079 | 4.341429 | -6.08526 | 2.56E-08 | 1.61E-07 | 8.657381 |
| BEST2 | -0.24218 | 0.845465 | 5.011978 | -6.08433 | 2.57E-08 | 1.62E-07 | 8.653347 |
| RGS10 | 0.564246 | 1.478615 | 6.14011 | 6.083902 | 2.57E-08 | 1.62E-07 | 8.6515 |
| GLI3 | -0.31434 | 0.80422 | 7.119341 | -6.0836 | 2.58E-08 | 1.62E-07 | 8.650175 |
| FOS | -1.52414 | 0.347688 | 6.82022 | -6.08139 | 2.60E-08 | 1.64E-07 | 8.640585 |
| KLK11 | -0.22285 | 0.856874 | 5.341758 | -6.07464 | 2.68E-08 | 1.69E-07 | 8.611274 |
| APOLD1 | -0.67522 | 0.626236 | 5.721099 | -6.07359 | 2.69E-08 | 1.69E-07 | 8.606742 |
| SFXN1 | -0.36832 | 0.774685 | 7.983516 | -6.07108 | 2.72E-08 | 1.71E-07 | 8.595844 |
| USP2 | -0.70921 | 0.611656 | 7.044945 | -6.07085 | 2.73E-08 | 1.71E-07 | 8.594843 |
| FBXO9 | -0.17622 | 0.88502 | 8.143077 | -6.07025 | 2.73E-08 | 1.72E-07 | 8.592225 |
| PP13 | -0.22185 | 0.857465 | 6.597033 | -6.06856 | 2.76E-08 | 1.73E-07 | 8.584909 |
| CIAPIN1 | 0.219101 | 1.164008 | 7.417912 | 6.067538 | 2.77E-08 | 1.73E-07 | 8.580482 |
| KIAA1467 | -0.27206 | 0.828135 | 6.452637 | -6.06522 | 2.80E-08 | 1.75E-07 | 8.570448 |
| ZBTB7B | -0.24839 | 0.841835 | 5.37956 | -6.06486 | 2.80E-08 | 1.75E-07 | 8.568851 |
| SLC13A4 | -0.22576 | 0.855145 | 5.319231 | -6.05939 | 2.87E-08 | 1.79E-07 | 8.545154 |
| LPO | -0.20083 | 0.870049 | 4.823297 | -6.05696 | 2.90E-08 | 1.81E-07 | 8.534624 |
| GCA | 0.309319 | 1.239123 | 6.142308 | 6.056072 | 2.91E-08 | 1.82E-07 | 8.530765 |
| CFI | 0.353126 | 1.277325 | 10.54527 | 6.056065 | 2.91E-08 | 1.82E-07 | 8.530739 |
| NFKBIA | -0.48726 | 0.713378 | 9.602308 | -6.0543 | 2.94E-08 | 1.83E-07 | 8.523102 |
| ADAMTS1 | 1.047802 | 2.067378 | 6.999231 | 6.052669 | 2.96E-08 | 1.84E-07 | 8.51602 |
| GRIP2 | -0.24775 | 0.842207 | 6.518571 | -6.0502 | 2.99E-08 | 1.86E-07 | 8.505316 |
| LIPF | -0.18983 | 0.876708 | 4.253736 | -6.05011 | 2.99E-08 | 1.86E-07 | 8.504924 |
| DNASE2 | 0.241768 | 1.182441 | 6.509121 | 6.049743 | 3.00E-08 | 1.86E-07 | 8.503343 |

| | | | | | | | |
|---|---|---|---|---|---|---|---|
| CD1B | -0.22905 | 0.853198 | 5.77956 | -6.04868 | 3.01E-08 | 1.87E-07 | 8.498745 |
| PARD6A | -0.19921 | 0.871026 | 6.526923 | -6.04819 | 3.02E-08 | 1.87E-07 | 8.496617 |
| PRKACG | -0.21483 | 0.861647 | 5.146374 | -6.04682 | 3.04E-08 | 1.88E-07 | 8.490703 |
| OLFM4 | 1.60585 | 3.043751 | 5.697363 | 6.046428 | 3.04E-08 | 1.88E-07 | 8.488983 |
| MYOZ3 | -0.2162 | 0.860831 | 6.074176 | -6.04554 | 3.05E-08 | 1.89E-07 | 8.485152 |
| SPTBN4 | -0.17626 | 0.884994 | 5.581099 | -6.04405 | 3.07E-08 | 1.90E-07 | 8.478695 |
| EI24 | 0.179971 | 1.132861 | 7.687692 | 6.043437 | 3.08E-08 | 1.90E-07 | 8.47603 |
| FOXO1 | -0.27759 | 0.824968 | 7.391209 | -6.04126 | 3.11E-08 | 1.92E-07 | 8.466594 |
| SSBP1 | 0.225527 | 1.169204 | 9.47022 | 6.040523 | 3.12E-08 | 1.93E-07 | 8.463414 |
| CGA | -0.56532 | 0.675806 | 5.421099 | -6.04034 | 3.12E-08 | 1.93E-07 | 8.462638 |
| NDRG1 | -0.32257 | 0.799644 | 11.17484 | -6.03956 | 3.14E-08 | 1.93E-07 | 8.45923 |
| LLGL1 | -0.22547 | 0.855317 | 6.362418 | -6.03951 | 3.14E-08 | 1.93E-07 | 8.459049 |
| ACTG1 | 0.34644 | 1.271419 | 12.2711 | 6.039499 | 3.14E-08 | 1.93E-07 | 8.458984 |
| ASF1A | 0.29956 | 1.230769 | 6.898352 | 6.039357 | 3.14E-08 | 1.93E-07 | 8.458366 |
| GML | -0.24254 | 0.845258 | 4.955934 | -6.03765 | 3.16E-08 | 1.94E-07 | 8.450964 |
| DCSTAMP | -0.1901 | 0.876544 | 4.661429 | -6.037 | 3.17E-08 | 1.95E-07 | 8.44817 |
| FGL2 | 0.639826 | 1.558141 | 8.198571 | 6.035609 | 3.19E-08 | 1.96E-07 | 8.442143 |
| ZNF384 | -0.16827 | 0.889912 | 7.834835 | -6.03552 | 3.19E-08 | 1.96E-07 | 8.441749 |
| MAP3K1 | 0.2797 | 1.213943 | 5.244835 | 6.035266 | 3.20E-08 | 1.96E-07 | 8.440661 |
| CHRM5 | -0.25946 | 0.835398 | 4.455824 | -6.03481 | 3.20E-08 | 1.96E-07 | 8.438706 |
| ELL2 | -0.58695 | 0.665748 | 6.246923 | -6.03472 | 3.20E-08 | 1.96E-07 | 8.438311 |
| YIPF1 | 0.223522 | 1.16758 | 7.739011 | 6.033474 | 3.22E-08 | 1.97E-07 | 8.432904 |
| OVOL1 | -0.27591 | 0.825927 | 6.357473 | -6.03257 | 3.23E-08 | 1.98E-07 | 8.429006 |
| PNP | -0.37875 | 0.769102 | 9.978791 | -6.03117 | 3.25E-08 | 1.99E-07 | 8.422956 |
| THBS2 | 0.817821 | 1.762742 | 6.986374 | 6.027322 | 3.31E-08 | 2.02E-07 | 8.406292 |
| BLOC1S1 | -0.22514 | 0.855515 | 8.987582 | -6.0273 | 3.31E-08 | 2.02E-07 | 8.406217 |
| CCL16 | -0.22779 | 0.853944 | 4.416923 | -6.02246 | 3.38E-08 | 2.06E-07 | 8.385288 |
| TRIM48 | -0.42196 | 0.746409 | 5.203077 | -6.02212 | 3.39E-08 | 2.06E-07 | 8.383799 |
| CRYBB1 | -0.24828 | 0.841902 | 5.144835 | -6.02151 | 3.40E-08 | 2.07E-07 | 8.38116 |
| RBM28 | 0.232232 | 1.174651 | 6.015275 | 6.019834 | 3.42E-08 | 2.08E-07 | 8.373916 |
| DOT1L | -0.22867 | 0.853423 | 4.736923 | -6.01949 | 3.43E-08 | 2.08E-07 | 8.372429 |
| CCT3 | 0.218899 | 1.163845 | 9.828681 | 6.017919 | 3.45E-08 | 2.10E-07 | 8.365636 |
| THOC5 | 0.264575 | 1.201282 | 7.146703 | 6.015204 | 3.49E-08 | 2.12E-07 | 8.353904 |
| HNRNPAB | 0.310488 | 1.240127 | 9.435495 | 6.014709 | 3.50E-08 | 2.12E-07 | 8.351763 |
| SACS | 0.396169 | 1.316009 | 4.253516 | 6.012201 | 3.54E-08 | 2.14E-07 | 8.340926 |
| YOD1 | -0.23733 | 0.848315 | 4.262857 | -6.012 | 3.54E-08 | 2.15E-07 | 8.340064 |
| CARHSP1 | 0.387377 | 1.308013 | 8.323516 | 6.011505 | 3.55E-08 | 2.15E-07 | 8.337921 |
| ALPPL2 | -0.23832 | 0.84773 | 5.467582 | -6.0103 | 3.57E-08 | 2.16E-07 | 8.332707 |
| PPWD1 | 0.236725 | 1.178314 | 5.64967 | 6.008592 | 3.60E-08 | 2.17E-07 | 8.325336 |
| ACOX2 | -0.48175 | 0.716109 | 9.253077 | -6.00769 | 3.61E-08 | 2.18E-07 | 8.321431 |
| TMEM185 | 0.28372 | 1.21733 | 5.097692 | 6.006693 | 3.63E-08 | 2.19E-07 | 8.317134 |
| USP22 | 0.226691 | 1.170148 | 8.473187 | 6.003803 | 3.68E-08 | 2.22E-07 | 8.304655 |
| DECR2 | -0.45785 | 0.72807 | 8.93033 | -6.00181 | 3.71E-08 | 2.23E-07 | 8.296058 |
| PRC1 | 0.526522 | 1.440452 | 4.563846 | 6.001553 | 3.71E-08 | 2.23E-07 | 8.294942 |
| HOXC8 | -0.22913 | 0.853149 | 4.995824 | -5.99937 | 3.75E-08 | 2.25E-07 | 8.285538 |
| PRKCA | -0.18159 | 0.881731 | 5.808681 | -5.99729 | 3.78E-08 | 2.27E-07 | 8.276525 |
| MAN1B1 | 0.187686 | 1.138935 | 6.908681 | 5.996232 | 3.80E-08 | 2.28E-07 | 8.271974 |

| | | | | | | | |
|---|---|---|---|---|---|---|---|
| KCNN1 | -0.20712 | 0.866264 | 5.647143 | -5.99601 | 3.81E-08 | 2.28E-07 | 8.270995 |
| KLK7 | -0.43736 | 0.738486 | 5.472637 | -5.99581 | 3.81E-08 | 2.28E-07 | 8.27016 |
| ALOX15B | -0.20814 | 0.865653 | 5.200769 | -5.99553 | 3.81E-08 | 2.28E-07 | 8.268929 |
| PRPF18 | 0.266222 | 1.202654 | 7.231648 | 5.994494 | 3.83E-08 | 2.29E-07 | 8.264476 |
| SLC35D2 | 0.238271 | 1.179578 | 7.32 | 5.994487 | 3.83E-08 | 2.29E-07 | 8.264445 |
| MED6 | 0.250246 | 1.18941 | 5.714835 | 5.993726 | 3.84E-08 | 2.30E-07 | 8.26116 |
| TACC1 | -0.37892 | 0.769012 | 8.351099 | -5.9937 | 3.84E-08 | 2.30E-07 | 8.261036 |
| TYK2 | -0.17872 | 0.883484 | 7.699011 | -5.99208 | 3.87E-08 | 2.31E-07 | 8.254068 |
| WDR62 | -0.17767 | 0.884129 | 5.456923 | -5.98723 | 3.96E-08 | 2.36E-07 | 8.233147 |
| SLC1A6 | -0.18225 | 0.881327 | 5.168571 | -5.98597 | 3.98E-08 | 2.37E-07 | 8.227715 |
| COL11A2 | -0.2031 | 0.868684 | 5.70022 | -5.98494 | 4.00E-08 | 2.38E-07 | 8.223252 |
| PRKAR1B | -0.16098 | 0.89442 | 5.486484 | -5.98289 | 4.03E-08 | 2.40E-07 | 8.214442 |
| PSMA4 | 0.209304 | 1.156131 | 9.152198 | 5.982103 | 4.05E-08 | 2.41E-07 | 8.211034 |
| CLEC1A | -0.27883 | 0.824261 | 5.004945 | -5.9781 | 4.12E-08 | 2.45E-07 | 8.193778 |
| ITPKB | -0.28296 | 0.821905 | 5.507033 | -5.97808 | 4.12E-08 | 2.45E-07 | 8.193686 |
| ASIC4 | -0.22717 | 0.85431 | 5.465055 | -5.97805 | 4.12E-08 | 2.45E-07 | 8.193563 |
| MAN2A1 | 0.302816 | 1.23355 | 8.276484 | 5.977614 | 4.13E-08 | 2.45E-07 | 8.191684 |
| PROS1 | 0.366271 | 1.289016 | 7.583297 | 5.976575 | 4.15E-08 | 2.46E-07 | 8.187207 |
| BEGAIN | -0.26741 | 0.830812 | 5.294505 | -5.97625 | 4.15E-08 | 2.46E-07 | 8.185825 |
| GPR143 | 0.450836 | 1.366832 | 5.870549 | 5.975093 | 4.18E-08 | 2.47E-07 | 8.18082 |
| MAP4K2 | -0.22325 | 0.856633 | 5.053297 | -5.97489 | 4.18E-08 | 2.47E-07 | 8.179927 |
| THOC1 | 0.200812 | 1.149345 | 7.355824 | 5.974654 | 4.18E-08 | 2.47E-07 | 8.178928 |
| PRSS53 | -0.22636 | 0.85479 | 5.711978 | -5.97219 | 4.23E-08 | 2.50E-07 | 8.168294 |
| UTP3 | 0.23371 | 1.175855 | 7.732527 | 5.97152 | 4.24E-08 | 2.50E-07 | 8.165427 |
| KPNA2 | 0.589145 | 1.504355 | 7.822857 | 5.969655 | 4.28E-08 | 2.52E-07 | 8.157397 |
| IFNAR2 | 0.223633 | 1.16767 | 5.604066 | 5.96567 | 4.35E-08 | 2.57E-07 | 8.140233 |
| PPIEL | -0.2344 | 0.850038 | 4.546044 | -5.96487 | 4.37E-08 | 2.57E-07 | 8.136804 |
| AZIN1 | 0.282372 | 1.216193 | 8.156374 | 5.964271 | 4.38E-08 | 2.58E-07 | 8.13421 |
| HADH | -0.37666 | 0.77022 | 10.14418 | -5.96418 | 4.38E-08 | 2.58E-07 | 8.133802 |
| WDR3 | 0.227488 | 1.170795 | 6.989451 | 5.96011 | 4.46E-08 | 2.62E-07 | 8.116298 |
| ELOVL6 | 0.320899 | 1.249108 | 5.269121 | 5.959215 | 4.48E-08 | 2.63E-07 | 8.112446 |
| MYO15A | -0.18678 | 0.878566 | 4.242637 | -5.9591 | 4.48E-08 | 2.63E-07 | 8.111941 |
| GPR12 | -0.22694 | 0.854447 | 5.085604 | -5.95895 | 4.48E-08 | 2.63E-07 | 8.111319 |
| TBX2 | -0.25324 | 0.839009 | 6.468901 | -5.9589 | 4.49E-08 | 2.63E-07 | 8.111101 |
| MED4 | 0.224357 | 1.168257 | 6.597033 | 5.955955 | 4.54E-08 | 2.66E-07 | 8.098418 |
| KIF1BP | 0.239353 | 1.180463 | 7.749231 | 5.955481 | 4.55E-08 | 2.67E-07 | 8.09638 |
| APLP1 | -0.24514 | 0.843737 | 5.387692 | -5.9485 | 4.70E-08 | 2.75E-07 | 8.066355 |
| MITF | 0.269106 | 1.205061 | 7.572857 | 5.947732 | 4.71E-08 | 2.76E-07 | 8.063048 |
| CNOT8 | 0.224923 | 1.168715 | 6.652747 | 5.947141 | 4.73E-08 | 2.76E-07 | 8.060504 |
| FAM193A | 0.257908 | 1.195744 | 5.963407 | 5.946692 | 4.74E-08 | 2.76E-07 | 8.058576 |
| PPT2 | -0.189 | 0.877216 | 7.721758 | -5.94591 | 4.75E-08 | 2.77E-07 | 8.055203 |
| HAUS4 | -0.38184 | 0.76746 | 7.798571 | -5.94494 | 4.77E-08 | 2.78E-07 | 8.051032 |
| SOWAHC | -0.39796 | 0.75893 | 7.344945 | -5.94335 | 4.81E-08 | 2.80E-07 | 8.044229 |
| GDF9 | -0.22653 | 0.854687 | 5.174066 | -5.94275 | 4.82E-08 | 2.81E-07 | 8.041636 |
| APBA1 | -0.19785 | 0.871849 | 5.278901 | -5.94173 | 4.84E-08 | 2.82E-07 | 8.037259 |
| MAP3K12 | -0.18189 | 0.881545 | 6.534835 | -5.94118 | 4.85E-08 | 2.82E-07 | 8.034877 |
| CCDC70 | -0.20686 | 0.866424 | 4.119231 | -5.94117 | 4.85E-08 | 2.82E-07 | 8.034845 |

| | | | | | | | |
|---|---|---|---|---|---|---|---|
| NDUFS6 | 0.192797 | 1.142978 | 10.07121 | 5.940826 | 4.86E-08 | 2.82E-07 | 8.033359 |
| INPP5A | 0.229188 | 1.172175 | 7.736813 | 5.93807 | 4.92E-08 | 2.85E-07 | 8.021515 |
| P3H1 | 0.215802 | 1.161349 | 6.477802 | 5.937724 | 4.93E-08 | 2.85E-07 | 8.02003 |
| RARRES1 | 1.135348 | 2.196715 | 5.862088 | 5.937702 | 4.93E-08 | 2.85E-07 | 8.019935 |
| PCDHGA1 | -0.24662 | 0.84287 | 4.49022 | -5.93734 | 4.94E-08 | 2.86E-07 | 8.018381 |
| DDX39A | 0.295333 | 1.227168 | 6.896044 | 5.935427 | 4.98E-08 | 2.88E-07 | 8.010158 |
| OCEL1 | -0.24692 | 0.842695 | 8.266593 | -5.93475 | 4.99E-08 | 2.89E-07 | 8.007258 |
| PKM | 0.463734 | 1.379107 | 8.497363 | 5.933942 | 5.01E-08 | 2.89E-07 | 8.003781 |
| LOC72916 | -0.17912 | 0.883241 | 4.810989 | -5.93376 | 5.01E-08 | 2.90E-07 | 8.003019 |
| SCAF8 | 0.2293 | 1.172266 | 7.651868 | 5.931518 | 5.06E-08 | 2.92E-07 | 7.993367 |
| RGR | -0.20816 | 0.865641 | 4.50989 | -5.93133 | 5.07E-08 | 2.92E-07 | 7.992582 |
| PROL1 | -0.22493 | 0.855635 | 4.676813 | -5.92995 | 5.10E-08 | 2.94E-07 | 7.986634 |
| NCK2 | 0.187406 | 1.138714 | 7.875934 | 5.928548 | 5.13E-08 | 2.95E-07 | 7.980612 |
| CEP83 | 0.219406 | 1.164254 | 4.451758 | 5.92688 | 5.17E-08 | 2.97E-07 | 7.973454 |
| TREML2 | -0.2286 | 0.853463 | 5.51 | -5.9264 | 5.18E-08 | 2.98E-07 | 7.971407 |
| C11orf16 | -0.16957 | 0.889108 | 6.375495 | -5.92444 | 5.22E-08 | 3.00E-07 | 7.962978 |
| NEK11 | 0.218643 | 1.163638 | 6.057033 | 5.923843 | 5.24E-08 | 3.01E-07 | 7.960414 |
| PFKP | 0.633401 | 1.551217 | 7.866264 | 5.922111 | 5.28E-08 | 3.03E-07 | 7.952979 |
| METTL1 | -0.34138 | 0.789285 | 8.139011 | -5.92106 | 5.30E-08 | 3.04E-07 | 7.94849 |
| CTRL | -0.21354 | 0.862421 | 4.755275 | -5.92078 | 5.31E-08 | 3.04E-07 | 7.947275 |
| KLF15 | -0.29842 | 0.813145 | 6.843736 | -5.9204 | 5.32E-08 | 3.05E-07 | 7.945625 |
| ROGDI | -0.35086 | 0.784119 | 7.688022 | -5.92018 | 5.32E-08 | 3.05E-07 | 7.944684 |
| OR10J1 | -0.19146 | 0.875717 | 5.069451 | -5.91945 | 5.34E-08 | 3.05E-07 | 7.941568 |
| COMMD9 | 0.177585 | 1.130989 | 8.259121 | 5.918866 | 5.35E-08 | 3.06E-07 | 7.939057 |
| PLD2 | -0.17521 | 0.88564 | 6.009011 | -5.91873 | 5.36E-08 | 3.06E-07 | 7.938492 |
| PELO | 0.237271 | 1.17876 | 7.254505 | 5.917884 | 5.38E-08 | 3.07E-07 | 7.934841 |
| GPR162 | -0.18863 | 0.877437 | 5.338242 | -5.91725 | 5.39E-08 | 3.08E-07 | 7.932102 |
| RTEL1 | -0.21814 | 0.859673 | 7.365824 | -5.91648 | 5.41E-08 | 3.08E-07 | 7.92881 |
| NXPH3 | -0.24042 | 0.846499 | 6.782198 | -5.91363 | 5.48E-08 | 3.12E-07 | 7.916613 |
| SPAST | 0.210932 | 1.157436 | 4.866264 | 5.91219 | 5.51E-08 | 3.14E-07 | 7.910416 |
| NOS1 | -0.19147 | 0.875711 | 5.509011 | -5.91168 | 5.53E-08 | 3.14E-07 | 7.90825 |
| TBX6 | -0.22326 | 0.85663 | 5.313077 | -5.91076 | 5.55E-08 | 3.16E-07 | 7.904298 |
| CDS1 | 0.271826 | 1.207335 | 5.996593 | 5.909556 | 5.58E-08 | 3.17E-07 | 7.899124 |
| MCM2 | 0.335469 | 1.261787 | 6.311978 | 5.90675 | 5.65E-08 | 3.21E-07 | 7.887092 |
| TIMM17A | 0.153841 | 1.112527 | 7.588901 | 5.906439 | 5.66E-08 | 3.21E-07 | 7.885759 |
| PIP | -0.15543 | 0.897868 | 6.139011 | -5.90442 | 5.71E-08 | 3.24E-07 | 7.877111 |
| SNW1 | 0.194024 | 1.14395 | 7.557033 | 5.903299 | 5.74E-08 | 3.25E-07 | 7.872301 |
| FERMT2 | -0.27712 | 0.825236 | 7.782527 | -5.90261 | 5.75E-08 | 3.26E-07 | 7.869354 |
| RMI1 | 0.251676 | 1.19059 | 6.18989 | 5.902225 | 5.76E-08 | 3.26E-07 | 7.867697 |
| AQR | 0.213865 | 1.159791 | 7.13967 | 5.902092 | 5.77E-08 | 3.26E-07 | 7.867128 |
| FGF1 | -0.77661 | 0.583737 | 6.638571 | -5.90172 | 5.78E-08 | 3.26E-07 | 7.865525 |
| BTNL2 | -0.21934 | 0.858957 | 4.371099 | -5.90166 | 5.78E-08 | 3.26E-07 | 7.865281 |
| ITGB1BP1 | 0.263469 | 1.200361 | 6.226374 | 5.901632 | 5.78E-08 | 3.26E-07 | 7.865157 |
| AASDHPPT | 0.246406 | 1.186248 | 8.45011 | 5.900289 | 5.81E-08 | 3.28E-07 | 7.859404 |
| TSC22D3 | -0.80617 | 0.571897 | 8.823516 | -5.89795 | 5.87E-08 | 3.31E-07 | 7.849399 |
| SUCLG1 | -0.26855 | 0.830153 | 12.21242 | -5.89724 | 5.89E-08 | 3.32E-07 | 7.84635 |
| TUBB4A | -0.2776 | 0.824963 | 5.410769 | -5.89711 | 5.89E-08 | 3.32E-07 | 7.845771 |

| | | | | | | | |
|---|---|---|---|---|---|---|---|
| CD151 | 0.455705 | 1.371453 | 7.802088 | 5.895303 | 5.94E-08 | 3.34E-07 | 7.83804 |
| CLPB | -0.24741 | 0.842407 | 6.054176 | -5.89442 | 5.96E-08 | 3.35E-07 | 7.834279 |
| CTF1 | -0.28475 | 0.820882 | 6.195275 | -5.89383 | 5.98E-08 | 3.36E-07 | 7.831724 |
| KIRREL | -0.17214 | 0.887528 | 4.828791 | -5.89315 | 6.00E-08 | 3.37E-07 | 7.828839 |
| DLGAP5 | 0.392531 | 1.312695 | 3.778022 | 5.891722 | 6.04E-08 | 3.39E-07 | 7.822708 |
| ZCCHC6 | 0.220836 | 1.165409 | 6.516813 | 5.889215 | 6.10E-08 | 3.42E-07 | 7.811973 |
| S1PR4 | -0.206 | 0.86694 | 5.848352 | -5.8878 | 6.14E-08 | 3.44E-07 | 7.805929 |
| EIF2B1 | 0.176246 | 1.12994 | 8.158242 | 5.887303 | 6.15E-08 | 3.45E-07 | 7.803788 |
| SERPINA6 | -0.81437 | 0.568658 | 8.630769 | -5.88596 | 6.19E-08 | 3.46E-07 | 7.798052 |
| GNG12 | 0.240164 | 1.181127 | 9.376154 | 5.885875 | 6.19E-08 | 3.46E-07 | 7.797675 |
| MKL2 | 0.267377 | 1.203617 | 7.384176 | 5.885504 | 6.20E-08 | 3.47E-07 | 7.796088 |
| IMPDH2 | 0.340014 | 1.265769 | 9.048791 | 5.885274 | 6.21E-08 | 3.47E-07 | 7.795102 |
| CYBA | 0.391005 | 1.311306 | 9.818571 | 5.885009 | 6.22E-08 | 3.47E-07 | 7.79397 |
| CRTC3 | 0.219884 | 1.16464 | 7.803516 | 5.884368 | 6.23E-08 | 3.48E-07 | 7.791224 |
| RHOBTB1 | 0.514348 | 1.428348 | 8.7 | 5.884203 | 6.24E-08 | 3.48E-07 | 7.79052 |
| CRMP1 | -0.2076 | 0.865977 | 6.615385 | -5.88405 | 6.24E-08 | 3.48E-07 | 7.789853 |
| FRAT2 | 0.316043 | 1.244912 | 5.468242 | 5.883849 | 6.25E-08 | 3.48E-07 | 7.789006 |
| MMD | 0.496367 | 1.410657 | 7.351978 | 5.883297 | 6.26E-08 | 3.48E-07 | 7.786643 |
| NFE2L2 | 0.262454 | 1.199517 | 9.32022 | 5.879166 | 6.38E-08 | 3.55E-07 | 7.768968 |
| GPR3 | -0.24644 | 0.842974 | 5.108352 | -5.87842 | 6.40E-08 | 3.55E-07 | 7.765779 |
| SLC39A1 | 0.212058 | 1.158339 | 7.487473 | 5.878342 | 6.40E-08 | 3.55E-07 | 7.765441 |
| MT1HL1 | -0.28883 | 0.818565 | 7.63978 | -5.87766 | 6.42E-08 | 3.56E-07 | 7.762515 |
| ZNF771 | -0.2341 | 0.850214 | 6.87967 | -5.87655 | 6.45E-08 | 3.58E-07 | 7.757781 |
| ZNF33B | 0.311256 | 1.240787 | 6.12044 | 5.87641 | 6.46E-08 | 3.58E-07 | 7.75718 |
| ALAD | -0.41276 | 0.751183 | 7.571538 | -5.87559 | 6.48E-08 | 3.59E-07 | 7.753691 |
| RPIA | 0.2597 | 1.19723 | 6.874945 | 5.874376 | 6.52E-08 | 3.60E-07 | 7.748482 |
| HSD17B14 | -0.41883 | 0.748031 | 7.915495 | -5.87436 | 6.52E-08 | 3.60E-07 | 7.748413 |
| USP33 | 0.19042 | 1.141096 | 8.163077 | 5.874051 | 6.52E-08 | 3.61E-07 | 7.747091 |
| RHOF | 0.312415 | 1.241785 | 6.443187 | 5.873682 | 6.54E-08 | 3.61E-07 | 7.745513 |
| ACTR6 | 0.277411 | 1.212018 | 6.990659 | 5.872203 | 6.58E-08 | 3.63E-07 | 7.739189 |
| FPGT | 0.283406 | 1.217065 | 5.943407 | 5.872109 | 6.58E-08 | 3.63E-07 | 7.738787 |
| SEMA4G | -0.21471 | 0.861722 | 5.792088 | -5.87171 | 6.59E-08 | 3.63E-07 | 7.737075 |
| METTL18 | 0.266652 | 1.203013 | 5.721209 | 5.871328 | 6.60E-08 | 3.64E-07 | 7.735447 |
| GDAP2 | -0.20665 | 0.866546 | 5.438462 | -5.87097 | 6.61E-08 | 3.64E-07 | 7.733932 |
| CCT8 | 0.219604 | 1.164414 | 11.33077 | 5.870948 | 6.61E-08 | 3.64E-07 | 7.733823 |
| APOBEC3F | 0.243638 | 1.183974 | 4.254176 | 5.86987 | 6.65E-08 | 3.65E-07 | 7.729215 |
| SRD5A2 | -0.22554 | 0.855277 | 5.379341 | -5.869 | 6.67E-08 | 3.66E-07 | 7.725513 |
| GNAS | 0.198082 | 1.147172 | 11.18165 | 5.868732 | 6.68E-08 | 3.67E-07 | 7.724351 |
| SLC34A1 | -0.7245 | 0.605208 | 8.949341 | -5.86686 | 6.73E-08 | 3.69E-07 | 7.716356 |
| PYCR1 | -0.20647 | 0.866656 | 6.406813 | -5.86675 | 6.74E-08 | 3.69E-07 | 7.715859 |
| RHOC | 0.268425 | 1.204492 | 9.991868 | 5.866488 | 6.75E-08 | 3.69E-07 | 7.714759 |
| NPC1L1 | -0.19824 | 0.871612 | 4.801099 | -5.86641 | 6.75E-08 | 3.69E-07 | 7.71444 |
| USP1 | 0.260357 | 1.197775 | 7.244835 | 5.866321 | 6.75E-08 | 3.69E-07 | 7.714046 |
| MAPK8IP2 | -0.19485 | 0.873667 | 4.935604 | -5.86474 | 6.80E-08 | 3.72E-07 | 7.707311 |
| CYFIP2 | -0.44208 | 0.736074 | 9.087912 | -5.86433 | 6.81E-08 | 3.72E-07 | 7.705545 |
| ATP9A | 0.407657 | 1.32653 | 6.726154 | 5.863753 | 6.83E-08 | 3.73E-07 | 7.703072 |
| KIF2A | 0.199729 | 1.148483 | 5.386593 | 5.859955 | 6.94E-08 | 3.79E-07 | 7.686847 |

| | | | | | | | |
|---|---|---|---|---|---|---|---|
| GUCA1B | -0.16989 | 0.888908 | 4.870769 | -5.85751 | 7.02E-08 | 3.83E-07 | 7.676385 |
| HOXB1 | -0.27835 | 0.824532 | 5.816484 | -5.85598 | 7.06E-08 | 3.85E-07 | 7.669855 |
| RBFOX2 | 0.22471 | 1.168542 | 7.293077 | 5.853834 | 7.13E-08 | 3.88E-07 | 7.660706 |
| C1orf105 | -0.23772 | 0.848082 | 5.224835 | -5.85265 | 7.17E-08 | 3.90E-07 | 7.655662 |
| ST3GAL6 | -0.22155 | 0.857646 | 5.180879 | -5.85263 | 7.17E-08 | 3.90E-07 | 7.655583 |
| TCF12 | 0.244464 | 1.184652 | 8.141758 | 5.851805 | 7.20E-08 | 3.91E-07 | 7.652044 |
| FSCN3 | -0.20927 | 0.864974 | 5.199341 | -5.85097 | 7.22E-08 | 3.92E-07 | 7.64849 |
| CDC25B | 0.412874 | 1.331336 | 6.643516 | 5.850388 | 7.24E-08 | 3.93E-07 | 7.645995 |
| RBM4B | 0.308444 | 1.238372 | 6.922527 | 5.850365 | 7.24E-08 | 3.93E-07 | 7.6459 |
| DENND5B | -0.18075 | 0.882242 | 4.346703 | -5.84806 | 7.31E-08 | 3.97E-07 | 7.636073 |
| CLUHP3 | -0.27373 | 0.827176 | 6.536593 | -5.84743 | 7.34E-08 | 3.98E-07 | 7.633367 |
| PDIA5 | 0.363937 | 1.286933 | 7.237143 | 5.845817 | 7.39E-08 | 4.00E-07 | 7.62649 |
| OLFML2A | -0.29958 | 0.812489 | 6.080769 | -5.8438 | 7.45E-08 | 4.03E-07 | 7.617899 |
| STK26 | 0.313488 | 1.242708 | 4.251978 | 5.843577 | 7.46E-08 | 4.04E-07 | 7.616934 |
| ARMCX1 | 0.309826 | 1.239558 | 7.745385 | 5.841225 | 7.54E-08 | 4.07E-07 | 7.6069 |
| LGALS3BP | 0.414918 | 1.333223 | 8.616484 | 5.841031 | 7.54E-08 | 4.08E-07 | 7.606075 |
| KRT19 | 0.785918 | 1.724189 | 7.904945 | 5.838737 | 7.62E-08 | 4.11E-07 | 7.596288 |
| CFAP20 | 0.197179 | 1.146454 | 8.660549 | 5.837928 | 7.65E-08 | 4.13E-07 | 7.592838 |
| SUN2 | -0.23042 | 0.852387 | 8.177143 | -5.83426 | 7.77E-08 | 4.19E-07 | 7.57721 |
| UNC5C | -0.15619 | 0.89739 | 4.264066 | -5.83406 | 7.78E-08 | 4.19E-07 | 7.576358 |
| WARS2 | 0.283058 | 1.216771 | 6.117582 | 5.833141 | 7.81E-08 | 4.21E-07 | 7.572431 |
| PNLIP | -0.16754 | 0.890362 | 4.278022 | -5.83228 | 7.84E-08 | 4.22E-07 | 7.56876 |
| SEC24C | 0.205778 | 1.153308 | 7.501758 | 5.830016 | 7.92E-08 | 4.26E-07 | 7.55911 |
| LSM7 | 0.311101 | 1.240655 | 8.033407 | 5.829417 | 7.94E-08 | 4.27E-07 | 7.556559 |
| FASTKD1 | 0.316333 | 1.245162 | 8.031429 | 5.828544 | 7.97E-08 | 4.28E-07 | 7.552841 |
| CDK2AP1 | 0.250169 | 1.189346 | 9.931319 | 5.825938 | 8.06E-08 | 4.33E-07 | 7.541735 |
| SPECC1L | 0.215058 | 1.160751 | 6.873956 | 5.825675 | 8.07E-08 | 4.33E-07 | 7.540615 |
| ZBTB32 | -0.17581 | 0.885269 | 4.061538 | -5.82551 | 8.08E-08 | 4.33E-07 | 7.539922 |
| ARL2 | -0.20581 | 0.867054 | 9.096923 | -5.82472 | 8.10E-08 | 4.34E-07 | 7.536548 |
| TPD52L2 | 0.205536 | 1.153115 | 8.110769 | 5.82451 | 8.11E-08 | 4.34E-07 | 7.535652 |
| ALDOA | 0.186227 | 1.137784 | 11.23231 | 5.823137 | 8.16E-08 | 4.37E-07 | 7.529805 |
| GSTM5 | -0.29428 | 0.815482 | 5.802088 | -5.82207 | 8.20E-08 | 4.38E-07 | 7.525253 |
| DCT | -0.18117 | 0.881985 | 4.747582 | -5.8207 | 8.25E-08 | 4.41E-07 | 7.519419 |
| DLEU1 | -0.28188 | 0.822519 | 7.566044 | -5.82026 | 8.26E-08 | 4.41E-07 | 7.517544 |
| F11R | 0.208729 | 1.15567 | 7.136044 | 5.820237 | 8.27E-08 | 4.41E-07 | 7.517456 |
| CPSF6 | 0.195285 | 1.14495 | 6.474396 | 5.816759 | 8.39E-08 | 4.48E-07 | 7.502647 |
| CDC20 | 0.633966 | 1.551825 | 5.061978 | 5.813667 | 8.51E-08 | 4.53E-07 | 7.489486 |
| VSIG4 | 0.778261 | 1.715062 | 6.346593 | 5.812906 | 8.54E-08 | 4.55E-07 | 7.486246 |
| MALT1 | 0.436203 | 1.353039 | 6.214835 | 5.810839 | 8.61E-08 | 4.59E-07 | 7.477449 |
| AKAP13 | -0.27781 | 0.824844 | 6.461319 | -5.81021 | 8.64E-08 | 4.59E-07 | 7.474775 |
| IPO4 | 0.314415 | 1.243508 | 6.194176 | 5.810163 | 8.64E-08 | 4.59E-07 | 7.474574 |
| SNX7 | 0.264952 | 1.201596 | 9.073846 | 5.806093 | 8.79E-08 | 4.67E-07 | 7.45726 |
| APOBEC2 | -0.23386 | 0.850357 | 5.160659 | -5.80422 | 8.87E-08 | 4.71E-07 | 7.449276 |
| THEM6 | -0.2536 | 0.838801 | 6.812637 | -5.80277 | 8.92E-08 | 4.73E-07 | 7.443134 |
| GJC2 | -0.22429 | 0.856013 | 6.235824 | -5.80272 | 8.92E-08 | 4.73E-07 | 7.442918 |
| ALDH7A1 | -0.272 | 0.828173 | 6.325714 | -5.80218 | 8.95E-08 | 4.74E-07 | 7.440617 |
| NGF | -0.24081 | 0.846272 | 5.104615 | -5.80115 | 8.99E-08 | 4.76E-07 | 7.436255 |

| | | | | | | | |
|---|---|---|---|---|---|---|---|
| ADIPOR1 | -0.24601 | 0.843225 | 8.307912 | -5.8003 | 9.02E-08 | 4.77E-07 | 7.432642 |
| ECI2 | -0.31032 | 0.806461 | 10.60198 | -5.7973 | 9.14E-08 | 4.83E-07 | 7.419856 |
| TFAP4 | -0.22774 | 0.853972 | 6.529121 | -5.7966 | 9.17E-08 | 4.85E-07 | 7.416897 |
| C8orf33 | 0.329763 | 1.256807 | 6.022418 | 5.794288 | 9.26E-08 | 4.89E-07 | 7.407072 |
| DTX4 | 0.374976 | 1.296818 | 8.079341 | 5.793595 | 9.29E-08 | 4.90E-07 | 7.404124 |
| TBC1D16 | 0.333894 | 1.260411 | 6.44033 | 5.793042 | 9.31E-08 | 4.91E-07 | 7.401776 |
| PVALB | -0.98908 | 0.5038 | 7.762418 | -5.79034 | 9.42E-08 | 4.97E-07 | 7.390282 |
| WNT2 | -0.2124 | 0.8631 | 5.496923 | -5.7899 | 9.44E-08 | 4.98E-07 | 7.388425 |
| CSNK1G3 | 0.259072 | 1.196709 | 6.866374 | 5.787115 | 9.56E-08 | 5.03E-07 | 7.376599 |
| CRNKL1 | 0.294019 | 1.226051 | 7.176264 | 5.780682 | 9.83E-08 | 5.17E-07 | 7.349286 |
| PLXNA2 | -0.22181 | 0.857488 | 5.718791 | -5.77771 | 9.96E-08 | 5.24E-07 | 7.336651 |
| NXN | 0.290188 | 1.2228 | 6.271978 | 5.776807 | 1.00E-07 | 5.26E-07 | 7.33284 |
| HOXA10 | 0.234638 | 1.176611 | 6.954725 | 5.775526 | 1.01E-07 | 5.28E-07 | 7.327402 |
| CD53 | 0.747502 | 1.678884 | 7.697253 | 5.775012 | 1.01E-07 | 5.29E-07 | 7.325221 |
| PDE10A | -0.17121 | 0.888099 | 4.800989 | -5.77163 | 1.02E-07 | 5.37E-07 | 7.310858 |
| PNPLA2 | -0.17743 | 0.884277 | 7.347912 | -5.76885 | 1.04E-07 | 5.43E-07 | 7.299066 |
| CXCL6 | 1.506957 | 2.842098 | 4.768242 | 5.767982 | 1.04E-07 | 5.45E-07 | 7.295404 |
| SDPR | -0.31864 | 0.801827 | 5.323736 | -5.76747 | 1.04E-07 | 5.45E-07 | 7.293247 |
| IL1RAPL1 | -0.23517 | 0.849583 | 4.820879 | -5.76739 | 1.04E-07 | 5.45E-07 | 7.292881 |
| RNASET2 | 0.347343 | 1.272215 | 8.122198 | 5.766221 | 1.05E-07 | 5.48E-07 | 7.287937 |
| STARD8 | -0.29918 | 0.812715 | 6.939011 | -5.76499 | 1.05E-07 | 5.50E-07 | 7.282705 |
| RTP4 | 0.514517 | 1.428516 | 5.517692 | 5.764732 | 1.05E-07 | 5.51E-07 | 7.281622 |
| PRLH | -0.28961 | 0.818121 | 5.434176 | -5.76399 | 1.06E-07 | 5.52E-07 | 7.278471 |
| MSH6 | 0.308232 | 1.238189 | 6.682857 | 5.758956 | 1.08E-07 | 5.64E-07 | 7.257141 |
| ATP13A3 | 0.25056 | 1.189669 | 7.629121 | 5.75847 | 1.08E-07 | 5.65E-07 | 7.255082 |
| LDLR | -0.5087 | 0.702858 | 5.257143 | -5.753 | 1.11E-07 | 5.78E-07 | 7.231895 |
| IDH3B | -0.16903 | 0.889441 | 9.25011 | -5.75281 | 1.11E-07 | 5.78E-07 | 7.231091 |
| UBE2A | 0.175783 | 1.129577 | 8.307143 | 5.751562 | 1.12E-07 | 5.81E-07 | 7.225819 |
| C1orf68 | -0.20073 | 0.87011 | 5.447912 | -5.75109 | 1.12E-07 | 5.82E-07 | 7.223839 |
| RAB3IL1 | -0.22644 | 0.854742 | 7.088242 | -5.75052 | 1.12E-07 | 5.83E-07 | 7.221398 |
| SSB | 0.248816 | 1.188232 | 7.62978 | 5.749368 | 1.13E-07 | 5.86E-07 | 7.21653 |
| ZNF43 | 0.206565 | 1.153938 | 5.717582 | 5.749208 | 1.13E-07 | 5.86E-07 | 7.215852 |
| FN3K | -0.22123 | 0.857836 | 4.205385 | -5.74871 | 1.13E-07 | 5.87E-07 | 7.213729 |
| TRAJ28 | -0.28069 | 0.823197 | 7.50011 | -5.74774 | 1.13E-07 | 5.89E-07 | 7.209655 |
| TMEM123 | 0.332754 | 1.259415 | 8.628462 | 5.746963 | 1.14E-07 | 5.90E-07 | 7.206348 |
| TRPV5 | -0.24313 | 0.844913 | 5.249121 | -5.74686 | 1.14E-07 | 5.90E-07 | 7.205903 |
| NTRK3 | -0.19699 | 0.872369 | 5.618022 | -5.74664 | 1.14E-07 | 5.90E-07 | 7.204964 |
| COL5A2 | 0.234541 | 1.176532 | 4.71033 | 5.746226 | 1.14E-07 | 5.91E-07 | 7.203229 |
| GPI | 0.230836 | 1.173515 | 9.831758 | 5.744267 | 1.15E-07 | 5.96E-07 | 7.194938 |
| MS4A6A | 0.667585 | 1.588411 | 6.741429 | 5.742896 | 1.16E-07 | 5.99E-07 | 7.189135 |
| OVOL3 | -0.21288 | 0.862814 | 5.295165 | -5.74136 | 1.17E-07 | 6.03E-07 | 7.182635 |
| SEMA3B | -0.32027 | 0.80092 | 7.249451 | -5.73833 | 1.18E-07 | 6.10E-07 | 7.16983 |
| COLEC11 | -0.52405 | 0.695418 | 9.168681 | -5.73823 | 1.18E-07 | 6.10E-07 | 7.169393 |
| VLDLR | 0.333889 | 1.260406 | 6.52011 | 5.736814 | 1.19E-07 | 6.14E-07 | 7.163401 |
| SNAP23 | 0.26001 | 1.197487 | 6.789011 | 5.732665 | 1.21E-07 | 6.24E-07 | 7.145858 |
| KITLG | 0.309116 | 1.238948 | 4.693077 | 5.730965 | 1.22E-07 | 6.29E-07 | 7.13867 |
| ATAT1 | -0.23626 | 0.848943 | 5.611429 | -5.7307 | 1.22E-07 | 6.29E-07 | 7.137565 |

| | | | | | | | |
|---|---|---|---|---|---|---|---|
| FAAH | -0.30929 | 0.807042 | 6.469231 | -5.73023 | 1.22E-07 | 6.30E-07 | 7.135566 |
| MTNR1A | -0.47877 | 0.717588 | 5.088462 | -5.73002 | 1.23E-07 | 6.30E-07 | 7.134681 |
| DDX47 | 0.172208 | 1.126781 | 9.054505 | 5.729299 | 1.23E-07 | 6.32E-07 | 7.131625 |
| CNTNAP1 | -0.19794 | 0.871793 | 5.584725 | -5.72804 | 1.24E-07 | 6.35E-07 | 7.126303 |
| HCLS1 | 0.654222 | 1.573767 | 6.083516 | 5.727784 | 1.24E-07 | 6.35E-07 | 7.125223 |
| CIC | -0.21567 | 0.861145 | 7.078132 | -5.72725 | 1.24E-07 | 6.36E-07 | 7.122969 |
| TCIRG1 | 0.327981 | 1.255255 | 5.431319 | 5.726771 | 1.24E-07 | 6.37E-07 | 7.120943 |
| ABCC8 | -0.24249 | 0.845284 | 5.047912 | -5.72499 | 1.25E-07 | 6.42E-07 | 7.113403 |
| MYL7 | -0.22418 | 0.856082 | 5.761099 | -5.72442 | 1.26E-07 | 6.43E-07 | 7.110987 |
| TM4SF5 | -0.39149 | 0.76234 | 8.299231 | -5.72122 | 1.27E-07 | 6.52E-07 | 7.097507 |
| LYPD1 | 0.492145 | 1.406534 | 5.22989 | 5.72082 | 1.28E-07 | 6.52E-07 | 7.095798 |
| AHR | 0.308217 | 1.238177 | 6.552198 | 5.720507 | 1.28E-07 | 6.53E-07 | 7.094476 |
| C1QB | 0.940889 | 1.919711 | 6.005275 | 5.720037 | 1.28E-07 | 6.54E-07 | 7.092488 |
| PDE1B | -0.20701 | 0.866331 | 5.522198 | -5.71795 | 1.29E-07 | 6.59E-07 | 7.083667 |
| KCNA6 | -0.22393 | 0.856231 | 5.232527 | -5.71696 | 1.30E-07 | 6.62E-07 | 7.079474 |
| ACTR10 | 0.204855 | 1.152571 | 9.71978 | 5.715486 | 1.31E-07 | 6.66E-07 | 7.07327 |
| PIGL | -0.28663 | 0.819813 | 5.69978 | -5.71533 | 1.31E-07 | 6.66E-07 | 7.072609 |
| CEBPA-AS1 | -0.24571 | 0.843398 | 6.491319 | -5.71079 | 1.33E-07 | 6.79E-07 | 7.053457 |
| CAPG | 0.522628 | 1.43657 | 8.156703 | 5.709646 | 1.34E-07 | 6.81E-07 | 7.048618 |
| SAMD4B | -0.22342 | 0.856532 | 5.935604 | -5.70955 | 1.34E-07 | 6.81E-07 | 7.048223 |
| CYP2C8 | -0.55683 | 0.679796 | 4.157473 | -5.70785 | 1.35E-07 | 6.86E-07 | 7.041035 |
| CCL25 | -0.26489 | 0.83226 | 4.668791 | -5.70705 | 1.35E-07 | 6.88E-07 | 7.03767 |
| LRRC16A | 0.379246 | 1.300662 | 7.623626 | 5.704738 | 1.37E-07 | 6.95E-07 | 7.02791 |
| HLA-DPA1 | 0.76901 | 1.7041 | 9.625714 | 5.704186 | 1.37E-07 | 6.96E-07 | 7.025582 |
| ROBO4 | -0.16745 | 0.890416 | 5.651978 | -5.70366 | 1.38E-07 | 6.97E-07 | 7.023378 |
| ACTL6A | 0.287295 | 1.22035 | 5.94033 | 5.702667 | 1.38E-07 | 7.00E-07 | 7.019172 |
| UGCG | 0.593749 | 1.509163 | 7.962308 | 5.699581 | 1.40E-07 | 7.09E-07 | 7.006158 |
| ITPA | 0.227353 | 1.170685 | 8.363297 | 5.696342 | 1.42E-07 | 7.18E-07 | 6.992502 |
| CLCN1 | -0.20411 | 0.868073 | 5.074066 | -5.69439 | 1.43E-07 | 7.24E-07 | 6.984291 |
| P3H4 | 0.232367 | 1.174761 | 5.321429 | 5.694326 | 1.43E-07 | 7.24E-07 | 6.984007 |
| NOS2 | -0.30857 | 0.807442 | 5.831758 | -5.69337 | 1.44E-07 | 7.26E-07 | 6.979981 |
| P2RY4 | -0.18901 | 0.877205 | 4.600879 | -5.69216 | 1.45E-07 | 7.30E-07 | 6.974858 |
| THBS4 | -0.26687 | 0.831118 | 5.388681 | -5.68844 | 1.47E-07 | 7.41E-07 | 6.959191 |
| PIWIL2 | -0.20041 | 0.870303 | 4.442418 | -5.68815 | 1.47E-07 | 7.42E-07 | 6.958001 |
| C5orf15 | 0.19858 | 1.147568 | 9.704286 | 5.685482 | 1.49E-07 | 7.50E-07 | 6.946744 |
| ADI1 | -0.39646 | 0.759721 | 11.4433 | -5.68509 | 1.49E-07 | 7.51E-07 | 6.945089 |
| NKX2-5 | -0.19667 | 0.872564 | 5.202747 | -5.68217 | 1.51E-07 | 7.60E-07 | 6.932786 |
| CAV3 | -0.1933 | 0.874603 | 6.405934 | -5.68189 | 1.51E-07 | 7.60E-07 | 6.931601 |
| PCDH1 | -0.19025 | 0.876456 | 5.224835 | -5.67855 | 1.53E-07 | 7.71E-07 | 6.917574 |
| DIMT1 | 0.186986 | 1.138383 | 7.086813 | 5.67839 | 1.53E-07 | 7.71E-07 | 6.916882 |
| HOPX | 0.657261 | 1.577085 | 5.911758 | 5.675398 | 1.55E-07 | 7.81E-07 | 6.904291 |
| ATIC | 0.217324 | 1.162575 | 8.817033 | 5.67448 | 1.56E-07 | 7.83E-07 | 6.900429 |
| HPRT1 | 0.298696 | 1.230032 | 8.419011 | 5.673876 | 1.57E-07 | 7.85E-07 | 6.897886 |
| KLK6 | -0.46502 | 0.724461 | 5.664176 | -5.6734 | 1.57E-07 | 7.86E-07 | 6.895898 |
| DSCR4 | -0.22615 | 0.854911 | 5.411209 | -5.67278 | 1.57E-07 | 7.88E-07 | 6.893292 |
| TMEM186 | 0.202715 | 1.150862 | 6.882418 | 5.67256 | 1.57E-07 | 7.88E-07 | 6.892348 |
| POP7 | 0.210053 | 1.156731 | 7.656264 | 5.671718 | 1.58E-07 | 7.90E-07 | 6.888808 |

| | | | | | | | |
|---|---|---|---|---|---|---|---|
| MPZL2 | 0.246203 | 1.186081 | 4.870879 | 5.671599 | 1.58E-07 | 7.90E-07 | 6.888306 |
| PDCD2 | 0.199353 | 1.148183 | 5.779451 | 5.671481 | 1.58E-07 | 7.90E-07 | 6.88781 |
| F3 | -0.62829 | 0.646945 | 6.116484 | -5.67109 | 1.58E-07 | 7.91E-07 | 6.886181 |
| C3orf36 | -0.17547 | 0.885477 | 4.706923 | -5.66979 | 1.59E-07 | 7.95E-07 | 6.88071 |
| LHFP | 0.531937 | 1.445869 | 6.82022 | 5.668769 | 1.60E-07 | 7.98E-07 | 6.876402 |
| PDCD1 | -0.22636 | 0.854787 | 5.741758 | -5.66857 | 1.60E-07 | 7.98E-07 | 6.875553 |
| CRYAB | 0.625338 | 1.542572 | 11.49945 | 5.663574 | 1.64E-07 | 8.15E-07 | 6.854559 |
| ZFYVE21 | 0.244174 | 1.184414 | 6.778571 | 5.662114 | 1.65E-07 | 8.20E-07 | 6.84842 |
| ERN2 | -0.22714 | 0.85433 | 5.856593 | -5.66156 | 1.65E-07 | 8.21E-07 | 6.846079 |
| KCNQ1DN | -0.19508 | 0.873526 | 5.535055 | -5.66125 | 1.65E-07 | 8.22E-07 | 6.844785 |
| GNMT | -0.24146 | 0.845887 | 4.894725 | -5.66101 | 1.65E-07 | 8.22E-07 | 6.843793 |
| HBB | 1.683222 | 3.211444 | 8.817363 | 5.658005 | 1.68E-07 | 8.32E-07 | 6.831154 |
| HRAS | 0.269995 | 1.205804 | 6.633297 | 5.657992 | 1.68E-07 | 8.32E-07 | 6.831097 |
| FEZ2 | 0.198527 | 1.147526 | 8.841868 | 5.657508 | 1.68E-07 | 8.34E-07 | 6.829065 |
| YBX2 | -0.19686 | 0.872445 | 5.393736 | -5.65707 | 1.68E-07 | 8.35E-07 | 6.827204 |
| FHL1 | 0.529928 | 1.443857 | 6.338791 | 5.654916 | 1.70E-07 | 8.42E-07 | 6.818173 |
| LEPROTL1 | 0.258899 | 1.196565 | 8.328462 | 5.65454 | 1.70E-07 | 8.43E-07 | 6.816597 |
| TACR2 | -0.2257 | 0.855183 | 5.362088 | -5.65385 | 1.71E-07 | 8.45E-07 | 6.813683 |
| PTS | 0.201667 | 1.150026 | 9.204725 | 5.653424 | 1.71E-07 | 8.46E-07 | 6.811908 |
| ITGA7 | -0.2184 | 0.859518 | 6.153956 | -5.65264 | 1.72E-07 | 8.48E-07 | 6.808629 |
| RFC2 | 0.219952 | 1.164695 | 5.906593 | 5.652011 | 1.72E-07 | 8.50E-07 | 6.805975 |
| CAV2 | 0.537019 | 1.450972 | 6.281429 | 5.651671 | 1.72E-07 | 8.51E-07 | 6.804545 |
| LOC10192 | -0.20831 | 0.865548 | 4.562857 | -5.6504 | 1.73E-07 | 8.55E-07 | 6.799204 |
| SPRY4 | -0.3205 | 0.800794 | 6.399121 | -5.65031 | 1.73E-07 | 8.55E-07 | 6.798843 |
| LCN2 | 1.011734 | 2.016334 | 5.828352 | 5.647363 | 1.76E-07 | 8.65E-07 | 6.786457 |
| ADM2 | -0.22968 | 0.852823 | 6.640769 | -5.64439 | 1.78E-07 | 8.76E-07 | 6.773977 |
| KPNB1 | 0.243826 | 1.184129 | 8.772747 | 5.643348 | 1.79E-07 | 8.80E-07 | 6.769604 |
| ARFGAP3 | 0.28013 | 1.214305 | 7.884396 | 5.642227 | 1.80E-07 | 8.83E-07 | 6.7649 |
| SMCO4 | 0.369285 | 1.291713 | 7.93044 | 5.64111 | 1.80E-07 | 8.87E-07 | 6.760211 |
| AP2B1 | 0.231357 | 1.173939 | 8.435495 | 5.640104 | 1.81E-07 | 8.90E-07 | 6.755991 |
| PSORS1C2 | -0.22204 | 0.857353 | 5.268462 | -5.6381 | 1.83E-07 | 8.98E-07 | 6.74758 |
| CCDC91 | 0.248763 | 1.188188 | 6.557363 | 5.637473 | 1.83E-07 | 9.00E-07 | 6.744953 |
| CHI3L1 | -0.80729 | 0.571452 | 5.762527 | -5.63675 | 1.84E-07 | 9.02E-07 | 6.741903 |
| NFKB1 | 0.249092 | 1.188459 | 7.592308 | 5.633375 | 1.87E-07 | 9.15E-07 | 6.727768 |
| LTBP2 | -0.27953 | 0.823861 | 6.503077 | -5.63288 | 1.87E-07 | 9.16E-07 | 6.72568 |
| SEMA6B | -0.18139 | 0.881852 | 5.507692 | -5.63252 | 1.87E-07 | 9.17E-07 | 6.724179 |
| OGFOD1 | 0.219647 | 1.164449 | 7.322747 | 5.63226 | 1.87E-07 | 9.17E-07 | 6.723092 |
| RUVBL1 | 0.201092 | 1.149568 | 7.708571 | 5.631972 | 1.88E-07 | 9.18E-07 | 6.721885 |
| MORN1 | -0.21389 | 0.862207 | 5.299011 | -5.63153 | 1.88E-07 | 9.19E-07 | 6.720036 |
| RTN3 | 0.253517 | 1.19211 | 9.493626 | 5.627895 | 1.91E-07 | 9.33E-07 | 6.704795 |
| PTPN3 | 0.223 | 1.167158 | 8.335275 | 5.627732 | 1.91E-07 | 9.33E-07 | 6.704111 |
| RAI2 | 0.340454 | 1.266155 | 6.088791 | 5.62559 | 1.93E-07 | 9.42E-07 | 6.695139 |
| POSTN | 1.146807 | 2.214233 | 6.133407 | 5.625345 | 1.93E-07 | 9.42E-07 | 6.694112 |
| SLC6A4 | -0.18974 | 0.876761 | 5.177692 | -5.62528 | 1.93E-07 | 9.42E-07 | 6.693829 |
| ITGB2 | 0.64099 | 1.559399 | 5.891538 | 5.624119 | 1.94E-07 | 9.46E-07 | 6.688974 |
| PGC | -0.18461 | 0.879888 | 4.951319 | -5.62342 | 1.95E-07 | 9.48E-07 | 6.686039 |
| LTK | -0.18751 | 0.878119 | 4.859231 | -5.62268 | 1.95E-07 | 9.51E-07 | 6.682945 |

| | | | | | | | |
|---|---|---|---|---|---|---|---|
| CUL1 | 0.211628 | 1.157994 | 7.117912 | 5.621911 | 1.96E-07 | 9.53E-07 | 6.679723 |
| PLOD3 | 0.21928 | 1.164153 | 8.606044 | 5.619335 | 1.98E-07 | 9.63E-07 | 6.668932 |
| ISOC2 | -0.31706 | 0.802705 | 8.905604 | -5.61867 | 1.99E-07 | 9.65E-07 | 6.666158 |
| C19orf73 | -0.16677 | 0.890833 | 4.812747 | -5.61866 | 1.99E-07 | 9.65E-07 | 6.666125 |
| KIAA0196 | 0.189372 | 1.140267 | 7.555385 | 5.617683 | 2.00E-07 | 9.69E-07 | 6.662014 |
| TWF2 | 0.276826 | 1.211527 | 6.694066 | 5.616886 | 2.00E-07 | 9.71E-07 | 6.658676 |
| HBQ1 | -0.2296 | 0.852869 | 5.104286 | -5.61679 | 2.00E-07 | 9.71E-07 | 6.658271 |
| PCOLCE2 | -0.96361 | 0.512771 | 5.620879 | -5.61665 | 2.00E-07 | 9.71E-07 | 6.657703 |
| TM9SF2 | 0.215213 | 1.160875 | 10.35099 | 5.611706 | 2.05E-07 | 9.92E-07 | 6.636993 |
| EMX1 | -0.26905 | 0.829867 | 6.24978 | -5.61136 | 2.05E-07 | 9.93E-07 | 6.635535 |
| DUSP22 | 0.228826 | 1.171881 | 7.70033 | 5.611109 | 2.05E-07 | 9.93E-07 | 6.634494 |
| CALM1 | -0.23306 | 0.85083 | 10.21714 | -5.6081 | 2.08E-07 | 1.00E-06 | 6.621889 |
| MAN2B1 | 0.338488 | 1.264431 | 7.089341 | 5.608019 | 2.08E-07 | 1.00E-06 | 6.621567 |
| ZNF574 | -0.19348 | 0.874492 | 6.727582 | -5.60799 | 2.08E-07 | 1.00E-06 | 6.62143 |
| MPPE1 | 0.213164 | 1.159228 | 8.517802 | 5.607342 | 2.09E-07 | 1.01E-06 | 6.618734 |
| WT1 | -0.4294 | 0.74257 | 5.674615 | -5.60553 | 2.10E-07 | 1.01E-06 | 6.611164 |
| ANKH | -0.16123 | 0.894261 | 4.614835 | -5.60389 | 2.12E-07 | 1.02E-06 | 6.604276 |
| PBLD | -0.36455 | 0.776713 | 10.95516 | -5.60287 | 2.13E-07 | 1.03E-06 | 6.600041 |
| HSPA4 | 0.227889 | 1.17112 | 7.447692 | 5.602402 | 2.13E-07 | 1.03E-06 | 6.59807 |
| PPL | 0.385541 | 1.30635 | 6.6 | 5.601449 | 2.14E-07 | 1.03E-06 | 6.594087 |
| ETV7 | -0.21951 | 0.858856 | 5.633407 | -5.59952 | 2.16E-07 | 1.04E-06 | 6.586037 |
| VSIG10 | 0.208585 | 1.155554 | 5.189451 | 5.597959 | 2.17E-07 | 1.04E-06 | 6.579495 |
| RRBP1 | 0.219386 | 1.164238 | 7.370879 | 5.597747 | 2.18E-07 | 1.04E-06 | 6.578608 |
| ATP4A | -0.19711 | 0.872296 | 4.492527 | -5.59675 | 2.18E-07 | 1.05E-06 | 6.57446 |
| PDIA3 | 0.365232 | 1.288089 | 9.286044 | 5.596254 | 2.19E-07 | 1.05E-06 | 6.572366 |
| ZNF839 | -0.15202 | 0.899987 | 5.653736 | -5.59362 | 2.21E-07 | 1.06E-06 | 6.561367 |
| LTBP4 | -0.19472 | 0.873743 | 4.931319 | -5.59301 | 2.22E-07 | 1.06E-06 | 6.558823 |
| REG1A | 1.471242 | 2.772604 | 5.733407 | 5.592496 | 2.23E-07 | 1.07E-06 | 6.556664 |
| ITGA10 | -0.21901 | 0.859155 | 5.636264 | -5.59195 | 2.23E-07 | 1.07E-06 | 6.554378 |
| JUND | -0.37485 | 0.771188 | 6.086593 | -5.59139 | 2.24E-07 | 1.07E-06 | 6.552033 |
| MMP17 | -0.20268 | 0.868937 | 5.339341 | -5.58983 | 2.25E-07 | 1.08E-06 | 6.545508 |
| A2M | 0.550947 | 1.465047 | 9.795055 | 5.589236 | 2.26E-07 | 1.08E-06 | 6.543045 |
| UPK2 | -0.18845 | 0.877546 | 4.466374 | -5.58891 | 2.26E-07 | 1.08E-06 | 6.541686 |
| KLK14 | -0.19715 | 0.872272 | 5.690769 | -5.58853 | 2.26E-07 | 1.08E-06 | 6.540079 |
| ZNF195 | 0.22772 | 1.170983 | 5.76 | 5.588106 | 2.27E-07 | 1.08E-06 | 6.538325 |
| PTGER3 | -0.30606 | 0.808849 | 6.976044 | -5.58757 | 2.27E-07 | 1.08E-06 | 6.536092 |
| KHDRBS2 | -0.22842 | 0.853569 | 4.918132 | -5.58698 | 2.28E-07 | 1.09E-06 | 6.533618 |
| CLN3 | 0.210232 | 1.156874 | 7.124396 | 5.585068 | 2.30E-07 | 1.09E-06 | 6.525639 |
| PIGT | 0.165541 | 1.121587 | 8.411209 | 5.584545 | 2.30E-07 | 1.10E-06 | 6.523455 |
| CFAP74 | -0.16519 | 0.891809 | 5.144615 | -5.58134 | 2.33E-07 | 1.11E-06 | 6.510085 |
| PKLR | -0.37541 | 0.770889 | 6.591099 | -5.58007 | 2.35E-07 | 1.12E-06 | 6.504788 |
| KCNA5 | -0.25149 | 0.84003 | 4.948681 | -5.57803 | 2.37E-07 | 1.13E-06 | 6.496243 |
| NUDT15 | 0.221188 | 1.165693 | 7.912857 | 5.573567 | 2.41E-07 | 1.15E-06 | 6.477641 |
| TNPO1 | 0.223754 | 1.167768 | 8.28956 | 5.573002 | 2.42E-07 | 1.15E-06 | 6.475283 |
| SLC2A5 | -0.5858 | 0.666281 | 8.259451 | -5.57177 | 2.43E-07 | 1.15E-06 | 6.470126 |
| ERAP1 | 0.212585 | 1.158762 | 5.171429 | 5.567275 | 2.48E-07 | 1.18E-06 | 6.451401 |
| P3H3 | -0.17457 | 0.886029 | 4.837802 | -5.56696 | 2.48E-07 | 1.18E-06 | 6.4501 |

| | | | | | | | |
|---|---|---|---|---|---|---|---|
| IFIT3 | 0.566435 | 1.48086 | 7.29967 | 5.5657 | 2.50E-07 | 1.18E-06 | 6.444838 |
| INPP5F | 0.267527 | 1.203742 | 6.390989 | 5.565593 | 2.50E-07 | 1.18E-06 | 6.444392 |
| PDZD3 | -0.29629 | 0.814344 | 7.20044 | -5.56555 | 2.50E-07 | 1.18E-06 | 6.444206 |
| APOA1 | -0.24912 | 0.841412 | 5.126593 | -5.56547 | 2.50E-07 | 1.18E-06 | 6.443879 |
| BCL7A | -0.19818 | 0.871647 | 6.303736 | -5.56493 | 2.51E-07 | 1.18E-06 | 6.441619 |
| CASK | 0.219406 | 1.164254 | 6.601758 | 5.563487 | 2.52E-07 | 1.19E-06 | 6.435616 |
| CER1 | -0.22299 | 0.856791 | 4.985385 | -5.56342 | 2.52E-07 | 1.19E-06 | 6.435324 |
| CDK4 | 0.170227 | 1.125236 | 8.894396 | 5.561208 | 2.55E-07 | 1.20E-06 | 6.426118 |
| KCNH6 | -0.31988 | 0.801134 | 5.077033 | -5.56082 | 2.55E-07 | 1.20E-06 | 6.424516 |
| TAS2R1 | -0.237 | 0.848505 | 5.177582 | -5.56071 | 2.55E-07 | 1.20E-06 | 6.424038 |
| SNRPF | 0.236435 | 1.178078 | 8.546484 | 5.560242 | 2.56E-07 | 1.20E-06 | 6.422092 |
| AURKA | 0.307976 | 1.23797 | 5.141209 | 5.55998 | 2.56E-07 | 1.20E-06 | 6.421 |
| FIBP | 0.190541 | 1.141192 | 8.598571 | 5.559923 | 2.56E-07 | 1.20E-06 | 6.420763 |
| RAP1GDS1 | 0.216072 | 1.161567 | 7.09011 | 5.559842 | 2.56E-07 | 1.20E-06 | 6.420428 |
| FAM110B | 0.312937 | 1.242234 | 5.976923 | 5.558614 | 2.57E-07 | 1.21E-06 | 6.415311 |
| ABCE1 | 0.212546 | 1.158731 | 6.73967 | 5.555173 | 2.61E-07 | 1.23E-06 | 6.400979 |
| CPLX3 | -0.17778 | 0.884061 | 4.861868 | -5.55155 | 2.65E-07 | 1.25E-06 | 6.3859 |
| HK1 | 0.354773 | 1.278784 | 7.83022 | 5.54982 | 2.67E-07 | 1.25E-06 | 6.378692 |
| SHOX | -0.21566 | 0.861151 | 5.468571 | -5.54958 | 2.68E-07 | 1.26E-06 | 6.377699 |
| ECM2 | -0.50255 | 0.705858 | 7.446703 | -5.54726 | 2.70E-07 | 1.27E-06 | 6.368057 |
| CD44 | 0.416444 | 1.334634 | 5.775934 | 5.54634 | 2.71E-07 | 1.27E-06 | 6.36421 |
| SYNCRIP | 0.204174 | 1.152027 | 7.688791 | 5.545271 | 2.73E-07 | 1.28E-06 | 6.359759 |
| PRPH | -0.23954 | 0.847015 | 6.362198 | -5.54343 | 2.75E-07 | 1.29E-06 | 6.352089 |
| FABP6 | -0.2175 | 0.860056 | 4.475055 | -5.54299 | 2.75E-07 | 1.29E-06 | 6.350251 |
| HTR5A | -0.21722 | 0.860223 | 4.397802 | -5.54089 | 2.78E-07 | 1.30E-06 | 6.341548 |
| EFNA3 | -0.18791 | 0.877875 | 5.900989 | -5.5397 | 2.79E-07 | 1.30E-06 | 6.336581 |
| RPH3A | -0.18384 | 0.880356 | 4.936264 | -5.53933 | 2.80E-07 | 1.31E-06 | 6.335042 |
| CENPM | 0.375372 | 1.297174 | 5.077363 | 5.53871 | 2.80E-07 | 1.31E-06 | 6.33247 |
| POP5 | 0.230425 | 1.173181 | 8.953077 | 5.537455 | 2.82E-07 | 1.31E-06 | 6.327251 |
| SMIM10L1 | 0.241836 | 1.182496 | 7.652198 | 5.534864 | 2.85E-07 | 1.33E-06 | 6.31648 |
| SHFM1 | 0.191169 | 1.141689 | 7.057143 | 5.531457 | 2.89E-07 | 1.35E-06 | 6.302319 |
| SSTR4 | -0.18676 | 0.878578 | 4.863516 | -5.53032 | 2.91E-07 | 1.35E-06 | 6.297584 |
| TBC1D2B | 0.384184 | 1.305121 | 7.058242 | 5.528925 | 2.92E-07 | 1.36E-06 | 6.291799 |
| PLA2R1 | -0.34526 | 0.787168 | 4.172747 | -5.52857 | 2.93E-07 | 1.36E-06 | 6.290321 |
| PLG | -0.77788 | 0.583222 | 10.49055 | -5.52851 | 2.93E-07 | 1.36E-06 | 6.290085 |
| LSM14B | -0.19052 | 0.876292 | 5.392527 | -5.5282 | 2.93E-07 | 1.36E-06 | 6.288773 |
| ARL6IP1 | 0.209676 | 1.156429 | 9.929121 | 5.52596 | 2.96E-07 | 1.38E-06 | 6.279481 |
| QTRTD1 | 0.226986 | 1.170387 | 5.606593 | 5.524256 | 2.98E-07 | 1.38E-06 | 6.272406 |
| ART1 | -0.19849 | 0.871463 | 5.16989 | -5.52273 | 3.00E-07 | 1.39E-06 | 6.266063 |
| GNB3 | -0.2068 | 0.866459 | 6.261868 | -5.5221 | 3.01E-07 | 1.40E-06 | 6.263439 |
| PLCB4 | 0.492121 | 1.406511 | 4.578791 | 5.520774 | 3.03E-07 | 1.40E-06 | 6.257949 |
| TTC26 | 0.209807 | 1.156533 | 5.635055 | 5.520268 | 3.04E-07 | 1.41E-06 | 6.255848 |
| LAMTOR3 | 0.259488 | 1.197054 | 8.455275 | 5.519489 | 3.05E-07 | 1.41E-06 | 6.252615 |
| BICC1 | 0.300647 | 1.231697 | 7.907802 | 5.519055 | 3.05E-07 | 1.41E-06 | 6.250813 |
| FRMD1 | -0.19508 | 0.873526 | 5.675055 | -5.51795 | 3.07E-07 | 1.42E-06 | 6.24623 |
| DNASE1L1 | -0.2063 | 0.866755 | 7.654286 | -5.51741 | 3.07E-07 | 1.42E-06 | 6.243997 |
| LSR | 0.322435 | 1.250439 | 6.319011 | 5.516275 | 3.09E-07 | 1.43E-06 | 6.239272 |

| | | | | | | | |
|---|---|---|---|---|---|---|---|
| ADCY1 | -0.15806 | 0.896231 | 4.929231 | -5.51622 | 3.09E-07 | 1.43E-06 | 6.239028 |
| MYBPH | -0.21727 | 0.860194 | 5.615604 | -5.50907 | 3.18E-07 | 1.47E-06 | 6.209384 |
| PLA2G2D | -0.21229 | 0.863166 | 5.981978 | -5.50743 | 3.21E-07 | 1.48E-06 | 6.202567 |
| SKIV2L2 | 0.189696 | 1.140523 | 7.87011 | 5.505588 | 3.23E-07 | 1.49E-06 | 6.194946 |
| PLAC4 | -0.18718 | 0.878319 | 5.474176 | -5.50554 | 3.23E-07 | 1.49E-06 | 6.19473 |
| CAAP1 | 0.224565 | 1.168425 | 7.686484 | 5.505433 | 3.23E-07 | 1.49E-06 | 6.194304 |
| ARHGAP12 | 0.240005 | 1.180997 | 6.758901 | 5.505358 | 3.24E-07 | 1.49E-06 | 6.193992 |
| UTP14C | 0.223527 | 1.167584 | 8.199231 | 5.505353 | 3.24E-07 | 1.49E-06 | 6.193973 |
| TCEAL4 | 0.205855 | 1.15337 | 9.495275 | 5.505349 | 3.24E-07 | 1.49E-06 | 6.193958 |
| JADE3 | 0.24058 | 1.181467 | 5.255055 | 5.503739 | 3.26E-07 | 1.50E-06 | 6.187281 |
| CD177 | -0.23419 | 0.85016 | 5.805495 | -5.50238 | 3.28E-07 | 1.50E-06 | 6.181644 |
| HEYL | -0.18774 | 0.877978 | 5.908681 | -5.50179 | 3.29E-07 | 1.51E-06 | 6.179196 |
| SH3BP1 | -0.25299 | 0.839158 | 5.940549 | -5.50122 | 3.29E-07 | 1.51E-06 | 6.176829 |
| ADAM5 | -0.20922 | 0.865003 | 4.601538 | -5.49979 | 3.31E-07 | 1.52E-06 | 6.170903 |
| CNTD2 | -0.25031 | 0.840713 | 6.752088 | -5.4986 | 3.33E-07 | 1.53E-06 | 6.165989 |
| LDB1 | -0.17222 | 0.887475 | 6.744835 | -5.49739 | 3.35E-07 | 1.53E-06 | 6.16096 |
| GJD2 | -0.20291 | 0.868797 | 5.648791 | -5.4955 | 3.38E-07 | 1.54E-06 | 6.153142 |
| DKFZP434A | -0.18406 | 0.880221 | 6.076154 | -5.49314 | 3.41E-07 | 1.56E-06 | 6.143368 |
| BAZ2B | 0.23687 | 1.178433 | 6.706264 | 5.49166 | 3.43E-07 | 1.57E-06 | 6.137239 |
| MFHAS1 | 0.330778 | 1.257691 | 6.938571 | 5.491481 | 3.43E-07 | 1.57E-06 | 6.136498 |
| ITGAE | 0.23244 | 1.17482 | 8.614725 | 5.49113 | 3.44E-07 | 1.57E-06 | 6.135047 |
| MS4A4A | 0.692275 | 1.61583 | 5.174725 | 5.490736 | 3.44E-07 | 1.57E-06 | 6.133415 |
| DMBT1 | -0.20465 | 0.867751 | 6.11967 | -5.48798 | 3.49E-07 | 1.59E-06 | 6.122006 |
| E2F4 | -0.19714 | 0.872281 | 8.211429 | -5.48771 | 3.49E-07 | 1.59E-06 | 6.12089 |
| PADI3 | -0.18858 | 0.877466 | 5.17044 | -5.48551 | 3.52E-07 | 1.60E-06 | 6.111801 |
| PLEKHA1 | 0.400155 | 1.319649 | 8.494835 | 5.485483 | 3.52E-07 | 1.60E-06 | 6.111674 |
| SEMA6C | -0.22371 | 0.856357 | 6.482198 | -5.48322 | 3.56E-07 | 1.62E-06 | 6.102323 |
| KRT31 | -0.14333 | 0.905428 | 4.199341 | -5.48318 | 3.56E-07 | 1.62E-06 | 6.102163 |
| PLCE1 | -0.3583 | 0.780081 | 5.129121 | -5.48271 | 3.56E-07 | 1.62E-06 | 6.100202 |
| FZD1 | 0.331932 | 1.258698 | 6.241099 | 5.480804 | 3.59E-07 | 1.63E-06 | 6.092315 |
| GSN-AS1 | -0.18043 | 0.88244 | 4.401429 | -5.48035 | 3.60E-07 | 1.63E-06 | 6.090437 |
| GABARAPL | -0.18498 | 0.879661 | 4.594396 | -5.47994 | 3.61E-07 | 1.64E-06 | 6.088745 |
| CKS2 | 0.644314 | 1.562996 | 8.802747 | 5.47825 | 3.63E-07 | 1.65E-06 | 6.081751 |
| LIN7B | -0.2534 | 0.838916 | 6.061648 | -5.47748 | 3.65E-07 | 1.65E-06 | 6.078561 |
| PKP1 | -0.17486 | 0.885851 | 4.794615 | -5.47496 | 3.68E-07 | 1.67E-06 | 6.068138 |
| SP140L | 0.332729 | 1.259394 | 5.307363 | 5.474764 | 3.69E-07 | 1.67E-06 | 6.067336 |
| ST3GAL4 | -0.3224 | 0.799738 | 5.812527 | -5.47457 | 3.69E-07 | 1.67E-06 | 6.066529 |
| CCNL1 | -0.3361 | 0.792182 | 7.509451 | -5.47427 | 3.70E-07 | 1.67E-06 | 6.065297 |
| SH3BP5 | -0.33992 | 0.790086 | 8.345604 | -5.47028 | 3.76E-07 | 1.70E-06 | 6.048816 |
| SARDH | -0.25162 | 0.839951 | 5.402527 | -5.46983 | 3.77E-07 | 1.70E-06 | 6.046961 |
| BPIFA1 | -0.22668 | 0.854599 | 5.257253 | -5.46598 | 3.83E-07 | 1.73E-06 | 6.031035 |
| CTSO | 0.276623 | 1.211356 | 7.854835 | 5.46335 | 3.87E-07 | 1.75E-06 | 6.020176 |
| ACSF2 | -0.36983 | 0.773873 | 9.624725 | -5.46003 | 3.93E-07 | 1.77E-06 | 6.006458 |
| SAMSN1 | 0.568019 | 1.482487 | 4.291758 | 5.458739 | 3.95E-07 | 1.78E-06 | 6.001139 |
| HACD1 | 0.374386 | 1.296288 | 5.152527 | 5.458245 | 3.96E-07 | 1.78E-06 | 5.999101 |
| SLC16A5 | 0.365155 | 1.28802 | 5.862527 | 5.458077 | 3.96E-07 | 1.78E-06 | 5.998406 |
| AGR2 | 0.649198 | 1.568296 | 5.414945 | 5.456403 | 3.99E-07 | 1.80E-06 | 5.991497 |

| | | | | | | | |
|---|---|---|---|---|---|---|---|
| GNG3 | -0.1893 | 0.877031 | 5.537912 | -5.45518 | 4.01E-07 | 1.80E-06 | 5.986453 |
| ZNF747 | -0.20533 | 0.867341 | 7.138681 | -5.45448 | 4.02E-07 | 1.81E-06 | 5.983582 |
| IRF7 | 0.450836 | 1.366832 | 5.900549 | 5.450676 | 4.09E-07 | 1.84E-06 | 5.967866 |
| XYLT2 | 0.201367 | 1.149787 | 6.061099 | 5.45009 | 4.10E-07 | 1.84E-06 | 5.96545 |
| SLC25A36 | -0.25927 | 0.835513 | 8.434835 | -5.44964 | 4.10E-07 | 1.84E-06 | 5.963582 |
| RAB4A | 0.171783 | 1.126449 | 7.585165 | 5.44935 | 4.11E-07 | 1.84E-06 | 5.962399 |
| LOC10050( | -0.20218 | 0.869234 | 5.651758 | -5.44913 | 4.11E-07 | 1.84E-06 | 5.961496 |
| MIP | -0.16545 | 0.891651 | 4.252967 | -5.44901 | 4.12E-07 | 1.84E-06 | 5.961007 |
| UBE2Z | 0.160232 | 1.117467 | 8.38967 | 5.448521 | 4.12E-07 | 1.85E-06 | 5.95898 |
| SNCG | -0.23739 | 0.848281 | 6.60022 | -5.44696 | 4.15E-07 | 1.86E-06 | 5.952545 |
| CCDC92 | 0.190527 | 1.14118 | 8.097912 | 5.440882 | 4.26E-07 | 1.91E-06 | 5.927487 |
| GALNT8 | -0.21423 | 0.862005 | 4.443626 | -5.44053 | 4.27E-07 | 1.91E-06 | 5.926056 |
| GIMAP6 | 0.435213 | 1.35211 | 5.24978 | 5.439556 | 4.29E-07 | 1.92E-06 | 5.922024 |
| FSTL4 | -0.17803 | 0.88391 | 4.590659 | -5.43922 | 4.29E-07 | 1.92E-06 | 5.920624 |
| SCNN1B | -0.49986 | 0.707175 | 6.379121 | -5.43791 | 4.32E-07 | 1.93E-06 | 5.915261 |
| PPP1R2P9 | -0.18004 | 0.882676 | 4.419011 | -5.43715 | 4.33E-07 | 1.93E-06 | 5.912095 |
| ALDH1A3 | 0.418826 | 1.336839 | 5.974286 | 5.433813 | 4.39E-07 | 1.96E-06 | 5.898367 |
| PRDM11 | -0.21137 | 0.863715 | 5.093736 | -5.43318 | 4.40E-07 | 1.96E-06 | 5.89576 |
| GAST | -0.20692 | 0.866386 | 5.076374 | -5.43131 | 4.44E-07 | 1.98E-06 | 5.888062 |
| LINC00094 | 0.230957 | 1.173613 | 6.637253 | 5.430288 | 4.46E-07 | 1.98E-06 | 5.883854 |
| SYT11 | 0.201768 | 1.150107 | 4.329341 | 5.430253 | 4.46E-07 | 1.98E-06 | 5.88371 |
| CHRND | -0.18418 | 0.880147 | 4.350659 | -5.42962 | 4.47E-07 | 1.99E-06 | 5.881118 |
| GDPD5 | -0.16423 | 0.892404 | 5.778352 | -5.42633 | 4.53E-07 | 2.02E-06 | 5.867547 |
| YIF1A | 0.245367 | 1.185394 | 7.852857 | 5.426298 | 4.53E-07 | 2.02E-06 | 5.867428 |
| NECAB2 | -0.53428 | 0.690503 | 7.553187 | -5.42604 | 4.54E-07 | 2.02E-06 | 5.866384 |
| BMP7 | -0.17599 | 0.885163 | 5.193626 | -5.42527 | 4.55E-07 | 2.02E-06 | 5.863195 |
| USP14 | 0.18385 | 1.135911 | 8.024176 | 5.425091 | 4.56E-07 | 2.02E-06 | 5.86246 |
| SUB1 | 0.160266 | 1.117493 | 9.561209 | 5.424822 | 4.56E-07 | 2.02E-06 | 5.861354 |
| SELENBP1 | -0.466 | 0.723969 | 9.01956 | -5.42433 | 4.57E-07 | 2.03E-06 | 5.859311 |
| MOXD1 | 0.584913 | 1.499949 | 6.12033 | 5.422844 | 4.60E-07 | 2.04E-06 | 5.853218 |
| NGB | -0.19833 | 0.87156 | 4.687143 | -5.42267 | 4.60E-07 | 2.04E-06 | 5.852509 |
| GDPD2 | -0.25428 | 0.838408 | 6.321868 | -5.4225 | 4.61E-07 | 2.04E-06 | 5.851822 |
| GTF2IRD1 | 0.225063 | 1.168828 | 8.039121 | 5.421792 | 4.62E-07 | 2.04E-06 | 5.848888 |
| PDHX | 0.253411 | 1.192022 | 7.818791 | 5.421443 | 4.63E-07 | 2.05E-06 | 5.847452 |
| RHCG | -0.8111 | 0.569947 | 8.249341 | -5.42032 | 4.65E-07 | 2.06E-06 | 5.842833 |
| GCK | -0.19925 | 0.871005 | 5.175385 | -5.41888 | 4.68E-07 | 2.07E-06 | 5.836927 |
| ADM | -0.7923 | 0.577421 | 9.254505 | -5.41263 | 4.80E-07 | 2.12E-06 | 5.811199 |
| CHN2 | -0.30194 | 0.811162 | 6.273516 | -5.41121 | 4.83E-07 | 2.13E-06 | 5.805389 |
| SET | 0.222435 | 1.166701 | 10.38956 | 5.409377 | 4.87E-07 | 2.15E-06 | 5.797847 |
| IQSEC3 | -0.21071 | 0.864115 | 5.474066 | -5.40857 | 4.89E-07 | 2.15E-06 | 5.79455 |
| CLTB | -0.23409 | 0.850223 | 7.81033 | -5.40813 | 4.90E-07 | 2.16E-06 | 5.792746 |
| DRD5 | -0.27364 | 0.827228 | 5.090769 | -5.40783 | 4.90E-07 | 2.16E-06 | 5.791473 |
| RHBG | -0.35684 | 0.780875 | 6.775934 | -5.40781 | 4.90E-07 | 2.16E-06 | 5.791413 |
| C17orf53 | -0.20289 | 0.868806 | 5.329451 | -5.40781 | 4.90E-07 | 2.16E-06 | 5.791395 |
| NMD3 | 0.32971 | 1.256761 | 7.84 | 5.407796 | 4.90E-07 | 2.16E-06 | 5.791352 |
| YIPF5 | 0.217256 | 1.16252 | 7.403956 | 5.405178 | 4.96E-07 | 2.18E-06 | 5.780599 |
| CLDN11 | -0.27823 | 0.824604 | 5.272198 | -5.40407 | 4.98E-07 | 2.19E-06 | 5.776033 |

| | | | | | | | |
|---|---|---|---|---|---|---|---|
| EPN1 | -0.23229 | 0.851286 | 6.352308 | -5.40403 | 4.98E-07 | 2.19E-06 | 5.775902 |
| EXOSC8 | 0.231705 | 1.174222 | 6.271319 | 5.403018 | 5.01E-07 | 2.20E-06 | 5.77173 |
| RPGRIP1 | -0.16562 | 0.891546 | 5.095275 | -5.40109 | 5.05E-07 | 2.21E-06 | 5.7638 |
| PRELP | -0.19861 | 0.871391 | 6.484396 | -5.40046 | 5.06E-07 | 2.22E-06 | 5.76121 |
| PRRX2 | -0.22918 | 0.853118 | 6.363407 | -5.39947 | 5.08E-07 | 2.22E-06 | 5.75715 |
| SYNPO | -0.32385 | 0.798935 | 6.396593 | -5.39945 | 5.08E-07 | 2.22E-06 | 5.757086 |
| SOX15 | -0.18099 | 0.882097 | 5.135934 | -5.39696 | 5.14E-07 | 2.25E-06 | 5.746869 |
| BTBD1 | 0.267957 | 1.204101 | 8.940549 | 5.396919 | 5.14E-07 | 2.25E-06 | 5.746694 |
| COX6A2 | -0.35325 | 0.782818 | 5.939011 | -5.39501 | 5.18E-07 | 2.26E-06 | 5.738844 |
| IFIT2 | 0.460478 | 1.375998 | 6.389231 | 5.394505 | 5.19E-07 | 2.27E-06 | 5.736789 |
| AIMP2 | 0.202884 | 1.150997 | 9.40011 | 5.391923 | 5.25E-07 | 2.29E-06 | 5.726197 |
| MAFF | -1.296 | 0.407254 | 6.799121 | -5.39065 | 5.27E-07 | 2.30E-06 | 5.720955 |
| TUBA1B | 0.443121 | 1.359542 | 11.12956 | 5.388937 | 5.31E-07 | 2.32E-06 | 5.71395 |
| C7orf43 | -0.17225 | 0.88746 | 6.183736 | -5.38832 | 5.33E-07 | 2.32E-06 | 5.711404 |
| NCR3 | -0.1206 | 0.919803 | 4.693187 | -5.38827 | 5.33E-07 | 2.32E-06 | 5.711201 |
| DNAH6 | -0.15663 | 0.897119 | 4.774286 | -5.38759 | 5.34E-07 | 2.33E-06 | 5.708435 |
| MRPL3 | 0.235522 | 1.177332 | 9.794945 | 5.386978 | 5.36E-07 | 2.33E-06 | 5.705918 |
| SOX4 | 0.622242 | 1.539265 | 7.428571 | 5.385791 | 5.38E-07 | 2.34E-06 | 5.701051 |
| HTR1E | -0.18894 | 0.877249 | 4.454176 | -5.38467 | 5.41E-07 | 2.35E-06 | 5.696474 |
| CMAS | 0.229995 | 1.172831 | 6.083516 | 5.38293 | 5.45E-07 | 2.37E-06 | 5.689324 |
| MARS | 0.173348 | 1.127672 | 7.326374 | 5.380805 | 5.50E-07 | 2.39E-06 | 5.680614 |
| IK | 0.176899 | 1.130451 | 9.037912 | 5.379305 | 5.53E-07 | 2.40E-06 | 5.674469 |
| NUBP1 | 0.183763 | 1.135843 | 7.60022 | 5.37926 | 5.54E-07 | 2.40E-06 | 5.674287 |
| RALYL | -0.58285 | 0.667643 | 7.473516 | -5.37779 | 5.57E-07 | 2.42E-06 | 5.668247 |
| NTSR2 | -0.21772 | 0.859921 | 4.464725 | -5.37489 | 5.64E-07 | 2.44E-06 | 5.656369 |
| P2RX4 | 0.251353 | 1.190323 | 8.205165 | 5.372118 | 5.71E-07 | 2.47E-06 | 5.645036 |
| MGC2889 | -0.18703 | 0.87841 | 4.490989 | -5.37192 | 5.71E-07 | 2.47E-06 | 5.644216 |
| CLCA1 | -0.19693 | 0.872407 | 4.860879 | -5.37124 | 5.73E-07 | 2.48E-06 | 5.641461 |
| CSNK1D | 0.210126 | 1.156789 | 7.39956 | 5.370257 | 5.75E-07 | 2.49E-06 | 5.637416 |
| NAT6 | -0.21258 | 0.86299 | 5.788571 | -5.3684 | 5.80E-07 | 2.51E-06 | 5.629797 |
| NUP153 | 0.263575 | 1.20045 | 7.431209 | 5.36657 | 5.84E-07 | 2.52E-06 | 5.622326 |
| SOCS7 | -0.16959 | 0.889096 | 5.094615 | -5.36626 | 5.85E-07 | 2.53E-06 | 5.62107 |
| TMED5 | 0.225647 | 1.169302 | 8.885714 | 5.365013 | 5.88E-07 | 2.54E-06 | 5.615959 |
| TNFRSF13I | -0.2209 | 0.858028 | 5.65011 | -5.36319 | 5.93E-07 | 2.56E-06 | 5.608508 |
| CLASRP | -0.21401 | 0.862138 | 8.083736 | -5.36245 | 5.94E-07 | 2.56E-06 | 5.605453 |
| EVX1 | -0.27662 | 0.825524 | 5.045385 | -5.36157 | 5.97E-07 | 2.57E-06 | 5.601863 |
| CSK | 0.192106 | 1.14243 | 7.01978 | 5.36066 | 5.99E-07 | 2.58E-06 | 5.598154 |
| DLEC1 | -0.15946 | 0.895358 | 5.425275 | -5.36023 | 6.00E-07 | 2.58E-06 | 5.596406 |
| UCP2 | 0.449614 | 1.365674 | 6.854945 | 5.360232 | 6.00E-07 | 2.58E-06 | 5.596402 |
| GOLPH3L | 0.233609 | 1.175772 | 8.727912 | 5.360156 | 6.00E-07 | 2.58E-06 | 5.596092 |
| CLTCL1 | -0.21043 | 0.864277 | 5.906374 | -5.35972 | 6.01E-07 | 2.59E-06 | 5.594309 |
| SLC34A2 | 0.512638 | 1.426656 | 5.442198 | 5.358459 | 6.04E-07 | 2.60E-06 | 5.589154 |
| NEUROG1 | -0.16844 | 0.889802 | 5.036703 | -5.3575 | 6.07E-07 | 2.61E-06 | 5.585228 |
| SPG11 | 0.196942 | 1.146266 | 7.20978 | 5.355744 | 6.11E-07 | 2.63E-06 | 5.578054 |
| LOC10050 | -0.20486 | 0.867626 | 4.38022 | -5.35509 | 6.13E-07 | 2.63E-06 | 5.575375 |
| DTL | 0.300725 | 1.231763 | 3.871319 | 5.352862 | 6.19E-07 | 2.65E-06 | 5.566275 |
| CATSPERG | -0.16258 | 0.893426 | 4.253516 | -5.35233 | 6.20E-07 | 2.66E-06 | 5.564102 |

| | | | | | | | |
|---|---|---|---|---|---|---|---|
| RSPH6A | -0.21315 | 0.862652 | 4.862857 | -5.35226 | 6.21E-07 | 2.66E-06 | 5.56382 |
| CELA3B | -0.17638 | 0.884923 | 6.535824 | -5.35212 | 6.21E-07 | 2.66E-06 | 5.563253 |
| ALOX12P2 | -0.20874 | 0.86529 | 5.293297 | -5.35097 | 6.24E-07 | 2.67E-06 | 5.558553 |
| EFNA4 | 0.263821 | 1.200655 | 5.502418 | 5.349373 | 6.28E-07 | 2.69E-06 | 5.55202 |
| GTF2A2 | 0.218121 | 1.163217 | 8.113297 | 5.348783 | 6.30E-07 | 2.69E-06 | 5.54961 |
| HIST1H1C | 0.530367 | 1.444297 | 8.148791 | 5.348545 | 6.30E-07 | 2.69E-06 | 5.548636 |
| RPA2 | 0.289024 | 1.221814 | 7.789011 | 5.345632 | 6.38E-07 | 2.73E-06 | 5.536742 |
| ZNF267 | 0.313082 | 1.242359 | 5.133516 | 5.343564 | 6.44E-07 | 2.75E-06 | 5.528299 |
| ALX3 | -0.20513 | 0.867463 | 4.517912 | -5.34274 | 6.46E-07 | 2.76E-06 | 5.524946 |
| PIK3C2G | -0.25021 | 0.840775 | 5.196923 | -5.34228 | 6.47E-07 | 2.76E-06 | 5.523054 |
| CUL9 | -0.14904 | 0.901851 | 5.87956 | -5.34183 | 6.48E-07 | 2.76E-06 | 5.521227 |
| LHB | -0.24231 | 0.845391 | 5.616264 | -5.33961 | 6.55E-07 | 2.79E-06 | 5.512152 |
| FAM49B | 0.17799 | 1.131307 | 7.057582 | 5.338538 | 6.58E-07 | 2.80E-06 | 5.507781 |
| LTB | 0.56143 | 1.475731 | 6.091978 | 5.338237 | 6.58E-07 | 2.80E-06 | 5.506557 |
| TUB | -0.17448 | 0.886085 | 6.181978 | -5.33756 | 6.60E-07 | 2.81E-06 | 5.503812 |
| TRIM21 | 0.209507 | 1.156293 | 6.561429 | 5.336364 | 6.64E-07 | 2.82E-06 | 5.498915 |
| TOB1 | -0.35737 | 0.780585 | 9.891538 | -5.33591 | 6.65E-07 | 2.83E-06 | 5.497051 |
| LRRC42 | 0.174111 | 1.128269 | 6.431099 | 5.335859 | 6.65E-07 | 2.83E-06 | 5.496853 |
| CHRM2 | -0.23662 | 0.84873 | 4.884945 | -5.33403 | 6.70E-07 | 2.85E-06 | 5.489375 |
| AP2S1 | 0.216005 | 1.161513 | 9.117033 | 5.332891 | 6.73E-07 | 2.86E-06 | 5.484746 |
| EIF3D | 0.19701 | 1.14632 | 9.432857 | 5.332286 | 6.75E-07 | 2.86E-06 | 5.482281 |
| LHX3 | -0.18934 | 0.877005 | 5.325934 | -5.33179 | 6.77E-07 | 2.87E-06 | 5.480243 |
| IL36A | -0.20644 | 0.866673 | 4.998132 | -5.33128 | 6.78E-07 | 2.87E-06 | 5.478165 |
| YWHAE | -0.23961 | 0.846975 | 9.399121 | -5.33091 | 6.79E-07 | 2.88E-06 | 5.476659 |
| ZNF268 | 0.278343 | 1.212801 | 6.143077 | 5.3307 | 6.80E-07 | 2.88E-06 | 5.475811 |
| ZNF83 | 0.233222 | 1.175457 | 5.63033 | 5.328904 | 6.85E-07 | 2.90E-06 | 5.468489 |
| ZCCHC10 | 0.243541 | 1.183895 | 5.69978 | 5.328644 | 6.86E-07 | 2.90E-06 | 5.467431 |
| SLC9A5 | -0.17352 | 0.886679 | 5.345934 | -5.32781 | 6.88E-07 | 2.91E-06 | 5.464014 |
| LY6G6D | -0.17775 | 0.884078 | 4.553187 | -5.32757 | 6.89E-07 | 2.91E-06 | 5.463045 |
| CSDC2 | -0.29272 | 0.816359 | 7.052637 | -5.32675 | 6.91E-07 | 2.92E-06 | 5.459726 |
| CA9 | -0.21687 | 0.86043 | 5.523626 | -5.32667 | 6.91E-07 | 2.92E-06 | 5.459372 |
| GPRC5D | -0.21669 | 0.86054 | 5.301978 | -5.32584 | 6.94E-07 | 2.93E-06 | 5.45598 |
| DPEP1 | -0.65112 | 0.636788 | 10.4278 | -5.32486 | 6.97E-07 | 2.94E-06 | 5.452025 |
| TTLL1 | 0.20685 | 1.154166 | 5.950549 | 5.321798 | 7.06E-07 | 2.97E-06 | 5.439532 |
| MEST | 0.596401 | 1.51194 | 8.112967 | 5.321382 | 7.07E-07 | 2.98E-06 | 5.437835 |
| HOXB7 | 0.322831 | 1.250782 | 7.777033 | 5.316328 | 7.22E-07 | 3.04E-06 | 5.417253 |
| IQSEC1 | -0.17072 | 0.888396 | 6.742967 | -5.31544 | 7.25E-07 | 3.05E-06 | 5.413644 |
| MASP2 | -0.14159 | 0.90652 | 5.008462 | -5.31399 | 7.29E-07 | 3.07E-06 | 5.40772 |
| CALU | 0.182357 | 1.134737 | 7.076264 | 5.312565 | 7.34E-07 | 3.08E-06 | 5.401932 |
| KIF3A | 0.169778 | 1.124885 | 5.063956 | 5.311519 | 7.37E-07 | 3.10E-06 | 5.397676 |
| CEACAM3 | -0.18663 | 0.878657 | 5.339451 | -5.31016 | 7.41E-07 | 3.11E-06 | 5.39214 |
| PTK2 | 0.165454 | 1.121519 | 7.867253 | 5.310109 | 7.41E-07 | 3.11E-06 | 5.391938 |
| EVI2A | 0.650106 | 1.569284 | 5.156264 | 5.309971 | 7.42E-07 | 3.11E-06 | 5.391379 |
| HNF1B | 0.281763 | 1.21568 | 9.488681 | 5.308936 | 7.45E-07 | 3.12E-06 | 5.387166 |
| KLK3 | -0.14417 | 0.904897 | 5.020879 | -5.30835 | 7.47E-07 | 3.13E-06 | 5.38479 |
| TBL1XR1 | 0.290889 | 1.223394 | 6.613846 | 5.308066 | 7.48E-07 | 3.13E-06 | 5.383627 |
| KIR3DL2 | -0.19871 | 0.871329 | 5.00978 | -5.30775 | 7.49E-07 | 3.13E-06 | 5.382342 |

| | | | | | | | |
|---|---|---|---|---|---|---|---|
| SLC6A16 | -0.23145 | 0.851776 | 4.98011 | -5.3073 | 7.50E-07 | 3.14E-06 | 5.380509 |
| CDH18 | -0.19686 | 0.872447 | 4.923956 | -5.30658 | 7.52E-07 | 3.15E-06 | 5.377571 |
| EMID1 | -0.18545 | 0.879375 | 6.553077 | -5.30605 | 7.54E-07 | 3.15E-06 | 5.375417 |
| MLLT11 | 0.30601 | 1.236284 | 5.781758 | 5.303792 | 7.61E-07 | 3.18E-06 | 5.366243 |
| STAT5A | -0.27904 | 0.82414 | 7.115275 | -5.30283 | 7.64E-07 | 3.19E-06 | 5.36235 |
| APBB1 | -0.20173 | 0.869508 | 5.752418 | -5.30237 | 7.66E-07 | 3.20E-06 | 5.360467 |
| SPRY2 | -0.30372 | 0.810161 | 6.612418 | -5.302 | 7.67E-07 | 3.20E-06 | 5.358955 |
| MIA2 | -0.20621 | 0.86681 | 4.398462 | -5.3015 | 7.69E-07 | 3.21E-06 | 5.356921 |
| R3HDM2 | -0.30337 | 0.810359 | 8.298462 | -5.29859 | 7.78E-07 | 3.24E-06 | 5.345102 |
| STMN4 | -0.16664 | 0.890917 | 4.678901 | -5.29694 | 7.83E-07 | 3.26E-06 | 5.338374 |
| LOC10050 | -0.17769 | 0.884117 | 4.976044 | -5.29291 | 7.97E-07 | 3.32E-06 | 5.322025 |
| CNTFR | -0.23099 | 0.852053 | 5.851429 | -5.29278 | 7.97E-07 | 3.32E-06 | 5.321503 |
| MAN2A2 | -0.16812 | 0.890001 | 6.301429 | -5.29219 | 7.99E-07 | 3.33E-06 | 5.319104 |
| PPP1CA | 0.178628 | 1.131807 | 9.526593 | 5.29115 | 8.03E-07 | 3.34E-06 | 5.314864 |
| ARF1 | 0.164841 | 1.121042 | 9.819341 | 5.290988 | 8.03E-07 | 3.34E-06 | 5.314207 |
| SNN | 0.190396 | 1.141077 | 5.361978 | 5.290123 | 8.06E-07 | 3.35E-06 | 5.310696 |
| PMEL | -0.1793 | 0.883132 | 5.702857 | -5.28903 | 8.10E-07 | 3.36E-06 | 5.306255 |
| ARR3 | -0.24596 | 0.843257 | 6.17033 | -5.28877 | 8.11E-07 | 3.37E-06 | 5.305188 |
| TP73 | -0.20005 | 0.870521 | 4.468901 | -5.28845 | 8.12E-07 | 3.37E-06 | 5.303903 |
| PDCD5 | 0.333797 | 1.260326 | 6.155934 | 5.286313 | 8.19E-07 | 3.40E-06 | 5.295224 |
| MUTYH | -0.20486 | 0.867623 | 7.15 | -5.28616 | 8.20E-07 | 3.40E-06 | 5.294606 |
| SFT2D2 | 0.32528 | 1.252908 | 6.658462 | 5.286119 | 8.20E-07 | 3.40E-06 | 5.294436 |
| RRN3 | 0.247319 | 1.186999 | 5.931648 | 5.285712 | 8.21E-07 | 3.40E-06 | 5.292784 |
| NOC3L | 0.267304 | 1.203557 | 6.620879 | 5.284577 | 8.25E-07 | 3.41E-06 | 5.288178 |
| NKAIN1 | -0.19726 | 0.872205 | 5.175714 | -5.28232 | 8.33E-07 | 3.44E-06 | 5.279 |
| SPRR3 | -0.20921 | 0.865012 | 5.032198 | -5.28229 | 8.33E-07 | 3.44E-06 | 5.278897 |
| MLLT4-AS1 | -0.27361 | 0.827245 | 5.092088 | -5.28172 | 8.35E-07 | 3.45E-06 | 5.276581 |
| BAG2 | 0.382377 | 1.303488 | 6.046044 | 5.281605 | 8.36E-07 | 3.45E-06 | 5.276118 |
| CD52 | 0.776986 | 1.713547 | 5.568571 | 5.280529 | 8.39E-07 | 3.46E-06 | 5.271752 |
| TMEM243 | 0.280454 | 1.214577 | 8.259121 | 5.280453 | 8.40E-07 | 3.46E-06 | 5.271443 |
| RUNDC3B | -0.34783 | 0.785765 | 5.795604 | -5.27741 | 8.50E-07 | 3.51E-06 | 5.25911 |
| SPTB | -0.17817 | 0.883824 | 5.684286 | -5.27678 | 8.53E-07 | 3.51E-06 | 5.256548 |
| ARMT1 | 0.35986 | 1.283301 | 8.481648 | 5.275759 | 8.56E-07 | 3.53E-06 | 5.252406 |
| DLG5 | 0.240193 | 1.181151 | 6.627473 | 5.275715 | 8.57E-07 | 3.53E-06 | 5.252228 |
| RDH8 | -0.1654 | 0.891681 | 5.025165 | -5.2751 | 8.59E-07 | 3.53E-06 | 5.249747 |
| TMBIM1 | 0.251768 | 1.190665 | 8.704066 | 5.273779 | 8.64E-07 | 3.55E-06 | 5.244377 |
| ARVCF | -0.19385 | 0.874272 | 7.341099 | -5.2728 | 8.67E-07 | 3.57E-06 | 5.240413 |
| MYOC | -0.18925 | 0.877064 | 4.97033 | -5.27243 | 8.68E-07 | 3.57E-06 | 5.238918 |
| CIDEC | -0.24181 | 0.845686 | 5.309121 | -5.26856 | 8.83E-07 | 3.63E-06 | 5.223226 |
| ARMCX5 | 0.227565 | 1.170857 | 6.462967 | 5.26789 | 8.85E-07 | 3.63E-06 | 5.220509 |
| CCIN | -0.18694 | 0.878469 | 5.105385 | -5.26694 | 8.89E-07 | 3.65E-06 | 5.216661 |
| ST14 | 0.286792 | 1.219925 | 6.527473 | 5.266767 | 8.89E-07 | 3.65E-06 | 5.215958 |
| TNNC1 | -0.40957 | 0.75285 | 4.997033 | -5.26645 | 8.91E-07 | 3.65E-06 | 5.214665 |
| PPP1R10 | -0.23644 | 0.848838 | 7.103297 | -5.26409 | 8.99E-07 | 3.69E-06 | 5.205119 |
| JRKL | 0.215836 | 1.161377 | 6.609341 | 5.263776 | 9.01E-07 | 3.69E-06 | 5.203842 |
| HCP5 | 0.446889 | 1.363098 | 5.900989 | 5.26363 | 9.01E-07 | 3.69E-06 | 5.20325 |
| SLC25A44 | -0.21934 | 0.858959 | 8.011319 | -5.26261 | 9.05E-07 | 3.70E-06 | 5.199132 |

| | | | | | | | |
|---|---|---|---|---|---|---|---|
| PPP1R12A | 0.193826 | 1.143793 | 6.298022 | 5.262434 | 9.06E-07 | 3.70E-06 | 5.198406 |
| ARHGEF15 | -0.19256 | 0.875051 | 5.23956 | -5.2605 | 9.13E-07 | 3.73E-06 | 5.190564 |
| OR1G1 | -0.17571 | 0.885332 | 5.026154 | -5.25985 | 9.15E-07 | 3.74E-06 | 5.18795 |
| GRAMD4 | 0.198696 | 1.14766 | 5.59956 | 5.257756 | 9.24E-07 | 3.77E-06 | 5.179466 |
| KRAS | 0.279406 | 1.213695 | 7.621429 | 5.256876 | 9.27E-07 | 3.78E-06 | 5.175906 |
| PCSK1N | -0.44765 | 0.733238 | 8.354505 | -5.25662 | 9.28E-07 | 3.79E-06 | 5.17487 |
| WARS | 0.234899 | 1.176824 | 8.416593 | 5.253825 | 9.39E-07 | 3.83E-06 | 5.163558 |
| ENTPD7 | 0.256449 | 1.194535 | 5.547033 | 5.252805 | 9.43E-07 | 3.84E-06 | 5.159432 |
| GABARAPL | -0.27844 | 0.824482 | 8.292527 | -5.25176 | 9.47E-07 | 3.86E-06 | 5.155187 |
| UBXN4 | 0.186135 | 1.137712 | 8.308132 | 5.251505 | 9.48E-07 | 3.86E-06 | 5.154172 |
| C10orf2 | 0.22943 | 1.172372 | 5.987802 | 5.2512 | 9.49E-07 | 3.86E-06 | 5.15294 |
| SLC48A1 | 0.264449 | 1.201177 | 6.560989 | 5.249369 | 9.57E-07 | 3.89E-06 | 5.145532 |
| RBP1 | 0.381618 | 1.302802 | 5.961538 | 5.248173 | 9.61E-07 | 3.91E-06 | 5.140699 |
| CIT | 0.300957 | 1.231961 | 7.581868 | 5.247983 | 9.62E-07 | 3.91E-06 | 5.139931 |
| SGCA | -0.17817 | 0.883821 | 4.204066 | -5.24755 | 9.64E-07 | 3.92E-06 | 5.138168 |
| PTPRC | 0.455981 | 1.371715 | 4.814615 | 5.245227 | 9.73E-07 | 3.95E-06 | 5.128786 |
| NCF2 | 0.533101 | 1.447037 | 4.893187 | 5.24513 | 9.74E-07 | 3.95E-06 | 5.128394 |
| PPM1D | 0.235357 | 1.177198 | 5.777473 | 5.245079 | 9.74E-07 | 3.95E-06 | 5.128189 |
| HEXA | 0.196121 | 1.145614 | 9.352418 | 5.244478 | 9.76E-07 | 3.96E-06 | 5.12576 |
| HHLA1 | -0.21003 | 0.86452 | 6.294835 | -5.24429 | 9.77E-07 | 3.96E-06 | 5.12501 |
| FAM127A | 0.171425 | 1.12617 | 9.608901 | 5.242969 | 9.83E-07 | 3.98E-06 | 5.119658 |
| KCNE5 | -0.1669 | 0.890753 | 4.186813 | -5.24293 | 9.83E-07 | 3.98E-06 | 5.11949 |
| PCDHGB5 | -0.19877 | 0.871294 | 5.747143 | -5.24288 | 9.83E-07 | 3.98E-06 | 5.119304 |
| MYOG | -0.20675 | 0.866488 | 5.364066 | -5.24034 | 9.93E-07 | 4.02E-06 | 5.109035 |
| RAI14 | 0.265527 | 1.202075 | 8.16 | 5.239768 | 9.96E-07 | 4.03E-06 | 5.106723 |
| CACNB4 | -0.21479 | 0.86167 | 4.928132 | -5.23918 | 9.98E-07 | 4.03E-06 | 5.104341 |
| KEL | -0.20494 | 0.867576 | 6.656484 | -5.23872 | 1.00E-06 | 4.04E-06 | 5.102497 |
| JUNB | -0.66672 | 0.629935 | 6.547692 | -5.23866 | 1.00E-06 | 4.04E-06 | 5.102266 |
| SNF8 | 0.174652 | 1.128692 | 8.315714 | 5.237594 | 1.00E-06 | 4.05E-06 | 5.097943 |
| YAP1 | 0.236807 | 1.178382 | 7.033407 | 5.2373 | 1.01E-06 | 4.06E-06 | 5.096754 |
| DOC2B | -0.1988 | 0.871277 | 5.565824 | -5.23704 | 1.01E-06 | 4.06E-06 | 5.095688 |
| BAG1 | -0.17098 | 0.888242 | 8.491538 | -5.2364 | 1.01E-06 | 4.07E-06 | 5.093138 |
| CUTC | -0.21493 | 0.861589 | 6.671978 | -5.23635 | 1.01E-06 | 4.07E-06 | 5.092924 |
| CASQ1 | -0.18855 | 0.87749 | 4.112198 | -5.23617 | 1.01E-06 | 4.07E-06 | 5.092205 |
| CLIP3 | -0.20087 | 0.870026 | 6.391538 | -5.23592 | 1.01E-06 | 4.07E-06 | 5.091198 |
| RIT1 | 0.406019 | 1.325025 | 6.751648 | 5.235755 | 1.01E-06 | 4.07E-06 | 5.090514 |
| VEZF1 | 0.188546 | 1.139615 | 7.737802 | 5.23484 | 1.02E-06 | 4.09E-06 | 5.086821 |
| KIF20A | 0.396 | 1.315855 | 4.365824 | 5.234157 | 1.02E-06 | 4.10E-06 | 5.08406 |
| DIO1 | -0.51363 | 0.700456 | 10.50253 | -5.22913 | 1.04E-06 | 4.18E-06 | 5.063766 |
| OSBP2 | -0.19403 | 0.874158 | 5.632527 | -5.228 | 1.05E-06 | 4.20E-06 | 5.059221 |
| PAPPA | 0.487372 | 1.401889 | 5.962747 | 5.227919 | 1.05E-06 | 4.20E-06 | 5.058881 |
| LMX1B | -0.19356 | 0.874445 | 5.184066 | -5.22775 | 1.05E-06 | 4.20E-06 | 5.058206 |
| GAS6 | -0.24563 | 0.843449 | 7.625275 | -5.22633 | 1.05E-06 | 4.22E-06 | 5.05245 |
| EIF5 | -0.188 | 0.877825 | 8.757253 | -5.22585 | 1.06E-06 | 4.23E-06 | 5.050552 |
| RSRC1 | 0.191575 | 1.14201 | 5.365604 | 5.225786 | 1.06E-06 | 4.23E-06 | 5.050275 |
| LUM | 0.820237 | 1.765696 | 8.296264 | 5.224287 | 1.06E-06 | 4.25E-06 | 5.044226 |
| SYNE2 | 0.395314 | 1.315229 | 7.214615 | 5.223584 | 1.07E-06 | 4.26E-06 | 5.04139 |

| | | | | | | | |
|---|---|---|---|---|---|---|---|
| OAT | -0.39989 | 0.757917 | 10.67725 | -5.22341 | 1.07E-06 | 4.26E-06 | 5.040687 |
| DCXR | -0.54305 | 0.686317 | 11.53407 | -5.22224 | 1.07E-06 | 4.28E-06 | 5.035973 |
| CNR2 | -0.21241 | 0.863094 | 5.416484 | -5.22212 | 1.07E-06 | 4.28E-06 | 5.035492 |
| RAD54L2 | -0.16878 | 0.889596 | 5.951538 | -5.22183 | 1.07E-06 | 4.29E-06 | 5.034307 |
| ORC2 | 0.20556 | 1.153134 | 5.291868 | 5.221045 | 1.08E-06 | 4.30E-06 | 5.031151 |
| PAM | 0.182952 | 1.135204 | 10.1833 | 5.220087 | 1.08E-06 | 4.31E-06 | 5.02729 |
| PDHA1 | 0.158425 | 1.116068 | 9.087473 | 5.219587 | 1.08E-06 | 4.32E-06 | 5.025274 |
| GLUD1 | -0.30042 | 0.812016 | 10.23253 | -5.21628 | 1.10E-06 | 4.38E-06 | 5.011929 |
| DDX60 | 0.49799 | 1.412245 | 6.565824 | 5.215963 | 1.10E-06 | 4.38E-06 | 5.010664 |
| PRRC2A | -0.21906 | 0.859123 | 6.463846 | -5.21505 | 1.10E-06 | 4.40E-06 | 5.006971 |
| ADGRB1 | -0.16255 | 0.893447 | 4.705055 | -5.21378 | 1.11E-06 | 4.42E-06 | 5.00188 |
| TXNRD1 | 0.323826 | 1.251646 | 8.752308 | 5.212996 | 1.11E-06 | 4.43E-06 | 4.99871 |
| CRLF2 | -0.22062 | 0.858195 | 4.972857 | -5.21241 | 1.12E-06 | 4.44E-06 | 4.996363 |
| DENND1C | -0.23445 | 0.850007 | 5.933626 | -5.21175 | 1.12E-06 | 4.45E-06 | 4.993687 |
| REEP2 | -0.20627 | 0.866778 | 6.916044 | -5.21128 | 1.12E-06 | 4.46E-06 | 4.991811 |
| LEPR | 0.23572 | 1.177494 | 5.123956 | 5.210126 | 1.13E-06 | 4.48E-06 | 4.987147 |
| RNF39 | -0.22616 | 0.854908 | 5.950989 | -5.20828 | 1.14E-06 | 4.51E-06 | 4.979696 |
| KAT2A | -0.19413 | 0.8741 | 7.258132 | -5.20587 | 1.15E-06 | 4.55E-06 | 4.970005 |
| PTPRU | -0.17987 | 0.882783 | 5.626923 | -5.20548 | 1.15E-06 | 4.56E-06 | 4.968449 |
| MT1X | -0.68889 | 0.620329 | 11.70912 | -5.20483 | 1.15E-06 | 4.57E-06 | 4.965821 |
| GTF3C3 | 0.205464 | 1.153057 | 6.457473 | 5.204164 | 1.16E-06 | 4.58E-06 | 4.96314 |
| MRPL18 | 0.163039 | 1.119643 | 9.477363 | 5.203831 | 1.16E-06 | 4.58E-06 | 4.961798 |
| NTAN1 | 0.214242 | 1.160094 | 6.856813 | 5.203756 | 1.16E-06 | 4.58E-06 | 4.961499 |
| SPINK4 | -0.18956 | 0.876876 | 5.106264 | -5.20185 | 1.17E-06 | 4.62E-06 | 4.953821 |
| B3GALNT1 | 0.308671 | 1.238567 | 5.512857 | 5.201453 | 1.17E-06 | 4.62E-06 | 4.95223 |
| ILF2 | 0.312797 | 1.242114 | 8.990549 | 5.200758 | 1.17E-06 | 4.63E-06 | 4.949435 |
| NFASC | -0.33129 | 0.794828 | 5.038352 | -5.20056 | 1.17E-06 | 4.63E-06 | 4.948618 |
| PSEN1 | 0.213976 | 1.15988 | 7.514725 | 5.20014 | 1.17E-06 | 4.64E-06 | 4.946947 |
| SLC17A7 | -0.19136 | 0.875781 | 5.864286 | -5.19883 | 1.18E-06 | 4.66E-06 | 4.941694 |
| EDRF1 | 0.15399 | 1.112643 | 5.265714 | 5.197368 | 1.19E-06 | 4.69E-06 | 4.935795 |
| CALM3 | 0.163232 | 1.119793 | 7.696154 | 5.196337 | 1.19E-06 | 4.71E-06 | 4.931649 |
| STAT3 | 0.282676 | 1.216449 | 8.83022 | 5.195815 | 1.20E-06 | 4.71E-06 | 4.929548 |
| TSPAN7 | -0.35852 | 0.779963 | 7.779231 | -5.19552 | 1.20E-06 | 4.72E-06 | 4.928348 |
| ACTR8 | 0.165169 | 1.121297 | 6.974286 | 5.192374 | 1.21E-06 | 4.78E-06 | 4.91571 |
| CSRP2 | 0.346531 | 1.2715 | 8.995275 | 5.189557 | 1.23E-06 | 4.83E-06 | 4.904387 |
| PMFBP1 | -0.17645 | 0.884875 | 4.632308 | -5.18768 | 1.24E-06 | 4.87E-06 | 4.896826 |
| BAHCC1 | -0.18196 | 0.881507 | 5.761978 | -5.18662 | 1.24E-06 | 4.89E-06 | 4.892589 |
| ZNF271P | 0.253981 | 1.192493 | 6.114725 | 5.186131 | 1.25E-06 | 4.90E-06 | 4.890622 |
| NUP155 | 0.186488 | 1.13799 | 5.404176 | 5.182975 | 1.26E-06 | 4.96E-06 | 4.877941 |
| COL1A1 | 0.425903 | 1.343413 | 5.166264 | 5.182137 | 1.27E-06 | 4.97E-06 | 4.874579 |
| ITIH1 | -0.16724 | 0.890544 | 4.531429 | -5.18053 | 1.27E-06 | 5.00E-06 | 4.868144 |
| GSK3A | -0.15053 | 0.900922 | 7.651868 | -5.18034 | 1.28E-06 | 5.01E-06 | 4.86738 |
| TPPP3 | -0.37354 | 0.771888 | 5.796154 | -5.17973 | 1.28E-06 | 5.02E-06 | 4.864894 |
| PSMA5 | 0.192589 | 1.142813 | 8.811758 | 5.178715 | 1.28E-06 | 5.03E-06 | 4.860835 |
| FLOT1 | 0.218628 | 1.163626 | 8.676374 | 5.178672 | 1.28E-06 | 5.03E-06 | 4.860666 |
| RHCE | -0.24046 | 0.846476 | 4.84044 | -5.17772 | 1.29E-06 | 5.05E-06 | 4.85684 |
| POLL | -0.1705 | 0.888533 | 5.993077 | -5.17625 | 1.30E-06 | 5.08E-06 | 4.850951 |

| | | | | | | | |
|---|---|---|---|---|---|---|---|
| FAM117A | -0.25898 | 0.835678 | 7.457802 | -5.17623 | 1.30E-06 | 5.08E-06 | 4.850845 |
| CYP11B2 | -0.27758 | 0.824974 | 5.961648 | -5.17508 | 1.30E-06 | 5.10E-06 | 4.846238 |
| RNASEH1 | 0.167976 | 1.123481 | 7.001978 | 5.173699 | 1.31E-06 | 5.13E-06 | 4.840704 |
| MRPL33 | 0.19728 | 1.146535 | 10.07516 | 5.172468 | 1.32E-06 | 5.15E-06 | 4.835765 |
| MYOM2 | -0.36008 | 0.77912 | 7.478242 | -5.17242 | 1.32E-06 | 5.15E-06 | 4.835562 |
| CDV3 | 0.230034 | 1.172862 | 6.815275 | 5.172271 | 1.32E-06 | 5.15E-06 | 4.834974 |
| NHLH1 | -0.21429 | 0.861967 | 5.650769 | -5.17207 | 1.32E-06 | 5.15E-06 | 4.834177 |
| IL6R | -0.4118 | 0.751684 | 6.265275 | -5.17196 | 1.32E-06 | 5.15E-06 | 4.833732 |
| HNF1A | -0.23291 | 0.850915 | 6.463736 | -5.17182 | 1.32E-06 | 5.15E-06 | 4.833176 |
| FAM57A | 0.256531 | 1.194603 | 5.840769 | 5.171518 | 1.32E-06 | 5.16E-06 | 4.831955 |
| CCDC176 | 0.233483 | 1.17567 | 6.832198 | 5.170675 | 1.33E-06 | 5.17E-06 | 4.828574 |
| PRR34 | -0.19676 | 0.872509 | 6.328571 | -5.16943 | 1.34E-06 | 5.20E-06 | 4.823585 |
| CEP57 | 0.184908 | 1.136745 | 6.492308 | 5.166686 | 1.35E-06 | 5.25E-06 | 4.812576 |
| MXI1 | -0.23154 | 0.851727 | 9.236374 | -5.16612 | 1.35E-06 | 5.26E-06 | 4.810296 |
| NR2F1 | -0.27431 | 0.826843 | 7.01022 | -5.16564 | 1.36E-06 | 5.27E-06 | 4.808402 |
| SLC7A10 | -0.21441 | 0.861898 | 5.125495 | -5.16418 | 1.36E-06 | 5.30E-06 | 4.802538 |
| S1PR2 | -0.16173 | 0.89395 | 4.851978 | -5.1628 | 1.37E-06 | 5.33E-06 | 4.79701 |
| HSD3B2 | -0.18577 | 0.879178 | 4.938352 | -5.16264 | 1.37E-06 | 5.33E-06 | 4.796353 |
| KANSL1L | 0.266778 | 1.203118 | 5.936923 | 5.162185 | 1.38E-06 | 5.34E-06 | 4.794535 |
| C12orf43 | 0.163961 | 1.120359 | 5.949341 | 5.161905 | 1.38E-06 | 5.34E-06 | 4.793413 |
| F12 | 0.369966 | 1.292323 | 5.811429 | 5.161187 | 1.38E-06 | 5.36E-06 | 4.790533 |
| TIA1 | 0.22087 | 1.165436 | 6.108352 | 5.160574 | 1.39E-06 | 5.37E-06 | 4.788079 |
| RNF111 | 0.200266 | 1.14891 | 7.060989 | 5.159232 | 1.39E-06 | 5.39E-06 | 4.782703 |
| NELL2 | -0.29652 | 0.814213 | 5.23989 | -5.15875 | 1.40E-06 | 5.40E-06 | 4.78078 |
| PDCD4 | 0.245512 | 1.185513 | 6.429451 | 5.157467 | 1.40E-06 | 5.43E-06 | 4.77563 |
| OR2C1 | -0.17717 | 0.884437 | 5.06978 | -5.15698 | 1.41E-06 | 5.44E-06 | 4.773665 |
| EIF4A3 | 0.204981 | 1.152671 | 9.105495 | 5.156273 | 1.41E-06 | 5.45E-06 | 4.770847 |
| PER2 | -0.33853 | 0.790849 | 6.268901 | -5.15619 | 1.41E-06 | 5.45E-06 | 4.770508 |
| FUT4 | 0.277961 | 1.21248 | 5.475714 | 5.155898 | 1.41E-06 | 5.45E-06 | 4.769345 |
| ZNF646 | -0.20098 | 0.869962 | 5.356703 | -5.15456 | 1.42E-06 | 5.48E-06 | 4.763995 |
| SP110 | 0.25101 | 1.19004 | 6.35956 | 5.15381 | 1.42E-06 | 5.50E-06 | 4.760986 |
| PTTG1 | 0.397454 | 1.317181 | 7.501978 | 5.153537 | 1.43E-06 | 5.50E-06 | 4.759893 |
| PTGES2 | -0.15035 | 0.90103 | 7.88978 | -5.15223 | 1.43E-06 | 5.53E-06 | 4.754667 |
| DCTN4 | 0.19514 | 1.144835 | 7.127802 | 5.152083 | 1.43E-06 | 5.53E-06 | 4.75407 |
| CYP2A13 | -0.23368 | 0.850465 | 5.629011 | -5.15164 | 1.44E-06 | 5.54E-06 | 4.752311 |
| NEUROD4 | -0.19721 | 0.872237 | 4.258132 | -5.15043 | 1.44E-06 | 5.56E-06 | 4.747441 |
| EXPH5 | -0.25086 | 0.840393 | 5.687033 | -5.15003 | 1.45E-06 | 5.57E-06 | 4.745853 |
| SND1 | 0.188343 | 1.139454 | 8.398571 | 5.149934 | 1.45E-06 | 5.57E-06 | 4.745469 |
| YY2 | -0.21127 | 0.863776 | 5.578352 | -5.14894 | 1.45E-06 | 5.59E-06 | 4.741493 |
| AVP | -0.23342 | 0.850619 | 5.530879 | -5.14796 | 1.46E-06 | 5.61E-06 | 4.737587 |
| HAND1 | -0.25138 | 0.840091 | 5.293516 | -5.14569 | 1.47E-06 | 5.66E-06 | 4.728479 |
| HAT1 | 0.248053 | 1.187603 | 7.685055 | 5.142935 | 1.49E-06 | 5.72E-06 | 4.717468 |
| NLGN1 | 0.266676 | 1.203033 | 5.162308 | 5.141045 | 1.50E-06 | 5.76E-06 | 4.709908 |
| TKT | 0.424792 | 1.342379 | 8.595714 | 5.140826 | 1.50E-06 | 5.77E-06 | 4.709036 |
| ESF1 | 0.223179 | 1.167303 | 6.663407 | 5.137641 | 1.52E-06 | 5.84E-06 | 4.696303 |
| ZC3H7B | -0.17439 | 0.886141 | 6.646154 | -5.13731 | 1.53E-06 | 5.85E-06 | 4.694959 |
| HLA-DQB1 | 0.884444 | 1.846054 | 6.487363 | 5.135062 | 1.54E-06 | 5.90E-06 | 4.685994 |

| | | | | | | | |
|---|---|---|---|---|---|---|---|
| ZNF281 | 0.241575 | 1.182283 | 5.98033 | 5.134964 | 1.54E-06 | 5.90E-06 | 4.685604 |
| MAP2K3 | -0.24146 | 0.845889 | 7.874945 | -5.13477 | 1.54E-06 | 5.90E-06 | 4.684816 |
| UBE2V2 | 0.168971 | 1.124256 | 7.587253 | 5.134116 | 1.55E-06 | 5.91E-06 | 4.682215 |
| TOR3A | 0.229372 | 1.172325 | 6.725165 | 5.132279 | 1.56E-06 | 5.96E-06 | 4.674875 |
| MLLT3 | 0.210217 | 1.156862 | 5.553736 | 5.131496 | 1.56E-06 | 5.97E-06 | 4.67175 |
| FRY | -0.30435 | 0.809805 | 4.973626 | -5.13071 | 1.57E-06 | 5.99E-06 | 4.668625 |
| PPARA | -0.17695 | 0.884573 | 5.54989 | -5.13056 | 1.57E-06 | 5.99E-06 | 4.667993 |
| VBP1 | 0.154213 | 1.112814 | 9.085824 | 5.130099 | 1.57E-06 | 6.00E-06 | 4.666169 |
| TFPI | 0.348362 | 1.273115 | 6.888571 | 5.129383 | 1.58E-06 | 6.01E-06 | 4.663308 |
| WDR73 | 0.161174 | 1.118197 | 7.352527 | 5.126473 | 1.60E-06 | 6.08E-06 | 4.651689 |
| PNPLA3 | -0.39138 | 0.762402 | 6.164505 | -5.12593 | 1.60E-06 | 6.09E-06 | 4.649529 |
| CAMKV | -0.22976 | 0.852775 | 5.837033 | -5.12523 | 1.60E-06 | 6.11E-06 | 4.646746 |
| IL18 | 0.36656 | 1.289275 | 5.156484 | 5.12474 | 1.61E-06 | 6.12E-06 | 4.644772 |
| ITPKC | -0.22434 | 0.855988 | 6.173846 | -5.12343 | 1.62E-06 | 6.15E-06 | 4.639549 |
| DCHS2 | -0.15227 | 0.899836 | 4.892747 | -5.12239 | 1.62E-06 | 6.17E-06 | 4.635404 |
| SRPK3 | -0.23617 | 0.848994 | 5.315385 | -5.12033 | 1.64E-06 | 6.22E-06 | 4.627176 |
| PRKX | 0.3333 | 1.259892 | 4.883297 | 5.120252 | 1.64E-06 | 6.22E-06 | 4.626864 |
| BCAT2 | 0.298628 | 1.229974 | 7.735934 | 5.119702 | 1.64E-06 | 6.23E-06 | 4.624668 |
| C14orf169 | 0.224386 | 1.16828 | 7.628352 | 5.118614 | 1.65E-06 | 6.26E-06 | 4.620328 |
| NMRK1 | -0.3161 | 0.803238 | 8.309121 | -5.11708 | 1.66E-06 | 6.30E-06 | 4.614211 |
| MRPL49 | -0.1456 | 0.904001 | 10.01582 | -5.11669 | 1.66E-06 | 6.30E-06 | 4.612659 |
| BAAT | -0.18089 | 0.882159 | 5.230549 | -5.1159 | 1.67E-06 | 6.32E-06 | 4.609519 |
| MAP6D1 | -0.17121 | 0.888099 | 5.590989 | -5.11472 | 1.67E-06 | 6.35E-06 | 4.604783 |
| FAM63A | 0.230391 | 1.173153 | 7.961538 | 5.114381 | 1.68E-06 | 6.36E-06 | 4.603448 |
| OR7A5 | -0.18937 | 0.876987 | 5.014615 | -5.11221 | 1.69E-06 | 6.41E-06 | 4.594797 |
| COL18A1 | 0.293599 | 1.225694 | 7.637143 | 5.109294 | 1.71E-06 | 6.48E-06 | 4.583171 |
| ALDH3A1 | -0.2269 | 0.85447 | 4.927363 | -5.10818 | 1.72E-06 | 6.51E-06 | 4.578749 |
| DSG2 | 0.281923 | 1.215814 | 8.275934 | 5.10796 | 1.72E-06 | 6.51E-06 | 4.577858 |
| CXXC4 | -0.19676 | 0.872506 | 4.508352 | -5.1078 | 1.72E-06 | 6.52E-06 | 4.577219 |
| SH2D2A | -0.23721 | 0.848383 | 5.948132 | -5.1071 | 1.73E-06 | 6.53E-06 | 4.574447 |
| SHC1 | 0.186763 | 1.138207 | 7.296703 | 5.106581 | 1.73E-06 | 6.54E-06 | 4.572362 |
| HSD11B2 | -0.52824 | 0.693399 | 9.547912 | -5.10623 | 1.73E-06 | 6.55E-06 | 4.570948 |
| ANKRD55 | -0.34033 | 0.789859 | 4.666703 | -5.10518 | 1.74E-06 | 6.58E-06 | 4.566764 |
| ZNRF4 | -0.17487 | 0.885848 | 5.914396 | -5.10408 | 1.75E-06 | 6.60E-06 | 4.562398 |
| PDE4C | -0.17481 | 0.885886 | 5.647253 | -5.10238 | 1.76E-06 | 6.64E-06 | 4.555626 |
| OR52A1 | -0.17351 | 0.886684 | 4.326374 | -5.10234 | 1.76E-06 | 6.64E-06 | 4.555484 |
| BTN3A3 | 0.341971 | 1.267487 | 4.687802 | 5.101886 | 1.77E-06 | 6.65E-06 | 4.553663 |
| DPP6 | -0.43797 | 0.738175 | 5.584945 | -5.1016 | 1.77E-06 | 6.66E-06 | 4.552543 |
| ZBTB22 | -0.19417 | 0.874073 | 6.266154 | -5.10151 | 1.77E-06 | 6.66E-06 | 4.552161 |
| LOC10013 | -0.18834 | 0.877616 | 4.921648 | -5.10128 | 1.77E-06 | 6.66E-06 | 4.551256 |
| PSME3 | 0.188232 | 1.139366 | 7.313516 | 5.099168 | 1.79E-06 | 6.72E-06 | 4.542845 |
| WRAP73 | 0.169343 | 1.124546 | 5.074176 | 5.099053 | 1.79E-06 | 6.72E-06 | 4.542387 |
| ZC3H4 | 0.230604 | 1.173326 | 8.481209 | 5.098071 | 1.79E-06 | 6.74E-06 | 4.538478 |
| HSPA12A | -0.27463 | 0.826663 | 7.795934 | -5.09719 | 1.80E-06 | 6.76E-06 | 4.534954 |
| GRIA3 | -0.16295 | 0.893199 | 4.156813 | -5.09699 | 1.80E-06 | 6.77E-06 | 4.53417 |
| CTSA | 0.260527 | 1.197916 | 11.03253 | 5.09536 | 1.81E-06 | 6.81E-06 | 4.527691 |
| ZNF34 | -0.17777 | 0.884067 | 5.052308 | -5.09498 | 1.82E-06 | 6.82E-06 | 4.526181 |

| | | | | | | | |
|---|---|---|---|---|---|---|---|
| PEA15 | 0.314681 | 1.243737 | 7.476264 | 5.094725 | 1.82E-06 | 6.82E-06 | 4.525163 |
| LINC0113€ | -0.18595 | 0.879069 | 4.94022 | -5.09404 | 1.82E-06 | 6.84E-06 | 4.522453 |
| TRMT2A | -0.15928 | 0.895472 | 6.523626 | -5.09258 | 1.84E-06 | 6.87E-06 | 4.516612 |
| PHYH | -0.37334 | 0.771992 | 11.07495 | -5.09246 | 1.84E-06 | 6.87E-06 | 4.51614 |
| GALNT2 | 0.270826 | 1.206498 | 6.551099 | 5.092323 | 1.84E-06 | 6.88E-06 | 4.51561 |
| SLC7A7 | -0.49496 | 0.709581 | 11.84198 | -5.09217 | 1.84E-06 | 6.88E-06 | 4.515018 |
| ZNF292 | 0.216014 | 1.16152 | 5.927473 | 5.08985 | 1.86E-06 | 6.94E-06 | 4.505772 |
| BTF3 | 0.213681 | 1.159643 | 10.05132 | 5.088384 | 1.87E-06 | 6.98E-06 | 4.499944 |
| ABL1 | 0.22713 | 1.170504 | 7.843187 | 5.087306 | 1.88E-06 | 7.01E-06 | 4.495658 |
| MAN2C1 | -0.19839 | 0.871522 | 6.904286 | -5.08725 | 1.88E-06 | 7.01E-06 | 4.495438 |
| ALOX5AP | 0.580599 | 1.49547 | 6.434066 | 5.086895 | 1.88E-06 | 7.01E-06 | 4.494025 |
| SLC22A6 | -0.46508 | 0.72443 | 9.761319 | -5.08616 | 1.88E-06 | 7.03E-06 | 4.491095 |
| LRRC3 | -0.14852 | 0.902174 | 4.183077 | -5.08584 | 1.89E-06 | 7.04E-06 | 4.48984 |
| KCNS1 | -0.21257 | 0.863001 | 6.439451 | -5.08551 | 1.89E-06 | 7.04E-06 | 4.488507 |
| SFN | 0.529502 | 1.443431 | 7.189451 | 5.08465 | 1.90E-06 | 7.07E-06 | 4.485101 |
| DDX50 | 0.279174 | 1.2135 | 7.940879 | 5.083966 | 1.90E-06 | 7.08E-06 | 4.482383 |
| PSMA6 | 0.262894 | 1.199883 | 10.36022 | 5.083772 | 1.90E-06 | 7.08E-06 | 4.481612 |
| NUP54 | 0.206575 | 1.153945 | 7.228022 | 5.083008 | 1.91E-06 | 7.10E-06 | 4.478579 |
| BCAS3 | -0.19481 | 0.873687 | 6.897143 | -5.07997 | 1.93E-06 | 7.19E-06 | 4.466515 |
| SLC26A10 | -0.19429 | 0.874 | 4.500659 | -5.07883 | 1.94E-06 | 7.22E-06 | 4.461971 |
| DPY19L1 | 0.25229 | 1.191096 | 5.377802 | 5.078029 | 1.95E-06 | 7.24E-06 | 4.458801 |
| MS4A12 | -0.14995 | 0.901281 | 3.848022 | -5.07754 | 1.95E-06 | 7.25E-06 | 4.456878 |
| NAP1L4 | 0.191778 | 1.14217 | 7.804835 | 5.075591 | 1.97E-06 | 7.31E-06 | 4.449122 |
| GLRX | -0.18163 | 0.881707 | 10.32692 | -5.07478 | 1.97E-06 | 7.32E-06 | 4.445886 |
| GRIK3 | -0.17342 | 0.886738 | 4.17033 | -5.07472 | 1.98E-06 | 7.32E-06 | 4.44565 |
| CYP24A1 | 1.157329 | 2.23044 | 7.002088 | 5.07471 | 1.98E-06 | 7.32E-06 | 4.445622 |
| ERGIC3 | 0.15015 | 1.109685 | 9.530989 | 5.069443 | 2.02E-06 | 7.48E-06 | 4.42472 |
| WT1-AS | -0.19383 | 0.874284 | 5.341978 | -5.06934 | 2.02E-06 | 7.48E-06 | 4.424315 |
| AURKC | -0.22051 | 0.858264 | 4.938132 | -5.06911 | 2.02E-06 | 7.49E-06 | 4.423409 |
| WNT7B | -0.22462 | 0.855821 | 5.481099 | -5.06732 | 2.04E-06 | 7.54E-06 | 4.416288 |
| COL4A3BP | 0.322633 | 1.250611 | 6.608022 | 5.066061 | 2.05E-06 | 7.57E-06 | 4.411304 |
| ZBP1 | -0.14851 | 0.902183 | 4.473736 | -5.06564 | 2.05E-06 | 7.58E-06 | 4.409637 |
| SLURP1 | -0.1515 | 0.900312 | 4.117473 | -5.06386 | 2.07E-06 | 7.64E-06 | 4.402575 |
| SH3GL2 | -0.50926 | 0.702582 | 7.201429 | -5.06331 | 2.07E-06 | 7.65E-06 | 4.400409 |
| HIST1H2Bk | 0.526749 | 1.440679 | 7.034176 | 5.063045 | 2.07E-06 | 7.65E-06 | 4.399346 |
| SATB2 | 0.55543 | 1.469607 | 7.059011 | 5.062445 | 2.08E-06 | 7.67E-06 | 4.396967 |
| CSTA | 0.590469 | 1.505736 | 4.683077 | 5.0624 | 2.08E-06 | 7.67E-06 | 4.39679 |
| PTMS | -0.29225 | 0.81663 | 6.354396 | -5.06201 | 2.08E-06 | 7.68E-06 | 4.395241 |
| RAN | 0.239647 | 1.180704 | 8.232637 | 5.061841 | 2.08E-06 | 7.68E-06 | 4.394573 |
| TMPO | 0.18044 | 1.133229 | 5.349011 | 5.060002 | 2.10E-06 | 7.73E-06 | 4.387282 |
| CCDC106 | -0.15243 | 0.899737 | 5.445495 | -5.05968 | 2.10E-06 | 7.74E-06 | 4.38599 |
| CDH17 | -0.16955 | 0.88912 | 5.026374 | -5.05953 | 2.10E-06 | 7.74E-06 | 4.385424 |
| CD48 | 0.732802 | 1.661864 | 6.178462 | 5.05913 | 2.11E-06 | 7.75E-06 | 4.383828 |
| ME1 | 0.41313 | 1.331572 | 4.955165 | 5.057961 | 2.12E-06 | 7.79E-06 | 4.379197 |
| DAO | -0.62706 | 0.647493 | 9.272088 | -5.05743 | 2.12E-06 | 7.80E-06 | 4.377099 |
| IGFBP4 | -0.41961 | 0.747625 | 9.50989 | -5.05526 | 2.14E-06 | 7.87E-06 | 4.368509 |
| DYNC1LI2 | 0.206314 | 1.153737 | 8.236154 | 5.054144 | 2.15E-06 | 7.90E-06 | 4.364074 |

| | | | | | | | |
|---|---|---|---|---|---|---|---|
| IL10RA | 0.444348 | 1.360699 | 5.425385 | 5.053277 | 2.16E-06 | 7.92E-06 | 4.360643 |
| DNPH1 | -0.21344 | 0.862476 | 9.919451 | -5.05193 | 2.17E-06 | 7.96E-06 | 4.355323 |
| CLDN1 | 0.48985 | 1.404299 | 5.305495 | 5.051931 | 2.17E-06 | 7.96E-06 | 4.35531 |
| DLG3 | 0.155932 | 1.114141 | 6.124066 | 5.051639 | 2.17E-06 | 7.97E-06 | 4.354156 |
| BCKDHA | -0.18692 | 0.878478 | 8.796044 | -5.05158 | 2.17E-06 | 7.97E-06 | 4.353938 |
| NPHS2 | -1.09789 | 0.4672 | 8.462088 | -5.04943 | 2.19E-06 | 8.03E-06 | 4.345419 |
| RBFOX1 | -0.17509 | 0.885711 | 4.784286 | -5.04942 | 2.19E-06 | 8.03E-06 | 4.345356 |
| PAPOLA | 0.186304 | 1.137845 | 7.985824 | 5.04926 | 2.19E-06 | 8.03E-06 | 4.344736 |
| EPB42 | -0.19336 | 0.874565 | 4.533077 | -5.0486 | 2.20E-06 | 8.05E-06 | 4.342123 |
| GRB10 | -0.23959 | 0.846984 | 7.90978 | -5.0484 | 2.20E-06 | 8.05E-06 | 4.34134 |
| NRBF2 | 0.265638 | 1.202167 | 5.075055 | 5.047973 | 2.21E-06 | 8.06E-06 | 4.339643 |
| MXRA8 | -0.24835 | 0.841857 | 7.281319 | -5.04739 | 2.21E-06 | 8.08E-06 | 4.33735 |
| SERPINA1( | -0.17597 | 0.885172 | 4.174286 | -5.04714 | 2.21E-06 | 8.08E-06 | 4.336349 |
| ABCC3 | 0.473913 | 1.388871 | 6.63044 | 5.046576 | 2.22E-06 | 8.10E-06 | 4.334115 |
| DHCR24 | 0.331391 | 1.258226 | 7.396484 | 5.044937 | 2.23E-06 | 8.15E-06 | 4.327628 |
| HNMT | -0.216 | 0.860952 | 7.663407 | -5.04472 | 2.23E-06 | 8.15E-06 | 4.326756 |
| PLBD1 | 0.264715 | 1.201399 | 8.563077 | 5.043779 | 2.24E-06 | 8.18E-06 | 4.323049 |
| THOC6 | 0.230367 | 1.173133 | 5.92044 | 5.04302 | 2.25E-06 | 8.20E-06 | 4.320045 |
| FGF6 | -0.16903 | 0.889441 | 5.99011 | -5.04183 | 2.26E-06 | 8.24E-06 | 4.315341 |
| CLN5 | 0.195903 | 1.145441 | 7.942527 | 5.040683 | 2.27E-06 | 8.28E-06 | 4.3108 |
| LASP1 | 0.206575 | 1.153945 | 9.068022 | 5.040422 | 2.27E-06 | 8.28E-06 | 4.309768 |
| DHRS3 | -0.2998 | 0.812364 | 8.920659 | -5.03976 | 2.28E-06 | 8.30E-06 | 4.307156 |
| LINC01565 | -0.17691 | 0.884594 | 4.921429 | -5.03761 | 2.30E-06 | 8.37E-06 | 4.298635 |
| ASIC2 | -0.17615 | 0.885062 | 4.626154 | -5.03559 | 2.32E-06 | 8.44E-06 | 4.290665 |
| CHMP4A | 0.165966 | 1.121917 | 8.180549 | 5.035196 | 2.32E-06 | 8.45E-06 | 4.28911 |
| ABCA1 | 0.418604 | 1.336633 | 8.524176 | 5.033085 | 2.34E-06 | 8.52E-06 | 4.280768 |
| PDE6A | -0.22859 | 0.853466 | 4.86022 | -5.03232 | 2.35E-06 | 8.54E-06 | 4.277755 |
| HOXB6 | 0.30701 | 1.237141 | 6.427253 | 5.031576 | 2.36E-06 | 8.56E-06 | 4.274805 |
| HMP19 | -0.174 | 0.886385 | 4.544176 | -5.03075 | 2.37E-06 | 8.59E-06 | 4.271526 |
| HBEGF | -0.46984 | 0.722047 | 5.745055 | -5.02987 | 2.38E-06 | 8.61E-06 | 4.268063 |
| SLC8A2 | -0.18012 | 0.882629 | 4.935495 | -5.02922 | 2.38E-06 | 8.63E-06 | 4.265505 |
| MAST3 | -0.2011 | 0.869886 | 6.350989 | -5.02713 | 2.40E-06 | 8.70E-06 | 4.257233 |
| SLC22A14 | -0.22255 | 0.857049 | 7.165165 | -5.02537 | 2.42E-06 | 8.76E-06 | 4.250306 |
| ITGA8 | -0.21762 | 0.859981 | 4.999341 | -5.02231 | 2.45E-06 | 8.87E-06 | 4.238221 |
| KDM5C | -0.17163 | 0.88784 | 6.681868 | -5.02109 | 2.46E-06 | 8.91E-06 | 4.233422 |
| NPFF | -0.14189 | 0.906332 | 4.614835 | -5.02026 | 2.47E-06 | 8.94E-06 | 4.230121 |
| MEPE | -0.20057 | 0.87021 | 5.355385 | -5.01924 | 2.48E-06 | 8.97E-06 | 4.226097 |
| SS18L1 | -0.25059 | 0.84055 | 6.809341 | -5.01674 | 2.51E-06 | 9.06E-06 | 4.216242 |
| ATP1A3 | -0.13964 | 0.907747 | 4.277253 | -5.0159 | 2.52E-06 | 9.09E-06 | 4.212919 |
| ADH6 | -0.38803 | 0.764173 | 8.536813 | -5.01462 | 2.53E-06 | 9.13E-06 | 4.207885 |
| GZMA | 0.602802 | 1.518663 | 4.994176 | 5.013893 | 2.54E-06 | 9.15E-06 | 4.205018 |
| UBE3C | 0.233614 | 1.175776 | 6.828132 | 5.013773 | 2.54E-06 | 9.15E-06 | 4.204545 |
| IFT122 | 0.242304 | 1.182881 | 7.373516 | 5.013523 | 2.54E-06 | 9.16E-06 | 4.203558 |
| FJX1 | 0.253391 | 1.192006 | 5.567912 | 5.012585 | 2.55E-06 | 9.19E-06 | 4.199861 |
| FGF5 | -0.15832 | 0.896069 | 4.557363 | -5.0118 | 2.56E-06 | 9.22E-06 | 4.196753 |
| CRAT | -0.18162 | 0.881713 | 7.737363 | -5.01149 | 2.56E-06 | 9.22E-06 | 4.195525 |
| GNG13 | -0.14388 | 0.905082 | 4.004286 | -5.01136 | 2.56E-06 | 9.23E-06 | 4.195023 |

| | | | | | | | |
|---|---|---|---|---|---|---|---|
| PKP2 | -0.21603 | 0.860929 | 5.271648 | -5.0109 | 2.57E-06 | 9.24E-06 | 4.193211 |
| HGFAC | -0.19301 | 0.874776 | 4.478901 | -5.00979 | 2.58E-06 | 9.28E-06 | 4.188835 |
| ZFP30 | 0.186541 | 1.138032 | 6.036593 | 5.008659 | 2.59E-06 | 9.32E-06 | 4.184388 |
| TULP2 | -0.17293 | 0.887038 | 5.162527 | -5.00803 | 2.60E-06 | 9.34E-06 | 4.181913 |
| FCGRT | -0.27 | 0.829322 | 9.506703 | -5.00729 | 2.61E-06 | 9.36E-06 | 4.178989 |
| CCNJL | -0.20961 | 0.864769 | 5.433736 | -5.00691 | 2.61E-06 | 9.37E-06 | 4.177515 |
| RAX | -0.21587 | 0.861027 | 5.949121 | -5.0069 | 2.61E-06 | 9.37E-06 | 4.177471 |
| ECHDC2 | -0.22147 | 0.857689 | 10.50418 | -5.00634 | 2.62E-06 | 9.38E-06 | 4.175234 |
| TTF2 | 0.222072 | 1.166408 | 4.063077 | 5.005727 | 2.62E-06 | 9.40E-06 | 4.172837 |
| SCFD1 | 0.174198 | 1.128337 | 6.695055 | 5.004875 | 2.63E-06 | 9.43E-06 | 4.169482 |
| LOC10050 | -0.17322 | 0.88686 | 5.989341 | -5.00381 | 2.64E-06 | 9.47E-06 | 4.165281 |
| CHRNB2 | -0.15446 | 0.898469 | 6.882967 | -5.00377 | 2.64E-06 | 9.47E-06 | 4.165142 |
| FGF16 | -0.17872 | 0.883484 | 4.449011 | -5.00337 | 2.65E-06 | 9.48E-06 | 4.163537 |
| N6AMT1 | 0.270778 | 1.206458 | 5.228901 | 5.001853 | 2.66E-06 | 9.53E-06 | 4.157579 |
| RCBTB1 | 0.365647 | 1.28846 | 6.384945 | 4.999403 | 2.69E-06 | 9.63E-06 | 4.147934 |
| HTR1F | -0.16546 | 0.891642 | 4.242308 | -4.99815 | 2.70E-06 | 9.67E-06 | 4.143012 |
| INTS12 | 0.190874 | 1.141455 | 7.303736 | 4.996889 | 2.72E-06 | 9.72E-06 | 4.138042 |
| PPP6R1 | -0.14495 | 0.90441 | 7.075495 | -4.9922 | 2.77E-06 | 9.90E-06 | 4.119583 |
| R3HCC1 | -0.1549 | 0.898196 | 8.182967 | -4.99151 | 2.78E-06 | 9.92E-06 | 4.116867 |
| SMARCD3 | 0.265 | 1.201636 | 5.616044 | 4.989315 | 2.80E-06 | 1.00E-05 | 4.10825 |
| BAZ1A | 0.28244 | 1.21625 | 4.809451 | 4.989057 | 2.81E-06 | 1.00E-05 | 4.107235 |
| PRORY | -0.20804 | 0.865713 | 5.425385 | -4.98861 | 2.81E-06 | 1.00E-05 | 4.105479 |
| PEX19 | -0.14819 | 0.90238 | 7.858022 | -4.98717 | 2.83E-06 | 1.01E-05 | 4.099814 |
| VTCN1 | 0.610671 | 1.52697 | 6.492198 | 4.98676 | 2.83E-06 | 1.01E-05 | 4.098207 |
| SAFB2 | -0.16156 | 0.894058 | 7.63989 | -4.98622 | 2.84E-06 | 1.01E-05 | 4.096066 |
| ZNF862 | -0.18562 | 0.879272 | 6.875385 | -4.98607 | 2.84E-06 | 1.01E-05 | 4.095476 |
| TAF1 | -0.1789 | 0.883377 | 6.191099 | -4.98427 | 2.86E-06 | 1.02E-05 | 4.08843 |
| RNF122 | -0.2002 | 0.870431 | 6.552088 | -4.98289 | 2.88E-06 | 1.02E-05 | 4.083005 |
| ALG6 | 0.216855 | 1.162197 | 7.005714 | 4.982682 | 2.88E-06 | 1.02E-05 | 4.082183 |
| ABCG4 | -0.17481 | 0.885886 | 5.337253 | -4.9813 | 2.90E-06 | 1.03E-05 | 4.076748 |
| FOXN3-AS | -0.17891 | 0.883371 | 4.490659 | -4.98017 | 2.91E-06 | 1.03E-05 | 4.072327 |
| UPK1A | -0.17158 | 0.887867 | 4.373846 | -4.97945 | 2.92E-06 | 1.04E-05 | 4.069506 |
| GLB1L2 | -0.33527 | 0.792638 | 8.767253 | -4.97896 | 2.92E-06 | 1.04E-05 | 4.067559 |
| CRK | 0.171768 | 1.126438 | 8.264505 | 4.978391 | 2.93E-06 | 1.04E-05 | 4.06533 |
| CACNA1S | -0.18182 | 0.881592 | 4.578352 | -4.97754 | 2.94E-06 | 1.04E-05 | 4.062 |
| PFKFB1 | -0.22523 | 0.855458 | 5.603187 | -4.97718 | 2.95E-06 | 1.04E-05 | 4.060576 |
| SLC5A7 | -0.15523 | 0.897988 | 4.477802 | -4.97585 | 2.96E-06 | 1.05E-05 | 4.055366 |
| C22orf46 | -0.13721 | 0.909277 | 4.417802 | -4.9751 | 2.97E-06 | 1.05E-05 | 4.052389 |
| NRIP2 | -0.21847 | 0.859474 | 5.020659 | -4.97429 | 2.98E-06 | 1.06E-05 | 4.049216 |
| PSG4 | -0.40329 | 0.756135 | 5.112747 | -4.97376 | 2.99E-06 | 1.06E-05 | 4.047134 |
| CCT6B | -0.35111 | 0.783983 | 5.466593 | -4.97241 | 3.00E-06 | 1.06E-05 | 4.04184 |
| CTLA4 | -0.17514 | 0.885684 | 4.552308 | -4.97109 | 3.02E-06 | 1.07E-05 | 4.036681 |
| CYP2B6 | -0.4242 | 0.74525 | 5.761099 | -4.97098 | 3.02E-06 | 1.07E-05 | 4.036238 |
| PPFIA2 | -0.15981 | 0.895142 | 4.829451 | -4.97064 | 3.03E-06 | 1.07E-05 | 4.034903 |
| CBX5 | 0.242981 | 1.183435 | 5.894286 | 4.970272 | 3.03E-06 | 1.07E-05 | 4.033464 |
| CCDC87 | -0.185 | 0.879652 | 5.393736 | -4.96905 | 3.05E-06 | 1.08E-05 | 4.02868 |
| ARHGAP32 | -0.15118 | 0.900511 | 5.721978 | -4.96763 | 3.06E-06 | 1.08E-05 | 4.023098 |

| | | | | | | | |
|---|---|---|---|---|---|---|---|
| ITGA9 | -0.16146 | 0.89412 | 4.674505 | -4.96737 | 3.07E-06 | 1.08E-05 | 4.022083 |
| GRAP2 | -0.17263 | 0.887225 | 4.876374 | -4.96691 | 3.07E-06 | 1.08E-05 | 4.020294 |
| CCL1 | -0.16628 | 0.89114 | 4.495385 | -4.96641 | 3.08E-06 | 1.09E-05 | 4.018306 |
| TRAM2 | 0.217517 | 1.162731 | 6.135824 | 4.965681 | 3.09E-06 | 1.09E-05 | 4.015456 |
| TCEB1 | 0.146918 | 1.107202 | 7.253956 | 4.96406 | 3.11E-06 | 1.09E-05 | 4.009101 |
| DTX2P1-UI | -0.20695 | 0.866366 | 9.754835 | -4.96371 | 3.11E-06 | 1.10E-05 | 4.007728 |
| HYAL1 | -0.23356 | 0.850536 | 9.564505 | -4.96322 | 3.12E-06 | 1.10E-05 | 4.005793 |
| PDZD7 | -0.1818 | 0.881604 | 5.509231 | -4.96164 | 3.14E-06 | 1.10E-05 | 3.99961 |
| ZNF76 | -0.1487 | 0.902066 | 7.015165 | -4.96022 | 3.16E-06 | 1.11E-05 | 3.994064 |
| GLYAT | -0.37658 | 0.770259 | 10.47747 | -4.95833 | 3.18E-06 | 1.12E-05 | 3.986665 |
| FN1 | 0.341203 | 1.266812 | 6.082857 | 4.957904 | 3.19E-06 | 1.12E-05 | 3.984975 |
| RORA | -0.33043 | 0.795297 | 6.367033 | -4.95787 | 3.19E-06 | 1.12E-05 | 3.984848 |
| TRA2A | 0.278589 | 1.213008 | 8.184286 | 4.957864 | 3.19E-06 | 1.12E-05 | 3.984819 |
| PECAM1 | 0.351068 | 1.275504 | 6.861648 | 4.95626 | 3.21E-06 | 1.13E-05 | 3.978536 |
| SMPDL3B | 0.211174 | 1.15763 | 5.807253 | 4.955369 | 3.22E-06 | 1.13E-05 | 3.975049 |
| NUP210 | -0.12645 | 0.91608 | 5.677033 | -4.95485 | 3.23E-06 | 1.13E-05 | 3.973035 |
| FKBP1A | 0.212048 | 1.158332 | 6.337033 | 4.954777 | 3.23E-06 | 1.13E-05 | 3.972729 |
| IL1RAPL2 | -0.19029 | 0.87643 | 5.072857 | -4.95475 | 3.23E-06 | 1.13E-05 | 3.972607 |
| PRPF19 | 0.21158 | 1.157955 | 7.765714 | 4.953506 | 3.24E-06 | 1.14E-05 | 3.967754 |
| LUZP4 | -0.15135 | 0.900406 | 5.844286 | -4.95076 | 3.28E-06 | 1.15E-05 | 3.957016 |
| BIN3-IT1 | -0.17646 | 0.884869 | 4.891868 | -4.95048 | 3.28E-06 | 1.15E-05 | 3.955922 |
| ABCG2 | -0.45269 | 0.730679 | 5.975055 | -4.94968 | 3.29E-06 | 1.15E-05 | 3.952772 |
| TDRD7 | 0.279778 | 1.214008 | 6.558352 | 4.949468 | 3.30E-06 | 1.15E-05 | 3.951946 |
| UFD1L | 0.241092 | 1.181887 | 7.908352 | 4.949046 | 3.30E-06 | 1.15E-05 | 3.950294 |
| ESRRA | -0.18285 | 0.880964 | 6.391538 | -4.94899 | 3.30E-06 | 1.15E-05 | 3.950065 |
| GALNT10 | 0.187063 | 1.138444 | 6.13033 | 4.947659 | 3.32E-06 | 1.16E-05 | 3.944867 |
| PLS3 | 0.212092 | 1.158367 | 9.909011 | 4.947583 | 3.32E-06 | 1.16E-05 | 3.944571 |
| CHP2 | -0.68058 | 0.623912 | 6.427143 | -4.94728 | 3.33E-06 | 1.16E-05 | 3.943371 |
| SLC12A2 | -0.20882 | 0.865244 | 7.00978 | -4.94635 | 3.34E-06 | 1.16E-05 | 3.939738 |
| APOB | -0.18001 | 0.882694 | 4.25033 | -4.9462 | 3.34E-06 | 1.17E-05 | 3.939166 |
| TFPI2 | 0.780314 | 1.717505 | 5.44 | 4.945037 | 3.36E-06 | 1.17E-05 | 3.934611 |
| MX1 | 0.718676 | 1.645671 | 7.085824 | 4.944746 | 3.36E-06 | 1.17E-05 | 3.93347 |
| GJB5 | -0.18921 | 0.877084 | 4.411868 | -4.94315 | 3.38E-06 | 1.18E-05 | 3.927223 |
| GADD45G | -0.44532 | 0.73442 | 5.32022 | -4.94284 | 3.39E-06 | 1.18E-05 | 3.926007 |
| DPY19L4 | 0.25771 | 1.19558 | 6.514396 | 4.942013 | 3.40E-06 | 1.18E-05 | 3.922782 |
| GLRB | 0.203531 | 1.151514 | 4.12956 | 4.941973 | 3.40E-06 | 1.18E-05 | 3.922629 |
| CST6 | 0.332889 | 1.259533 | 4.474615 | 4.94079 | 3.42E-06 | 1.19E-05 | 3.918004 |
| IZUMO4 | -0.19295 | 0.874817 | 5.501978 | -4.94053 | 3.42E-06 | 1.19E-05 | 3.916999 |
| MAPK10 | -0.17109 | 0.88817 | 6.446264 | -4.94027 | 3.42E-06 | 1.19E-05 | 3.915966 |
| STMN1 | 0.196039 | 1.145549 | 6.358681 | 4.939485 | 3.43E-06 | 1.19E-05 | 3.9129 |
| THG1L | 0.199614 | 1.148391 | 6.191319 | 4.939082 | 3.44E-06 | 1.19E-05 | 3.911326 |
| PLEKHM1 | 0.192643 | 1.142855 | 7.344176 | 4.937448 | 3.46E-06 | 1.20E-05 | 3.90494 |
| SEC23A | 0.276865 | 1.211559 | 7.115824 | 4.936854 | 3.47E-06 | 1.20E-05 | 3.902619 |
| KRT75 | -0.17979 | 0.88283 | 5.01044 | -4.93543 | 3.49E-06 | 1.21E-05 | 3.897051 |
| CAMK2N1 | -0.21617 | 0.860845 | 9.735275 | -4.93508 | 3.50E-06 | 1.21E-05 | 3.895686 |
| CYP2A6 | -0.16176 | 0.893935 | 5.690879 | -4.93437 | 3.51E-06 | 1.21E-05 | 3.892923 |
| SOX21 | -0.20904 | 0.865111 | 4.90967 | -4.93415 | 3.51E-06 | 1.21E-05 | 3.892055 |

| | | | | | | | |
|---|---|---|---|---|---|---|---|
| TLR6 | -0.13559 | 0.910298 | 4.251429 | -4.93359 | 3.52E-06 | 1.22E-05 | 3.889862 |
| IRF8 | 0.526502 | 1.440433 | 6.832967 | 4.932718 | 3.53E-06 | 1.22E-05 | 3.886459 |
| FNDC8 | -0.18589 | 0.879107 | 4.763077 | -4.93259 | 3.53E-06 | 1.22E-05 | 3.885942 |
| RPL13A | 0.15356 | 1.112311 | 13.66615 | 4.932167 | 3.54E-06 | 1.22E-05 | 3.884308 |
| PTGES3 | 0.182386 | 1.134759 | 10.98758 | 4.931712 | 3.54E-06 | 1.22E-05 | 3.882532 |
| MEAF6 | 0.215126 | 1.160805 | 6.727033 | 4.931371 | 3.55E-06 | 1.23E-05 | 3.881199 |
| ADAM15 | -0.17343 | 0.886735 | 6.08011 | -4.92983 | 3.57E-06 | 1.23E-05 | 3.875163 |
| PRAMEF12 | -0.19031 | 0.876415 | 4.701758 | -4.9244 | 3.65E-06 | 1.26E-05 | 3.853977 |
| MC4R | -0.14046 | 0.907231 | 4.27989 | -4.92419 | 3.65E-06 | 1.26E-05 | 3.853181 |
| FUT1 | -0.17474 | 0.885925 | 5.18011 | -4.92418 | 3.65E-06 | 1.26E-05 | 3.853117 |
| ADAM11 | -0.20113 | 0.869869 | 5.51967 | -4.92272 | 3.68E-06 | 1.27E-05 | 3.847416 |
| CCNA2 | 0.288367 | 1.221257 | 4.419121 | 4.922487 | 3.68E-06 | 1.27E-05 | 3.846524 |
| NGLY1 | 0.15101 | 1.110346 | 8.17011 | 4.920417 | 3.71E-06 | 1.28E-05 | 3.83845 |
| DHX9 | 0.187643 | 1.138901 | 6.226703 | 4.920317 | 3.71E-06 | 1.28E-05 | 3.838058 |
| EGR3 | -0.47874 | 0.717605 | 4.42 | -4.9198 | 3.72E-06 | 1.28E-05 | 3.836029 |
| PSG11 | -0.21491 | 0.861598 | 4.682637 | -4.91958 | 3.72E-06 | 1.28E-05 | 3.835195 |
| ABHD11 | 0.239034 | 1.180202 | 7.074725 | 4.919291 | 3.73E-06 | 1.28E-05 | 3.834058 |
| GJA8 | -0.19346 | 0.874504 | 5.508462 | -4.9186 | 3.74E-06 | 1.28E-05 | 3.831344 |
| GJA3 | -0.17777 | 0.88407 | 4.782527 | -4.91725 | 3.76E-06 | 1.29E-05 | 3.826086 |
| COL15A1 | 0.55615 | 1.47034 | 5.351758 | 4.915988 | 3.78E-06 | 1.30E-05 | 3.82118 |
| EPB41L2 | 0.24157 | 1.182279 | 6.10011 | 4.915277 | 3.79E-06 | 1.30E-05 | 3.818407 |
| GYPB | -0.16385 | 0.89264 | 3.835714 | -4.91389 | 3.81E-06 | 1.31E-05 | 3.813014 |
| RABGGTB | -0.25157 | 0.839982 | 8.094945 | -4.91377 | 3.81E-06 | 1.31E-05 | 3.81253 |
| CLDN9 | -0.18259 | 0.88112 | 5.063187 | -4.91317 | 3.82E-06 | 1.31E-05 | 3.810192 |
| R3HDM1 | 0.168976 | 1.12426 | 7.607473 | 4.912959 | 3.82E-06 | 1.31E-05 | 3.809376 |
| WDR11 | 0.260411 | 1.19782 | 6.997253 | 4.910599 | 3.86E-06 | 1.32E-05 | 3.800178 |
| PREPL | 0.184077 | 1.13609 | 8.114505 | 4.910556 | 3.86E-06 | 1.32E-05 | 3.800011 |
| KCNE2 | -0.16751 | 0.890377 | 5.479121 | -4.91051 | 3.86E-06 | 1.32E-05 | 3.799818 |
| BMP5 | -0.25547 | 0.837715 | 5.147582 | -4.9102 | 3.87E-06 | 1.32E-05 | 3.798641 |
| NDUFA2 | -0.13235 | 0.912346 | 9.728901 | -4.90894 | 3.89E-06 | 1.33E-05 | 3.793702 |
| SLC16A7 | -0.37571 | 0.770729 | 5.777473 | -4.90721 | 3.91E-06 | 1.34E-05 | 3.78698 |
| FUT7 | -0.14978 | 0.901386 | 4.495714 | -4.90715 | 3.92E-06 | 1.34E-05 | 3.786741 |
| TBC1D10B | -0.16427 | 0.89238 | 6.826593 | -4.90626 | 3.93E-06 | 1.34E-05 | 3.783285 |
| RHD | -0.1807 | 0.882275 | 4.909121 | -4.90598 | 3.93E-06 | 1.34E-05 | 3.78218 |
| POLE2 | 0.301459 | 1.23239 | 4.964725 | 4.904352 | 3.96E-06 | 1.35E-05 | 3.775854 |
| CINP | 0.253589 | 1.19217 | 7.136923 | 4.903995 | 3.97E-06 | 1.35E-05 | 3.774465 |
| CDKN1B | 0.295498 | 1.227308 | 8.093516 | 4.903867 | 3.97E-06 | 1.35E-05 | 3.773966 |
| MT4 | -0.4312 | 0.741643 | 9.622637 | -4.90371 | 3.97E-06 | 1.35E-05 | 3.773366 |
| NME5 | 0.2423 | 1.182877 | 8.743297 | 4.903428 | 3.97E-06 | 1.35E-05 | 3.772255 |
| TNK1 | -0.17924 | 0.88317 | 6.965714 | -4.90227 | 3.99E-06 | 1.36E-05 | 3.767761 |
| HAX1 | -0.16101 | 0.894396 | 9.094725 | -4.90072 | 4.02E-06 | 1.37E-05 | 3.761702 |
| CA1 | -0.19234 | 0.875186 | 5.86967 | -4.90061 | 4.02E-06 | 1.37E-05 | 3.761303 |
| CTGF | -0.74868 | 0.595147 | 8.719121 | -4.90046 | 4.02E-06 | 1.37E-05 | 3.760709 |
| NETO2 | 0.221696 | 1.166103 | 4.265934 | 4.899957 | 4.03E-06 | 1.37E-05 | 3.758749 |
| MPC1 | -0.26604 | 0.8316 | 10.6367 | -4.89974 | 4.03E-06 | 1.37E-05 | 3.757912 |
| RWDD2A | 0.181092 | 1.133742 | 5.738681 | 4.899174 | 4.04E-06 | 1.37E-05 | 3.755702 |
| GABRA2 | -0.23951 | 0.847032 | 4.143516 | -4.8982 | 4.06E-06 | 1.38E-05 | 3.751919 |

| | | | | | | | |
|---|---|---|---|---|---|---|---|
| ARHGEF1 | -0.18364 | 0.880477 | 6.525275 | -4.8976 | 4.07E-06 | 1.38E-05 | 3.749576 |
| ACAP1 | -0.19036 | 0.876389 | 5.70978 | -4.89645 | 4.09E-06 | 1.39E-05 | 3.74511 |
| SEMA7A | -0.19162 | 0.875623 | 5.322418 | -4.89484 | 4.12E-06 | 1.40E-05 | 3.738829 |
| CNIH4 | 0.243879 | 1.184172 | 7.665165 | 4.894523 | 4.12E-06 | 1.40E-05 | 3.737615 |
| CDKN3 | 0.374324 | 1.296232 | 4.79967 | 4.8928 | 4.15E-06 | 1.41E-05 | 3.730914 |
| TRIM29 | -0.16906 | 0.889423 | 4.888791 | -4.89197 | 4.16E-06 | 1.41E-05 | 3.727694 |
| PPP1CB | 0.187396 | 1.138707 | 8.165495 | 4.891576 | 4.17E-06 | 1.41E-05 | 3.726156 |
| SIL1 | 0.183271 | 1.135455 | 7.667802 | 4.891129 | 4.18E-06 | 1.41E-05 | 3.724421 |
| L2HGDH | -0.25976 | 0.835228 | 5.562418 | -4.89003 | 4.20E-06 | 1.42E-05 | 3.720158 |
| RPLP2 | 0.269541 | 1.205424 | 11.44264 | 4.888744 | 4.22E-06 | 1.43E-05 | 3.715151 |
| CHAC1 | -0.24503 | 0.843796 | 5.702308 | -4.88774 | 4.24E-06 | 1.43E-05 | 3.711235 |
| ALG5 | 0.196386 | 1.145825 | 9.514505 | 4.887354 | 4.24E-06 | 1.43E-05 | 3.709752 |
| ZNF688 | -0.14964 | 0.901474 | 5.832088 | -4.88671 | 4.25E-06 | 1.44E-05 | 3.707249 |
| DENND5A | 0.195087 | 1.144793 | 7.775385 | 4.885243 | 4.28E-06 | 1.44E-05 | 3.701553 |
| METAP2 | 0.160739 | 1.11786 | 6.432747 | 4.883207 | 4.31E-06 | 1.46E-05 | 3.693647 |
| MDK | 0.256754 | 1.194787 | 5.580879 | 4.883133 | 4.31E-06 | 1.46E-05 | 3.693357 |
| ADAR | 0.201217 | 1.149668 | 9.744286 | 4.882019 | 4.33E-06 | 1.46E-05 | 3.689031 |
| PUS7 | 0.240251 | 1.181198 | 6.37011 | 4.88184 | 4.34E-06 | 1.46E-05 | 3.688338 |
| SLC1A7 | -0.14117 | 0.906784 | 4.797582 | -4.87993 | 4.37E-06 | 1.47E-05 | 3.68094 |
| SNCAIP | -0.1992 | 0.871032 | 4.807363 | -4.87676 | 4.43E-06 | 1.49E-05 | 3.668618 |
| ADGRB2 | -0.18005 | 0.88267 | 4.638571 | -4.87596 | 4.44E-06 | 1.50E-05 | 3.665503 |
| PTPRB | -0.15825 | 0.896114 | 5.150659 | -4.87456 | 4.47E-06 | 1.50E-05 | 3.660076 |
| CACNA1I | -0.15704 | 0.896864 | 5.005604 | -4.87422 | 4.47E-06 | 1.51E-05 | 3.658761 |
| AGMAT | -0.36952 | 0.774039 | 11.41879 | -4.87278 | 4.50E-06 | 1.51E-05 | 3.653174 |
| PDIA2 | -0.17439 | 0.886141 | 4.946154 | -4.87274 | 4.50E-06 | 1.51E-05 | 3.653026 |
| MRPL15 | 0.197918 | 1.147042 | 9.314176 | 4.868447 | 4.58E-06 | 1.54E-05 | 3.636385 |
| THOC7 | 0.2093 | 1.156127 | 8.651978 | 4.868343 | 4.58E-06 | 1.54E-05 | 3.63598 |
| MYBL2 | -0.17698 | 0.884555 | 5.628571 | -4.86783 | 4.59E-06 | 1.54E-05 | 3.633986 |
| KHDRBS3 | 0.246338 | 1.186193 | 6.367033 | 4.867414 | 4.60E-06 | 1.54E-05 | 3.63238 |
| FOXH1 | -0.21518 | 0.861437 | 4.72033 | -4.86665 | 4.61E-06 | 1.55E-05 | 3.629413 |
| FUS | 0.231662 | 1.174187 | 7.279341 | 4.865591 | 4.63E-06 | 1.55E-05 | 3.625318 |
| CHCHD3 | 0.202169 | 1.150427 | 7.687582 | 4.864669 | 4.65E-06 | 1.56E-05 | 3.621745 |
| TNR | -0.16215 | 0.893689 | 4.482857 | -4.86242 | 4.69E-06 | 1.57E-05 | 3.613025 |
| FAM21A | 0.288295 | 1.221196 | 9.615824 | 4.859364 | 4.75E-06 | 1.59E-05 | 3.6012 |
| KIR2DS4 | -0.1842 | 0.880135 | 5.01978 | -4.85932 | 4.75E-06 | 1.59E-05 | 3.60101 |
| PIWIL1 | -0.15531 | 0.89794 | 3.734286 | -4.85926 | 4.75E-06 | 1.59E-05 | 3.600814 |
| HTRA1 | -0.47034 | 0.721793 | 8.841978 | -4.85921 | 4.75E-06 | 1.59E-05 | 3.600602 |
| EBNA1BP2 | 0.172918 | 1.127336 | 7.596813 | 4.858796 | 4.76E-06 | 1.59E-05 | 3.598999 |
| TESK1 | -0.19802 | 0.871744 | 7.220989 | -4.85815 | 4.77E-06 | 1.60E-05 | 3.596504 |
| ACACB | -0.23012 | 0.852566 | 6.930989 | -4.85781 | 4.78E-06 | 1.60E-05 | 3.59519 |
| ACKR2 | -0.16907 | 0.889417 | 5.298352 | -4.85763 | 4.78E-06 | 1.60E-05 | 3.594472 |
| PGAP3 | -0.14699 | 0.903133 | 7.392747 | -4.85588 | 4.82E-06 | 1.61E-05 | 3.587706 |
| BHLHB9 | 0.294464 | 1.226429 | 5.526484 | 4.853189 | 4.87E-06 | 1.63E-05 | 3.577302 |
| ARMC1 | 0.220734 | 1.165327 | 7.782198 | 4.853137 | 4.87E-06 | 1.63E-05 | 3.577101 |
| LIMS2 | -0.19175 | 0.875541 | 5.806264 | -4.85302 | 4.87E-06 | 1.63E-05 | 3.576635 |
| MEX3C | 0.172242 | 1.126808 | 5.246044 | 4.852362 | 4.88E-06 | 1.63E-05 | 3.574102 |
| SLC1A2 | -0.1372 | 0.90928 | 4.568022 | -4.85228 | 4.89E-06 | 1.63E-05 | 3.573788 |

| | | | | | | | |
|---|---|---|---|---|---|---|---|
| LRRC2 | -0.22249 | 0.857083 | 4.077802 | -4.85078 | 4.92E-06 | 1.64E-05 | 3.567964 |
| UBA3 | 0.190778 | 1.141379 | 7.469341 | 4.850712 | 4.92E-06 | 1.64E-05 | 3.567718 |
| CCL17 | -0.18867 | 0.877416 | 4.946703 | -4.84903 | 4.95E-06 | 1.65E-05 | 3.561213 |
| BHLHE41 | 0.239937 | 1.180941 | 5.015824 | 4.846025 | 5.01E-06 | 1.67E-05 | 3.549596 |
| TEX10 | 0.154309 | 1.112889 | 6.67022 | 4.844553 | 5.04E-06 | 1.68E-05 | 3.543906 |
| SUPV3L1 | -0.15358 | 0.899017 | 7.172967 | -4.84422 | 5.05E-06 | 1.68E-05 | 3.542616 |
| NT5C2 | 0.189913 | 1.140695 | 6.88 | 4.843725 | 5.06E-06 | 1.68E-05 | 3.540706 |
| FDXR | 0.259179 | 1.196797 | 5.451209 | 4.843236 | 5.07E-06 | 1.68E-05 | 3.538819 |
| GP9 | -0.16913 | 0.889379 | 5.825495 | -4.84294 | 5.07E-06 | 1.69E-05 | 3.537662 |
| CLCN7 | -0.14344 | 0.905355 | 7.114066 | -4.84284 | 5.07E-06 | 1.69E-05 | 3.537281 |
| SERPINI1 | 0.455135 | 1.370911 | 8.766154 | 4.841231 | 5.11E-06 | 1.70E-05 | 3.531071 |
| EBLN2 | -0.19416 | 0.874079 | 5.546593 | -4.83946 | 5.14E-06 | 1.71E-05 | 3.524222 |
| STAB2 | -0.20798 | 0.865748 | 4.738022 | -4.83908 | 5.15E-06 | 1.71E-05 | 3.522746 |
| NCAPG2 | 0.178222 | 1.131489 | 5.298132 | 4.838788 | 5.16E-06 | 1.71E-05 | 3.521637 |
| PLXNB3 | -0.18234 | 0.881271 | 5.564396 | -4.83865 | 5.16E-06 | 1.71E-05 | 3.521109 |
| BHMT | -0.48127 | 0.716349 | 12.41505 | -4.83817 | 5.17E-06 | 1.71E-05 | 3.519236 |
| HMGB2 | 0.369657 | 1.292046 | 7.907363 | 4.837957 | 5.18E-06 | 1.71E-05 | 3.518427 |
| MAML1 | 0.164899 | 1.121087 | 6.801978 | 4.835779 | 5.22E-06 | 1.73E-05 | 3.510015 |
| TM2D3 | 0.173947 | 1.128141 | 8.703626 | 4.835032 | 5.24E-06 | 1.73E-05 | 3.507132 |
| OR7C2 | -0.18784 | 0.877919 | 5.404286 | -4.83421 | 5.25E-06 | 1.74E-05 | 3.503965 |
| CAPNS1 | 0.180343 | 1.133153 | 10.25462 | 4.834071 | 5.26E-06 | 1.74E-05 | 3.503423 |
| BTG2 | -0.54086 | 0.687359 | 7.203626 | -4.83235 | 5.29E-06 | 1.75E-05 | 3.496794 |
| PAH | -0.58713 | 0.665668 | 11.90901 | -4.83026 | 5.34E-06 | 1.76E-05 | 3.488702 |
| PBX2 | -0.16516 | 0.891827 | 5.605934 | -4.82933 | 5.36E-06 | 1.77E-05 | 3.485124 |
| CEACAM4 | -0.1507 | 0.900816 | 5.924176 | -4.82808 | 5.38E-06 | 1.78E-05 | 3.480322 |
| AQP5 | -0.1865 | 0.878733 | 5.255165 | -4.82789 | 5.39E-06 | 1.78E-05 | 3.47957 |
| ACYP2 | 0.220464 | 1.165108 | 6.52989 | 4.827491 | 5.40E-06 | 1.78E-05 | 3.478035 |
| HDHD3 | -0.25461 | 0.838212 | 8.056484 | -4.8272 | 5.40E-06 | 1.78E-05 | 3.476904 |
| ASCC1 | 0.195546 | 1.145157 | 6.736264 | 4.826425 | 5.42E-06 | 1.79E-05 | 3.473925 |
| MYCL | -0.20181 | 0.869461 | 5.658901 | -4.82602 | 5.43E-06 | 1.79E-05 | 3.472346 |
| MAT1A | -0.18774 | 0.877981 | 5.738901 | -4.82593 | 5.43E-06 | 1.79E-05 | 3.472005 |
| SMYD5 | -0.17924 | 0.88317 | 6.645714 | -4.82522 | 5.45E-06 | 1.79E-05 | 3.469296 |
| OTOR | -0.2006 | 0.870186 | 4.543626 | -4.82399 | 5.47E-06 | 1.80E-05 | 3.464546 |
| CNTN2 | -0.14575 | 0.90391 | 4.939231 | -4.82394 | 5.48E-06 | 1.80E-05 | 3.464337 |
| LIN37 | 0.17714 | 1.13064 | 5.198901 | 4.823644 | 5.48E-06 | 1.80E-05 | 3.463203 |
| ACVR2B-A! | -0.20493 | 0.867579 | 4.316703 | -4.82304 | 5.49E-06 | 1.81E-05 | 3.460874 |
| ATF1 | 0.204295 | 1.152123 | 6.784286 | 4.819031 | 5.58E-06 | 1.83E-05 | 3.445425 |
| PAMR1 | -0.29723 | 0.813815 | 5.407802 | -4.81871 | 5.59E-06 | 1.84E-05 | 3.444181 |
| ACTR1A | 0.185594 | 1.137285 | 7.563516 | 4.818653 | 5.59E-06 | 1.84E-05 | 3.44397 |
| PICALM | 0.191923 | 1.142285 | 8.331429 | 4.817345 | 5.62E-06 | 1.84E-05 | 3.438931 |
| GLUL | 0.279575 | 1.213837 | 8.319121 | 4.81631 | 5.65E-06 | 1.85E-05 | 3.434946 |
| RHOB | -0.50842 | 0.702994 | 9.49989 | -4.81615 | 5.65E-06 | 1.85E-05 | 3.434344 |
| CYP4F12 | -0.31466 | 0.804042 | 6.624835 | -4.81491 | 5.68E-06 | 1.86E-05 | 3.429543 |
| WWP1 | 0.237787 | 1.179183 | 7.228022 | 4.81402 | 5.70E-06 | 1.87E-05 | 3.426128 |
| LPP | 0.226662 | 1.170124 | 7.231868 | 4.81216 | 5.74E-06 | 1.88E-05 | 3.418967 |
| CPZ | -0.18003 | 0.882685 | 5.32967 | -4.81148 | 5.76E-06 | 1.88E-05 | 3.416354 |
| ADGRF5 | -0.54858 | 0.683691 | 8.622418 | -4.811 | 5.77E-06 | 1.89E-05 | 3.414519 |

| | | | | | | | |
|---|---|---|---|---|---|---|---|
| CASP3 | 0.27914 | 1.213471 | 6.299341 | 4.810856 | 5.77E-06 | 1.89E-05 | 3.413948 |
| BRD3 | 0.228855 | 1.171905 | 7.631648 | 4.8106 | 5.78E-06 | 1.89E-05 | 3.412963 |
| CTSF | -0.24296 | 0.845012 | 8.106813 | -4.80968 | 5.80E-06 | 1.90E-05 | 3.409419 |
| IFI44 | 0.762783 | 1.69676 | 5.582418 | 4.809373 | 5.80E-06 | 1.90E-05 | 3.408239 |
| COL5A3 | -0.1391 | 0.908088 | 5.551868 | -4.80785 | 5.84E-06 | 1.91E-05 | 3.402382 |
| KCNJ6 | -0.16243 | 0.893522 | 4.980549 | -4.80769 | 5.84E-06 | 1.91E-05 | 3.401765 |
| RNASE6 | 0.607043 | 1.523135 | 5.867143 | 4.804788 | 5.91E-06 | 1.93E-05 | 3.390602 |
| GPSM3 | -0.17466 | 0.885975 | 6.153846 | -4.80422 | 5.93E-06 | 1.93E-05 | 3.388433 |
| GRPEL1 | -0.19771 | 0.87193 | 8.805055 | -4.8041 | 5.93E-06 | 1.93E-05 | 3.387955 |
| TNNC2 | -0.21517 | 0.861445 | 5.390989 | -4.80363 | 5.94E-06 | 1.94E-05 | 3.386133 |
| EPHB3 | -0.15519 | 0.898015 | 5.61978 | -4.8036 | 5.94E-06 | 1.94E-05 | 3.386046 |
| NT5M | -0.17918 | 0.883203 | 5.108132 | -4.80332 | 5.95E-06 | 1.94E-05 | 3.384943 |
| NFKBIE | 0.367662 | 1.29026 | 6.056593 | 4.802744 | 5.96E-06 | 1.94E-05 | 3.382742 |
| ARMC6 | -0.15445 | 0.898475 | 6.183407 | -4.80177 | 5.98E-06 | 1.95E-05 | 3.37899 |
| SEMA6D | -0.28405 | 0.82128 | 5.157143 | -4.7986 | 6.06E-06 | 1.97E-05 | 3.36683 |
| GM2A | 0.182174 | 1.134592 | 6.807912 | 4.797749 | 6.08E-06 | 1.98E-05 | 3.363543 |
| CXorf36 | -0.16528 | 0.891758 | 5.530879 | -4.79758 | 6.08E-06 | 1.98E-05 | 3.362893 |
| BMP3 | -0.15265 | 0.899598 | 4.375385 | -4.79677 | 6.10E-06 | 1.98E-05 | 3.359765 |
| NYX | -0.15499 | 0.898141 | 5.059011 | -4.79575 | 6.13E-06 | 1.99E-05 | 3.355881 |
| ANKRD2 | 0.440377 | 1.356959 | 5.594725 | 4.795554 | 6.13E-06 | 1.99E-05 | 3.355112 |
| CUBN | -0.4941 | 0.710004 | 11.3811 | -4.79535 | 6.14E-06 | 1.99E-05 | 3.354323 |
| STUB1 | -0.15327 | 0.899213 | 9.477253 | -4.79386 | 6.18E-06 | 2.00E-05 | 3.348599 |
| TLE1 | 0.196754 | 1.146116 | 5.971209 | 4.793632 | 6.18E-06 | 2.01E-05 | 3.347728 |
| FAM131A | -0.20081 | 0.870064 | 5.964396 | -4.79208 | 6.22E-06 | 2.02E-05 | 3.341784 |
| PSIP1 | 0.23771 | 1.17912 | 7.174505 | 4.791906 | 6.22E-06 | 2.02E-05 | 3.3411 |
| PPIL6 | -0.13963 | 0.907753 | 4.067692 | -4.79168 | 6.23E-06 | 2.02E-05 | 3.340239 |
| PDZD2 | -0.43903 | 0.737628 | 6.486374 | -4.79058 | 6.26E-06 | 2.03E-05 | 3.335992 |
| USP46 | 0.173918 | 1.128118 | 5.532308 | 4.789268 | 6.29E-06 | 2.04E-05 | 3.330975 |
| RUNX1-IT1 | -0.15974 | 0.895184 | 5.192527 | -4.78839 | 6.31E-06 | 2.04E-05 | 3.327613 |
| USO1 | 0.179623 | 1.132588 | 8.311868 | 4.788216 | 6.32E-06 | 2.04E-05 | 3.326937 |
| TMEM248 | 0.149841 | 1.109447 | 8.536923 | 4.788215 | 6.32E-06 | 2.04E-05 | 3.326934 |
| LPAL2 | -0.23747 | 0.848232 | 4.716484 | -4.7873 | 6.34E-06 | 2.05E-05 | 3.323409 |
| RAC2 | 0.397169 | 1.316921 | 6.479011 | 4.786288 | 6.37E-06 | 2.06E-05 | 3.319539 |
| CWF19L1 | -0.15482 | 0.898244 | 6.476484 | -4.78482 | 6.40E-06 | 2.07E-05 | 3.31389 |
| PRG2 | -0.18056 | 0.882363 | 4.585714 | -4.78364 | 6.43E-06 | 2.08E-05 | 3.309392 |
| FABP2 | -0.1756 | 0.885397 | 4.670989 | -4.78261 | 6.46E-06 | 2.09E-05 | 3.305422 |
| AGRN | 0.187246 | 1.138588 | 6.788681 | 4.782527 | 6.46E-06 | 2.09E-05 | 3.305112 |
| PYY | -0.20902 | 0.865125 | 5.870769 | -4.78106 | 6.50E-06 | 2.10E-05 | 3.299473 |
| IQCA1 | 0.232087 | 1.174533 | 5.308681 | 4.780712 | 6.51E-06 | 2.10E-05 | 3.298151 |
| MYO5C | 0.352333 | 1.276624 | 7.699231 | 4.779981 | 6.53E-06 | 2.10E-05 | 3.295349 |
| CACNG3 | -0.18733 | 0.87823 | 4.857582 | -4.77983 | 6.53E-06 | 2.10E-05 | 3.294754 |
| DRD2 | -0.14898 | 0.90189 | 5.412418 | -4.7781 | 6.58E-06 | 2.12E-05 | 3.288147 |
| C1orf115 | -0.22362 | 0.856412 | 9.146374 | -4.77636 | 6.62E-06 | 2.13E-05 | 3.281459 |
| LAMP1 | 0.156324 | 1.114444 | 10.78187 | 4.775957 | 6.63E-06 | 2.13E-05 | 3.279924 |
| PLEKHG6 | -0.16882 | 0.889572 | 4.69978 | -4.77307 | 6.71E-06 | 2.16E-05 | 3.268853 |
| BMX | -0.17086 | 0.88831 | 4.266593 | -4.77298 | 6.71E-06 | 2.16E-05 | 3.268531 |
| DHRS7B | 0.215072 | 1.160762 | 8.024615 | 4.772601 | 6.72E-06 | 2.16E-05 | 3.267069 |

| | | | | | | | |
|---|---|---|---|---|---|---|---|
| CCNC | 0.161705 | 1.118609 | 9.786703 | 4.77196 | 6.74E-06 | 2.17E-05 | 3.264613 |
| AIMP1 | 0.150222 | 1.10974 | 7.974286 | 4.771888 | 6.74E-06 | 2.17E-05 | 3.264337 |
| TMEM184 | 0.179609 | 1.132577 | 7.861209 | 4.771616 | 6.75E-06 | 2.17E-05 | 3.263296 |
| LSM8 | 0.177884 | 1.131224 | 6.612747 | 4.771567 | 6.75E-06 | 2.17E-05 | 3.263106 |
| RIN1 | -0.1899 | 0.876667 | 6.110659 | -4.76992 | 6.80E-06 | 2.18E-05 | 3.256811 |
| FOSL1 | -0.3409 | 0.789547 | 4.090769 | -4.7699 | 6.80E-06 | 2.18E-05 | 3.256714 |
| GPR153 | -0.19358 | 0.874433 | 6.923187 | -4.76937 | 6.81E-06 | 2.18E-05 | 3.254693 |
| HSPA6 | -0.75286 | 0.593426 | 6.409011 | -4.76572 | 6.91E-06 | 2.21E-05 | 3.240741 |
| HERC1 | -0.22743 | 0.854155 | 6.483187 | -4.76561 | 6.91E-06 | 2.21E-05 | 3.240293 |
| FZD5 | -0.22919 | 0.853112 | 6.982967 | -4.76509 | 6.93E-06 | 2.22E-05 | 3.2383 |
| TRPM2 | -0.20012 | 0.870478 | 5.755604 | -4.76442 | 6.95E-06 | 2.22E-05 | 3.235764 |
| TSHZ2 | -0.13949 | 0.907841 | 5.344066 | -4.76433 | 6.95E-06 | 2.22E-05 | 3.235409 |
| PTPRO | -0.83802 | 0.559411 | 6.114725 | -4.76409 | 6.95E-06 | 2.22E-05 | 3.2345 |
| KPNA4 | 0.165623 | 1.12165 | 6.004945 | 4.76103 | 7.04E-06 | 2.25E-05 | 3.22278 |
| GAPDHS | -0.15953 | 0.895319 | 3.992418 | -4.761 | 7.04E-06 | 2.25E-05 | 3.222672 |
| C12orf49 | 0.203449 | 1.151448 | 7.905824 | 4.760115 | 7.07E-06 | 2.26E-05 | 3.219281 |
| TDRD1 | -0.15614 | 0.897426 | 3.936703 | -4.75976 | 7.08E-06 | 2.26E-05 | 3.21794 |
| ZNF189 | 0.25558 | 1.193815 | 7.667473 | 4.759661 | 7.08E-06 | 2.26E-05 | 3.217543 |
| TEX11 | -0.17608 | 0.885106 | 4.849451 | -4.75877 | 7.10E-06 | 2.27E-05 | 3.214152 |
| TBX21 | -0.16596 | 0.891334 | 4.97967 | -4.75754 | 7.14E-06 | 2.28E-05 | 3.209431 |
| DDX25 | -0.25233 | 0.83954 | 5.54044 | -4.75523 | 7.20E-06 | 2.30E-05 | 3.200597 |
| MELK | 0.346681 | 1.271632 | 4.892088 | 4.755218 | 7.21E-06 | 2.30E-05 | 3.200559 |
| WDR13 | -0.18104 | 0.882065 | 8.873516 | -4.75499 | 7.21E-06 | 2.30E-05 | 3.199697 |
| APOC1 | 0.890527 | 1.853853 | 8.174066 | 4.753991 | 7.24E-06 | 2.31E-05 | 3.19587 |
| PPA1 | 0.257077 | 1.195055 | 10.8156 | 4.752504 | 7.28E-06 | 2.32E-05 | 3.190188 |
| C11orf73 | 0.202333 | 1.150558 | 4.445055 | 4.751488 | 7.31E-06 | 2.33E-05 | 3.186308 |
| SLC6A20 | 0.371048 | 1.293292 | 4.540659 | 4.751052 | 7.33E-06 | 2.33E-05 | 3.184641 |
| CAPN10 | -0.17024 | 0.888694 | 6.204945 | -4.74923 | 7.38E-06 | 2.35E-05 | 3.177667 |
| LINC00472 | -0.30485 | 0.809529 | 4.981209 | -4.74813 | 7.41E-06 | 2.36E-05 | 3.173499 |
| FNBP1L | 0.329386 | 1.256479 | 8.315275 | 4.747842 | 7.42E-06 | 2.36E-05 | 3.17238 |
| PPP1R1A | 0.684816 | 1.607497 | 8.065385 | 4.746712 | 7.45E-06 | 2.37E-05 | 3.168066 |
| SPTLC1 | 0.156918 | 1.114903 | 7.998901 | 4.74456 | 7.52E-06 | 2.39E-05 | 3.159853 |
| TIPRL | 0.197502 | 1.146711 | 6.655275 | 4.744202 | 7.53E-06 | 2.39E-05 | 3.158488 |
| SCN10A | -0.16949 | 0.889158 | 4.979231 | -4.74356 | 7.55E-06 | 2.40E-05 | 3.156035 |
| SERINC3 | 0.164256 | 1.120588 | 8.622747 | 4.742674 | 7.57E-06 | 2.40E-05 | 3.152656 |
| AASS | -0.28803 | 0.81902 | 6.186264 | -4.7423 | 7.59E-06 | 2.41E-05 | 3.151229 |
| RBX1 | 0.162981 | 1.119598 | 9.694725 | 4.74128 | 7.62E-06 | 2.41E-05 | 3.147337 |
| SNRNP70 | 0.246251 | 1.186121 | 7.553077 | 4.740126 | 7.65E-06 | 2.42E-05 | 3.142935 |
| SLC26A1 | -0.15464 | 0.898355 | 4.584615 | -4.73884 | 7.69E-06 | 2.44E-05 | 3.138023 |
| AGO1 | -0.18999 | 0.876615 | 6.726703 | -4.73608 | 7.77E-06 | 2.46E-05 | 3.127506 |
| CDC7 | 0.239333 | 1.180447 | 4.418352 | 4.735665 | 7.79E-06 | 2.46E-05 | 3.125924 |
| PGD | 0.255589 | 1.193823 | 7.717912 | 4.734775 | 7.82E-06 | 2.47E-05 | 3.122531 |
| CRYBB2 | -0.19937 | 0.870933 | 5.15989 | -4.73428 | 7.83E-06 | 2.48E-05 | 3.120628 |
| TMEM131 | 0.192715 | 1.142913 | 6.417473 | 4.733754 | 7.85E-06 | 2.48E-05 | 3.118638 |
| LY6G6C | -0.17878 | 0.883448 | 5.246374 | -4.73153 | 7.92E-06 | 2.50E-05 | 3.110156 |
| SQSTM1 | 0.221208 | 1.165709 | 7.563736 | 4.731049 | 7.93E-06 | 2.51E-05 | 3.108332 |
| GLRX2 | 0.270184 | 1.205961 | 7.101868 | 4.727819 | 8.03E-06 | 2.54E-05 | 3.096027 |

| | | | | | | | |
|---|---|---|---|---|---|---|---|
| KIAA0040 | -0.16507 | 0.891887 | 5.71033 | -4.72759 | 8.04E-06 | 2.54E-05 | 3.095139 |
| PRPF38B | 0.192575 | 1.142802 | 6.171099 | 4.726144 | 8.09E-06 | 2.55E-05 | 3.089648 |
| GBP1 | 0.501575 | 1.415758 | 6.318901 | 4.725638 | 8.10E-06 | 2.56E-05 | 3.08772 |
| KCNH4 | -0.1782 | 0.883803 | 4.662747 | -4.72398 | 8.16E-06 | 2.57E-05 | 3.081428 |
| EPHA7 | 0.302937 | 1.233653 | 5.151978 | 4.722453 | 8.21E-06 | 2.59E-05 | 3.075598 |
| UBR4 | 0.413797 | 1.332187 | 9.115495 | 4.722276 | 8.21E-06 | 2.59E-05 | 3.074922 |
| LGI2 | -0.17904 | 0.883288 | 6.294505 | -4.72066 | 8.27E-06 | 2.60E-05 | 3.068775 |
| HSPA14 | 0.20457 | 1.152343 | 6.686813 | 4.719781 | 8.29E-06 | 2.61E-05 | 3.065429 |
| RHOQ | 0.19485 | 1.144605 | 6.454615 | 4.717981 | 8.35E-06 | 2.63E-05 | 3.058583 |
| KLHL20 | -0.18137 | 0.881864 | 5.928571 | -4.71783 | 8.36E-06 | 2.63E-05 | 3.058 |
| MLF2 | -0.18218 | 0.881368 | 8.461648 | -4.71683 | 8.39E-06 | 2.64E-05 | 3.054221 |
| ADRA2B | -0.19971 | 0.870725 | 5.424286 | -4.71539 | 8.44E-06 | 2.65E-05 | 3.048723 |
| GTF2I | 0.245541 | 1.185537 | 5.610769 | 4.715155 | 8.45E-06 | 2.65E-05 | 3.047834 |
| WISP2 | -0.14566 | 0.903968 | 4.493407 | -4.71439 | 8.47E-06 | 2.66E-05 | 3.044927 |
| PMM2 | 0.244058 | 1.184319 | 6.073297 | 4.714034 | 8.49E-06 | 2.66E-05 | 3.043573 |
| MUC5B | -0.14914 | 0.901785 | 4.654725 | -4.71374 | 8.50E-06 | 2.67E-05 | 3.042462 |
| FABP1 | -0.57871 | 0.669564 | 11.14209 | -4.70791 | 8.69E-06 | 2.73E-05 | 3.020294 |
| GIN1 | 0.193483 | 1.143521 | 6.292418 | 4.707143 | 8.72E-06 | 2.74E-05 | 3.017388 |
| PDCD6 | 0.175483 | 1.129343 | 8.763516 | 4.707035 | 8.72E-06 | 2.74E-05 | 3.016975 |
| MST1R | -0.17946 | 0.883034 | 5.225604 | -4.70606 | 8.76E-06 | 2.75E-05 | 3.013269 |
| PSTPIP2 | 0.248812 | 1.188228 | 4.65956 | 4.70582 | 8.77E-06 | 2.75E-05 | 3.012363 |
| KRT32 | -0.15715 | 0.896792 | 4.91033 | -4.70436 | 8.82E-06 | 2.76E-05 | 3.006818 |
| SCGB1D2 | 0.712971 | 1.639176 | 5.676264 | 4.699395 | 8.99E-06 | 2.82E-05 | 2.987974 |
| GP2 | -0.2597 | 0.835264 | 5.045275 | -4.69583 | 9.12E-06 | 2.85E-05 | 2.97444 |
| GNAT1 | -0.14098 | 0.906906 | 4.546374 | -4.69544 | 9.13E-06 | 2.86E-05 | 2.972965 |
| MUC6 | -0.17171 | 0.887787 | 4.817912 | -4.69512 | 9.14E-06 | 2.86E-05 | 2.971756 |
| RAD50 | 0.20957 | 1.156344 | 6.024286 | 4.694267 | 9.18E-06 | 2.87E-05 | 2.96852 |
| TERF2IP | -0.16052 | 0.894704 | 8.697363 | -4.69325 | 9.21E-06 | 2.88E-05 | 2.964673 |
| RAP2C | 0.212937 | 1.159045 | 6.967473 | 4.69292 | 9.22E-06 | 2.88E-05 | 2.963413 |
| PPARG | 0.186039 | 1.137636 | 5.823736 | 4.692161 | 9.25E-06 | 2.89E-05 | 2.960534 |
| TM9SF3 | 0.203435 | 1.151436 | 9.025165 | 4.691919 | 9.26E-06 | 2.89E-05 | 2.959619 |
| PYGM | -0.19544 | 0.873307 | 5.528571 | -4.69153 | 9.28E-06 | 2.89E-05 | 2.958157 |
| SSBP3 | -0.2849 | 0.820799 | 6.918681 | -4.69151 | 9.28E-06 | 2.89E-05 | 2.958052 |
| C16orf71 | -0.15664 | 0.89711 | 4.663626 | -4.69134 | 9.28E-06 | 2.89E-05 | 2.957439 |
| BMS1 | 0.155836 | 1.114067 | 6.85967 | 4.690545 | 9.31E-06 | 2.90E-05 | 2.954409 |
| ANGPTL2 | -0.21926 | 0.859005 | 5.294835 | -4.68746 | 9.43E-06 | 2.94E-05 | 2.942737 |
| BST1 | -0.35754 | 0.780496 | 6.034066 | -4.68574 | 9.49E-06 | 2.96E-05 | 2.936215 |
| CHRNB4 | -0.14437 | 0.904776 | 3.772088 | -4.68567 | 9.49E-06 | 2.96E-05 | 2.935927 |
| MYOD1 | -0.14788 | 0.902576 | 5.382308 | -4.68446 | 9.54E-06 | 2.97E-05 | 2.931359 |
| FDX1 | -0.21704 | 0.860327 | 9.185714 | -4.6831 | 9.59E-06 | 2.98E-05 | 2.926193 |
| GREB1L | -0.18904 | 0.87719 | 5.23978 | -4.68227 | 9.62E-06 | 2.99E-05 | 2.923055 |
| UBE2M | -0.2141 | 0.862086 | 7.63978 | -4.68202 | 9.63E-06 | 2.99E-05 | 2.922103 |
| ITGA6 | 0.243831 | 1.184133 | 8.492967 | 4.681545 | 9.65E-06 | 3.00E-05 | 2.920319 |
| FAM111A | 0.193024 | 1.143157 | 7.491538 | 4.680611 | 9.68E-06 | 3.01E-05 | 2.916782 |
| KIF1A | -0.14538 | 0.90414 | 4.635934 | -4.6799 | 9.71E-06 | 3.02E-05 | 2.914093 |
| CCL22 | -0.16108 | 0.894357 | 5.591868 | -4.67942 | 9.73E-06 | 3.02E-05 | 2.912294 |
| ATAD5 | -0.17916 | 0.883217 | 5.509231 | -4.67938 | 9.73E-06 | 3.02E-05 | 2.912119 |

| | | | | | | | |
|---|---|---|---|---|---|---|---|
| ACAT1 | -0.21231 | 0.863155 | 11.2411 | -4.67927 | 9.74E-06 | 3.02E-05 | 2.911703 |
| OSBPL10 | 0.251821 | 1.190709 | 7.146484 | 4.679061 | 9.74E-06 | 3.02E-05 | 2.910915 |
| XPO7 | -0.21637 | 0.860727 | 7.726264 | -4.6745 | 9.92E-06 | 3.08E-05 | 2.89368 |
| WNT6 | -0.15911 | 0.89558 | 4.521538 | -4.67424 | 9.93E-06 | 3.08E-05 | 2.892676 |
| SV2A | -0.23147 | 0.851767 | 5.259451 | -4.67396 | 9.94E-06 | 3.08E-05 | 2.891632 |
| ARFIP2 | -0.16989 | 0.888908 | 8.290769 | -4.67169 | 1.00E-05 | 3.11E-05 | 2.88302 |
| RAB5A | 0.134783 | 1.097927 | 9.411868 | 4.666272 | 1.02E-05 | 3.17E-05 | 2.862557 |
| AKR1C2 | 0.70886 | 1.634512 | 6.819231 | 4.66568 | 1.03E-05 | 3.18E-05 | 2.860323 |
| TCEA2 | -0.2126 | 0.862978 | 6.547692 | -4.66545 | 1.03E-05 | 3.18E-05 | 2.859452 |
| SNRPD1 | 0.171841 | 1.126495 | 7.337802 | 4.665042 | 1.03E-05 | 3.18E-05 | 2.857913 |
| GPN2 | 0.183899 | 1.135949 | 6.236374 | 4.664592 | 1.03E-05 | 3.19E-05 | 2.856214 |
| SH2B3 | 0.31215 | 1.241556 | 7.021099 | 4.663142 | 1.04E-05 | 3.21E-05 | 2.850737 |
| ADO | 0.152357 | 1.111384 | 7.191429 | 4.662407 | 1.04E-05 | 3.21E-05 | 2.847958 |
| KLK5 | -0.17409 | 0.886325 | 4.99978 | -4.66166 | 1.04E-05 | 3.22E-05 | 2.845153 |
| GLA | 0.227884 | 1.171116 | 8.277473 | 4.661562 | 1.04E-05 | 3.22E-05 | 2.84477 |
| APLNR | -0.23513 | 0.849611 | 6.163077 | -4.66133 | 1.04E-05 | 3.22E-05 | 2.843891 |
| STC2 | -0.19361 | 0.874413 | 5.781648 | -4.66122 | 1.05E-05 | 3.22E-05 | 2.843463 |
| TIMM17B | -0.16553 | 0.891603 | 7.839451 | -4.6598 | 1.05E-05 | 3.24E-05 | 2.838104 |
| GRINA | -0.21912 | 0.859089 | 7.031209 | -4.65679 | 1.06E-05 | 3.28E-05 | 2.826753 |
| CCDC33 | -0.15419 | 0.898638 | 4.575275 | -4.65592 | 1.07E-05 | 3.29E-05 | 2.823462 |
| PCNT | -0.16766 | 0.890284 | 6.722308 | -4.65562 | 1.07E-05 | 3.29E-05 | 2.822362 |
| PAOX | -0.18489 | 0.879714 | 6.478352 | -4.65562 | 1.07E-05 | 3.29E-05 | 2.822329 |
| TRIP6 | 0.258285 | 1.196056 | 8.450549 | 4.655611 | 1.07E-05 | 3.29E-05 | 2.822311 |
| SPHK2 | -0.20131 | 0.869761 | 7.141538 | -4.6556 | 1.07E-05 | 3.29E-05 | 2.822273 |
| TAOK3 | 0.198116 | 1.147199 | 7.173187 | 4.655286 | 1.07E-05 | 3.29E-05 | 2.821084 |
| C1orf116 | 0.19257 | 1.142798 | 5.260879 | 4.654946 | 1.07E-05 | 3.29E-05 | 2.819799 |
| SEC24D | 0.199377 | 1.148202 | 4.140549 | 4.654877 | 1.07E-05 | 3.29E-05 | 2.819541 |
| RFC5 | 0.16757 | 1.123165 | 5.463516 | 4.653449 | 1.08E-05 | 3.31E-05 | 2.814155 |
| DPF2 | 0.16457 | 1.120832 | 7.467033 | 4.653089 | 1.08E-05 | 3.31E-05 | 2.812795 |
| ZNF84 | 0.185763 | 1.137419 | 5.531209 | 4.650198 | 1.09E-05 | 3.35E-05 | 2.801896 |
| SNAI1 | -0.16773 | 0.89024 | 5.409011 | -4.64999 | 1.09E-05 | 3.35E-05 | 2.801127 |
| CX3CR1 | 0.666628 | 1.587359 | 5.747912 | 4.647778 | 1.10E-05 | 3.38E-05 | 2.792773 |
| CYB5R4 | 0.242111 | 1.182722 | 6.684725 | 4.647482 | 1.10E-05 | 3.38E-05 | 2.791659 |
| TIGAR | 0.370222 | 1.292552 | 5.783077 | 4.647443 | 1.10E-05 | 3.38E-05 | 2.79151 |
| ELAC2 | 0.172879 | 1.127306 | 7.035055 | 4.646638 | 1.11E-05 | 3.39E-05 | 2.788478 |
| SSX3 | -0.19627 | 0.872807 | 5.020989 | -4.64654 | 1.11E-05 | 3.39E-05 | 2.788109 |
| GUCY1A2 | -0.19821 | 0.871633 | 4.422637 | -4.6461 | 1.11E-05 | 3.40E-05 | 2.786453 |
| CTDP1 | -0.13955 | 0.907802 | 6.311209 | -4.64559 | 1.11E-05 | 3.40E-05 | 2.784541 |
| NCALD | -0.28161 | 0.82267 | 6.868132 | -4.64542 | 1.11E-05 | 3.40E-05 | 2.783886 |
| LTA | -0.14788 | 0.902576 | 4.742308 | -4.64526 | 1.11E-05 | 3.40E-05 | 2.783271 |
| EFNB1 | -0.16962 | 0.889075 | 5.113077 | -4.64508 | 1.11E-05 | 3.41E-05 | 2.782599 |
| FRAS1 | -0.21493 | 0.861589 | 5.751978 | -4.64499 | 1.11E-05 | 3.41E-05 | 2.782285 |
| PRRG3 | -0.20556 | 0.867205 | 5.288352 | -4.64492 | 1.11E-05 | 3.41E-05 | 2.781986 |
| SLC4A1 | -0.45553 | 0.729244 | 8.316044 | -4.64411 | 1.12E-05 | 3.42E-05 | 2.778945 |
| DOC2A | -0.15611 | 0.897444 | 4.508022 | -4.64399 | 1.12E-05 | 3.42E-05 | 2.778487 |
| TCL1B | -0.16943 | 0.889194 | 5.311868 | -4.64362 | 1.12E-05 | 3.42E-05 | 2.77711 |
| CBFB | 0.215087 | 1.160774 | 7.095275 | 4.643191 | 1.12E-05 | 3.42E-05 | 2.775491 |

| | | | | | | | |
|---|---|---|---|---|---|---|---|
| RCL1 | -0.16496 | 0.891955 | 7.365385 | -4.64247 | 1.13E-05 | 3.43E-05 | 2.772772 |
| RAD23B | 0.13401 | 1.097339 | 8.686703 | 4.642197 | 1.13E-05 | 3.43E-05 | 2.771749 |
| ZMIZ1 | 0.228319 | 1.171469 | 9.077253 | 4.641866 | 1.13E-05 | 3.44E-05 | 2.770502 |
| CTSG | -0.1676 | 0.89032 | 5.224945 | -4.64111 | 1.13E-05 | 3.45E-05 | 2.767672 |
| PODXL | -0.7634 | 0.589108 | 9.30967 | -4.6411 | 1.13E-05 | 3.45E-05 | 2.767617 |
| LAMB2 | -0.28151 | 0.822728 | 7.142747 | -4.64007 | 1.14E-05 | 3.46E-05 | 2.763756 |
| SLITRK3 | -0.13403 | 0.91128 | 3.902198 | -4.63962 | 1.14E-05 | 3.46E-05 | 2.762035 |
| C7orf49 | 0.165507 | 1.12156 | 7.86967 | 4.638345 | 1.14E-05 | 3.48E-05 | 2.757244 |
| CCNB2 | 0.413874 | 1.332259 | 4.839011 | 4.637611 | 1.15E-05 | 3.49E-05 | 2.754482 |
| ELOVL1 | 0.179932 | 1.132831 | 7.985934 | 4.63596 | 1.15E-05 | 3.51E-05 | 2.74827 |
| AIPL1 | -0.14426 | 0.904846 | 4.737143 | -4.63565 | 1.16E-05 | 3.51E-05 | 2.747112 |
| CAT | -0.2337 | 0.850451 | 11.12791 | -4.63541 | 1.16E-05 | 3.51E-05 | 2.746214 |
| ARL4C | 0.58442 | 1.499436 | 6.107912 | 4.634308 | 1.16E-05 | 3.53E-05 | 2.742053 |
| HIST1H2AI | 0.239841 | 1.180862 | 4.101429 | 4.633012 | 1.17E-05 | 3.55E-05 | 2.737179 |
| AAAS | -0.14649 | 0.903447 | 6.985604 | -4.63256 | 1.17E-05 | 3.55E-05 | 2.735481 |
| ZC3H15 | 0.174705 | 1.128734 | 8.748132 | 4.631846 | 1.17E-05 | 3.56E-05 | 2.732792 |
| PARVA | 0.136768 | 1.099439 | 6.942198 | 4.629504 | 1.18E-05 | 3.59E-05 | 2.723988 |
| ELAVL1 | 0.140314 | 1.102145 | 7.653516 | 4.629494 | 1.18E-05 | 3.59E-05 | 2.723948 |
| PIK3IP1 | 0.203874 | 1.151787 | 7.405165 | 4.628681 | 1.19E-05 | 3.60E-05 | 2.720891 |
| CAND1 | 0.171 | 1.125839 | 7.32956 | 4.6273 | 1.19E-05 | 3.62E-05 | 2.715703 |
| SPATA2L | -0.34494 | 0.78734 | 7.427033 | -4.62683 | 1.20E-05 | 3.62E-05 | 2.713938 |
| KCNC3 | -0.15366 | 0.898969 | 4.709451 | -4.62623 | 1.20E-05 | 3.63E-05 | 2.711683 |
| ZNF140 | 0.195164 | 1.144854 | 5.868901 | 4.624939 | 1.21E-05 | 3.65E-05 | 2.706828 |
| PGLS | 0.182783 | 1.135071 | 8.315604 | 4.623778 | 1.21E-05 | 3.66E-05 | 2.702468 |
| ACTR1B | -0.12404 | 0.917615 | 7.836923 | -4.62223 | 1.22E-05 | 3.68E-05 | 2.696652 |
| REEP5 | 0.140097 | 1.101979 | 10.14363 | 4.620405 | 1.23E-05 | 3.71E-05 | 2.689798 |
| THTPA | -0.16615 | 0.891218 | 7.771099 | -4.6204 | 1.23E-05 | 3.71E-05 | 2.689792 |
| CREB3L1 | -0.15982 | 0.895136 | 5.399011 | -4.62018 | 1.23E-05 | 3.71E-05 | 2.688958 |
| CYP26A1 | -0.15187 | 0.900083 | 4.660769 | -4.61908 | 1.23E-05 | 3.72E-05 | 2.684831 |
| TNFRSF10I | -0.22774 | 0.853972 | 5.319121 | -4.61887 | 1.23E-05 | 3.72E-05 | 2.684017 |
| MTHFD2 | 0.43286 | 1.349907 | 5.732747 | 4.61886 | 1.23E-05 | 3.72E-05 | 2.683998 |
| TLN2 | -0.33867 | 0.790772 | 8.322527 | -4.61718 | 1.24E-05 | 3.75E-05 | 2.677701 |
| HNRNPF | 0.179208 | 1.132262 | 8.202967 | 4.617129 | 1.24E-05 | 3.75E-05 | 2.677499 |
| LINC00574 | -0.22234 | 0.857172 | 5.904615 | -4.61621 | 1.25E-05 | 3.76E-05 | 2.674051 |
| ASB8 | -0.15244 | 0.899728 | 6.894835 | -4.61583 | 1.25E-05 | 3.76E-05 | 2.672642 |
| FOXP3 | -0.13815 | 0.908681 | 4.884725 | -4.61444 | 1.26E-05 | 3.78E-05 | 2.667405 |
| BMP8B | -0.20204 | 0.869318 | 4.938132 | -4.61355 | 1.26E-05 | 3.80E-05 | 2.664062 |
| NFIL3 | -0.49204 | 0.71102 | 8.064945 | -4.61122 | 1.27E-05 | 3.83E-05 | 2.655331 |
| TAPBP | 0.175633 | 1.12946 | 6.89033 | 4.611 | 1.27E-05 | 3.83E-05 | 2.654504 |
| DNAJC3 | 0.202787 | 1.15092 | 5.965714 | 4.609718 | 1.28E-05 | 3.85E-05 | 2.649698 |
| TP53BP2 | 0.256304 | 1.194415 | 6.66044 | 4.609641 | 1.28E-05 | 3.85E-05 | 2.649408 |
| PPP1R3C | 0.388343 | 1.308889 | 7.097473 | 4.609242 | 1.28E-05 | 3.85E-05 | 2.647914 |
| RFPL3 | -0.22563 | 0.85522 | 5.854945 | -4.60888 | 1.28E-05 | 3.86E-05 | 2.646567 |
| SNPH | -0.16255 | 0.893444 | 5.524835 | -4.60853 | 1.29E-05 | 3.86E-05 | 2.645235 |
| CASP7 | 0.21915 | 1.164047 | 6.36011 | 4.608152 | 1.29E-05 | 3.87E-05 | 2.643828 |
| TOR1B | 0.158314 | 1.115982 | 6.912418 | 4.607264 | 1.29E-05 | 3.88E-05 | 2.640498 |
| ADGRG3 | -0.15277 | 0.899523 | 4.97989 | -4.60575 | 1.30E-05 | 3.90E-05 | 2.634841 |

| | | | | | | | |
|---|---|---|---|---|---|---|---|
| GCGR | -0.19615 | 0.872874 | 5.516044 | -4.60237 | 1.32E-05 | 3.95E-05 | 2.622179 |
| KATNA1 | 0.216729 | 1.162096 | 7.27 | 4.601782 | 1.32E-05 | 3.96E-05 | 2.619958 |
| ELP3 | 0.166266 | 1.12215 | 7.224176 | 4.600648 | 1.33E-05 | 3.97E-05 | 2.615709 |
| LOC10192 | -0.14018 | 0.907407 | 4.102637 | -4.60057 | 1.33E-05 | 3.97E-05 | 2.615429 |
| MMP26 | -0.18454 | 0.879932 | 4.524615 | -4.60019 | 1.33E-05 | 3.98E-05 | 2.613984 |
| LMBR1L | -0.17797 | 0.883945 | 6.143297 | -4.59958 | 1.33E-05 | 3.99E-05 | 2.611706 |
| TPK1 | 0.228005 | 1.171214 | 5.182967 | 4.597467 | 1.34E-05 | 4.02E-05 | 2.603801 |
| EVI2B | 0.503034 | 1.417191 | 4.655275 | 4.596934 | 1.35E-05 | 4.02E-05 | 2.601805 |
| CCR4 | -0.18776 | 0.877966 | 4.747802 | -4.59677 | 1.35E-05 | 4.03E-05 | 2.601204 |
| PPP1R17 | -0.14072 | 0.907063 | 4.427802 | -4.59669 | 1.35E-05 | 4.03E-05 | 2.600897 |
| AGXT | -0.82857 | 0.563089 | 7.284835 | -4.59538 | 1.35E-05 | 4.04E-05 | 2.595997 |
| EFHC1 | 0.250551 | 1.189661 | 5.328681 | 4.59438 | 1.36E-05 | 4.06E-05 | 2.592247 |
| PAK1IP1 | 0.165734 | 1.121737 | 5.28 | 4.59425 | 1.36E-05 | 4.06E-05 | 2.59176 |
| SST | -0.36076 | 0.778752 | 4.757253 | -4.59109 | 1.38E-05 | 4.11E-05 | 2.579947 |
| BMPR1B | -0.22472 | 0.855761 | 5.336484 | -4.58908 | 1.39E-05 | 4.14E-05 | 2.57242 |
| MPRIP | -0.15664 | 0.897113 | 6.053846 | -4.58689 | 1.40E-05 | 4.17E-05 | 2.56424 |
| DSG3 | -0.1327 | 0.912126 | 4.293077 | -4.58669 | 1.40E-05 | 4.18E-05 | 2.563497 |
| LMAN1L | -0.17905 | 0.883283 | 5.714066 | -4.58597 | 1.40E-05 | 4.19E-05 | 2.560786 |
| GULP1 | 0.191411 | 1.14188 | 7.448132 | 4.584408 | 1.41E-05 | 4.21E-05 | 2.554958 |
| ZMAT3 | 0.202401 | 1.150612 | 6.528132 | 4.583801 | 1.42E-05 | 4.22E-05 | 2.55269 |
| PCBP3 | -0.17632 | 0.884955 | 5.378242 | -4.5825 | 1.42E-05 | 4.24E-05 | 2.547849 |
| GYPC | 0.27201 | 1.207489 | 7.614945 | 4.58063 | 1.43E-05 | 4.27E-05 | 2.540846 |
| TRAV12-2 | -0.22453 | 0.855873 | 4.625055 | -4.58031 | 1.44E-05 | 4.27E-05 | 2.539655 |
| AQP8 | -0.18216 | 0.881383 | 5.282747 | -4.57938 | 1.44E-05 | 4.29E-05 | 2.536183 |
| NAA15 | 0.20442 | 1.152223 | 5.42 | 4.578987 | 1.44E-05 | 4.29E-05 | 2.53471 |
| GPS1 | -0.19761 | 0.871995 | 7.85989 | -4.57887 | 1.44E-05 | 4.29E-05 | 2.534287 |
| IPO13 | -0.18174 | 0.881637 | 7.261648 | -4.57872 | 1.44E-05 | 4.29E-05 | 2.53373 |
| TAAR3 | -0.14084 | 0.906994 | 4.202747 | -4.57809 | 1.45E-05 | 4.30E-05 | 2.531347 |
| HYPM | -0.1562 | 0.897387 | 4.493846 | -4.57649 | 1.46E-05 | 4.33E-05 | 2.525396 |
| TTLL7 | 0.295473 | 1.227288 | 4.702418 | 4.575972 | 1.46E-05 | 4.33E-05 | 2.523456 |
| JPH2 | -0.14379 | 0.90514 | 4.518462 | -4.57229 | 1.48E-05 | 4.39E-05 | 2.509698 |
| TBC1D13 | -0.20435 | 0.867928 | 7.723077 | -4.57079 | 1.49E-05 | 4.42E-05 | 2.504103 |
| RTCA | 0.274889 | 1.209901 | 9.255934 | 4.570327 | 1.49E-05 | 4.43E-05 | 2.502395 |
| ESPL1 | -0.4625 | 0.725729 | 6.448901 | -4.5681 | 1.51E-05 | 4.46E-05 | 2.494093 |
| HS3ST1 | 0.404155 | 1.323313 | 4.976813 | 4.56794 | 1.51E-05 | 4.46E-05 | 2.493493 |
| KHK | -0.30496 | 0.809464 | 9.885934 | -4.56586 | 1.52E-05 | 4.50E-05 | 2.485742 |
| GNL1 | -0.13688 | 0.909481 | 5.112527 | -4.56504 | 1.52E-05 | 4.51E-05 | 2.482683 |
| KLC2 | -0.13338 | 0.911692 | 5.201868 | -4.5642 | 1.53E-05 | 4.52E-05 | 2.479571 |
| S100A2 | 0.777594 | 1.71427 | 9.506264 | 4.564157 | 1.53E-05 | 4.52E-05 | 2.479392 |
| ABCC4 | 0.303159 | 1.233844 | 7.632088 | 4.562874 | 1.54E-05 | 4.54E-05 | 2.474613 |
| NAMPT | 0.31343 | 1.242659 | 7.839341 | 4.562425 | 1.54E-05 | 4.55E-05 | 2.472942 |
| CFD | 0.384029 | 1.304981 | 5.591209 | 4.561339 | 1.55E-05 | 4.57E-05 | 2.468893 |
| TPCN1 | -0.24373 | 0.844556 | 8.491429 | -4.56093 | 1.55E-05 | 4.57E-05 | 2.467385 |
| BID | 0.226628 | 1.170097 | 5.96033 | 4.56069 | 1.55E-05 | 4.58E-05 | 2.466476 |
| LINC00341 | -0.23179 | 0.851576 | 5.704725 | -4.55948 | 1.56E-05 | 4.60E-05 | 2.46197 |
| NDUFB2 | 0.149164 | 1.108927 | 11.57615 | 4.559311 | 1.56E-05 | 4.60E-05 | 2.461341 |
| ALOX12 | -0.12726 | 0.915571 | 4.360549 | -4.5589 | 1.56E-05 | 4.60E-05 | 2.459817 |

| | | | | | | | |
|---|---|---|---|---|---|---|---|
| LEP | -0.13163 | 0.912798 | 4.051429 | -4.55849 | 1.56E-05 | 4.61E-05 | 2.458277 |
| ISG15 | 0.842174 | 1.792749 | 7.094286 | 4.557095 | 1.57E-05 | 4.63E-05 | 2.45309 |
| TAS2R4 | -0.18055 | 0.882369 | 4.336154 | -4.55675 | 1.57E-05 | 4.64E-05 | 2.451821 |
| TSPAN8 | 0.791024 | 1.730302 | 8.007253 | 4.555983 | 1.58E-05 | 4.65E-05 | 2.448952 |
| CYP3A5 | -0.59863 | 0.660379 | 5.945495 | -4.55475 | 1.59E-05 | 4.67E-05 | 2.444353 |
| DLX5 | -0.13857 | 0.908422 | 4.436044 | -4.55429 | 1.59E-05 | 4.68E-05 | 2.442649 |
| SLC22A4 | -0.34815 | 0.785588 | 7.810879 | -4.55344 | 1.59E-05 | 4.69E-05 | 2.439485 |
| MYBPC3 | -0.16275 | 0.893318 | 6.325604 | -4.55215 | 1.60E-05 | 4.71E-05 | 2.434679 |
| DNAJC2 | 0.176319 | 1.129997 | 7.391538 | 4.552084 | 1.60E-05 | 4.71E-05 | 2.434442 |
| TFF2 | -0.13351 | 0.91161 | 4.285934 | -4.55207 | 1.60E-05 | 4.71E-05 | 2.4344 |
| SERTAD3 | -0.24746 | 0.842376 | 6.761758 | -4.55113 | 1.61E-05 | 4.73E-05 | 2.430913 |
| CYBB | 0.45572 | 1.371467 | 5.282747 | 4.549357 | 1.62E-05 | 4.76E-05 | 2.424302 |
| EIF3M | 0.201778 | 1.150115 | 8.82978 | 4.549287 | 1.62E-05 | 4.76E-05 | 2.42404 |
| RGS7 | -0.52583 | 0.694561 | 4.907802 | -4.54888 | 1.62E-05 | 4.76E-05 | 2.422517 |
| LPL | -0.5314 | 0.691885 | 6.064396 | -4.54886 | 1.62E-05 | 4.76E-05 | 2.422463 |
| SH3YL1 | -0.35463 | 0.782071 | 10.18637 | -4.5483 | 1.63E-05 | 4.77E-05 | 2.420377 |
| LIMD2 | -0.1715 | 0.88792 | 4.867802 | -4.54768 | 1.63E-05 | 4.78E-05 | 2.418053 |
| TMPRSS2 | -0.29795 | 0.813409 | 7.155055 | -4.5469 | 1.63E-05 | 4.79E-05 | 2.41517 |
| GHRH | -0.1614 | 0.894159 | 4.387363 | -4.5464 | 1.64E-05 | 4.80E-05 | 2.41331 |
| DPT | -0.17187 | 0.887692 | 4.180879 | -4.54634 | 1.64E-05 | 4.80E-05 | 2.413099 |
| SYNGR4 | -0.15378 | 0.898894 | 4.713956 | -4.54582 | 1.64E-05 | 4.81E-05 | 2.411139 |
| NELFB | 0.168314 | 1.123744 | 7.017363 | 4.54527 | 1.65E-05 | 4.82E-05 | 2.409105 |
| CSAD | -0.18304 | 0.880846 | 5.152747 | -4.54459 | 1.65E-05 | 4.83E-05 | 2.406567 |
| LINC0126( | -0.17174 | 0.887772 | 5.656813 | -4.54269 | 1.66E-05 | 4.86E-05 | 2.399537 |
| DTX2 | -0.16588 | 0.891385 | 4.593407 | -4.54265 | 1.66E-05 | 4.86E-05 | 2.399365 |
| TRPV4 | -0.27332 | 0.827414 | 5.695495 | -4.54155 | 1.67E-05 | 4.88E-05 | 2.395277 |
| PHF21A | 0.147 | 1.107265 | 5.957692 | 4.539785 | 1.68E-05 | 4.91E-05 | 2.38873 |
| NELL1 | -0.44136 | 0.736439 | 6.31044 | -4.53822 | 1.69E-05 | 4.94E-05 | 2.382905 |
| MB | -0.18197 | 0.881501 | 5.161538 | -4.53762 | 1.69E-05 | 4.95E-05 | 2.38068 |
| PSMD10 | 0.163367 | 1.119898 | 8.392308 | 4.533617 | 1.72E-05 | 5.03E-05 | 2.365835 |
| TONSL | -0.18705 | 0.878398 | 5.10011 | -4.53347 | 1.72E-05 | 5.03E-05 | 2.365289 |
| PLIN3 | 0.285541 | 1.218867 | 7.960549 | 4.533065 | 1.72E-05 | 5.03E-05 | 2.363785 |
| GRHPR | -0.265 | 0.832202 | 10.53418 | -4.53229 | 1.73E-05 | 5.05E-05 | 2.360922 |
| ACO1 | -0.28033 | 0.823404 | 10.21659 | -4.53215 | 1.73E-05 | 5.05E-05 | 2.360375 |
| ZNF562 | 0.184932 | 1.136764 | 5.773407 | 4.529464 | 1.75E-05 | 5.10E-05 | 2.350429 |
| MFAP3L | 0.186787 | 1.138226 | 5.937802 | 4.529341 | 1.75E-05 | 5.10E-05 | 2.34997 |
| LCT | -0.169 | 0.889456 | 4.861209 | -4.52923 | 1.75E-05 | 5.10E-05 | 2.349548 |
| FXYD3 | -0.19792 | 0.871805 | 6.465604 | -4.52918 | 1.75E-05 | 5.10E-05 | 2.349391 |
| VSTM4 | -0.30486 | 0.809524 | 5.890769 | -4.52601 | 1.77E-05 | 5.16E-05 | 2.337619 |
| DNAJC15 | 0.232319 | 1.174722 | 9.089231 | 4.526 | 1.77E-05 | 5.16E-05 | 2.337585 |
| ACTR2 | 0.246111 | 1.186006 | 8.606703 | 4.525868 | 1.77E-05 | 5.16E-05 | 2.337097 |
| TUBB6 | 0.496261 | 1.410553 | 7.197143 | 4.524556 | 1.78E-05 | 5.18E-05 | 2.332234 |
| BANF1 | 0.20385 | 1.151768 | 9.174066 | 4.52448 | 1.78E-05 | 5.18E-05 | 2.331952 |
| SH3BGRL | 0.172406 | 1.126936 | 9.093516 | 4.523239 | 1.79E-05 | 5.21E-05 | 2.327354 |
| NOTCH2 | 0.130242 | 1.094477 | 6.765275 | 4.521525 | 1.80E-05 | 5.24E-05 | 2.321003 |
| CACNA1E | -0.17231 | 0.887418 | 4.330659 | -4.52141 | 1.80E-05 | 5.24E-05 | 2.320594 |
| MCUR1 | -0.25475 | 0.838133 | 8.73033 | -4.5192 | 1.82E-05 | 5.28E-05 | 2.312376 |

| | | | | | | | |
|---|---|---|---|---|---|---|---|
| FOXRED2 | -0.18235 | 0.881265 | 5.543956 | -4.51714 | 1.83E-05 | 5.32E-05 | 2.304782 |
| DDB2 | 0.248184 | 1.187711 | 5.700989 | 4.514441 | 1.85E-05 | 5.38E-05 | 2.294775 |
| DGKZ | 0.31287 | 1.242176 | 7.783846 | 4.512775 | 1.87E-05 | 5.41E-05 | 2.288613 |
| NTF3 | -0.17828 | 0.883756 | 4.969231 | -4.51262 | 1.87E-05 | 5.41E-05 | 2.288037 |
| PTP4A2 | 0.140604 | 1.102366 | 8.896703 | 4.512556 | 1.87E-05 | 5.41E-05 | 2.287803 |
| ZC3HAV1 | 0.168164 | 1.123628 | 7.090549 | 4.512551 | 1.87E-05 | 5.41E-05 | 2.287783 |
| TSPO2 | -0.17537 | 0.885542 | 5.121758 | -4.51197 | 1.87E-05 | 5.42E-05 | 2.285629 |
| TLE6 | -0.16427 | 0.89238 | 4.486593 | -4.51116 | 1.88E-05 | 5.44E-05 | 2.282628 |
| KCNN2 | -0.73577 | 0.600496 | 5.306374 | -4.5088 | 1.89E-05 | 5.48E-05 | 2.273903 |
| PHKB | 0.196203 | 1.145679 | 7.736154 | 4.508735 | 1.90E-05 | 5.48E-05 | 2.273668 |
| FAM129A | 0.286362 | 1.219561 | 4.737912 | 4.508213 | 1.90E-05 | 5.49E-05 | 2.27174 |
| ACKR3 | -0.50172 | 0.706264 | 6.304505 | -4.50813 | 1.90E-05 | 5.49E-05 | 2.271429 |
| GDE1 | 0.194435 | 1.144276 | 8.065714 | 4.507594 | 1.90E-05 | 5.50E-05 | 2.269453 |
| ANPEP | 0.337444 | 1.263516 | 10.30187 | 4.506325 | 1.91E-05 | 5.53E-05 | 2.264762 |
| RALB | 0.165145 | 1.121279 | 8.353187 | 4.506255 | 1.91E-05 | 5.53E-05 | 2.264503 |
| NUCB2 | 0.271633 | 1.207173 | 6.797802 | 4.50538 | 1.92E-05 | 5.54E-05 | 2.261266 |
| DHRS11 | -0.25589 | 0.837468 | 6.288242 | -4.50435 | 1.93E-05 | 5.56E-05 | 2.257469 |
| TMEM8A | 0.256097 | 1.194243 | 6.610989 | 4.504129 | 1.93E-05 | 5.57E-05 | 2.256645 |
| EIF4G1 | 0.16856 | 1.123936 | 6.988571 | 4.503631 | 1.93E-05 | 5.58E-05 | 2.254806 |
| TMEM74B | -0.14842 | 0.902241 | 4.907912 | -4.49986 | 1.96E-05 | 5.66E-05 | 2.240863 |
| CASQ2 | -0.22343 | 0.856527 | 5.115165 | -4.49946 | 1.96E-05 | 5.66E-05 | 2.2394 |
| ABTB2 | -0.19453 | 0.873857 | 6.53989 | -4.49673 | 1.99E-05 | 5.72E-05 | 2.229321 |
| TNS1 | -0.20432 | 0.867948 | 7.524615 | -4.49621 | 1.99E-05 | 5.73E-05 | 2.227412 |
| NPTX1 | -0.1717 | 0.887796 | 4.588571 | -4.49383 | 2.01E-05 | 5.78E-05 | 2.218617 |
| STX7 | 0.177357 | 1.130811 | 8.938791 | 4.493623 | 2.01E-05 | 5.78E-05 | 2.217853 |
| RLBP1 | -0.18545 | 0.879375 | 5.053077 | -4.49234 | 2.02E-05 | 5.81E-05 | 2.213117 |
| TRAV10 | -0.14077 | 0.907033 | 4.425604 | -4.49105 | 2.03E-05 | 5.84E-05 | 2.208377 |
| OR7E14P | 0.336763 | 1.26292 | 7.120879 | 4.489944 | 2.04E-05 | 5.86E-05 | 2.204284 |
| CHRNA6 | -0.13357 | 0.911576 | 4.893516 | -4.48915 | 2.04E-05 | 5.87E-05 | 2.20137 |
| BNIP3L | -0.17993 | 0.882747 | 9.434286 | -4.48913 | 2.04E-05 | 5.87E-05 | 2.201297 |
| HLA-G | 0.176304 | 1.129986 | 8.280879 | 4.48886 | 2.05E-05 | 5.88E-05 | 2.200286 |
| ADIPOQ | -0.18377 | 0.880401 | 3.91956 | -4.48845 | 2.05E-05 | 5.89E-05 | 2.198774 |
| MYO16 | -0.14384 | 0.905106 | 4.886044 | -4.48803 | 2.05E-05 | 5.89E-05 | 2.197221 |
| AKIP1 | 0.160179 | 1.117426 | 6.047253 | 4.488016 | 2.05E-05 | 5.89E-05 | 2.197175 |
| BDH2 | -0.21822 | 0.859627 | 10.90231 | -4.48781 | 2.05E-05 | 5.89E-05 | 2.196431 |
| SLC12A9 | -0.15462 | 0.89837 | 6.405714 | -4.48712 | 2.06E-05 | 5.91E-05 | 2.193863 |
| HYI | -0.18756 | 0.878089 | 7.517033 | -4.48304 | 2.09E-05 | 6.00E-05 | 2.178818 |
| SPCS3 | 0.190884 | 1.141463 | 7.794176 | 4.4823 | 2.10E-05 | 6.01E-05 | 2.17611 |
| CD5 | -0.13805 | 0.908748 | 5.89956 | -4.48183 | 2.10E-05 | 6.02E-05 | 2.174395 |
| XRCC2 | -0.24903 | 0.84146 | 6.00033 | -4.48138 | 2.11E-05 | 6.03E-05 | 2.172731 |
| MC5R | -0.15875 | 0.895802 | 4.487802 | -4.47966 | 2.12E-05 | 6.07E-05 | 2.166372 |
| PPP1R26 | -0.18679 | 0.87856 | 6.462198 | -4.47929 | 2.12E-05 | 6.08E-05 | 2.165021 |
| POT1 | 0.161188 | 1.118208 | 6.353187 | 4.478655 | 2.13E-05 | 6.09E-05 | 2.162684 |
| SHARPIN | -0.13684 | 0.909512 | 7.004725 | -4.47772 | 2.14E-05 | 6.11E-05 | 2.159231 |
| CHPT1 | -0.23732 | 0.848318 | 9.613077 | -4.47618 | 2.15E-05 | 6.14E-05 | 2.153563 |
| PEX7 | -0.20111 | 0.86988 | 7.260549 | -4.47477 | 2.16E-05 | 6.17E-05 | 2.148389 |
| AGTPBP1 | 0.170411 | 1.125379 | 5.932747 | 4.473876 | 2.17E-05 | 6.19E-05 | 2.145095 |

| | | | | | | | |
|---|---|---|---|---|---|---|---|
| FMNL1 | -0.18229 | 0.881306 | 5.437033 | -4.47339 | 2.17E-05 | 6.20E-05 | 2.143305 |
| APOD | -0.50186 | 0.706193 | 6.497912 | -4.47332 | 2.17E-05 | 6.20E-05 | 2.143045 |
| FURIN | -0.18286 | 0.880958 | 6.941099 | -4.47262 | 2.18E-05 | 6.22E-05 | 2.14049 |
| HCFC1R1 | 0.199314 | 1.148152 | 7.897692 | 4.471222 | 2.19E-05 | 6.25E-05 | 2.135334 |
| GOLGA3 | 0.177087 | 1.130599 | 6.706484 | 4.469952 | 2.20E-05 | 6.28E-05 | 2.130662 |
| SLC28A3 | -0.1373 | 0.909216 | 4.793407 | -4.46979 | 2.20E-05 | 6.28E-05 | 2.130065 |
| HRC | -0.15687 | 0.896966 | 4.293077 | -4.46857 | 2.21E-05 | 6.31E-05 | 2.125571 |
| TGM1 | -0.20323 | 0.868603 | 4.694066 | -4.46851 | 2.21E-05 | 6.31E-05 | 2.12537 |
| TRMT1L | 0.223691 | 1.167717 | 6.006703 | 4.468401 | 2.21E-05 | 6.31E-05 | 2.124959 |
| TBP | 0.139671 | 1.101654 | 6.654286 | 4.467933 | 2.22E-05 | 6.32E-05 | 2.123238 |
| CD33 | -0.20016 | 0.870454 | 5.563846 | -4.46709 | 2.23E-05 | 6.33E-05 | 2.120154 |
| FHL3 | -0.17068 | 0.888423 | 6.084945 | -4.46708 | 2.23E-05 | 6.33E-05 | 2.120106 |
| DARS | 0.136188 | 1.098998 | 8.995824 | 4.46674 | 2.23E-05 | 6.34E-05 | 2.118854 |
| ESRRG | -0.24243 | 0.845317 | 8.080549 | -4.46664 | 2.23E-05 | 6.34E-05 | 2.118482 |
| IL25 | -0.15759 | 0.896522 | 5.380549 | -4.4642 | 2.25E-05 | 6.40E-05 | 2.10953 |
| FNDC3B | 0.318865 | 1.247349 | 7.616593 | 4.463317 | 2.26E-05 | 6.41E-05 | 2.106279 |
| SLC38A10 | -0.2052 | 0.867417 | 7.134396 | -4.46328 | 2.26E-05 | 6.41E-05 | 2.106136 |
| BRCC3 | 0.249391 | 1.188705 | 6.055934 | 4.458802 | 2.30E-05 | 6.52E-05 | 2.089697 |
| GPR65 | 0.30099 | 1.23199 | 4.193407 | 4.458798 | 2.30E-05 | 6.52E-05 | 2.089681 |
| TMEM147 | -0.171 | 0.88823 | 5.870659 | -4.45729 | 2.31E-05 | 6.56E-05 | 2.084164 |
| RPS6KC1 | 0.251676 | 1.19059 | 5.98989 | 4.452212 | 2.36E-05 | 6.68E-05 | 2.065515 |
| LYRM1 | -0.15657 | 0.897158 | 9.737143 | -4.4508 | 2.37E-05 | 6.72E-05 | 2.060348 |
| PTCH2 | -0.146 | 0.90375 | 5.067582 | -4.45008 | 2.38E-05 | 6.73E-05 | 2.057692 |
| ZNF552 | 0.213802 | 1.15974 | 6.706813 | 4.449552 | 2.38E-05 | 6.75E-05 | 2.055759 |
| SPINK5 | -0.19558 | 0.873219 | 4.841978 | -4.44887 | 2.39E-05 | 6.76E-05 | 2.053244 |
| MYO1A | -0.15922 | 0.895511 | 4.656484 | -4.44837 | 2.39E-05 | 6.77E-05 | 2.051437 |
| CANX | 0.195947 | 1.145476 | 9.874505 | 4.44768 | 2.40E-05 | 6.79E-05 | 2.048899 |
| WWOX | -0.14026 | 0.907355 | 7.228901 | -4.44733 | 2.40E-05 | 6.79E-05 | 2.047629 |
| GNL3 | 0.250889 | 1.18994 | 7.144066 | 4.446926 | 2.40E-05 | 6.80E-05 | 2.046133 |
| EPPIN | -0.13401 | 0.911295 | 4.003297 | -4.44613 | 2.41E-05 | 6.82E-05 | 2.043235 |
| TMX2 | 0.157643 | 1.115463 | 9.621868 | 4.44378 | 2.43E-05 | 6.88E-05 | 2.034606 |
| HMOX1 | 0.499821 | 1.414038 | 9.499121 | 4.443228 | 2.44E-05 | 6.89E-05 | 2.032586 |
| ELN | -0.16296 | 0.893193 | 5.096374 | -4.44306 | 2.44E-05 | 6.90E-05 | 2.031974 |
| PRB4 | -0.17666 | 0.884748 | 4.892857 | -4.44256 | 2.45E-05 | 6.90E-05 | 2.030122 |
| PPP2R3C | 0.263449 | 1.200345 | 6.835495 | 4.442527 | 2.45E-05 | 6.90E-05 | 2.030019 |
| GNB2L1 | 0.166565 | 1.122383 | 12.3178 | 4.442508 | 2.45E-05 | 6.90E-05 | 2.029948 |
| PUF60 | 0.159889 | 1.117201 | 8.774066 | 4.441163 | 2.46E-05 | 6.94E-05 | 2.025023 |
| IFIT5 | 0.274758 | 1.209792 | 7.16 | 4.44081 | 2.46E-05 | 6.94E-05 | 2.023729 |
| ARHGEF38 | -0.18974 | 0.876764 | 4.957912 | -4.43927 | 2.48E-05 | 6.98E-05 | 2.018103 |
| PLK3 | -0.2333 | 0.850684 | 5.165934 | -4.43887 | 2.48E-05 | 6.99E-05 | 2.016643 |
| AP2M1 | 0.247812 | 1.187405 | 9.624066 | 4.437973 | 2.49E-05 | 7.01E-05 | 2.013342 |
| FCER1G | 0.575072 | 1.489752 | 7.222637 | 4.435318 | 2.51E-05 | 7.08E-05 | 2.003627 |
| DNAJB9 | -0.22232 | 0.857184 | 9.255495 | -4.43504 | 2.52E-05 | 7.09E-05 | 2.002595 |
| PPY2P | -0.14307 | 0.905592 | 4.211209 | -4.43404 | 2.53E-05 | 7.11E-05 | 1.998936 |
| COBL | 0.206415 | 1.153818 | 7.900769 | 4.430486 | 2.56E-05 | 7.21E-05 | 1.985953 |
| RSG1 | -0.16136 | 0.894183 | 5.349121 | -4.42859 | 2.58E-05 | 7.26E-05 | 1.979022 |
| APOM | -0.48576 | 0.714119 | 11.12044 | -4.42713 | 2.59E-05 | 7.30E-05 | 1.973672 |

| | | | | | | | |
|---|---|---|---|---|---|---|---|
| INE1 | -0.15717 | 0.896783 | 5.72967 | -4.42703 | 2.59E-05 | 7.30E-05 | 1.973339 |
| HIST1H4D | -0.20899 | 0.865145 | 4.072308 | -4.42654 | 2.60E-05 | 7.31E-05 | 1.971522 |
| INS | -0.15034 | 0.901039 | 3.78044 | -4.42538 | 2.61E-05 | 7.34E-05 | 1.967305 |
| CD70 | -0.15999 | 0.895031 | 6.211319 | -4.42509 | 2.61E-05 | 7.34E-05 | 1.966223 |
| MRPS16 | 0.148957 | 1.108767 | 8.126703 | 4.423371 | 2.63E-05 | 7.39E-05 | 1.959953 |
| KCNK5 | -0.46491 | 0.724515 | 8.399011 | -4.42178 | 2.65E-05 | 7.43E-05 | 1.954151 |
| CYP7B1 | 0.16257 | 1.119279 | 4.656044 | 4.42096 | 2.66E-05 | 7.45E-05 | 1.951149 |
| ZNF350 | 0.152362 | 1.111388 | 5.021648 | 4.420261 | 2.66E-05 | 7.47E-05 | 1.948598 |
| HAO1 | -0.31517 | 0.803757 | 4.451538 | -4.42013 | 2.66E-05 | 7.47E-05 | 1.948112 |
| METTL3 | 0.174589 | 1.128643 | 7.342857 | 4.416904 | 2.70E-05 | 7.56E-05 | 1.936346 |
| ATP6V1C1 | 0.185329 | 1.137076 | 7.401429 | 4.416407 | 2.70E-05 | 7.57E-05 | 1.934534 |
| TIPIN | 0.216063 | 1.161559 | 4.94967 | 4.41437 | 2.72E-05 | 7.63E-05 | 1.927102 |
| GTPBP8 | 0.165053 | 1.121207 | 7.609011 | 4.412517 | 2.74E-05 | 7.68E-05 | 1.920342 |
| MIS12 | 0.173812 | 1.128035 | 6.347473 | 4.412024 | 2.75E-05 | 7.69E-05 | 1.918548 |
| NMNAT2 | -0.14182 | 0.906374 | 4.757912 | -4.41001 | 2.77E-05 | 7.75E-05 | 1.911199 |
| CD163 | 0.54186 | 1.455848 | 5.261648 | 4.409999 | 2.77E-05 | 7.75E-05 | 1.911162 |
| FLRT1 | -0.19896 | 0.871178 | 5.398352 | -4.40796 | 2.79E-05 | 7.81E-05 | 1.903727 |
| C1orf159 | -0.15353 | 0.899047 | 5.275165 | -4.40676 | 2.80E-05 | 7.84E-05 | 1.899347 |
| LYRM4 | 0.196725 | 1.146093 | 7.29989 | 4.405186 | 2.82E-05 | 7.88E-05 | 1.893627 |
| SIT1 | -0.15468 | 0.898331 | 6.032857 | -4.40469 | 2.83E-05 | 7.89E-05 | 1.891823 |
| RFPL2 | -0.17187 | 0.887692 | 3.860879 | -4.40465 | 2.83E-05 | 7.89E-05 | 1.891688 |
| STAMBP | 0.142217 | 1.1036 | 7.70011 | 4.404642 | 2.83E-05 | 7.89E-05 | 1.891643 |
| NOD1 | -0.15588 | 0.897585 | 6.138352 | -4.40313 | 2.84E-05 | 7.94E-05 | 1.886123 |
| FBXO28 | 0.214604 | 1.160385 | 7.783297 | 4.402281 | 2.85E-05 | 7.96E-05 | 1.883044 |
| LIPC | 0.564686 | 1.479066 | 7.55011 | 4.401509 | 2.86E-05 | 7.98E-05 | 1.880234 |
| ANKRD17 | 0.153546 | 1.1123 | 7.465495 | 4.401292 | 2.86E-05 | 7.98E-05 | 1.879445 |
| ANKRD49 | 0.194072 | 1.143988 | 7.409231 | 4.401272 | 2.86E-05 | 7.98E-05 | 1.879374 |
| RPAP1 | -0.13133 | 0.91299 | 6.175275 | -4.40101 | 2.87E-05 | 7.99E-05 | 1.878426 |
| WBP2 | -0.16886 | 0.889545 | 7.087802 | -4.40085 | 2.87E-05 | 7.99E-05 | 1.877852 |
| FILIP1L | -0.3502 | 0.784474 | 5.747692 | -4.4006 | 2.87E-05 | 7.99E-05 | 1.876921 |
| ASIC3 | -0.18703 | 0.878413 | 4.381209 | -4.39986 | 2.88E-05 | 8.01E-05 | 1.874223 |
| MGST3 | 0.158623 | 1.116221 | 11.79648 | 4.399565 | 2.88E-05 | 8.02E-05 | 1.87316 |
| KLF3 | 0.19015 | 1.140882 | 6.470769 | 4.399281 | 2.89E-05 | 8.03E-05 | 1.872124 |
| SIGLEC9 | -0.17997 | 0.882724 | 4.812527 | -4.39696 | 2.91E-05 | 8.09E-05 | 1.863664 |
| MGEA5 | 0.233343 | 1.175556 | 8.115824 | 4.396048 | 2.92E-05 | 8.12E-05 | 1.860362 |
| INA | -0.14063 | 0.907124 | 5.182198 | -4.3959 | 2.92E-05 | 8.12E-05 | 1.859841 |
| IGSF6 | 0.304512 | 1.235001 | 3.903626 | 4.394662 | 2.94E-05 | 8.16E-05 | 1.855321 |
| CBX7 | -0.22322 | 0.85665 | 7.864615 | -4.3944 | 2.94E-05 | 8.16E-05 | 1.85435 |
| USP47 | 0.160034 | 1.117313 | 7.740659 | 4.393113 | 2.95E-05 | 8.20E-05 | 1.849688 |
| CCDC177 | -0.16236 | 0.893564 | 4.393626 | -4.39248 | 2.96E-05 | 8.22E-05 | 1.847381 |
| TROVE2 | 0.254285 | 1.192744 | 8.458571 | 4.392182 | 2.96E-05 | 8.22E-05 | 1.846304 |
| HARS | 0.149705 | 1.109343 | 8.120769 | 4.39207 | 2.97E-05 | 8.23E-05 | 1.845895 |
| MYH10 | 0.173812 | 1.128035 | 8.697473 | 4.391776 | 2.97E-05 | 8.23E-05 | 1.844826 |
| NSUN3 | 0.186744 | 1.138192 | 7.145824 | 4.390362 | 2.98E-05 | 8.27E-05 | 1.839686 |
| PSMC4 | 0.157971 | 1.115717 | 7.886813 | 4.390252 | 2.99E-05 | 8.27E-05 | 1.839286 |
| CEBPE | -0.16881 | 0.889575 | 4.84 | -4.38947 | 2.99E-05 | 8.30E-05 | 1.836448 |
| HMGB3 | 0.234164 | 1.176225 | 7.033187 | 4.385296 | 3.04E-05 | 8.43E-05 | 1.821283 |

| | | | | | | | |
|---|---|---|---|---|---|---|---|
| KCNN4 | -0.15399 | 0.898761 | 4.884286 | -4.3845 | 3.05E-05 | 8.45E-05 | 1.818383 |
| IQCC | -0.17351 | 0.886682 | 5.256154 | -4.38447 | 3.05E-05 | 8.45E-05 | 1.818285 |
| ENDOD1 | 0.267947 | 1.204093 | 6.98011 | 4.384166 | 3.06E-05 | 8.45E-05 | 1.817177 |
| RNASE3 | -0.17797 | 0.883945 | 5.223297 | -4.38383 | 3.06E-05 | 8.46E-05 | 1.815956 |
| SUV39H1 | -0.16286 | 0.893256 | 5.710989 | -4.38327 | 3.07E-05 | 8.48E-05 | 1.81393 |
| SNX27 | -0.13929 | 0.907963 | 7.122857 | -4.38262 | 3.07E-05 | 8.50E-05 | 1.811564 |
| SLC10A2 | -0.50449 | 0.704908 | 8.078352 | -4.38195 | 3.08E-05 | 8.51E-05 | 1.809121 |
| FAM76A | -0.15924 | 0.895496 | 4.845385 | -4.38099 | 3.09E-05 | 8.54E-05 | 1.805647 |
| PDYN | -0.14271 | 0.905816 | 4.287473 | -4.37982 | 3.11E-05 | 8.58E-05 | 1.801401 |
| HSD17B4 | 0.168024 | 1.123519 | 9.534176 | 4.378191 | 3.13E-05 | 8.63E-05 | 1.795491 |
| NDC80 | 0.287367 | 1.220411 | 3.853626 | 4.375867 | 3.15E-05 | 8.70E-05 | 1.787059 |
| TPP2 | 0.157329 | 1.11522 | 6.867582 | 4.375186 | 3.16E-05 | 8.72E-05 | 1.784592 |
| TSSK2 | -0.13958 | 0.907784 | 4.55989 | -4.37442 | 3.17E-05 | 8.74E-05 | 1.781827 |
| TRAV8-6 | -0.12828 | 0.914925 | 5.264176 | -4.37305 | 3.19E-05 | 8.79E-05 | 1.77683 |
| VDR | -0.30721 | 0.808202 | 7.213516 | -4.37265 | 3.19E-05 | 8.80E-05 | 1.775414 |
| NEDD4L | -0.18834 | 0.877616 | 7.631648 | -4.37252 | 3.19E-05 | 8.80E-05 | 1.77493 |
| TYMS | 0.38729 | 1.307934 | 7.99956 | 4.372451 | 3.20E-05 | 8.80E-05 | 1.774674 |
| PHIP | 0.171913 | 1.126551 | 6.841099 | 4.372413 | 3.20E-05 | 8.80E-05 | 1.774537 |
| NR2F2 | 0.200179 | 1.148841 | 7.607033 | 4.372099 | 3.20E-05 | 8.80E-05 | 1.773401 |
| CA10 | -0.23099 | 0.85205 | 4.601209 | -4.37117 | 3.21E-05 | 8.83E-05 | 1.770018 |
| IER2 | -0.44277 | 0.735719 | 9.516264 | -4.37115 | 3.21E-05 | 8.83E-05 | 1.769945 |
| TOMM70A | 0.177019 | 1.130546 | 8.253407 | 4.36958 | 3.23E-05 | 8.88E-05 | 1.764271 |
| PIGF | 0.218116 | 1.163214 | 7.283077 | 4.36958 | 3.23E-05 | 8.88E-05 | 1.76427 |
| ATOH1 | -0.15687 | 0.896969 | 4.693297 | -4.36943 | 3.23E-05 | 8.88E-05 | 1.763713 |
| NSMF | -0.21934 | 0.858957 | 6.811099 | -4.36935 | 3.23E-05 | 8.88E-05 | 1.763443 |
| TNMD | 0.763087 | 1.697118 | 5.156264 | 4.369309 | 3.23E-05 | 8.88E-05 | 1.76329 |
| C9orf91 | 0.164005 | 1.120393 | 4.611319 | 4.368991 | 3.24E-05 | 8.89E-05 | 1.762138 |
| MLH3 | 0.166213 | 1.122109 | 5.901758 | 4.368316 | 3.25E-05 | 8.91E-05 | 1.759691 |
| DPYSL2 | 0.241961 | 1.182599 | 9.447912 | 4.367815 | 3.25E-05 | 8.92E-05 | 1.757875 |
| NRGN | -0.14756 | 0.902776 | 5.526813 | -4.36723 | 3.26E-05 | 8.94E-05 | 1.755771 |
| DCTN5 | 0.116507 | 1.084107 | 6.98044 | 4.365014 | 3.29E-05 | 9.01E-05 | 1.747734 |
| AXIN1 | -0.14115 | 0.906793 | 7.028242 | -4.36455 | 3.29E-05 | 9.02E-05 | 1.746053 |
| B3GNT3 | -0.19601 | 0.872959 | 6.252418 | -4.36421 | 3.30E-05 | 9.03E-05 | 1.74481 |
| CA14 | -0.19288 | 0.874858 | 5.155055 | -4.36388 | 3.30E-05 | 9.03E-05 | 1.743633 |
| EMC7 | 0.119826 | 1.086604 | 9.191429 | 4.363861 | 3.30E-05 | 9.03E-05 | 1.743559 |
| MSN | 0.267686 | 1.203875 | 8.508242 | 4.363831 | 3.30E-05 | 9.03E-05 | 1.74345 |
| FOXF1 | -0.1624 | 0.89354 | 4.291868 | -4.36379 | 3.30E-05 | 9.03E-05 | 1.743301 |
| DMTN | -0.147 | 0.90313 | 7.192527 | -4.36067 | 3.34E-05 | 9.14E-05 | 1.73202 |
| WNT11 | -0.16452 | 0.892224 | 5.685165 | -4.36036 | 3.35E-05 | 9.15E-05 | 1.730879 |
| IDI1 | -0.22506 | 0.855558 | 8.790879 | -4.35904 | 3.36E-05 | 9.19E-05 | 1.726112 |
| C8B | -0.17914 | 0.883226 | 5.52989 | -4.3581 | 3.37E-05 | 9.22E-05 | 1.722696 |
| CCL20 | 0.917396 | 1.888703 | 5.066484 | 4.357992 | 3.38E-05 | 9.22E-05 | 1.722319 |
| MMP1 | 0.243502 | 1.183863 | 3.738022 | 4.357371 | 3.38E-05 | 9.24E-05 | 1.720074 |
| PDE6D | 0.130841 | 1.094931 | 7.062527 | 4.355038 | 3.41E-05 | 9.32E-05 | 1.711639 |
| APP | 0.226734 | 1.170183 | 9.585165 | 4.354966 | 3.41E-05 | 9.32E-05 | 1.711378 |
| CST8 | -0.15008 | 0.901199 | 4.662088 | -4.35416 | 3.42E-05 | 9.34E-05 | 1.708474 |
| SPTBN2 | -0.20327 | 0.868579 | 5.522308 | -4.35274 | 3.44E-05 | 9.39E-05 | 1.70333 |

| | | | | | | | |
|---|---|---|---|---|---|---|---|
| RANBP6 | 0.262758 | 1.19977 | 6.834066 | 4.3502 | 3.48E-05 | 9.48E-05 | 1.694154 |
| DPEP3 | -0.1528 | 0.899505 | 4.758571 | -4.34985 | 3.48E-05 | 9.49E-05 | 1.692878 |
| ODC1 | 0.201082 | 1.14956 | 10.28813 | 4.349145 | 3.49E-05 | 9.51E-05 | 1.690341 |
| C12orf4 | 0.193662 | 1.143663 | 5.690549 | 4.347844 | 3.51E-05 | 9.55E-05 | 1.685642 |
| CASP8AP2 | 0.167304 | 1.122958 | 5.301429 | 4.34748 | 3.51E-05 | 9.56E-05 | 1.684327 |
| TMPRSS4 | 0.324087 | 1.251872 | 6.544176 | 4.346633 | 3.52E-05 | 9.59E-05 | 1.681271 |
| TMEM260 | 0.185546 | 1.137247 | 6.101319 | 4.345244 | 3.54E-05 | 9.64E-05 | 1.676256 |
| THRB | -0.17502 | 0.885753 | 6.497363 | -4.34469 | 3.55E-05 | 9.66E-05 | 1.674269 |
| MRAS | 0.253039 | 1.191714 | 6.301868 | 4.344299 | 3.56E-05 | 9.67E-05 | 1.672844 |
| ZFAND1 | -0.22418 | 0.856082 | 7.771099 | -4.34423 | 3.56E-05 | 9.67E-05 | 1.672585 |
| CRABP2 | -0.20715 | 0.866247 | 6.145824 | -4.34402 | 3.56E-05 | 9.67E-05 | 1.671843 |
| SLN | -0.15454 | 0.898418 | 5.139231 | -4.34376 | 3.56E-05 | 9.68E-05 | 1.670895 |
| TRMT2B | -0.18373 | 0.880424 | 6.421319 | -4.34321 | 3.57E-05 | 9.69E-05 | 1.668916 |
| MFSD7 | -0.18818 | 0.87771 | 6.148681 | -4.3431 | 3.57E-05 | 9.70E-05 | 1.668518 |
| RBMX2 | 0.146208 | 1.106657 | 6.501648 | 4.34292 | 3.57E-05 | 9.70E-05 | 1.667867 |
| FHOD3 | 0.301406 | 1.232345 | 7.182308 | 4.342855 | 3.57E-05 | 9.70E-05 | 1.667632 |
| LRRC17 | -0.15709 | 0.896834 | 4.473407 | -4.33997 | 3.61E-05 | 9.80E-05 | 1.657241 |
| CDC73 | 0.159507 | 1.116906 | 6.696703 | 4.339582 | 3.62E-05 | 9.81E-05 | 1.655823 |
| GNG11 | 0.364981 | 1.287864 | 7.834615 | 4.338263 | 3.64E-05 | 9.86E-05 | 1.651066 |
| GOLIM4 | -0.16099 | 0.894411 | 5.885824 | -4.33683 | 3.66E-05 | 9.91E-05 | 1.645917 |
| TBCC | 0.182957 | 1.135208 | 7.563516 | 4.336801 | 3.66E-05 | 9.91E-05 | 1.645797 |
| OAS1 | 0.560729 | 1.475015 | 6.23011 | 4.336426 | 3.66E-05 | 9.92E-05 | 1.644444 |
| DR1 | 0.155 | 1.113422 | 6.341648 | 4.336039 | 3.67E-05 | 9.93E-05 | 1.643047 |
| PCDHA10 | -0.15599 | 0.897519 | 4.503516 | -4.33596 | 3.67E-05 | 9.93E-05 | 1.642754 |
| HAPLN2 | -0.16986 | 0.888926 | 4.672088 | -4.3356 | 3.67E-05 | 9.94E-05 | 1.641476 |

**Table S2. CKD-Blue enriched geneset**

| *NAME* | *SIZE* | *ES* | *NES* | *NOM p-val* | *FDR q-val* |
|---|---|---|---|---|---|
| HALLMARK_INTERFERON_ALPHA_RESPONSE | 73 | 0.70697904 | 2.401329 | 0 | 0 |
| REACTOME_IMMUNOREGULATORY_INTERACTIONS_BETWEEN_A_LYMPHOID_AND_A_NON_LYMPHOID_CELL | 88 | 0.6503452 | 2.2540326 | 0 | 0 |
| REACTOME_INTERFERON_ALPHA_BETA_SIGNALING | 55 | 0.6127595 | 1.9683331 | 0 | 0.0209609 |
| GO_CELLULAR_DEFENSE_RESPONSE | 36 | 0.66562515 | 1.9735125 | 0 | 0.02286122 |
| GO_NEGATIVE_REGULATION_OF_VIRAL_GENOME_REPLICATION | 39 | 0.6566933 | 1.9766963 | 0 | 0.02875609 |
| KEGG_TYPE_I_DIABETES_MELLITUS | 28 | 0.68634254 | 1.9211932 | 0 | 0.03665807 |
| HALLMARK_ALLOGRAFT_REJECTION | 148 | 0.50134385 | 1.9084237 | 0 | 0.039381754 |
| KEGG_ALLOGRAFT_REJECTION | 24 | 0.7093952 | 1.893494 | 0 | 0.03938945 |
| REACTOME_ANTIVIRAL_MECHANISM_BY_IFN_STIMULATED_GENES | 54 | 0.5924195 | 1.8861372 | 0 | 0.040561426 |
| REACTOME_INTERFERON_SIGNALING | 132 | 0.5130978 | 1.898133 | 0 | 0.041903697 |
| HALLMARK_TNFA_SIGNALING_VIA_NFKB | 175 | -0.83062816 | -2.7448096 | 0 | 0 |
| HALLMARK_HYPOXIA | 153 | -0.69562817 | -2.2672408 | 0 | 0 |
| HALLMARK_UV_RESPONSE_UP | 116 | -0.70585686 | -2.239725 | 0 | 0 |
| PID_ATF2_PATHWAY | 35 | -0.8456175 | -2.2285554 | 0 | 0 |
| HALLMARK_APOPTOSIS | 123 | -0.6905139 | -2.172021 | 0 | 2.32E-04 |
| REACTOME_NUCLEAR_EVENTS_KINASE_AND_TRANSCRIPTION_FACTOR_ACTIVATION | 35 | -0.81615317 | -2.1120372 | 0 | 2.58E-04 |
| REACTOME_NGF_STIMULATED_TRANSCRIPTION | 27 | -0.8399939 | -2.1341226 | 0 | 2.91E-04 |
| PID_AP1_PATHWAY | 48 | -0.77188134 | -2.1419587 | 0 | 3.32E-04 |
| GO_P38MAPK_CASCADE | 27 | -0.8545522 | -2.163108 | 0 | 3.88E-04 |
| GO_REGULATION_OF_EPITHELIAL_CELL_DIFFERENTIATION | 94 | -0.68354857 | -2.0781128 | 0 | 5.79E-04 |
| GO_CELLULAR_RESPONSE_TO_STARVATION | 84 | -0.69371736 | -2.0881803 | 0 | 6.32E-04 |
| GO_RESPONSE_TO_STARVATION | 111 | -0.6752055 | -2.0894158 | 0 | 6.95E-04 |
| GO_SKELETAL_MUSCLE_CELL_DIFFERENTIATION | 27 | -0.83449644 | -2.068933 | 0 | 8.00E-04 |
| GO_NEGATIVE_REGULATION_OF_PROTEIN_SERINE_THREONINE_KINASE_ACTIVITY | 82 | -0.6720453 | -2.044121 | 0 | 0.001239647 |
| PID_SMAD2_3NUCLEAR_PATHWAY | 49 | -0.7398351 | -2.0355604 | 0 | 0.001618643 |
| GO_FAT_CELL_DIFFERENTIATION | 121 | -0.6333734 | -2.0297682 | 0 | 0.001803759 |
| GO_NEGATIVE_REGULATION_OF_TRANSFERASE_ACTIVITY | 157 | -0.6236511 | -2.0040205 | 0 | 0.002919317 |
| HALLMARK_IL2_STAT5_SIGNALING | 137 | -0.62587684 | -2.0053234 | 0 | 0.00295514 |
| GO_MONOSACCHARIDE_BIOSYNTHETIC_PROCESS | 64 | -0.6794318 | -2.007393 | 0 | 0.003128972 |
| GO_MUSCLE_TISSUE_DEVELOPMENT | 202 | -0.59344834 | -1.9936025 | 0 | 0.003350122 |
| GO_SKELETAL_MUSCLE_ORGAN_DEVELOPMENT | 75 | -0.67626023 | -1.9874661 | 0 | 0.003901186 |
| REACTOME_THE_CITRIC_ACID_TCA_CYCLE_AND_RESPIRATORY_ELECTRON_TRANSPORT | 93 | -0.6394663 | -1.9807293 | 0 | 0.004180828 |
| PID_FOXO_PATHWAY | 36 | -0.7465937 | -1.9818312 | 0 | 0.004262608 |
| HALLMARK_P53_PATHWAY | 148 | -0.6119882 | -1.9832422 | 0 | 0.004350977 |
| PID_IL6_7_PATHWAY | 35 | -0.75684047 | -1.9704227 | 0 | 0.005038227 |
| PID_ERBB1_DOWNSTREAM_PATHWAY | 75 | -0.662648 | -1.9712172 | 0 | 0.005143076 |
| GO_NEGATIVE_REGULATION_OF_KINASE_ACTIVITY | 143 | -0.615533 | -1.9674637 | 0 | 0.005188041 |
| GO_KINASE_INHIBITOR_ACTIVITY | 38 | -0.7390166 | -1.9668684 | 0 | 0.00520724 |
| HALLMARK_ESTROGEN_RESPONSE_EARLY | 141 | -0.61476165 | -1.9716511 | 0 | 0.005303158 |
| REACTOME_SIGNALING_BY_NTRKS | 84 | -0.65229577 | -1.963201 | 0 | 0.0054552 |
| PID_TAP63_PATHWAY | 39 | -0.7247743 | -1.9577416 | 0 | 0.006064393 |
| GO_DNA_BINDING_TRANSCRIPTION_FACTOR_BINDING | 198 | -0.58109826 | -1.9538983 | 0 | 0.006561243 |
| HALLMARK_ANDROGEN_RESPONSE | 74 | -0.6481518 | -1.9421616 | 0 | 0.008386841 |
| GO_NEGATIVE_REGULATION_OF_MAP_KINASE_ACTIVITY | 49 | -0.69819593 | -1.9371995 | 0 | 0.008917522 |
| GO_GLUCOSE_METABOLIC_PROCESS | 125 | -0.6091817 | -1.9329743 | 0 | 0.009254478 |
| HALLMARK_CHOLESTEROL_HOMEOSTASIS | 50 | -0.69416386 | -1.9332062 | 0 | 0.009486132 |
| GO_RNA_POLYMERASE_II_SPECIFIC_DNA_BINDING_TRANSCRIPTION_FACTOR_BINDING | 159 | -0.5894504 | -1.9290084 | 0 | 0.009877053 |
| GO_TRANSCRIPTION_FACTOR_BINDING | 356 | -0.54382837 | -1.9229456 | 0 | 0.010983478 |

| | | | | | |
|---|---|---|---|---|---|
| GO_PLATELET_DERIVED_GROWTH_FACTOR_RECEPTOR_SIGNALING_PATHWAY | 36 | -0.7289513 | -1.9130274 | 0 | 0.012882965 |
| GO_REGULATION_OF_EPIDERMAL_CELL_DIFFERENTIATION | 34 | -0.7434667 | -1.9141777 | 0.001631321 | 0.012946786 |
| GO_NEGATIVE_REGULATION_OF_PROTEIN_MODIFICATION_PROCESS | 343 | -0.5469002 | -1.9013855 | 0 | 0.013005317 |
| GO_PROTEIN_TYROSINE_SERINE_THREONINE_PHOSPHATASE_ACTIVITY | 21 | -0.79851407 | -1.8996131 | 0 | 0.013020961 |
| HALLMARK_OXIDATIVE_PHOSPHORYLATION | 143 | -0.585117 | -1.8981546 | 0 | 0.013192682 |
| KEGG_MAPK_SIGNALING_PATHWAY | 166 | -0.58063966 | -1.9014492 | 0 | 0.013247353 |
| GO_RESPONSE_TO_CAMP | 69 | -0.6402604 | -1.8967516 | 0 | 0.013465758 |
| GO_NEGATIVE_REGULATION_OF_MRNA_METABOLIC_PROCESS | 38 | -0.7176737 | -1.9016762 | 0 | 0.013498969 |
| GO_NEGATIVE_REGULATION_OF_DNA_BINDING_TRANSCRIPTION_FACTOR_ACTIVITY | 86 | -0.6230947 | -1.9083968 | 0 | 0.013612621 |
| PID_P53_DOWNSTREAM_PATHWAY | 92 | -0.62016773 | -1.9068199 | 0 | 0.013725613 |
| GO_REGULATION_OF_FAT_CELL_DIFFERENTIATION | 69 | -0.65297747 | -1.9089084 | 0.001526718 | 0.013746833 |
| GO_CELLULAR_RESPONSE_TO_IONIZING_RADIATION | 35 | -0.74278885 | -1.9049602 | 0 | 0.013781395 |
| GO_REGULATION_OF_GLUCOSE_METABOLIC_PROCESS | 65 | -0.6576233 | -1.9017991 | 0 | 0.013786182 |
| GO_NUCLEAR_RECEPTOR_BINDING | 61 | -0.65563416 | -1.9038528 | 0.001540832 | 0.013860978 |
| GO_INACTIVATION_OF_MAPK_ACTIVITY | 18 | -0.8054818 | -1.9018669 | 0.001851852 | 0.014060816 |
| GO_CELLULAR_RESPONSE_TO_EXTRACELLULAR_STIMULUS | 160 | -0.57528716 | -1.8905765 | 0 | 0.014711431 |
| GO_REGULATION_OF_OSSIFICATION | 101 | -0.60701764 | -1.8880504 | 0 | 0.015095102 |
| GO_CIRCADIAN_RHYTHM | 121 | -0.59083664 | -1.8834752 | 0 | 0.015106598 |
| GO_EMBRYONIC_PLACENTA_MORPHOGENESIS | 16 | -0.8354626 | -1.8838549 | 0.001912046 | 0.015307354 |
| GO_GTPASE_ACTIVITY | 168 | -0.57291836 | -1.886213 | 0 | 0.015340773 |
| GO_NEGATIVE_REGULATION_OF_MAPK_CASCADE | 104 | -0.6091863 | -1.8838776 | 0 | 0.015575904 |
| GO_POSITIVE_REGULATION_OF_P38MAPK_CASCADE | 18 | -0.80663437 | -1.8708954 | 0 | 0.017990027 |
| REACTOME_NEGATIVE_REGULATION_OF_MAPK_PATHWAY | 20 | -0.7978409 | -1.8712628 | 0 | 0.018150233 |
| GO_NEGATIVE_REGULATION_OF_PHOSPHORYLATION | 267 | -0.5459901 | -1.873349 | 0 | 0.018176721 |
| HALLMARK_TGF_BETA_SIGNALING | 41 | -0.7021484 | -1.8717458 | 0 | 0.01829561 |
| REACTOME_PYRUVATE_METABOLISM_AND_CITRIC_ACID_TCA_CYCLE | 40 | -0.68700486 | -1.8684243 | 0 | 0.01880626 |
| GO_RESPONSE_TO_THYROID_HORMONE | 20 | -0.8017767 | -1.8661026 | 0 | 0.019262884 |
| HALLMARK_MTORC1_SIGNALING | 150 | -0.57299834 | -1.8585124 | 0 | 0.021235706 |
| GO_CELLULAR_RESPONSE_TO_HORMONE_STIMULUS | 411 | -0.52183646 | -1.8593156 | 0 | 0.021276645 |
| GO_PROTEIN_SERINE_THREONINE_KINASE_INHIBITOR_ACTIVITY | 16 | -0.82539296 | -1.859634 | 0.001876173 | 0.021473186 |
| REACTOME_TOLL_LIKE_RECEPTOR_10_TLR10_CASCADE | 39 | -0.69346017 | -1.8598784 | 0.00166113 | 0.021729186 |
| REACTOME_RAF_INDEPENDENT_MAPK1_3_ACTIVATION | 17 | -0.8282311 | -1.8564632 | 0 | 0.021870077 |
| REACTOME_RESPONSE_OF_EIF2AK4_GCN2_TO_AMINO_ACID_DEFICIENCY | 37 | -0.69311905 | -1.8492026 | 0 | 0.025152793 |
| GO_MUSCLE_CELL_PROLIFERATION | 131 | -0.5757095 | -1.8452152 | 0 | 0.026519116 |
| GO_NEGATIVE_REGULATION_OF_PHOSPHORUS_METABOLIC_PROCESS | 319 | -0.5302398 | -1.842547 | 0 | 0.026641332 |
| GO_POSITIVE_REGULATION_OF_OSSIFICATION | 52 | -0.64713603 | -1.842928 | 0.001623377 | 0.026767699 |
| GO_NUCLEAR_HORMONE_RECEPTOR_BINDING | 90 | -0.6052684 | -1.8440144 | 0 | 0.026818251 |
| GO_POSITIVE_REGULATION_OF_OSTEOBLAST_DIFFERENTIATION | 37 | -0.696446 | -1.8392848 | 0 | 0.027580457 |
| GO_PERK_MEDIATED_UNFOLDED_PROTEIN_RESPONSE | 16 | -0.8105782 | -1.8400416 | 0.001855288 | 0.027596964 |
| GO_CELLULAR_RESPONSE_TO_EXTERNAL_STIMULUS | 211 | -0.54565525 | -1.8384356 | 0 | 0.02762593 |
| PID_HIF1_TFPATHWAY | 53 | -0.6533825 | -1.8369089 | 0.001594896 | 0.02813603 |
| HALLMARK_UNFOLDED_PROTEIN_RESPONSE | 74 | -0.6107912 | -1.8347086 | 0 | 0.028302422 |
| GO_MONOSACCHARIDE_METABOLIC_PROCESS | 172 | -0.55361366 | -1.8359715 | 0 | 0.028331753 |
| GO_RESPONSE_TO_AMINO_ACID_STARVATION | 26 | -0.74365324 | -1.8347828 | 0 | 0.028623104 |
| GO_HORMONE_RECEPTOR_BINDING | 104 | -0.5954186 | -1.8303574 | 0 | 0.028928872 |
| GO_CIS_REGULATORY_REGION_BINDING | 308 | -0.5242206 | -1.8306017 | 0 | 0.029093463 |
| REACTOME_RHO_GTPASES_ACTIVATE_ROCKS | 15 | -0.8081595 | -1.8306732 | 0.001828154 | 0.02938482 |
| GO_RESPONSE_TO_PEPTIDE | 318 | -0.5204943 | -1.8311347 | 0 | 0.029503081 |

| | | | | | |
|---|---|---|---|---|---|
| GO_INTRINSIC_APOPTOTIC_SIGNALING_PATHWAY | 176 | -0.5467616 | -1.8264143 | 0 | 0.030262794 |
| BIOCARTA_KERATINOCYTE_PATHWAY | 35 | -0.69199324 | -1.8268498 | 0 | 0.030385412 |
| REACTOME_PERK_REGULATES_GENE_EXPRESSION | 16 | -0.82316774 | -1.8243421 | 0.001964637 | 0.031036165 |
| GO_FACE_DEVELOPMENT | 28 | -0.73653626 | -1.823684 | 0 | 0.031139567 |
| REACTOME_INTERLEUKIN_4_AND_INTERLEUKIN_13_SIGNALING | 77 | -0.6105357 | -1.820077 | 0.001445087 | 0.031795837 |
| GO_ENDOTHELIAL_CELL_CHEMOTAXIS | 17 | -0.7879509 | -1.8203732 | 0.003731343 | 0.031990863 |
| GO_CELLULAR_RESPONSE_TO_CALCIUM_ION | 47 | -0.657398 | -1.819018 | 0 | 0.03204498 |
| REACTOME_TRANSCRIPTIONAL_REGULATION_OF_GRANULOPOIESIS | 18 | -0.7906674 | -1.8208548 | 0 | 0.032075346 |
| GO_CIRCADIAN_REGULATION_OF_GENE_EXPRESSION | 41 | -0.67421335 | -1.8145554 | 0 | 0.03416801 |
| GO_CORE_PROMOTER_BINDING | 26 | -0.72226983 | -1.8137445 | 0.003636364 | 0.034257933 |
| GO_POSITIVE_REGULATION_OF_FAT_CELL_DIFFERENTIATION | 32 | -0.71536744 | -1.8116666 | 0.003490401 | 0.034383345 |
| GO_REGULATION_OF_STEROID_BIOSYNTHETIC_PROCESS | 66 | -0.62129366 | -1.8111413 | 0.00304414 | 0.034454316 |
| GO_RESPONSE_TO_EXTRACELLULAR_STIMULUS | 325 | -0.5165384 | -1.8120238 | 0 | 0.034510065 |
| GO_REGULATION_OF_KERATINOCYTE_DIFFERENTIATION | 19 | -0.7715206 | -1.8104521 | 0.001821494 | 0.034546785 |
| GO_RESPONSE_TO_COLD | 33 | -0.6943842 | -1.808575 | 0.001748252 | 0.0346486 |
| GO_RESPONSE_TO_PEPTIDE_HORMONE | 268 | -0.5290487 | -1.8124175 | 0 | 0.0346754 |
| PID_REG_GR_PATHWAY | 51 | -0.6431772 | -1.809151 | 0 | 0.034682527 |
| REACTOME_ESTROGEN_DEPENDENT_GENE_EXPRESSION | 52 | -0.6488219 | -1.8095019 | 0 | 0.034740023 |
| GO_REGULATION_OF_STEROID_METABOLIC_PROCESS | 84 | -0.5957946 | -1.8023158 | 0.00141844 | 0.037416242 |
| PID_MTOR_4PATHWAY | 47 | -0.6597635 | -1.8028271 | 0.001628664 | 0.037453786 |
| GO_MRNA_3_UTR_BINDING | 43 | -0.6682957 | -1.8015141 | 0.001620746 | 0.037639655 |
| GO_KETONE_BIOSYNTHETIC_PROCESS | 30 | -0.71884555 | -1.7998961 | 0.001776199 | 0.037784018 |
| GO_CORE_PROMOTER_SEQUENCE_SPECIFIC_DNA_BINDING | 19 | -0.7724514 | -1.7990643 | 0 | 0.03786039 |
| GO_REGULATION_OF_CELLULAR_CARBOHYDRATE_METABOLIC_PROCESS | 82 | -0.6041915 | -1.8002603 | 0 | 0.03799553 |
| REACTOME_FOXO_MEDIATED_TRANSCRIPTION | 43 | -0.66676396 | -1.7980205 | 0 | 0.038141947 |
| GO_REGULATION_OF_MAP_KINASE_ACTIVITY | 198 | -0.53645825 | -1.7971041 | 0 | 0.038315 |
| PID_S1P_S1P2_PATHWAY | 18 | -0.7886778 | -1.7955271 | 0.007259528 | 0.038750347 |
| REACTOME_UNFOLDED_PROTEIN_RESPONSE_UPR | 55 | -0.64547247 | -1.79435 | 0 | 0.03915964 |
| GO_BROWN_FAT_CELL_DIFFERENTIATION | 26 | -0.72054094 | -1.7922918 | 0.003669725 | 0.03986524 |
| GO_NEGATIVE_REGULATION_OF_INTRINSIC_APOPTOTIC_SIGNALING_PATHWAY | 61 | -0.6167787 | -1.7927958 | 0 | 0.03989101 |
| PID_IL3_PATHWAY | 16 | -0.7988491 | -1.7888 | 0 | 0.041752513 |
| GO_REGULATION_OF_HEART_GROWTH | 44 | -0.6434592 | -1.7808244 | 0 | 0.046851072 |
| REACTOME_PYRUVATE_METABOLISM | 22 | -0.7370138 | -1.779453 | 0.005681818 | 0.047051936 |
| PID_NOTCH_PATHWAY | 29 | -0.7192823 | -1.7797679 | 0 | 0.047195192 |
| REACTOME_TOLL_LIKE_RECEPTOR_TLR1_TLR2_CASCADE | 48 | -0.63790137 | -1.7782223 | 0 | 0.047578283 |
| REACTOME_RESPIRATORY_ELECTRON_TRANSPORT | 54 | -0.63224 | -1.7744325 | 0 | 0.04986309 |

**Table S2. CKD-Gold enriched geneset**

| *NAME* | *SIZE* | *ES* | *NES* | *NOM p-val* | *FDR q-val* |
|---|---|---|---|---|---|
| HALLMARK_INTERFERON_GAMMA_RESPONSE | 158 | 0.72248185 | 2.7589803 | 0 | 0 |
| HALLMARK_ALLOGRAFT_REJECTION | 148 | 0.71396226 | 2.7121916 | 0 | 0 |
| HALLMARK_INTERFERON_ALPHA_RESPONSE | 73 | 0.77763826 | 2.638582 | 0 | 0 |
| REACTOME_INTERFERON_SIGNALING | 132 | 0.68524617 | 2.5763793 | 0 | 0 |
| REACTOME_IMMUNOREGULATORY_INTERACTIONS_BETWEEN_A_LYMPHOID_AND_A_NON_LYMPHOID_CELL | 88 | 0.721665 | 2.5371282 | 0 | 0 |
| REACTOME_INTERFERON_ALPHA_BETA_SIGNALING | 55 | 0.7552968 | 2.4802504 | 0 | 0 |
| GO_B_CELL_MEDIATED_IMMUNITY | 94 | 0.68875885 | 2.468712 | 0 | 0 |
| GO_ADAPTIVE_IMMUNE_RESPONSE | 283 | 0.5946575 | 2.3951645 | 0 | 0 |
| GO_ACTIVATION_OF_IMMUNE_RESPONSE | 402 | 0.56711304 | 2.3847802 | 0 | 0 |
| REACTOME_INTEGRIN_CELL_SURFACE_INTERACTIONS | 65 | 0.7085641 | 2.371726 | 0 | 0 |
| GO_LYMPHOCYTE_MEDIATED_IMMUNITY | 178 | 0.6122509 | 2.3692725 | 0 | 0 |
| KEGG_CELL_ADHESION_MOLECULES_CAMS | 94 | 0.65757346 | 2.3681655 | 0 | 0 |
| REACTOME_INTERFERON_GAMMA_SIGNALING | 61 | 0.7121832 | 2.3598552 | 0 | 0 |
| GO_IMMUNE_RESPONSE_REGULATING_CELL_SURFACE_RECEPTOR_SIGNALING_PATHWAY | 262 | 0.5836106 | 2.359081 | 0 | 0 |
| GO_ADAPTIVE_IMMUNE_RESPONSE_BASED_ON_SOMATIC_RECOMBINATION_OF_IMMUNE_RECEPTORS_BUILT_FROM_IMMUNOGLOBULIN_SUPERFAMILY_DOMAINS | 181 | 0.59985775 | 2.3533556 | 0 | 0 |
| KEGG_SYSTEMIC_LUPUS_ERYTHEMATOSUS | 37 | 0.77003413 | 2.3486228 | 0 | 0 |
| KEGG_VIRAL_MYOCARDITIS | 51 | 0.7261803 | 2.3374794 | 0 | 0 |
| GO_PHAGOCYTOSIS | 190 | 0.59491885 | 2.3340962 | 0 | 0 |
| GO_CYTOKINE_MEDIATED_SIGNALING_PATHWAY | 483 | 0.5421821 | 2.329726 | 0 | 0 |
| GO_HUMORAL_IMMUNE_RESPONSE_MEDIATED_BY_CIRCULATING_IMMUNOGLOBULIN | 53 | 0.71120507 | 2.3203897 | 0 | 0 |
| GO_INTERFERON_GAMMA_MEDIATED_SIGNALING_PATHWAY | 59 | 0.70186967 | 2.3080623 | 0 | 0 |
| GO_RESPONSE_TO_TYPE_I_INTERFERON | 67 | 0.6813517 | 2.3062305 | 0 | 0 |
| GO_ANTIGEN_RECEPTOR_MEDIATED_SIGNALING_PATHWAY | 165 | 0.6001997 | 2.3048716 | 0 | 0 |
| GO_EXTRACELLULAR_MATRIX_STRUCTURAL_CONSTITUENT | 103 | 0.63823676 | 2.3024452 | 0 | 0 |
| GO_LEUKOCYTE_PROLIFERATION | 189 | 0.5856273 | 2.2867763 | 0 | 0 |
| GO_RESPONSE_TO_INTERFERON_GAMMA | 128 | 0.61133206 | 2.2828174 | 0 | 0 |
| GO_IMMUNE_RESPONSE_REGULATING_SIGNALING_PATHWAY | 365 | 0.547853 | 2.271127 | 0 | 0 |
| GO_REGULATION_OF_LEUKOCYTE_PROLIFERATION | 138 | 0.60507023 | 2.2657256 | 0 | 0 |
| GO_POSITIVE_REGULATION_OF_CELL_ACTIVATION | 222 | 0.5638499 | 2.2610543 | 0 | 0 |
| GO_PHAGOCYTIC_VESICLE | 86 | 0.6434419 | 2.2501915 | 0 | 0 |
| GO_REGULATION_OF_RESPONSE_TO_CYTOKINE_STIMULUS | 110 | 0.6202597 | 2.2481072 | 0 | 0 |
| GO_POSITIVE_REGULATION_OF_TUMOR_NECROSIS_FACTOR_SUPERFAMILY_CYTOKINE_PRODUCTION | 57 | 0.6782891 | 2.247096 | 0 | 0 |
| GO_B_CELL_RECEPTOR_SIGNALING_PATHWAY | 48 | 0.7162396 | 2.2436428 | 0 | 0 |
| GO_MEMBRANE_INVAGINATION | 56 | 0.68579096 | 2.2381368 | 0 | 0 |
| GO_HUMORAL_IMMUNE_RESPONSE | 163 | 0.5861149 | 2.2359755 | 0 | 0 |
| REACTOME_ECM_PROTEOGLYCANS | 50 | 0.6859998 | 2.234548 | 0 | 0 |
| KEGG_LEISHMANIA_INFECTION | 42 | 0.72038966 | 2.2193074 | 0 | 0 |
| HALLMARK_INFLAMMATORY_RESPONSE | 156 | 0.5809337 | 2.2176056 | 0 | 0 |
| GO_NEGATIVE_REGULATION_OF_VIRAL_GENOME_REPLICATION | 39 | 0.72393405 | 2.2168684 | 0 | 0 |
| GO_POSITIVE_REGULATION_OF_LEUKOCYTE_PROLIFERATION | 85 | 0.6312752 | 2.215714 | 0 | 0 |
| KEGG_ANTIGEN_PROCESSING_AND_PRESENTATION | 57 | 0.6785498 | 2.2115698 | 0 | 0 |
| KEGG_ALLOGRAFT_REJECTION | 24 | 0.8130395 | 2.2059782 | 0 | 0 |
| KEGG_ECM_RECEPTOR_INTERACTION | 59 | 0.6697777 | 2.2059617 | 0 | 0 |

| | | | | | |
|---|---|---|---|---|---|
| GO_TUMOR_NECROSIS_FACTOR_SUPERFAMILY_CYTOKINE_PRODUCTION | 97 | 0.6119537 | 2.2045088 | 0 | 0 |
| GO_POSITIVE_REGULATION_OF_CYTOKINE_BIOSYNTHETIC_PROCESS | 40 | 0.7172684 | 2.2031846 | 0 | 0 |
| REACTOME_TCR_SIGNALING | 73 | 0.63900286 | 2.1990075 | 0 | 0 |
| GO_POSITIVE_REGULATION_OF_T_CELL_PROLIFERATION | 59 | 0.65837723 | 2.197328 | 0 | 0 |
| KEGG_AUTOIMMUNE_THYROID_DISEASE | 33 | 0.7461788 | 2.1942272 | 0 | 0 |
| REACTOME_ANTIVIRAL_MECHANISM_BY_IFN_STIMULATED_GENES | 54 | 0.6732106 | 2.1888206 | 0 | 0 |
| KEGG_GRAFT_VERSUS_HOST_DISEASE | 28 | 0.76720786 | 2.1845777 | 0 | 0 |
| GO_RESPONSE_TO_INTERFERON_BETA | 21 | 0.8094829 | 2.1827152 | 0 | 0 |
| GO_REGULATION_OF_ADAPTIVE_IMMUNE_RESPONSE | 99 | 0.606733 | 2.1768272 | 0 | 0 |
| GO_ORGANIC_ACID_CATABOLIC_PROCESS | 190 | -0.7486109 | -2.989512 | 0 | 0 |
| GO_MONOCARBOXYLIC_ACID_CATABOLIC_PROCESS | 89 | -0.7941556 | -2.828716 | 0 | 0 |
| GO_SMALL_MOLECULE_CATABOLIC_PROCESS | 296 | -0.6835396 | -2.827341 | 0 | 0 |
| GO_MITOCHONDRIAL_MATRIX | 283 | -0.6732215 | -2.774104 | 0 | 0 |
| GO_CELLULAR_AMINO_ACID_CATABOLIC_PROCESS | 89 | -0.7722317 | -2.753503 | 0 | 0 |
| HALLMARK_OXIDATIVE_PHOSPHORYLATION | 143 | -0.7195928 | -2.749675 | 0 | 0 |
| GO_FATTY_ACID_CATABOLIC_PROCESS | 73 | -0.7921725 | -2.737742 | 0 | 0 |
| GO_ALPHA_AMINO_ACID_METABOLIC_PROCESS | 130 | -0.712034 | -2.698991 | 0 | 0 |
| GO_ALPHA_AMINO_ACID_CATABOLIC_PROCESS | 72 | -0.7733498 | -2.670438 | 0 | 0 |
| GO_FATTY_ACID_BETA_OXIDATION | 54 | -0.7927936 | -2.655567 | 0 | 0 |
| GO_LIPID_OXIDATION | 77 | -0.7565406 | -2.639187 | 0 | 0 |
| GO_COENZYME_BINDING | 213 | -0.6502039 | -2.599875 | 0 | 0 |
| REACTOME_PROTEIN_LOCALIZATION | 97 | -0.7045046 | -2.570181 | 0 | 0 |
| GO_COFACTOR_BINDING | 345 | -0.6018902 | -2.540683 | 0 | 0 |
| GO_CELLULAR_RESPIRATION | 101 | -0.6950992 | -2.536949 | 0 | 0 |
| GO_CELLULAR_AMINO_ACID_METABOLIC_PROCESS | 203 | -0.6376154 | -2.528089 | 0 | 0 |
| KEGG_VALINE_LEUCINE_AND_ISOLEUCINE_DEGRADATION | 38 | -0.8146973 | -2.527015 | 0 | 0 |
| REACTOME_THE_CITRIC_ACID_TCA_CYCLE_AND_RESPIRATORY_ELECTRON_TRANSPORT | 93 | -0.696444 | -2.512391 | 0 | 0 |
| GO_MITOCHONDRIAL_PROTEIN_COMPLEX | 120 | -0.6629022 | -2.481446 | 0 | 0 |
| REACTOME_RESPIRATORY_ELECTRON_TRANSPORT | 54 | -0.7404892 | -2.439099 | 0 | 0 |
| GO_CELLULAR_LIPID_CATABOLIC_PROCESS | 137 | -0.6406441 | -2.435594 | 0 | 0 |
| REACTOME_MITOCHONDRIAL_FATTY_ACID_BETA_OXIDATION | 24 | -0.8526335 | -2.416459 | 0 | 0 |
| GO_COENZYME_METABOLIC_PROCESS | 160 | -0.6320059 | -2.405803 | 0 | 0 |
| GO_LIPID_MODIFICATION | 145 | -0.6326164 | -2.403342 | 0 | 0 |
| GO_INNER_MITOCHONDRIAL_MEMBRANE_PROTEIN_COMPLEX | 51 | -0.7402148 | -2.386925 | 0 | 0 |
| GO_OXIDOREDUCTASE_ACTIVITY | 455 | -0.5554379 | -2.375537 | 0 | 0 |
| GO_MICROBODY | 84 | -0.6618079 | -2.365251 | 0 | 0 |
| KEGG_PEROXISOME | 60 | -0.6964842 | -2.357178 | 0 | 0 |
| GO_DICARBOXYLIC_ACID_METABOLIC_PROCESS | 61 | -0.7058827 | -2.356236 | 0 | 0 |
| GO_COFACTOR_METABOLIC_PROCESS | 281 | -0.5711477 | -2.35424 | 0 | 0 |
| GO_MICROBODY_LUMEN | 34 | -0.7851597 | -2.349446 | 0 | 0 |
| KEGG_FATTY_ACID_METABOLISM | 33 | -0.7949361 | -2.348957 | 0 | 0 |
| KEGG_BUTANOATE_METABOLISM | 29 | -0.8034571 | -2.347224 | 0 | 0 |
| REACTOME_PEROXISOMAL_PROTEIN_IMPORT | 45 | -0.7470961 | -2.340099 | 0 | 0 |
| GO_AEROBIC_RESPIRATION | 52 | -0.7206347 | -2.337018 | 0 | 0 |
| KEGG_ARGININE_AND_PROLINE_METABOLISM | 41 | -0.7508987 | -2.335903 | 0 | 0 |
| GO_MITOCHONDRIAL_GENE_EXPRESSION | 85 | -0.6488451 | -2.333363 | 0 | 0 |
| GO_RESPIRATORY_CHAIN_COMPLEX | 37 | -0.7661789 | -2.331075 | 0 | 0 |

| | | | | | |
|---|---|---|---|---|---|
| REACTOME_RESPIRATORY_ELECTRON_TRANSPORT_ATP_SYNTHESIS_BY_CHEMIOSMOTIC_COUPLING_AND_HEAT_PRODUCTION_BY_UNCOUPLING_PROTEINS | 55 | -0.7018408 | -2.316378 | 0 | 0 |
| HALLMARK_FATTY_ACID_METABOLISM | 125 | -0.6209004 | -2.315157 | 0 | 0 |
| GO_PEROXISOMAL_TRANSPORT | 50 | -0.7156619 | -2.314535 | 0 | 0 |
| GO_RESPIRASOME | 48 | -0.7095763 | -2.308672 | 0 | 0 |
| GO_MITOCHONDRIAL_RESPIRATORY_CHAIN_COMPLEX_ASSEMBLY | 40 | -0.7413091 | -2.300767 | 0 | 0 |
| GO_ELECTRON_TRANSPORT_CHAIN | 108 | -0.6257538 | -2.300532 | 0 | 0 |
| GO_RESPIRATORY_ELECTRON_TRANSPORT_CHAIN | 57 | -0.6922551 | -2.290569 | 0 | 0 |
| HALLMARK_XENOBIOTIC_METABOLISM | 169 | -0.5940884 | -2.287118 | 0 | 0 |
| GO_ENERGY_DERIVATION_BY_OXIDATION_OF_ORGANIC_COMPOUNDS | 160 | -0.599901 | -2.285412 | 0 | 0 |
| REACTOME_BRANCHED_CHAIN_AMINO_ACID_CATABOLISM | 19 | -0.8759688 | -2.280634 | 0 | 0 |
| GO_PEROXISOME_ORGANIZATION | 57 | -0.6868153 | -2.279625 | 0 | 0 |
| REACTOME_SULFUR_AMINO_ACID_METABOLISM | 20 | -0.8420649 | -2.276723 | 0 | 0 |
| KEGG_TRYPTOPHAN_METABOLISM | 30 | -0.779476 | -2.274525 | 0 | 0 |
| GO_CELLULAR_MODIFIED_AMINO_ACID_METABOLIC_PROCESS | 120 | -0.6139822 | -2.266874 | 0 | 0 |
| REACTOME_GLUCONEOGENESIS | 26 | -0.8018687 | -2.26687 | 0 | 0 |
| GO_MITOCHONDRIAL_ENVELOPE | 364 | -0.5409133 | -2.260006 | 0 | 0 |
| HALLMARK_ADIPOGENESIS | 143 | -0.5856509 | -2.255389 | 0 | 0 |
| GO_SULFUR_COMPOUND_METABOLIC_PROCESS | 235 | -0.5576224 | -2.24919 | 0 | 0 |
| GO_OXIDOREDUCTASE_ACTIVITY_ACTING_ON_THE_CH_CH_GROUP_OF_DONORS | 40 | -0.6958904 | -2.170412 | 0 | 8.85E-06 |
| GO_NADH_DEHYDROGENASE_COMPLEX_ASSEMBLY | 31 | -0.7467698 | -2.173984 | 0 | 8.97E-06 |
| GO_WATER_SOLUBLE_VITAMIN_METABOLIC_PROCESS | 54 | -0.6560282 | -2.174612 | 0 | 9.08E-06 |
| KEGG_OXIDATIVE_PHOSPHORYLATION | 63 | -0.6465343 | -2.179238 | 0 | 9.20E-06 |
| KEGG_GLYCINE_SERINE_AND_THREONINE_METABOLISM | 24 | -0.7881349 | -2.181455 | 0 | 9.32E-06 |
| REACTOME_GLYOXYLATE_METABOLISM_AND_GLYCINE_DEGRADATION | 26 | -0.7760737 | -2.182416 | 0 | 9.45E-06 |
| REACTOME_PYRUVATE_METABOLISM_AND_CITRIC_ACID_TCA_CYCLE | 40 | -0.7043501 | -2.184156 | 0 | 9.58E-06 |
| GO_LIPID_CATABOLIC_PROCESS | 194 | -0.5525947 | -2.185464 | 0 | 9.71E-06 |
| GO_MITOCHONDRIAL_ELECTRON_TRANSPORT_NADH_TO_UBIQUINONE | 31 | -0.7457333 | -2.18938 | 0 | 9.85E-06 |
| GO_DRUG_METABOLIC_PROCESS | 377 | -0.5204105 | -2.193111 | 0 | 9.99E-06 |
| GO_FATTY_ACID_METABOLIC_PROCESS | 238 | -0.5420886 | -2.195032 | 0 | 1.01E-05 |
| GO_BRANCHED_CHAIN_AMINO_ACID_METABOLIC_PROCESS | 18 | -0.8692683 | -2.196108 | 0 | 1.03E-05 |
| GO_MITOCHONDRIAL_TRANSLATION | 71 | -0.6512105 | -2.209394 | 0 | 1.04E-05 |
| GO_FLAVIN_ADENINE_DINUCLEOTIDE_BINDING | 63 | -0.6578865 | -2.221233 | 0 | 1.06E-05 |
| GO_GENERATION_OF_PRECURSOR_METABOLITES_AND_ENERGY | 313 | -0.5307103 | -2.226903 | 0 | 1.08E-05 |
| REACTOME_METABOLISM_OF_AMINO_ACIDS_AND_DERIVATIVES | 215 | -0.5616351 | -2.227199 | 0 | 1.09E-05 |
| GO_XENOBIOTIC_METABOLIC_PROCESS | 80 | -0.631865 | -2.228262 | 0 | 1.11E-05 |
| GO_SULFUR_AMINO_ACID_METABOLIC_PROCESS | 26 | -0.7961816 | -2.235969 | 0 | 1.13E-05 |
| GO_ORGANELLE_INNER_MEMBRANE | 250 | -0.5525092 | -2.241343 | 0 | 1.15E-05 |
| KEGG_CITRATE_CYCLE_TCA_CYCLE | 26 | -0.7958706 | -2.241673 | 0 | 1.17E-05 |
| GO_ASPARTATE_FAMILY_AMINO_ACID_METABOLIC_PROCESS | 34 | -0.7480921 | -2.241683 | 0 | 1.19E-05 |
| KEGG_PROPANOATE_METABOLISM | 24 | -0.8130571 | -2.242377 | 0 | 1.21E-05 |
| GO_VITAMIN_BINDING | 87 | -0.63441 | -2.247405 | 0 | 1.23E-05 |
| GO_REGULATION_OF_B_CELL_ACTIVATION | 83 | 0.6190978 | 2.1717718 | 0 | 2.35E-05 |

| | | | | | |
|---|---|---|---|---|---|
| GO_CYTOKINE_PRODUCTION | 473 | 0.5118066 | 2.1733425 | 0 | 2.40E-05 |
| KEGG_BETA_ALANINE_METABOLISM | 18 | -0.8350814 | -2.153786 | 0 | 2.47E-05 |
| REACTOME_NUCLEAR_RECEPTOR_TRANSCRIPTION_PATHWAY | 31 | -0.7233114 | -2.154029 | 0 | 2.50E-05 |
| GO_NADH_DEHYDROGENASE_COMPLEX | 26 | -0.7531539 | -2.154314 | 0 | 2.53E-05 |
| REACTOME_BIOLOGICAL_OXIDATIONS | 130 | -0.5729873 | -2.158347 | 0 | 2.56E-05 |
| GO_ATP_SYNTHESIS_COUPLED_ELECTRON_TRANSPORT | 48 | -0.6663019 | -2.160417 | 0 | 2.59E-05 |
| GO_MONOCARBOXYLIC_ACID_METABOLIC_PROCESS | 403 | -0.5096181 | -2.162393 | 0 | 2.63E-05 |
| GO_OXIDOREDUCTASE_ACTIVITY_ACTING_ON_NAD_P_H_QUINONE_OR_SIMILAR_COMPOUND_AS_ACCEPTOR | 35 | -0.7034621 | -2.138901 | 0 | 3.08E-05 |
| REACTOME_COMPLEX_I_BIOGENESIS | 30 | -0.7426813 | -2.141709 | 0 | 3.12E-05 |
| KEGG_LYSINE_DEGRADATION | 29 | -0.7456541 | -2.143954 | 0 | 3.15E-05 |
| REACTOME_PHASE_I_FUNCTIONALIZATION_OF_COMPOUNDS | 69 | -0.6318396 | -2.145784 | 0 | 3.19E-05 |
| REACTOME_PEROXISOMAL_LIPID_METABOLISM | 19 | -0.8160441 | -2.148363 | 0 | 3.23E-05 |
| GO_DRUG_CATABOLIC_PROCESS | 94 | -0.5878354 | -2.132867 | 0 | 3.74E-05 |
| GO_HYDRO_LYASE_ACTIVITY | 33 | -0.6943777 | -2.133658 | 0 | 3.78E-05 |
| GO_THIOESTER_METABOLIC_PROCESS | 68 | -0.6320365 | -2.136775 | 0 | 3.83E-05 |
| REACTOME_MITOCHONDRIAL_TRANSLATION | 47 | -0.6676238 | -2.117025 | 0 | 4.30E-05 |
| GO_ORGANIC_ACID_TRANSMEMBRANE_TRANSPORTER_ACTIVITY | 78 | -0.5971049 | -2.117777 | 0 | 4.35E-05 |
| GO_TRICARBOXYLIC_ACID_CYCLE | 26 | -0.7577686 | -2.121675 | 0 | 4.39E-05 |
| GO_CELLULAR_AMINO_ACID_BIOSYNTHETIC_PROCESS | 53 | -0.6446444 | -2.125155 | 0 | 4.44E-05 |
| REACTOME_FATTY_ACID_METABOLISM | 116 | -0.5752508 | -2.11504 | 0 | 4.96E-05 |
| HALLMARK_BILE_ACID_METABOLISM | 81 | -0.6027807 | -2.11139 | 0 | 5.55E-05 |
| GO_GLUCOSE_METABOLIC_PROCESS | 125 | -0.5581976 | -2.113277 | 0 | 5.61E-05 |
| GO_CELLULAR_ALDEHYDE_METABOLIC_PROCESS | 44 | -0.6642549 | -2.107485 | 0 | 6.20E-05 |
| GO_AROMATIC_AMINO_ACID_FAMILY_METABOLIC_PROCESS | 22 | -0.760559 | -2.104193 | 0 | 6.82E-05 |
| GO_VITAMIN_METABOLIC_PROCESS | 87 | -0.5898089 | -2.102827 | 0 | 7.44E-05 |
| GO_REGULATION_OF_PHAGOCYTOSIS | 62 | 0.65082866 | 2.1468284 | 0 | 7.47E-05 |
| HALLMARK_IL6_JAK_STAT3_SIGNALING | 70 | 0.6342547 | 2.1485069 | 0 | 7.58E-05 |
| REACTOME_EXTRACELLULAR_MATRIX_ORGANIZATION | 193 | 0.5483118 | 2.1508996 | 0 | 7.69E-05 |
| GO_CELL_SUBSTRATE_JUNCTION | 281 | 0.5274285 | 2.1525497 | 0 | 7.81E-05 |
| REACTOME_ANTIGEN_PROCESSING_CROSS_PRESENTATION | 67 | 0.6423929 | 2.1530993 | 0 | 7.93E-05 |
| GO_POSITIVE_REGULATION_OF_LEUKOCYTE_CELL_CELL_ADHESION | 146 | 0.5792788 | 2.1532602 | 0 | 8.05E-05 |
| GO_NEGATIVE_REGULATION_OF_LEUKOCYTE_MEDIATED_IMMUNITY | 37 | 0.7183214 | 2.1553903 | 0 | 8.18E-05 |
| GO_B_CELL_ACTIVATION | 147 | 0.565624 | 2.1558044 | 0 | 8.31E-05 |
| GO_LYMPHOCYTE_ACTIVATION | 410 | 0.5129904 | 2.1570787 | 0 | 8.45E-05 |
| GO_REGULATION_OF_LYMPHOCYTE_ACTIVATION | 272 | 0.53153 | 2.1579664 | 0 | 8.59E-05 |
| GO_COMPLEMENT_ACTIVATION | 69 | 0.6262067 | 2.1350532 | 0 | 8.72E-05 |
| KEGG_TYPE_I_DIABETES_MELLITUS | 28 | 0.7584741 | 2.1591837 | 0 | 8.74E-05 |
| GO_REGULATION_OF_IMMUNE_EFFECTOR_PROCESS | 267 | 0.5283779 | 2.135166 | 0 | 8.84E-05 |
| GO_ANTIGEN_BINDING | 55 | 0.6669835 | 2.1606731 | 0 | 8.89E-05 |
| GO_NUCLEAR_TRANSCRIBED_MRNA_CATABOLIC_PROCESS_NONSENSE_MEDIATED_DECAY | 47 | 0.67593753 | 2.1371515 | 0 | 8.96E-05 |
| GO_POSITIVE_REGULATION_OF_CYTOKINE_PRODUCTION | 275 | 0.53089476 | 2.162929 | 0 | 9.04E-05 |
| KEGG_INTESTINAL_IMMUNE_NETWORK_FOR_IGA_PRODUCTION | 26 | 0.7668281 | 2.1415234 | 0 | 9.09E-05 |
| GO_LEUKOCYTE_CELL_CELL_ADHESION | 223 | 0.5414095 | 2.1651022 | 0 | 9.20E-05 |
| REACTOME_GENERATION_OF_SECOND_MESSENGER_MOLECULES | 26 | 0.7641959 | 2.1433682 | 0 | 9.22E-05 |
| GO_REGULATION_OF_VIRAL_GENOME_REPLICATION | 58 | 0.66424596 | 2.1684613 | 0 | 9.37E-05 |

| | | | | | |
|---|---|---|---|---|---|
| BIOCARTA_CASPASE_PATHWAY | 19 | 0.8177482 | 2.129408 | 0.00203252 | 1.03E-04 |
| GO_SERINE_FAMILY_AMINO_ACID_METABOLIC_PROCESS | 26 | -0.7554327 | -2.088206 | 0 | 1.07E-04 |
| KEGG_PPAR_SIGNALING_PATHWAY | 48 | -0.6449691 | -2.089534 | 0 | 1.08E-04 |
| PID_INTEGRIN1_PATHWAY | 52 | 0.649067 | 2.1111507 | 0 | 1.10E-04 |
| HALLMARK_EPITHELIAL_MESENCHYMAL_TRANSITION | 168 | 0.5511911 | 2.1155632 | 0 | 1.11E-04 |
| REACTOME_SRP_DEPENDENT_COTRANSLATIONAL_PROTEIN_TARGETING_TO_MEMBRANE | 47 | 0.67011404 | 2.1163058 | 0 | 1.12E-04 |
| REACTOME_NON_INTEGRIN_MEMBRANE_ECM_INTERACTIONS | 43 | 0.677785 | 2.119221 | 0 | 1.14E-04 |
| PID_INTEGRIN3_PATHWAY | 34 | 0.7127371 | 2.1231484 | 0 | 1.15E-04 |
| GO_REGULATION_OF_MULTI_ORGANISM_PROCESS | 450 | 0.5010253 | 2.1236856 | 0 | 1.17E-04 |
| GO_ELECTRON_TRANSFER_ACTIVITY | 74 | -0.6121476 | -2.085407 | 0 | 1.18E-04 |
| GO_NEGATIVE_REGULATION_OF_IMMUNE_RESPONSE | 98 | 0.59953415 | 2.125662 | 0 | 1.18E-04 |
| GO_OXIDOREDUCTASE_COMPLEX | 65 | -0.6251103 | -2.085543 | 0 | 1.19E-04 |
| GO_INTERLEUKIN_1_PRODUCTION | 61 | 0.6234719 | 2.1053278 | 0 | 1.21E-04 |
| GO_MATURE_B_CELL_DIFFERENTIATION | 16 | 0.83443755 | 2.108066 | 0 | 1.22E-04 |
| GO_T_CELL_ACTIVATION | 290 | 0.5163027 | 2.1082225 | 0 | 1.24E-04 |
| GO_MONOSACCHARIDE_METABOLIC_PROCESS | 172 | -0.5306606 | -2.079588 | 0 | 1.42E-04 |
| KEGG_HISTIDINE_METABOLISM | 19 | -0.8047929 | -2.076803 | 0 | 1.46E-04 |
| GO_NAD_BINDING | 46 | -0.6565214 | -2.077886 | 0 | 1.48E-04 |
| GO_OXIDATIVE_PHOSPHORYLATION | 61 | -0.6123714 | -2.075134 | 0 | 1.57E-04 |
| GO_MONOSACCHARIDE_BIOSYNTHETIC_PROCESS | 64 | -0.6100166 | -2.068972 | 0 | 1.61E-04 |
| GO_STEROID_HORMONE_RECEPTOR_ACTIVITY | 34 | -0.6904094 | -2.072712 | 0 | 1.62E-04 |
| GO_COFACTOR_BIOSYNTHETIC_PROCESS | 141 | -0.5369382 | -2.06595 | 0 | 1.71E-04 |
| GO_NUCLEOSIDE_BISPHOSPHATE_METABOLIC_PROCESS | 86 | -0.5757176 | -2.061711 | 0 | 1.75E-04 |
| KEGG_GLYCOLYSIS_GLUCONEOGENESIS | 42 | -0.6581132 | -2.062446 | 0 | 1.76E-04 |
| GO_GLUTAMINE_FAMILY_AMINO_ACID_METABOLIC_PROCESS | 46 | -0.6463163 | -2.05675 | 0 | 1.91E-04 |
| GO_LYMPHOCYTE_ACTIVATION_INVOLVED_IN_IMMUNE_RESPONSE | 103 | 0.57970685 | 2.0972955 | 0 | 1.91E-04 |
| GO_DEFENSE_RESPONSE_TO_VIRUS | 143 | 0.55355275 | 2.0980725 | 0 | 1.93E-04 |
| GO_MITOCHONDRIAL_TRANSLATIONAL_TERMINATION | 45 | -0.6585794 | -2.055933 | 0 | 1.95E-04 |
| GO_ORGANELLAR_RIBOSOME | 45 | -0.6436018 | -2.048115 | 0 | 2.10E-04 |
| GO_OXIDOREDUCTASE_ACTIVITY_ACTING_ON_CH_OH_GROUP_OF_DONORS | 79 | -0.5908949 | -2.050785 | 0 | 2.11E-04 |
| GO_POSITIVE_REGULATION_OF_INTERLEUKIN_12_PRODUCTION | 21 | 0.7843035 | 2.09356 | 0 | 2.13E-04 |
| GO_INTEGRIN_BINDING | 91 | 0.5917659 | 2.0936282 | 0 | 2.16E-04 |
| REACTOME_ADAPTIVE_IMMUNE_SYSTEM | 450 | 0.49426925 | 2.093949 | 0 | 2.18E-04 |
| GO_T_CELL_PROLIFERATION | 121 | 0.56670576 | 2.0907462 | 0 | 2.20E-04 |
| GO_MYD88_DEPENDENT_TOLL_LIKE_RECEPTOR_SIGNALING_PATHWAY | 20 | 0.791781 | 2.0914166 | 0 | 2.22E-04 |
| GO_INTERFERON_GAMMA_PRODUCTION | 65 | 0.62284213 | 2.091917 | 0 | 2.25E-04 |
| GO_T_CELL_RECEPTOR_SIGNALING_PATHWAY | 130 | 0.5641788 | 2.0887399 | 0 | 2.26E-04 |
| GO_RESPONSE_TO_VIRUS | 196 | 0.532527 | 2.0887845 | 0 | 2.29E-04 |
| REACTOME_NONSENSE_MEDIATED_DECAY_NMD | 46 | 0.66561264 | 2.089227 | 0 | 2.31E-04 |
| GO_REGULATION_OF_HUMORAL_IMMUNE_RESPONSE | 65 | 0.6180855 | 2.0867019 | 0 | 2.37E-04 |
| REACTOME_SIGNALING_BY_ROBO_RECEPTORS | 104 | 0.5821847 | 2.0824883 | 0 | 2.48E-04 |
| GO_NADH_DEHYDROGENASE_ACTIVITY | 24 | -0.7398388 | -2.038073 | 0 | 2.72E-04 |
| GO_REGULATION_OF_T_CELL_ACTIVATION | 199 | 0.53134036 | 2.0766377 | 0 | 2.97E-04 |
| GO_OXIDOREDUCTASE_ACTIVITY_ACTING_ON_THE_CH_NH_GROUP_OF_DONORS | 16 | -0.7897101 | -2.030131 | 0 | 3.01E-04 |
| GO_MITOCHONDRIAL_TRANSMEMBRANE_TRANSPORT | 42 | -0.6512229 | -2.031225 | 0 | 3.04E-04 |
| REACTOME_COLLAGEN_CHAIN_TRIMERIZATION | 24 | 0.7385227 | 2.0745482 | 0 | 3.04E-04 |
| KEGG_PROXIMAL_TUBULE_BICARBONATE_RECLAMATION | 18 | -0.7919112 | -2.026364 | 0 | 3.05E-04 |

| | | | | | |
|---|---|---|---|---|---|
| REACTOME_COLLAGEN_DEGRADATION | 39 | 0.67471695 | 2.0745676 | 0 | 3.07E-04 |
| GO_POSITIVE_REGULATION_OF_PHAGOCYTOSIS | 47 | 0.645222 | 2.0700405 | 0 | 3.07E-04 |
| GO_COLLAGEN_CONTAINING_EXTRACELLULAR_MATRIX | 264 | 0.5162326 | 2.0720556 | 0 | 3.10E-04 |
| GO_ANTIGEN_PROCESSING_AND_PRESENTATION | 161 | 0.5370435 | 2.0727592 | 0 | 3.13E-04 |
| GO_CELL_ACTIVATION_INVOLVED_IN_IMMUNE_RESPONSE | 472 | 0.48411295 | 2.0654907 | 0 | 3.38E-04 |
| REACTOME_DEGRADATION_OF_THE_EXTRACELLULAR_MATRIX | 85 | 0.5882512 | 2.0661504 | 0 | 3.41E-04 |
| GO_REGULATION_OF_ANTIGEN_RECEPTOR_MEDIATED_SIGNALING_PATHWAY | 41 | 0.66894585 | 2.0638218 | 0 | 3.58E-04 |
| GO_PTERIDINE_CONTAINING_COMPOUND_METABOLIC_PROCESS | 22 | -0.7372885 | -2.01718 | 0 | 3.59E-04 |
| GO_DENDRITIC_CELL_DIFFERENTIATION | 28 | 0.718199 | 2.0610182 | 0 | 3.67E-04 |
| GO_PRODUCTION_OF_MOLECULAR_MEDIATOR_OF_IMMUNE_RESPONSE | 142 | 0.5451029 | 2.0580025 | 0 | 3.75E-04 |
| GO_ORGANIC_HYDROXY_COMPOUND_CATABOLIC_PROCESS | 43 | -0.6338819 | -2.014646 | 0 | 3.79E-04 |
| GO_TRANSLATIONAL_TERMINATION | 55 | -0.6143643 | -2.012025 | 0 | 4.03E-04 |
| GO_REGULATION_OF_RESPONSE_TO_BIOTIC_STIMULUS | 307 | 0.50201774 | 2.0563035 | 0 | 4.07E-04 |
| GO_SMALL_MOLECULE_BIOSYNTHETIC_PROCESS | 457 | -0.4716843 | -2.011035 | 0 | 4.11E-04 |
| GO_MYELOID_LEUKOCYTE_ACTIVATION | 452 | 0.48320314 | 2.054604 | 0 | 4.15E-04 |
| GO_DEFENSE_RESPONSE_TO_BACTERIUM | 135 | 0.546216 | 2.0532935 | 0 | 4.34E-04 |
| GO_COENZYME_BIOSYNTHETIC_PROCESS | 91 | -0.5554664 | -2.004891 | 0 | 4.53E-04 |
| GO_SECONDARY_ACTIVE_TRANSMEMBRANE_TRANSPORTER_ACTIVITY | 134 | -0.5282381 | -2.005355 | 0 | 4.56E-04 |
| GO_MHC_PROTEIN_COMPLEX | 17 | 0.81694853 | 2.0507607 | 0 | 4.64E-04 |
| GO_B_CELL_ACTIVATION_INVOLVED_IN_IMMUNE_RESPONSE | 40 | 0.6740689 | 2.0487535 | 0 | 4.71E-04 |
| GO_EXTRACELLULAR_MATRIX_STRUCTURAL_CONSTITUENT_CONFERRING_TENSILE_STRENGTH | 24 | 0.7385229 | 2.0477283 | 0 | 4.74E-04 |
| KEGG_DRUG_METABOLISM_CYTOCHROME_P450 | 41 | -0.6461679 | -2.001484 | 0 | 4.76E-04 |
| GO_FC_RECEPTOR_MEDIATED_STIMULATORY_SIGNALING_PATHWAY | 69 | 0.59629303 | 2.0445147 | 0 | 4.76E-04 |
| GO_IMMUNE_RECEPTOR_ACTIVITY | 87 | 0.5786091 | 2.0480702 | 0 | 4.78E-04 |
| GO_INTERLEUKIN_6_PRODUCTION | 87 | 0.576535 | 2.0460293 | 0 | 4.80E-04 |
| GO_CELLULAR_DEFENSE_RESPONSE | 36 | 0.67193365 | 2.040859 | 0 | 4.90E-04 |
| GO_REGULATION_OF_DEFENSE_RESPONSE | 444 | 0.48074895 | 2.0409708 | 0 | 4.94E-04 |
| GO_CELLULAR_BIOGENIC_AMINE_METABOLIC_PROCESS | 32 | -0.6688463 | -1.998607 | 0 | 4.96E-04 |
| GO_ASPARTATE_FAMILY_AMINO_ACID_CATABOLIC_PROCESS | 15 | -0.8156954 | -1.998789 | 0 | 5.00E-04 |
| GO_ALPHA_AMINO_ACID_BIOSYNTHETIC_PROCESS | 44 | -0.6313474 | -1.999009 | 0 | 5.04E-04 |
| GO_LEUKOCYTE_MIGRATION | 305 | 0.50042397 | 2.0364535 | 0 | 5.20E-04 |
| GO_REGULATION_OF_LEUKOCYTE_APOPTOTIC_PROCESS | 57 | 0.61798567 | 2.0366538 | 0 | 5.24E-04 |
| GO_I_KAPPAB_KINASE_NF_KAPPAB_SIGNALING | 167 | 0.5243533 | 2.0351138 | 0 | 5.26E-04 |
| HALLMARK_G2M_CHECKPOINT | 122 | 0.5519363 | 2.037113 | 0 | 5.28E-04 |
| GO_POSITIVE_REGULATION_OF_I_KAPPAB_KINASE_NF_KAPPAB_SIGNALING | 118 | 0.56032294 | 2.0345984 | 0 | 5.32E-04 |
| REACTOME_SLC_MEDIATED_TRANSMEMBRANE_TRANSPORT | 142 | -0.5204597 | -1.995085 | 0 | 5.39E-04 |
| GO_CELLULAR_RESPONSE_TO_XENOBIOTIC_STIMULUS | 117 | -0.5348817 | -1.992034 | 0 | 5.50E-04 |
| GO_NEGATIVE_REGULATION_OF_VIRAL_PROCESS | 65 | 0.60560226 | 2.0309997 | 0 | 5.55E-04 |
| GO_CELL_KILLING | 96 | 0.56700677 | 2.0326402 | 0 | 5.58E-04 |
| PID_SYNDECAN_1_PATHWAY | 33 | 0.68438375 | 2.0311308 | 0 | 5.59E-04 |
| NABA_COLLAGENS | 24 | 0.73852265 | 2.0313504 | 0 | 5.63E-04 |
| GO_MYELOID_LEUKOCYTE_MEDIATED_IMMUNITY | 389 | 0.4799269 | 2.0283685 | 0 | 5.75E-04 |
| GO_POSITIVE_REGULATION_OF_ADAPTIVE_IMMUNE_RESPONSE | 64 | 0.6009637 | 2.0295103 | 0 | 5.80E-04 |
| GO_EXTRACELLULAR_STRUCTURE_ORGANIZATION | 242 | 0.50440216 | 2.0279746 | 0 | 5.81E-04 |
| REACTOME_MITOCHONDRIAL_PROTEIN_IMPORT | 31 | -0.6816427 | -1.98678 | 0 | 5.97E-04 |

| | | | | | |
|---|---|---|---|---|---|
| GO_REGULATION_OF_LEUKOCYTE_MEDIATED_IMMUNITY | 130 | 0.5442821 | 2.0226068 | 0 | 6.16E-04 |
| GO_REGULATION_OF_SYMBIOSIS_ENCOMPASSING_MUTUALISM_THROUGH_PARASITISM | 136 | 0.5399299 | 2.0254092 | 0 | 6.20E-04 |
| GO_REGULATION_OF_PRODUCTION_OF_MOLECULAR_MEDIATOR_OF_IMMUNE_RESPONSE | 96 | 0.56307787 | 2.0228925 | 0 | 6.20E-04 |
| GO_PROGRAMMED_NECROTIC_CELL_DEATH | 25 | 0.72679085 | 2.0255587 | 0 | 6.25E-04 |
| GO_POSITIVE_REGULATION_OF_CELL_CELL_ADHESION | 169 | 0.52908844 | 2.024526 | 0 | 6.25E-04 |
| REACTOME_CITRIC_ACID_CYCLE_TCA_CYCLE | 18 | -0.7750162 | -1.980282 | 0 | 6.33E-04 |
| GO_OXIDOREDUCTASE_ACTIVITY_ACTING_ON_PAIRED_DONORS_WITH_INCORPORATION_OR_REDUCTION_OF_MOLECULAR_OXYGEN_NAD_P_H_AS_ONE_DONOR_AND_INCORPORATION_OF_ONE_ATOM_OF_OXYGEN | 30 | -0.6661966 | -1.979183 | 0 | 6.39E-04 |
| REACTOME_NEUTROPHIL_DEGRANULATION | 334 | 0.48882136 | 2.0194082 | 0 | 6.76E-04 |
| NABA_CORE_MATRISOME | 145 | 0.52967125 | 2.0201297 | 0 | 6.76E-04 |
| GO_ANTIGEN_PROCESSING_AND_PRESENTATION_OF_EXOGENOUS_PEPTIDE_ANTIGEN_VIA_MHC_CLASS_I | 50 | 0.6362913 | 2.0195591 | 0 | 6.81E-04 |
| REACTOME_SYNTHESIS_OF_BILE_ACIDS_AND_BILE_SALTS | 20 | -0.734449 | -1.97601 | 0 | 6.84E-04 |
| GO_COMPLEX_OF_COLLAGEN_TRIMERS | 16 | 0.80637866 | 2.0108812 | 0 | 7.00E-04 |
| GO_POSITIVE_REGULATION_OF_B_CELL_ACTIVATION | 55 | 0.6275533 | 2.013245 | 0 | 7.01E-04 |
| REACTOME_PARASITE_INFECTION | 53 | 0.62265563 | 2.009735 | 0 | 7.04E-04 |
| KEGG_HEMATOPOIETIC_CELL_LINEAGE | 57 | 0.61069524 | 2.011786 | 0 | 7.05E-04 |
| REACTOME_CELL_CYCLE_CHECKPOINTS | 126 | 0.5464391 | 2.0132012 | 0 | 7.05E-04 |
| GO_INTERFERON_BETA_PRODUCTION | 31 | 0.69854265 | 2.0134237 | 0 | 7.06E-04 |
| REACTOME_INTERLEUKIN_10_SIGNALING | 35 | 0.6580926 | 2.0123045 | 0 | 7.09E-04 |
| GO_POSITIVE_REGULATION_OF_DEFENSE_RESPONSE | 293 | 0.49658543 | 2.013908 | 0 | 7.11E-04 |
| GO_NEGATIVE_REGULATION_OF_VIRAL_LIFE_CYCLE | 52 | 0.6330413 | 2.0148244 | 0 | 7.16E-04 |
| GO_REGULATION_OF_VIRAL_LIFE_CYCLE | 87 | 0.5765351 | 2.0161731 | 0 | 7.17E-04 |
| GO_POSITIVE_REGULATION_OF_INTERLEUKIN_6_PRODUCTION | 56 | 0.6200052 | 2.0069194 | 0 | 7.20E-04 |
| GO_INTERLEUKIN_1_BETA_PRODUCTION | 54 | 0.617846 | 2.0150762 | 0 | 7.21E-04 |
| GO_BASEMENT_MEMBRANE | 55 | 0.6290896 | 2.0071352 | 0 | 7.25E-04 |
| GO_REGULATION_OF_CELL_ACTIVATION | 360 | 0.4835262 | 2.0043998 | 0 | 7.58E-04 |
| GO_MYELOID_LEUKOCYTE_DIFFERENTIATION | 126 | 0.53453314 | 1.9975432 | 0 | 7.68E-04 |
| GO_NEGATIVE_REGULATION_OF_LYMPHOCYTE_MEDIATED_IMMUNITY | 28 | 0.707576 | 1.9978553 | 0 | 7.73E-04 |
| GO_PHAGOCYTIC_VESICLE_MEMBRANE | 50 | 0.6234795 | 2.0004392 | 0 | 7.77E-04 |
| GO_CYTOKINE_SECRETION | 142 | 0.5269137 | 1.9955804 | 0 | 7.95E-04 |
| REACTOME_COMPLEMENT_CASCADE | 53 | 0.618695 | 1.9952196 | 0 | 7.98E-04 |
| GO_FATTY_ACYL_COA_BINDING | 16 | -0.795861 | -1.968451 | 0 | 8.08E-04 |
| GO_VITAMIN_B6_BINDING | 33 | -0.6716303 | -1.967307 | 0 | 8.12E-04 |
| REACTOME_EUKARYOTIC_TRANSLATION_ELONGATION | 32 | 0.683572 | 1.9920024 | 0 | 8.41E-04 |
| GO_INFLAMMATORY_RESPONSE | 447 | 0.47177336 | 1.9900228 | 0 | 8.41E-04 |
| GO_POSITIVE_REGULATION_OF_RESPONSE_TO_CYTOKINE_STIMULUS | 37 | 0.65711755 | 1.9910126 | 0 | 8.44E-04 |
| GO_ACTIVATION_OF_INNATE_IMMUNE_RESPONSE | 186 | 0.5125591 | 1.9905716 | 0 | 8.47E-04 |
| PID_INTEGRIN_A4B1_PATHWAY | 21 | 0.7422398 | 1.9892068 | 0 | 8.55E-04 |
| GO_LUMENAL_SIDE_OF_ENDOPLASMIC_RETICULUM_MEMBRANE | 17 | 0.7991514 | 1.9892145 | 0 | 8.60E-04 |
| GO_REGULATION_OF_B_CELL_PROLIFERATION | 42 | 0.6353078 | 1.9874798 | 0 | 8.96E-04 |
| GO_NEGATIVE_REGULATION_OF_MULTI_ORGANISM_PROCESS | 141 | 0.5202538 | 1.9857062 | 0 | 9.08E-04 |
| GO_FC_RECEPTOR_SIGNALING_PATHWAY | 125 | 0.533168 | 1.9859109 | 0 | 9.13E-04 |
| GO_AMINO_ACID_BINDING | 37 | -0.6459281 | -1.958637 | 0 | 9.33E-04 |
| HALLMARK_COMPLEMENT | 163 | 0.51203185 | 1.9837354 | 0 | 9.40E-04 |

| | | | | | |
|---|---|---|---|---|---|
| GO_CYTOKINE_METABOLIC_PROCESS | 72 | 0.5730254 | 1.9820607 | 0 | 9.42E-04 |
| GO_POSITIVE_REGULATION_OF_IMMUNE_EFFECTOR_PROCESS | 142 | 0.5286947 | 1.9816153 | 0 | 9.59E-04 |
| GO_REGULATION_OF_LYMPHOCYTE_MEDIATED_IMMUNITY | 86 | 0.5647398 | 1.9806303 | 0 | 9.63E-04 |
| GO_POSITIVE_REGULATION_OF_B_CELL_MEDIATED_IMMUNITY | 19 | 0.7425922 | 1.9807087 | 0 | 9.69E-04 |
| GO_CARBON_CARBON_LYASE_ACTIVITY | 37 | -0.6419026 | -1.956747 | 0 | 9.71E-04 |
| GO_ORGANIC_ACID_BIOSYNTHETIC_PROCESS | 217 | -0.486579 | -1.956555 | 0 | 9.74E-04 |
| KEGG_ALANINE_ASPARTATE_AND_GLUTAMATE_METABOLISM | 22 | -0.7180977 | -1.955206 | 0.002083333 | 9.75E-04 |
| GO_ANION_HOMEOSTASIS | 42 | -0.6190319 | -1.955577 | 0 | 9.77E-04 |
| GO_POSITIVE_REGULATION_OF_PRODUCTION_OF_MOLECULAR_MEDIATOR_OF_IMMUNE_RESPONSE | 66 | 0.59314835 | 1.9794295 | 0 | 9.80E-04 |
| GO_MYELOID_LEUKOCYTE_MIGRATION | 131 | 0.5213908 | 1.9783223 | 0 | 9.96E-04 |
| GO_CELLULAR_CARBOHYDRATE_CATABOLIC_PROCESS | 31 | -0.6603433 | -1.953489 | 0 | 9.97E-04 |
| GO_POSITIVE_REGULATION_OF_RESPONSE_TO_BIOTIC_STIMULUS | 229 | 0.49895126 | 1.9772573 | 0 | 0.001006435 |
| GO_CELL_RECOGNITION | 80 | 0.5739764 | 1.9766084 | 0 | 0.001007963 |
| KEGG_CHEMOKINE_SIGNALING_PATHWAY | 119 | 0.5363139 | 1.9775375 | 0 | 0.001012154 |
| GO_REGULATION_OF_FATTY_ACID_OXIDATION | 21 | -0.7274227 | -1.950097 | 0.00407332 | 0.001050925 |
| REACTOME_EPH_EPHRIN_SIGNALING | 64 | 0.5915918 | 1.9747441 | 0 | 0.001073882 |
| PID_AVB3_INTEGRIN_PATHWAY | 53 | 0.6048353 | 1.9719923 | 0 | 0.001084701 |
| REACTOME_MET_PROMOTES_CELL_MOTILITY | 26 | 0.70889145 | 1.9713854 | 0 | 0.001085708 |
| GO_T_CELL_MEDIATED_IMMUNITY | 71 | 0.5850766 | 1.9735557 | 0 | 0.001089552 |
| REACTOME_HOST_INTERACTIONS_OF_HIV_FACTORS | 78 | 0.5788041 | 1.9721363 | 0 | 0.001090694 |
| GO_INTERLEUKIN_12_PRODUCTION | 35 | 0.65079725 | 1.9699681 | 0 | 0.001114412 |
| GO_GLUTAMINE_FAMILY_AMINO_ACID_CATABOLIC_PROCESS | 21 | -0.7167017 | -1.945956 | 0 | 0.001132854 |
| GO_MHC_PROTEIN_COMPLEX_BINDING | 23 | 0.7154551 | 1.9677479 | 0 | 0.001170711 |
| GO_SIDE_OF_MEMBRANE | 343 | 0.47473577 | 1.9677358 | 0 | 0.00117129 |
| GO_POSITIVE_REGULATION_OF_HEMOPOIESIS | 119 | 0.53431845 | 1.9670714 | 0 | 0.001178799 |
| GO_NEUTROPHIL_MIGRATION | 75 | 0.5730711 | 1.9659736 | 0 | 0.001213402 |
| GO_SECRETORY_GRANULE_MEMBRANE | 189 | 0.50351197 | 1.9657794 | 0 | 0.001213739 |
| KEGG_LEUKOCYTE_TRANSENDOTHELIAL_MIGRATION | 81 | 0.5634489 | 1.9652039 | 0 | 0.001220824 |
| GO_REGULATION_OF_NATURAL_KILLER_CELL_ACTIVATION | 20 | 0.742387 | 1.96476 | 0 | 0.001221315 |
| GO_REGULATION_OF_B_CELL_MEDIATED_IMMUNITY | 28 | 0.684015 | 1.9627144 | 0 | 0.001241502 |
| GO_LYMPHOCYTE_MIGRATION | 68 | 0.57040405 | 1.9587449 | 0 | 0.001300735 |
| KEGG_PATHOGENIC_ESCHERICHIA_COLI_INFECTION | 40 | 0.6388041 | 1.9588022 | 0 | 0.00130744 |
| GO_REGULATION_OF_CYTOKINE_PRODUCTION_INVOLVED_IN_IMMUNE_RESPONSE | 63 | 0.58240485 | 1.9573946 | 0 | 0.001313503 |
| GO_TOLL_LIKE_RECEPTOR_SIGNALING_PATHWAY | 88 | 0.5533282 | 1.96 | 0 | 0.001314214 |
| REACTOME_INFLUENZA_INFECTION | 72 | 0.5755982 | 1.9571267 | 0 | 0.001320032 |
| GO_ORGANIC_ANION_TRANSMEMBRANE_TRANSPORTER_ACTIVITY | 110 | -0.5269566 | -1.937741 | 0 | 0.00132526 |
| GO_COTRANSLATIONAL_PROTEIN_TARGETING_TO_MEMBRANE | 38 | 0.65451646 | 1.9566386 | 0 | 0.001332932 |
| GO_LYASE_ACTIVITY | 120 | -0.5253762 | -1.937842 | 0 | 0.001334037 |
| GO_OSTEOCLAST_DIFFERENTIATION | 54 | 0.5925779 | 1.9536186 | 0 | 0.001357346 |
| REACTOME_INITIAL_TRIGGERING_OF_COMPLEMENT | 26 | 0.69004655 | 1.9542449 | 0 | 0.001357713 |
| GO_CALCIUM_DEPENDENT_PHOSPHOLIPID_BINDING | 28 | 0.6797488 | 1.9544214 | 0 | 0.001364502 |
| GO_CYTOKINE_PRODUCTION_INVOLVED_IN_IMMUNE_RESPONSE | 74 | 0.56653523 | 1.9547322 | 0 | 0.001371358 |
| GO_AROMATIC_AMINO_ACID_FAMILY_CATABOLIC_PROCESS | 17 | -0.7665316 | -1.934114 | 0 | 0.001413073 |
| GO_LEUKOCYTE_DIFFERENTIATION | 305 | 0.47270405 | 1.9512128 | 0 | 0.001432753 |
| GO_NEUTRAL_LIPID_METABOLIC_PROCESS | 67 | -0.5628683 | -1.932597 | 0 | 0.001435829 |
| REACTOME_METABOLISM_OF_WATER_SOLUBLE_VITAMINS_AND_COFACTORS | 73 | -0.5618602 | -1.932358 | 0 | 0.001440116 |

| | | | | | |
|---|---|---|---|---|---|
| GO_CELL_ADHESION_MOLECULE_BINDING | 308 | 0.4707432 | 1.9480804 | 0 | 0.001463396 |
| SIG_REGULATION_OF_THE_ACTIN_CYTOSKELETON_BY_RHO_GTPASES | 21 | 0.734466 | 1.9468508 | 0 | 0.001481218 |
| REACTOME_ASSEMBLY_OF_COLLAGEN_FIBRILS_AND_OTHER_MULTIMERIC_STRUCTURES | 37 | 0.6396472 | 1.9459008 | 0.002087683 | 0.001511376 |
| GO_POSITIVE_REGULATION_OF_INTERLEUKIN_1_PRODUCTION | 30 | 0.67961174 | 1.9452329 | 0 | 0.001522537 |
| GO_POSITIVE_REGULATION_OF_INTERLEUKIN_8_PRODUCTION | 40 | 0.63379085 | 1.9432762 | 0 | 0.001558357 |
| GO_ORGANIC_ACID_TRANSPORT | 193 | -0.4857485 | -1.926172 | 0 | 0.001573182 |
| GO_BILE_ACID_BIOSYNTHETIC_PROCESS | 19 | -0.7237598 | -1.926919 | 0 | 0.001574233 |
| REACTOME_NERVOUS_SYSTEM_DEVELOPMENT | 335 | 0.46755987 | 1.9413283 | 0 | 0.001600106 |
| REACTOME_BILE_ACID_AND_BILE_SALT_METABOLISM | 26 | -0.6814162 | -1.92429 | 0 | 0.001616316 |
| GO_TUMOR_NECROSIS_FACTOR_BIOSYNTHETIC_PROCESS | 17 | 0.7695063 | 1.9373356 | 0 | 0.001677992 |
| HALLMARK_E2F_TARGETS | 134 | 0.5213378 | 1.9376329 | 0 | 0.001679836 |
| GO_EPHRIN_RECEPTOR_SIGNALING_PATHWAY | 59 | 0.59125 | 1.9378489 | 0 | 0.001687797 |
| GO_INTEGRIN_MEDIATED_SIGNALING_PATHWAY | 74 | 0.5590656 | 1.9369186 | 0 | 0.001688138 |
| GO_EXTERNAL_SIDE_OF_PLASMA_MEMBRANE | 231 | 0.48284367 | 1.9380097 | 0 | 0.001695834 |
| GO_NEGATIVE_REGULATION_OF_LEUKOCYTE_APOPTOTIC_PROCESS | 36 | 0.64639044 | 1.9355803 | 0 | 0.001739937 |
| REACTOME_FCERI_MEDIATED_NF_KB_ACTIVATION | 55 | 0.5938408 | 1.9346054 | 0 | 0.001773501 |
| GO_ALCOHOL_METABOLIC_PROCESS | 241 | -0.4708178 | -1.918842 | 0 | 0.001848298 |
| GO_ANTIGEN_PROCESSING_AND_PRESENTATION_OF_PEPTIDE_ANTIGEN | 134 | 0.5176235 | 1.9307913 | 0 | 0.001871915 |
| GO_VIRAL_GENOME_REPLICATION | 75 | 0.5613908 | 1.9305422 | 0 | 0.001872098 |
| GO_RESPONSE_TO_INTERFERON_ALPHA | 17 | 0.77542263 | 1.9306316 | 0 | 0.001880685 |
| GO_CELL_CELL_ADHESION | 482 | 0.4506791 | 1.9302077 | 0 | 0.001887037 |
| GO_REGULATION_OF_CELL_ADHESION | 425 | 0.45527384 | 1.9296012 | 0 | 0.001907441 |
| GO_POSITIVE_REGULATION_OF_CELL_ADHESION | 256 | 0.48051816 | 1.9283189 | 0 | 0.001939062 |
| GO_CHEMOKINE_BINDING | 22 | 0.7043095 | 1.9266505 | 0 | 0.001981959 |
| GO_ORGANIC_ACID_BINDING | 132 | -0.5049404 | -1.913266 | 0 | 0.002076983 |
| GO_NECROTIC_CELL_DEATH | 30 | 0.6528963 | 1.9225802 | 0 | 0.002133249 |
| GO_POSITIVE_REGULATION_OF_INFLAMMATORY_RESPONSE | 89 | 0.53809834 | 1.922243 | 0 | 0.002135092 |
| GO_GRANULOCYTE_MIGRATION | 90 | 0.5441637 | 1.921722 | 0 | 0.002148154 |
| REACTOME_MET_ACTIVATES_PTK2_SIGNALING | 20 | 0.7406839 | 1.9182223 | 0 | 0.002149311 |
| GO_TUMOR_NECROSIS_FACTOR_MEDIATED_SIGNALING_PATHWAY | 96 | 0.53339523 | 1.9214753 | 0 | 0.002150025 |
| GO_POSITIVE_REGULATION_OF_SMALL_GTPASE_MEDIATED_SIGNAL_TRANSDUCTION | 37 | 0.6328691 | 1.9210914 | 0 | 0.002151963 |
| GO_B_CELL_DIFFERENTIATION | 69 | 0.5707003 | 1.9187634 | 0 | 0.002156737 |
| GO_T_CELL_MIGRATION | 41 | 0.61957026 | 1.9183071 | 0 | 0.002158458 |
| REACTOME_EPHB_MEDIATED_FORWARD_SIGNALING | 28 | 0.68094957 | 1.919032 | 0 | 0.002160562 |
| GO_CHEMOKINE_PRODUCTION | 62 | 0.58461887 | 1.9199249 | 0 | 0.002162635 |
| GO_NEGATIVE_REGULATION_OF_INNATE_IMMUNE_RESPONSE | 38 | 0.63113916 | 1.9201078 | 0 | 0.002166359 |
| GO_REGULATION_OF_ANTIGEN_PROCESSING_AND_PRESENTATION | 17 | 0.768122 | 1.919396 | 0 | 0.002169875 |
| GO_EXTRACELLULAR_MATRIX | 319 | 0.4682656 | 1.9202225 | 0 | 0.002170224 |
| KEGG_NATURAL_KILLER_CELL_MEDIATED_CYTOTOXICITY | 86 | 0.54662186 | 1.917404 | 0 | 0.002178357 |
| KEGG_TYROSINE_METABOLISM | 28 | -0.659578 | -1.908959 | 0 | 0.002220694 |
| GO_INTERLEUKIN_8_PRODUCTION | 54 | 0.5857559 | 1.91474 | 0 | 0.002282903 |
| GO_FOLIC_ACID_CONTAINING_COMPOUND_METABOLIC_PROCESS | 17 | -0.7549537 | -1.90641 | 0 | 0.002301841 |
| GO_LEUKOCYTE_MEDIATED_CYTOTOXICITY | 70 | 0.5646322 | 1.914104 | 0 | 0.002310694 |
| GO_MACROPHAGE_ACTIVATION | 66 | 0.5717389 | 1.9138631 | 0 | 0.00231188 |

| | | | | | |
|---|---|---|---|---|---|
| GO_CELLULAR_RESPONSE_TO_BIOTIC_STIMULUS | 150 | 0.5030143 | 1.9134148 | 0 | 0.00231288 |
| GO_LIPOPOLYSACCHARIDE_BINDING | 18 | 0.7306326 | 1.9131062 | 0 | 0.002319239 |
| HALLMARK_PEROXISOME | 79 | -0.5434679 | -1.90536 | 0 | 0.002335103 |
| REACTOME_REGULATION_OF_EXPRESSION_OF_SLITS_AND_ROBOS | 77 | 0.5548074 | 1.9118587 | 0 | 0.002343392 |
| GO_POSITIVE_REGULATION_OF_MULTI_ORGANISM_PROCESS | 307 | 0.46489567 | 1.9124386 | 0 | 0.002346686 |
| GO_COLLAGEN_TRIMER | 42 | 0.61520165 | 1.9115243 | 0 | 0.0023504 |
| REACTOME_PD_1_SIGNALING | 16 | 0.7772079 | 1.9120072 | 0 | 0.002352996 |
| GO_MONOSACCHARIDE_CATABOLIC_PROCESS | 44 | -0.6106731 | -1.90483 | 0 | 0.002355508 |
| GO_OXIDOREDUCTASE_ACTIVITY_ACTING_ON_NAD_P_H | 68 | -0.5659488 | -1.90418 | 0 | 0.002358161 |
| GO_INTERLEUKIN_8_SECRETION | 21 | 0.7268214 | 1.9115696 | 0 | 0.002359954 |
| GO_OXIDOREDUCTASE_ACTIVITY_ACTING_ON_THE_ALDEHYDE_OR_OXO_GROUP_OF_DONORS | 33 | -0.6401753 | -1.903341 | 0 | 0.002365077 |
| GO_CARBON_OXYGEN_LYASE_ACTIVITY | 46 | -0.6143063 | -1.90297 | 0 | 0.002367521 |
| GO_FAD_BINDING | 27 | -0.6610065 | -1.901088 | 0 | 0.002419773 |
| GO_POSITIVE_REGULATION_OF_LEUKOCYTE_DIFFERENTIATION | 97 | 0.53179085 | 1.907974 | 0 | 0.002491137 |
| REACTOME_FCERI_MEDIATED_MAPK_ACTIVATION | 35 | 0.6327763 | 1.9066699 | 0 | 0.002527694 |
| REACTOME_TRANSPORT_OF_MATURE_TRANSCRIPT_TO_CYTOPLASM | 50 | 0.5905959 | 1.9060949 | 0 | 0.002532934 |
| GO_PROTEIN_TRIMERIZATION | 20 | 0.7155428 | 1.9051878 | 0 | 0.002538075 |
| GO_POSITIVE_REGULATION_OF_RESPONSE_TO_EXTERNAL_STIMULUS | 377 | 0.455889 | 1.9052163 | 0 | 0.002548147 |
| GO_BONE_CELL_DEVELOPMENT | 19 | 0.7348847 | 1.9053464 | 0 | 0.002553212 |
| KEGG_GLYCEROLIPID_METABOLISM | 29 | -0.6498272 | -1.896643 | 0 | 0.002587229 |
| HALLMARK_KRAS_SIGNALING_UP | 152 | 0.50504833 | 1.9040183 | 0 | 0.002598718 |
| GO_BILE_ACID_METABOLIC_PROCESS | 25 | -0.6670209 | -1.894605 | 0 | 0.002679149 |
| GO_ANION_TRANSMEMBRANE_TRANSPORTER_ACTIVITY | 176 | -0.4854138 | -1.893458 | 0 | 0.002700597 |
| GO_PEPTIDE_ANTIGEN_BINDING | 17 | 0.74259114 | 1.9022998 | 0.003861004 | 0.002703807 |
| REACTOME_SIGNALING_BY_INTERLEUKINS | 271 | 0.47030362 | 1.9019558 | 0 | 0.002713544 |
| PID_CASPASE_PATHWAY | 38 | 0.6397996 | 1.9006242 | 0 | 0.002720066 |
| GO_ANCHORING_JUNCTION | 499 | 0.4423401 | 1.9016914 | 0 | 0.002727657 |
| REACTOME_CELL_SURFACE_INTERACTIONS_AT_THE_VASCULAR_WALL | 117 | 0.514651 | 1.9007168 | 0 | 0.002730528 |
| GO_POSITIVE_REGULATION_OF_INTERFERON_GAMMA_PRODUCTION | 37 | 0.6363449 | 1.9008777 | 0 | 0.002731129 |
| GO_DNA_CONFORMATION_CHANGE | 127 | 0.5106425 | 1.9013053 | 0 | 0.002731875 |
| GO_ORGANIC_HYDROXY_COMPOUND_METABOLIC_PROCESS | 347 | -0.4505794 | -1.890282 | 0 | 0.002827297 |
| GO_POSITIVE_REGULATION_OF_INTERLEUKIN_2_PRODUCTION | 21 | 0.6957716 | 1.8977125 | 0.00204918 | 0.002871515 |
| GO_PATTERN_RECOGNITION_RECEPTOR_ACTIVITY | 17 | 0.75216246 | 1.8964726 | 0 | 0.002933481 |
| GO_DICARBOXYLIC_ACID_TRANSPORT | 59 | -0.5698445 | -1.887949 | 0 | 0.002936455 |
| GO_ANTIPORTER_ACTIVITY | 43 | -0.5997776 | -1.887437 | 0 | 0.002947695 |
| GO_ACETYL_COA_METABOLIC_PROCESS | 29 | -0.6493198 | -1.885883 | 0.002057613 | 0.002985003 |
| REACTOME_TRANSPORT_OF_BILE_SALTS_AND_ORGANIC_ACIDS_METAL_IONS_AND_AMINE_COMPOUNDS | 46 | -0.5847555 | -1.88616 | 0 | 0.002995077 |
| GO_MICROBODY_MEMBRANE | 39 | -0.6011162 | -1.886004 | 0 | 0.002997938 |
| REACTOME_RHO_GTPASE_EFFECTORS | 148 | 0.50207204 | 1.8942707 | 0 | 0.003046451 |
| REACTOME_COSTIMULATION_BY_THE_CD28_FAMILY | 41 | 0.6190852 | 1.894379 | 0 | 0.003053145 |
| GO_PLATELET_ACTIVATION | 104 | 0.51922166 | 1.8936002 | 0 | 0.003063884 |
| GO_AZUROPHIL_GRANULE | 105 | 0.5217162 | 1.8932687 | 0 | 0.003071395 |
| REACTOME_METABOLISM_OF_VITAMINS_AND_COFACTORS | 120 | -0.5129451 | -1.882062 | 0 | 0.003095991 |
| GO_MONOVALENT_INORGANIC_CATION_TRANSMEMBRANE_TRANSPORTER_ACTIVITY | 190 | -0.4758462 | -1.882288 | 0 | 0.003113384 |

| | | | | | |
|---|---|---|---|---|---|
| GO_LEUKOCYTE_HOMEOSTASIS | 54 | 0.58043617 | 1.8904544 | 0 | 0.003184574 |
| GO_NEGATIVE_REGULATION_OF_CYTOKINE_PRODUCTION | 167 | 0.496483 | 1.8899355 | 0 | 0.003210975 |
| REACTOME_SUMOYLATION_OF_DNA_REPLICATION_PROTEINS | 31 | 0.6464422 | 1.8898672 | 0 | 0.003213334 |
| KEGG_RIBOSOME | 31 | 0.6455981 | 1.8895444 | 0 | 0.003225496 |
| REACTOME_SYNTHESIS_OF_BILE_ACIDS_AND_BILE_SALTS_VIA_7ALPHA_HYDROXYCHOLESTEROL | 15 | -0.7558383 | -1.878961 | 0 | 0.003268981 |
| GO_INTRINSIC_COMPONENT_OF_MITOCHONDRIAL_INNER_MEMBRANE | 24 | -0.6819692 | -1.876994 | 0.003891051 | 0.003336258 |
| GO_BRUSH_BORDER_MEMBRANE | 39 | -0.6095166 | -1.874397 | 0 | 0.003422156 |
| GO_PURINE_CONTAINING_COMPOUND_METABOLIC_PROCESS | 272 | -0.4546028 | -1.873993 | 0 | 0.00342247 |
| GO_INTRINSIC_COMPONENT_OF_MITOCHONDRIAL_MEMBRANE | 39 | -0.6060885 | -1.874782 | 0 | 0.003425492 |
| REACTOME_PLATELET_AGGREGATION_PLUG_FORMATION | 26 | 0.673194 | 1.8858029 | 0 | 0.003468205 |
| GO_POSITIVE_REGULATION_OF_CHEMOKINE_PRODUCTION | 41 | 0.6126402 | 1.8855711 | 0 | 0.003488018 |
| GO_NEGATIVE_REGULATION_OF_REGULATED_SECRETORY_PATHWAY | 18 | 0.73747545 | 1.8844863 | 0 | 0.003536105 |
| GO_MHC_CLASS_II_PROTEIN_COMPLEX_BINDING | 16 | 0.7524027 | 1.8838422 | 0.001972387 | 0.003555837 |
| GO_LEUKOCYTE_ADHESION_TO_VASCULAR_ENDOTHELIAL_CELL | 29 | 0.6740821 | 1.8828613 | 0 | 0.003598648 |
| KEGG_PARKINSONS_DISEASE | 56 | -0.5627036 | -1.869385 | 0 | 0.003607471 |
| GO_HYDROLASE_ACTIVITY_ACTING_ON_CARBON_NITROGEN_BUT_NOT_PEPTIDE_BONDS_IN_LINEAR_AMIDES | 38 | -0.6074406 | -1.869763 | 0 | 0.003611897 |
| GO_POSITIVE_REGULATION_OF_CYTOKINE_PRODUCTION_INVOLVED_IN_IMMUNE_RESPONSE | 42 | 0.60602444 | 1.882244 | 0 | 0.003613247 |
| REACTOME_DEGRADATION_OF_BETA_CATENIN_BY_THE_DESTRUCTION_COMPLEX | 42 | 0.60847604 | 1.8815566 | 0 | 0.003627998 |
| GO_ESTABLISHMENT_OF_PROTEIN_LOCALIZATION_TO_ENDOPLASMIC_RETICULUM | 43 | 0.60382414 | 1.8805621 | 0.001908397 | 0.003661382 |
| GO_ARGININE_METABOLIC_PROCESS | 15 | -0.7650853 | -1.867396 | 0 | 0.003662992 |
| KEGG_ASTHMA | 20 | 0.71279246 | 1.8803021 | 0.002061856 | 0.003666735 |
| GO_MRNA_EXPORT_FROM_NUCLEUS | 59 | 0.5671179 | 1.8798989 | 0 | 0.003667833 |
| REACTOME_CD22_MEDIATED_BCR_REGULATION | 17 | 0.74339694 | 1.8800801 | 0.002020202 | 0.003671751 |
| GO_VIRAL_LIFE_CYCLE | 199 | 0.4802477 | 1.8790144 | 0 | 0.003673652 |
| GO_POSITIVE_REGULATION_OF_CYTOKINE_SECRETION | 83 | 0.54196054 | 1.8806669 | 0 | 0.003674505 |
| GO_NEGATIVE_REGULATION_OF_EXOCYTOSIS | 25 | 0.68428737 | 1.8794229 | 0 | 0.003677512 |
| REACTOME_G2_M_CHECKPOINTS | 72 | 0.55387205 | 1.8777561 | 0 | 0.003697487 |
| PID_TCR_PATHWAY | 47 | 0.597934 | 1.8769251 | 0 | 0.003702214 |
| REACTOME_DISEASES_ASSOCIATED_WITH_GLYCOSAMINOGLYCAN_METABOLISM | 25 | 0.68480617 | 1.8767465 | 0 | 0.003702697 |
| NABA_BASEMENT_MEMBRANES | 21 | 0.6906184 | 1.877812 | 0.003853565 | 0.00371037 |
| GO_B_CELL_PROLIFERATION | 61 | 0.56842595 | 1.8779467 | 0 | 0.003723344 |
| REACTOME_PROGRAMMED_CELL_DEATH | 109 | 0.51158124 | 1.8738527 | 0 | 0.003883467 |
| GO_LEUKOCYTE_ACTIVATION_INVOLVED_IN_INFLAMMATORY_RESPONSE | 34 | 0.6232885 | 1.8732831 | 0 | 0.003896314 |
| GO_POSITIVE_REGULATION_OF_ANION_TRANSPORT | 34 | -0.6236492 | -1.863398 | 0 | 0.00390054 |
| GO_POSITIVE_REGULATION_OF_LYMPHOCYTE_DIFFERENTIATION | 65 | 0.5587458 | 1.8719372 | 0 | 0.003944141 |
| GO_IMMUNOGLOBULIN_COMPLEX | 26 | 0.65773153 | 1.8710903 | 0.003960396 | 0.003978756 |
| GO_REGULATION_OF_CELL_KILLING | 59 | 0.56583935 | 1.8705186 | 0 | 0.004012946 |
| KEGG_CYTOKINE_CYTOKINE_RECEPTOR_INTERACTION | 158 | 0.49168873 | 1.8699671 | 0 | 0.004037381 |
| GO_NATURAL_KILLER_CELL_MEDIATED_IMMUNITY | 36 | 0.6282535 | 1.8700532 | 0 | 0.004038136 |
| KEGG_RETINOL_METABOLISM | 32 | -0.6314837 | -1.860294 | 0 | 0.004046662 |
| GO_CYTOKINE_BINDING | 83 | 0.5372416 | 1.8694243 | 0 | 0.004075505 |

| | | | | | |
|---|---|---|---|---|---|
| GO_REGULATION_OF_LEUKOCYTE_DIFFERENTIATION | 167 | 0.48709252 | 1.8688226 | 0 | 0.004083172 |
| GO_GLUTATHIONE_METABOLIC_PROCESS | 36 | -0.6152528 | -1.857716 | 0 | 0.004165387 |
| GO_POSITIVE_REGULATION_OF_TYPE_I_INTERFERON_PRODUCTION | 47 | 0.5871418 | 1.8669811 | 0 | 0.004167248 |
| REACTOME_SIGNALING_BY_RHO_GTPASES | 210 | 0.47104067 | 1.8657756 | 0 | 0.004190472 |
| GO_NEGATIVE_REGULATION_OF_IMMUNE_EFFECTOR_PROCESS | 85 | 0.53134495 | 1.8658084 | 0 | 0.004204394 |
| GO_NEURON_CELLULAR_HOMEOSTASIS | 21 | -0.6862416 | -1.85695 | 0 | 0.004205682 |
| GO_IMMUNOGLOBULIN_PRODUCTION | 74 | 0.5491948 | 1.8650126 | 0 | 0.004206525 |
| GO_NEGATIVE_REGULATION_OF_CELL_KILLING | 16 | 0.75073344 | 1.8643813 | 0 | 0.004247525 |
| GO_ANTIGEN_PROCESSING_AND_PRESENTATION_OF_PEPTIDE_ANTIGEN_VIA_MHC_CLASS_I | 67 | 0.54843205 | 1.8628465 | 0 | 0.004300361 |
| GO_LUMENAL_SIDE_OF_MEMBRANE | 22 | 0.711561 | 1.8633596 | 0.002020202 | 0.004304973 |
| REACTOME_MITOTIC_G1_PHASE_AND_G1_S_TRANSITION | 86 | 0.5282641 | 1.8629761 | 0 | 0.004310218 |
| GO_RESPONSE_TO_CHEMOKINE | 70 | 0.55529547 | 1.8631462 | 0 | 0.00431169 |
| REACTOME_RESPONSE_OF_EIF2AK4_GCN2_TO_AMINO_ACID_DEFICIENCY | 37 | 0.6111798 | 1.8615701 | 0.002057613 | 0.004352982 |
| REACTOME_MITOTIC_PROPHASE | 47 | 0.5890039 | 1.8597956 | 0.001926782 | 0.004379138 |
| KEGG_FOCAL_ADHESION | 139 | 0.49505237 | 1.860267 | 0 | 0.004382466 |
| REACTOME_DEFECTS_IN_VITAMIN_AND_COFACTOR_METABOLISM | 16 | -0.7303231 | -1.853358 | 0 | 0.004384226 |
| GO_DENDRITIC_CELL_MIGRATION | 19 | 0.7072877 | 1.8607657 | 0.001904762 | 0.004384299 |
| GO_NEUROINFLAMMATORY_RESPONSE | 47 | 0.58882433 | 1.8598193 | 0 | 0.004393174 |
| REACTOME_CHEMOKINE_RECEPTORS_BIND_CHEMOKINES | 43 | 0.60316646 | 1.8590609 | 0.002008032 | 0.004406027 |
| GO_STEROID_BIOSYNTHETIC_PROCESS | 132 | -0.4898218 | -1.851188 | 0 | 0.004454267 |
| KEGG_PYRUVATE_METABOLISM | 29 | -0.6383074 | -1.851275 | 0 | 0.00447373 |
| GO_DNA_GEOMETRIC_CHANGE | 67 | 0.5464533 | 1.8575627 | 0 | 0.004532051 |
| PID_IL2_STAT5_PATHWAY | 19 | 0.72241676 | 1.8576509 | 0 | 0.00453426 |
| REACTOME_INFECTIOUS_DISEASE | 387 | 0.44734395 | 1.8564544 | 0 | 0.004594475 |
| GO_LEUKOCYTE_APOPTOTIC_PROCESS | 73 | 0.54619896 | 1.8551819 | 0 | 0.004640475 |
| GO_MONOCARBOXYLIC_ACID_BINDING | 46 | -0.5752016 | -1.846926 | 0 | 0.004678551 |
| HALLMARK_APICAL_JUNCTION | 134 | 0.49193147 | 1.8542447 | 0 | 0.004682229 |
| GO_TERTIARY_GRANULE | 113 | 0.5135837 | 1.8519222 | 0 | 0.004860573 |
| GO_ENDOCYTIC_VESICLE_MEMBRANE | 97 | 0.514233 | 1.8519683 | 0 | 0.004867798 |
| GO_APOPTOTIC_CELL_CLEARANCE | 27 | 0.6490846 | 1.8507957 | 0 | 0.00488269 |
| GO_CELLULAR_RESPONSE_TO_INTERFERON_BETA | 15 | 0.7523828 | 1.8508292 | 0 | 0.004893822 |
| PID_CD8_TCR_PATHWAY | 40 | 0.6008279 | 1.8508298 | 0 | 0.00490902 |
| GO_CELLULAR_EXTRAVASATION | 45 | 0.58671516 | 1.8496717 | 0 | 0.004923418 |
| GO_ORGANIC_ACID_TRANSMEMBRANE_TRANSPORT | 80 | -0.5230595 | -1.843043 | 0 | 0.004937092 |
| GO_RESPIRATORY_BURST | 22 | 0.6806771 | 1.849707 | 0 | 0.004938567 |
| GO_TYPE_I_INTERFERON_PRODUCTION | 72 | 0.5380754 | 1.8488048 | 0.001908397 | 0.004967011 |
| GO_ANTIBACTERIAL_HUMORAL_RESPONSE | 21 | 0.69932014 | 1.847935 | 0 | 0.004994941 |
| GO_METAL_CLUSTER_BINDING | 42 | -0.5852802 | -1.841886 | 0 | 0.00501855 |
| GO_ALCOHOL_CATABOLIC_PROCESS | 30 | -0.628474 | -1.841082 | 0.002066116 | 0.005049814 |
| REACTOME_SWITCHING_OF_ORIGINS_TO_A_POST_REPLICATIVE_STATE | 45 | 0.5916848 | 1.8468579 | 0 | 0.005073594 |
| PID_TOLL_ENDOGENOUS_PATHWAY | 16 | 0.75765604 | 1.8464363 | 0.001926782 | 0.005093214 |
| HALLMARK_ANGIOGENESIS | 36 | 0.60656935 | 1.8451651 | 0.001838235 | 0.005131959 |
| GO_REGULATION_OF_TOLL_LIKE_RECEPTOR_SIGNALING_PATHWAY | 41 | 0.59601474 | 1.845369 | 0.001876173 | 0.005147464 |
| GO_NEGATIVE_REGULATION_OF_IMMUNE_SYSTEM_PROCESS | 288 | 0.45394167 | 1.8442992 | 0 | 0.005170491 |
| GO_PLASMA_MEMBRANE_SIGNALING_RECEPTOR_COMPLEX | 121 | 0.49239883 | 1.8443319 | 0 | 0.005186018 |
| GO_ACETYL_COA_BIOSYNTHETIC_PROCESS | 18 | -0.7012864 | -1.838196 | 0.002132196 | 0.005230089 |

| | | | | | |
|---|---|---|---|---|---|
| REACTOME_METABOLISM_OF_LIPIDS | 440 | -0.4284234 | -1.838639 | 0 | 0.005231448 |
| GO_ORGANIC_ANION_TRANSPORT | 275 | -0.4453558 | -1.837015 | 0 | 0.005308169 |
| GO_CHROMATIN_ASSEMBLY | 45 | 0.5903303 | 1.8421751 | 0 | 0.005331069 |
| REACTOME_SYNDECAN_INTERACTIONS | 23 | 0.6696713 | 1.8416697 | 0 | 0.00536462 |
| GO_PROTEIN_TRANSMEMBRANE_IMPORT_INTO_INTRACELLULAR_ORGANELLE | 16 | -0.7371217 | -1.834697 | 0 | 0.005441852 |
| REACTOME_DISEASES_OF_IMMUNE_SYSTEM | 15 | 0.7540804 | 1.8397864 | 0 | 0.005473718 |
| REACTOME_THE_ROLE_OF_NEF_IN_HIV_1_REPLICATION_AND_DISEASE_PATHOGENESIS | 21 | 0.6799508 | 1.8393481 | 0.002012072 | 0.005480514 |
| REACTOME_CELL_CYCLE | 315 | 0.44639105 | 1.8374931 | 0 | 0.005653687 |
| GO_GLIAL_CELL_ACTIVATION | 38 | 0.61161035 | 1.8365663 | 0 | 0.005699198 |
| GO_NEGATIVE_REGULATION_OF_LEUKOCYTE_PROLIFERATION | 51 | 0.56892425 | 1.8367622 | 0 | 0.005704572 |
| GO_POSITIVE_REGULATION_OF_INTERFERON_BETA_PRODUCTION | 21 | 0.69796664 | 1.8360243 | 0 | 0.005727188 |
| REACTOME_ANTIGEN_PRESENTATION_FOLDING_ASSEMBLY_AND_PEPTIDE_LOADING_OF_CLASS_I_MHC | 23 | 0.67151123 | 1.8357722 | 0.001919386 | 0.005733083 |
| PID_AVB3_OPN_PATHWAY | 26 | 0.6540243 | 1.8352424 | 0.001984127 | 0.005735206 |
| GO_REGULATION_OF_CELL_CELL_ADHESION | 251 | 0.4557978 | 1.8346157 | 0 | 0.005766787 |
| GO_ENDODERM_FORMATION | 28 | 0.63922477 | 1.8343753 | 0 | 0.005783437 |
| REACTOME_CELL_CYCLE_MITOTIC | 265 | 0.44998667 | 1.8337945 | 0 | 0.005829661 |
| REACTOME_SEMAPHORIN_INTERACTIONS | 42 | 0.587494 | 1.832915 | 0 | 0.005905082 |
| REACTOME_DISEASES_OF_CARBOHYDRATE_METABOLISM | 24 | -0.6559328 | -1.827554 | 0 | 0.005947209 |
| GO_REGULATION_OF_T_CELL_MEDIATED_IMMUNITY | 49 | 0.5698808 | 1.8312645 | 0 | 0.006075013 |
| REACTOME_EUKARYOTIC_TRANSLATION_INITIATION | 48 | 0.57738954 | 1.8311999 | 0 | 0.006083511 |
| GO_NEGATIVE_REGULATION_OF_ADAPTIVE_IMMUNE_RESPONSE | 31 | 0.63257384 | 1.8307351 | 0 | 0.006085326 |
| REACTOME_CASPASE_ACTIVATION_VIA_EXTRINSIC_APOPTOTIC_SIGNALLING_PATHWAY | 18 | 0.7177797 | 1.8309209 | 0.001941748 | 0.006087945 |
| BIOCARTA_CSK_PATHWAY | 17 | 0.71609557 | 1.8287401 | 0.001984127 | 0.006249169 |
| GO_DICARBOXYLIC_ACID_TRANSMEMBRANE_TRANSPORTER_ACTIVITY | 22 | -0.6791351 | -1.822935 | 0 | 0.006363778 |
| REACTOME_MITOTIC_METAPHASE_AND_ANAPHASE | 122 | 0.49667528 | 1.827227 | 0.001814882 | 0.006372224 |
| PID_ATR_PATHWAY | 21 | 0.6881359 | 1.8273814 | 0 | 0.006382943 |
| REACTOME_DAP12_INTERACTIONS | 29 | 0.6341069 | 1.8267044 | 0.001858736 | 0.006415246 |
| GO_POSITIVE_REGULATION_OF_AMYLOID_PRECURSOR_PROTEIN_CATABOLIC_PROCESS | 15 | 0.73009926 | 1.8260372 | 0.002083333 | 0.006472695 |
| SA_CASPASE_CASCADE | 16 | 0.73951584 | 1.8255911 | 0 | 0.006486715 |
| GO_ENDOCYTIC_VESICLE | 175 | 0.47094634 | 1.8246738 | 0 | 0.006540112 |
| GO_IMMUNOGLOBULIN_PRODUCTION_INVOLVED_IN_IMMUNOGLOBULIN_MEDIATED_IMMUNE_RESPONSE | 28 | 0.6376654 | 1.8233571 | 0.003717472 | 0.006642843 |
| GO_NEGATIVE_REGULATION_OF_LYMPHOCYTE_ACTIVATION | 96 | 0.5121954 | 1.8228024 | 0 | 0.006662953 |
| GO_LYMPHOCYTE_COSTIMULATION | 39 | 0.60202575 | 1.8229483 | 0 | 0.006663786 |
| GO_ANTIBIOTIC_METABOLIC_PROCESS | 79 | -0.5223505 | -1.818837 | 0 | 0.006745469 |
| GO_TRANSLATIONAL_ELONGATION | 62 | -0.5417367 | -1.816022 | 0 | 0.006860492 |
| GO_TRIGLYCERIDE_METABOLIC_PROCESS | 55 | -0.5515726 | -1.817113 | 0 | 0.006862422 |
| GO_NUCLEOBASE_CONTAINING_SMALL_MOLECULE_METABOLIC_PROCESS | 376 | -0.4300331 | -1.816624 | 0 | 0.006863031 |
| GO_OXIDOREDUCTASE_ACTIVITY_ACTING_ON_PAIRED_DONORS_WITH_INCORPORATION_OR_REDUCTION_OF_MOLECULAR_OXYGEN_REDUCED_FLAVIN_OR_FLAVOPROTEIN_AS_ONE_DONOR_AND_INCORPORATION_OF_ONE_ATOM_OF_OXYGEN | 22 | -0.6707935 | -1.816279 | 0.00204499 | 0.006876773 |
| GO_LEUKOCYTE_TETHERING_OR_ROLLING | 20 | 0.6804414 | 1.8194633 | 0.002020202 | 0.00693632 |

| | | | | | |
|---|---|---|---|---|---|
| REACTOME_CYTOSOLIC_SENSORS_OF_PATHOGEN_ASSOCIATED_DNA | 32 | 0.6140157 | 1.8191098 | 0.001841621 | 0.006942004 |
| GO_INTERFERON_ALPHA_PRODUCTION | 17 | 0.710266 | 1.81947 | 0.001923077 | 0.006955428 |
| BIOCARTA_FAS_PATHWAY | 25 | 0.6552678 | 1.8180157 | 0 | 0.007017886 |
| GO_RNA_EXPORT_FROM_NUCLEUS | 74 | 0.5342862 | 1.8181518 | 0 | 0.00702296 |
| GO_PODOSOME | 16 | 0.7269542 | 1.8182777 | 0.002020202 | 0.007031558 |
| PID_BETA_CATENIN_NUC_PATHWAY | 45 | 0.5673678 | 1.8173006 | 0 | 0.007072037 |
| GO_INNER_MITOCHONDRIAL_MEMBRANE_ORGANIZATION | 16 | -0.7007189 | -1.813138 | 0.00422833 | 0.007093727 |
| REACTOME_ORC1_REMOVAL_FROM_CHROMATIN | 38 | 0.6045481 | 1.8167167 | 0 | 0.00712546 |
| REACTOME_HIV_INFECTION | 133 | 0.4850139 | 1.8162671 | 0 | 0.0071338 |
| GO_AMMONIUM_TRANSMEMBRANE_TRANSPORTER_ACTIVITY | 17 | -0.7129486 | -1.811156 | 0.003883495 | 0.007238688 |
| GO_CELL_CHEMOTAXIS | 184 | 0.4669102 | 1.8146781 | 0 | 0.007242385 |
| GO_CELLULAR_RESPONSE_TO_COPPER_ION | 20 | -0.6786845 | -1.809976 | 0 | 0.007299055 |
| GO_LEUKOCYTE_CHEMOTAXIS | 139 | 0.48276192 | 1.8140587 | 0 | 0.007302082 |
| GO_CELLULAR_MODIFIED_AMINO_ACID_BIOSYNTHETIC_PROCESS | 32 | -0.602761 | -1.810054 | 0.002004008 | 0.007320263 |
| GO_REGULATION_OF_IMMUNOGLOBULIN_PRODUCTION | 39 | 0.57778597 | 1.8137004 | 0.001883239 | 0.007320433 |
| GO_COPII_COATED_ER_TO_GOLGI_TRANSPORT_VESICLE | 57 | 0.55670387 | 1.8127408 | 0 | 0.007389587 |
| GO_POSITIVE_REGULATION_OF_NF_KAPPAB_TRANSCRIPTION_FACTOR_ACTIVITY | 96 | 0.50437796 | 1.8116136 | 0 | 0.007475102 |
| GO_REGULATION_OF_MYELOID_LEUKOCYTE_MEDIATED_IMMUNITY | 47 | 0.5670843 | 1.8105953 | 0.001949318 | 0.007533532 |
| REACTOME_INTERACTIONS_OF_REV_WITH_HOST_CELLULAR_PROTEINS | 23 | 0.6405576 | 1.8104428 | 0.001824818 | 0.007533972 |
| GO_REGULATION_OF_HEMOPOIESIS | 260 | 0.4509797 | 1.810183 | 0 | 0.007551226 |
| REACTOME_NS1_MEDIATED_EFFECTS_ON_HOST_PATHWAYS | 25 | 0.6488481 | 1.8086427 | 0 | 0.00771664 |
| REACTOME_METABOLIC_DISORDERS_OF_BIOLOGICAL_OXIDATION_ENZYMES | 23 | -0.6482832 | -1.805439 | 0.001988072 | 0.00771983 |
| GO_MYELOID_CELL_DIFFERENTIATION | 229 | 0.45503825 | 1.8082857 | 0 | 0.007740074 |
| REACTOME_CREATION_OF_C4_AND_C2_ACTIVATORS | 21 | 0.6670426 | 1.8078175 | 0.001988072 | 0.00776313 |
| GO_STEROID_METABOLIC_PROCESS | 214 | -0.4511363 | -1.804314 | 0 | 0.007791968 |
| GO_MONOOXYGENASE_ACTIVITY | 62 | -0.5365728 | -1.802939 | 0 | 0.007946898 |
| GO_ORGANIC_CATION_TRANSPORT | 19 | -0.6935481 | -1.801471 | 0.003913894 | 0.008055529 |
| GO_RNA_DEPENDENT_DNA_BIOSYNTHETIC_PROCESS | 33 | 0.6122596 | 1.8048507 | 0.002012072 | 0.008060598 |
| GO_WOUND_HEALING | 355 | 0.43466786 | 1.8045862 | 0 | 0.008063118 |
| KEGG_TOLL_LIKE_RECEPTOR_SIGNALING_PATHWAY | 70 | 0.53228736 | 1.8039811 | 0 | 0.00807098 |
| GO_REGULATION_OF_MYELOID_LEUKOCYTE_DIFFERENTIATION | 70 | 0.5252759 | 1.804009 | 0 | 0.008091944 |
| GO_ANION_TRANSPORT | 353 | -0.4305647 | -1.800319 | 0 | 0.00810034 |
| GO_PROTEIN_COMPLEX_INVOLVED_IN_CELL_ADHESION | 23 | 0.65254295 | 1.8033429 | 0.002 | 0.008132584 |
| GO_OXIDOREDUCTASE_ACTIVITY_ACTING_ON_PAIRED_DONORS_WITH_INCORPORATION_OR_REDUCTION_OF_MOLECULAR_OXYGEN | 93 | -0.5041464 | -1.80041 | 0 | 0.008134305 |
| REACTOME_NUCLEAR_PORE_COMPLEX_NPC_DISASSEMBLY | 23 | 0.67026865 | 1.8020684 | 0 | 0.00828968 |
| GO_CATALYTIC_STEP_2_SPLICEOSOME | 43 | 0.56931305 | 1.8000506 | 0 | 0.008475858 |
| GO_MYELOID_CELL_DEVELOPMENT | 41 | 0.5843202 | 1.7995642 | 0.001945525 | 0.00850322 |
| GO_POSITIVE_REGULATION_OF_CELL_KILLING | 40 | 0.5810219 | 1.799184 | 0 | 0.008537209 |
| REACTOME_CYTOCHROME_P450_ARRANGED_BY_SUBSTRATE_TYPE | 42 | -0.5794048 | -1.794793 | 0 | 0.008617147 |
| GO_POSITIVE_REGULATION_OF_LYMPHOCYTE_MEDIATED_IMMUNITY | 61 | 0.5465996 | 1.7982309 | 0 | 0.008636211 |
| GO_INNATE_IMMUNE_RESPONSE_ACTIVATING_CELL_SURFACE_RECEPTOR_SIGNALING_PATHWAY | 62 | 0.5382991 | 1.797281 | 0 | 0.008643181 |

| | | | | | |
|---|---|---|---|---|---|
| GO_CELL_MATRIX_ADHESION | 138 | 0.47979015 | 1.7979485 | 0 | 0.008653296 |
| GO_POSITIVE_REGULATION_OF_NIK_NF_KAPPAB_SIGNALING | 49 | 0.56424546 | 1.7973245 | 0 | 0.008661817 |
| GO_RESPONSE_TO_WOUNDING | 425 | 0.4242234 | 1.7969698 | 0 | 0.008663098 |
| GO_AZUROPHIL_GRANULE_MEMBRANE | 36 | 0.5939268 | 1.7966639 | 0 | 0.008664524 |
| GO_CYTOKINE_RECEPTOR_ACTIVITY | 67 | 0.5322765 | 1.7967544 | 0 | 0.008676627 |
| GO_REGULATION_OF_LEUKOCYTE_MEDIATED_CYTOTOXICITY | 48 | 0.5651169 | 1.7974191 | 0 | 0.008677249 |
| GO_REGULATION_OF_LEUKOCYTE_MIGRATION | 135 | 0.47428307 | 1.796158 | 0 | 0.008678138 |
| GO_MYD88_INDEPENDENT_TOLL_LIKE_RECEPTOR_SIGNALING_PATHWAY | 19 | 0.7000581 | 1.7974368 | 0.001912046 | 0.008692833 |
| GO_POSITIVE_REGULATION_OF_T_CELL_MIGRATION | 21 | 0.68877095 | 1.7958722 | 0 | 0.008694793 |
| GO_FICOLIN_1_RICH_GRANULE_MEMBRANE | 35 | 0.5986547 | 1.7956654 | 0.003780718 | 0.008705054 |
| REACTOME_SIGNALING_BY_PDGF | 44 | 0.56618696 | 1.7955766 | 0.001945525 | 0.008709041 |
| GO_IRON_ION_BINDING | 89 | -0.4937702 | -1.792232 | 0 | 0.008866061 |
| GO_RUFFLE_MEMBRANE | 55 | 0.5445377 | 1.7928365 | 0 | 0.008950111 |
| GO_CELL_ADHESION_MEDIATED_BY_INTEGRIN | 49 | 0.5640111 | 1.7924681 | 0 | 0.008956591 |
| GO_GLUTAMATE_METABOLIC_PROCESS | 19 | -0.6810638 | -1.790934 | 0.002028398 | 0.008977446 |
| GO_PROTEIN_DNA_COMPLEX_SUBUNIT_ORGANIZATION | 114 | 0.48963904 | 1.790718 | 0 | 0.009120597 |
| GO_CARBOHYDRATE_BIOSYNTHETIC_PROCESS | 120 | -0.4807083 | -1.788734 | 0 | 0.009238484 |
| GO_REGULATION_OF_INFLAMMATORY_RESPONSE | 207 | 0.45003125 | 1.7889309 | 0 | 0.009280657 |
| REACTOME_SIGNALING_BY_THE_B_CELL_RECEPTOR_BCR | 76 | 0.5159654 | 1.7881864 | 0 | 0.009287954 |
| GO_RESPONSE_TO_BACTERIUM | 367 | 0.42727834 | 1.7884735 | 0 | 0.009294919 |
| REACTOME_APC_C_MEDIATED_DEGRADATION_OF_CELL_CYCLE_PROTEINS | 44 | 0.5717625 | 1.7884923 | 0.001980198 | 0.009314466 |
| REACTOME_FCGR_ACTIVATION | 19 | 0.6921594 | 1.7878002 | 0.001960784 | 0.009321244 |
| REACTOME_FCERI_MEDIATED_CA_2_MOBILIZATION | 33 | 0.6015926 | 1.7862061 | 0.003809524 | 0.009478954 |
| GO_TISSUE_REMODELING | 111 | 0.48429003 | 1.7848556 | 0 | 0.009607201 |
| REACTOME_PYRUVATE_METABOLISM | 22 | -0.6403963 | -1.785021 | 0.006410257 | 0.009612306 |
| KEGG_REGULATION_OF_ACTIN_CYTOSKELETON | 138 | 0.47183713 | 1.7837685 | 0 | 0.009696567 |
| GO_INTERLEUKIN_6_BIOSYNTHETIC_PROCESS | 17 | 0.6971457 | 1.7838874 | 0.002012072 | 0.009701351 |
| GO_PATTERN_RECOGNITION_RECEPTOR_SIGNALING_PATHWAY | 117 | 0.4793498 | 1.7830663 | 0 | 0.009744167 |
| GO_HOST_CELLULAR_COMPONENT | 42 | 0.5733153 | 1.7821639 | 0 | 0.009818866 |
| REACTOME_SEMA4D_IN_SEMAPHORIN_SIGNALING | 17 | 0.700698 | 1.7798057 | 0.005928854 | 0.010113544 |
| GO_POSITIVE_REGULATION_OF_NEUTROPHIL_MIGRATION | 21 | 0.6758559 | 1.7799802 | 0.001923077 | 0.010119459 |
| GO_NUCLEOSOME_ORGANIZATION | 55 | 0.54217744 | 1.7792504 | 0 | 0.010123473 |
| GO_REGULATION_OF_T_HELPER_1_TYPE_IMMUNE_RESPONSE | 16 | 0.69973326 | 1.7785287 | 0.009823183 | 0.010130335 |
| GO_POSITIVE_REGULATION_OF_LYMPHOCYTE_MIGRATION | 25 | 0.64478844 | 1.7794329 | 0.001897533 | 0.010134935 |
| GO_IMMUNOGLOBULIN_RECEPTOR_BINDING | 15 | 0.7431819 | 1.7784224 | 0 | 0.010139692 |
| GO_REGULATION_OF_T_CELL_MIGRATION | 27 | 0.64187 | 1.7792804 | 0 | 0.010144448 |
| GO_REGULATION_OF_LYMPHOCYTE_MIGRATION | 39 | 0.57309055 | 1.7789584 | 0 | 0.010144843 |
| KEGG_CYTOSOLIC_DNA_SENSING_PATHWAY | 32 | 0.60753226 | 1.77862 | 0 | 0.010151146 |
| REACTOME_TRANSPORT_OF_MATURE_MRNAS_DERIVED_FROM_INTRONLESS_TRANSCRIPTS | 24 | 0.64779955 | 1.7786707 | 0.001956947 | 0.010172023 |
| GO_POSITIVE_REGULATION_OF_LEUKOCYTE_MIGRATION | 89 | 0.5079082 | 1.7777436 | 0 | 0.010184966 |
| GO_CHROMATIN_ASSEMBLY_OR_DISASSEMBLY | 58 | 0.547014 | 1.7775028 | 0.002004008 | 0.010200051 |
| GO_BENZENE_CONTAINING_COMPOUND_METABOLIC_PROCESS | 16 | -0.7019629 | -1.779401 | 0.002155172 | 0.010372362 |
| GO_REGULATION_OF_TYPE_I_INTERFERON_MEDIATED_SIGNALING_PATHWAY | 23 | 0.64355713 | 1.775071 | 0.005940594 | 0.010509428 |
| GO_SODIUM_ION_TRANSMEMBRANE_TRANSPORTER_ACTIVITY | 89 | -0.4972163 | -1.777582 | 0 | 0.010559179 |
| GO_ER_TO_GOLGI_TRANSPORT_VESICLE_MEMBRANE | 38 | 0.57888263 | 1.7742687 | 0 | 0.010618453 |
| GO_REGULATION_OF_NATURAL_KILLER_CELL_MEDIATED_IMMUNITY | 22 | 0.66482925 | 1.7739489 | 0.01183432 | 0.010644137 |
| GO_NEGATIVE_REGULATION_OF_CELL_ACTIVATION | 134 | 0.47610906 | 1.7737956 | 0 | 0.010649176 |

| | | | | | |
|---|---|---|---|---|---|
| GO_TOLL_LIKE_RECEPTOR_4_SIGNALING_PATHWAY | 23 | 0.6498282 | 1.7731646 | 0.001996008 | 0.010694211 |
| GO_TRANSFORMING_GROWTH_FACTOR_BETA_PRODUCTION | 31 | 0.61020535 | 1.7732034 | 0.004140787 | 0.01070993 |
| REACTOME_DIGESTION_AND_ABSORPTION | 16 | -0.7030869 | -1.775882 | 0 | 0.010719122 |
| GO_MITOCHONDRIAL_LARGE_RIBOSOMAL_SUBUNIT | 29 | -0.6225713 | -1.774786 | 0.002096436 | 0.010843786 |
| BIOCARTA_NUCLEARRS_PATHWAY | 25 | -0.6340479 | -1.773776 | 0.008264462 | 0.010936707 |
| GO_T_CELL_ACTIVATION_INVOLVED_IN_IMMUNE_RESPONSE | 64 | 0.5202159 | 1.7709132 | 0 | 0.010968997 |
| GO_POSITIVE_REGULATION_OF_IMMUNOGLOBULIN_PRODUCTION | 25 | 0.64520985 | 1.7701758 | 0 | 0.011021363 |
| REACTOME_BINDING_AND_UPTAKE_OF_LIGANDS_BY_SCAVENGER_RECEPTORS | 42 | 0.5660968 | 1.7702341 | 0 | 0.011040596 |
| GO_REGULATION_OF_INTRINSIC_APOPTOTIC_SIGNALING_PATHWAY_IN_RESPONSE_TO_DNA_DAMAGE | 23 | 0.6338945 | 1.7696068 | 0.005660377 | 0.01106343 |
| GO_NEGATIVE_REGULATION_OF_SIGNAL_TRANSDUCTION_BY_P53_CLASS_MEDIATOR | 18 | 0.6936956 | 1.7681127 | 0.001923077 | 0.011221292 |
| GO_RAC_PROTEIN_SIGNAL_TRANSDUCTION | 25 | 0.63475597 | 1.7671977 | 0.003853565 | 0.01132919 |
| GO_POSITIVE_REGULATION_OF_MYELOID_CELL_DIFFERENTIATION | 54 | 0.54323393 | 1.7667719 | 0 | 0.01133292 |
| GO_REGULATION_OF_NEUROINFLAMMATORY_RESPONSE | 22 | 0.650582 | 1.7669364 | 0.006097561 | 0.011341081 |
| GO_INTERLEUKIN_1_SECRETION | 33 | 0.58690107 | 1.7661003 | 0.003824092 | 0.011439742 |
| REACTOME_SUMOYLATION_OF_DNA_DAMAGE_RESPONSE_AND_REPAIR_PROTEINS | 41 | 0.5642523 | 1.7648242 | 0 | 0.011618169 |
| GO_MONOCARBOXYLIC_ACID_TRANSPORT | 102 | -0.4841091 | -1.768433 | 0 | 0.011685844 |
| GO_COLLAGEN_FIBRIL_ORGANIZATION | 36 | 0.5908755 | 1.763519 | 0.004056795 | 0.011741132 |
| GO_PHAGOCYTOSIS_RECOGNITION | 20 | 0.6686002 | 1.7635474 | 0.007751938 | 0.011764473 |
| PID_CXCR4_PATHWAY | 68 | 0.5290884 | 1.7631501 | 0 | 0.011775064 |
| REACTOME_SUMOYLATION_OF_UBIQUITINYLATION_PROTEINS | 22 | 0.645972 | 1.7623106 | 0.002057613 | 0.011891376 |
| PID_HNF3B_PATHWAY | 35 | -0.58903 | -1.764668 | 0 | 0.01212409 |
| KEGG_ALZHEIMERS_DISEASE | 92 | -0.4892317 | -1.764907 | 0.00204499 | 0.012127907 |
| GO_ORGANELLE_ENVELOPE_LUMEN | 57 | -0.530922 | -1.764939 | 0 | 0.012165162 |
| REACTOME_MITOTIC_SPINDLE_CHECKPOINT | 49 | 0.54876643 | 1.7597274 | 0.001964637 | 0.012337104 |
| GO_SPLICEOSOMAL_COMPLEX | 91 | 0.4932805 | 1.7590064 | 0 | 0.01241475 |
| REACTOME_FC_EPSILON_RECEPTOR_FCERI_SIGNALING | 92 | 0.49194625 | 1.758513 | 0 | 0.012475075 |
| PID_ILK_PATHWAY | 29 | 0.60724205 | 1.758026 | 0.001876173 | 0.012482024 |
| GO_LYMPHOCYTE_DIFFERENTIATION | 205 | 0.44227502 | 1.7582144 | 0 | 0.012484165 |
| GO_ENDODERM_DEVELOPMENT | 43 | 0.5696615 | 1.7574986 | 0 | 0.012567474 |
| GO_T_CELL_MEDIATED_CYTOTOXICITY | 31 | 0.6024553 | 1.7553166 | 0.003663004 | 0.012871271 |
| NABA_ECM_GLYCOPROTEINS | 103 | 0.48270774 | 1.7547929 | 0 | 0.01289586 |
| GO_REGULATORY_T_CELL_DIFFERENTIATION | 20 | 0.67182493 | 1.754844 | 0 | 0.01291559 |
| GO_NUCLEAR_TRANSCRIBED_MRNA_CATABOLIC_PROCESS | 82 | 0.5078205 | 1.7541586 | 0 | 0.012962633 |
| REACTOME_STIMULI_SENSING_CHANNELS | 46 | -0.550761 | -1.758334 | 0 | 0.012999379 |
| GO_NEUTRAL_LIPID_CATABOLIC_PROCESS | 22 | -0.6541477 | -1.757352 | 0.002118644 | 0.013148708 |
| GO_DETECTION_OF_BIOTIC_STIMULUS | 21 | 0.65392274 | 1.7527351 | 0.003868472 | 0.013200823 |
| GO_ACTIVE_TRANSMEMBRANE_TRANSPORTER_ACTIVITY | 207 | -0.4390776 | -1.756766 | 0 | 0.013211611 |
| GO_T_CELL_DIFFERENTIATION | 146 | 0.45726708 | 1.752136 | 0 | 0.013272251 |
| GO_NATURAL_KILLER_CELL_ACTIVATION | 48 | 0.55166596 | 1.7497706 | 0.00189394 | 0.013348261 |
| GO_CHEMOKINE_ACTIVITY | 32 | 0.589123 | 1.7495764 | 0 | 0.013355323 |
| PID_INTEGRIN2_PATHWAY | 24 | 0.6294019 | 1.7493919 | 0.001930502 | 0.013359412 |
| GO_INTRINSIC_COMPONENT_OF_ENDOPLASMIC_RETICULUM_MEMBRANE | 92 | 0.4882445 | 1.749152 | 0 | 0.013360998 |
| REACTOME_FCGAMMA_RECEPTOR_FCGR_DEPENDENT_PHAGOCYTOSIS | 70 | 0.5151949 | 1.7498382 | 0 | 0.013363081 |

| | | | | | |
|---|---|---|---|---|---|
| GO_POSITIVE_REGULATION_OF_LEUKOCYTE_MEDIATED_IMMUNITY | 86 | 0.49648687 | 1.7502176 | 0 | 0.013373482 |
| GO_U2_TYPE_SPLICEOSOMAL_COMPLEX | 42 | 0.56183314 | 1.7498938 | 0.00390625 | 0.013377871 |
| REACTOME_LAMININ_INTERACTIONS | 23 | 0.6471307 | 1.7484292 | 0.001992032 | 0.013384263 |
| GO_B_CELL_HOMEOSTASIS | 18 | 0.6716714 | 1.7506629 | 0.00625 | 0.013387995 |
| GO_MHC_PROTEIN_BINDING | 23 | 0.64274603 | 1.7508034 | 0.002008032 | 0.013389286 |
| REACTOME_TRANSPORT_OF_THE_SLBP_DEPENDANT_MATURE_MRNA | 23 | 0.64251196 | 1.750467 | 0.001937985 | 0.013389494 |
| GO_COLLAGEN_BINDING | 47 | 0.5601815 | 1.7486598 | 0.001992032 | 0.01339217 |
| GO_SPECIFIC_GRANULE | 116 | 0.47248897 | 1.7485305 | 0 | 0.013396327 |
| GO_CHEMOKINE_RECEPTOR_BINDING | 40 | 0.5702765 | 1.7510524 | 0.001934236 | 0.013398754 |
| GO_VESICLE_MEMBRANE | 468 | 0.41021836 | 1.7502186 | 0 | 0.013402119 |
| REACTOME_HEMOSTASIS | 415 | 0.41247654 | 1.7475965 | 0 | 0.013486397 |
| GO_CYTOSOLIC_LARGE_RIBOSOMAL_SUBUNIT | 19 | 0.66782176 | 1.7476332 | 0.003976143 | 0.013509269 |
| KEGG_PRIMARY_IMMUNODEFICIENCY | 23 | 0.6403496 | 1.7471155 | 0.002012072 | 0.013511885 |
| REACTOME_INTERLEUKIN_4_AND_INTERLEUKIN_13_SIGNALING | 77 | 0.5045052 | 1.7461901 | 0 | 0.013598611 |
| GO_POSITIVE_REGULATION_OF_INTERLEUKIN_1_SECRETION | 19 | 0.66744274 | 1.7454202 | 0.003944773 | 0.013681741 |
| GO_ANTIGEN_PROCESSING_AND_PRESENTATION_OF_ENDOGENOUS_ANTIGEN | 20 | 0.6578534 | 1.7449242 | 0.002024292 | 0.013730197 |
| GO_INTRINSIC_APOPTOTIC_SIGNALING_PATHWAY_IN_RESPONSE_TO_DNA_DAMAGE | 62 | 0.52187085 | 1.7439991 | 0 | 0.013854951 |
| GO_CARBOHYDRATE_CATABOLIC_PROCESS | 135 | -0.4598354 | -1.751856 | 0 | 0.01394188 |
| REACTOME_INTERLEUKIN_2_FAMILY_SIGNALING | 25 | 0.6356844 | 1.7417544 | 0.003853565 | 0.014164561 |
| GO_INTERFERON_GAMMA_SECRETION | 16 | 0.69633156 | 1.7412665 | 0.004106776 | 0.014204252 |
| GO_STEROID_HYDROXYLASE_ACTIVITY | 23 | -0.6218666 | -1.749942 | 0.00867679 | 0.014215243 |
| GO_LIGAND_ACTIVATED_TRANSCRIPTION_FACTOR_ACTIVITY | 29 | -0.6059482 | -1.749174 | 0.002155172 | 0.014278861 |
| GO_REGULATION_OF_EPITHELIAL_CELL_DIFFERENTIATION_INVOLVED_IN_KIDNEY_DEVELOPMENT | 15 | 0.69920295 | 1.7403276 | 0.001851852 | 0.014351461 |
| GO_RNA_CATABOLIC_PROCESS | 183 | 0.44624707 | 1.7397088 | 0 | 0.014424467 |
| GO_REGULATION_OF_T_CELL_RECEPTOR_SIGNALING_PATHWAY | 30 | 0.59924185 | 1.7382486 | 0.005769231 | 0.014591442 |
| GO_POSITIVE_REGULATION_OF_LEUKOCYTE_MEDIATED_CYTOTOXICITY | 34 | 0.5766268 | 1.7384597 | 0.001980198 | 0.014604005 |
| REACTOME_C_TYPE_LECTIN_RECEPTORS_CLRS | 80 | 0.496283 | 1.7382658 | 0 | 0.014618627 |
| REACTOME_ELASTIC_FIBRE_FORMATION | 34 | 0.58272123 | 1.7372041 | 0.001901141 | 0.01471126 |
| REACTOME_EXPORT_OF_VIRAL_RIBONUCLEOPROTEINS_FROM_NUCLEUS | 22 | 0.6278371 | 1.7373267 | 0.003921569 | 0.014720329 |
| GO_HOMOTYPIC_CELL_CELL_ADHESION | 57 | 0.52415705 | 1.7349658 | 0.00204918 | 0.014996194 |
| GO_MEMBRANE_RAFT_ORGANIZATION | 19 | 0.6833251 | 1.7350098 | 0.001945525 | 0.015013521 |
| GO_ACTIN_FILAMENT_ORGANIZATION | 222 | 0.43866095 | 1.7351197 | 0 | 0.015028438 |
| GO_TOXIN_METABOLIC_PROCESS | 18 | -0.6779697 | -1.743621 | 0.004065041 | 0.015104622 |
| GO_POSITIVE_REGULATION_OF_ORGANIC_ACID_TRANSPORT | 25 | -0.6186417 | -1.743843 | 0.002024292 | 0.015121008 |
| GO_ANION_TRANSMEMBRANE_TRANSPORT | 163 | -0.4480861 | -1.742534 | 0 | 0.015235642 |
| REACTOME_ANTIMICROBIAL_PEPTIDES | 28 | 0.6054803 | 1.7334163 | 0.001890359 | 0.015236286 |
| GO_RESPONSE_TO_X_RAY | 19 | 0.6688233 | 1.7332189 | 0.001988072 | 0.015236569 |
| GO_RECEPTOR_COMPLEX | 239 | 0.43550164 | 1.732442 | 0 | 0.015341581 |
| GO_FC_EPSILON_RECEPTOR_SIGNALING_PATHWAY | 79 | 0.50137 | 1.7318411 | 0 | 0.015426477 |
| GO_REGULATION_OF_ACTIN_FILAMENT_BASED_PROCESS | 222 | 0.43523625 | 1.7314488 | 0 | 0.015477064 |
| REACTOME_RHO_GTPASES_ACTIVATE_WASPS_AND_WAVES | 25 | 0.62911236 | 1.730885 | 0.005791506 | 0.015507275 |
| REACTOME_NUCLEAR_ENVELOPE_BREAKDOWN | 36 | 0.58225185 | 1.7310176 | 0.005660377 | 0.015507622 |
| REACTOME_DNA_REPLICATION | 70 | 0.50490206 | 1.7300869 | 0 | 0.015624032 |
| GO_AMINO_ACID_TRANSMEMBRANE_TRANSPORTER_ACTIVITY | 39 | -0.5558227 | -1.737451 | 0 | 0.015840074 |

| | | | | | |
|---|---|---|---|---|---|
| GO_CELLULAR_RESPONSE_TO_CAMP | 36 | -0.5770848 | -1.737621 | 0 | 0.015867341 |
| GO_MONOVALENT_INORGANIC_ANION_HOMEOSTASIS | 18 | -0.667864 | -1.738134 | 0.00203252 | 0.015914345 |
| GO_NADP_BINDING | 40 | -0.5600087 | -1.737758 | 0.002150538 | 0.015914964 |
| KEGG_METABOLISM_OF_XENOBIOTICS_BY_CYTOCHROME_P450 | 36 | -0.5828493 | -1.738324 | 0.004123712 | 0.015930975 |
| GO_TETRAPYRROLE_BINDING | 88 | -0.4830247 | -1.736217 | 0 | 0.01595436 |
| GO_THIOLESTER_HYDROLASE_ACTIVITY | 17 | -0.68898 | -1.736524 | 0.004081633 | 0.015962556 |
| GO_RIBOSE_PHOSPHATE_METABOLIC_PROCESS | 246 | -0.4234478 | -1.735565 | 0 | 0.016002763 |
| BIOCARTA_HIVNEF_PATHWAY | 44 | 0.55103946 | 1.726695 | 0.003577818 | 0.01627975 |
| GO_REGULATION_OF_ACTIN_FILAMENT_ORGANIZATION | 143 | 0.46066338 | 1.7263892 | 0 | 0.016300688 |
| GO_RESPONSE_TO_XENOBIOTIC_STIMULUS | 185 | -0.4405798 | -1.733928 | 0 | 0.016325483 |
| REACTOME_RHO_GTPASES_ACTIVATE_PKNS | 22 | 0.6391379 | 1.7245789 | 0.001968504 | 0.016637258 |
| REACTOME_NUCLEAR_IMPORT_OF_REV_PROTEIN | 22 | 0.6315089 | 1.724029 | 0.001890359 | 0.016733032 |
| GO_SOLUTE_SODIUM_SYMPORTER_ACTIVITY | 47 | -0.5360341 | -1.731516 | 0 | 0.016743239 |
| GO_MACROPHAGE_MIGRATION | 33 | 0.588556 | 1.7233459 | 0.001937985 | 0.016848687 |
| GO_MEMBRANE_REGION | 208 | 0.43665704 | 1.7217547 | 0 | 0.017004633 |
| REACTOME_SNRNP_ASSEMBLY | 28 | 0.61209834 | 1.7217656 | 0.001915709 | 0.01703535 |
| GO_NIK_NF_KAPPAB_SIGNALING | 102 | 0.47274438 | 1.7219758 | 0 | 0.017036015 |
| PID_EPHB_FWD_PATHWAY | 27 | 0.611388 | 1.7220553 | 0.002053388 | 0.017056772 |
| GO_HETEROTYPIC_CELL_CELL_ADHESION | 43 | 0.54503393 | 1.7209498 | 0.001980198 | 0.01711514 |
| GO_PROTEIN_LOCALIZATION_TO_CHROMOSOME | 32 | 0.59077275 | 1.7210696 | 0.001838235 | 0.017123548 |
| GO_HORMONE_METABOLIC_PROCESS | 152 | -0.4517011 | -1.728464 | 0 | 0.017240884 |
| GO_REGULATION_OF_DEFENSE_RESPONSE_TO_VIRUS_BY_HOST | 18 | 0.6698461 | 1.7200502 | 0.007984032 | 0.017249854 |
| PID_RB_1PATHWAY | 36 | 0.5735227 | 1.7201079 | 0 | 0.017268198 |
| GO_MULTI_ORGANISM_LOCALIZATION | 36 | 0.56980914 | 1.7191292 | 0.003868472 | 0.017385365 |
| GO_ANTIMICROBIAL_HUMORAL_RESPONSE | 59 | 0.5182842 | 1.7191918 | 0.004264392 | 0.01740177 |
| REACTOME_S_PHASE | 87 | 0.48255366 | 1.718273 | 0 | 0.01745865 |
| GO_ACTIN_POLYMERIZATION_OR_DEPOLYMERIZATION | 113 | 0.46559805 | 1.7182943 | 0 | 0.017492224 |
| GO_P53_BINDING | 38 | 0.5589396 | 1.717332 | 0.001824818 | 0.017607458 |
| PID_FANCONI_PATHWAY | 25 | 0.6137004 | 1.7170546 | 0.001956947 | 0.017634813 |
| REACTOME_GPVI_MEDIATED_ACTIVATION_CASCADE | 26 | 0.6171333 | 1.7157227 | 0.009861933 | 0.017824486 |
| GO_REGULATION_OF_LEUKOCYTE_ADHESION_TO_VASCULAR_ENDOTHELIAL_CELL | 15 | 0.69976133 | 1.7159754 | 0.004246285 | 0.017826762 |
| GO_CARBOHYDRATE_METABOLIC_PROCESS | 375 | -0.406332 | -1.725009 | 0 | 0.017936679 |
| GO_ONE_CARBON_METABOLIC_PROCESS | 18 | -0.664619 | -1.724506 | 0.003952569 | 0.017987147 |
| REACTOME_METABOLISM_OF_STEROIDS | 101 | -0.4730768 | -1.723882 | 0 | 0.01807577 |
| GO_POSITIVE_REGULATION_OF_ALPHA_BETA_T_CELL_ACTIVATION | 44 | 0.5479378 | 1.7141314 | 0.004048583 | 0.018129326 |
| GO_POSITIVE_REGULATION_OF_TOLL_LIKE_RECEPTOR_SIGNALING_PATHWAY | 15 | 0.6992984 | 1.713508 | 0.005628518 | 0.018216096 |
| GO_RAS_PROTEIN_SIGNAL_TRANSDUCTION | 235 | 0.4282045 | 1.7126923 | 0 | 0.018310033 |
| HALLMARK_HEME_METABOLISM | 141 | -0.4538485 | -1.722184 | 0 | 0.018337274 |
| GO_ARP2_3_COMPLEX_MEDIATED_ACTIN_NUCLEATION | 18 | 0.6706848 | 1.7119555 | 0.009242144 | 0.01840867 |
| REACTOME_DNA_REPLICATION_PRE_INITIATION | 48 | 0.5285092 | 1.7115415 | 0 | 0.018485278 |
| GO_DNA_PACKAGING | 63 | 0.5143872 | 1.7108079 | 0 | 0.018562218 |
| GO_CELL_SUBSTRATE_ADHESION | 208 | 0.43328717 | 1.710618 | 0 | 0.018580418 |
| GO_ANAPHASE_PROMOTING_COMPLEX_DEPENDENT_CATABOLIC_PROCESS | 42 | 0.54591155 | 1.710831 | 0 | 0.01858763 |
| GO_REGULATION_OF_SYNCYTIUM_FORMATION_BY_PLASMA_MEMBRANE_FUSION | 22 | 0.63565695 | 1.710312 | 0.004032258 | 0.018620068 |
| GO_INTERLEUKIN_10_PRODUCTION | 31 | 0.5839433 | 1.7100401 | 0.006122449 | 0.01865012 |
| REACTOME_COLLAGEN_FORMATION | 55 | 0.53014433 | 1.7093813 | 0 | 0.018687889 |
| GO_METHYLTRANSFERASE_COMPLEX | 47 | 0.54203796 | 1.7095352 | 0.003759399 | 0.01869415 |

| | | | | | |
|---|---|---|---|---|---|
| GO_AZUROPHIL_GRANULE_LUMEN | 62 | 0.513894 | 1.708709 | 0 | 0.018705258 |
| GO_RHO_PROTEIN_SIGNAL_TRANSDUCTION | 94 | 0.48124743 | 1.7090887 | 0 | 0.018715212 |
| GO_REGULATION_OF_TELOMERE_MAINTENANCE_VIA_TELOMERE_LENGTHENING | 27 | 0.59400064 | 1.7087841 | 0.005847953 | 0.018737586 |
| GO_SYMPORTER_ACTIVITY | 84 | -0.4925414 | -1.719594 | 0 | 0.018764533 |
| GO_POSITIVE_REGULATION_OF_LIPID_KINASE_ACTIVITY | 23 | 0.6220794 | 1.7069287 | 0.005905512 | 0.019083396 |
| GO_HORMONE_BIOSYNTHETIC_PROCESS | 60 | -0.5063064 | -1.717752 | 0.002096436 | 0.019088516 |
| GO_REGULATION_OF_LEUKOCYTE_DEGRANULATION | 40 | 0.55796486 | 1.7067391 | 0.001964637 | 0.01909775 |
| REACTOME_TRANSPORT_OF_INORGANIC_CATIONS_ANIONS_AND_AMINO_ACIDS_OLIGOPEPTIDES | 64 | -0.5130606 | -1.717967 | 0 | 0.019100036 |
| REACTOME_SEPARATION_OF_SISTER_CHROMATIDS | 93 | 0.48118278 | 1.7058742 | 0 | 0.019241767 |
| REACTOME_COLLAGEN_BIOSYNTHESIS_AND_MODIFYING_ENZYMES | 42 | 0.5380042 | 1.7048862 | 0.004016064 | 0.019415397 |
| REACTOME_TRANSPORT_OF_SMALL_MOLECULES | 406 | -0.4036948 | -1.716117 | 0 | 0.019428808 |
| GO_NUCLEOID | 25 | -0.6132253 | -1.715729 | 0.003883495 | 0.019431895 |
| GO_LIGASE_ACTIVITY | 70 | -0.495098 | -1.715297 | 0 | 0.019450855 |
| GO_REGULATION_OF_LYMPHOCYTE_DIFFERENTIATION | 102 | 0.469148 | 1.7037238 | 0 | 0.019622112 |
| GO_SYNCYTIUM_FORMATION | 34 | 0.5820542 | 1.7025582 | 0.001904762 | 0.01971461 |
| GO_PROTEIN_LOCALIZATION_TO_ENDOPLASMIC_RETICULUM | 57 | 0.5311912 | 1.7027017 | 0 | 0.019724978 |
| GO_REGULATION_OF_T_CELL_DIFFERENTIATION | 84 | 0.48624858 | 1.702734 | 0.001956947 | 0.019756375 |
| GO_ACTIN_CYTOSKELETON_REORGANIZATION | 66 | 0.50576025 | 1.7027588 | 0 | 0.019787889 |
| REACTOME_SUMOYLATION_OF_SUMOYLATION_PROTEINS | 23 | 0.6182354 | 1.7017927 | 0 | 0.019860024 |
| PID_RXR_VDR_PATHWAY | 15 | -0.683107 | -1.713091 | 0.006198347 | 0.019872596 |
| GO_NEGATIVE_REGULATION_OF_MYELOID_LEUKOCYTE_DIFFERENTIATION | 30 | 0.5814878 | 1.7010523 | 0.007984032 | 0.019984335 |
| GO_ALPHA_BETA_T_CELL_PROLIFERATION | 23 | 0.62332267 | 1.7008579 | 0 | 0.019987606 |
| GO_REGULATION_OF_DEFENSE_RESPONSE_TO_VIRUS_BY_VIRUS | 22 | 0.6280562 | 1.6997366 | 0.008316008 | 0.02017854 |
| REACTOME_M_PHASE | 181 | 0.44161123 | 1.6998116 | 0 | 0.020201124 |
| GO_POSITIVE_REGULATION_OF_DNA_BINDING_TRANSCRIPTION_FACTOR_ACTIVITY | 154 | 0.44036636 | 1.6984742 | 0 | 0.02037765 |
| GO_TOLERANCE_INDUCTION | 15 | 0.690063 | 1.6986076 | 0.005952381 | 0.020386644 |
| GO_NEGATIVE_REGULATION_OF_B_CELL_ACTIVATION | 26 | 0.6032256 | 1.6981194 | 0.001937985 | 0.020439696 |
| GO_ION_TRANSMEMBRANE_TRANSPORTER_ACTIVITY | 426 | -0.3990083 | -1.710101 | 0 | 0.020473443 |
| GO_REGULATION_OF_VIRAL_ENTRY_INTO_HOST_CELL | 20 | 0.64029866 | 1.6973132 | 0.008032128 | 0.020614292 |
| REACTOME_PLATELET_ACTIVATION_SIGNALING_AND_AGGREGATION | 181 | 0.43690464 | 1.6967962 | 0 | 0.020694118 |
| GO_CELL_SUBSTRATE_JUNCTION_ORGANIZATION | 66 | 0.5010454 | 1.6956283 | 0.005454545 | 0.020929307 |
| REACTOME_NUCLEOTIDE_BINDING_DOMAIN_LEUCINE_RICH_REPEAT_CONTAINING_RECEPTOR_NLR_SIGNALING_PATHWAYS | 27 | 0.59600735 | 1.6950967 | 0.008097166 | 0.021021899 |
| GO_DNA_DEPENDENT_ATPASE_ACTIVITY | 58 | 0.51485425 | 1.6945528 | 0 | 0.021134786 |
| REACTOME_ROLE_OF_LAT2_NTAL_LAB_ON_CALCIUM_MOBILIZATION | 22 | 0.6226121 | 1.6938533 | 0.005882353 | 0.021242749 |
| GO_STEROID_DEHYDROGENASE_ACTIVITY | 23 | -0.618545 | -1.705797 | 0.004048583 | 0.021314163 |
| GO_CENTROMERE_COMPLEX_ASSEMBLY | 20 | 0.6494599 | 1.6932243 | 0.007889546 | 0.021348601 |
| GO_EPOXYGENASE_P450_PATHWAY | 16 | -0.6862529 | -1.704855 | 0.004056795 | 0.021457104 |
| GO_REGULATION_OF_TUMOR_NECROSIS_FACTOR_MEDIATED_SIGNALING_PATHWAY | 35 | 0.574664 | 1.6917504 | 0.003773585 | 0.021607991 |
| REACTOME_MOLECULES_ASSOCIATED_WITH_ELASTIC_FIBRES | 30 | 0.58564585 | 1.6917944 | 0 | 0.021639414 |
| GO_SITE_OF_DNA_DAMAGE | 30 | 0.58314615 | 1.6913314 | 0.005780347 | 0.021684866 |
| GO_VESICLE_LUMEN | 235 | 0.4228141 | 1.6909518 | 0 | 0.021723604 |
| GO_FATTY_ACID_DERIVATIVE_BINDING | 21 | -0.6357935 | -1.703245 | 0.00625 | 0.021750353 |

| | | | | | |
|---|---|---|---|---|---|
| GO_DNA_REPLICATION_INDEPENDENT_NUCLEOSOME_ORGANIZATION | 21 | 0.63339216 | 1.6902903 | 0.0056926 | 0.021868045 |
| GO_REGULATION_OF_LYMPHOCYTE_APOPTOTIC_PROCESS | 34 | 0.56208974 | 1.6894041 | 0.007782101 | 0.02204553 |
| GO_STEROID_HORMONE_BIOSYNTHETIC_PROCESS | 43 | -0.535329 | -1.701605 | 0.004149378 | 0.022054467 |
| GO_STRUCTURAL_CONSTITUENT_OF_NUCLEAR_PORE | 16 | 0.6751597 | 1.6887785 | 0.006072875 | 0.022092998 |
| GO_T_HELPER_1_TYPE_IMMUNE_RESPONSE | 25 | 0.6121235 | 1.6888067 | 0.001834862 | 0.022116018 |
| BIOCARTA_STATHMIN_PATHWAY | 16 | 0.68765396 | 1.6888173 | 0.011881189 | 0.022152582 |
| GO_CATION_TRANSMEMBRANE_TRANSPORTER_ACTIVITY | 316 | -0.4085113 | -1.699009 | 0 | 0.022543734 |
| PID_IL4_2PATHWAY | 40 | 0.5577841 | 1.6863905 | 0.001945525 | 0.022677004 |
| GO_ATP_METABOLIC_PROCESS | 161 | -0.4405173 | -1.697629 | 0 | 0.022827353 |
| GO_REGULATION_OF_CYTOSKELETON_ORGANIZATION | 289 | 0.40936306 | 1.6855781 | 0 | 0.022840688 |
| GO_CELL_ADHESION_MEDIATOR_ACTIVITY | 35 | 0.5561966 | 1.6853932 | 0.005882353 | 0.022863315 |
| GO_ANTIGEN_PROCESSING_AND_PRESENTATION_OF_PEPTIDE_OR_POLYSACCHARIDE_ANTIGEN_VIA_MHC_CLASS_II | 76 | 0.48253965 | 1.6849443 | 0 | 0.022925982 |
| GO_REGULATION_OF_PEPTIDASE_ACTIVITY | 256 | 0.4160501 | 1.6841068 | 0 | 0.02301537 |
| GO_MEMBRANE_PROTEIN_PROTEOLYSIS | 33 | 0.5702751 | 1.6841645 | 0.001992032 | 0.02304621 |
| GO_REGULATION_OF_CENTROSOME_CYCLE | 24 | 0.6075114 | 1.6835492 | 0.0041841 | 0.023112783 |
| GO_DEFENSE_RESPONSE_TO_GRAM_NEGATIVE_BACTERIUM | 38 | 0.56033796 | 1.6820844 | 0.00407332 | 0.023436677 |
| GO_ACTIVE_ION_TRANSMEMBRANE_TRANSPORTER_ACTIVITY | 144 | -0.4480317 | -1.694868 | 0 | 0.023451755 |
| REACTOME_ASSEMBLY_OF_THE_PRE_REPLICATIVE_COMPLEX | 36 | 0.5549041 | 1.6820966 | 0.007366483 | 0.023477016 |
| GO_INTRINSIC_APOPTOTIC_SIGNALING_PATHWAY_IN_RESPONSE_TO_DNA_DAMAGE_BY_P53_CLASS_MEDIATOR | 26 | 0.6018673 | 1.6814866 | 0.00990099 | 0.023487804 |
| GO_GROWTH_PLATE_CARTILAGE_DEVELOPMENT | 21 | 0.62590677 | 1.6816144 | 0.014799154 | 0.023501705 |
| REACTOME_INTEGRIN_SIGNALING | 18 | 0.656464 | 1.6812582 | 0.011952192 | 0.023502542 |
| GO_MEGAKARYOCYTE_DIFFERENTIATION | 35 | 0.5649942 | 1.6805964 | 0.005714286 | 0.023619357 |
| GO_REGULATION_OF_MYELOID_CELL_DIFFERENTIATION | 127 | 0.45219064 | 1.6799878 | 0 | 0.023702452 |
| NABA_ECM_AFFILIATED | 78 | 0.4860753 | 1.6800374 | 0.001795332 | 0.023727512 |
| GO_DETOXIFICATION | 86 | -0.4730031 | -1.693282 | 0 | 0.023753159 |
| REACTOME_SIGNALING_BY_NOTCH4 | 49 | 0.5362325 | 1.6792799 | 0.001941748 | 0.023860235 |
| GO_POSITIVE_REGULATION_OF_RHO_PROTEIN_SIGNAL_TRANSDUCTION | 15 | 0.6854918 | 1.6786959 | 0.010121457 | 0.02389693 |
| GO_REGULATION_OF_TYPE_2_IMMUNE_RESPONSE | 24 | 0.60287625 | 1.6788011 | 0.01369863 | 0.023902781 |
| GO_RECEPTOR_SIGNALING_PATHWAY_VIA_STAT | 89 | 0.47673452 | 1.6788723 | 0 | 0.023927987 |
| REACTOME_HCMV_EARLY_EVENTS | 47 | 0.5293545 | 1.6781845 | 0.003752345 | 0.023968542 |
| GO_ACTIN_FILAMENT | 68 | 0.49845013 | 1.6773127 | 0.001949318 | 0.024167396 |
| GO_NEGATIVE_REGULATION_OF_CELL_ADHESION | 182 | 0.4313139 | 1.6768949 | 0 | 0.024245316 |
| GO_NEGATIVE_REGULATION_OF_TUMOR_NECROSIS_FACTOR_SUPERFAMILY_CYTOKINE_PRODUCTION | 36 | 0.5490038 | 1.6765596 | 0.003824092 | 0.024256341 |
| REACTOME_RESOLUTION_OF_SISTER_CHROMATID_COHESION | 61 | 0.51010615 | 1.6765906 | 0.002 | 0.024294844 |
| REACTOME_SUMOYLATION_OF_CHROMATIN_ORGANIZATION_PROTEINS | 35 | 0.5577472 | 1.6755427 | 0.007575758 | 0.024456032 |
| GO_U2_TYPE_CATALYTIC_STEP_2_SPLICEOSOME | 16 | 0.6715388 | 1.6742799 | 0.006160164 | 0.024636555 |
| GO_NEGATIVE_REGULATION_OF_LEUKOCYTE_DIFFERENTIATION | 67 | 0.49863476 | 1.6746165 | 0.001941748 | 0.024637453 |
| GO_GROWTH_FACTOR_BINDING | 91 | 0.47434017 | 1.674348 | 0 | 0.024656365 |
| REACTOME_SUMOYLATION_OF_RNA_BINDING_PROTEINS | 27 | 0.59146446 | 1.6735038 | 0.003649635 | 0.024776958 |
| GO_AMMONIUM_ION_METABOLIC_PROCESS | 115 | -0.4576749 | -1.688987 | 0 | 0.02480693 |
| GO_MEMBRANE_PROTEIN_ECTODOMAIN_PROTEOLYSIS | 28 | 0.5900332 | 1.6717826 | 0.011363637 | 0.025195071 |
| REACTOME_LEISHMANIA_INFECTION | 155 | 0.44170275 | 1.6713641 | 0 | 0.025263514 |
| REACTOME_TNFR2_NON_CANONICAL_NF_KB_PATHWAY | 60 | 0.50111026 | 1.670837 | 0.00204499 | 0.025321702 |
| GO_TELOMERE_MAINTENANCE_VIA_TELOMERE_LENGTHENING | 38 | 0.5485078 | 1.6708436 | 0.001923077 | 0.025363557 |
| GO_TRIGLYCERIDE_CATABOLIC_PROCESS | 17 | -0.6581905 | -1.685867 | 0.006437768 | 0.025593976 |
| GO_ENTRY_INTO_HOST | 88 | 0.47623435 | 1.6691828 | 0 | 0.025761737 |

| | | | | | |
|---|---|---|---|---|---|
| GO_HELICASE_ACTIVITY | 77 | 0.48549792 | 1.6689686 | 0 | 0.025776384 |
| REACTOME_METABOLISM_OF_PORPHYRINS | 19 | -0.6499764 | -1.684268 | 0.006465518 | 0.025990862 |
| GO_REGULATION_OF_GLUCOSE_METABOLIC_PROCESS | 65 | -0.4967241 | -1.683882 | 0.002020202 | 0.026005458 |
| KEGG_PENTOSE_PHOSPHATE_PATHWAY | 20 | -0.6385914 | -1.68329 | 0.012121212 | 0.02606809 |
| GO_EPIBOLY | 18 | 0.6534677 | 1.6674231 | 0.003891051 | 0.026158063 |
| GO_TERTIARY_GRANULE_MEMBRANE | 49 | 0.51948285 | 1.6667968 | 0.005847953 | 0.026287587 |
| GO_POTASSIUM_ION_TRANSMEMBRANE_TRANSPORTER_ACTIVITY | 74 | -0.4825526 | -1.682061 | 0.002173913 | 0.02632065 |
| GO_CALCIUM_DEPENDENT_PROTEIN_BINDING | 41 | 0.5481855 | 1.6663119 | 0.00390625 | 0.026380956 |
| GO_PRIMARY_ALCOHOL_METABOLIC_PROCESS | 56 | -0.5149146 | -1.680574 | 0 | 0.026599173 |
| GO_AROMATASE_ACTIVITY | 17 | -0.6541303 | -1.680651 | 0.010460251 | 0.026671799 |
| PID_IL12_2PATHWAY | 44 | 0.5271382 | 1.6647032 | 0 | 0.026739782 |
| GO_ASPARTATE_FAMILY_AMINO_ACID_BIOSYNTHETIC_PROCESS | 17 | -0.6621228 | -1.679754 | 0.01004016 | 0.026749985 |
| GO_EXTRACELLULAR_MATRIX_BINDING | 40 | 0.554153 | 1.6644546 | 0.005882353 | 0.026771484 |
| REACTOME_SIGNALING_BY_MET | 49 | 0.52339387 | 1.6636944 | 0.003676471 | 0.026925553 |
| GO_SUBSTRATE_DEPENDENT_CELL_MIGRATION | 19 | 0.639548 | 1.6633731 | 0.004008016 | 0.026942315 |
| GO_ACID_THIOL_LIGASE_ACTIVITY | 16 | -0.6677406 | -1.677818 | 0.012552301 | 0.027140979 |
| GO_ACTIVATION_OF_PROTEIN_KINASE_B_ACTIVITY | 24 | -0.5969382 | -1.677407 | 0.004065041 | 0.027163396 |
| GO_PYRIDINE_CONTAINING_COMPOUND_METABOLIC_PROCESS | 20 | -0.6316068 | -1.677928 | 0.010752688 | 0.027202083 |
| PID_AJDISS_2PATHWAY | 32 | 0.56771666 | 1.6619453 | 0.007366483 | 0.02731837 |
| GO_LYMPHOCYTE_CHEMOTAXIS | 38 | 0.5437863 | 1.6614903 | 0.005813954 | 0.027370797 |
| REACTOME_REGULATION_OF_GLUCOKINASE_BY_GLUCOKINASE_REGULATORY_PROTEIN | 20 | 0.6247465 | 1.6616005 | 0.004158004 | 0.027373804 |
| GO_POSITIVE_REGULATION_OF_B_CELL_PROLIFERATION | 27 | 0.5830027 | 1.6610013 | 0.018 | 0.027424812 |
| GO_CHROMATIN_REMODELING | 66 | 0.50036156 | 1.661102 | 0.001968504 | 0.027433846 |
| GO_POSITIVE_REGULATION_OF_CYTOSOLIC_CALCIUM_ION_CONCENTRATION_INVOLVED_IN_PHOSPHOLIPASE_C_ACTIVATING_G_PROTEIN_COUPLED_SIGNALING_PATHWAY | 15 | 0.6778151 | 1.6605473 | 0.010245902 | 0.027485646 |
| REACTOME_GLUTATHIONE_CONJUGATION | 22 | -0.6182438 | -1.675791 | 0.008316008 | 0.02754587 |
| GO_POSITIVE_REGULATION_OF_T_CELL_MEDIATED_IMMUNITY | 36 | 0.5500166 | 1.65928 | 0.004008016 | 0.027746927 |
| BIOCARTA_IL2RB_PATHWAY | 25 | 0.60011923 | 1.6594102 | 0.009727626 | 0.027758416 |
| PID_FCER1_PATHWAY | 42 | 0.54421324 | 1.6589795 | 0.00591716 | 0.02780514 |
| GO_NAD_METABOLIC_PROCESS | 32 | -0.561177 | -1.673434 | 0.004201681 | 0.028139286 |
| GO_PLATELET_AGGREGATION | 44 | 0.52881545 | 1.6576602 | 0.001901141 | 0.028146572 |
| GO_PROTEASE_BINDING | 79 | 0.47766152 | 1.657006 | 0.001883239 | 0.028298333 |
| REACTOME_PROCESSING_OF_CAPPED_INTRON_CONTAINING_PRE_MRNA | 129 | 0.44954106 | 1.6556801 | 0 | 0.028596144 |
| REACTOME_INTERACTIONS_OF_VPR_WITH_HOST_CELLULAR_PROTEINS | 24 | 0.6074439 | 1.6557341 | 0.01459854 | 0.028617326 |
| GO_REGULATION_OF_INTRINSIC_APOPTOTIC_SIGNALING_PATHWAY_BY_P53_CLASS_MEDIATOR | 16 | 0.66174453 | 1.6552981 | 0.010548524 | 0.028660288 |
| GO_NUCLEAR_CHROMOSOME_TELOMERIC_REGION | 53 | 0.51572186 | 1.6549635 | 0.001890359 | 0.028708553 |
| GO_RESPONSE_TO_CADMIUM_ION | 46 | -0.522063 | -1.671012 | 0.006198347 | 0.028747227 |
| GO_RNA_LOCALIZATION | 120 | 0.44192514 | 1.6544493 | 0 | 0.028819472 |
| REACTOME_PHASE_II_CONJUGATION_OF_COMPOUNDS | 57 | -0.5047249 | -1.669879 | 0.00204499 | 0.029023852 |
| KEGG_CELL_CYCLE | 73 | 0.48795527 | 1.653131 | 0.001934236 | 0.029179372 |
| REACTOME_INTERLEUKIN_3_INTERLEUKIN_5_AND_GM_CSF_SIGNALING | 29 | 0.5823517 | 1.6527202 | 0.01004016 | 0.029247236 |
| GO_TISSUE_HOMEOSTASIS | 123 | 0.44728768 | 1.6527749 | 0 | 0.02926307 |
| GO_PROTEIN_CONTAINING_COMPLEX_LOCALIZATION | 155 | 0.43692103 | 1.6522349 | 0 | 0.02932027 |
| GO_POSITIVE_REGULATION_OF_GLIOGENESIS | 49 | 0.5167256 | 1.6522555 | 0 | 0.029364418 |

| | | | | | |
|---|---|---|---|---|---|
| GO_POSITIVE_REGULATION_OF_GRANULOCYTE_CHEMOTAXIS | 20 | 0.628544 | 1.6514325 | 0.007905139 | 0.029405117 |
| GO_TYPE_2_IMMUNE_RESPONSE | 27 | 0.5834718 | 1.6517044 | 0.01372549 | 0.029414903 |
| GO_NCRNA_EXPORT_FROM_NUCLEUS | 25 | 0.59304374 | 1.651551 | 0.010080645 | 0.029414933 |
| GO_CD8_POSITIVE_ALPHA_BETA_T_CELL_ACTIVATION | 15 | 0.6641966 | 1.6507384 | 0.009708738 | 0.029545076 |
| PID_IL6_7_PATHWAY | 35 | 0.5462061 | 1.6507871 | 0.005940594 | 0.029575257 |
| PID_BARD1_PATHWAY | 18 | 0.6491995 | 1.6489297 | 0.011494253 | 0.029872658 |
| GO_POSITIVE_REGULATION_OF_STRIATED_MUSCLE_CELL_DIFFERENTIATION | 29 | 0.5739834 | 1.6490667 | 0.011650485 | 0.0298893 |
| BIOCARTA_NKT_PATHWAY | 16 | 0.6548335 | 1.6491778 | 0.02020202 | 0.02991195 |
| HALLMARK_IL2_STAT5_SIGNALING | 137 | 0.43947825 | 1.6479244 | 0 | 0.030002715 |
| REACTOME_RHO_GTPASE_CYCLE | 70 | 0.48099992 | 1.6480062 | 0.003868472 | 0.030013315 |
| GO_REGULATION_OF_ACTOMYOSIN_STRUCTURE_ORGANIZATION | 56 | 0.5043977 | 1.6480678 | 0 | 0.03004196 |
| GO_NEGATIVE_REGULATION_OF_LEUKOCYTE_CELL_CELL_ADHESION | 77 | 0.4842787 | 1.6481202 | 0.001876173 | 0.030066663 |
| GO_BLOOD_MICROPARTICLE | 91 | 0.47186673 | 1.6475191 | 0 | 0.030094763 |
| GO_SITE_OF_DOUBLE_STRAND_BREAK | 22 | 0.6229223 | 1.6472013 | 0.005836576 | 0.03012005 |
| GO_MITOTIC_SPINDLE_ORGANIZATION | 68 | 0.48606208 | 1.6473087 | 0 | 0.030127225 |
| GO_DNA_REPLICATION | 132 | 0.4369921 | 1.6462177 | 0 | 0.030322231 |
| GO_SUPRAMOLECULAR_FIBER_ORGANIZATION | 374 | 0.3949852 | 1.6462625 | 0 | 0.030356903 |
| GO_PROTON_TRANSMEMBRANE_TRANSPORT | 69 | -0.4808828 | -1.665026 | 0 | 0.030372726 |
| REACTOME_METABOLISM_OF_COFACTORS | 15 | -0.6853202 | -1.665265 | 0.009578544 | 0.0303882 |
| GO_CELL_DIVISION | 269 | 0.40670043 | 1.6453578 | 0 | 0.030398691 |
| PID_EPO_PATHWAY | 19 | 0.62937474 | 1.6455576 | 0.007604563 | 0.03043282 |
| REACTOME_REGULATION_OF_RUNX3_EXPRESSION_AND_ACTIVITY | 29 | 0.5673454 | 1.6453688 | 0.003891051 | 0.030445028 |
| REACTOME_POSTMITOTIC_NUCLEAR_PORE_COMPLEX_NPC_REFORMATION | 17 | 0.6468761 | 1.6449718 | 0.01183432 | 0.030463535 |
| REACTOME_NCAM1_INTERACTIONS | 27 | 0.5837777 | 1.6455748 | 0.005847953 | 0.03047545 |
| GO_NEGATIVE_REGULATION_OF_OSTEOCLAST_DIFFERENTIATION | 18 | 0.64731294 | 1.6438049 | 0.016563147 | 0.030749628 |
| GO_CYTOKINETIC_PROCESS | 19 | 0.62526304 | 1.6430713 | 0.007590133 | 0.03085016 |
| PID_PDGFRB_PATHWAY | 91 | 0.4574334 | 1.6431482 | 0.001996008 | 0.03086777 |
| GO_REGULATION_OF_DNA_DAMAGE_RESPONSE_SIGNAL_TRANSDUCTION_BY_P53_CLASS_MEDIATOR | 20 | 0.61647075 | 1.6432831 | 0.007561437 | 0.030867858 |
| REACTOME_ANTIGEN_ACTIVATES_B_CELL_RECEPTOR_BCR_LEADING_TO_GENERATION_OF_SECOND_MESSENGERS | 34 | 0.54438573 | 1.6424474 | 0.006097561 | 0.030956248 |
| REACTOME_RHO_GTPASES_ACTIVATE_FORMINS | 69 | 0.4855962 | 1.6418437 | 0 | 0.030970514 |
| GO_NEGATIVE_REGULATION_OF_RESPONSE_TO_CYTOKINE_STIMULUS | 38 | 0.5281351 | 1.6421542 | 0.008179959 | 0.030971384 |
| GO_REGULATION_OF_EXTRINSIC_APOPTOTIC_SIGNALING_PATHWAY_VIA_DEATH_DOMAIN_RECEPTORS | 32 | 0.56113386 | 1.6418937 | 0 | 0.031003723 |
| GO_REGULATION_OF_ISOTYPE_SWITCHING | 15 | 0.66412085 | 1.6414666 | 0.015748031 | 0.031027723 |
| GO_T_CELL_CYTOKINE_PRODUCTION | 30 | 0.5663076 | 1.6410623 | 0.013618677 | 0.03103059 |
| GO_REGULATION_OF_ALPHA_BETA_T_CELL_ACTIVATION | 58 | 0.5016621 | 1.6411732 | 0 | 0.031040734 |
| GO_MITOCHONDRIAL_SMALL_RIBOSOMAL_SUBUNIT | 15 | -0.6767206 | -1.662535 | 0.008179959 | 0.031058433 |
| GO_UBIQUITIN_LIKE_PROTEIN_CONJUGATING_ENZYME_ACTIVITY | 24 | 0.5925576 | 1.6402733 | 0.009652509 | 0.031219352 |
| GO_REGULATION_OF_OSTEOCLAST_DIFFERENTIATION | 35 | 0.5481051 | 1.640371 | 0.007920792 | 0.031229496 |
| PID_DELTA_NP63_PATHWAY | 28 | 0.5702191 | 1.6399176 | 0.004040404 | 0.031279728 |
| GO_AMINO_ACID_TRANSPORT | 79 | -0.4688059 | -1.661327 | 0 | 0.031322796 |
| PID_INTEGRIN_CS_PATHWAY | 18 | 0.64209443 | 1.6391772 | 0.011976048 | 0.03145955 |

| | | | | | |
|---|---|---|---|---|---|
| REACTOME_GLYCEROPHOSPHOLIPID_BIOSYNTHESIS | 64 | -0.4851106 | -1.660419 | 0 | 0.031478554 |
| GO_REGULATION_OF_PROTEIN_MATURATION | 41 | 0.52414757 | 1.6388953 | 0.003929273 | 0.031500343 |
| GO_LIPOPOLYSACCHARIDE_MEDIATED_SIGNALING_PATHWAY | 43 | 0.52341 | 1.6376456 | 0.003795066 | 0.03181987 |
| GO_GLYCOSAMINOGLYCAN_BINDING | 139 | 0.4352392 | 1.6377804 | 0 | 0.031832643 |
| PID_IL12_STAT4_PATHWAY | 24 | 0.5974241 | 1.6367272 | 0.015209125 | 0.032034032 |
| GO_ORGANOPHOSPHATE_BIOSYNTHETIC_PROCESS | 339 | -0.3947318 | -1.657597 | 0 | 0.032228097 |
| GO_VIRAL_GENE_EXPRESSION | 95 | 0.45232698 | 1.6359367 | 0 | 0.03225509 |
| GO_OXIDOREDUCTASE_ACTIVITY_ACTING_ON_THE_ALDEHYDE_OR_OXO_GROUP_OF_DONORS_NAD_OR_NADP_AS_ACCEPTOR | 27 | -0.5853839 | -1.656927 | 0.006355932 | 0.03231578 |
| GO_ENDOPLASMIC_RETICULUM_LUMEN | 198 | 0.4192515 | 1.6350224 | 0 | 0.032378357 |
| REACTOME_DAP12_SIGNALING | 24 | 0.59907246 | 1.6350676 | 0.013944224 | 0.032403316 |
| GO_PLASMA_MEMBRANE_PROTEIN_COMPLEX | 327 | 0.3962256 | 1.6351405 | 0 | 0.032424547 |
| GO_POSITIVE_REGULATION_OF_CHEMOTAXIS | 90 | 0.46642783 | 1.6345491 | 0.005434783 | 0.032471694 |
| GO_TAXIS | 364 | 0.38955513 | 1.6341765 | 0 | 0.032542385 |
| KEGG_NOD_LIKE_RECEPTOR_SIGNALING_PATHWAY | 39 | 0.54051137 | 1.6338311 | 0.00754717 | 0.03261451 |
| GO_VITAMIN_TRANSPORT | 23 | -0.6002029 | -1.654818 | 0.007984032 | 0.032886844 |
| GO_PHOSPHATE_ION_TRANSPORT | 17 | -0.6591628 | -1.65439 | 0.016494846 | 0.032908324 |
| PID_KIT_PATHWAY | 37 | 0.5393688 | 1.6319063 | 0 | 0.033135515 |
| GO_NEGATIVE_REGULATION_OF_CYTOKINE_SECRETION | 40 | 0.53594 | 1.6319208 | 0.002004008 | 0.033180203 |
| GO_RUFFLE | 109 | 0.44689873 | 1.6312058 | 0.001831502 | 0.03328109 |
| GO_POSITIVE_REGULATION_OF_INTERLEUKIN_10_PRODUCTION | 19 | 0.6141385 | 1.6310635 | 0.016064256 | 0.033286832 |
| GO_CARBOXY_LYASE_ACTIVITY | 26 | -0.5728464 | -1.652138 | 0.01443299 | 0.03353499 |
| GO_RESPONSE_TO_MOLECULE_OF_BACTERIAL_ORIGIN | 221 | 0.41362956 | 1.6298712 | 0 | 0.03363266 |
| GO_INTERLEUKIN_4_PRODUCTION | 21 | 0.60352343 | 1.628937 | 0.013358778 | 0.03392588 |
| GO_REGULATION_OF_ACUTE_INFLAMMATORY_RESPONSE | 27 | 0.575804 | 1.6285123 | 0.015625 | 0.034032244 |
| GO_CADHERIN_BINDING | 198 | 0.4118432 | 1.6282868 | 0 | 0.034042396 |
| GO_CHROMOSOMAL_REGION | 151 | 0.42594764 | 1.6271517 | 0 | 0.034180533 |
| GO_MITOCHONDRIAL_TRANSPORT | 135 | -0.4362725 | -1.650041 | 0 | 0.034187943 |
| PID_RAC1_PATHWAY | 35 | 0.54393953 | 1.6276546 | 0.013307985 | 0.03420806 |
| KEGG_P53_SIGNALING_PATHWAY | 46 | 0.5149562 | 1.6272374 | 0.005649718 | 0.034211285 |
| GO_RECEPTOR_TYROSINE_KINASE_BINDING | 42 | 0.5255388 | 1.6274996 | 0.003809524 | 0.034227036 |
| REACTOME_APC_C_CDH1_MEDIATED_DEGRADATION_OF_CDC20_AND_OTHER_APC_C_CDH1_TARGETED_PROTEINS_IN_LATE_MITOSIS_EARLY_G1 | 37 | 0.53527874 | 1.6273237 | 0.005836576 | 0.034233015 |
| GO_POSITIVE_REGULATION_OF_CELL_CYCLE_PHASE_TRANSITION | 53 | 0.5014605 | 1.6264329 | 0.005791506 | 0.03438263 |
| GO_SMALL_GTPASE_MEDIATED_SIGNAL_TRANSDUCTION | 299 | 0.39655417 | 1.6253704 | 0 | 0.03466542 |
| PID_BCR_5PATHWAY | 47 | 0.5132193 | 1.6247544 | 0 | 0.034734827 |
| GO_POLYOL_METABOLIC_PROCESS | 74 | -0.4785259 | -1.647974 | 0 | 0.034739148 |
| REACTOME_G2_M_DNA_DAMAGE_CHECKPOINT | 33 | 0.5539405 | 1.6249839 | 0.005535055 | 0.03474417 |
| GO_ALPHA_BETA_T_CELL_ACTIVATION | 89 | 0.45929885 | 1.6248474 | 0.001883239 | 0.03475513 |
| GO_APOPTOTIC_SIGNALING_PATHWAY | 364 | 0.3894582 | 1.623684 | 0 | 0.035035394 |
| REACTOME_RNA_POLYMERASE_II_TRANSCRIPTION_TERMINATION | 35 | 0.55482125 | 1.622764 | 0.005847953 | 0.035182204 |
| GO_PROTEIN_TYROSINE_KINASE_BINDING | 59 | 0.49371836 | 1.6228666 | 0.003968254 | 0.03520669 |
| REACTOME_TRANSCRIPTIONAL_REGULATION_BY_RUNX3 | 52 | 0.49751356 | 1.6229993 | 0.003891051 | 0.035207454 |
| GO_DOUBLE_STRANDED_RNA_BINDING | 45 | 0.51841855 | 1.6223491 | 0.003533569 | 0.035223845 |
| HALLMARK_APOPTOSIS | 123 | 0.43799427 | 1.6224346 | 0 | 0.035242934 |
| GO_PROTEIN_HOMOTETRAMERIZATION | 64 | -0.4809521 | -1.646007 | 0.002079002 | 0.035287995 |
| HALLMARK_UV_RESPONSE_DN | 103 | 0.44605187 | 1.6208917 | 0 | 0.03564079 |
| GO_EXTRINSIC_COMPONENT_OF_CYTOPLASMIC_SIDE_OF_PLASMA_MEMBRANE | 49 | 0.5057868 | 1.6207061 | 0.008179959 | 0.035657387 |

| | | | | | |
|---|---|---|---|---|---|
| GO_NEGATIVE_REGULATION_OF_INTRINSIC_APOPTOTIC_SIGNALING_PATHWAY_IN_RESPONSE_TO_DNA_DAMAGE | 19 | 0.62344754 | 1.6208954 | 0.013806706 | 0.035691056 |
| PID_NOTCH_PATHWAY | 29 | 0.5538943 | 1.620174 | 0.009596929 | 0.03575667 |
| GO_POSITIVE_REGULATION_OF_CD4_POSITIVE_ALPHA_BETA_T_CELL_ACTIVATION | 26 | 0.58169454 | 1.6202042 | 0.00591716 | 0.03580333 |
| GO_CHROMATIN_REMODELING_AT_CENTROMERE | 15 | 0.65095437 | 1.6196016 | 0.022088353 | 0.035896704 |
| GO_POSITIVE_REGULATION_OF_SUPRAMOLECULAR_FIBER_ORGANIZATION | 116 | 0.44106877 | 1.6171997 | 0 | 0.03650841 |
| KEGG_SMALL_CELL_LUNG_CANCER | 54 | 0.49714977 | 1.6172752 | 0.003868472 | 0.036532436 |
| GO_CYTOKINE_RECEPTOR_BINDING | 160 | 0.4224501 | 1.6174729 | 0 | 0.03656298 |
| GO_MACROPHAGE_CHEMOTAXIS | 24 | 0.5788178 | 1.6173296 | 0.009487666 | 0.036567237 |
| REACTOME_EPHRIN_SIGNALING | 15 | 0.66208947 | 1.6175774 | 0.011673152 | 0.03657459 |
| GO_EXTRINSIC_COMPONENT_OF_PLASMA_MEMBRANE | 88 | 0.4591833 | 1.6162782 | 0.001919386 | 0.036684252 |
| GO_POSITIVE_REGULATION_OF_LOCOMOTION | 353 | 0.3901379 | 1.6164501 | 0 | 0.036687165 |
| REACTOME_TOLL_LIKE_RECEPTOR_CASCADES | 89 | 0.46181047 | 1.6158267 | 0.002053388 | 0.03674768 |
| GO_SINGLE_STRANDED_RNA_BINDING | 44 | 0.5169246 | 1.6158758 | 0.005649718 | 0.03678087 |
| GO_THIOESTER_BIOSYNTHETIC_PROCESS | 37 | -0.5343512 | -1.641197 | 0.00631579 | 0.0368379 |
| GO_RECEPTOR_MEDIATED_ENDOCYTOSIS | 173 | 0.41384614 | 1.6148843 | 0 | 0.03693993 |
| GO_MICROTUBULE_CYTOSKELETON_ORGANIZATION_INVOLVED_IN_MITOSIS | 84 | 0.45888123 | 1.6150625 | 0 | 0.03697138 |
| GO_AXONAL_GROWTH_CONE | 17 | 0.63925946 | 1.6149253 | 0.013944224 | 0.036978506 |
| GO_MITOCHONDRIAL_RNA_METABOLIC_PROCESS | 26 | -0.5775701 | -1.640423 | 0.013645224 | 0.037016585 |
| GO_NEGATIVE_REGULATION_OF_ANTIGEN_RECEPTOR_MEDIATED_SIGNALING_PATHWAY | 21 | 0.6034495 | 1.6144099 | 0.019157087 | 0.037038855 |
| REACTOME_INTRINSIC_PATHWAY_FOR_APOPTOSIS | 30 | 0.5567535 | 1.6141622 | 0.005976096 | 0.037095573 |
| GO_CELL_VOLUME_HOMEOSTASIS | 18 | -0.6218422 | -1.639549 | 0.014522822 | 0.037237104 |
| GO_REGULATION_OF_T_CELL_MEDIATED_CYTOTOXICITY | 24 | 0.59760606 | 1.6134889 | 0.015065913 | 0.03729924 |
| GO_POSITIVE_REGULATION_OF_ACTIN_FILAMENT_POLYMERIZATION | 49 | 0.505505 | 1.6130904 | 0.005825243 | 0.03737961 |
| GO_NEGATIVE_REGULATION_OF_EXTRINSIC_APOPTOTIC_SIGNALING_PATHWAY_VIA_DEATH_DOMAIN_RECEPTORS | 22 | 0.6050856 | 1.6123259 | 0.015810277 | 0.03762152 |
| GO_NEGATIVE_REGULATION_OF_RESPONSE_TO_DNA_DAMAGE_STIMULUS | 37 | 0.5363682 | 1.6109275 | 0.007858546 | 0.038029104 |
| GO_VASCULAR_ENDOTHELIAL_GROWTH_FACTOR_RECEPTOR_SIGNALING_PATHWAY | 66 | 0.47912037 | 1.6107032 | 0 | 0.03805908 |
| GO_SODIUM_ION_TRANSPORT | 132 | -0.4362909 | -1.635797 | 0.002178649 | 0.038126606 |
| GO_RECOMBINATIONAL_REPAIR | 49 | 0.49860835 | 1.6103325 | 0.003883495 | 0.03815036 |
| GO_ORGANIC_HYDROXY_COMPOUND_BIOSYNTHETIC_PROCESS | 176 | -0.4203679 | -1.636821 | 0 | 0.038159043 |
| GO_TRANSFERASE_ACTIVITY_TRANSFERRING_SULFUR_CONTAINING_GROUPS | 38 | -0.5358778 | -1.636411 | 0.00845666 | 0.038185485 |
| GO_STEROID_BINDING | 69 | -0.4674282 | -1.635839 | 0.00203252 | 0.038236786 |
| REACTOME_G1_S_DNA_DAMAGE_CHECKPOINTS | 32 | 0.5482114 | 1.6097124 | 0.007751938 | 0.03832659 |
| GO_G_PROTEIN_ALPHA_SUBUNIT_BINDING | 17 | 0.63892394 | 1.6094191 | 0.014227643 | 0.038385812 |
| GO_RESPONSE_TO_TUMOR_NECROSIS_FACTOR | 184 | 0.41045377 | 1.6085163 | 0 | 0.0385454 |
| GO_POSITIVE_REGULATION_OF_RECEPTOR_MEDIATED_ENDOCYTOSIS | 34 | 0.5432347 | 1.6082209 | 0.012635379 | 0.03856226 |
| SIG_BCR_SIGNALING_PATHWAY | 32 | 0.5496022 | 1.608558 | 0.01192843 | 0.038578596 |
| GO_LIPID_HOMEOSTASIS | 85 | -0.4616696 | -1.634314 | 0.002283105 | 0.038593624 |
| GO_PRODUCTION_OF_MOLECULAR_MEDIATOR_INVOLVED_IN_INFLAMMATORY_RESPONSE | 50 | 0.50569135 | 1.6082592 | 0 | 0.03859363 |
| GO_BONE_REMODELING | 58 | 0.48695788 | 1.6085836 | 0 | 0.038615298 |
| KEGG_UBIQUITIN_MEDIATED_PROTEOLYSIS | 71 | 0.4659358 | 1.607626 | 0.001980198 | 0.038710427 |

| | | | | | |
|---|---|---|---|---|---|
| GO_ORGANOPHOSPHATE_CATABOLIC_PROCESS | 79 | -0.4700698 | -1.633433 | 0 | 0.038811613 |
| GO_POLYOL_BIOSYNTHETIC_PROCESS | 46 | -0.5100702 | -1.632708 | 0.004310345 | 0.03887939 |
| GO_INORGANIC_ANION_TRANSMEMBRANE_TRANSPORTER_ACTIVITY | 77 | -0.4689728 | -1.632299 | 0.002079002 | 0.03891414 |
| GO_POSITIVE_REGULATION_OF_LIPID_CATABOLIC_PROCESS | 16 | -0.6561223 | -1.632786 | 0.009920635 | 0.038965836 |
| GO_SPECIFIC_GRANULE_LUMEN | 46 | 0.5127445 | 1.6063665 | 0.003846154 | 0.0391333 |
| GO_FOCAL_ADHESION_ASSEMBLY | 55 | 0.4933232 | 1.606002 | 0.007532957 | 0.03916056 |
| GO_POSITIVE_REGULATION_OF_PEPTIDE_SECRETION | 173 | 0.41476566 | 1.6061049 | 0 | 0.039170254 |
| REACTOME_METABOLISM_OF_RNA | 323 | 0.3903906 | 1.605335 | 0 | 0.0392218 |
| GO_DEFENSE_RESPONSE_TO_GRAM_POSITIVE_BACTERIUM | 41 | 0.52031004 | 1.6056498 | 0.009784736 | 0.03925074 |
| GO_MITOTIC_SPINDLE_ASSEMBLY | 38 | 0.53248066 | 1.6053722 | 0.005952381 | 0.03926228 |
| GO_REGULATION_OF_SYNAPSE_STRUCTURE_OR_ACTIVITY | 123 | 0.43083322 | 1.6053865 | 0 | 0.039311424 |
| GO_SULFUR_COMPOUND_BIOSYNTHETIC_PROCESS | 124 | -0.437171 | -1.630015 | 0 | 0.0396165 |
| GO_STEROL_METABOLIC_PROCESS | 108 | -0.4503009 | -1.629135 | 0.002040816 | 0.0398128 |
| GO_EXOGENOUS_DRUG_CATABOLIC_PROCESS | 15 | -0.6577488 | -1.6284 | 0.017208412 | 0.04002033 |
| REACTOME_CONSTITUTIVE_SIGNALING_BY_AKT1_E17K_IN_CANCER | 16 | -0.6506354 | -1.627785 | 0.022 | 0.04011454 |
| KEGG_B_CELL_RECEPTOR_SIGNALING_PATHWAY | 53 | 0.4959875 | 1.6018319 | 0.005952381 | 0.040475037 |
| REACTOME_RRNA_PROCESSING | 90 | 0.4508374 | 1.6013 | 0 | 0.04054117 |
| PID_EPHRINB_REV_PATHWAY | 23 | 0.58407366 | 1.601409 | 0.014553014 | 0.04056609 |
| GO_NUCLEUS_LOCALIZATION | 15 | -0.6437534 | -1.626193 | 0.012658228 | 0.040613063 |
| REACTOME_MAPK6_MAPK4_SIGNALING | 48 | 0.50661784 | 1.6009412 | 0.003809524 | 0.040655967 |
| REACTOME_PEPTIDE_LIGAND_BINDING_RECEPTORS | 113 | 0.43261304 | 1.6006613 | 0 | 0.040714223 |
| HALLMARK_WNT_BETA_CATENIN_SIGNALING | 23 | 0.5914528 | 1.5989043 | 0.008421052 | 0.041263983 |
| GO_HISTONE_METHYLTRANSFERASE_COMPLEX | 34 | 0.54170066 | 1.5989963 | 0.006012024 | 0.04128303 |
| REACTOME_INFECTION_WITH_MYCOBACTERIUM_TUBERCULOSIS | 15 | 0.6441156 | 1.5982711 | 0.017578125 | 0.04141259 |
| REACTOME_TRANSCRIPTIONAL_REGULATION_BY_RUNX1 | 90 | 0.44938174 | 1.598329 | 0.002016129 | 0.04144518 |
| KEGG_FC_GAMMA_R_MEDIATED_PHAGOCYTOSIS | 65 | 0.47194463 | 1.5978522 | 0.001890359 | 0.041525576 |
| GO_ACID_SECRETION | 78 | -0.461407 | -1.62371 | 0 | 0.041613452 |
| GO_GLYCEROLIPID_METABOLIC_PROCESS | 220 | -0.4049437 | -1.622099 | 0 | 0.04210309 |
| REACTOME_HCMV_INFECTION | 56 | 0.4847025 | 1.5957859 | 0.002012072 | 0.042276405 |
| PID_SYNDECAN_4_PATHWAY | 27 | 0.56511563 | 1.5943755 | 0.016853932 | 0.042687546 |
| GO_REGULATION_OF_ACTIN_FILAMENT_BUNDLE_ASSEMBLY | 59 | 0.48892364 | 1.5944076 | 0.007766991 | 0.04273858 |
| GO_GOLGI_ASSOCIATED_VESICLE_MEMBRANE | 64 | 0.47445837 | 1.5939385 | 0.005607477 | 0.042765006 |
| KEGG_T_CELL_RECEPTOR_SIGNALING_PATHWAY | 67 | 0.47236532 | 1.5944214 | 0.005964215 | 0.04278804 |
| GO_ENDOCYTOSIS | 349 | 0.38563508 | 1.5939392 | 0 | 0.042820983 |
| GO_CYTOSOLIC_RIBOSOME | 37 | 0.52975917 | 1.5933164 | 0.008163265 | 0.04288883 |
| REACTOME_THE_ROLE_OF_GTSE1_IN_G2_M_PROGRESSION_AFTER_G2_CHECKPOINT | 47 | 0.50225216 | 1.5934625 | 0.005758158 | 0.0428912 |
| GO_EYE_PHOTORECEPTOR_CELL_DEVELOPMENT | 17 | -0.6335522 | -1.618824 | 0.019455252 | 0.043273095 |
| REACTOME_NUCLEAR_ENVELOPE_NE_REASSEMBLY | 50 | 0.49230915 | 1.5918959 | 0.007707129 | 0.043377 |
| GO_MYOBLAST_FUSION | 24 | 0.5881102 | 1.5916802 | 0.010080645 | 0.043405645 |
| REACTOME_CROSS_PRESENTATION_OF_SOLUBLE_EXOGENOUS_ANTIGENS_ENDOSOMES | 29 | 0.54638827 | 1.5919424 | 0.012145749 | 0.043413416 |
| GO_POSITIVE_REGULATION_OF_PHOSPHOLIPID_METABOLIC_PROCESS | 31 | 0.53890836 | 1.5909486 | 0.009433962 | 0.043638278 |
| GO_REGULATION_OF_T_CELL_CYTOKINE_PRODUCTION | 26 | 0.5686777 | 1.5906981 | 0.021868788 | 0.043689422 |
| GO_NEGATIVE_REGULATION_OF_INTERLEUKIN_6_PRODUCTION | 22 | 0.58985466 | 1.589986 | 0.007677543 | 0.043869976 |
| GO_PHAGOCYTIC_CUP | 18 | 0.614565 | 1.5897448 | 0.016949153 | 0.043897834 |
| KEGG_JAK_STAT_SIGNALING_PATHWAY | 83 | 0.44736877 | 1.589308 | 0.001960784 | 0.044021126 |
| GO_MYOSIN_BINDING | 44 | 0.5143315 | 1.5889033 | 0.0056926 | 0.04411619 |

| | | | | | |
|---|---|---|---|---|---|
| GO_KILLING_OF_CELLS_OF_OTHER_ORGANISM | 26 | 0.56176335 | 1.5886132 | 0.02008032 | 0.044184487 |
| GO_PLATELET_DEGRANULATION | 97 | 0.43916255 | 1.5878943 | 0 | 0.044232555 |
| GO_ATP_DEPENDENT_CHROMATIN_REMODELING | 34 | 0.52445924 | 1.5880272 | 0.012219959 | 0.044233542 |
| GO_DNA_HELICASE_ACTIVITY | 37 | 0.52870786 | 1.5883119 | 0.008097166 | 0.044257764 |
| GO_POSITIVE_REGULATION_OF_RECEPTOR_SIGNALING_PATHWAY_VIA_STAT | 55 | 0.4841426 | 1.588041 | 0.003921569 | 0.04428694 |
| GO_TERTIARY_GRANULE_LUMEN | 43 | 0.5096086 | 1.5880815 | 0.017964073 | 0.044322416 |
| PID_P53_DOWNSTREAM_PATHWAY | 92 | 0.44819924 | 1.5873886 | 0.001930502 | 0.044420008 |
| GO_SOLUTE_CATION_SYMPORTER_ACTIVITY | 64 | -0.4807924 | -1.61509 | 0 | 0.04442331 |
| GO_RESPONSE_TO_CAMP | 69 | -0.4713409 | -1.615731 | 0.002141328 | 0.04444921 |
| GO_MONOCARBOXYLIC_ACID_TRANSMEMBRANE_TRANSPORTER_ACTIVITY | 28 | -0.5721867 | -1.615274 | 0.014767933 | 0.044482753 |
| GO_LONG_TERM_MEMORY | 20 | -0.6026228 | -1.614488 | 0.014553014 | 0.04456126 |
| GO_MONOVALENT_INORGANIC_CATION_TRANSPORT | 271 | -0.396786 | -1.613915 | 0 | 0.044660646 |
| GO_ORGANIC_ACID_SODIUM_SYMPORTER_ACTIVITY | 20 | -0.6128277 | -1.612577 | 0.023454158 | 0.045031905 |
| BIOCARTA_DEATH_PATHWAY | 23 | 0.5821906 | 1.5847611 | 0.015473888 | 0.045421906 |
| GO_INORGANIC_ANION_TRANSPORT | 94 | -0.4541389 | -1.611048 | 0.002053388 | 0.045556843 |
| GO_ACTIN_FILAMENT_BUNDLE_ORGANIZATION | 95 | 0.44579247 | 1.5839455 | 0.001824818 | 0.04573932 |
| REACTOME_RUNX1_REGULATES_TRANSCRIPTION_OF_GENES_INVOLVED_IN_DIFFERENTIATION_OF_HSCS | 39 | 0.513748 | 1.5835465 | 0.019267824 | 0.04585561 |
| GO_POSITIVE_REGULATION_OF_CELLULAR_COMPONENT_BIOGENESIS | 288 | 0.38656375 | 1.5832547 | 0 | 0.045911465 |
| REACTOME_DEGRADATION_OF_GLI1_BY_THE_PROTEASOME | 31 | 0.53411037 | 1.5828143 | 0.011881189 | 0.046014223 |
| GO_ANTIBIOTIC_CATABOLIC_PROCESS | 38 | -0.5258972 | -1.609479 | 0.008016032 | 0.046067785 |
| GO_POSITIVE_REGULATION_OF_MYELOID_LEUKOCYTE_CYTOKINE_PRODUCTION_INVOLVED_IN_IMMUNE_RESPONSE | 18 | 0.62016284 | 1.581007 | 0.029469548 | 0.046672847 |
| REACTOME_SIGNALING_BY_RETINOIC_ACID | 27 | -0.5608186 | -1.60696 | 0.004056795 | 0.046966612 |
| GO_OXIDOREDUCTASE_ACTIVITY_ACTING_ON_SINGLE_DONORS_WITH_INCORPORATION_OF_MOLECULAR_OXYGEN | 18 | -0.6196461 | -1.606644 | 0.016260162 | 0.046970587 |
| GO_POSITIVE_REGULATION_OF_NOTCH_SIGNALING_PATHWAY | 23 | 0.5775052 | 1.5797 | 0.025641026 | 0.047179762 |
| GO_REGULATION_OF_INTERLEUKIN_1_SECRETION | 28 | 0.5564492 | 1.5793158 | 0.013435701 | 0.04729674 |
| GO_ENDOPEPTIDASE_REGULATOR_ACTIVITY | 109 | 0.43524975 | 1.5779954 | 0 | 0.047795456 |
| GO_POSITIVE_REGULATION_OF_PEPTIDASE_ACTIVITY | 113 | 0.42595038 | 1.5771855 | 0.001897533 | 0.048042227 |
| GO_STRUCTURAL_MOLECULE_ACTIVITY | 366 | 0.37739527 | 1.5765615 | 0 | 0.04819855 |
| GO_T_CELL_SELECTION | 29 | 0.5495833 | 1.5766119 | 0.011976048 | 0.048238408 |
| GO_HYDROLASE_ACTIVITY_ACTING_ON_CARBON_NITROGEN_BUT_NOT_PEPTIDE_BONDS | 71 | -0.4594715 | -1.600794 | 0.001964637 | 0.048325177 |
| GO_KETONE_BIOSYNTHETIC_PROCESS | 30 | -0.5482359 | -1.600357 | 0.018556701 | 0.048403505 |
| GO_LONG_CHAIN_FATTY_ACID_METABOLIC_PROCESS | 68 | -0.4692149 | -1.600928 | 0.002087683 | 0.048410125 |
| GO_FATTY_ACID_BINDING | 26 | -0.5606852 | -1.601238 | 0.014227643 | 0.048416894 |
| GO_VACUOLAR_LUMEN | 123 | 0.4253197 | 1.5758046 | 0 | 0.048434354 |
| GO_LIPID_BIOSYNTHETIC_PROCESS | 422 | -0.3783905 | -1.601321 | 0 | 0.048527475 |
| REACTOME_GLUCOSE_METABOLISM | 62 | -0.4812385 | -1.602443 | 0.004140787 | 0.04862924 |
| GO_4_IRON_4_SULFUR_CLUSTER_BINDING | 30 | -0.5577638 | -1.60134 | 0.02 | 0.048657354 |
| GO_MYELOID_LEUKOCYTE_CYTOKINE_PRODUCTION | 25 | 0.55538255 | 1.5751016 | 0.022774328 | 0.048661362 |
| GO_CALCIUM_ION_IMPORT | 49 | -0.5012454 | -1.601504 | 0.009881423 | 0.048727397 |
| GO_PROTON_TRANSMEMBRANE_TRANSPORTER_ACTIVITY | 58 | -0.4856287 | -1.601793 | 0.007722008 | 0.048751846 |
| GO_CHROMOSOME_SEPARATION | 35 | 0.5292608 | 1.5737274 | 0.005747126 | 0.049041998 |
| GO_REGULATION_OF_MRNA_METABOLIC_PROCESS | 160 | 0.41380635 | 1.57375 | 0 | 0.049096994 |
| GO_GLIAL_CELL_PROLIFERATION | 31 | 0.5398731 | 1.5737576 | 0.013645224 | 0.04915532 |
| GO_CELL_CYCLE_CHECKPOINT | 96 | 0.4402221 | 1.5730414 | 0 | 0.049186576 |
| GO_PYRUVATE_METABOLIC_PROCESS | 104 | -0.4366395 | -1.598208 | 0.004175365 | 0.04921322 |

| | | | | | |
|---|---|---|---|---|---|
| GO_REGULATION_OF_BONE_REMODELING | 29 | 0.54796416 | 1.5730696 | 0.020754717 | 0.049240023 |
| REACTOME_CLASS_I_MHC_MEDIATED_ANTIGEN_PROCESSING_PRESENTATION | 192 | 0.40140653 | 1.5725089 | 0 | 0.049387697 |
| GO_REGULATION_OF_NIK_NF_KAPPAB_SIGNALING | 67 | 0.46347833 | 1.572158 | 0.005535055 | 0.049477883 |
| REACTOME_STABILIZATION_OF_P53 | 27 | 0.55498713 | 1.571851 | 0.007662835 | 0.049545493 |
| GO_APICAL_PLASMA_MEMBRANE | 207 | -0.3968272 | -1.597122 | 0 | 0.049593575 |
| HALLMARK_MYC_TARGETS_V1 | 124 | 0.42452788 | 1.5712198 | 0 | 0.04976393 |
| GO_EXTRACELLULAR_MATRIX_DISASSEMBLY | 49 | 0.4896171 | 1.5701091 | 0.007920792 | 0.049984846 |

**Table S2. CKD-Olive enriched geneset**

| *NAME* | *SIZE* | *ES* | *NES* | *NOM p-val* | *FDR q-val* |
|---|---|---|---|---|---|
| GO_MITOCHONDRIAL_GENE_EXPRESSION | 85 | 0.65229046 | 2.267848 | 0 | 0 |
| HALLMARK_TNFA_SIGNALING_VIA_NFKB | 175 | -0.71899724 | -3.1513178 | 0 | 0 |
| REACTOME_NGF_STIMULATED_TRANSCRIPTION | 27 | -0.8094353 | -2.52837 | 0 | 0 |
| REACTOME_CLASS_A_1_RHODOPSIN_LIKE_RECEPTORS | 168 | -0.5631937 | -2.502836 | 0 | 0 |
| REACTOME_NUCLEAR_EVENTS_KINASE_AND_TRANSCRIPTION_FACTOR_ACTIVATION | 35 | -0.75876415 | -2.4958098 | 0 | 0 |
| HALLMARK_KRAS_SIGNALING_DN | 136 | -0.58310866 | -2.4956658 | 0 | 0 |
| REACTOME_GPCR_LIGAND_BINDING | 225 | -0.55271363 | -2.492893 | 0 | 0 |
| GO_GLOMERULUS_DEVELOPMENT | 47 | -0.6737668 | -2.3901112 | 0 | 0 |
| GO_G_PROTEIN_COUPLED_RECEPTOR_ACTIVITY | 202 | -0.5309663 | -2.3873925 | 0 | 0 |
| GO_TRNA_METABOLIC_PROCESS | 73 | 0.6391042 | 2.2022629 | 0 | 2.29E-04 |
| GO_MITOCHONDRIAL_MATRIX | 283 | 0.5427342 | 2.1672316 | 0 | 2.70E-04 |
| GO_NCRNA_METABOLIC_PROCESS | 191 | 0.5626319 | 2.1718395 | 0 | 3.38E-04 |
| GO_PEROXISOME_ORGANIZATION | 57 | 0.65827864 | 2.1625917 | 0 | 3.38E-04 |
| GO_MITOCHONDRIAL_TRANSLATION | 71 | 0.65156645 | 2.2221332 | 0 | 3.43E-04 |
| REACTOME_MITOTIC_SPINDLE_CHECKPOINT | 49 | 0.67890716 | 2.1527152 | 0 | 3.88E-04 |
| REACTOME_PEROXISOMAL_PROTEIN_IMPORT | 45 | 0.67779905 | 2.1389458 | 0 | 4.24E-04 |
| HALLMARK_PROTEIN_SECRETION | 72 | 0.6175955 | 2.1079254 | 0 | 5.52E-04 |
| GO_PEROXISOMAL_TRANSPORT | 50 | 0.6482501 | 2.1065464 | 0 | 5.63E-04 |
| GO_NEGATIVE_REGULATION_OF_MAP_KINASE_ACTIVITY | 49 | -0.6412498 | -2.3081656 | 0 | 5.73E-04 |
| GO_NCRNA_PROCESSING | 159 | 0.56081283 | 2.1202424 | 0 | 6.01E-04 |
| REACTOME_DNA_REPAIR | 147 | 0.56028867 | 2.1131804 | 0 | 6.07E-04 |
| NABA_SECRETED_FACTORS | 166 | -0.5275348 | -2.2887585 | 0 | 6.13E-04 |
| GO_P38MAPK_CASCADE | 27 | -0.7268468 | -2.2751486 | 0 | 6.46E-04 |
| REACTOME_PEPTIDE_LIGAND_BINDING_RECEPTORS | 113 | -0.5453449 | -2.2899451 | 0 | 6.60E-04 |
| GO_INACTIVATION_OF_MAPK_ACTIVITY | 18 | -0.81544054 | -2.2856953 | 0 | 6.89E-04 |
| GO_NEUROPEPTIDE_SIGNALING_PATHWAY | 39 | -0.6750704 | -2.2696898 | 0 | 7.11E-04 |
| GO_REGULATION_OF_EPITHELIAL_CELL_DIFFERENTIATION | 94 | -0.55625445 | -2.290903 | 0 | 7.15E-04 |
| GO_KINETOCHORE | 63 | 0.61986965 | 2.0851672 | 0 | 7.65E-04 |
| GO_CELLULAR_AMINO_ACID_METABOLIC_PROCESS | 203 | 0.53815955 | 2.0869596 | 0 | 7.72E-04 |
| GO_PROTEIN_LOCALIZATION_TO_CHROMOSOME | 32 | 0.6989692 | 2.0881674 | 0 | 7.80E-04 |
| KEGG_NEUROACTIVE_LIGAND_RECEPTOR_INTERACTION | 138 | -0.5471579 | -2.295804 | 0 | 7.80E-04 |
| REACTOME_HIV_INFECTION | 133 | 0.56130385 | 2.0773504 | 0 | 8.02E-04 |
| PID_ATF2_PATHWAY | 35 | -0.6962275 | -2.3008952 | 0 | 8.58E-04 |
| GO_SKELETAL_MUSCLE_CELL_DIFFERENTIATION | 27 | -0.72919786 | -2.258769 | 0 | 8.59E-04 |
| PID_FRA_PATHWAY | 27 | -0.7179398 | -2.2364116 | 0 | 9.99E-04 |
| GO_ANTIMICROBIAL_HUMORAL_IMMUNE_RESPONSE_MEDIATED_BY_ANTIMICROBIAL_PEPTIDE | 36 | -0.6748693 | -2.2327883 | 0 | 0.00103673 |
| REACTOME_MITOTIC_METAPHASE_AND_ANAPHASE | 122 | 0.5547193 | 2.0601268 | 0 | 0.001152018 |
| GO_MITOCHONDRIAL_PROTEIN_COMPLEX | 120 | 0.5688479 | 2.0548766 | 0 | 0.001163366 |
| GO_CALCIUM_CHANNEL_REGULATOR_ACTIVITY | 24 | -0.73085445 | -2.218277 | 0 | 0.001253356 |
| REACTOME_FORMATION_OF_THE_CORNIFIED_ENVELOPE | 49 | -0.6028737 | -2.2232037 | 0 | 0.001313039 |
| GO_KERATINIZATION | 54 | -0.6019733 | -2.2067044 | 0 | 0.001320545 |
| GO_MONONUCLEAR_CELL_MIGRATION | 54 | -0.5998211 | -2.2162757 | 0 | 0.001343985 |
| REACTOME_TRANSLATION | 134 | 0.54412204 | 2.0365582 | 0 | 0.001353025 |
| REACTOME_ANTIGEN_PROCESSING_UBIQUITINATION_PROTEASOME_DEGRADATION | 140 | 0.54684204 | 2.0445826 | 0 | 0.001353196 |
| GO_RIBONUCLEOPROTEIN_COMPLEX_BIOGENESIS | 198 | 0.5258416 | 2.0418377 | 0 | 0.001353417 |

| | | | | | |
|---|---|---|---|---|---|
| KEGG_LEISHMANIA_INFECTION | 42 | -0.6317497 | -2.2087188 | 0 | 0.001373367 |
| REACTOME_TRANSCRIPTIONAL_REGULATION_BY_SMALL_RNAS | 30 | 0.70608044 | 2.0343776 | 0 | 0.00138173 |
| GO_RNA_PROCESSING | 426 | 0.4941372 | 2.037449 | 0 | 0.001383536 |
| GO_LIGASE_ACTIVITY | 70 | 0.60498637 | 2.0450664 | 0 | 0.001424417 |
| NABA_CORE_MATRISOME | 145 | -0.518924 | -2.2105315 | 0 | 0.00143059 |
| HALLMARK_HYPOXIA | 153 | -0.5001534 | -2.194429 | 0 | 0.001462697 |
| GO_TRANSCRIPTION_COUPLED_NUCLEOTIDE_EXCISION_REPAIR | 40 | 0.65247834 | 1.9945294 | 0 | 0.001469452 |
| REACTOME_TRNA_PROCESSING | 56 | 0.6147914 | 1.9954349 | 0 | 0.001472361 |
| GO_MICROBODY | 84 | 0.57162404 | 1.9900677 | 0 | 0.001479546 |
| GO_NCRNA_EXPORT_FROM_NUCLEUS | 25 | 0.7144898 | 2.0034745 | 0 | 0.001509109 |
| GO_RNA_POLYMERASE_II_HOLOENZYME | 47 | 0.6297023 | 1.9956646 | 0 | 0.001510114 |
| REACTOME_GENE_SILENCING_BY_RNA | 41 | 0.645742 | 2.0120876 | 0 | 0.001512377 |
| REACTOME_VIRAL_MESSENGER_RNA_SYNTHESIS | 27 | 0.71238726 | 1.9909594 | 0 | 0.001514773 |
| KEGG_VALINE_LEUCINE_AND_ISOLEUCINE_DEGRADATION | 38 | 0.6455556 | 1.9884077 | 0 | 0.001522666 |
| GO_NUCLEOID | 25 | 0.72565264 | 2.019589 | 0 | 0.001529308 |
| GO_GENE_SILENCING | 110 | 0.55500287 | 1.9972806 | 0 | 0.001532498 |
| REACTOME_CELL_CYCLE_CHECKPOINTS | 126 | 0.5393703 | 2.0133343 | 0 | 0.001540932 |
| REACTOME_MITOCHONDRIAL_TRANSLATION | 47 | 0.6264513 | 2.0004754 | 0 | 0.001542477 |
| GO_TRANSLATIONAL_TERMINATION | 55 | 0.6024557 | 2.0036683 | 0 | 0.001553495 |
| REACTOME_POSTMITOTIC_NUCLEAR_PORE_COMPLEX_NPC_REFORMATION | 17 | 0.7930977 | 2.020829 | 0 | 0.001561957 |
| REACTOME_HOST_INTERACTIONS_OF_HIV_FACTORS | 78 | 0.58720654 | 2.0250251 | 0 | 0.001570087 |
| GO_MITOCHONDRIAL_RNA_METABOLIC_PROCESS | 26 | 0.7033798 | 1.997946 | 0 | 0.001573917 |
| GO_RIBONUCLEOPROTEIN_COMPLEX | 326 | 0.4931239 | 1.98569 | 0 | 0.001578707 |
| GO_INTERSTRAND_CROSS_LINK_REPAIR | 19 | 0.77160656 | 1.982238 | 0 | 0.001578916 |
| REACTOME_METABOLISM_OF_RNA | 323 | 0.4955354 | 2.005309 | 0 | 0.001587094 |
| GO_HORMONE_ACTIVITY | 59 | -0.5874385 | -2.1847432 | 0 | 0.001593894 |
| GO_RIBOSOME_BIOGENESIS | 136 | 0.538512 | 2.0148637 | 0 | 0.001595966 |
| REACTOME_TRNA_PROCESSING_IN_THE_NUCLEUS | 33 | 0.67277104 | 2.004072 | 0 | 0.001600571 |
| PID_IL6_7_PATHWAY | 35 | -0.6649413 | -2.1750374 | 0 | 0.001600724 |
| KEGG_PEROXISOME | 60 | 0.60370445 | 1.9826615 | 0 | 0.001603552 |
| GO_EPIDERMIS_DEVELOPMENT | 184 | -0.48912862 | -2.1694512 | 0 | 0.001603976 |
| REACTOME_FORMATION_OF_TC_NER_PRE_INCISION_COMPLEX | 25 | 0.6968985 | 1.9823202 | 0 | 0.00161251 |
| GO_CATALYTIC_ACTIVITY_ACTING_ON_RNA | 157 | 0.5326801 | 2.006467 | 0 | 0.00163829 |
| REACTOME_KERATINIZATION | 50 | -0.60733 | -2.176042 | 0 | 0.001655921 |
| GO_MITOCHONDRIAL_TRANSLATIONAL_TERMINATION | 45 | 0.6329663 | 1.9791116 | 0 | 0.001671856 |
| REACTOME_FANCONI_ANEMIA_PATHWAY | 15 | 0.79838437 | 1.9785333 | 0 | 0.001679177 |
| GO_CHROMOSOMAL_REGION | 151 | 0.5219997 | 1.9752727 | 0 | 0.001765868 |
| REACTOME_SEPARATION_OF_SISTER_CHROMATIDS | 93 | 0.55402917 | 1.9738085 | 0 | 0.001784061 |
| REACTOME_PROTEIN_UBIQUITINATION | 39 | 0.63619405 | 1.9672147 | 0 | 0.001900767 |
| REACTOME_SWITCHING_OF_ORIGINS_TO_A_POST_REPLICATIVE_STATE | 45 | 0.61961466 | 1.9674287 | 0 | 0.00193471 |
| REACTOME_E3_UBIQUITIN_LIGASES_UBIQUITINATE_TARGET_PROTEINS | 26 | 0.6946658 | 1.9681125 | 0 | 0.001957463 |
| REACTOME_ANTIVIRAL_MECHANISM_BY_IFN_STIMULATED_GENES | 54 | 0.59909695 | 1.9631771 | 0 | 0.001961309 |
| GO_TRNA_PROCESSING | 53 | 0.60660374 | 1.9681923 | 0 | 0.001981129 |
| GO_HOST_CELLULAR_COMPONENT | 42 | 0.63496965 | 1.9612634 | 0 | 0.001985464 |

| | | | | | |
|---|---|---|---|---|---|
| GO_NUCLEOTIDE_EXCISION_REPAIR_PREINCISION_COMPLEX_ASSEMBLY | 19 | 0.7393532 | 1.9608723 | 0 | 0.001986007 |
| GO_MRNA_PROCESSING | 242 | 0.5020404 | 1.9684877 | 0 | 0.002018509 |
| REACTOME_PROTEIN_LOCALIZATION | 97 | 0.5479566 | 1.9544846 | 0 | 0.002117348 |
| GO_MICROBODY_LUMEN | 34 | 0.6603143 | 1.9553509 | 0 | 0.002140522 |
| REACTOME_NUCLEOTIDE_EXCISION_REPAIR | 59 | 0.5917051 | 1.9559382 | 0 | 0.002142406 |
| GO_UBIQUITIN_LIGASE_COMPLEX | 106 | 0.5397005 | 1.9513243 | 0 | 0.00221148 |
| REACTOME_SNRNP_ASSEMBLY | 28 | 0.6746407 | 1.9444945 | 0 | 0.002321016 |
| KEGG_RNA_DEGRADATION | 24 | 0.71847457 | 1.9457409 | 0 | 0.002339793 |
| GO_DNA_REPAIR | 227 | 0.4969211 | 1.9447259 | 0 | 0.002355658 |
| REACTOME_INTERACTIONS_OF_REV_WITH_HOST_CELLULAR_PROTEINS | 23 | 0.70665085 | 1.9463167 | 0.001574803 | 0.002365233 |
| GO_RNA_SPLICING_VIA_TRANSESTERIFICATION_REACTIONS | 168 | 0.50371987 | 1.9428974 | 0 | 0.002385419 |
| REACTOME_TRANSPORT_OF_THE_SLBP_DEPENDANT_MATURE_MRNA | 23 | 0.7010189 | 1.9414167 | 0 | 0.002429132 |
| GO_CORNIFICATION | 50 | -0.6008756 | -2.1379766 | 0 | 0.002491423 |
| GO_NEUROTRANSMITTER_RECEPTOR_ACTIVITY | 44 | -0.6247998 | -2.1426077 | 0 | 0.002515487 |
| PID_AP1_PATHWAY | 48 | -0.59911454 | -2.1406705 | 0 | 0.002541119 |
| GO_NEGATIVE_REGULATION_OF_PROTEIN_SERINE_THREONINE_KINASE_ACTIVITY | 82 | -0.53761894 | -2.1385374 | 0 | 0.002564701 |
| REACTOME_NUCLEAR_IMPORT_OF_REV_PROTEIN | 22 | 0.69992787 | 1.9379562 | 0.001557632 | 0.002623475 |
| GO_RRNA_METABOLIC_PROCESS | 100 | 0.54298073 | 1.9368082 | 0 | 0.002653063 |
| GO_LIPID_TRANSFER_ACTIVITY | 24 | 0.69630635 | 1.9362935 | 0 | 0.00267276 |
| GO_EXTRACELLULAR_MATRIX_STRUCTURAL_CONSTITUENT | 103 | -0.50942755 | -2.124916 | 0 | 0.002802216 |
| GO_ALPHA_BETA_T_CELL_DIFFERENTIATION | 65 | -0.5577301 | -2.1212935 | 0 | 0.002878034 |
| REACTOME_DNA_REPLICATION | 70 | 0.5690401 | 1.9317497 | 0 | 0.002908418 |
| GO_DNA_TEMPLATED_TRANSCRIPTION_TERMINATION | 35 | 0.6395991 | 1.9324008 | 0 | 0.002929387 |
| GO_ANTIBACTERIAL_HUMORAL_RESPONSE | 21 | -0.7136905 | -2.1214395 | 0 | 0.002955819 |
| KEGG_NUCLEOTIDE_EXCISION_REPAIR | 33 | 0.6450871 | 1.9274141 | 0 | 0.00300889 |
| GO_MULTI_ORGANISM_LOCALIZATION | 36 | 0.6340833 | 1.92785 | 0 | 0.0030395 |
| GO_RECEPTOR_REGULATOR_ACTIVITY | 244 | -0.46133462 | -2.1085029 | 0 | 0.003048891 |
| HALLMARK_OXIDATIVE_PHOSPHORYLATION | 143 | 0.5143699 | 1.9255736 | 0 | 0.003053175 |
| REACTOME_HIV_LIFE_CYCLE | 80 | 0.56144565 | 1.9260042 | 0 | 0.003074793 |
| REACTOME_HIV_TRANSCRIPTION_INITIATION | 31 | 0.6553008 | 1.9247303 | 0 | 0.003082579 |
| GO_CYTOKINE_ACTIVITY | 109 | -0.5078009 | -2.10911 | 0 | 0.003125113 |
| REACTOME_APC_C_MEDIATED_DEGRADATION_OF_CELL_CYCLE_PROTEINS | 44 | 0.6116639 | 1.921756 | 0 | 0.003139642 |
| GO_PROTEIN_DNA_COMPLEX_SUBUNIT_ORGANIZATION | 114 | 0.52617705 | 1.9224122 | 0 | 0.003144792 |
| GO_OLFACTORY_RECEPTOR_ACTIVITY | 17 | -0.76283556 | -2.1117249 | 0 | 0.003159044 |
| REACTOME_CLASS_I_MHC_MEDIATED_ANTIGEN_PROCESSING_PRESENTATION | 192 | 0.5007449 | 1.9204808 | 0 | 0.003199749 |
| REACTOME_EXPORT_OF_VIRAL_RIBONUCLEOPROTEINS_FROM_NUCLEUS | 22 | 0.70852494 | 1.9184166 | 0 | 0.003218191 |
| REACTOME_INTERACTIONS_OF_VPR_WITH_HOST_CELLULAR_PROTEINS | 24 | 0.6919042 | 1.9179026 | 0 | 0.003220388 |
| GO_NUCLEAR_CHROMOSOME_TELOMERIC_REGION | 53 | 0.59131885 | 1.9148966 | 0 | 0.003332835 |
| GO_G_PROTEIN_COUPLED_RECEPTOR_SIGNALING_PATHWAY | 481 | -0.42407453 | -2.0981855 | 0 | 0.003338453 |
| GO_TRANSLATION_REGULATOR_ACTIVITY_NUCLEIC_ACID_BINDING | 45 | 0.61152196 | 1.9152724 | 0 | 0.003363729 |

| | | | | | |
|---|---|---|---|---|---|
| GO_OXIDOREDUCTASE_ACTIVITY_ACTING_ON_THE_CH_CH_GROUP_OF_DONORS | 40 | 0.6199597 | 1.9135305 | 0 | 0.003418692 |
| REACTOME_MRNA_CAPPING | 17 | 0.7488566 | 1.9115434 | 0.001633987 | 0.003448203 |
| GO_TRNA_MODIFICATION | 34 | 0.63289106 | 1.9120675 | 0.001547988 | 0.003471688 |
| GO_RNA_CAPPING | 17 | 0.7488566 | 1.9086165 | 0 | 0.003559204 |
| GO_NEGATIVE_REGULATION_OF_CELL_CYCLE_G2_M_PHASE_TRANSITION | 48 | 0.5923575 | 1.9044408 | 0 | 0.003599946 |
| REACTOME_TRANSCRIPTION_COUPLED_NUCLEOTIDE_EXCISION_REPAIR_TC_NER | 43 | 0.6171787 | 1.9037976 | 0 | 0.003611592 |
| GO_TRANSLATION_INITIATION_FACTOR_ACTIVITY | 28 | 0.6705602 | 1.9051875 | 0 | 0.003616337 |
| GO_PROTEIN_POLYUBIQUITINATION | 146 | 0.5113581 | 1.9064765 | 0 | 0.003621277 |
| GO_ORGANELLAR_RIBOSOME | 45 | 0.6067157 | 1.9055039 | 0 | 0.003640305 |
| GO_FEMALE_PREGNANCY | 121 | -0.49991155 | -2.0910814 | 0 | 0.003654935 |
| REACTOME_FORMATION_OF_INCISION_COMPLEX_IN_GG_NER | 27 | 0.6773167 | 1.9067044 | 0 | 0.003660638 |
| GO_COENZYME_METABOLIC_PROCESS | 160 | 0.50130635 | 1.9021559 | 0 | 0.003675475 |
| REACTOME_M_PHASE | 181 | 0.49613526 | 1.902552 | 0 | 0.003692297 |
| GO_SENSORY_PERCEPTION_OF_CHEMICAL_STIMULUS | 56 | -0.5763994 | -2.0937872 | 0 | 0.003700363 |
| GO_RESPONSE_TO_CHEMOKINE | 70 | -0.5464491 | -2.0832741 | 0 | 0.00371117 |
| GO_POSTSYNAPTIC_NEUROTRANSMITTER_RECEPTOR_ACTIVITY | 32 | -0.62846524 | -2.0885143 | 0 | 0.003726528 |
| KEGG_MAPK_SIGNALING_PATHWAY | 166 | -0.47252116 | -2.0838385 | 0 | 0.003791847 |
| REACTOME_SIGNALING_BY_GPCR | 426 | -0.43265623 | -2.0788693 | 0 | 0.003955875 |
| HALLMARK_MYC_TARGETS_V1 | 124 | 0.5117034 | 1.897323 | 0 | 0.003957928 |
| GO_PHOSPHOTRANSFERASE_ACTIVITY_PHOSPHATE_GROUP_AS_ACCEPTOR | 24 | 0.68445957 | 1.8956003 | 0.001594896 | 0.003979804 |
| NABA_MATRISOME_ASSOCIATED | 379 | -0.43142262 | -2.0771508 | 0 | 0.003981365 |
| GO_COFACTOR_BIOSYNTHETIC_PROCESS | 141 | 0.507877 | 1.8961748 | 0 | 0.003992469 |
| GO_POSITIVE_REGULATION_OF_OSTEOBLAST_DIFFERENTIATION | 37 | -0.6203968 | -2.0730062 | 0 | 0.004039671 |
| GO_GENE_SILENCING_BY_RNA | 79 | 0.54217494 | 1.893584 | 0 | 0.00410545 |
| GO_ANAPHASE_PROMOTING_COMPLEX_DEPENDENT_CATABOLIC_PROCESS | 42 | 0.60952127 | 1.8924488 | 0 | 0.004157477 |
| GO_MONOCYTE_CHEMOTAXIS | 38 | -0.61249584 | -2.0661464 | 0 | 0.004185712 |
| REACTOME_SIGNALING_BY_NTRKS | 84 | -0.5279017 | -2.0712368 | 0 | 0.004196858 |
| GO_TRANSMITTER_GATED_CHANNEL_ACTIVITY | 25 | -0.669004 | -2.060613 | 0 | 0.004225343 |
| GO_POSITIVE_REGULATION_OF_OSSIFICATION | 52 | -0.5778159 | -2.0681672 | 0 | 0.0042332 |
| REACTOME_RAF_INDEPENDENT_MAPK1_3_ACTIVATION | 17 | -0.76847494 | -2.0634198 | 0 | 0.004234088 |
| GO_SKELETAL_MUSCLE_ORGAN_DEVELOPMENT | 75 | -0.53048426 | -2.0612414 | 0 | 0.004240084 |
| GO_AZUROPHIL_GRANULE | 105 | 0.5258156 | 1.8911104 | 0 | 0.004253427 |
| REACTOME_TRANSPORT_OF_MATURE_MRNAS_DERIVED_FROM_INTRONLESS_TRANSCRIPTS | 24 | 0.67862475 | 1.8904963 | 0 | 0.004270942 |
| GO_CD4_POSITIVE_ALPHA_BETA_T_CELL_ACTIVATION | 57 | -0.5594862 | -2.0686545 | 0 | 0.004314608 |
| GO_TELOMERE_MAINTENANCE_VIA_TELOMERE_LENGTHENING | 38 | 0.6248933 | 1.8883904 | 0 | 0.004314873 |
| GO_REGULATION_OF_FAT_CELL_DIFFERENTIATION | 69 | -0.5425325 | -2.0621521 | 0 | 0.0043158 |
| REACTOME_DUAL_INCISION_IN_TC_NER | 38 | 0.62826145 | 1.8886265 | 0 | 0.004317239 |
| GO_NEGATIVE_REGULATION_OF_TELOMERE_MAINTENANCE | 16 | 0.7417157 | 1.8873924 | 0 | 0.00433259 |
| REACTOME_NS1_MEDIATED_EFFECTS_ON_HOST_PATHWAYS | 25 | 0.6916261 | 1.8888971 | 0 | 0.004338358 |
| GO_CHROMOSOME_CENTROMERIC_REGION | 86 | 0.54085267 | 1.8875464 | 0 | 0.004365575 |

| | | | | | |
|---|---|---|---|---|---|
| GO_ENDOPEPTIDASE_COMPLEX | 33 | 0.6376735 | 1.8877996 | 0 | 0.004380433 |
| GO_GLOBAL_GENOME_NUCLEOTIDE_EXCISION_REPAIR | 17 | 0.7252804 | 1.8860856 | 0 | 0.004383958 |
| REACTOME_APC_C_CDH1_MEDIATED_DEGRADATION_OF_CDC20_AND_OTHER_APC_C_CDH1_TARGETED_PROTEINS_IN_LATE_MITOSIS_EARLY_G1 | 37 | 0.6171228 | 1.8843778 | 0 | 0.004536316 |
| GO_NUCLEOTIDE_EXCISION_REPAIR | 60 | 0.5737302 | 1.8813828 | 0 | 0.004644789 |
| GO_PROTEIN_CONTAINING_COMPLEX_LOCALIZATION | 155 | 0.49525163 | 1.8814744 | 0 | 0.004673321 |
| REACTOME_TRANSCRIPTION_OF_THE_HIV_GENOME | 40 | 0.61725825 | 1.8773019 | 0.001470588 | 0.004894362 |
| GO_NUCLEOTIDE_EXCISION_REPAIR_DNA_INCISION | 27 | 0.6678177 | 1.8762584 | 0 | 0.004933022 |
| REACTOME_GLYOXYLATE_METABOLISM_AND_GLYCINE_DEGRADATION | 26 | 0.66727716 | 1.8750308 | 0 | 0.00497714 |
| NABA_ECM_GLYCOPROTEINS | 103 | -0.49340373 | -2.036812 | 0 | 0.005085269 |
| GO_REGULATION_OF_EPIDERMIS_DEVELOPMENT | 50 | -0.56030226 | -2.0372992 | 0 | 0.005137422 |
| GO_REGULATION_OF_LEUKOCYTE_PROLIFERATION | 138 | -0.48305985 | -2.0400298 | 0 | 0.005137584 |
| GO_ALPHA_BETA_T_CELL_ACTIVATION | 89 | -0.5042754 | -2.038164 | 0 | 0.005165631 |
| REACTOME_MUSCLE_CONTRACTION | 117 | -0.4949417 | -2.042169 | 0 | 0.005165807 |
| GO_VOLTAGE_GATED_CALCIUM_CHANNEL_COMPLEX | 21 | -0.6961228 | -2.0428774 | 0 | 0.005166105 |
| REACTOME_SUMOYLATION_OF_UBIQUITINYLATION_PROTEINS | 22 | 0.67428833 | 1.8715179 | 0 | 0.005169444 |
| GO_TRANSLATIONAL_ELONGATION | 62 | 0.562065 | 1.8715552 | 0 | 0.005206815 |
| GO_ACTIVATION_OF_MAPK_ACTIVITY | 83 | -0.5186619 | -2.0454652 | 0 | 0.005225088 |
| GO_MICROBODY_MEMBRANE | 39 | 0.61357415 | 1.8694882 | 0.001490313 | 0.00523409 |
| REACTOME_INFLUENZA_INFECTION | 72 | 0.545982 | 1.8696296 | 0 | 0.005276993 |
| GO_UBIQUITIN_LIKE_PROTEIN_TRANSFERASE_ACTIVITY | 174 | 0.49165118 | 1.8678001 | 0 | 0.00532287 |
| GO_MUSCLE_ORGAN_DEVELOPMENT | 201 | -0.4498735 | -2.0310032 | 0 | 0.005416917 |
| GO_CHONDROCYTE_DEVELOPMENT | 21 | -0.6907847 | -2.0294557 | 0 | 0.00548577 |
| KEGG_HYPERTROPHIC_CARDIOMYOPATHY_HCM | 56 | -0.5497783 | -2.0312521 | 0 | 0.005498992 |
| REACTOME_G_ALPHA_S_SIGNALLING_EVENTS | 90 | -0.5108022 | -2.0279448 | 0 | 0.005553208 |
| REACTOME_MITOCHONDRIAL_FATTY_ACID_BETA_OXIDATION | 24 | 0.672503 | 1.8632102 | 0 | 0.005795198 |
| GO_CORECEPTOR_ACTIVITY | 26 | -0.6530976 | -2.017563 | 0 | 0.00582776 |
| GO_DNA_BINDING_TRANSCRIPTION_ACTIVATOR_ACTIVITY | 230 | -0.44974196 | -2.0185206 | 0 | 0.005837346 |
| GO_CD4_POSITIVE_ALPHA_BETA_T_CELL_DIFFERENTIATION | 47 | -0.5628817 | -2.023334 | 0 | 0.005864386 |
| GO_PEPTIDASE_COMPLEX | 44 | 0.5793685 | 1.8587615 | 0 | 0.005866022 |
| GO_CELL_PROLIFERATION_INVOLVED_IN_KIDNEY_DEVELOPMENT | 16 | -0.75836074 | -2.0187354 | 0 | 0.005869411 |
| GO_PHOSPHOLIPID_TRANSPORTER_ACTIVITY | 25 | 0.6637966 | 1.8620046 | 0.003246753 | 0.005872945 |
| GO_CELLULAR_AMINO_ACID_CATABOLIC_PROCESS | 89 | 0.5316016 | 1.861098 | 0 | 0.005874903 |
| GO_TRANSLATION_FACTOR_ACTIVITY_RNA_BINDING | 40 | 0.6089048 | 1.8599415 | 0 | 0.005878302 |
| GO_RNA_DEPENDENT_DNA_BIOSYNTHETIC_PROCESS | 33 | 0.6326763 | 1.8591287 | 0.001470588 | 0.005895592 |
| GO_CELL_FATE_DETERMINATION | 23 | -0.6826698 | -2.0207639 | 0 | 0.005901465 |
| GO_REGULATION_OF_TELOMERE_MAINTENANCE_VIA_TELOMERE_LENGTHENING | 27 | 0.6433276 | 1.8600187 | 0 | 0.005913686 |
| GO_EPIDERMAL_CELL_DIFFERENTIATION | 132 | -0.4769538 | -2.0194347 | 0 | 0.005937115 |
| GO_NEUROTRANSMITTER_RECEPTOR_ACTIVITY_INVOLVED_IN_REGULATION_OF_POSTSYNAPTIC_MEMBRANE_POTENTIAL | 26 | -0.6578429 | -2.0188794 | 0 | 0.005949814 |
| GO_MYOFIBRIL_ASSEMBLY | 33 | -0.5998155 | -2.0148294 | 0 | 0.005996564 |
| GO_POSITIVE_REGULATION_OF_BLOOD_CIRCULATION | 46 | -0.56322306 | -2.013604 | 0 | 0.006116236 |
| GO_GLOMERULAR_EPITHELIAL_CELL_DIFFERENTIATION | 19 | -0.6955448 | -2.0109186 | 0 | 0.006211437 |

| | | | | | |
|---|---|---|---|---|---|
| PID_DELTA_NP63_PATHWAY | 28 | -0.628234 | -2.0086772 | 0 | 0.006368953 |
| GO_RNA_MODIFICATION | 64 | 0.5548116 | 1.8536756 | 0 | 0.006371109 |
| REACTOME_REGULATION_OF_PTEN_STABILITY_AND_ACTIVITY | 39 | 0.61316466 | 1.8527236 | 0 | 0.006394534 |
| GO_MULTI_MULTICELLULAR_ORGANISM_PROCESS | 144 | -0.47801033 | -2.0072536 | 0 | 0.006394567 |
| KEGG_MISMATCH_REPAIR | 18 | 0.7085162 | 1.8516693 | 0.001692047 | 0.006458129 |
| GO_NEGATIVE_REGULATION_OF_MAPK_CASCADE | 104 | -0.49172017 | -2.0039573 | 0 | 0.006712338 |
| GO_ANTIMICROBIAL_HUMORAL_RESPONSE | 59 | -0.53919256 | -2.0022924 | 0 | 0.006774284 |
| GO_NEGATIVE_REGULATION_OF_ERK1_AND_ERK2_CASCADE | 43 | -0.5805972 | -1.9905481 | 0 | 0.00688505 |
| GO_ENDOCRINE_PANCREAS_DEVELOPMENT | 24 | -0.6478059 | -1.98973 | 0 | 0.006918946 |
| GO_REGULATION_OF_OSSIFICATION | 101 | -0.48970968 | -1.9912589 | 0 | 0.006923081 |
| GO_REGULATION_OF_BLOOD_CIRCULATION | 173 | -0.44290102 | -1.9924393 | 0 | 0.006925036 |
| GO_MUSCLE_TISSUE_DEVELOPMENT | 202 | -0.44566387 | -1.9977231 | 0 | 0.006951585 |
| BIOCARTA_KERATINOCYTE_PATHWAY | 35 | -0.5804216 | -1.9928827 | 0 | 0.006963257 |
| GO_CELL_CHEMOTAXIS | 184 | -0.44519535 | -1.9940941 | 0 | 0.006964863 |
| GO_REGULATION_OF_CARDIOCYTE_DIFFERENTIATION | 24 | -0.65377134 | -1.9999944 | 0.002702703 | 0.006977361 |
| GO_PEPTIDE_RECEPTOR_ACTIVITY | 70 | -0.5161934 | -1.9985024 | 0 | 0.006995121 |
| GO_GLOMERULAR_EPITHELIUM_DEVELOPMENT | 22 | -0.6817036 | -1.9879291 | 0 | 0.007004168 |
| GO_REGULATION_OF_HEART_GROWTH | 44 | -0.5596703 | -1.9958521 | 0 | 0.007004827 |
| PID_SMAD2_3NUCLEAR_PATHWAY | 49 | -0.55055785 | -1.9884481 | 0 | 0.007006464 |
| GO_RESPONSE_TO_MECHANICAL_STIMULUS | 132 | -0.46826553 | -1.9964644 | 0 | 0.007007095 |
| GO_POTASSIUM_ION_TRANSMEMBRANE_TRANSPORTER_ACTIVITY | 74 | -0.5042151 | -1.9977273 | 0 | 0.007032418 |
| GO_TRANSFERASE_COMPLEX | 371 | 0.4509893 | 1.8457967 | 0 | 0.00706292 |
| KEGG_LYSOSOME | 92 | 0.52340275 | 1.8438601 | 0 | 0.007076414 |
| REACTOME_REGULATION_OF_GLUCOKINASE_BY_GLUCOKINASE_REGULATORY_PROTEIN | 20 | 0.685364 | 1.8442618 | 0 | 0.007093366 |
| GO_SPLICEOSOMAL_COMPLEX | 91 | 0.51855636 | 1.8445472 | 0 | 0.007120594 |
| GO_MUSCLE_CONTRACTION | 218 | -0.43703547 | -1.98489 | 0 | 0.007126275 |
| GO_SNAP_RECEPTOR_ACTIVITY | 15 | 0.7572375 | 1.8410302 | 0 | 0.007150594 |
| GO_COFACTOR_METABOLIC_PROCESS | 281 | 0.45747268 | 1.8418093 | 0 | 0.007156308 |
| REACTOME_ASSEMBLY_OF_THE_PRE_REPLICATIVE_COMPLEX | 36 | 0.61453617 | 1.8428192 | 0.001545595 | 0.007157153 |
| GO_SCF_DEPENDENT_PROTEASOMAL_UBIQUITIN_DEPENDENT_PROTEIN_CATABOLIC_PROCESS | 46 | 0.5766347 | 1.841986 | 0 | 0.007192837 |
| GO_RNA_POLYMERASE_COMPLEX | 60 | 0.5674155 | 1.8410536 | 0 | 0.00720095 |
| GO_ORGANIC_ACID_CATABOLIC_PROCESS | 190 | 0.47579455 | 1.8396686 | 0 | 0.007251093 |
| GO_N_ACETYLTRANSFERASE_ACTIVITY | 40 | 0.5980567 | 1.8387288 | 0 | 0.007299561 |
| GO_CHROMOSOME_TELOMERIC_REGION | 67 | 0.54554766 | 1.8379124 | 0 | 0.007333102 |
| GO_POSITIVE_REGULATION_OF_ALPHA_BETA_T_CELL_ACTIVATION | 44 | -0.56470865 | -1.9808512 | 0 | 0.007343523 |
| GO_MITOCHONDRIAL_RESPIRATORY_CHAIN_COMPLEX_ASSEMBLY | 40 | 0.5898431 | 1.8374351 | 0.001501502 | 0.00734789 |
| REACTOME_GLOBAL_GENOME_NUCLEOTIDE_EXCISION_REPAIR_GG_NER | 48 | 0.5742983 | 1.8363531 | 0.001445087 | 0.007371641 |
| GO_G_PROTEIN_COUPLED_RECEPTOR_SIGNALING_PATHWAY_COUPLED_TO_CYCLIC_NUCLEOTIDE_SECOND_MESSENGER | 128 | -0.46639058 | -1.980991 | 0 | 0.0074177 |
| REACTOME_SUMOYLATION_OF_SUMOYLATION_PROTEINS | 23 | 0.66908646 | 1.8354247 | 0 | 0.007431331 |

| | | | | | |
|---|---|---|---|---|---|
| GO_NUCLEAR_TRANSCRIBED_MRNA_CATABOLIC_PROCESS_EXONUCLEOLYTIC | 17 | 0.70447165 | 1.8348101 | 0 | 0.007436186 |
| REACTOME_DEFECTS_IN_VITAMIN_AND_COFACTOR_METABOLISM | 16 | 0.71860236 | 1.8340596 | 0.003367003 | 0.007503731 |
| REACTOME_ORC1_REMOVAL_FROM_CHROMATIN | 38 | 0.60022545 | 1.8325019 | 0.001428571 | 0.007591134 |
| HALLMARK_FATTY_ACID_METABOLISM | 125 | 0.49675155 | 1.8325205 | 0 | 0.007641076 |
| GO_PROTEIN_MODIFICATION_BY_SMALL_PROTEIN_CONJUGATION | 407 | 0.4470453 | 1.8313997 | 0 | 0.007708927 |
| GO_POSITIVE_REGULATION_OF_EPIDERMIS_DEVELOPMENT | 22 | -0.6593259 | -1.9763776 | 0.002557545 | 0.007742699 |
| GO_CHROMATIN_ORGANIZATION_INVOLVED_IN_REGULATION_OF_TRANSCRIPTION | 33 | 0.61114883 | 1.8292178 | 0.001485884 | 0.007956462 |
| GO_CATALYTIC_ACTIVITY_ACTING_ON_A_TRNA | 43 | 0.59048927 | 1.8285238 | 0 | 0.008005273 |
| GO_GOLGI_ORGANIZATION | 66 | 0.54040015 | 1.8276029 | 0 | 0.00803635 |
| GO_EXTRACELLULAR_MATRIX | 319 | -0.40981242 | -1.969931 | 0 | 0.008424562 |
| KEGG_UBIQUITIN_MEDIATED_PROTEOLYSIS | 71 | 0.53783756 | 1.8233962 | 0 | 0.008484924 |
| GO_REGULATION_OF_CALCIUM_ION_TRANSMEMBRANE_TRANSPORTER_ACTIVITY | 53 | -0.5238759 | -1.9653672 | 0 | 0.008504963 |
| GO_NUCLEOBASE_CONTAINING_COMPOUND_KINASE_ACTIVITY | 24 | 0.64706767 | 1.8237597 | 0.003034901 | 0.008508571 |
| PID_CD8_TCR_DOWNSTREAM_PATHWAY | 42 | -0.5595287 | -1.9645473 | 0 | 0.008537542 |
| GO_REGULATION_OF_SYSTEM_PROCESS | 335 | -0.40784022 | -1.965811 | 0 | 0.008553066 |
| GO_REGULATION_OF_MAP_KINASE_ACTIVITY | 198 | -0.43807378 | -1.9678297 | 0 | 0.008555131 |
| REACTOME_G_ALPHA_I_SIGNALLING_EVENTS | 222 | -0.4315862 | -1.9671581 | 0 | 0.00856985 |
| GO_REGULATION_OF_GENE_SILENCING | 63 | 0.5462308 | 1.8223295 | 0 | 0.008605559 |
| GO_SMAD_PROTEIN_SIGNAL_TRANSDUCTION | 48 | -0.53983825 | -1.9678311 | 0 | 0.008638191 |
| GO_AMINO_ACID_ACTIVATION | 18 | 0.70809364 | 1.8213378 | 0.003367003 | 0.008703521 |
| BIOCARTA_IL1R_PATHWAY | 19 | -0.700473 | -1.9603696 | 0 | 0.008739511 |
| GO_NEPHRON_DEVELOPMENT | 97 | -0.48359096 | -1.9620379 | 0 | 0.008740866 |
| GO_OXIDOREDUCTASE_ACTIVITY_ACTING_ON_THE_ALDEHYDE_OR_OXO_GROUP_OF_DONORS | 33 | 0.6080834 | 1.8202238 | 0.003007519 | 0.008779418 |
| GO_MRNA_METABOLIC_PROCESS | 380 | 0.4483905 | 1.8203638 | 0 | 0.008808556 |
| GO_GOLGI_VESICLE_TRANSPORT | 206 | 0.46471432 | 1.8194715 | 0 | 0.008833149 |
| GO_ANTIGEN_PROCESSING_AND_PRESENTATION_OF_PEPTIDE_ANTIGEN_VIA_MHC_CLASS_I | 67 | 0.5381541 | 1.818199 | 0 | 0.008906409 |
| GO_RNA_SPLICING | 214 | 0.46490815 | 1.8182204 | 0 | 0.008956293 |
| GO_TRABECULA_MORPHOGENESIS | 29 | -0.60788393 | -1.9580495 | 0 | 0.008967541 |
| GO_SM_LIKE_PROTEIN_FAMILY_COMPLEX | 32 | 0.6196485 | 1.8163177 | 0.001547988 | 0.009100249 |
| GO_CULLIN_RING_UBIQUITIN_LIGASE_COMPLEX | 56 | 0.5472322 | 1.8158804 | 0 | 0.009102662 |
| REACTOME_DEFECTIVE_CFTR_CAUSES_CYSTIC_FIBROSIS | 31 | 0.61013454 | 1.8128203 | 0 | 0.009406909 |
| GO_CHROMOSOME_SEGREGATION | 126 | 0.4919189 | 1.8136551 | 0 | 0.009417678 |
| GO_AEROBIC_RESPIRATION | 52 | 0.5576912 | 1.8128897 | 0.003067485 | 0.009458314 |
| REACTOME_CHEMOKINE_RECEPTORS_BIND_CHEMOKINES | 43 | -0.55904686 | -1.9530009 | 0 | 0.009509749 |
| GO_U2_TYPE_SPLICEOSOMAL_COMPLEX | 42 | 0.5859198 | 1.8112147 | 0.001402525 | 0.009561141 |
| GO_REGULATION_OF_HORMONE_BIOSYNTHETIC_PROCESS | 21 | -0.6614501 | -1.9497521 | 0 | 0.009567097 |
| GO_PROTEIN_TYROSINE_SERINE_THREONINE_PHOSPHATASE_ACTIVITY | 21 | -0.67059135 | -1.9500388 | 0.002604167 | 0.009621938 |
| GO_ENDOPLASMIC_RETICULUM_TO_GOLGI_VESICLE_MEDIATED_TRANSPORT | 121 | 0.49759075 | 1.8101677 | 0 | 0.009658581 |
| HALLMARK_INFLAMMATORY_RESPONSE | 156 | -0.44872072 | -1.9456002 | 0 | 0.009796137 |

| | | | | | |
|---|---|---|---|---|---|
| GO_SENSORY_PERCEPTION_OF_SMELL | 30 | -0.60675144 | -1.9451574 | 0 | 0.009829696 |
| GO_SINGLE_STRANDED_DNA_BINDING | 62 | 0.5449585 | 1.8081453 | 0 | 0.009858414 |
| PID_FANCONI_PATHWAY | 25 | 0.64330083 | 1.8082178 | 0.001587302 | 0.009907274 |
| GO_COENZYME_BIOSYNTHETIC_PROCESS | 91 | 0.5078087 | 1.8053493 | 0 | 0.010149414 |
| GO_ALPHA_AMINO_ACID_METABOLIC_PROCESS | 130 | 0.48970535 | 1.8034296 | 0 | 0.010352281 |
| REACTOME_INTERLEUKIN_4_AND_INTERLEUKIN_13_SIGNALING | 77 | -0.507107 | -1.9400634 | 0 | 0.01044701 |
| GO_KERATINOCYTE_DIFFERENTIATION | 97 | -0.48007637 | -1.9367229 | 0 | 0.010579836 |
| GO_RENAL_SYSTEM_VASCULATURE_DEVELOPMENT | 21 | -0.6567393 | -1.938977 | 0 | 0.010603725 |
| GO_REGULATION_OF_ENDOTHELIAL_CELL_DIFFERENTIATION | 27 | -0.6100973 | -1.9368578 | 0 | 0.010653899 |
| GO_GROWTH_FACTOR_ACTIVITY | 80 | -0.48839608 | -1.9349307 | 0 | 0.010688055 |
| GO_CELL_DIFFERENTIATION_INVOLVED_IN_KIDNEY_DEVELOPMENT | 42 | -0.55161524 | -1.9370567 | 0 | 0.010729739 |
| REACTOME_ACTIVATION_OF_ATR_IN_RESPONSE_TO_REPLICATION_STRESS | 20 | 0.6651733 | 1.7959538 | 0.00487013 | 0.011470369 |
| GO_TAXIS | 364 | -0.39765015 | -1.9282025 | 0 | 0.01177936 |
| GO_BRANCHED_CHAIN_AMINO_ACID_METABOLIC_PROCESS | 18 | 0.6902902 | 1.7918103 | 0.004854369 | 0.011892466 |
| REACTOME_STABILIZATION_OF_P53 | 27 | 0.62562436 | 1.7919117 | 0.002994012 | 0.01194694 |
| GO_MUSCLE_SYSTEM_PROCESS | 272 | -0.40899387 | -1.9256126 | 0 | 0.011960006 |
| GO_DNA_DAMAGE_RESPONSE_DETECTION_OF_DNA_DAMAGE | 23 | 0.6401427 | 1.7921007 | 0 | 0.011979396 |
| GO_REGULATION_OF_GENE_EXPRESSION_EPIGENETIC | 136 | 0.4825811 | 1.7924552 | 0 | 0.011981956 |
| HALLMARK_EPITHELIAL_MESENCHYMAL_TRANSITION | 168 | -0.4331752 | -1.9260166 | 0 | 0.012001022 |
| GO_GATED_CHANNEL_ACTIVITY | 154 | -0.44430512 | -1.9238331 | 0 | 0.012271289 |
| GO_TELOMERE_ORGANIZATION | 69 | 0.52924347 | 1.7888807 | 0 | 0.012338247 |
| REACTOME_NUCLEAR_PORE_COMPLEX_NPC_DISASSEMBLY | 23 | 0.64867723 | 1.7884285 | 0 | 0.012345002 |
| GO_VESICLE_TETHERING_COMPLEX | 25 | 0.6442992 | 1.7877934 | 0 | 0.012388057 |
| GO_EPITHELIAL_CELL_DEVELOPMENT | 124 | -0.458995 | -1.9214529 | 0 | 0.012401589 |
| KEGG_CYTOKINE_CYTOKINE_RECEPTOR_INTERACTION | 158 | -0.44570246 | -1.9223311 | 0 | 0.012431212 |
| GO_RESPONSE_TO_FIBROBLAST_GROWTH_FACTOR | 83 | -0.48260054 | -1.9187119 | 0 | 0.012433007 |
| GO_VOLTAGE_GATED_CATION_CHANNEL_ACTIVITY | 61 | -0.5175986 | -1.9198972 | 0 | 0.012453399 |
| GO_NEPHRON_EPITHELIUM_DEVELOPMENT | 74 | -0.4999765 | -1.9189496 | 0 | 0.012462321 |
| GO_HEART_TRABECULA_MORPHOGENESIS | 21 | -0.6405385 | -1.9202683 | 0 | 0.012470382 |
| GO_CATION_CHANNEL_ACTIVITY | 145 | -0.44002262 | -1.9180264 | 0 | 0.01249359 |
| GO_POSITIVE_REGULATION_OF_T_CELL_PROLIFERATION | 59 | -0.51567304 | -1.917592 | 0 | 0.012503622 |
| GO_EXTRACELLULAR_LIGAND_GATED_ION_CHANNEL_ACTIVITY | 34 | -0.5891157 | -1.9203588 | 0 | 0.012540673 |
| REACTOME_DISORDERS_OF_TRANSMEMBRANE_TRANSPORTERS | 105 | 0.49642152 | 1.7854438 | 0 | 0.012689888 |
| GO_NEGATIVE_REGULATION_OF_LEUKOCYTE_APOPTOTIC_PROCESS | 36 | -0.5578643 | -1.9145255 | 0 | 0.012713629 |
| GO_NEGATIVE_REGULATION_OF_NOTCH_SIGNALING_PATHWAY | 21 | -0.656335 | -1.914539 | 0.002923977 | 0.012793971 |
| GO_LABYRINTHINE_LAYER_DEVELOPMENT | 34 | -0.57256866 | -1.9147514 | 0 | 0.012850844 |
| GO_G_PROTEIN_COUPLED_AMINE_RECEPTOR_ACTIVITY | 21 | -0.6497415 | -1.9094156 | 0 | 0.013066417 |
| GO_REGULATION_OF_EPIDERMAL_CELL_DIFFERENTIATION | 34 | -0.5852781 | -1.9111621 | 0 | 0.013138789 |
| NABA_PROTEOGLYCANS | 18 | -0.688051 | -1.9094517 | 0.00257732 | 0.013146989 |

| | | | | | |
|---|---|---|---|---|---|
| GO_SODIUM_ION_TRANSMEMBRANE_TRANSPORTER_ACTIVITY | 89 | -0.47590783 | -1.9096202 | 0 | 0.013216655 |
| GO_ACUTE_PHASE_RESPONSE | 29 | -0.6004407 | -1.9062278 | 0.002747253 | 0.013463715 |
| GO_REGULATION_OF_MONONUCLEAR_CELL_MIGRATION | 29 | -0.60819113 | -1.904014 | 0 | 0.013677218 |
| GO_LIPOPOLYSACCHARIDE_MEDIATED_SIGNALING_PATHWAY | 43 | -0.5512617 | -1.9047668 | 0 | 0.013678353 |
| GO_ADENYLATE_CYCLASE_ACTIVATING_G_PROTEIN_COUPLED_RECEPTOR_SIGNALING_PATHWAY | 68 | -0.4957215 | -1.9033848 | 0 | 0.013710627 |
| GO_DNA_REPAIR_COMPLEX | 23 | 0.6509176 | 1.7794026 | 0.001569859 | 0.013712969 |
| GO_OXIDOREDUCTASE_ACTIVITY_ACTING_ON_THE_ALDEHYDE_OR_OXO_GROUP_OF_DONORS_NAD_OR_NADP_AS_ACCEPTOR | 27 | 0.62651885 | 1.7788657 | 0.001501502 | 0.013726437 |
| GO_PROTEIN_MODIFICATION_BY_SMALL_PROTEIN_CONJUGATION_OR_REMOVAL | 478 | 0.43045953 | 1.7771645 | 0 | 0.013972755 |
| GO_REGULATION_OF_MUSCLE_CONTRACTION | 110 | -0.45476702 | -1.9012053 | 0 | 0.014022192 |
| GO_MISMATCH_REPAIR | 23 | 0.645497 | 1.7764806 | 0.001550388 | 0.014031099 |
| REACTOME_FORMATION_OF_RNA_POL_II_ELONGATION_COMPLEX | 31 | 0.60883313 | 1.7759811 | 0.003058104 | 0.0140534 |
| REACTOME_FORMATION_OF_THE_EARLY_ELONGATION_COMPLEX | 22 | 0.6453328 | 1.7751867 | 0.001602564 | 0.014135665 |
| GO_TRANSLATION_REGULATOR_ACTIVITY | 58 | 0.546479 | 1.7737389 | 0 | 0.014283847 |
| GO_COENZYME_BINDING | 213 | 0.4495609 | 1.774064 | 0 | 0.014305129 |
| GO_TELOMERIC_DNA_BINDING | 21 | 0.6492382 | 1.7711163 | 0.003139718 | 0.014463687 |
| GO_VESICLE_MEDIATED_TRANSPORT_TO_THE_PLASMA_MEMBRANE | 46 | 0.5581218 | 1.7719768 | 0 | 0.014473281 |
| KEGG_FATTY_ACID_METABOLISM | 33 | 0.5981525 | 1.7722025 | 0.004385965 | 0.014485148 |
| GO_CYTOPLASMIC_TRANSLATION | 44 | 0.57413334 | 1.7712927 | 0 | 0.01450612 |
| GO_NUCLEOTIDE_EXCISION_REPAIR_DNA_INCISION_3_TO_LESION | 16 | 0.70706505 | 1.769549 | 0.003262643 | 0.014596412 |
| GO_SKIN_DEVELOPMENT | 159 | -0.43942767 | -1.897268 | 0 | 0.014606915 |
| REACTOME_SUMOYLATION_OF_DNA_REPLICATION_PROTEINS | 31 | 0.60492355 | 1.7697774 | 0.003034901 | 0.014622149 |
| REACTOME_NEGATIVE_REGULATION_OF_MAPK_PATHWAY | 20 | -0.657477 | -1.8967073 | 0.005221932 | 0.014623432 |
| HALLMARK_APOPTOSIS | 123 | -0.45507953 | -1.8964316 | 0 | 0.014628401 |
| GO_REGULATION_OF_MAPK_CASCADE | 417 | -0.39046237 | -1.889045 | 0 | 0.01510893 |
| GO_GTP_METABOLIC_PROCESS | 16 | 0.699257 | 1.7663375 | 0.006768189 | 0.015137916 |
| REACTOME_BRANCHED_CHAIN_AMINO_ACID_CATABOLISM | 19 | 0.6772058 | 1.765943 | 0 | 0.015143588 |
| GO_POSITIVE_REGULATION_OF_LEUKOCYTE_PROLIFERATION | 85 | -0.47410256 | -1.8892771 | 0 | 0.015173189 |
| GO_REGULATION_OF_OSTEOBLAST_DIFFERENTIATION | 65 | -0.49469036 | -1.8880732 | 0 | 0.015218999 |
| GO_COLLAGEN_CONTAINING_EXTRACELLULAR_MATRIX | 264 | -0.40621063 | -1.8876969 | 0 | 0.015231589 |
| REACTOME_REGULATION_OF_RAS_BY_GAPS | 32 | 0.6038214 | 1.7652178 | 0 | 0.015235784 |
| GO_EMBRYONIC_PLACENTA_MORPHOGENESIS | 16 | -0.7025221 | -1.8900423 | 0.002754821 | 0.015259264 |
| GO_NEUROPEPTIDE_RECEPTOR_ACTIVITY | 17 | -0.67681706 | -1.8893901 | 0.002638523 | 0.015271081 |
| GO_LEUKOCYTE_CELL_CELL_ADHESION | 223 | -0.4132854 | -1.8915584 | 0 | 0.015289564 |
| GO_ERK1_AND_ERK2_CASCADE | 169 | -0.43025213 | -1.8912216 | 0 | 0.015290741 |
| HALLMARK_IL2_STAT5_SIGNALING | 137 | -0.43995008 | -1.8903222 | 0 | 0.015325598 |
| GO_PROTEIN_MONOUBIQUITINATION | 33 | 0.5855031 | 1.7642695 | 0.003076923 | 0.015364065 |
| GO_CALCIUM_CHANNEL_COMPLEX | 31 | -0.5942639 | -1.8847089 | 0 | 0.015446357 |

| | | | | | |
|---|---|---|---|---|---|
| GO_EMBRYONIC_PLACENTA_DEVELOPMENT | 53 | -0.5236325 | -1.8852146 | 0 | 0.015446802 |
| GO_REGULATION_OF_TELOMERE_MAINTENANCE | 36 | 0.5828294 | 1.7634368 | 0.00155521 | 0.01544768 |
| GO_ANTERIOR_POSTERIOR_AXIS_SPECIFICATION | 27 | -0.60042775 | -1.8853918 | 0.002680965 | 0.015511027 |
| GO_EPITHELIAL_CELL_DIFFERENTIATION_INVOLVED_IN_KIDNEY_DEVELOPMENT | 36 | -0.55947363 | -1.8811935 | 0 | 0.0158957 |
| GO_GLIAL_CELL_ACTIVATION | 38 | -0.55190367 | -1.8784211 | 0 | 0.016017057 |
| GO_MUSCLE_FILAMENT_SLIDING | 23 | -0.61734027 | -1.8796567 | 0.002739726 | 0.016060172 |
| GO_REGULATION_OF_CELLULAR_AMINO_ACID_METABOLIC_PROCESS | 37 | 0.5803683 | 1.7601061 | 0.001506024 | 0.016112527 |
| GO_KERATIN_FILAMENT | 16 | -0.70195544 | -1.8784598 | 0.002739726 | 0.016113546 |
| GO_MITOCHONDRIAL_ENVELOPE | 364 | 0.43230698 | 1.7587173 | 0 | 0.016114874 |
| GO_STRUCTURAL_CONSTITUENT_OF_NUCLEAR_PORE | 16 | 0.69870436 | 1.7583045 | 0.001602564 | 0.016115801 |
| GO_NEGATIVE_REGULATION_OF_LEUKOCYTE_PROLIFERATION | 51 | -0.51888114 | -1.8788309 | 0 | 0.016128588 |
| GO_VESICLE_DOCKING | 33 | 0.5900621 | 1.7588931 | 0.001515152 | 0.016159497 |
| GO_MODIFICATION_DEPENDENT_MACROMOLECULE_CATABOLIC_PROCESS | 305 | 0.43878365 | 1.7595412 | 0 | 0.016162077 |
| GO_FLAVIN_ADENINE_DINUCLEOTIDE_BINDING | 63 | 0.523986 | 1.7591093 | 0.001428571 | 0.016185135 |
| GO_SMALL_MOLECULE_CATABOLIC_PROCESS | 296 | 0.43835112 | 1.756995 | 0 | 0.016311329 |
| REACTOME_DEGRADATION_OF_GLI1_BY_THE_PROTEASOME | 31 | 0.6018632 | 1.7562215 | 0.001594896 | 0.0164028 |
| REACTOME_PROCESSING_OF_CAPPED_INTRON_CONTAINING_PRE_MRNA | 129 | 0.47370562 | 1.7554586 | 0 | 0.016484335 |
| GO_INTEGRIN_ACTIVATION | 16 | -0.6774438 | -1.8751273 | 0 | 0.016646069 |
| GO_NUCLEAR_PORE | 48 | 0.5550238 | 1.7536063 | 0 | 0.016714148 |
| GO_RNA_LOCALIZATION | 120 | 0.47480842 | 1.7539221 | 0.001285347 | 0.016747868 |
| GO_N_ACYLTRANSFERASE_ACTIVITY | 48 | 0.55137396 | 1.7524552 | 0.002923977 | 0.016855335 |
| REACTOME_CELLULAR_RESPONSE_TO_HYPOXIA | 39 | 0.57078254 | 1.7509478 | 0 | 0.016871972 |
| GO_TRANSCRIPTION_FACTOR_TFIID_COMPLEX | 22 | 0.6429887 | 1.7513654 | 0.001675042 | 0.016884144 |
| GO_OXIDOREDUCTASE_ACTIVITY_ACTING_ON_A_SULFUR_GROUP_OF_DONORS | 40 | 0.5545465 | 1.7515926 | 0.001488095 | 0.016905423 |
| GO_NUCLEAR_CHROMOSOME_SEGREGATION | 98 | 0.489486 | 1.7519269 | 0 | 0.016917652 |
| GO_INNER_MITOCHONDRIAL_MEMBRANE_ORGANIZATION | 16 | 0.6906567 | 1.7502285 | 0 | 0.016972806 |
| GO_REGULATION_OF_LYMPHOCYTE_ACTIVATION | 272 | -0.39963704 | -1.8722851 | 0 | 0.017083788 |
| GO_REGULATION_OF_STRIATED_MUSCLE_CELL_DIFFERENTIATION | 51 | -0.52236295 | -1.8700546 | 0 | 0.017365895 |
| GO_POSITIVE_REGULATION_OF_VIRAL_LIFE_CYCLE | 32 | 0.5859955 | 1.7481697 | 0.003062787 | 0.01740701 |
| REACTOME_POTASSIUM_CHANNELS | 50 | -0.5136193 | -1.8694186 | 0.002994012 | 0.017425383 |
| GO_ARACHIDONIC_ACID_SECRETION | 18 | -0.6489829 | -1.8702117 | 0 | 0.017468046 |
| GO_GLYCOSYL_COMPOUND_METABOLIC_PROCESS | 82 | 0.5029329 | 1.7474968 | 0 | 0.017495478 |
| GO_CELL_FATE_COMMITMENT | 125 | -0.4428935 | -1.8678691 | 0 | 0.017674277 |
| GO_POSTREPLICATION_REPAIR | 26 | 0.62022406 | 1.7452204 | 0.00309119 | 0.017945465 |
| REACTOME_ORGANELLE_BIOGENESIS_AND_MAINTENANCE | 145 | 0.46194667 | 1.7431151 | 0 | 0.01803792 |
| GO_PURINE_NUCLEOSIDE_METABOLIC_PROCESS | 36 | 0.58245236 | 1.7434946 | 0 | 0.018039856 |
| REACTOME_ABC_TRANSPORTER_DISORDERS | 41 | 0.56076825 | 1.7444644 | 0.001501502 | 0.018051779 |
| GO_POSTTRANSCRIPTIONAL_REGULATION_OF_GENE_EXPRESSION | 295 | 0.4345837 | 1.7440513 | 0 | 0.018058741 |
| GO_CHROMOSOME_LOCALIZATION | 36 | 0.5803406 | 1.7436644 | 0.001510574 | 0.01808333 |
| GO_POSITIVE_REGULATION_OF_EPITHELIAL_CELL_DIFFERENTIATION | 40 | -0.542404 | -1.8645176 | 0 | 0.018135538 |

| | | | | | |
|---|---|---|---|---|---|
| GO_REGULATION_OF_MONOCYTE_CHEMOTAXIS | 16 | -0.68380415 | -1.863756 | 0 | 0.018138122 |
| GO_TRANSCRIPTION_PREINITIATION_COMPLEX_ASSEMBLY | 23 | 0.6360431 | 1.7421759 | 0.001607717 | 0.01818871 |
| GO_T_CELL_PROLIFERATION | 121 | -0.43880042 | -1.8628409 | 0 | 0.018188804 |
| GO_CUL4_RING_E3_UBIQUITIN_LIGASE_COMPLEX | 15 | 0.6955625 | 1.7413188 | 0.006644518 | 0.018207107 |
| GO_POSITIVE_REGULATION_OF_ACUTE_INFLAMMATORY_RESPONSE | 16 | -0.68204594 | -1.8647062 | 0.004901961 | 0.018210031 |
| GO_POST_GOLGI_VESICLE_MEDIATED_TRANSPORT | 57 | 0.5247226 | 1.7416621 | 0 | 0.018215558 |
| GO_REGULATION_OF_MYELOID_CELL_DIFFERENTIATION | 127 | -0.43249637 | -1.8649454 | 0 | 0.018226897 |
| GO_LEUKOCYTE_PROLIFERATION | 189 | -0.4141077 | -1.8612542 | 0 | 0.018384881 |
| GO_NEGATIVE_REGULATION_OF_LYMPHOCYTE_ACTIVATION | 96 | -0.47138986 | -1.8605267 | 0 | 0.018492602 |
| GO_SENSORY_PERCEPTION | 258 | -0.3946183 | -1.8597388 | 0 | 0.018597461 |
| REACTOME_STRIATED_MUSCLE_CONTRACTION | 21 | -0.64133656 | -1.858852 | 0.005277045 | 0.018675288 |
| GO_METANEPHROS_DEVELOPMENT | 60 | -0.49539807 | -1.8578901 | 0 | 0.018684711 |
| GO_REGULATION_OF_ORGAN_GROWTH | 62 | -0.48644748 | -1.8576059 | 0 | 0.018685363 |
| GO_NEGATIVE_REGULATION_OF_INTERLEUKIN_6_PRODUCTION | 22 | -0.6168472 | -1.8580679 | 0.002793296 | 0.018730383 |
| GO_VESICLE_TARGETING | 56 | 0.529704 | 1.738012 | 0.004207574 | 0.018782025 |
| REACTOME_IMMUNOREGULATORY_INTERACTIONS_BETWEEN_A_LYMPHOID_AND_A_NON_LYMPHOID_CELL | 88 | -0.45881718 | -1.8567364 | 0 | 0.018785492 |
| REACTOME_TRNA_AMINOACYLATION | 15 | 0.7072465 | 1.7386051 | 0.008156607 | 0.0188038 |
| GO_VACUOLE_ORGANIZATION | 85 | 0.49518904 | 1.7380172 | 0.001369863 | 0.01886195 |
| REACTOME_DEADENYLATION_DEPENDENT_MRNA_DECAY | 22 | 0.64237684 | 1.7365618 | 0.004918033 | 0.018892007 |
| REACTOME_CELL_CYCLE | 315 | 0.42963377 | 1.7360619 | 0 | 0.01891793 |
| HALLMARK_ADIPOGENESIS | 143 | 0.46355283 | 1.7357562 | 0 | 0.018926809 |
| GO_REGULATION_OF_VASOCONSTRICTION | 43 | -0.51577455 | -1.8554919 | 0 | 0.018931331 |
| GO_SISTER_CHROMATID_SEGREGATION | 77 | 0.49838772 | 1.7366375 | 0 | 0.018954275 |
| GO_HISTONE_UBIQUITINATION | 18 | 0.68122333 | 1.7348877 | 0 | 0.019033464 |
| GO_CILIUM_ORGANIZATION | 146 | 0.46270522 | 1.7366376 | 0 | 0.019034252 |
| HALLMARK_P53_PATHWAY | 148 | -0.43002358 | -1.8467236 | 0 | 0.01937803 |
| GO_CYCLIC_NUCLEOTIDE_METABOLIC_PROCESS | 22 | -0.61902225 | -1.846899 | 0 | 0.019424057 |
| GO_KIDNEY_EPITHELIUM_DEVELOPMENT | 95 | -0.464867 | -1.8459358 | 0 | 0.019441944 |
| KEGG_SNARE_INTERACTIONS_IN_VESICULAR_TRANSPORT | 20 | 0.64081913 | 1.7322505 | 0.003372681 | 0.01946423 |
| GO_REGULATION_OF_T_CELL_ACTIVATION | 199 | -0.40989473 | -1.8453549 | 0 | 0.019471783 |
| GO_POSITIVE_T_CELL_SELECTION | 18 | -0.65229833 | -1.8470756 | 0.005333333 | 0.019478884 |
| GO_CELL_ADHESION_MEDIATOR_ACTIVITY | 35 | -0.5518729 | -1.8520396 | 0.003003003 | 0.01949294 |
| REACTOME_RNA_POLYMERASE_II_TRANSCRIBES_SNRNA_GENES | 36 | 0.56717813 | 1.7324384 | 0.006060606 | 0.019496612 |
| GO_OXIDOREDUCTASE_ACTIVITY_ACTING_ON_NAD_P_H_QUINONE_OR_SIMILAR_COMPOUND_AS_ACCEPTOR | 35 | 0.57788557 | 1.7318101 | 0.003174603 | 0.019503651 |
| GO_CYTOKINE_RECEPTOR_BINDING | 160 | -0.42685625 | -1.8471886 | 0 | 0.019525658 |
| GO_DNA_CONFORMATION_CHANGE | 127 | 0.46381328 | 1.7325772 | 0 | 0.01955731 |
| GO_POSITIVE_REGULATION_OF_LEUKOCYTE_CELL_CELL_ADHESION | 146 | -0.42523924 | -1.848347 | 0 | 0.019589243 |
| PID_IL23_PATHWAY | 26 | -0.6009779 | -1.8472469 | 0.002564103 | 0.019624772 |
| GO_MESENCHYMAL_CELL_PROLIFERATION | 35 | -0.5583865 | -1.8503741 | 0 | 0.019638222 |
| GO_NEGATIVE_REGULATION_OF_KINASE_ACTIVITY | 143 | -0.42714566 | -1.8483611 | 0 | 0.01969022 |
| GO_ADENYLATE_CYCLASE_INHIBITING_G_PROTEIN_COUPLED_RECEPTOR_SIGNALING_PATHWAY | 46 | -0.5271379 | -1.8416262 | 0.003289474 | 0.019694751 |

| | | | | | |
|---|---|---|---|---|---|
| GO_TRANSFORMING_GROWTH_FACTOR_BETA_RECEPTOR_BINDING | 30 | -0.57461035 | -1.8504515 | 0 | 0.019714786 |
| GO_STRIATED_MUSCLE_CONTRACTION | 104 | -0.43897718 | -1.8473178 | 0 | 0.019716127 |
| KEGG_PROPANOATE_METABOLISM | 24 | 0.62758297 | 1.730608 | 0.003215434 | 0.019751124 |
| HALLMARK_MYOGENESIS | 156 | -0.42450222 | -1.8405374 | 0 | 0.019757092 |
| GO_ANCHORED_COMPONENT_OF_MEMBRANE | 82 | -0.4711813 | -1.848564 | 0 | 0.01977403 |
| GO_EXTRACELLULAR_STRUCTURE_ORGANIZATION | 242 | -0.3996081 | -1.8407578 | 0 | 0.019787624 |
| GO_NEGATIVE_REGULATION_OF_CHEMOTAXIS | 34 | -0.5585958 | -1.8416489 | 0.002793296 | 0.019788984 |
| GO_NEGATIVE_REGULATION_OF_DNA_BINDING | 30 | -0.57880163 | -1.8429723 | 0 | 0.019794507 |
| GO_MEMBRANE_REPOLARIZATION_DURING_ACTION_POTENTIAL | 15 | -0.6844992 | -1.841906 | 0.005076142 | 0.019834546 |
| REACTOME_OLFACTORY_SIGNALING_PATHWAY | 24 | -0.5994757 | -1.8485821 | 0.00248139 | 0.019868275 |
| GO_MESENCHYME_MORPHOGENESIS | 34 | -0.546958 | -1.8431175 | 0.00286533 | 0.019882346 |
| GO_METANEPHRIC_NEPHRON_DEVELOPMENT | 24 | -0.6130286 | -1.8420306 | 0.002840909 | 0.019889373 |
| GO_SODIUM_ION_HOMEOSTASIS | 40 | -0.54260725 | -1.848804 | 0.002816901 | 0.019954642 |
| REACTOME_CARDIAC_CONDUCTION | 71 | -0.48946163 | -1.8432817 | 0 | 0.019962925 |
| REACTOME_CLASS_B_2_SECRETIN_FAMILY_RECEPTORS | 48 | -0.50909984 | -1.8388801 | 0 | 0.020025056 |
| REACTOME_DEGRADATION_OF_AXIN | 30 | 0.6006133 | 1.7287382 | 0.001545595 | 0.020100027 |
| GO_MACROAUTOPHAGY | 150 | 0.45706385 | 1.728376 | 0 | 0.020125577 |
| REACTOME_TRANSCRIPTIONAL_REGULATION_OF_GRANULOPOIESIS | 18 | -0.6575907 | -1.8381693 | 0.002538071 | 0.020139452 |
| GO_POTASSIUM_CHANNEL_COMPLEX | 42 | -0.5197448 | -1.8376628 | 0 | 0.020156747 |
| GO_NEGATIVE_REGULATION_OF_SMOOTH_MUSCLE_CELL_PROLIFERATION | 32 | -0.56074864 | -1.8369408 | 0.002638523 | 0.020167094 |
| GO_HUMORAL_IMMUNE_RESPONSE | 163 | -0.42214218 | -1.8363708 | 0 | 0.020170175 |
| GO_TRICARBOXYLIC_ACID_CYCLE | 26 | 0.6009178 | 1.72714 | 0.0064 | 0.020181654 |
| GO_RNA_EXPORT_FROM_NUCLEUS | 74 | 0.50937635 | 1.7277544 | 0 | 0.020191437 |
| GO_REGULATION_OF_SMOOTH_MUSCLE_CONTRACTION | 40 | -0.53416795 | -1.8346366 | 0 | 0.02019812 |
| GO_CALCIUM_INDEPENDENT_CELL_CELL_ADHESION_VIA_PLASMA_MEMBRANE_CELL_ADHESION_MOLECULES | 16 | -0.6654321 | -1.8364962 | 0 | 0.020215401 |
| GO_DNA_GEOMETRIC_CHANGE | 67 | 0.51122093 | 1.7271593 | 0.00136612 | 0.020262059 |
| HALLMARK_ESTROGEN_RESPONSE_EARLY | 141 | -0.42475492 | -1.8346716 | 0 | 0.020274185 |
| GO_SMALL_NUCLEAR_RIBONUCLEOPROTEIN_COMPLEX | 27 | 0.6134495 | 1.7263799 | 0.004622496 | 0.020278491 |
| GO_ORGANELLE_FUSION | 61 | 0.515311 | 1.7259413 | 0 | 0.020315731 |
| KEGG_CARDIAC_MUSCLE_CONTRACTION | 42 | -0.5298168 | -1.8352118 | 0 | 0.020326585 |
| GO_REGULATION_OF_T_CELL_APOPTOTIC_PROCESS | 21 | -0.6180533 | -1.8349324 | 0.002762431 | 0.020326963 |
| GO_REGULATION_OF_CHEMOTAXIS | 132 | -0.43428013 | -1.8259838 | 0 | 0.020727254 |
| GO_POSITIVE_REGULATION_OF_P38MAPK_CASCADE | 18 | -0.6506764 | -1.8318026 | 0.002506266 | 0.020781038 |
| PID_NFAT_TFPATHWAY | 26 | -0.58701026 | -1.8260483 | 0.002808989 | 0.020800795 |
| GO_NEGATIVE_REGULATION_OF_FAT_CELL_DIFFERENTIATION | 25 | -0.6133651 | -1.8252958 | 0.002801121 | 0.020821093 |
| GO_POSITIVE_REGULATION_OF_EPITHELIAL_CELL_PROLIFERATION | 118 | -0.4447715 | -1.8263592 | 0 | 0.02082416 |
| GO_MESONEPHROS_DEVELOPMENT | 64 | -0.47451735 | -1.831206 | 0 | 0.020839952 |
| GO_PHOSPHATE_ION_TRANSPORT | 17 | -0.6615938 | -1.8267114 | 0.002590674 | 0.020848162 |
| GO_POSITIVE_REGULATION_OF_ALPHA_BETA_T_CELL_DIFFERENTIATION | 33 | -0.5630678 | -1.8278525 | 0.002985075 | 0.02087555 |
| GO_HOMOPHILIC_CELL_ADHESION_VIA_PLASMA_MEMBRANE_ADHESION_MOLECULES | 65 | -0.47493827 | -1.826814 | 0 | 0.020908121 |
| GO_TRANSCRIPTION_COFACTOR_BINDING | 23 | -0.6002115 | -1.828412 | 0 | 0.02094617 |
| GO_POSITIVE_REGULATION_OF_HEART_CONTRACTION | 23 | -0.6091344 | -1.8279096 | 0.005263158 | 0.02095895 |
| GO_LEUKOCYTE_CHEMOTAXIS | 139 | -0.42154792 | -1.826859 | 0 | 0.020998243 |

| | | | | | |
|---|---|---|---|---|---|
| GO_PASSIVE_TRANSMEMBRANE_TRANSPORTER_ACTIVITY | 203 | -0.40696776 | -1.8295085 | 0 | 0.021011971 |
| GO_GLAND_MORPHOGENESIS | 75 | -0.46808878 | -1.8285689 | 0 | 0.021015342 |
| GO_CYTOPLASMIC_TRANSLATIONAL_INITIATION | 20 | 0.6580612 | 1.7230135 | 0.001628664 | 0.02102257 |
| GO_POSITIVE_REGULATION_OF_ENDOTHELIAL_CELL_PROLIFERATION | 60 | -0.48799828 | -1.8240491 | 0.003436426 | 0.021048108 |
| GO_LEUKOCYTE_ACTIVATION_INVOLVED_IN_INFLAMMATORY_RESPONSE | 34 | -0.5466276 | -1.8287144 | 0 | 0.021092845 |
| GO_FLUID_TRANSPORT | 18 | -0.65280104 | -1.8295691 | 0.004889976 | 0.021105358 |
| GO_MUSCLE_CELL_PROLIFERATION | 131 | -0.43116164 | -1.8228403 | 0 | 0.021187786 |
| GO_ACTOMYOSIN_STRUCTURE_ORGANIZATION | 105 | -0.4389938 | -1.821779 | 0 | 0.021370174 |
| GO_AUTOPHAGOSOME | 49 | 0.5342944 | 1.7204157 | 0 | 0.02168679 |
| GO_REGULATION_OF_HETEROTYPIC_CELL_CELL_ADHESION | 16 | -0.67716676 | -1.8190105 | 0.004987531 | 0.021904714 |
| GO_LYMPHOCYTE_DIFFERENTIATION | 205 | -0.40701136 | -1.817393 | 0 | 0.02229321 |
| GO_REGULATION_OF_CARDIAC_MUSCLE_CELL_DIFFERENTIATION | 18 | -0.6364772 | -1.8155757 | 0 | 0.022471683 |
| GO_POSITIVE_REGULATION_OF_CANONICAL_WNT_SIGNALING_PATHWAY | 79 | 0.49877203 | 1.7173448 | 0 | 0.022520348 |
| KEGG_HEMATOPOIETIC_CELL_LINEAGE | 57 | -0.49014613 | -1.8157111 | 0 | 0.022521524 |
| GO_REGULATION_OF_VASCULATURE_DEVELOPMENT | 219 | -0.39553088 | -1.815763 | 0 | 0.022606952 |
| GO_CATION_CHANNEL_COMPLEX | 93 | -0.44575298 | -1.8140665 | 0 | 0.022612993 |
| GO_DEFENSE_RESPONSE_TO_BACTERIUM | 135 | -0.42673174 | -1.8142966 | 0 | 0.02263542 |
| GO_GRANULOCYTE_MIGRATION | 90 | -0.44729134 | -1.8130182 | 0 | 0.022637373 |
| GO_CELLULAR_RESPONSE_TO_VASCULAR_ENDOTHELIAL_GROWTH_FACTOR_STIMULUS | 34 | -0.54887277 | -1.8130264 | 0 | 0.022721577 |
| GO_REGULATION_OF_INTERLEUKIN_1_SECRETION | 28 | -0.56806415 | -1.8109093 | 0.002680965 | 0.022889948 |
| GO_MITOCHONDRIAL_SMALL_RIBOSOMAL_SUBUNIT | 15 | 0.6951907 | 1.7157826 | 0.006734007 | 0.02289533 |
| GO_POSITIVE_REGULATION_OF_LEUKOCYTE_DIFFERENTIATION | 97 | -0.43707404 | -1.8111827 | 0 | 0.022900403 |
| GO_REGULATION_OF_ASTROCYTE_DIFFERENTIATION | 19 | -0.637939 | -1.8114388 | 0.002747253 | 0.022902483 |
| GO_REGULATION_OF_POSTTRANSCRIPTIONAL_GENE_SILENCING | 52 | 0.52410436 | 1.7152863 | 0 | 0.022974612 |
| REACTOME_DISEASES_ASSOCIATED_WITH_GLYCOSYLATION_PRECURSOR_BIOSYNTHESIS | 17 | 0.67750806 | 1.7148722 | 0.008223685 | 0.023000838 |
| REACTOME_ABC_FAMILY_PROTEINS_MEDIATED_TRANSPORT | 54 | 0.522442 | 1.7139419 | 0 | 0.023180244 |
| GO_SMOOTH_MUSCLE_CELL_PROLIFERATION | 96 | -0.44829518 | -1.8094423 | 0 | 0.02318038 |
| GO_EPITHELIAL_CELL_DIFFERENTIATION | 378 | -0.3778311 | -1.8097397 | 0 | 0.023184452 |
| GO_GOLGI_LUMEN | 56 | -0.4971282 | -1.8088278 | 0.003322259 | 0.023242963 |
| GO_TRANSLATIONAL_INITIATION | 83 | 0.49625683 | 1.713192 | 0.001402525 | 0.023293748 |
| GO_RESPONSE_TO_MINERALOCORTICOID | 22 | -0.61508244 | -1.8071827 | 0.005376344 | 0.023396105 |
| GO_NEUROINFLAMMATORY_RESPONSE | 47 | -0.5126582 | -1.8078758 | 0 | 0.023418868 |
| GO_ASTROCYTE_DIFFERENTIATION | 48 | -0.5010519 | -1.8075451 | 0 | 0.023420395 |
| KEGG_PROTEASOME | 24 | 0.6175765 | 1.711341 | 0.00152207 | 0.023746617 |
| GO_OXIDOREDUCTASE_COMPLEX | 65 | 0.5058043 | 1.7090299 | 0 | 0.024191849 |
| REACTOME_CELL_CYCLE_MITOTIC | 265 | 0.42801258 | 1.7093279 | 0 | 0.02420398 |
| REACTOME_DUAL_INCISION_IN_GG_NER | 27 | 0.5962728 | 1.709621 | 0.00462963 | 0.024223963 |
| GO_T_CELL_ACTIVATION_INVOLVED_IN_IMMUNE_RESPONSE | 64 | -0.46681315 | -1.8020422 | 0.003597122 | 0.024397554 |
| GO_T_CELL_DIFFERENTIATION_INVOLVED_IN_IMMUNE_RESPONSE | 40 | -0.5206889 | -1.8025618 | 0 | 0.024399633 |

| | | | | | |
|---|---|---|---|---|---|
| HALLMARK_UV_RESPONSE_UP | 116 | -0.4306259 | -1.8028598 | 0 | 0.02441479 |
| GO_CARTILAGE_DEVELOPMENT | 114 | -0.4345809 | -1.800866 | 0 | 0.02446547 |
| GO_HEART_GROWTH | 56 | -0.48746094 | -1.800586 | 0 | 0.024470696 |
| GO_POSITIVE_REGULATION_OF_SMOOTH_MUSCLE_CELL_PROLIFERATION | 62 | -0.47652712 | -1.8013514 | 0 | 0.024480164 |
| REACTOME_HIV_TRANSCRIPTION_ELONGATION | 24 | 0.61962694 | 1.7065867 | 0.001547988 | 0.024831422 |
| GO_MONOCARBOXYLIC_ACID_CATABOLIC_PROCESS | 89 | 0.4860476 | 1.7060754 | 0.001340483 | 0.02487524 |
| GO_PRERIBOSOME | 36 | 0.5644962 | 1.7056085 | 0.006042296 | 0.024926612 |
| REACTOME_RRNA_MODIFICATION_IN_THE_NUCLEUS_AND_CYTOSOL | 32 | 0.5832469 | 1.705275 | 0 | 0.02495753 |
| GO_DEFENSE_RESPONSE_TO_GRAM_POSITIVE_BACTERIUM | 41 | -0.5163303 | -1.7963414 | 0 | 0.025126828 |
| WNT_SIGNALING | 56 | -0.47749168 | -1.7965004 | 0 | 0.025157984 |
| GO_CYCLIC_NUCLEOTIDE_MEDIATED_SIGNALING | 112 | -0.43472365 | -1.796764 | 0 | 0.0252068 |
| PID_IL12_STAT4_PATHWAY | 24 | -0.5848556 | -1.7968657 | 0.002702703 | 0.025250496 |
| GO_SLEEP | 19 | -0.62478167 | -1.7941532 | 0.005050505 | 0.0252603 |
| GO_MEMBRANE_REPOLARIZATION | 28 | -0.56245446 | -1.7929103 | 0.002824859 | 0.02530685 |
| GO_CIS_REGULATORY_REGION_BINDING | 308 | -0.38210228 | -1.7932541 | 0 | 0.025318002 |
| PID_FGF_PATHWAY | 29 | -0.5722531 | -1.7925208 | 0.002702703 | 0.025326667 |
| GO_MYOTUBE_DIFFERENTIATION | 54 | -0.48659176 | -1.794259 | 0 | 0.025334254 |
| GO_MITOTIC_METAPHASE_PLATE_CONGRESSION | 25 | 0.60852253 | 1.7013203 | 0.0048 | 0.025370155 |
| BIOCARTA_IL12_PATHWAY | 16 | -0.6539258 | -1.7943324 | 0.007352941 | 0.025396096 |
| HALLMARK_KRAS_SIGNALING_UP | 152 | -0.40893576 | -1.7946943 | 0 | 0.025414139 |
| GO_MEMBRANE_COAT | 49 | 0.53127784 | 1.7014025 | 0 | 0.025426969 |
| REACTOME_REGULATION_OF_LIPID_METABOLISM_BY_PPARALPHA | 71 | 0.49465734 | 1.7015527 | 0 | 0.025457386 |
| KEGG_BASAL_TRANSCRIPTION_FACTORS | 21 | 0.6428405 | 1.7019933 | 0 | 0.025504466 |
| REACTOME_BASE_EXCISION_REPAIR | 31 | 0.58214676 | 1.7016898 | 0 | 0.02550775 |
| GO_REGULATION_OF_CHROMOSOME_ORGANIZATION | 149 | 0.44808418 | 1.7003522 | 0 | 0.025516534 |
| REACTOME_CARGO_TRAFFICKING_TO_THE_PERICILIARY_MEMBRANE | 26 | 0.6054993 | 1.7021897 | 0.003169572 | 0.02552347 |
| GO_ENDONUCLEASE_ACTIVITY_ACTIVE_WITH_EITHER_RIBO_OR_DEOXYRIBONUCLEIC_ACIDS_AND_PRODUCING_5_PHOSPHOMONOESTERS | 18 | 0.6575456 | 1.7024454 | 0.006557377 | 0.025532663 |
| REACTOME_RESOLUTION_OF_SISTER_CHROMATID_COHESION | 61 | 0.50433844 | 1.7033452 | 0.001449275 | 0.02553295 |
| GO_OSTEOCLAST_DIFFERENTIATION | 54 | -0.49175903 | -1.7887869 | 0 | 0.025545241 |
| GO_REGULATION_OF_MACROPHAGE_ACTIVATION | 39 | -0.53070474 | -1.7910821 | 0.008955224 | 0.025549801 |
| GO_TRANSPORTER_COMPLEX | 151 | -0.41467217 | -1.7902611 | 0 | 0.02556866 |
| REACTOME_MITOCHONDRIAL_BIOGENESIS | 43 | 0.5397625 | 1.7026552 | 0.002998501 | 0.025571486 |
| REACTOME_NONHOMOLOGOUS_END_JOINING_NHEJ | 19 | 0.6493012 | 1.6999645 | 0.005128205 | 0.025577081 |
| REACTOME_SYNTHESIS_OF_ACTIVE_UBIQUITIN_ROLES_OF_E1_AND_E2_ENZYMES | 19 | 0.6449134 | 1.7004511 | 0.007861636 | 0.025583247 |
| GO_NEGATIVE_REGULATION_OF_STRIATED_MUSCLE_CELL_DIFFERENTIATION | 20 | -0.63531023 | -1.7895851 | 0 | 0.025593378 |
| GO_ORGAN_OR_TISSUE_SPECIFIC_IMMUNE_RESPONSE | 16 | -0.6592298 | -1.7905247 | 0.005012531 | 0.025593488 |
| REACTOME_DNA_REPLICATION_PRE_INITIATION | 48 | 0.53524417 | 1.7028266 | 0 | 0.025610719 |
| GO_CARDIAC_VENTRICLE_MORPHOGENESIS | 47 | -0.5057939 | -1.7888055 | 0.002949853 | 0.025623871 |
| GO_LABYRINTHINE_LAYER_BLOOD_VESSEL_DEVELOPMENT | 15 | -0.6557767 | -1.7889212 | 0.00486618 | 0.025696697 |
| GO_ALPHA_AMINO_ACID_CATABOLIC_PROCESS | 72 | 0.49953505 | 1.6993531 | 0 | 0.02570164 |
| GO_NADP_METABOLIC_PROCESS | 23 | 0.625165 | 1.6984032 | 0.004731861 | 0.025880065 |

| | | | | | |
|---|---|---|---|---|---|
| KEGG_CITRATE_CYCLE_TCA_CYCLE | 26 | 0.59227717 | 1.6974185 | 0.001592357 | 0.026104094 |
| GO_PEPTIDE_BIOSYNTHETIC_PROCESS | 306 | 0.42346632 | 1.6968807 | 0 | 0.026178177 |
| GO_CARDIAC_MUSCLE_TISSUE_MORPHOGENESIS | 38 | -0.528224 | -1.7856039 | 0 | 0.026191944 |
| GO_NEGATIVE_REGULATION_OF_AXON_EXTENSION | 21 | -0.59283423 | -1.7852907 | 0.002604167 | 0.026212791 |
| GO_SARCOMERE_ORGANIZATION | 21 | -0.612252 | -1.7856252 | 0 | 0.026260119 |
| KEGG_DILATED_CARDIOMYOPATHY | 59 | -0.47396046 | -1.7856382 | 0 | 0.026352584 |
| GO_POSITIVE_REGULATION_OF_VIRAL_PROCESS | 60 | 0.5120428 | 1.6959709 | 0 | 0.02636525 |
| GO_PML_BODY | 51 | 0.5182684 | 1.6943252 | 0.001490313 | 0.026512323 |
| REACTOME_AUF1_HNRNP_D0_BINDS_AND_DESTABILIZES_MRNA | 29 | 0.5910248 | 1.6948447 | 0.004846527 | 0.026526738 |
| GO_SERINE_FAMILY_AMINO_ACID_METABOLIC_PROCESS | 26 | 0.59421057 | 1.6944904 | 0.003164557 | 0.026549853 |
| REACTOME_METABOLISM_OF_POLYAMINES | 36 | 0.565687 | 1.6950859 | 0.004658385 | 0.02656446 |
| GO_POSITIVE_REGULATION_OF_VIRAL_GENOME_REPLICATION | 17 | 0.661138 | 1.6936713 | 0.008196721 | 0.026628086 |
| REACTOME_METABOLISM_OF_AMINO_ACIDS_AND_DERIVATIVES | 215 | 0.43196058 | 1.693379 | 0 | 0.026629834 |
| GO_4_IRON_4_SULFUR_CLUSTER_BINDING | 30 | 0.5776285 | 1.6929164 | 0.004636785 | 0.026665967 |
| GO_ACETYLTRANSFERASE_ACTIVITY | 56 | 0.5174341 | 1.6922348 | 0.007183908 | 0.02668524 |
| GO_POSITIVE_REGULATION_OF_MYELOID_LEUKOCYTE_DIFFERENTIATION | 33 | -0.5440642 | -1.7827232 | 0.002717391 | 0.026685532 |
| GO_ORGANELLE_ENVELOPE_LUMEN | 57 | 0.5152689 | 1.6924936 | 0.002735978 | 0.026690438 |
| GO_AP_TYPE_MEMBRANE_COAT_ADAPTOR_COMPLEX | 17 | 0.65865505 | 1.6917593 | 0.006655574 | 0.026722968 |
| REACTOME_MITOTIC_PROPHASE | 47 | 0.52931345 | 1.6912771 | 0.004366812 | 0.026781132 |
| GO_RESPONSE_TO_MORPHINE | 19 | -0.628484 | -1.7816666 | 0.007712083 | 0.026823966 |
| GO_REGULATION_OF_CELL_ACTIVATION | 360 | -0.37123093 | -1.7817888 | 0 | 0.026881235 |
| REACTOME_PLASMA_LIPOPROTEIN_ASSEMBLY | 15 | 0.68189216 | 1.6904244 | 0.01141925 | 0.026946934 |
| GO_REGULATION_OF_HORMONE_METABOLIC_PROCESS | 27 | -0.5710483 | -1.77948 | 0.005602241 | 0.027191792 |
| GO_NEGATIVE_REGULATION_OF_CELL_ACTIVATION | 134 | -0.41665128 | -1.7800263 | 0 | 0.027231626 |
| GO_POSITIVE_REGULATION_OF_VASCULATURE_DEVELOPMENT | 127 | -0.41744694 | -1.7796222 | 0 | 0.027243607 |
| GO_UBIQUITIN_LIKE_PROTEIN_LIGASE_ACTIVITY | 92 | 0.48082653 | 1.6887623 | 0 | 0.027411025 |
| GO_CELLULAR_PROTEIN_CATABOLIC_PROCESS | 388 | 0.41188857 | 1.6882379 | 0 | 0.027452927 |
| GO_CONDENSED_CHROMOSOME_CENTROMERIC_REGION | 53 | 0.51847804 | 1.687706 | 0 | 0.027525676 |
| GO_CILIARY_BASAL_BODY | 42 | 0.5364541 | 1.6855905 | 0.00141844 | 0.027879667 |
| GO_POSITIVE_REGULATION_OF_TELOMERE_MAINTENANCE | 25 | 0.59776485 | 1.6864042 | 0.00913242 | 0.027887253 |
| GO_NEGATIVE_REGULATION_OF_TRANSFERASE_ACTIVITY | 157 | -0.40496832 | -1.7755843 | 0 | 0.027910292 |
| REACTOME_MITOTIC_PROMETAPHASE | 98 | 0.4712176 | 1.6857357 | 0 | 0.02792647 |
| GO_VASCULAR_ENDOTHELIAL_GROWTH_FACTOR_PRODUCTION | 20 | -0.61019206 | -1.7760358 | 0.002762431 | 0.027948173 |
| GO_COLLAGEN_METABOLIC_PROCESS | 67 | -0.46697468 | -1.7757034 | 0 | 0.027952598 |
| GO_NEGATIVE_REGULATION_OF_CELL_GROWTH | 111 | -0.42839 | -1.7749561 | 0 | 0.027956929 |
| KEGG_GLYCINE_SERINE_AND_THREONINE_METABOLISM | 24 | 0.6151656 | 1.6858877 | 0.004731861 | 0.0279625 |
| GO_REGULATION_OF_BONE_REMODELING | 29 | -0.56220794 | -1.7761179 | 0.002840909 | 0.028002597 |
| GO_T_CELL_DIFFERENTIATION | 146 | -0.4031172 | -1.7742544 | 0 | 0.028010622 |
| PID_P53_DOWNSTREAM_PATHWAY | 92 | -0.43976605 | -1.7750857 | 0 | 0.028010758 |
| GO_VESICLE_ORGANIZATION | 175 | 0.44235992 | 1.6846765 | 0 | 0.028062005 |
| GO_PERK_MEDIATED_UNFOLDED_PROTEIN_RESPONSE | 16 | -0.65363824 | -1.773637 | 0.005102041 | 0.02807119 |
| GO_POSITIVE_REGULATION_OF_ORGAN_GROWTH | 33 | -0.5309551 | -1.7714753 | 0 | 0.028576117 |

| | | | | | |
|---|---|---|---|---|---|
| GO_CHEMOKINE_ACTIVITY | 32 | -0.54090834 | -1.7710785 | 0.005509642 | 0.028616779 |
| GO_NEGATIVE_REGULATION_OF_LYMPHOCYTE_DIFFERENTIATION | 36 | -0.5227331 | -1.7689929 | 0.005970149 | 0.028959565 |
| GO_REGULATION_OF_KIDNEY_DEVELOPMENT | 41 | -0.5175068 | -1.7693669 | 0.003076923 | 0.028965402 |
| BIOCARTA_NFKB_PATHWAY | 15 | -0.6555387 | -1.7675747 | 0.013157895 | 0.029204566 |
| GO_C21_STEROID_HORMONE_BIOSYNTHETIC_PROCESS | 19 | -0.6095283 | -1.7676563 | 0.008108108 | 0.029255437 |
| GO_REGULATION_OF_ALPHA_BETA_T_CELL_ACTIVATION | 58 | -0.48236218 | -1.7679447 | 0.006514658 | 0.029256182 |
| GO_DENDRITE_MEMBRANE | 22 | -0.58932865 | -1.7663352 | 0.005571031 | 0.029490123 |
| GO_CARDIOVASCULAR_SYSTEM_DEVELOPMENT | 465 | -0.35557577 | -1.7655089 | 0 | 0.029536353 |
| GO_LEUKOCYTE_DIFFERENTIATION | 305 | -0.37388182 | -1.7656604 | 0 | 0.029582871 |
| GO_NEGATIVE_REGULATION_OF_SMOOTH_MUSCLE_CELL_MIGRATION | 16 | -0.6387959 | -1.7647889 | 0.004987531 | 0.029622365 |
| GO_PITUITARY_GLAND_DEVELOPMENT | 22 | -0.58390665 | -1.7633216 | 0.002785515 | 0.029840246 |
| GO_RESPONSE_TO_MOLECULE_OF_BACTERIAL_ORIGIN | 221 | -0.38603643 | -1.763624 | 0 | 0.02984327 |
| GO_ANTIGEN_PROCESSING_AND_PRESENTATION_OF_EXOGENOUS_PEPTIDE_ANTIGEN_VIA_MHC_CLASS_I | 50 | 0.52725446 | 1.6789153 | 0.00143472 | 0.029896405 |
| GO_T_HELPER_1_TYPE_IMMUNE_RESPONSE | 25 | -0.5645089 | -1.7617337 | 0.002638523 | 0.029921882 |
| GO_MEDIATOR_COMPLEX | 16 | 0.6719817 | 1.6778175 | 0.006472492 | 0.029944796 |
| GO_CYTOKINE_METABOLIC_PROCESS | 72 | -0.45678592 | -1.7622155 | 0 | 0.029965607 |
| GO_CARDIAC_MUSCLE_CONTRACTION | 85 | -0.4400263 | -1.7613027 | 0 | 0.029972803 |
| GO_PEPTIDE_METABOLIC_PROCESS | 400 | 0.40905097 | 1.677944 | 0 | 0.029986354 |
| GO_MORPHOGENESIS_OF_A_BRANCHING_STRUCTURE | 116 | -0.4177339 | -1.7617762 | 0 | 0.03 |
| GO_COLLAGEN_TRIMER | 42 | -0.5032165 | -1.762402 | 0.003164557 | 0.03000709 |
| GO_REGULATION_OF_MYELOID_LEUKOCYTE_DIFFERENTIATION | 70 | -0.47103545 | -1.7627108 | 0 | 0.030010158 |
| GO_RESPONSE_TO_BMP | 89 | -0.44030973 | -1.7606487 | 0 | 0.030038683 |
| GO_SECOND_MESSENGER_MEDIATED_SIGNALING | 244 | -0.38218334 | -1.7597588 | 0 | 0.030074282 |
| GO_MATURATION_OF_SSU_RRNA | 22 | 0.61784464 | 1.6779442 | 0.008250825 | 0.030082772 |
| REACTOME_G2_M_CHECKPOINTS | 72 | 0.4915988 | 1.6780516 | 0 | 0.03015147 |
| GO_REGULATION_OF_MUSCLE_ORGAN_DEVELOPMENT | 72 | -0.44780296 | -1.7598088 | 0.003424658 | 0.03016241 |
| GO_POSITIVE_REGULATION_OF_FIBROBLAST_PROLIFERATION | 35 | -0.5390175 | -1.7586403 | 0.003095975 | 0.030280277 |
| GO_MOLTING_CYCLE | 49 | -0.49392253 | -1.7575315 | 0 | 0.03029523 |
| GO_RENAL_FILTRATION | 19 | -0.6201755 | -1.7576553 | 0.015228426 | 0.030356703 |
| GO_VOLTAGE_GATED_ION_CHANNEL_ACTIVITY | 84 | -0.43895695 | -1.7579602 | 0 | 0.030392766 |
| GO_FATTY_ACID_BETA_OXIDATION | 54 | 0.5168463 | 1.6744368 | 0.002902758 | 0.03050376 |
| GO_GLYCOSIDE_METABOLIC_PROCESS | 15 | 0.6768792 | 1.6755811 | 0.00990099 | 0.03055569 |
| REACTOME_MITOCHONDRIAL_PROTEIN_IMPORT | 31 | 0.57289016 | 1.6758617 | 0.004601227 | 0.030564379 |
| GO_FATTY_ACID_CATABOLIC_PROCESS | 73 | 0.48874378 | 1.6744772 | 0.001414427 | 0.03059511 |
| GO_PROTEASOMAL_PROTEIN_CATABOLIC_PROCESS | 239 | 0.42396462 | 1.6745126 | 0 | 0.030674249 |
| GO_NEUTROPHIL_MIGRATION | 75 | -0.4542426 | -1.7558483 | 0 | 0.030718217 |
| GO_NADH_DEHYDROGENASE_COMPLEX_ASSEMBLY | 31 | 0.5712025 | 1.674607 | 0.001519757 | 0.03073473 |
| GO_REGULATION_OF_TRANSLATIONAL_INITIATION | 43 | 0.53928065 | 1.674795 | 0.007407407 | 0.030743904 |
| GO_DOUBLE_STRAND_BREAK_REPAIR | 87 | 0.48159993 | 1.6733247 | 0.002666667 | 0.030759282 |
| GO_VESICLE_MEDIATED_TRANSPORT_BETWEEN_ENDOSOMAL_COMPARTMENTS | 26 | 0.5897212 | 1.6728303 | 0.007824726 | 0.030830257 |
| GO_BONE_MORPHOGENESIS | 66 | -0.4528096 | -1.7547951 | 0 | 0.030847425 |
| GO_MEMORY | 73 | -0.44716316 | -1.7537597 | 0 | 0.030866463 |
| GO_NEGATIVE_REGULATION_OF_PHOSPHORYLATION | 267 | -0.37877542 | -1.7535363 | 0 | 0.03087127 |
| GO_PROTEIN_SERINE_THREONINE_KINASE_INHIBITOR_ACTIVITY | 16 | -0.64215237 | -1.7550473 | 0.012886598 | 0.030873127 |
| GO_RENAL_SYSTEM_DEVELOPMENT | 192 | -0.39710936 | -1.7539439 | 0 | 0.030923529 |

| | | | | | |
|---|---|---|---|---|---|
| GO_DETECTION_OF_STIMULUS_INVOLVED_IN_SENSORY_PERCEPTION | 56 | -0.48023733 | -1.7541184 | 0 | 0.030945096 |
| GO_SMOOTH_MUSCLE_CONTRACTION | 67 | -0.4585582 | -1.7527465 | 0 | 0.03100385 |
| GO_G_PROTEIN_COUPLED_PURINERGIC_RECEPTOR_SIGNALING_PATHWAY | 15 | -0.6536939 | -1.7516528 | 0.004950495 | 0.03106785 |
| GO_SENSORY_PERCEPTION_OF_LIGHT_STIMULUS | 93 | -0.43470398 | -1.7519467 | 0 | 0.031068763 |
| GO_NUCLEOCYTOPLASMIC_CARRIER_ACTIVITY | 15 | 0.67641145 | 1.6717621 | 0.006745363 | 0.031108087 |
| GO_KIDNEY_MORPHOGENESIS | 57 | -0.4797531 | -1.752044 | 0 | 0.031125091 |
| GO_TRABECULA_FORMATION | 18 | -0.6147793 | -1.7507399 | 0.01010101 | 0.0311866 |
| GO_RESPONSE_TO_ANESTHETIC | 43 | -0.499221 | -1.7510076 | 0.002994012 | 0.031222926 |
| GO_CIRCULATORY_SYSTEM_PROCESS | 338 | -0.37081394 | -1.7500349 | 0 | 0.031310078 |
| PID_REG_GR_PATHWAY | 51 | -0.4870587 | -1.7484723 | 0.003215434 | 0.031519786 |
| GO_RESPONSE_TO_BACTERIUM | 367 | -0.365264 | -1.7486695 | 0 | 0.031552453 |
| REACTOME_RRNA_PROCESSING | 90 | 0.46818307 | 1.6696124 | 0.002628121 | 0.031646397 |
| GO_AZUROPHIL_GRANULE_LUMEN | 62 | 0.5063927 | 1.6696389 | 0.00141844 | 0.031739876 |
| GO_HEART_PROCESS | 170 | -0.40056807 | -1.7472305 | 0 | 0.03183508 |
| REACTOME_S_PHASE | 87 | 0.47733408 | 1.6680282 | 0 | 0.032127045 |
| GO_MEMBRANE_DOCKING | 89 | 0.4666947 | 1.6673409 | 0 | 0.0322632 |
| GO_GUANOSINE_CONTAINING_COMPOUND_METABOLIC_PROCESS | 23 | 0.60504 | 1.666644 | 0.006441224 | 0.032392517 |
| GO_REGULATION_OF_LEUKOCYTE_MIGRATION | 135 | -0.40508777 | -1.7427981 | 0 | 0.032439493 |
| GO_ASTROCYTE_DEVELOPMENT | 24 | -0.56323075 | -1.7442468 | 0.002857143 | 0.03247494 |
| REACTOME_ECM_PROTEOGLYCANS | 50 | -0.47999257 | -1.7440274 | 0 | 0.03248934 |
| GO_NEURON_RECOGNITION | 25 | -0.5541648 | -1.7428203 | 0.005277045 | 0.03252719 |
| GO_NEGATIVE_REGULATION_OF_TUMOR_NECROSIS_FACTOR_SUPERFAMILY_CYTOKINE_PRODUCTION | 36 | -0.53976417 | -1.7443633 | 0.005934718 | 0.032529287 |
| GO_PRI_MIRNA_TRANSCRIPTION_BY_RNA_POLYMERASE_II | 28 | -0.55279505 | -1.7430238 | 0.008219178 | 0.03254704 |
| GO_POSITIVE_REGULATION_OF_FAT_CELL_DIFFERENTIATION | 32 | -0.5337456 | -1.7430897 | 0.00591716 | 0.032605678 |
| GO_FAT_CELL_DIFFERENTIATION | 121 | -0.41055933 | -1.743288 | 0 | 0.032645397 |
| SIG_CD40PATHWAYMAP | 22 | -0.5725623 | -1.7414746 | 0.00802139 | 0.03276378 |
| GO_DNA_METABOLIC_PROCESS | 405 | 0.40326053 | 1.6651647 | 0 | 0.03286281 |
| GO_EPITHELIAL_CELL_APOPTOTIC_PROCESS | 64 | -0.45161965 | -1.7390356 | 0 | 0.03335047 |
| REACTOME_INTERLEUKIN_10_SIGNALING | 35 | -0.54620475 | -1.7391528 | 0.005714286 | 0.033405893 |
| GO_RESPONSE_TO_CAMP | 69 | -0.4585644 | -1.7382705 | 0 | 0.033539023 |
| PID_IL12_2PATHWAY | 44 | -0.4944826 | -1.7372576 | 0.002881844 | 0.033822622 |
| GO_MITOTIC_SISTER_CHROMATID_SEGREGATION | 64 | 0.4983752 | 1.6618831 | 0.004178273 | 0.033933464 |
| REACTOME_SUMOYLATION_OF_DNA_DAMAGE_RESPONSE_AND_REPAIR_PROTEINS | 41 | 0.5442355 | 1.6615505 | 0.007541478 | 0.033957843 |
| GO_REGULATION_OF_PROTEIN_SERINE_THREONINE_KINASE_ACTIVITY | 295 | -0.36779857 | -1.7364181 | 0 | 0.034019828 |
| GO_ORGAN_GROWTH | 109 | -0.4189833 | -1.7350348 | 0 | 0.03433338 |
| GO_REGULATION_OF_BICELLULAR_TIGHT_JUNCTION_ASSEMBLY | 16 | -0.6229915 | -1.7346859 | 0.012690355 | 0.03433791 |
| GO_REGULATION_OF_ALPHA_BETA_T_CELL_DIFFERENTIATION | 40 | -0.5049582 | -1.7335781 | 0 | 0.034610845 |
| GO_ENTEROENDOCRINE_CELL_DIFFERENTIATION | 15 | -0.63641626 | -1.7320964 | 0.005 | 0.03502888 |
| GO_NEGATIVE_REGULATION_OF_CYTOKINE_BIOSYNTHETIC_PROCESS | 20 | -0.58978933 | -1.7315772 | 0.010723861 | 0.03509294 |
| GO_RIBONUCLEOPROTEIN_COMPLEX_SUBUNIT_ORGANIZATION | 87 | 0.47727302 | 1.6580384 | 0.002670227 | 0.03529794 |

| | | | | | |
|---|---|---|---|---|---|
| GO_NEGATIVE_REGULATION_OF_GENE_EXPRESSION_EPIGENETIC | 34 | 0.56166995 | 1.6569378 | 0.004573171 | 0.035508763 |
| GO_UBIQUITIN_LIKE_PROTEIN_CONJUGATING_ENZYME_ACTIVITY | 24 | 0.5873507 | 1.6570711 | 0.012882448 | 0.03556648 |
| GO_PROTEIN_N_LINKED_GLYCOSYLATION | 51 | 0.5146225 | 1.6562337 | 0.002919708 | 0.035567794 |
| REACTOME_NEDDYLATION | 104 | 0.4600334 | 1.6562383 | 0.004 | 0.035673965 |
| KEGG_INTESTINAL_IMMUNE_NETWORK_FOR_IGA_PRODUCTION | 26 | -0.5595598 | -1.7290797 | 0.005617978 | 0.035690654 |
| REACTOME_DNA_DAMAGE_BYPASS | 28 | 0.57940567 | 1.6548383 | 0.007633588 | 0.0357311 |
| REACTOME_AMINE_LIGAND_BINDING_RECEPTORS | 20 | -0.6121406 | -1.7291155 | 0.007894737 | 0.035788435 |
| GO_SEMI_LUNAR_VALVE_DEVELOPMENT | 25 | -0.5629354 | -1.7294115 | 0 | 0.035792895 |
| REACTOME_METABOLISM_OF_COFACTORS | 15 | 0.6598722 | 1.6549028 | 0.009966778 | 0.035812553 |
| REACTOME_TELOMERE_MAINTENANCE | 26 | 0.58298635 | 1.6551273 | 0.008090615 | 0.03581821 |
| GO_PROTEIN_TARGETING | 219 | 0.42377824 | 1.6551858 | 0.001179245 | 0.035890426 |
| GO_MANGANESE_ION_BINDING | 38 | 0.5334284 | 1.6538124 | 0.007716049 | 0.036020365 |
| GO_CELLULAR_COMPONENT_ASSEMBLY_INVOLVED_IN_MORPHOGENESIS | 57 | -0.47145438 | -1.727248 | 0 | 0.036120582 |
| GO_ORGANELLE_MEMBRANE_FUSION | 49 | 0.51030856 | 1.6525397 | 0.004366812 | 0.036161184 |
| GO_TRANSFERASE_ACTIVITY_TRANSFERRING_ACYL_GROUPS | 118 | 0.4486472 | 1.6530352 | 0 | 0.036208477 |
| GO_MITOCHONDRIAL_LARGE_RIBOSOMAL_SUBUNIT | 29 | 0.57290787 | 1.652614 | 0.009448819 | 0.036242947 |
| REACTOME_CHROMOSOME_MAINTENANCE | 38 | 0.54341406 | 1.650435 | 0.005970149 | 0.036302812 |
| REACTOME_UCH_PROTEINASES | 40 | 0.5341103 | 1.6506654 | 0.001470588 | 0.03632741 |
| REACTOME_ASPARAGINE_N_LINKED_GLYCOSYLATION | 199 | 0.4252523 | 1.6516621 | 0 | 0.03632815 |
| REACTOME_NUCLEAR_ENVELOPE_NE_REASSEMBLY | 50 | 0.51156485 | 1.6500198 | 0.00443787 | 0.03635972 |
| REACTOME_NEGATIVE_EPIGENETIC_REGULATION_OF_RRNA_EXPRESSION | 29 | 0.5583069 | 1.6507515 | 0.010920437 | 0.036396537 |
| REACTOME_RNA_POLYMERASE_III_TRANSCRIPTION | 22 | 0.6059822 | 1.6517017 | 0.0208 | 0.03641774 |
| GO_QUINONE_METABOLIC_PROCESS | 23 | 0.59624076 | 1.6511239 | 0.004807693 | 0.036436267 |
| REACTOME_RUNX1_REGULATES_TRANSCRIPTION_OF_GENES_INVOLVED_IN_DIFFERENTIATION_OF_HSCS | 39 | 0.53789914 | 1.650828 | 0.007692308 | 0.03646605 |
| GO_NEGATIVE_REGULATION_OF_LEUKOCYTE_CELL_CELL_ADHESION | 77 | -0.44697788 | -1.7238559 | 0.003802281 | 0.03651708 |
| GO_PANCREAS_DEVELOPMENT | 42 | -0.4922214 | -1.7258773 | 0 | 0.036524337 |
| GO_REGULATION_OF_LEUKOCYTE_CHEMOTAXIS | 79 | -0.43387136 | -1.7231135 | 0 | 0.036524486 |
| GO_POTASSIUM_CHANNEL_ACTIVITY | 51 | -0.4756045 | -1.7241064 | 0 | 0.036527604 |
| GO_GLANDULAR_EPITHELIAL_CELL_DIFFERENTIATION | 24 | -0.55872196 | -1.7231636 | 0.008219178 | 0.036581066 |
| GO_INTERLEUKIN_4_PRODUCTION | 21 | -0.5794388 | -1.7226287 | 0.016393442 | 0.03659454 |
| GO_NEPHRON_MORPHOGENESIS | 46 | -0.48148817 | -1.7244056 | 0 | 0.036595732 |
| GO_CELL_SUBSTRATE_ADHESION | 208 | -0.38502866 | -1.7241174 | 0 | 0.03662553 |
| GO_CHONDROITIN_SULFATE_BIOSYNTHETIC_PROCESS | 16 | -0.62485456 | -1.7232939 | 0.002450981 | 0.0366511 |
| GO_BASEMENT_MEMBRANE | 55 | -0.4685168 | -1.7244879 | 0.006779661 | 0.036653187 |
| REACTOME_NUCLEAR_ENVELOPE_BREAKDOWN | 36 | 0.55797815 | 1.6487024 | 0.010719755 | 0.036653742 |
| GO_CHONDROITIN_SULFATE_PROTEOGLYCAN_BIOSYNTHETIC_PROCESS | 18 | -0.6098977 | -1.725111 | 0.01058201 | 0.036656864 |
| GO_SPLICEOSOMAL_TRI_SNRNP_COMPLEX | 19 | 0.6227202 | 1.6489797 | 0.011784512 | 0.03665953 |
| PID_MAPK_TRK_PATHWAY | 21 | -0.57001966 | -1.7246909 | 0.0025 | 0.03669228 |
| GO_CELLULAR_PROTEIN_COMPLEX_DISASSEMBLY | 116 | 0.45332843 | 1.6480985 | 0.001321004 | 0.03670284 |
| GO_INTRACELLULAR_PROTEIN_TRANSMEMBRANE_TRANSPORT | 25 | 0.59414005 | 1.6483268 | 0.011217949 | 0.036708925 |
| GO_VACUOLAR_MEMBRANE | 234 | 0.41724473 | 1.6476147 | 0 | 0.03680121 |
| GO_ORGANELLE_INNER_MEMBRANE | 250 | 0.4175828 | 1.6472995 | 0 | 0.03683462 |

| | | | | | |
|---|---|---|---|---|---|
| GO_MYELOID_LEUKOCYTE_MIGRATION | 131 | -0.40559906 | -1.7215515 | 0 | 0.036838822 |
| GO_REGULATION_OF_MUSCLE_SYSTEM_PROCESS | 152 | -0.3962365 | -1.7204381 | 0 | 0.036963314 |
| GO_CCR_CHEMOKINE_RECEPTOR_BINDING | 24 | -0.5680323 | -1.7207464 | 0.01396648 | 0.036970917 |
| GO_POSITIVE_REGULATION_OF_MAPK_CASCADE | 297 | -0.36570957 | -1.7200342 | 0 | 0.036996327 |
| GO_REGULATION_OF_HEART_MORPHOGENESIS | 24 | -0.56889194 | -1.7185723 | 0.01591512 | 0.03728948 |
| GO_LYMPHOCYTE_ACTIVATION | 410 | -0.3543713 | -1.7182013 | 0 | 0.037294623 |
| GO_COLUMNAR_CUBOIDAL_EPITHELIAL_CELL_DIFFERENTIATION | 60 | -0.46169704 | -1.7178184 | 0 | 0.037295256 |
| GO_REGULATION_OF_ANIMAL_ORGAN_FORMATION | 25 | -0.57145107 | -1.7187071 | 0 | 0.037306927 |
| GO_CHONDROCYTE_DIFFERENTIATION | 63 | -0.45931417 | -1.716579 | 0.003236246 | 0.03756365 |
| GO_INTRACILIARY_TRANSPORT_INVOLVED_IN_CILIUM_ASSEMBLY | 22 | 0.6056719 | 1.6451023 | 0.014516129 | 0.037596088 |
| GO_INFLAMMATORY_RESPONSE | 447 | -0.35529944 | -1.7161107 | 0 | 0.037682038 |
| GO_POSITIVE_CHEMOTAXIS | 38 | -0.49711978 | -1.7150455 | 0 | 0.037846692 |
| GO_CARDIAC_CONDUCTION | 83 | -0.43387127 | -1.7153053 | 0 | 0.03784765 |
| GO_RNA_CATABOLIC_PROCESS | 183 | 0.42564896 | 1.6437018 | 0 | 0.038043004 |
| GO_HEAD_MORPHOGENESIS | 19 | -0.6098493 | -1.7138602 | 0.015503876 | 0.038044844 |
| GO_MESENCHYMAL_CELL_DIFFERENTIATION | 112 | -0.41316143 | -1.7141384 | 0 | 0.038085118 |
| GO_RIBONUCLEOPROTEIN_COMPLEX_BINDING | 76 | 0.4858884 | 1.64381 | 0.002754821 | 0.038103454 |
| REACTOME_SMOOTH_MUSCLE_CONTRACTION | 30 | -0.53979945 | -1.7130711 | 0.008174387 | 0.038186617 |
| GO_LIGASE_ACTIVITY_FORMING_CARBON_SULFUR_BONDS | 25 | 0.59137684 | 1.6425397 | 0.00952381 | 0.03822682 |
| GO_NUCLEOSIDE_MONOPHOSPHATE_METABOLIC_PROCESS | 44 | 0.5232402 | 1.6422558 | 0.004360465 | 0.03824087 |
| GO_OXIDOREDUCTASE_ACTIVITY | 455 | 0.39617926 | 1.642727 | 0 | 0.038244344 |
| GO_POSITIVE_REGULATION_OF_VASCULAR_ENDOTHELIAL_GROWTH_FACTOR_PRODUCTION | 16 | -0.6323935 | -1.7125219 | 0.01 | 0.038253203 |
| REACTOME_PROCESSING_OF_DNA_DOUBLE_STRAND_BREAK_ENDS | 31 | 0.5531371 | 1.6429216 | 0.013392857 | 0.038273364 |
| GO_SENSORY_PERCEPTION_OF_TASTE | 24 | -0.56796354 | -1.7119699 | 0.007978723 | 0.03834193 |
| GO_POSITIVE_REGULATION_OF_MAP_KINASE_ACTIVITY | 149 | -0.3928335 | -1.7106222 | 0 | 0.03868303 |
| REACTOME_DEGRADATION_OF_DVL | 32 | 0.5578569 | 1.6408176 | 0.010606061 | 0.038715802 |
| GO_CHROMOSOME_ORGANIZATION_INVOLVED_IN_MEIOTIC_CELL_CYCLE | 20 | 0.6147816 | 1.640378 | 0.014950166 | 0.03878959 |
| HALLMARK_E2F_TARGETS | 134 | 0.44227746 | 1.6399761 | 0 | 0.038864706 |
| GO_SNRNA_TRANSCRIPTION | 36 | 0.54343146 | 1.6394002 | 0.009463723 | 0.0390108 |
| GO_MYOBLAST_FUSION | 24 | -0.57257783 | -1.7090374 | 0.007894737 | 0.039104592 |
| GO_RESPIRATORY_SYSTEM_DEVELOPMENT | 122 | -0.3993058 | -1.7091382 | 0 | 0.039177164 |
| GO_NUCLEIC_ACID_PHOSPHODIESTER_BOND_HYDROLYSIS | 133 | 0.44335946 | 1.6380949 | 0.001267427 | 0.039448947 |
| GO_PEPTIDYL_ASPARAGINE_MODIFICATION | 24 | 0.5816216 | 1.6378071 | 0.009661836 | 0.039455645 |
| GO_MESODERM_MORPHOGENESIS | 38 | -0.50111896 | -1.7074374 | 0.005988024 | 0.039510384 |
| GO_NEGATIVE_REGULATION_OF_HEMOPOIESIS | 91 | -0.4306432 | -1.7068424 | 0 | 0.03960018 |
| GO_NEGATIVE_REGULATION_OF_GROWTH | 147 | -0.3911918 | -1.7066274 | 0 | 0.039616 |
| REACTOME_TOLL_LIKE_RECEPTOR_10_TLR10_CASCADE | 39 | -0.4903329 | -1.7058451 | 0.003367003 | 0.039794665 |
| GO_NUCLEAR_ENVELOPE | 256 | 0.40986925 | 1.636619 | 0 | 0.039859854 |
| REACTOME_NR1H2_AND_NR1H3_MEDIATED_SIGNALING | 25 | 0.5856071 | 1.6359246 | 0.009771987 | 0.039947852 |
| GO_T_CELL_ACTIVATION | 290 | -0.3698367 | -1.7050588 | 0 | 0.039967105 |
| KEGG_VASCULAR_SMOOTH_MUSCLE_CONTRACTION | 72 | -0.43768835 | -1.7047213 | 0 | 0.04000418 |
| KEGG_BUTANOATE_METABOLISM | 29 | 0.566587 | 1.6360013 | 0.009569378 | 0.040020414 |
| GO_BASAL_TRANSCRIPTION_MACHINERY_BINDING | 32 | 0.54595715 | 1.635287 | 0.006279435 | 0.040128622 |
| REACTOME_METABOLISM_OF_NUCLEOTIDES | 67 | 0.48767215 | 1.6347169 | 0.002820875 | 0.040142138 |

| | | | | | |
|---|---|---|---|---|---|
| REACTOME_EXTENSION_OF_TELOMERES | 26 | 0.58298635 | 1.6347647 | 0.006578947 | 0.0402327 |
| GO_NEGATIVE_REGULATION_OF_DEVELOPMENTAL_GROWTH | 61 | -0.45859134 | -1.7021245 | 0 | 0.040371347 |
| GO_RIBOSOME_BINDING | 31 | 0.5546329 | 1.632818 | 0.012288787 | 0.040412486 |
| GO_REGULATION_OF_MEMBRANE_REPOLARIZATION | 21 | -0.58550787 | -1.7016048 | 0.012919896 | 0.040456507 |
| REACTOME_COMPLEX_I_BIOGENESIS | 30 | 0.56390494 | 1.633764 | 0.012658228 | 0.040467978 |
| GO_REGULATION_OF_INTERLEUKIN_4_PRODUCTION | 18 | -0.62178004 | -1.7021346 | 0.012690355 | 0.040470053 |
| GO_CELL_JUNCTION_ASSEMBLY | 155 | -0.3880258 | -1.702502 | 0 | 0.040473435 |
| GO_SKELETAL_SYSTEM_MORPHOGENESIS | 135 | -0.3955634 | -1.7030237 | 0 | 0.040478833 |
| GO_NON_MOTILE_CILIUM_ASSEMBLY | 21 | 0.5997823 | 1.6328603 | 0.004830918 | 0.040495157 |
| GO_POSITIVE_REGULATION_OF_CD4_POSITIVE_ALPHA_BETA_T_CELL_ACTIVATION | 26 | -0.539736 | -1.702602 | 0.005464481 | 0.040530432 |
| PID_ERBB1_RECEPTOR_PROXIMAL_PATHWAY | 24 | 0.57687443 | 1.6329135 | 0.0192 | 0.04057468 |
| GO_INNER_MITOCHONDRIAL_MEMBRANE_PROTEIN_COMPLEX | 51 | 0.51332086 | 1.633102 | 0.001477105 | 0.040583167 |
| GO_RNA_3_END_PROCESSING | 67 | 0.49066547 | 1.6332628 | 0.005617978 | 0.040602777 |
| GO_BLOOD_VESSEL_MORPHOGENESIS | 392 | -0.3514186 | -1.7008595 | 0 | 0.04064394 |
| GO_MUSCLE_STRUCTURE_DEVELOPMENT | 340 | -0.35770923 | -1.7005666 | 0 | 0.040677402 |
| GO_BRANCHING_MORPHOGENESIS_OF_AN_EPITHELIAL_TUBE | 90 | -0.4232671 | -1.6998004 | 0.003875969 | 0.040781867 |
| GO_AUTONOMIC_NERVOUS_SYSTEM_DEVELOPMENT | 22 | -0.56357485 | -1.6998339 | 0.010526316 | 0.04085586 |
| GO_REGULATION_OF_METANEPHROS_DEVELOPMENT | 17 | -0.6053672 | -1.6983639 | 0.010416667 | 0.04088371 |
| GO_CONNECTIVE_TISSUE_DEVELOPMENT | 152 | -0.38680217 | -1.6985917 | 0 | 0.040928993 |
| GO_LYMPHOCYTE_CHEMOTAXIS | 38 | -0.49954575 | -1.698774 | 0.006230529 | 0.040974207 |
| GO_NEUROPEPTIDE_RECEPTOR_BINDING | 15 | -0.6491911 | -1.6987972 | 0.015463918 | 0.041052368 |
| REACTOME_RECOGNITION_OF_DNA_DAMAGE_BY_PCNA_CONTAINING_REPLICATION_COMPLEX | 19 | 0.61787754 | 1.630403 | 0.009917355 | 0.041327152 |
| GO_RNA_HELICASE_ACTIVITY | 34 | 0.55982214 | 1.6289477 | 0.003039514 | 0.041847114 |
| GO_CELL_JUNCTION_ORGANIZATION | 179 | -0.38560086 | -1.6951765 | 0 | 0.041847307 |
| GO_SEQUENCE_SPECIFIC_DOUBLE_STRANDED_DNA_BINDING | 431 | -0.34424165 | -1.6954559 | 0 | 0.041853327 |
| KEGG_NOD_LIKE_RECEPTOR_SIGNALING_PATHWAY | 39 | -0.4862451 | -1.6942086 | 0.005865103 | 0.042128786 |
| GO_MRNA_EXPORT_FROM_NUCLEUS | 59 | 0.49619472 | 1.6280223 | 0.005822416 | 0.04215793 |
| GO_POTASSIUM_ION_IMPORT_ACROSS_PLASMA_MEMBRANE | 30 | -0.5242157 | -1.6935912 | 0.00621118 | 0.04222234 |
| GO_CELLULAR_RESPONSE_TO_IONIZING_RADIATION | 35 | -0.5119044 | -1.6932251 | 0.002949853 | 0.042255484 |
| REACTOME_SUMOYLATION_OF_RNA_BINDING_PROTEINS | 27 | 0.5724405 | 1.6267048 | 0.006359301 | 0.042625483 |
| GO_NEGATIVE_REGULATION_OF_PHOSPHORUS_METABOLIC_PROCESS | 319 | -0.35716823 | -1.6915123 | 0 | 0.04265337 |
| GO_GOLGI_TO_PLASMA_MEMBRANE_TRANSPORT | 31 | 0.56839967 | 1.626088 | 0.010989011 | 0.042678103 |
| GO_POSITIVE_REGULATION_OF_TYROSINE_PHOSPHORYLATION_OF_STAT_PROTEIN | 40 | -0.501054 | -1.691524 | 0.003355705 | 0.042749684 |
| GO_POSITIVE_REGULATION_OF_HEART_GROWTH | 25 | -0.56134313 | -1.6904899 | 0.002590674 | 0.042772785 |
| GO_ORGANELLE_LOCALIZATION | 336 | 0.40205908 | 1.6260997 | 0 | 0.042782858 |
| GO_SPECIFICATION_OF_ANIMAL_ORGAN_IDENTITY | 21 | -0.5700414 | -1.6907277 | 0.005434783 | 0.042785015 |
| GO_POSITIVE_REGULATION_OF_ANIMAL_ORGAN_MORPHOGENESIS | 52 | -0.46709442 | -1.69092 | 0 | 0.042785045 |
| GO_REGULATION_OF_RENAL_SYSTEM_PROCESS | 26 | -0.54565597 | -1.6893721 | 0.007792208 | 0.043020386 |
| GO_CARDIAC_MUSCLE_CELL_MEMBRANE_REPOLARIZATION | 22 | -0.5591016 | -1.6890085 | 0.013661202 | 0.04304742 |
| GO_MACROPHAGE_ACTIVATION | 66 | -0.45352718 | -1.6895232 | 0.003636364 | 0.043061256 |

| | | | | | |
|---|---|---|---|---|---|
| GO_RIBOSOMAL_SUBUNIT | 79 | 0.47609186 | 1.6228224 | 0 | 0.043927945 |
| GO_REGULATION_OF_EPITHELIAL_CELL_APOPTOTIC_PROCESS | 55 | -0.46358165 | -1.6862904 | 0 | 0.043983646 |
| GO_DEOXYRIBONUCLEOTIDE_METABOLIC_PROCESS | 24 | 0.57239246 | 1.6228902 | 0.007898894 | 0.044006 |
| GO_BODY_MORPHOGENESIS | 25 | -0.5451487 | -1.6856326 | 0.011627907 | 0.04412155 |
| GO_ADRENERGIC_RECEPTOR_SIGNALING_PATHWAY | 16 | -0.62010306 | -1.6852895 | 0.002557545 | 0.044152774 |
| GO_CELLULAR_AMINO_ACID_BIOSYNTHETIC_PROCESS | 53 | 0.50535625 | 1.6216469 | 0.011494253 | 0.044354986 |
| REACTOME_REGULATION_OF_RUNX3_EXPRESSION_AND_ACTIVITY | 29 | 0.5554836 | 1.6211773 | 0.004587156 | 0.04449213 |
| REACTOME_HEDGEHOG_LIGAND_BIOGENESIS | 34 | 0.5465829 | 1.6207196 | 0.006015038 | 0.044551186 |
| HALLMARK_INTERFERON_ALPHA_RESPONSE | 73 | 0.47321078 | 1.6204228 | 0.001367989 | 0.044563543 |
| GO_LEUKOCYTE_APOPTOTIC_PROCESS | 73 | -0.43958196 | -1.6836828 | 0 | 0.044671733 |
| GO_ENDOTHELIAL_CELL_APOPTOTIC_PROCESS | 38 | -0.48821852 | -1.6823144 | 0.009146341 | 0.04472104 |
| GO_NEURON_PROJECTION_GUIDANCE | 146 | -0.3967125 | -1.6824081 | 0 | 0.044783503 |
| GO_METANEPHROS_MORPHOGENESIS | 20 | -0.5764129 | -1.6825473 | 0.005128205 | 0.04482264 |
| GO_NEGATIVE_REGULATION_OF_T_CELL_PROLIFERATION | 38 | -0.49174917 | -1.6827611 | 0.003058104 | 0.044831336 |
| REACTOME_ADORA2B_MEDIATED_ANTI_INFLAMMATORY_CYTOKINES_PRODUCTION | 62 | -0.4519837 | -1.6830076 | 0 | 0.04483169 |
| GO_REGULATION_OF_RECEPTOR_SIGNALING_PATHWAY_VIA_STAT | 77 | -0.42833903 | -1.6814501 | 0 | 0.044942178 |
| GO_ALPHA_AMINO_ACID_BIOSYNTHETIC_PROCESS | 44 | 0.511145 | 1.6190175 | 0.007836991 | 0.04504631 |
| GO_UROGENITAL_SYSTEM_DEVELOPMENT | 216 | -0.36623588 | -1.6806185 | 0 | 0.04520438 |
| GO_HELICASE_ACTIVITY | 77 | 0.46898264 | 1.6180637 | 0.002857143 | 0.045371465 |
| GO_NEGATIVE_REGULATION_OF_AXON_GUIDANCE | 18 | -0.5985322 | -1.6798434 | 0.007614213 | 0.045375615 |
| GO_CELL_CELL_ADHESION_VIA_PLASMA_MEMBRANE_ADHESION_MOLECULES | 124 | -0.40423733 | -1.6789694 | 0 | 0.045426138 |
| GO_NEGATIVE_REGULATION_OF_GLIOGENESIS | 24 | -0.55691814 | -1.6786952 | 0.008356546 | 0.045446556 |
| REACTOME_CELL_JUNCTION_ORGANIZATION | 59 | -0.44423696 | -1.6793426 | 0 | 0.045469113 |
| GO_MULTICELLULAR_ORGANISMAL_SIGNALING | 117 | -0.40546662 | -1.6790658 | 0 | 0.045474343 |
| GO_SKELETAL_MUSCLE_CONTRACTION | 26 | -0.54511 | -1.6781546 | 0.002832861 | 0.045557935 |
| REACTOME_RESPIRATORY_ELECTRON_TRANSPORT | 54 | 0.49279127 | 1.616998 | 0.00913242 | 0.045778822 |
| REACTOME_G_ALPHA_Q_SIGNALLING_EVENTS | 109 | -0.40770492 | -1.6770632 | 0 | 0.045784276 |
| GO_ENDOCHONDRAL_BONE_MORPHOGENESIS | 43 | -0.4903062 | -1.6772543 | 0.009202454 | 0.04581029 |
| GO_CYTOSOLIC_TRANSPORT | 82 | 0.46470818 | 1.6161895 | 0.004032258 | 0.046020523 |
| GO_DNA_BINDING_TRANSCRIPTION_FACTOR_ACTIVITY_RNA_POLYMERASE_II_SPECIFIC | 478 | -0.3438494 | -1.6761706 | 0 | 0.046037327 |
| GO_PROTEIN_HETEROTETRAMERIZATION | 17 | 0.6312713 | 1.6157919 | 0.013422819 | 0.04606789 |
| GO_REGULATION_OF_NEUROINFLAMMATORY_RESPONSE | 22 | -0.567762 | -1.6758226 | 0.01861702 | 0.046123832 |
| GO_REGULATION_OF_LIPOPOLYSACCHARIDE_MEDIATED_SIGNALING_PATHWAY | 19 | -0.5813051 | -1.6748428 | 0.014634146 | 0.04634986 |
| GO_NEGATIVE_REGULATION_OF_MYELOID_CELL_DIFFERENTIATION | 52 | -0.47017822 | -1.6744355 | 0.003311258 | 0.046371695 |
| GO_ENDOCARDIAL_CUSHION_MORPHOGENESIS | 23 | -0.55202675 | -1.67417 | 0.016574586 | 0.046381958 |
| HALLMARK_CHOLESTEROL_HOMEOSTASIS | 50 | -0.46369317 | -1.6739327 | 0.00308642 | 0.046389073 |
| GO_TRANSCRIPTION_INITIATION_FROM_RNA_POLYMERASE_I_PROMOTER | 20 | 0.61427253 | 1.6146431 | 0.020477816 | 0.04640331 |
| KEGG_PORPHYRIN_AND_CHLOROPHYLL_METABOLISM | 17 | 0.6357729 | 1.6148168 | 0.011456628 | 0.046422735 |
| GO_REGULATION_OF_SPINDLE_ORGANIZATION | 25 | 0.57300967 | 1.6142242 | 0.012882448 | 0.046488248 |
| GO_NEGATIVE_REGULATION_OF_CHROMOSOME_ORGANIZATION | 62 | 0.4824131 | 1.6131824 | 0 | 0.04656852 |

| | | | | | |
|---|---|---|---|---|---|
| GO_RAN_GTPASE_BINDING | 17 | 0.6290839 | 1.6138018 | 0.021666666 | 0.04658099 |
| REACTOME_REGULATION_OF_HSF1_MEDIATED_HEAT_SHOCK_RESPONSE | 57 | 0.48825878 | 1.613381 | 0.006934813 | 0.046584796 |
| GO_ASPARTATE_FAMILY_AMINO_ACID_METABOLIC_PROCESS | 34 | 0.53597116 | 1.6134461 | 0.007763975 | 0.04664801 |
| GO_HETEROTYPIC_CELL_CELL_ADHESION | 43 | -0.4821646 | -1.6727667 | 0 | 0.046720847 |
| GO_SUBSTRATE_ADHESION_DEPENDENT_CELL_SPREADING | 68 | -0.43924853 | -1.6719382 | 0.003144654 | 0.046988167 |
| GO_ESTABLISHMENT_OF_RNA_LOCALIZATION | 105 | 0.44669238 | 1.6120319 | 0.001356852 | 0.04701086 |
| GO_POSITIVE_REGULATION_OF_CHEMOTAXIS | 90 | -0.4147404 | -1.6711589 | 0 | 0.047065448 |
| GO_POSITIVE_REGULATION_OF_KIDNEY_DEVELOPMENT | 30 | -0.528964 | -1.6713843 | 0.006134969 | 0.047089756 |
| GO_MITOTIC_CYTOKINESIS | 38 | 0.52841234 | 1.6106144 | 0.013910355 | 0.047342148 |
| GO_REGULATION_OF_CELL_CYCLE_G2_M_PHASE_TRANSITION | 105 | 0.44654334 | 1.6100496 | 0 | 0.04741292 |
| GO_DIGESTIVE_SYSTEM_DEVELOPMENT | 86 | -0.41910017 | -1.6696386 | 0 | 0.047438532 |
| PID_LYSOPHOSPHOLIPID_PATHWAY | 43 | -0.46918046 | -1.6691904 | 0.003125 | 0.04745488 |
| GO_AMIDE_BIOSYNTHETIC_PROCESS | 389 | 0.3932275 | 1.6106286 | 0 | 0.04745599 |
| REACTOME_CILIUM_ASSEMBLY | 102 | 0.44904777 | 1.6107348 | 0.001347709 | 0.04750568 |
| GO_METAPHASE_PLATE_CONGRESSION | 30 | 0.5463968 | 1.6101012 | 0.009360374 | 0.04750666 |
| GO_POSITIVE_REGULATION_OF_MYOTUBE_DIFFERENTIATION | 18 | -0.592927 | -1.6697202 | 0.015075377 | 0.04751528 |
| GO_NEGATIVE_REGULATION_OF_CELL_ADHESION | 182 | -0.37749168 | -1.6692724 | 0 | 0.04752758 |
| GO_INTRACILIARY_TRANSPORT | 24 | 0.58976924 | 1.6083896 | 0.018604651 | 0.047560465 |
| GO_TELOMERE_CAPPING | 19 | 0.60607535 | 1.608753 | 0.009615385 | 0.047619846 |
| GO_DOUBLE_STRANDED_RNA_BINDING | 45 | 0.5042573 | 1.6090053 | 0.009022556 | 0.04764142 |
| GO_RNA_PHOSPHODIESTER_BOND_HYDROLYSIS | 69 | 0.47266838 | 1.6084313 | 0.002770083 | 0.047648486 |
| GO_SNARE_COMPLEX | 23 | 0.60026205 | 1.6092358 | 0.011164274 | 0.04765993 |
| REACTOME_HDR_THROUGH_SINGLE_STRAND_ANNEALING_SSA | 20 | 0.6072416 | 1.6078569 | 0.01433121 | 0.04767818 |
| GO_METAL_ION_TRANSMEMBRANE_TRANSPORTER_ACTIVITY | 225 | -0.3610945 | -1.6670237 | 0 | 0.04789749 |
| GO_MUSCLE_ORGAN_MORPHOGENESIS | 47 | -0.46528578 | -1.6671084 | 0.003184713 | 0.04795624 |
| GO_RESPONSE_TO_GONADOTROPIN | 22 | -0.55428934 | -1.6660953 | 0.017721519 | 0.04800062 |
| GO_IMMUNE_RECEPTOR_ACTIVITY | 87 | -0.41989675 | -1.6674908 | 0.003921569 | 0.048008516 |
| REACTOME_DISEASES_ASSOCIATED_WITH_GLYCOSAMINOGLYCAN_METABOLISM | 25 | -0.5375361 | -1.666337 | 0.005763689 | 0.048009276 |
| GO_FORMATION_OF_PRIMARY_GERM_LAYER | 67 | -0.43423527 | -1.6672386 | 0 | 0.04802592 |
| REACTOME_NCAM1_INTERACTIONS | 27 | -0.5375299 | -1.6663513 | 0.013812155 | 0.048104107 |
| GO_LIVER_MORPHOGENESIS | 15 | -0.63170457 | -1.6644256 | 0.007518797 | 0.04858731 |
| GO_CELL_DIVISION | 269 | 0.402619 | 1.6053486 | 0 | 0.048705876 |
| GO_POSITIVE_REGULATION_OF_TELOMERE_MAINTENANCE_VIA_TELOMERE_LENGTHENING | 19 | 0.6211174 | 1.6055014 | 0.019769358 | 0.04874604 |
| GO_DNA_REPLICATION_INDEPENDENT_NUCLEOSOME_ORGANIZATION | 21 | 0.59774256 | 1.6046218 | 0.014950166 | 0.04889883 |
| KEGG_LINOLEIC_ACID_METABOLISM | 18 | -0.588301 | -1.6635078 | 0.013661202 | 0.048898913 |
| GO_POSITIVE_REGULATION_OF_STRIATED_MUSCLE_CELL_DIFFERENTIATION | 29 | -0.5230452 | -1.6627986 | 0.002688172 | 0.049134113 |
| GO_PRERIBOSOME_LARGE_SUBUNIT_PRECURSOR | 16 | 0.6288377 | 1.6036634 | 0.024 | 0.04913453 |
| GO_DNA_DEPENDENT_ATPASE_ACTIVITY | 58 | 0.48984092 | 1.6037953 | 0.002902758 | 0.049199477 |
| GO_RESPIRATORY_ELECTRON_TRANSPORT_CHAIN | 57 | 0.48495877 | 1.6031147 | 0.004267425 | 0.049318895 |
| GO_CELL_FATE_SPECIFICATION | 41 | -0.4760854 | -1.6609482 | 0.00295858 | 0.049379833 |

| | | | | | |
|---|---|---|---|---|---|
| GO_NEGATIVE_REGULATION_OF_IMMUNE_SYSTEM_PROCESS | 288 | -0.35462293 | -1.6619345 | 0 | 0.049404237 |
| REACTOME_BIOSYNTHESIS_OF_THE_N_GLYCAN_PRECURSOR_DOLICHOL_LIPID_LINKED_OLIGOSACCHARIDE_LLO_AND_TRANSFER_TO_A_NASCENT_PROTEIN | 51 | 0.501263 | 1.6023973 | 0 | 0.04940991 |
| REACTOME_NR1H3_NR1H2_REGULATE_GENE_EXPRESSION_LINKED_TO_CHOLESTEROL_TRANSPORT_AND_EFFLUX | 17 | 0.62626535 | 1.602618 | 0.0224 | 0.049433682 |
| GO_REGULATION_OF_CYTOSOLIC_CALCIUM_ION_CONCENTRATION | 201 | -0.36857048 | -1.6610609 | 0 | 0.049440324 |
| GO_NCRNA_TRANSCRIPTION | 55 | 0.49156204 | 1.6021043 | 0.004341534 | 0.04945937 |
| GO_REGULATION_OF_ENDOCRINE_PROCESS | 30 | -0.51058424 | -1.6605202 | 0.002754821 | 0.049469173 |
| GO_ANCHORED_COMPONENT_OF_PLASMA_MEMBRANE | 29 | -0.50680953 | -1.6612033 | 0.005390836 | 0.049472053 |
| GO_REGULATION_OF_LEUKOCYTE_DIFFERENTIATION | 167 | -0.37849465 | -1.6600358 | 0 | 0.04949723 |
| GO_EOSINOPHIL_MIGRATION | 16 | -0.608542 | -1.6613551 | 0.010282776 | 0.04952966 |
| GO_RENAL_TUBULE_DEVELOPMENT | 59 | -0.4469888 | -1.6601503 | 0.003663004 | 0.04956087 |

**Table S2. CKD-Plum enriched geneset**

| *NAME* | *SIZE* | *ES* | *NES* | *NOM p-val* | *FDR q-val* |
|---|---|---|---|---|---|
| REACTOME_HOST_INTERACTIONS_OF_HIV_FACTORS | 78 | 0.6683543 | 2.413475 | 0 | 0 |
| HALLMARK_PROTEIN_SECRETION | 72 | 0.6724704 | 2.392642 | 0 | 0 |
| REACTOME_MITOTIC_METAPHASE_AND_ANAPHASE | 122 | 0.6235686 | 2.3769555 | 0 | 0 |
| HALLMARK_INTERFERON_ALPHA_RESPONSE | 73 | 0.67290586 | 2.3672793 | 0 | 0 |
| REACTOME_CELL_CYCLE_CHECKPOINTS | 126 | 0.61805594 | 2.3556974 | 0 | 0 |
| GO_ANTIGEN_PROCESSING_AND_PRESENTATION_OF_EXOGENOUS_PEPTIDE_ANTIGEN_VIA_MHC_CLASS_I | 50 | 0.7093177 | 2.3514268 | 0 | 0 |
| REACTOME_ANTIGEN_PROCESSING_CROSS_PRESENTATION | 67 | 0.66880304 | 2.3394752 | 0 | 0 |
| HALLMARK_INTERFERON_GAMMA_RESPONSE | 158 | 0.5894067 | 2.3162444 | 0 | 0 |
| HALLMARK_MYC_TARGETS_V1 | 124 | 0.602848 | 2.310885 | 0 | 0 |
| REACTOME_HIV_INFECTION | 133 | 0.59932905 | 2.302423 | 0 | 0 |
| REACTOME_SEPARATION_OF_SISTER_CHROMATIDS | 93 | 0.6188613 | 2.260279 | 0 | 0 |
| HALLMARK_E2F_TARGETS | 134 | 0.5881925 | 2.2591953 | 0 | 0 |
| GO_ANTIGEN_PROCESSING_AND_PRESENTATION_OF_PEPTIDE_ANTIGEN_VIA_MHC_CLASS_I | 67 | 0.63533163 | 2.2587836 | 0 | 0 |
| KEGG_VIRAL_MYOCARDITIS | 51 | 0.663247 | 2.247386 | 0 | 0 |
| GO_ANAPHASE_PROMOTING_COMPLEX_DEPENDENT_CATABOLIC_PROCESS | 42 | 0.6940226 | 2.2457716 | 0 | 0 |
| REACTOME_APC_C_MEDIATED_DEGRADATION_OF_CELL_CYCLE_PROTEINS | 44 | 0.6933524 | 2.2398477 | 0 | 0 |
| GO_AZUROPHIL_GRANULE | 105 | 0.5958775 | 2.2323115 | 0 | 0 |
| GO_PHAGOCYTIC_VESICLE | 86 | 0.6120066 | 2.230073 | 0 | 0 |
| GO_ANTIGEN_PROCESSING_AND_PRESENTATION_OF_PEPTIDE_ANTIGEN | 134 | 0.57415617 | 2.2149522 | 0 | 0 |
| REACTOME_M_PHASE | 181 | 0.55884045 | 2.2048626 | 0 | 0 |
| REACTOME_METABOLISM_OF_RNA | 323 | 0.5263727 | 2.201286 | 0 | 0 |
| REACTOME_SWITCHING_OF_ORIGINS_TO_A_POST_REPLICATIVE_STATE | 45 | 0.6668859 | 2.188853 | 0 | 0 |
| REACTOME_CLASS_I_MHC_MEDIATED_ANTIGEN_PROCESSING_PRESENTATION | 192 | 0.54258734 | 2.1795783 | 0 | 0 |
| REACTOME_G2_M_CHECKPOINTS | 72 | 0.61519164 | 2.1743748 | 0 | 0 |
| HALLMARK_KRAS_SIGNALING_DN | 136 | -0.5605149 | -2.460704 | 0 | 0 |
| GO_SODIUM_ION_TRANSMEMBRANE_TRANSPORTER_ACTIVITY | 89 | -0.5845436 | -2.412706 | 0 | 0 |
| GO_RIBONUCLEOPROTEIN_COMPLEX | 326 | 0.5139593 | 2.155488 | 0 | 3.12E-05 |
| REACTOME_INTERFERON_SIGNALING | 132 | 0.55430675 | 2.1557667 | 0 | 3.23E-05 |
| REACTOME_ANTIVIRAL_MECHANISM_BY_IFN_STIMULATED_GENES | 54 | 0.64387107 | 2.1563647 | 0 | 3.34E-05 |
| REACTOME_PROCESSING_OF_CAPPED_INTRON_CONTAINING_PRE_MRNA | 129 | 0.5698105 | 2.1568973 | 0 | 3.46E-05 |
| GO_ANTIGEN_PROCESSING_AND_PRESENTATION | 161 | 0.552038 | 2.1625018 | 0 | 3.59E-05 |
| GO_RNA_EXPORT_FROM_NUCLEUS | 74 | 0.6091065 | 2.165959 | 0 | 3.72E-05 |
| HALLMARK_ALLOGRAFT_REJECTION | 148 | 0.5600111 | 2.168533 | 0 | 3.87E-05 |
| GO_SPLICEOSOMAL_COMPLEX | 91 | 0.5883961 | 2.144017 | 0 | 6.10E-05 |
| GO_KINETOCHORE | 63 | 0.61786777 | 2.1375122 | 0 | 1.48E-04 |
| REACTOME_CELL_CYCLE_MITOTIC | 265 | 0.51810133 | 2.1336925 | 0 | 1.73E-04 |
| REACTOME_POSTMITOTIC_NUCLEAR_PORE_COMPLEX_NPC_REFORMATION | 17 | 0.81021106 | 2.1323197 | 0 | 1.97E-04 |
| REACTOME_FCERI_MEDIATED_NF_KB_ACTIVATION | 55 | 0.6243203 | 2.1280785 | 0 | 2.18E-04 |

| | | | | | |
|---|---|---|---|---|---|
| REACTOME_ORC1_REMOVAL_FROM_CHROMATIN | 38 | 0.6731818 | 2.1245267 | 0 | 2.39E-04 |
| REACTOME_MITOTIC_SPINDLE_CHECKPOINT | 49 | 0.64322543 | 2.1211483 | 0 | 2.84E-04 |
| REACTOME_MITOTIC_PROPHASE | 47 | 0.6414262 | 2.1095634 | 0 | 3.11E-04 |
| GO_RNA_LOCALIZATION | 120 | 0.5554457 | 2.1131606 | 0 | 3.19E-04 |
| REACTOME_CELL_CYCLE | 315 | 0.5057555 | 2.1074698 | 0 | 3.19E-04 |
| REACTOME_APC_C_CDH1_MEDIATED_DEGRADATION_OF_CDC20_AND_OTHER_APC_C_CDH1_TARGETED_PROTEINS_IN_LATE_MITOSIS_EARLY_G1 | 37 | 0.6849326 | 2.1089606 | 0 | 3.27E-04 |
| REACTOME_REGULATION_OF_RAS_BY_GAPS | 32 | 0.69699717 | 2.1134174 | 0 | 3.27E-04 |
| REACTOME_THE_ROLE_OF_GTSE1_IN_G2_M_PROGRESSION_AFTER_G2_CHECKPOINT | 47 | 0.6263809 | 2.0998967 | 0 | 3.70E-04 |
| GO_PHAGOCYTIC_VESICLE_MEMBRANE | 50 | 0.63741475 | 2.1003132 | 0 | 3.79E-04 |
| REACTOME_UCH_PROTEINASES | 40 | 0.66195476 | 2.095549 | 0 | 3.83E-04 |
| GO_NEGATIVE_REGULATION_OF_CELL_CYCLE_G2_M_PHASE_TRANSITION | 48 | 0.6289131 | 2.093986 | 0 | 3.95E-04 |
| REACTOME_DEFECTIVE_CFTR_CAUSES_CYSTIC_FIBROSIS | 31 | 0.68754554 | 2.0906463 | 0 | 4.38E-04 |
| GO_U2_TYPE_SPLICEOSOMAL_COMPLEX | 42 | 0.6452494 | 2.0915813 | 0 | 4.47E-04 |
| REACTOME_RESOLUTION_OF_SISTER_CHROMATID_COHESION | 61 | 0.6049552 | 2.0875359 | 0 | 4.87E-04 |
| REACTOME_ANTIGEN_PRESENTATION_FOLDING_ASSEMBLY_AND_PEPTIDE_LOADING_OF_CLASS_I_MHC | 23 | 0.73955226 | 2.07766 | 0 | 5.05E-04 |
| GO_PROGRAMMED_NECROTIC_CELL_DEATH | 25 | 0.71164817 | 2.0836613 | 0 | 5.06E-04 |
| GO_CHROMOSOME_CENTROMERIC_REGION | 86 | 0.5661198 | 2.0798142 | 0 | 5.15E-04 |
| GO_RIBONUCLEOPROTEIN_COMPLEX_BIOGENESIS | 198 | 0.52023375 | 2.083983 | 0 | 5.16E-04 |
| REACTOME_DNA_REPLICATION | 70 | 0.59661293 | 2.0715652 | 0 | 5.57E-04 |
| GO_CATALYTIC_STEP_2_SPLICEOSOME | 43 | 0.63545907 | 2.071652 | 0 | 5.67E-04 |
| REACTOME_S_PHASE | 87 | 0.5710918 | 2.068202 | 0 | 5.81E-04 |
| GO_FATTY_ACID_CATABOLIC_PROCESS | 73 | -0.5657588 | -2.286948 | 0 | 5.97E-04 |
| GO_RNA_SPLICING_VIA_TRANSESTERIFICATION_REACTIONS | 168 | 0.52566296 | 2.0634673 | 0 | 6.76E-04 |
| REACTOME_ASSEMBLY_OF_THE_PRE_REPLICATIVE_COMPLEX | 36 | 0.6681778 | 2.064384 | 0 | 6.88E-04 |
| GO_CELLULAR_AMINO_ACID_CATABOLIC_PROCESS | 89 | -0.5506228 | -2.286953 | 0 | 6.97E-04 |
| HALLMARK_G2M_CHECKPOINT | 122 | 0.53573596 | 2.06152 | 0 | 7.14E-04 |
| REACTOME_PROGRAMMED_CELL_DEATH | 109 | 0.54637504 | 2.0468585 | 0 | 8.34E-04 |
| GO_SENSORY_PERCEPTION_OF_SMELL | 30 | -0.6915641 | -2.289806 | 0 | 8.36E-04 |
| GO_AZUROPHIL_GRANULE_MEMBRANE | 36 | 0.66228294 | 2.0471241 | 0 | 8.48E-04 |
| GO_ENDOPEPTIDASE_COMPLEX | 33 | 0.66031414 | 2.0483294 | 0 | 8.61E-04 |
| GO_HOST_CELLULAR_COMPONENT | 42 | 0.638308 | 2.0447505 | 0 | 8.67E-04 |
| GO_NECROTIC_CELL_DEATH | 30 | 0.68386763 | 2.0418549 | 0 | 8.85E-04 |
| REACTOME_STABILIZATION_OF_P53 | 27 | 0.69522655 | 2.0393019 | 0 | 8.86E-04 |
| GO_POSITIVE_REGULATION_OF_TYPE_I_INTERFERON_PRODUCTION | 47 | 0.6182728 | 2.042477 | 0 | 8.98E-04 |
| KEGG_NEUROACTIVE_LIGAND_RECEPTOR_INTERACTION | 138 | -0.5221464 | -2.268368 | 0 | 9.14E-04 |
| GO_HORMONE_ACTIVITY | 59 | -0.5840501 | -2.241135 | 0 | 9.30E-04 |
| PID_ERBB1_RECEPTOR_PROXIMAL_PATHWAY | 24 | 0.69912606 | 2.0348592 | 0 | 9.87E-04 |
| REACTOME_REGULATION_OF_RUNX3_EXPRESSION_AND_ACTIVITY | 29 | 0.69113886 | 2.033812 | 0 | 0.001001003 |
| KEGG_UBIQUITIN_MEDIATED_PROTEOLYSIS | 71 | 0.58129984 | 2.0352154 | 0 | 0.001001705 |
| REACTOME_VIRAL_MESSENGER_RNA_SYNTHESIS | 27 | 0.69172996 | 2.03148 | 0 | 0.001014219 |
| GO_DNA_CONFORMATION_CHANGE | 127 | 0.52750814 | 2.0296998 | 0 | 0.001040358 |
| GO_ALPHA_AMINO_ACID_CATABOLIC_PROCESS | 72 | -0.5748448 | -2.297001 | 0 | 0.001045281 |

| | | | | | |
|---|---|---|---|---|---|
| GO_DNA_GEOMETRIC_CHANGE | 67 | 0.5758989 | 2.025008 | 0 | 0.001073774 |
| GO_MRNA_PROCESSING | 242 | 0.4890593 | 2.026046 | 0 | 0.001076277 |
| GO_MRNA_METABOLIC_PROCESS | 380 | 0.4770255 | 2.026261 | 0 | 0.001077923 |
| REACTOME_NEUTROPHIL_DEGRANULATION | 334 | 0.4791263 | 2.025177 | 0 | 0.001087903 |
| REACTOME_TRANSLATION | 134 | 0.5221553 | 2.0263937 | 0 | 0.001092689 |
| GO_NUCLEAR_CHROMOSOME_TELOMERIC_REGION | 53 | 0.606775 | 2.0152323 | 0 | 0.00117406 |
| REACTOME_SYNTHESIS_OF_ACTIVE_UBIQUITIN_ROLES_OF_E1_AND_E2_ENZYMES | 19 | 0.73846656 | 2.0157442 | 0 | 0.001176338 |
| KEGG_ANTIGEN_PROCESSING_AND_PRESENTATION | 57 | 0.59665555 | 2.0161102 | 0 | 0.001190178 |
| REACTOME_DEGRADATION_OF_BETA_CATENIN_BY_THE_DESTRUCTION_COMPLEX | 42 | 0.62988997 | 2.0132687 | 0 | 0.001193809 |
| GO_NCRNA_EXPORT_FROM_NUCLEUS | 25 | 0.6940861 | 2.020393 | 0 | 0.001197141 |
| GO_MRNA_EXPORT_FROM_NUCLEUS | 59 | 0.5931361 | 2.016747 | 0 | 0.001204347 |
| REACTOME_MRNA_SPLICING | 96 | 0.5454254 | 2.0044754 | 0 | 0.001218812 |
| GO_CHROMOSOMAL_REGION | 151 | 0.52038366 | 2.0168073 | 0 | 0.001218857 |
| REACTOME_NUCLEAR_ENVELOPE_NE_REASSEMBLY | 50 | 0.6115079 | 2.0171525 | 0 | 0.001221961 |
| REACTOME_G1_S_DNA_DAMAGE_CHECKPOINTS | 32 | 0.65968525 | 2.017503 | 0 | 0.001228022 |
| GO_TYPE_I_INTERFERON_PRODUCTION | 72 | 0.57174516 | 2.0181453 | 0 | 0.001231318 |
| KEGG_PROTEASOME | 24 | 0.70372576 | 2.0049481 | 0 | 0.001231778 |
| KEGG_ALLOGRAFT_REJECTION | 24 | 0.7059225 | 2.017169 | 0 | 0.001237046 |
| GO_NUCLEAR_EXPORT | 104 | 0.5401977 | 2.0052204 | 0 | 0.001245023 |
| GO_CONDENSED_CHROMOSOME_CENTROMERIC_REGION | 53 | 0.59363914 | 2.002783 | 0 | 0.001246803 |
| REACTOME_PROTEIN_UBIQUITINATION | 39 | 0.62446404 | 2.0085537 | 0 | 0.00125085 |
| REACTOME_DOWNSTREAM_SIGNALING_EVENTS_OF_B_CELL_RECEPTOR_BCR | 43 | 0.62929213 | 2.011065 | 0 | 0.001256932 |
| GO_ENDOSOME_ORGANIZATION | 36 | 0.6318954 | 2.0055974 | 0 | 0.001258556 |
| GO_SENSORY_PERCEPTION_OF_CHEMICAL_STIMULUS | 56 | -0.5819722 | -2.216829 | 0 | 0.001259764 |
| REACTOME_CROSS_PRESENTATION_OF_SOLUBLE_EXOGENOUS_ANTIGENS_ENDOSOMES | 29 | 0.6714386 | 2.0088847 | 0.001605136 | 0.001264748 |
| REACTOME_SRP_DEPENDENT_COTRANSLATIONAL_PROTEIN_TARGETING_TO_MEMBRANE | 47 | 0.610097 | 1.9983505 | 0 | 0.001339632 |
| GO_PEPTIDASE_COMPLEX | 44 | 0.61614156 | 2.0000222 | 0.001490313 | 0.00135091 |
| REACTOME_INTERACTIONS_OF_REV_WITH_HOST_CELLULAR_PROTEINS | 23 | 0.70071656 | 1.9990311 | 0 | 0.001353029 |
| REACTOME_INFLUENZA_INFECTION | 72 | 0.5707184 | 1.9993372 | 0 | 0.001356855 |
| REACTOME_TRANSCRIPTIONAL_REGULATION_BY_SMALL_RNAS | 30 | 0.6686289 | 2.0000768 | 0 | 0.001364837 |
| GO_POTASSIUM_ION_TRANSMEMBRANE_TRANSPORTER_ACTIVITY | 74 | -0.5765718 | -2.30041 | 0 | 0.001393708 |
| REACTOME_AUF1_HNRNP_D0_BINDS_AND_DESTABILIZES_MRNA | 29 | 0.66961646 | 1.997122 | 0 | 0.001422124 |
| GO_PROTEIN_CONTAINING_COMPLEX_LOCALIZATION | 155 | 0.51193315 | 1.9965429 | 0 | 0.001427157 |
| REACTOME_SCF_SKP2_MEDIATED_DEGRADATION_OF_P27_P21 | 34 | 0.65159315 | 1.9954381 | 0 | 0.001431983 |
| GO_INTERFERON_BETA_PRODUCTION | 31 | 0.65729177 | 1.9944184 | 0 | 0.001432334 |
| GO_MONOVALENT_INORGANIC_CATION_TRANSMEMBRANE_TRANSPORTER_ACTIVITY | 190 | -0.477648 | -2.202571 | 0 | 0.001435763 |
| GO_COPII_COATED_ER_TO_GOLGI_TRANSPORT_VESICLE | 57 | 0.58376306 | 1.9945245 | 0 | 0.001436896 |
| REACTOME_DNA_REPLICATION_PRE_INITIATION | 48 | 0.6091872 | 1.9922173 | 0 | 0.001500539 |
| REACTOME_TRNA_PROCESSING_IN_THE_NUCLEUS | 33 | 0.6474962 | 1.9910709 | 0 | 0.001513663 |
| REACTOME_SUMOYLATION_OF_DNA_REPLICATION_PROTEINS | 31 | 0.66345 | 1.9879491 | 0 | 0.001562276 |

| | | | | | |
|---|---|---|---|---|---|
| PID_INTEGRIN_A4B1_PATHWAY | 21 | 0.7109409 | 1.9869119 | 0.003327787 | 0.001565795 |
| GO_RESPONSE_TO_INTERFERON_BETA | 21 | 0.7169579 | 1.9856813 | 0 | 0.001578085 |
| GO_STEROID_HORMONE_RECEPTOR_ACTIVITY | 34 | -0.6448715 | -2.191557 | 0 | 0.001657305 |
| GO_G_PROTEIN_COUPLED_RECEPTOR_ACTIVITY | 202 | -0.4700158 | -2.197719 | 0 | 0.00166928 |
| REACTOME_DEGRADATION_OF_AXIN | 30 | 0.6542184 | 1.9801168 | 0 | 0.001694291 |
| GO_ORGANIC_ACID_CATABOLIC_PROCESS | 190 | -0.4783824 | -2.192588 | 0 | 0.00178479 |
| REACTOME_RHO_GTPASES_ACTIVATE_FORMINS | 69 | 0.56543046 | 1.9759989 | 0 | 0.001791979 |
| REACTOME_TRANSPORT_OF_MATURE_TRANSCRIPT_TO_CYTOPLASM | 50 | 0.59973985 | 1.9763703 | 0 | 0.001807837 |
| REACTOME_ADAPTIVE_IMMUNE_SYSTEM | 450 | 0.46047106 | 1.9749211 | 0 | 0.001810212 |
| GO_MONOCARBOXYLIC_ACID_CATABOLIC_PROCESS | 89 | -0.5269705 | -2.176902 | 0 | 0.001831304 |
| KEGG_LEISHMANIA_INFECTION | 42 | 0.6137698 | 1.9733098 | 0 | 0.001853466 |
| GO_AZUROPHIL_GRANULE_LUMEN | 62 | 0.5679229 | 1.9719062 | 0 | 0.001895882 |
| GO_RNA_CATABOLIC_PROCESS | 183 | 0.49263284 | 1.9699306 | 0 | 0.001921028 |
| GO_MHC_PROTEIN_COMPLEX | 17 | 0.73082024 | 1.9704585 | 0 | 0.001929204 |
| REACTOME_SIGNALING_BY_ROBO_RECEPTORS | 104 | 0.5279906 | 1.9676455 | 0 | 0.001945341 |
| PID_NECTIN_PATHWAY | 18 | 0.7418624 | 1.9687015 | 0 | 0.001953455 |
| GO_GDP_BINDING | 47 | 0.5960626 | 1.9677721 | 0 | 0.001961287 |
| REACTOME_TCR_SIGNALING | 73 | 0.56164145 | 1.9687763 | 0 | 0.001969734 |
| REACTOME_TRANSPORT_OF_MATURE_MRNAS_DERIVED_FROM_INTRONLESS_TRANSCRIPTS | 24 | 0.6879771 | 1.9656267 | 0 | 0.001984597 |
| GO_SCF_DEPENDENT_PROTEASOMAL_UBIQUITIN_DEPENDENT_PROTEIN_CATABOLIC_PROCESS | 46 | 0.5973283 | 1.9611667 | 0 | 0.002021237 |
| REACTOME_NOD1_2_SIGNALING_PATHWAY | 16 | 0.7662903 | 1.9616158 | 0 | 0.002021794 |
| GO_DEFENSE_RESPONSE_TO_VIRUS | 143 | 0.5062812 | 1.9619421 | 0 | 0.002037714 |
| GO_RNA_SPLICING | 214 | 0.48537934 | 1.9626162 | 0 | 0.002038427 |
| GO_VACUOLAR_MEMBRANE | 234 | 0.48266897 | 1.9607816 | 0 | 0.002043405 |
| GO_CADHERIN_BINDING | 198 | 0.48964214 | 1.9627916 | 0 | 0.002054734 |
| REACTOME_NUCLEAR_RECEPTOR_TRANSCRIPTION_PATHWAY | 31 | -0.6551102 | -2.153754 | 0 | 0.00211145 |
| GO_CATION_CHANNEL_COMPLEX | 93 | -0.5237495 | -2.160498 | 0 | 0.002111707 |
| GO_PROTEIN_LOCALIZATION_TO_CHROMOSOME | 32 | 0.6360091 | 1.9543889 | 0 | 0.002170101 |
| REACTOME_NUCLEAR_PORE_COMPLEX_NPC_DISASSEMBLY | 23 | 0.6804927 | 1.9534518 | 0 | 0.002175698 |
| REACTOME_REGULATION_OF_PTEN_STABILITY_AND_ACTIVITY | 39 | 0.62203604 | 1.954784 | 0 | 0.002179047 |
| GO_PIGMENT_GRANULE | 80 | 0.5442307 | 1.9557768 | 0 | 0.00218072 |
| REACTOME_HIV_LIFE_CYCLE | 80 | 0.5488813 | 1.9561527 | 0 | 0.002182553 |
| PID_FANCONI_PATHWAY | 25 | 0.67534906 | 1.9562125 | 0 | 0.002199213 |
| GO_CELL_SUBSTRATE_JUNCTION | 281 | 0.47149956 | 1.9514256 | 0 | 0.002202265 |
| GO_NUCLEAR_PORE | 48 | 0.58513963 | 1.9510076 | 0 | 0.00220744 |
| GO_REGULATION_OF_SYMBIOSIS_ENCOMPASSING_MUTUALISM_THROUGH_PARASITISM | 136 | 0.504837 | 1.9479407 | 0 | 0.002259274 |
| PID_INTEGRIN3_PATHWAY | 34 | 0.6287587 | 1.9484311 | 0 | 0.002263725 |
| PID_BARD1_PATHWAY | 18 | 0.7225129 | 1.9480375 | 0 | 0.002275184 |
| GO_MULTI_ORGANISM_LOCALIZATION | 36 | 0.6272253 | 1.9484799 | 0 | 0.002279895 |
| KEGG_SPLICEOSOME | 69 | 0.55618626 | 1.9464375 | 0 | 0.002288356 |
| GO_REGULATION_OF_ESTABLISHMENT_OF_PLANAR_POLARITY | 64 | 0.56321174 | 1.9486918 | 0 | 0.002289328 |
| GO_INTRINSIC_APOPTOTIC_SIGNALING_PATHWAY_IN_RESPONSE_TO_DNA_DAMAGE_BY_P53_CLASS_MEDIATOR | 26 | 0.67009497 | 1.9465585 | 0 | 0.002297482 |

| | | | | | |
|---|---|---|---|---|---|
| GO_NEGATIVE_REGULATION_OF_VIRAL_GENOME_REPLICATION | 39 | 0.6171654 | 1.9417236 | 0 | 0.002359385 |
| GO_MIDBODY | 91 | 0.5268361 | 1.9418963 | 0 | 0.002368441 |
| KEGG_SYSTEMIC_LUPUS_ERYTHEMATOSUS | 37 | 0.6236773 | 1.943554 | 0 | 0.002376546 |
| GO_GOLGI_VESICLE_TRANSPORT | 206 | 0.47802287 | 1.9424427 | 0 | 0.002384126 |
| GO_MITOTIC_CYTOKINESIS | 38 | 0.6190112 | 1.9437814 | 0 | 0.002386203 |
| GO_PROTEIN_FOLDING | 131 | 0.49929637 | 1.9354018 | 0 | 0.002396582 |
| REACTOME_NUCLEOTIDE_BINDING_DOMAIN_LEUCINE_RICH_REPEAT_CONTAINING_RECEPTOR_NLR_SIGNALING_PATHWAYS | 27 | 0.6622143 | 1.9431028 | 0 | 0.002399883 |
| REACTOME_MITOTIC_G1_PHASE_AND_G1_S_TRANSITION | 86 | 0.5326739 | 1.9424627 | 0 | 0.00240002 |
| REACTOME_SNRNP_ASSEMBLY | 28 | 0.65419376 | 1.9429327 | 0 | 0.002403221 |
| GO_MHC_CLASS_II_PROTEIN_COMPLEX_BINDING | 16 | 0.76275444 | 1.93569 | 0 | 0.002411468 |
| REACTOME_NUCLEAR_IMPORT_OF_REV_PROTEIN | 22 | 0.69014525 | 1.934612 | 0 | 0.002411633 |
| REACTOME_RHO_GTPASE_EFFECTORS | 148 | 0.4976135 | 1.9368273 | 0 | 0.002423487 |
| GO_REGULATION_OF_HEMATOPOIETIC_PROGENITOR_CELL_DIFFERENTIATION | 50 | 0.58338815 | 1.9360948 | 0 | 0.00242654 |
| REACTOME_RUNX1_REGULATES_TRANSCRIPTION_OF_GENES_INVOLVED_IN_DIFFERENTIATION_OF_HSCS | 39 | 0.60344887 | 1.9343209 | 0 | 0.002426729 |
| REACTOME_CYTOSOLIC_SENSORS_OF_PATHOGEN_ASSOCIATED_DNA | 32 | 0.63295496 | 1.9369661 | 0 | 0.002438826 |
| REACTOME_DEUBIQUITINATION | 120 | 0.509128 | 1.9386634 | 0 | 0.002438944 |
| GO_POSITIVE_REGULATION_OF_I_KAPPAB_KINASE_NF_KAPPAB_SIGNALING | 118 | 0.5054073 | 1.9374415 | 0 | 0.002439043 |
| REACTOME_SIGNALING_BY_NOTCH4 | 49 | 0.5944206 | 1.937016 | 0 | 0.002448214 |
| REACTOME_REGULATION_OF_EXPRESSION_OF_SLITS_AND_ROBOS | 77 | 0.54209906 | 1.937962 | 0 | 0.002454778 |
| GO_RIBOSOME_BIOGENESIS | 136 | 0.50074947 | 1.9327089 | 0 | 0.002485502 |
| GO_DNA_DEPENDENT_ATPASE_ACTIVITY | 58 | 0.5597963 | 1.9316314 | 0 | 0.002490737 |
| GO_MHC_PROTEIN_COMPLEX_BINDING | 23 | 0.68576616 | 1.9317605 | 0 | 0.002499806 |
| GO_REGULATION_OF_VIRAL_LIFE_CYCLE | 87 | 0.5272255 | 1.9327185 | 0 | 0.002500566 |
| GO_UBIQUITIN_LIKE_PROTEIN_CONJUGATING_ENZYME_ACTIVITY | 24 | 0.6749868 | 1.9305564 | 0 | 0.002516119 |
| REACTOME_SUMOYLATION_OF_DNA_DAMAGE_RESPONSE_AND_REPAIR_PROTEINS | 41 | 0.60050553 | 1.9296255 | 0 | 0.002535647 |
| GO_ERAD_PATHWAY | 48 | 0.5886717 | 1.9289852 | 0 | 0.002566368 |
| REACTOME_TRANSCRIPTIONAL_REGULATION_BY_RUNX3 | 52 | 0.57391006 | 1.9277834 | 0 | 0.002596396 |
| REACTOME_INTERLEUKIN_1_SIGNALING | 53 | 0.57693774 | 1.9270319 | 0 | 0.002609363 |
| GO_MODIFICATION_DEPENDENT_MACROMOLECULE_CATABOLIC_PROCESS | 305 | 0.4638428 | 1.9262484 | 0 | 0.002633506 |
| REACTOME_NEGATIVE_REGULATION_OF_NOTCH4_SIGNALING | 33 | 0.6278123 | 1.9252969 | 0 | 0.002640555 |
| REACTOME_SIGNALING_BY_THE_B_CELL_RECEPTOR_BCR | 76 | 0.53971654 | 1.9221085 | 0 | 0.002754397 |
| REACTOME_SUMOYLATION_OF_UBIQUITINYLATION_PROTEINS | 22 | 0.6879254 | 1.9222535 | 0 | 0.002759081 |
| GO_CELLULAR_PROTEIN_CATABOLIC_PROCESS | 388 | 0.45474252 | 1.9225366 | 0 | 0.002769254 |
| REACTOME_SUMOYLATION_OF_SUMOYLATION_PROTEINS | 23 | 0.6822891 | 1.9208729 | 0 | 0.002783036 |
| REACTOME_SUMOYLATION_OF_CHROMATIN_ORGANIZATION_PROTEINS | 35 | 0.6123164 | 1.9204304 | 0.001567398 | 0.002789297 |
| GO_REGULATION_OF_CELL_CYCLE_G2_M_PHASE_TRANSITION | 105 | 0.5192802 | 1.9202278 | 0 | 0.002789948 |

| | | | | | |
|---|---|---|---|---|---|
| GO_POSITIVE_REGULATION_OF_WNT_SIGNALING_PATHWAY | 103 | 0.5106695 | 1.9209391 | 0 | 0.002793194 |
| GO_MYELOID_LEUKOCYTE_MEDIATED_IMMUNITY | 389 | 0.45028362 | 1.9190408 | 0 | 0.002849075 |
| GO_RNA_DEPENDENT_DNA_BIOSYNTHETIC_PROCESS | 33 | 0.6208147 | 1.917503 | 0 | 0.002886393 |
| REACTOME_HCMV_EARLY_EVENTS | 47 | 0.57935673 | 1.9169625 | 0 | 0.002896797 |
| GO_PROTEASOMAL_PROTEIN_CATABOLIC_PROCESS | 239 | 0.46717048 | 1.9161325 | 0 | 0.002912631 |
| KEGG_LYSOSOME | 92 | 0.5260706 | 1.9162414 | 0 | 0.002912673 |
| REACTOME_SUMOYLATION_OF_RNA_BINDING_PROTEINS | 27 | 0.64446086 | 1.9164059 | 0 | 0.002922951 |
| REACTOME_INTEGRIN_CELL_SURFACE_INTERACTIONS | 65 | 0.5542226 | 1.9164076 | 0 | 0.002938666 |
| GO_PROTEIN_MODIFICATION_BY_SMALL_PROTEIN_REMOVAL | 123 | 0.49972913 | 1.9138857 | 0 | 0.002947686 |
| GO_ESTABLISHMENT_OF_PROTEIN_LOCALIZATION_TO_ENDOPLASMIC_RETICULUM | 43 | 0.5942194 | 1.9140451 | 0 | 0.002957893 |
| REACTOME_UB_SPECIFIC_PROCESSING_PROTEASES | 90 | 0.521659 | 1.9150809 | 0 | 0.002958273 |
| REACTOME_INTERACTIONS_OF_VPR_WITH_HOST_CELLULAR_PROTEINS | 24 | 0.65915966 | 1.9143323 | 0 | 0.00297338 |
| GO_AXONAL_GROWTH_CONE | 17 | 0.72240376 | 1.91312 | 0 | 0.002982491 |
| PID_CASPASE_PATHWAY | 38 | 0.6040622 | 1.9096655 | 0 | 0.003150451 |
| GO_RIBONUCLEOPROTEIN_COMPLEX_SUBUNIT_ORGANIZATION | 87 | 0.52487624 | 1.908705 | 0 | 0.003159082 |
| GO_REGULATION_OF_VIRAL_GENOME_REPLICATION | 58 | 0.56789076 | 1.9097884 | 0 | 0.003161674 |
| GO_PROTEIN_K48_LINKED_UBIQUITINATION | 30 | 0.64388335 | 1.9068406 | 0 | 0.003200188 |
| REACTOME_HEDGEHOG_LIGAND_BIOGENESIS | 34 | 0.6261041 | 1.9069037 | 0 | 0.003216351 |
| REACTOME_NUCLEAR_ENVELOPE_BREAKDOWN | 36 | 0.61204135 | 1.9056691 | 0 | 0.003237747 |
| REACTOME_METABOLISM_OF_POLYAMINES | 36 | 0.6130805 | 1.9050634 | 0 | 0.003249255 |
| REACTOME_CELLULAR_RESPONSE_TO_HYPOXIA | 39 | 0.6083603 | 1.9052819 | 0.001531394 | 0.003250851 |
| GO_REGULATION_OF_PROTEIN_MATURATION | 41 | 0.5956331 | 1.9044148 | 0 | 0.00329077 |
| GO_INTRINSIC_COMPONENT_OF_ENDOPLASMIC_RETICULUM_MEMBRANE | 92 | 0.5230191 | 1.9031708 | 0 | 0.0033061 |
| GO_GLYCOSPHINGOLIPID_METABOLIC_PROCESS | 40 | 0.5859541 | 1.903891 | 0 | 0.00330797 |
| GO_RNA_3_END_PROCESSING | 67 | 0.54325616 | 1.9009006 | 0 | 0.003398546 |
| KEGG_O_GLYCAN_BIOSYNTHESIS | 17 | 0.72155845 | 1.9001521 | 0 | 0.003403404 |
| GO_PROTEIN_DNA_COMPLEX_SUBUNIT_ORGANIZATION | 114 | 0.50537235 | 1.8997962 | 0 | 0.003405628 |
| GO_LUMENAL_SIDE_OF_MEMBRANE | 22 | 0.68043214 | 1.9003808 | 0 | 0.003415151 |
| REACTOME_DIGESTION_AND_ABSORPTION | 16 | -0.7560031 | -2.115295 | 0 | 0.003416061 |
| REACTOME_TRANSCRIPTIONAL_REGULATION_BY_RUNX2 | 71 | 0.5283359 | 1.8991886 | 0 | 0.003435552 |
| REACTOME_EPH_EPHRIN_SIGNALING | 64 | 0.5419067 | 1.8971877 | 0 | 0.003448516 |
| GO_SKELETAL_MUSCLE_CONTRACTION | 26 | -0.6537805 | -2.107689 | 0 | 0.003449147 |
| GO_ANTIGEN_PROCESSING_AND_PRESENTATION_OF_PEPTIDE_OR_POLYSACCHARIDE_ANTIGEN_VIA_MHC_CLASS_II | 76 | 0.5297659 | 1.8972112 | 0 | 0.003460175 |
| GO_NUCLEAR_MATRIX | 52 | 0.5588168 | 1.8972205 | 0 | 0.003476497 |
| REACTOME_DEGRADATION_OF_DVL | 32 | 0.62641627 | 1.8978139 | 0 | 0.003479275 |
| KEGG_GRAFT_VERSUS_HOST_DISEASE | 28 | 0.6411647 | 1.8958963 | 0.00155521 | 0.003490576 |
| REACTOME_NS1_MEDIATED_EFFECTS_ON_HOST_PATHWAYS | 25 | 0.65897423 | 1.8961043 | 0 | 0.003497779 |
| REACTOME_INTERLEUKIN_12_FAMILY_SIGNALING | 34 | 0.6085888 | 1.8947626 | 0 | 0.003507669 |
| GO_VESICLE_ORGANIZATION | 175 | 0.47856992 | 1.896205 | 0 | 0.003514048 |
| GO_RNA_PROCESSING | 426 | 0.442856 | 1.8949058 | 0 | 0.0035193 |
| GO_POTASSIUM_CHANNEL_ACTIVITY | 51 | -0.5680606 | -2.125893 | 0 | 0.003524866 |
| GO_POTASSIUM_CHANNEL_COMPLEX | 42 | -0.5991178 | -2.122056 | 0 | 0.003563936 |
| GO_LIPID_OXIDATION | 77 | -0.5235485 | -2.115817 | 0 | 0.003578731 |
| REACTOME_PCP_CE_PATHWAY | 52 | 0.5639666 | 1.8933051 | 0 | 0.003589151 |

| | | | | | |
|---|---|---|---|---|---|
| GO_OLFACTORY_RECEPTOR_ACTIVITY | 17 | -0.7292876 | -2.118874 | 0 | 0.003598762 |
| GO_CELL_DIVISION | 269 | 0.4609738 | 1.8929135 | 0 | 0.003603934 |
| REACTOME_DEGRADATION_OF_GLI1_BY_THE_PROTEASOME | 31 | 0.627757 | 1.8918602 | 0 | 0.003666612 |
| GO_HELICASE_ACTIVITY | 77 | 0.53406036 | 1.8911108 | 0 | 0.003676361 |
| GO_UBIQUITIN_DEPENDENT_ERAD_PATHWAY | 39 | 0.59983015 | 1.8901368 | 0 | 0.003694546 |
| REACTOME_PTEN_REGULATION | 79 | 0.5221036 | 1.8894447 | 0 | 0.003721418 |
| GO_ESTABLISHMENT_OF_RNA_LOCALIZATION | 105 | 0.5046963 | 1.8878573 | 0 | 0.003749233 |
| GO_POSITIVE_REGULATION_OF_TUMOR_NECROSIS_FACTOR_SUPERFAMILY_CYTOKINE_PRODUCTION | 57 | 0.5504187 | 1.8880147 | 0 | 0.003757161 |
| GO_ENDOPLASMIC_RETICULUM_TO_GOLGI_VESICLE_MEDIATED_TRANSPORT | 121 | 0.49002898 | 1.8885067 | 0 | 0.003760907 |
| GO_RESPONSE_TO_TYPE_I_INTERFERON | 67 | 0.53988117 | 1.8870661 | 0 | 0.003766759 |
| REACTOME_SIGNALING_BY_PDGF | 44 | 0.581463 | 1.8862653 | 0 | 0.003779867 |
| REACTOME_CYCLIN_A_CDK2_ASSOCIATED_EVENTS_AT_S_PHASE_ENTRY | 47 | 0.5794455 | 1.8854336 | 0 | 0.003814485 |
| GO_RETROGRADE_VESICLE_MEDIATED_TRANSPORT_GOLGI_TO_ENDOPLASMIC_RETICULUM | 58 | 0.5536157 | 1.8821996 | 0 | 0.003817279 |
| GO_SPECIFIC_GRANULE | 116 | 0.5029824 | 1.8856075 | 0 | 0.003818402 |
| REACTOME_EXPORT_OF_VIRAL_RIBONUCLEOPROTEINS_FROM_NUCLEUS | 22 | 0.6808843 | 1.8822062 | 0.001675042 | 0.003833118 |
| GO_REGULATION_OF_VACUOLE_ORGANIZATION | 26 | 0.64494497 | 1.8824666 | 0 | 0.003836976 |
| REACTOME_MAP2K_AND_MAPK_ACTIVATION | 26 | 0.6443896 | 1.8829402 | 0.001579779 | 0.003840842 |
| GO_MORPHOGENESIS_OF_A_POLARIZED_EPITHELIUM | 83 | 0.5177105 | 1.8846924 | 0 | 0.00384391 |
| GO_FICOLIN_1_RICH_GRANULE | 94 | 0.5098986 | 1.8844091 | 0 | 0.003848191 |
| PID_SYNDECAN_1_PATHWAY | 33 | 0.6203615 | 1.8829516 | 0 | 0.00385698 |
| GO_DNA_TEMPLATED_TRANSCRIPTION_TERMINATION | 35 | 0.61073107 | 1.8829633 | 0 | 0.003873254 |
| GO_REGULATION_OF_DEFENSE_RESPONSE_TO_VIRUS_BY_HOST | 18 | 0.70007545 | 1.8830498 | 0 | 0.003885571 |
| REACTOME_RRNA_PROCESSING | 90 | 0.5151297 | 1.8832854 | 0 | 0.003889719 |
| REACTOME_ASPARAGINE_N_LINKED_GLYCOSYLATION | 199 | 0.47169706 | 1.8804754 | 0 | 0.003909765 |
| GO_INTERFERON_GAMMA_MEDIATED_SIGNALING_PATHWAY | 59 | 0.5491767 | 1.8792824 | 0 | 0.003941015 |
| GO_INTERFERON_ALPHA_PRODUCTION | 17 | 0.70642054 | 1.878829 | 0.003430532 | 0.003952656 |
| GO_NEGATIVE_REGULATION_OF_CELL_CYCLE_PHASE_TRANSITION | 123 | 0.4848991 | 1.8792944 | 0 | 0.003957036 |
| GO_COTRANSLATIONAL_PROTEIN_TARGETING_TO_MEMBRANE | 38 | 0.59324944 | 1.8793306 | 0 | 0.003969221 |
| GO_GOLGI_ASSOCIATED_VESICLE | 101 | 0.50774896 | 1.8795881 | 0 | 0.003977628 |
| GO_SPECIFIC_GRANULE_LUMEN | 46 | 0.58112687 | 1.877564 | 0 | 0.004026565 |
| GO_RESPONSE_TO_AXON_INJURY | 45 | 0.5776555 | 1.8766334 | 0 | 0.004084429 |
| REACTOME_TRANSPORT_OF_THE_SLBP_DEPENDANT_MATURE_MRNA | 23 | 0.67740244 | 1.8747001 | 0 | 0.004120559 |
| REACTOME_ANTIGEN_PROCESSING_UBIQUITINATION_PROTEASOME_DEGRADATION | 140 | 0.488334 | 1.8751174 | 0 | 0.004121433 |
| HALLMARK_DNA_REPAIR | 99 | 0.5112772 | 1.8753873 | 0 | 0.004130126 |
| GO_CYTOKINESIS | 73 | 0.52767545 | 1.8740867 | 0 | 0.004150301 |
| GO_POSITIVE_REGULATION_OF_INTERFERON_BETA_PRODUCTION | 21 | 0.6785599 | 1.873405 | 0 | 0.004176021 |
| GO_ENDOPLASMIC_RETICULUM_GOLGI_INTERMEDIATE_COMPARTMENT | 82 | 0.5147624 | 1.8730036 | 0 | 0.00419778 |
| GO_STRUCTURAL_CONSTITUENT_OF_NUCLEAR_PORE | 16 | 0.7177557 | 1.8719347 | 0.003294893 | 0.004310202 |

| | | | | | |
|---|---|---|---|---|---|
| GO_POSTTRANSCRIPTIONAL_REGULATION_OF_GENE_EXPRESSION | 295 | 0.4532142 | 1.8704273 | 0 | 0.004399218 |
| GO_TRANSLATIONAL_INITIATION | 83 | 0.51930964 | 1.8694117 | 0 | 0.004414166 |
| GO_COMPLEMENT_ACTIVATION | 69 | 0.52604246 | 1.8695115 | 0 | 0.00443121 |
| GO_MRNA_3_END_PROCESSING | 52 | 0.5586015 | 1.868813 | 0 | 0.004438072 |
| KEGG_PATHOGENIC_ESCHERICHIA_COLI_INFECTION | 40 | 0.5890896 | 1.8679066 | 0.001474926 | 0.004480628 |
| REACTOME_DECTIN_1_MEDIATED_NONCANONICAL_NF_KB_SIGNALING | 35 | 0.60210687 | 1.8675092 | 0 | 0.004485703 |
| GO_REGULATION_OF_TELOMERE_MAINTENANCE_VIA_TELOMERE_LENGTHENING | 27 | 0.6381587 | 1.8668054 | 0 | 0.004523834 |
| GO_NEGATIVE_REGULATION_OF_VIRAL_LIFE_CYCLE | 52 | 0.55909586 | 1.8661319 | 0 | 0.004547264 |
| PID_EPHB_FWD_PATHWAY | 27 | 0.63557273 | 1.8643768 | 0 | 0.004676372 |
| REACTOME_G2_M_DNA_DAMAGE_CHECKPOINT | 33 | 0.600103 | 1.8635015 | 0 | 0.004706857 |
| GO_VIRAL_GENOME_REPLICATION | 75 | 0.5184054 | 1.8636665 | 0.001414427 | 0.004713553 |
| GO_CYTOSKELETON_DEPENDENT_CYTOKINESIS | 42 | 0.5828407 | 1.8630897 | 0 | 0.004714691 |
| REACTOME_REGULATION_OF_MRNA_STABILITY_BY_PROTEINS_THAT_BIND_AU_RICH_ELEMENTS | 43 | 0.5838437 | 1.8589092 | 0 | 0.005025089 |
| REACTOME_ASSEMBLY_OF_COLLAGEN_FIBRILS_AND_OTHER_MULTIMERIC_STRUCTURES | 37 | 0.5958348 | 1.8580767 | 0 | 0.005048239 |
| GO_CELL_ADHESION_MOLECULE_BINDING | 308 | 0.44531038 | 1.8584441 | 0 | 0.005049779 |
| GO_CHROMOSOME_LOCALIZATION | 36 | 0.6034446 | 1.8581824 | 0 | 0.005056079 |
| GO_PROTEIN_LOCALIZATION_TO_ENDOPLASMIC_RETICULUM | 57 | 0.5427779 | 1.857054 | 0 | 0.005086702 |
| GO_INTERLEUKIN_1_MEDIATED_SIGNALING_PATHWAY | 54 | 0.5475092 | 1.8568149 | 0 | 0.005092901 |
| KEGG_N_GLYCAN_BIOSYNTHESIS | 32 | 0.6092356 | 1.8544052 | 0.0015625 | 0.005317153 |
| GO_BASEMENT_MEMBRANE | 55 | 0.556488 | 1.8542048 | 0 | 0.005329351 |
| REACTOME_RNA_POLYMERASE_II_TRANSCRIBES_SNRNA_GENES | 36 | 0.6008661 | 1.8539082 | 0 | 0.005348628 |
| REACTOME_INTERFERON_ALPHA_BETA_SIGNALING | 55 | 0.56303287 | 1.852905 | 0 | 0.005406445 |
| GO_POSITIVE_REGULATION_OF_CANONICAL_WNT_SIGNALING_PATHWAY | 79 | 0.52052635 | 1.8513587 | 0 | 0.005505115 |
| GO_ER_TO_GOLGI_TRANSPORT_VESICLE_MEMBRANE | 38 | 0.5872549 | 1.8507323 | 0 | 0.005537506 |
| REACTOME_FGFR2_ALTERNATIVE_SPLICING | 18 | 0.6868312 | 1.8501486 | 0 | 0.005566221 |
| GO_NUCLEAR_ENVELOPE | 256 | 0.45048237 | 1.8480804 | 0 | 0.005675053 |
| REACTOME_COMPLEMENT_CASCADE | 53 | 0.55642897 | 1.8478991 | 0 | 0.005682517 |
| GO_SISTER_CHROMATID_SEGREGATION | 77 | 0.5128881 | 1.848164 | 0 | 0.005684745 |
| REACTOME_MHC_CLASS_II_ANTIGEN_PRESENTATION | 92 | 0.50618595 | 1.8482579 | 0 | 0.005694532 |
| GO_REGULATION_OF_CELLULAR_AMINO_ACID_METABOLIC_PROCESS | 37 | 0.59323895 | 1.8469527 | 0 | 0.005727196 |
| GO_PROTEIN_CATABOLIC_PROCESS | 474 | 0.42977566 | 1.8466984 | 0 | 0.005734279 |
| GO_HYDROLASE_ACTIVITY_ACTING_ON_ACID_ANHYDRIDES | 434 | 0.4332029 | 1.8456885 | 0 | 0.005792138 |
| REACTOME_STIMULI_SENSING_CHANNELS | 46 | -0.5617295 | -2.070854 | 0 | 0.005794624 |
| GO_GATED_CHANNEL_ACTIVITY | 154 | -0.4565033 | -2.064953 | 0 | 0.005798019 |
| GO_MYD88_INDEPENDENT_TOLL_LIKE_RECEPTOR_SIGNALING_PATHWAY | 19 | 0.6828862 | 1.8459446 | 0 | 0.005798327 |
| REACTOME_HCMV_INFECTION | 56 | 0.54469913 | 1.8457997 | 0 | 0.00580191 |
| GO_VOLTAGE_GATED_POTASSIUM_CHANNEL_ACTIVITY | 33 | -0.6163493 | -2.071948 | 0 | 0.005814679 |
| GO_GLUTAMATE_SECRETION | 29 | -0.6249579 | -2.060814 | 0 | 0.005896285 |
| GO_POSTSYNAPTIC_NEUROTRANSMITTER_RECEPTOR_ACTIVITY | 32 | -0.6101362 | -2.066263 | 0 | 0.005897857 |
| GO_LIGAND_GATED_ION_CHANNEL_ACTIVITY | 69 | -0.5139356 | -2.052628 | 0 | 0.005975494 |

| | | | | | |
|---|---|---|---|---|---|
| REACTOME_RUNX1_INTERACTS_WITH_CO_FACTORS_WHOSE_PRECISE_EFFECT_ON_RUNX1_TARGETS_IS_NOT_KNOWN | 19 | 0.6809651 | 1.8433292 | 0 | 0.005982167 |
| GO_ACID_SECRETION | 78 | -0.5080401 | -2.055977 | 0 | 0.005984694 |
| GO_MITOTIC_METAPHASE_PLATE_CONGRESSION | 25 | 0.6471768 | 1.8423643 | 0.003311258 | 0.006013585 |
| PID_AVB3_INTEGRIN_PATHWAY | 53 | 0.5482405 | 1.8420835 | 0.001459854 | 0.006019541 |
| GO_CYTOKINETIC_PROCESS | 19 | 0.66714114 | 1.8424548 | 0.001602564 | 0.00602739 |
| GO_NUCLEAR_TRANSCRIBED_MRNA_CATABOLIC_PROCESS_EXONUCLEOLYTIC | 17 | 0.7141822 | 1.8426106 | 0 | 0.006031435 |
| GO_NEUROTRANSMITTER_RECEPTOR_ACTIVITY | 44 | -0.5768768 | -2.053527 | 0 | 0.006032059 |
| GO_CATION_CHANNEL_ACTIVITY | 145 | -0.4679935 | -2.04765 | 0 | 0.006062689 |
| GO_FATTY_ACID_BETA_OXIDATION | 54 | -0.5388148 | -2.048222 | 0 | 0.006118031 |
| GO_ESTABLISHMENT_OF_PROTEIN_LOCALIZATION_TO_MEMBRANE | 153 | 0.47442892 | 1.8402523 | 0 | 0.006143563 |
| KEGG_FATTY_ACID_METABOLISM | 33 | -0.593353 | -2.044572 | 0 | 0.006166234 |
| REACTOME_ASYMMETRIC_LOCALIZATION_OF_PCP_PROTEINS | 35 | 0.5985943 | 1.8397968 | 0 | 0.006178202 |
| GO_PROTEIN_TARGETING_TO_VACUOLE | 23 | 0.6520799 | 1.8394759 | 0.001589825 | 0.006190063 |
| GO_PEPTIDYL_ASPARAGINE_MODIFICATION | 24 | 0.6383832 | 1.8375975 | 0 | 0.006331488 |
| GO_RESPONSE_TO_INTERLEUKIN_12 | 30 | 0.6145372 | 1.8364034 | 0 | 0.006410575 |
| REACTOME_MITOTIC_PROMETAPHASE | 98 | 0.49743626 | 1.8358865 | 0 | 0.006444117 |
| GO_LUMENAL_SIDE_OF_ENDOPLASMIC_RETICULUM_MEMBRANE | 17 | 0.69845915 | 1.8356354 | 0 | 0.006445358 |
| GO_POSITIVE_REGULATION_OF_TOLL_LIKE_RECEPTOR_SIGNALING_PATHWAY | 15 | 0.72200084 | 1.8329663 | 0 | 0.006520984 |
| REACTOME_E3_UBIQUITIN_LIGASES_UBIQUITINATE_TARGET_PROTEINS | 26 | 0.6284933 | 1.8339481 | 0 | 0.006521221 |
| REACTOME_EPHB_MEDIATED_FORWARD_SIGNALING | 28 | 0.62424004 | 1.8333551 | 0.001517451 | 0.006526322 |
| REACTOME_INTERFERON_GAMMA_SIGNALING | 61 | 0.5396848 | 1.8340818 | 0 | 0.006536223 |
| GO_CHROMOSOME_TELOMERIC_REGION | 67 | 0.52364796 | 1.8334553 | 0 | 0.00654446 |
| REACTOME_REGULATION_OF_RUNX2_EXPRESSION_AND_ACTIVITY | 41 | 0.5707504 | 1.8318979 | 0.00462963 | 0.006557421 |
| GO_SM_LIKE_PROTEIN_FAMILY_COMPLEX | 32 | 0.6050223 | 1.8315487 | 0.001607717 | 0.006558231 |
| REACTOME_GOLGI_TO_ER_RETROGRADE_TRANSPORT | 88 | 0.4982653 | 1.8319913 | 0 | 0.00656603 |
| GO_INNATE_IMMUNE_RESPONSE_ACTIVATING_CELL_SURFACE_RECEPTOR_SIGNALING_PATHWAY | 62 | 0.53140205 | 1.8295461 | 0 | 0.006680771 |
| GO_EPHRIN_RECEPTOR_SIGNALING_PATHWAY | 59 | 0.54135126 | 1.8291824 | 0 | 0.006687345 |
| PID_PLK1_PATHWAY | 30 | 0.6088238 | 1.8285569 | 0 | 0.006712603 |
| GO_POLYSOME | 35 | 0.5839743 | 1.827651 | 0.001529052 | 0.006722703 |
| GO_TRANSLATION_INITIATION_FACTOR_ACTIVITY | 28 | 0.63119227 | 1.8280749 | 0 | 0.006728399 |
| PID_AJDISS_2PATHWAY | 32 | 0.5966162 | 1.8277698 | 0.006125575 | 0.006731694 |
| REACTOME_TRNA_PROCESSING | 56 | 0.5288768 | 1.8268217 | 0 | 0.006781021 |
| GO_GTPASE_ACTIVITY | 168 | 0.45870116 | 1.825746 | 0 | 0.006805653 |
| GO_VACUOLE | 475 | 0.42428565 | 1.8261497 | 0 | 0.006811553 |
| GO_REGULATION_OF_MRNA_CATABOLIC_PROCESS | 92 | 0.4945158 | 1.8262945 | 0 | 0.006817611 |
| GO_REGULATION_OF_MRNA_METABOLIC_PROCESS | 160 | 0.4677586 | 1.8258027 | 0 | 0.006817732 |
| GO_REGULATION_OF_CYTOKINESIS | 40 | 0.5745705 | 1.8253152 | 0.00148368 | 0.00682058 |
| GO_CELL_CYCLE_G2_M_PHASE_TRANSITION | 136 | 0.4742028 | 1.8250588 | 0 | 0.006823469 |
| REACTOME_THE_ROLE_OF_NEF_IN_HIV_1_REPLICATION_AND_DISEASE_PATHOGENESIS | 21 | 0.648469 | 1.8238015 | 0 | 0.006826102 |
| REACTOME_PARASITE_INFECTION | 53 | 0.5457241 | 1.825375 | 0 | 0.00683269 |
| GO_PROTEIN_N_LINKED_GLYCOSYLATION | 51 | 0.54905826 | 1.8238202 | 0.001443001 | 0.006843938 |

| | | | | | |
|---|---|---|---|---|---|
| GO_NUCLEOSOME_ORGANIZATION | 55 | 0.53306377 | 1.824203 | 0 | 0.00685021 |
| REACTOME_SIGNALING_BY_MET | 49 | 0.5522008 | 1.8243133 | 0 | 0.006856354 |
| GO_VIRAL_LIFE_CYCLE | 199 | 0.45401594 | 1.8238326 | 0 | 0.006864804 |
| GO_GOLGI_ASSOCIATED_VESICLE_MEMBRANE | 64 | 0.52899826 | 1.8228624 | 0 | 0.006890869 |
| GO_TRANSPORTER_COMPLEX | 151 | -0.4518395 | -2.03452 | 0 | 0.006958029 |
| GO_REGULATION_OF_MULTI_ORGANISM_PROCESS | 450 | 0.42179498 | 1.8214469 | 0 | 0.006960702 |
| GO_REGULATION_OF_SISTER_CHROMATID_SEGREGATION | 29 | 0.614872 | 1.8212996 | 0.001550388 | 0.006966059 |
| GO_MRNA_TRANSPORT | 80 | 0.50641716 | 1.821655 | 0 | 0.006967046 |
| GO_HEMATOPOIETIC_STEM_CELL_DIFFERENTIATION | 52 | 0.54416525 | 1.8197799 | 0 | 0.00708463 |
| REACTOME_NEGATIVE_REGULATORS_OF_DDX58_IFIH1_SIGNALING | 17 | 0.6987067 | 1.8191013 | 0.001669449 | 0.007108771 |
| GO_TRANSFERASE_ACTIVITY_TRANSFERRING_HEXOSYL_GROUPS | 109 | 0.49100828 | 1.8192143 | 0 | 0.007118399 |
| GO_REGULATION_OF_FAT_CELL_DIFFERENTIATION | 69 | -0.5081091 | -2.031697 | 0 | 0.007149052 |
| REACTOME_HCMV_LATE_EVENTS | 28 | 0.60612136 | 1.8181489 | 0.001594896 | 0.007194441 |
| GO_REGULATION_OF_SIGNAL_TRANSDUCTION_BY_P53_CLASS_MEDIATOR | 95 | 0.4890825 | 1.8162502 | 0 | 0.007323433 |
| GO_CELL_CYCLE_CHECKPOINT | 96 | 0.4909545 | 1.8164129 | 0 | 0.007333595 |
| REACTOME_FC_EPSILON_RECEPTOR_FCERI_SIGNALING | 92 | 0.48946574 | 1.8158206 | 0 | 0.007342064 |
| GO_TRANSLATION_FACTOR_ACTIVITY_RNA_BINDING | 40 | 0.56537193 | 1.8151447 | 0 | 0.007406066 |
| GO_INTRINSIC_APOPTOTIC_SIGNALING_PATHWAY_BY_P53_CLASS_MEDIATOR | 44 | 0.5550358 | 1.8141432 | 0 | 0.007492939 |
| KEGG_RNA_DEGRADATION | 24 | 0.6310946 | 1.814201 | 0 | 0.007511905 |
| REACTOME_GPCR_LIGAND_BINDING | 225 | -0.4282256 | -2.026856 | 0 | 0.007644526 |
| GO_SODIUM_CHANNEL_ACTIVITY | 23 | -0.6492099 | -2.024627 | 0 | 0.00772391 |
| GO_RUFFLE | 109 | 0.47944722 | 1.8110361 | 0 | 0.007736458 |
| REACTOME_CLEC7A_DECTIN_1_SIGNALING | 59 | 0.5316518 | 1.8111796 | 0 | 0.00774475 |
| GO_RESPONSE_TO_VIRUS | 196 | 0.44991547 | 1.8114065 | 0 | 0.007750212 |
| GO_PEPTIDE_ANTIGEN_BINDING | 17 | 0.693206 | 1.8114358 | 0 | 0.007761394 |
| REACTOME_NUCLEOTIDE_EXCISION_REPAIR | 59 | 0.5282598 | 1.8088949 | 0 | 0.007959375 |
| REACTOME_COPI_MEDIATED_ANTEROGRADE_TRANSPORT | 70 | 0.51253897 | 1.8082101 | 0 | 0.008025562 |
| GO_GUANOSINE_CONTAINING_COMPOUND_METABOLIC_PROCESS | 23 | 0.6286864 | 1.8078284 | 0 | 0.008041389 |
| GO_VESICLE_BUDDING_FROM_MEMBRANE | 64 | 0.5225293 | 1.8060503 | 0 | 0.008173277 |
| GO_UBIQUITIN_LIKE_PROTEIN_LIGASE_BINDING | 182 | 0.4546803 | 1.8042455 | 0 | 0.008328927 |
| GO_CHROMATIN_ASSEMBLY | 45 | 0.5528711 | 1.8035092 | 0 | 0.008373933 |
| GO_CELL_ACTIVATION_INVOLVED_IN_IMMUNE_RESPONSE | 472 | 0.42061245 | 1.8032923 | 0 | 0.008377791 |
| GO_SYMPORTER_ACTIVITY | 84 | -0.4821473 | -2.018373 | 0 | 0.008394307 |
| GO_REGULATION_OF_TELOMERE_MAINTENANCE | 36 | 0.5705694 | 1.8025035 | 0 | 0.008409016 |
| GO_TELOMERE_MAINTENANCE_VIA_TELOMERE_LENGTHENING | 38 | 0.577429 | 1.8019145 | 0 | 0.008456332 |
| GO_VACUOLAR_LUMEN | 123 | 0.47284573 | 1.8011225 | 0 | 0.008485045 |
| HALLMARK_EPITHELIAL_MESENCHYMAL_TRANSITION | 168 | 0.4550236 | 1.8013374 | 0 | 0.00849517 |
| GO_CHROMOSOME_SEGREGATION | 126 | 0.46798223 | 1.7995316 | 0 | 0.00860991 |
| GO_ACTIVATION_OF_INNATE_IMMUNE_RESPONSE | 186 | 0.44964296 | 1.798913 | 0 | 0.008645298 |
| REACTOME_DISEASES_OF_GLYCOSYLATION | 81 | 0.49583474 | 1.798725 | 0 | 0.008653716 |
| GO_SOLUTE_SODIUM_SYMPORTER_ACTIVITY | 47 | -0.5480288 | -2.013069 | 0 | 0.008687062 |
| GO_NEGATIVE_REGULATION_OF_APOPTOTIC_SIGNALING_PATHWAY | 144 | 0.4604054 | 1.7969971 | 0 | 0.008863232 |
| GO_CALCIUM_ION_IMPORT | 49 | -0.5404947 | -2.007683 | 0 | 0.008914036 |

| | | | | | |
|---|---|---|---|---|---|
| GO_FC_RECEPTOR_SIGNALING_PATHWAY | 125 | 0.46769193 | 1.7954283 | 0 | 0.009020451 |
| GO_GOLGI_ORGANIZATION | 66 | 0.5133697 | 1.7949908 | 0.002967359 | 0.009037744 |
| GO_POSITIVE_REGULATION_OF_RESPONSE_TO_CYTOKINE_STIMULUS | 37 | 0.5770872 | 1.7950257 | 0.003174603 | 0.009059814 |
| GO_DNA_PACKAGING | 63 | 0.520852 | 1.7943511 | 0 | 0.009100719 |
| GO_VACUOLE_ORGANIZATION | 85 | 0.49555856 | 1.7937533 | 0 | 0.009134266 |
| REACTOME_COLLAGEN_DEGRADATION | 39 | 0.5639906 | 1.792166 | 0 | 0.009277105 |
| GO_NEGATIVE_REGULATION_OF_CELL_CYCLE_PROCESS | 170 | 0.44966376 | 1.7922055 | 0 | 0.009299595 |
| GO_MYELOID_LEUKOCYTE_ACTIVATION | 452 | 0.41957346 | 1.7911941 | 0 | 0.009331767 |
| GO_REGULATION_OF_INTRINSIC_APOPTOTIC_SIGNALING_PATHWAY_BY_P53_CLASS_MEDIATOR | 16 | 0.68527484 | 1.7912033 | 0.001636661 | 0.009354289 |
| GO_RIBONUCLEOPROTEIN_COMPLEX_BINDING | 76 | 0.49924168 | 1.7903779 | 0 | 0.009382234 |
| GO_ENDOMEMBRANE_SYSTEM_ORGANIZATION | 207 | 0.44345224 | 1.7896204 | 0 | 0.00942584 |
| GO_NUCLEAR_TRANSCRIBED_MRNA_CATABOLIC_PROCESS_NONSENSE_MEDIATED_DECAY | 47 | 0.55394405 | 1.789823 | 0 | 0.009427503 |
| GO_RUFFLE_MEMBRANE | 55 | 0.5353038 | 1.7889822 | 0 | 0.009471641 |
| GO_I_KAPPAB_KINASE_NF_KAPPAB_SIGNALING | 167 | 0.45574957 | 1.7891358 | 0 | 0.009478817 |
| REACTOME_CARDIAC_CONDUCTION | 71 | -0.5067142 | -1.998432 | 0 | 0.009507187 |
| GO_POTASSIUM_ION_IMPORT_ACROSS_PLASMA_MEMBRANE | 30 | -0.6027751 | -1.999279 | 0 | 0.00953116 |
| GO_DNA_REPAIR | 227 | 0.4374417 | 1.7881919 | 0 | 0.009536617 |
| GO_METAL_ION_TRANSMEMBRANE_TRANSPORTER_ACTIVITY | 225 | -0.4218741 | -2.000487 | 0 | 0.009582287 |
| GO_PASSIVE_TRANSMEMBRANE_TRANSPORTER_ACTIVITY | 203 | -0.4314342 | -1.994843 | 0 | 0.009602537 |
| REACTOME_C_TYPE_LECTIN_RECEPTORS_CLRS | 80 | 0.49331492 | 1.787053 | 0 | 0.009637051 |
| GO_SMALL_NUCLEAR_RIBONUCLEOPROTEIN_COMPLEX | 27 | 0.6119978 | 1.7859001 | 0 | 0.009744834 |
| GO_NUCLEAR_TRANSPORT | 192 | 0.44658393 | 1.7859852 | 0 | 0.009749915 |
| GO_B_CELL_MEDIATED_IMMUNITY | 94 | 0.4884941 | 1.7846973 | 0 | 0.009908078 |
| REACTOME_RNA_POLYMERASE_II_TRANSCRIPTION_TERMINATION | 35 | 0.5728947 | 1.7843198 | 0.001508296 | 0.009932973 |
| GO_PROTEIN_MODIFICATION_BY_SMALL_PROTEIN_CONJUGATION_OR_REMOVAL | 478 | 0.41483277 | 1.7827069 | 0 | 0.010021112 |
| GO_SITE_OF_DNA_DAMAGE | 30 | 0.598659 | 1.7833688 | 0 | 0.010031325 |
| GO_ATPASE_COMPLEX | 37 | 0.5679583 | 1.7827101 | 0 | 0.010042018 |
| REACTOME_NON_INTEGRIN_MEMBRANE_ECM_INTERACTIONS | 43 | 0.5468725 | 1.7828861 | 0.004587156 | 0.010060555 |
| GO_GOLGI_MEMBRANE | 391 | 0.41772503 | 1.7819158 | 0 | 0.010100487 |
| GO_ATPASE_ACTIVITY | 216 | 0.43757346 | 1.7813351 | 0 | 0.010139383 |
| GO_INTRINSIC_APOPTOTIC_SIGNALING_PATHWAY | 176 | 0.44694042 | 1.7808465 | 0 | 0.010178072 |
| GO_LOW_DENSITY_LIPOPROTEIN_RECEPTOR_PARTICLE_METABOLIC_PROCESS | 16 | 0.6809161 | 1.7796916 | 0.005154639 | 0.010305889 |
| GO_SIGNAL_TRANSDUCTION_BY_P53_CLASS_MEDIATOR | 148 | 0.45582724 | 1.7792293 | 0 | 0.010358984 |
| GO_DICARBOXYLIC_ACID_TRANSPORT | 59 | -0.517043 | -1.987075 | 0 | 0.010363461 |
| PID_ECADHERIN_NASCENT_AJ_PATHWAY | 28 | 0.6010157 | 1.7786206 | 0 | 0.010421738 |
| GO_G_PROTEIN_COUPLED_AMINE_RECEPTOR_ACTIVITY | 21 | -0.6594841 | -1.985054 | 0 | 0.010434528 |
| GO_DICARBOXYLIC_ACID_TRANSMEMBRANE_TRANSPORTER_ACTIVITY | 22 | -0.6707228 | -1.987805 | 0 | 0.010474707 |
| REACTOME_DNA_REPAIR | 147 | 0.45437124 | 1.7767034 | 0 | 0.010630131 |
| GO_ATP_DEPENDENT_CHROMATIN_REMODELING | 34 | 0.57571346 | 1.7759396 | 0 | 0.01071626 |
| GO_ACTIVATION_OF_IMMUNE_RESPONSE | 402 | 0.4169537 | 1.7753837 | 0 | 0.010762843 |
| PID_ATR_PATHWAY | 21 | 0.65336436 | 1.7750591 | 0.004830918 | 0.010774892 |
| REACTOME_GENE_SILENCING_BY_RNA | 41 | 0.55434185 | 1.7739561 | 0.003134796 | 0.010901469 |

| | | | | | |
|---|---|---|---|---|---|
| GO_REGULATION_OF_PROTEIN_MODIFICATION_BY_SMALL_PROTEIN_CONJUGATION_OR_REMOVAL | 118 | 0.46113467 | 1.7735131 | 0 | 0.010934996 |
| GO_SNRNA_TRANSCRIPTION | 36 | 0.56791955 | 1.7724231 | 0 | 0.011072869 |
| GO_CELLULAR_RESPONSE_TO_DSRNA | 17 | 0.6689603 | 1.7717274 | 0 | 0.011178891 |
| GO_METAPHASE_PLATE_CONGRESSION | 30 | 0.586914 | 1.7715234 | 0.00152439 | 0.01118024 |
| GO_P38MAPK_CASCADE | 27 | -0.6287217 | -1.978744 | 0 | 0.011215203 |
| GO_ORGANELLE_FUSION | 61 | 0.51829326 | 1.7707655 | 0 | 0.01122475 |
| REACTOME_ABC_FAMILY_PROTEINS_MEDIATED_TRANSPORT | 54 | 0.5239951 | 1.770846 | 0.001408451 | 0.011242895 |
| GO_VESICLE_LUMEN | 235 | 0.43353233 | 1.7709527 | 0 | 0.011244263 |
| GO_VIRAL_GENE_EXPRESSION | 95 | 0.48211 | 1.7702475 | 0 | 0.01126185 |
| GO_CHROMATIN_ASSEMBLY_OR_DISASSEMBLY | 58 | 0.5188883 | 1.7684047 | 0.001449275 | 0.011480192 |
| REACTOME_MITOTIC_G2_G2_M_PHASES | 107 | 0.47269663 | 1.7676038 | 0 | 0.011599579 |
| REACTOME_POTASSIUM_CHANNELS | 50 | -0.5330643 | -1.972622 | 0 | 0.011691293 |
| GO_VOLTAGE_GATED_CALCIUM_CHANNEL_COMPLEX | 21 | -0.6624246 | -1.973917 | 0 | 0.011691555 |
| HALLMARK_MTORC1_SIGNALING | 150 | 0.44893712 | 1.766622 | 0 | 0.011697046 |
| GO_REGULATION_OF_RESPONSE_TO_BIOTIC_STIMULUS | 307 | 0.42305133 | 1.7666273 | 0 | 0.011723253 |
| GO_VESICLE_TARGETING | 56 | 0.5221358 | 1.7650622 | 0 | 0.011926611 |
| GO_GUANYL_NUCLEOTIDE_BINDING | 199 | 0.43624088 | 1.7647558 | 0 | 0.011940029 |
| REACTOME_COLLAGEN_FORMATION | 55 | 0.53296125 | 1.7627363 | 0 | 0.011999284 |
| GO_REGULATION_OF_GENE_SILENCING | 63 | 0.50433064 | 1.7639235 | 0.001420455 | 0.01200225 |
| REACTOME_TRNA_AMINOACYLATION | 15 | 0.68736726 | 1.7632548 | 0.00317965 | 0.012003769 |
| REACTOME_NONSENSE_MEDIATED_DECAY_NMD | 46 | 0.5276515 | 1.7634469 | 0 | 0.012004497 |
| PID_TOLL_ENDOGENOUS_PATHWAY | 16 | 0.6944157 | 1.7640837 | 0 | 0.012021848 |
| PID_FAK_PATHWAY | 39 | 0.54851586 | 1.7627385 | 0.00154321 | 0.012027853 |
| GO_GTP_METABOLIC_PROCESS | 16 | 0.6677885 | 1.7634536 | 0.001694915 | 0.012033355 |
| GO_MITOTIC_SISTER_CHROMATID_SEGREGATION | 64 | 0.50382304 | 1.7635888 | 0.001416431 | 0.012038908 |
| GO_PROTEIN_MODIFICATION_BY_SMALL_PROTEIN_CONJUGATION | 407 | 0.4159966 | 1.7627602 | 0 | 0.012056559 |
| GO_NCRNA_TRANSCRIPTION | 55 | 0.5148364 | 1.762057 | 0 | 0.012057187 |
| GO_P_BODY | 44 | 0.5388193 | 1.7621839 | 0.003134796 | 0.012069709 |
| GO_PROTEASE_BINDING | 79 | 0.49535847 | 1.7607242 | 0 | 0.01219492 |
| KEGG_RETINOL_METABOLISM | 32 | -0.5837947 | -1.967608 | 0 | 0.012196162 |
| GO_CAJAL_BODY | 26 | 0.5921033 | 1.760756 | 0.006359301 | 0.012209863 |
| GO_NUCLEAR_PERIPHERY | 63 | 0.5100437 | 1.7602345 | 0 | 0.012230317 |
| REACTOME_ABC_TRANSPORTER_DISORDERS | 41 | 0.55383444 | 1.7593637 | 0.001453488 | 0.012322324 |
| GO_NUCLEOLUS | 471 | 0.4081805 | 1.7590386 | 0 | 0.012338801 |
| GO_TUMOR_NECROSIS_FACTOR_BIOSYNTHETIC_PROCESS | 17 | 0.6738867 | 1.757512 | 0.006079027 | 0.012525509 |
| GO_CYTOPLASMIC_TRANSLATIONAL_INITIATION | 20 | 0.63957566 | 1.7566371 | 0.003236246 | 0.012596252 |
| GO_MEMBRANE_DOCKING | 89 | 0.48246184 | 1.7566996 | 0.001390821 | 0.012620982 |
| GO_POSITIVE_REGULATION_OF_TELOMERE_MAINTENANCE | 25 | 0.61126244 | 1.7556375 | 0.004580153 | 0.012726787 |
| GO_REGULATION_OF_INTRINSIC_APOPTOTIC_SIGNALING_PATHWAY | 100 | 0.47586623 | 1.7550547 | 0 | 0.012734912 |
| REACTOME_ER_TO_GOLGI_ANTEROGRADE_TRANSPORT | 100 | 0.47496897 | 1.7553709 | 0 | 0.012740037 |
| GO_TOXIN_TRANSPORT | 22 | 0.6277943 | 1.75513 | 0.0015625 | 0.012753147 |
| GO_CATION_TRANSMEMBRANE_TRANSPORTER_ACTIVITY | 316 | -0.4083999 | -1.959679 | 0 | 0.012838787 |
| PID_HNF3B_PATHWAY | 35 | -0.5697399 | -1.958176 | 0 | 0.012892064 |
| GO_REGULATION_OF_CARDIOCYTE_DIFFERENTIATION | 24 | -0.6272429 | -1.956896 | 0 | 0.012944405 |
| GO_BASEMENT_MEMBRANE_ORGANIZATION | 21 | 0.6337186 | 1.7531576 | 0 | 0.012991881 |
| GO_NIK_NF_KAPPAB_SIGNALING | 102 | 0.4738157 | 1.7533185 | 0 | 0.013001549 |
| GO_AMINO_ACID_ACTIVATION | 18 | 0.6536456 | 1.7534579 | 0.001631321 | 0.013004577 |

| | | | | | |
|---|---|---|---|---|---|
| GO_ORGANIC_ACID_TRANSMEMBRANE_TRANSPORTER_ACTIVITY | 78 | -0.4894358 | -1.960081 | 0 | 0.013021871 |
| GO_RAS_PROTEIN_SIGNAL_TRANSDUCTION | 235 | 0.42750987 | 1.7512707 | 0 | 0.013270322 |
| REACTOME_INTRA_GOLGI_AND_RETROGRADE_GOLGI_TO_ER_TRAFFIC | 121 | 0.45655683 | 1.7508001 | 0 | 0.013324032 |
| GO_NUCLEAR_CHROMOSOME_SEGREGATION | 98 | 0.47713494 | 1.7505443 | 0 | 0.013331281 |
| REACTOME_RHO_GTPASES_ACTIVATE_WASPS_AND_WAVES | 25 | 0.6067365 | 1.749229 | 0.007824726 | 0.013514102 |
| REACTOME_REGULATION_OF_GLUCOKINASE_BY_GLUCOKINASE_REGULATORY_PROTEIN | 20 | 0.6425431 | 1.7483464 | 0 | 0.013615223 |
| REACTOME_SIGNALING_BY_PDGFR_IN_DISEASE | 16 | 0.6703067 | 1.7480292 | 0.003344482 | 0.013639368 |
| GO_RESPONSE_TO_INTERFERON_GAMMA | 128 | 0.45636183 | 1.7474675 | 0 | 0.013735624 |
| GO_NEGATIVE_REGULATION_OF_MITOTIC_CELL_CYCLE | 166 | 0.44090447 | 1.7469733 | 0 | 0.013776857 |
| GO_POSITIVE_REGULATION_OF_TELOMERE_MAINTENANCE_VIA_TELOMERE_LENGTHENING | 19 | 0.64715266 | 1.7457952 | 0.003424658 | 0.013944047 |
| GO_POSITIVE_REGULATION_OF_VIRAL_PROCESS | 60 | 0.51174635 | 1.7453371 | 0 | 0.013977105 |
| GO_NCRNA_PROCESSING | 159 | 0.44640335 | 1.7454237 | 0 | 0.013995287 |
| GO_MULTIVESICULAR_BODY_SORTING_PATHWAY | 17 | 0.6687427 | 1.7446834 | 0.003257329 | 0.013996986 |
| PID_LIS1_PATHWAY | 20 | 0.6417908 | 1.744714 | 0.008223685 | 0.014023793 |
| BIOCARTA_CERAMIDE_PATHWAY | 17 | 0.66079533 | 1.7424427 | 0.006655574 | 0.014212498 |
| GO_RESPONSE_TO_ENDOPLASMIC_RETICULUM_STRESS | 157 | 0.44248495 | 1.7432339 | 0 | 0.014213327 |
| GO_REGULATION_OF_CHROMOSOME_SEGREGATION | 38 | 0.54831904 | 1.7427428 | 0 | 0.01422164 |
| GO_NCRNA_METABOLIC_PROCESS | 191 | 0.43509334 | 1.7429261 | 0 | 0.014229369 |
| GO_REGULATION_OF_CHROMOSOME_SEPARATION | 26 | 0.6012138 | 1.7424755 | 0.001618123 | 0.014239467 |
| KEGG_PROXIMAL_TUBULE_BICARBONATE_RECLAMATION | 18 | -0.6620701 | -1.947086 | 0 | 0.014260383 |
| KEGG_VALINE_LEUCINE_AND_ISOLEUCINE_DEGRADATION | 38 | -0.5530377 | -1.94283 | 0 | 0.014410847 |
| GO_INORGANIC_ION_IMPORT_ACROSS_PLASMA_MEMBRANE | 54 | -0.5246319 | -1.943704 | 0 | 0.014456393 |
| GO_PRECATALYTIC_SPLICEOSOME | 27 | 0.5880922 | 1.7401721 | 0.001615509 | 0.014553537 |
| REACTOME_CLASS_A_1_RHODOPSIN_LIKE_RECEPTORS | 168 | -0.4233533 | -1.944704 | 0 | 0.014579391 |
| GO_ATPASE_ACTIVITY_COUPLED | 148 | 0.4480012 | 1.7391924 | 0 | 0.014649 |
| GO_VESICLE_TARGETING_TO_FROM_OR_WITHIN_GOLGI | 43 | 0.53494215 | 1.7392931 | 0 | 0.014666165 |
| REACTOME_MAPK6_MAPK4_SIGNALING | 48 | 0.5298782 | 1.7393118 | 0.00143472 | 0.01469177 |
| GO_PROTEIN_POLYUBIQUITINATION | 146 | 0.44435558 | 1.7387352 | 0 | 0.01470172 |
| REACTOME_PHASE_0_RAPID_DEPOLARISATION | 23 | -0.6354948 | -1.939331 | 0 | 0.014937999 |
| GO_GLYCOSYLATION | 142 | 0.44688293 | 1.7367208 | 0 | 0.015034664 |
| GO_PROTEIN_SUMOYLATION | 46 | 0.5321461 | 1.7365308 | 0 | 0.015037753 |
| GO_U2_TYPE_CATALYTIC_STEP_2_SPLICEOSOME | 16 | 0.66618425 | 1.7368274 | 0.008183306 | 0.015046124 |
| GO_REGULATION_OF_RECEPTOR_BINDING | 16 | 0.6767955 | 1.7360697 | 0.005008347 | 0.015107743 |
| KEGG_TYPE_I_DIABETES_MELLITUS | 28 | 0.58333224 | 1.7353612 | 0.004893964 | 0.0151646 |
| GO_PROTEIN_TARGETING_TO_MEMBRANE | 79 | 0.4870127 | 1.7356311 | 0 | 0.015167031 |
| GO_CYTOPLASMIC_TRANSLATION | 44 | 0.5333337 | 1.7354723 | 0.002894356 | 0.015172117 |
| GO_REGULATION_OF_RESPONSE_TO_CYTOKINE_STIMULUS | 110 | 0.46352133 | 1.7348394 | 0 | 0.015231791 |
| GO_BONE_CELL_DEVELOPMENT | 19 | 0.63600284 | 1.7345978 | 0.009966778 | 0.015255389 |
| BIOCARTA_ECM_PATHWAY | 15 | 0.6832841 | 1.7334288 | 0.008474576 | 0.015359506 |
| KEGG_CELL_CYCLE | 73 | 0.48636365 | 1.7335889 | 0 | 0.015373501 |
| REACTOME_COLLAGEN_CHAIN_TRIMERIZATION | 24 | 0.5986499 | 1.7325429 | 0.001636661 | 0.015434111 |
| GO_RECEPTOR_CATABOLIC_PROCESS | 22 | 0.61794406 | 1.7327867 | 0.004724409 | 0.015435969 |
| REACTOME_INTERLEUKIN_12_SIGNALING | 28 | 0.5887341 | 1.7319539 | 0.004769475 | 0.015475171 |
| REACTOME_TRANSCRIPTIONAL_REGULATION_BY_E2F6 | 16 | 0.65509546 | 1.7320114 | 0.009852217 | 0.015501703 |

| | | | | | |
|---|---|---|---|---|---|
| REACTOME_INTERCONVERSION_OF_NUCLEOTIDE_DI_AND_TRIPHOSPHATES | 21 | 0.629709 | 1.7310199 | 0.004862237 | 0.015560486 |
| GO_POSITIVE_REGULATION_OF_MULTI_ORGANISM_PROCESS | 307 | 0.41768324 | 1.7312092 | 0 | 0.015562627 |
| GO_REGULATION_OF_TRANSCRIPTION_FROM_RNA_POLYMERASE_II_PROMOTER_IN_RESPONSE_TO_HYPOXIA | 40 | 0.5384263 | 1.7313086 | 0.003081664 | 0.015578985 |
| GO_TRANSMISSION_OF_NERVE_IMPULSE | 41 | -0.5515648 | -1.931404 | 0 | 0.015659241 |
| REACTOME_GENE_AND_PROTEIN_EXPRESSION_BY_JAK_STAT_SIGNALING_AFTER_INTERLEUKIN_12_STIMULATION | 23 | 0.6056284 | 1.7296625 | 0.0048 | 0.015754255 |
| REACTOME_INFECTIOUS_DISEASE | 387 | 0.40881598 | 1.7296788 | 0 | 0.015787076 |
| GO_POTASSIUM_ION_HOMEOSTASIS | 24 | -0.6147937 | -1.93162 | 0.002906977 | 0.015807616 |
| REACTOME_DOWNSTREAM_SIGNAL_TRANSDUCTION | 19 | 0.64248043 | 1.7288169 | 0.008130081 | 0.015891338 |
| GO_REGULATION_OF_DEFENSE_RESPONSE_TO_VIRUS_BY_VIRUS | 22 | 0.61665046 | 1.7277538 | 0.004854369 | 0.016018603 |
| REACTOME_VOLTAGE_GATED_POTASSIUM_CHANNELS | 15 | -0.7012827 | -1.931988 | 0 | 0.016028043 |
| REACTOME_GLYCOSPHINGOLIPID_METABOLISM | 28 | 0.5821744 | 1.7277792 | 0 | 0.016049735 |
| GO_DNA_REPLICATION_INDEPENDENT_NUCLEOSOME_ORGANIZATION | 21 | 0.6217025 | 1.7273457 | 0.012461059 | 0.016077565 |
| GO_XENOBIOTIC_METABOLIC_PROCESS | 80 | -0.4829398 | -1.926424 | 0 | 0.016233938 |
| GO_TRANSLATION_REGULATOR_ACTIVITY_NUCLEIC_ACID_BINDING | 45 | 0.53880244 | 1.7256159 | 0 | 0.016394556 |
| GO_REGULATION_OF_TYPE_I_INTERFERON_MEDIATED_SIGNALING_PATHWAY | 23 | 0.609104 | 1.7249275 | 0.003058104 | 0.016500529 |
| GO_VESICLE_DOCKING | 33 | 0.55803055 | 1.7244985 | 0.004511278 | 0.016558295 |
| REACTOME_NGF_STIMULATED_TRANSCRIPTION | 27 | -0.5930063 | -1.923575 | 0 | 0.016577661 |
| REACTOME_MRNA_CAPPING | 17 | 0.64751685 | 1.7238563 | 0.006557377 | 0.01665956 |
| GO_POSITIVE_REGULATION_OF_RESPONSE_TO_BIOTIC_STIMULUS | 229 | 0.42312756 | 1.7229513 | 0 | 0.016774211 |
| GO_LIGAND_GATED_CATION_CHANNEL_ACTIVITY | 54 | -0.5173893 | -1.915744 | 0 | 0.016958414 |
| GO_OXIDOREDUCTASE_ACTIVITY_ACTING_ON_PAIRED_DONORS_WITH_INCORPORATION_OR_REDUCTION_OF_MOLECULAR_OXYGEN_REDUCED_FLAVIN_OR_FLAVOPROTEIN_AS_ONE_DONOR_AND_INCORPORATION_OF_ONE_ATOM_OF_OXYGEN | 22 | -0.6217964 | -1.916672 | 0.002688172 | 0.017066084 |
| KEGG_BETA_ALANINE_METABOLISM | 18 | -0.6627562 | -1.918581 | 0 | 0.01719691 |
| REACTOME_N_GLYCAN_TRIMMING_IN_THE_ER_AND_CALNEXIN_CALRETICULIN_CYCLE | 18 | 0.64604336 | 1.7206186 | 0.006622517 | 0.017229512 |
| GO_NITRIC_OXIDE_MEDIATED_SIGNAL_TRANSDUCTION | 19 | -0.6514934 | -1.916926 | 0.004819277 | 0.01728588 |
| REACTOME_DEGRADATION_OF_THE_EXTRACELLULAR_MATRIX | 85 | 0.48000723 | 1.7197838 | 0 | 0.017340615 |
| GO_SINGLE_STRANDED_RNA_BINDING | 44 | 0.53809756 | 1.7189991 | 0.001552795 | 0.017451486 |
| GO_HUMORAL_IMMUNE_RESPONSE_MEDIATED_BY_CIRCULATING_IMMUNOGLOBULIN | 53 | 0.5068446 | 1.7182628 | 0.001461988 | 0.017534386 |
| GO_INTERSTRAND_CROSS_LINK_REPAIR | 19 | 0.63145274 | 1.7179912 | 0.00952381 | 0.017546166 |
| GO_NUCLEOBASE_CONTAINING_COMPOUND_TRANSPORT | 129 | 0.45157146 | 1.7176092 | 0 | 0.017585253 |
| GO_TERTIARY_GRANULE | 113 | 0.46271443 | 1.716638 | 0 | 0.017760845 |
| REACTOME_CELLULAR_RESPONSES_TO_EXTERNAL_STIMULI | 315 | 0.41108096 | 1.7141651 | 0 | 0.018239137 |
| GO_NUCLEOCYTOPLASMIC_CARRIER_ACTIVITY | 15 | 0.67553693 | 1.7135463 | 0.006688963 | 0.018356549 |
| GO_NEGATIVE_REGULATION_OF_EXTRINSIC_APOPTOTIC_SIGNALING_PATHWAY_VIA_DEATH_DOMAIN_RECEPTORS | 22 | 0.60550666 | 1.7128808 | 0.006451613 | 0.018456038 |

| | | | | | |
|---|---|---|---|---|---|
| GO_PROTEIN_EXIT_FROM_ENDOPLASMIC_RETICULUM | 24 | 0.601294 | 1.7126548 | 0.004862237 | 0.018477479 |
| GO_IMPORT_ACROSS_PLASMA_MEMBRANE | 78 | -0.4795016 | -1.905305 | 0 | 0.018506454 |
| GO_PROTEIN_HETEROTETRAMERIZATION | 17 | 0.64995116 | 1.7117902 | 0.011437909 | 0.018585866 |
| GO_CELLULAR_RESPONSE_TO_INTERFERON_BETA | 15 | 0.67077935 | 1.7111217 | 0.009771987 | 0.018699616 |
| GO_MRNA_BINDING | 118 | 0.44932246 | 1.7105246 | 0.001321004 | 0.018741032 |
| KEGG_AUTOIMMUNE_THYROID_DISEASE | 33 | 0.5601105 | 1.7106105 | 0.003184713 | 0.01877049 |
| REACTOME_PROCESSING_OF_DNA_DOUBLE_STRAND_BREAK_ENDS | 31 | 0.5698096 | 1.7098862 | 0.001470588 | 0.018797537 |
| GO_PURINE_NUCLEOSIDE_TRIPHOSPHATE_METABOLIC_PROCESS | 39 | 0.53939635 | 1.7099609 | 0.00304414 | 0.018825071 |
| NABA_BASEMENT_MEMBRANES | 21 | 0.6078259 | 1.7069365 | 0.009819968 | 0.019428344 |
| GO_GENE_SILENCING | 110 | 0.45481566 | 1.7066565 | 0 | 0.019451313 |
| NABA_COLLAGENS | 24 | 0.5986499 | 1.7056332 | 0.007849294 | 0.01965904 |
| REACTOME_DEADENYLATION_DEPENDENT_MRNA_DECAY | 22 | 0.6013801 | 1.7042595 | 0.001607717 | 0.01988263 |
| GO_EXTRACELLULAR_MATRIX_STRUCTURAL_CONSTITUENT_CONFERRING_TENSILE_STRENGTH | 24 | 0.5986499 | 1.7038566 | 0.006420546 | 0.01989983 |
| GO_LAMELLIPODIUM | 123 | 0.4531386 | 1.7043512 | 0 | 0.019900668 |
| GO_TOLL_LIKE_RECEPTOR_SIGNALING_PATHWAY | 88 | 0.47068122 | 1.704024 | 0 | 0.0199026 |
| GO_POSITIVE_REGULATION_OF_VIRAL_LIFE_CYCLE | 32 | 0.56327355 | 1.7034574 | 0.001538462 | 0.019953797 |
| GO_POSTREPLICATION_REPAIR | 26 | 0.5860348 | 1.7032616 | 0.003267974 | 0.01996215 |
| GO_TRANSFERASE_ACTIVITY_TRANSFERRING_GLYCOSYL_GROUPS | 139 | 0.44457763 | 1.7026819 | 0.00132626 | 0.020010734 |
| BIOCARTA_HIVNEF_PATHWAY | 44 | 0.5249399 | 1.7027303 | 0.001466276 | 0.020040043 |
| GO_TRNA_METABOLIC_PROCESS | 73 | 0.4844348 | 1.7021017 | 0 | 0.020104975 |
| GO_POSITIVE_REGULATION_OF_DEFENSE_RESPONSE | 293 | 0.4090068 | 1.7009804 | 0 | 0.02033738 |
| REACTOME_INITIAL_TRIGGERING_OF_COMPLEMENT | 26 | 0.5864486 | 1.7003121 | 0.004909984 | 0.020449445 |
| GO_ORGANELLE_MEMBRANE_FUSION | 49 | 0.51423204 | 1.7001039 | 0 | 0.020464197 |
| GO_TRANSFORMING_GROWTH_FACTOR_BETA_PRODUCTION | 31 | 0.5630284 | 1.699374 | 0.001652893 | 0.020620687 |
| REACTOME_EXTRACELLULAR_MATRIX_ORGANIZATION | 193 | 0.4239264 | 1.6988356 | 0 | 0.020722328 |
| GO_REGULATION_OF_PROTEIN_STABILITY | 167 | 0.42752174 | 1.6985135 | 0 | 0.020758731 |
| KEGG_CALCIUM_SIGNALING_PATHWAY | 108 | -0.4487457 | -1.892511 | 0 | 0.02094582 |
| GO_RIBONUCLEOSIDE_TRIPHOSPHATE_METABOLIC_PROCESS | 38 | 0.53793985 | 1.6972722 | 0 | 0.020990808 |
| GO_SOLUTE_CATION_SYMPORTER_ACTIVITY | 64 | -0.4845229 | -1.890833 | 0 | 0.021239303 |
| REACTOME_SIGNALING_BY_NOTCH | 99 | 0.45534703 | 1.6960253 | 0 | 0.021281358 |
| REACTOME_RRNA_MODIFICATION_IN_THE_NUCLEUS_AND_CYTOSOL | 32 | 0.5505299 | 1.6957664 | 0.004477612 | 0.021307435 |
| GO_DNA_DAMAGE_RESPONSE_SIGNAL_TRANSDUCTION_BY_P53_CLASS_MEDIATOR | 62 | 0.48594296 | 1.6953589 | 0.001451379 | 0.021388477 |
| GO_COPI_COATED_VESICLE | 16 | 0.6485811 | 1.6950135 | 0.006734007 | 0.021452736 |
| GO_RNA_CAPPING | 17 | 0.6475169 | 1.6947619 | 0.006700167 | 0.021472767 |
| GO_TELOMERE_ORGANIZATION | 69 | 0.4857019 | 1.6943413 | 0.001412429 | 0.021531219 |
| REACTOME_SUMOYLATION | 96 | 0.4587807 | 1.6936482 | 0.001345895 | 0.021563215 |
| REACTOME_SIGNALLING_TO_ERKS | 23 | 0.59199286 | 1.6937667 | 0.011589404 | 0.021579985 |
| REACTOME_INTRINSIC_PATHWAY_FOR_APOPTOSIS | 30 | 0.5621093 | 1.6937835 | 0.006163328 | 0.021616805 |
| GO_CONDENSED_CHROMOSOME | 96 | 0.46394628 | 1.6928157 | 0 | 0.021718862 |
| GO_REGULATION_OF_CENTROSOME_CYCLE | 24 | 0.58459514 | 1.6916298 | 0.011475409 | 0.021852097 |
| GO_NUCLEOSIDE_TRIPHOSPHATE_METABOLIC_PROCESS | 53 | 0.4963559 | 1.6916976 | 0.001470588 | 0.021881865 |
| GO_REGULATION_OF_OXIDATIVE_STRESS_INDUCED_INTRINSIC_APOPTOTIC_SIGNALING_PATHWAY | 17 | 0.6444537 | 1.6917005 | 0.003418803 | 0.021922614 |
| GO_DNA_METABOLIC_PROCESS | 405 | 0.3991285 | 1.6907427 | 0 | 0.022018807 |

| | | | | | |
|---|---|---|---|---|---|
| PID_PI3KCI_PATHWAY | 39 | 0.52598506 | 1.6895235 | 0.002985075 | 0.022247571 |
| GO_LIPOPOLYSACCHARIDE_BINDING | 18 | 0.6298577 | 1.6888518 | 0.009646302 | 0.022378767 |
| GO_GENE_SILENCING_BY_RNA | 79 | 0.47443578 | 1.6883811 | 0.001440922 | 0.022450492 |
| REACTOME_DISEASES_OF_SIGNAL_TRANSDUCTION_BY_GROWTH_FACTOR_RECEPTORS_AND_SECOND_MESSENGERS | 207 | 0.4207989 | 1.6880056 | 0 | 0.022502307 |
| REACTOME_SIGNALING_BY_RHO_GTPASES | 210 | 0.4214268 | 1.6874179 | 0 | 0.022565998 |
| GO_REGULATION_OF_CENTROSOME_DUPLICATION | 18 | 0.62292963 | 1.687546 | 0.014851485 | 0.022571754 |
| GO_ENDOSOMAL_TRANSPORT | 96 | 0.45409355 | 1.6871569 | 0 | 0.0225868 |
| REACTOME_NERVOUS_SYSTEM_DEVELOPMENT | 335 | 0.3998638 | 1.6865526 | 0 | 0.022633541 |
| PID_S1P_S1P1_PATHWAY | 17 | 0.6518664 | 1.6865951 | 0.00499168 | 0.022667758 |
| GO_PERIKARYON | 71 | -0.4791151 | -1.882953 | 0 | 0.022792943 |
| HALLMARK_UNFOLDED_PROTEIN_RESPONSE | 74 | 0.47660503 | 1.6844838 | 0.001388889 | 0.023040581 |
| GO_APOPTOTIC_SIGNALING_PATHWAY | 364 | 0.3991009 | 1.684532 | 0 | 0.023070004 |
| GO_NEGATIVE_REGULATION_OF_CHROMOSOME_ORGANIZATION | 62 | 0.48924989 | 1.684619 | 0.002840909 | 0.02308717 |
| GO_RRNA_METABOLIC_PROCESS | 100 | 0.45152923 | 1.6835526 | 0 | 0.0232767 |
| GO_REGULATION_OF_CELLULAR_AMINE_METABOLIC_PROCESS | 49 | 0.5033928 | 1.6820898 | 0 | 0.023506187 |
| HALLMARK_COMPLEMENT | 163 | 0.42778715 | 1.6821182 | 0 | 0.02353976 |
| BIOCARTA_CASPASE_PATHWAY | 19 | 0.62606686 | 1.6821775 | 0.0046875 | 0.023568198 |
| PID_INTEGRIN1_PATHWAY | 52 | 0.50707597 | 1.6811935 | 0.001533742 | 0.023590159 |
| PID_P53_REGULATION_PATHWAY | 30 | 0.55441314 | 1.6809931 | 0.004823151 | 0.023603583 |
| GO_ENDOCYTIC_VESICLE | 175 | 0.42317402 | 1.6812478 | 0 | 0.02361849 |
| GO_IMMUNOGLOBULIN_PRODUCTION_INVOLVED_IN_IMMUNOGLOBULIN_MEDIATED_IMMUNE_RESPONSE | 28 | 0.5665006 | 1.6813344 | 0.003344482 | 0.023629412 |
| GO_CHROMATIN_REMODELING | 66 | 0.48178187 | 1.680629 | 0.001464129 | 0.02363598 |
| GO_ANTIGEN_RECEPTOR_MEDIATED_SIGNALING_PATHWAY | 165 | 0.42238617 | 1.6814342 | 0 | 0.023642132 |
| GO_SIGNAL_TRANSDUCTION_IN_RESPONSE_TO_DNA_DAMAGE | 73 | 0.47741774 | 1.6800244 | 0 | 0.02374071 |
| GO_FIBRILLAR_CENTER | 72 | 0.474981 | 1.6793599 | 0.004103967 | 0.023855634 |
| REACTOME_DUAL_INCISION_IN_TC_NER | 38 | 0.5276519 | 1.6780027 | 0.004731861 | 0.02415244 |
| GO_ENDOSOME_MEMBRANE | 261 | 0.4030701 | 1.6767989 | 0 | 0.024427755 |
| GO_DOUBLE_STRANDED_RNA_BINDING | 45 | 0.512321 | 1.6765358 | 0.003025719 | 0.024472175 |
| GO_REGULATION_OF_SUBSTRATE_ADHESION_DEPENDENT_CELL_SPREADING | 38 | 0.5283577 | 1.6761593 | 0.00619195 | 0.024523411 |
| GO_PROTEIN_MONOUBIQUITINATION | 33 | 0.5444889 | 1.6749125 | 0.006299213 | 0.024758019 |
| PID_ECADHERIN_STABILIZATION_PATHWAY | 30 | 0.56735545 | 1.6750182 | 0.007763975 | 0.024781117 |
| GO_POSITIVE_REGULATION_OF_PROTEIN_MODIFICATION_BY_SMALL_PROTEIN_CONJUGATION_OR_REMOVAL | 62 | 0.48712793 | 1.6743238 | 0.004291846 | 0.02478604 |
| GO_CENTROMERE_COMPLEX_ASSEMBLY | 20 | 0.60777235 | 1.6744028 | 0.013468013 | 0.024803948 |
| REACTOME_RMTS_METHYLATE_HISTONE_ARGININES | 15 | 0.6645514 | 1.6741087 | 0.005042017 | 0.024808986 |
| REACTOME_SPHINGOLIPID_METABOLISM | 47 | 0.51553905 | 1.6744033 | 0.001468429 | 0.024847388 |
| REACTOME_POSITIVE_EPIGENETIC_REGULATION_OF_RRNA_EXPRESSION | 25 | 0.57506853 | 1.6735516 | 0.003333333 | 0.024945106 |
| REACTOME_REGULATION_OF_HSF1_MEDIATED_HEAT_SHOCK_RESPONSE | 57 | 0.48747817 | 1.6725482 | 0.001430615 | 0.025146509 |
| GO_VESICLE_MEDIATED_TRANSPORT_BETWEEN_ENDOSOMAL_COMPARTMENTS | 26 | 0.57917213 | 1.672278 | 0.004724409 | 0.025178755 |
| REACTOME_HEDGEHOG_OFF_STATE | 66 | 0.4860779 | 1.6725805 | 0.004219409 | 0.02518172 |
| GO_REGULATION_OF_STEM_CELL_DIFFERENTIATION | 63 | 0.47741055 | 1.6715319 | 0.002805049 | 0.02521319 |

| | | | | | |
|---|---|---|---|---|---|
| REACTOME_DAP12_INTERACTIONS | 29 | 0.5598096 | 1.6716449 | 0.008143323 | 0.02522645 |
| HALLMARK_PI3K_AKT_MTOR_SIGNALING | 69 | 0.47334036 | 1.6716548 | 0 | 0.02526827 |
| GO_NUCLEAR_MEMBRANE | 157 | 0.42295763 | 1.6717727 | 0 | 0.025273181 |
| GO_POSITIVE_REGULATION_OF_CYTOKINE_PRODUCTION_INVOLVED_IN_IMMUNE_RESPONSE | 42 | 0.5216152 | 1.6705965 | 0.00304414 | 0.025399942 |
| GO_INTRINSIC_APOPTOTIC_SIGNALING_PATHWAY_IN_RESPONSE_TO_OXIDATIVE_STRESS | 27 | 0.5799983 | 1.6707245 | 0.001626016 | 0.02540679 |
| GO_REPLICATION_FORK | 38 | 0.5236722 | 1.6681639 | 0.00619195 | 0.026032913 |
| GO_NUCLEOTIDE_SUGAR_BIOSYNTHETIC_PROCESS | 19 | 0.61931354 | 1.6669472 | 0.009819968 | 0.026355773 |
| REACTOME_INTEGRIN_SIGNALING | 18 | 0.6223634 | 1.6667248 | 0.008517887 | 0.026372157 |
| REACTOME_TRANSLOCATION_OF_SLC2A4_GLUT4_TO_THE_PLASMA_MEMBRANE | 51 | 0.4964066 | 1.6662561 | 0 | 0.02646608 |
| REACTOME_COPI_DEPENDENT_GOLGI_TO_ER_RETROGRADE_TRAFFIC | 65 | 0.47409368 | 1.665772 | 0.007042253 | 0.026553074 |
| REACTOME_REGULATION_OF_TP53_ACTIVITY_THROUGH_PHOSPHORYLATION | 51 | 0.49743745 | 1.6655774 | 0.001438849 | 0.026577333 |
| REACTOME_RAB_GERANYLGERANYLATION | 33 | 0.54233927 | 1.6644922 | 0.003021148 | 0.026865926 |
| GO_RESPONSE_TO_X_RAY | 19 | 0.6273041 | 1.6642041 | 0.006622517 | 0.026891196 |
| GO_CHROMOSOME_SEPARATION | 35 | 0.54177725 | 1.6633856 | 0.001536098 | 0.026940705 |
| REACTOME_HEDGEHOG_ON_STATE | 41 | 0.5127065 | 1.6634101 | 0.001557632 | 0.026977863 |
| GO_B_CELL_HOMEOSTASIS | 18 | 0.6159997 | 1.6630007 | 0.005128205 | 0.026991567 |
| GO_MITOTIC_CELL_CYCLE_CHECKPOINT | 79 | 0.46777108 | 1.6634505 | 0 | 0.02700849 |
| PID_PDGFRB_PATHWAY | 91 | 0.45940414 | 1.6635189 | 0.001335114 | 0.027036043 |
| GO_HEMATOPOIETIC_PROGENITOR_CELL_DIFFERENTIATION | 98 | 0.44783583 | 1.6626586 | 0.001356852 | 0.02704244 |
| GO_REGULATION_OF_TRANSLATIONAL_INITIATION | 43 | 0.5133539 | 1.6623259 | 0.004531722 | 0.027083416 |
| GO_SECONDARY_ACTIVE_TRANSMEMBRANE_TRANSPORTER_ACTIVITY | 134 | -0.4262708 | -1.864296 | 0 | 0.027212048 |
| GO_DNA_REPLICATION | 132 | 0.43489918 | 1.6609408 | 0.00129199 | 0.027420621 |
| GO_CELLULAR_RESPONSE_TO_DNA_DAMAGE_STIMULUS | 398 | 0.38924274 | 1.6593504 | 0 | 0.027786803 |
| GO_RAB_PROTEIN_SIGNAL_TRANSDUCTION | 35 | 0.5402068 | 1.6593672 | 0.00624025 | 0.027828163 |
| GO_ANION_CATION_SYMPORTER_ACTIVITY | 16 | -0.6597636 | -1.858822 | 0.005025126 | 0.02817584 |
| GO_REGULATION_OF_POSTTRANSCRIPTIONAL_GENE_SILENCING | 52 | 0.5001324 | 1.6572024 | 0.004511278 | 0.02836352 |
| GO_SODIUM_ION_TRANSPORT | 132 | -0.4142743 | -1.859376 | 0 | 0.028380426 |
| REACTOME_OVARIAN_TUMOR_DOMAIN_PROTEASES | 20 | 0.6002019 | 1.6569632 | 0.012658228 | 0.028395586 |
| GO_ORGANELLE_SUBCOMPARTMENT | 194 | 0.41465673 | 1.6572497 | 0 | 0.02840248 |
| GO_NUCLEOBASE_CONTAINING_SMALL_MOLECULE_INTERCONVERSION | 21 | 0.6085008 | 1.6560605 | 0.009677419 | 0.028604003 |
| GO_VOLTAGE_GATED_CATION_CHANNEL_ACTIVITY | 61 | -0.4846008 | -1.85146 | 0 | 0.028664114 |
| GO_DETECTION_OF_STIMULUS_INVOLVED_IN_SENSORY_PERCEPTION | 56 | -0.4830886 | -1.852443 | 0 | 0.02882378 |
| PID_VEGFR1_PATHWAY | 17 | 0.63586587 | 1.6550874 | 0.005025126 | 0.028825078 |
| REACTOME_APOPTOTIC_EXECUTION_PHASE | 37 | 0.5269682 | 1.6548626 | 0.004615385 | 0.028841471 |
| REACTOME_RECYCLING_PATHWAY_OF_L1 | 32 | 0.5400054 | 1.6551409 | 0.014173228 | 0.028866174 |
| GO_NEUROTRANSMITTER_RECEPTOR_ACTIVITY_INVOLVED_IN_REGULATION_OF_POSTSYNAPTIC_MEMBRANE_POTENTIAL | 26 | -0.5883157 | -1.855707 | 0.002808989 | 0.02896859 |
| GO_TRANSMITTER_GATED_CHANNEL_ACTIVITY | 25 | -0.5851778 | -1.852922 | 0 | 0.029039145 |
| GO_RUFFLE_ORGANIZATION | 36 | 0.5241069 | 1.6539214 | 0.002989537 | 0.02908927 |
| GO_REGULATION_OF_APOPTOTIC_SIGNALING_PATHWAY | 247 | 0.4056983 | 1.6533247 | 0 | 0.029186327 |
| REACTOME_SIGNALING_BY_HEDGEHOG | 81 | 0.46072552 | 1.6534355 | 0.001388889 | 0.029194409 |

| | | | | | |
|---|---|---|---|---|---|
| REACTOME_ECM_PROTEOGLYCANS | 50 | 0.500285 | 1.6530236 | 0.00304878 | 0.029237086 |
| GO_CALCIUM_DEPENDENT_PHOSPHOLIPID_BINDING | 28 | 0.5571658 | 1.6526855 | 0.011345219 | 0.029254282 |
| GO_REGULATION_OF_VIRAL_ENTRY_INTO_HOST_CELL | 20 | 0.6112548 | 1.6527257 | 0.004731861 | 0.029292472 |
| GO_ENDOCYTIC_VESICLE_MEMBRANE | 97 | 0.44616607 | 1.6519897 | 0.001369863 | 0.029318567 |
| GO_REGULATION_OF_UBIQUITIN_PROTEIN_TRANSFERASE_ACTIVITY | 24 | 0.5738712 | 1.6522827 | 0.004950495 | 0.029323785 |
| GO_REGULATION_OF_GLUCONEOGENESIS | 26 | -0.5705273 | -1.853139 | 0.002583979 | 0.029326055 |
| PID_S1P_S1P3_PATHWAY | 19 | 0.6143657 | 1.6520281 | 0.01 | 0.02935983 |
| GO_METAPHASE_ANAPHASE_TRANSITION_OF_CELL_CYCLE | 23 | 0.5785311 | 1.6513098 | 0.011589404 | 0.029483506 |
| KEGG_SPHINGOLIPID_METABOLISM | 22 | 0.594262 | 1.6511387 | 0.001615509 | 0.029487597 |
| GO_UDP_GLYCOSYLTRANSFERASE_ACTIVITY | 71 | 0.47446638 | 1.6506964 | 0.001390821 | 0.029563794 |
| GO_FC_EPSILON_RECEPTOR_SIGNALING_PATHWAY | 79 | 0.4618246 | 1.6497692 | 0.001386963 | 0.029730204 |
| GO_GOLGI_VESICLE_BUDDING | 48 | 0.5061613 | 1.650067 | 0.004601227 | 0.029736577 |
| GO_DNA_REPAIR_COMPLEX | 23 | 0.5830564 | 1.6495402 | 0.011146497 | 0.029738763 |
| GO_REGULATION_OF_GENE_EXPRESSION_EPIGENETIC | 136 | 0.42526323 | 1.6498095 | 0 | 0.029760692 |
| GO_ANTERIOR_POSTERIOR_AXIS_SPECIFICATION | 27 | -0.5787761 | -1.84648 | 0.002808989 | 0.029910322 |
| GO_LIPOSACCHARIDE_METABOLIC_PROCESS | 65 | 0.4747939 | 1.6486447 | 0.001443001 | 0.029977104 |
| REACTOME_COLLAGEN_BIOSYNTHESIS_AND_MODIFYING_ENZYMES | 42 | 0.52283794 | 1.6478988 | 0.006006006 | 0.030155504 |
| GO_ION_TRANSMEMBRANE_TRANSPORTER_ACTIVITY | 426 | -0.3665341 | -1.842496 | 0 | 0.030589182 |
| GO_T_CELL_RECEPTOR_SIGNALING_PATHWAY | 130 | 0.43248913 | 1.6459966 | 0 | 0.030736068 |
| GO_MULTICELLULAR_ORGANISMAL_SIGNALING | 117 | -0.4250834 | -1.843178 | 0 | 0.030752158 |
| GO_NEGATIVE_REGULATION_OF_VIRAL_PROCESS | 65 | 0.46645463 | 1.6455444 | 0.004310345 | 0.030826207 |
| GO_LATE_ENDOSOME_MEMBRANE | 71 | 0.46333843 | 1.6446264 | 0.002785515 | 0.030975148 |
| GO_DOUBLE_STRAND_BREAK_REPAIR | 87 | 0.4523774 | 1.6446944 | 0 | 0.03100422 |
| GO_REGULATION_OF_PROTEOLYSIS | 411 | 0.3901872 | 1.644779 | 0 | 0.03102574 |
| GO_EXTRACELLULAR_LIGAND_GATED_ION_CHANNEL_ACTIVITY | 34 | -0.5417783 | -1.840054 | 0 | 0.031032708 |
| KEGG_HISTIDINE_METABOLISM | 19 | -0.6268019 | -1.837043 | 0 | 0.031048585 |
| REACTOME_COPII_MEDIATED_VESICLE_TRANSPORT | 40 | 0.5157585 | 1.643813 | 0.00295421 | 0.031106165 |
| GO_SPLICEOSOMAL_TRI_SNRNP_COMPLEX | 19 | 0.6032983 | 1.6440395 | 0.017973857 | 0.031115346 |
| GO_SITE_OF_DOUBLE_STRAND_BREAK | 22 | 0.58188635 | 1.6438402 | 0.0144 | 0.03114447 |
| GO_GLANDULAR_EPITHELIAL_CELL_DIFFERENTIATION | 24 | -0.5856792 | -1.838621 | 0.002702703 | 0.031152051 |
| REACTOME_GLOBAL_GENOME_NUCLEOTIDE_EXCISION_REPAIR_GG_NER | 48 | 0.4909652 | 1.6429482 | 0.010130246 | 0.03137379 |
| REACTOME_SIGNALING_BY_GPCR | 426 | -0.3650066 | -1.837166 | 0 | 0.03140141 |
| GO_NEGATIVE_REGULATION_OF_GLIOGENESIS | 24 | -0.5806089 | -1.833491 | 0.007731959 | 0.03182312 |
| GO_DNA_INTEGRITY_CHECKPOINT | 72 | 0.4677669 | 1.6409832 | 0.002773925 | 0.031847585 |
| REACTOME_CARBOXYTERMINAL_POST_TRANSLATIONAL_MODIFICATIONS_OF_TUBULIN | 21 | 0.5854585 | 1.6412808 | 0.00792393 | 0.031854533 |
| GO_REGULATION_OF_DNA_DAMAGE_RESPONSE_SIGNAL_TRANSDUCTION_BY_P53_CLASS_MEDIATOR | 20 | 0.600993 | 1.6410073 | 0.013422819 | 0.031894475 |
| GO_POSITIVE_REGULATION_OF_DNA_BIOSYNTHETIC_PROCESS | 36 | 0.5310177 | 1.6402189 | 0.006069803 | 0.031908218 |
| GO_UNFOLDED_PROTEIN_BINDING | 72 | 0.470065 | 1.6405557 | 0 | 0.03193014 |
| GO_NON_CANONICAL_WNT_SIGNALING_PATHWAY | 84 | 0.45585862 | 1.6402677 | 0.004166667 | 0.03193512 |
| GO_DNA_BINDING_TRANSCRIPTION_ACTIVATOR_ACTIVITY | 230 | -0.3906844 | -1.830096 | 0 | 0.03194764 |
| GO_SPHINGOLIPID_METABOLIC_PROCESS | 86 | 0.45397747 | 1.6403214 | 0.002666667 | 0.031960722 |
| GO_REGULATION_OF_NEUROTRANSMITTER_SECRETION | 57 | -0.4795173 | -1.830285 | 0 | 0.032190647 |
| KEGG_P53_SIGNALING_PATHWAY | 46 | 0.50350904 | 1.6391358 | 0.001533742 | 0.03225608 |

| | | | | | |
|---|---|---|---|---|---|
| GO_REGULATION_OF_PHAGOCYTOSIS | 62 | 0.47562456 | 1.6388372 | 0.001512859 | 0.03232056 |
| GO_PHAGOCYTOSIS | 190 | 0.4106393 | 1.6386918 | 0 | 0.032329157 |
| KEGG_TRYPTOPHAN_METABOLISM | 30 | -0.540733 | -1.830802 | 0.010781671 | 0.032335676 |
| GO_PROTEIN_LOCALIZATION_TO_MEMBRANE | 328 | 0.39044708 | 1.6378874 | 0 | 0.032492947 |
| GO_TERTIARY_GRANULE_LUMEN | 43 | 0.5101754 | 1.6379185 | 0.006389776 | 0.03253119 |
| KEGG_SMALL_CELL_LUNG_CANCER | 54 | 0.49353242 | 1.6374582 | 0.001426534 | 0.032593116 |
| GO_LYTIC_VACUOLE_ORGANIZATION | 39 | 0.52150905 | 1.6370512 | 0.007507508 | 0.032613896 |
| GO_REGULATION_OF_PEPTIDASE_ACTIVITY | 256 | 0.40134314 | 1.636808 | 0 | 0.03262222 |
| GO_RESPONSE_TO_AMINO_ACID_STARVATION | 26 | 0.56255454 | 1.6371015 | 0.01618123 | 0.032649055 |
| GO_MEDIATOR_COMPLEX | 16 | 0.6294389 | 1.6361692 | 0.012006861 | 0.0328211 |
| REACTOME_INTERLEUKIN_1_FAMILY_SIGNALING | 75 | 0.46256882 | 1.6360155 | 0.002777778 | 0.032821532 |
| REACTOME_ACTIVATION_OF_ATR_IN_RESPONSE_TO_REPLICATION_STRESS | 20 | 0.5938668 | 1.6355824 | 0.008196721 | 0.03293332 |
| REACTOME_TRANSCRIPTIONAL_REGULATION_BY_TP53 | 194 | 0.40459135 | 1.6350027 | 0 | 0.033003062 |
| GO_TRANSFERASE_COMPLEX | 371 | 0.38693672 | 1.6351447 | 0 | 0.033014886 |
| GO_PROTEIN_LOCALIZATION_TO_ORGANELLE | 493 | 0.37738493 | 1.6348056 | 0 | 0.03301805 |
| GO_HISTONE_UBIQUITINATION | 18 | 0.6150471 | 1.6345987 | 0.017944535 | 0.033040248 |
| GO_ORGANIC_ACID_TRANSPORT | 193 | -0.3943451 | -1.824928 | 0 | 0.033442445 |
| BIOCARTA_RACCYCD_PATHWAY | 18 | 0.60984015 | 1.6332468 | 0.014446228 | 0.033495564 |
| GO_EXTRINSIC_COMPONENT_OF_PLASMA_MEMBRANE | 88 | 0.44907212 | 1.6324701 | 0 | 0.03367528 |
| REACTOME_SEMA4D_IN_SEMAPHORIN_SIGNALING | 17 | 0.62701577 | 1.6325299 | 0.009661836 | 0.03371152 |
| GO_REGULATION_OF_RECEPTOR_RECYCLING | 19 | 0.59445524 | 1.6321028 | 0.017799353 | 0.033748917 |
| GO_PML_BODY | 51 | 0.4875049 | 1.6312711 | 0.001443001 | 0.034004565 |
| REACTOME_TNFR2_NON_CANONICAL_NF_KB_PATHWAY | 60 | 0.47329244 | 1.6310471 | 0.005755396 | 0.034033794 |
| GO_LYMPHOCYTE_MEDIATED_IMMUNITY | 178 | 0.40989488 | 1.6301018 | 0 | 0.03433313 |
| GO_CYTOKINE_MEDIATED_SIGNALING_PATHWAY | 483 | 0.37884563 | 1.6296204 | 0 | 0.03441517 |
| REACTOME_COPI_INDEPENDENT_GOLGI_TO_ER_RETROGRADE_TRAFFIC | 36 | 0.5222479 | 1.6294802 | 0.003076923 | 0.034418907 |
| GO_NEGATIVE_REGULATION_OF_SIGNAL_TRANSDUCTION_BY_P53_CLASS_MEDIATOR | 18 | 0.612571 | 1.6293263 | 0.015679443 | 0.034425747 |
| GO_ACTIVATED_T_CELL_PROLIFERATION | 21 | 0.5884771 | 1.6296991 | 0.02003339 | 0.034434784 |
| GO_NEGATIVE_REGULATION_OF_AUTOPHAGY | 41 | 0.51055723 | 1.629095 | 0.005873715 | 0.03446432 |
| GO_RAC_PROTEIN_SIGNAL_TRANSDUCTION | 25 | 0.5676506 | 1.6286608 | 0.010989011 | 0.034562 |
| GO_REGULATION_OF_PROTEIN_CATABOLIC_PROCESS | 217 | 0.40116957 | 1.6279012 | 0 | 0.03478536 |
| GO_TRANSLATION_REGULATOR_ACTIVITY | 58 | 0.4858442 | 1.6271089 | 0.001420455 | 0.034970544 |
| GO_MACROAUTOPHAGY | 150 | 0.41722256 | 1.626948 | 0 | 0.034972113 |
| GO_REGULATION_OF_CHROMOSOME_ORGANIZATION | 149 | 0.41891298 | 1.6271213 | 0 | 0.035019547 |
| GO_REGULATION_OF_SPINDLE_ORGANIZATION | 25 | 0.5571972 | 1.6264066 | 0.010785825 | 0.03510578 |
| GO_POSITIVE_REGULATION_OF_LAMELLIPODIUM_ORGANIZATION | 16 | 0.6270376 | 1.6260619 | 0.022988506 | 0.035170145 |
| REACTOME_G_ALPHA_S_SIGNALLING_EVENTS | 90 | -0.4443101 | -1.818981 | 0 | 0.035229765 |
| REACTOME_DNA_DAMAGE_BYPASS | 28 | 0.543842 | 1.6252867 | 0.007763975 | 0.035406128 |
| REACTOME_SYNDECAN_INTERACTIONS | 23 | 0.58345336 | 1.6240604 | 0.01255887 | 0.035794597 |
| GO_REGULATION_OF_HUMORAL_IMMUNE_RESPONSE | 65 | 0.46637338 | 1.6233224 | 0.001464129 | 0.035980355 |
| GO_REGULATION_OF_CELLULAR_RESPONSE_TO_HEAT | 55 | 0.47921106 | 1.6233823 | 0 | 0.036006022 |
| GO_IMMUNE_RESPONSE_REGULATING_SIGNALING_PATHWAY | 365 | 0.38188958 | 1.6219691 | 0 | 0.036411673 |
| GO_COMPLEX_OF_COLLAGEN_TRIMERS | 16 | 0.62785625 | 1.6216571 | 0.017713366 | 0.036465287 |
| GO_NEGATIVE_REGULATION_OF_MULTI_ORGANISM_PROCESS | 141 | 0.41682976 | 1.6205122 | 0 | 0.036815133 |
| GO_CYSTEINE_TYPE_PEPTIDASE_ACTIVITY | 76 | 0.45723236 | 1.6205354 | 0 | 0.0368646 |
| GO_POSITIVE_REGULATION_OF_TRANSLATION | 69 | 0.46471193 | 1.6184819 | 0.002762431 | 0.037494566 |

| | | | | | |
|---|---|---|---|---|---|
| REACTOME_SCAVENGING_BY_CLASS_A_RECEPTORS | 15 | 0.6315865 | 1.6181259 | 0.013490725 | 0.037552807 |
| GO_AMYLOID_PRECURSOR_PROTEIN_CATABOLIC_PROCESS | 34 | 0.5244642 | 1.617911 | 0.004680187 | 0.03757447 |
| GO_SECRETORY_GRANULE_MEMBRANE | 189 | 0.40083385 | 1.6175712 | 0 | 0.037658013 |
| KEGG_ECM_RECEPTOR_INTERACTION | 59 | 0.4749676 | 1.6155157 | 0.005813954 | 0.038043242 |
| GO_NEGATIVE_REGULATION_OF_CELLULAR_PROTEIN_CATABOLIC_PROCESS | 45 | 0.50516766 | 1.615654 | 0.008928572 | 0.03804343 |
| GO_FC_RECEPTOR_MEDIATED_STIMULATORY_SIGNALING_PATHWAY | 69 | 0.464962 | 1.6153257 | 0.005586592 | 0.038052816 |
| GO_FILOPODIUM | 64 | 0.47254473 | 1.6162333 | 0.001449275 | 0.038058028 |
| REACTOME_CHROMATIN_MODIFYING_ENZYMES | 87 | 0.4430934 | 1.6156904 | 0.001449275 | 0.038091253 |
| GO_REGULATION_OF_CELL_CYCLE_PHASE_TRANSITION | 230 | 0.39529097 | 1.6156962 | 0 | 0.038143452 |
| GO_SPLICEOSOMAL_SNRNP_ASSEMBLY | 16 | 0.6210476 | 1.6149405 | 0.016474465 | 0.038145933 |
| REACTOME_TRANSCRIPTION_OF_THE_HIV_GENOME | 40 | 0.49954584 | 1.6157328 | 0.005988024 | 0.03818456 |
| REACTOME_FANCONI_ANEMIA_PATHWAY | 15 | 0.6268142 | 1.6143576 | 0.012820513 | 0.03833079 |
| GO_CATALYTIC_ACTIVITY_ACTING_ON_RNA | 157 | 0.41079715 | 1.6135097 | 0 | 0.03846893 |
| GO_CELLULAR_PROTEIN_CONTAINING_COMPLEX_ASSEMBLY | 484 | 0.37398955 | 1.6136603 | 0 | 0.038471118 |
| GO_PROTEIN_TRIMERIZATION | 20 | 0.5730363 | 1.6136643 | 0.010135135 | 0.03852468 |
| GO_MEMBRANE_ORGANIZATION | 444 | 0.37857157 | 1.6124614 | 0 | 0.038638312 |
| REACTOME_TRANSPORT_TO_THE_GOLGI_AND_SUBSEQUENT_MODIFICATION | 122 | 0.4242084 | 1.6126046 | 0.002587322 | 0.03864611 |
| GO_REGULATION_OF_PROTEIN_OLIGOMERIZATION | 18 | 0.6019878 | 1.6126131 | 0.025125628 | 0.03869409 |
| REACTOME_SIGNALING_BY_PTK6 | 30 | 0.53480226 | 1.6126767 | 0.01255887 | 0.03872558 |
| GO_MANGANESE_ION_BINDING | 38 | 0.51328117 | 1.6115377 | 0.001515152 | 0.038850453 |
| GO_NUCLEAR_TRANSCRIBED_MRNA_CATABOLIC_PROCESS | 82 | 0.446547 | 1.611613 | 0.002853067 | 0.03887103 |
| REACTOME_BIOSYNTHESIS_OF_THE_N_GLYCAN_PRECURSOR_DOLICHOL_LIPID_LINKED_OLIGOSACCHARIDE_LLO_AND_TRANSFER_TO_A_NASCENT_PROTEIN | 51 | 0.4901231 | 1.6116853 | 0.007451565 | 0.03890681 |
| GO_CALCIUM_CHANNEL_COMPLEX | 31 | -0.5322194 | -1.802515 | 0.003058104 | 0.039215233 |
| GO_TRANSLESION_SYNTHESIS | 21 | 0.569767 | 1.609871 | 0.006644518 | 0.03943121 |
| GO_SENSORY_PERCEPTION_OF_LIGHT_STIMULUS | 93 | -0.4315745 | -1.806007 | 0 | 0.039523233 |
| GO_NEURON_CELLULAR_HOMEOSTASIS | 21 | -0.5920275 | -1.803552 | 0 | 0.039559163 |
| GO_PITUITARY_GLAND_DEVELOPMENT | 22 | -0.5870276 | -1.802616 | 0.005494506 | 0.039590165 |
| GO_NEGATIVE_REGULATION_OF_FAT_CELL_DIFFERENTIATION | 25 | -0.5762386 | -1.804436 | 0.002617801 | 0.039669827 |
| GO_REGULATION_OF_SYSTEM_PROCESS | 335 | -0.3643834 | -1.796948 | 0 | 0.03983545 |
| GO_TERTIARY_GRANULE_MEMBRANE | 49 | 0.4854756 | 1.6082313 | 0.007496252 | 0.039969318 |
| GO_MULTICELLULAR_ORGANISMAL_MOVEMENT | 33 | -0.5318728 | -1.797412 | 0 | 0.04000536 |
| GO_STEROID_HORMONE_BIOSYNTHETIC_PROCESS | 43 | -0.5027383 | -1.798246 | 0.003003003 | 0.040056843 |
| GO_NUCLEOSOME_BINDING | 26 | 0.54688704 | 1.607599 | 0.012638231 | 0.04008345 |
| GO_GLYCOPROTEIN_BIOSYNTHETIC_PROCESS | 184 | 0.40120906 | 1.6077023 | 0 | 0.040108297 |
| GO_DIGESTION | 86 | -0.4357521 | -1.798499 | 0 | 0.04033825 |
| GO_NEGATIVE_REGULATION_OF_INTRINSIC_APOPTOTIC_SIGNALING_PATHWAY | 61 | 0.46546397 | 1.6067181 | 0.002915452 | 0.04035693 |
| BIOCARTA_FAS_PATHWAY | 25 | 0.5630482 | 1.605286 | 0.01642036 | 0.040817577 |
| GO_PROTEIN_LOCALIZATION_TO_VACUOLE | 37 | 0.50852835 | 1.6050831 | 0.007429421 | 0.040842157 |
| REACTOME_ION_CHANNEL_TRANSPORT | 96 | -0.4344789 | -1.79255 | 0 | 0.040937223 |
| GO_POSITIVE_REGULATION_OF_CHROMOSOME_ORGANIZATION | 81 | 0.4501771 | 1.60443 | 0.002832861 | 0.04094225 |
| GO_GLYCOPROTEIN_METABOLIC_PROCESS | 223 | 0.39334688 | 1.6044699 | 0 | 0.040988535 |

| | | | | | |
|---|---|---|---|---|---|
| GO_MICROBODY_LUMEN | 34 | -0.5212005 | -1.793081 | 0 | 0.041085806 |
| GO_POSITIVE_REGULATION_OF_DNA_METABOLIC_PROCESS | 91 | 0.43801636 | 1.6031524 | 0.002717391 | 0.041345797 |
| REACTOME_EUKARYOTIC_TRANSLATION_INITIATION | 48 | 0.48565117 | 1.603265 | 0.001519757 | 0.04136416 |
| GO_REGULATION_OF_TOLL_LIKE_RECEPTOR_SIGNALING_PATHWAY | 41 | 0.50045174 | 1.60135 | 0.004552352 | 0.04205631 |
| GO_ESTABLISHMENT_OF_ORGANELLE_LOCALIZATION | 237 | 0.39126027 | 1.6000053 | 0 | 0.042421404 |
| GO_TOLL_LIKE_RECEPTOR_4_SIGNALING_PATHWAY | 23 | 0.5656719 | 1.6000744 | 0.014634146 | 0.042443793 |
| KEGG_CELL_ADHESION_MOLECULES_CAMS | 94 | 0.4324113 | 1.5998069 | 0 | 0.04244488 |
| GO_ANCHORING_JUNCTION | 499 | 0.3708808 | 1.6001178 | 0 | 0.042486418 |
| GO_POSITIVE_REGULATION_OF_ORGANELLE_ORGANIZATION | 326 | 0.38200486 | 1.5990367 | 0 | 0.042659655 |
| BIOCARTA_DEATH_PATHWAY | 23 | 0.54462194 | 1.5986363 | 0.014308427 | 0.042763 |
| GO_CYSTEINE_TYPE_ENDOPEPTIDASE_ACTIVITY | 57 | 0.47213522 | 1.5984296 | 0.008633094 | 0.042782865 |
| PID_CXCR3_PATHWAY | 34 | 0.5155455 | 1.5982426 | 0.007656968 | 0.042793483 |
| GO_NEURON_PROJECTION_EXTENSION_INVOLVED_IN_NEURON_PROJECTION_GUIDANCE | 22 | -0.5796378 | -1.786352 | 0 | 0.043013852 |
| GO_RIBONUCLEOPROTEIN_GRANULE | 117 | 0.42098287 | 1.5974942 | 0.001308901 | 0.043057404 |
| GO_NEUROPEPTIDE_HORMONE_ACTIVITY | 15 | -0.6408447 | -1.78512 | 0.007462686 | 0.043205146 |
| GO_REGULATION_OF_DEFENSE_RESPONSE | 444 | 0.37255353 | 1.5968342 | 0 | 0.043271385 |
| GO_EARLY_ENDOSOME_MEMBRANE | 76 | 0.4487953 | 1.5963267 | 0.005698006 | 0.04335497 |
| GO_VACUOLAR_TRANSPORT | 80 | 0.45444956 | 1.5960469 | 0.006756757 | 0.04340328 |
| REACTOME_DDX58_IFIH1_MEDIATED_INDUCTION_OF_INTERFERON_ALPHA_BETA | 38 | 0.5054731 | 1.59635 | 0.004470939 | 0.04340359 |
| REACTOME_DNA_DAMAGE_TELOMERE_STRESS_INDUCED_SENESCENCE | 16 | 0.6013704 | 1.5958669 | 0.015100671 | 0.04341713 |
| GO_SMALL_GTPASE_MEDIATED_SIGNAL_TRANSDUCTION | 299 | 0.3813788 | 1.5956008 | 0 | 0.04347959 |
| REACTOME_ASSOCIATION_OF_TRIC_CCT_WITH_TARGET_PROTEINS_DURING_BIOSYNTHESIS | 16 | 0.61647874 | 1.5954458 | 0.016313214 | 0.043495756 |
| GO_SODIUM_ION_HOMEOSTASIS | 40 | -0.5118488 | -1.781806 | 0 | 0.043701787 |
| GO_PROTEIN_INSERTION_INTO_MEMBRANE | 36 | 0.51132274 | 1.594602 | 0.006144393 | 0.043812357 |
| KEGG_DRUG_METABOLISM_CYTOCHROME_P450 | 41 | -0.5002871 | -1.782192 | 0 | 0.044033863 |
| GO_INORGANIC_ANION_TRANSPORT | 94 | -0.4162954 | -1.771266 | 0 | 0.04419022 |
| GO_REGULATION_OF_CARDIAC_MUSCLE_CELL_DIFFERENTIATION | 18 | -0.6140541 | -1.771842 | 0.004950495 | 0.044295903 |
| GO_IMMUNE_RESPONSE_REGULATING_CELL_SURFACE_RECEPTOR_SIGNALING_PATHWAY | 262 | 0.38841546 | 1.592751 | 0 | 0.04454563 |
| GO_PROTEIN_HETEROOLIGOMERIZATION | 53 | 0.4756081 | 1.5925759 | 0.01510574 | 0.044558816 |
| GO_REGULATION_OF_BLOOD_CIRCULATION | 173 | -0.3903243 | -1.779075 | 0 | 0.044579607 |
| GO_ADAPTIVE_IMMUNE_RESPONSE_BASED_ON_SOMATIC_RECOMBINATION_OF_IMMUNE_RECEPTORS_BUILT_FROM_IMMUNOGLOBULIN_SUPERFAMILY_DOMAINS | 181 | 0.40215802 | 1.592201 | 0 | 0.04465212 |
| GO_SMALL_MOLECULE_CATABOLIC_PROCESS | 296 | -0.3683727 | -1.771877 | 0 | 0.044653274 |
| REACTOME_COOPERATION_OF_PREFOLDIN_AND_TRIC_CCT_IN_ACTIN_AND_TUBULIN_FOLDING | 25 | 0.558762 | 1.591982 | 0.02027027 | 0.044681072 |
| REACTOME_SLC_MEDIATED_TRANSMEMBRANE_TRANSPORT | 142 | -0.3998243 | -1.771883 | 0 | 0.045034926 |
| GO_RUFFLE_ASSEMBLY | 27 | 0.5438054 | 1.5909795 | 0.015479876 | 0.045055147 |
| REACTOME_TRANSPORT_OF_INORGANIC_CATIONS_ANIONS_AND_AMINO_ACIDS_OLIGOPEPTIDES | 64 | -0.4599974 | -1.77679 | 0 | 0.04506091 |
| GO_CELLULAR_RESPONSE_TO_XENOBIOTIC_STIMULUS | 117 | -0.4051574 | -1.77269 | 0 | 0.045112416 |
| GO_DRUG_TRANSPORT | 104 | -0.4131524 | -1.76683 | 0 | 0.04512687 |

| | | | | | |
|---|---|---|---|---|---|
| GO_DNA_DAMAGE_RESPONSE_DETECTION_OF_DNA_DAMAGE | 23 | 0.5594941 | 1.5904945 | 0.020134227 | 0.045135356 |
| GO_REGULATION_OF_CELLULAR_CATABOLIC_PROCESS | 436 | 0.37475374 | 1.5906402 | 0 | 0.045139734 |
| GO_HORMONE_METABOLIC_PROCESS | 152 | -0.4029413 | -1.775427 | 0 | 0.045220938 |
| GO_NEUTRAL_LIPID_METABOLIC_PROCESS | 67 | -0.4407115 | -1.767373 | 0 | 0.045254506 |
| GO_AROMATASE_ACTIVITY | 17 | -0.6321937 | -1.765581 | 0.004938272 | 0.04532702 |
| GO_LIGAND_ACTIVATED_TRANSCRIPTION_FACTOR_ACTIVITY | 29 | -0.5475035 | -1.758994 | 0.008 | 0.045448937 |
| REACTOME_PEROXISOMAL_LIPID_METABOLISM | 19 | -0.6053978 | -1.767694 | 0 | 0.04547197 |
| GO_PHOSPHATE_ION_TRANSPORT | 17 | -0.6205833 | -1.772768 | 0 | 0.045476727 |
| GO_ENDOPLASMIC_RETICULUM_LUMEN | 198 | 0.39611787 | 1.5894519 | 0 | 0.04551138 |
| GO_POSITIVE_REGULATION_OF_LIPID_CATABOLIC_PROCESS | 16 | -0.6335639 | -1.773654 | 0.010178117 | 0.045522198 |
| GO_L_AMINO_ACID_TRANSMEMBRANE_TRANSPORTER_ACTIVITY | 30 | -0.5399169 | -1.759734 | 0.008474576 | 0.045523126 |
| GO_ORGANELLE_LOCALIZATION | 336 | 0.37593928 | 1.58917 | 0 | 0.045574337 |
| GO_THYROID_HORMONE_METABOLIC_PROCESS | 15 | -0.6507825 | -1.761954 | 0.012787724 | 0.04570785 |
| GO_ARGININE_METABOLIC_PROCESS | 15 | -0.6316903 | -1.759978 | 0.012722646 | 0.045735907 |
| GO_SENSORY_PERCEPTION | 258 | -0.3718487 | -1.760729 | 0 | 0.045812137 |
| GO_INORGANIC_ANION_TRANSMEMBRANE_TRANSPORT | 66 | -0.4468605 | -1.762204 | 0 | 0.046036914 |
| GO_CELL_FATE_SPECIFICATION | 41 | -0.4962939 | -1.762877 | 0.002949853 | 0.046091318 |
| GO_REGULATION_OF_RESPONSE_TO_DNA_DAMAGE_STIMULUS | 99 | 0.42916715 | 1.5878026 | 0.003916449 | 0.04614249 |
| GO_ZYMOGEN_ACTIVATION | 40 | 0.4961847 | 1.5871564 | 0.010852713 | 0.04615665 |
| GO_AMYLOID_BETA_FORMATION | 26 | 0.550562 | 1.587503 | 0.012987013 | 0.046159342 |
| GO_DISULFIDE_OXIDOREDUCTASE_ACTIVITY | 29 | 0.52246755 | 1.5876156 | 0.0171875 | 0.04617293 |
| GO_CELLULAR_PIGMENTATION | 25 | 0.5527779 | 1.5872077 | 0.01650165 | 0.04619347 |
| GO_MHC_PROTEIN_BINDING | 23 | 0.56611 | 1.5865935 | 0.009646302 | 0.046325415 |
| GO_REGULATION_OF_RUFFLE_ASSEMBLY | 20 | 0.58554715 | 1.5861813 | 0.014240506 | 0.04638301 |
| REACTOME_SIGNALING_BY_INTERLEUKINS | 271 | 0.38580447 | 1.5862364 | 0 | 0.04642517 |
| GO_AMINO_ACID_TRANSMEMBRANE_TRANSPORTER_ACTIVITY | 39 | -0.5032964 | -1.755011 | 0.003215434 | 0.046740837 |
| PID_ILK_PATHWAY | 29 | 0.53178185 | 1.5848733 | 0.018691588 | 0.04689887 |
| KEGG_BASAL_TRANSCRIPTION_FACTORS | 21 | 0.55800587 | 1.5837731 | 0.020547945 | 0.047252424 |
| GO_REGULATION_OF_RECEPTOR_MEDIATED_ENDOCYTOSIS | 61 | 0.46183455 | 1.5838549 | 0.00591716 | 0.04728276 |
| GO_PROTEIN_LOCALIZATION_TO_LYSOSOME | 23 | 0.5553502 | 1.5831027 | 0.022187004 | 0.04747176 |
| GO_IRE1_MEDIATED_UNFOLDED_PROTEIN_RESPONSE | 39 | 0.49907082 | 1.5820284 | 0.012084592 | 0.04768508 |
| GO_REGULATION_OF_TELOMERASE_ACTIVITY | 23 | 0.5518105 | 1.5820633 | 0.020833334 | 0.04773614 |
| GO_TRANSCRIPTION_COUPLED_NUCLEOTIDE_EXCISION_REPAIR | 40 | 0.49606597 | 1.5817455 | 0.009375 | 0.047749825 |
| GO_REGULATION_OF_EPITHELIAL_CELL_DIFFERENTIATION_INVOLVED_IN_KIDNEY_DEVELOPMENT | 15 | 0.6289511 | 1.5815632 | 0.024096385 | 0.047764853 |
| GO_POSITIVE_REGULATION_OF_CELLULAR_AMIDE_METABOLIC_PROCESS | 88 | 0.43384886 | 1.5821308 | 0 | 0.047769453 |
| GO_NEGATIVE_REGULATION_OF_MACROAUTOPHAGY | 17 | 0.5945564 | 1.5822417 | 0.016528925 | 0.047802966 |
| GO_PROTEIN_LOCALIZATION_TO_NUCLEUS | 153 | 0.40493637 | 1.5805938 | 0 | 0.048071727 |
| GO_NEGATIVE_REGULATION_OF_PROTEIN_CATABOLIC_PROCESS | 72 | 0.4501517 | 1.5806586 | 0.005665722 | 0.048104003 |
| GO_TETRAPYRROLE_BINDING | 88 | -0.4289933 | -1.74575 | 0.003831418 | 0.04824443 |
| GO_LONG_TERM_MEMORY | 20 | -0.5824333 | -1.750679 | 0.002590674 | 0.048282094 |
| BIOCARTA_EDG1_PATHWAY | 18 | 0.5911238 | 1.579967 | 0.026845638 | 0.0483208 |

| | | | | | |
|---|---|---|---|---|---|
| GO_HORMONE_BIOSYNTHETIC_PROCESS | 60 | -0.4617131 | -1.746289 | 0 | 0.048325107 |
| GO_AMINO_ACID_TRANSPORT | 79 | -0.433673 | -1.747008 | 0 | 0.048361402 |
| REACTOME_FORMATION_OF_TUBULIN_FOLDING_INTERMEDIATES_BY_CCT_TRIC | 19 | 0.5777201 | 1.5796008 | 0.031932775 | 0.048449222 |
| HALLMARK_UV_RESPONSE_DN | 103 | 0.42234525 | 1.5792805 | 0.004048583 | 0.048525326 |
| GO_TUMOR_NECROSIS_FACTOR_SUPERFAMILY_CYTOKINE_PRODUCTION | 97 | 0.42169037 | 1.5791173 | 0.00136612 | 0.048526753 |
| GO_PRERIBOSOME | 36 | 0.51569194 | 1.578897 | 0.014446228 | 0.048567295 |
| GO_REGULATION_OF_NECROTIC_CELL_DEATH | 15 | 0.60218567 | 1.5785763 | 0.025423728 | 0.048621394 |
| GO_PROTEIN_REFOLDING | 24 | 0.54771054 | 1.5782032 | 0.017001545 | 0.048670348 |
| REACTOME_PHASE_I_FUNCTIONALIZATION_OF_COMPOUNDS | 69 | -0.4456147 | -1.747073 | 0 | 0.04869397 |
| GO_SPINDLE | 172 | 0.3999209 | 1.5782253 | 0 | 0.048729435 |
| GO_REGULATION_OF_ATPASE_ACTIVITY | 57 | 0.46476227 | 1.577801 | 0.001477105 | 0.04879429 |
| GO_REGULATION_OF_KERATINOCYTE_DIFFERENTIATION | 19 | -0.5950397 | -1.74856 | 0.007334963 | 0.048799325 |
| GO_HIPPO_SIGNALING | 22 | 0.565595 | 1.5775759 | 0.015974442 | 0.04885048 |
| GO_REGULATION_OF_CELLULAR_PROTEIN_CATABOLIC_PROCESS | 142 | 0.410157 | 1.577326 | 0 | 0.04888769 |
| GO_APICAL_JUNCTION_ASSEMBLY | 36 | 0.50050646 | 1.5766693 | 0.017054264 | 0.048892356 |
| GO_NEGATIVE_REGULATION_OF_INNATE_IMMUNE_RESPONSE | 38 | 0.49379528 | 1.5769926 | 0.012403101 | 0.048928153 |
| KEGG_APOPTOSIS | 59 | 0.45298764 | 1.5768766 | 0.005873715 | 0.0489317 |
| GO_ENDODERM_FORMATION | 28 | 0.5335855 | 1.5770847 | 0.016447369 | 0.04893599 |
| REACTOME_O_LINKED_GLYCOSYLATION_OF_MUCINS | 31 | 0.5168904 | 1.5766835 | 0.020155039 | 0.048946146 |
| GO_CELL_FATE_DETERMINATION | 23 | -0.5682038 | -1.747183 | 0.005141388 | 0.048999537 |
| REACTOME_VESICLE_MEDIATED_TRANSPORT | 400 | 0.37280664 | 1.5761992 | 0 | 0.04906452 |
| REACTOME_TCF_DEPENDENT_SIGNALING_IN_RESPONSE_TO_WNT | 86 | 0.4315425 | 1.575816 | 0.004076087 | 0.049128972 |
| GO_REGULATION_OF_MITOTIC_CELL_CYCLE | 330 | 0.3747898 | 1.5759101 | 0 | 0.049150724 |
| GO_PEPTIDE_BIOSYNTHETIC_PROCESS | 306 | 0.3780835 | 1.575217 | 0 | 0.04933479 |
| KEGG_NITROGEN_METABOLISM | 18 | -0.6091809 | -1.742683 | 0 | 0.049377173 |
| REACTOME_O_LINKED_GLYCOSYLATION | 51 | 0.47208104 | 1.5747064 | 0.007173601 | 0.049504288 |
| GO_TRANS_GOLGI_NETWORK | 117 | 0.41975677 | 1.5745336 | 0 | 0.049515404 |
| GO_POSITIVE_REGULATION_OF_INTERLEUKIN_8_PRODUCTION | 40 | 0.4945122 | 1.5737171 | 0.012139605 | 0.04975425 |
| GO_MEMBRANE_RAFT_ORGANIZATION | 19 | 0.5827545 | 1.5738322 | 0.024875622 | 0.049761828 |
| PID_CDC42_PATHWAY | 42 | 0.491159 | 1.5734025 | 0.013274336 | 0.04983011 |
| KEGG_FOCAL_ADHESION | 139 | 0.40867123 | 1.5730376 | 0.002541296 | 0.049936485 |

### Table S3. Blue

| Upstream Regulator | Expr Log Ratio | Molecule Type | Predicted Activation State | Bias-corrected z-score | Activation z-score | Bias Term | p-value of overlap | B-H corrected p-value | Target Molecules in Dataset |
|---|---|---|---|---|---|---|---|---|---|
| IL1B | 0,005 | cytokine | Inhibited | -3,891 | -5,500 | -0,225 | 5,46E-32 | 1,19E-28 | ADM,ATF3,BTG2,CCN2,CD55,CD83,CDKN1A,CEBPB,CEBPD,CHI3L1,CISH,CYP27B1,DDX5,DUSP1,DUSP5,EFNA1,EGR1,ETS2,F3,FKBP5,FOS,FOSB,FOXO1,GADD45A,GADD45B,GDF15,GEM,HBEGF,HES1,HNF1B,HOMER1,IER2,IER3,IFIT3,IFRD1,IL6R,JUN,JUNB,JUND,KLF10,LDLR,LIFR,MCL1,NAMPT,NFIL3,NFKBIA,NR4A1,NR4A3,PIM1,PTP4A1,RCAN1,RHOB,SERPINE1,SLC20A1,SMAD7,SOCS2,TSC22D3,UAP1,VEGFA,ZFP36 |
| beta-estradiol | | chemical - endogenous mammalian | Inhibited | -2,462 | -4,023 | -0,193 | 1,87E-30 | 2,05E-27 | ACKR3,ADM,ATF3,BHLHE40,BTG2,CCN2,CCNA2,CD55,CDKN1A,CDKN1C,CEBPB,CEBPD,CHGB,CHI3L1,CITED2,CYP51A1,DLG3,DUSP1,DUSP5,DUSP6,EFNA1,EGR1,ERF,F3,FOS,FOSB,FOSL2,FOXO1,GADD45A,GADD45B,GEM,GUCY1A1,HBEGF,HES1,HMGCR,HNF1B,HOMER1,IARS1,IER2,IER3,IFRD1,IL6R,JAG1,JUN,JUNB,JUND,KLF10,KLF6,KLF9,LDLR,MAP2K3,MCM2,NAMPT,NEDD9,NFKBIA,NR4A1,NR4A3,PDK4,PPP1R15A,PPP1R3C,PTP4A1,RBBP8,RGS2,RGS3,RND3,RYBP,SERPINE1,SGK1,SLC20A1,SLC38A2,SMAD7,SOCS2,SPRY2,SRSF2,TNFRSF1A,TRIB1,TSC22D3,VEGFA,YWHAZ,ZFP36,ZNF638 |
| TNF | 0,072 | cytokine | Inhibited | -4,904 | -6,526 | -0,201 | 6,91E-30 | 6,03E-27 | ACKR3,ADM,ATF3,BHLHE40,BTG2,BTG3,CCN2,CD55,CD83,CDKN1A,CEBPB,CEBPD,CHI3L1,CISH,CITED2,CRY1,CYP27B1,DDX5,DUSP1,DUSP5,DUSP6,EFNA1,EGR1,ETS2,F3,FOS,FOSB,FOSL2,FOXO1,FZD5,GADD45A,GADD45B,GCLM,GDF15,GEM,HBEGF,HES1,HMGCR,IER2,IER3,IFIT3,JAG1,JUN,JUNB,JUND,KLF10,KLF5,KLF6,LDLR,LIFR,MAFF,MAP2K3,MCL1,NAMPT,NEDD9,NFKBIA,NR4A1,NR4A3,PIM1,PPP1R15A,PPP1R3C,RCAN1,RGS2,RGS3,RHOB,RND3,RRAD,SERPINE1,SGK1,SLC20A1,SMAD7,SOCS2,TFRC,TNFRSF1A,TSC22D3,VEGFA,VMP1,ZFP36 |
| TGFB1 | | growth factor | Inhibited | -4,599 | -5,911 | -0,186 | 4,26E-25 | 1,69E-22 | ADM,BHLHE40,CAVIN2,CCN2,CCNA2,CD55,CD83,CDKN1A,CDKN1C,CEBPB,CHI3L1,CITED2,CYP27B1,DDX5,DUSP1,EGR1,ERF,F3,FERMT2,FILIP1L,FOS,FOSB,FOSL2,FOXO1,GADD45A,GADD45B,GDF15,GEM,GNA13,HBEGF,HES1,HNRNPDL,IARS1,IDI1,IER2,IER3,IFIT3,IFRD1,IL6R,JAG1,JUN,JUNB,JUND,KLF10,KLF9,LDLR,LIFR,MAP2K3,MCM2,NAMPT,NEDD9,NFKBIA,NR4A1,NR4A3,PIM1,PPID,PPP1R3C,RHOB,RRAD,RYBP,SERPINE1,SGK1,SLC20A1,SLC39A14,SMAD7,SRSF2,TFRC,TSC22D3,TUBB2A,VEGFA,ZFP36 |
| CREB1 | | transcription regulator | Inhibited | -4,207 | -5,328 | -0,200 | 1,06E-24 | 3,85E-22 | ADM,APOLD1,ATF3,BAG3,BHLHE40,BTG2,CCNA2,CDKN1A,CEBPB,CEBPD,CHGB,CYP51A1,DUSP1,EGR1,FOS,FOSB,GADD45A,GADD45B,GEM,HECA,HMGCR,HOMER1,IDI1,IER2,IER3,JUN,JUNB,KLF5,LDLR,MCL1,NFIL3,NR4A1,NR4A3,PER1,PPP1R15A,RGS2,SGK1,SOCS2,TFRC,TIPARP,VEGFA,ZFP36 |
| KRAS | -0,059 | enzyme | Inhibited | -2,356 | -2,364 | -0,002 | 2,39E-24 | 8,01E-22 | ATF3,CAVIN2,CCN2,CCNA2,CDKN1A,CEBPB,CHI3L1,CHSY1,DUSP1,DUSP5,DUSP6,EFNA1,EGR1,EIF5,F3,FOS,FOSB,GADD45A,GCLM,GMNN,GNA13,HBEGF,HES1,HNRNPDL,IER3,JUN,JUNB,KLF6,LIPG,NFKBIA,NR4A1,PIM1,RAB20,RHOB,SLC20A1,SLC25A32,SLCO4A1,SPRY2,TRIB1,VEGFA,VMP1,WEE1 |
| NR3C1 | 0,081 | ligand-dependent nuclear receptor | | -1,211 | -1,503 | -0,064 | 1,29E-22 | 3,30E-20 | ADM,BAG3,BHLHE40,CAVIN2,CD83,CDKN1A,CDKN1C,CEBPB,CHIC2,DDX5,DUSP1,EFNA1,EGR1,F3,FKBP5,FOS,FOXO1,GADD45A,GADD45B,GCLM,GEM,IER2,IER3,IFIT2,JADE1,JUN,JUND,KLF9,MCL1,NAMPT,NFIL3,NFKBIA,PER1,PPP1R15A,RGS2,RHOB,SERPINE1,SGK1,SLC19A2,TIPARP,TNFRSF1A,TSC22D3,YWHAZ |
| FSH | | complex | | -0,830 | -1,566 | -0,178 | 7,20E-21 | 1,49E-18 | ADM,BTG2,CCN2,CD55,CDKN1C,DUSP1,EFNA1,ELL2,FILIP1L,FKBP5,FOS,FOSB,FOSL2,FOXO1,GATA6,GEM,IL6R,JUN,JUNB,KLF10,LDLR,MAP2K3,MCL1,NR4A1,PTP4A1,RHOB,SERPINE1,SGK1,SMAD7,TFRC,TRIB1,VEGFA,ZNF331 |
| CREM | | transcription regulator | Inhibited | -2,453 | -2,951 | -0,116 | 1,01E-20 | 1,92E-18 | APOLD1,ATF3,BHLHE40,BTG2,CEBPB,CYP51A1,DUSP1,EGR1,FOS,GADD45B,HMGCR,IDI1,JUNB,KLF5,LDLR,MCL1,NFIL3,NR4A1,PER1,PPP1R15A,RHOB,TIPARP,VEGFA |
| TP53 | 0,089 | transcription regulator | Inhibited | -3,479 | -3,924 | -0,067 | 4,10E-19 | 6,17E-17 | ATF3,BHLHE40,BTG2,CCN2,CCNA2,CCNL1,CDKN1A,CEBPB,CEBPD,CITED2,CYP51A1,DDX3X,DDX5,DUSP1,DUSP5,DUSP6,EFNA1,EGR1,EIF2AK3,EIF5,ERF,F3,FERMT2,FKBP5,FOS,FOXO1,GADD45A,GADD45B,GATA6,GDF15,GMNN,GNA13,HBEGF,HMGCR,IER3,JUN,JUNB,JUND,KLF6,LDLR,MAP2K3,MCL1,MCM2,MLF2,NAMPT,NFKBIA,PIM1,PPP1R15A,PTP4A1,RHOB,RND3,RRAD,SERPINE1,SGK1,SLC19A2,SMAD7,SOCS2,TFRC,TNFRSF1A,TSC22D3,VEGFA,VMP1,WEE1,ZFP36 |
| IFNG | | cytokine | Inhibited | -2,238 | -3,894 | -0,233 | 4,39E-19 | 6,39E-17 | ADM,ARG2,ATF3,CCN2,CCNA2,CD55,CD83,CDKN1A,CEBPB,CEBPD,CISH,DDX5,DUSP1,DUSP5,EGR1,ETS2,F3,FKBP5,FOS,FOSB,FOXO1,GDF15,GNA13,HBEGF,HMGCR,IDI1,IER2,IER3,IFIT2,IFIT3,IL6R,JAG1,JUN,JUNB,JUND,KLF10,KLF6,LIFR,MAFF,NAMPT,NEDD9,NFKBIA,PCDH17,PIM1,RAB20,RHOB,SERPINE1,SMAD7,SOCS2,TFRC,TNFRSF1A,TSC22D3,VEGFA,ZFP36,ZNF638 |

| | | | | | | | | | |
|---|---|---|---|---|---|---|---|---|---|
| INS | 0,009 | other | Inhibited | -2,187 | -2,510 | -0,085 | 4,13E-18 | 5,30E-16 | ADM,CCN2,CDKN1A,CEBPB,CITED2,DUSP1,EGR1,F3,FOS,FOSB,FOSL2,FOXO1,GABARAPL1,HMGCR,IER2,IER3,JUN,JUNB,JUND,NR4A1,VEGFA |
| Growth hormone | | group | Inhibited | -1,330 | -2,448 | -0,241 | 7,49E-18 | 8,88E-16 | CEBPB,CEBPD,CISH,CITED2,CYP27B1,EGR1,FKBP5,FOS,FOSL2,FZD5,GDF15,IER3,JUN,JUNB,KLF10,LDLR,NAMPT,NEDD9,NFKBIA,PDK4,PPP1R15A,SGK1,SOCS2,YWHAZ |
| IGF2 | | growth factor | | -0,664 | -1,103 | -0,225 | 2,88E-17 | 3,07E-15 | CCN2,CDKN1A,CRY1,DUSP1,EGR1,GADD45A,GATA6,HBEGF,IER2,IER3,JUNB,KLF9,MCM2,NFKBIA,PIM1,RHOB,SERPINE1,SLC20A1,SLC38A2,VEGFA,ZFAND5 |
| ESR1 | 0,031 | ligand-dependent nuclear receptor | | 0,739 | 0,258 | -0,085 | 1,09E-16 | 1,06E-14 | ACKR3,ATF3,CCNA2,CD55,CD83,CDKN1A,CDKN1C,CEBPB,CEBPD,DUSP5,EFNA1,EGR1,F3,FAM102A,FOS,FOSB,FOSL2,FOXO1,GADD45B,GDF15,GEM,GNA13,GUCY1A1,HBEGF,HMGCR,IER3,IFRD1,JUN,JUNB,JUND,LDLR,LIFR,NEDD9,NFKBIA,PDK4,PPP1R3C,RGS3,RND3,SERPINE1,SGK1,SLC38A2,SOCS2,STEAP1,TACC1,TFRC,TRIB1,TSC22D3,TSPYL2,VEGFA,WEE1 |
| progesterone | | chemical - endogenous mammalian | Inhibited | -1,647 | -2,385 | -0,143 | 3,95E-16 | 3,38E-14 | ADM,ATF3,BHLHE40,CCNA2,CDKN1A,CEBPB,CEBPD,CITED2,EFNA1,EGR1,ELL2,F3,FKBP5,FOS,FOXO1,GABARAPL1,HBEGF,HMGCR,IL6R,JUN,KLF10,KLF5,KLF9,MAP2K3,MCM2,NEDD9,NFIL3,NFKBIA,NR4A1,SGK1,STEAP1,TIPARP,TNFRSF1A,VEGFA |
| NFKBIA | -0,778 | transcription regulator | Inhibited | -2,130 | -2,939 | -0,164 | 6,56E-16 | 5,30E-14 | ACKR3,BTG2,CCNA2,CDKN1A,CEBPB,CEBPD,CHI3L1,DDX3X,DUSP6,F3,FOS,GADD45A,GADD45B,HES1,IER3,JUN,JUNB,KLF5,LIFR,NFKBIA,NR4A1,PIM1,RCAN1,RGS3,SGK1,TFRC,VEGFA,YWHAZ,ZNF638 |
| Akt | | group | | 1,054 | 0,160 | -0,194 | 1,12E-15 | 8,85E-14 | ADM,AMOT,BHLHE40,BTG2,CCN2,CDKN1A,CDKN1C,CEBPB,F3,FKBP5,FOS,FOXO1,HES1,IER3,JUN,MCL1,NFIL3,NFKBIA,PDK4,RHOB,SERPINE1,SGK1,TIPARP,VEGFA |
| ADRB | | group | | -0,161 | 0,639 | 0,191 | 3,64E-15 | 2,69E-13 | ATF3,BHLHE40,CAVIN2,CCNA2,CDKN1A,CYP51A1,FOS,GADD45A,GCLM,GDF15,IFIT3,JUN,JUNB,NFKBIA,NR4A1,NR4A3,SLCO4A1,WEE1 |
| FAS | -0,014 | transmembrane receptor | Activated | 2,088 | 2,197 | 0,036 | 6,66E-15 | 4,47E-13 | BTG3,CCN2,CD83,CDKN1A,CEBPD,EFNA1,EGR1,F3,FOS,FOSB,FOXO1,IER3,JUN,JUND,LDLR,NFKBIA,NR4A1,PPP1R15A,PTP4A1,RGS3,SERPINE1,TFRC,TNFRSF1A,TRIB1,VEGFA,ZFP36 |
| prostaglandin E2 | | chemical - endogenous mammalian | Inhibited | -1,919 | -2,805 | -0,212 | 9,06E-15 | 5,99E-13 | ATF3,CCN2,CD55,CD83,CDKN1A,CEBPB,CEBPD,DUSP1,EGR1,FOS,FOSB,FOSL2,FOXO1,IER3,JUN,MCL1,NEDD9,NFIL3,NFKBIA,NR4A1,RGS2,SOCS2,TSC22D3,VEGFA |
| NRG1 | | growth factor | Inhibited | -1,400 | -2,026 | -0,244 | 1,17E-14 | 7,61E-13 | CCN2,CDKN1A,DUSP1,DUSP6,EGR1,FOS,FOSB,HES1,HMGCR,IDI1,IER2,IER3,JUN,JUNB,JUND,LIFR,MCL1,NR4A1,NR4A3,VEGFA,ZFP36 |
| FOXO3 | -0,179 | transcription regulator | Inhibited | -3,163 | -3,567 | -0,090 | 2,38E-14 | 1,46E-12 | CCN2,CDKN1A,CDKN1C,EGR1,FOS,FOSB,FOXO1,GABARAPL1,GADD45A,GADD45B,GMNN,HMGCR,IARS1,IER3,JAG1,JUNB,MCM2,NAMPT,NFKBIA,PPP1R15A,SERPINE1,SGK1,TNFRSF1A,VEGFA |
| OSM | | cytokine | Inhibited | -3,500 | -4,701 | -0,230 | 2,49E-14 | 1,49E-12 | ARF6,ATF3,BAIAP2,BHLHE40,CDKN1A,CEBPD,CHD1,CHI3L1,CISH,CRY1,EGR1,ETS2,F3,FOS,FOSB,HBEGF,IARS1,IER3,IL6R,JAG1,JUN,JUNB,KLF10,LDLR,LIFR,NAMPT,PTP4A1,SERPINE1,UAP1,VEGFA,ZFP36 |
| Raf | | group | | -1,518 | -1,948 | -0,221 | 1,13E-13 | 6,18E-12 | CDKN1A,DUSP6,EGR1,FOS,GDF15,IER2,IER3,JUN,MAFF,MCL1,RND3,SLC20A1,SPRY2 |
| MAPK1 | -0,012 | kinase | Inhibited | -3,694 | -3,730 | -0,007 | 1,27E-13 | 6,84E-12 | ARG2,ATF3,CCN2,CDKN1A,CISH,DUSP1,EGR1,FOS,FOSB,FOSL2,GMNN,HOMER1,IFIT2,IFIT3,JUN,JUNB,JUND,MCL1,NR4A1,RGS2,SOCS2,SPRY2,UTP18,ZNF184 |
| Insulin | | group | Inhibited | -2,774 | -3,579 | -0,168 | 2,37E-13 | 1,22E-11 | ARG2,ATF3,CCN2,CDKN1A,CEBPB,DUSP1,EGR1,FOS,FOSB,FOXO1,HES1,HMGCR,IFIT2,IFIT3,JUN,NFIL3,NFKBIA,NR4A1,NR4A3,PDK4,PPP1R15A,SERPINE1,SGK1,SLC38A2,SLC39A14,SMAD7,VEGFA |
| TP63 | | transcription regulator | Inhibited | -2,228 | -3,246 | -0,246 | 2,38E-13 | 1,22E-11 | ADM,ADRM1,BHLHE40,CCNA2,CDKN1A,CDKN1C,CITED2,DUSP6,F3,FOS,GADD45A,HBEGF,HES1,IER3,JAG1,JUNB,KLF6,MAFF,MAP2K3,RBBP8,SERPINE1,TNFRSF1A,VEGFA,VMP1,WEE1 |
| FOXO4 | -0,086 | transcription regulator | Inhibited | -2,453 | -2,145 | 0,090 | 4,70E-13 | 2,28E-11 | CCN2,CDKN1A,CYP51A1,GADD45A,GADD45B,HMGCR,IDI1,IER3,JAG1,NAMPT,SERPINE1,SGK1,VEGFA,ZFAND5 |
| CEBPA | 0,144 | transcription regulator | | -0,365 | -1,172 | -0,196 | 4,93E-13 | 2,36E-11 | ARG2,BTG2,CCNA2,CDKN1A,CEBPB,CEBPD,CHI3L1,FOS,FOXO1,GADD45A,GATA6,HMGCR,HNF1B,IER3,IL6R,JUN,JUNB,KLF5,NFIL3,PPP1R3C,RGS2,SERPINE1,TNFRSF1A,TRIB1,TSC22D3,TUBB2A |
| Mek | | group | Inhibited | -2,561 | -3,265 | -0,200 | 1,37E-12 | 5,84E-11 | CCN2,CDKN1A,DUSP5,DUSP6,EGR1,FOS,GDF15,HOMER1,IER2,IER3,JUN,JUNB,MAFF,MCL1,NR4A1,RND3,SERPINE1,SLC20A1,SPRY2 |
| HRAS | -0,041 | enzyme | | -0,581 | -1,353 | -0,177 | 2,50E-12 | 1,03E-10 | ADM,ARF6,BTG2,CCN2,CCNA2,CDKN1A,CEBPB,CEBPD,CISH,DUSP1,DUSP6,EGR1,FERMT2,FOS,FOSB,FOSL2,FOXO1,GNA13,HES1,JUN,JUNB,KLF5,KLF6,LIPG,NFKBIA,RHOB,SERPINE1,SOCS2,SPRY2,VEGFA |
| NUPR1 | 0,033 | transcription regulator | Inhibited | -4,808 | -3,922 | 0,174 | 6,38E-12 | 2,42E-10 | ADM,ATF3,AVPI1,CCNA2,CEBPB,CITED2,DUSP5,ELL2,GADD45A,GDF15,HBEGF,IFIT2,IL6R,JOSD1,KLF6,LMAN2L,NFIL3,PIM1,PPP1R15A,RAB20,RYBP,SERPINE1,SPTSSA,TRIB1,UAP1,WDCP |

| | | | | | | | | | |
|---|---|---|---|---|---|---|---|---|---|
| ethanol | | chemical - endogenous mammalian | Inhibited | -1,485 | -2,214 | -0,164 | 6,75E-12 | 2,51E-10 | ACKR3,ADM,ATF3,CCN2,CDKN1A,CEBPB,CEBPD,DUSP1,DUSP6,EGR1,FOS,FOSB,FOSL2,GADD45A,JUN,KLF6,PPP1R3C,PRKRA,RHOB,SERPINE1,SGK1,TSC22D3,WEE1 |
| tretinoin | | chemical - endogenous mammalian | | -0,206 | -1,384 | -0,195 | 1,01E-11 | 3,74E-10 | ADM,BHLHE40,BTG2,CCN2,CCNA2,CDKN1A,CEBPB,CHGB,CITED2,DNAAF2,DUSP1,EGR1,EIF5,F3,FOS,FOSL2,GATA6,GDF15,GMNN,GNA13,HBEGF,HES1,HNF1B,IER3,IFIT2,IFIT3,IL6R,JUN,KLF5,KLF9,MAP2K3,MCL1,MLF2,NDUFAF4,PIM1,PNRC1,RAE1,SERPINE1,SLC38A2,SMAD7,TFRC,TNFRSF1A,TSC22D3,VEGFA,VMP1,YWHAZ,ZFAND5 |
| D-glucose | | chemical - endogenous mammalian | Inhibited | -2,387 | -3,226 | -0,173 | 1,31E-11 | 4,77E-10 | ATF3,CCN2,CD55,CDKN1A,CEBPB,DUSP1,EGR1,EIF2AK3,FOS,GABARAPL1,GDF15,HMGCR,IER2,JAG1,JUN,JUNB,KLF10,LDLR,MCL1,NAMPT,NR4A1,NR4A3,RGS2,SERPINE1,SGK1,SLC39A14,SRSF2,TFRC,VEGFA |
| MAPK8 | 0,000 | kinase | Inhibited | -1,370 | -2,230 | -0,226 | 1,48E-11 | 5,29E-10 | CCN2,CCNA2,CDKN1A,CEBPB,DUSP1,DUSP6,EGR1,FOS,FOSL2,FOXO1,GADD45A,JUN,JUND,KLF6,TSC22D3,ZFP36 |
| CDKN2A | | transcription regulator | | -0,288 | -0,082 | 0,054 | 1,85E-11 | 6,41E-10 | BTG2,BTG3,CCN2,CCNA2,CDKN1A,CEBPB,CEBPD,CITED2,DDX5,DUSP1,EFNA1,EGR1,FOS,GNA13,JUN,MCL1,SERPINE1,SLBP,TNFRSF1A,VEGFA |
| LIF | -0,118 | cytokine | Inhibited | -2,483 | -3,059 | -0,153 | 5,75E-11 | 1,86E-09 | CCNA2,CDKN1A,CEBPB,CEBPD,CISH,EGR1,FOS,GATA6,HBEGF,HES1,JUN,JUNB,KLF10,LIFR,PIM1,SOCS2,VEGFA |
| nitric oxide | | chemical - endogenous mammalian | Inhibited | -2,120 | -2,861 | -0,214 | 6,35E-11 | 2,04E-09 | ATF3,CCN2,CDKN1A,DUSP1,DUSP6,EGR1,FOS,GADD45A,GCLM,IER3,JUN,JUNB,SGK1,TFRC,VEGFA |
| MAPK9 | | kinase | | 0,113 | -0,758 | -0,248 | 9,04E-11 | 2,84E-09 | CCN2,CDKN1A,CEBPB,EGR1,FOS,FOSL2,GADD45A,GADD45B,IER2,JUN,JUNB,JUND,KLF6,PPP1R15A,ZFP36 |
| GH1 | | growth factor | Inhibited | -2,559 | -2,907 | -0,103 | 1,31E-10 | 4,00E-09 | ATF3,CEBPB,CEBPD,CISH,EGR1,FOS,GADD45A,JUN,JUNB,LDLR,SOCS2,TNFRSF1A,ZFP36 |
| L-triiodothyronine | | chemical - endogenous mammalian | | 1,640 | 0,897 | -0,173 | 3,26E-10 | 9,22E-09 | ADM,CCN2,CDKN1A,CHD1,CYP27B1,DUSP1,DUSP5,EFNA1,EGR1,ETF1,FOS,GDF15,HOMER1,KLF9,LDLR,NR4A1,PDK4,PER1,RGS2,SERPINE1,SMAD7 |
| PTEN | 0,088 | phosphatase | | 1,236 | 1,301 | 0,015 | 3,47E-10 | 9,75E-09 | ADM,ATF3,BTG2,CDKN1A,CEBPB,CITED2,DUSP1,ETS2,F3,FOS,FOXO1,HMGCR,IDI1,IFRD1,JAG1,JUND,KLF5,KLF6,MCL1,NFIL3,PIM1,SERPINE1,SLC20A1,SPRY2,STEAP1,TSC22D3,VEGFA |
| KLF2 | -0,138 | transcription regulator | Activated | 1,999 | 2,344 | 0,099 | 3,95E-10 | 1,10E-08 | ACKR3,ADM,CCN2,CD55,CDKN1A,EFNA1,F3,GADD45A,IL6R,JUNB,NFKBIA,RGS3,SERPINE1,TFRC,WEE1 |
| MYC | -0,549 | transcription regulator | | 0,270 | -0,724 | -0,205 | 5,51E-10 | 1,47E-08 | ADM,CCNA2,CDKN1A,CEBPD,DDX3X,DDX5,DUSP1,DUSP5,DUSP6,EGR1,F3,FKBP5,FOS,GADD45A,GADD45B,GATA6,GCLM,HES1,IER3,IFIT2,IFIT3,JUN,KLF10,KLF6,MCL1,MFAP1,NFKBIA,PPID,RHOB,SERPINE1,SGK1,SRSF2,TFRC,VEGFA,YRDC,ZFP36 |
| TP73 | 0,061 | transcription regulator | Inhibited | -1,343 | -2,103 | -0,170 | 7,39E-10 | 1,91E-08 | ATF3,BHLHE40,BTG2,CCNA2,CD55,CD83,CDKN1A,CDKN1C,EGR1,GADD45A,HBEGF,HES1,IER3,JAG1,KLF5,MIR22HG,PNRC1,SERPINE1,TNFRSF1A,VEGFA,VMP1 |
| NELFB | -0,014 | other | Activated | 2,212 | 2,449 | 0,097 | 7,56E-10 | 1,93E-08 | ATF3,CCN2,CDKN1A,GADD45A,GADD45B,VEGFA |
| ESR2 | | ligand-dependent nuclear receptor | | -1,115 | -1,373 | -0,093 | 7,81E-10 | 1,98E-08 | ATF3,CCN2,CCNA2,CDKN1A,CDKN1C,CEBPD,EGR1,FOS,FOXO1,GADD45B,JAG1,JUNB,MAP2K3,NEDD9,NFKBIA,PDK4,PPP1R3C,SERPINE1,SGK1,SMAD7,SOCS2,TRIB1,VEGFA |
| ID2 | -0,211 | transcription regulator | Inhibited | -2,162 | -2,111 | 0,015 | 8,33E-10 | 2,09E-08 | ADM,BTG2,CD83,CDKN1A,CEBPB,DUSP1,ETS2,FOXO1,GADD45B,IL6R,KLF6,NR4A1,NR4A3,SMAD7,TNFRSF1A |
| LEP | 0,022 | growth factor | | -0,592 | -1,551 | -0,232 | 8,86E-10 | 2,20E-08 | CCNA2,CDKN1A,CEBPB,CYP27B1,DUSP6,EGR1,F3,FOS,GADD45A,HES1,HMGCR,JUN,JUNB,JUND,LDLR,LIFR,MCL1,NAMPT,SERPINE1,TNFRSF1A,VEGFA,ZFP36 |
| TGFB2 | | growth factor | Inhibited | -1,414 | -2,108 | -0,226 | 1,16E-09 | 2,80E-08 | ALX1,BHLHE40,CCN2,EGR1,FOS,HBEGF,JAG1,LIPG,PDK4,RGS3,SERPINE1,VEGFA |
| ID3 | 0,127 | transcription regulator | | -2,066 | -1,943 | 0,038 | 1,37E-09 | 3,26E-08 | ADM,BTG2,CD83,CDKN1A,CEBPB,DUSP1,ETS2,FOXO1,GADD45B,IL6R,KLF6,NR4A1,NR4A3,SMAD7,TNFRSF1A |
| CSF3 | 0,069 | cytokine | Inhibited | -2,358 | -2,909 | -0,162 | 2,45E-09 | 5,48E-08 | CEBPB,CISH,DUSP6,EGR1,FOS,GADD45A,GADD45B,JUN,MCL1,NFIL3,PIM1,TFRC,TNFRSF1A,VEGFA |
| BDNF | 0,075 | growth factor | Inhibited | -2,235 | -3,055 | -0,235 | 2,60E-09 | 5,78E-08 | BAG3,BHLHE40,CDKN1A,DUSP1,DUSP6,EGR1,FOS,FOSB,HMGCR,HOMER1,IFRD1,JUN,KLF10,NR4A1,NR4A3,SPRY2,TFRC,TIPARP |
| AHR | -0,077 | ligand-dependent nuclear receptor | | -1,697 | -1,921 | -0,068 | 3,42E-09 | 7,46E-08 | ADM,ARG2,BTG2,CCNA2,CDKN1A,DUSP6,FOS,GADD45A,HES1,HMGCR,JAG1,JUN,JUNB,NEDD9,PPP1R15A,SERPINE1,TIPARP,TSPYL2,VEGFA |
| IL10 | | cytokine | | -0,679 | -0,915 | -0,056 | 4,69E-09 | 9,78E-08 | ADM,ARG2,CD55,CD83,CDKN1A,DUSP1,ETS2,F3,FKBP5,FOS,IFIT2,JUN,NEDD9,NR4A3,TFRC,TNFRSF1A,TSC22D3,VEGFA,ZFP36 |

| | | | | | | | | | |
|---|---|---|---|---|---|---|---|---|---|
| SFTPA1 | | transporter | Activated | 2,470 | 3,162 | 0,219 | 5,64E-09 | 1,17E-07 | AMOT,CCN2,EGR1,FOS,FOSB,GUCY1A1,HBEGF,NAMPT,PDK4,SERPINE1 |
| PDX1 | | transcription regulator | Inhibited | -2,385 | -2,885 | -0,145 | 6,16E-09 | 1,26E-07 | ATF3,CHGB,DUSP5,DUSP6,EFNA1,EGR1,F3,GCLM,IER3,JUN,JUNB,KLF10,KLF6,PTP4A1 |
| glutamine | | chemical - endogenous mammalian | | 0,429 | 0,970 | 0,184 | 6,66E-09 | 1,36E-07 | CEBPB,DUSP1,FOS,GADD45A,JUN,JUNB,MCL1,NFKBIA,NR4A1,SRSF2,VEGFA |
| FOS | -1,659 | transcription regulator | | -0,182 | -0,706 | -0,154 | 6,93E-09 | 1,41E-07 | ATF3,ATF7IP,CCN2,CCNA2,CHI3L1,DDX3X,EGR1,F3,FOS,FOSB,FZD5,JUN,JUNB,KLF6,NFIL3,NR4A1,PTP4A1,SERPINE1,SMAD7,TIPARP,VEGFA,WEE1,YWHAZ |
| TNFRSF1A | -0,309 | transmembrane receptor | | -0,069 | -0,523 | -0,139 | 7,15E-09 | 1,44E-07 | CCN2,CD83,CDKN1A,CEBPB,EFNA1,GCLM,IER3,JUN,PIM1,SERPINE1,TFRC,TNFRSF1A,VEGFA |
| VHL | | transcription regulator | | 0,260 | 0,765 | 0,211 | 9,97E-09 | 1,92E-07 | BHLHE40,CCN2,CDKN1A,CDKN1C,EIF5,F3,FILIP1L,IER3,JADE1,NEDD9,TFRC,VEGFA |
| NR3C2 | 0,012 | ligand-dependent nuclear receptor | | -1,303 | -1,723 | -0,142 | 9,97E-09 | 1,92E-07 | CCN2,EGR1,FKBP5,GUCY1A1,JUNB,KLF9,RGS2,RRAD,SERPINE1,SGK1,TSC22D3,VEGFA |
| STAT6 | | transcription regulator | Inhibited | -1,907 | -2,266 | -0,097 | 1,06E-08 | 2,03E-07 | APOLD1,CDKN1A,CISH,DNAAF2,EGR1,FKBP5,GADD45B,HMGCR,IER2,IFIT3,JAG1,LIFR,NEDD9,NFIL3,NFKBIA,PDK4,SERPINE1,SOCS2,ZFP36 |
| Immunoglobulin | | complex | | -0,381 | -0,403 | -0,005 | 1,07E-08 | 2,03E-07 | ADM,CD83,CDKN1C,CEBPD,CISH,CYP27B1,DUSP1,EGR1,EIF2AK3,ETS2,F3,FOS,FOSB,MAFF,NAMPT,NFIL3,NFKBIA,NR4A1,PDK4,RGS2,TFRC,VEGFA |
| SRF | | transcription regulator | | 0,448 | -0,090 | -0,213 | 1,18E-08 | 2,21E-07 | APOLD1,CCN2,CDKN1A,CEBPD,DUSP5,EGR1,FERMT2,FILIP1L,FOS,FOSB,GADD45A,IER2,JUN,JUNB,LDLR,MCL1,NR4A1,RND3 |
| Hdac | | group | Activated | 2,327 | 2,467 | 0,040 | 1,20E-08 | 2,24E-07 | ATF3,CDKN1A,EGR1,FOS,GADD45B,HNF1B,IL6R,JUN,KLF10,KLF6,KLF9,NR4A1,SPRY2 |
| WT1 | -0,178 | transcription regulator | | -1,018 | -1,256 | -0,079 | 1,32E-08 | 2,45E-07 | BTG2,CCN2,CDKN1A,DDX3X,EFNA1,EGR1,FOSB,HBEGF,IDI1,JUNB,JUND,MCL1,SERPINE1,SLC20A1,VEGFA |
| NFATC2 | | transcription regulator | | 1,839 | 1,026 | -0,249 | 1,36E-08 | 2,48E-07 | CCNA2,CD83,CDKN1A,CISH,CITED2,IER3,IFIT2,IFIT3,JUN,NFKBIA,NR4A1,RCAN1,RGS2,SOCS2 |
| SMAD4 | | transcription regulator | Inhibited | -2,724 | -3,527 | -0,228 | 1,75E-08 | 3,12E-07 | CCN2,CDKN1A,CITED2,FOS,GADD45A,GADD45B,IER3,JAG1,NAMPT,NFKBIA,SERPINE1,SGK1,SMAD7,VEGFA,ZFP36 |
| butyric acid | | chemical - endogenous mammalian | | -0,696 | -0,890 | -0,056 | 2,20E-08 | 3,81E-07 | CCNA2,CD83,CDKN1A,CDKN1C,DUSP1,FOS,GADD45A,GATA6,IFIT3,KLF5,MCL1,NAMPT,NDUFAF4,NFKBIA,NR4A1,SERPINE1,SGK1,SRSF2,VEGFA,WEE1 |
| oleic acid | | chemical - endogenous mammalian | | -0,114 | -0,588 | -0,152 | 2,32E-08 | 3,98E-07 | DUSP1,FOS,GADD45A,GADD45B,GDF15,HMGCR,LDLR,NFKBIA,NR4A1,PDK4,SERPINE1 |
| STAT5A | -0,073 | transcription regulator | | -1,444 | -1,803 | -0,122 | 3,04E-08 | 5,07E-07 | CDKN1A,CEBPB,CISH,CYP27B1,DUSP5,FOS,FOSB,FOSL2,GADD45A,IL6R,KLF10,MCL1,PIM1,SOCS2,TNFRSF1A,ZFP36 |
| 15-deoxy-delta-12,14-PGJ 2 | | chemical - endogenous mammalian | | -1,245 | -0,569 | 0,192 | 3,58E-08 | 5,83E-07 | CCN2,CDKN1A,CEBPD,EGR1,FOS,GADD45A,GCLM,GDF15,JUN,MCL1,NFKBIA,SERPINE1,VEGFA |
| arachidonic acid | | chemical - endogenous mammalian | Inhibited | -1,752 | -2,053 | -0,099 | 5,66E-08 | 8,69E-07 | CD83,EGR1,F3,FOS,JUN,KLF6,LDLR,NFKBIA,RGS2,TNFRSF1A |
| HNF4A | 0,020 | transcription regulator | | -0,444 | -0,881 | -0,133 | 6,00E-08 | 9,18E-07 | ARG2,AVPI1,CCNA2,CD55,CDKN1A,CEBPB,CEBPD,CHI3L1,CHIC2,COQ10B,CRY1,CUTC,CYP51A1,DUSP6,EFNA1,EGR1,EIF5,FOSL2,FOXO1,GADD45B,GATA6,GDF15,HBEGF,HNF1B,HNRNPA0,JUN,JUNB,LDLR,LMAN2L,NAMPT,PDK4,POGLUT2,PPP1R3C,SERPINE1,SERTAD3,SGK1,SLC25A32,SPTSSA,SRSF2,TFRC,TMEM140,TXNL4B,UBE2M,UTP18,VMP1,WDCP,WDR13,ZFP36 |
| Il3 | | cytokine | Inhibited | -1,843 | -2,287 | -0,151 | 6,02E-08 | 9,18E-07 | BHLHE40,CDKN1A,CISH,EGR1,FOS,MCL1,NFIL3,PIM1,TNFRSF1A |
| HDAC1 | -0,015 | transcription regulator | | -0,533 | -0,244 | 0,094 | 6,52E-08 | 9,84E-07 | ATF3,CAVIN2,CCNA2,CDKN1A,DUSP1,EGR1,FOS,GABARAPL1,HES1,NFKBIA,RHOB,SERPINE1,SGK1,SMAD7,TSC22D3 |

| | | | | | | | | | |
|---|---|---|---|---|---|---|---|---|---|
| RB1 | -0,015 | transcription regulator | | -0,182 | -0,588 | -0,125 | 6,60E-08 | 9,90E-07 | BTG3,CCN2,CCNA2,CDKN1A,CITED2,CYP27B1,EGR1,F3,FOS,GMNN,HOMER1,MAP2K3,MCM2,MFAP1,PIM1,SERPINE1,SLBP,VEGFA,ZNF638 |
| IL13 | -0,012 | cytokine | Inhibited | -1,459 | -2,448 | -0,246 | 9,12E-08 | 1,32E-06 | ACKR3,ARG2,ATF3,AVPI1,CCN2,CEBPB,CHI3L1,CISH,EGR1,F3,IARS1,JAG1,JUN,KLF6,MCL1,SERPINE1,TFRC,TNFRSF1A |
| Lh | | complex | Inhibited | -2,161 | -2,577 | -0,161 | 2,17E-07 | 2,89E-06 | CD55,CDKN1C,DUSP1,FKBP5,FOS,GEM,MAP2K3,MCL1,PER1,PTP4A1,RHOB,SGK1,TRIB1,VEGFA |
| BRCA1 | | transcription regulator | | 1,153 | 0,578 | -0,178 | 2,43E-07 | 3,20E-06 | CDKN1A,EGR1,FKBP5,FOXO1,GADD45A,GADD45B,GDF15,IFIT2,IFIT3,NEDD9,VEGFA,WEE1 |
| RASSF1 | | other | | -0,484 | -0,765 | -0,096 | 2,91E-07 | 3,77E-06 | BHLHE40,CCNA2,CDKN1A,EGR1,FOS,GDF15,HBEGF,KLF10,TUBB2A |
| SMAD7 | -0,352 | transcription regulator | | 0,428 | 0,919 | 0,164 | 2,99E-07 | 3,86E-06 | CCN2,CCNA2,CDKN1A,CITED2,FOXO1,GADD45A,HBEGF,NFKBIA,SERPINE1,SMAD7,VEGFA |
| palmitic acid | | chemical - endogenous mammalian | Inhibited | -1,304 | -2,103 | -0,235 | 3,59E-07 | 4,52E-06 | ATF3,CCN2,CDKN1A,FOS,FOXO1,GDF15,IL6R,MCL1,NFKBIA,NR4A1,NR4A3,PDK4,SERPINE1 |
| NOTCH1 | | transcription regulator | | -0,360 | -0,592 | -0,078 | 4,19E-07 | 5,22E-06 | CCN2,CDKN1A,CDKN1C,CEBPB,CEBPD,EPHA4,FOS,FOSB,HES1,JAG1,RND3,SERPINE1,SPRY2,VEGFA |
| TCF4 | 0,086 | transcription regulator | | 2,390 | 1,796 | -0,165 | 5,03E-07 | 6,21E-06 | BHLHE40,CCNA2,CD55,CDKN1A,CDKN1C,FOS,FOXO1,HES1,JAG1,JUN,MCL1,NEDD9,PPID,SGK1,SMAD7,TMEM140,YWHAZ |
| NGFR | | transmembrane receptor | | 0,186 | -0,314 | -0,228 | 5,15E-07 | 6,33E-06 | CCNA2,FOS,JUN,LDLR,RHOB,SERPINE1 |
| 15(S)-HETE | | chemical - endogenous mammalian | Inhibited | -2,054 | -2,215 | -0,073 | 6,88E-07 | 8,25E-06 | EGR1,F3,FOSB,HMGCR,JUN |
| estrogen receptor | | group | | 0,352 | 0,389 | 0,011 | 7,88E-07 | 9,36E-06 | CCN2,EGR1,FOS,HBEGF,JAG1,JUN,KLF10,LDLR,NEDD9,RGS2,SERPINE1,TSC22D3,VEGFA |
| dinoprost | | chemical - endogenous mammalian | | -1,383 | -1,939 | -0,216 | 9,24E-07 | 1,08E-05 | ADM,FOS,FOSL2,JUN,NFIL3,NR4A1,VEGFA,YWHAZ |
| levodopa | | chemical - endogenous mammalian | Inhibited | -2,734 | -3,031 | -0,065 | 1,00E-06 | 1,15E-05 | ARG2,DUSP1,DUSP6,ETS2,FKBP5,FOS,FOSB,FOXO1,GABARAPL1,GADD45B,GNA13,GUCY1A1,HBEGF,HOMER1,IER3,JUNB,KLF6,KLF9,PER1,SLC29A3,SLC38A2,VMP1 |
| STAT5B | | transcription regulator | | 0,511 | 0,302 | -0,063 | 1,04E-06 | 1,19E-05 | BAIAP2,CDKN1A,CISH,FOS,FOSL2,GADD45A,GADD45B,MCL1,PDK4,PIM1,PPP1R3C,SOCS2,TNFRSF1A,ZFP36 |
| SETD2 | | enzyme | | 0,206 | 0,816 | 0,249 | 1,10E-06 | 1,24E-05 | CCN2,CITED2,GNA13,JUN,RHOB,SERPINE1 |
| ACVRL1 | 0,049 | kinase | | 1,358 | 1,000 | -0,179 | 1,31E-06 | 1,45E-05 | CDKN1A,EFNA1,GDF15,SERPINE1,SMAD7,VEGFA |
| PPARG | 0,078 | ligand-dependent nuclear receptor | | -1,243 | -1,835 | -0,169 | 1,58E-06 | 1,73E-05 | CAVIN2,CCN2,CDKN1A,CEBPB,GDF15,HES1,HMGCR,JUN,KLF6,LDLR,NR4A1,PDK4,RAB20,RHOB,SERPINE1,SGK1,TSC22D3,VEGFA |
| CNTF | | cytokine | | -0,749 | -0,958 | -0,106 | 1,65E-06 | 1,80E-05 | CEBPD,CISH,EGR1,FOS,JUNB,LIFR,SOCS2,ZFP36 |
| DUSP1 | -1,398 | phosphatase | | 1,506 | 1,614 | 0,037 | 1,98E-06 | 2,12E-05 | DUSP1,DUSP6,GDF15,IER3,IFIT3,JUN,PNRC1,SERPINE1,ZFP36 |
| ERBB4 | 0,074 | kinase | | 0,002 | -0,113 | -0,041 | 2,36E-06 | 2,49E-05 | BTG2,CDKN1A,DUSP1,EGR1,GADD45A,HBEGF,JUN,MAP2K3 |
| RHOA | 0,007 | enzyme | | -1,194 | -1,701 | -0,205 | 2,36E-06 | 2,49E-05 | CCN2,CDKN1A,CEBPB,FOS,GDF15,JUN,RHOB,RND3 |
| NR1H3 | 0,002 | ligand-dependent nuclear receptor | | -0,081 | -0,299 | -0,079 | 2,47E-06 | 2,60E-05 | ARG2,ATF3,CCNA2,CD55,CDKN1A,HMGCR,IFIT2,LDLR,MYLIP,NFKBIA,TRIB1,VEGFA |
| Ins1 | | other | Inhibited | -2,109 | -2,914 | -0,224 | 2,89E-06 | 3,01E-05 | CDKN1A,CEBPB,CEBPD,EGR1,FOS,HMGCR,JUN,JUNB,LDLR,LIFR,PER1,SERPINE1,SOCS2,VEGFA |
| RARA | | ligand-dependent nuclear receptor | | -0,108 | -0,102 | 0,002 | 3,05E-06 | 3,16E-05 | CCNA2,CDKN1A,CEBPB,DUSP1,EGR1,ELL2,FOS,FOSL2,GDF15,HNF1B,IL6R,JUN,LIFR,NEDD9,RGS3 |
| MACROH2A1 | -0,017 | other | | -0,775 | -0,378 | 0,150 | 3,53E-06 | 3,59E-05 | CDKN1A,CDKN1C,CITED2,EGR1,GADD45A,SERPINE1,SOCS2 |

| | | | | | | | | | |
|---|---|---|---|---|---|---|---|---|---|
| amino acids | | chemical - endogenous mammalian | Activated | 1,786 | 2,164 | 0,174 | 4,03E-06 | 4,05E-05 | FOS,FOSB,JUN,JUNB,SLC38A2 |
| APP | -0,110 | other | Inhibited | -1,312 | -2,178 | -0,203 | 4,18E-06 | 4,19E-05 | ARF6,CCN2,CITED2,EGR1,ETS2,FOS,FOSB,GDF15,HES1,HMGCR,HOMER1,IFIT2,JUN,JUNB,JUND,LDLR,MAFF,NAMPT,RCAN1,RNASEL,SERPINE1,SMAD7,TUBB2A,VEGFA,YWHAZ |
| ADCYAP1 | | other | | -1,393 | -1,853 | -0,136 | 4,23E-06 | 4,24E-05 | BTG2,CCNA2,CEBPB,CEBPD,CHGB,EGR1,FOS,GCLM,IER3,MCM2,NFKBIA,RHOB |
| RPS6KA4 | | kinase | | -1,988 | -1,988 | 0,000 | 4,29E-06 | 4,28E-05 | DUSP1,FOS,JUN,JUNB |
| NME1 | 0,110 | kinase | | 1,479 | 1,904 | 0,224 | 4,44E-06 | 4,41E-05 | CCN2,CDKN1A,RAE1,SERPINE1,SLBP,SMAD7 |
| SOX2 | | transcription regulator | | 1,155 | 1,497 | 0,084 | 4,77E-06 | 4,72E-05 | AMOT,ATF3,BHLHE40,CCN2,CDKN1A,CITED2,DUSP1,DUSP5,ETS2,FOS,GADD45B,GATA6,JAG1,JUN,KLF6,SERPINE1,VEGFA |
| quinolinic acid | | chemical - endogenous mammalian | Inhibited | -2,208 | -2,376 | -0,071 | 5,08E-06 | 4,97E-05 | EGR1,FOS,FOSB,JUN,JUNB,JUND |
| 10E,12Z-octadecadienoic acid | | chemical - endogenous mammalian | | -1,726 | -1,941 | -0,083 | 5,84E-06 | 5,63E-05 | ATF3,GDF15,LDLR,NAMPT,NR4A1,NR4A3,PPP1R15A |
| POMC | 0,034 | other | Inhibited | -2,038 | -2,581 | -0,210 | 6,24E-06 | 5,92E-05 | DUSP1,FOS,FOSL2,JUN,JUNB,LDLR,NR4A1,NR4A3 |
| RXRA | -0,076 | ligand-dependent nuclear receptor | | 0,174 | -0,147 | -0,116 | 8,60E-06 | 7,95E-05 | ARG2,CCNA2,CDKN1A,CEBPD,CYP27B1,FOS,HBEGF,HNF1B,IER3,KLF9,NEDD9,VEGFA,WEE1 |
| BCL6 | 0,069 | transcription regulator | | 0,712 | 1,067 | 0,179 | 8,84E-06 | 8,13E-05 | CDKN1A,CISH,DUSP6,GABARAPL1,GADD45A,IL6R,JUNB,KLF6,LDLR,SERPINE1,SOCS2 |
| TERT | 0,041 | enzyme | | 0,527 | 0,333 | -0,116 | 9,01E-06 | 8,24E-05 | ATF3,CCN2,CDKN1A,DUSP6,GATA6,GNA13,IFIT2,LIPG,NFKBIA |
| FOSL1 | -0,100 | transcription regulator | Activated | 2,390 | 2,390 | 0,000 | 9,27E-06 | 8,42E-05 | ADM,EGR1,FOS,FOSB,JUN,JUNB,SERPINE1 |
| TBX2 | -0,100 | transcription regulator | | 1,699 | 1,238 | -0,178 | 1,01E-05 | 9,13E-05 | ATF3,BHLHE40,CCNA2,CCNL1,CDKN1A,MCM2,NFIL3 |
| leucine | | chemical - endogenous mammalian | | -0,109 | -0,200 | -0,042 | 1,04E-05 | 9,33E-05 | ATF3,CDKN1A,FOS,NR4A1,PPP1R15A |
| DCN | -0,258 | other | | -0,726 | -0,589 | 0,059 | 1,34E-05 | 1,18E-04 | CCN2,CDKN1A,CDKN1C,DUSP1,SERPINE1,VEGFA |
| PRKG1 | | kinase | | -0,750 | -0,651 | 0,045 | 1,45E-05 | 1,26E-04 | CCN2,CDKN1A,FOS,GUCY1A1,SERPINE1 |
| SPI1 | | transcription regulator | | 2,536 | 1,947 | -0,227 | 1,47E-05 | 1,28E-04 | CCNA2,CDKN1A,CHI3L1,DUSP6,ETS2,FOS,FOXO1,HES1,IFIT2,IFIT3,JUN,MCL1 |
| HMGA1 | -0,045 | transcription regulator | | 0,360 | -0,156 | -0,174 | 1,77E-05 | 1,50E-04 | EGR1,FOS,FOSL2,HMGCR,IDI1,IER2,IER3,JUNB,RHOB |
| NDRG1 | -0,029 | kinase | | -0,249 | -0,249 | 0,000 | 1,97E-05 | 1,64E-04 | ATF3,EIF2AK3,FOS,JUN,VEGFA |
| DDIT3 | -0,239 | transcription regulator | | -0,041 | -0,213 | -0,062 | 2,14E-05 | 1,77E-04 | ATF3,CEBPD,FOXO1,JAG1,PER1,PPP1R15A,SRSF2,TSC22D3 |
| CTBP1 | -0,151 | enzyme | | 1,509 | 1,977 | 0,237 | 2,62E-05 | 2,07E-04 | ACKR3,CDKN1A,FKBP5,FOS,SPRY2 |
| KDM1A | | enzyme | | 0,765 | 1,174 | 0,133 | 2,63E-05 | 2,08E-04 | ADM,CCNA2,CDKN1A,ELL2,FOXO1,MCM2,SERPINE1,SMAD7,SOCS2,ZNF165 |
| ERBB3 | 0,029 | kinase | | 0,925 | 1,091 | 0,069 | 2,66E-05 | 2,10E-04 | CDKN1A,DUSP1,EGR1,F3,HBEGF,IER2,VEGFA |
| corticosterone | | chemical - endogenous mammalian | | -1,815 | -1,858 | -0,017 | 2,73E-05 | 2,15E-04 | CCN2,CDKN1A,FKBP5,FOS,JUN,KLF9,PDK4,SGK1 |
| PML | | transcription regulator | | 0,246 | 0,131 | -0,039 | 2,96E-05 | 2,31E-04 | CDKN1A,GADD45A,GCLM,HMGCR,IFIT3,LDLR,MAFF,PDK4,UBE2M |
| TRH | 0,049 | other | Inhibited | -1,749 | -2,169 | -0,194 | 3,01E-05 | 2,33E-04 | DUSP1,FOS,JUN,JUNB,NR4A1 |
| XDH | 0,089 | enzyme | | 0,134 | 0,277 | 0,065 | 3,01E-05 | 2,33E-04 | CCN2,CEBPB,DUSP1,F3,SERPINE1 |
| PIAS1 | | transcription regulator | | 0,634 | 0,762 | 0,065 | 3,01E-05 | 2,33E-04 | CDKN1A,GADD45A,KLF5,MCL1,RRAD |
| Pde4 | | group | Activated | 1,572 | 2,000 | 0,214 | 3,18E-05 | 2,44E-04 | CDKN1A,CYP51A1,HMGCR,LDLR |

| | | | | | | | | | |
|---|---|---|---|---|---|---|---|---|---|
| mevalonic acid | | chemical - endogenous mammalian | | 0,942 | 0,812 | -0,061 | 3,43E-05 | 2,58E-04 | CD55,HMGCR,LDLR,SERPINE1,VEGFA |
| FGF1 | -0,567 | growth factor | | -2,132 | -1,853 | 0,110 | 3,45E-05 | 2,59E-04 | CCN2,CDKN1A,DUSP6,EGR1,FOS,GCLM,JUN,VEGFA |
| HDAC4 | | transcription regulator | | -0,225 | 0,192 | 0,210 | 3,65E-05 | 2,73E-04 | ATF3,CDKN1A,FOS,GADD45A,HOMER1,NR4A1,RGS2,VEGFA |
| NR2F1 | 0,082 | ligand-dependent nuclear receptor | | -0,620 | -0,811 | -0,097 | 3,91E-05 | 2,89E-04 | EGR1,GDF15,HNF1B,JAG1,SPRY2 |
| LY6E | 0,188 | other | | -0,116 | 0,000 | 0,058 | 3,94E-05 | 2,89E-04 | CCN2,CD83,NFKBIA,SERPINE1 |
| AGTR1 | -0,044 | G-protein coupled receptor | | -0,974 | -1,400 | -0,194 | 4,43E-05 | 3,19E-04 | DUSP1,FOS,JUN,SERPINE1,SGK1 |
| ATF3 | -1,461 | transcription regulator | | -0,541 | -0,248 | 0,122 | 4,67E-05 | 3,36E-04 | ATF3,CD83,CDKN1A,JUN,LDLR,PPP1R15A,SERPINE1 |
| ADIPOQ | 0,054 | other | | 1,262 | 1,141 | -0,044 | 4,78E-05 | 3,42E-04 | CCN2,CDKN1A,DUSP1,EGR1,F3,FOS,HBEGF,LDLR,NFKBIA |
| IGFBP5 | 0,066 | other | | -1,812 | -1,976 | -0,083 | 4,83E-05 | 3,42E-04 | BHLHE40,F3,SERPINE1,TNFRSF1A |
| RPS6KA5 | | kinase | | -1,528 | -1,990 | -0,233 | 4,83E-05 | 3,42E-04 | DUSP1,FOS,JUN,JUNB |
| Pkg | | group | Inhibited | -1,502 | -2,000 | -0,249 | 5,86E-05 | 4,04E-04 | EGR1,FOS,FOSB,JUN |
| MAPK13 | 0,073 | kinase | | -0,807 | -1,114 | -0,159 | 5,86E-05 | 4,04E-04 | CDKN1A,CITED2,FOS,VEGFA |
| DYRK1A | | kinase | | -1,498 | -1,000 | 0,249 | 5,86E-05 | 4,04E-04 | CDKN1A,RCAN1,SERPINE1,SRSF2 |
| TNFSF10 | 0,253 | cytokine | Activated | 2,233 | 2,023 | -0,090 | 6,07E-05 | 4,18E-04 | FOS,JUN,MCL1,NFKBIA,RCAN1,TFRC,VEGFA |
| THRB | 0,005 | ligand-dependent nuclear receptor | | -0,594 | -0,816 | -0,091 | 6,22E-05 | 4,25E-04 | BTG2,CYP27B1,FOS,JUN,KLF9,LDLR,NR4A1,PDK4,PER1,SERPINE1 |
| ADORA2A | | G-protein coupled receptor | | -0,191 | -0,426 | -0,088 | 6,29E-05 | 4,28E-04 | DUSP1,FOS,NFKBIA,NR4A1,NR4A3,TSC22D3,TUBB2A,VEGFA |
| S100A8 | -0,493 | other | | -0,381 | -0,788 | -0,144 | 7,15E-05 | 4,81E-04 | ARG2,ATF3,BHLHE40,EFNA1,NFKBIA,NR4A3,PER1,RAB20,SGK1 |
| GLI3 | 0,051 | transcription regulator | Inhibited | -2,125 | -2,449 | -0,133 | 7,22E-05 | 4,84E-04 | DUSP1,DUSP5,EGR1,FOSB,FOXO1,JUNB |
| KLF6 | -1,235 | transcription regulator | | 0,642 | 0,486 | -0,073 | 7,81E-05 | 5,14E-04 | ATF3,CDKN1A,MCL1,NFKBIA,NR4A1,VEGFA |
| K+ | | chemical - endogenous mammalian | | 0,125 | -0,420 | -0,249 | 7,90E-05 | 5,18E-04 | ATF3,EGR1,FOS,JUN,NR4A1 |
| OGA | 0,193 | enzyme | | 1,296 | 1,499 | 0,055 | 8,09E-05 | 5,29E-04 | ADRM1,CAVIN2,CD55,CDKN1A,CEBPB,EPHA4,FERMT2,GADD45A,HMGCR,IFIT3,IL6R,JUN,RHOB,SOCS2 |
| VDR | -0,079 | transcription regulator | | 0,368 | 0,671 | 0,126 | 8,11E-05 | 5,29E-04 | CDKN1A,CEBPB,CYP27B1,F3,GADD45A,IER3,JUN,LDLR |
| SP110 | 0,179 | transcription regulator | Activated | 1,783 | 2,121 | 0,120 | 8,11E-05 | 5,29E-04 | ATF3,F3,GNA13,IFIT3,IL6R,MCL1,NFIL3,TNFRSF1A |
| AGER | | transmembrane receptor | Inhibited | -1,735 | -2,187 | -0,207 | 8,44E-05 | 5,45E-04 | CDKN1A,CHGB,EGR1,F3,SERPINE1,VEGFA |
| SPHK1 | 0,035 | kinase | | -0,751 | -1,159 | -0,185 | 8,80E-05 | 5,65E-04 | CCN2,CDKN1A,EGR1,JUN,SERPINE1 |
| ACOX1 | -0,057 | enzyme | Activated | 2,500 | 2,813 | 0,111 | 9,40E-05 | 5,99E-04 | CDKN1A,ETS2,GADD45A,KLF6,PDK4,RGS2,SGK1,SLC20A1 |
| thyroid hormone | | chemical - endogenous mammalian | | -1,221 | -1,195 | 0,010 | 9,61E-05 | 6,06E-04 | BTG2,CCNA2,CDKN1A,CHGB,EGR1,FOS,HMGCR,KLF9,PER1 |
| PTPN11 | | phosphatase | | -0,096 | -0,232 | -0,069 | 9,82E-05 | 6,12E-04 | CDKN1A,EGR1,FOS,JUN,MCL1,PIM1 |
| BCL2 | | transporter | | 1,499 | 0,946 | -0,217 | 9,90E-05 | 6,15E-04 | CDKN1A,CHGB,FOS,GADD45A,MCL1,NFKBIA,VEGFA |
| WWTR1 | -0,100 | transcription regulator | | -0,877 | -1,387 | -0,230 | 1,08E-04 | 6,64E-04 | CCN2,CCNA2,CDKN1A,SERPINE1,SMAD7 |
| PRKAA1 | -0,053 | kinase | | -0,980 | -0,378 | 0,228 | 1,09E-04 | 6,66E-04 | ACKR3,ATF3,CDKN1A,CHGB,FKBP5,NFKBIA,RGS2,RND3 |
| YY1 | | transcription regulator | | 1,315 | 1,423 | 0,045 | 1,12E-04 | 6,88E-04 | BTG2,CCNA2,CDKN1A,CEBPB,EGR1,FOS,HMGCR,LDLR,PPP1R15A,RND3,VEGFA |

| | | | | | | | | | |
|---|---|---|---|---|---|---|---|---|---|
| NOS2 | -0,049 | enzyme | | -0,048 | -0,251 | -0,068 | 1,13E-04 | 6,91E-04 | CDKN1A,CHI3L1,DDX3X,FOXO1,IFIT3,KLF6,NFKBIA,TSPYL2,VEGFA |
| LTBP1 | 0,107 | other | | 0,378 | 0,000 | -0,194 | 1,16E-04 | 7,03E-04 | MCL1,PIM1,SERPINE1,SMAD7 |
| DTX1 | | transcription regulator | | 0,113 | -0,152 | -0,134 | 1,16E-04 | 7,03E-04 | DUSP6,EGR1,MCL1,VEGFA |
| MAP3K8 | -0,061 | kinase | | -1,095 | -1,311 | -0,084 | 1,31E-04 | 7,84E-04 | CEBPD,DUSP6,FOS,IER3,JUN,JUNB,RGS3,SLCO4A1 |
| BTK | 0,052 | kinase | | -1,768 | -1,969 | -0,077 | 1,55E-04 | 9,09E-04 | CDKN1A,CEBPB,CISH,FOXO1,IFIT3,JUN,PIM1 |
| ZBTB20 | -0,099 | transcription regulator | Activated | 2,249 | 2,200 | -0,022 | 1,59E-04 | 9,28E-04 | CISH,NEDD9,NFKBIA,PDK4,SOCS2 |

### Table S3. Gold

| Upstream Regulator | Expr Log Ratio | Molecule Type | Predicted Activation State | Bias-corrected z-score | Activation z-score | Bias Term | p-value of overlap | B-H corrected p-value | Target Molecules in Dataset |
|---|---|---|---|---|---|---|---|---|---|
| beta-estradiol | | chemical - endogenous mammalian | | -0,606 | -0,534 | 0,003 | 2,92E-69 | 8,24E-66 | A2M,ABCC5,ABCG2,ACAA1,ACAA2,ACADM,ACADVL,ACE,ACKR3,ACOX1,ACOX2,ACP5,ACSL1,ACSM3,ACTB,ADAMDEC1,ADAMTS1,ADCY9,ADIRF,ADM,ADRB2,AEBP1,AFP,AGRN,AHR,ALB,ALDH18A1,ALDH4A1,ALDH7A1,ALPG,ANKH,ANXA1,ANXA3,ANXA4,APBB1,APOA1,APOA4,APOE,AQP1,ARAF,ARC,ARNT2,ASB9,ATF3,AURKA,AVP,BAG1,BAX,BAZ1A,BCAS3,BCL2A1,BCL6,BHLHE40,BID,BIN1,BIRC3,BLNK,BNIP2,BNIP3L,C3,CA2,CALB1,CALM1 (includes others),CALR,CAMK2A,CAP2,CASP3,CASP4,CAST,CAV1,CAV2,CBX6,CCL20,CCL21,CCL5,CCNA2,CCNB1,CCNB2,CCND1,CCND2,CCND3,CCNG1,CCR2,CCR5,CCT2,CD14,CD24,CD40,CD44,CD55,CD69,CD74,CD86,CDC123,CDH11,CDH15,CDK1,CDK18,CDK5,CDKN1B,CDKN1C,CELF2,CENPM,CFB,CGA,CHUK,CKM,CLCN3,CLDN1,CLIC4,CLU,CNN2,COA4,COL1A1,COL1A2,COL3A1,COLEC12,COMT,COX6A2,CRADD,CRYZ,CSE1L,CSHL1,CSTB,CTPS1,CTSB,CTSD,CTSK,CUBN,CXADR,CXCL1,CXCL10,CXCL11,CXCL12,CXCR4,CYB5A,CYBB,CYP17A1,CYP1B1,CYP2A6 (includes others),CYP2B6,CYP3A4,DAB2,DBN1,DCLK1,DCN,DDB2,DDC,DDIT3,DDOST,DDR1,DECR1,DEFB1,DERL1,DHCR7,DHRS9,DIO1,DLG3,DMTN,DNPH1,DUSP1,DUSP3,DYNLL1,DYRK2,ECH1,ECHDC2,EFNB2,EGF,EIF3D,ELF1,ENC1,ENO1,ENPP2,EPB41L3,ERBB3,ERF,ERH,ESR1,ESRRA,EXT2,F12,F2RL1,F3,FAAH,FAM111A,FARP1,FAS,FBN1,FCGR2B,FCGRT,FEZ2,FGF1,FGF7,FGFR3,FHL1,FHL2,FMO1,FMO5,FN1,FNBP1,FOLH1,FOLR1,FOS,FOSB,FOSL1,FOXO1,FSCN1,FSTL1,FXYD5,FZD2,G6PC,GAB2,GADD45A,GADD45B,GADD45G,GALNT1,GALNT7,GARS1,GBP2,GCH1,GCHFR,GCLC,GHR,GIPC2,GJA1,GJB1,GK,GLRB,GM2A,GMFB,GNA11,GNB1,GNL3,GPER1,GPR137B,GPSM2,GPX3,GRB10,GSR,GSTA1,GSTP1,GTF2IRD1,GUCY1A1,GUCY1B1,GYPA,HADH,HBB,HDAC1,HIF1A,HLA-A,HLA-DQA1,HLA-DRB1,HLA-E,HNF1B,HOXA10,HPGD,HSD11B2,HSD17B11,HSP90B1,HSPA5,HSPA8,HSPA9,HSPB8,HTR6,HTRA1,ICAM1,ID1,ID2,IDO1,IFI30,IFNGR2,IFRD1,IFRD2,IFT122,IGFBP4,IGFBP6,IGSF6,IL13,IL15,IL18,IL1R1,IL4R,IL6R,IL7,IL7R,IMPDH2,INHA,INPP4B,INPP5J,IQGAP1,IRF1,IRF7,ISG15,ITGA2,ITGAV,ITGB2,ITGB3,JAK1,JUND,KCNN2,KDELR3,KITLG,KLF9,KLK1,KRAS,KRT19,KRT4,KRT7,KRT81,KYNU,LAMB1,LAPTM5,LCN2,LEPR,LEPROT,LGALS3BP,LHB,LHFPL2,LIMA1,LIN7A,LITAF,LPIN1,LPO,LTB,LTF,LUM,LYZ,LZTS3,MAD2L1,MAF,MAL,MAOA,MAP2K3,MAPT,MCAM,MCM2,MCM3,MCM5,MCM6,MCM7,MEIS2,MET,MGP,MMD,MME,MMP1,MMP2,MMP7,MMP9,MPC1,MPDZ,MRC1,MSMO1,MT1F,MT1G,MT1X,MTOR,MTTP,MUC1,MX1,MYB,MYL12A,MYO1B,MYOF,NAMPT,NCF2,NDRG1,NEDD4L,NEDD9,NET1,NFE2L2,NFKB1,NIPSNAP1,NME1,NOS2,NPPA,NPR3,NPY1R,NR0B2,NR1H3,NR4A1,NR4A3,NRP1,NSDHL,NUCB2,NUDT1,OGDH,OSTF1,OXT,P4HB,PARP1,PCBD1,PCNA,PCYOX1,PDCD4,PDGFRA,PDIA3,PDI[illegible] |

| | | | | | | | | |
|---|---|---|---|---|---|---|---|---|
| TGFB1 | growth factor | Activated | 2,761 | 2,823 | 0,003 | 1,61E-63 | 3,40E-60 | AASS,ABCC2,ABCG1,ABCG2,ACAA2,ACE,ACKR1,ACSM3,ACTN1,ADAM10,ADAM17,ADCY9,ADI1,ADM,ADRA2A,AFM,AFP,AGR2,AHNAK,AHR,AIM2,ALB,ALDH18A1,ALDH2,ALDH3A2,ALDH5A1,ALDOB,ALOX5,ALOX5AP,ANKH,ANXA11,ANXA2,APOE,AQP1,ARF4,ARNT2,ARPC2,ASNS,ASS1,AVP,AXL,B2M,B4GAT1,BACE1,BARD1,BAX,BCL6,BHLHE40,BIN1,BMP7,BNIP3L,BUD31,C1QA,C1QB,C1R,C1S,C2,C3,C3AR1,CALM1 (includes others),CALML3,CALML4,CAMK2G,CAMK2N1,CAP1,CAPRIN1,CASP1,CASP3,CASP4,CASP8,CAT,CAV1,CAV2,CBFB,CBR3,CCDC28A,CCL18,CCL20,CCL5,CCL8,CCN3,CCNA2,CCNB1,CCNB2,CCND1,CCND2,CCND3,CCR2,CCR5,CCR7,CCT2,CCT5,CCT6A,CD14,CD163,CD300A,CD36,CD40,CD44,CD55,CD59,CD69,CD83,CD86,CDC42SE1,CDC7,CDH11,CDH5,CDHR2,CDK1,CDK2AP1,CDKN1B,CDKN1C,CDKN2C,CDKN3,CELF2,CELSR2,CFB,CFD,CFH,CHST8,CIDEC,CKM,CLDN1,CLIC4,CLU,CNN2,COL16A1,COL18A1,COL1A1,COL1A2,COL3A1,COL4A1,COL4A2,COL5A1,COL6A3,CORO1C,COTL1,CRIP2,CRMP1,CSF1R,CTCF,CTPS1,CTSB,CTSD,CTSK,CTSS,CUL5,CX3CR1,CXADR,CXCL1,CXCL10,CXCL12,CXCR4,CYBA,CYBB,CYC1,CYP17A1,CYP27B1,CYP3A7,DAB2,DCN,DDB2,DKK3,DMTN,DNMT1,DNPEP,DOCK2,DOK1,DSG2,DUSP1,DYNLL1,DYRK2,EGF,EIF4A3,ELF4,ELK3,ELMO1,ENG,ENO1,ENO3,ERF,ERLIN2,ESM1,ESPL1,ESRP2,EXT1,EXT2,F12,F2RL1,F3,FAM110B,FAS,FBN1,FCER1A,FCER1G,FCGR3A/FCGR3B,FERMT2,FGF7,FHL1,FLI1,FLNA,FLRT1,FN1,FNBP1,FNDC3B,FOS,FOSB,FOXO1,FSCN1,FTL,FUBP1,FUS,FXYD5,FYN,FZD2,G6PC,GAD1,GADD45A,GADD45B,GARS1,GATM,GBP1,GBP2,GCLC,GJA1,GLS,GNAI2,GNAS,GNB5,GNG7,GPER1,GPR21,GPT,GZMA,GZMB,H1-10,HABP2,HADH,HEXB,HIF1A,HK1,HLA-DMA,HLA-DQA1,HLA-DQB1,HMOX2,HNF1A,HNF4A,HNMT,HOXB5,HPGD,HSD17B10,HSD3B2,HSPA5,HTR2B,HTRA1,ICAM1,ICAM2,ID1,ID2,ID3,IDI1,IFI16,IFI27,IFI30,IFIH1,IFIT3,IFRD1,IGFBP4,IGFBP6,IGFBP7,IGHM,IL10RA,IL13,IL15,IL18,IL1R1,IL2RB,IL32,IL33,IL36A,IL4R,IL6R,IMPDH1,INHA,INPP5D,IRF1,ITGA2,ITGA3,ITGA7,ITGAL,ITGAM,ITGAV,ITGB2,ITGB3,ITGB6,JUND,KDELR3,KDM5B,KITLG,KLF15,KLF9,KLRB1,KLRD1,KNG1,KRAS,KRT10,KRT19,KRT7,KRT81,LAMC1,LBR,LCN2,LEMD3,LITAF,LOC102724788/PRODH,LOXL1,LPAR1,LTBP3,MAF,MAOA,MAP2K3,MAP3K1,MAPK13,MCM2,ME2,MEF2C,MET,MFAP2,MGAT3,MGP,MIA2,MIA3,MID1,MLXIPL,MMP1,MMP2,MMP7,MMP9,MRC1,MSH6,MSMO1,MSN,MTHFD2,MTRR,MXI1,MYB,MYD88,MYH9,MYL12A,MYL3,MYL6,MYOF,NAB2,NAMPT,NBEA,NCF2,NDC80,NDUFS4,NEDD9,NET1,NGF,NIT1,NME1,NNMT,NOS2,NOX4,NPPA,NR1H3,NR4A1,NR4A3,NRP1,NUAK1,NUCB2,NUP62,OSTF1,OVOL1,P2RY14,P4HA1,PAPPA,PCCB,PCGF2,PCNA,PCOLCE2,PDCD4,PDGFRA,PDK2,PDLIM4,PDZK1IP1,PECAM1,PFKL,PFKP,PGAM2,PGRMC1,PGRMC2,PHG[illegible] |

| | | | | | | | | | |
|---|---|---|---|---|---|---|---|---|---|
| HNF4A | -0,438 | transcription regulator | Inhibited | -6,325 | -6,298 | 0,002 | 1,06E-61 | 1,79E-58 | A1CF,AAMDC,AASS,ABCC2,ABCC3,ABCC6,ABCG1,ABHD10,ABHD6,ACAA2,ACAD8,ACAT1,ACO1,ACO2,ACOT13,ACOX1,ACOX2,ACOX3,ACSL1,ACSS3,ACTN1,ACTR3,ADCY7,ADH1B,ADH6,ADIPOR1,ADRB2,AFP,AGPAT1,AHNAK,AK2,AKR1B1,ALB,ALDH1L1,ALDH2,ALDH3A1,ALDH5A1,ALDH8A1,ALDOB,AMT,ANAPC15,ANG,ANXA5,ANXA9,AOC1,APEH,APOA1,APOA4,APOC1,APOC3,APOE,APOH,APOM,AQP1,AQP3,ARFIP2,ARG2,ARL4C,ARPC5,ASNS,ATOX1,ATP6V1D,AVPI1,BAAT,BACE1,BARD1,BAZ1A,BCCIP,BCKDHA,BCL6,BCLAF1,BHMT,BLOC1S1,BLVRB,BMP7,BOLA1,BPHL,BST1,BTN2A1,BUD31,C11orf71,C1S,C2,C3,CALML4,CAP1,CAPZA2,CAT,CAV1,CBR3,CBX3,CCDC25,CCDC90B,CCNA2,CCNB1,CCND1,CCNG1,CCNH,CD1D,CD55,CDC123,CDHR2,CDK1,CDK5,CDKN1B,CENPU,CEP83,CES2,CETN3,CHP1,CIDEC,CLCN6,CLDN1,CLDN3,CLPB,CLPTM1,CLPX,CLTCL1,CMC2,COA4,COA7,COASY,COPB1,COPS7A,COQ3,COQ4,COQ6,COQ8A,COX11,COX8A,CPT2,CRADD,CRAT,CRIP1,CRYZ,CRYZL1,CSK,CSTF2T,CTSB,CTSZ,CUL5,CUTC,CXADR,CYP1B1,CYP27A1,CYP2A6 (includes others),CYP2B6,CYP2C8,CYP2J2,CYP3A4,CYP3A7,CYP4F2,DCAF11,DCAF8,DCK,DDOST,DDX10,DDX25,DDX47,DEPP1,DERL1,DLST,DNAJA3,DNAJB9,DNPH1,DPAGT1,DTL,DUSP3,DYNC1LI1,EBP,ECI2,EED,EIF4EBP2,EIF4ENIF1,EIF5,ELMO1,ENC1,EPHX1,ESYT1,ETFDH,ETNK2,ETNPPL,EVPL,F11,F12,FABP1,FABP2,FAHD2A,FAM117A,FAM50B,FANCF,FAS,FBP1,FDX1,FGF13,FGF7,FH,FHIT,FHL1,FKBP3,FLNA,FMO1,FOXO1,FUT3,FUT6,FUZ,FZD6,G0S2,G6PC,GADD45B,GADD45G,GATB,GCHFR,GDAP2,GGCX,GHITM,GINS2,GIPC2,GJB1,GLYAT,GMDS,GNAI3,GNL3,GOLGA4,GOLIM4,GOT1,GPATCH3,GPT,GPX7,GRHPR,GSS,GSTA1,GSTK1,GSTM4,GSTO1,GSTZ1,GTF2E1,HAAO,HADHA,HAO1,HAO2,HBQ1,HDAC6,HGD,HIF1A,HIKESHI,HIRIP3,HLA-A,HLA-B,HLA-C,HLA-F,HLA-G,HMGB2,HMGN4,HMOX2,HNF1A,HNF1B,HNF4A,HNRNPR,HPD,HPF1,HPN,HSDL2,HSP90B1,HSPA4L,HSPA5,HSPA8,IER5,IFI30,IFITM2,IFNGR1,IFT122,IL10RA,IL15,IL2RB,INTS9,INVS,IPO13,ISOC1,ISOC2,ITGAL,KCNN2,KIF20A,KIF22,KLF15,KLHL20,KNG1,KPNB1,KPTN,KRT10,KRT7,L2HGDH,LAMTOR5,LARP4,LCN2,LEPR,LIPA,LOC102724788/PRODH,LPGAT1,LRP5,LSM4,LTF,MAIP1,MAOA,MAOB,MAP7,MAPK8IP2,MAT2A,MCCC1,MCM3,MDH2,MFSD1,MGST2,MICU1,MID1,MLXIPL,MMUT,MPP1,MPZL2,MRM3,MRPL2,MRPL24,MRPL34,MRPL4,MRPL44,MRPL46,MRPL49,MRPS12,MRPS15,MRPS35,MRPS7,MSN,MSRB1,MSRB2,MT1X,MTHFD1,MTNR1A,MTTP,MYL6,NAMPT,NAPA,NARS2,NAT10,NAT8,NBEA,NCK1,NDC80,NDRG1,NDUFA3,NDUFA7,NDUFS1,NDUFS3,NDUFS4,NDUFV1,NFE2L1,NFE2L2,NFKBIL1,NFS1,NME1,NOD2,NOP16,NOP53,NOX4,NR0B2,NR1H2,NR1H3,NR1I3,NR2F2,NSF,NTRK2,NUAK1,NUCB1,NUDT6,NUP54,NUP62,OAS3,OGDH,OGFOD2,OPA1,ORMDL2,OSMR,OVOL1,P2RY14,P[illegible] |

| | | | | | | | | |
|---|---|---|---|---|---|---|---|---|
| IFNG | cytokine | Activated | 8,670 | 8,748 | 0,004 | 2,79E-59 | 3,93E-56 | A2M,ABCC1,ACE,ACO1,ACTB,ADA2,ADAM17,ADCY7,ADGRG1,ADM,ADRA2A,AGPAT1,AGRN,AHCY,AHR,AIM2,ALDH1A3,ALDH1L1,ALDH7A1,ALOX5AP,ALPL,APOBEC3G,APOL1,AQP1,ARAP2,ARC,ARG2,ARL6IP5,ARNT2,ASNS,ASS1,ATF3,AZGP1,B2M,BACE1,BACH2,BAX,BCL2A1,BCL2L2,BID,BIRC2,BIRC3,BLNK,BST1,BST2,BTN3A1,BTN3A2,C1QA,C1QB,C1R,C2,C3,CALB1,CAPN3,CASP1,CASP3,CASP4,CASP6,CASP8,CAT,CAV1,CCL18,CCL19,CCL20,CCL5,CCL8,CCNA2,CCND1,CCND2,CCND3,CCR2,CCR5,CD14,CD163,CD1D,CD2,CD200,CD36,CD38,CD40,CD44,CD55,CD63,CD74,CD83,CD86,CDH5,CDKN1B,CDX2,CELSR2,CFB,CHRND,CHST8,CIRBP,CISH,CLEC10A,CLEC7A,CLIC4,CLIC5,COL1A1,COL1A2,COL5A2,CORO1A,CRABP1,CSE1L,CSF1R,CSF2RB,CSK,CTSB,CTSD,CTSK,CTSS,CTSV,CTSZ,CX3CR1,CXADR,CXCL1,CXCL10,CXCL11,CXCL12,CXCL6,CXCL9,CXCR4,CYB5A,CYBA,CYBB,CYP27A1,CYP3A4,DDB2,DDIT3,DDX58,DDX60,DECR1,DEFB1,DEPP1,DHX9,DIO1,DOK1,DPP4,DUSP1,DUSP9,EFNB2,EGF,EGOT,EHF,ENO1,ERAP1,ESM1,ETV7,F2RL1,F3,FAAH,FAH,FANCF,FAS,FBP1,FCER1G,FCGR2B,FCGR3A/FCGR3B,FCN1,FGF1,FGL2,FHL2,FKBP5,FKBP6,FN1,FOS,FOSB,FOXO1,FTL,FZD2,GAD1,GAL3ST1,GBP1,GBP2,GCH1,GCHFR,GJA1,GLS,GLUL,GNAI2,GNAS,GNB5,GNG7,GPER1,GPT,GRK2,GSTP1,GZMB,HAAO,HCK,HCP5,HERC6,HIF1A,HLA-A,HLA-B,HLA-C,HLA-DMA,HLA-DMB,HLA-DOA,HLA-DOB,HLA-DQA1,HLA-DQB1,HLA-DRA,HLA-DRB1,HLA-E,HLA-F,HLA-G,HSPA5,HSPA8,HTRA1,ICAM1,ID1,IDH3B,IDI1,IDO1,IFI16,IFI27,IFI30,IFI35,IFI44,IFI44L,IFI6,IFIH1,IFIT2,IFIT3,IFIT5,IFITM1,IFITM2,IFNGR1,IFNGR2,IGFBP4,IL10RA,IL13,IL15,IL15RA,IL18,IL1R1,IL32,IL4R,IL6R,IL7,IL7R,INSR,IRAK1,IRF1,IRF2,IRF7,IRF8,IRF9,ISG15,ISG20,ITGAL,ITGAM,ITGAV,ITGB2,ITGB3,JUND,KCTD12,KITLG,KMO,KRAS,KRT19,KYAT1,KYNU,LAMC1,LAMP3,LCN2,LCP2,LGALS3BP,LST1,LTB,LY96,LYN,MAFF,MARCKSL1,MAT2A,MIF,MMP1,MMP2,MMP9,MNDA,MRAS,MRC1,MSH2,MT1F,MT1G,MUC1,MX1,MX2,MYB,MYD88,MYH9,NAMPT,NCALD,NCF2,NEDD9,NFE2L1,NFKB1,NGF,NMI,NOD2,NOS2,NOX4,NR1H2,NTRK2,OAS1,OAS2,OAS3,OPTN,P2RY14,PANX1,PAPPA,PBK,PCDH17,PCK1,PCTP,PEA15,PECAM1,PFKFB3,PFKL,PHGDH,PIGR,PKM,PLA2G7,PLAAT3,PLAAT4,PLD1,PLEK,PLSCR1,PPARA,PPARGC1A,PRDX2,PRKCA,PRKCD,PRKCQ,PRNP,PRSS8,PSMA4,PSMA6,PSMB10,PSMB8,PSMB9,PSME1,PTGES,PTGES2,PTPN6,RAB17,RAB27A,RAB38,RAC2,RACK1,RAP2B,RARRES1,RB1,RBBP7,RGN,RHOB,RORC,RSAD2,RSPH14,RTP4,RUNX3,S100A10,S100A8,S100A9,SAMD9,SAMHD1,SCNN1B,SDC1,SEC11A,SELL,SELP,SERPINA5,SERPINB9,SERPING1,SFRP1,SGPL1,SHARPIN,SLC12A2,SLC15A3,SLC16A3,SLC1A3,SLC25A5,SLC28A1,SLC29A1,SLC2A11,SLC2A2,SLC37A4,SLC4A4,SLC5A2,SLC9A3,SLPI,SMAD1,SMAD7,SMAGP,SMTN,SOAT1,SOCS2,SOD2,SP110,SP3,SPON1,SP[illegible] |

| | | | | | | | | | |
|---|---|---|---|---|---|---|---|---|---|
| TP53 | 0,368 | transcription regulator | Activated | 2,950 | 2,974 | 0,001 | 1,46E-56 | 1,76E-53 | A2M,ABAT,ABCC1,ABCC2,ABCG2,ACAA2,ACACB,ACADL,ACADM,ACADS,ACADVL,ACAT1,ACE,ACO1,ACO2,ACOT11,ACSF2,ACSL1,ACSM5,ACTB,ACTN1,ADA,ADD3,ADGRE5,ADH5,ADRB2,AFP,AHCY,AKAP12,AKR1B1,ALB,ALDH18A1,ALDH1B1,ALDH4A1,ALDH9A1,ALOX5,ANKH,ANXA1,ANXA2,ANXA3,ANXA4,APOA1,APOE,APOL1,AQP3,ARAP2,ARPC1B,ASL,ASNS,ASS1,ATF3,ATG4A,AURKA,BAG1,BAX,BCAS3,BCL2A1,BHLHE40,BHLHE41,BID,BIRC2,BIRC3,BNIP3,C2,CALCRL,CALU,CAMK2N1,CAP1,CARHSP1,CASP1,CASP3,CASP4,CASP6,CASP8,CAT,CAV1,CAV2,CCL5,CCNA2,CCNB1,CCNB1IP1,CCNB2,CCND1,CCND2,CCND3,CCNG1,CCS,CD247,CD36,CD44,CD47,CD59,CD81,CDC25B,CDC7,CDH3,CDK1,CDKN1B,CDKN3,CEL,CEP55,CES2,CHST12,CHUK,CKAP2,CKM,CLIC4,CLPP,CLU,CLUH,CNN2,COL18A1,COL1A1,COL1A2,COL3A1,COL4A1,COL4A2,COL5A2,COMT,COPB1,COQ2,COQ3,COQ4,COQ6,COQ8A,CORO1A,COX11,COX15,COX6A2,CPT2,CSF1R,CSHL1,CSK,CSTB,CTF1,CTSB,CTSD,CTSF,CTSK,CXCL1,CXCL10,CXCL12,CYB5A,CYFIP2,DCK,DDB2,DDIT3,DDR1,DDX39A,DDX56,DEK,DHCR7,DHRS9,DICER1,DKK3,DLAT,DLD,DLGAP5,DLST,DNAJA3,DNAJB5,DNAJC12,DNM1L,DNMT1,DOK1,DPEP1,DPP4,DRAM1,DSTN,DTYMK,DUSP1,EBAG9,ECH1,EGF,EIF4G2,EIF4G3,EIF5,ELF4,ENG,ENO3,ENPP2,EPHX1,ERF,ESPL1,ESR1,ESRRG,ETFA,EZH2,F3,FAS,FERMT2,FGF13,FGF7,FH,FHL1,FHL2,FKBP3,FKBP4,FKBP5,FKBP6,FMO1,FN1,FOS,FOSL1,FOXO1,FSTL1,FUBP1,FYN,FZD6,G0S2,G6PC,GADD45A,GADD45B,GADD45G,GATM,GBP1,GJA1,GLB1,GLIPR1,GLRX,GLUL,GNAI1,GNAI2,GNL3,GOLGA4,GPD1,GPD1L,GPR162,GPX3,GSR,GSTP1,HADH,HADHA,HIF1A,HLA-B,HLA-DQA1,HMGB2,HS3ST1,HSPA4L,HSPA8,HYAL1,ICAM1,ID1,ID2,ID3,IDH1,IDH3G,IER5,IFI16,IFI30,IFI35,IGFBP4,IGFBP7,IL10RA,IL4R,IL7,ING2,INKA2,INSR,IRF7,IRF8,IRF9,ISG15,ITGA2,ITGAV,JUND,KCNJ2,KIF22,KITLG,KPNB1,LAMA5,LAMC1,LBR,LIMA1,LMAN2,LOC102724788/PRODH,LPIN1,LRRC3,LSP1,LYZ,MACROH2A1,MAD2L1,MAFB,MAP2K3,MAP4K2,MAPK12,MCAM,MCAT,MCM2,MCM3,MCM5,MCM6,MCM7,MDH2,ME1,ME2,MELK,MET,MFAP2,MGST2,MLF2,MMP1,MMP2,MMP9,MPDZ,MPI,MPZL2,MRPL16,MRPL2,MRPL34,MRPL46,MRPS2,MSH2,MSH6,MSRB1,MX1,MYH9,MYL3,MYOF,NAMPT,NAP1L1,NCK2,NDC80,NDRG1,NDUFAF1,NDUFS1,NDUFS3,NFKB1,NFS1,NID2,NKX2-5,NME1,NOS2,NOX4,NR0B2,NRP1,OAS1,OAT,OGDH,OPA1,OSGIN1,OXA1L,P2RY14,P4HA1,P4HB,PALLD,PBK,PCCA,PCGF2,PCK1,PCLAF,PCNA,PDE6A,PDGFRA,PDIA5,PDIA6,PDK2,PDK3,PDLIM1,PDSS2,PEA15,PECAM1,PERP,PFKFB3,PFKP,PFN1,PGAM2,PHGDH,PHLDA3,PIK3CB,PIK3R3,PINK1,PKM,PLAAT3,PLAAT4,PLEKHB2,PLK2,PLOD2,PLPBP,PLTP,PMM1,POLE2,POSTN,PPARA,PPARGC1A,PPFIBP1,PPIC,PPM1D,PPP1R15A,PPT1,PRC1,PRDX2,PRDX3,PRI[illegible] |

| | | | | | | | | | |
|---|---|---|---|---|---|---|---|---|---|
| TNF | 0,121 | cytokine | Activated | 8,219 | 8,293 | 0,003 | 2,37E-56 | 2,51E-53 | A2M,ABCC1,ABCC2,ABCC3,ABCG2,ABTB2,ACACA,ACADM,ACADS,ACADVL,ACE,ACKR3,ACOX1,ACP5,ACSL1,ACTB,ADAM17,ADM,ADRB2,AEBP1,AK2,AKAP12,AKR1B1,ALAD,ALB,ALDH1A3,ALDH2,ALDH3A1,ALOX5,ALOX5AP,ANXA1,ANXA11,APOA1,APOBEC3B,APOE,APOL1,AQP1,AQP3,ARC,ARF4,ARHGDIB,ARHGEF17,ARL6IP5,ASS1,ATF3,ATP2B4,AVP,B2M,BACE1,BAX,BBOX1,BCKDHA,BCKDHB,BCL2A1,BCL2L13,BCL2L2,BCL6,BHLHE40,BHLHE41,BID,BIRC2,BIRC3,BLVRA,BST2,BTN3A3,C3,C3AR1,CA2,CALR,CARD8,CASP1,CASP3,CASP4,CASP6,CASP8,CAT,CAV1,CAVIN3,CBR3,CCL18,CCL19,CCL20,CCL5,CCN3,CCND1,CCND2,CCND3,CCR2,CCR5,CCR7,CD14,CD163,CD1C,CD200,CD247,CD36,CD38,CD40,CD44,CD47,CD55,CD59,CD69,CD83,CD86,CDH11,CDH3,CDH5,CDKN2C,CFB,CFD,CHRND,CIB2,CIDEC,CISH,CKM,CLIC4,CLTCL1,CLU,COL15A1,COL16A1,COL1A1,COL1A2,COL3A1,COTL1,CPA3,CPT2,CRADD,CSF1R,CSF2RB,CSGALNACT1,CST7,CTSB,CTSF,CTSK,CTSS,CTSZ,CX3CR1,CXCL1,CXCL10,CXCL11,CXCL12,CXCL6,CXCL9,CXCR4,CYBA,CYBB,CYP17A1,CYP1B1,CYP27A1,CYP27B1,CYP2C8,CYTIP,DCN,DDIT3,DDX58,DENND2D,DEPP1,DGAT1,DHRS3,DIO1,DLEU1,DNMT1,DPP4,DPYS,DUSP1,DUSP9,DVL1,ECH1,EFNB2,EGOT,EHD1,EHF,EIF2B2,ELK3,ENG,ENPP2,ENTPD5,ESM1,ESR1,EXT1,F2RL1,F3,FABP1,FAS,FCER1G,FCGR2B,FCGRT,FMO1,FN1,FNBP1,FOS,FOSB,FOSL1,FOXO1,FOXO4,FPR1,FRZB,FSCN1,FUT3,FUT4,FYN,FZD5,G0S2,GADD45A,GADD45B,GADD45G,GASK1B,GBP1,GBP2,GCH1,GCLC,GCLM,GHR,GJA1,GLS,GM2A,GNAI1,GNAI2,GNAI3,GOSR2,GPD1,GRN,GSTA1,GSTP1,HDAC1,HERC1,HEXB,HIF1A,HIPK3,HLA-A,HLA-B,HLA-DRA,HLA-E,HLA-F,HLA-J,HMGN3,HNF4A,HPGD,HPGDS,HPRT1,HSD3B2,HSP90B1,HSPA5,HSPA8,ICAM1,ICAM2,ID1,ID3,IDO1,IFI16,IFI27,IFI6,IFIH1,IFIT3,IFIT5,IFITM1,IFNAR2,IFNGR1,IFNGR2,IGFBP4,IGFBP6,IGKC,IL10RA,IL13,IL15,IL15RA,IL18,IL1R1,IL32,IL33,IL36A,IL4R,IL7,IL7R,INHA,INPP5D,INSR,IRF1,IRF7,IRF8,ISG15,ITGA2,ITGAL,ITGAM,ITGAV,ITGB2,ITGB3,ITGB6,ITGB8,JAK1,JUND,KCNJ2,KIF20A,KITLG,KL,KLRB1,KMO,KYNU,LAMC1,LAMP3,LCN2,LITAF,LPAR6,LRRC8D,LTB,LXN,LY96,LYN,MAFF,MAP2K3,MAP4K4,MAPKAP1,MARCKSL1,MAT2A,MCAM,MCM3,MECOM,MEF2C,MET,MFHAS1,MGP,MGST2,MIF,MMD,MMP1,MMP2,MMP7,MMP9,MPC1,MSH2,MTTP,MUC1,MX1,MYD88,MYH9,MYL6,NAGLU,NAMPT,NCF2,NCK1,NEDD9,NET1,NFE2L2,NFKB1,NFKBIE,NFS1,NGF,NME1,NNMT,NOCT,NOD2,NOS2,NOX4,NPHS1,NPPA,NR0B2,NR1H3,NR1I3,NR4A1,NR4A3,NRP1,NUAK1,NUCB2,OAS1,OAS2,OAS3,OLFML2B,OPTN,OSMR,P4HB,PAK2,PAPPA,PARP1,PC,PCBP2,PCK1,PCK2,PCNA,PDGFRA,PDIA4,PDK3,PDZD2,PECAM1,PEMT,PHB,PHGDH,PIGR,PIK3C2B,PIK3CB,PKM,PLA2G4A,PLA2G4C,PLAAT3,PLAAT4,PLAGL2,PLIN2,PLK2,PLOD2,PLSCR1,POSTN,POU2AF1,PPARA,PPARGC1A,PPP1R15A,PPP1R3C,PRKCA,PRK [illegible] |

| | | | | | | | | |
|---|---|---|---|---|---|---|---|---|
| tretinoin | chemical - endogenous mammalian | Activated | 4,365 | 4,429 | 0,003 | 5,38E-50 | 5,05E-47 | A2M,ABCG1,ACACB,ACOX1,ACSL1,ADA,ADAM10,ADAR,ADD3,ADGRE2,ADM,AFP,AGO1,AHNAK,AHR,AKAP12,AKR1A1,ALDH1A3,ALDH3A1,ALDH9A1,ALOX5,ALPL,ANXA1,ANXA2,ANXA5,AP1S2,APOC3,APOE,AQP1,ARHGAP29,ASNS,ATP6V1B2,ATR,AURKA,BACE1,BCL2A1,BCL6,BHLHE40,BHMT,BID,BIRC2,BIRC3,BNIP3,BNIP3L,BPI,C12orf29,C1QA,C3,C3AR1,CA2,CA8,CALB1,CALM1 (includes others),CALML3,CALR,CAPN2,CASP1,CASP3,CASP6,CAVIN3,CBFB,CCL19,CCL20,CCN3,CCNA2,CCNB1,CCND1,CCND2,CCND3,CCNG1,CCR5,CCT5,CD14,CD1C,CD1D,CD36,CD38,CD44,CD47,CD52,CD53,CD63,CD74,CD86,CD9,CD93,CDH5,CDH6,CDK5,CDKN1B,CDKN2C,CDKN3,CDX2,CEBPE,CGA,CHN2,CLDN1,CLPS,COL1A1,COL1A2,COL3A1,COL4A1,COL4A2,COL6A3,COTL1,COX11,CPE,CRABP1,CRAT,CSE1L,CSF1R,CSHL1,CSNK1A1,CTSB,CTSD,CTSK,CTSS,CUBN,CX3CR1,CXCL1,CXCL10,CXCL6,CXCR4,CYBB,CYP17A1,CYP2B6,CYP4A11,CYP4F2,DAB2,DAPK2,DCN,DDB2,DDIT3,DDX10,DDX58,DDX60,DGAT1,DHPS,DHRS3,DHRS9,DHX15,DKK3,DLEU1,DLGAP5,DNMT1,DPP4,DTL,DUSP1,DYNLL1,ECI2,EHD1,EIF4EBP2,EIF5,EIF5B,EIF6,ELF4,EMP3,ENPP2,EPHX1,EYA2,F3,FCER1G,FGR,FLRT1,FN1,FOLR2,FOS,FOSL1,FRAT2,FUT4,G0S2,GARS1,GAS2,GCA,GDF2,GHR,GIMAP6,GJA1,GJB1,GJB4,GM2A,GNAS,GPR21,GRB10,GSTP1,GZMA,HADHA,HDAC1,HK1,HLA-A,HLA-B,HLA-C,HLA-DMB,HMGCS2,HNF1A,HNF1B,HNF4A,HNRNPF,HOXA1,HOXA10,HOXB1,HOXB2,HOXB5,HOXB6,HOXB7,HPRT1,HS3ST1,HSD17B11,HSF4,ICAM1,ID1,ID2,ID3,IFI16,IFI27,IFI35,IFI44,IFI44L,IFI6,IFIH1,IFIT2,IFIT3,IFIT5,IFITM1,IFNGR1,IGFBP4,IGFBP6,IGFBP7,IGHM,IL10RA,IL13,IL15,IL1R1,IL6R,IMPDH2,IRF1,IRF9,ISG15,ITGA2,ITGAL,ITGAM,ITGB2,ITGB3,IVD,JAK1,JCHAIN,KCNJ2,KITLG,KLF15,KLF9,KRT19,KRT7,LAMB1,LCN2,LEPR,LGALS1,LGALS3BP,LILRB2,LIPA,LRP5,LSP1,LTF,LY6G6D,LYN,LYPD1,MAFB,MAOB,MAP2K3,MAP3K1,MAP3K10,MAP7,MAPK10,MAPK8,MARCKSL1,MDK,MECOM,MEF2C,MEIS2,METTL7A,MFNG,MGP,MID1,MLF2,MMP1,MMP7,MMP9,MNDA,MOK,MRAS,MS4A1,MSH3,MTHFD2,MYB,MYH9,NAE1,NCF2,NCL,NDRG1,NDUFAF4,NDUFB7,NDUFV1,NFE2L2,NFKB1,NID2,NKX2-5,NMI,NOS2,NPHS1,NPHS2,NPPA,NR0B2,NR2F2,NRP1,NTRK2,OAS1,OAS2,OAS3,OAT,OLFM4,OPTN,OXT,P4HA2,PALLD,PCK1,PDCD4,PDGFRA,PDLIM4,PEX1,PFN1,PKP2,PLA2G7,PLAAT4,PLCE1,PLD1,PLEK,PLEKHO2,PLIN2,PLK2,PLS3,PLSCR1,PNP,POR,POSTN,PPARA,PPARGC1A,PPP1R16B,PRDX2,PRKCA,PRKCD,PRKCH,PRMT1,PRNP,PROM1,PSMB10,PSMB8,PSMB9,PTEN,PTGES,PTGS1,PTPRC,RABGGTB,RAE1,RARG,RARRES1,RB1,RBP1,RFC2,RHOA,RNASE1,RORC,RPA1,RPL3,RPS16,RPS19,RPS20,RPS3,RTP4,RUNX3,RXRA,S100A10,S100A8,S100A9,SACS,SAMHD1,SCD,SCML1,SDCBP,SELL,SELP,SERPINB9,SERPINE2,SERPING1,SFRP1,SHANK2,SKAP2,SLA,SLC12A2,SLC25A20,SLC25A36,SLC27A2,SLCO2A1,[illegible] |

| | | | | | | | | |
|---|---|---|---|---|---|---|---|---|
| IL4 | cytokine | | 0,872 | 0,953 | 0,005 | 1,01E-42 | 8,52E-40 | ACOX1,ACP5,ACSL1,ACTG1,ADAM17,AHNAK,AHR,AK2,ALDH2,ALOX5,ALOX5AP,ANXA1,ANXA2,ANXA4,APOE,APOLD1,APRT,ARG2,ARNTL2,ARSB,ASCC3,ASS1,ATP1A3,AUH,BAX,BCL11A,BCL6,BHLHE40,BIN1,BIRC2,BNIP3,BNIP3L,C3,CA2,CAPG,CASP3,CASP6,CAT,CBFB,CBX3,CCL18,CCL20,CCL5,CCL8,CCND1,CCND2,CCND3,CCR5,CCR7,CCT3,CD14,CD163,CD1C,CD2,CD200,CD300A,CD36,CD38,CD40,CD44,CD55,CD58,CD69,CD74,CD83,CD86,CD8A,CDK1,CDKN1B,CELF2,CISH,CKAP4,CLCN3,CLEC10A,CLEC7A,CLUH,COL18A1,COL1A1,COL1A2,COL6A3,CRADD,CRIP1,CSF1R,CSF2RB,CST7,CTNS,CTSK,CXCL1,CXCL10,CXCL11,CXCL6,CXCL9,CXCR4,CYB5A,CYBB,DARS2,DDAH1,DECR1,DHODH,DOCK10,DPP4,EPHX1,EXT1,F2RL1,F3,FAAH,FAM49A,FAS,FCER1G,FCGR2B,FCGR3A/FCGR3B,FKBP5,FLNA,FN1,FOS,FOSB,FSCN1,GADD45G,GALK2,GATB,GATM,GBP2,GCAT,GJA1,GPM6B,GPT,GZMA,HCK,HCLS1,HDAC1,HDDC2,HLA-DMA,HLA-DMB,HPGD,HRH1,HSD3B2,ICAM1,ID3,IDO1,IFI16,IFI30,IFI44,IFIH1,IFIT3,IFNAR2,IFNGR2,IGFBP4,IL10RA,IL13,IL13RA1,IL15,IL15RA,IL18,IL1R1,IL2RB,IL33,IL4R,IL7R,IRF1,IRF7,IRF8,IRF9,ISG15,ISG20,ITGA2,ITGAL,ITGAV,ITGB2,ITGB3,ITPA,JAK1,JCHAIN,JUND,KCNAB2,KCTD12,KITLG,KLHDC2,KLRB1,KLRD1,KMO,KRT10,LCP2,LGALS1,LGALS2,LGALS3BP,LGALSL,LMAN2,LRPPRC,LSP1,LTB,LY75,MAF,MAFB,MAL,MAOA,MCM5,MMP1,MMP2,MMP9,MRC1,MS4A1,MSH2,MUC1,MYH9,MYOF,NAB2,NAP1L1,NCL,NDRG1,NEDD9,NFIL3,NFKB1,NOCT,NOS2,NR4A3,NRP1,NUP155,NUP214,OAS3,PFKL,PFKP,PHB,PIGR,PLA2G4C,PLD1,PLIN2,PLOD2,PLSCR1,PMP22,PMS1,PMVK,PNP,POLE2,POSTN,POU2AF1,PPARGC1A,PRF1,PRKCA,PRKCD,PRMT1,PRNP,PRSS8,PSMA6,PSMB10,PSMB6,PSMB8,PSME1,PSMG1,PTPRCAP,RHOA,RIDA,RIN1,RNF19B,RNPS1,S100A10,S100A8,S100A9,SAMSN1,SDF2L1,SEC31A,SELL,SELP,SEMA4D,SH2D1A,SLC29A1,SMC3,SNX4,SOCS2,SOX13,SPHK2,SPINT2,SPP1,ST14,ST6GAL1,ST7,STAB1,STAT1,STAT4,STK39,STUB1,SWAP70,SYK,SYNGR2,SYT11,TAP2,TAPBP,TBC1D17,TES,TFEC,TFF3,TGFBI,TGFBR2,THY1,TIMP1,TIMP2,TLE1,TLN2,TLR2,TLR4,TM7SF2,TNC,TNFRSF11B,TNFRSF17,TP53,TREM2,TXLNA,TYRO3,UBL3,UQCRC1,USP2,VCAM1,VDR,VEGFA,VIM,VPS13C,VPS35,VTCN1,XBP1,XPO |
| OSM | cytokine | Activated | 6,761 | 6,818 | 0,004 | 7,97E-42 | 6,13E-39 | A2M,ABCC1,ABCC3,ABCC5,ABCC8,ABCG1,ACAA1,ACE,ACSL5,ADAM17,ADAMTS1,ADH5,AFP,AHR,AKAP3,ALB,ALDH3B1,ANXA1,ANXA11,ANXA2,ANXA3,ANXA9,ARL4C,ASNS,ATF3,ATP2B4,ATP9A,B2M,BAIAP2,BBOX1,BHLHE40,BST1,C1R,C1S,CAP1,CASP4,CAT,CBX3,CCL20,CCL5,CCND1,CCND2,CCT2,CDA,CDH3,CDKN1B,CGA,CISH,CKM,CLEC2B,CLPP,CLPX,COL1A1,COL1A2,COL3A1,COX7B,CST6,CTSV,CXADR,CXCL1,CXCL10,CXCL12,CYP1B1,DENND5A,DHRS3,DHX9,DNM1L,ECM2,EMP3,ERBB3,ETFB,EVPL,EXOSC10,F3,FLNA,FMO5,FN1,FOS,FOSB,FOSL1,GADD45G,GBP1,GBP2,GCA,GFPT1,GLUL,GPNMB,GRHPR,HIF1A,HIPK3,HLA-A,HLA-B,HLA-C,HLA-F,HMG20B,HMGN4,HSD11B2,HSF4,HSPA5,ICAM1,ID1,ID2,IFI35,IFNGR1,IGFBP6,IL13,IL13RA1,IL15,IL15RA,IL18,IL32,IL33,IL4R,IL6R,IL7,IRAK1,IRF1,IRF7,IRF9,ISG20,ITGA7,ITGAL,KLK1,KPNB1,KRT19,LCN2,LITAF,LRRFIP1,LSM4,MAOA,MARCKS,MCM7,MLLT11,MMP1,MMP2,MMP9,MPDU1,MPZL2,MRC1,MT1X,MX1,MYD88,NAMPT,NEDD4L,NMT2,NOS2,NR1I3,NUAK1,OAS1,OGDH,OSMR,PAK2,PCNA,PDCD4,PDZK1IP1,PEPD,PFKFB2,PFKFB3,PIGR,PKIG,PLA2G4A,PLCB4,PRDX2,PSMB8,PSMB9,PTEN,PTGES,PTOV1,PTP4A1,PYGL,QSOX1,RAB31,RARG,RBFOX2,RORA,RYK,S100A8,S100A9,SCNN1B,SELP,SERPINA3,SERPINB1,SERPING1,SLC16A3,SLC7A8,SLPI,SMPD1,SPP1,STAT1,STAT3,STK25,SULT2B1,TAP1,TAP2,TAPBP,TDO2,TDP2,TERT,TIMP1,TLR2,TLR3,TM4SF1,TM7SF2,TMBIM6,TNC,TNFAIP2,TNFRSF11B,TNNC1,TOP2A,TP53,TPBG,TPM1,TPX2,TRIM22,TSTA3,TUBA1A,TYRO3,UAP1,UBE2L6,UBQLN2,VCAM1,VDR,VEGFA,VRK2,ZFP36,ZNF266 |

| | | | | | | | | | |
|---|---|---|---|---|---|---|---|---|---|
| MYC | 0,375 | transcription regulator | | -1,575 | -1,518 | 0,003 | 2,07E-41 | 1,46E-38 | ABCC1,ABCC3,ACACA,ACAT1,ACTB,ACTN1,ADAMTS1,ADIPOR1,ADM,AFP,AHCY,AK2,AKAP1,AKAP12,ALB,ALDH18A1,ALOX5,ANXA4,ANXA5,APOC3,AQP1,ARAF,ARF3,ARHGAP25,ASNS,ASS1,AXL,BAG1,BAX,BCKDHB,BCL2A1,BCL6,BIN1,BIRC2,BZW2,C1QBP,CAMK2N1,CAPN2,CASP1,CASP3,CASP8,CAST,CAV1,CCNA2,CCNB1,CCNB2,CCND1,CCND2,CCND3,CCT3,CD151,CD44,CD47,CD48,CD69,CD86,CD9,CDC25B,CDK1,CDKN1B,CDX2,CEBPE,CHP1,CHST15,CIDEC,CLIC4,CLPB,CLU,CLUH,COL15A1,COL1A1,COL1A2,COL3A1,COL4A1,COL4A2,COL5A1,COL5A2,COL6A3,COX5B,CPD,CPT2,CRIP1,CSE1L,CSF1R,CST6,CSTB,CTSB,CTSD,CTSV,CUL1,CXCL10,CXCL11,CXCL9,CYFIP2,DBN1,DCTPP1,DDB2,DDIT3,DEK,DLEU1,DNMT1,DNPH1,DROSHA,DUSP1,ECSIT,EED,EFEMP1,EIF3D,ENO1,ERAP1,EVPL,EVX1,EXOSC8,EXT2,EZH2,F3,FABP1,FABP2,FARP1,FAS,FBN1,FCGRT,FKBP5,FLNA,FN1,FOS,FOSL1,FRZB,FSTL1,FUT3,G6PC,GADD45A,GADD45B,GADD45G,GAMT,GBP2,GCLC,GCLM,GFPT1,GGH,GGTLC1,GJA1,GLS,GLUD1,GLUL,GLYR1,GM2A,GOT1,GOT2,GPC3,GPT,GRPEL1,GSR,HIF1A,HLA-A,HLA-B,HLA-E,HMOX2,HSD11B2,HSPA9,ICAM1,ID1,ID2,ID3,IDH1,IER5,IFI16,IFI27,IFI35,IFI44,IFI44L,IFI6,IFIH1,IFIT2,IFIT3,IFIT5,IFNA10,IMPA2,IQGAP2,IRF7,IRF9,ISG20,ITGA3,ITGAL,ITGAM,KLK1,KRAS,KRT7,LGALS1,LIMA1,LOC102724788/PRODH,LSM4,LUM,LXN,LYZ,MAD2L1,MAT2A,MCM5,MCM6,MCM7,ME2,MFAP1,MGP,MIF,MMP7,MMP9,MRC1,MSH2,MSN,MTHFD1,MX1,MX2,MYD88,MYO1B,NAP1L1,NCL,NDRG1,NDUFS4,NFE2L2,NGF,NIBAN1,NME1,NMI,NOCT,NPPA,NRP1,NUCB1,OAS1,OAT,OVOL1,PARP1,PCK1,PCNA,PDCD4,PDGFRA,PECAM1,PERP,PEX19,PFKFB3,PFKL,PFKP,PHB,PHF21A,PKLR,PKM,PLS3,PLSCR1,PMP22,POLR2G,PPARA,PPID,PPL,PRDX2,PRDX3,PRDX4,PRKACB,PRMT1,PROM1,PSAT1,PSMB8,PTEN,PTPRC,RAB40B,RARG,RARRES1,RARS1,RB1,RBBP7,RBP1,RHOA,RHOB,RIOX2,ROCK1,RPL23,RPL3,RPS16,RPS19,RPS20,RRM2,RSAD2,S100A10,S100A6,SAE1,SARDH,SDCBP,SEPHS2,SERINC3,SERPINE2,SFRP1,SFXN1,SGK1,SHMT1,SHMT2,SIRT2,SLC16A3,SLC16A7,SLC1A5,SLC22A4,SLC25A5,SLC2A2,SLC38A1,SLC3A2,SMAD1,SNRPD1,SNRPD2,SOD2,SP3,SPARC,SPP1,SQOR,ST3GAL1,ST3GAL4,STAT1,STAT3,STMN1,SUCLA2,TAGLN2,TERT,TES,TGFBR2,THBS2,THY1,TIAM1,TIMM23,TIMP1,TIMP2,TKT,TLE1,TLR7,TMSB10/TMSB4X,TNC,TNFSF10,TOB1,TP53,TPI1,TPP2,TRAP1,TRIM5,TRIP12,TSPAN7,TSPO,TXN,TXNIP,UBE2C,UBE2I,VCAM1,VEGFA,VEGFC,VIM,VLDLR,WNT8B,XPO1,YBX3,YRDC,YTHDC1,ZFP36 |
| ERBB2 | | kinase | Activated | 2,530 | 2,591 | 0,005 | 2,09E-39 | 1,36E-36 | ABCG2,ACAA2,ACTB,ADAM17,AGR2,AHNAK,ALAD,ANG,ANXA2,AURKA,BACE1,BARD1,BCKDHB,BHLHE40,BIRC3,BMP7,BNIP2,BNIP3,CAPZA2,CAVIN1,CAVIN3,CCL20,CCL5,CCNA2,CCNB1,CCNB2,CCND1,CCND2,CCND3,CD36,CD47,CD59,CD9,CDC14B,CDC25B,CDC37L1,CDC42SE1,CDC7,CDCP1,CDH11,CDH3,CDK1,CDKN1B,CDKN2C,CDKN3,CHUK,CIDEC,CLDN3,CLU,COL18A1,COL1A1,COL3A1,COL4A1,COL5A1,COL5A2,COL6A3,CPT2,CRADD,CRIP1,CSE1L,CSF1R,CTSB,CUL1,CXCL10,CXCL12,CYP2J2,DDIT3,DDX10,DECR1,DERL1,DGAT1,DOK1,DPEP1,DSG2,EBP,EHF,EHHADH,EIF6,ELF2,ELK3,ELL2,EMP3,EPHX2,ERBB3,ESPL1,ESR1,ETFB,FEZ2,FGF7,FHIT,FN1,FOLR1,FOS,FOXO1,FSCN1,FSTL1,FZD4,GALNT3,GDPD3,GHR,GINS1,GINS2,GJA1,GLS,GNAI1,GPX3,GSTZ1,GZMH,HEPH,HIF1A,HMGB2,HSD17B11,HSPB8,HTRA1,ID1,ID2,IDH1,IDI1,IGFBP4,IGFBP6,IL6R,ITGA2,ITGB3,JAK1,JAM2,JCHAIN,KDM5B,KRT7,KRT81,LAMB1,LCN2,LGALS1,LITAF,LPCAT1,LRRFIP1,LTBP3,LUM,LXN,MACF1,MAF,MAOA,MAP2K3,MAPK13,MCCC1,MCM2,MCM3,MCM5,MCM6,MCM7,MFAP2,MFNG,MIF,MME,MMP1,MMP7,MMP9,MRC1,MUC1,MXI1,NDC80,NDRG1,NDUFAB1,NDUFV1,NEDD9,NET1,NFKB1,NNMT,NOS2,NOX4,NR1H2,NR1H3,NRP1,NUCB2,ORC6,P4HA2,PCK2,PCNA,PDCD4,PDE8A,PDIA4,PDK4,PDLIM4,PFKFB3,PGM1,PHYH,PINK1,PLAC8,PLIN2,PMVK,POLD3,POLDIP2,POLE2,POLR2G,PRC1,PRDX2,PRIM1,PROM1,PRSS3,PSMB9,PTEN,PTGES,PTGS1,PTPRK,RAB31,RARG,RECK,RETREG1,RFC2,RHOB,RPA1,RPA2,RRM1,RRM2,S100A4,S100A6,SDC1,SERPINA3,SFRP1,SHMT1,SLC66A2,SLPI,SMAD2,SMAD7,SMTN,SOCS2,SOWAHC,SOX4,SPAG5,SPARC,SPINT2,SQOR,SRPX,ST3GAL4,ST3GAL6,STAT3,STK17B,TAP1,TAP2,TAPBP,TERT,TGFBI,THBS4,THY1,TJP1,TMEM43,TNC,TOP2A,TOPBP1,TP53,TPD52L1,TPM1,TSC22D1,TUBA1A,UBE2C,USP14,VCAN,VDR,VEGFA,VEGFC,VIM,VWF,WT1,WWC1,XBP1,ZWINT |

| | | | | | | | | |
|---|---|---|---|---|---|---|---|---|
| ESR1 | -0,197 | ligand-dependent nuclear receptor | Activated | 2,337 | 2,361 | 0,001 | 1,50E-38 | 9,04E-36 | AASS,ABCC3,ABCC5,ABCG2,ACE,ACKR3,ACOX1,ACSL1,ACTR2,ADA,ADAM17,ADD3,AHNAK,AKAP1,ALOX5,ANXA3,ANXA4,APOA1,APOE,AQP1,AQP3,ARAF,ARFIP2,ARHGEF9,ASS1,ATF3,ATP6V0D1,AURKA,AVP,BAG1,BAX,BCAS3,BCCIP,BCL2L2,BCL6,BCLAF1,BIN1,BIRC3,C3,CA2,CALB1,CALML4,CAP2,CAPN2,CASP6,CASP8AP2,CAV1,CAV2,CCL20,CCN3,CCNA2,CCNB1,CCND1,CCND3,CCNG1,CCNH,CD55,CD59,CD69,CD83,CDC42SE1,CDK1,CDK5,CDKN1B,CDKN1C,CDKN3,CELSR2,CENPM,CENPU,CEP135,CGA,CHFR,CLCN3,CLDN1,CLEC7A,CLTCL1,COL16A1,COL5A2,COMT,COTL1,CPE,CRADD,CSNK1A1,CTNS,CTSB,CTSD,CTSS,CXADR,CXCL1,CXCL12,CYFIP1,CYFIP2,CYP17A1,CYP1B1,CYP4A11,DAB2,DDB2,DECR1,DHCR7,DLG5,DMTN,DSG2,DUSP9,DYRK2,EFEMP1,EFNB2,EGF,ENAH,ENO1,ENPP2,ERBB3,ERGIC2,ESR1,ESRRA,F12,F2RL1,F3,FABP1,FAM102A,FAS,FHL1,FIS1,FKBP4,FLNA,FOLR1,FOS,FOSB,FOSL1,FOXO1,FOXO4,FRAT2,FZD2,GADD45B,GALNT7,GGA2,GHR,GJA1,GJB1,GLB1,GLRX,GM2A,GNAS,GNB1,GPER1,GPSM2,GUCY1A1,GUCY1B1,HACD1,HAX1,HCP5,HIF1A,HIP1R,HLA-B,HLA-C,HLA-E,HLA-G,HLA-J,HMGCS2,HOMER3,HOOK2,HPGDS,HSD3B2,HSP90B1,HTRA1,HYAL1,ICAM1,ID1,IFI27,IFI44,IFI44L,IFI6,IFITM1,IFITM2,IFRD1,IGFBP4,IMPDH2,INSR,IQGAP1,IRF1,ISG20,ITGB1BP1,JAK1,JUND,KIF20A,KL,KLHL20,KPNB1,KRAS,KRT19,KRT7,LAMB1,LAPTM5,LCK,LCN1,LCN2,LEMD3,LEPR,LGALS1,LGALS3BP,LHB,LIMA1,LTB,LTF,LYPD1,MAD2L1,MAP3K1,MAP3K12,MAP3K13,MAP7,MAPK12,MAPK8,MAPT,MARCKS,MAST2,MCF2L,METTL7A,MMD,MME,MMP1,MMP9,MPC1,MPDZ,MR1,MX1,MYB,NCF2,NEDD9,NFKB1,NFKBIE,NFYB,NIPSNAP1,NLGN4X,NME1,NOS2,NPR3,NR0B2,NR1H2,NR1H3,NUCB1,NUP214,OIP5,OPA1,OSTF1,OXT,PAK2,PALLD,PBK,PCBD1,PCGF2,PCNA,PCYOX1,PDCD4,PDCD5,PDK4,PGRMC2,PHB,PIBF1,PLA2G4C,PLCE1,PLIN2,PLOD2,PLS1,POR,PPARGC1A,PPP1R12A,PPP1R3C,PPP6R2,PRKACB,PRKCD,PRKCH,PRKCI,PRKD3,PRKX,PRLR,PROM1,PSAT1,PSIP1,PTEN,PTGER3,PTP4A2,PTPRT,PTTG1,PTTG1IP,PYGL,RAB27A,RAB31,RAB5C,RAI2,RAPGEF6,RB1,RBM22,RDX,RENBP,RGS19,RND3,ROBO1,ROCK1,RORC,RSAD2,RTP4,S100A9,SAMD9,SAMHD1,SCD,SCNN1B,SEC31A,SELL,SEMA3B,SERPINB9,SFXN3,SGK1,SHANK2,SIGIRR,SIPA1L1,SLC16A3,SLC25A36,SLC26A2,SLC2A4RG,SLC39A4,SLC3A2,SLC7A8,SLC9A3,SLC9A3R1,SMAD2,SMC3,SMC4,SMPD1,SNN,SOCS2,SOX4,SP110,SPAG9,SPP1,SPTLC2,SRPK2,STAT1,STAT3,STC2,SUCLA2,SULT1C2,TAPBP,TCIRG1,TDO2,TEAD4,TERT,TESK1,TFF3,THBS4,THRB,TIMM17B,TIMP2,TJP1,TM4SF1,TMED10,TMSB15A,TNFAIP2,TNFAIP6,TNFRSF11B,TNFSF10,TOB1,TP53,TP53BP1,TPD52L1,TPM1,TPP2,TRAM1,TRIB3,TSKU,TSPO,TSPYL2,TTK,TXNIP,TYK2,UBE2I,UBL3,VEGFA,VEGFC,VEZF1,VIM,VPS26C,VPS35,VRK2,WARS1,WASF2,WNK1,WTAP,XB |
| progesterone | | chemical - endogenous mammalian | | -1,082 | -1,049 | 0,002 | 5,90E-34 | 3,12E-31 | ABCG2,ACAT1,ACE,ACOT7,ACOX1,ACP5,ACSL1,ACTB,ADAMDEC1,ADAMTS1,ADH5,ADM,ADRA2A,ADRB2,AEBP1,ALPL,ANKH,APOE,AQP1,ATF3,ATP6V1B2,AURKA,BAX,BCL6,BHLHE40,BIRC3,C3,CA2,CASP3,CAV1,CCNA2,CCNB1,CCNB2,CCND1,CCND3,CCNG1,CCR2,CCR5,CD1D,CD36,CD40,CD86,CDH11,CDKN1B,CFB,CFD,CIDEC,CLDN3,COL15A1,COMT,CTSF,CTSV,CXCL10,CXCL11,CXCL12,CXCR4,CYP1B1,CYP2A6 (includes others),CYP2B6,CYP2C8,CYP3A4,DDIT3,DICER1,DPP4,EGF,EIF5B,ELL2,ENC1,ENG,ENPEP,ENPP2,ESPL1,ESR1,F2RL1,F3,FAAH,FAM49A,FKBP3,FKBP4,FKBP5,FMO5,FN1,FOS,FOXO1,FRAT2,GABARAPL1,GAD1,GARS1,GCH1,GHR,GJA1,GNMT,GRB10,GRINA,GSTA3,HIF1A,HLA-DQB1,HLA-G,HOPX,HOXA10,HSD11B2,HSPA8,ICAM1,ID1,ID3,IHH,IL13,IL15,IL6R,INHA,INSR,ISG20,ITGA2,JAM2,KLF9,KLK1,KRT7,LCN2,LEPR,LEPROT,LGALS1,LHB,LITAF,LPIN1,LTF,LXN,MAD2L1,MAOA,MAP2K3,MCM2,MCM3,MCM5,MCM6,MCM7,ME1,MME,MMP1,MMP2,MMP7,MMP9,MRC1,MUC1,MYB,MYD88,NDRG1,NEDD9,NET1,NFIL3,NOP16,NOS2,NPL,NPY1R,NR1H3,NR4A1,NRP1,NSUN5,OXT,PARM1,PBX3,PCDH1,PCK1,PCNA,PDK3,PDLIM1,PDXDC1,PDZK1IP1,PFKFB3,PGRMC1,PHF21A,PIGR,PKM,PLA2G7,PLK2,PMP22,POSTN,PRLR,PRNP,PTGER3,PTGS1,RAB11FIP5,RASSF2,RBP1,RRM2,S100A6,SCD,SCNN1B,SFN,SGK1,SLC1A5,SLC31A2,SMAD2,SMPDL3A,SOD2,SPP1,SPSB1,SRSF7,ST3GAL1,ST3GAL4,STAT1,STAT3,STX18,TACSTD2,TBX2,TDO2,TERT,TGFBI,TGFBR3,TIAM1,TIMP1,TIPARP,TNFRSF21,TNFSF10,TP53,TRAF5,UGCG,VDR,VEGFA,VIM,WFDC2 |

| | | | | | | | | | |
|---|---|---|---|---|---|---|---|---|---|
| Interferon alpha | | group | Activated | 9,087 | 9,160 | 0,006 | 3,03E-32 | 1,35E-29 | ADA2,ADAR,AFP,AIM2,ANXA5,APOBEC3A,APOBEC3B,APOBEC3F,APOBEC3G,APOL1,APOL3,AQP3,ASCC2,ATF3,AXL,B2M,BCL2A1,BCL2L13,BCL6,BST2,C3,C3AR1,CASP1,CASP3,CASP8,CCL19,CCL5,CCND1,CCND2,CCND3,CCR5,CCR7,CD1D,CD38,CD40,CD69,CD81,CD83,CD86,CD8A,CDKN1B,CISH,CLTCL1,CSF2RB,CTH,CXCL1,CXCL10,CXCL11,CXCL9,CYP3A4,DDX58,DEFB1,DYNLT1,ENPP2,F3,FAS,FCER1A,FCGR2B,FOS,GBP1,GBP2,GIMAP4,GLS,GZMB,GZMH,HERC6,HLA-A,HLA-B,HLA-C,HLA-F,HLA-G,ICAM1,IDO1,IFI16,IFI27,IFI35,IFI44,IFI44L,IFI6,IFIH1,IFIT2,IFIT3,IFITM1,IFITM2,IFNAR2,IGFBP4,IL10RA,IL13,IL15,IL15RA,IL1R1,IL32,IL4R,IL6R,IL7,IL7R,INSR,IRF1,IRF7,IRF8,IRF9,ISG15,ISG20,ITGAM,JAK1,LAMP3,LCK,MNDA,MX1,MX2,MYD88,MYLIP,NFIL3,NKG7,NMI,NOS2,OAS1,OAS2,OAS3,PARP12,PHF11,PLAAT4,PLSCR1,PRF1,PSMB8,PSMB9,PTGES,RB1,RBX1,RNASEL,RSAD2,RSPH14,RTP4,S100A9,SAMD9,SERPINB1,SERPINB9,SIGLEC1,SLC1A5,SLC2A11,SOCS2,SP110,SPRY1,SPRY2,STAT1,STAT3,STAT4,TAP1,TAP2,TAPBP,TDRD7,TENT4A,TERT,TLR1,TLR2,TLR3,TLR4,TLR7,TLR8,TMEM140,TNFSF10,TOPBP1,TRANK1,TRIL,TRIM21,TRIM22,TRIM5,TRIOBP,UBA7,UBE2L6,VCAM1,WARS1 |
| IL6 | -0,135 | cytokine | Activated | 3,486 | 3,556 | 0,005 | 1,05E-31 | 4,44E-29 | A2M,ABCC1,ABCC2,ABCC3,ABCC5,ABCG2,ACOX1,ADAMTS1,ADGRE5,ADGRL2,ADM,AFP,AHNAK,AHR,ALB,ANG,ANXA1,APOA1,APOE,ARL4C,ATF3,AVP,BAX,BCL6,BST2,C1QL1,C3,CAPRIN1,CASP1,CASP3,CASP4,CCL20,CCL5,CCNA2,CCNB1,CCNB2,CCND1,CCR2,CCR5,CCR7,CD14,CD163,CD36,CD38,CD40,CD44,CD48,CD53,CD74,CD83,CD86,CDK1,CDKN1B,CDKN3,CDX2,CES2,CFD,CFH,CIB2,CISH,CLEC10A,CLU,COL1A1,COL3A1,CSF1R,CSF2RB,CTSK,CX3CR1,CXCL1,CXCL10,CXCL6,CXCR4,CYBB,CYP1B1,CYP2B6,CYP2C8,CYP3A4,CYTIP,DCN,DIO1,DNMT1,DUSP1,DVL1,ENPP2,EZH2,F12,F3,FAH,FAS,FCGRT,FLI1,FN1,FOS,FOSL1,FSCN1,G6PC,GADD45A,GADD45B,GADD45G,GALE,GBP2,GCH1,GINS1,GLRX,GRK2,GSTA1,GZMB,HIF1A,HLA-A,HLA-DMA,HLA-DQA1,HOMER3,HPGD,HSPA5,ICAM1,ID1,ID2,IDO1,IFI16,IFIT2,IGFBP4,IGFBP6,IGHM,IK,IL13,IL15,IL1R1,IL4R,IL6R,IL7,IL7R,IRF1,IRF9,ITGAM,ITGAV,ITGB3,JAK1,JCHAIN,JUND,KIF22,KLRB1,KLRD1,LCN2,LGALS1,LTF,LY86,LY96,LYZ,MACROD1,MAD2L1,MAF,MCM3,MET,MIA2,MMD,MMP1,MMP2,MMP7,MMP9,MRAS,MTOR,MTTP,MUC1,MYB,MYD88,NAMPT,NCF2,NFKB1,NGF,NOS2,NUCB2,OIP5,OSBPL1A,PCLAF,PCNA,PECAM1,PHB,PLG,PLK2,PPP1R15A,PRF1,PRNP,PROM1,PSMB10,PSMB8,PSMB9,PSME1,PTGES,PTPRC,PTTG1,RAB27A,RB1,REG1A,RNASE6,RORA,RORC,RRM2,S100A9,SC5D,SENP2,SERPINA3,SGK1,SH3GLB1,SLC12A2,SLC2A2,SLC39A14,SLC5A2,SLC7A7,SMAD7,SNX10,SOCS2,SOD2,SP110,SPP1,SST,STAT1,STAT3,STAT4,STEAP4,STS,TAC1,TAP1,TBC1D9,TBXAS1,TERT,TIMP1,TLR1,TLR2,TLR3,TLR4,TLR5,TLR7,TLR8,TNC,TNFRSF11B,TNFRSF17,TNFRSF1B,TNFSF10,TOP2A,TP53,TTK,TYK2,UBE2C,VCAM1,VEGFA,VIM,VLDLR,VTCN1,WARS1,XBP1,XPNPEP2,XRCC5 |
| IFNA2 | | cytokine | Activated | 7,235 | 7,313 | 0,008 | 2,43E-31 | 9,77E-29 | ADAM17,ALOX5,ANXA1,ARL5A,B2M,BAG3,BAX,BST2,C1R,C1S,CAV1,CBFB,CCL5,CCL8,CD69,CD86,CDKN1B,CISH,CXCL10,CXCL11,CXCL9,CYB5A,DDX56,DDX58,DDX60,DPP4,EPB41L3,FAS,GBP1,GBP2,GNAS,GOT1,HERC6,HIF1A,HLA-A,HLA-B,HLA-C,HLA-E,IDO1,IFI16,IFI27,IFI35,IFI44,IFI44L,IFI6,IFIH1,IFIT2,IFIT3,IFIT5,IFITM1,IFITM2,IFNAR2,IL10RB,IL2RB,IRF1,IRF7,IRF9,ISG15,ISG20,LAMP3,LAPTM4B,LGALS3BP,LILRB2,LPAR6,LYN,MAL,MET,MMP9,MT1F,MT1G,MT1X,MX1,MX2,NPDC1,OAS1,OAS2,OAS3,PARP12,PHF11,PLAAT4,PLSCR1,PSME1,PTTG1,RB1,RIPOR2,RSAD2,SAMD9,SAMHD1,SAMSN1,SDC1,SERPINB9,SH2D1A,SLC15A3,SOCS2,SP110,STAT1,STAT4,TAP1,TDRD7,THEMIS2,TLR3,TNFRSF10D,TNFSF10,TP53,TRIM21,TRIM22,UBA7,UBD,UBE2L6,VCAM1,VEGFA,VEGFC,XAF1,XBP1 |

| | | | | | | | | | |
|---|---|---|---|---|---|---|---|---|---|
| CD40LG | 0,101 | cytokine | Activated | 2,918 | 2,996 | 0,008 | 8,06E-28 | 2,72E-25 | ADAM17,AHR,APOBEC3G,APRT,ARID4B,ARSB,ASCC3,ATF3,AUH,AXL,B2M,BAX,BCL2A1,BCL6,BIRC2,BIRC3,BNIP3,CA2,CASP1,CAST,CBX3,CCL18,CCL19,CCL20,CCL5,CCND3,CCNG1,CCR2,CCR5,CCR7,CD1C,CD1D,CD27,CD38,CD40,CD44,CD63,CD69,CD83,CD86,CD8A,CDC25B,CDKN1B,CELF2,CLU,CLUH,CXCL1,CXCL10,CXCR4,CYP1B1,CYTIP,DECR1,DUSP1,F3,FAS,FCGR2B,FOS,FOSB,FSCN1,G0S2,GADD45A,GATB,GCH1,GPR183,GZMB,HDDC2,HIF1A,HLA-DQA1,HLA-DQB1,ICAM1,ID2,ID3,IDO1,IFI44,IFI6,IFIT2,IFIT3,IFITM1,IL10RA,IL13RA1,IL15,IL2RB,IL4R,IL7R,IRF1,IRF2,ISG15,ITGAM,ITPA,JUND,KIF2A,LAMP3,LMAN2,LTB,MAP2K3,MARCKS,MARCKSL1,MIF,MSH2,MT1G,MX1,NAMPT,NAP1L1,NET1,NFKB1,NFKBIE,NOS2,NR4A3,PLA2G4C,PLEK,PMVK,PSMB10,PSMB8,PSMB9,PSME1,PSMG1,PTPN12,RASSF2,RB1,RIDA,RUNX3,SAMSN1,SELL,SELP,SEMA4D,SLC29A1,SNX4,SOD2,SPHK2,STAT1,STAT3,STAT4,SWAP70,TAP1,TAP2,TM7SF2,TNFAIP2,TNFAIP6,TNFRSF11B,TNFRSF17,TNFSF10,TP53,TRAF5,TREM2,TUBB,UBD,VCAM1,VEGFA,XPO7,XRCC5,YBX3,ZZZ3 |
| IL1B | 0,211 | cytokine | Activated | 6,036 | 6,093 | 0,004 | 1,67E-27 | 5,28E-25 | A2M,ABCC2,ABCC3,ABCG2,ADAMTS1,ADM,ADRB2,AKR1B1,ALDH7A1,ALPL,ANKH,ANXA1,ANXA9,APOE,ARC,ARL6IP5,ASS1,ATF3,AVP,B2M,BACE1,BAX,BCL2A1,BCL6,BIRC2,BIRC3,C1R,C3,CALB1,CASP3,CASP4,CAT,CCL20,CCL5,CCL8,CCR2,CCR5,CCR7,CD14,CD40,CD44,CD55,CD69,CD74,CD83,CD86,CFB,CISH,COL1A1,CSF2RB,CTSB,CTSF,CTSS,CTSZ,CX3CR1,CXCL1,CXCL10,CXCL11,CXCL12,CXCL6,CXCL9,CXCR4,CYBA,CYBB,CYP27B1,CYP2B6,CYP3A4,CYTIP,DAB2,DCN,DDIT3,DIO1,DNMT1,DPP4,DUSP1,EGOT,EHF,ENG,ENPP2,ESR1,ESRRA,F2RL1,F3,FABP1,FAS,FCGR2B,FGF7,FGFR3,FKBP5,FN1,FOS,FOSB,FOSL1,FOXO1,G0S2,GAD1,GADD45A,GADD45B,GBP1,GBP2,GCH1,GCHFR,GCLC,GHR,GJA1,GM2A,GNAS,GRN,GSTA1,GYS1,H1-10,HDAC1,HEXB,HIF1A,HLA-A,HLA-DRA,HLA-E,HNF1B,HNF4A,HPGDS,HPRT1,HSPA5,ICAM1,ID3,IDO1,IFI16,IFIT3,IFNGR1,IFRD1,IGFBP4,IGFBP6,IL10RA,IL15,IL15RA,IL18,IL1R1,IL32,IL33,IL36A,IL6R,IL7,INSR,IRAK1,IRF1,IRF7,ISG15,ISG20,ITGAM,ITGAV,ITGB3,ITGB8,JUND,KLRB1,KRT19,LCN2,LCP1,LHB,LY96,MAPT,MARCKSL1,MEF2C,MFAP2,MIF,MMP1,MMP2,MMP7,MMP9,MUC1,MX1,MYD88,NAMPT,NFE2L2,NFIL3,NFKB1,NFYB,NGF,NIBAN1,NMI,NOCT,NOS2,NPPA,NR0B2,NR1H3,NR1I3,NR4A1,NR4A3,NRP1,OAS2,OSMR,P4HB,PAPPA,PCNA,PDGFRA,PFKP,PIGR,PLA2G4A,PLD1,PLSCR1,POSTN,PPARA,PPARGC1A,PRKCD,PSMB10,PSMB8,PSMB9,PTGES,PTGS1,PTP4A1,RAC2,RCAN1,RHOB,RNASE1,RORA,RORC,RSAD2,RXRA,S100A10,S100A6,S100A8,S100A9,SCNN1B,SDC1,SERPINA3,SERPINB9,SERPINF2,SGPL1,SIRT3,SLC10A2,SLC1A3,SLC22A4,SLC2A2,SLC2A9,SLC37A4,SLC5A2,SLC9A3,SMAD7,SOCS2,SOD2,SPARC,SPP1,SRGN,SSTR2,STAT1,STAT3,STAT4,TAC1,TAP2,TBP,TFPI2,TGFBR2,THY1,TIMP1,TIMP2,TJP1,TLR2,TLR3,TLR4,TLR7,TLR8,TNFAIP2,TNFAIP6,TNFRSF11B,TNFRSF1B,TNFSF10,TOB1,TREM2,TXNIP,UAP1,UBD,UBE2L6,UGCG,VCAM1,VCA |
| MAPK1 | 0,203 | kinase | Inhibited | -6,175 | -6,173 | 0,000 | 4,57E-27 | 1,38E-24 | ACO1,ADAM17,ADAR,APBB3,APOE,ARC,ARG2,ARSB,ATF3,AURKA,BCL11A,BST2,BTN3A3,C1S,CAPN2,CAV1,CCL5,CCNB1,CCND1,CD69,CD84,CDKN1B,CFB,CGA,CISH,CPPED1,CRYL1,CSTA,CTSB,CXCL11,DAB2,DDX58,DNMT1,DSE,DUSP1,EHD1,ENO3,ERAP1,ESR1,EXOC2,FCMR,FHL2,FN1,FOS,FOSB,GBP1,GBP2,GDPD3,GLS,GRAMD1B,HBB,HLA-B,HLA-C,HLA-DMA,HLA-E,IFI16,IFI27,IFI35,IFI44,IFI6,IFIH1,IFIT2,IFIT3,IFIT5,IFITM1,IGFBP7,IL13,IRF7,IRF9,ISG15,ISG20,ITGAV,ITGB3,JUND,LAMP3,LGALS1,LGALS3BP,LHB,MAOB,MAPK13,MAST3,MMP1,MMP2,MMP9,MNS1,MX2,NFE2L2,NKG7,NMI,NOS2,NOX5,NPPA,NR4A1,NRP1,OAS1,OAS2,OAS3,PARP12,PHF11,PLA2G4A,PLD1,PLSCR1,PPT2,PSMB8,PSMB9,PTPRK,SAMHD1,SECTM1,SHANK2,SOCS2,SP110,SPP1,SPRY2,SPSB1,STAT1,SUN2,SUOX,TAP1,TDRD7,TJP1,TLR3,TM4SF5,TMEM87A,TNFRSF1B,TNFSF10,TP53,TRANK1,TRIM21,TRIM22,TRIM38,TRIM5,TSKU,UBE2C,UBE2I,UBE2L6,VDR,VIM,WIPF1,ZNF184 |

| | | | | | | | | | |
|---|---|---|---|---|---|---|---|---|---|
| PRL | -0,187 | cytokine | Activated | 5,860 | 5,930 | 0,007 | 1,92E-26 | 5,40E-24 | A2M,ABCG2,ACTB,ACTR3,ADAR,ANXA2,ANXA3,ANXA5,ARF3,B2M,BAG1,BCL6,BST2,CAMK2N1,CAPN2,CAV1,CCL20,CCNB1,CCND1,CCND3,CD40,CD69,CISH,CLDN3,CLU,COL1A1,COL1A2,COL3A1,COL5A1,CPD,CTSB,CTSD,CTSS,CTSV,CXCL10,CXCL11,CXCL9,DCN,DDX58,DVL1,EGF,ENO1,ERBB3,ESR1,FN1,FOS,GPNMB,HERC6,HIPK3,HSD11B2,ID1,ID2,ID3,IFI35,IFI44,IFI44L,IFI6,IFIH1,IFIT3,IFIT5,IFITM1,IRF1,IRF7,IRF9,ISG15,KRT19,LAMC1,LAMP3,MAGED1,MGP,MME,MSN,MX2,NFKB1,NOS2,OAS1,OAS2,OAS3,OXT,P4HB,PARM1,PARP12,PC,PCNA,PCOLCE,PDGFRA,PDIA4,PDIA5,PDK2,PDK4,PDLIM4,PGRMC1,PKIG,PLSCR1,PRLR,PSME1,RHOG,RSAD2,SAMD9,SAMHD1,SCP2,SDC1,SERPINA3,SND1,SOCS2,SOD1,SP110,SPARC,SPP1,STAT1,TDRD7,TIMP1,TIMP2,TMEM140,TP53,TUBA1B,VDR,VIM,XAF1 |
| IL2 | | cytokine | Activated | 3,866 | 3,937 | 0,006 | 5,12E-26 | 1,35E-23 | ABCC1,ADCY9,AHR,ANXA1,APOBEC3G,ARHGAP25,ARHGAP32,ARHGEF6,ARNT2,AVP,BAG1,BAX,BCCIP,BCL2A1,BCL6,BHLHE40,BIRC3,C3,CAMK2N1,CAPG,CASP1,CASP3,CASP4,CCL5,CCND1,CCND2,CCND3,CCNG1,CCR2,CCR5,CCR7,CD2,CD247,CD38,CD3D,CD3G,CD40,CD44,CD47,CD58,CD59,CD69,CD74,CD83,CD86,CD8A,CD9,CDKN1B,CDKN1C,CDKN2C,CHN2,CISH,CPD,CRMP1,CSF1R,CSF2RB,CTCF,CTPS1,CX3CR1,CXCL10,CXCL12,CXCR4,CYP27A1,CYP2B6,CYTIP,DAPK2,DARS2,DENND2D,DHODH,DLST,DPP4,DSG2,ELF1,ELMO3,ENPP2,EVL,FAM13B,FAS,FCER1G,FCGR3A/FCGR3B,FCMR,FGF13,FGF7,FGL2,FOS,FOXO1,FYN,GADD45B,GADD45G,GARS1,GNL3,GRN,GZMA,GZMB,GZMK,HCK,HDAC1,HSP90B1,HSPA5,ICAM1,IDI1,IDO1,IFNGR1,IFNGR2,IGFBP4,IL10RA,IL13,IL18,IL1R1,IL2RB,IL2RG,IL4R,IL7R,IRF1,ISG20,ITGA2,ITGB8,ITK,JAK1,JUND,KLRB1,KLRD1,LAPTM5,LCK,LCP2,LTB,LTBP3,MAF,MLLT3,MMD,MME,MMP2,MMP25,MMP9,MX1,MYB,NAB2,NDRG1,NET1,NETO2,NFKB1,NME1,NMI,NOS2,NRP1,PCNA,PDCD4,PDIA3,PDLIM4,PEA15,PECAM1,PHB,PIK3R3,PLAC8,PNP,POLE2,PRF1,PRKD2,PRKX,PTPN6,PTPRC,RFTN1,RHOB,RNF128,RORC,RRAS2,RUFY1,RXRA,S100A4,S100A8,SCP2,SELL,SELP,SERPINB9,SIRPG,SOCS2,SPP1,STAM,STAT3,STAT4,STC2,STK17B,SWAP70,TFDP2,TGFBR3,TIMP1,TLR2,TNFAIP8,TNFRSF10D,TNFRSF11B,TNFRSF17,TNFRSF1B,TNFRSF21,TNFSF10,TP53,TPI1,TRAF5,TRIB3,VAV1,VEGFA,VIM,WD[illegible] |
| PPARA | -0,276 | ligand-dependent nuclear receptor | Inhibited | -3,242 | -3,214 | 0,003 | 2,70E-25 | 6,91E-23 | ACAA1,ACAA2,ACACA,ACADL,ACADM,ACADS,ACADVL,ACAT1,ACOT8,ACOX1,ACSL1,ACSL5,ADAM10,AGXT,AHCY,ALDH2,ALDH3A2,ALDH9A1,ALDOB,ANXA2,APOA1,APOA4,APOC3,APOE,APOM,AQP3,ASL,ASNS,ASS1,AURKA,BAX,C1QA,C1QB,C1R,C1S,C2,C3,CAT,CCNA2,CCNB1,CCND1,CD36,CD63,CDK1,CDKN2C,CFH,CIDEC,CPT2,CTH,CXCL12,CYBA,CYBB,CYP27A1,CYP2B6,CYP2C8,CYP4A11,DECR1,DECR2,DHCR7,DIO1,DYNLL1,ECH1,ECI1,ECI2,EHHADH,ELOVL6,ENDOG,FABP1,FABP2,FBXO21,FOS,FTCD,G0S2,G6PC,GLS,GLUD1,GLUL,GNMT,GOT2,GPD1,GPT,GSTK1,GSTP1,GSTZ1,HADH,HADHA,HIF1A,HLA-DQA1,HLA-E,HMGCS2,HNF4A,HPD,HSD17B10,HSD17B11,IDI1,IGFBP6,KHK,KIF20A,KYAT1,MAD2L1,MAT2A,ME2,MLYCD,MMD,MMP9,MSMO1,MTNR1A,MTTP,NCF2,NFE2L2,NFKB1,NOS2,NPPA,NR1H3,NSDHL,OGG1,OXT,PAH,PBLD,PC,PCK1,PCNA,PCTP,PDK4,PKLR,PLA2G7,PLIN2,PLTP,PMVK,PNPLA2,POR,PPARA,PPARGC1A,PPM1D,PRC1,QPCT,RETSAT,RNF186,S100A8,S100A9,SARDH,SC5D,SCD,SCP2,SELENBP1,SLC10A2,SLC25A20,SLC27A2,SMC4,SOCS2,TERT,TJP1,TLR2,TLR5,TOP2A,TSPO,TXN,UCP2,UQCRC1,VCAM1,VEGFA |
| NFKBIA | -0,188 | transcription regulator | | 0,954 | 0,985 | 0,003 | 5,15E-25 | 1,21E-22 | A2M,ACKR3,ADGRE5,AEBP1,AK2,APOA1,ATP6V1B1,AXL,AZGP1,BAX,BCCIP,BCL11A,BCL2A1,BCL2L2,BID,BIRC2,BIRC3,BNIP3,C3,CAPZA2,CASP4,CASP8,CCL20,CCL5,CCNA2,CCNB1,CCND1,CCND2,CD40,CD69,CD86,CDC25B,CDH3,CLU,COL1A2,COL3A1,COL5A2,CSK,CTSB,CTSF,CTSZ,CXCL1,CXCL10,CXCL11,CXCL12,CXCL6,CXCR4,CYBA,CYP1B1,DCN,DDIT3,EIF4G2,ENPP2,ERAP1,ESM1,F3,FAS,FCGRT,FHL2,FN1,FOS,FSCN1,GADD45A,GADD45B,GADD45G,GBP2,GCH1,GM2A,GPX3,GRK5,GRN,GZMB,GZMK,HIF1A,HLA-A,HOXA10,HOXB2,HOXB5,HPSE,HSPA5,HSPA8,ICAM1,IFI16,IFI6,IGFBP6,IL13,IL15,IL15RA,IL33,IL7R,IRAK1,IRF1,ISG15,ITGA2,ITGA3,ITGAV,LAMA5,LCN2,LIMA1,LITAF,MGP,MMP1,MMP9,MPDU1,MR1,MSH2,NAGLU,NFKB1,NFKBIE,NGF,NOCT,NOD2,NOS2,NPPA,NR1I3,NR4A1,PCNA,PDLIM1,PEA15,PTEN,PTGES,PTP4A2,RAC1,RBP1,RCAN1,ROCK1,RPL8,RRM1,S100A8,S100A9,SDC1,SERPINE2,SFRP1,SGK1,SLC12A2,SOAT1,SOD1,SOD2,TAC1,TCF12,TIMP1,TIMP2,TLR2,TLR4,TNC,TNFAIP2,TNFAIP8,TNFRSF11B,TNFRSF1B,TNFSF10,TOLLIP,TP53,VCAM1,VEGFA,VEGFC,XRCC5,ZNF638 |

| | | | | | | | | |
|---|---|---|---|---|---|---|---|---|
| STAT3 | 0,302 | transcription regulator | | 1,765 | 1,818 | 0,005 | 1,24E-24 | 2,75E-22 | A2M,ADM,AHR,ALDH1A3,APOA4,ARG2,BCL6,BNIP3L,BST2,CASP1,CASP3,CAT,CCL20,CCL5,CCNB1,CCND1,CCND2,CCR5,CD40,CD74,CD83,CD86,CD9,CDH5,CDK1,CDKN1B,CFB,CLIC5,CLN6,COL1A1,COL1A2,COL3A1,COL5A1,CTF1,CTSB,CXCL10,CXCL6,CXCL9,CXCR4,DDIT3,DNMT1,DPP4,ECT2,EED,ENPP2,ESR1,F3,FAS,FCER1G,FERMT2,FGL2,FLRT3,FN1,FOS,FSCN1,FUT4,G6PC,GADD45A,GADD45G,GBP2,GJA1,GLIPR1,GPR65,GZMB,HBB,HERC6,HIF1A,HLA-A,HLA-DMA,HLA-DQA1,HNF4A,HOXA10,ICAM1,ID1,ID2,IFI16,IFI27,IFI30,IFI35,IFI44,IFI6,IFIH1,IFIT2,IFIT3,IFIT5,IFITM1,IFITM2,IHH,IL13,IL1R1,IL2RB,IL4R,IL6R,IL7,IRF1,IRF7,IRF9,ISG15,ISG20,ITGAM,ITGAV,ITGB2,ITGB6,LEPR,LILRB2,LTF,LYZ,MAF,MAFB,MMP2,MMP7,MMP9,MRAS,MUC1,MX1,MX2,MXI1,MYB,MYD88,NAMPT,NDRG1,NFKB1,NOS2,NOX5,NPPA,NRP1,OAS1,OAS2,OAS3,PCK1,PDIA4,PECAM1,PEX1,PFKL,PHB,PLA2G4A,PLSCR1,PPARGC1A,PRF1,PROCR,PROM1,PSIP1,PSMB5,PSMB8,PSMB9,PTPN6,PTTG1,RAB27A,RALGDS,REG1A,RORA,RORC,RRAS2,RSAD2,S100A9,S1PR1,SERPINA3,SERPINB1,SERPINB9,SERPINE2,SGK1,SLC1A3,SLC9A3,SMAD7,SOD2,SP110,STAT1,STAT3,STC2,TAP1,TCF4,TERT,TFF3,TFPI2,TIMP1,TLR3,TNFRSF1B,TNFSF10,TNS1,TP53,TRIM22,TRIM5,TRPM3,UBD,VCAN,VEGF[illegible] |
| KRAS | 0,289 | enzyme | Inhibited | -2,602 | -2,602 | 0,000 | 2,44E-24 | 5,30E-22 | ABCC5,ACTR3,AGRN,AHNAK,AHR,AOX1,ATF3,AXL,B2M,BAG1,BIRC3,BNIP3,BZW2,CASP1,CASP3,CAV1,CAVIN1,CCNA2,CCNB1,CCND1,CCND3,CCT5,CD44,CD74,CDH3,CDKN1B,CDKN2C,CLEC10A,CLIC5,CLU,COL1A1,COL3A1,COL4A1,CSE1L,CXADR,CXCL10,CXCL14,CXCL6,DBN1,DCLK1,DDAH1,DDIT3,DNMT1,DPYSL2,DUSP1,EFEMP1,EIF5,ENO1,EPM2A,EZH2,F2RL1,F3,FAH,FAS,FBN1,FGF7,FKBP11,FLII,FN1,FOS,FOSB,FOSL1,FRAT1,FSTL1,GADD45A,GCLC,GCLM,GCNT3,GFPT1,GJA1,GLUL,GPC3,GRN,GSTA3,GZMK,HIF1A,HLA-DRB1,HOPX,HSPA5,ID1,IDO1,IFI6,IFITM1,IFITM2,IL33,IQGAP2,IRF1,IRF2,IRF9,ISG15,ITGB3,ITM2A,KAT5,KIF22,KPNB1,KRAS,LAMA5,LAMB1,LAMP3,LCN2,LMNB1,LYZ,MAPK12,MAPK8,MCAM,MET,MICB,MMD,MMP2,MMP7,MMP9,MSN,MTHFD1,MTRR,MX1,MX2,NCL,NETO2,NFE2L2,NFKB1,NFYB,NOS2,NR4A1,NSF,NXN,OAS1,PALLD,PCOLCE,PDGFRA,PDIA3,PECAM1,PFKL,PGAM2,PKM,PLD1,PLEKHO2,PMP22,PPFIBP1,PRC1,PRDX4,PRKCD,PRKCI,PTEN,PTGS1,PTTG1,QTRT1,RAB27A,RBBP7,RBM7,RHOA,RHOB,ROCK1,S100A4,S100A6,SAMHD1,SCD,SERPINA3,SERPINE2,SLC25A32,SLC29A1,SLC34A2,SLC35B1,SLC3A2,SLC43A3,SOAT1,SPARC,SPP1,SPRY2,ST5,STAT1,STAT3,TAP1,TESC,TGFBR2,THY1,TIMM10,TIMP2,TNC,TNFAIP2,TNFRSF11B,TOP2A,TP53,UBE2I,UQCRC2,VCAN,VEGFA,V[illegible] |
| D-glucose | | chemical - endogenous mammalian | | 1,774 | 1,813 | 0,003 | 3,05E-24 | 6,44E-22 | ABCG1,ACAA1,ACAA2,ACACA,ACADL,ACADM,ACE,ACO2,ACOX1,ACSL1,ACTB,ACTN1,ADA,ADAM17,ADIPOR1,AKR1B1,ALDOB,ANXA1,ANXA3,APOC3,AQP1,ARF3,ARF4,ASNS,ASS1,ATF3,ATP2B4,ATP5MC2,BAX,C3,CALB1,CALR,CAP1,CAPZA2,CASP1,CASP3,CAT,CAV1,CBR3,CCL20,CCNB1,CCND1,CCND2,CCND3,CD36,CD55,CD59,CDK1,CDKN1B,CLU,COL1A1,COL3A1,COL4A1,COX6A2,CPD,CPT2,CSF1R,CTSB,CTSV,CXCR4,CYBA,CYBB,DCN,DDIT3,DECR1,DUSP1,EFHD1,ELOVL6,ENO1,FABP2,FAS,FBP1,FN1,FOS,FTO,G6PC,GABARAPL1,GCDH,GCH1,GFPT1,GJA1,GLUL,GOT1,GPD1,GRK2,GRN,HIF1A,HK1,HNF4A,HPRT1,HSPA5,ICAM1,ID1,IDH1,INSR,ITPR3,LAMB1,LAMC1,LCP1,LOC102724788/PRODH,LY96,MAFB,MCAM,MDH2,MIF,MLXIPL,MMP1,MMP2,MMP9,MSRB1,MTOR,NAMPT,NEU1,NFE2L2,NOS2,NOX4,NPHS1,NPY1R,NR4A1,NR4A3,NUAK1,OAT,OGDH,PARP1,PBK,PC,PCCB,PCK1,PCK2,PCNA,PCSK1N,PDCD4,PDGFRA,PDIA3,PDIA4,PDIA6,PFKL,PFKP,PGM1,PHB,PINK1,PKLR,PKM,PLIN3,PNPLA3,PON2,PPARA,PPARGC1A,PPIB,PRDX3,PRKCZ,PTPN6,RRM2,RTN3,S100A4,SCD,SDHB,SEPHS2,SGK1,SHC1,SIRT2,SIRT3,SIRT4,SLC16A4,SLC1A3,SLC25A1,SLC25A10,SLC25A11,SLC25A5,SLC29A1,SLC2A2,SLC33A1,SLC35B1,SLC37A4,SLC39A14,SLC39A6,SLC3A2,SOD2,SPHK2,SPP1,SSR1,SST,STUB1,STXBP1,SUCLA2,TACSTD2,TAGLN2,TIMP1,TKT,TLR2,TLR4,TP53,TPI1,TRPV4,TRPV5,TXNIP,UCP2,UQCRC2,VAMP2,VCAM1,VEGFA,VIL1,W[illegible] |

| | | | | | | | | | |
|---|---|---|---|---|---|---|---|---|---|
| FAS | 0,651 | transmembrane receptor | | -0,326 | -0,330 | -0,001 | 3,38E-24 | 6,97E-22 | AIFM1,ANXA2,ARAP2,BAX,BCL11A,BCL2A1,BID,BIRC2,BIRC3,BTN3A3,CAMK2G,CASP3,CASP8,CAST,CCL20,CCL5,CD1D,CD200,CD44,CD48,CD59,CD83,CD86,CD93,CDKN1B,CLTB,COL15A1,COL18A1,COL1A1,COL1A2,COL3A1,COL4A1,COL4A2,COL5A1,COL6A3,CXCL1,CXCL10,CXCL11,CXCL6,CXCL9,F2RL1,F3,FAS,FBN1,FCGR2B,FGL2,FOS,FOSB,FOXO1,FPR1,FSCN1,GADD45G,GBP1,GBP2,GLIPR1,GNAI2,GNG7,GPNMB,GRK5,GTPBP6,HLA-DQB1,HLA-F,ICAM1,IL10RA,IL13RA1,IL15RA,IL18,IMPA2,INPP5D,IQGAP2,IRF1,ITGA7,ITGAV,ITGB8,JUND,LAMP3,LAT,LILRB2,LST1,LTB,LUM,LY75,MAP4K4,MEF2C,MET,MGP,MIF,MME,MMP1,MMP9,MPZL1,NFKBIE,NOS2,NR4A1,OPTN,PAK2,PECAM1,PIK3C2B,PINK1,PLD1,PLIN2,PPP1R15A,PRKCI,PTP4A1,PTP4A2,PTPRN2,PVALB,RDX,RSAD2,SDC1,SELP,SERPINB9,SHC1,SLPI,SOD2,SPP1,STK17B,STK38,SYK,TAGLN2,TLN2,TLR2,TLR8,TNC,TNFAIP6,TNFRSF11B,TNFRSF1B,TNFSF10,TP53BP1,TPM1,TWF2,TYK2,UGCG,VCAM1,VEGFA,VRK2,WNK1,XPC,ZFP36 |
| HRAS | 0,086 | enzyme | | 1,544 | 1,571 | 0,003 | 9,57E-24 | 1,88E-21 | A2M,ACTB,ACTG1,ADGRE5,ADM,ADRA2A,AFP,ALDH1A3,ANXA2,ANXA3,ASAP1,ASNS,ATP1A3,AURKA,B2M,BAX,BIRC3,BMP7,BNIP3,CALCRL,CALU,CAPG,CAPN2,CAV1,CCL5,CCNA2,CCNB1,CCND1,CCND2,CCND3,CCNG1,CD24,CD44,CD69,CD81,CD96,CDH3,CDK1,CDKN1B,CDX2,CFB,CFH,CGA,CHST12,CISH,CLU,CNN2,COL18A1,COL1A1,COL1A2,COL3A1,COL4A1,COL5A2,CSTA,CTSB,CTSS,CTSV,CTSZ,CXCL1,CXCL10,CXCL12,DDIT3,DNMT1,DOK1,DPF1,DPP4,DTYMK,DUSP1,DUSP9,EFEMP1,EIF2D,EIF4G3,ELK3,EPHX1,FAS,FERMT2,FHL1,FKBP6,FLNA,FN1,FOS,FOSB,FOSL1,FOXO1,FSTL1,FZD2,GADD45G,GJA1,GNB1,GPX3,GRN,HDHD5,HIF1A,HLA-B,HOXB6,HSPA8,HTRA1,ICAM1,ID2,IFI30,IFITM2,IGFBP4,IGKC,IL13RA1,IL7R,IQGAP2,ITGAM,ITGAV,KRAS,KRT19,LMNB1,LOXL1,LSP1,LXN,LYZ,MAOB,MAPK12,MCAM,MET,MMP1,MMP2,MMP9,MSMO1,MSRB1,MUC13,MYL12A,MYL3,NID2,NOS2,NPPA,NRP1,P4HB,PAFAH1B3,PCK1,PCNA,PDCD4,PDGFRA,PDIA5,PDLIM1,PFN1,PLA2G4A,PLOD1,PLOD2,PLS3,PLTP,POLR2G,POSTN,PPIC,PRDX2,PRKCA,PRKCD,PRNP,PRR5,PSMB9,PTEN,PTGS1,PTPRE,RASGRP1,RBBP7,RBPMS,RECK,RHOA,RHOB,RNF19B,RPL23,RPS3,RRM1,RRM2,RSU1,S100A4,SASH1,SEM1,SERPINA3,SERPINB9,SERPINE2,SLC2A2,SMC3,SOAT1,SOCS2,SOD1,SOX4,SP3,SPARC,SPON1,SPP1,SPRY2,STAU1,STK10,TACC3,TAP1,TAP2,TBP,TERT,TG,TLN2,TMSB10/TMSB4X,TOP2A,TP53,TRPV1,TSC22D1,TYRO3,VCAN,VEGFA,VIM |
| butyric acid | | chemical - endogenous mammalian | | -0,582 | -0,572 | 0,001 | 1,02E-23 | 1,96E-21 | A2M,ABCC2,ADGRE5,ADGRG1,ADH6,ALAD,ALB,ALOX5,ALPG,ANXA1,ANXA2,ANXA5,AQP1,ATP2B4,BAG1,BAX,BCL2A1,C7,CALB1,CALM1 (includes others),CAP1,CASP3,CCL20,CCNA2,CCNB1,CCNB2,CCND1,CCND2,CCNG1,CCNH,CCR5,CD44,CD81,CD83,CD86,CDH11,CDK1,CDK5,CDKN1B,CDKN1C,CGA,CLCN3,CLDN1,CLU,COL4A1,COL4A2,COL5A1,COL5A2,COL6A3,CPA3,CUL1,CXCR4,CYP2B6,CYP4A11,DDR1,DEK,DTYMK,DUSP1,EIF4G2,ELK3,EMP3,ERCC2,ESR1,FOS,FSTL1,FYN,G0S2,G6PC,GADD45A,GAK,GBP2,GCLC,GLI3,GNAI2,GRN,GSTA1,GSTP1,HBB,HK1,HLA-DRA,HLA-DRB1,HMGB2,ICAM1,IFI16,IFI30,IFIT3,IFITM1,IFITM2,IFNAR2,IFNGR2,IL1R1,IL32,INPP5D,INPP5J,IQGAP1,IRF1,IRF2,ITGAM,ITPR3,JCHAIN,KRAS,MAPK13,MMP7,MMP9,MUC1,MYB,NAMPT,NCL,NDUFAF4,NEU1,NFATC4,NFKB1,NKX2-5,NME1,NOS2,NR4A1,NRP1,NUCB1,OAS2,PCK1,PCNA,PDIA3,PDK2,PDK3,PFKL,PGRMC2,PHB,PLSCR1,PRKCD,PRLR,PTEN,PTGS1,PTPRM,PYGL,RB1,RHOA,RXRA,SGK1,SLC26A2,SMPD1,SPP1,STAT3,SYK,TFF3,TGFBI,TGFBR2,THBS4,TIMP1,TNFAIP2,TOP2A,TP53,TP53BP2,TRPV1,UTRN,VEGFA,WT1 |
| PML | | transcription regulator | Activated | 4,475 | 4,480 | 0,001 | 1,44E-23 | 2,71E-21 | ACACA,ACACB,ACADL,ACADM,ACADS,ACOX1,ACTG1,AKR1A1,AKR7A2,ANXA4,APOA1,APOE,BAX,BST2,CAT,CCND1,CEBPE,CPE,CXCL1,CXCL11,CXCR4,DDX60,DNAJB9,FABP1,FAS,FTL,GADD45A,GCLM,HBB,HERC6,HLA-DMB,HLA-DRA,ID2,IFI27,IFI35,IFI44,IFI44L,IFIH1,IFIT3,IFITM1,IRF7,ISG15,ISG20,LYZ,MAF,MAFF,MCM6,MX1,NDRG1,NFE2L2,OAS1,OAS2,OAS3,PCBP2,PDK4,PIK3CB,PLSCR1,PPARGC1A,PRKCA,PRKCH,PSMB8,PSMB9,RRAS2,RSU1,RXRA,SCD,SLC25A20,STAT1,STMN1,TAP1,TAP2,TBX2,TOPBP1,TP53,TXN,TXN2,UBE2M,UCP2,VIM |

| | | | | | | | | | |
|---|---|---|---|---|---|---|---|---|---|
| IL5 | | cytokine | Activated | 2,644 | 2,743 | 0,009 | 2,52E-23 | 4,60E-21 | A2M,ACAA2,ACP5,ANXA2,AOC1,ASNS,ASS1,BCL6,BIRC2,BNIP3,BNIP3L,CA4,CASP4,CCND2,CCND3,CCNH,CCR2,CCR5,CCR7,CD24,CD40,CD55,CD63,CD69,CD9,CDC123,CFHR2,CHST12,CISH,CKAP4,CLEC2B,CRELD2,CRIP1,CSF2RB,CX3CR1,CXCR4,EAF2,EBP,EGOT,ELL2,ENDOG,ENO1,FAS,FCGR2B,FGL2,GADD45A,GADD45G,GALK1,GBP2,GCAT,GCLM,GK,HIF1A,HSP90B1,HSPA5,ICAM1,IDI1,IFI30,IGHM,IL13,IL13RA1,IL2RG,IL4R,IL6R,IRF7,ITGAM,ITGAV,ITGB6,JCHAIN,KCNAB2,KLHDC2,LCP2,LMNB1,LTB,LUM,MANF,MAT2A,MMD,MMP9,NDRG1,NFIL3,NOS2,P4HA1,PDIA6,PDLIM1,PFKP,PGM1,PKM,PMP22,PRDX4,PSAT1,PTPRC,QSOX1,RAB21,RBPMS,RHOH,RIPOR2,RRAS2,S100A4,S100A6,SDF2L1,SGK1,SLC16A3,SLC1A5,SOCS2,SRI,ST7,STK39,THY1,TLR1,TPI1,TRA2A,UPB1,VIM,XBP1 |
| Immunoglobulin | | complex | | -0,155 | -0,154 | 0,000 | 2,56E-23 | 4,60E-21 | ACACA,ACO1,ACO2,ACSL1,ADIPOR1,ADM,BCL2A1,BCL6,BIRC3,BLVRB,C3AR1,CALR,CAPG,CASP4,CCL18,CCL5,CCL8,CCND2,CCND3,CCR2,CCR7,CD14,CD40,CD44,CD69,CD74,CD83,CD86,CDKN1B,CDKN1C,CHN2,CISH,COX11,CSF1R,CSF2RB,CTH,CTSZ,CXCL1,CXCL10,CXCL11,CXCL12,CXCL6,CXCL9,CXCR4,CYP1B1,CYP27B1,DLAT,DLD,DLST,DPYSL2,DUSP1,ENO1,ENO3,EXT1,EZH2,F3,FAS,FBP1,FCER1G,FCGR3A/FCGR3B,FOS,FOSB,FPR1,FTO,G6PC,GYS1,GZMB,HCK,HLA-DQB1,HSD11B2,ICAM1,IDO1,IFI30,IFNGR2,IGFBP4,IL10RA,IL10RB,IL13,IL18,IL2RB,IL32,IL33,IL4R,IL7R,IMPDH2,IRAK1,IRF1,IRF7,ITGB8,JCHAIN,KCTD12,KLRD1,LCK,LCN2,LILRB2,LST1,LTBP3,MAF,MAFF,MAPT,MDH2,MEF2C,MICAL2,MMP1,MMP2,MMP9,MTHFD1,MTHFD2,MTOR,MZB1,NAMPT,NCF2,NDUFA10,NDUFA3,NDUFA7,NDUFB7,NDUFS1,NDUFS7,NDUFS8,NFIL3,NOS2,NR4A1,NRP1,OGDH,PC,PCK1,PCK2,PDIA6,PDK3,PDK4,PDLIM4,PFKL,PGAM2,PGM1,PKLR,PLSCR1,PRF1,RAB31,RB1,RBKS,RNF128,RORC,RPIA,RSAD2,S100A8,S100A9,SCD,SCP2,SDC1,SDHD,SELL,SOD1,SOD2,SPP1,STAB1,STAT3,SUCLA2,SUCLG1,SUCLG2,TGFBR3,TIMP1,TLR2,TLR7,TLR8,TNFRSF1B,TXN,UBD,VCAM1,VCAN,VEGFA,XBP1,YBX3 |
| IL13 | -0,264 | cytokine | | 1,858 | 1,904 | 0,004 | 5,84E-23 | 1,01E-20 | ACADVL,ACKR3,ACOX1,ACSL1,ADA,ADAMDEC1,ALOX5AP,ARG2,ARNTL2,ATF3,AVPI1,BAX,BCL2A1,BID,BIRC2,BLVRA,BZW2,C3,C3AR1,CA2,CAPN2,CASP1,CASP3,CASP6,CASP8,CCL18,CCL20,CCL5,CCR5,CD14,CD163,CD1C,CD247,CD36,CD37,CD40,CD44,CD48,CD52,CD86,CHN2,CIDEB,CISH,CLEC10A,CLEC4A,CLEC7A,COL18A1,COL1A1,COL1A2,COL3A1,CTSB,CTSK,CTSS,CTSV,CXCL1,CXCL10,CXCL6,CXCR4,CYBB,DNASE2,DYNLT3,ENPP2,EVL,F3,FAS,FCGR2B,G0S2,GNAI2,GPNMB,GPX3,HK1,HPSE,HRH1,HTRA1,ICAM1,IDO1,IFNGR1,IFNGR2,IL13,IL13RA1,IL18,IL1R1,JAK1,KITLG,LILRB2,LIPA,MAF,MAOA,MCUR1,MMP1,MMP2,MMP9,MRC1,MS4A4A,MSMO1,MT1X,NOS2,PAPSS1,PDLIM2,PFKP,PLA2G4A,PLIN2,POSTN,PPARGC1A,PRSS8,PTGES,QSOX1,RFTN1,RHOA,RIN2,RORC,RYK,S100A8,SAMSN1,SELP,SERPINF1,SLA,SLC26A6,SLC43A3,SLC7A7,SLPI,SPINT2,SPP1,ST14,ST3GAL6,ST8SIA4,STXBP1,SWAP70,SYNGR2,TFEC,TFF3,TIMP1,TIMP2,TLR1,TLR4,TMC6,TNFRSF11B,TNFRSF1B,TNS1,TRPV1,TYRO3,VCAM1 |
| IL15 | 0,170 | cytokine | Activated | 3,471 | 3,526 | 0,006 | 2,16E-22 | 3,65E-20 | ACADS,AHR,ANXA1,AQP3,ARHGDIB,CALM1 (includes others),CAMK2G,CASP3,CCL18,CCL19,CCL5,CCND1,CCND2,CCND3,CCR2,CD2,CD38,CD40,CD44,CD53,CD55,CD59,CD69,CD74,CD83,CD84,CD86,CD8A,CDK2AP1,CDKN1B,CISH,CORO1A,CX3CR1,CXCL10,CXCL12,CXCR4,DARS2,DPP4,EBP,ELK3,ENO1,ENO3,EXOSC8,FAS,FBP1,FCGR3A/FCGR3B,FYB1,FYN,GARS1,GLDC,GNAS,GNLY,GZMB,GZMK,HK1,HNRNPF,HSPA8,ICAM2,IFI35,IFITM1,IL13,IL15,IL15RA,IL1R1,IL2RB,IL2RG,IL32,IL7R,IRF1,KLRB1,KLRD1,LCK,LSP1,LYN,MACROH2A1,MAF,MCM5,MICB,MIF,MLLT3,MT1G,MX1,MYD88,NFKB1,NME1,NOS2,NUDT1,PAFAH1B3,PCBP2,PCNA,PDCD4,PDCD5,PDIA3,PDIA4,PFKFB2,PFKFB3,PFKL,PFKP,PFN1,PGAM2,PGM1,PIK3R3,PKM,PLA2G4A,PLD1,PLEK,PNP,PPARGC1A,PRDX4,PRF1,PSMB10,PSMB8,PSMB9,PTP4A1,PTP4A2,PTPRC,PYGL,RAB8A,RAC2,RBX1,RPIA,S100A11,S1PR1,SELL,SET,SLC16A3,SLC25A5,SMAD2,SMAD7,SOCS2,SRGN,SRSF7,TFDP2,TGFBR2,TIMP1,TJP1,TKT,TLR2,TNFRSF17,TNFSF10,TNNI1,TOP2A,TPI1,TRIM5,TSPAN7,TYROBP,VCAM1,VDR,VEGFA,XBP1,ZFP36 |

| | | | | | | | | | |
|---|---|---|---|---|---|---|---|---|---|
| FSH | | complex | | -0,352 | -0,329 | 0,003 | 3,92E-22 | 6,50E-20 | ACP5,ACTB,ACTN1,ACTR2,ADAM17,ADAMTS1,ADCY7,ADCY9,ADGRG1,ADM,AHR,AKAP12,ALDH3A2,ALPL,AQP3,ARL4C,ARL6IP5,ATP9A,AXL,BAG1,BCL11A,BCL2L2,BNIP3L,CAMK2G,CAP2,CASP4,CCND1,CCND2,CD55,CDK14,CDKN1C,CHUK,COL15A1,COL18A1,CPE,CRYM,CSNK1A1,CTSV,CXCR4,CYFIP2,CYP17A1,DAB2,DHCR7,DHRS3,DHX15,DNAJB9,DUSP1,DUSP3,DUSP9,ELL2,ENC1,ESR1,FDX1,FKBP5,FOS,FOSB,FOSL1,FOXO1,GBP1,GCLC,GK,GNAS,GNB1,GNLY,GOT1,GPER1,GRK5,GZMB,HSD3B2,ID1,IGFBP4,IL6R,ING2,INHA,ISG20,ITGA3,KITLG,LGALS1,MAP2K3,MAP4K4,MMP2,MMP9,MSMO1,MT1X,NPY1R,NR4A1,P4HA2,PCK1,PCNA,PGRMC1,PHKA2,PKM,PLCL1,PLIN3,PLK2,PPARA,PPIH,PRKD1,PRKX,PRLR,PRNP,PSIP1,PSMD1,PTEN,PTGS1,PTP4A1,PTPRE,PTPRF,RAB27A,RAB31,RAB5C,RASSF2,RGS7,RHOB,SGK1,SHBG,SMAD1,SMAD2,SMAD7,SMARCA4,SSTR2,STAT1,STK25,STX4,TFPI2,TGFBR2,TGFBR3,TIMP1,TIMP2,TMBIM6,TNFAIP6,TNFRSF11B,TOB1,TP53,TPM1,TUBA1A,TYRO3,VEGFA,VEGFC,WT1,YBX3,ZNF331 |
| NR3C1 | 0,164 | ligand-dependent nuclear receptor | Inhibited | -2,713 | -2,704 | 0,001 | 4,48E-22 | 7,29E-20 | A2M,ACAT1,ACTB,ACTN1,ADM,ALOX5AP,ANXA1,APOE,APOL3,AQP1,ARID4B,ARPC2,AVP,B2M,BAG1,BAG3,BARD1,BAX,BCKDHA,BCL2L2,BCL6,BHLHE40,BIN1,BIRC2,BIRC3,BNIP3L,C1R,CACNA2D3,CAPN2,CASP4,CASP6,CAT,CAV1,CCL5,CCND1,CD38,CD47,CD69,CD83,CDKN1C,CGA,CIDEC,CKAP2,COMT,COQ8A,CORO1C,CUL1,CXCL10,CXCR4,CYP27A1,CYP2A6 (includes others),CYP2C8,CYP3A4,CYP3A7,DAPK2,DDC,DUSP1,DUSP22,DYNLL1,EBAG9,ENC1,ENDOG,ETNK2,F2RL1,F3,FAM89B,FCER1G,FKBP4,FKBP5,FN1,FNTA,FOS,FOXO1,G6PC,GADD45A,GADD45B,GBP2,GCLM,GLIPR1,GLUL,GNMT,GPR65,GPX3,GSTA1,GSTP1,GZMA,HDAC1,HGD,HMGCS2,HSD11B2,ICAM1,IDH1,IFI6,IFIH1,IFIT2,IFT122,IL13,IL15,IL15RA,IL18,IL33,IL7R,ING2,INSR,IRF1,IRF8,ISG15,ITGAV,ITGB2,ITGB6,JUND,KAT5,KLF9,LHB,LTB,MAL,MALT1,MAOA,MMP1,MMP9,MSRB1,MTCH2,MTHFD1,MYD88,MZB1,NAMPT,NFIL3,NFKB1,NOD2,NOS2,NPPA,NR0B2,NR1I3,NSMAF,OAT,PAK2,PCK1,PDCD4,PEA15,PER1,PIGR,PIK3CB,PIK3R3,PLA2G4A,PLEKHF1,PLIN2,PLK2,PPARA,PPP1R15A,PRKACB,PRKCD,PRKCI,PRLR,PSMA6,PSMG2,PTEN,PTP4A2,RAB11A,RABGGTB,RB1,RHOB,ROCK1,SERINC3,SERPINA6,SERPINB9,SGK1,SGPL1,SGPP1,SH3GLB1,SLC10A2,SLC19A2,SLC2A5,SLC38A1,SMAD1,SOX4,SPP1,SRD5A2,STAT3,STK17B,STK39,STUB1,TAC1,THAP1,TIPARP,TLR1,TLR2,TLR5,TNFAIP2,TNFAIP6,TNFAIP8,TNFRSF11B,TNFRSF1B,TNFRSF21,TP53,TP53BP2,TRAF5,TRIB3,TRIM45,TXN,UGCG,UNC13B,VCAM1,VDR,WD[illegible] |
| APP | 0,254 | other | Activated | 2,168 | 2,207 | 0,003 | 1,03E-21 | 1,65E-19 | ABCG1,ABCG2,ACO2,ACTB,ACTG1,AIFM1,ALB,ANXA5,APOE,ARC,ARF4,ARL6IP5,ATOX1,ATP1A3,ATP5F1D,ATP6V0D1,ATP6V1E1,AXL,BACE1,BASP1,BAX,BCL2L2,C1QBP,C1R,C1S,C3,CALB1,CALML3,CALML4,CALR,CAP1,CAPN1,CASP3,CAST,CAT,CCDC25,CCL20,CCL5,CCND1,CCND2,CCR5,CD151,CD40,CD44,CD59,CD69,CD74,CD86,CDH3,CDH5,CDK1,CDK5,CLDN1,CLIC1,CLTB,CLU,COL18A1,CPNE6,CRABP1,CRYL1,CRYM,CSF1R,CSF2RB,CTSB,CTSD,CTSV,CXCL1,CXCL10,CXCL12,CXCL6,CXCR4,CYBB,CYFIP1,CYFIP2,DBN1,DCN,DDAH1,DDX58,DNM1L,DPYSL2,DSTN,ECH1,ENG,ENO1,ENPP2,ERMP1,ESR1,EVL,EXT1,F2RL1,FAM49A,FCGR2B,FLNA,FN1,FOLR1,FOS,FOSB,FUZ,FYN,GALK1,GBP2,GHR,GNB1,GOT2,GPM6B,GSTP1,HDAC1,HECW1,HERC6,HK1,HLA-E,HSPA5,HSPA8,ICAM1,ICAM2,IDO1,IFI16,IFI35,IFIH1,IFIT2,IL13,IL15,IL18,IL7,INSR,IRF1,IRF7,ISG20,ITGA2,ITGA3,ITGAL,ITGAM,ITGB3,ITPR3,JUND,KIF2A,KL,KPNB1,KYNU,LHFPL2,LPAR6,LRP5,MAF,MAFB,MAFF,MAPT,MDK,MEF2C,MGAT3,MME,MMP1,MMP2,MMP9,MRC1,MTOR,MXI1,MYD88,NAMPT,NARS2,NDUFB11,NDUFS4,NDUFS8,NFKB1,NME1,NOS2,NRP1,PDCD4,PDGFRA,PDK2,PEA15,PEBP1,PFKFB3,PFKP,PITX3,PJA1,PKM,POU2AF1,PRDX2,PRKCA,PRLR,PRMT1,PRNP,PSME1,PTDSS1,PTPRC,PVALB,QDPR,RASSF2,RCAN1,RHOA,RNASEL,ROCK1,RSAD2,RTP4,S100A8,S1PR1,SERPINF1,SET,SH3GL2,SHC1,SLAMF8,SLC13A3,SLIT2,SMAD7,SMARCA4,SOD1,SOD2,SSRP1,ST3GAL5,ST6GAL1,ST8SIA4,STAT3,STMN1,STUB1,STXBP1,SUCLG1,TERT,TGFBR2,THY1,TJP1,TLR2,TLR3,TLR4,TLR5,TLR7,TLR8,TNFAIP2,TNFSF10,TOP2A,TOP2B,TOR3A,TP53,TPD52L1,TPI1,TPM1,TRIB3,TRIM21,TUBA1A,TUBA1B,TUBB,TUBB2A,TUBB4B,TUBB6,TUFM,TXN,UCHL1,VAMP2,VCAM1,VCAN,VEGFA,[illegible] |

| | | | | | | | | | |
|---|---|---|---|---|---|---|---|---|---|
| OGA | 0,160 | enzyme | | 0,717 | 0,706 | -0,001 | 1,30E-21 | 2,04E-19 | ACADM,ACO2,ACTG1,ADGRL2,AIFM1,ALOX5AP,APOBEC3G,APOE,ARAF,ARHGAP29,ASS1,AZGP1,BAX,BCL2L13,BNIP2,C1QA,C1QB,C1RL,C3AR1,C6orf15,CACNA2D3,CALB1,CALCRL,CAPN1,CASP1,CAV1,CCL20,CCND1,CCR5,CD14,CD151,CD24,CD36,CD40,CD44,CD47,CD48,CD53,CD55,CD86,CD93,CDH15,CDK10,CDK5,CDKN2C,CLDN1,COLEC11,CPE,CRABP1,CREB5,CSF1R,CSF2RB,CX3CR1,CXCL6,CYFIP2,CYP2B6,DAB2,DCLK1,DHCR7,DLST,DPEP1,EFNB2,ELF1,ERBB3,FAS,FBP1,FERMT2,FGF13,FLNA,FN1,FSCN1,FZD6,GADD45A,GADD45G,GALK2,GALNT6,GALNT7,GLI3,GLUL,GPER1,GPR21,GTF2IRD1,HPGD,HPGDS,IFIT3,IFNGR2,IGBP1,IGFBP4,IGFBP7,IL18,IL6R,ITGA2,JAK1,KRT81,LAMB1,LPXN,MCAM,MICALL2,MSMO1,NDUFA2,NDUFB11,NDUFS1,NDUFS4,NOTCH2,NPHS1,NSDHL,OGDH,PECR,PHKA2,PLA2G4A,PLAAT3,PLCE1,PLIN2,PLIN3,PRDX4,PRF1,PROM1,PYGL,RAB27A,RB1,RHOB,S100A2,S100A6,SERPINB9,SERPINE2,SIGIRR,SLC2A9,SLC33A1,SOCS2,ST14,STAT1,SUCLG2,TBP,TGFBI,TGFBR2,TGFBR3,TIMP1,TIMP2,TNC,TNFRSF11B,TP53,TP53BP1,TPD52L1,TUBA1A,VIM |
| STAT1 | 1,186 | transcription regulator | Activated | 7,452 | 7,520 | 0,007 | 1,50E-21 | 2,31E-19 | A2M,ALDH1A3,APOBEC3B,APOBEC3G,APOE,AXL,B2M,BAX,BCL6,BST2,C1R,C1S,C3,CAPN1,CASP1,CASP3,CASP4,CASP6,CASP8,CCL19,CCL20,CCL5,CCND1,CCND2,CCND3,CCR7,CD14,CD40,CD86,CDKN1B,CFB,CLIC5,CTSS,CX3CR1,CXCL10,CXCL11,CXCL9,CYP2C8,CYP4A11,DDX60,DPP4,FAS,FCER1G,FCGR2B,FGF7,FGL2,FOS,FOXO1,GBP1,GBP2,GLS,HERC6,HIF1A,HLA-DQA1,HLA-E,HTRA1,ICAM1,IDO1,IFI16,IFI27,IFI30,IFI35,IFI44,IFI44L,IFI6,IFIH1,IFIT2,IFIT3,IFITM1,IFITM2,IL15,IL15RA,IL18,IL1R1,IRF1,IRF2,IRF7,IRF8,IRF9,ISG15,KCTD12,LCN2,LY96,MMP9,MUC1,MX1,NOS2,NOX4,NOX5,NPPA,OAS1,OAS2,OAS3,PCDH17,PECAM1,PLSCR1,PRF1,PSMB10,PSMB8,PSMB9,PSME1,RORC,RSAD2,RTP4,S100A10,SAMHD1,SERPINA3,SERPING1,SLAMF8,SLC2A2,SMAD2,SMAD7,SMAGP,SOAT1,SP110,STAT1,STAT3,TAP1,TAPBPL,TLR3,TLR4,TLR8,TNFSF10,TP53,TP53BP2,TRIM21,TRIM22,UBD,ULK1,WARS1,XAF1 |
| PGR | | ligand-dependent nuclear receptor | | -0,677 | -0,616 | 0,006 | 2,68E-21 | 4,04E-19 | ABCG2,ACOX1,ACP5,ACSL1,ADAMTS1,ALDH1A3,BCL6,BIRC2,BIRC3,CAPN2,CCL8,CCNB1,CCNB2,CCND1,CCND3,CD36,CD44,CD59,CDK1,CDKN1B,CDKN1C,CGA,CKAP4,COMT,CPT2,CTSD,DDC,DPYSL2,DYNLL1,EIF5B,ELL2,ENC1,ESR1,ESRRA,F2RL1,F3,FAM49A,FAM89B,FKBP5,FN1,FOS,FOXO1,G6PC,GLS,GLUL,GOT1,GPER1,GRB10,HIF1A,HPCAL1,HPGD,HSD11B2,ICAM1,ID1,IFI27,IL1R1,KLF9,LHB,MAD2L1,MAP2K3,MAT2A,MUC1,NDRG1,NEDD9,NET1,NOP16,NQO2,NSUN5,OCEL1,OGG1,P4HA2,PCDH1,PCNA,PDLIM1,PFKFB3,PFKP,PGRMC1,PHF21A,PLK2,POSTN,PRKCH,PRLR,PRRX1,PTGES,PTP4A1,RASSF2,S100A6,S100A8,SDC1,SERPINA5,SGK1,SIK3,SLC31A2,SLC9A3R1,SPRY2,SRSF7,ST3GAL1,ST3GAL4,STAT1,STAT3,TGFBI,TIPARP,TM4SF1,TNC,TNFRSF21,TRAF5,UGCG,VCAN,VDR |
| FOXO1 | -0,278 | transcription regulator | | 0,050 | 0,096 | 0,004 | 6,84E-21 | 1,01E-18 | A2M,ACACA,ACSL5,ADIPOR1,AGXT,ALB,ALDOB,ANXA3,APOC3,APOM,ATP6V1D,BACH2,BANK1,BAX,BCL2A1,BCL2L13,BCL2L2,BID,BIRC3,BLNK,C1QA,CA2,CASP6,CASP8,CAT,CAV1,CCL20,CCN3,CCNA2,CCNB1,CCNB2,CCND1,CCR7,CD36,CD40,CDH5,CDK1,CDKN1B,CDKN1C,CDKN2C,COL4A1,COL4A2,CPT2,CTSK,CTSV,CXCL10,DLGAP5,EFHD1,ELOVL6,ENPEP,FABP2,FAS,FBXO21,FN1,FOLR1,FOS,FOSB,FOXO1,FYN,G6PC,GABARAPL1,GADD45A,GADD45B,GPD1,GRHPR,GSTK1,GZMA,HIF1A,HPD,HSPA5,ICAM1,IGFBP7,IL10RB,IL15,IL18,IL1R1,IL7R,INSR,ITGA3,ITGAM,ITGAV,ITGB2,ITGB6,LCP2,LEMD3,LHB,MCM5,ME1,MFN1,MLXIPL,MMP1,MMP9,MRPL34,MRPL46,MRPS12,MRPS2,MRPS22,MRPS7,MTTP,MXI1,MYL3,NAMPT,NDN,NDUFA7,NDUFAF1,NDUFAF4,OVOL1,PBK,PCK1,PCK2,PDK4,PIK3IP1,PKLR,PLSCR1,PNPLA2,PPARGC1A,PPP1R15A,PRC1,PRF1,PRKCD,PSMB6,PSMB8,PTEN,PTGS1,PTPRC,RIPOR2,RPIA,S1PR1,SCD,SELL,SFN,SGK1,SLC25A20,SLC25A24,SLC2A2,SOD2,SPRY2,SPTLC2,TDO2,TKT,TNFSF10,TNNC1,TP53,TRIB3,TSPO,TXNIP,VCAM1 |

| | | | | | | | | | |
|---|---|---|---|---|---|---|---|---|---|
| SMARCA4 | 0,194 | transcription regulator | Activated | 3,689 | 3,783 | 0,008 | 7,97E-21 | 1,16E-18 | A2M,ACE,ACE2,ACSL5,ADGRG1,AFP,AGMAT,AGR2,AHNAK,AHR,AIM2,ALB,ALDH1A3,ALDH2,AP1S2,AP3M2,APOA1,APOBEC3G,APOL3,ARHGDIB,ARL4C,ASNS,ATP2B4,ATP6V1B1,AZGP1,BCL2A1,BIN1,BNIP3L,C1orf54,CASP1,CASP3,CCNA2,CCND1,CCR2,CD44,CD47,CD52,CD63,CD74,CDH11,CDKN1B,CKM,CLIC2,CLPX,COL1A1,COTL1,CTSB,CTSS,CXCR4,CYP3A4,DHRS9,DYNLT3,ECM2,EHF,ELOVL6,EMP3,ENO3,F3,FAS,FBP1,FCHSD2,FHL2,FKBP5,FN1,FOLR1,FOS,GADD45A,GADD45B,GADD45G,GBP1,GCAT,GCHFR,GCLC,GMFG,GNAI3,GPR183,GSTO1,GSTP1,GYS1,H1-10,HBB,HCLS1,HCP5,HEPH,HEXB,HLA-B,HLA-C,HLA-DRA,HLA-E,HLA-F,HLA-J,HNF4A,HNMT,HPGD,HS3ST1,HYI,ICAM1,ICAM2,ID3,IFI16,IFI27,IFI30,IFITM1,IFITM2,IGFBP4,IGFBP7,IL13,IL15RA,IL7R,IRF1,ITGA3,ITGA7,ITGAV,KCNJ2,LGALS1,LPXN,LUM,MAFF,MAOB,MAPK13,MEF2C,MGP,MICB,MMP1,MMP2,MMP7,MUC1,MYB,MYL5,NAP1L3,NCALD,NR2F2,NRP1,PCDH1,PERP,PLEKHO1,PLIN2,PLS1,PSMB9,PTP4A1,PTP4A2,RAB27A,RAB38,RAC2,RBM4B,RBP1,ROCK1,RRM2,S100A2,S100A6,SEL1L3,SEMA3B,SERPINE2,SKAP2,SLC35G2,SLC43A3,SMARCA4,SNAPC2,SOCS2,SPP1,SPRY2,SRPX,ST3GAL1,ST6GALNAC4,ST7,SYK,TACSTD2,TAOK3,TAP1,TAPBPL,TBX2,TCF4,THY1,TIMP2,TLR2,TNFAIP8,TNFSF10,TNNT2,TP53,TPK1,TPM1,TREM2,TRIM15,TSPAN13,TSPAN8,TUBB,UBD,VIM,WDR45,WIPF1,WT1 |
| IFNL1 | | cytokine | Activated | 6,607 | 6,671 | 0,009 | 1,85E-20 | 2,57E-18 | ATF3,BST2,CCR7,CD40,CD83,CXCL10,CXCL11,CXCL9,DDX58,DDX60,GBP1,HERC6,HLA-B,HLA-C,IFI27,IFI35,IFI44,IFI44L,IFI6,IFIH1,IFIT2,IFIT3,IFIT5,IFITM1,IFITM2,IL13,IRF9,ISG15,ISG20,LAMP3,LGALS3BP,MX1,OAS1,OAS2,OAS3,PHF11,PLAAT4,PLSCR1,PSMB9,RSAD2,RTP4,SAMD9,SLC15A3,SP110,STAT1,TDRD7,TLR2,TMEM140,TRIM22,UBE2L6,XAF1 |
| Ifnar | | group | Activated | 6,432 | 6,499 | 0,009 | 2,41E-20 | 3,29E-18 | AIM2,APOBEC3B,AXL,B2M,C3,CASP1,CCL5,CD40,CD74,CD86,CD8A,CXCL10,DDIT3,DDX58,FCER1A,FCER1G,GBP2,HLA-A,HLA-E,HLA-G,IDO1,IFI16,IFI35,IFIH1,IFIT2,IFIT3,IFNGR1,IL15,IRF1,IRF7,IRF8,IRF9,ISG15,ISG20,MMP9,MYD88,OAS1,OAS2,PSMB8,PSMB9,RSAD2,SPP1,STAT1,TAP1,TAP2,TAPBP,TLR2,TLR3,TLR7,TNFSF10,TRIM21,UBE2L6,VCAM1,XAF1 |
| PTEN | 0,445 | phosphatase | | -0,588 | -0,592 | -0,000 | 2,49E-20 | 3,34E-18 | ACAA1,ACAA2,ACACA,ACO2,ACOX1,ACSL1,ADAMDEC1,ADM,AGFG1,AGPAT4,AKAP1,ALOX5AP,ANXA1,ARF4,ATF3,AURKA,BAX,C3,CA2,CA8,CALCRL,CCL5,CCNB1,CCNB2,CCND1,CCND3,CCR7,CD40,CD44,CD86,CDH5,CDK10,CDKN1B,CENPS/CENPS-CORT,CFD,COX8A,CST6,CTSB,CTSV,CTSZ,CX3CR1,CXCL10,CXCL11,CXCR4,CYP27A1,DENR,DHRS11,DLAT,DLD,DNPH1,DUSP1,EBP,ECI2,EED,EFNB2,EGF,EHHADH,ERBB3,ESR1,F3,FAS,FGF1,FGF7,FGFR3,FN3K,FOS,FOXO1,FZD6,G0S2,GALK1,GCAT,GMDS,GPR162,GPX3,GSR,GSS,GTPBP6,GYS1,HADH,HADHA,HCLS1,HDDC2,HEXB,HIF1A,HLA-DMA,HOXA10,IDH3B,IDH3G,IDI1,IFI30,IFNGR2,IFRD1,IGFBP6,IL4R,INPPL1,ISG15,ITPR3,JUND,KIAA0930,KRT19,LEPROT,LHPP,LRRFIP1,MACROH2A1,MAD2L1,MAPK8,MAT2A,MCAM,MET,MME,MMP2,MMP9,MRC1,MTOR,NDRG1,NDUFC1,NDUFS3,NEU1,NFIL3,NFKB1,NKX2-5,NME1,NOS2,NPPA,NR2F2,NUDT1,P2RY14,PARP1,PCBP2,PDCD4,PECAM1,PECR,PFKFB3,PFKL,PGM1,PHGDH,PINK1,PITX3,PKLR,PLOD1,PPARGC1A,PTEN,PTGS1,PTP4A2,PTPRC,PTPRF,PTPRK,PYGL,RAB38,RB1,RMI1,RRM1,SCD,SDC1,SDHB,SDHD,SELL,SEPHS2,SERINC3,SFRP1,SLC25A1,SLC35D1,SLC7A8,SMAD1,SMAD2,SMARCA4,SNRPF,SOAT1,SOX4,SPP1,SPRY2,SQOR,STARD8,STAT3,SUCLA2,SUCLG1,SUCLG2,SYK,TG,TGFBR2,TLE2,TNFRSF10D,TNFRSF11B,TNFSF10,TP53,TP53BP2,TRAM1,UBA7,UCKL1,VCAM1,[illegible] |

| | | | | | | | | | |
|---|---|---|---|---|---|---|---|---|---|
| NFkB (complex) | | complex | Activated | 4,683 | 4,781 | 0,009 | 3,11E-20 | 3,99E-18 | A2M,ABCG1,ACKR3,ADM,ADRA2A,AHR,ALDH1A3,ALDH1B1,ALDH7A1,APOE,AQP1,ATF3,BACE1,BAX,BCL11A,BCL2A1,BCL2L2,BID,BIRC2,BIRC3,BLVRA,C1R,C3,CALB1,CARD8,CASP4,CASP8,CBR3,CCL20,CCL5,CCL8,CCNB1,CCNB2,CCND1,CCND2,CCNH,CCR2,CCR7,CD36,CD40,CD44,CD69,CD74,CD83,CD86,CD99,CDC25B,CDX2,CFB,CLU,COL18A1,COL1A2,CRADD,CST7,CTCF,CXCL1,CXCL10,CXCL11,CXCL12,CXCL6,CXCL9,CXCR4,CYBB,CYP3A4,DDIT3,DIO1,DOK5,EHF,ELL2,ERAP1,F2RL1,F3,FAS,FGF1,FN1,FOS,G0S2,GAD1,GADD45A,GADD45B,GBP2,GCH1,GCLC,GNAS,GRK5,GZMK,HDAC6,HIF1A,HLA-A,HLA-DMB,HLA-F,HNF4A,HSPA9,ICAM1,IDO1,IFNA10,IFNAR2,IGKC,IL10RA,IL13,IL15,IL15RA,IL18,IL32,IL7R,IRF1,IRF7,ISG15,ITGAL,ITGAM,ITGAV,ITGB8,KRT19,KYNU,LCN2,LITAF,LSP1,LTB,MFHAS1,MMP1,MMP2,MMP9,MSH2,NAMPT,NCF2,NFKB1,NFKBIE,NGF,NOS2,NPPA,NUP62,PEA15,PECAM1,PLD1,PLK2,PPARA,PPM1D,PRKCD,PSMB9,PTGES,RFTN1,RNF19B,RRAS,RRAS2,RSAD2,SENP2,SERPINA3,SLC16A3,SLC22A4,SLC2A2,SLC2A5,SLC37A4,SLIT2,SLPI,SMAD7,SOAT1,SOD2,SPP1,STAT4,STK10,STX4,TAC1,TAP1,TAP2,TCF12,TDO2,TERT,TFEC,TFPI2,TIMP1,TIMP2,TLR2,TLR4,TNFAIP2,TNFAIP6,TNFAIP8,TNFRSF11B,TNFSF10,TOLLIP,TP53,TPMT,TRIB3,TRIM38,UBD,VCAM1,VEGFA,VEGFC,VIM,WTAP,XRCC5,ZFP36 |
| AGT | -0,502 | growth factor | Activated | 3,586 | 3,644 | 0,005 | 4,56E-20 | 5,57E-18 | ACE,ACE2,ADAM17,ADD3,ADIPOR1,ADM,ANXA1,ANXA3,ARHGEF9,ASL,ATF3,AVP,BAX,CALR,CASP3,CASP8,CAT,CAV1,CCL5,CCL8,CCND1,CCND2,CCND3,CCR7,CD36,CD44,CD86,CDH5,CDK1,CDKN1B,COL1A1,COL1A2,COL3A1,COL4A1,CREB5,CTF1,CTSS,CXCL10,CYBA,CYBB,CYP17A1,CYP2J2,CYP4A11,DAB2,DUSP1,EBP,EFEMP1,EFNB2,ENG,F3,FBXO21,FDX1,FGF13,FGF7,FN1,FOS,FRAT2,GAS2,GCH1,GFPT1,GJA1,GRK5,GSS,GSTA3,GSTP1,HIF1A,HMGB2,HSD3B2,ICAM1,ID3,IDH1,IDI1,IGFBP4,IL13,IL15,IL18,INSR,ITGA2,ITGA7,ITGAV,ITGB3,KCNJ2,KITLG,KL,KRAS,LAMB1,LEPR,LZTS3,MAD2L1,MAPT,MIF,MMP1,MMP2,MMP9,MSMO1,MYB,NAMPT,NCALD,NCF2,NCL,NDRG1,NGF,NOS2,NOX4,NOX5,NPHS1,NPPA,NPR3,NR4A1,OXT,PALLD,PARP1,PECAM1,PFN1,POSTN,PPARGC1A,PPFIBP1,PPP1R3C,PSMB10,PSMB8,PTEN,PTGS1,RAC1,RCAN1,RHOA,RRAS,SELL,SGK1,SHC1,SLC1A1,SLC5A2,SMAD7,SOD2,SOX4,SPP1,TBX2,TGFBR2,TIMP1,TJP1,TLN2,TNFRSF1B,TP53,TXNIP,UCP2,UGCG,VCAM1,VEGFA,VEGFC,VIM,ZEB2,ZFP36 |
| CD3 | | complex | Activated | 3,004 | 3,089 | 0,007 | 4,56E-20 | 5,57E-18 | ACADS,ACTR3,AHR,ANG,ANXA1,AQP3,ARHGDIB,BCL2A1,BCL6,BIRC3,BST2,C1R,C2,CALM1 (includes others),CAMK2A,CASP3,CASP6,CASP8,CCL18,CCL20,CCL5,CCND1,CCND2,CCND3,CCNH,CCR2,CCR5,CD2,CD247,CD27,CD44,CD53,CD69,CD74,CD83,CD86,CD8A,CDK1,CDK2AP1,CDKN1B,CISH,CLCN1,CORO1A,CXCL10,CXCR4,CYBA,CYC1,DOCK2,DPP4,DUSP22,EBP,EIF4A3,ELK3,EXOSC8,FAS,FCER1G,FKBP3,FOS,FTL,FYB1,FYN,GBP1,GLDC,GNAS,GTF2IRD1,GTPBP4,GUCY1B1,GYS1,GZMA,GZMB,GZMK,HAX1,HIF1A,HLA-DQB1,HNRNPF,HNRNPR,HSP90B1,HSPA5,HSPA8,ICAM1,ICAM2,ID3,IFI35,IFITM1,IFNGR1,IFRD2,IGFBP6,IL10RB,IL13,IL15RA,IL1R1,IL2RB,IL2RG,IL32,IL4R,IL7R,IRF1,IRF8,IRF9,JAK1,LCK,LCP1,LONP1,LSP1,LYN,MACROH2A1,MAF,MAP3K10,MCM5,MDH2,MMP2,MT1G,MTHFD2,NAMPT,NASP,NCL,NFE2L2,NFKB1,NIT1,NME1,NOS2,NR4A1,NUDT1,OGDH,ORAI3,P4HB,PAFAH1B3,PCBP2,PCNA,PCYOX1,PCYT2,PDCD5,PDIA4,PFN1,PHB,PKLR,PLA2R1,PLEK,PLIN2,PNP,PRDX4,PRF1,PSMA6,PTEN,PTP4A1,PTP4A2,PTPRC,PYGL,RAB8A,RAC2,RARS1,RB1,RHOH,RNF128,RORA,RORC,RPL35A,RXRA,RYK,S100A11,SHC1,SLC25A5,SLPI,SNRPB,SOCS2,SRGN,SRSF7,STAT1,STMN1,TFDP2,TFEC,TFPI2,THY1,TLR2,TNFSF10,TOM1L1,TOP2A,TPST1,TRA2A,TRAF5,TRIM24,TRIM5,TSPAN7,TSTA3,TTF1,UQCR11,VEGFA,VPS4B,XAF1,ZFP36,ZNF76 |

| | | | | | | | | | |
|---|---|---|---|---|---|---|---|---|---|
| HGF | | growth factor | Activated | 2,403 | 2,479 | 0,007 | 4,61E-20 | 5,57E-18 | A2M,ABCC6,ABCG2,ACKR3,ADAMTS1,ADGRG1,ADGRL2,AFP,AHR,AIFM1,AKAP12,ARPC5,ASNS,ATF3,AURKA,BCL2A1,BIRC2,BIRC3,CALCRL,CALM1 (includes others),CASP1,CASP8,CAV1,CCL5,CCND1,CCNG1,CD40,CD44,CD93,CDC25B,CDK1,CDKN1B,CDKN2C,CDKN3,CGRRF1,COL1A1,COL1A2,COL3A1,COL4A1,COPB1,CSE1L,CTSK,CXCR4,CYP17A1,CYP2B6,DDIT3,DHPS,DOK5,DTYMK,DYNLL1,EBP,EFNB2,EHF,ESM1,F3,FANCG,FGF7,FKBP5,FLNA,FN1,FOS,FOSB,FOSL1,G6PC,GAK,GPSM2,GRB10,GRK5,GUCY1B1,HK1,ICAM1,ID1,IGFBP4,IL13,IL15RA,INPPL1,IRF8,ISG15,ISG20,ITGA2,ITGB8,ITPR3,KIF22,KITLG,KRT19,LCN2,LY75,MAD2L1,MAP2K3,MAT2A,MCM2,MCM5,MELK,MET,MID1,MMP1,MMP2,MMP9,MPC1,NAB2,NAMPT,NDC80,NFKB1,NID2,NOS2,NR0B2,NR4A1,NR4A3,NRP1,OSMR,PCK1,PCNA,PEMT,PEX5,PHB,PIK3CB,PLA2R1,PLOD1,PMP22,PRC1,PTGS1,PTPRF,PTTG1,RAC1,RFC2,RHOB,RIN1,S100A9,SFRP1,SGK1,SLC19A2,SLC2A2,SLC9A3R1,SNRNP70,SOCS2,SOD2,SPP1,SPRY2,ST14,STAB1,STK10,TACSTD2,TAGLN2,TGFBR2,TIMP1,TJP1,TNFAIP2,TNFAIP6,TOPBP1,TP53,TPX2,TTK,UBE2C,VCAM1,VDR,VEGFA,VEGFC,VIM,WT1,YWHAH |
| PPARG | 0,055 | ligand-dependent nuclear receptor | Inhibited | -4,127 | -4,097 | 0,003 | 5,03E-20 | 5,96E-18 | ABCG1,ABCG2,ACAA1,ACAA2,ACACA,ACADL,ACADM,ACADS,ACOT8,ACOX1,ACSL1,ACSL5,ADRB2,AGPAT2,ALDH9A1,APOA1,APOE,AQP3,ASAP1,ATP6V1D,BACE1,BAX,BCL6,BIN1,C3,CA2,CAT,CAV1,CAV2,CCL5,CCND1,CCR7,CD36,CD40,CDKN2C,CFD,CIDEC,COL1A1,COL1A2,CPT2,CRAT,CTPS1,CXCL1,CXCL14,CXCL6,CYBA,CYP4A11,DGAT1,DHRS3,DLAT,EHHADH,ELOVL6,EPHX1,ESRRA,FABP1,FABP2,FGF1,FN1,FOSL1,G6PC,GAK,GPD1,GPT,GSTA1,HADHA,HMGCS2,HR,ICAM1,IDH1,IGFBP6,IGFBP7,IHH,IL13,IL33,INSR,IVD,KRT19,LHPP,LOC102724788/PRODH,MCM7,MCTP1,ME1,MLYCD,MMP9,MRC1,MTOR,MTTP,MUC1,MUC13,MYD88,NDRG1,NFE2L2,NOS2,NOX4,NPPA,NR1H2,NR1H3,NR2F2,NR4A1,NSF,OXR1,PC,PCK1,PCK2,PCNA,PCTP,PDK4,PEA15,PECAM1,PEPD,PFKFB3,PLIN2,PMM1,PMVK,PPARA,PPARGC1A,PPIC,PTEN,PTGES,PTPRF,PYGL,RHOB,RORC,S100A8,SAP18,SCD,SCP2,SDC1,SGK1,SHBG,SLC25A20,SLC27A3,SLC2A2,SNCG,SOD1,SPP1,TBXAS1,TCF4,TGFBR2,TJP1,TKT,TLR4,TNFSF10,TP53,TRIB3,TUBB,UCP2,VAMP2,VAMP5,VCAM1,VDR,VEGFA,VIM,VLDLR,WIPF1,YWHAE |
| FOXO3 | -0,006 | transcription regulator | Inhibited | -3,148 | -3,135 | 0,002 | 5,08E-20 | 5,96E-18 | AGFG1,AHR,ALDH3A1,ANXA1,ANXA3,AURKA,BAX,BCL2L13,BCL6,BNIP2,BNIP3,BNIP3L,C1QA,CASP4,CASP8,CAT,CAV1,CCNB1,CCNB2,CCND1,CCND2,CCND3,CDH5,CDKN1B,CDKN1C,CDKN3,CEP55,CLDN1,COL4A1,COL4A2,CTSB,CTSV,CXCL10,CXCR4,DDIT3,DLGAP5,DNM1L,DOK5,ENPEP,ESR1,FN1,FOS,FOSB,FOXO1,FOXO4,FYN,GABARAPL1,GABRR2,GADD45A,GADD45B,GCLC,GLUL,GYPA,HIF1A,HMGCS2,HTATIP2,ICAM1,IFIH1,IGFBP7,IL10RB,IL13,IL1R1,IMPDH2,ITGAM,ITGB2,JAK1,LCN2,LCP2,LEMD3,MCAM,MCM2,MCM7,MEF2C,MET,MMP9,MTHFD2,MXI1,NAMPT,NDRG1,NOS2,OPTN,OVOL1,PBK,PCNA,PECAM1,PIK3IP1,PPARGC1A,PPP1R15A,PRDX3,PRKCD,PRNP,PTEN,PTGS1,PTPRC,PTTG1,RTN3,SELL,SGK1,SH2B3,SH3GLB1,SLIT2,SOD1,SOD2,SPAG5,SPTLC2,TNFRSF1B,TNFSF10,TP53,TPX2,TXN2,TXNIP,TXNRD2,UBE2C,UCP2,ULK1,VCAM1,VEGFA,VIM,ZWINT |
| Insulin | | group | | 0,942 | 0,972 | 0,003 | 5,83E-20 | 6,75E-18 | ACACA,ACADM,ACAT1,ACOX1,ACSL1,ACTB,ADAM10,ADIPOR1,AEBP1,ALB,ALDOB,APOA1,APOE,APOM,ARG2,ATF3,ATP5F1D,ATP5MC2,AVP,BACE1,CCND1,CCND3,CCT2,CCT5,CCT6A,CD36,CDKN1B,CDX2,CIDEC,COL1A1,COL1A2,COL3A1,COL4A1,COL5A1,COX6A2,CSHL1,CTH,CXCL1,CYBB,CYP17A1,DEFB1,DUSP1,ELOVL6,ETFA,FKBP8,FN1,FOS,FOSB,FOXO1,FTL,G6PC,GCLC,GRB10,GRK2,GSTP1,HIF1A,HNF1A,HPSE,HSD3B2,HSP90B1,HSPA5,HSPA8,HSPA9,ICAM1,IDH1,IFIT2,IFIT3,IGFBP4,INSR,IRF7,ITGA3,ITGB3,LAMB1,LEPR,LGALS3BP,LONP1,LPIN1,MAOA,MAOB,MAPT,MCM6,MCM7,MDH2,ME1,MLXIPL,MMP2,MMP9,MTCH2,MTOR,MTTP,NDUFS1,NDUFS2,NDUFV1,NFE2L1,NFIL3,NOS2,NR1H3,NR4A1,NR4A3,NUCB2,P4HB,PCCB,PCK1,PCNA,PDGFRA,PDK2,PDK4,PFKFB3,PKM,PLAAT3,PLIN3,PNPLA2,PPARA,PPARGC1A,PPIB,PPIC,PPP1R15A,PRKCA,PRKCD,PRKCZ,PTEN,RAC1,RETSAT,RPS16,SCD,SDHB,SGK1,SLC10A2,SLC25A11,SLC25A5,SLC29A1,SLC2A2,SLC2A5,SLC2A8,SLC37A4,SLC38A3,SLC39A14,SLC5A2,SMAD7,SOD1,STAT1,STAT3,TGFBI,THBS2,TJP1,TLR2,TNC,TRPV1,UCP2,UQCRC1,UQCRC2,VEGFA,VLDLR,XBP1 |

| | | | | | | | | | |
|---|---|---|---|---|---|---|---|---|---|
| RXRA | -0,422 | ligand-dependent nuclear receptor | Inhibited | -3,631 | -3,616 | 0,002 | 6,66E-20 | 7,60E-18 | ABCC2,ABCC3,ABCG1,ABCG2,ACACA,ACACB,ACADM,ACOX1,ACSL1,ADH1B,AHCY,ANGPTL3,APOA1,APOC3,APOE,APOM,ARG2,ARL4C,ASNS,BAAT,C1QA,C3,CALB1,CAT,CCL20,CCNA2,CCNB1,CCND1,CCND3,CD36,CD38,CEBPE,CLMN,COL1A1,COLEC12,CPT2,CSHL1,CTSB,CTSS,CXCL10,CYP27B1,CYP2B6,CYP3A4,CYP4A11,EHHADH,ENG,EPHX1,FABP1,FABP2,FLRT3,FOS,FSTL1,FTL,GCLC,GPD1,GPT,HADHA,HCK,HIF1A,HMGCS2,HNF1B,HNF4A,HOXA1,HOXB1,IGFBP6,IL13,KLF9,KNG1,LCN2,LTF,LYZ,MAOB,MGP,MLXIPL,MLYCD,MMP2,MYL7,NEDD9,NME1,NPPA,NR0B2,NR1H3,PC,PCK1,PCK2,PKLR,PLD1,PLTP,PMP22,PPARA,RARG,RBP1,RGR,RORA,RORC,RXRA,S100A4,S100A8,SCD,SDC1,SEMA4D,SLC10A2,SLC27A5,SLC2A2,SLC34A1,SPP1,STRA6,TXN,VCAN,VDR,VEGFA |
| SP1 | 0,037 | transcription regulator | | 1,256 | 1,337 | 0,009 | 8,33E-20 | 9,39E-18 | ABCC8,ACSL1,ADA,ADAMTS1,ADD3,ADH5,AEBP1,AKAP12,ALDH3A2,ALOX5,ALPL,ANKH,APOA1,APOC3,APOE,ASNS,ASS1,ATF3,B4GALT5,BACE1,BAX,BMP7,BNIP3L,C1QBP,CASP3,CAT,CAV1,CCL20,CCL5,CCNA2,CCND1,CCND2,CCND3,CD40,CD55,CD59,CD99,CDC25B,CDK1,CDKN1B,CDKN2C,CIRBP,COL1A1,COL1A2,CRYBB3,CTH,CTSD,CXCL10,CXCL12,CXCR4,CYBRD1,CYC1,CYP17A1,CYP1B1,CYP2B6,ENG,ESR1,ESRRA,F3,FAS,FDX1,FLNA,FN1,FOLR2,FOS,FOSL1,FYN,GHR,GJA1,GNAI2,GNAS,GSS,HADHA,HBB,HDAC1,HIF1A,HNF4A,HPSE,HSD11B2,HSPA5,ICAM1,IFNGR1,IGFBP4,IL15,IL2RB,IL4R,INSR,IRF1,ITGA2,ITGAM,ITGAV,ITGB3,JAM2,KRT19,KRT4,KRT81,LHB,LIPA,LTB,LTBP3,MAOA,MAOB,MAT2A,MET,MMP1,MMP2,MMP9,MRC1,MSH6,NAB2,NCF2,NDRG1,NFKB1,NOX4,NOX5,NPPA,OGG1,PARP1,PCK1,PDGFRA,PEMT,PKM,PLA2G4A,PRKCA,PRKCD,PRLR,PRNP,PROCR,PROM1,PSIP1,PTDSS1,PTGS1,PTH1R,PTTG1,RB1,RECK,RRAS,SELL,SERPINE2,SGK1,SIGIRR,SLC22A4,SLC25A5,SLC31A1,SLC9A3,SMAD7,SMPD1,SNCG,SOAT1,SOD1,SOD2,SPARC,SPP1,STAT1,STAT3,TERT,TFPI2,TGFBR2,TGFBR3,THRB,TIMP1,TIMP2,TLR2,TNC,TNFSF10,TNNC1,TP53,TSPO,TUG1,UGCG,VDR,VEGFA,VIM,VWF,[illegible] |
| Ifn | | group | Activated | 5,245 | 5,293 | 0,007 | 1,12E-19 | 1,25E-17 | ADAMTS1,APOBEC3A,B2M,BAG1,BCL6,CASP8,CD40,CD44,CD58,CD69,CD83,CD86,CFH,CXCL10,CXCL11,CYP4A11,DDX58,FAS,FOS,HIF1A,HLA-A,ICAM1,IDO1,IFI16,IFIH1,IFITM1,IL10RA,IL15,IL15RA,IL1R1,IL32,IL33,IL7,IRF1,IRF7,IRF8,ISG15,ISG20,LCN2,MMP9,MX1,NOS2,NTRK2,OAS2,OAS3,PECAM1,PLAAT4,RNASEL,RSAD2,S100A8,SECTM1,SPP1,STAT1,TAP1,TLR1,TLR2,TLR3,TLR5,TLR7,TP53,TRIM21,TRIM22 |
| FOS | -1,302 | transcription regulator | | 0,080 | 0,103 | 0,003 | 1,21E-19 | 1,33E-17 | A2M,ABCC1,ACACA,ACOX1,ACTG1,ADNP,AGPS,ALDH1A3,ANXA4,AQP3,ARPC5,ASAP1,ASL,ATF3,ATF7IP,C3AR1,CA2,CALU,CAPN2,CASP3,CAST,CAT,CCL20,CCL5,CCNA2,CCND1,CCNG1,CD14,CD164,CD44,CELF2,CGA,CLU,COL16A1,COL18A1,COL1A1,CRABP1,CSTA,CTH,CTSB,CTSV,CYP17A1,CYP2J2,DAPK2,DNMT1,EIF4G2,ELOVL6,ENAH,ENO1,EPHX2,ESM1,EVX1,F3,FBN1,FEZ2,FN1,FOLR2,FOS,FOSB,FOSL1,FRZB,FSTL1,FZD5,FZD6,G6PC,GAK,GAS2,GJA1,GM2A,GPER1,GPSM2,GSR,GSTP1,HDAC6,HIPK3,HLA-B,HOXA1,HPGD,HPGDS,HSD17B10,HSD3B2,HSP90B1,HSPA5,HSPB8,HTATIP2,ICAM1,IGFBP6,KIF2A,KITLG,LETMD1,LGALS3BP,LHB,LTBP3,MAOA,MAP4K4,MCM5,MET,MMP1,MMP2,MMP7,MMP9,MPZL2,MYH9,NAE1,NCF2,NDUFS7,NET1,NFE2L2,NFIL3,NGF,NIPSNAP1,NPLOC4,NPPA,NR1H3,NR4A1,NSDHL,PAOX,PCYT2,PFN1,PGAM2,PIK3CB,PLA2G4A,PLD1,PLK2,PTH1R,PTP4A1,PTP4A2,PTPN12,PTPRD,PTTG1,RARG,RBBP7,RBP1,RFNG,RPS24,RXRA,S100A10,S100A8,S100A9,SAP18,SCD,SDC1,SELENBP1,SEMA3B,SEMA4D,SERPINB9,SERPINE2,SLC10A2,SLPI,SMAD7,SNCG,SNN,SPP1,STMN1,SULT2B1,SWAP70,TAC1,TGFBI,TIMP1,TIPARP,TNFRSF11B,TP53,TUBB,TXN,UGCG,UTRN,VCAM1,VEGFA,VIM,WNT10B,XAF1 |
| MAP4K4 | 0,499 | kinase | Activated | 5,840 | 5,768 | -0,009 | 1,24E-19 | 1,34E-17 | ACAA2,ACACA,ACACB,ACADS,ACADVL,ACO2,ACSL5,AGPAT3,ALDH3B1,ALDH4A1,ARAF,AUH,BCKDHB,CHPT1,COQ3,COX11,CYC1,DHODH,DLAT,DLD,DLST,GCDH,GLUL,GOT1,GRHPR,GYS1,HADH,HADHA,HMGCL,HYAL2,IMPDH1,IVD,MAFB,MLXIPL,MPI,NDUFA3,NDUFAB1,NDUFB7,NDUFS1,NDUFS4,NDUFS8,NDUFV1,NT5M,OGDH,PEX19,PEX3,PGM1,PHYH,PLA2G12A,PLCB4,PMM1,PNPLA3,PON2,PTDSS2,PTGES,PXMP2,SCD,SCP2,ST3GAL4,SUCLG1,UQCRC1 |

| | | | | | | | | | |
|---|---|---|---|---|---|---|---|---|---|
| TCR | | complex | | 0,220 | 0,274 | 0,006 | 1,99E-19 | 2,13E-17 | ADM,AIM2,AK2,APOA4,APOC3,APOE,BACH2,BCL2A1,BIN1,BIRC3,CARD8,CCL5,CCND1,CCND3,CCR2,CCR5,CCR7,CD200,CD27,CD38,CD44,CD69,CDKN1B,CISH,CTSB,CXCL10,CXCL11,CXCL9,CXCR4,CYTIP,DDX58,ECH1,ELL2,ENO1,FAS,FLNA,FOS,FYN,GADD45B,GADD45G,GLUD1,GLUD2,GNPDA1,GZMA,GZMB,HADH,HIF1A,HSD17B10,HSP90B1,HSPA5,HSPA9,ID3,IFI44,IFI44L,IFI6,IFIH1,IFIT2,IFIT3,IFNGR1,IL13,IL4R,IL7,IL7R,IRF1,IRF7,ISG15,ISG20,KCNK5,LAT,LBR,LCK,LCP2,LIME1,LIPA,LMNB1,MAF,MAPK8,MDH2,MX1,MX2,MYD88,NAB2,NDUFA10,NFKB1,NME1,NR4A1,NRP1,OAS1,P4HB,PDCD4,PDIA4,PHB,PIK3R3,PRF1,PRKACB,PRMT1,PTEN,PTPRC,RASGRP1,RNF128,RPL13A,RPL3,RPS16,RPS3,RSAD2,SDCBP,SELL,SERPINA3,SLC1A5,SLC25A3,SMAD7,SOCS2,SPP1,STAT1,STAT3,STAT4,TLR3,TLR4,TLR7,TNFRSF1B,TOB1,TPI1,TRAF5,TSC22D1,TUFM,UQCRC1,VAV1,VDR,ZBTB7B |
| EGF | -1,786 | growth factor | | 1,813 | 1,885 | 0,006 | 5,08E-19 | 5,17E-17 | ABCG2,ACTN1,ADA,ADAM17,AFP,APOA1,AQP3,ARC,ATF3,AURKA,B4GALT5,BIRC2,CASP1,CASP3,CAV1,CCL20,CCNB1,CCND1,CCND2,CCND3,CD44,CDH11,CDK1,CDK5,CDKN1B,CDX2,CEBPE,CGA,CISH,CLCN3,CLDN3,CLU,COL1A1,COL1A2,COL3A1,CSF1R,CTCF,CTSD,CXCL14,CXCL6,CXCR4,CYP2B6,DCN,DDIT3,DPP4,DUSP1,EFNB2,ENAH,ESR1,F3,FAM89B,FN1,FOS,FOSB,FOSL1,FSTL1,FUT3,GAB2,GADD45A,GAK,GAL3ST1,GDF2,GFPT1,GJA1,GPER1,GPS2,GSTP1,HIF1A,HNF4A,HOXB5,HPGD,HSD3B2,ICAM1,ID1,ID2,IDH1,IDI1,IGFBP4,IL1R1,IL7R,ITGA2,ITGAM,ITGB3,JUND,KRAS,KRT19,LCN2,LTF,MAD2L1,MET,MMP1,MMP2,MMP9,MUC1,NCL,NFE2L1,NGF,NOS2,NPPA,NR4A1,NR4A3,NRP1,PBK,PCNA,PDCD4,PDK4,PER1,PLSCR1,PPIB,PRKCH,PROM1,PSMB5,PSMB6,PSMB7,PTGES,PTGS1,PTPN12,PTTG1,RAC1,RHOA,RHOB,RRM1,RRM2,RXRA,S100A10,S100A11,S100A4,S100A9,SCD,SDC1,SERPINA3,SH2B3,SHC1,SLC37A1,SMAD1,SOX4,SPARC,SPP1,SPRY2,ST3GAL1,ST3GAL4,STAT1,TBP,TERT,TFPI2,TGFBR2,TIMP1,TJP1,TM4SF5,TNC,TP53,TUFM,ULK1,VAMP2,VCAM1,VCAN,VDR,VEGFA,VEGFC,VIL1,VIM,ZFP36 |
| Vegf | | group | Activated | 2,786 | 2,875 | 0,007 | 5,36E-19 | 5,40E-17 | A2M,ACE,ACKR3,ADAM10,ADAMTS1,ADGRG1,ADGRL2,AHR,AKAP12,ALOX5AP,APOM,ARHGEF17,ASNS,ATF3,AURKA,BAX,BCL2A1,BMP7,CA2,CALB1,CALCRL,CALM1 (includes others),CASP1,CASP3,CASP8,CD40,CD44,CD93,CDC25B,CDH5,CDK1,CDKN2C,CDKN3,CLDN1,CPA3,CRABP1,CSE1L,CSGALNACT1,CXCL1,CXCL10,CXCL11,CXCL12,CXCR4,CYP4A11,DDIT3,DENND2D,DHPS,DNAJB9,DOK5,DTYMK,EBP,EFNB2,EHF,EIF2B2,ENPP2,ERBB3,ESM1,F3,FANCG,FAS,FKBP5,FLNA,FN1,FOSB,FOSL1,GASK1B,GPSM2,GRB10,GRK5,GUCY1B1,HIF1A,HMGCS2,HOPX,HPSE,ICAM1,ID1,IGFBP4,IHH,IL15RA,IL18,IL1R1,IRF1,ISG20,ITGA2,ITGB8,ITPR3,JAM2,KIF22,KITLG,LCN2,LPAR1,LPAR6,LY75,LYN,MAD2L1,MAOB,MAP2K3,MCM2,MCM5,MEF2C,MELK,MET,MGP,MGST2,MID1,MIF,MMP2,MMP9,MPC1,MT1G,NAB2,NAMPT,NDC80,NFKB1,NID2,NPL,NR2F2,NR4A1,NR4A3,NRP1,OSMR,P2RY14,PC,PEMT,PEX5,PIK3CB,PLOD2,PMP22,PRC1,RAC1,RCAN1,RECK,SFN,SFRP1,SGK1,SLC19A2,SOCS2,ST8SIA4,STAT3,STK10,TACSTD2,TGFBR2,TIMP1,TJP1,TLR3,TNC,TNFRSF11B,TPST1,TPX2,TRAF5,TTK,UBE2C,UGCG,VASH1,VCAM1,VEGFA,VEGFC,VIM,VWF |
| dihydrotesto sterone | | chemical - endogeno us mammalia n | | 0,237 | 0,285 | 0,005 | 2,17E-18 | 2,06E-16 | ABCG1,ACACA,ACADVL,ACKR3,ACO2,ACTB,ADAMTS1,ADM,ALDH1A3,APOE,APOM,AQP1,AVP,AZGP1,B2M,BASP1,BAX,C1orf116,CASP1,CASP3,CASP8,CCNA2,CCNB1,CCND1,CCND2,CCND3,CCT3,CD36,CD63,CD74,CDKN1B,CGA,CKM,CLU,COL3A1,COL4A1,COL5A2,CST6,CYBB,CYP17A1,CYP4A11,DCN,DCTPP1,DPP4,DTL,DYNLL1,EGF,ENO1,ESR1,FAM89B,FGF7,FKBP4,FKBP5,FN1,FOS,FOXO1,FTL,FZD6,G0S2,GHR,GLUL,GNMT,GPD1,GPX3,GPX5,GRB10,GRN,GSTP1,HLA-A,HPGD,HPGDS,HSD11B2,HSD3B2,ID2,IGFBP4,IGFBP7,INSR,IQGAP2,ITGA2,KLK1,KRT19,LAMA5,LGALS1,LHB,LYZ,MAF,MAFF,MCAM,MCM5,ME1,MME,MMP1,MMP2,MMP7,MMP9,MUC1,NAT1,NDRG1,NEDD4L,NID2,NOS2,NR4A1,NUCB1,OXT,PC,PCK1,PDCD4,PEA15,PER1,PFN1,PHB,PHF21A,PHGDH,POSTN,PPARA,PRNP,RAB27A,RARG,RB1,RGN,RHOB,RPL28,RPL3,RPL8,RPLP2,RPS20,RPS24,SCD,SCP2,SDC3,SELENBP1,SERPINE2,SGK1,SHC1,SLC16A7,SLC19A2,SLC1A3,SLC1A5,SLC25A16,SLC2A5,SLC2A9,SLC3A2,SLC4A4,SLC7A7,SLC7A8,SLC7A9,SLCO3A1,SPARC,SPP1,SRD5A2,SSRP1,STARD8,STXBP1,TACC2,TERT,TGFBR2,TGFBR3,TIMP2,TKT,TMPRSS2,TNFSF10,TP53,UAP1,VCAM1,VCAN,VEGFA,VIM,VLDLR,WIF1,YTHDC1 |

| | | | | | | | | | |
|---|---|---|---|---|---|---|---|---|---|
| IL10 | | cytokine | | 0,044 | 0,054 | 0,001 | 6,60E-18 | 6,20E-16 | ADM,ALOX5AP,ARAP2,ARG2,ARNT2,BAX,BCL2A1,CASP3,CASP8,CCL18,CCL19,CCL20,CCL5,CCND2,CCND3,CCR2,CCR5,CCR7,CD14,CD163,CD2,CD40,CD44,CD55,CD83,CD86,CDKN1B,CHUK,CLEC7A,COL1A1,CSF1R,CSF2RB,CTSB,CTSS,CTSZ,CX3CR1,CXCL10,CXCL11,CXCL12,CXCL6,CXCL9,CXCR4,DCN,DEPP1,DUSP1,EHF,F3,FAAH,FAS,FCER1G,FCGR2B,FCGR3A/FCGR3B,FKBP5,FOS,FPR1,GZMA,GZMB,HLA-G,HPGD,HTATIP2,ICAM1,IDO1,IFIT2,IGFBP4,IL10RA,IL13,IL18,IL1R1,IL33,IL4R,IL7,IL7R,INPP5D,IRAK1,IRF1,ITGAL,ITGAM,KITLG,KLRD1,LAMP3,LGALS1,LILRB2,LTB,LY75,MAF,MMP1,MMP2,MMP9,MRC1,MUC1,MYD88,NEDD9,NOD2,NOS2,NPHS1,NR4A3,PHGDH,PSMB9,RARRES1,S100A8,SELL,SLC1A3,STAT1,STAT3,STAT4,TAP1,TAP2,TERT,TIMP1,TLR2,TLR3,TLR4,TLR5,TLR7,TLR8,TNFRSF17,TNFRSF1B,TNFSF10,TUBB4B,VCAM1,VCAN,VEGFA,VTCN1,ZFP36 |
| IL3 | | cytokine | | 0,593 | 0,641 | 0,007 | 6,76E-18 | 6,28E-16 | ACOX1,ADA,ADGRE5,AKR1A1,ALDH2,ALOX5AP,ARHGAP29,ARHGDIB,BIRC3,BNIP3L,C1QA,CALR,CASP3,CAT,CBFB,CCL19,CCN3,CCND1,CCND2,CCND3,CCNG1,CCT5,CD14,CD247,CD3D,CD3G,CD40,CD48,CD53,CD63,CD69,CD83,CD86,CDKN1B,CISH,CRAT,CSF1R,CSNK1A1,CXCL9,CXCR4,DDIT3,DHPS,ECH1,EMP3,FAS,FCER1A,FCGR2B,FKBP3,FOS,FUS,GADD45A,GADD45B,GADD45G,GNAS,GPR21,GZMA,HK1,HLA-A,HNF1A,HNF4A,HSP90B1,HSPA5,HSPA9,ICAM1,IK,IL13,IL33,IMPDH2,ITGAM,ITGAV,ITGB3,JCHAIN,KLF9,KPNB1,LAMP3,LCN2,LY75,LYN,MAN2B1,MCM5,MID1,MLF2,MMP1,NCL,NEDD9,NRP1,PFKL,PHGDH,PLA2G7,PSMB7,PSME1,RARS1,RB1,RIDA,RPL3,RPS16,RPS19,RPS20,RPS3,SELL,SERPINA3,SLC3A2,SNRPD2,SOD1,SOX4,STAT4,STT3A,TARS1,TERT,TLR2,TLR4,TNFSF10,TYK2,UCP2,VCAM1,VEGFA,WNK1,XBP1 |
| Growth hormone | | group | Inhibited | -2,154 | -2,123 | 0,004 | 7,03E-18 | 6,46E-16 | ACAA1,ACACA,ACSL1,ACTB,ADRB2,AKR1A1,AKR1B1,ALB,APOE,ATP5MC2,BCL6,BMP7,BPHL,C3,CALB1,CAT,CCND1,CCND2,CD36,CDH3,CISH,CLDN15,CLU,COL18A1,COL1A1,COL3A1,COMT,CSHL1,CTH,CYP27B1,CYP2C8,CYP3A4,DDIT3,DDR1,DECR1,DYRK2,FGFR3,FKBP5,FN1,FOS,FZD5,G6PC,GCDH,GCLC,GHR,GLUD1,GPT,GSTA1,GSTP1,HADH,HBB,HOXA1,ID1,IGFBP6,INSR,IRF1,LEPR,MMP9,NAMPT,NEDD9,NOX4,NPPA,NR2F6,PCK1,PCNA,PDK4,PFKFB3,PIK3C2B,PLAAT3,PLK2,PPARA,PPP1R15A,PRDX3,PRKAG2,PRLR,RAB3A,SGK1,SLC7A9,SOCS2,SOD1,SPP1,SST,TFF3,TGFBR2,TKT,TNFSF10,TSC22D1,TUBA1A,TXNIP,ZEB2 |
| NFE2L2 | 0,255 | transcription regulator | | -1,188 | -1,114 | 0,007 | 1,08E-17 | 9,86E-16 | ABCC1,ABCC2,ABCC3,ABCG2,ACSL5,ACTG1,AGPAT3,AHR,AKR1A1,AKR7A2,ALDH3A1,ANG,AOX1,APOA4,ATF3,BHMT,BNIP3,CA11,CALB1,CAT,CCL5,CCT3,CD36,CD86,CDKN2C,CFD,CLPP,COL1A1,COL3A1,COL4A2,CTSD,CTSK,CUL1,CXCL10,CXCL6,CYBB,DDIT3,DDX50,DHCR7,DNAJB5,DSTN,DYNLL1,EIF4G2,ENTPD5,EPHX1,FABP1,FGF13,FKBP5,FMO1,FN1,FTL,GADD45G,GAS2,GCLC,GCLM,GHR,GNA11,GNAI2,GPX3,GSR,GSS,GSTA1,GSTA3,GSTM4,GSTO1,GSTP1,HAX1,HNF1A,HPRT1,HSP90B1,HSPA9,HTATIP2,IDH1,IFNGR2,IFRD1,IL13,IMPDH1,MAFF,MAPK10,ME1,MEF2C,MMP9,MORF4L2,MSMO1,NAT8B,NCF2,NFE2L1,NFE2L2,NOCT,NOS2,NR0B2,NR1I3,NSMF,NUCB2,OAT,OSGIN1,PDIA3,PDIA4,PDIA6,PHGDH,PLIN2,PPARGC1A,PPIB,PSAT1,PSMA4,PSMA6,PSMB5,PSMB6,PSMD1,RACK1,RARS1,RBBP7,RPS16,S100A13,SERINC3,SERPINA3,SERPINA6,SHC1,SHMT2,SLC1A1,SLC35B1,SLC38A1,SLC38A3,SLC7A8,SOD1,SOD2,SRGN,TBXAS1,TCN2,TKT,TP53,TPI1,TXN,UBQLN2,USP14,VCAM1,VEGFA,XBP1 |
| Lh | | complex | | -1,168 | -1,152 | 0,003 | 1,18E-17 | 1,06E-15 | ACP5,ACTB,ACTN1,ACTR2,ADAMTS1,ADCY7,ADCY9,ADGRG1,AKAP12,ALPL,ARL4C,ARL6IP5,ATP9A,AXL,BAG1,BNIP3L,CAMK2G,CAP2,CASP4,CCND2,CD55,CDK14,CDK5,CDKN1C,CHUK,COL15A1,COL18A1,CSNK1A1,CXCR4,CYFIP2,CYP17A1,DAB2,DHCR7,DHRS3,DUSP1,DUSP3,DUSP9,FDX1,FKBP5,FOS,GBP1,GJA1,GK,GNAS,GNB1,GNLY,GRK5,HSD3B2,INHA,ITGA3,MAP2K3,MAP4K4,MMP2,MMP9,MSMO1,P4HA2,PER1,PGRMC1,PHB,PHKA2,PLCL1,PLIN3,PPIH,PRKD1,PRKX,PSIP1,PSMD1,PTGS1,PTP4A1,PTPRE,PTPRF,RAB27A,RAB31,RAB5C,RGS7,RHOB,SGK1,SLIT2,SMARCA4,SOD2,SRD5A2,STAT1,STK25,TFPI2,THBS2,TMBIM6,TNFAIP6,TNFRSF11B,TP53,TPM1,TUBA1A,TYRO3,VEGFA,VEGFC,YBX3 |
| IRF1 | 0,758 | transcription regulator | Activated | 6,314 | 6,372 | 0,007 | 1,29E-17 | 1,14E-15 | B2M,BAX,C1R,CASP1,CASP3,CASP8,CCL19,CCL20,CCL5,CCNB1,CCND1,CD40,CFB,CTSS,CXCL10,CXCL11,CYBB,DDX58,EIF4A3,ERAP1,FGL2,GBP2,HLA-G,IDO1,IFI27,IFI35,IFI44L,IFI6,IFIH1,IFIT2,IFIT3,IFIT5,IFITM1,IL13,IL15,IL18,IL7,IRF1,IRF2,IRF7,IRF9,ISG15,LTB,MAP4K4,MMP9,MX1,MYB,NOS2,OAS1,OAS2,PCNA,PIGR,PLAAT3,PLAAT4,PSMB10,PSMB8,PSMB9,PSME1,RSAD2,SELL,SLPI,SOCS7,SP110,STAT1,STAT3,TAP1,TAP2,TAPBP,TERT,TLR3,TNFSF10,TP53,TRIM21,TRIM22,UBD,VCAM1,XAF1 |

| | | | | | | | | | |
|---|---|---|---|---|---|---|---|---|---|
| RELA | | transcription regulator | Activated | 4,000 | 4,063 | 0,007 | 2,10E-17 | 1,81E-15 | A2M,ABCG2,AFP,AHR,ALOX5AP,APOE,ARHGDIB,B2M,BACE1,BACH2,BAX,BCL2A1,BHLHE40,BIRC2,BIRC3,BLVRA,BNIP3L,C3,CARD8,CASP3,CASP8,CAV1,CCL19,CCL20,CCL5,CCND1,CCR7,CD14,CD40,CD44,CD59,CD69,CDKN1B,CFB,COL1A1,COL1A2,CTCF,CTSB,CXCL1,CXCL10,CXCL11,CXCL6,CXCL9,CXCR4,CYBB,CYP17A1,CYP2B6,CYP3A4,DDIT3,DUSP1,EHF,ELMO1,ERAP1,F3,FAS,FCGRT,FN1,FOS,FOSB,FSCN1,GBP1,GCH1,GRK5,GSTA1,HIF1A,HLA-B,HNF4A,HSP90B1,ICAM1,ID1,IFNGR2,IGHM,IGKC,IL13,IL15RA,IL32,IL7R,IRF1,IRF7,ISG15,LTB,LUM,LYN,MAT2A,MIF,MMP1,MMP9,MUC1,MYB,NAMPT,NFE2L2,NFKB1,NFKBIE,NMI,NOD2,NOS2,NOX5,NR1I3,NR4A1,NR4A3,OAS2,PCK1,PECAM1,PKM,PLD1,POU2AF1,PPM1D,PRKCD,PSMB10,PSMB9,PTEN,PTGES,PTPN6,RORC,S100A6,SELP,SERPINE2,SHBG,SLC2A5,SMAD7,SOD2,TAC1,TAP1,TAP2,TAPBP,TERT,TFPI2,THOC1,TLR2,TNFAIP2,TNFRSF11B,TP53,TPMT,UBD,VCAM1,VEGFA,VIM,WT1 |
| HIF1A | 0,653 | transcription regulator | | 1,427 | 1,498 | 0,007 | 7,92E-17 | 6,57E-15 | ABCC8,ACO1,ADAM10,ADAM17,ADAMTS1,ADM,AFP,AKAP12,ALOX5AP,APOE,ARPC2,ASNS,AURKA,AXL,BACE1,BACH2,BAX,BHLHE40,BHLHE41,BID,BIRC2,BNIP3,BNIP3L,CAV1,CCL5,CCN3,CCND1,CCR2,CCR5,CCR7,CD164,CD24,CD36,CD44,CDCP1,CDKN1B,CLDN1,CORO1A,CTPS1,CTSD,CXCL12,CXCR4,CYB5A,CYP46A1,EED,ENG,ENO1,ESRRG,FHL1,FHL2,FN1,FOS,FSCN1,FYN,FZD2,GADD45B,GCHFR,GHR,GJA1,GLYR1,GPER1,H2BC12,HIF1A,HMGCL,HSPA5,ID1,ID2,IL15,ITGAV,ITGB2,ITGB3,KITLG,KLHL20,KRT19,LGALS1,MAFF,MANF,MEF2C,MET,MIF,MMP1,MMP2,MMP7,MMP9,MUC1,NAE1,NDRG1,NOS2,NOX4,NPPA,NR4A1,NR4A3,NTRK2,P4HA1,P4HA2,PFKFB3,PFKL,PKM,PLAC8,PLOD2,PPARA,PRKCA,PROM1,PTEN,RAPGEF6,RIOK3,RORC,SFRP1,SIRT2,SLC16A3,SLC16A4,SLC29A1,SMAD7,SOD2,ST3GAL1,STAT3,STC2,TAC1,TCF4,TERT,TLR2,TMEM45A,TNFAIP2,TNFRSF11B,TP53,TPI1,TRIM21,TXN,VCAM1,VEGFA,VEGFC,VIM,WNT10B,WT1,ZNF267 |
| EGFR | | kinase | | 1,160 | 1,207 | 0,004 | 1,09E-16 | 8,91E-15 | ABCG2,ACOT7,ACTB,ANXA1,BAX,BCL6,BZW2,CAPZA2,CASP3,CASP4,CAV1,CCL5,CCNA2,CCND1,CCT5,CD47,CDCP1,CDK1,CDKN1B,COL1A1,COL3A1,COL5A2,CRMP1,CTSK,CUL1,CXCL10,CXCL12,CXCL14,CXCR4,DDIT3,DSG2,DUSP1,EFNB2,EGF,EIF6,ERBB3,ESR1,F2RL1,F3,FBN1,FGF7,FKBP11,FOS,FOSL1,GADD45A,GBP1,GHR,GJA1,GRN,GZMB,HIF1A,HMGB3,HSD11B2,HSP90B1,HSPA5,ICAM1,ID1,IGBP1,IGFBP4,IL33,IL36A,IQGAP2,IRF1,ITGA2,JUND,KPNB1,LCN2,MACF1,MARCKS,MET,MME,MMP1,MMP2,MMP9,MT1F,MTHFD1,MUC1,MXI1,NCL,NOS2,NPPA,NSF,OSMR,PCNA,PKM,PLA2G4A,POSTN,PRF1,PRKCZ,PROM1,PRORP,PSMB5,PSMB6,PSMB7,PTEN,PTGES,PTGS1,PTPRE,QTRT1,RB1,RBBP7,RHOB,RRAS,S100A2,SDF2L1,SDHB,SERPINA3,SERPINE2,SFRP1,SLC25A32,SLPI,SPRY2,STAT3,TBXAS1,TIMM10,TM4SF5,TNC,TNFAIP6,TNFRSF11B,TPST1,TTF1,TUBB4B,TUBG1,TXNIP,UQCRC2,VCAN,VDR,VEGFA,VIM,WNT10B,ZEB2 |
| IL10RA | 1,302 | transmembrane receptor | Inhibited | -2,510 | -2,507 | 0,000 | 1,31E-16 | 1,06E-14 | ABAT,ABCG1,ACSL1,ACSM3,ADD3,ADGRL2,ALDH2,ALDOB,ALOX5,ANKH,AOC1,ARG2,ARL5A,ASS1,BAX,BNIP3,C3,CA2,CCL5,CCN3,CCR7,CD1D,CD36,CD40,CD69,CFB,CLCN7,CST7,CTH,CYFIP2,DCN,DPEP1,DRAM1,EFEMP1,EGF,EHF,ENTPD5,FAS,FBN1,FHL1,FN1,FOLR2,FSTL1,GBP2,GHR,GPR171,GPR18,HAO2,HLA-A,HPGD,HPSE,HSPA8,IFI16,IL15RA,IL2RG,IRF1,IRF7,ITGAM,KITLG,LCN2,LUM,MAOB,MECOM,MECR,MMP25,MSRB1,MYB,NAMPT,NOS2,NPL,NR1H3,PCK1,PCOLCE,PDGFRA,PLAAT3,PMP22,POSTN,PPARGC1A,PPT2,PROCR,PSMB8,PSMB9,PYGL,RSAD2,S1PR1,SAMHD1,SECTM1,SELENBP1,SELL,SERPINB9,SLAMF8,SLC16A3,SORBS2,SPARC,SPON1,ST3GAL6,STARD8,STAT1,STAT3,SUCLG2,SULT1C2,TAC1,TAP1,TAP2,TFEC,THBS4,TLR2,WFDC2 |
| STAT6 | | transcription regulator | | -1,345 | -1,329 | 0,002 | 2,12E-16 | 1,69E-14 | ACOX1,ACP5,ACSL1,ALDH2,ANXA1,APOE,APOLD1,ARNTL2,ASS1,AUH,BAX,BCL6,CASP6,CCL20,CCL5,CCL8,CCND2,CCR5,CCR7,CD2,CD38,CD40,CD69,CDKN1B,CEL,CIDEB,CIDEC,CISH,CLPS,COL1A1,COL1A2,CRADD,CRIP1,CST7,CTNS,CTSB,CTSS,CXCL10,CYBB,CYP4A11,DDAH1,DOCK10,EPHX1,EXT1,FABP1,FAM49A,FCER1G,FCGR3A/FCGR3B,FKBP5,FSCN1,GADD45B,GADD45G,GALK2,GATM,GBP2,GPM6B,HCK,HLA-DMA,HLA-DMB,HMGCS2,HRH1,IFI16,IFI44,IFIH1,IFIT3,IFNAR2,IL10RA,IL13,IL15,IL33,IL4R,IRF1,IRF7,IRF9,ISG15,ISG20,KCTD12,KMO,LCP2,LGALS3BP,LTB,LY75,MAF,MMP2,MMP9,MYB,NEDD9,NFIL3,OAS3,PDK4,PLD1,PLSCR1,PNPLA2,POLE2,POU2AF1,PPARA,PPARGC1A,PRKCA,PRNP,RNF19B,S100A10,SAMSN1,SELENBP1,SELP,SERPINF1,SOCS2,SOX13,ST6GAL1,STAT4,STK39,SYK,SYNGR2,TBC1D17,TES,TFEC,TFF3,THY1,TIMP1,TLE1,UBL3,USP2,VCAM1,VIM,XBP1,ZFP36 |

| | | | | | | | | | |
|---|---|---|---|---|---|---|---|---|---|
| MTOR | -0,241 | kinase | | -0,084 | -0,059 | 0,003 | 2,40E-16 | 1,90E-14 | ACACA,ACACB,ACADL,ACADM,ACOX1,ADM,AIFM1,AKR1A1,AKR1B1,ALDH1A3,ARL4C,BNIP3,CASP1,CASP3,CASP8,CCND1,CCND3,CCR7,CD40,CD86,CPT2,CRADD,CSF1R,CTCF,CXCL1,CXCR4,CYP17A1,DDIT3,DEFB1,ENO1,ETFA,FGFR3,FN1,GLUD1,GPHN,GSTZ1,HADHA,HIF1A,HK1,HMGCS2,HSP90B1,IFI16,IL13,IL1R1,IL2RB,IL4R,INHA,IQSEC2,IRAK1,IRF1,IRF8,MAPT,MDH2,MLXIPL,MLYCD,MMP9,MTOR,MUC1,NAB2,NFKBIL1,NOS2,NOX4,OAS1,PCNA,PDK2,PDK3,PDK4,PECAM1,PFKFB2,PFKP,PFN1,PGRMC1,PKM,PPARA,PPARGC1A,PPIB,PRKCZ,PRX,PSMB5,PTEN,PTH1R,PTPN6,PYGB,RHOA,RORC,RPL23,S1PR1,SELL,SERPINB1,SGCA,SLC12A2,SLC16A3,SMPD1,SNCG,SND1,SOAT1,SOD1,SOX4,SRPX,ST6GAL1,STAT1,STAT3,TCF4,TLR4,TLR5,TMPRSS2,TNFRSF11B,TNFSF10,TP53,TXNIP,UBE2B,UBE2C,UQCRC2,UTRN,VCAM1,VEGFA |
| TCF3 | | transcription regulator | Activated | 1,972 | 2,007 | 0,005 | 2,84E-16 | 2,18E-14 | ABCC8,ACACB,ADA,ARAF,ARSA,ATF3,AURKA,AZGP1,BACH2,BCL6,BID,BLNK,CA2,CASP3,CASP6,CCNA2,CCNB1,CCNB2,CCND2,CCND3,CCS,CD2,CD55,CDC25B,CDH5,CDK1,CDKN2C,CDKN3,CEP55,CKAP4,CKM,COL1A1,CRELD2,CSF1R,CTSV,DCK,DDX58,DSTN,EAF2,ECT2,FGFR3,FKBP11,FOXO1,GADD45A,GADD45B,GIMAP4,GM2A,GYPA,HADHA,HMGB2,HS3ST1,HSD17B14,HSP90B1,ID2,ID3,IFI16,IGHM,IL7R,JUND,KLHDC2,MAD2L1,MAL,MANF,MEF2C,MORF4L2,MSMO1,MZB1,NDC80,NDN,NEIL1,NELL1,NFIL3,NPPA,NR0B2,NR4A3,NTRK2,OPTN,PAFAH1B3,PARP8,PDIA4,PDIA6,PFKP,PLAC8,POU2AF1,PRC1,PRDX2,PRDX4,PRSS3,PTDSS1,PTGS1,PTPRCAP,RIPOR2,RORA,RORC,RRM2,SCN4A,SDF2L1,SEC24D,SELL,SERPINB1,SLC12A2,SLC35B1,SMAGP,SUB1,SULT2B1,TAPBPL,TMEM184B,TOP2A,TOX,TRAF5,TTK,XBP1,ZP2 |
| ERK1/2 | | group | | 0,537 | 0,595 | 0,006 | 3,06E-16 | 2,33E-14 | ABCC6,ALOX5,AOC1,ATP1A3,AVP,B2M,BAX,BCL2A1,BHMT,BIRC2,BIRC3,C3,CALR,CAT,CAV1,CCL20,CCL5,CCL8,CCND1,CCND2,CCR7,CD40,CD44,CD86,CDK1,CDKN1B,COL1A1,COL3A1,CTSB,CTSV,CXCL1,CXCL10,CXCL12,CXCL6,CXCR4,CYBB,CYP17A1,CYP46A1,DDIT3,DUSP1,EFNB2,ENO1,ESR1,F3,FKBP5,FN1,FOS,FOSB,FOSL1,FOXO1,GAD1,GJA1,GRK5,HIF1A,HLA-A,HPGD,HSPA5,ICAM1,ID1,ID3,ITGA2,ITGA3,ITGB3,JUND,KIF2A,LEPR,MAP3K10,MAPT,MMP1,MMP2,MMP9,NAMPT,NKX2-5,NOS2,NOX4,NPPA,NR4A3,PCNA,PDGFRA,PKM,POSTN,PRKCZ,PSMB10,PSMB6,PSMB8,PSMB9,PTGES,PTPRE,RAC1,RB1,S100A6,SGK1,SLC12A6,SOD1,SPP1,TAC1,TAP1,TAP2,TAPBP,TBP,TERT,TIMP1,TIMP2,VCAM1,VCAN,VEGFA |
| LDL | | complex | Activated | 2,353 | 2,397 | 0,005 | 4,08E-16 | 3,05E-14 | ABCG1,AKAP1,APOBEC3B,APOE,ARG2,ATF3,AXL,BCL2A1,C3,CAPN1,CAPN2,CASP1,CASP3,CASP6,CASP8,CCL20,CCL5,CCL8,CCND1,CCR2,CCR7,CD36,CD40,CDH5,CDK1,CTSK,CX3CR1,CXCL10,CXCL11,CXCL6,CYBB,CYP2C8,CYP2J2,DDIT3,DNMT1,DUSP1,F3,FAS,FCGR2B,FOS,FOSB,G0S2,GCLC,GCLM,GNAI1,GPR183,HIF1A,HSP90B1,HSPA5,ICAM1,IDO1,IFIT2,IFITM1,IGFBP4,IL10RA,IRF2,IRF7,ITGAV,ITGB3,MMP1,MMP2,MMP9,MX1,NAMPT,NFKB1,NME1,NOS2,NR0B2,NR1H3,NR4A1,NR4A3,PANX1,PARP1,PECAM1,PLIN2,PTEN,PTPRF,RCAN1,RND3,S100A8,SELP,SHC1,SIGLEC1,SOD2,STAT3,TGFBR2,TGFBR3,TLR4,TNFAIP6,TNFSF10,TP53,VCAM1,VEGFA,XBP1 |
| CEBPA | 0,150 | transcription regulator | | -0,742 | -0,707 | 0,003 | 4,55E-16 | 3,37E-14 | A2M,ACACA,ACSL1,ADCY7,ADH1B,ADH6,AGPAT2,AKAP12,AKR1B1,ALB,ALOX5AP,ANXA1,APOA4,APOC3,ARG2,ARL4C,ARL6IP5,ASL,ASNS,ASS1,BCL2A1,BHMT,BIN1,C3,C3AR1,CA2,CCL20,CCNA2,CCNB2,CCND1,CCND2,CD14,CD3G,CEBPE,CFD,CKAP4,COL1A1,COL1A2,CRADD,CSF1R,CTSK,CXCR4,CYP2A6 (includes others),CYP2B6,CYP3A4,CYP3A7,DHCR7,DYNLT3,EFNB2,EPHX1,EVPL,FHL1,FOS,FOXO1,FTO,G0S2,G6PC,GADD45A,GBP1,GCH1,GGH,GJA1,GLIPR1,GLRX,GSTP1,H1-10,HDAC1,HLA-B,HNF1A,HNF1B,HPGD,HSPA5,ICAM1,ICAM2,ID1,ID2,IFI27,IFI6,IGFBP4,IL6R,INSR,ISG15,ITGAL,ITGAM,KRT7,LCK,LCN2,LEPR,LGALS1,LITAF,LST1,LTF,MALT1,MNDA,NFIL3,NRP1,OAS2,OVOL1,PCK1,PCNA,PLIN2,PLOD2,PLXND1,PPARA,PPARGC1A,PPL,PPP1R3C,PRLR,PTGS1,PTPRC,PTPRE,RAB31,RIN1,RORA,RUNX3,S100A8,S100A9,SCD,SECTM1,SERPINB1,SERPINF1,SMPDL3A,SOD1,SOD2,SPINT2,SPP1,STEAP4,TAC1,TBXAS1,THRB,TIAM1,TNFAIP6,TNFSF10,TUBB2A,UBE2I,VCAN,VDR,VLDLR,ZBTB48 |

| | | | | | | | | | |
|---|---|---|---|---|---|---|---|---|---|
| E2F1 | | transcription regulator | | 0,868 | 0,920 | 0,006 | 4,67E-16 | 3,43E-14 | ABCG2,ACADL,ACSL5,ACTR3,ALG5,ALPL,AURKA,BAX,BID,BNIP3,CA2,CALM1 (includes others),CALR,CASP3,CASP6,CASP8,CAV1,CCNA2,CCNB1,CCNB2,CCND1,CCND2,CCND3,CCT2,CD2AP,CD44,CD9,CDC25B,CDK1,CDKN1B,CDKN1C,CDKN2C,CLIC1,CRADD,CRIP2,CSE1L,CSHL1,CTNNBIP1,CTSB,CYB5A,CYP27B1,DDB2,DDIT3,DDX1,DLEU1,DNMT1,DUSP1,ECT2,EED,ERH,EYA2,EZH2,FAS,FGFR3,FHIT,FHL1,FKBP3,FOS,FOXO1,GINS1,GM2A,GPS2,GRAMD4,HADHA,HMGB2,HNRNPR,HSD11B2,HSP90B1,HSPA8,ICAM1,ID3,JPT1,KRAS,KRT10,LCK,MAD2L1,MAF,MCM2,MCM3,MCM5,MCM6,MCM7,MFAP1,MMP1,MSH2,MT1G,MTHFD1,MYB,NCL,NDC80,NDUFC1,NMI,NRP1,OXT,PCLAF,PCNA,PDCD5,PDK4,PHB,PLK2,PMPCB,PPARGC1A,PPP1R26,PPP1R8,PPT2,PSAT1,PSMB9,RB1,RBFOX2,REV3L,RFC2,RND3,RPA2,RPA3,RPS16,RRM1,RRM2,SERPINF2,SLC22A18AS,SLC3A2,SMARCA5,SMC4,SOD2,STAM,STK17B,STMN1,TAP1,TAP2,TAPBP,TERT,TOP2A,TOP2B,TOPBP1,TP53,TP53BP2,TRAP1,UCP2,ULK1,UQCR11,VCAM1,VEGFA,VIM,VRK1,WWOX,XPC,YBX3,YWHAE,ZFP36,ZNF22 |
| IRF7 | 0,848 | transcription regulator | Activated | 7,741 | 7,822 | 0,010 | 5,47E-16 | 3,99E-14 | ADAR,APOBEC3G,BCL2L13,CASP4,CCL19,CCL5,CCL8,CD40,CD69,CXCL10,CXCL9,DDX58,GBP1,IDO1,IFI16,IFI35,IFI44,IFI44L,IFI6,IFIH1,IFIT2,IFIT3,IFITM1,IFITM2,IFNA10,IL15,IL15RA,IL33,IRF1,IRF7,IRF8,IRF9,ISG15,ISG20,ITGAM,MICB,MX1,MX2,NAMPT,NMI,OAS1,OAS2,OAS3,PARP12,PHF11,PLAC8,PLSCR1,PSMB10,PSMB8,PSMB9,PSME1,RSAD2,RTP4,S100A8,STAT1,TAP1,TAP2,TDRD7,TLR4,TLR8,TMBIM6,TNFAIP8,TNFSF10,TOR1B,TPST1,TRIM21,TRIM22,TRIM5,UBA7,UBE2L6,XAF1 |
| CSF2 | | cytokine | Activated | 4,376 | 4,459 | 0,008 | 7,62E-16 | 5,51E-14 | ABCG1,ACE,ACP5,ADA,ADAM17,ADGRE5,AKR1A1,ALOX5,ALOX5AP,ANXA1,AOC1,AURKA,BAX,BCL2A1,BID,BIRC3,C3,CASP1,CASP3,CCL18,CCNA2,CCNB1,CCND1,CCND2,CCND3,CCR5,CCR7,CD14,CD163,CD180,CD1C,CD24,CD38,CD40,CD63,CD69,CD74,CD83,CD86,CD8A,CDC123,CDK1,CDKN1B,CDKN2C,CFH,CISH,CLEC7A,CSF1R,CSF2RB,CXCL1,CXCL10,CXCR4,CYBB,DDIT3,F2RL1,F3,FANCL,FAS,FCGR2B,FOLR2,FOS,FOSL1,GADD45GIP1,GCH1,GCLM,GK,HIF1A,HLA-DQB1,ICAM1,ID2,ID3,IDO1,IK,IL13,IL15,IL1R1,IL32,INPP5D,IRAK1,IRF2,ITGAM,ITGB3,LAMP3,LCP1,LTB,LY75,LY96,MAT2A,MCM3,MCM5,MCM6,MET,MMD,MME,MMP1,MMP2,MMP9,MNS1,MRC1,MYB,NET1,NFKB1,NOS2,NR1H3,NR4A1,NRP1,OSMR,PHGDH,PLAGL2,PLD1,PMP22,PPP1R15A,PRC1,PRNP,PROZ,QSOX1,RB1,RHOH,RIPOR2,RRM2,SERPINB9,SGK1,SLC1A5,SOCS2,SOD2,SPAG5,SPP1,STMN1,TCF4,TERT,TLR1,TLR2,TLR4,TLR5,TNFRSF10D,TNFRSF1B,TOP2A,TP53,TPI1,TPX2,UBD,UBE2C,VDR,VTCN1,XBP1,ZFP36 |
| ethanol | | chemical - endogenous mammalian | | 1,379 | 1,408 | 0,003 | 8,66E-16 | 6,20E-14 | ABCG1,ACACA,ACADL,ACADM,ACKR3,ACOX1,ADAM17,ADIPOR1,ADM,ALDH1A3,ALOX5,APOA1,ATF3,AVP,BAX,BHMT,C3AR1,CAPN1,CASP3,CAT,CCL5,CCND1,CD14,CD36,CDKN1B,CHUK,CLIC4,CLU,COL1A2,COL3A1,CXCR4,CYP2A6 (includes others),CYP2B6,CYP2C8,CYP3A4,CYP3A7,CYP4A11,DDIT3,DUSP1,EPHX1,ESR1,ETNPPL,FAAH,FABP1,FABP2,FAS,FOS,FOSB,G6PC,GADD45A,GNAI1,GNAI2,GNAS,GPT,GRK2,GSR,GZMB,HAO2,HIF1A,HLA-A,HOXA1,HSBP1,HSD17B10,HSP90B1,HSPA5,HTATIP2,ICAM1,ID1,IGHM,IL15,IL1R1,IRF7,ITGAM,LGALS1,LPIN1,MAPK8,MAT2A,MLXIPL,MME,MMP2,MMP9,MYD88,NCF2,NMI,NOS2,NPPA,NR1I3,NSDHL,NTRK2,PARP1,PCK1,PCNA,PHB,PNPLA2,PPARA,PPARGC1A,PPP1R3C,PRF1,PTEN,RBP1,RHOB,RORA,SCD,SGK1,SLC2A2,SOD1,SOD2,STAT1,STAT3,TBP,TFF3,TFPI2,TIMP1,TIMP2,TLR1,TLR2,TLR4,TLR7,TLR8,TNFAIP6,TP53,TRIB3,VAMP2,VCAM1 |
| VEGFA | -0,804 | growth factor | | -0,094 | -0,021 | 0,007 | 9,23E-16 | 6,56E-14 | ABCC1,ACADM,ACE,ACSL1,ADAMTS1,ADH5,ALDH2,ARG2,ATP5MC2,BAX,BCL2A1,CASP3,CAV1,CBFB,CCND1,CD36,CD40,CD55,CD83,CD86,CDH5,CDKN1B,CLIC4,COL1A1,COX5B,COX8A,CSTB,CTSB,CTSS,CTSV,CXCL10,CXCL12,CXCL6,CXCR4,DECR1,EFNB2,ERAP1,ESM1,ETFA,F3,FN1,FOS,FOSB,GBP1,GJA1,GRB10,HIF1A,HK1,HLA-DQB1,HSPA5,ICAM1,ID1,ID3,IGFBP4,INPP5D,ITGA2,ITGAV,ITGB3,MDH2,ME2,MEF2C,MMP1,MMP2,MMP9,MRPS12,NME1,NOS2,NPHS1,NR2F2,NR4A1,NRP1,PARP1,PCNA,PECAM1,PNPLA2,PPARGC1A,PRKCA,PTGS1,RCAN1,SDHB,SDHD,SET,SHMT2,SLC12A6,SLC25A5,SMTN,SNCG,SOD2,SPARC,ST5,STARD8,TERT,TFPI2,TIMP1,TLR1,TMSB10/TMSB4X,TNC,TP53,TSPO,TUFM,TXN2,VASH1,VCAM1,VEGFA |

| | | | | | | | | | |
|---|---|---|---|---|---|---|---|---|---|
| ACOX1 | -0,586 | enzyme | Inhibited | -2,377 | -2,391 | -0,002 | 1,19E-15 | 8,37E-14 | ACAA1,ACADL,ACADM,ACADVL,ACOX1,ACSL1,ADAMTS1,ANXA2,BAAT,CAT,CD36,CD63,COL4A2,CRAT,CSTB,CTSV,CXCL12,CYP2C8,CYP4A11,CYP4F12,DDIT3,DIO1,DPP4,EHHADH,FABP2,FAH,G6PC,GADD45A,GNMT,GOT1,GSTP1,HDAC1,HLA-DQA1,HPD,HSPA5,LCN2,LGALS3BP,LHPP,LONP1,NFE2L2,NR1H3,OAT,PAH,PDK4,PIGP,PIPOX,PLIN2,PLSCR1,PXMP2,QPCT,RGN,RPS24,SELENBP1,SGK1,SLC27A2,SLC37A4,SLC4A4,TAGLN2,TP53,TRIB3,UCP2,UGCG,XBP1 |
| TRIM24 | 0,331 | transcription regulator | Inhibited | -4,784 | -4,845 | -0,009 | 1,39E-15 | 9,72E-14 | AGRN,BLNK,CA2,CALB1,CNOT6,CXCL10,DDX58,DDX60,FAM110B,FBN1,FLRT3,GBP2,GLUL,HERC6,HNF1B,IFI35,IFI44,IFIH1,IFIT2,IFIT3,IFNGR1,IFNGR2,IRF1,IRF7,IRF9,ISG15,LGALS3BP,NMI,OAS1,PARP12,PCLAF,PHF11,PLAC8,PRKCQ,PSMB10,PSMB8,PSMB9,RTP4,SAMHD1,SOCS2,SPP1,STAT1,TAP1,TGFBI,TLR2,TP53,TRIM26,TRPV5,UBA7,VEGFA |
| JUN | -0,404 | transcription regulator | | -0,488 | -0,438 | 0,005 | 1,67E-15 | 1,15E-13 | A2M,ACP5,AFP,AKAP12,ANXA1,ANXA2,APOC3,APOE,APOM,ASNS,ATF3,AXL,BAX,BCL2A1,BIRC2,BIRC3,C3AR1,CALM1 (includes others),CAPN2,CAV1,CCL20,CCL8,CCN3,CCNA2,CCND1,CCND2,CCND3,CD14,CD164,CD247,CD44,CDK1,CDKN1B,CGA,CLU,COL1A1,COL1A2,COMT,CSF1R,CSTA,CXCL1,CXCL10,CYP1B1,CYP2J2,DICER1,DNMT1,DROSHA,DUSP1,EHD4,EPHX2,EVX1,F3,FAS,FN1,FOLR2,FOS,FOSL1,FRZB,FUS,GADD45A,GCLC,GJA1,GOT1,GPT,GSTP1,HLA-B,HLA-G,HPGD,ICAM1,ID2,IFI16,IGFBP4,IGFBP6,IGFBP7,IL7R,ITGAM,ITGAV,ITGB8,LHB,LPAR1,MAF,MAOB,MAT2A,MCF2L,MET,MMP1,MMP2,MMP7,MMP9,MSH2,MXI1,MYB,NCF2,NGF,NKX2-5,NOS2,NOX5,NPPA,NR0B2,NR4A1,PDCD4,PEA15,PLA2G4A,PPM1D,PTEN,PTGES,PTPN6,PTPRD,PXDN,RARG,RGN,RHOB,RXRA,S100A10,SCD,SDC1,SERPINB9,SGK1,SLC10A2,SMAD7,SNCG,SNRNP70,SOD2,SPARC,SPP1,SST,STAT1,STMN1,SYK,TIMP1,TNC,TNNC1,TP53,TXN,VCAM1,VDR,VEGFA,VIM,ZEB2,ZFP36 |
| NRAS | 0,127 | enzyme | | -1,649 | -1,676 | -0,004 | 1,70E-15 | 1,17E-13 | ACADM,AGRN,AKAP12,ANXA4,B2M,BAX,CALCRL,CCL5,CCNB1,CCNB2,CCND1,CCNG1,CD48,CD86,CDKN3,CFH,CKM,COL1A2,CX3CR1,CYBB,EIF2D,ENPEP,EPHX1,FHL1,FN1,GALE,GBP2,GJA1,HIF1A,HLA-A,HOMER3,HTRA1,IFI16,IFI35,IFIH1,IGHM,IL13RA1,ISG15,ITGA2,JCHAIN,KCTD12,KIF22,KRAS,LCN2,LGALS1,LSP1,LY86,MCM3,MGP,NCF2,NPPA,NPR3,OIP5,OSBPL1A,P3H4,PERP,PHLDA3,PLEKHO1,PLTP,PRDX2,PSMB8,RBM38,RBPMS,RHOB,RNASE6,SARDH,SDC1,SERPINE2,STAT1,STAT3,TAP1,TMEM43,TUBB2A,UBE2C,VEGFA,XPNPEP2 |
| PAX3 | | transcription regulator | | 0,905 | 0,917 | 0,004 | 1,80E-15 | 1,23E-13 | ALDH3A2,ANOS1,ANXA1,AP3M2,ARC,ARHGDIB,ASNS,ASS1,ATP2B4,BST2,CAPN1,CD151,CDH3,CLU,COL1A2,CPN2,CXCR4,CYP1B1,DBN1,DPEP1,EYA2,F2RL1,F3,FHL1,FUBP1,G0S2,GLB1,GLDC,GPT,GUCY1A1,HLA-DRB1,HNRNPA3,HTR6,ICAM2,ID2,ID3,IFITM2,IL6R,IL7R,MCAM,MET,MGP,MICB,MMP9,NEDD9,NNMT,NUDT1,PAK2,PCNA,PEA15,PHGDH,PPIB,PSME1,PTGER3,PTP4A1,PTPN12,PXDN,RAB31,RASSF2,RHOB,RUNX3,SFN,SHMT1,SLA,SOX4,SRGN,TCN2,TFPI2,TIA1,TNC,TNNI1,TP53,TSC22D1,TSPAN7,TYRO3,VCAN,YWHAE |
| CNOT7 | 0,001 | transcription regulator | Inhibited | -2,136 | -2,157 | -0,007 | 2,19E-15 | 1,48E-13 | B2M,BST2,CLDN1,CSTA,HERC6,IFI27,IFI35,IFI44L,IFI6,IFIT5,IFITM1,ISG15,LGALS3BP,OAS1,OAS2,OAS3,PARP12,PLSCR1,PSMB8,SP110,STAT1,TAP1,TAP2,TFF3,UBE2L6 |
| CREBBP | | transcription regulator | | -1,019 | -0,978 | 0,007 | 2,55E-15 | 1,70E-13 | AKAP12,AP3M2,ASL,B2M,BANK1,BCL6,BMP7,CA2,CAMK2G,CAPG,CCL5,CCND1,CCND2,CCND3,CCR5,CCR7,CD44,CD53,CD59,CDH5,CDKN1B,CGA,CKM,COL1A2,CST7,CXCL10,CXCL12,CXCL9,CXCR4,CYBB,DDC,DHRS3,DUSP1,DUSP22,EHHADH,EPN1,EVI2A,EVL,EYA2,FDX1,FGFR3,FNBP1,FNDC3B,FOS,FOSB,G6PC,GIMAP4,GM2A,GNAS,GPR18,HBB,HLA-B,HLA-DRA,HLA-G,HMGCS2,HSD3B2,IL10RA,IL2RB,IL7R,INHA,IRF1,ISG15,JUND,KMO,LGALS3BP,MAF,MCF2L,MCM3,MEF2C,MFHAS1,MGA,MMP9,MSH2,MYO1F,NDRG1,NFE2L2,NOS2,NPPA,NR4A1,NR4A3,OAS3,PCK1,PCNA,PDE10A,PDLIM4,PHIP,PIPOX,PKLR,PLEKHF1,PPARGC1A,PRF1,PRKCD,PSMB9,PTPRF,RB1,RHOA,RNASEL,RSAD2,RTP4,SDC1,SELP,SERPINA3,SERPINE2,SH2D1A,SPSB1,SST,ST6GAL1,STAT4,TAP2,TAPBP,TESC,TIPARP,TLE2,TLR2,TMPRSS2,TMPRSS4,TNC,TNFSF10,TRAT1,TRIM26,UTRN,VCAM1,VEGFA,WNT8B |

| | | | | | | | | | |
|---|---|---|---|---|---|---|---|---|---|
| PPARGC1A | -0,374 | transcription regulator | Inhibited | -6,098 | -6,076 | 0,002 | 2,56E-15 | 1,70E-13 | ACACA,ACACB,ACADL,ACADM,ACADVL,ACAT1,ACOX1,ADAMTS1,ADGRG1,ANG,APOA4,APOC3,BAX,BNIP3,C2,C3,CALM1 (includes others),CAT,CD36,CD40,CIDEB,COL1A2,COL6A3,COX5B,COX7B,CXCL10,CXCL14,CYP2A6 (includes others),CYP2B6,CYP3A4,DCN,DGAT1,DIO1,DLAT,ENDOG,ESRRA,G6PC,GK,GOT2,HMGCL,HMGCS2,HSD3B2,IGFBP6,IGFBP7,IL18,IL4R,INHA,INPP5J,ITGB1BP1,KYAT1,LAMB1,LIPA,LPIN1,LUM,MAL,MAOB,MCM3,MCM5,MCM6,MDH2,ME1,MMP2,MSMO1,NDUFS1,NDUFS8,NPPA,NR1H3,NR1I3,NSDHL,PCK1,PCK2,PCSK5,PDIA5,PDK2,PDK4,PHYH,PLA2G7,PLA2R1,PLIN2,PNPLA2,POSTN,PPARA,PPARGC1A,PRDX3,PVALB,RXRA,SCD,SDHB,SERPINA3,SERPINE2,SIRT3,SLC25A20,SLC39A14,SMAD7,SOD1,SOD2,TGFBR3,THY1,TIMP1,TIMP2,TNFRSF11B,TNFSF10,TNNI1,TP53,TRIB3,TXN2,TXNRD2,UCP2,UQCRC2,VAMP2,VCAN,VEGFA |
| INSR | -0,426 | kinase | Inhibited | -5,966 | -5,906 | 0,008 | 3,58E-15 | 2,29E-13 | ACAA2,ACADL,ACADM,ACADS,ACADVL,ACO2,ACOT13,ACSL1,ACSL5,ALAD,ALB,ALDH2,ALDH6A1,AQP1,ATF3,ATP5F1D,BAG1,BCL2A1,C1QA,C2,CASP6,CCND1,CCND2,CD36,CD83,CDK1,CDKN2C,COL6A3,CORO1A,COX6A2,CPT2,CRAT,CSTB,CTSB,CTSV,CXCR4,CYC1,CYP1B1,DECR1,DEK,DHCR7,DUSP1,ECHS1,ECI1,ETFA,ETFB,ETFDH,FBP1,FH,FHL1,FOS,GADD45A,GCH1,GNB1,GOT2,GRB10,GRN,HADHA,HSD17B10,IDH1,IDI1,IFITM2,IFNAR2,INSR,IQGAP1,KLF9,LCP1,LEPR,LGALS1,LRP10,LYZ,MAOA,MAOB,MARCKSL1,MCM2,MCM5,MCM7,MDH2,MMP2,MMP9,MSH6,MSMO1,MXI1,MYOM2,NAMPT,NDN,NDRG1,NDUFA3,NFE2L2,NGF,NPPA,NR1H3,NSDHL,OGDH,PDCD4,PDK4,PECR,PKLR,PMS1,PPARA,PPARGC1A,PRIM1,PRKCQ,PRKD1,RHOB,RPS16,RRM1,RRM2,SC5D,SCD,SCP2,SEZ6L2,SIRT2,SKAP2,SLC25A20,SLC29A1,SOCS2,SUCLA2,THBS2,TNNI1,TSC22D1,UCP2,UQCRC1,UQCRC2,VCAM1 |
| hydrogen peroxide | | chemical - endogenous mammalian | | 1,004 | 1,066 | 0,005 | 6,27E-15 | 3,93E-13 | ACE,ADAM17,ADAM9,AEBP1,ALOX5,ANXA2,ATF3,AURKA,BAX,BCL2A1,BIRC3,CASP1,CASP3,CAT,CAV1,CCNA2,CCNB1,CCND1,CCND3,CCNH,CCR2,CCR5,CD247,CD55,CD59,CD63,CD81,CD86,CDK1,CDKN1B,CDKN2C,CFD,CLU,COL1A1,COL1A2,CTSB,CTSD,CTSS,CXCL6,CXCR4,DDIT3,DHCR7,DNAJB1,DNMT1,DPAGT1,DUSP1,ENC1,ENG,ENPP2,EZH2,F2RL1,FAS,FN1,FOS,FOSL1,FOXO4,FZD2,GADD45A,GADD45B,GLRX,GRK2,GSTA1,GZMB,HIF1A,HLA-DRA,HMGCS2,HSPA5,ICAM1,ID3,IER5,IFI6,IFIT2,IL10RB,IL13,IL32,ITGA2,ITGA3,ITGB3,KITLG,KRAS,LCN1,LCN2,MAPK8,MAPKAP1,MCAM,MCM2,MDK,MIF,MMP1,MMP2,MMP9,MSMO1,MUC1,NAT10,NDC80,NFE2L2,NFIL3,NGF,NOS2,NOX5,NR4A1,OSGIN1,PAH,PARP1,PCBP2,PDCD10,PDCD4,PDGFRA,PDIA3,PDIA4,PPARGC1A,PPM1D,PRDX2,PRKCI,PSTPIP2,PTEN,PTP4A2,RAB27A,RAC1,RIOK3,RNF19B,RTN3,RUNX3,SC5D,SGK1,SHC1,SOD1,SOD2,SPARC,SPP1,SRGN,STAT3,STMN1,TAP1,TGFBR3,TLR4,TP53,TRIB3,TUBB2A,TXN,TXNIP,UCP2,VCAM1,VDR,VEGFA,VIM,WNK1,XAF1,XBP1 |
| Akt | | group | Activated | 2,134 | 2,165 | 0,003 | 7,02E-15 | 4,37E-13 | A2M,ADM,AQP3,B4GALT5,BAX,BCL2A1,BCL6,BHLHE40,BID,BIRC2,CCND1,CCND2,CCND3,CCR7,CD36,CD44,CDKN1B,CDKN1C,COL3A1,CTSD,CXCL10,CXCL12,CXCR4,ERBB3,ESR1,EZH2,F3,FAS,FHIT,FKBP5,FN1,FOS,FOSL1,FOXO1,GSR,GSS,GSTK1,HIF1A,HPGD,IL13,IL1R1,IL2RB,IL32,IL7R,INSR,KL,LCN2,MCAM,MMP2,MMP9,MT1X,MUC1,MYB,NDRG1,NDUFV1,NET1,NFE2L2,NFIL3,NFKB1,NOS2,NOTCH2,NOX4,NPPA,NR0B2,OSTF1,PCK1,PCNA,PDK4,PEA15,PLA2G4A,PLG,PPARGC1A,PTGS1,RB1,RHOB,RRM2,SDHB,SGK1,SOD2,SOX4,SPP1,TERT,THBS2,TIPARP,TJP1,TLR2,TMED10,TNC,TNFSF10,TP53,UQCRC2,UTRN,VCAM1,VCAN,VEGFA,WT1,WTAP,XBP1 |
| TP73 | -0,171 | transcription regulator | | -0,447 | -0,416 | 0,003 | 7,51E-15 | 4,63E-13 | ABCC1,ADA,ADAM10,ADAM17,AFP,ATF3,ATG4A,AUH,AURKA,B2M,BAX,BHLHE40,BID,BIRC2,BMP7,C1R,C3,CASP1,CASP3,CASP4,CCNA2,CCNB1,CCND3,CCNG1,CD55,CD83,CDC25B,CDK1,CDKN1B,CDKN1C,CDKN3,CLEC7A,CLMN,COL18A1,COL1A1,COL5A2,CTH,CTSD,CXCL1,CXCL10,DDIT3,DHRS3,DIMT1,DRAM1,DTYMK,ENG,ERCC2,FAS,FGFR3,FN1,G0S2,GAB2,GADD45A,GPNMB,GPR137B,GRK5,GRN,HAGH,HIF1A,HPSE,HSPB8,HTATIP2,ID2,IGFBP4,IGSF3,IL4R,IL7,KIF22,KRT10,LCP1,LYZ,MCM6,MCM7,MDK,MIR22HG,MMP2,MPZL2,MTHFD1,NAP1L3,NFKB1,NRP1,PERP,PIK3R3,PLK2,PMP22,PPL,PRKAG2,PROCR,PTEN,PTGES,RB1,RBX1,S100A2,SERPINA3,SERPINA5,SERPINE2,SERPINF1,SERPING1,SFN,SMUG1,SPHK2,SPP1,STMN1,TAP1,TERT,THBS2,THY1,TIMP1,TNFAIP2,TNFRSF1B,TNNI1,TP53,TPX2,TREM2,TRIM22,TRIM32,TSPAN1,UBE2C,UBE2D1,UBL3,ULK1,VDR,VEGFA,VEGFC,YWHAB,ZMAT3 |

| | | | | | | | | | |
|---|---|---|---|---|---|---|---|---|---|
| TCF4 | 0,649 | transcription regulator | | 0,562 | 0,589 | 0,003 | 8,96E-15 | 5,45E-13 | ADA,ADAM17,APOBEC3G,ARHGEF6,BACH2,BARD1,BCL2L13,BCL6,BHLHE40,BMP7,BNIP3,CARD8,CASP1,CASP3,CASP4,CASP8,CCNA2,CCNB2,CCND1,CCT6A,CD2,CD24,CD38,CD40,CD55,CD9,CDC25B,CDK1,CDKN1C,CDKN3,CEP55,CKAP4,CRELD2,DCK,DDX58,DIMT1,DNAJC12,DSTN,EAF2,ELF1,ELF4,FAM111A,FAS,FGF1,FKBP11,FKBP8,FN1,FOS,FOXO1,GEMIN6,GIMAP4,GRPEL1,HIF1A,HMGB2,HSP90B1,HTR2B,ID2,ID3,IDO1,IFI16,IGFBP4,IL15,MAD2L1,MANF,MEF2C,MMP1,MORF4L2,MYB,MZB1,NDC80,NEDD9,NEIL1,NELL1,NOS2,NOTCH2,OPTN,OSBPL1A,PARP8,PDIA4,PDIA6,PIK3C2B,PLAC8,PLK2,PPID,PPIH,PRC1,PRDX4,PSD3,RIPOR2,RPL35A,RRM2,SACS,SDF2L1,SEC24D,SELL,SERPINB1,SERPINF1,SGK1,SH3GLB1,SLC35B1,SLC9A3R1,SMAD1,SMAD2,SMAD7,SMAGP,SORBS2,SP140L,SPP1,STAT3,SUB1,TAPBPL,TERT,TIA1,TMEM140,TMEM184B,TNFSF10,TRAF5,TRIM38,TXNIP,VCAN,WARS1,XBP1,ZEB2,ZP2 |
| IGF1 | 0,033 | growth factor | | 0,513 | 0,566 | 0,005 | 9,10E-15 | 5,50E-13 | A2M,ACACA,ACTR1B,ADA,ADM,AKR1B1,BAX,BCL2A1,BHLHE40,BIRC3,C1QA,CASP3,CCL5,CCNA2,CCNB1,CCND1,CCND2,CCND3,CCNG1,CD44,CDK1,CDKN1B,CISH,CLU,COL1A1,COL3A1,CSHL1,CTSD,CXCL6,CYBA,CYBB,CYFIP2,CYP17A1,CYP27B1,DCLK1,DDIT3,DDX50,DUSP1,EFNB2,EGF,ESR1,F3,FN1,FOS,FOSB,FOXO1,GADD45A,GADD45G,GHR,GPD1,GPER1,HBB,HIF1A,HSD11B2,HSD17B11,HSPA5,ICAM1,ID1,ID2,IFNGR2,IGFBP4,IGFBP6,IHH,IL18,IL1R1,IL4R,INHA,INSR,ITGA3,ITGAV,LCN2,LEPR,MAPT,MGA,MIF,MMP1,MMP2,MXI1,MYB,NCF2,NFE2L2,NOS2,NOX4,NPPA,NR4A1,NTRK2,OSMR,OXT,PARP1,PBK,PCK2,PCNA,PCOLCE,PDGFRA,PDLIM2,PITX3,PKM,PLK2,PRLR,PRNP,PSMB8,PSMB9,PTEN,PTH1R,PTPRC,SCD,SFN,SGK1,SHBG,SIRT2,SLC12A6,SLC2A8,SLPI,SNCG,SPP1,SST,TAP1,TAP2,TBX2,TG,TGFBI,THY1,TJP1,TLR4,TNFRSF11B,TP53,TRPV1,TTF2,TXNIP,UBE2B,UBE2D2,UGCG,VCAM1,VEGFA,VEGFC,VIM,WDR45,WT1,WTAP,XBP1,YBX3,ZFP36 |
| AHR | 0,505 | ligand-dependent nuclear receptor | | -0,817 | -0,807 | 0,001 | 9,99E-15 | 5,99E-13 | A2M,ABCG2,ACACA,ACOX1,ACTN1,ADM,AKR1A1,ALB,ALDH1B1,ALDH2,ALDH3A1,ALOX5,ARG2,BAX,BZW2,CAMK2N1,CASP8,CBR3,CCL21,CCL5,CCNA2,CCNB1,CCND1,CCND3,CD14,CD36,CD38,CDC25B,CDK1,CDKN1B,CDKN2C,COL16A1,COL1A1,COL1A2,COL3A1,COL4A1,COL4A2,COL5A1,COL5A2,COL6A3,COX8A,CTSD,CXCL10,CYP17A1,CYP1B1,CYP2A6 (includes others),CYP2B6,CYP2C8,CYP3A4,DCN,DIO1,DNAJB1,EFEMP1,EHD1,ENO1,ENPEP,ESR1,ESRRG,FAS,FN1,FOS,FOSL1,GADD45A,GHITM,GOT1,GSTA1,HIF1A,HSPA5,IDO1,IGHM,IRF1,ITGA7,ITGAM,LTBP3,MMP2,NEDD9,NFE2L2,NFYB,NID2,NMT2,NNMT,NOS2,OPA1,PCK1,PDGFRA,PJA1,PKLR,PLA2G12A,PMM1,PPP1R15A,RBP1,RORC,SCD,SDC1,SLC16A5,SLC1A1,SLC22A7,SNTA1,SOD1,SOD2,SPP1,STAT3,STEAP4,TDO2,TFF3,TGFBI,TGFBR2,TIMP2,TIPARP,TP53,TSPYL2,TXN,VEGFA,VEGFC,VIM |
| SPI1 | | transcription regulator | Activated | 3,749 | 3,780 | 0,004 | 1,10E-14 | 6,54E-13 | ACTB,BAX,BCL11A,BCL6,BCL7A,BLNK,C3AR1,CCNA2,CCNB1,CCNB2,CCND1,CCND2,CCR7,CD14,CD180,CD1D,CDK1,CEBPE,CSF1R,CSF2RB,CTSK,CTSS,CXCL10,CYBB,DAB2,EPB41L2,FCER1A,FCER1G,FCGR2B,FCGRT,FLI1,FOS,FOXO1,HSPA8,ID2,ID3,IFI27,IFI44,IFI44L,IFI6,IFIT2,IFIT3,IFITM1,IL13,IL18,IL7R,IRF7,IRF9,ISG15,ISG20,ITGAM,ITGB2,KLK1,LAMP3,LBR,LIMD1,LTF,LYZ,MEF2C,MME,MMP1,MMP2,MRC1,MS4A1,MT1F,MT1G,MT1X,MX1,MYB,NCF2,NCF4,NKX2-5,NOS2,PARP12,PCNA,PSMB10,PSMB8,PSMB9,PTEN,PTPN6,PTPRC,RACK1,RSAD2,RUNX3,SP110,SPP1,TFEC,TLR2,TLR4,TNFSF10,TRIM22,VAV1,VIM |
| CEBPB | 0,081 | transcription regulator | | 1,344 | 1,409 | 0,006 | 1,27E-14 | 7,53E-13 | ADM,ALB,ALOX5AP,AOC1,APOC3,ASNS,ATR,BCL2A1,BHMT,BLNK,C3,CCL5,CCNA2,CCNB2,CCND1,CCND2,CCR5,CD14,CD1D,CDC25B,CDH3,CDKN1B,CFD,CIRBP,CLPP,COL1A1,COL1A2,COL5A2,CSF1R,CXCR4,CYB5A,CYP17A1,CYP27B1,CYP2A6 (includes others),CYP3A4,DAB2,DCN,DDB2,DDIT3,DHCR7,DHRS1,DHX9,DYNLT1,EFNB2,FAS,FHL1,FHL2,FKBP6,FOS,FTL,G6PC,GADD45A,GNAS,GNLY,GSTA1,HBB,HLA-A,HSD3B2,HSPA8,ICAM1,ID1,IDH3B,IGFBP4,IGKC,IL7,INSR,IRF9,ITGAL,LAMC1,LCN2,LCP2,LEMD3,LEPR,LY96,LYN,MAPT,MCM3,MGP,MMP1,MSH2,NFE2L1,NOS2,NR1H3,NRP1,PCK1,PCNA,PCTP,PDGFRA,PDK4,PEA15,PLA2G4A,PLG,PLXND1,PPARA,PPARGC1A,PPP1R15A,PRKCD,PRLR,PRSS8,PSMA6,PTGES,PTGS1,RAC2,RBBP7,RCAN1,RORA,SCD,SEC11A,SGK1,SLC12A2,SMAD2,SPP1,SQOR,SST,STAT3,TAC1,TLR8,TM4SF1,TMEM50B,TNFAIP6,TP53,TRIB3,USP33,VCAN,VDR,VIM,VLDLR,XBP1,ZNF638 |

| | | | | | | | | | |
|---|---|---|---|---|---|---|---|---|---|
| IFNB1 | -0,046 | cytokine | Activated | 4,309 | 4,346 | 0,004 | 1,42E-14 | 8,26E-13 | ADAM17,ANXA5,APOL1,APOL3,AXL,BHLHE40,BST2,BTN3A3,CAPN2,CASP1,CASP3,CASP8,CCL5,CCR5,CD14,CD38,CD40,CD83,CD86,CDK10,CTSK,CXCL10,CXCL11,CXCL6,CXCL9,DDX58,DEFB1,DICER1,DKK3,FBN1,FOS,GBP2,GPC4,GZMH,HLA-A,HMGB2,IDI1,IDO1,IFI16,IFI27,IFI6,IFIH1,IFIT2,IFIT3,IFITM1,IFNGR1,IL13,IL18,IL1R1,IRF1,IRF7,IRF9,ISG15,ISG20,ITGAV,ITGB3,LAMB1,MCM6,MGST2,MMP2,MMP9,MX1,MYD88,NFKBIE,NID2,NMI,NNMT,NOCT,NOD2,NOS2,OAS1,OAS2,PARP12,PDK4,PKP2,PLAAT4,RNASEL,RND3,RSAD2,SLC39A14,SPP1,SPRY1,SPRY2,STAT1,STAT4,THBS2,TIMP1,TIMP2,TLR3,TLR7,TLR8,TNFSF10,TRIM21,TRIM38,UBA7,UCHL1,VDR,XAF1,ZDHHC14 |
| IKBKB | | kinase | Activated | 3,234 | 3,289 | 0,006 | 1,56E-14 | 9,04E-13 | AEBP1,AFP,ATF3,BAX,BIRC2,BIRC3,C3,CASP8,CCL19,CCL20,CCL5,CCNA2,CCND1,CCR7,CD44,CDC25B,CLU,CSF1R,CTSB,CTSF,CTSK,CTSZ,CXCL1,CXCL10,CXCL12,CXCL6,CXCR4,CYBA,CYP1B1,DCN,ECT2,ENPP2,EZH2,F3,FAS,FOS,FOXO1,FSCN1,FYN,GADD45A,GBP2,GCH1,GM2A,GRK5,GRN,HIF1A,HLA-A,ICAM1,IFI16,IGFBP6,IGFBP7,IL13,IRF1,ISG15,ITGB3,KIF20A,LCN2,LMNB1,LTB,MAT2A,MGP,MMP1,MMP2,MMP9,MX1,NAGLU,NFKB1,NGF,NOCT,NOS2,NPPA,PLA2G4A,POSTN,POU2AF1,PPM1D,PTEN,RBP1,RCAN1,S100A4,SERPINE2,SFRP1,SGK1,SMAD7,SOCS2,SOD2,STAT3,TGFBR2,TIMP2,TLR3,TNC,TNFAIP2,TNFRSF11B,TNFRSF1B,TP53,VCAM1,VCAN,VEGFA |
| CDKN1A | -0,136 | kinase | | -1,726 | -1,744 | -0,003 | 2,35E-14 | 1,34E-12 | ACTB,AURKA,BAX,BIRC2,CASP3,CCNA2,CCNB1,CCND1,CCND3,CDC25B,CDK1,CDKN1B,CDKN3,CEP55,COL1A2,CPA3,CTSB,CTSK,DLGAP5,DSE,DTL,DUSP1,ECSIT,FANCG,FAS,FN1,GLB1,GRN,HMGB2,HSPA5,IL1R1,IL32,ITGAM,ITGAV,ITGB3,KIF20A,LBR,LGALS3BP,LIMA1,LUC7L3,MAD2L1,MAF,MCM2,MCM3,MCM6,MCM7,MMP1,MMP9,MTTP,P4HA1,PBK,PCLAF,PCNA,PHGDH,PLK2,PRC1,PTPRC,RB1,RPA3,RRM1,SHC1,SMC4,SOD2,SPAG5,SSPN,STMN1,TCN2,TERT,TNFAIP6,TOP2A,TP53,TPX2,TTK,TUBG2,UBE2C,VEGFA,VGLL4,VIM,YTHDC1,ZBED5,ZNF84 |
| PDGF BB | | complex | | -0,020 | 0,034 | 0,006 | 2,82E-14 | 1,60E-12 | ACKR3,ADD3,ADM,ADRB2,AKAP12,AKR7A2,ANXA11,ARAF,ARG2,ATF3,BHLHE40,C3,CALM1 (includes others),CASP4,CAV2,CCL20,CCL8,CCND1,CCND3,CD44,CDK1,CDKN1B,COL18A1,COL3A1,CSF1R,CTH,DAB2,DCN,DDIT3,DGAT1,DUSP1,F3,FGF7,FHL1,FHL2,FLNA,FMO1,FN1,FOS,FOSB,FOSL1,FRZB,FXYD5,FZD2,GADD45A,GBP2,GHR,GJA1,GLUL,GPR183,GSS,GUCY1B1,HLA-E,ID3,IGFBP4,KRAS,MAT2A,MCM6,MGST2,MMP1,MMP2,MMP9,NAMPT,NFIL3,NOS2,NOX4,NPR3,NR4A1,NR4A3,NXF1,PCGF2,PCNA,PCOLCE,PDGFRA,PDLIM1,PLK2,POSTN,PPP1R15A,QARS1,RACK1,RBP1,RHOB,RND3,RPN2,SCD,SERPINA3,SGK1,SLC1A1,SLC25A11,SLC29A1,SLC7A7,SOD2,SRSF7,STAT3,SYK,TEAD4,TIMP1,TNC,TOB1,TP53,TRA2B,TRIB3,TRIOBP,VCAM1,VCAN,VEGFA,ZFP36 |
| TGM2 | 0,350 | enzyme | Activated | 5,529 | 5,567 | 0,004 | 3,12E-14 | 1,76E-12 | ADGRE2,ALPL,BCL2A1,BCL6,BIRC3,BNIP3,C12orf29,C3,CA2,CCND1,CD36,CD74,CD86,CD93,COX11,CX3CR1,CXCL10,DAPK2,DDX60,DLEU1,EHD1,FCER1G,FN1,GCA,GIMAP6,GJA1,GM2A,HIF1A,HLA-B,IFI35,IFI6,IFIT2,IFIT3,IFIT5,IL10RA,IRF9,ITGAM,ITGB3,LILRB2,LSP1,MAFB,MGP,MMP2,MMP9,NCF2,OAS1,OAS2,OAS3,OLFM4,PLAAT4,PLEKHO2,PLSCR1,POR,PPP1R16B,PTGES,PTGS1,S100A8,SACS,SAMHD1,SDCBP,SELL,SELP,SLC25A20,SLC27A2,SLPI,SMPD1,SP110,SPP1,STAT1,TAP1,TNFAIP2,TNFRSF11B,TOR1B,TP53,TPCN1,TRIM22,TYROBP,UBA7,VSIG4,WDR3,XAF1,ZNF146 |
| SREBF1 | -0,051 | transcription regulator | | -0,195 | -0,141 | 0,007 | 4,79E-14 | 2,68E-12 | ACACA,ACACB,ACADS,ACSL1,ACSL5,ADGRG1,AGMAT,APOBEC3F,APOC3,ARF4,ATOX1,BAX,BEST1,BHLHE40,BHLHE41,CAP2,CD14,CFD,CXADR,CXCL10,CYB5A,CYP27B1,CYP4F2,DEPP1,DHCR7,EHHADH,ELOVL6,FAS,FN1,FOXO1,G6PC,GLB1,GPNMB,GPX3,GSR,HNF4A,HSPA5,IDH1,IDI1,IFI30,IGHM,LPIN1,MDK,MEF2C,MLYCD,MSMO1,MTTP,NCF2,NFIL3,NOS2,NR0B2,NR1H3,NSDHL,OAT,P4HA2,PCK1,PCK2,PIK3R3,PKLR,PLS3,PLTP,PMP22,PMPCA,PMVK,PNPLA3,RBP1,RDH11,RETREG1,RNASET2,RPS24,S100A13,SC5D,SCD,SERPINA3,SEZ6L2,SLC22A4,SLC22A5,SLC2A2,SMPD1,SORBS3,STXBP1,SUCLG1,TM7SF2,TNFRSF1B,TNNI1,TP53,UCP2,VEGFA |
| VCAN | 2,007 | other | | 1,522 | 1,529 | 0,001 | 8,45E-14 | 4,70E-12 | ADM,AEBP1,AGRN,ANG,APOE,ASS1,C1R,C1S,C3,CASP1,CASP3,CBX6,CCL20,CD247,CD59,CLU,CPE,CXCL12,CXCL6,CYBA,DCN,ENPP2,F3,FAS,FN1,GLB1,GUCY1B1,ICAM1,IFI44,IFI44L,IFI6,IFIT2,IFITM1,IGFBP4,IRF8,ITGA3,LAMB1,LCN2,MEF2C,MIF,MMP9,MX1,MX2,NFKB1,OAS2,OAS3,OLFML2B,PCSK1N,PCSK5,RBP1,ROCK1,RXRA,SERPING1,SOD2,ST6GAL1,STAT1,TIMP1,TP53,TRIM22,VCAM1,VCAN,VEGFA,XAF1,YBX3 |

| | | | | | | | | | |
|---|---|---|---|---|---|---|---|---|---|
| RB1 | 0,275 | transcription regulator | Inhibited | -4,906 | -4,887 | 0,002 | 8,74E-14 | 4,83E-12 | ACO2,ACTB,ALG5,ALPL,ATP6V1D,BARD1,BAX,BCKDHA,BNIP3,CALCRL,CASP3,CASP4,CASP6,CASP8,CCNA2,CCNB1,CCND1,CCND3,CDK1,CDKN1B,CDX2,CKM,CLDN3,CRADD,CYP27B1,DCK,DDB2,DDIT3,DEK,DLAT,DLST,DMPK,DNMT1,EED,EZH2,F3,FABP2,FAM111A,FANCL,FAS,FN1,FOS,FOSL1,FOXN1,FZD6,GADD45GIP1,GHITM,GPR162,GPS2,GRPEL1,HMGB2,HSBP1,ID1,IDH3B,KANK2,KRT10,KRT4,LCK,MAF,MAP2K3,MCAT,MCM2,MCM3,MCM5,MCM6,MCM7,MECR,MEF2C,MELK,MET,MFAP1,MLYCD,MMP2,MRPL58,MT1G,MTOR,MYB,MYOM2,NDUFA10,NDUFA2,NDUFA3,NDUFS2,NDUFS4,NDUFS7,NDUFV1,NIT1,NKX2-5,NME1,OPA1,P2RY14,PARP1,PBX3,PCLAF,PCNA,PGAM2,PGRMC1,PMPCB,POLD3,PPARGC1A,PPFIBP1,PPT2,PTGS1,RAB27A,RB1,RMI1,RRM1,RRM2,SDHB,SDHD,SH3BP2,SLC22A18AS,SMARCA5,SOD2,TERT,TG,TNNC1,TNNT2,TOPBP1,TP53,TUBA1A,TUBG1,TXN2,ULK1,UQCRC1,VEGFA,VRK1,YWHAE,ZEB2,ZNF638 |
| EP300 | | transcription regulator | | -0,737 | -0,682 | 0,008 | 9,52E-14 | 5,23E-12 | ACACB,ACADL,ACADM,AKAP12,AP3M2,B2M,BANK1,BAX,BCL6,BCLAF1,CA2,CAPG,CCND1,CCND3,CCR7,CD36,CD53,CD59,CDKN1B,CHPT1,CKM,COL1A2,COMT,CPT2,CST7,CXCR4,DDC,DHRS3,DIO1,DUSP1,DUSP22,EPN1,EVI2A,EVL,EYA2,FDX1,FN1,FNBP1,FNDC3B,FOS,FOXO1,G6PC,GIMAP4,GM2A,GPR18,HBB,HLA-B,HLA-G,HMGCS2,HSD11B2,ID1,ID2,ID3,IGFBP6,IL10RA,IL15RA,IL1R1,IL7R,ITGB2,KMO,LGALS3BP,LHB,MAF,MAPT,MCF2L,MCM3,MEF2C,MFHAS1,MMD,MME,MMP1,MMP7,MSH2,MYO1F,NDRG1,NOS2,NPPA,NR4A1,OAS3,PARP1,PCK1,PDE10A,PDK4,PDLIM4,PERP,PHIP,PIPOX,PKLR,PLA2G4A,PLA2G7,PLEKHF1,PPARA,PPARGC1A,PRKCD,PRLR,PTEN,PTPRF,RB1,RBP1,RHOA,RNASEL,RORC,RSAD2,RTP4,SDC1,SH2D1A,SMAD7,SOD2,SPSB1,ST6GAL1,STAT4,TBXAS1,TERT,TESC,TGFBR2,TIPARP,TLE2,TLR2,TMPRSS2,TMPRSS4,TNC,TP53,TRAT1,TRIM26,TXNIP,UBE2C,UTRN,VEGFA,WNT8B,XBP1 |
| F2 | -0,163 | peptidase | Activated | 3,522 | 3,605 | 0,009 | 1,18E-13 | 6,44E-12 | AKR1B1,ALDH1B1,BAX,BIRC2,BIRC3,CALR,CASP1,CASP3,CBR3,CCL20,CCL8,CCNA2,CCND1,CCND2,CD44,CD55,CD63,CDKN1B,CKAP4,CLIC1,CLU,COL1A1,COL4A1,CORO1C,COTL1,CTPS1,CXCL1,CXCL10,CXCR4,CYBB,CYP2B6,DHRS9,DKK3,DOCK10,DOK5,DUSP1,EHD4,ELL2,ESM1,F2RL1,F3,FHL1,FN1,FOS,FOSB,FOSL1,HIF1A,ICAM1,IFRD2,IGFBP4,IL13,IL15RA,IL32,MFHAS1,MIF,MMP1,MMP2,MMP9,MYH9,NCF2,NOS2,NR4A3,NUP62,PDGFRA,PDIA4,PLA2G4A,PROCR,RAB3A,RAC1,RAC2,RCAN1,RHOA,RHOB,SELP,SERPINC1,SLC16A5,SLIT2,SMTN,SOD2,SRGN,TAGLN2,TAP2,TFPI2,TIMP2,TMED9,TNFSF10,TNNT2,TXNRD2,VCAM1,VEGFA,VWF |
| EGR1 | -0,859 | transcription regulator | | 0,566 | 0,620 | 0,007 | 1,23E-13 | 6,67E-12 | ACE,ALOX5AP,APOA1,ARC,ATF3,BACE1,BAIAP2,BAX,CASP3,CASP6,CASP8,CAV1,CAVIN1,CCND1,CCND2,CCR2,CD44,CLU,COL1A1,COL1A2,CSHL1,CXCL6,CYP17A1,ENPEP,F3,FAS,FN1,FOSL1,FTL,FYN,GAD1,GADD45A,GADD45B,GNAS,HIF1A,HPGD,HPSE,ICAM1,ID1,IL13,IL6R,ITGAM,JUND,LHB,LYZ,MAOB,ME1,MMP9,MTNR1A,MYB,NAB2,NDRG1,NFKB1,NR4A1,PNPLA2,PTEN,PTGES,RB1,SGK1,SLC12A2,SOD1,SOD2,SPP1,TGFBR2,TIAM1,TIMP1,TLR4,TNFSF10,TP53,VCAM1,VEGFA |
| P38 MAPK | | group | Activated | 2,249 | 2,325 | 0,008 | 1,54E-13 | 8,19E-12 | ADAM17,ANXA5,ARG2,ASS1,ATF3,BAX,BCL2A1,BNIP3,BNIP3L,CAV1,CCL5,CCL8,CCND1,CCND2,CCR7,CD1D,CD36,CD40,CD44,CD55,CD69,CD83,CD86,CDH5,CDX2,COL3A1,CTSK,CXCL1,CXCL10,CXCL11,CXCL12,CXCL6,CXCL9,CXCR4,CYBB,DDB2,DDC,DDIT3,DUSP1,F3,FAS,FLNA,FN1,FOS,GBP1,GCLC,GJA1,HIF1A,HSPA5,HTRA1,ICAM1,ID3,IL13,IL32,IRF7,ITGAM,ITGAV,ITGB2,ITGB3,ITGB8,JAM2,KDELR3,KLF15,MAL,MAP4K4,MEF2C,MIF,MMP1,MMP2,MMP9,MSH2,NFATC4,NFE2L2,NFKB1,NGF,NKX2-5,NOS2,NPPA,NR4A1,NR4A3,PCK1,PCNA,PLA2G4A,PLA2G7,POSTN,PPARGC1A,PPM1D,PTEN,PTPN12,RB1,RBP1,RCAN1,RHOB,RND3,SGK1,SMAD2,STAT1,TBP,TERT,TIMP1,TIMP2,TLR2,TNFSF10,TNNC1,TNNI1,TOP2A,TP53,ULK1,VCAM1,VDR,VEGFA,VEGFC,VLDLR,ZFP36,ZMAT3 |
| Hbb-b1 | | transporter | | -1,099 | -1,048 | 0,008 | 1,66E-13 | 8,73E-12 | ATP5MC2,C1QA,CCR2,CCR5,CD44,CD74,CLEC7A,COX15,COX6A2,COX8A,CXCL10,CYBA,CYBB,CYC1,FN1,FTL,HIF1A,ICAM1,IL10RA,IL2RB,IL2RG,ITGAM,LTB,LY86,NCF2,NDUFA3,NDUFA7,NDUFB11,NDUFB7,NDUFC1,NDUFS3,NDUFS4,NDUFS7,NDUFS8,NDUFV1,PECAM1,PRLR,SERPINF2,SURF1,TLR2,UCP2,UQCRC1,UQCRC2,VCAM1,VEGFC |

| | | | | | | | | | |
|---|---|---|---|---|---|---|---|---|---|
| HTT | | transcription regulator | Activated | 2,670 | 2,683 | 0,002 | 2,76E-13 | 1,40E-11 | ABCC10,ACADM,ACKR1,AGRN,AKAP12,APOA1,APOE,AQP1,ATF3,ATP1A3,ATP6V0E2,AVP,AZIN1,B2M,B4GAT1,BAIAP2,BASP1,BCL6,BHLHE40,BID,BIRC2,C1R,CA4,CACNA2D3,CALML4,CAMK2A,CASP1,CASP3,CAST,CAVIN3,CCL5,CCND1,CD74,CD9,CDH11,CDHR5,CDK2AP1,CDK5,CHUK,CKM,CLCN1,CLDN1,COL16A1,COL18A1,COL4A1,COL4A2,COL6A3,COX15,COX6A2,CSF1R,CTSD,CYC1,CYFIP2,DCN,DDIT3,DDX50,DHCR7,DKK3,DLG3,DLST,DNAJB1,ECI1,EMP3,ENO3,ENPP2,EPB41L3,ETNPPL,EVL,FGF1,FHL1,FKBP4,FN1,FOLR1,FOS,FOSL1,GAD1,GADD45G,GCLC,GLUL,GNB5,GNG7,GPX3,GRK2,GRK5,GSR,GSS,HIF1A,HMGB2,HOXB1,HOXB2,HPCAL1,HSPA5,HSPA8,HSPA9,IFI16,IL15,KCNJ2,KL,KLF9,LTF,MAST3,MCAM,MDH2,MEIS2,MGP,MICU1,MLF2,MMP2,MTNR1A,MTOR,NAB2,NCS1,NDUFA3,NDUFA7,NDUFS3,NDUFS4,NDUFS7,NFE2L2,NFIL3,NOS2,NR4A1,NSF,NTRK2,OSBPL8,OXT,PC,PCNA,PCTP,PDE10A,PDK2,PDK3,PDLIM4,PEA15,PER1,PFN1,POR,PPARA,PPARGC1A,PPP1R16B,PRKACB,PRKCD,PROM1,PSMB8,PSMB9,PSME1,PVALB,RAB17,RNF34,SDHB,SERPINA3,SERPINF1,SERPING1,SFRP1,SGK1,SIRT3,SLC12A2,SLC1A3,SNN,SOCS2,SOD1,SOD2,SP3,SPP1,SST,STAU1,STMN1,STX4,STXBP1,TAC1,TESC,TGFBI,THBS2,THRB,TIAM1,TM4SF1,TP53,TPI1,TRAP1,TUBB,TYRO3,UCHL1,UQCR11,USP2,VCAM1,VTI1B,XBP1 |
| CXCL12 | 0,814 | cytokine | | 1,892 | 1,940 | 0,006 | 3,14E-13 | 1,57E-11 | ACP5,ADAM17,AFP,ARAF,AVP,AXL,BAX,CA2,CCL5,CCND1,CD14,CD36,CD3G,CD44,CD47,CD69,CD9,CDKN1C,COL1A1,CTSK,CXCL10,CXCL12,CXCR4,CYP2A6 (includes others),DMPK,DPP4,FAS,FLNA,FN1,FOS,FYN,GAD1,GAS2,GSR,HCLS1,HIPK3,ICAM1,ID1,IFI27,IFNAR2,IFNGR2,IGFBP7,ITGAL,ITGB3,LCP2,MAPK12,MAPK8IP2,MARCKS,MMP2,MMP9,MSH6,NCK1,NPHS1,NR2F6,NR4A1,PCDH1,PPEF1,PRKCD,ROCK1,RRAS,RUNX3,SOCS2,SORBS3,TAC1,TIMP1,TNFRSF10D,TNFRSF11B,TNFRSF1B,TNFSF10,TNNC1,TP53,VIM,XRCC5 |
| palmitic acid | | chemical - endogenous mammalian | | -0,224 | -0,191 | 0,004 | 3,94E-13 | 1,96E-11 | ACAA1,ACACA,ACADL,ACADM,ACSL1,AHR,APOM,ATF3,BACE1,BAX,BIRC3,CASP3,CCL19,CCL5,CD36,CD86,CKM,CPE,CTSB,CXCL10,CYBA,CYBB,CYP4F12,CYP4F2,DDIT3,ELOVL6,EPHX2,F2RL1,FABP1,FABP2,FN1,FOLR2,FOS,FOXO1,G6PC,GCLC,HNF1A,HNF4A,HSPA5,HTR2B,ICAM1,IDO1,IL6R,INSR,LCN2,MAFB,MET,MLXIPL,MTOR,NCF2,NCF4,NFE2L2,NOS2,NR4A1,NR4A3,PCK1,PDK4,PKLR,PLIN2,PMVK,PNPLA2,PNPLA3,PPARA,PPARGC1A,PROCR,PTEN,SCD,SLC25A1,SLC2A2,TLR2,TLR4,TNFSF10,TP53,TRIB3,TXNIP,UCHL1,UCP2,VCAM1,XBP1 |
| IRF2 | 0,260 | transcription regulator | | 1,184 | 1,219 | 0,005 | 5,23E-13 | 2,59E-11 | ANG,B2M,BNIP3L,C1S,CASP1,CCND1,CD14,CDKN1B,CFB,CIDEC,CLEC10A,CLIC1,CTSS,CXCL10,DPY19L1,ERAP1,FAM111A,GBP1,HGD,ID1,IDH1,IFI35,IL7,IRF1,IRF7,IRF8,ISG15,LCN2,MMP7,MYH9,NAMPT,NOS2,OAS1,PFKFB3,PSMB10,PSMB8,PSMB9,PSME1,RARRES1,SDC1,SETD3,SRGN,SUPT16H,TAP1,TAP2,TAPBP,TLR3,TLR4,TLR5,TNFSF10,TPM1,TRIM21,UBE2L6,VCAM1 |
| CD38 | 0,912 | enzyme | | 0,590 | 0,659 | 0,009 | 6,01E-13 | 2,94E-11 | ANXA2,ASNS,ASS1,BCL6,BNIP3,BNIP3L,CCND2,CD44,CD86,CHST12,CISH,CKAP4,CLCN3,COL1A1,CRELD2,CRIP1,CSF2RB,EAF2,ELL2,ENDOG,FGL2,FN1,GADD45G,GALK1,GBP2,GCAT,ICAM1,IFI30,IL6R,JCHAIN,KCNAB2,KLHDC2,LGALSL,MANF,NDRG1,NFIL3,P4HA1,PDIA6,PDLIM1,PFKL,PFKP,PGM1,PKM,PPARGC1A,PRDX4,PSAT1,RBPMS,RRAS2,S100A4,S100A6,SDC1,SDF2L1,SELL,SLC16A3,SOCS2,ST7,STK39,THY1,TPI1,UPB1,VCAM1,VIM,XBP1 |
| IFN Beta | | group | Activated | 4,888 | 4,941 | 0,007 | 8,28E-13 | 3,98E-11 | ATF3,BST2,C3AR1,CASP4,CCL5,CCR7,CD247,CD40,CD69,CD74,CD83,CD86,CXCL1,CXCL10,CXCL11,CYP3A4,DDX58,DPP4,DUSP1,ENPEP,FN1,HLA-A,HLA-B,HLA-G,IDO1,IFI16,IFI27,IFI35,IFI44,IFI6,IFIH1,IFIT2,IFIT3,IFITM1,IFNAR2,IL15,IL15RA,IRF1,IRF7,IRF9,ISG15,MMP9,MX1,MX2,NOS2,OAS1,OAS2,OAS3,PRKCA,RSAD2,SPRY1,SPRY2,STAT1,TAPBP,TLR3,TNFSF10,TP53,XAF1 |
| APOE | -0,901 | transporter | Inhibited | -2,523 | -2,558 | -0,006 | 1,00E-12 | 4,78E-11 | ACAT1,ACOX1,ACSL1,ACTB,ADAM10,ADGRG1,ADNP,ANXA11,APOA1,APOE,ATF3,BAX,BEGAIN,C1QA,C1QB,C2,C3AR1,CALR,CASP1,CASP3,CAT,CAV1,CCL5,CCND1,CCND2,CCR5,CD36,CD40,CD44,CD86,CLU,COL18A1,COL1A1,CTSB,CTSD,CTSK,CTSS,CTSV,CYBB,DDIT3,ESR1,F2RL1,F3,FCGR2B,FKBP4,FKBP5,FOS,GLUL,GPX3,HABP2,HPCAL1,HSP90B1,HSPA5,ICAM1,IGFBP6,IL10RA,IL10RB,INSR,ITGA3,ITGAM,JAK1,LCN2,LIPA,MAP2K3,MAP3K1,MAPT,MGP,MMP2,MMP9,MYLIP,MYO1B,NCF2,NOS2,NOX4,NQO2,NR1H3,P4HB,PC,PEA15,PHF21A,PPP1R15A,PRKCQ,PRPSAP2,PTGES,S100A8,S100A9,SERPINA3,SLC25A44,SMAD1,SOD1,SOD2,SPP1,STAT4,TCIRG1,THY1,TIAM1,TIMP1,TREM2,TXNIP,UCP2,VCAM1,WBP2,XBP1 |

| | | | | | | | | | |
|---|---|---|---|---|---|---|---|---|---|
| HNF1A | -0,407 | transcription regulator | Inhibited | -4,242 | -4,170 | 0,009 | 1,13E-12 | 5,35E-11 | A1CF,ABCC2,ACE2,ADH1B,ADH6,AFP,AK2,ALB,ALDOB,ANGPTL3,ANXA4,APOC3,APOH,APOM,AQP3,BACH2,BCL6,BTN2A1,C1orf50,C1S,C2,C8B,CCND1,CD55,CDHR2,CDKN1B,CLIC5,CMC2,COL3A1,COX6A2,CSHL1,CTSZ,CYB5A,CYP27A1,DDC,DPP4,DTL,EIF4EBP2,ENPEP,F11,FABP1,FAS,FBP1,FH,FMO1,FOLR1,FUT3,FUT6,G0S2,G6PC,GARS1,GATM,GCNT3,GJB1,GLUL,GMDS,GOT1,GPR137B,GRHPR,GSTA1,GTF2E1,HABP2,HAO1,HGD,HNF1A,HNF1B,HNF4A,HNMT,HOPX,HPGDS,HSPA5,IFNA10,IHH,IL36A,INSR,KIF20A,KNG1,LIPA,MCCC1,ME3,MIA2,MLXIPL,MSH6,MT1X,MTHFD1,MTTP,NAPA,NAT10,NDUFS2,NOP16,NR0B2,OGDH,PAH,PATJ,PCK1,PFKFB2,PFKL,PFKP,PIGR,PKLR,PLG,PRLR,PROC,RACK1,RNF186,SERPING1,SGK2,SLC10A2,SLC22A11,SLC26A1,SLC2A2,SLC37A4,SLC9A3R1,SLCO3A1,SLPI,SRI,SULT1C2,SUPV3L1,TARS1,TBC1D8B,TEF,TMED10,TMEM140,TMPRSS2,TMPRSS4,TP53TG1,TPI1,TRAPPC13,UCP2,UGT2A3,UQCRC2,UROD,VIL1,VLDLR,XBP1 |
| IL27 | | cytokine | Activated | 5,645 | 5,684 | 0,005 | 1,27E-12 | 6,00E-11 | AHR,APOL1,B2M,BST2,CCL20,CCL5,CCND2,CCND3,CD14,CD163,CD38,CD63,CD69,CD74,CD86,CKLF,CXCL10,CXCL11,CXCL9,DEPP1,FOS,GADD45G,GBP2,GZMB,HLA-A,HLA-B,HLA-C,HLA-DMA,HLA-DMB,HLA-DOA,HLA-DOB,HLA-DPA1,HLA-DQA1,HLA-DQB1,HLA-DRA,HLA-E,HPGD,ICAM1,ID2,IL10RB,IL13,IL15RA,IL18,IRF1,ITGAM,MAF,MRC1,MX1,NOS2,OAS1,PLAAT4,RORC,STAT1,TAP1,TLR3,TNFSF10,UBD,VCAM1,VIM |
| NR4A1 | -1,108 | ligand-dependent nuclear receptor | | 1,408 | 1,407 | -0,000 | 1,56E-12 | 7,28E-11 | ACACA,ACO1,ACO2,ADM,ALDOB,APOE,ATF3,CCL5,CCND1,CCND2,CD36,CD69,CDH5,CDKN1B,COL16A1,COL1A1,COL1A2,COX11,CST7,CXCL12,CYP17A1,DDIT3,DLAT,DLD,DLST,EHF,EHHADH,ELK3,ENO1,ENO3,FBP1,G0S2,G6PC,GPD1,GYS1,HDAC1,HDAC6,HIF1A,HSD3B2,ICAM1,IL13,KITLG,LPIN1,MAPK8,MARCKSL1,MAT2A,MDH2,MMP7,MMP9,NDUFA10,NDUFA3,NDUFA7,NDUFB7,NDUFS1,NDUFS7,NDUFS8,NR4A3,NUCB2,OGDH,PC,PCK1,PCK2,PCNA,PDIA3,PDK3,PDK4,PEBP1,PFKL,PGAM2,PGM1,PKLR,PPP1R3C,RBKS,RPIA,RXRA,SCD,SDHD,SMAD7,SOAT1,SPINT2,SUCLA2,SUCLG1,SUCLG2,TJP1,TNFSF10,UBE2C,UCP2,VCAM1 |
| IL12 (complex) | | complex | Activated | 3,528 | 3,575 | 0,006 | 1,70E-12 | 7,91E-11 | ACADL,AHR,CASP1,CASP3,CCL19,CCL20,CCL5,CCR2,CCR5,CCR7,CD44,CD63,CD69,CD86,CISH,CXCL10,CXCL11,CXCL6,CYTIP,DDB2,FAAH,FAS,FOS,GADD45B,GADD45G,GCNT2,GJA1,GPT,GZMA,GZMB,ICAM1,IDO1,IFIH1,IFIT2,IFNGR1,IFNGR2,IL13,IL15,IL18,IL32,IL7R,IRF1,IRF8,ISG15,ISG20,ITGAL,ITGB2,KLRB1,KLRD1,LTB,MIF,MMP2,MMP9,MYD88,NOS2,PDCD4,PLAC8,PRF1,RPA1,RPA2,RPA3,SELL,SELP,SET,SH2D1A,SMAD2,SOCS2,STAT4,TGFBR2,TIMP1,TLR2,TLR4,VEGFA,XPC |
| CSF3 | -0,136 | cytokine | | -0,112 | -0,091 | 0,003 | 1,73E-12 | 7,97E-11 | ARHGEF10,BAX,BIRC3,C3AR1,CAPG,CCL5,CCND2,CCND3,CCR7,CD14,CD44,CDKN1B,CEBPE,CISH,CTSD,CX3CR1,CXCL12,CXCR4,CYBB,ENPP2,FOS,FPR1,FUS,GADD45A,GADD45B,GADD45G,GJA1,GNLY,GPR183,GZMB,GZMH,HLA-DPA1,HLA-DQA1,HLA-DRA,HLA-DRB1,HOXB7,ICAM1,IK,IL7,ITGAM,ITGB2,KITLG,LCN2,LTB,LTF,LY96,LYZ,MET,MMP2,MMP9,NFIL3,PLSCR1,PRKCA,RAB27A,RORC,SELL,STAT3,TLR2,TLR4,TLR5,TLR8,TNFRSF1B,TNFSF10,TP53,VCAM1,VEGFA |
| IL1 | | group | Activated | 4,795 | 4,856 | 0,006 | 1,75E-12 | 8,03E-11 | ABCG2,ACACA,ADAM17,ADAMTS1,ADRB2,AHR,ALB,ALDH3A1,APOC3,APOE,AVP,BAX,BHLHE40,C2,C3,CASP1,CBR3,CCL20,CCL5,CCR7,CD40,CD55,CD59,CFB,CLEC7A,COL1A1,CSF1R,CSF2RB,CTSB,CXCL1,CXCL10,CXCL11,CXCL9,CYP17A1,CYP27A1,CYP2B6,DCN,DDC,DUSP1,F3,FAS,FGF7,FN1,FOS,FPR1,FZD5,GJA1,GLS,HIF1A,ICAM1,IDO1,IL1R1,IL2RB,IL33,IL4R,IL7,IRF1,ITGAM,ITGB2,KITLG,KYNU,LAMB1,LYZ,MAP2K3,MMP1,MMP2,MMP7,MMP9,MTTP,NGF,NOS2,NR1H3,PANX1,PDGFRA,PIGR,PLA2G4A,PLD1,PPARA,PPARGC1A,PTGS1,RORA,RORC,RXRA,S100A9,SELL,SELP,SHBG,SOD2,SPARC,TAC1,TG,THY1,TIMP1,TLR2,TLR4,TLR7,TNC,TNFAIP6,TNFRSF11B,TNFRSF1B,TNFSF10,TP53,UGCG,VCAM1,VCAN,VEGFA |
| STK11 | | kinase | Inhibited | -3,454 | -3,428 | 0,003 | 2,03E-12 | 9,30E-11 | A2M,ACE,ADGRE2,AHCY,AK2,AKR1A1,ALB,ALDH9A1,ASS1,ATF3,ATP5F1D,ATP5MC2,BTF3,CCL18,CCNA2,CCND1,CCT6A,CDKN1B,COL3A1,COX8A,CTPS1,CTSD,CXCL6,CYC1,CYP2B6,DCTPP1,DLGAP2,DNMT1,ECM2,ENO1,FBXO9,FN1,FOSL1,GADD45A,GADD45B,GOT2,HIF1A,ID1,IL2RB,IL33,IMPDH2,LTB,LUM,MCAM,MDH2,MMP2,MMP9,MPST,MRPS15,NDUFA10,NDUFA2,NDUFA3,NDUFB11,NDUFB7,NDUFS7,NDUFS8,NDUFV1,NEDD9,NET1,NOS2,PAPSS1,PCK2,PGM1,PHGDH,PKM,PNP,PSAT1,PSMB5,PTEN,RASGRP1,RASSF2,RB1,RPIA,RRM1,RRM2,SARDH,SDHB,SDHD,SHMT2,SLC16A4,SPP1,STK17B,TARS1,TKT,TNFSF10,TOP2A,TPI1,TWF2,UQCR11,UQCRC1,VEGFA |

| | | | | | | | | | |
|---|---|---|---|---|---|---|---|---|---|
| TNFSF11 | | cytokine | | 1,249 | 1,317 | 0,008 | 2,65E-12 | 1,20E-10 | ACP5,AHR,AURKA,BCL2A1,BST1,C3AR1,CA2,CAPN2,CAV1,CCL5,CCND1,CCND3,CD14,CD40,CD44,CD83,CDK14,CDKN1B,CSF1R,CTSK,DAB2,DMPK,DUSP1,ECT2,EHD1,ESR1,F3,FAS,FGR,FHL2,FOS,FOSL1,FOXO1,FPR1,GADD45B,GCH1,GCLC,GLRX,GRB10,GRN,HCK,HCLS1,ICAM1,IFRD1,IL13,IL13RA1,IL15,IL15RA,IRF8,ITGAL,ITGAM,ITGAV,ITGB3,LAT,MAFB,MARCKSL1,MMP1,MMP9,NAB2,NFIL3,NFKB1,NOS2,NR1H3,PHLDA3,PLD1,POR,PTPRE,RALGDS,RCAN1,RHOB,RRAS,SDC1,SENP2,SERPINB9,SLC7A7,SOD2,SPP1,STAT1,STMN1,TLR2,TLR4,TNFRSF11B,TNFRSF1B,TSPAN13,TTK,VCAM1,VRK1 |
| LEP | -0,067 | growth factor | | -0,051 | -0,011 | 0,004 | 3,09E-12 | 1,38E-10 | ABCC3,ACACA,ACADL,ACADM,ACADVL,ACAT1,ACO1,ACOX1,ACSL5,ALDOB,APOA1,APOA4,APOH,APOM,AQP3,ASL,ASS1,BAX,BCL2A1,BID,CASP3,CASP6,CAV1,CCL5,CCNA2,CCND1,CD14,CD36,CD40,CD63,CDKN1B,CEL,CFD,COL1A1,COL3A1,COL4A1,CSHL1,CTSV,CXCL10,CYB5A,CYBA,CYBB,CYP17A1,CYP27B1,CYP2C8,CYP4A11,DGAT1,ECH1,EHHADH,ELOVL6,EPHX2,ESR1,ETFDH,F3,FAAH,FAS,FDX1,FMO5,FOS,FTO,G6PC,GADD45A,GCLC,GNAS,GOT2,GSTA3,HGD,HSPA5,ICAM1,IDH1,IL13,IL1R1,ITGAM,JUND,LEPR,LIPA,MLX,MLXIPL,MMP1,MMP2,MMP7,MSMO1,NAMPT,NCF2,NOS2,NPPA,NPY1R,OPLAH,PCK1,PCNA,PECAM1,PEMT,PLIN2,PNPLA2,PPARA,PPARGC1A,PRF1,RAC1,RORC,SC5D,SCD,SCP2,SELP,SLC13A2,SLC25A5,SLC27A5,SLC2A2,SMPD1,SOD1,SOD2,SPARC,SPP1,SST,STAT3,STEAP4,TERT,TG,TIMP1,TMED9,TNFRSF11B,TNFSF10,TP53,TRPV1,TSPO,UCP2,VCAM1,VEGFA,VIM,ZFP36 |
| IRF8 | 1,414 | transcription regulator | | 1,933 | 1,963 | 0,005 | 4,00E-12 | 1,78E-10 | ATF3,B2M,BANK1,BAX,BCL6,CASP3,CCL5,CCR2,CCR7,CD14,CD37,CD40,CD74,CD83,CD86,CEBPE,CFB,CSF1R,CTSS,CYBB,DAB2,DDX58,FANCF,FAS,FOS,GBP1,HOXB1,ICAM1,ID3,IDO1,IFI44L,IFI6,IFIT2,IFIT3,IL15,IL18,IRF8,ISG15,ITGAM,JAK1,LSP1,MAF,MAFB,MAP4K4,MMP9,MS4A1,NOS2,OAS1,SOCS7,STAT1,TERT,TLR3,TLR4,TP53,TRIM21,TYROBP |
| TLR4 | 0,334 | transmembrane receptor | Activated | 5,315 | 5,379 | 0,007 | 4,16E-12 | 1,84E-10 | ADM,APOBEC3B,ATF3,BAX,BPI,C3,CASP8,CAST,CCL20,CCL5,CCL8,CCND2,CCR5,CCR7,CD14,CD163,CD200,CD38,CD40,CD44,CD86,CERS2,CFB,CISH,CTSK,CXCL10,CXCL6,CYP27B1,DAB2,DDIT3,EGF,F3,FAS,FCGR2B,FGL2,GADD45G,GBP2,GJA1,GRK2,HDAC1,HHEX,HLA-DQA1,HPSE,HSPA5,HTRA1,ICAM1,IFI16,IFIT2,IFIT3,IFNA10,IL13,IL15,IL15RA,IL18,IL2RB,IRF1,IRF2,IRF7,IRF8,ISG15,ISG20,ITGAM,LCN2,LHB,MET,METTL1,MIF,MMP1,MMP9,MX1,MYD88,NFKB1,NMI,NOD2,NOS2,NR1I3,NR4A1,NR4A3,NSMAF,PLEK,PLK2,PTGES,RAC1,RSAD2,RXRA,SELP,SGPL1,SIGIRR,SIRT3,SLA,SLCO3A1,SMPD1,SMPDL3A,SOD2,SPP1,SPTLC2,ST3GAL1,STAT1,STAT3,TCF4,TLR2,TLR3,TLR4,TNFSF10,TPM1,TREM2,TRIM21,TRIM38,TSC22D1,TSPO,UGCG,VCAM1,XBP1 |
| prostaglandin E2 | | chemical - endogenous mammalian | | 0,750 | 0,780 | 0,004 | 4,34E-12 | 1,91E-10 | ABCG1,ADAMTS1,AHR,ANXA2,AQP3,ARC,ATF3,BAX,BIRC3,CASP1,CASP3,CASP4,CASP6,CCL20,CCL5,CCL8,CCNB1,CCND1,CCR5,CCR7,CD14,CD40,CD44,CD55,CD69,CD83,CD86,CNNM2,COL1A1,CST7,CXCL1,CXCL10,CXCL12,CXCR4,CYTIP,DCN,DGAT1,DUSP1,DYNLL1,ESRRA,F2RL1,FAS,FN1,FOS,FOSB,FOSL1,FOXO1,GJA1,HIF1A,HSD3B2,ICAM1,ID1,IDO1,IGFBP4,IGFBP7,IL15RA,IL1R1,IL2RG,IL7R,IRF1,ITGAM,MAFB,MMP1,MMP2,MMP9,MRC1,MXI1,NEDD9,NFIL3,NOS2,NR1H3,NR4A1,NRP1,PKLR,PLA2G4A,PLK2,PTGES,PTGS1,RIN1,RORC,S100A8,SELL,SOCS2,SPP1,ST3GAL1,ST3GAL4,TCF4,TGFBR3,TJP1,TNFAIP6,TNFRSF11B,TP53,TRAP1,UCP2,VEGFA,VEGFC,VIM,XRCC5 |
| HNF1B | 0,329 | transcription regulator | Inhibited | -2,631 | -2,626 | 0,001 | 4,39E-12 | 1,92E-10 | ACADVL,ACTN1,AFP,ALB,ALDOB,ANXA4,BHLHE40,CA2,CD44,CD99,CDK1,CLDN1,COL5A1,COL5A2,CXCL1,DEFB1,DUSP1,EXT1,FN1,FZD2,GSTA1,HNF1A,HNF4A,ID1,ID3,IHH,ITGA3,ITGB6,LAMB1,LUM,MAFB,MET,MMP2,MMP7,MPZL2,NID2,PDZK1IP1,PLEK2,PMP22,PSTPIP2,PTPRM,S100A2,S100A4,S100A6,SGK2,SLC29A1,SLC2A2,SPARC,TACSTD2,ZEB2 |
| RARA | | ligand-dependent nuclear receptor | | 0,405 | 0,405 | -0,000 | 4,69E-12 | 2,05E-10 | AASS,ABCC2,ACACA,ACACB,ADAM10,ADAM17,ALOX5,APOA1,APOC3,APOM,ARNT2,CALB1,CCNA2,CCND1,CD14,CD24,CD38,CD44,CD53,CD84,CD9,CD93,CDK1,CDKN1B,CDKN3,CEBPE,CENPM,CENPU,CLMN,CSHL1,CTSS,DHRS3,DUSP1,ELL2,ENAH,F12,FOLR2,FOS,FUT4,GDF2,GHR,GJB4,HCP5,HLA-B,HLA-C,HLA-G,HLA-J,HNF1B,HOXA1,HOXA10,HOXB1,HOXB7,IFI44,IFI44L,IGFBP6,IL13,IL1R1,IL6R,IRF1,LAMB1,LGALS1,LGALS3BP,LTF,LY6G6D,LYPD1,MAD2L1,MAFB,MAML1,MAOB,MAP3K10,METTL7A,MGP,NEDD9,NRP1,OAS1,OXT,PBK,PCK1,PTTG1,RARG,RARRES1,RBP1,RORC,RSAD2,RTP4,SAMD9,SAMHD1,SEC31A,SELL,SHANK2,SIPA1L1,SLC25A36,SLC2A2,SMC4,SMPD1,SP110,SPP1,STAT1,STRA6,TAPBP,TERT,TG,TMSB15A,TNFRSF11B,UBA7,UBL3,WNT8B,XBP1 |

| | | | | | | | | | |
|---|---|---|---|---|---|---|---|---|---|
| TP63 | | transcription regulator | | -0,183 | -0,146 | 0,004 | 4,75E-12 | 2,06E-10 | ADA,ADA2,ADAMTS1,ADM,AGR2,AHR,ALDH1A3,ANOS1,ARAF,ATG4A,AXL,BAX,BHLHE40,BMP7,BST2,CA4,CASP3,CAST,CCNA2,CCNB1,CCND1,CCND2,CCND3,CCNG1,CDK1,CDK18,CDKN1B,CDKN1C,CLDN3,COL4A1,COL5A1,CPT2,CSTA,CTSD,CXCR4,CYP2A6 (includes others),DDB2,DICER1,DNMT1,DOK1,EFNA4,ENG,F3,FAS,FBN1,FGFR3,FN1,FOS,FOSL1,FUBP1,GADD45A,GCNT3,HAGH,HSPA8,ID1,ID3,IGFBP6,IGFBP7,IHH,INSR,ITGA2,ITGA3,ITGA7,ITGB8,KHK,KRT10,KRT7,LYN,MAD2L1,MAFF,MAP2K3,MAP4K4,MAPK8,MPZL2,MYB,PCNA,PERP,PIK3R3,PKLR,PKP2,POSTN,PPARA,PRKAG2,PRKCZ,PRNP,PRPF4,PTEN,PTPN12,RAC2,RIN1,RPA1,S100A2,S100A4,S100A8,SERPINF1,SFN,SH2B3,SMAD2,SSPN,TERT,TGFBR2,THY1,TMBIM6,TNC,TNFAIP8,TNFSF10,TP53,TPM1,ULK1,UPK2,VDR,VEGFA,VIL1,VIM,WFDC2,XPC,ZEB2 |
| RAF1 | -0,147 | kinase | | -0,879 | -0,810 | 0,009 | 6,22E-12 | 2,63E-10 | ACOT7,AKAP12,ANXA1,ATP6V1B1,BCL6,CA2,CCL19,CCND1,CD37,CD44,CD74,CDKN1B,CHN1,CLDN1,CSTA,DHRS3,EMP3,ESR1,FAS,FOS,GRN,HIF1A,HMGB3,HPCAL1,HSD11B2,ID3,IGBP1,IL1R1,ITGA2,ITGAM,ITGB3,LHB,LPGAT1,MAOB,MCF2L,MET,MMP1,MMP9,MT1F,NFKB1,NGF,NPPA,OSMR,PC,PCK1,PLD1,PNP,PPP1R10,PRORP,PRSS3,RAC1,RND3,RRM2,SDC1,SELENBP1,SH2D1A,SLC16A7,SLC26A2,SMAGP,SNRPD1,SPRY2,ST3GAL4,TBP,TERT,TFPI2,TIAM1,TIMP1,TNNI1,TSC22D1,TSPOAP1,TUBB2A,TUBG1,TXNIP,VASH1,VEGFA |
| AKT1 | 0,023 | kinase | | 1,697 | 1,736 | 0,005 | 7,11E-12 | 2,98E-10 | ANKH,APOC3,ASS1,ATP6V1E1,BAX,BCL2A1,BIRC3,CASP3,CCL5,CCND1,CCND2,CCND3,CD40,CD69,CD74,CD86,CDKN1B,CISH,CLU,COL4A1,CRYZ,CTSV,CXCL10,CXCL6,CXCR4,DHCR7,EBP,ENO1,ESR1,FAS,FN1,FOS,FOSB,FOXO1,FSTL1,FXYD5,G6PC,GATM,GCLC,GCLM,GOSR2,GOT2,GSR,GSS,HIF1A,HIPK3,HMGB2,HNRNPM,JUND,KLF15,LAMB1,LCN2,LIPA,ME1,MMP2,MMP9,MMUT,MSMO1,MYB,NDC80,NFE2L2,NFKB1,NOS2,NOX4,NPPA,PCK1,PCOLCE,PDGFRA,PKM,PLA2G4A,PLG,PPARA,PPARGC1A,PTGS1,RAB11A,RHOB,SCD,SERPINF1,SLC3A2,SPP1,STUB1,TGFBR3,THBS2,TIMP1,TNFSF10,TP53,TPD52L1,UBE2C,VCAN,VEGFA,VIM,ZWINT |
| CSF1 | | cytokine | Activated | 2,111 | 2,170 | 0,007 | 8,04E-12 | 3,33E-10 | ACP5,ADGRE5,APOE,BAX,CCL5,CCNB1,CCND1,CCND2,CCND3,CCR5,CD163,CDKN1B,CSF1R,CTSK,CXCL6,CYBB,CYP17A1,DHCR7,DUSP1,DUSP9,EBP,ENO1,F2RL1,FAS,FCER1G,FCGR3A/FCGR3B,FHL2,FN1,FOLR2,FOS,GLS,GOT2,GPT,HSP90B1,HSPA5,IDI1,IFNGR1,IL10RA,IL18,IRF7,IRF8,ITGAM,ITGAV,ITGB3,LBR,MAFB,MMP9,MRC1,MSMO1,NCF2,NSDHL,PCNA,PFKFB3,PFKL,PKM,PMVK,PTPRC,RHOA,SC5D,SFRP1,SIGLEC1,SLC16A3,SLC1A5,SLC29A1,STAB1,STAT1,TLR1,TLR2,TLR5,TNFRSF1B,TP53,TPI1,VCAM1,VEGFA,XBP1 |
| IFN alpha/beta | | group | Activated | 5,798 | 5,844 | 0,007 | 8,21E-12 | 3,39E-10 | ARHGEF10,AXL,BAX,BID,CCL5,CCR2,CCR7,CD40,CD69,CD83,CD86,CLEC10A,CXCL10,CXCL14,FGL2,HLA-A,IDO1,IFI16,IFIT2,IFIT3,IL15,IL15RA,IL7,IRF1,IRF7,IRF8,MRC1,NOS2,RSAD2,SERPINB9,SLPI,STAT1,TLR1,TLR2,TLR3,TLR5,TLR7,TNFSF10,TRIM21 |
| Ige | | complex | Activated | 4,136 | 4,207 | 0,007 | 1,11E-11 | 4,53E-10 | ABAT,ACTN1,ALOX5,ANXA1,AQP3,BAIAP2,BCL2A1,BCL6,C3,CAMK2G,CAPN2,CARHSP1,CAV1,CCL5,CCND2,CCND3,CCR5,CD40,CD53,CD74,CD86,CHST8,CISH,CLEC7A,COL18A1,CTSK,CXADR,DUSP3,EIF4G2,ELMO1,FAS,FCER1A,FCER1G,FCGR2B,FHL2,FYN,GADD45B,HIP1R,HLA-DMA,HSPA9,ICAM1,IL13,IL15,IL33,IL7R,ITGAV,ITK,KLC1,KRAS,LAMA5,LCP2,MALT1,MARCKSL1,MCF2L,MDFIC,MGST2,MIF,MMD,MTOR,NAPA,NCF4,NFKB1,NFKBIE,NPHS2,NRP1,PEA15,PEBP1,PECAM1,PFN1,PIK3CB,PIK3IP1,PILRA,PLA2G12A,PRKCA,PTPN6,PTPRF,PXMP2,RALGDS,RASGRP1,RSAD2,SCD,SDC3,SERPINB1,SNRPD1,SPP1,TCIRG1,TLR2,TLR7,TNC,TNFRSF11B,TNFRSF1B,TNFSF10,TPPP3,TRAF5,TUBB6,UGCG,VCAM1,VDR,VEGFC,VIM,WNT10B |
| CHUK | -0,292 | kinase | Activated | 3,497 | 3,560 | 0,008 | 1,45E-11 | 5,84E-10 | ACKR3,AEBP1,ARL6IP5,ATF3,C3,CASP4,CCL19,CCL21,CCL5,CCN3,CCNB1,CCND1,CDKN1B,CHUK,CLU,COL18A1,CTSB,CTSF,CTSZ,CXCL10,CXCL12,CXCL6,CYBA,DCN,DNMT1,EGF,ENAH,ENPP2,F3,FAS,FOS,GADD45B,GBP2,GCH1,GM2A,GRN,HLA-A,ICAM1,IFI16,IFI35,IFIT2,IGFBP6,IL13,ISG15,ITGB3,LCN2,MAT2A,MGP,MMP2,MMP9,NAGLU,NGF,NOCT,OVOL1,PCNA,PLA2G4A,PLSCR1,RB1,RBP1,RCAN1,SERPINE2,SFRP1,SGK1,SNX10,SPP1,TLR3,TNFAIP2,TNFRSF11B,TNFRSF1B,TP53,VCAM1,VEGFA,VEGFC |

| | | | | | | | | | |
|---|---|---|---|---|---|---|---|---|---|
| SIRT1 | -0,165 | transcription regulator | Inhibited | -5,156 | -5,194 | -0,004 | 1,53E-11 | 6,13E-10 | ABCC2,ABCG1,ACADM,ACTB,ADAR,AFP,ANXA4,ARNT2,BAX,BCLAF1,BIRC3,BNIP3,CAT,CAV1,CCL5,CCNB2,CCND1,CCND2,CCND3,CD36,CD74,CD83,CDKN1B,CDKN2C,CORO1A,CYP2B6,DDX58,DDX60,ERMP1,ESR1,EVPL,FAS,FCMR,FGF7,FGFR3,FN1,FOXO1,G6PC,GABARAPL1,GADD45A,GFPT1,GSTA3,HCK,HIF1A,HLA-A,HLA-DQA1,HLA-DQB1,ICAM1,ID1,IFI44,IFIT3,IGHM,IL13,IL2RG,IRF7,JCHAIN,KDM5B,KIF13B,LAMB1,LAPTM5,LCK,LGALS3BP,MAOA,MAPT,MEF2C,MGAT2,MMP1,MMP2,MMP9,MYL6,NAT1,NEDD4L,NFKBIE,NKG7,NTRK2,OAS1,OAS2,PARP1,PARP12,PC,PCK1,PCK2,PDGFRA,PDK4,PER1,PHB,PLAGL2,PLEKHA6,PNPLA3,PPARGC1A,PRKCI,PSMB9,RAC1,ROCK1,RSAD2,RTP4,SHANK2,SIRT3,SLC29A1,SLC43A3,SNRNP70,SOD2,SP110,SSBP3,STAT1,STAT3,TACSTD2,TAP1,TERT,TIMP2,TMPRSS4,TP53,UBA7,UCP2,UQCRC2,XPC |
| PARP9 | | enzyme | Activated | 3,326 | 3,361 | 0,009 | 1,54E-11 | 6,16E-10 | BCL6,CD74,HLA-DOB,HLA-DQA1,HLA-DQB1,HLA-E,IFI44,IFIT2,IFIT3,IRF1,IRF2,IRF7,ISG15,OAS2,SP110,STAT1 |
| BRCA1 | | transcription regulator | Activated | 2,115 | 2,137 | 0,003 | 1,77E-11 | 7,04E-10 | ACTB,ALDH1A3,BAX,BCKDHB,CCL5,CCNB1,CCNB2,CCND1,CCNG1,CDH3,CDKN1B,COL18A1,CTSD,CTSF,CYP1B1,CYP3A4,DDB2,DDIT3,DDX58,DUSP3,ENPP2,ESR1,FAS,FHIT,FHL2,FKBP5,FOXO1,GADD45A,GADD45B,HBB,HSD11B2,IFI27,IFI6,IFIT2,IFIT3,IFITM1,IRF7,JAK1,LPIN1,MAD2L1,MX1,NEDD9,NFE2L2,PCBP2,PCNA,PERP,PLSCR1,PLTP,PTEN,RB1,ROBO1,S100A2,SERPINE2,SFN,STAT1,TAP1,TBP,TERT,TNFAIP2,TP53,UBE2I,VCAN,VEGFA,VIM,XPC |
| ESR2 | | ligand-dependent nuclear receptor | | -0,382 | -0,371 | 0,002 | 2,05E-11 | 7,98E-10 | ADAMTS1,AKAP1,AKAP12,ANXA4,APOE,AQP3,ARNT2,ATF3,AVP,BAG1,BID,BIN1,C3,CA2,CAPN2,CAV1,CCN3,CCNA2,CCND1,CD93,CDKN1B,CDKN1C,CGA,CHST15,CHUK,CLDN1,CLIC5,COL16A1,COL1A1,COL1A2,CRYZ,CTSD,CXCL12,CYP17A1,CYP2A6 (includes others),DAB2,DDAH1,DDB2,DHCR7,DMTN,ENPP2,ESR1,F2RL1,FHL1,FN1,FOS,FOXO1,FSCN1,FZD2,GADD45B,GALNT7,GCLC,GJB1,GLB1,GLRX,GM2A,GPSM2,GRPR,GSTP1,HPGD,IGFBP4,IMPDH2,ITGA2,JAK1,KCNK15,KRT19,LAPTM5,LHB,MAOA,MAP2K3,ME2,MGAT3,MME,MMP1,MMP2,MPC1,MUC1,NDRG1,NEDD9,NFKB1,NID2,NIPSNAP1,NOS2,OXT,PCBD1,PCYOX1,PDK4,PERP,PGRMC2,PIBF1,PLIN2,PPP1R12A,PPP1R3C,PRKCH,PSAT1,PTEN,PTH1R,PYGL,RAI2,RAPGEF6,RASSF2,ROCK1,RORC,S100A9,SEMA3B,SFXN3,SGK1,SHC1,SLC2A4RG,SLC9A3R1,SMAD2,SMAD7,SNN,SOCS2,SOX4,SPP1,ST8SIA4,SUCLA2,TAC1,TEAD4,TERT,TESK1,TGFBR2,THBS4,TIMP1,TIMP2,TNFRSF11B,TPM1,TSC22D1,TSPAN13,TSPO,UBL3,VEGFA,VIM,VWF |
| ADIPOQ | -0,154 | other | | -0,275 | -0,270 | 0,001 | 2,39E-11 | 9,22E-10 | ACACA,ACACB,ACOX1,ACSL1,ACSL5,ACSM5,ADIPOR1,CASP6,CCL20,CCL5,CCND1,CD163,CD36,CDKN1B,CGA,CLEC10A,COL3A1,CXCL1,CXCL10,CXCL11,CXCL9,CYBA,CYBB,DGAT1,DUSP1,F3,FN1,FOS,G6PC,GPD1,HMGCS2,ICAM1,IL4R,LEPR,MMP1,MMP2,NCF2,NCF4,NOS2,NOX4,NPPA,NR1H3,PCK1,PCNA,PDCD4,PPARA,PPARGC1A,PRDX3,PTEN,ROCK1,SCD,SHC1,SLC27A2,SLC27A5,SOAT1,SOD2,STAT3,TIMP1,TP53,UCP2,VCAM1,VEGFC |
| CLDN7 | 0,302 | other | | -0,130 | -0,129 | 0,000 | 2,77E-11 | 1,05E-09 | ALDH3A1,ANXA3,APOE,ARHGDIB,ASAP1,C1S,C3,CCL20,CKAP4,DENND5A,F3,FOSL1,FZD2,GLS,GRINA,HLA-B,IFI44,IFI6,IGFBP7,ITPR3,LIMA1,MAGED1,MMP2,MMP9,MRPL40,MRPS15,MT1X,MUC1,MUC13,MX1,NETO2,NNMT,PHGDH,PLAAT3,PRKCZ,PRNP,PRSS8,PSMG2,RNASET2,RPP40,RRAS2,SERPINB1,SLC35B1,SLCO2A1,TIMP2,TJP1,TPK1,UBE2L6,WIPI1,ZCCHC14 |
| NR1H3 | -0,242 | ligand-dependent nuclear receptor | | 0,563 | 0,571 | 0,001 | 3,56E-11 | 1,33E-09 | ABCG1,ACACA,ACOX1,ADRB2,ANGPTL3,APOA1,APOC3,APOE,APOM,ARG2,ARL4C,ATF3,AURKA,C1QA,CAT,CCL5,CCNA2,CCNB1,CCNB2,CCR2,CCR5,CCR7,CD38,CD40,CD47,CD55,CDC25B,CDK1,CDKN1B,CFB,CX3CR1,CXCL10,CYTIP,DDX60,DHRS9,FBP1,FGL2,FPR1,G6PC,GPT,GPX3,GSTP1,HK1,IFIT2,IRF7,ITGAL,ITGB3,KIF20A,LAIR1,LCP2,LSP1,LY86,LYZ,MELK,MIF,MLXIPL,MMP9,MYLIP,NOS2,NR0B2,NR1H3,PKLR,PLTP,PPARGC1A,PRC1,RB1,SCD,TAP1,THRB,TLR1,TOP2A,UBE2C,VCAN,VEGFA,VEGFC |
| IgG | | complex | | -0,499 | -0,536 | -0,005 | 3,64E-11 | 1,35E-09 | ADGRG1,ADM,AFM,AOC1,APOE,ARSB,ATP6V0D1,B2M,C3,CALR,CCL5,CCND2,CCR2,CD40,CD86,CD9,COL3A1,CXCL10,CYP27A1,DDIT3,DHCR7,DUSP1,FCGR2B,FN1,FOLR2,FOSB,GNAS,GUK1,HLA-E,HSPA5,HTR2B,IFITM1,IL13,INPP5D,ISG15,ITGAM,ITGB8,JUND,KRT10,LCN2,LGALS1,MAF,MAFB,MMP9,MS4A1,NGF,PEA15,PLS3,PPL,PPP1R15A,PRSS8,PTPN6,RND3,S1PR1,SERPINF1,ST14,STAB1,TACSTD2,TAPBP,TLR4,TLR7,TNC,TNFRSF17,TOB1,TUBB2A,UGCG,VCAM1,VEGFA,ZFP36 |

| | | | | | | | | | |
|---|---|---|---|---|---|---|---|---|---|
| ANGPT2 | | growth factor | Activated | 2,653 | 2,707 | 0,007 | 3,64E-11 | 1,35E-09 | ABCC1,ADA,ARL4C,BAG1,C1R,CALR,CASP3,CAT,CCL5,CFB,COL1A1,COL1A2,COL3A1,COMT,CRYBB3,CYBA,CYBB,DDIT3,EPHX2,EXT2,FAS,FGF7,FKBP4,FN1,HIF1A,HSPA5,ICAM1,LARP4,LEPR,LYN,MAPK10,MAPK12,MIF,MMP1,MMP2,MMP7,MMP9,NME1,NOX4,NPHS1,NPPA,P4HB,PDCD4,PDIA3,PDIA5,PDIA6,PLG,POSTN,PPARA,PTGES,RCAN1,RFC2,RHOB,RPA2,RPA3,SERPING1,SMYD5,SOCS7,SOD1,SOD2,ST13,TCN2,TGFBR2,THBS2,TLR3,TP53,TXNIP,VCAM1.VIM |
| INS | -0,118 | other | | 0,985 | 0,995 | 0,001 | 4,15E-11 | 1,53E-09 | ACACA,ACADL,ACADM,ACADVL,ADAM10,ADIPOR1,ADM,ALDH3A2,APOA1,AQP3,AVP,CAT,CCND1,CD36,CIDEC,CSHL1,CUL5,CYP17A1,DDIT3,DUSP1,ERBB3,F3,FAS,FOS,FOSB,FOSL1,FOXO1,G6PC,GABARAPL1,GRB10,HIF1A,HSPA5,ICAM1,IGFBP4,IGKC,INSR,JUND,NDUFV1,NOX4,NR1H3,NR4A1,OXT,PCK1,PLIN2,PLK2,PPARGC1A,PTGS1,SCD,SELP,SLC2A2,SMPD1,SOD1,TP53,TRIB3,UGCG,VEGFA.VLDLR.XBP1 |
| CD44 | 0,962 | other | Activated | 4,431 | 4,482 | 0,007 | 4,34E-11 | 1,59E-09 | ABCA5,ABCG2,ACKR3,ACTB,ADRB2,ANXA11,BIRC2,BIRC3,CALR,CCL19,CCL5,CCND1,CCND2,CCNG1,CCR2,CCR5,CCR7,CD36,CD44,CD63,CD69,CD8A,CDH5,CDKN1B,CDX2,CIDEC,CLEC7A,CLU,COL18A1,COL1A1,COL3A1,COL5A2,COMMD3,CTSK,CX3CR1,CXADR,CXCL6,DDIT3,DNMT1,FAS,FKBP4,FKBP5,FN1,GADD45A,IFITM2,ITGA3,ITGAV,LAMB1,LAMC1,LGALS1,MAPK8,MET,MMP2,MMP7,MMP9,MYO1B,NECAP2,NOX4,PDCD4,PECAM1,SELL,SMAD1,SPP1,TCIRG1.THBS2.THY1.TIAM1.TLR8.VCAM1.VIM.WBP2 |
| CG | | complex | | 1,585 | 1,648 | 0,006 | 4,44E-11 | 1,63E-09 | ACSL1,ACTB,ADAMTS1,ADM,B2M,BARD1,BAX,BHLHE40,BHMT,CCND2,CCND3,CD93,CDKN1B,CLU,CTSV,CXCL10,CXCR4,CYP17A1,DDX25,DHCR7,DTL,DUSP1,EFEMP1,EFNB2,ELK3,ENPP2,ESM1,F2RL1,FAS,FGL2,FHL1,FOS,FYN,FZD4,G0S2,G6PC,GASK1B,GJA1,GM2A,GTF2E1,HIF1A,HLA-A,HLA-DQA1,HPGD,HPSE,HSD11B2,HSD3B2,ICAM1,IFIT3,IGFBP4,IL33,IL4R,INHA,ITGAV,ITGB3,KHK,KLHDC4,LEPR,LGALS3BP,MAF,MCAM,MMP1,MMP2,MMP9,NOS2,NPR3,NR1H2,NR2F6,NR4A1,NRP1,NUCB2,OXT,PAPPA,PCNA,PECAM1,PFKFB3,PLXND1,PPARA,PRLR,PTGS1,RASSF2,RECQL,RORA,S100A10,S100A6,SACS,SDC1,SLC4A4,SMAD2,SMARCA1,SMARCA5,SPP1,SPRY2,SRD5A2,ST8SIA4,TAC1,TFPI2,TGFBR3,TIMP1,TM4SF1,TNFAIP6,TP53,TSPAN13,VCAN,VEGFA VEGFC |
| E2F4 | -0,023 | transcription regulator | | -1,768 | -1,762 | 0,003 | 4,53E-11 | 1,65E-09 | ALG5,ANAPC15,BARD1,BNIP3,CALM1 (includes others),CASP4,CASP6,CCNA2,CCND1,CDK1,CDKN2C,CRADD,DHPS,DROSHA,ECT2,EFHC1,EIF2B2,EXOSC8,FAS,FGF7,FOXN1,GINS1,HMGB3,HSP90B1,ID3,IER5,KIF22,LPAR1,MAD2L1,MCM3,MCM5,MCM6,MSH2,MT1G,MTHFD1,MYB,NASP,NCL,NDC80,NFE2L1,OIP5,PCLAF,PCNA,PCSK1N,PGRMC1,PLSCR1,PPARGC1A,PPP1R8,PRC1,PSAT1,PTTG1,RB1,RFC2,RPA3,RRM1,RRM2,SFRP1,SLC3A2,SMARCA5,SMC4,STAM,STMN1,TEAD4,TERT,TOP2A,TOP2B,TOPBP1,TP53,TP53BP2,TPX2,TTK,UBE2C,XPC,ZNF267,ZNF91 |
| THRB | -0,215 | ligand-dependent nuclear receptor | | -0,926 | -0,916 | 0,002 | 5,04E-11 | 1,83E-09 | ACACA,ACOX1,ANGPTL3,APOA1,APOC3,APOM,ATP1A3,CASP1,CCNB1,CCND1,CCND2,CCND3,CDKN1B,CFD,CGA,CSHL1,CSNK1A1,CTSD,CXCL10,CXCR4,CYP27B1,DCN,DDC,DIO1,EGF,EHHADH,ENG,ERBB3,F12,FGFR3,FOS,FOSL1,FOXN1,FRAT1,FRZB,FSTL1,FZD4,FZD6,G6PC,GCH1,HIF1A,ID1,IGHM,IL7,KLF9,LYZ,MAPK8,ME1,MET,MLXIPL,MMP1,MMP2,MMP7,MMP9,NME1,NPPA,NR1H3,NR4A1,PCK1,PDK4,PER1,PHYH,POR,PROM1,PTTG1,SFRP1,TGFBI,THRB,TLE2.TP53.WIF1.WNT8B.YWHAE |
| UCP1 | | transporter | Inhibited | -4,326 | -4,313 | 0,002 | 5,67E-11 | 2,04E-09 | ACACA,AGAP1,ALDH18A1,ARHGAP33,ASNS,BCKDHA,BCKDHB,BTF3,BZW2,CAPN3,CASP3,CLCN3,CTH,CTSV,CXCL10,CYB5R1,DDIT3,DDX58,DKK3,ESRRG,FGF7,GADD45A,GARS1,GHITM,GNMT,GOT1,GPX3,GSR,GSTO1,GSTZ1,IFIT2,IFIT3,LRRC8D,MTHFD1,MTHFD2,NDUFA10,NDUFS3,OAS3,PCK1,PCK2,PHGDH,PSAT1,PSD3,RAB40B,RNASEL,SDHB,SHMT1,SHMT2,SLC3A2,SMYD2,SND1,TLR3,TLR7.TRIB3.TTLL7.TXN2.UCP2.UQCRC1.UQCRC2.USH1C |
| mir-21 | | microRNA | Inhibited | -3,516 | -3,577 | -0,007 | 5,83E-11 | 2,09E-09 | AOAH,ARF4,C1R,CASP4,CCL19,CCNA2,CCNB1,CCND1,CD180,CDH5,CLPB,CLU,COL1A1,COL3A1,CRY2,CSE1L,CXCL10,DDAH1,DLGAP5,ECT2,FAS,FGF1,FGL2,FN1,FUBP1,GBP2,HIPK3,ICAM1,IDO1,IFI16,IGHM,IL18,IL6R,INPP5D,IRAK1,IRF1,ITK,LEPR,LTB,MMP2,MMP9,MSH2,MYD88,NME1,NMT2,NOS2,OAS2,OAS3,PBK,PDCD4,PECAM1,PIP4K2A,PPARA,PRC1,PTEN,RECK,RHOB,SIGLEC1,SLPI,SMAD2,SMAD7,SMARCA4,SOD2,SPRY1,SPRY2,SSR1,STAT1,STAT3,STMN1,TACC3,TAP1,TERT,TIAM1,TLR1,TLR2,TLR4,TOP2A,TP53,TPM1,TREM2,TRIM37,UBA7,UBE2L6,VCAM1,VRK1,YWHAB,ZWILCH |

| | | | | | | | | | |
|---|---|---|---|---|---|---|---|---|---|
| IL1RN | | cytokine | Inhibited | -5,934 | -5,987 | -0,008 | 5,97E-11 | 2,13E-09 | ATF3,BTN3A1,CCL20,CD44,CTSS,CXCL1,CXCL11,DDX58,GBP1,HERC6,HLA-DQB1,ICAM1,IFI27,IFI44,IFI44L,IFI6,IFIH1,IFIT3,IFIT5,IGFBP4,IL32,IRF1,IRF7,IRF9,ISG20,LAMP3,MMP1,MMP9,MX1,MX2,NGF,NOS2,NR0B2,OAS1,OAS2,OAS3,RARRES1,RSAD2,RTP4,S100A9,SAMD9,SLC15A3,SPP1,TAC1,THEMIS2,TIMP1,TNFSF10,TRIM22,UBD,VEGFA,VEGFC |
| TLR9 | | transmembrane receptor | Activated | 4,009 | 4,077 | 0,008 | 6,00E-11 | 2,13E-09 | ARG2,ATF3,CAST,CCL5,CCNA2,CCNB1,CCND1,CCR7,CD27,CD300A,CD38,CD40,CD69,CD83,CD86,CDKN1B,CISH,CTH,CXCL10,CXCL6,CXCL9,DUSP1,FAS,FCMR,GADD45B,GCH1,IDO1,IFI16,IFI35,IFI44L,IFIT2,IFIT3,IFITM1,IFNA10,IL10RA,IL18,IL33,IL36A,IRF1,IRF7,IRF9,ISG15,ISG20,KDELR3,LAIR1,LCN2,MX1,MX2,NAMPT,NOS2,NR0B2,NR4A1,NSMAF,OAS2,OAS3,ORMDL2,PFKFB3,PROCR,RSAD2,S100A8,SDC1,SGPL1,SMPD1,SMPDL3A,SPRY1,SPTLC2,ST3GAL5,STAT1,TIMP1,TLR4,TLR7,TNC,TNFRSF1B,TNFSF10,TP53,UAP1,UGCG,XBP1 |
| NKX2-3 | | transcription regulator | | -1,022 | -1,021 | 0,000 | 6,43E-11 | 2,27E-09 | ACO1,ADM,ALPL,APOBEC3G,ARHGDIB,ASNS,CASP1,CCNB2,CD36,CD38,CDKN3,CEP55,CXADR,DCLK1,DDX58,DDX60,F2RL1,FAR2,GASK1B,GBP1,GBP2,GCA,GCH1,GHR,HCP5,HLA-B,HLA-C,HLA-F,IDO1,IL7R,KRT81,LYPD1,MCTP1,MFNG,MMP7,MYD88,NETO2,NT5DC2,PARP12,PHGDH,PLD1,PLSCR1,POSTN,PSAT1,PSMB8,PSMB9,PTPRE,RPL23,RTP4,SAMD9,SERPINE2,SHMT2,SLC25A28,SMAD7,SP110,SRPX,SRPX2,STAT1,SYT11,TAP1,TAP2,TFF3,TFPI2,TIPARP,TNFRSF10D,TRIM21,TRIM22,TRIM5,UBA7,UBE2L6,VCAM1,XAF1,ZMAT3 |
| SP110 | 0,627 | transcription regulator | Inhibited | -2,979 | -2,994 | -0,002 | 6,93E-11 | 2,44E-09 | ANXA5,AQP3,ATF3,BST2,CD74,CLU,CTSB,CXCL10,CXCL11,DICER1,F3,FLII,FUS,GRN,IFI27,IFI6,IFIH1,IFIT3,IFITM1,IL18,IL1R1,IL4R,IL6R,IRF9,ITGAV,LY96,MAOA,MX1,MYH9,NCK1,NFIL3,OAS1,OAS3,PANX1,PERP,PLSCR1,PRKCA,PTPN12,RAC1,RPA1,RXRA,SMAD2,SMARCA4,SMARCD3,SOX4,SP3,STAT1,TGFBR3,TIA1,TIMP1,TNFRSF21,TRIM5,TXNIP,WWOX |
| ARNT | | transcription regulator | | 0,030 | 0,070 | 0,008 | 7,27E-11 | 2,54E-09 | ADAM17,ADM,AHR,ALDH3A1,ALDH7A1,BHLHE40,BHLHE41,BNIP3,CAV1,CCND2,CCR5,CCR7,CD81,CDCP1,CLDN1,CTSD,CXCR4,CYP1B1,CYP2A6 (includes others),EIF6,ENG,ENO1,ESRRG,G6PC,GSTA1,GZMH,HIF1A,HNF1A,HNF4A,ID2,IGHM,INSR,ITGAV,ITGB3,KIF20A,MPP1,MYL6,NDUFS2,NDUFS3,NDUFS4,PCK2,PRF1,S1PR1,SELL,TP53,TPI1,UBE2C,VEGFA,VIM |
| MAPT | -0,922 | other | | -1,546 | -1,542 | 0,002 | 7,30E-11 | 2,54E-09 | ABCG1,ABCG2,ACAT1,ACO2,ACTB,ACTG1,ACTN1,ALB,ALDH5A1,ANXA5,ARF4,ATOX1,ATP1A3,ATP5F1D,ATP6V0D1,ATP6V1E1,BACE1,BASP1,C1QBP,C1R,C1S,C3,CALML3,CAMK2A,CAMK2G,CAP1,CAPN1,CAPN2,CASP3,CAST,CCDC25,CCNB1,CCND1,CDK5,CDKN1B,CLTB,CORO1A,CRYM,CTSD,CYFIP2,DLD,DNAJC6,DNM1L,DPYSL2,DSTN,DYNC1LI1,ENO1,ERMP1,FGFR3,FOS,FUZ,FYN,GAD1,GOT2,GPM6B,GSTP1,GUCY1B1,HK1,HSPA4L,HSPA5,HSPA8,KLC1,MAPK8,MAPT,MTHFD1,MTOR,NAPA,NDUFS3,NDUFS4,NDUFS8,NFKB1,NME1,NOS2,NTRK2,OGDH,PEA15,PEBP1,PKM,PRDX3,PRNP,PVALB,QDPR,RPL23,RPS16,SEC31A,SH3GL2,SLC1A3,SOD1,STMN1,STUB1,STXBP1,SUCLG1,THY1,TLN2,TOP2B,TPI1,TUBA1A,TUBA1B,TUBB,TUBB2A,TUBB4B,TUBB6,TUFM,TXN,UCHL1,UQCRC2,VAMP2,YWHAE |
| STAT5A | -0,216 | transcription regulator | | -1,906 | -1,894 | 0,002 | 7,41E-11 | 2,57E-09 | A2M,AGR2,ALDH3A1,APOE,BAX,BCL11A,BCL2A1,BCL6,BIRC3,CASP3,CASP4,CASP6,CASP8,CCND1,CCND2,CCND3,CCR5,CD24,CDC7,CDKN1B,CISH,CYP1B1,CYP27B1,CYP2J2,ESR1,EZH2,FAS,FCER1G,FOS,FOSB,GADD45A,GADD45G,GASK1B,GCHFR,GCNT3,GDPD3,GJB1,GZMA,GZMK,HOXB6,HSD3B2,ICAM1,ICAM2,ID1,ID2,IFITM2,IL2RB,IL6R,IL7R,IRF1,KLRB1,LCN2,LTB,LY6G6D,MAF,MMP7,MUC13,NR1H3,OAS1,PLAAT4,PRF1,PRLR,PSMB8,QPCT,QPRT,RIN2,RORA,RPA1,SELENBP1,SERPINA3,SH3GL2,SLC2A10,SLC34A2,SLC7A7,SLC7A8,SOCS2,TACSTD2,TFF3,TGFBI,TGFBR2,TM4SF1,TNNC1,TNNI1,TP53,TSPAN13,TSPAN8,TXNIP,WARS1,ZFP36 |
| CLOCK | 0,011 | transcription regulator | | -0,386 | -0,392 | -0,001 | 8,74E-11 | 3,00E-09 | ACOX1,ARHGEF10,ARHGEF28,ARNT2,AVP,BHLHE40,BPHL,C1QBP,CBFB,CCND1,CD86,CPD,DDIT3,DHTKD1,DNAJB9,DNAJC16,EFHC1,EIF4G2,ELOVL6,ESYT1,FGF1,FKBP5,FZD6,GADD45A,GADD45B,GCC2,GIMAP4,GJA1,GLIPR1,GNPTAB,GPR3,GTF2E1,HIPK3,HLA-DQB1,HMGCS2,HSD17B8,ICAM1,ID1,IHH,ITM2C,KANK2,KAT5,KIF20A,LPIN1,MAP2K3,MAPK8,MGAT2,MGP,MMP7,MOXD1,MSMO1,NDUFA3,NFIL3,NOCT,NR0B2,OAS3,PER1,POMGNT1,POR,PPARA,PRDX3,RAP2B,RIOX2,SCD,SLC12A3,ST3GAL5,ST6GALNAC4,TBRG4,TEF,TMED3,TMX1,TNFRSF21,TOR1B,TP53BP1,TPST1,TRIL,UBL3,USP2,VSTM4,WNK1,WT1,ZZZ3 |

| | | | | | | | | | |
|---|---|---|---|---|---|---|---|---|---|
| 15-deoxy-delta-12,14 -PGJ 2 | | chemical - endogenous mammalian | Inhibited | -2,550 | -2,575 | -0,003 | 1,22E-10 | 4,17E-09 | BAX,BIRC2,BIRC3,CASP3,CAV2,CCL5,CCNB1,CCND1,CCND2,CCNG1,CD200,CD36,COL1A1,COL1A2,CTSB,CTSD,CXCL10,CXCL11,CXCL9,CYBB,DDIT3,DDX58,DNAJB1,EHF,ESR1,FABP2,FKBP4,FN1,FOS,GADD45A,GCLC,GCLM,GSR,GSTP1,HLA-DRA,ICAM1,IRF1,ITGAM,MMP1,MMP9,MUC1,MYB,NFE2L2,NFKB1,NOS2,NOX4,P4HA1,PARP1,PDGFRA,PLA2G4A,PLAAT4,PTGES,PTGES2,RORA,SOAT1,SOD1,SPP1,STAT1,TBXAS1,TG,TMPRSS2,TP53,VCAM1,VEGFA |
| CTNNB1 | 0,011 | transcription regulator | | 1,042 | 1,104 | 0,007 | 1,24E-10 | 4,22E-09 | ABCC1,ABCD4,ACACA,ACAT1,ACTB,ACTR3,ADAM17,AFP,AGRN,AHR,ALDH1A3,ALDH3A2,ANXA1,AURKA,BACE1,BHLHE40,BMP7,BOLA1,BTF3,CALM1 (includes others),CAPN1,CCNA2,CCND1,CCND2,CD36,CD44,CD47,CD99,CDH11,CDKN1B,CDX2,CENPM,CFD,CIDEC,CLU,CNN2,COL1A1,COL4A1,COL4A2,CPT2,CRAT,CRIP1,CTSF,CTSZ,CXCL12,CXCL6,CYB5A,CYFIP2,CYP1B1,CYP2A6 (includes others),CYP4F12,DDIT3,DDX50,DIO1,DNAJC6,DOK1,DPEP1,EHD1,ELMO1,ENO3,ENPP2,ERBB3,EVX1,EYA2,FAS,FBRS,FCER1G,FN1,FOS,FOSL1,GAD1,GADD45B,GADD45G,GCH1,GHR,GJA1,GJB1,GLI3,GLUL,GPR137B,GRAMD4,GTPBP6,HHEX,HMG20B,HMGCS2,HNF1A,HNF4A,HOXB7,HTATIP2,HTRA1,ID1,ID2,ID3,IDO1,IFITM1,IGHM,IHH,IL13,IL1R1,IL7,IMPDH2,INHA,IRF8,ITGAL,ITGB2,JAM2,JCHAIN,KITLG,KRT7,LAMB1,LCK,LHB,LIPA,LSP1,MAN2B1,ME1,MECOM,MME,MMP1,MMP2,MMP7,MMP9,MPZL2,NAT8,NKX2-5,NOP53,NOS2,NPHS2,NR4A1,OAT,OGG1,OSBPL1A,PARD3,PCCA,PCNA,PECAM1,PFN1,PLS3,PMP22,PRDM14,PRX,PTEN,QPCT,RAB3A,RGN,S100A4,S100A8,SCD,SCRN1,SERPINA3,SERPINA5,SERPINE2,SFN,SFRP1,SGK1,SLC12A2,SLC1A5,SLC25A46,SLC26A2,SLC39A14,SLC9A3R1,SMAD7,SNRNP70,SORBS3,SOX4,SPP1,STAT3,STXBP1,SYK,TCF12,TCF4,TERT,TIA1,TIMP1,TJP1,TMSB15A,TNC,TNFRSF11B,TNFSF10,TNNC1,TP53,TRAPPC2L,TSC22D1,TSPAN8,UGCG,UPB1,VAV1,VCAM1,VCAN,VEGFA,VIM,WIF1,WT1,XAF1,ZEB2 |
| Alpha catenin | | group | Inhibited | -5,241 | -5,307 | -0,010 | 1,34E-10 | 4,55E-09 | ADAMTS1,BIRC3,CCN3,CDH11,CDKN1B,COL15A1,COL1A1,COL1A2,COL3A1,COL5A1,COL5A2,COL6A3,CTSB,CTSV,CXCL10,CXCL12,CXCL6,ELK3,ENG,FSTL1,ICAM2,IGFBP4,IGFBP6,IGFBP7,IL2RG,IL36A,IRF1,ITGAM,LUM,LYN,LYZ,MMP2,MMP9,PDGFRA,S100A8,S100A9,SGK1,STEAP4,THBS2,TIMP1,TNC,TNFAIP2,TNFAIP6,TSPAN8,VCAM1,VIM,WIPF1,ZEB2 |
| Ifn gamma | | complex | Activated | 3,516 | 3,548 | 0,005 | 1,44E-10 | 4,86E-09 | ADAR,APOBEC3G,BAG1,CASP1,CASP3,CCL5,CD86,CDKN1B,COL1A1,CSE1L,CXCL10,CXCL9,CYBB,ESR1,FAS,FN1,GBP1,HLA-DRA,ICAM1,IFI44L,IGFBP4,IL32,IL33,IRF8,LY96,MAP4K4,MMP9,MYD88,NOS2,NR4A3,PCK1,PSME1,SH2D1A,SOCS7,STAT1,TACR2,TERT,TLR3,TLR4,TNFSF10,VSIG4,XAF1 |
| estrogen receptor | | group | Inhibited | -3,193 | -3,194 | -0,000 | 1,59E-10 | 5,34E-09 | ABCG1,ABCG2,AGO1,ANXA1,ANXA9,AXL,BAX,BMP7,C3,CALB1,CAPG,CAV1,CAV2,CCND1,CCND2,CD24,CD44,CD59,CD99,CDH11,CDH15,CDH3,CDH5,CDH9,CLDN3,COL4A1,COL4A2,COL5A1,CXCL1,CXCL12,CXCR4,CYP2B6,DICER1,DNMT1,ERBB3,F12,FGF1,FGF13,FGFR3,FN1,FOS,HOXC10,ICAM1,ICAM2,ITGA3,KRT19,KRT7,LTF,LYN,MAPK12,MAPT,MMP1,MMP9,MSN,MUC1,NEDD9,PCDH1,PLEK2,POR,PRLR,SELL,SLC9A3R1,SMAD1,SPARC,TERT,TFF3,TGFBR2,TGFBR3,TIMP1,TIMP2,TLR4,TNC,TP53,VEGFA,VEGFC,VIM,XBP1 |
| MYCN | | transcription regulator | Inhibited | -2,938 | -2,919 | 0,002 | 1,62E-10 | 5,45E-09 | ABCB7,ABCC1,ABCC3,ABCD4,ABCG2,ACTB,ACTG1,ARPC1B,B2M,BASP1,BAX,BID,CAV1,CCND1,CCND2,CCNH,CDH11,CDH5,CDKN1B,CHPF2,CKAP4,CLU,COL18A1,COL1A1,COL4A1,COL5A2,DKK3,EED,EIF4G2,EZH2,FAU,FBN1,FN1,HLA-A,ID2,IGFBP7,ITGA2,ITGA3,KDM5B,LGALS1,MMP2,MXI1,MYH9,MYL12A,NCL,NDRG1,NME1,NUCB1,P3H1,PDIA4,PGRMC2,PHB,PHGDH,PRKN,PSMB6,PSMB7,PTDSS1,RBBP7,RPL13A,RPL23,RPL28,RPL3,RPL35A,RPL8,RPLP2,RPS16,RPS19,RPS20,RPS24,RPS3,S100A10,SLC16A7,SPARC,TERT,TIMP2,TM6SF1,TMED9,TMPRSS2,TP53,TPI1,TPM1,TUBA1B,TUBB,TUFM,UCHL1,VIM,ZEB2 |

| | | | | | | | | | |
|---|---|---|---|---|---|---|---|---|---|
| levodopa | | chemical - endogenous mammalian | Inhibited | -2,350 | -2,336 | 0,001 | 1,70E-10 | 5,68E-09 | ACKR1,ADAR,ADD3,AGPAT1,ALPL,AMZ2,ARAF,ARC,AREL1,ARG2,ASPSCR1,AXL,BAX,C1GALT1C1,C1QA,C8B,CACNA2D3,CACNB2,CAP1,CAPG,CASP3,CBR3,CCND1,CCS,CD200,CD63,CHST8,CIB2,CLN6,COA4,COL4A1,COMT,COTL1,CRY2,CRYM,CTNNBIP1,CTSB,CTSZ,CXCL12,CYP2A6 (includes others),DDOST,DMTN,DUSP1,EIF4EBP2,EIF4G2,EIF6,FAAH,FAH,FAM3A,FCGRT,FKBP5,FOS,FOSB,FOSL1,FOXO1,FRAT2,FTL,FXYD5,GABARAPL1,GADD45B,GLB1,GNAS,GNB5,GNG7,GOLIM4,GPC3,GPD1,GPS1,GRB10,GUCY1A1,HCLS1,HECW1,HEXB,HLA-A,HTATIP2,HTR1B,IDO1,IFNAR2,IQGAP1,ITGB1BP1,KCNA6,KDM5B,KLF9,KYAT1,LAPTM5,LRRC1,LYPD1,LZTS3,MAP3K1,MAPKAP1,MAT2A,MEF2C,MEIS2,MRPL48,MSMO1,MSRB1,MYO1B,NAB2,NAGLU,NASP,NCF4,NDUFS4,NEDD4L,NIT1,NOCT,NUAK1,PALM,PAOX,PARD6A,PARM1,PCYT2,PDCD4,PDZD2,PER1,PFKP,PGPEP1,PKP2,PLEK,PLEKHO2,PLK2,PRKD3,PTPRT,PVALB,RAB8A,RASGRP1,RENBP,RESF1,RFC2,RUNDC3B,S100A10,SCRN1,SDR39U1,SFXN3,SH3BGRL3,SH3GL2,SHC1,SLC22A17,SLC22A18,SLC25A44,SMUG1,SOBP,ST3GAL5,STAB1,STXBP1,SUB1,TAC1,TBP,TCF4,TCIRG1,TIAM1,TLE1,TMC6,TMED10,TP53,TRIB3,TRIOBP,UAP1L1,UBFD1,UBL3,UBXN6,UPB1,VLDLR,VPS13A,VTI1B,WDR1,WIPF1,ZEB2,ZMYND8,ZNF76 |
| FGF2 | | growth factor | | 0,990 | 1,036 | 0,005 | 2,01E-10 | 6,68E-09 | ACE,ADGRL2,ADM,AFP,AKR1B1,ALB,ANG,AQP3,ARC,BAX,BCL6,C1QL1,CALB1,CAPZA2,CAV1,CBFB,CCL20,CCN3,CCND1,CCND2,CCND3,CDH11,CDKN1B,CDX2,CIDEC,COL1A1,COL1A2,COL3A1,CTSK,CXCL12,CXCL6,CXCR4,DCN,DDIT3,EFNB2,ENO1,ENPP2,F3,FAH,FAS,FBN1,FDX1,FGFR3,FLNA,FN1,FOS,FOSL1,FOXO1,FOXO4,FUT3,GADD45A,GBP1,GCH1,GJA1,GPNMB,HIF1A,HPSE,ICAM1,ID2,ID3,IGFBP4,ITGA3,KL,KRAS,MACROD1,MAPT,MCAM,MET,MGP,MIF,MMD,MMP1,MMP2,MMP9,NKX2-5,NOS2,NR4A1,PCNA,PDGFRA,PLK2,PPP1R12A,PRKCD,PTPRE,PTTG1,RB1,S100A10,S100A4,S100A8,SCD,SDC1,SFRP1,SHBG,SLC34A1,SPARC,SPP1,SPRY1,SPRY2,ST3GAL1,ST3GAL4,TAC1,TEAD4,TGFBR3,TIMP1,TNFSF10,TOP2A,TP53,VCAM1,VEGFA,VEGFC,VIM,VWF,WNT10B,XRCC5,ZEB2 |
| ID3 | 0,581 | transcription regulator | | -0,243 | -0,248 | -0,001 | 2,97E-10 | 9,73E-09 | ADGRG1,ADM,BCL6,BMP7,CCL5,CCNB1,CCND2,CCNG1,CCR5,CCR7,CD44,CD83,CDC25B,CDK1,CDKN1B,CDKN2C,CSF2RB,CXCL1,CXCR4,DUSP1,DUSP22,ELOVL6,FCER1G,FOXO1,GADD45B,GADD45G,GNL3,HIF1A,ICAM1,ID3,IFI16,IFNGR1,IFNGR2,IGFBP4,IL10RA,IL2RB,IL4R,IL6R,IL7R,IRF8,ITGB3,LEPR,LGALS1,LTB,MAF,MMP2,MMP9,MYB,NPPA,NR4A1,NR4A3,PIK3C2B,PIK3IP1,RORA,SDC1,SELL,SH2D1A,SMAD7,SOX13,SOX4,STAT4,TCF4,TGFBR3,TNFSF10,TP53,TRAF5,VCAM1 |
| CCN1 | | other | | 1,677 | 1,715 | 0,007 | 3,64E-10 | 1,18E-08 | BAX,C3,CCND1,CCND2,COL1A1,COL1A2,CXCL10,DVL1,ESR1,FCER1G,FN1,HIF1A,IL10RB,IL18,ITGA2,ITGA3,ITGAV,ITGB3,MIF,MMP1,MMP2,MMP9,NOS2,NR4A1,PECAM1,SENP2,SFRP1,TIMP1,TIMP2,TLE1,TLR4,TLR7,TNFSF10,TOLLIP,TP53,VEGFA,VEGFC,WIF1 |
| PRDM1 | 0,092 | transcription regulator | | -1,747 | -1,765 | -0,002 | 3,84E-10 | 1,24E-08 | ACOX2,ADA,ADGRG1,AFP,AIM2,ALPL,APOM,AQP3,ARG2,ATP9A,BCL2A1,BCL6,CCL20,CCR7,CD44,CD53,CD74,CD86,CFH,CLDN1,CXCL6,CXCR4,ERAP1,ESR1,FAR2,FGR,FOS,FOSL1,FZD2,GAD1,GNLY,GPX5,HLA-DQA1,HLA-DRA,ID3,IDO1,IGHM,IL7R,IRF8,JCHAIN,KRT81,LGALS1,MAFB,ME1,METTL7A,MMP25,MS4A4A,NOS2,PLA2G4A,PLAAT4,PLAC8,PRF1,PSMB10,PSMB8,RASSF2,REG1A,RSAD2,S100A8,S100A9,SELL,SERPINA3,SLC16A5,SLC2A5,ST14,SWAP70,TAPBP,TNFAIP6,TNFRSF10D,TNFRSF17,TNFSF10,TREH,TRIM38,XBP1,XRCC5 |
| IFNA1/IFNA13 | | cytokine | Activated | 4,826 | 4,860 | 0,007 | 4,24E-10 | 1,37E-08 | CCL8,CD40,CD69,CD83,CD86,CXCL10,CXCL11,CXCL9,IFI27,IFI6,IFIH1,IFIT2,IFITM1,IRF1,IRF7,ISG15,MX1,OAS1,OAS2,PLAAT4,PLSCR1,RSAD2,SIGLEC1,STAT1,STAT4,TLR3,UBE2D1,UBE2E1,UBE2L6 |
| IL1A | | cytokine | Activated | 3,666 | 3,722 | 0,007 | 4,44E-10 | 1,43E-08 | ABCC2,ADAMTS1,ALDH1A3,ALDH3A2,ATF3,BAX,BIRC2,BIRC3,BTN3A3,CA2,CCL20,CCL5,CCL8,CCR2,CD40,CD44,CD83,CD86,CTSK,CXCL1,CXCL10,CXCL12,CXCL6,CYP17A1,CYP2B6,CYP3A4,DPP4,F2RL1,FAM89B,FAS,FGF7,FOS,FOSB,FOSL1,GBP1,GCH1,HSD11B2,ICAM1,IFNGR1,IFNGR2,IK,IL13,IL18,IL32,IRAK1,IRF1,ITGAV,ITGB3,JAM2,KITLG,LCN2,MCAM,MMP1,MMP2,MMP9,NFKB1,NGF,NOS2,NR2F2,PDZK1IP1,PLA2G4A,PTGES,RHCG,S100A8,S100A9,SERPINA3,SOD2,SPARC,SPP1,SSTR2,TAC1,TLR2,TNFRSF11B,VCAM1,VEGFC |

| | | | | | | | | | |
|---|---|---|---|---|---|---|---|---|---|
| AR | 0,022 | ligand-dependent nuclear receptor | | 1,854 | 1,912 | 0,006 | 4,72E-10 | 1,51E-08 | ABCG1,ACTR3,ADM,AQP3,ATP1A3,AZGP1,BASP1,C3,CA4,CALU,CAMK2A,CAST,CAV1,CAV2,CCNA2,CCND1,CCT3,CDC25B,CDH11,CDK14,CEND1,CGA,CKAP4,CLCN1,CLDN3,COL3A1,CTSO,CXCL12,CYC1,CYP17A1,DCTPP1,DHCR7,DNAJB9,DNM1L,DSG2,EBP,ELMO1,EPHX1,ERBB3,ESPL1,FAM89B,FGF7,FKBP4,FKBP5,GJA1,GPD1,GPX5,GSTP1,GUCY1A1,HOPX,HSD3B2,HSPA5,IL1R1,IMPDH2,INHA,INPP4B,ITGA2,KCNJ2,KIF22,KITLG,LAMA5,MAD2L1,MAOA,MCM2,MECOM,MEF2C,MME,MRAS,MSMO1,MT1X,MTHFD2,MUC1,MYB,MYL3,MYO1B,NAP1L1,NDRG1,P4HA1,PBK,PCBD1,PCSK1N,PDGFRA,PDIA5,PEMT,PER1,PFKFB2,PIGR,PLOD2,PLS3,PRKCA,PRKCD,PTEN,PTGS1,RARRES1,RGN,RHOB,SCN4A,SERPINA3,SERPINA5,SGK1,SHBG,SLC22A4,SLCO3A1,SMTN,SPP1,SRD5A2,STAT1,STK39,TACC2,TACSTD2,TBXAS1,TERT,TFF3,TGFBR2,TIMP2,TJP1,TMPRSS2,TNFAIP2,TNFSF10,TNNT2,TPD52L1,TPX2,TSPAN8,UBE2C,VCAN,VEGFA,VIM,WIPF1,WT1,XBP1,ZWINT |
| IRF3 | 0,066 | transcription regulator | Activated | 4,689 | 4,747 | 0,008 | 4,81E-10 | 1,53E-08 | ABCC2,ADAM9,ADAR,AHNAK,ANOS1,ANXA4,APOBEC3B,ARG2,B2M,B4GALT5,BIRC2,BIRC3,CCL19,CCL5,CD40,CD58,CD69,CD86,CDH11,CLIC4,CXCL1,CXCL10,DDX58,DDX60,FAS,FGF1,FN1,GBP1,HLA-F,IFI16,IFI27,IFI44,IFI6,IFIH1,IFIT2,IFIT3,IFNA10,IL15,IRF1,IRF7,ISG15,ISG20,LCN2,MFAP2,MMP9,NOS2,OAS1,OAS2,OAS3,PAH,PARP12,PHF11,PLAC8,PLAG1,PNP,PRNP,PTMS,RSAD2,STAT1,TAP1,TDRD7,TLR3,TLR4,TNFSF10,TPST1,UBE2L6,VIM |
| FOXO4 | -0,393 | transcription regulator | | -1,622 | -1,632 | -0,002 | 5,07E-10 | 1,61E-08 | ACACA,ANXA3,BCL6,BNIP3,C1QA,CASP8,CAV1,CCND1,CCND2,CDH5,CDKN1B,COL4A1,COL4A2,ELOVL6,ENPEP,FYN,GADD45A,GADD45B,HIF1A,IDI1,IGFBP7,IL10RB,IL1R1,ITGAM,ITGB2,LCP2,LEMD3,MMP9,NAMPT,OVOL1,PRKCD,PTGS1,PTPRC,SCD,SELL,SGK1,SOD2,SPTLC2,VCAM1,VEGFA |
| TSC2 | -0,151 | other | | -0,296 | -0,328 | -0,005 | 5,44E-10 | 1,72E-08 | A2M,ANXA1,ANXA2,ARC,ATF3,ATP6V1B2,ATP6V1D,BAX,CASP1,CCND1,CCND2,CCND3,CDKN1B,CRIP1,CST6,CXCL10,DDIT3,EMP3,FOS,FZD2,GGA2,GPNMB,HIF1A,HMGCS2,HSPA5,MAOB,METRN,MGP,NEU1,NFE2L1,NIBAN1,NSDHL,OGG1,OLFM4,PDGFRA,PDK4,PLP2,PRKCA,PSMB5,PSMD1,PTEN,PVALB,S100A11,S100A6,SCD,SMPDL3A,SOD2,STAT1,UCP2,ZFP36 |
| SOCS1 | 0,094 | other | Inhibited | -3,852 | -3,902 | -0,007 | 5,57E-10 | 1,74E-08 | A2M,BAX,BCL2A1,BID,CCND1,CCND2,CCND3,CCR7,CD1D,CD40,CD44,CD69,CD86,CISH,CXCL10,DDX58,DUSP1,FAS,FCER1G,FKBP5,FOS,ICAM1,IFI16,IFI44,IFIH1,IFIT2,IFIT3,IL15,IL2RB,IL4R,IRF1,IRF7,ISG15,ISG20,MMP1,MUC1,MX1,MYD88,NOS2,OAS1,OAS2,RALGDS,RB1,RSAD2,STAT1,STAT3,TLR3,TRIM22,TXN,VAV1,VCAM1 |
| KDM5B | 0,407 | transcription regulator | Inhibited | -4,162 | -4,196 | -0,005 | 5,57E-10 | 1,74E-08 | ARL6IP5,ASF1A,AURKA,CAV1,CCNB1,CCND1,CDK1,CDKN1B,CLDN1,CYB5A,DDIT3,DLGAP5,DPY19L1,ECT2,EHD1,EIF5B,ERBB3,ESR1,FHL1,GADD45A,GCA,HACD1,HSD17B8,ISG15,ITGB1BP1,LGALS3BP,MCAM,MCM2,MCM3,MIA3,MPC1,MT1F,MT1X,NDC80,NEDD9,NKX2-5,OIP5,PBK,PLS3,PRLR,PSD3,PSIP1,RECQL,RNASET2,SNRPD1,SWAP70,TMEM14A,TOP2A,TTK,TUBB2A,XBP1 |
| NOS2 | -0,356 | enzyme | | 1,864 | 1,873 | 0,001 | 5,63E-10 | 1,76E-08 | ACO1,ADH5,ALDH3A1,AVP,AZGP1,BAX,BCL2A1,BNIP3,CASP3,CD14,CD44,CD53,CD86,CFD,COL4A1,COX6A2,FAS,FOXO1,G6PC,GJA1,GPD1,GPT,HIF1A,HMGB2,IFIT3,IL32,IRAK1,ISG15,KRT4,LCK,LCN2,LGALS3BP,LPAR1,MAPK8,MCM5,MMP9,MYL3,NKX2-5,NOS2,NPPA,PC,PCNA,PERP,PIGR,PLSCR1,PPARGC1A,PRF1,PTGS1,S100A8,SERPINA3,SFXN1,SLC1A5,STXBP1,THY1,TIMP1,TKT,TLR4,TNFSF10,TNNC1,TNNT2,TOR3A,TSPYL2,TYROBP,VEGFA |
| PI3K (complex) | | complex | Activated | 1,998 | 2,050 | 0,007 | 5,79E-10 | 1,80E-08 | ADIPOR1,ADRB2,ALOX5AP,ATF3,BIRC3,CASP3,CCNB1,CCND1,CCND2,CCND3,CCT6A,CD2,CD44,CD55,CD86,CDKN1B,COL1A1,CTCF,CTSD,CXCL10,CXCR4,CYBB,DDIT3,DUSP1,FHIT,FLI1,FN1,FOS,FOSL1,FOXO1,GCLC,GCLM,GJA1,GSTA1,GSTP1,HIF1A,HK1,ICAM1,IL33,IL7R,INHA,IRF2,ITGAM,ITGB2,KRAS,LCN2,LSP1,MCAM,MEF2C,MMP1,MMP2,MMP9,MRC1,MTHFD2,NCL,NFE2L2,NME1,NOS2,NOX4,NPPA,OSTF1,PDCD4,PLA2G4A,POSTN,PRDX2,PRKCZ,PTPN12,RHOB,RND3,RPL23,SAMHD1,SELL,SGK1,SLC12A6,SNCG,SOAT1,SPP1,SST,STMN1,TLR4,TNFSF10,TP53,TXNIP,UGCG,VCAM1,VEGFA,WT1,XBP1 |
| HOXA10 | 0,338 | transcription regulator | | -0,375 | -0,400 | -0,003 | 6,16E-10 | 1,90E-08 | ACTB,ADM,ALOX5AP,ALPL,APOE,AQP1,ARL2,CD1C,CFD,COL15A1,COL3A1,CSTA,CTSV,CYBB,DHRS9,DPP4,ENPP2,FCER1G,FKBP3,FN1,FZD5,GJA1,GSTA3,HLA-DQA1,HLA-DQB1,ID1,ID2,IL1R1,ITGB3,KAT5,KLF9,KRT4,LCN2,LEPR,LTF,LYZ,MAPK12,ME1,MEIS2,MFNG,MSN,NCF2,NR4A1,NUP214,P4HB,PBX3,PHGDH,PIGR,PRKCD,PROM1,PTGER3,RBP1,RHOG,SCD,SSPN,ST6GAL1,STK17B,TDO2,TGFBR3,TP53,WNT10B,ZNF22 |

| | | | | | | | | | |
|---|---|---|---|---|---|---|---|---|---|
| Jnk | | group | Activated | 2,291 | 2,340 | 0,006 | 6,36E-10 | 1,96E-08 | A2M,ANXA5,BAX,BIRC3,CASP3,CCL5,CCND1,CCR7,CD40,CD44,CD55,CD69,CD83,CD86,CELF2,CLEC10A,COL1A1,COL3A1,CRIP2,CXCL12,CXCL6,CXCR4,DDIT3,DNMT1,DUSP1,F12,F2RL1,F3,FAS,FLI1,FN1,FOS,FOSB,GADD45A,GJA1,HNF4A,HSPA5,ICAM1,IL13,JUND,LHB,MMP1,MMP2,MMP9,MRC1,MSH2,NFE2L2,NOS2,NOX4,NR4A1,PLG,PLSCR1,PON2,POSTN,PPM1D,PTEN,PTGES,RARG,RBP1,RXRA,SOD2,STRA6,TBP,TERT,TGFBR2,TIMP1,TP53,VCAM1,VCAN,ZFP36 |
| GDF2 | -0,204 | growth factor | | 1,069 | 1,123 | 0,009 | 6,46E-10 | 1,98E-08 | ABCG2,ANKH,ANXA3,BID,CAV1,CDH11,CDH6,COL1A1,CRABP1,CXCL12,CXCR4,EFNB2,ENG,ENPP2,FGFR3,FOS,GADD45B,HIF1A,HSPB8,HTRA1,ICAM1,ID1,ID2,ID3,MMP9,NGF,NRP1,POSTN,SEMA3B,SERPINE2,SERPINF1,SLC3A2,SMAD1,SMAD7,SOCS2,SPP1,TLR4,VCAM1,YWHAH |
| ERK | | group | Activated | 2,021 | 2,070 | 0,006 | 7,80E-10 | 2,37E-08 | ACP5,ACSL5,ACTN1,ADAM10,APOBEC3G,ARG2,ARPC2,AXL,BAX,CAV1,CCND1,CCND3,CD1D,CDKN1B,CGA,CLU,COL1A1,COL3A1,CTSK,CXCL1,CXCL10,CXCL6,DPYSL2,DUSP1,ESR1,ESRRA,EZH2,FAS,FN1,FOS,FOSL1,FOXO1,GCLC,GK,GOT1,HOXA1,ICAM1,IFNAR2,ITGAM,ITGAV,ITGB2,ITGB3,KYNU,LAMB1,LCN2,MAFB,MAFF,MAL,MAP4K4,MCAM,MMP1,MMP2,MMP9,MS4A1,MUC1,NDRG1,NFE2L2,PDK4,PECAM1,PTGES,RECK,ROCK1,RORC,SELL,SFN,SOX4,STAT1,TERT,TIMP1,TNFAIP8,TP53,TUBA1A,UGCG,VCAN,VEGFA,ZFP36 |
| PSEN1 | 0,039 | peptidase | | -1,706 | -1,713 | -0,001 | 7,93E-10 | 2,40E-08 | ABCC6,ABCG1,ABCG2,ACO2,ACSL1,ACTB,ACTG1,ALB,APOE,ARC,ARF4,ATOX1,ATP1A3,ATP5F1D,ATP6V1E1,AXL,BACE1,BASP1,BAX,C1QA,C1QB,C1QBP,C1R,C1S,C3,C3AR1,CALML3,CAP1,CASP3,CCDC25,CCL5,CCND1,CCR5,CCS,CD74,CD9,CLTB,COX8A,CRYM,CTSD,CTSS,CTSZ,CYFIP2,CYTIP,DNM1L,DPYSL2,DSTN,DUSP1,ENO1,ENPP2,EPM2A,ERMP1,FOS,FZD4,GADD45B,GJA1,GNB1,GOT2,GPM6B,GPX5,GSTP1,HK1,HLA-E,HSPA5,HSPA8,INSR,IRF7,KCNAB2,MAPT,MME,MMP9,MTOR,NDUFS4,NME1,NOS2,OCEL1,PDGFRA,PDXDC1,PEA15,PEBP1,PKM,PRC1,PSENEN,PTPRF,PVALB,QDPR,SH3GL2,SOD1,STAB1,STMN1,STXBP1,SUCLG1,TGFBR2,TIPARP,TOP2B,TP53,TPI1,TUBA1A,TUBA1B,TUBB,TUBB2A,TUBB4B,TUBB6,TUFM,TXN,UCHL1,VAMP2,WARS1,YWHAE |
| SP3 | 0,291 | transcription regulator | | 1,497 | 1,523 | 0,005 | 7,98E-10 | 2,41E-08 | ACSL1,ADD3,ADH5,AEBP1,AKAP12,ASNS,ATF3,CCL20,CCND1,CD2AP,CD40,CDC25B,CDKN1B,CDKN2C,COL1A1,COL1A2,CYP17A1,CYP46A1,DUSP1,ESR1,F3,FAS,FDX1,FOS,FOSL1,GHR,GNAI2,HPSE,HSD11B2,HSPA5,IL2RB,ITGA2,ITGB8,JAM2,KRT81,MAOB,MAT2A,MET,MMP2,MSH6,NCF2,NFKB1,PEMT,PKM,PRKCD,PRLR,PROCR,PTDSS1,PTGS1,PTTG1,RECK,RRAS,SGK1,SLC9A3,SOD2,SPARC,STAT1,STAT3,TERT,TGFBR2,TGFBR3,TIMP1,TIMP2,TLR2,TNC,TNNC1,TP53,TSPO,VEGFA,VIM |
| SAFB | | other | Inhibited | -4,013 | -4,048 | -0,007 | 8,15E-10 | 2,45E-08 | B2M,BCCIP,CCL5,CCN3,CD74,CXCL10,CXCL11,FOS,HLA-C,HLA-DPA1,HLA-DPB1,HLA-DRA,HLA-E,ISG20,LTB,MLLT3,NEDD9,PPP1R12A,PSMB9,RPL35A,S100A8,SAFB2,TBX2,TNFRSF11B,TNFSF10,UBD,UBE2B,UQCRC2 |
| SRC | | kinase | | -0,070 | -0,035 | 0,006 | 8,32E-10 | 2,50E-08 | ACAT1,CA2,CAV1,CAVIN1,CCND1,CCND2,CDK1,CGA,CKM,COL1A2,CTSB,CTSV,CXCL10,DOK1,DUSP1,ENO1,F3,FGR,FN1,FOS,FOSL1,GBP1,GJA1,HIF1A,ICAM1,ID1,LHB,MMP1,MMP2,MMP9,NFKB1,NPPA,POSTN,PRKCA,PRKCD,PTH1R,RETSAT,SPP1,STAT3,TFPI2,TJP1,VCAM1,VEGFA,VEGFC,VIM |
| IFNAR2 | 0,380 | transmembrane receptor | Activated | 3,230 | 3,257 | 0,008 | 9,69E-10 | 2,86E-08 | CXCL10,CXCL11,DDX58,IDO1,IFI44,IFI6,IFIH1,IFITM1,IL18,ISG15,LAMP3,MX2,OAS1,OAS2,PSMB10,PSMB8,PSMB9,RIPOR2,TLR3,TNFSF10,TRIM22,UBA7,UBD,UBE2L6,XAF1 |
| TGFB3 | -0,082 | growth factor | | 1,788 | 1,832 | 0,007 | 1,09E-09 | 3,17E-08 | BHLHE40,CCN3,CD59,CDH6,CDKN1B,COL1A1,COL1A2,COL3A1,COL5A1,ENG,F2RL1,FN1,FOS,FOSB,GJA1,HTRA1,ITGAV,ITGB3,JUND,MGP,MICAL2,MMP1,MMP2,MMP9,MS4A1,PALLD,PLOD2,PPARGC1A,PTEN,SCD,SDC3,SERPINE2,SMAD2,SMAD7,STAT4,TGFBR3,TJP1,TNC,TNFRSF11B,TNS1,TPM1,TSPAN13,VEGFA,ZEB2 |
| IGF1R | -0,136 | transmembrane receptor | | -2,024 | -1,976 | 0,006 | 1,25E-09 | 3,62E-08 | ACADM,ACADS,ACSL1,ANXA3,ATP5F1D,BAX,CALU,CAPG,CAVIN3,CBFB,CCL5,CD300A,CD36,CD37,CD38,CD48,CD53,CD84,CD93,CDK1,CDKN1B,CLU,COL18A1,COL1A1,COL3A1,COL4A1,COL4A2,COX6A2,CPT2,CSHL1,CTSD,DDIT3,ESR1,FOS,GPER1,GRB10,HADHA,HIF1A,HSPA5,HTRA1,ID2,IFNGR2,IGFBP4,ITGAM,KIF20A,KRT19,MYOM2,NDN,NDUFA3,NME1,NR4A1,PDCD10,PDLIM2,PECR,PKM,PROM1,RAC1,RBBP7,RFC2,RHOA,RPL3,RPL35A,RPS21,RPS24,S100A4,SCP2,SLC25A20,SLC2A2,SNCG,SST,STAT3,TGFBI,TJP1,TMEM87A,TNFSF10,TNNI1,TP53,TXN,UQCRC1,UQCRC2,VEGFA,VEGFC |

| | | | | | | | | | |
|---|---|---|---|---|---|---|---|---|---|
| GATA1 | 0,025 | transcription regulator | | -0,204 | -0,174 | 0,004 | 1,35E-09 | 3,88E-08 | ACTN1,ALOX5,ALOX5AP,AQP1,BACH2,BLVRB,BZW2,CA2,CALR,CAVIN1,CCL5,CCND1,CCND2,CCND3,CCR5,CD164,CD36,CDKN1B,CDKN2C,CPA3,CPD,CYBB,CYFIP1,DICER1,DMTN,EXOC1,EZH2,FCER1A,FHL1,FLI1,FOSB,FYB1,GNAS,GUCY1B1,GYPA,HBB,HHEX,HPSE,HSPA9,IL13,IL6R,IL7R,INHA,IRF8,ITGB3,LEPR,LYZ,MAGED1,MCM3,MCM5,MECOM,MGST2,MS4A1,MSH2,MTHFD1,MYB,NAB2,NCL,NFIL3,NFKB1,NPPA,PCNA,PECAM1,PLEK,PLXNA1,PSTPIP2,PTPN6,RRM1,RUNX3,SASH1,SELP,SLC3A2,SLC4A1,SMARCE1,SP140,SSBP3,ST3GAL6,STAT4,TAC3,TCF4,TFDP2,TFEB,TFF3,TOP2A,UAP1,VDR,VWF |
| PIK3R1 | 0,168 | kinase | | 1,401 | 1,441 | 0,007 | 1,64E-09 | 4,66E-08 | CCND1,CCND2,CD36,CDKN1B,COL1A2,CXCL10,DDIT3,DNAJB9,ENG,F3,FOS,FOXO1,GCH1,HIF1A,HSP90B1,HSPA5,ICAM1,IRF7,ISG15,MMP2,NFE2L1,NFE2L2,NOS2,PCK1,PDGFRA,PIK3CB,PKM,PTEN,SELP,SOD2,TRIB3,TXNIP,VCAM1,VEGFA,VEGFC |
| ERG | | transcription regulator | Activated | 2,804 | 2,857 | 0,008 | 1,93E-09 | 5,45E-08 | ACOT8,ACTN1,ADGRG1,ADRB2,APOC1,ARHGAP17,ARHGAP28,ARHGEF6,BIRC3,CCNA2,CD164,CD59,CHTOP,CXCL1,CXCR4,CYFIP1,DBN1,DOCK10,DOCK2,DSTN,ELMO1,EPB41L3,EXT1,EZH2,FHL1,FKBP5,FOLH1,FOSB,FYN,GAK,GDF2,GMFB,GUCY1A1,GUCY1B1,HDAC6,HPCAL1,HPGD,ICAM1,ICAM2,LAMA5,MET,MMP1,MMP9,NDRG1,NFIL3,ORC6,PECAM1,PHACTR2,PLEK,PRKCI,PSTPIP2,PTPN6,RHOA,RPA1,RRAS,RSU1,SLIT2,SOX4,SPARC,SPP1,TGFBR2,TMPRSS2,TRIOBP,UTRN,VIM,VWF,WIPF1 |
| oleic acid | | chemical - endogenous mammalian | Inhibited | -2,128 | -2,111 | 0,003 | 2,22E-09 | 6,21E-08 | ACACA,ACADL,ACADM,ACSL1,ALOX5,APOE,AQP3,BAX,CD36,CLU,CYP4F12,CYP4F2,DDIT3,DUSP1,FABP1,FABP2,FOS,G6PC,GADD45A,GADD45B,GCLC,HSPA5,ICAM1,LCN2,LHB,MLXIPL,MTTP,NOS2,NR1H3,NR4A1,PCK1,PDK4,PIK3IP1,PLIN2,PLIN3,PNPLA3,PPARA,PPARGC1A,PTEN,RXRA,SCD,SOAT1,TLR2,UCP2,VCAM1 |
| EIF2AK2 | 0,249 | kinase | Activated | 3,020 | 3,074 | 0,009 | 2,24E-09 | 6,25E-08 | ATF3,BAX,BCL2A1,BHLHE40,BIRC2,CCND1,DDIT3,DDX58,EGOT,FAS,FOS,IFI27,IFI35,IFI6,IFIT5,IFITM1,IFITM2,IRF1,ISG15,ISG20,LGALS3BP,MMP9,NEDD9,NMI,NNMT,NOS2,OAS1,OAS3,PARP12,PLSCR1,PPP1R15A,PSMB10,SAMHD1,SOD2,SP140,STAT1,TP53,TUBA1A,UBE2L6,ZNF292 |
| PPARD | | ligand-dependent nuclear receptor | Inhibited | -2,167 | -2,134 | 0,004 | 2,40E-09 | 6,64E-08 | ACAA1,ACAA2,ACACA,ACACB,ACADL,ACADVL,ACOX1,ALDH2,ALDH3A2,ALDH9A1,ANGPTL3,APOA1,APOA4,APOE,AQP3,C1QA,C1QB,CCND1,CCND2,CD36,CDK1,CDKN1B,CLEC7A,CPT2,CYP2B6,ECH1,ECI2,FABP1,FBP1,FN1,GOT2,GPD1,HMGCS2,HSD11B2,ICAM1,IFI44L,KYAT1,LRP5,MLYCD,MMP9,MYOM2,NOS2,NPPA,OCEL1,PCNA,PCYOX1,PDK4,PKLR,PLIN2,PNPLA2,PPARGC1A,PTEN,RASGRP1,RBP1,S100A6,SCD,SLC10A2,SLC25A20,SLC27A2,SLC2A2,ST3GAL5,STAT1,TLR5,TNFRSF11B,TP53,UCP2,VEGFA,VLDLR,YWHAE,ZBED5 |
| LDLR | -0,410 | transporter | | -0,838 | -0,847 | -0,003 | 2,59E-09 | 7,16E-08 | A1CF,ABCG1,ACACA,APOE,AURKA,C1QA,CCL5,CCNA2,CCNB1,CCNB2,CCND1,CCR2,CCR5,CD36,CD40,CD47,CD55,CDC25B,CDK1,CFB,CX3CR1,CYP27A1,CYTIP,DDIT3,DHRS9,DUSP1,ELOVL6,F3,FGL2,FPR1,G6PC,GADD45G,GATM,GJA1,GPT,HSPA5,ICAM1,IRF7,ITGB3,KIF20A,LAIR1,LCN2,LCP2,LSP1,LY86,LYZ,MIF,MMP2,MMP9,NR1H2,NR1H3,PCYT2,PLTP,PPARA,PPARGC1A,PPP1R15A,PRC1,PSMB7,PSMB9,SC5D,SCD,SPP1,TAP1,TLR1,TOP2A,UBE2C,VCAM1,VCAN,VLDLR |
| EDN1 | 0,329 | cytokine | Activated | 2,443 | 2,497 | 0,008 | 2,78E-09 | 7,66E-08 | ACTB,ANXA1,ANXA4,ANXA5,AVP,BCL2A1,CALCRL,CCND1,CCR7,CDK1,CDKN1B,COL1A2,COL4A1,COL5A1,CXCR4,CYBB,ENAH,FN1,FOS,FOSB,FOSL1,GJA1,GRN,HIF1A,HSPA5,HSPA8,ICAM1,IL18,INSR,ITGA2,ITGAM,ITGAV,ITGB3,LEPR,MARCKSL1,MCAM,MMP1,MMP2,MMP7,MMP9,MSN,MYL6,NOX5,NPPA,PDIA6,PLCB4,PLOD1,PPARA,PRKCA,PRKCD,PTGS1,RAB3A,SDC1,SELL,SOD1,TIMP1,TNC,TP53,VCAM1,VCAN,VEGFA,VEGFC,VIM,VWF,YWHAE |
| WT1 | -0,458 | transcription regulator | | 0,738 | 0,747 | 0,001 | 2,84E-09 | 7,81E-08 | ACTN1,ADGRE5,AHCY,ANXA11,APRT,ARL2,ASNS,BCL2A1,CAPRIN1,CCND1,CD2AP,CDKN2C,CIRBP,CKM,CLIC1,CLIC4,COL4A1,CSF1R,CTSV,CXCL10,ESR1,ETFA,EXT2,FGF1,FOSB,GRB10,GSR,HSP90B1,IDI1,IFRD2,IL2RB,INHA,INSR,IRF8,ITGAM,ITGB1BP1,JUND,KRAS,MAPK8,MMP2,MMP9,MPP1,MT1X,MYL6,NDRG1,NME1,NPHS1,NQO2,NTRK2,NUCB1,PCK2,PDE8A,PDIA4,PECAM1,PPIB,ROCK1,SALL2,SDC1,SLC35G2,SPRY1,SRSF6,ST6GAL1,TARS1,TERT,TM4SF1,TRAP1,TRIM21,TSC22D1,TYROBP,VDR,VEGFA,WARS1,WT1 |
| EPAS1 | -0,039 | transcription regulator | | -0,983 | -0,935 | 0,007 | 3,21E-09 | 8,79E-08 | ABCG2,ACACA,ACP5,ADM,AKAP12,ARG2,AXL,BHLHE40,BIRC3,BNIP3,C1QA,CAT,CAV1,CCND1,CCR2,CCR5,CDCP1,CKM,CLDN1,CORO1A,CXCR4,CYBRD1,DDIT3,DECR2,DGAT1,DMXL1,ENO1,FABP2,FHL1,FMO5,FN1,FOS,GADD45B,GAL3ST1,GCHFR,GJA1,GLS,HIF1A,HSPA5,ITGAV,ITGB3,MAFF,MANF,MIF,MYO7A,MYOM2,NAMPT,NDRG1,NFIL3,PFKFB3,PLIN2,PLOD2,PRKCA,SLC16A4,SLC29A1,SOD1,SOD2,STC2,TERT,TGFBR2,TMEM45A,TNFRSF11B,TNNI1,TP53,TPP2,TRIM21,VEGFA,WNT10B,ZNF423 |

| | | | | | | | | | |
|---|---|---|---|---|---|---|---|---|---|
| ERN1 | 0,044 | kinase | | 1,801 | 1,831 | 0,005 | 3,23E-09 | 8,80E-08 | ACACA,ANG,AP1M2,ARF4,ATF3,BLOC1S1,CDH6,CYP1B1,DDIT3,DGAT1,DNAJB9,DNAJC10,FAS,FKBP11,G6PC,GOLGA3,GOT1,GPT,HSP90B1,HSPA5,IFITM1,ITGB6,MLXIPL,MORF4L2,MPZL2,NOS2,NOX4,NPPA,NR1H3,OAS2,P3H1,PLIN2,SCD,SCN3A,SDF2L1,SEC11A,SEC24D,SLC25A28,SLC35B1,SPARC,ST14,SYT11,TBCB,TMED3,TMPRSS2,UBE2J1,VEGFA,WARS1,XBP1 |
| STAT2 | | transcription regulator | Activated | 3,499 | 3,528 | 0,007 | 3,42E-09 | 9,28E-08 | CAV1,CCL19,CCL5,CD40,CD86,CXCL10,GBP1,IFI27,IFI35,IFI6,IFIT2,IFIT3,IFITM1,IFITM2,IRF1,IRF7,IRF9,ISG15,MX1,OAS1,OAS2,PSMB8,RBX1,RSAD2,RTP4,ST13,STAT1,TNFSF10,WARS1 |
| HNRNPA2B1 | 0,159 | other | | -0,771 | -0,762 | 0,005 | 4,11E-09 | 1,10E-07 | ABCC6,ACSL5,CACNB2,CD24,CIDEC,CLDN1,CPVL,CRIP1,CTSV,DCLK1,DDR1,DSE,ERBB3,FHL1,FN1,FSCN1,GJA1,HMGB3,HNMT,HYAL1,KCTD12,KRT4,LEPR,LPAR1,MEIS2,MFHAS1,MMD,MUC13,NPR3,OAS1,P4HA1,PAPPA,PHACTR2,PITPNC1,PKM,PLCB4,PLOD2,PPARGC1A,PRSS3,SEMA3B,SLC1A1,SRI,SRPX,ST8SIA4,STK32B,TFPI2,TLE1,USH1C,UTRN,VCAN,VIL1,WWOX |
| JAK2 | | kinase | | 1,845 | 1,887 | 0,008 | 4,11E-09 | 1,10E-07 | ACE,ACTB,ACTN1,AKAP12,BAX,BLNK,BMP7,CASP1,CCL5,CCND1,CCND2,CD36,CDKN1B,CST7,CXCL10,CYBB,DCXR,ENPP2,ESR1,FCGR2B,FOS,FOXO1,GBP2,GCNT2,ICAM1,IRF1,JCHAIN,LCN2,LYN,MYL12A,NDN,NKG7,NOS2,NR4A1,OSMR,PECAM1,PFN1,PHLDA3,PLK2,POU2AF1,PSMB9,RBP1,SELP,SET,SLC1A3,STAT1,STAT3,TAP1,TP53,TRAM1,TSC22D1,VLDLR |
| Mek | | group | | -0,034 | -0,008 | 0,003 | 4,15E-09 | 1,11E-07 | ABCG2,ACAT1,APOE,AXL,BID,CCND1,CCND2,CCND3,CD200,CD36,CDCP1,CDKN1B,CDX2,CLU,COL1A2,CSF1R,CXCL10,CXCL11,CXCL9,DDR1,DDX58,DNMT1,ERBB3,ESR1,FOS,FOSL1,GBP2,GNL3,GPER1,HIF1A,HRH1,ID1,ID2,IL15,IRF1,ITGB3,LHB,MAF,MAFF,MANSC1,MMP2,MMP9,MS4A1,NGF,NOX4,NR1H3,NR4A1,OSBPL8,PDGFRA,PPARGC1A,PTEN,RABGGTB,RETSAT,RND3,ROCK1,RPL23,SACS,SEMA4C,SH2B3,SLC43A3,SNCG,SPRY1,SPRY2,ST6GAL1,TFB2M,TG,TOP2A,TP53,VEGFC,VIM,XAF1 |
| cholesterol | | chemical - endogenous mammalian | Activated | 2,794 | 2,817 | 0,003 | 4,15E-09 | 1,11E-07 | ABCG1,ACAT1,ACSL1,APOE,ATF3,CASP1,CAV1,CAV2,CCR2,CCR5,CD14,CD1C,COL1A1,COL3A1,CTSK,CXCL10,CYBB,CYP27A1,DDC,DDIT3,DHCR7,EPHX1,F3,FCGR2B,FN1,GLI3,GNAI1,GNMT,HSP90B1,HSPA5,ICAM1,IDI1,IHH,IL10RA,IL10RB,ITGAM,JAK1,KITLG,LAPTM5,LEPR,LYZ,MANF,MAPT,ME1,MME,MMP1,MMP2,MSMO1,NCF2,NR0B2,NR1H3,NR4A1,NR4A3,NSDHL,PCYT2,PLTP,PMVK,PSAT1,RDH11,RHOA,S100A8,SC5D,SCD,SCP2,SLC1A3,TIMP1,TLR4,TM7SF2,TMEM123,VCAM1,XBP1 |
| SMARCB1 | 0,106 | transcription regulator | | -0,411 | -0,372 | 0,006 | 4,24E-09 | 1,13E-07 | A2M,ACAT1,ACSL1,ACTR2,ADD3,AURKA,BAG3,BNIP3,BNIP3L,CA8,CCNA2,CCND1,CD44,CDC7,CDK1,CDKN2C,CIDEB,COL18A1,COL1A1,COL1A2,COL5A2,CXCR4,CYP2C8,CYP3A7,CYP4A11,DSG2,F11,FAS,FBP1,FOS,G6PC,GADD45A,GJA1,GJB1,IFI16,IFITM1,IFNAR2,IL15RA,MCM2,MCM3,MCM5,MMP1,MX1,OAS1,OAS3,PC,PCK1,PLCL1,POSTN,PPP1R3C,PRIM1,PTMS,RAB5C,RND3,SLC37A4,SMC4,SPARC,TP53 |
| MAPK14 | | kinase | | 0,509 | 0,551 | 0,006 | 4,27E-09 | 1,13E-07 | ACP5,ALOX5AP,BACE1,BAX,CALR,CAT,CCL5,CCNA2,CCNB1,CCND1,CCR7,CD40,CD83,CD86,CKM,COL1A1,CXCL10,DAB2,DUSP1,EFEMP1,FAS,FN1,FOS,G6PC,GADD45A,GJA1,GNAS,ICAM1,IL13,IL18,ITGB8,JUND,KITLG,MAOB,MAPT,MEF2C,MMP1,MMP9,NOS2,NPPA,NRP1,PCK1,PECAM1,PPARGC1A,PTEN,PTGES,SGK1,SHC1,SIGIRR,SOD1,SOD2,TIMP1,TIMP2,TOP2A,TP53,TRPV1,VCAM1,VDR,VEGFA,VEGFC,WIF1,ZFP36 |
| MITF | 0,214 | transcription regulator | | 1,703 | 1,775 | 0,009 | 4,27E-09 | 1,13E-07 | ACO2,ACP5,ACSL1,APOE,APOLD1,ATP6V1B2,BEST1,CAPN3,CCNB1,CD151,CD44,CDC25B,CENPM,CEP55,CLCN7,COL1A1,CTSK,ECT2,ESPL1,FANCG,FN1,FOS,GM2A,GNPTAB,GPM6B,GPR137B,GZMB,HIF1A,HPGD,HPGDS,IL6R,INPP4B,ITGA3,KCNN2,KIF20A,MCM2,MCM5,MCM6,MET,MGAT4B,MGP,NDST2,ORC6,POLE2,POLM,PRF1,QDPR,RAB27A,RMI1,RPA3,SDC1,SERPINF1,SHTN1,SLC19A2,SLC7A8,SNW1,SOX13,SPAG5,ST3GAL6,STXBP1,TACC3,TBC1D16,TBX2,TDRD7,TERT,TMC6,TP53,TPX2,TXNIP,UBE2C,UBL3,UIMC1 |
| TAZ | | enzyme | | 0,876 | 0,872 | -0,001 | 4,51E-09 | 1,18E-07 | ADAMTS1,ADRB2,ANXA1,AOX1,ARHGAP29,ARNT2,AXL,BASP1,C7,CAMK2N1,CAV1,CAV2,CD44,COL1A1,COL5A1,COL5A2,CYP1B1,DICER1,DUSP1,ENPP2,ESM1,F2RL1,FN1,HIF1A,IDH1,IFI16,LCP1,LUM,MET,MYOF,NOS2,PGM1,PKM,PLK2,S1PR1,SORBS2,TFPI2,TPI1,VEGFA,VLDLR |
| POR | -0,443 | enzyme | | -1,238 | -1,256 | -0,004 | 4,83E-09 | 1,26E-07 | ABCC3,ACADL,ACADVL,ACSL1,ACTG1,AHR,ANXA5,APOA4,ASNS,CD36,CD74,CLDN3,CPA3,CPT2,CYP17A1,CYP2A6 (includes others),CYP2B6,CYP2C8,CYP4A11,DHCR7,ECI1,EHHADH,ELOVL6,ENTPD5,FABP2,FGF1,GADD45B,GBP2,GPHN,GSTA3,HIF1A,HLA-A,HLA-DMA,HLA-DQA1,HLA-DQB1,HSD17B10,HSD17B11,IDI1,IHH,LGALS1,LGALSL,MSMO1,NOCT,NR0B2,NSDHL,PDK4,PFKFB3,PLA2G7,PMVK,POR,PRNP,RETSAT,SC5D,SCD,SERPINA3,SLC22A5,STAT1,TM7SF2,UBD,VEGFA |

| | | | | | | | | | |
|---|---|---|---|---|---|---|---|---|---|
| LGALS3 | 0,209 | other | | 0,663 | 0,673 | 0,002 | 5,90E-09 | 1,53E-07 | ABCC1,BAX,CASP3,CCNB2,CCND1,CCR2,CDKN1B,CLEC2B,COL3A1,COL4A1,COL5A2,CTSF,CTSV,DDOST,ENPP2,FN1,FSCN1,GPT,ICAM1,IL13,ITGAM,ITGAV,ITPR3,KRAS,KRT19,LMNB1,MAP3K10,MUC1,NOS2,PLEK2,PNPLA2,PPARGC1A,PRKCZ,PRSS3,PTEN,RAC1,RB1,TCF4,VCAM1 |
| CD2 | 1,234 | transmembrane receptor | Activated | 3,429 | 3,464 | 0,008 | 6,62E-09 | 1,72E-07 | CCNA2,CCNB1,CCND1,CCR7,CD40,CD44,CD48,CD58,CD86,CD8A,FAS,HLA-DMA,HLA-DPA1,ICAM1,IL13,ITGAL,PTEN,PTPRC,RB1,SELL,STAT1,STAT4,TP53 |
| Nr1h | | group | Inhibited | -2,302 | -2,325 | -0,003 | 6,77E-09 | 1,75E-07 | ABCG1,ACACA,ACACB,ACADL,ACE,AGPAT1,ANGPTL3,APOC1,APOE,AQP3,ARG2,ARNT2,C2,C3,CAV1,CCL5,CCND2,CCR2,CCR5,CCR7,CD40,CPT2,CXCL1,CXCL10,ELOVL6,FGF1,G6PC,HCK,HLA-DOA,HLA-DOB,IFIH1,IL15,ITGAL,LTB,MARCKSL1,MLXIPL,MYLIP,NOS2,PLTP,PPARGC1A,PRKCQ,PTGES,RGN,RUNX3,SCD,SELP,SFRP1,SP110,TAP1,THY1,TLR4,TP53,VLDLR |
| Tgf beta | | group | | 0,360 | 0,397 | 0,004 | 7,28E-09 | 1,88E-07 | ADGRE5,ANKH,BASP1,BHLHE40,CASP1,CASP8,CCL5,CCND1,CCND2,CCR2,CCR7,CDH11,CDKN1B,CLU,COL1A1,COL1A2,COL4A1,COL4A2,COL5A1,CSF1R,CTF1,CTSK,CX3CR1,CXCL10,CXCR4,DAB2,DCN,ENG,FN1,FOS,FSCN1,FSTL1,GADD45B,GCLC,GZMB,HTATIP2,ID1,ID3,IDO1,IGFBP4,IL13,IL32,INPP5D,ITGA2,ITGAV,ITGB3,JAK1,MAF,MET,MIF,MMP1,MMP2,MMP7,MMP9,MUC1,NEDD9,NOS2,NOX4,POSTN,PPARGC1A,PROM1,PTEN,RHOB,RORA,RORC,RUNX3,SDC1,SGK1,SMAD2,SMAD7,SMTN,SPARC,SPP1,SSBP3,TERT,TGFBI,TIMP1,TIMP2,TLR2,TNFRSF11B,VCAM1,VEGFA,VIM |
| CD40 | 0,351 | transmembrane receptor | Activated | 3,293 | 3,340 | 0,007 | 7,45E-09 | 1,91E-07 | APOBEC3G,BAX,BCL2A1,BCL6,BIRC3,CALR,CCL18,CCL5,CCNA2,CCND1,CCND2,CCND3,CD27,CD38,CD40,CD44,CD69,CD83,CD86,CD8A,CDKN1B,CUL1,CXCL10,CXCL12,DUSP1,F3,FAS,FCGR2B,FOS,HLA-DQB1,HSP90B1,HSPA5,ICAM1,IL10RA,IL13RA1,IL15,IL15RA,IL18,IL2RB,IL4R,IL6R,IRF1,ITGAL,ITGAM,ITGB2,JCHAIN,KLF9,MET,MMP9,NFKB1,NOS2,POU2AF1,PSMB10,PSMB9,SAMSN1,SELL,SMAD7,TAP1,TAP2,TAPBP,TLR2,TNFRSF11B,TNFSF10,TP53,TRAF5,TXLNA,VCAM1,VEGFA,XBP1 |
| SPP1 | 0,481 | cytokine | | 1,508 | 1,554 | 0,006 | 7,70E-09 | 1,96E-07 | ABCG2,AURKA,BAX,CA2,CASP8,CCL18,CCL20,CCL5,CCR2,CCS,CD163,CD44,CD9,COL1A1,CXCL1,CXCL12,CXCL6,CXorf21,DHCR7,EGF,FN1,FOS,GCDH,GSTZ1,HMOX2,HTATIP2,ICAM1,ITGA2,ITGA3,ITGAV,ITGB3,MECR,MMP1,MMP2,MMP7,MMP9,NDUFS4,NDUFS7,NDUFS8,NDUFV1,NOS2,PLOD1,POR,RB1,S100A4,S100A6,SAMM50,SC5D,SEMA6C,SERPINF1,SPP1,TERT,TGFBR2,TIMP1,TM7SF2,TP53,TXNRD2,UQCRC2,VEGFA,VIM |
| IRF4 | | transcription regulator | Inhibited | -3,423 | -3,450 | -0,004 | 8,40E-09 | 2,13E-07 | ADA,ALPL,B2M,BCL6,BNIP3,CCL20,CCNB1,CDK1,CTSS,CX3CR1,CXCL10,CXCL11,CXCL9,CXCR4,DDIT3,DNASE2,ELL2,ERBB3,GBP1,HSPA8,IL13,IL15RA,IL1R1,IL33,IL7R,INPP5D,IRF1,IRF7,IRF9,ISG15,ITGAL,MANF,MS4A1,OAS1,PCNA,PHF11,PLSCR1,PPIB,PRKCA,PRKCQ,PROCR,RAC1,RHOA,RHOB,RORA,RORC,SMARCA4,SPP1,STAT1,SUB1,TNFSF10,TRIM21,XBP1,YWHAH |
| ID2 | -0,560 | transcription regulator | | 0,880 | 0,878 | -0,000 | 8,44E-09 | 2,14E-07 | ADM,BCL6,BMP7,CCL5,CCNB1,CCND1,CCND2,CCNG1,CCR5,CCR7,CD44,CD83,CDC25B,CDK1,CDKN2C,CSF2RB,CXCR4,DUSP1,DUSP22,FCER1G,FGFR3,FOXO1,GADD45B,GADD45G,HIF1A,ICAM1,IFI16,IFNGR1,IFNGR2,IGFBP4,IL10RA,IL2RB,IL4R,IL6R,IL7R,IRF8,ITGB3,LEPR,LGALS1,LTB,MAF,MMP2,MYB,NR4A1,NR4A3,NTRK2,PIK3C2B,PIK3IP1,RORA,SDC1,SELL,SH2D1A,SMAD7,SOX13,SOX4,STAT4,TCF4,TGFBR3,TNFSF10,TRAF5,TUBA1A,VCAM1 |
| INSIG1 | -0,258 | other | | -1,762 | -1,821 | -0,010 | 8,86E-09 | 2,24E-07 | ABCG1,ACACA,ACACB,ALOX5AP,APOE,CCL5,CCR5,CCR7,CLEC7A,CXCL10,CXCL6,CXCR4,CYP4F2,DHCR7,ELOVL6,HLA-DQA1,HMGCS2,IDI1,IRF8,LCN2,LIPA,LPIN1,MLXIPL,MRC1,PCYT2,PLA2G7,PLD1,PLIN2,PMVK,PRF1,PTGES,PTGS1,RAB27A,S100A8,S100A9,SCD,SLC2A2,STAT4,TM7SF2,UCP2 |
| inosine | | chemical - endogenous mammalian | Activated | 4,101 | 4,139 | 0,008 | 8,91E-09 | 2,24E-07 | A2M,ANXA1,ANXA2,ANXA5,ARPC1B,C1QA,C1QB,C1S,C2,C3,CD63,CFB,CFD,DPP4,EMP3,FTL,IRF1,LCP1,LGALS3BP,RNASET2,SERPINB1,SERPING1,TIMP1,TSPO,TUBA1A,VIM |
| TLR3 | 0,626 | transmembrane receptor | Activated | 4,318 | 4,384 | 0,009 | 9,37E-09 | 2,35E-07 | AIM2,ARG2,ATF3,C3,CAST,CCL20,CCL5,CD38,CD40,CD69,CD86,CFB,CLU,CTH,CTSK,CXCL10,CXCL11,DDX58,DEFB1,DNMT1,DUSP1,FAS,FOS,GADD45B,GBP2,GCH1,ICAM1,ID1,IFI16,IFI44,IFI44L,IFI6,IFIH1,IFIT2,IFIT3,IFNA10,IL13,IL15,IL33,IL36A,IRF1,IRF7,ISG15,ISG20,LCN2,LIPA,LMNB1,MMP1,MX1,MX2,MYD88,NFKB1,NMI,NOS2,NR4A1,OAS1,PECAM1,PFKFB3,PIK3R3,PLK2,PROCR,PTGES,RHOB,RSAD2,S100A8,SERPINB9,SLPI,SPRY1,STAT1,TIMP1,TLR3,TLR4,TLR7,TNC,TNFRSF1B,TNFSF10,TRIM38 |

| | | | | | | | | | |
|---|---|---|---|---|---|---|---|---|---|
| Rxr | | group | | 0,468 | 0,475 | 0,002 | 9,95E-09 | 2,48E-07 | ABCC2,ADAM10,APOE,AQP3,BMP7,C1QA,CD36,CEBPE,CRABP1,CSHL1,CTSB,CTSS,CYP2B6,CYP2C8,CYP3A4,CYP4F2,DIO1,EMX1,GADD45A,GAS2,GSTA1,HNF1B,HR,INSR,ITGB2,KNG1,LTF,MGP,MMP1,MMP2,NOS2,NR0B2,NR1H3,PCK1,PDK4,PLIN2,PLTP,POR,RXRA,SEMA4D,SERPINF1,SPP1,TERT |
| RICTOR | | other | Activated | 2,420 | 2,359 | -0,007 | 1,10E-08 | 2,74E-07 | ATP5F1D,ATP5MC2,ATP6V0D1,ATP6V0E2,ATP6V1B1,ATP6V1B2,ATP6V1D,ATP6V1E1,ATP6V1G2,BAX,CD69,COX11,COX15,COX6A2,COX7B,COX8A,CYC1,FAU,G6PC,HIF1A,IFI16,IFIT2,IL7R,ISG15,LEPR,LHPP,NAT8,NDUFA10,NDUFA2,NDUFA3,NDUFA7,NDUFAB1,NDUFB7,NDUFC1,NDUFS1,NDUFS2,NDUFS3,NDUFS4,NDUFS7,NDUFS8,NDUFV1,OAS2,PHF11,PPA2,PRKCA,PSMA4,PSMA6,PSMB10,PSMB5,PSMB6,PSMB7,PSMB8,PSMB9,PSMD1,PSME1,PTEN,RAC1,RORC,RPL13A,RPL14,RPL23,RPL28,RPL35A,RPL8,RPLP2,RPS19,RPS21,RPS24,RPS3,SDHB,SDHD,SEM1,SGK1,SLC10A2,TCIRG1,UQCR11,UQCRC1,UQCRC2,VCAM1 |
| PDX1 | | transcription regulator | | 1,106 | 1,123 | 0,002 | 1,13E-08 | 2,79E-07 | ACE2,ACTB,ACTG1,AKR1B1,AKR7A2,ALB,ALDOB,ANXA2,ASNS,ATF3,CAMK2N1,CAT,CCND1,CD44,COL1A1,COL3A1,CPT2,CRELD2,CXCR4,DNAJC10,ECH1,F3,FGF1,FH,G6PC,GCH1,GCLM,GNAS,GOT1,GRN,HK1,HNF4A,HSPA5,ID1,ID2,ID3,IDH1,LRRC49,LXN,MAOB,MAPT,NFKB1,PC,PCNA,PDIA4,PDIA6,PDZK1IP1,PHGDH,PNPO,PTGER3,PTP4A1,PTPRF,RSAD2,SLC2A2,SPP1,SSBP3,SST,TGFBR3,TRIB3,TSPAN8,TXNIP |
| CFTR | 0,201 | ion channel | Inhibited | -3,461 | -3,465 | -0,001 | 1,17E-08 | 2,88E-07 | ACAA1,ACAA2,AQP1,ASNS,BLNK,CA4,CAPG,CCL5,CFH,CPA3,CUBN,CYP2B6,CYP4A11,EPHX2,FAAH,FABP2,FAS,GSR,HADH,HPGD,HSD17B11,IDI1,MYL7,NFKB1,NOS2,OSBPL1A,POR,S100A8,SCD,SCP2,SLC27A2,SLC37A4,SLC7A8,SLC9A3,STAT1 |
| IL18 | 0,440 | cytokine | Activated | 3,526 | 3,576 | 0,007 | 1,30E-08 | 3,18E-07 | ADA2,AQP3,ATF3,BCL6,CCL20,CCL5,CCR5,CCR7,CD40,CD44,CD69,CD83,CD86,CTH,CXCL10,CXCL11,CXCL12,CXCL9,ENPP2,FAS,FCGR3A/FCGR3B,GADD45B,GADD45G,GIMAP4,GJA1,GZMB,GZMH,ICAM1,IDO1,IL13,IL18,IL32,INPP5D,IRF1,ITGAL,ITGAM,MMP1,MMP9,MSH2,NFIL3,NKG7,NOS2,NPPA,PRF1,PTEN,SAMHD1,SELL,SERPINB1,SET,SMAD2,SOCS2,SPP1,TGFBR2,TIMP1,TLR4,TNFRSF11B,VAMP2,VCAM1,VEGFA |
| PTP4A1 | -0,365 | phosphatase | | 1,058 | 1,074 | 0,002 | 1,36E-08 | 3,29E-07 | ACTN1,ACTR2,ACTR3,AHCY,ANXA5,BAX,CCL5,COL15A1,COL5A1,DNAJB1,EIF6,FLNA,FLRT1,GSTP1,HIF1A,HNF4A,HNRNPA3,HNRNPF,IFI16,IGFBP7,IMPDH2,ITGB8,MAP3K1,MAPK10,MAPK8,MMP2,NCK1,NME1,PCNA,PECAM1,PKM,PPP1R12A,PRDX2,ROCK1,SHC3,SLC35G2,SPARC,SPINT2,STMN1,TAGLN2,TKT,TP53,VCAN,VIM |
| IL21 | | cytokine | Activated | 3,507 | 3,555 | 0,007 | 1,39E-08 | 3,35E-07 | ARG2,BCL6,CASP1,CASP3,CCL20,CCL5,CCND2,CCR7,CD1D,CD2,CD27,CD38,CD69,CD86,CDKN1B,CLIC5,CXCL10,DPP4,FCER1G,FCGR2B,FGL2,GZMA,GZMB,HERC6,HIF1A,ID2,IDO1,IFI16,IFIT2,IFIT3,IL13,IL2RB,IL7R,IRF7,ISG15,KLRB1,KLRD1,MMP2,MMP9,MYD88,NOS2,OAS2,PCNA,PRF1,PSMB9,PTPRC,RORA,RORC,RSAD2,SELL,SGK1,SOCS2,STAT3,STAT4,TAP1,TLR4,TYROBP,XBP1 |
| Ins1 | | other | | -0,747 | -0,714 | 0,004 | 1,40E-08 | 3,37E-07 | ACACA,ACADM,ACTB,ADIPOR1,ALAD,ALB,APOA1,APOC3,BCKDHA,CCL5,CCND1,CCND3,CDKN1B,CFD,CKM,COL18A1,COL1A1,CRAT,CSHL1,CYB5A,DDIT3,DYNLL1,ECI1,ECI2,EHHADH,ELOVL6,FOS,G6PC,GPD1,GSTP1,HSD11B2,IGFBP4,IMPDH2,INSR,IVD,LGALS3BP,MAPT,MIF,MLXIPL,MMP1,MMP2,MTTP,NDUFS4,NGF,NOS2,NPPA,NR1H3,OXT,PCK1,PCNA,PER1,PIKFYVE,PKLR,PLIN2,PPARA,PPIB,PRKCA,PRKCQ,PRLR,PTEN,RETSAT,RGN,RHOA,RRM1,RRM2,RXRA,SCD,SDHB,SLC28A1,SLC2A2,SLC37A4,SOCS2,SOD2,SP3,SPARC,STAT3,STX4,THY1,TIMP1,TIMP2,TTF2,UBE2D2,UQCRC1,UQCRC2,VAMP2,VAMP5,VEGFA,VIM |
| DCN | 0,741 | other | | -0,598 | -0,603 | -0,001 | 1,49E-08 | 3,55E-07 | BAX,CASP3,CASP8,CDKN1B,CDKN1C,CKM,COL1A2,CXCL10,DUSP1,ENG,FBN1,HIF1A,HLA-DQA1,HLA-DQB1,ICAM1,ITGAV,MET,MMP1,MMP2,MMP9,NOS2,S1PR1,SDC1,TP53,VCAM1,VEGFA |
| IRF5 | | transcription regulator | Activated | 4,382 | 4,425 | 0,008 | 1,51E-08 | 3,59E-07 | BAX,BID,CASP3,CCL19,CCL21,CCL5,COL1A1,COL3A1,COL4A1,CXCL10,CXCL11,CXCR4,DAPK2,DDX58,FOXO1,IFI44,IFIH1,IFIT2,IFIT3,IRF7,ISG15,ISG20,NAMPT,OAS1,OAS2,PARP12,PLSCR1,RSAD2,SP110,STAT1,TNFSF10,UBE2L6,VEGFA,YWHAE |
| SOX4 | 0,823 | transcription regulator | Activated | 3,570 | 3,617 | 0,006 | 1,52E-08 | 3,62E-07 | ADGRE5,ALDH1B1,APBB1,BAX,CAPN2,CASP1,CCN3,CD1D,CD2,CD53,CD74,DICER1,ENPEP,EVI2A,EVL,EZH2,FCER1G,FGF13,FHL1,FN1,FOS,GCAT,GGA2,GLDC,GPR171,GPR183,H1-10,HLA-DQB1,IFI30,IGFBP4,IGKC,KDM7A,KYNU,LCK,LHFPL6,LYZ,MAGED1,MAP7,MARCKSL1,MGST2,NRXN2,NUAK1,PECAM1,PNP,PPIC,PYGL,RARRES1,SDR39U1,SEL1L3,SELL,SERPINE2,SMAD1,SNN,SNX10,ST6GAL1,TES,TIAM1,TUBA1A,UCHL1,VIM |
| S100A9 | 0,976 | other | Activated | 2,860 | 2,884 | 0,003 | 1,57E-08 | 3,73E-07 | A2M,ABCG1,ADAMTS1,AKAP12,ALPL,ATF3,BACE1,BHLHE40,C1QB,C1S,C3,CASP1,CCND1,CD27,CD69,CDH3,CORO1A,COX6A2,CSF2RB,CXCL10,CXCL14,DDC,ENPP2,FAS,FOSL1,GZMB,ICAM1,IL33,ISG15,ITGAM,KLF15,LOC102724788/PRODH,MCF2L,MMP1,MMP2,MMP9,NOS2,NR4A3,PCDHGB5,PER1,PIK3IP1,POSTN,PTGES,RNASE6,S100A8,S100A9,SEMA3B,SGK1,SHC3,SLC26A1,SOCS7,TNFAIP8,TXNIP,VCAM1,VCAN,WT1 |

| | | | | | | | | | |
|---|---|---|---|---|---|---|---|---|---|
| SYVN1 | | transporter | | 1,262 | 1,326 | 0,009 | 1,74E-08 | 4,09E-07 | ABCC3,ADAM9,ADCY9,AHR,AXL,BHLHE40,CALM1 (includes others),CAVIN1,CCND1,CD151,CD44,CDA,CRTAP,CTPS1,CYP1B1,DAB2,DUSP1,EXT2,F3,FANCG,FAS,FOLR1,GGCX,GNB2,HLA-A,HLA-C,HNRNPM,IFI44,IFITM2,IL7R,ITGA3,ITGAV,LGALS3BP,MCAM,MGRN1,MYOF,NFE2L2,PFN1,PNPLA2,RPN2,SETX,SLC12A3,SLC1A5,SLC27A3,SLC3A2,SLC43A3,TARS1,USP5 |
| S100A8 | 2,072 | other | Activated | 2,501 | 2,519 | 0,002 | 1,75E-08 | 4,11E-07 | A2M,ABCG1,ADAMTS1,AKAP12,ALPL,ARG2,ATF3,BHLHE40,BIRC2,C1QB,C1S,CASP1,CCL20,CCND1,CD69,CDH3,CORO1A,COX6A2,CSF2RB,CXCL10,CXCL14,CXCL9,DDC,ENPP2,FAS,FOSL1,GZMB,ICAM1,IDO1,IL33,ISG15,ITGAM,KLF15,LOC102724788/PRODH,MCF2L,MMP1,MMP2,MMP9,NFKB1,NOS2,NR4A3,PCDHGB5,PER1,PIK3IP1,POSTN,PTGES,RNASE6,S100A8,SEMA3B,SGK1,SHC3,SLC26A1,SOCS7,TNFAIP8,TXNIP,VCAM1,VCAN,WT1 |
| COL18A1 | 0,396 | other | Inhibited | -2,276 | -2,322 | -0,008 | 1,86E-08 | 4,35E-07 | CCND1,CDC25B,CDH5,CDKN2C,EFNB2,F11,F12,F2RL1,F3,FN1,FOS,HIF1A,ICAM1,ID1,ID3,IL18,ITGAV,ITGB2,ITGB3,KNG1,MAPK8,MMP1,MMP2,MXI1,NOS2,NRP1,PDCD5,PECAM1,SELP,SP3,STAT1,STAT3,TNFRSF21,TUBA1A,VEGFA,VWF |
| ETV5 | -0,165 | transcription regulator | | 1,486 | 1,496 | 0,002 | 2,14E-08 | 4,99E-07 | AGRN,AHNAK,BAX,CAV1,CAVIN1,CCND1,CLDN1,CLDN3,CLIC5,CXCL12,CXCR4,DPYSL2,FN1,FXYD5,HOPX,ICAM1,ICAM2,KRT19,KRT7,LAMP3,LCN2,LYZ,MAF,MET,MMP2,MSN,MYB,PKP2,PMP22,RAB27A,S100A6,SAMHD1,SCD,SLC34A2,SOAT1,SPP1,TJP1,VEGFA |
| folic acid | | chemical - endogenous mammalian | | 1,521 | 1,522 | 0,000 | 2,17E-08 | 5,05E-07 | ABCC1,ABCG2,CCL5,CLU,COL1A1,COL3A1,CSNK1A1,CYBB,DDIT3,DNMT1,FABP1,FN1,FZD6,HDAC6,HOXA1,IFI27,IFI6,MMP2,MMP9,MYD88,NOS2,NOX4,PCNA,PECAM1,PLG,PTH1R,SFRP1,TP53,VCAM1 |
| MKNK1 | 0,043 | kinase | | 0,805 | 0,867 | 0,010 | 2,24E-08 | 5,21E-07 | ACOT7,AGRN,ANXA2,ANXA5,ATP1A3,BAG3,BMP8B,CAMK2A,CRMP1,EPB41L2,FAM49A,FLNA,FOS,FOSB,GNAI1,GPC4,GPS1,HADHA,HSPA4L,IFIT2,IFRD1,ISG15,KLC1,LAMC1,LCN2,MAPT,NR4A3,PLTP,RAB3A,SFRP1,SMAD2,SPARC,STXBP1,TMBIM6,TP53BP1,TRIM32,TXNIP,VAMP2,VIM |
| TERT | -0,188 | enzyme | | 0,116 | 0,122 | 0,002 | 2,27E-08 | 5,26E-07 | ALDH1A3,ASAP1,ASNS,ATF3,B2M,BAX,BNIP3,CALU,CAV1,CCND1,CD24,COL5A2,EFEMP1,ELK3,FUT4,GLB1,HDHD5,HLA-B,HOXB6,HTRA1,ICAM1,IFI16,IFIT2,IGFBP4,IL13RA1,IQGAP2,ISG15,ITGAV,KRT19,MMP1,MMP2,MMP9,MX1,PLOD2,PLTP,PROM1,PTGS1,RB1,RRM2,SASH1,SOD2,SPON1,SPP1,TACC3,TERT,TP53,VIM,XBP1 |
| FOXM1 | 0,191 | transcription regulator | Activated | 2,732 | 2,787 | 0,009 | 2,33E-08 | 5,39E-07 | ACE,ACE2,ADAM17,ANXA1,CAT,CAV1,CCNA2,CCNB1,CCNB2,CCND1,CCND2,CDC25B,CDK1,CDKN1B,CDKN3,CXCL12,ESR1,FOS,FUT4,GLB1,HSD11B2,KIF20A,MAPK8,MET,MMP2,MMP7,MMP9,NFKB1,PCNA,PECAM1,PRC1,PRDX2,PROM1,PTTG1,RMI1,SFRP1,SOD2,STAT3,STMN1,TGFBR2,TJP1,TOP2A,TP53,VCAN,VEGFA,VIM,ZEB2 |
| L-methionine | | chemical - endogenous mammalian | | 1,044 | 1,039 | -0,002 | 2,44E-08 | 5,62E-07 | ALB,CCNB1,CCNB2,CDK1,CDKN2C,CLU,DDIT3,F3,FANCL,G6PC,GADD45B,GNA11,HSPA5,IFI16,IGFBP4,MAT2A,MTTP,PCK1,PCK2,PRC1,RHOB,SMAD7,SOD2,SPHK2,SPP1,TOP2A |
| Pkc(s) | | group | | 0,198 | 0,236 | 0,006 | 2,45E-08 | 5,63E-07 | ACAT1,ACO1,ADM,AHR,APOA1,AQP3,ARG2,ATF3,CCND3,CCR2,CCR5,CD36,CD55,CDKN1B,CDX2,CGA,CLDN1,COL3A1,CTSB,CXCR4,CYBB,CYP2A6 (includes others),DDIT3,DUSP1,F3,FAS,FN1,FOS,FOSL1,GADD45A,GADD45B,GPER1,HIF1A,HPGD,HSP90B1,HSPA5,ID2,KCNJ2,LHB,LIPA,MAOB,MMP1,MMP2,MMP9,MS4A1,NFKB1,NOS2,NOX4,NR4A1,NR4A3,OVOL1,P4HB,PDIA4,PER1,PPARA,PPP1R15A,PRKCA,PTPRE,RB1,S100A8,SPP1,STC2,TAC1,VCAM1,VDR,VEGFA,XBP1 |
| RETNLB | | other | Activated | 3,274 | 3,327 | 0,009 | 2,82E-08 | 6,46E-07 | CASP3,CCL5,CCR2,CCR5,CD86,COL1A1,COL1A2,FABP1,FN1,FRZB,FZD6,IL13,IL13RA1,IL4R,JAK1,KLF15,LMNB1,MID1,MMP2,MSN,NDN,NR4A1,PTEN,SFRP1,SMAD1,SOCS2,STK17B,TERT,THBS4,TIMP1,TLR2,TNNC1,UBE2C |
| NR1I2 | | ligand-dependent nuclear receptor | | 0,472 | 0,488 | 0,002 | 2,84E-08 | 6,49E-07 | ABCC2,ABCC3,ACOX1,AHCY,AHR,ALDH3A2,ANG,APOA4,ARAF,BHMT,CAT,CD36,CES2,CSHL1,CTH,CYP27A1,CYP2A6 (includes others),CYP2B6,CYP2C8,CYP3A4,CYP3A7,CYP4A11,DHCR7,ELOVL6,ENTPD5,FABP1,FMO5,GLUD1,GLUL,GOT1,GSTA1,GSTP1,HMGCS2,HNF4A,HSD3B2,LCN2,MGP,MTTP,MYB,NAT8,NAT8B,NR0B2,NR1I3,PDIA4,PGRMC2,PIPOX,POR,PPARA,S100A8,SCD,SLC22A7,STAT3,TCN2,TNFRSF11B |
| ELAVL1 | -0,037 | other | | 0,656 | 0,685 | 0,007 | 3,23E-08 | 7,31E-07 | ANXA11,ARAF,ARG2,ATF3,CALM1 (includes others),CCL8,CD83,CLDN1,CLDN3,DCN,DGAT1,DUSP1,EIF4ENIF1,FLNA,FOS,GSS,IL13,IRF1,NCL,NOS2,NXF1,PBK,PCGF2,PCOLCE,PIGR,PRC1,PROM1,QARS1,RACK1,RPN2,SLC25A11,SLC7A7,SRSF7,STAT3,TIA1,TJP1,TOP2A,TP53,TRIOBP,VEGFA,ZFP36 |

| | | | | | | | | | |
|---|---|---|---|---|---|---|---|---|---|
| ursodeoxycholic acid | | chemical - endogenous mammalian | | 0,236 | 0,248 | 0,002 | 3,33E-08 | 7,52E-07 | ABCC2,ABCC3,ABCG2,BAX,CCL20,CDX2,COL1A1,COL1A2,COL4A1,CXCL10,CYP27A1,CYP2B6,CYP3A4,CYP4A11,DDIT3,GCLC,GCLM,GPT,MAF,NFKB1,NOS2,PRKCZ,SLC10A2,SLC5A2,SPARC,SPP1,TIMP1,TP53,VDR |
| MAP2K1 | 0,166 | kinase | | 0,720 | 0,771 | 0,008 | 3,36E-08 | 7,58E-07 | ABCC1,ADGRG1,AHR,ALDH1L1,APOBEC3G,APOE,ATF3,BCL2A1,BIRC2,BIRC3,CAPN3,CCL5,CCNA2,CCND1,CD44,CDH5,CDKN1B,CKM,COL1A1,COL1A2,COL3A1,CSTA,CTSB,CYP17A1,CYP46A1,DAB2,DNMT1,DPP4,DUSP1,ENO1,F3,FLNA,FOS,FOSL1,HIF1A,HSPA5,ITGB3,JUND,LHB,MAOB,MMP1,MMP2,MMP9,NET1,NOS2,NOX5,OLFM4,PCNA,PDGFRA,PECAM1,PLA2G4A,RAB38,RAP2B,RRM2,SERPINE2,SMAD2,TFPI2,TNC,TNFSF10,VEGFA,ZWINT |
| NFYA | | transcription regulator | | -0,046 | -0,017 | 0,007 | 3,44E-08 | 7,73E-07 | ALB,CALR,CAT,CCNA2,CCNB1,CCNB2,CDC25B,CDK1,COL1A2,EPHX1,F12,G6PC,GADD45A,GADD45B,GUCY1B1,HLA-DQB1,HLA-DRA,HSPA5,ICAM1,IRF1,JUND,LPIN1,OGG1,PCK1,PKM,PPP1R15A,PTTG1,RHOB,RRM2,SCD,TGFBR2,TOP2A,TXNIP,UBE2I,VWF |
| EPO | | cytokine | | -1,531 | -1,499 | 0,005 | 3,82E-08 | 8,57E-07 | ACTB,ATP6V1B2,B2M,BAX,BCL6,BLVRB,BNIP3L,BZW2,CA2,CASP3,CCL19,CCND2,CCND3,CD36,CD52,CDKN1B,CISH,CLIC1,CTSK,CTSZ,CUL1,CXCL12,CXCR4,DDIT3,FABP1,FAS,FOS,FOSB,FPR1,G6PC,GCC2,GLUL,GYPA,HBB,HLA-DRB1,HNRNPF,IL18,KCNH6,KPNB1,LAPTM5,LCP1,MACROH2A1,MIA2,MMP2,MYB,NAB2,OSBPL1A,PCNA,PDCD4,PDLIM2,PIK3IP1,PRDM14,PRKCA,PRKCD,PRKCQ,PRKCZ,RHOA,RNF128,ROCK1,SAMSN1,SELP,SLC4A1,SOCS2,SOD1,SPP1,SSRP1,ST3GAL4,STAT3,TJP1,TLR4,TNFRSF11B,TNFSF10,TP53,TPI1,TXNIP,UCP2,UGCG,VEGFA,VEGFC,VWF,WDR1,XPC |
| Histone h3 | | group | Inhibited | -2,643 | -2,621 | 0,008 | 3,88E-08 | 8,67E-07 | ACKR3,ADAM10,ADD3,AFP,AKAP12,ARHGEF17,ARR3,ATF7IP,B2M,CA4,CAV1,CCL5,CCNA2,CCNB1,CCND1,CCND2,CD1D,CD40,CD69,CD9,CDK14,CDKN1B,CDKN2C,CDX2,CGA,CHFR,CKAP4,CLPS,COL1A2,CRIP2,CSF1R,CSHL1,CXCL10,DIO1,ENO3,ESR1,EVX1,FAM49A,FKBP5,FOS,GAD1,GAS2,GHR,HNF4A,HOXA10,HOXB1,HOXB5,HOXB6,HPSE,ICAM1,ID1,ID2,IFI6,IGFBP4,IL13,IL6R,INHA,IRF7,KAT5,KIAA0895,MAD2L1,MCM2,MCM3,MCM5,MEIS2,MET,MMP9,MT1G,MX1,NDUFA7,NDUFB7,NDUFC1,NDUFS1,NDUFS2,NDUFS3,NDUFS4,NDUFV1,NOS2,OPA1,OSGIN1,PALLD,PCNA,PKLR,PRLR,PTEN,PTGES,RAB38,RB1,RCAN1,RGS10,RSAD2,S100A8,S100A9,SCD,SEC31A,SFRP1,SGK1,SLC2A2,SOD2,SPP1,TACC2,TBX2,TERT,TGFBR2,TLR2,TMPRSS2,TNFSF10,TOP2A,TYRO3,UQCRC1,UQCRC2,USP3,VEGFA,WDR1,WNT8B,XBP1,ZNF423 |
| TYROBP | 2,117 | transmembrane receptor | Activated | 3,051 | 3,073 | 0,006 | 4,13E-08 | 9,20E-07 | CCR7,CD40,CD69,CD83,CD84,CD86,FCGR2B,ICAM1,IDO1,IL10RA,IL13RA1,IL18,IRF8,ITGAM,KLRD1,LHFPL2,NCF2,NCKAP1L,NOD2,PLEK,SAMSN1,SH2B3,SLC1A5,SPP1,TCIRG1,TNFRSF1B,TREM2,TYROBP |
| DNMT3A | | enzyme | | 1,521 | 1,512 | -0,001 | 4,28E-08 | 9,48E-07 | ACE,ACSL1,ADCY7,AKAP12,ATP2B4,BLNK,CAMK2A,CASP4,CASP6,CD38,CKM,CSF1R,CXCR4,CYBB,EFEMP1,EMX1,FCER1G,FOXO1,GPER1,GRINA,GSS,GUCY1A1,HOXA1,ITGAV,ITPR3,JCHAIN,MAPK13,MYL3,NR4A1,PIK3C2B,PLA2G7,PLCB4,PLK2,POU2AF1,PPP1R3C,PRKCI,PTGS1,PTPRC,RAP2B,RRAS2,S1PR1,SHC1,SLC12A8,SLIT2,SPP1,ST14,TNNC1,TNNI1,TNNT2,TPCN1,TSC22D1 |
| Esrra | | transcription regulator | Inhibited | -2,328 | -2,314 | 0,003 | 4,30E-08 | 9,48E-07 | ACADL,ACADM,ACAT1,ACE2,ACO1,ACOX1,ACSL5,ALDOB,APOA4,COX6A2,COX8A,CPT2,CRAT,CYC1,DLAT,ECH1,ENO1,ENO3,ESRRA,FABP1,FABP2,FBP1,GOT1,GYS1,IHH,LEPR,NDUFA2,NDUFS3,PCK1,PCK2,PDK4,PFKL,PGAM2,PGM1,PPARGC1A,PYGL,SDHB,SIRT3,SPP1,UQCR11 |
| CST5 | | other | Inhibited | -3,396 | -3,438 | -0,005 | 4,50E-08 | 9,85E-07 | ABCC1,AGO1,AHNAK,AKAP12,ANXA2,ANXA3,ANXA5,AP1M2,ARHGAP29,ATP2C2,ATR,BLOC1S1,CAMK2G,CAV1,CD44,CNOT8,COPS7A,CORO1A,CTSB,CYBRD1,DECR1,DHX9,DNAJA3,EIF2B2,EIF5B,ELK3,EMP3,ESRRA,EVPL,FHL1,GAR1,GNL3,HNRNPA3,ID3,IGBP1,IQGAP2,ITPR3,KDM5B,LAMC1,MALT1,MAPK13,MEF2C,MRM3,MSN,NCL,NDUFB7,NOP10,NOP53,NRP1,NUP107,NUP155,PCLAF,PIP4K2A,PLS3,PPIC,PPL,PRDX2,PRKACB,PRSS8,PYGB,RAB25,RBM28,RBP1,RRP1,RTN3,S100A11,SGPL1,SLC1A3,SMAD2,SOX4,SSRP1,SUPV3L1,TESC,TXN,VCAN,VIM,VTI1B,WDR3,ZFC3H1 |
| MYOD1 | -0,045 | transcription regulator | Inhibited | -2,170 | -2,129 | 0,006 | 4,51E-08 | 9,85E-07 | ACACA,ACACB,ACSL5,ADCY7,ASS1,BCL6,BIN1,CCND3,CDH15,CDKN1C,CDX2,CHRND,CKM,COL4A1,DAO,DUSP1,ENO3,ENPP2,ESR1,FOSL1,FSTL1,FYN,GADD45A,GADD45G,GMFG,GUCA2A,GYS1,HADH,HIPK3,HSPA4L,HSPB8,ID2,IFI16,ITGA7,LZTS3,MAFB,MEF2C,MMP2,PLEKHO1,PNP,POSTN,PPARGC1A,RB1,RBM38,RECK,RRAS,SCN4A,SGCA,SH3GLB1,SLC12A2,SMAD7,SST,STAM,TGFBR3,TJP1,TKT,TNNC1,TNNI1,TNNT2,TNS1 |

| | | | | | | | | | |
|---|---|---|---|---|---|---|---|---|---|
| BDNF | 0,071 | growth factor | | -1,918 | -1,887 | 0,004 | 4,86E-08 | 1,06E-06 | ACOT7,ADAM17,ADRB2,AGRN,ALDH7A1,ANXA2,ANXA5,ARC,ATP1A3,ATP5MC2,BAG3,BCL2L2,BHLHE40,BID,BMP7,CAMK2A,CASP3,CAV2,CCND2,CD36,CDADC1,CRMP1,CXCR4,DNAJB5,DUSP1,ELOVL6,EPB41L2,FAM49A,FLNA,FOS,FOSB,FSCN1,FTL,GLUL,GNAI1,GPC4,GPS1,HADHA,HSPA4L,HSPA5,IFRD1,KLC1,LAMC1,LCN2,LGALS1,LYPD1,MACROD1,MAPT,MMP1,NAB2,NECAB3,NOS2,NR4A1,NR4A3,NTRK2,P4HA1,PCNA,PFKFB3,PLG,PLTP,PRKCZ,PTEN,RAB3A,RHOA,RPL35A,S100A10,S100A9,SFRP1,SLC16A7,SOD2,SPARC,SPP1,SPRY2,SST,STXBP1,TAC1,TIAM1,TIPARP,TMBIM6,TMEM45A,TNFRSF11B,TP53BP1,TRIM32,TUBA1A,TXNIP,VAMP2,VIM |
| KITLG | 0,406 | growth factor | | 1,772 | 1,821 | 0,008 | 5,30E-08 | 1,15E-06 | ACTB,ALOX5,ALOX5AP,ATP6V1B2,AXL,B2M,BLVRB,BZW2,CA2,CASP1,CASP3,CCNA2,CCND3,CD14,CD247,CD3D,CD3G,CDKN1B,CISH,CLIC1,CXCL12,CXCR4,EFNB2,FOS,GCC2,GJA1,HCK,HNRNPF,ID2,IGHM,IK,IL13,IL18,IRF2,ITGAV,ITGB3,KPNB1,LAPTM5,LTF,MEF2C,MME,MMP2,MYB,NPPA,OGG1,OSBPL1A,PDCD4,PIK3IP1,PKM,PLSCR1,PRKCA,PRKCQ,PSMB5,RBBP7,RNF128,SAMSN1,SHC1,SLC4A1,ST3GAL4,STAT3,STK25,SYK,TCF12,TERT,TXNIP,UCP2,UGCG,WDR1,ZNF207 |
| IFNAR1 | 0,155 | transmembrane receptor | Activated | 3,669 | 3,700 | 0,006 | 5,76E-08 | 1,24E-06 | ATF3,AXL,B2M,CCL20,CCL5,CD40,CLIC5,CXCL10,CXCL11,CXCL6,HLA-A,IDI1,IDO1,IFI16,IFI44,IFI6,IFIH1,IFIT2,IFIT3,IFITM1,IFNGR1,IL15,IL18,IRF7,ISG15,ITGAV,LAMP3,MX2,NOS2,OAS1,OAS2,OAS3,PARP12,PLA2G7,RIPOR2,RSAD2,RTP4,SDC1,SOD2,STAT1,TAPBPL,TLR2,TLR3,TLR7,TNFAIP2,TNFSF10,TRIM22,UBD,VCAM1,XAF1 |
| PI3K (family) | | group | | 1,905 | 1,935 | 0,005 | 5,82E-08 | 1,25E-06 | ABCG1,ADM,BIRC3,CAT,CCND3,CDKN1B,CDX2,CTSD,CXCL1,CXCL10,CXCL11,CXCL12,CXCL9,CXCR4,DDIT3,DEFB1,ESR1,FOS,FOXO1,FOXO4,GCLC,GCLM,HIF1A,HK1,HSPA5,ISG15,MMP2,MMP9,NFE2L2,NOS2,PKM,PRKCZ,PTEN,PTGS1,SOX4,TJP1,TNC,TNFSF10,TXNIP,VEGFA,WTAP |
| HDAC1 | 0,381 | transcription regulator | | -0,540 | -0,550 | -0,002 | 6,25E-08 | 1,34E-06 | AKAP12,APOA1,ARC,ATF3,BAX,BCL6,CCL5,CCNA2,CCNB1,CCNB2,CCND1,CCND2,CD1D,CD27,CD44,CDK1,CDKN1B,COL1A1,COL1A2,CSF1R,CSHL1,CTSK,DUSP1,ESR1,FABP1,FABP2,FAS,FOS,GABARAPL1,GLUL,GNAS,GSTP1,ID1,IL13,LAT,LCN2,MCM3,MCM5,MCM7,MEF2C,MME,MMP9,MT1G,MYO1F,NASP,NCL,NKX2-5,NOS2,OLFM4,PCK1,PFKFB3,POLL,PPARGC1A,PTEN,PTPN6,RBBP7,RECK,RGS10,RHOB,RORC,RRM2,S100A9,SFRP1,SGK1,SLC35D1,SMAD7,SPP1,ST5,STAT3,TBX2,TERT,TGFBR2,TNS1,TOP2A,TP53,VDR |
| hydrocortisone | | chemical - endogenous mammalian | | -1,379 | -1,369 | 0,002 | 6,36E-08 | 1,36E-06 | ACO1,ADM,ALDH18A1,ALDH1L1,BACE1,BCLAF1,CCND1,CCND2,CCND3,CD163,CD40,CD86,CDH11,CDK1,CFD,CSHL1,CXCR4,CYP3A4,FAS,FGF7,FN1,GADD45B,GBP1,GLUL,GNAS,ICAM1,IGFBP4,IGFBP6,IGFBP7,ITGA2,ITGAM,LY75,NR1I3,NTRK2,PLA2G4A,PLG,PRLR,PTGS1,S100A8,SFRP1,STC2,TLE2,TLR2,TNFAIP2,TNFRSF11B,TP53,TUBB,VAMP2 |
| CDKN2A | | transcription regulator | Inhibited | -2,080 | -2,088 | -0,001 | 6,56E-08 | 1,40E-06 | A2M,BARD1,BAX,C3,CAPG,CASP3,CCL18,CCL19,CCL5,CCNA2,CCNB1,CCND1,CCND3,CCNG1,CCR7,CDC25B,CDK1,CDKN1B,CDKN2C,COL1A2,CUL1,CXCL10,CXCL14,CXCL6,DCK,DDR1,DEK,DUSP1,EED,EZH2,FAM111A,FANCL,FHL2,FOS,GADD45G,GLB1,GNAS,GNL3,GTF3C1,HMGB2,IFI35,IL15,IL2RG,IL36A,ITGAV,ITGB3,KANK2,KAT5,KRAS,MAD2L1,MCM5,MCM7,MELK,MMP1,MMP2,MRC1,MS4A1,P4HA2,PBX3,PCNA,POLD3,POU2AF1,PPFIBP1,PSMB9,PTEN,RAB27A,RB1,RBBP7,RMI1,RRM1,RRM2,S100A6,SAE1,SH3BP2,TLR4,TNFRSF1B,TOB1,TP53,TPM1,VEGFA,VIM,VRK1,ZMAT3 |
| MAPK8 | -0,265 | kinase | | -1,312 | -1,286 | 0,004 | 6,65E-08 | 1,42E-06 | APOA1,APOE,AQP1,B4GALT5,BIRC3,CASP3,CAT,CCL5,CCNA2,CCND1,CDKN1B,COL1A1,CSTA,CYBB,CYP27A1,DDIT3,DGAT1,DNMT1,DUSP1,EGF,FOS,FOSL1,FOXO1,FOXO4,FUS,G6PC,GADD45A,HDAC6,HIPK3,IL13,JUND,LHB,MAOB,MIF,MMP1,MMP2,MMP7,MMP9,MTTP,NOS2,NOX4,NPPA,NRP1,PPARGC1A,PTEN,PTGES,RBP1,SOD2,TAC1,TBP,TLR2,TP53,VDR,VEGFC,ZFP36 |
| WNT5A | 0,180 | cytokine | | 1,247 | 1,248 | 0,000 | 7,36E-08 | 1,56E-06 | ARHGDIB,CAPN1,CCND1,CD14,CD38,CD40,COL1A1,CXCL1,CXCL12,CXCL6,CXCR4,DAB2,ENPP2,ESR1,FLNA,FN1,FOXN1,ID2,ID3,IL15,IRF1,IRF2,IRF8,LCP1,MMP1,NFKB1,NOS2,NR0B2,PITX3,PRKCI,PRLR,PTEN,SDC1,SPARC,SPP1,STAT1,STAT3,TLR1,TLR3,TLR4,TLR5,TNFSF10,VEGFA,VIM |
| ELOVL5 | -0,122 | enzyme | | 1,149 | 1,123 | -0,006 | 8,06E-08 | 1,70E-06 | ACACA,ACADL,CYP4A11,EHHADH,ELOVL6,FOXO1,G6PC,HADHA,HMGCS2,MLXIPL,PCK1,PDK4,PLTP,PNPLA2,PPARGC1A,SCD,TRIB3 |
| MTPN | | transcription regulator | Activated | 3,028 | 3,084 | 0,010 | 8,31E-08 | 1,75E-06 | ANXA1,BAX,BCL2A1,BCL2L2,BNIP3,CASP3,CASP8,CCNA2,CCNB1,CCNB2,CCND1,CCND2,CCND3,CDK1,COL1A1,COL1A2,DDC,ENG,FAS,FN1,FOS,LTB,MIF,NFKB1,NPPA,S100A10,S100A6,S100A8,SPARC,SPP1,TNFSF10,TUBA1A,VCAM1 |

| | | | | | | | | | |
|---|---|---|---|---|---|---|---|---|---|
| BCL6 | 0,401 | transcription regulator | | 0,108 | 0,090 | -0,003 | 8,65E-08 | 1,82E-06 | A2M,ADCY9,BACH2,BAX,BCL2A1,BCL6,BLNK,CCND1,CCND2,CCR7,CD151,CD2,CD44,CD69,CDKN1B,CISH,CLEC10A,COL1A1,CPD,CTSK,CXCL10,DOCK10,DPYSL2,EPB41L2,FAS,FCGRT,GABARAPL1,GADD45A,HACD1,HERC6,IL13,IL13RA1,IL18,IL2RB,IL4R,IL6R,IL7R,IRF8,IRF9,ITGA3,KLK1,LITAF,LPIN1,MAF,MGP,MRC1,MSH2,MYB,NFKB1,NOS2,PLA2G4A,PTEN,RHOF,RORC,SCP2,SDC1,SMC3,SOCS2,SYK,TACSTD2,TP53,UBA7,VWA5A,XBP1 |
| linoleic acid | | chemical - endogenous mammalian | | -0,639 | -0,620 | 0,003 | 8,74E-08 | 1,83E-06 | ACADM,APOA1,CD36,CD47,CLU,CYP4F12,CYP4F2,DDIT3,DUSP1,FABP1,G6PC,HSP90B1,HSPA5,ICAM1,LHB,MLXIPL,MMP1,MTTP,NEDD4L,NOS2,PECAM1,PKLR,PLIN2,RXRA,S100A8,S100A9,SCD,UCP2,VCAM1,VEGFA |
| MAP2K7 | | kinase | | 0,168 | 0,181 | 0,004 | 9,76E-08 | 2,04E-06 | ATF3,BIRC3,CCL5,CCND1,CSTA,CXCL10,EFNB2,ENPP2,FAS,FCGRT,FN1,GJA1,HSPA5,IL1R1,KLC1,LSP1,MAOB,MAPK8,MTTP,NPPA,PDIA3,PLA2G4A,RHOB,SERPINC1,TFPI2,VDR |
| ATP | | chemical - endogenous mammalian | | -0,615 | -0,583 | 0,005 | 1,04E-07 | 2,16E-06 | AHR,BAX,CAPN1,CAPN2,CASP1,CCND1,CCR5,CCR7,CD14,CD200,CD83,CD86,CDKN1B,CLDN1,CXCL10,CXCR4,DUSP1,F3,FLNA,FOS,FOXO1,GCLC,GCLM,HIF1A,ICAM1,IL33,ITGAM,LCN2,MMP1,NOS2,NR4A1,NRP1,SELL,SLC12A3,TLR4,TNFRSF1B,TP53,TRPV1,TXN,VCAM1 |
| ZBTB16 | -0,736 | transcription regulator | Inhibited | -2,946 | -2,943 | 0,000 | 1,04E-07 | 2,16E-06 | BACH2,BCL6,BID,BMP7,CASP3,CCNA2,CCR2,CD14,CD180,CD2,CD3D,CD3G,CD44,CD55,CD74,CD81,CD83,CD86,CD8A,COL1A1,CXCL10,EYA2,F2RL1,FGFR3,ID2,ID3,IFNGR1,IL10RA,IL13,IL4R,IL7R,ITGAV,ITGB3,KCNJ8,KLRB1,LY86,LY96,MAF,MMP9,RARG,RORA,RORC,RUNX3,SELL,SELP,SOX13,SPP1,TSC22D1 |
| KLF2 | 0,390 | transcription regulator | | 1,622 | 1,612 | -0,002 | 1,09E-07 | 2,25E-06 | ACE,ACKR3,ADM,ASS1,CAV1,CCL5,CCND3,CCR2,CCR5,CCR7,CD36,CD40,CD44,CD55,CD86,CD93,COL4A1,CX3CR1,CXCR4,CYP1B1,DOCK2,F2RL1,F3,FGR,FUT4,GADD45A,HBB,HIF1A,HPSE,HYAL2,ID1,ID3,IL2RG,IL6R,IL7R,ITGB2,ITGB3,KITLG,LEPR,NDRG1,NOTCH2,PECAM1,PLOD2,PRF1,S1PR1,SELL,SERPINB9,SLCO2A1,STAT1,STAT3,TCF4,TERT,TFPI2,TIMP1,VAV1,VCAM1,VWF |
| TXN | -0,333 | enzyme | | 0,648 | 0,652 | 0,001 | 1,24E-07 | 2,55E-06 | ACADM,APOA1,CCND2,CD38,CD40,CD86,CYP1B1,ESR1,FOS,HIF1A,ICAM1,MIF,MMP9,MS4A1,NFKB1,NOS2,NSF,SELL,SOD2,TP53,TXN,VEGFA |
| L-triiodothyronine | | chemical - endogenous mammalian | Inhibited | -2,283 | -2,255 | 0,003 | 1,25E-07 | 2,56E-06 | A2M,ACOX1,ACSL1,ACTB,ADCY9,ADM,ADRB2,AFP,ALB,ANGPTL3,APOA1,APOC1,APOC3,APOM,ARSA,ATP1A3,BNIP3L,C1S,CCND1,CD36,CD44,CDKN1B,CGA,CLU,COL4A1,CSHL1,CTH,CXCL12,CYP17A1,CYP27B1,CYP4A11,DIO1,DUSP1,DVL1,EGF,EHHADH,EIF4EBP2,ENG,ENPP2,ESR1,FABP1,FAS,FH,FOS,G6PC,GNAI2,HIF1A,HR,ID1,ID2,ID3,IGFBP4,IGFBP6,INHA,ITGA7,ITGAV,ITGB3,KLF9,LYZ,ME1,MLXIPL,MMP9,NKX2-5,NME1,NOS2,NR1H3,NR4A1,OAT,PARP1,PCK1,PCK2,PDK2,PDK4,PER1,PITX3,POR,PPARA,PPARGC1A,PRDX2,PRDX3,PRKCQ,PRKCZ,PTH1R,PTTG1,PYGL,RASGRP1,RGN,ROCK1,RYK,SCD,SHBG,SLC2A2,SMAD7,SP3,ST3GAL4,ST6GAL1,THRB,TIAM1,TP53,TRHDE,UCP2 |
| miR-16-5p (and other miRNAs w/seed AGCAGCA) | | mature microRNA | | -1,027 | -1,103 | -0,010 | 1,27E-07 | 2,60E-06 | ABHD10,ARL2,BCL2L2,CACNA2D1,CAPRIN1,CARD8,CCND1,CCND2,CCND3,CDC14B,CSHL1,ECHDC1,EIF2B2,EIF4G2,FGF7,FNDC3B,GALNT7,GFPT1,GRB10,GRN,GSTM4,HERC6,HPF1,HSDL2,HSP90B1,IFRD1,ITGA2,KITLG,LAMC1,LAMTOR5,MLLT11,MSH2,MYB,NOTCH2,NPR3,PANX1,PDCD4,PLAG1,PMS1,PNP,PRIM1,PSAT1,RAB21,RARS1,RECK,RIDA,RNASEL,SERPINE2,SKAP2,SLC12A2,SLC16A3,SLC38A1,TIA1,TMEM43,TPI1,TPPP3,TXN2,UCP2,VEGFA,VTI1B,WIPF1,WT1 |
| ATF4 | | transcription regulator | | -1,341 | -1,286 | 0,008 | 1,29E-07 | 2,64E-06 | ABCG2,ANGPTL3,ASNS,ATF3,CALR,CDKN1B,CTH,CYP27A1,DDIT3,FN1,FYN,GADD45A,GARS1,GCH1,GHITM,GRB10,GYS1,HNF4A,HSP90B1,HSPA5,IGFBP7,IHH,KLF9,MRPS7,MTHFD2,NDRG1,NID2,NOX4,NR1H3,OSMR,PCK1,PCK2,PHGDH,PMP22,PPP1R15A,PRKN,PSAT1,SERPINF1,SHMT2,SLC1A5,SLC38A3,SLC3A2,SOD2,SSBP2,SST,STAT3,STC2,TKT,TNFRSF11B,TRIB3,VEGFA,WARS1 |
| RASSF1 | | other | | 1,368 | 1,378 | 0,002 | 1,30E-07 | 2,65E-06 | ABAT,APOE,BHLHE40,CARD8,CASP3,CASP4,CAV1,CAV2,CCNA2,CCND1,CCND2,CLU,COL4A1,COL4A2,CPE,CRIP2,CXCR4,DAB2,EFEMP1,EFNB2,EPB41L3,ESR1,FOS,GHR,HLA-DPA1,HOXA1,LITAF,MDK,MMP2,MRAS,PLK2,SPP1,SRPX,TUBB,TUBB2A |
| F2R | 0,190 | G-protein coupled receptor | Activated | 1,972 | 2,010 | 0,006 | 1,38E-07 | 2,80E-06 | BIRC2,CASP3,CASP4,CCND1,CD24,CD44,CD55,CDH11,CLDN3,CXCL12,DUSP1,ESR1,F11,F3,FBP1,FN1,FOS,GJA1,ICAM1,IGFBP7,KRT19,LAMB1,MCAM,MIF,MMP2,MMP9,NR4A1,RHOA,S100A4,SELP,TFPI2,TJP1,TP53,VCAM1,VEGFA,VEGFC,VIM,VWF |
| NEUROG1 | -0,079 | transcription regulator | Inhibited | -2,635 | -2,646 | -0,002 | 1,43E-07 | 2,88E-06 | ADD3,ASS1,C1orf115,C1S,C3,CCND2,CFH,COL3A1,CXCL1,FN1,GAS2,GASK1B,ITGB3,LCP1,MMP9,P4HA2,PAPPA,PLK2,PPIC,PXDN,RHOA,RIPOR2,S100A4,SERPINF1,SLC43A3,SQOR,TRIM22,TSPAN7 |

| | | | | | | | | | |
|---|---|---|---|---|---|---|---|---|---|
| NFKB1 | 0,370 | transcription regulator | Activated | 2,868 | 2,915 | 0,007 | 1,52E-07 | 3,06E-06 | A2M,AKR1B1,APOE,B2M,BAX,BCL2A1,BHLHE40,BIRC3,CCL19,CCL20,CCL5,CCND1,CCND2,CD3G,CD40,CD59,CD86,CFB,COL1A1,COL1A2,CRMP1,CTCF,CTSB,CXCL10,CXCL11,CXCL9,CYBB,DICER1,DROSHA,DUSP1,EHF,ENPP2,FAS,FCGRT,FOS,FOSB,FSCN1,GADD45A,GBP1,GJA1,GNAI3,GRK5,HLA-DMB,ICAM1,IFNGR2,IGHM,IGKC,IL13,IL18,IRF1,ISG15,LGALS1,LTB,MAT2A,MMP7,MMP9,MYB,NFKB1,NMI,NOD2,NOS2,NOX5,NR4A1,NR4A3,PRKCD,PTPN6,RACK1,SELP,SOD2,STAT1,TERT,TLR2,TNFSF10,TP53,VCAM1,VEGFA,WT1 |
| HMGA1 | -0,032 | transcription regulator | | 0,189 | 0,209 | 0,003 | 1,58E-07 | 3,16E-06 | ALPL,B2M,CAV1,CAV2,CCND1,CCND2,CD44,CLCN6,COL1A1,COL1A2,COL4A1,COL4A2,COX6A2,CPE,CSK,DAB2,DDIT3,DHCR7,ESR1,FOS,FSTL1,GHR,HCK,HS3ST1,ID3,IDI1,IGFBP4,IGFBP7,IL15,INSR,ITGAL,KITLG,KRT10,LAMB1,MAPT,MMP9,MYO1F,NKX2-5,NOS2,OXT,PTH1R,RHOB,SHC3,SLC27A2,SOX4,SPARC,STAT3,SYK,TFEB |
| miR-124-3p (and other miRNAs w/seed AAGGCAC) | | mature microRNA | | -1,476 | -1,556 | -0,010 | 1,63E-07 | 3,26E-06 | ACAA2,AHR,AK2,ALDH9A1,AP1M2,ARAF,ARFIP1,ARHGAP29,ARPC1B,CAV1,CD164,CD59,CDC14B,CERS2,CHP1,CYP1B1,DCTD,DRAM1,ECI2,ELF4,ELK3,ERH,GLI3,GNAI3,HADH,HTATIP2,IFRD2,IQGAP1,KLF15,LAMC1,LITAF,LMNB1,LRRC1,MAPK1IP1L,MDFIC,MMADHC,MTMR6,MYH9,NECAP2,OSBPL8,PGM1,PGRMC2,PLP2,PRKD1,PTPN12,PTTG1IP,RARG,RHOG,RYK,SEC11A,SLC17A5,SLC22A5,SMCO4,STAT3,SUCLG2,SWAP70,SYNGR2,TLCD3A,TMBIM1,TMEM248,TNFRSF21,TOM1L1,TSC22D4,TSKU,TSPAN15,TUBB6,ZCCHC24 |
| GAPDH | | enzyme | Inhibited | -2,731 | -2,748 | -0,004 | 1,66E-07 | 3,32E-06 | C2,CCL20,CXCL1,DUSP1,ENO1,FCER1G,HLA-C,IFI6,IFIT2,IFITM1,IFITM2,OAS1,OAS2,OAS3,SPP1,STAT1,TXNIP,UBE2L6,WARS1 |
| LIPE | | enzyme | | -1,007 | -1,000 | 0,002 | 1,68E-07 | 3,33E-06 | ACSL1,AQP1,ATF3,BCL6,CA2,CASP8,CD36,CD74,CIB2,COL1A1,COL1A2,DMPK,FKBP5,FOS,G0S2,GJA1,GRB10,HLA-DQA1,ID2,KITLG,MAPT,MEF2C,NET1,NR4A1,PC,PCK1,PDE8A,PDK4,PER1,PLIN2,POSTN,PPARGC1A,PRDX2,PVALB,RB1,RXRA,SCD,SMAD7,ST3GAL5,ST8SIA4,SUCLA2,TKT,TWF2 |
| let-7 | | microRNA | Inhibited | -3,355 | -3,428 | -0,010 | 1,70E-07 | 3,37E-06 | ALDH1B1,AURKA,CASP3,CBFB,CCNA2,CCNB1,CCND1,CCR7,CD44,CDK1,CLUH,COL1A1,COL1A2,COL3A1,DDX56,DICER1,DZIP1,EIF4G2,ERF,ETV2,EZH2,FARP1,FN1,FOSL1,GAB2,GPX7,ID2,IL6R,ITGB3,KRAS,MAD2L1,MCM2,MCM3,MCM5,MCM6,MCM7,MYD88,NAP1L1,NFKB1,ORC6,PBX2,PCGF2,PKM,PLAGL2,PROM1,RBM38,RCAN1,RFC2,RIOX2,RRM1,RRM2,S100A4,SCD,SLC25A13,SMAD2,SSR1,STAT3,TLR4,TRAP1,VIM,ZFP36 |
| CCR2 | 0,955 | G-protein coupled receptor | Activated | 3,893 | 3,932 | 0,006 | 1,82E-07 | 3,60E-06 | AEBP1,C1QA,C1QB,CCL5,CCR2,CCR5,CD163,CD40,CD86,COL1A1,COL1A2,COL4A1,COL4A2,COL6A3,CXCL10,CXCL12,CXCL6,CXCR4,DCN,FBN1,FN1,GPT,HTR2B,IL13,IL18,ITGAL,LUM,MMP2,MMP9,P4HA1,PCOLCE,PLOD1,SLPI,SPARC,TGFBI,TIMP1,TIMP2,TNC,TXLNA,VCAN |
| arachidonic acid | | chemical - endogenous mammalian | | -0,538 | -0,528 | 0,002 | 1,85E-07 | 3,63E-06 | ABCG1,ACACA,ACOX1,AQP3,BCL2A1,CD36,CD40,CD83,CD86,CLU,COL1A2,DDIT3,DNAJB9,ELOVL6,F3,FABP1,FAS,FOS,FOSL1,MRC1,PECAM1,PKLR,PLA2G4A,PLD1,PLTP,PNPLA2,PRKD1,PTGES,PTGS1,RXRA,SCD,SELP,SOAT1,TNFRSF1B,UCP2,VCAM1,VIM |
| NR1H4 | -0,097 | ligand-dependent nuclear receptor | | -1,145 | -1,148 | -0,000 | 1,86E-07 | 3,63E-06 | ABCC2,ABCC3,ACACA,ADH1B,ADRB2,APOA1,APOC3,APOM,BAAT,C3,CPT2,CRABP1,CYP2B6,CYP3A4,DDAH1,FABP1,FABP2,FAS,G6PC,GCLM,GPT,GSTA3,HNF1A,HNF4A,HSPA5,INSR,ITGAM,KNG1,LCN2,MMP2,MTTP,NOS2,NR0B2,NR1I3,PCK1,PGM1,PKLR,PLTP,PNPLA3,PPARA,PPARGC1A,PTGS1,RXRA,S1PR1,SDC1,SLC10A2,SLC27A5,SPP1,TCEA2,TIMP1,TIMP2,TPI1,TXNIP |
| SATB1 | 0,215 | transcription regulator | | -1,375 | -1,387 | -0,002 | 1,86E-07 | 3,63E-06 | ACTG1,ACTN1,BCL2A1,BLNK,CCND1,CD40,CD69,CD86,CD8A,CLEC2B,CTSD,CYBB,ENOSF1,EVI2A,FERMT2,FEZ2,FOSB,GADD45B,GLRX,GPR18,GTF2E1,HBB,HLA-DMB,HLA-DOB,HSPA8,ID3,IL10RA,IL13,IL4R,IL7,IL7R,IRF7,IRF8,KITLG,LAX1,MAF,MAL,NFE2L2,NIBAN1,PIK3IP1,RB1,RIPOR2,S1PR1,SELL,SGK1,SLC27A2,SS18L1,SUN2,TRIM22,UBE2L6,VIM,WARS1,XAF1 |
| FOXA1 | 0,206 | transcription regulator | Inhibited | -2,509 | -2,513 | -0,001 | 1,86E-07 | 3,63E-06 | AFP,AGR2,ALDH6A1,ANXA1,AQP3,ARHGEF9,CD58,CDX2,COL18A1,CSHL1,CXCL10,DDC,DSE,EFHD1,ELK3,EPHX1,ERBB3,ESR1,FABP2,FKBP4,FNDC3B,FSTL1,G6PC,GBP2,GLS,HADH,HK1,HNF1A,HNF1B,HNF4A,LHFPL6,LYN,MALT1,MGP,MLXIPL,MSH2,PCK1,PPARA,PRNP,RORA,RORC,S100A2,SLC12A2,SLPI,SOD1,SST,TFF3,TMPRSS2,TNNI1,XBP1 |
| FN1 | 0,617 | enzyme | | 1,829 | 1,869 | 0,006 | 1,87E-07 | 3,63E-06 | ACE,ACP5,ACTR2,AHCY,APOE,BIRC3,BNIP3L,CAPN2,CCL5,CCND1,CD83,CD8A,CDKN1B,CLIC4,COL1A1,CORO1A,CTSD,CXCL1,F3,FERMT2,FLNA,FN1,FOS,HK1,HLA-B,IL1R1,IL7,ITGAM,ITGAV,ITGB3,JUND,KRT7,MAPK8,MARCKS,MAT2A,MGP,MMP1,MMP2,MMP9,MTOR,NOS2,PCNA,PDGFRA,PPP6R2,RASSF2,RDX,RECK,SCRN1,SDC1,SDCBP,SMAD2,SMARCA4,SPARC,SPP1,SURF1,TCF4,TGFBR2,TIMP2,TLR2,TNFAIP2,UBE2B,VIM |

| | | | | | | | | | |
|---|---|---|---|---|---|---|---|---|---|
| BRD4 | -0,129 | kinase | Activated | 3,677 | 3,733 | 0,010 | 1,88E-07 | 3,65E-06 | ACSL5,ACSM3,ALDH1B1,BHLHE40,BTN3A2,CCNA2,CCND1,CCND2,CDKN1B,COL1A1,COL1A2,COL4A1,COL5A1,CXCR4,DCTPP1,FBN1,FKBP11,FN1,FOS,GINS1,ICAM1,ICAM2,ID2,IL7R,IRF1,IRF8,KAT5,LCK,LOXL1,MAL,MAP3K1,MCM3,MLF2,MYB,MYD88,NME1,POLE2,RRM2,SMARCA4,STC2,THBS2,TIMP1,TOP2A,TUBB,TYRO3,XBP1 |
| STAT5B | | transcription regulator | | -0,719 | -0,710 | 0,001 | 1,96E-07 | 3,79E-06 | A2M,ACOT11,AGPAT2,AHNAK,ALDH3A2,ASAP1,BAIAP2,BAX,BCL6,BIRC3,C8B,CA2,CASP3,CASP4,CASP6,CASP8,CCND1,CCND2,CCND3,CISH,COX6A2,CRADD,CSHL1,CYP17A1,CYP2A6 (includes others),CYP2B6,CYP2C8,CYP4A11,DDC,DDIT3,ESR1,FAS,FOS,G0S2,GADD45A,GADD45B,GJB1,GPD1,GPX7,GSTP1,HAO2,HSD3B2,IL2RB,IRF1,KRT19,LTB,MAF,MDH2,ME1,MOXD1,MPC2,MYL7,MZB1,NAT8,NNMT,NOX4,NPPA,NUCB1,PCSK5,PDK4,PLTP,PMM1,PPP1R3C,PRLR,PROM1,RPA1,SCD,SLC16A5,SLC34A2,SLC47A1,SOCS2,ST8SIA4,TLR2,TNNC1,TNNI1,TOX,TRIM24,WARS1,ZFP36 |
| XBP1 | 0,528 | transcription regulator | | 1,793 | 1,850 | 0,007 | 2,16E-07 | 4,17E-06 | ACACA,ACACB,ADAM10,AFP,APOA1,CALR,CAT,CCL5,COPB1,DDIT3,DDOST,DERL1,DNAJB9,DNAJC10,ESR1,FAS,FKBP11,FN1,GALK2,GGCX,GOLGA3,GOLGA4,GOSR2,HLA-DRA,HSP90B1,HSPA5,ICAM1,KDELR3,LMAN2,MGAT2,NOS2,NR1H3,NUCB2,P3H1,PDIA3,PDIA4,PDIA6,POU2AF1,PPIB,PRNP,RAB3A,RPN2,S100A6,S1PR1,SCD,SDF2L1,SEC11A,SEC24D,SEC31A,SEC61G,SLC1A5,SLC38A1,SMC3,SOD1,SPARC,SPCS3,SSR1,SSR4,STT3A,STX18,TRAM1,TXN,VAMP2,VCAM1,WNT10B,XBP1 |
| testosterone | | chemical - endogenous mammalian | | 0,606 | 0,647 | 0,005 | 2,22E-07 | 4,27E-06 | A2M,ACSL1,ADRA2A,ANXA1,ANXA3,ARAF,ARG2,AVP,BAX,BMP7,CASP3,CASP8,CAST,CAT,CAV1,CCND1,CDKN1B,CGA,CLCN3,CLU,COX6A2,CTSV,CYB5A,DCN,EGF,EIF4G3,ESR1,FAM89B,FAS,FGF7,FGFR3,FN1,FOS,FOSB,FOSL1,FOXO1,GADD45A,GMFB,GPX5,HIF1A,HMGCS2,HSD3B2,ID1,ID3,JUND,KRAS,LGALS1,LHB,MAL,MAOA,MMP7,MTTP,MX1,NDRG1,NGF,NOS2,OAT,PARP1,PCNA,PDIA4,PMP22,PPARA,PRKCZ,PRLR,PTP4A1,RARG,SDC1,SELP,SERPINA5,SHBG,SLC17A1,SLC25A10,SMAD2,SNRNP25,SOD2,SRD5A2,SST,SSTR2,STAT3,SYK,SYT11,TAC3,TAP2,TBXAS1,TG,TGFBR2,TLR4,TNFRSF11B,TOP2A,TUBB2A,UBE2B,VCAM1,VCAN,YWHAE |
| POU2F1 | | transcription regulator | | -1,779 | -1,769 | 0,003 | 2,25E-07 | 4,32E-06 | A2M,ACSL1,AOX1,BCL6,CCND1,CCNG1,CD55,CDX2,CSF1R,CYP2A6 (includes others),CYP2B6,CYP2C8,CYP2J2,DECR1,ELK3,ESR1,GADD45A,GBP2,GCLM,GPX3,GSTK1,GSTM4,HLA-DRA,HNF1B,HOMER3,ICAM1,IDH1,IGKC,JUND,NFKB1,NOS2,NXN,PRDX2,PTGES,SERPING1,SOD1,SPP1,TLR4,TMPRSS2,TNNC1,VCAM1,VWF |
| FGF19 | | growth factor | | 0,071 | 0,067 | -0,001 | 2,28E-07 | 4,35E-06 | ACACA,ACACB,ACSL1,AFP,APOE,CCND1,CD36,CYP17A1,ENO3,FBP1,FOS,G0S2,GADD45A,GAS2,HLA-DQB1,HSD3B2,ID1,KL,LCN2,LEPR,MCAM,MME,NR0B2,PKLR,PROM1,SCD,SERPINA3,SGK1,SLC10A2,SLC2A2,SLC2A5,SLPI,TNNI1 |
| RET | | kinase | | 1,774 | 1,804 | 0,006 | 2,39E-07 | 4,53E-06 | ANXA1,ANXA4,AUH,CALB1,CCL20,CCN3,CCND1,CD151,CDKN1B,CLU,COL1A1,COL1A2,CTSB,CTSV,CXCL12,CXCR4,DCN,DPYSL2,EIF4G3,FKBP4,FOS,GJA1,HSPA8,HSPA9,IL13,MAF,MET,MMP1,MMP7,MMP9,PLOD2,PRNP,S100A9,SELL,SST,ST13,STUB1,TACC3,TP53,UBQLN2 |
| KIT | 0,147 | transmembrane receptor | | -0,823 | -0,816 | 0,002 | 2,48E-07 | 4,69E-06 | ANKH,C3AR1,CCND2,CCND3,CCR2,CD164,CD44,CD53,CD63,CDKN1B,CISH,CPA3,CSF2RB,CXCR4,FCER1A,HSD17B10,ICAM1,IL13,IL15,IL15RA,IL4R,ITGAM,ITGB2,KITLG,MGST2,MTOR,PRKCQ,PSTPIP2,PTPN6,RAB32,SH3BP2,SHC1,SOCS2,SPP1,STAT1,STAT3,TJP1 |
| PTGER4 | 0,299 | G-protein coupled receptor | | -1,476 | -1,492 | -0,002 | 2,50E-07 | 4,72E-06 | CCL20,CCR7,CD40,CD44,CD69,CXCL10,CXCR4,CYBB,DDX58,FAM49A,GBP2,HERC6,ICAM1,IFI16,IFI35,IFIH1,IFIT2,IL18,IRF1,IRF7,ISG20,LHFPL2,LPAR6,MAFB,MAFF,MMP2,MMP9,MXI1,NCF2,NOS2,PDCD4,PDK2,PTGER3,RASSF2,RNASEL,RSAD2,RTP4,S1PR1,SLAMF8,SLC13A3,SPP1,ST6GAL1,ST8SIA4,TLR8,TNFSF10,TOR3A,TRIM21,XAF1,ZDHHC14 |
| APC | | enzyme | | 0,168 | 0,148 | -0,004 | 2,53E-07 | 4,77E-06 | CCND1,CCND2,CD44,CD55,CDX2,CLU,CXCL12,DHRS9,DPP4,FABP2,FH,GLI3,HIF1A,HNF1A,ID1,ID2,ID3,IDH3G,LCK,MDH2,MMP7,NOS2,OGDH,PARD3,PDK2,PDK4,PLA2G4A,PPARA,PRKCA,PRKCZ,PTEN,PTGS1,SDHB,SGK1,SPP1,SST,SUCLG1,SUCLG2,TFF3,TIAM1,TIMP1,TJP1,TNFRSF11B,TP53,VEGFA |
| ETS1 | 0,227 | transcription regulator | Activated | 3,501 | 3,545 | 0,007 | 2,66E-07 | 5,01E-06 | ARHGDIB,ARL4C,ARPC2,B4GALT5,BAX,BCL11A,CASP1,CAV1,CCDC90B,CCL5,CCND1,CCNG1,CD14,CD27,CD44,CD69,CDH5,CHUK,COL1A2,CSF1R,DNAJA3,FLI1,FN1,GZMA,GZMK,HPGD,HPSE,HSPA8,HSPB8,ICAM1,ID2,IL13,IL2RB,IL7R,ITGB2,ITGB3,ITGB6,ITK,JAK1,LAIR1,LCK,LTB,MET,MMP1,MMP2,MMP7,MMP9,MYB,NFIL3,PARP1,PERP,PRF1,PTPRN2,RUNX3,SELL,SPP1,SRGN,TBXAS1,TDP2,TGFBR2,THY1,TP53,VEGFA,ZEB2 |
| ATF6 | -0,007 | transcription regulator | | 0,282 | 0,318 | 0,008 | 2,70E-07 | 5,06E-06 | ACOX1,AURKA,CALR,CDKN3,CPT2,DDIT3,FOS,G6PC,HSP90B1,HSPA5,MANF,NPPA,NR0B2,NUCB2,PCK1,PDIA4,PLIN2,PPARA,PRDX2,PRNP,RCAN1,SLC35B1,SPARC,UBE2C,UNC13B,VEGFA,XBP1 |

| | | | | | | | | | |
|---|---|---|---|---|---|---|---|---|---|
| BIRC5 | 0,145 | other | | -1,728 | -1,720 | 0,003 | 2,87E-07 | 5,35E-06 | BAX,CASP3,CD53,CD74,CHUK,COL1A1,COL3A1,COL5A2,FBN1,FCGR2B,FN1,HLA-DQA1,HLA-DQB1,IGHM,IL13,IL2RG,JCHAIN,LUM,LYZ,MMP2,PTTG1,TP53,VEGFC,VIM,VWF |
| PRKCA | -0,237 | kinase | | 0,397 | 0,425 | 0,005 | 2,91E-07 | 5,43E-06 | APOBEC3G,APOM,ARHGDIB,ARL4C,ARPC2,AVP,BCL11A,BNIP3L,CAV1,CCND1,CD55,CHRND,COL1A1,CSTA,CTSB,CXCL10,CYBB,FN1,FOS,ICAM1,ID1,ID2,ID3,IFIT3,MMP1,MMP9,NFKB1,NOS2,NPPA,PLD1,PRKCD,PTEN,PTPRN2,RB1,RSAD2,S100A8,S100A9,SDCBP,SELP,TP53,VCAM1,VEGFA |
| 26s Proteasome | | complex | | 1,112 | 1,081 | -0,005 | 3,13E-07 | 5,82E-06 | AHR,ATG4A,ATP6V1D,BAG3,BAX,BIRC2,BIRC3,CCNA2,CDK1,CDKN1B,CLCN7,CTSB,CTSD,CTSF,DDIT3,DDX58,EGF,ESR1,FOSL1,FOXO1,GCH1,HMGB2,HSBP1,HSPB8,ICAM1,IFI35,MAD2L1,MICB,NFKB1,PRKCA,PRNP,PSMB5,PSMB6,PTEN,RB1,RUNX3,S1PR1,SMAD2,STAT1,TNFSF10,TOP2A,TP53,TXNIP,UBD,UBE2I,UBFD1,VIM,WBP2,WIPI1 |
| BAX | 0,426 | transporter | Activated | 2,312 | 2,346 | 0,008 | 3,57E-07 | 6,58E-06 | BAX,BIRC2,CASP3,CCL5,CCNB1,CDK1,CDKN1B,CTSB,CTSD,CTSK,CTSS,CXCL10,DNAJB9,HLA-A,IFITM2,IFNAR2,IL15,IRF7,JAK1,MMP9,PDIA6,TIMP1,TYK2,XBP1 |
| cholic acid | | chemical - endogenous mammalian | | 0,763 | 0,774 | 0,002 | 3,72E-07 | 6,81E-06 | ABCC2,ABCC3,ACE,ACOX1,APOA1,APOM,CCNG1,CDX2,CIRBP,CYP27A1,CYP2B6,CYP3A4,CYP4A11,DHCR7,EGF,EHHADH,EPHX1,FABP1,FBP1,G6PC,MDK,MYB,NR0B2,NR1H3,PCK1,PLTP,PPARGC1A,RBP1,SLC10A2,TM4SF5 |
| PTH | | other | | 0,859 | 0,895 | 0,006 | 4,15E-07 | 7,56E-06 | ADAMTS1,ADD3,AHNAK,ALPL,BHLHE40,CALB1,CCND1,CDKN1B,COL1A1,COL1A2,CXCL12,CXCR4,CYBA,CYP27B1,CYP3A4,DDIT3,DUSP1,EFNB2,FN1,FOS,FOSB,GALNT3,GJA1,HSPA5,IL33,IL6R,ITGAV,KL,LIMA1,MGP,MME,MMP1,MMP2,NFIL3,NR4A1,PALM,POSTN,PTH1R,SLC34A1,SLC9A3,SMAD7,SPP1,ST14,TFPI2,THBS4,TNFRSF11B,TNS1,TRPV5,VDR,WIF1,WNT10B |
| SCAP | -0,125 | other | | -1,416 | -1,370 | 0,010 | 4,16E-07 | 7,57E-06 | ACAA2,ACACA,ACACB,ACSL1,ACSL5,ALB,ASNS,ATF3,CSF1R,CYB5A,CYP2B6,CYP4F2,DDIT3,DHCR7,EHHADH,ELOVL6,HMGCS2,ID1,IDI1,IGHM,MSMO1,MTHFD2,NSDHL,PLA2G4C,PMVK,RDH11,REG1A,SC5D,SCD,TFF3,TM7SF2,TRIB3 |
| POU2F2 | | transcription regulator | | 1,323 | 1,374 | 0,010 | 4,39E-07 | 7,94E-06 | ADA,ALOX5,ARG2,ASB13,BANK1,BCL6,CCR5,CD69,DEPP1,DHRS9,ELL2,ENO1,GATM,GGA2,GPR183,HLA-DRA,HNF1B,IGKC,LCK,MCM7,MS4A1,NOS2,PFKFB3,SCD,SIRT3,SNRPB,SPP1,SSTR2,STAT3,TERT,TIMP2,TNFAIP2,TNFRSF21,XBP1 |
| CIITA | -0,091 | transcription regulator | Activated | 3,181 | 3,197 | 0,004 | 4,63E-07 | 8,34E-06 | B2M,BTN2A2,CCND1,CD74,COL1A1,COL1A2,GCNT2,HLA-A,HLA-B,HLA-DMA,HLA-DMB,HLA-DOA,HLA-DOB,HLA-DPA1,HLA-DQA1,HLA-DQB1,HLA-DRA,HLA-DRB1,IL13,MMP9 |
| SOD1 | -0,288 | enzyme | | -1,343 | -1,364 | -0,006 | 4,90E-07 | 8,81E-06 | ACO1,ACTB,APOE,ASL,ASS1,ATP1A3,ATP6V1G2,AURKAIP1,B2M,BAG3,BAX,BID,C1QA,CASP1,CASP3,CASP4,CAST,CCND1,CD14,CD36,CDK5,CGA,CLU,COL1A1,COMT,CSF1R,DAO,DCXR,FOS,GAK,GCLC,GCLM,GNAS,GNB1,GNG7,GPX3,GSTP1,GUK1,HPCAL1,HSBP1,HSPA8,HSPB8,ICAM1,IL15,JPT1,KCNAB2,MAOB,MET,MMP9,NDUFAF1,NETO2,NFE2L2,NFIL3,NOS2,PLCB4,PRRX1,PSMB5,PSMB8,RAC1,RDX,SH3BGRL3,SHANK2,SNCG,SOD1,SOD2,SPP1,STAT4,STMN1,TGFBR2,TP53,VCAM1 |
| IFNL4 | | cytokine | Activated | 2,950 | 2,979 | 0,010 | 5,07E-07 | 9,08E-06 | CXCL10,CXCL11,DDX58,IFIH1,ISG15,MX1,OAS1,OAS2,STAT1 |
| GLI1 | -0,186 | transcription regulator | | 1,839 | 1,900 | 0,008 | 5,23E-07 | 9,34E-06 | ABCG2,ACADVL,ARC,ARPC5,BCL2A1,BID,CAV1,CCND1,CCND2,CD24,CDH11,CLU,COL1A1,FAS,FGF1,FHL1,GMFG,GPC3,GPER1,HPSE,ID2,IGFBP6,IGFBP7,IMPA2,KRT19,MEF2C,MMP2,MMP9,NPDC1,NQO2,NR2F2,PCOLCE2,PDGFRA,PLOD1,PLPP2,PPT2,PRKCI,PSTPIP2,PTH1R,RGS10,RHOB,RNASEL,RRAS2,RXRA,S100A9,SFRP1,SH2B3,SH3GLB1,SPP1,TERT,TNC,TP53,TSC22D1,TTC13,TUFM,ULK1,VEGFA,VEGFC,VIL1,VIM,WIF1 |
| SMAD7 | -0,363 | transcription regulator | | -1,745 | -1,763 | -0,003 | 5,25E-07 | 9,37E-06 | ALDOB,APOC3,BIRC2,BMP7,BZW2,CCL5,CCNA2,CCNB1,CDK1,CDKN1B,CDKN2C,CKM,COL1A1,COL1A2,COL3A1,COL6A3,CXCL10,CXCL12,DCN,EFEMP1,FAS,FN1,FOXO1,GADD45A,ICAM1,ID1,ID2,ID3,ITGA7,LTBP3,MET,MMP1,MMP2,MMP9,NET1,NID2,NOS2,PCNA,PROM1,RAC1,SCD,SMAD7,TERT,TGFBI,TGFBR2,TIMP1,TXN,VEGFA |
| BTK | 0,188 | kinase | Inhibited | -2,494 | -2,487 | 0,001 | 5,27E-07 | 9,38E-06 | BAG1,BCL2A1,CCND2,CCND3,CCR7,CCT6A,CD2,CD38,CD40,CD69,CD86,CISH,CXCL10,CXCL9,CXCR4,ENPEP,FOXO1,IFI35,IFI44L,IFIT3,IFITM1,IL10RA,IL13,IL18,IRF1,IRF9,ISG15,ISG20,ITGAV,ITGB3,MTHFD2,MX1,MX2,NCL,OAS2,OAS3,RB1,SELP,STAT1,TP53 |
| NFAT5 | 0,054 | transcription regulator | | 1,344 | 1,387 | 0,008 | 5,91E-07 | 1,04E-05 | AKR1B1,AQP1,BCL2A1,C2,CD24,COL1A1,COL1A2,CXCL10,CYBB,F3,IGFBP7,IL6R,IRAK1,JAM2,LCN2,NFKB1,NOS2,NOX4,NUDT1,PFKP,PTGES,PTGS1,RRM2,S100A4,SEMA4C,SGK1,SPP1,TLR2,TNFAIP2,TNFRSF11B,TNFRSF21,VEGFA,VLDLR,VWF |
| ATF3 | -1,112 | transcription regulator | | -0,889 | -0,900 | -0,002 | 5,92E-07 | 1,04E-05 | ADIPOR1,ASNS,ATF3,AURKA,BHLHE41,BIRC3,CCL5,CCND1,CD69,CD83,CDK1,CGA,CXCL1,DDIT3,FBP1,G6PC,GBP2,HSPA5,ICAM1,ID1,KAT5,MMP1,MMP2,MSH2,NOS2,PCK1,PPP1R15A,PTGES,RSAD2,SLC15A3,SRI,STAT3,TLR2,TP53,XBP1 |

| | | | | | | | | | |
|---|---|---|---|---|---|---|---|---|---|
| CAV1 | 0,676 | transmembrane receptor | | 1,919 | 1,934 | 0,003 | 6,08E-07 | 1,07E-05 | ADAM17,AFP,CAV1,CAV2,CAVIN1,CCND1,CD14,CD36,CSF1R,CTSD,ESR1,FAS,FOLR1,FOS,GIPC2,HSPA8,ICAM1,ID1,IDO1,INSR,KRT19,KRT4,KRT7,MMP1,NEDD4L,NOS2,NPPA,NR4A1,NTRK2,PCNA,PDK2,PLA2G4A,PRLR,PROM1,PTEN,RAC1,SLC12A2,SLC1A1,SPAG9,ST6GAL1,ST6GALNAC4,TJP1,TMEM87A,TP53,VCAM1,WFDC2 |
| JAK1 | 0,628 | kinase | Activated | 3,052 | 3,095 | 0,009 | 6,37E-07 | 1,12E-05 | BID,CAPN1,CD14,CD40,CDKN1B,ENPP2,FOS,HIF1A,HLA-A,HLA-C,HLA-F,ICAM1,ID1,IFIT2,IL4R,IRF1,IRF7,IRF9,MMP2,MX1,OSMR,PSMB9,SLC1A3,SMAD7,STAT1,STAT3,TAP1,TNFRSF21 |
| HBA1/HBA2 | | transporter | Inhibited | -4,181 | -4,146 | 0,008 | 6,48E-07 | 1,13E-05 | ATP5MC2,COX15,COX6A2,COX8A,CYC1,HIF1A,NDUFA3,NDUFA7,NDUFB11,NDUFB7,NDUFC1,NDUFS3,NDUFS4,NDUFS7,NDUFS8,NDUFV1,SURF1,UCP2,UQCRC1,UQCRC2,VEGFC |
| IKBKG | | kinase | | 1,803 | 1,849 | 0,007 | 6,51E-07 | 1,13E-05 | AEBP1,BAX,BIRC3,C3,CCL5,CD247,CLU,CTSB,CTSF,CTSZ,CXCL10,CXCL12,CXCL6,DCN,ENPP2,FOS,GBP2,GCH1,GM2A,GPT,GRN,HIF1A,HLA-A,ICAM1,IFI16,IGFBP6,IRF7,ISG15,LCN2,MGP,NAGLU,NGF,NOCT,NPPA,PPARA,PTEN,RCAN1,SERPINE2,SFRP1,SGK1,TG,TNFAIP2,TNFRSF1B,TNFSF10,VEGFA |
| CREB1 | | transcription regulator | | -0,123 | -0,089 | 0,003 | 6,55E-07 | 1,14E-05 | ADM,AKAP1,APOE,APOLD1,ARC,ARHGAP33,ARL15,ATF3,ATP2B4,ATP6V0D1,AURKA,AVP,BAG3,BCAP29,BHLHE40,BNIP3,CA10,CACNA2D1,CCNA2,CCNB1,CCNB2,CCND1,CCND3,CD3D,CD9,CDA,CDH11,CDK14,CGA,COX8A,CPD,CRMP1,CRYM,CXCR4,CYFIP2,CYP46A1,DUSP1,EBP,EHD4,ESRRG,FABP2,FGF13,FGL2,FN1,FOS,FOSB,FRAT1,G6PC,GAD1,GADD45A,GADD45B,GADD45G,GALNT1,GLS,GPNMB,GPR3,HECA,HLA-A,HLA-DQB1,HLA-DRA,HLA-G,HPCAL1,HPGD,HSPA5,ID1,IDI1,INHA,IRF7,JPT1,KDELR3,KYAT1,LAPTM4B,LCN2,LITAF,MARCKSL1,MAT2A,MCAM,MCM5,MCM6,MCTP1,MMP1,MNS1,MRC1,MRPL49,MSMO1,MYO1B,NAB2,NDUFA10,NDUFV1,NFIL3,NOS2,NOX5,NR4A1,NR4A3,NRP1,OSTF1,OXT,PCK1,PCNA,PDGFRA,PER1,PLA2G4A,PPARGC1A,PPP1R15A,RAC3,RALYL,RB1,RIPOR2,ROBO1,RRAS2,SCD,SGK1,SHTN1,SIK3,SLC35G2,SOCS2,SOD2,SRPK2,SSBP2,SSR4,SST,STAT3,STMN1,SULT2B1,TAC1,TAGLN2,TG,TIPARP,TMBIM1,TSKU,TXN,VEGFA,VIM,ZFP36 |
| KLF3 | 0,160 | transcription regulator | | -1,940 | -1,968 | -0,003 | 6,84E-07 | 1,18E-05 | ACOT11,AHNAK,ALDH3B1,ANXA5,AP1M2,APOE,APRT,BCAP29,BCL2L2,BLOC1S1,BOLA1,CAPG,CARHSP1,CBFB,CCT3,CD1D,CD55,CD69,CD93,CDK10,CIRBP,CSNK1G3,ELMO1,EXTL1,FAM49A,FCGRT,FOS,GAD1,GALNT1,GSTK1,GSTP1,HBB,HEXB,HNRNPF,HS3ST1,IDH1,KBTBD2,LY96,MAGED1,MAN2A2,MAPK12,MCAT,MGST2,MIF,MRPL34,MSN,MTCH2,MZB1,NDRG1,NSMAF,OGFRL1,PATJ,PCYOX1,PDLIM1,PDLIM2,PECAM1,PFKP,PIK3IP1,PLPP2,PTP4A2,RMDN1,RNF167,SCP2,SDC1,SEC24D,SELL,SGK1,SIGIRR,SLC6A13,SMPDL3A,SP140,SSBP2,STAT4,TDRD7,TES,TNFRSF21,TOPORS,TRIM32,TUBA1A,TYROBP,UBL3,UGCG,VAV1,VIM,ZDHHC14,ZEB2,ZMAT5 |
| PRKAA1 | -0,043 | kinase | | -0,067 | -0,092 | -0,004 | 7,06E-07 | 1,22E-05 | ACKR3,ACOT7,ALB,ARHGDIB,ATF3,BAX,BIRC3,CA2,CAT,CD24,CDH3,CDKN1B,CLU,CXCL12,CYP2B6,DDAH1,DDIT3,DLGAP2,EAF2,EGF,FKBP5,FN1,G6PC,GHR,GSTA1,HSD3B2,HSPA5,ID2,IGFBP4,LITAF,MLYCD,MMP9,NCF2,OAT,PCOLCE,PDGFRA,PKLR,PPARGC1A,PRKACB,PSMB8,RND3,SIRT2,SIRT3,SOD2,TJP1,TP53,VCAM1 |
| KDM5A | | transcription regulator | Activated | 4,314 | 4,258 | -0,009 | 7,40E-07 | 1,28E-05 | ACO2,ATP6V1D,BCKDHA,CCND1,CD55,CDKN1B,COL1A2,DLAT,DLST,DMPK,GADD45GIP1,GHITM,GRPEL1,HSBP1,IDH3B,MAP2K3,MCAT,MCM2,MCM3,MLYCD,MRPL58,MTOR,MYOM2,NDUFA10,NDUFA2,NDUFA3,NDUFS2,NDUFS4,NDUFS7,NDUFV1,NIT1,NME1,OPA1,PGAM2,PMPCB,SDHB,SDHD,SOD2,TERT,TNNC1,TNNT2,TXN2,UQCRC1 |
| CCND1 | -0,313 | transcription regulator | Activated | 2,352 | 2,362 | 0,001 | 7,84E-07 | 1,35E-05 | AHNAK,ALDH1A3,AQP3,ARMCX1,AURKA,AZGP1,BCLAF1,C7,CALU,CCNA2,CCND1,CCND2,CCND3,CD44,CDKN1B,CDKN2C,CEP55,CLPB,COL1A1,COL5A2,CTNNBIP1,DNMT1,DTL,DYNC1LI1,EIF5,ENO1,ENPP2,FGF7,FLNA,GNMT,HSPA8,HSPB8,ID1,ID3,ITGAV,ITGB3,ITGB6,JPT1,KIF20A,KRT10,KRT4,LIN7B,MACF1,MAFF,MAGI2,MARCKS,MCM7,MEIS2,MELK,MMP2,MNS1,MYO7A,PAH,PBLD,PCLAF,PCNA,PDCD4,PDLIM4,POLD3,PTPRC,RB1,RRM2,SEC22A,SELENBP1,SERINC3,SLC4A4,SOX4,SPP1,STXBP1,TCF4,TGFBR2,TIAM1,TM7SF2,TMPRSS2,TMSB15A,TOR3A,TP53,TPI1,TPX2,TUBA1B,TUBB,TUFM,UAP1,USF1,UTRN,WWOX,ZNF423 |
| KLF15 | -0,313 | transcription regulator | Inhibited | -3,413 | -3,380 | 0,007 | 8,45E-07 | 1,45E-05 | ACADL,ACADM,ACADS,ACADVL,ACAT1,ACOX1,ACSL1,CD36,CPT2,DECR1,EHHADH,ESRRG,HADHA,MLYCD,NPHS1,NPHS2,NPPA,PDK4,PPARGC1A,PXMP2,SCD,SLC25A20 |
| CBX5 | 0,177 | transcription regulator | | -1,813 | -1,874 | -0,010 | 8,73E-07 | 1,49E-05 | AGR2,ALDH3A1,BCL11A,CD24,CKM,CYP1B1,CYP2J2,GASK1B,GCHFR,GCNT3,GDPD3,HBB,HOXB6,ICAM2,ITGA7,LCN2,LTB,LY6G6D,MMP7,MUC13,OAS1,PLAAT4,PSMB8,QPCT,QPRT,RIN2,RORA,SELENBP1,SERPINA3,SH3GL2,SLC2A10,SLC7A7,SLC7A8,TACSTD2,TFF3,TGFBI,TM4SF1,TSPAN13,TSPAN8,TXNIP |

| | | | | | | | | | |
|---|---|---|---|---|---|---|---|---|---|
| NR3C2 | 0,108 | ligand-dependent nuclear receptor | | 0,822 | 0,835 | 0,002 | 8,76E-07 | 1,49E-05 | ADAMTS1,APOE,ARPC2,BAX,BCL2A1,BID,CACNA2D3,CAT,COL1A1,COL3A1,CXCL6,CXCR4,CYFIP2,CYP1B1,DDC,ECHDC2,FKBP4,FKBP5,FN1,GHR,GPX3,GUCY1A1,HLA-DQB1,HMOX2,HSPA9,ICAM1,IDH3B,KLF9,LCN2,MMP1,MSRB1,NOX4,NPPA,PPIB,PRDX4,PROCR,PTPRCAP,SERPINA3,SGK1,TACC2,TGFBI,TP53,VEGFA,VLDLR |
| SUMO3 | 0,132 | other | | -1,385 | -1,393 | -0,002 | 8,96E-07 | 1,52E-05 | BACE1,CAV1,CAV2,CCN3,CCND2,CD38,CD59,CTH,CXCL10,HLA-G,HSPA8,ISG15,LAMP3,MOXD1,PNP,TAP1,TNC,TNFRSF21,XRCC5 |
| RIPK2 | | kinase | | 1,360 | 1,392 | 0,006 | 9,03E-07 | 1,53E-05 | ACACA,ACADL,ACADM,ACOX1,ACSL1,CCL5,CD36,CD40,CD69,CD86,CXCL10,DGAT1,FABP1,ICAM1,IL33,MARCKSL1,MTTP,NOS2,NR1H3,PNPLA2,PPARA,PPARGC1A,PTGES,RALGDS,RASGRP1,SCD,SOD2,VCAM1 |
| HRG | -1,390 | other | | 1,686 | 1,722 | 0,010 | 9,27E-07 | 1,57E-05 | ARF3,C1QA,C1QB,CD74,CD83,CXCL14,CXCR4,FCGR3A/FCGR3B,HLA-DQA1,MUC1,SPARC,SPP1,TIMP1 |
| PTGS2 | 0,230 | enzyme | Activated | 2,532 | 2,563 | 0,005 | 9,49E-07 | 1,60E-05 | ANG,ANXA1,ANXA2,AQP1,BAX,BIRC3,CCL5,CCND1,CCR7,CD163,CD44,CDK5,CDKN1B,CDKN2C,CLU,CTSD,CTSK,CXCL10,CXCL12,CXCL14,CXCR4,CYP1B1,DUSP1,DYNLL1,ERBB3,ESRRA,FN1,FOS,ICAM1,ID1,ITGAL,ITGAV,MMP1,MMP2,MMP7,MMP9,NOS2,PCNA,PLA2G4A,PTGER3,PTGS1,RAC1,RHOA,RPS19,SELL,ST3GAL1,TNFAIP6,TNFSF10,TP53,VEGFA,VEGFC |
| RAC1 | 0,256 | enzyme | | 0,695 | 0,729 | 0,007 | 9,53E-07 | 1,61E-05 | ACP5,APBB1,ATF3,BCL6,BID,CCND1,CD86,CKM,COL1A2,DPYSL2,DUSP1,ESR1,FAS,FN1,FOS,FOSL1,HIF1A,HOXA1,HSD11B2,ICAM1,IL13,IL33,IL6R,KIR3DL2,MAPK8,MMP1,MMP2,MMP9,NPPA,RAC1,RAC3,RHOA,SHC1,SOD2,STAT1,STAT3,VCAM1,VEGFA |
| miR-199a-5p (and other miRNAs w/seed CCAGUGU) | | mature microRNA | Inhibited | -3,552 | -3,603 | -0,010 | 9,62E-07 | 1,62E-05 | ALOX5AP,APOL1,C3,COL1A1,COL4A1,CSTB,DCN,DDR1,FN1,HIF1A,ID3,IFI27,ISG15,ITGA3,ITGB3,LCN2,MMP1,MMP7,MPP5,MX2,NEDD9,NFKB1,PIGR,RSAD2,SET,SMTN,TGFBR2,TGFBR3,TMPRSS2 |
| cytokine | | group | Activated | 2,606 | 2,657 | 0,008 | 1,00E-06 | 1,68E-05 | ALOX5,ANG,BAX,CCL5,CCN3,CCND1,CCR7,CD27,CD40,CD69,CD86,CD93,CDKN1B,CISH,CLU,CXCL1,DDIT3,DICER1,DUSP1,F3,FAS,FOS,HIF1A,ICAM1,IDO1,IRF1,MAPK8,MMP1,MMP2,MMP9,NGF,NOS2,PCNA,PLAAT4,S100A4,SELP,SGK1,SOCS2,SOD2,SPARC,STAT3,TIMP1,TLR2,TLR4,TNFAIP6,TNFRSF1B,VCAM1,VWF |
| Irgm1 | | other | Inhibited | -3,664 | -3,709 | -0,010 | 1,04E-06 | 1,74E-05 | CCL5,CCNA2,CCNB1,CCNB2,CXCL10,DTL,FAS,GINS1,ID2,ID3,IFI16,IFIT2,IFIT3,IRF7,KIF20A,OAS2,RRM2,RSAD2,SELP,SPAG5,UBE2C |
| CD28 | | transmembrane receptor | | 0,788 | 0,845 | 0,006 | 1,07E-06 | 1,78E-05 | ACTR3,AHR,ANXA1,ARHGDIB,BCL2A1,BHLHE40,BIN1,C2,CAMK2A,CASP3,CASP6,CASP8,CCND1,CCND2,CCND3,CCR5,CD247,CD27,CD69,CD86,CDKN1B,CISH,CXCR4,CYC1,DDX58,DUSP22,FCER1G,FOS,FYB1,FYN,GBP1,GBP2,GTF2IRD1,GTPBP4,GUCY1B1,GYS1,GZMA,GZMB,HAX1,HLA-DQB1,HSPA8,IFITM1,IFNGR1,IFRD2,IGFBP6,IL10RA,IL10RB,IL13,IL15RA,IL1R1,IL7,IL7R,IRF1,IRF8,ITGAL,LCP1,LSP1,MACROH2A1,MAF,MAP3K10,MMP2,NAMPT,NCL,NFKB1,NR4A1,ORAI3,PCYOX1,PHB,PKLR,PLA2R1,PNP,PRDX4,PSMA6,PTEN,RNF128,RORA,RORC,RYK,SLC16A3,SLC25A5,SNRPB,SOCS2,SRGN,STAT3,TFPI2,TNFRSF1B,TNFSF10,TOB1,TRIM24,TSC22D1,TSTA3 |
| IFNK | | cytokine | Activated | 2,684 | 2,714 | 0,009 | 1,11E-06 | 1,85E-05 | CD40,CD83,CD86,CXCL10,CXCL11,CXCL9,IFIH1,IRF1,MX1,OAS1,PLAAT4,STAT1,STAT4,TLR3 |
| NOSTRIN | | transcription regulator | Inhibited | -2,808 | -2,848 | -0,010 | 1,12E-06 | 1,86E-05 | ACE,ADAM17,ANXA5,CASP3,CAV1,CCL5,COL18A1,FGF1,FN1,ITGB3,MMP1,MMP2,MMP9,NFKB1,PECAM1,SELP,SOD1 |
| MYD88 | 0,260 | other | Activated | 5,053 | 5,125 | 0,009 | 1,16E-06 | 1,92E-05 | ACSL1,APOBEC3B,ATP6V1E1,ATR,C3,CASP4,CCL5,CD14,CD200,CD38,CD40,CD44,CD83,CD86,CISH,CLEC10A,CLIC4,CTSK,CXCL1,CXCL10,CXCL12,CXCL6,DUSP1,FANCF,FCER1G,FOS,FPR1,HCK,HIF1A,HLA-A,HSPA5,ICAM1,IFIT2,IFNA10,IL13,IL15,IL18,IL32,IL7,INPP5D,IRF1,IRF7,IRF8,ISG15,LPAR1,MAFF,MCM2,MET,MMP2,MMP7,MMP9,MRC1,MSH2,MSH3,NFKB1,NOD2,NOS2,PARP1,PILRA,PLA2G4A,PSTPIP2,PTGES,RSAD2,S100A8,SAMHD1,SLC16A3,SLCO3A1,SPP1,TFEC,TIMP1,TLR2,TSC22D1,VCAM1 |
| EBI3 | 0,197 | cytokine | Activated | 4,073 | 4,099 | 0,005 | 1,23E-06 | 2,03E-05 | B2M,CCND2,CCND3,CD86,FOS,GADD45G,GBP2,HLA-A,HLA-B,HLA-C,HLA-DMA,HLA-DMB,HLA-DOB,HLA-DPA1,HLA-DQA1,HLA-DRA,ICAM1,IL13,IRF1,MX1,RORC,STAT1,TAP1,TNFSF10 |
| KL | -1,014 | enzyme | | -0,534 | -0,546 | -0,003 | 1,25E-06 | 2,06E-05 | AXL,BAX,CAST,CCND1,CYBB,CYP27B1,CYP46A1,ICAM1,IHH,IRF7,KL,MMP7,MMP9,SELP,SLC34A1,SOD2,TNFRSF11B,TYRO3,VDR,VEGFA |
| DDX58 | 0,730 | enzyme | Activated | 2,955 | 2,998 | 0,009 | 1,27E-06 | 2,08E-05 | AHNAK,ALPL,CASP4,CCL5,CXCL10,DDX58,EGOT,IFI27,IFI35,IFI44,IFIH1,IFIT2,IFIT3,IRF1,IRF7,IRF8,ISG15,ISG20,LTBP3,MAT2A,NFATC4,NMI,OAS1,RSAD2,SPP1,STAT1,TLR3,TNFSF10 |

| | | | | | | | | | |
|---|---|---|---|---|---|---|---|---|---|
| Creb | | group | | -0,426 | -0,389 | 0,005 | 1,27E-06 | 2,09E-05 | AASS,ADM,ALOX5,ARHGEF9,ARPC5,ATF3,BACE1,C3AR1,CASP1,CCND1,CD14,CD59,CGA,CIDEC,CLMN,COMT,COX6A2,CSF1R,CTH,CTSB,CTSV,CXCL10,CYP17A1,DUSP1,EIF4EBP2,FAS,FCER1G,FOS,FOSB,FOSL1,FSCN1,G6PC,GABRR2,GDPD3,GJA1,GNAS,GSTM4,HLA-G,HPGD,HSPB8,IFI16,INHA,ITGAM,KLRB1,KPTN,LTBP3,MAPK8,MAT2A,MCAM,MCM3,MIF,MMP1,MMP2,MYB,NDN,NDUFA3,NEDD9,NOX5,NR4A1,NR4A3,NRL,PCK1,PCNA,PCOLCE,PER1,PHTF1,PLIN3,POU2AF1,PPARGC1A,PPFIBP1,PRKCA,PSENEN,SLC25A44,SLIT2,SOD1,SOX4,SST,SSTR2,TAC1,THAP1,TIA1,TNFRSF11B,VEGFA |
| MAP3K7 | | kinase | | 1,284 | 1,307 | 0,004 | 1,38E-06 | 2,25E-05 | BIRC3,CCL20,CCL5,CCR7,CD14,CFB,CXCL10,CXCL6,DDX58,GBP1,GBP2,GCH1,IFI44,IFIH1,IFIT2,IFIT3,IFITM1,IRF1,IRF7,MAP2K3,MMP1,NFKB1,NKX2-5,NPPA,OAS2,RNASEL,SLPI,TERT,TNFSF10,XAF1 |
| TFRC | -0,218 | transporter | | -1,920 | -1,961 | -0,008 | 1,41E-06 | 2,29E-05 | ADGRG1,ANXA2,ATF3,BNIP3,CASP3,CCNA2,CCNB1,CD300A,CDK1,CSTF2T,DDIT3,FOS,FTL,GADD45A,GPX3,GPX7,HSPA5,ID2,ITGAM,ITGB2,LY75,ME1,MS4A6A,PFKFB3,PHGDH,PPP1R15A,STC2,TNFSF10,TP53,UBE2E1,XBP1 |
| PNPLA2 | -0,199 | enzyme | Inhibited | -2,392 | -2,374 | 0,004 | 1,49E-06 | 2,41E-05 | ACAA2,ACADL,ACADM,ACADVL,ACOX1,ACSL1,CD36,GSTK1,HSPA5,ICAM1,INSR,MTTP,PCK1,PDK4,PPARA,PPARGC1A,UBE2B,UCP2,ULK1 |
| NRG1 | | growth factor | Activated | 2,123 | 2,147 | 0,004 | 1,50E-06 | 2,42E-05 | ACAT1,ACTN1,BDH2,BIN1,BNIP3,CAPN1,CASP3,CAV1,CAVIN1,CCND1,CCND2,CCND3,CDKN1B,CDX2,CHRND,COL1A1,COL6A3,CXCL6,CXCR4,DDIT3,DUSP1,ENO3,ERBB3,ETFB,FGF1,FN1,FOS,FOSB,FSTL1,GAD1,GPER1,GPS2,GPX3,HIF1A,ID1,IDH3B,IDI1,ITGAM,ITGAV,ITGB3,JUND,MAF,MMP9,NDUFS1,NKX2-5,NPPA,NR4A1,NR4A3,NUP214,PCCB,PDCD4,PGAM2,PKLR,PLG,RAB3A,SGSH,SHBG,SLC1A1,SOX4,UTRN,VEGFA,VEGFC,VIM,ZFP36 |
| SMAD3 | | transcription regulator | | 0,353 | 0,393 | 0,006 | 1,53E-06 | 2,47E-05 | ALB,APOA1,APOA4,APOC3,C3,CCND1,CCND2,CD59,CDH5,CKM,COL1A1,COL1A2,COL3A1,COL6A3,CXCL10,DDIT3,ENG,ESR1,F12,FN1,FOS,G6PC,GADD45B,GLS,GZMA,GZMB,HNF4A,HSP90B1,HSPA5,ID1,ID2,ID3,IL13,IL15,INHA,INPP5D,JAM2,JUND,LHB,LPAR1,LUM,MMP1,MMP9,NET1,NOS2,PC,PCNA,POR,PPARGC1A,PTPRK,RAC1,RHOB,S1PR1,SLC5A2,SMAD1,SMAD7,SPP1,SSTR2,STAT3,TERT,TIMP1,TNC,TPM1,VEGFA,VEGFC,VIM,ZEB2,ZFP36 |
| SFRP1 | 0,575 | transmembrane receptor | Inhibited | -2,635 | -2,630 | 0,001 | 1,57E-06 | 2,52E-05 | ACACA,BACE1,BAX,CASP3,CCND1,CD24,COL1A1,ELOVL6,FN1,G6PC,MLXIPL,MMP1,MMP2,MMP9,NAMPT,NR1H3,PCK1,PRLR,SCD,SFRP1,SLC2A2,TP53,VIM |
| Hbb-b2 | | other | Activated | 3,533 | 3,578 | 0,010 | 1,62E-06 | 2,60E-05 | C1QA,CCR2,CCR5,CD44,CD74,CLEC7A,CXCL10,CYBA,CYBB,FN1,FTL,IL10RA,IL2RB,IL2RG,ITGAM,LTB,LY86,NCF2,PRLR,SERPINF2,TLR2 |
| KDM1A | | enzyme | | 0,747 | 0,731 | -0,002 | 1,63E-06 | 2,61E-05 | ADM,AFP,ARHGDIB,ARNTL2,ATP6V0E2,AZGP1,BCL7A,C1orf116,CCL5,CCNA2,CCND2,CDA,CENPU,CHN1,COL1A1,COL1A2,CRYM,CSGALNACT1,CXADR,CXCL10,CXCL9,DNMT1,ELL2,ENTPD4,FIP1L1,FOXO1,GLT8D2,GNMT,HBB,HOXB7,ID2,IGFBP4,IQGAP2,KCNAB2,LCK,MCM2,MCM3,MCM6,MPC2,MYO1B,NCL,PPP1R12A,PROM1,PSAT1,PTEN,PTTG1,RARG,RBFOX2,RIN1,SCD,SERPINB1,SFRP1,SMAD7,SOCS2,TERT,TM4SF1,ZNF165 |
| Pro-inflammatory Cytokine | | group | | 0,414 | 0,443 | 0,008 | 1,67E-06 | 2,66E-05 | ABCC2,ADM,AKR1B1,APOE,ASS1,CCN3,CD47,CD55,CLU,F3,GCLC,GLUL,GSTA1,HDAC6,HSPA5,ICAM1,LCN2,LGALS1,MMP9,NFE2L2,NOS2,NR4A1,SELP,TIMP1,TLR2,UBD,VCAM1 |
| Mmp | | group | | -0,692 | -0,684 | 0,003 | 1,67E-06 | 2,66E-05 | AXL,CSF1R,DSG2,FAS,ITGAM,JAM2,MMP9,PLG,TJP1,VEGFA |
| E2F3 | 0,109 | transcription regulator | | 1,130 | 1,158 | 0,005 | 1,71E-06 | 2,71E-05 | CAV2,CCL20,CCNA2,CCNB1,CCNB2,CCND1,CCND3,CD9,CDK1,COL18A1,CSHL1,CXCL1,DEK,ECHDC2,EED,EZH2,FAM102A,FOXN1,HMGB2,ID3,JAM2,KRAS,LBR,MAD2L1,MCM2,MCM3,MCM5,MCM6,MCM7,MT1F,MT1G,MT1X,MYB,PCLAF,PCNA,PLCB4,PPP1R8,PTGER3,PTTG1,RB1,RBPMS,RRM1,RRM2,S100A8,TERT,TNFAIP2,TOP2B,TOPBP1,TP53BP2,TPX2,UBE2C,WFDC2,ZNF76 |
| glutamine | | chemical - endogenous mammalian | | -0,248 | -0,265 | -0,003 | 1,73E-06 | 2,74E-05 | ACTB,ADA,ASNS,ASS1,CCND1,COL1A1,CXCL1,DDIT3,DUSP1,FN1,FOS,GADD45A,GCDH,HIF1A,HK1,HSBP1,HSPA5,ICAM1,IDH1,IL15,IRF2,NR4A1,PCK1,PCK2,PCNA,PHB,PPARGC1A,PROM1,THBS2,TP53,VEGFA,VIM,WARS1,WT1,XBP1,ZMYND8 |
| Ca2+ | | chemical - endogenous mammalian | | -0,244 | -0,204 | 0,005 | 1,76E-06 | 2,79E-05 | AHR,ANKH,ARC,ATF3,CALR,CCND1,CCND2,CCND3,CCR2,CD83,CD86,CDKN1B,CGA,COL1A1,CUL9,CXCR4,CYP1B1,CYP27B1,DDIT3,DUSP1,FAAH,FN1,FOS,FOSL1,GJA1,GLS,GRK5,HOXA1,HSPA5,ICAM1,ID1,ID3,IL33,IL6R,IRF1,ITGA3,ITPR3,JUND,KRT10,LHB,MCAM,MGP,MMP9,MYB,NDRG1,NOX5,NPPA,NR4A1,NR4A3,NTRK2,PEBP1,PER1,PLS3,PPARA,PPARGC1A,PTTG1,S100A8,S100A9,SELP,SGK1,SLC12A3,SPP1,SST,SSTR2,TAC1,TCF4,TIMP2,TLR4,UGCG,VCAM1,VDR,VEGFA |

| | | | | | | | | | |
|---|---|---|---|---|---|---|---|---|---|
| miR-1-3p (and other miRNAs w/seed GGAAUGU) | | mature microRNA | Inhibited | -2,461 | -2,533 | -0,010 | 1,81E-06 | 2,86E-05 | ABHD11,ADAR,AGMAT,AGRN,ANP32B,ANXA2,ARF3,ARF4,ARHGAP29,ATP6V1B2,AXL,BCKDHB,BLCAP,CAP1,CDCP1,CDK14,CERS2,CLCN3,CNOT6,CORO1C,DHX15,DNAJB1,ESR1,FERMT2,FSTL1,GAK,GCH1,GJA1,IFT52,KCNJ2,KIF2A,LARP4,MET,MMD,MTHFD2,NELFCD,NETO2,NOTCH2,NRP1,OAT,PDCD4,PLEKHB2,PNP,PPIB,PTPRF,SFXN1,SH3BGRL3,SHTN1,SLC25A1,TAC1,TAGLN2,TLCD3A,TMSB10/TMSB4X,TPM1,UTRN,ZNF264 |
| RBL1 | | transcription regulator | | -1,046 | -1,075 | -0,007 | 1,83E-06 | 2,89E-05 | CASP4,CASP6,CCNA2,CCND1,CDK1,CDKN1B,CRADD,F3,FAS,FOS,FOXN1,KRT10,MCM2,MCM3,MCM5,MCM6,MCM7,MET,MT1G,MTOR,PCNA,PPARGC1A,PPP1R8,RB1,RRM1,RRM2,SFRP1,TERT,TP53 |
| ERBB3 | -0,315 | kinase | | 0,500 | 0,494 | -0,001 | 1,89E-06 | 2,97E-05 | ACTB,ADAM9,BAX,CCND1,CCND2,CD47,CDKN1B,CLU,COL1A1,CTSK,CXCL12,CXCR4,DUSP1,EIF6,ERBB3,F3,FBN1,FN1,GHR,GZMH,HIF1A,MXI1,PTEN,PTGS1,RRAS,SERPINA3,SFRP1,SLPI,SPARC,TNC,USP14,VEGFA |
| HSPA5 | 0,380 | enzyme | | -1,082 | -1,069 | 0,003 | 1,99E-06 | 3,10E-05 | ACADM,ACADVL,BAX,BCL2L2,CASP3,CCL5,CD47,CLU,CXCL10,DDIT3,ESR1,G6PC,HADHA,HSP90B1,HSPA5,HSPA8,IDI1,ISG15,P4HB,PTEN,SCD,XBP1 |
| IFNE | | cytokine | Activated | 3,135 | 3,171 | 0,009 | 1,99E-06 | 3,10E-05 | ANGPTL3,APOBEC3G,BST2,CD40,CD83,CD86,CXCL10,CXCL11,CXCL9,HEXB,IFIH1,IFIT2,ISG15,ISG20,MX2,PLAAT4,STAT1,STAT4,TLR3,TRIM5 |
| D-galactosamine | | chemical - endogenous mammalian | | 1,546 | 1,583 | 0,008 | 1,99E-06 | 3,10E-05 | ALB,BAX,CASP1,CASP8,CD14,CTSB,CXCL10,FOS,GPT,ICAM1,IL18,IL33,JUND,NFE2L2,NOS2,SDC3,SOD2,TIMP1,TLR4,TNFSF10 |
| VHL | | transcription regulator | | 0,272 | 0,255 | -0,004 | 2,30E-06 | 3,57E-05 | ADGRG1,BHLHE40,BNIP3,CAV1,CCND1,CDCP1,CDKN1B,CDKN1C,CPT2,CXCL12,CXCR4,DAB2,DECR1,EIF5,F3,FN1,FTL,GAL3ST1,HIF1A,HNF1A,HYAL1,MAD2L1,MAF,MIF,MXI1,NEDD9,NOX4,PCNA,PFKFB3,QPRT,RHOF,SEC61G,SLC16A3,SLC1A3,SOD2,SPARC,TMED3,TNFSF10,TP53,VCAM1,VEGFA,VEGFC,VIM |
| FANCC | -0,126 | other | | -0,202 | -0,213 | -0,004 | 2,33E-06 | 3,61E-05 | ANKRD6,CDK1,CXCL12,FHL2,FZD2,FZD4,FZD6,HHEX,HNF1B,IFI44L,IL1R1,MMP7,MX1,MYOF,NOS2,NUAK1,OAS1,OAT,PTPRC,RLN2,RND3,SFRP1,SKAP2,TACSTD2,TBC1D9,TOM1L1,TP53 |
| cholecalciferol | | chemical - endogenous mammalian | Activated | 3,200 | 3,232 | 0,005 | 2,43E-06 | 3,75E-05 | ADGRG1,ALOX5,ANXA2,CCL18,CCL5,CD14,CD300A,CD38,CD40,CD83,CD86,CDA,CDKN1B,COL1A1,CSTF2T,CTSK,CXCL10,FOS,ITGAM,ITGB2,LY75,ME1,MS4A6A,NDUFAF4,NR0B2,PDGFRA,PRKX,PTH1R,PVALB,RB1,RORC,RUNX3,SPP1,TGFBR2,TNFRSF11B,TNFSF10,UBE2E1,VDR,WT1 |
| TCL1A | | transcription regulator | Activated | 2,184 | 2,200 | 0,007 | 2,51E-06 | 3,86E-05 | C1QA,C1QB,C3AR1,CPA3,CTSB,CTSD,CTSS,CYBB,FCER1G,FCGR2B,FKBP5,FOS,FPR1,GPNMB,IGSF6,ITGAM,ITGB2,LCN2,LTF,MAFB,MAN2C1,MMP9,NETO2,NEU1,S100A8,S100A9,SLC15A3,STEAP4,TLR7,TNFAIP2,TREM2,UTRN |
| DUSP1 | -1,149 | phosphatase | | 0,843 | 0,839 | -0,001 | 2,58E-06 | 3,95E-05 | CCL20,CD40,CD86,CFB,CGA,CIDEC,CLEC10A,COL3A1,CXCL1,CXCL10,CYBB,CYP17A1,DUSP1,FOSL1,ICAM1,IFIT3,IRF1,ISG20,LHB,LTF,LUM,MMP1,MRC1,MX1,NOS2,NRP1,PLIN2,PTEN,PTPRK,RETSAT,S100A9,SOD2,SPRYD7,TLR2,UBD,VCAM1,VEGFC,ZFP36 |
| MAP2K4 | -0,074 | kinase | | 1,067 | 1,103 | 0,009 | 2,63E-06 | 4,01E-05 | ATF3,BIRC3,CCL5,CCND1,CD44,CD86,CXCL10,FAS,FLNA,FOS,GADD45A,KIF2A,MAPK8,MEF2C,MMP1,NOS2,NPPA,PAK2,PLA2G4A,ROCK1,STAT3,TP53,TUBB2A,VDR |
| NOTCH1 | | transcription regulator | | 1,790 | 1,801 | 0,001 | 2,67E-06 | 4,06E-05 | AQP3,BCL6,BIRC2,BIRC3,CCND1,CCND2,CCND3,CD14,CD44,CD84,CDKN1B,CDKN1C,CFD,CKM,COL1A1,COL1A2,CSF1R,CSF2RB,CXCL1,DLD,ECHS1,ECT2,EFNB2,ERBB3,ESM1,FCER1G,FCGR2B,FOS,FOSB,FOSL1,G6PC,GLS,GLUD1,GRN,GYPA,HIF1A,HSPA8,ICAM1,ID1,ID2,IFI16,IGFBP4,IL18,IL7R,IRF1,IRF8,ITGAM,MCAM,MMP1,MMP2,NCF4,NDUFS1,NFKB1,NOS2,NR2F2,OAT,PDIA3,PTEN,PTPRK,RB1,RND3,RORC,RUNX3,SPP1,SPRY2,TFF3,TGFBR2,TGFBR3,TP53,TUG1,VEGFA |
| thyroid hormone | | chemical - endogenous mammalian | Inhibited | -2,974 | -2,975 | -0,000 | 2,68E-06 | 4,07E-05 | ABCG1,ACE,ACSL5,AFP,ALDOB,APOA4,APOE,CCNA2,CCND1,CD2,CDKN1B,COMT,CPE,CRAT,DDAH1,DDC,ECH1,EGF,FOS,FZD6,G6PC,GCH1,GHR,HR,ID1,IGFBP4,IGHM,INHA,IRF2,KLF9,ME1,NPPA,NR1H2,NTRK2,OAT,OXT,PCK1,PER1,PHYH,PIP4K2A,PKLR,PPARA,PTEN,RYK,S100A10,SERPINE2,SSRP1,SST,ST14,STAT1,THRB,UCP2,VIM,YWHAE |
| HOXA9 | | transcription regulator | | 1,804 | 1,825 | 0,005 | 2,74E-06 | 4,16E-05 | ADD3,ADGRG1,ARPC5,BCL7A,CALB1,CALCRL,CBX6,CD36,CD93,CELF2,CISH,CLCN7,CLIC2,CSNK1A1,CTH,CYBB,DLGAP5,FCGRT,FZD5,GLI3,GLRX,GPR183,HOXB2,ID2,IL7R,INPPL1,LY86,LYN,LYZ,MAFB,MAN2A2,MYB,OSBPL8,PCDH1,PPP1R16B,PRKD3,PTPRC,PTPRCAP,RAB31,RDX,RPP40,S100A8,SDCBP,SERPINB9,SOX4,SP1,ST13,TSC22D1,USP1,VIM,WNT10B,ZNF22 |
| EIF2AK3 | -0,056 | kinase | | -0,379 | -0,344 | 0,008 | 2,87E-06 | 4,33E-05 | ALDH18A1,ANG,ASNS,ATF3,BIRC2,BIRC3,CCL20,CCND1,CLCN3,CYB5R1,DDIT3,DNAJB9,FOSL1,GADD45A,GCH1,HSPA5,HSPA9,LONP1,MECR,NDRG1,NFIL3,NOS2,PCK2,PON2,PPP1R15A,RNASEL,SHMT2,SNW1,TARS1,TP53,TXNIP,UIMC1,VEGFA,WARS1,XBP1,ZFP36 |

| | | | | | | | | | |
|---|---|---|---|---|---|---|---|---|---|
| PRKCD | -0,442 | kinase | | 1,743 | 1,777 | 0,005 | 2,87E-06 | 4,33E-05 | APOBEC3A,ARHGAP32,BCL2A1,BIRC2,BIRC3,BNIP3,CCL20,CCL5,CCNA2,CCND1,COL1A1,COL1A2,CTSB,CXCL10,CXCL12,DDIT3,FBN1,FOS,FOSL1,HRH1,ICAM1,IL13,IL2RG,IL32,ITGAM,ITPR3,JCHAIN,MMP2,MMP9,NOCT,NOS2,NOTCH2,NR0B2,NR4A1,OAS1,PDCD4,PLAGL2,PLSCR1,PPP1R15A,PROM1,PTPN6,RCAN1,SIK3,SLC22A1,SPRY1,STAT3,TLR2,TNFRSF11B,TP53,TPBG,TUBB2A,UGCG,VCAM1,VEGFA,ZFP36 |
| RHO | | G-protein coupled receptor | | -1,922 | -1,914 | 0,002 | 2,88E-06 | 4,33E-05 | ACADM,AQP1,B2M,C1QA,CACNB2,CASP3,CD44,CD74,COL1A2,CRABP1,CSF1R,CTSS,DDIT3,FOS,GNB1,HCLS1,HMGB2,IMPDH1,MEF2C,PCOLCE,PDE6A,PLA2G7,S100A6,SERPING1,SGK1 |
| MEF2D | | transcription regulator | | -0,982 | -0,959 | 0,005 | 2,88E-06 | 4,33E-05 | BACH2,BCL2L2,CCND1,CCND3,CKM,COL1A2,COL3A1,FN1,FOS,FOSB,JUND,MCM3,MCM5,MCM6,NPPA,NR4A1,PCNA,PPARA,PPARGC1A,PTEN,SYK,TNNC1,TNNI1,VIM,ZFP36 |
| N-cor | | group | | 0,832 | 0,801 | -0,008 | 3,00E-06 | 4,51E-05 | ACACA,ACACB,ACSL1,ASAP1,BIN1,C1QA,CCL5,CCND1,CD36,CIDEC,CSHL1,CTSB,CTSS,CXCL10,CXCL14,DHRS3,ELOVL6,ESR1,FOLR1,GSTP1,IDH1,INPP4B,LHPP,MCTP1,MGA,MMP2,MMP9,PCK1,PLIN2,RHOB,SCD,SEMA4D,SGK1,TBX2,WIPF1 |
| PARP1 | 0,295 | enzyme | | 0,826 | 0,855 | 0,005 | 3,07E-06 | 4,60E-05 | BAX,CASP4,CCL5,CCND1,COL3A1,CXCL1,CXCL10,CXCL12,DNAJC12,FAS,FKBP5,FOS,GBP2,HRH1,ICAM1,ITGAL,KRAS,LGALS3BP,MMP2,MMP9,NAT1,NOS2,PARP1,PVALB,RB1,REG1A,RPA1,SELP,SH3BP2,SOCS2,STAT1,TIMP2,TLR1,TLR4,TMPRSS2,TMSB15A,TP53,VCAM1,VEGFC |
| CEBPD | -0,369 | transcription regulator | | 0,450 | 0,474 | 0,005 | 3,12E-06 | 4,66E-05 | ALB,ALOX5AP,APOC3,ASNS,BCL2A1,C3,CCL20,CCNB1,CCNB2,CCND1,CD14,CDKN1B,CDKN1C,CLU,CSF1R,CTSV,CXCL1,CXCR4,CYP2A6 (includes others),DDIT3,FOS,HIF1A,ITGAM,NGF,NOS2,PDGFRA,RB1,SLC5A12,TLR8,TNFAIP6,TP53,VEGFC,VLDLR,XBP1 |
| PDLIM2 | -0,527 | other | | -0,111 | -0,115 | -0,001 | 3,23E-06 | 4,81E-05 | BST1,CCL20,CCND1,ENPP2,F2RL1,FBP1,FOXRED2,FSTL1,GHR,HLA-DOB,IFI44,IFIT3,IL7,KCTD12,KYNU,LGALS3BP,MECOM,MME,MPZL2,NPR3,OAS3,PCSK1N,PDLIM2,PHLDA3,RNF128,SH3GL2,SLC43A3,STAT1,STAT4,STC2,TNS1,TXNIP,WWOX |
| EGR2 | -0,321 | transcription regulator | | -0,166 | -0,113 | 0,008 | 3,38E-06 | 5,01E-05 | ABCC1,ASS1,BCL6,BHLHE40,BNIP3L,CAST,CDH6,CRABP1,CSF1R,DDN,DHCR7,DLG3,EIF5B,FOS,GHR,GJB1,HSPA5,ID2,ID3,IL6R,IL7R,KITLG,LHB,MME,MR1,MYB,NAB2,NCL,NSF,NUP54,PLCL1,PMP22,PPT2,PRKCD,PRX,ROBO1,RORA,RORC,SCD,SLC12A2,SLC1A3,SMAD1,TGFBR3,TNFSF10,TOP2B,TXNRD2,VEGFA,ZEB2 |
| CDH1 | 0,218 | other | | -0,987 | -0,982 | 0,001 | 3,40E-06 | 5,01E-05 | ACTB,APOA4,BIRC2,BMP7,CCND1,CD44,CDH3,ERBB3,FABP2,FN1,FOSL1,HOXA1,IFI27,IFITM1,IL13,ITGB3,KRT19,KRT7,LCN2,MCAM,MMP1,MMP9,MTOR,PFKFB3,PTEN,S100A4,TPBG,VEGFA,VIM,XPC |
| CLEC11A | | growth factor | Activated | 2,204 | 2,219 | 0,006 | 3,40E-06 | 5,01E-05 | ALOX5,ALOX5AP,ANXA1,CAP1,CCR5,CD74,CPA3,CXCL12,CYP17A1,FHL2,FOS,GNB2,IL13,MIF,NFKB1,NR4A1,PMP22,S100A6,SLPI,STAT4,TAC1,TKT,TSC22D1 |
| TAL1 | | transcription regulator | | -0,351 | -0,349 | 0,000 | 3,59E-06 | 5,26E-05 | ACOT8,ADGRG1,ARAF,ARSA,AZGP1,BCL6,BHMT2,C3,C3AR1,CCNB1,CD69,CDH5,CDKN3,CENPU,CIB2,DSG2,FBN1,FCER1A,GALNT7,GINS1,GUCY1A1,GYPA,HADHA,HBB,HLA-A,HLA-B,HLA-C,HLA-E,HLA-F,HMGB3,HSD17B14,ICAM1,ID2,IL10RA,KIF20A,MAP3K1,MARCKS,MCM2,MCTP1,MELK,MUC1,MYB,NDN,NFKB1,NR4A3,OIP5,OXR1,PLEKHO1,PRSS8,PTDSS1,PTPRCAP,RBM7,RNASE6,SOX4,SPRY2,SYK,TNFSF10,TOB1,TOX,TSPAN7,TUBB4B,USP34 |
| mir-223 | | microRNA | | 0,835 | 0,847 | 0,002 | 3,84E-06 | 5,62E-05 | BST1,C7,CA4,CD180,CD36,CD93,CYB5A,DNAJC6,EGF,FCER1A,FGL2,GASK1B,GFPT1,GPR65,HLA-DOB,HPGDS,ICAM1,ID2,IFI16,INPP5J,LY96,MEF2C,MMP25,MSMO1,MUC13,MYOF,PARP1,POSTN,RAB38,S100A6,S100A9,SGPL1,SLC16A7,SLC29A1,SOAT1,SP3,STAT3,STMN1,TGFBI,TLR4,TLR7,TOB1 |
| nitric oxide | | chemical - endogenous mammalian | | -1,307 | -1,286 | 0,004 | 4,09E-06 | 5,98E-05 | ACE,AHR,ATF3,AVP,BAX,BIRC2,BNIP3,CCND1,CCNG1,CD44,CDKN1B,CXCL10,CXCL12,DDIT3,DUSP1,FAS,FGF7,FOS,GADD45A,GCLC,GCLM,GSTP1,HPGDS,HPRT1,IDO1,IL18,IL7R,MMP2,MMP9,NDRG1,NFE2L2,NOS2,PDIA4,PECAM1,PPARGC1A,PPM1D,PTGS1,RHBDF2,SFN,SGK1,SLC16A4,SOD1,SPP1,TIMP2,TP53,UBE2C,VEGFA,WT1 |
| TCF7L2 | | transcription regulator | | 0,850 | 0,928 | 0,009 | 4,19E-06 | 6,11E-05 | ABHD17B,ACAA1,ACADL,ACSL1,ADGRG1,ALDH3A2,ALOX5,ANXA1,AQP3,ARAP2,ARHGEF10,ASPA,BACE1,BIN1,CAMK2N1,CAP1,CARHSP1,CCND1,CCND2,CD24,CDKN1B,CERS2,CPD,CREB5,CYP2C8,CYP3A7,DDC,DOCK10,EHHADH,ENPP2,ENTPD5,ERBB3,ERF,EVI2A,EVI2B,FBP1,FH,G6PC,GCLM,GJB1,GLUL,GSS,HMGCS2,ID1,ID2,ID3,IDH1,IPO13,KLHL2,MAL,MAP4K4,MAP7,MCAM,ME1,MMP1,MMP7,MTMR6,NINJ2,PAPSS1,PCK1,PIP4K2A,PPARGC1A,PPP1R16B,PROP1,PRRG1,RAB31,RALGDS,RHOA,RHOG,ROCK1,SC5D,SEMA4D,SIRT2,SLC12A2,SLC27A2,SMAD7,SORBS3,SPP1,STK17B,STK39,TMEM123,TNS1,TSPAN15,VEGFA |

| | | | | | | | | | |
|---|---|---|---|---|---|---|---|---|---|
| miR-29b-3p (and other miRNAs w/seed AGCACCA) | | mature microRNA | Inhibited | -3,542 | -3,591 | -0,010 | 4,29E-06 | 6,21E-05 | BACE1,CAV2,CCND1,CNOT8,COL15A1,COL1A1,COL1A2,COL3A1,COL4A1,COL4A2,COL5A2,DCP2,FBN1,FRAT2,GMFB,HMGN3,LAMC1,NASP,PMP22,PPIC,PPM1D,PTEN,SPARC,TP53,TUBB2A,VEGFA |
| sodium chloride | | chemical - endogenous mammalian | | 0,664 | 0,703 | 0,008 | 4,29E-06 | 6,21E-05 | ACE,ADRA2A,AKR1B1,AQP1,AVP,CALB1,CASP3,CCL5,CYBA,CYBB,FOS,HIF1A,ICAM1,MAPK8,MPDZ,NOS2,NOX4,NPPA,NPY1R,OXT,PTGES,SLC12A3,SOD1,SPP1,TRPV5,VWF |
| TGFB2 | | growth factor | | 0,654 | 0,676 | 0,004 | 4,41E-06 | 6,36E-05 | AHR,BHLHE40,CD36,CD40,CD59,CD86,CDH11,CDH5,COL1A1,COL1A2,COL3A1,COL4A1,FAS,FN1,FOS,JAM2,MLLT3,MMP2,MMP9,NGF,NOS2,PDK4,PECAM1,PLOD2,PTEN,S100A8,SCD,SELL,SMAD2,SPP1,TGFBR3,TIMP1,TIMP2,TNFRSF11B,VCAN,VDR,VEGFA |
| MAP2K3 | -0,419 | kinase | | 0,238 | 0,276 | 0,009 | 4,44E-06 | 6,39E-05 | ARG2,BAX,CCL8,CDKN1B,COL1A1,COL1A2,COL3A1,CTSK,CXCL10,CXCL9,DDIT3,GSTP1,ICAM1,IRF9,ISG15,MAOB,MMP1,MMP9,NOS2,NPPA,PPARGC1A,RGS7,SGK1,SLC1A3,STAT1,TNFSF10,VEGFA |
| IL7 | 0,492 | cytokine | Activated | 2,425 | 2,465 | 0,007 | 4,48E-06 | 6,44E-05 | AHR,BAX,BLNK,CASP3,CCND2,CCND3,CD2,CD247,CD3D,CD3G,CD69,CD86,CD8A,CDKN1B,CISH,COL1A1,CTSK,CXCL1,CXCR4,DARS2,DPP4,FAS,FN1,FOXO1,GARS1,GSR,IGHM,IL13,IL1R1,IL7,IL7R,LTB,LYN,MAF,MLLT3,MMP9,MX1,PIK3IP1,RORC,S1PR1,SELL,SMAD7,SYK,TNFSF10,XBP1 |
| IL17A | | cytokine | Activated | 4,433 | 4,489 | 0,007 | 4,69E-06 | 6,72E-05 | ACTB,BCL2A1,C3,CCL20,CCL5,CCND1,CD14,CD163,CD40,CD83,COL1A1,COTL1,CTSK,CXCL1,CXCL10,CXCL12,CXCL6,CXCR4,ECHS1,FAS,FMO1,FOS,FOSL1,GADD45A,GBP2,HEXB,HSPB8,ICAM1,IL13,IL32,IL33,ISG15,LCN2,MAP2K3,MAP3K1,MLXIPL,MMP1,MMP2,MMP9,MRC1,NOS2,NRP1,OSMR,PECAM1,PPARA,S100A8,S100A9,SELP,SPSB1,STAT3,STEAP4,TGFBR2,TIMP1,TIMP2,TLR2,TLR3,TLR4,TNC,TNFRSF11B,TNFRSF1B,TNNC1,VCAM1,VEGFA,VEGFC,VWF |
| SMAD4 | | transcription regulator | | -0,248 | -0,223 | 0,004 | 4,69E-06 | 6,72E-05 | ACKR1,AFP,AGR2,AIM2,APOA1,APOA4,APOC3,BMP7,CBFB,CCL20,CCND1,CCND2,CDH3,CDH5,CDKN1B,CELF2,COL1A2,CTSD,CYP17A1,ENG,FGFR3,FN1,FOS,FSCN1,GADD45A,GADD45B,GJA1,HHEX,HNF4A,ICAM1,ID1,ID2,IHH,INPP5D,JAM2,LEMD3,LOC102724788/PRODH,LPAR1,MET,MMP1,MTOR,NAMPT,NKX2-5,OVOL1,PGRMC2,PILRA,POR,PTEN,PTPRC,PTPRK,RAC1,RUNX3,SAE1,SCD,SGK1,SMAD2,SMAD7,SSTR2,TIMP1,TNC,TNFAIP6,TPM1,VEGFA,VIM,ZFP36 |
| EBF1 | | transcription regulator | Activated | 2,149 | 2,188 | 0,007 | 4,75E-06 | 6,80E-05 | ACACB,ARAF,BCL6,CCL5,CHUK,COL1A1,CRABP1,CXCL10,CXCL12,ESR1,FOXO1,G6PC,GYS1,IGHM,IGKC,INPP5D,INPPL1,INSR,IRF1,IRF7,KLRB1,KRAS,MAPK10,MAPK8,MTOR,MYD88,NFIL3,PCK2,PFKP,PHKA2,PIK3R3,POSTN,PPARGC1A,PPP1R3C,PRKAG2,PTPRF,PYGB,SCD,STAT1,TLR2,TLR3,TLR4 |
| TXNIP | 0,648 | other | | 1,263 | 1,261 | -0,000 | 4,78E-06 | 6,83E-05 | BCL6,CCNA2,CCND1,CD36,CDKN1B,COL1A2,DHCR7,FABP2,G0S2,ICAM1,IL2RB,MTTP,NFKB1,NOX4,NPHS2,PCNA,PPARA,PPARGC1A,TXNIP,VEGFA |
| bisphenol A | | chemical - endogenous mammalian | | 1,120 | 1,158 | 0,006 | 4,95E-06 | 7,06E-05 | ALDH5A1,APEH,ARNT2,BAX,C3,CASP3,CDKN1B,CFD,CLU,CTSD,CXCL10,CXCL12,DNM1L,ENO1,ESR1,FABP2,FOS,GPER1,HOXA10,HSP90B1,LCN2,LCP2,MAOA,MDH2,MMP9,NR4A1,NR4A3,OXT,PCNA,PEBP1,PKM,POSTN,PPP1R10,PRDX2,RBM22,RBM4B,RBPMS,RPN2,SLC1A5,SLC22A6,SLFN12,SPG21,SRD5A2,STAT3,STK17B,TP53,TRA2A,TWF2 |
| diacylglycerol | | chemical - endogenous mammalian | | -0,469 | -0,447 | 0,010 | 5,02E-06 | 7,14E-05 | ACADM,ACOX1,CD36,FABP1,PRKCA,PRKCD,PRKCH,PRKCQ,RASGRP1,UCP2 |
| CYP1A1 | -0,018 | enzyme | | -0,265 | -0,262 | 0,001 | 5,21E-06 | 7,39E-05 | ACOT11,ANXA3,ANXA4,ARAF,CBX3,CCND1,CHP1,CNN2,CYP17A1,CYP2B6,ECI2,GAMT,GPT,GSTA1,KIF2A,KITLG,KRT19,KRT7,MAPT,MID1,NDUFV1,PTP4A1,SDC1,SDHB,SHANK2,SMARCA4,SMTN,SOCS2,SPAG9,ST3GAL4,TACC3,TNNT2,TSC22D1,VLDLR |
| TNFSF10 | 0,956 | cytokine | Activated | 3,287 | 3,296 | 0,002 | 5,21E-06 | 7,39E-05 | BAX,BID,BIRC2,BIRC3,CASP3,CASP8,CD14,CD69,CSE1L,CTSB,CXCR4,FOS,HLA-A,HLA-C,HLA-F,IFI16,IFI27,IFI6,IFITM1,IFITM2,IL13,IRF9,ISG15,ITGAM,MMP2,MMP9,PCNA,RCAN1,STAT1,TIMP1,TNFAIP2,TNFSF10,TP53,VEGFA |
| TLR7 | 0,469 | transmembrane receptor | Activated | 5,194 | 5,260 | 0,010 | 5,29E-06 | 7,50E-05 | ATF3,BANK1,BCL2A1,CCL20,CCL5,CCNB1,CCND1,CCND3,CCR7,CD1D,CD300A,CD38,CD40,CD44,CD69,CD83,CD86,CDKN1B,CFH,CREB5,CXCL1,CXCL10,CXCL9,ESM1,FCER1G,FCMR,FOSL1,ICAM1,IDO1,IFI35,IFI44,IFI44L,IFIT3,IFITM1,IFNA10,IL10RA,IL13,IL1R1,INPP5D,IRF1,IRF7,IRF9,ISG15,ISG20,MX1,MX2,MYD88,NCF4,NFKB1,OAS2,OAS3,PHF11,PTPN6,RSAD2,SLC12A3,SOD2,STAT1,TLR7,TP53,TRIM38 |

| | | | | | | | | | |
|---|---|---|---|---|---|---|---|---|---|
| SOX11 | | transcription regulator | | 0,467 | 0,493 | 0,005 | 5,45E-06 | 7,70E-05 | ADAM10,ADAM9,BCL7A,BST2,CASP6,CAV1,CCL5,CCND1,CD24,CD52,CD58,CD86,CD9,CNOT6,CUL1,CXCR4,DBN1,FAS,FCMR,HMGB3,IFIH1,LCK,MAPK8,MS4A1,MYO1B,NIPSNAP1,SKAP2,SMAD1,STAT1,TFDP2,TIAM1,TMSB15A,TUBA1A |
| SNAI1 | -0,052 | transcription regulator | | 0,464 | 0,458 | -0,001 | 5,46E-06 | 7,71E-05 | ACTN1,ACTR2,ACTR3,AXL,CCND1,CCND2,CCND3,CD55,CDC25B,CDH5,CLDN1,CLDN3,COL1A1,COL1A2,CXADR,ELF4,FLNA,FN1,FOS,FOSL1,HNF4A,HPGD,ID1,INHA,ITGA2,LIMA1,MLXIPL,MMP9,MUC1,PEBP1,PFKP,PTEN,RAB25,SDC1,SLC27A2,SLC29A1,SMAD7,SPARC,TGFBR2,TGFBR3,THY1,TPM1,VDR,WIPF1,ZEB2 |
| RUNX3 | 0,469 | transcription regulator | | -0,921 | -0,960 | -0,007 | 5,59E-06 | 7,88E-05 | ABCC1,ABCC2,ABCC5,BAX,CAPN6,CCND1,CCR7,CDH11,CDX2,COL4A1,CPVL,DIP2C,EXT1,FBN1,FLNA,FSTL1,FZD2,GRK5,IL13,IL18,ITGAL,ITGAM,MACF1,MID1,MMP2,MMP9,NUAK1,PAPPA,PXDN,SGPP1,SLIT2,SPP1,ST3GAL5,STC2,THY1,TIMP2,UGCG,VIM,VWF,WT1 |
| YY1 | | transcription regulator | | 1,494 | 1,491 | -0,001 | 5,64E-06 | 7,93E-05 | AURKA,BLNK,BTF3,CALR,CAPG,CBX3,CCNA2,CCNB1,CCNB2,CCR5,CD3D,CD93,CDKN2C,CEND1,CLU,COL15A1,COL1A1,CRABP1,CXCL12,CXCR4,CYBB,DMPK,DROSHA,EIF4G2,FAS,FCER1A,FOS,FUZ,GADD45G,GBP1,GPC3,HLA-DRA,HSPA5,ID2,ID3,JCHAIN,KIF20A,LMNB1,MAD2L1,MAT2A,MCM5,MCM6,MCM7,MMP2,NIPSNAP1,NKX2-5,NOTCH2,PCNA,PDGFRA,PEMT,PERP,PPP1R15A,PRC1,RAB8B,RND3,RRM1,SFXN1,TACC3,TCF4,TGFBI,TLN2,TNNC1,TNNI1,TOP2A,TP53,TRA2A,USP14,VAV1,VEGFA,VEGFC,VWF,WT1,YWHAE |
| NR1I3 | -0,714 | ligand-dependent nuclear receptor | | -0,913 | -0,870 | 0,008 | 5,94E-06 | 8,33E-05 | ABCC2,ABCC3,ACOX1,AHR,APOA1,CSHL1,CYP27A1,CYP2A6 (includes others),CYP2B6,CYP2C8,CYP3A4,CYP3A7,DIO1,EPHX1,FBRS,G6PC,GADD45B,GSTM4,GSTP1,HK1,HNF4A,ICAM1,IL1R1,MAFF,NEDD9,PCK1,POR,PSMB7,RARRES1,RPA2,SCD,SLC29A1,STEAP4,SULT2B1,TMPRSS2,TNFAIP2 |
| SPARC | 0,704 | other | Inhibited | -2,260 | -2,269 | -0,002 | 5,94E-06 | 8,33E-05 | ARG2,BARD1,BHLHE40,CCL19,CDH3,CFD,CIDEC,CLDN1,COL1A1,COL1A2,CTSF,DVL1,FN1,GPD1,HSF4,MCM6,MMP2,MPZL1,MSH2,NDRG1,NIPSNAP2,NOTCH2,PCGF2,POLE2,PPIH,RIC8A,RRM1,SLC22A5,SLC34A2,SPARC,TMEM45A,TOPBP1,TRIM5,TSKU,USP2,VIM |
| TWIST1 | 0,104 | transcription regulator | Activated | 2,755 | 2,777 | 0,003 | 5,97E-06 | 8,36E-05 | ADAMTS1,AXL,BAX,C3,CASP3,CD40,CD44,CLU,COL1A1,CXCL12,DCN,DKK3,EFNB2,EMP3,ESR1,EZH2,FAS,FGFR3,FN1,FOS,FOSL1,FOXO1,IGFBP4,ITGB3,MET,MME,MMP1,MMP2,OSGIN1,PCOLCE,PDGFRA,PDZD2,PROM1,PXDN,RHOA,RUNX3,SDC3,SLC29A1,SPP1,TGFBI,TGFBR2,THBS2,TNFRSF21,TP53,VCAN,VEGFA,VIM,ZEB2 |
| glucosamine | | chemical - endogenous mammalian | | 1,921 | 1,909 | -0,003 | 6,40E-06 | 8,94E-05 | ACACA,ACADM,ASS1,ATP5MC2,COX6A2,CXCL1,DDIT3,GPD1,HSP90B1,ICAM1,INSR,LEPR,MDH2,NOS2,NPPA,PCCB,SLC25A11,SLC25A5,STAT3,TP53,TXNIP,UQCRC2 |
| mir-155 | | microRNA | Inhibited | -3,644 | -3,675 | -0,005 | 6,52E-06 | 9,10E-05 | ADAM10,ANXA2,BCL6,CCL18,CCL5,CCND1,CD47,CD69,CLDN1,CXCL10,DNAJB1,HIF1A,IL13,IL13RA1,IL7,INPP5D,IRF7,IRF8,IRF9,MAF,MME,MSH2,MSH6,MX1,MYB,MYD88,NOS2,PDCD4,PSIP1,PTEN,RORA,S1PR1,SDCBP,SMAD2,STAT1,STAT3,TP53,TRIM32 |
| ALDH1A2 | 0,220 | enzyme | | -0,420 | -0,432 | -0,003 | 6,56E-06 | 9,14E-05 | ABCG2,ALDH1A3,ATF3,CD44,COL1A2,COL3A1,HOXA1,IGFBP4,MAFB,NKX2-5,NPPA,PCOLCE,SPARC,SPP1,TGFBI,TIMP1 |
| Tnf (family) | | group | Activated | 2,735 | 2,770 | 0,006 | 6,65E-06 | 9,24E-05 | ABCC3,ATF3,BCL2A1,BIRC2,BIRC3,C3AR1,CAVIN3,CCL5,CCL8,CCND1,CCR5,CD163,CD40,CD86,CIRBP,CXCL1,CXCL10,CXCL11,CXCL12,CXCL9,CYP2B6,CYP3A4,ENPP2,ESM1,F3,GHR,GJA1,HIF1A,ICAM1,IL1R1,IL33,KL,LCN2,MMP9,NOS2,NOX4,NOX5,PCNA,PLEK2,PTGER3,SELL,SHBG,TLR2,TNFRSF1B,VCAM1 |
| HMOX1 | -0,101 | enzyme | | 0,625 | 0,615 | -0,002 | 6,94E-06 | 9,63E-05 | ABCG1,ANGPTL3,AQP1,CASP3,CCND1,CD14,CDKN1B,COL1A1,CXCL1,CXCL10,CXCL12,CXCL9,ENG,FGF1,FOS,GCLC,GPT,GPX3,ICAM1,ID1,ID3,ITGAM,ITGB3,MAPT,MMP9,NFE2L2,NOS2,NPPA,PCNA,PECAM1,PITX3,PROCR,RORC,SLC25A5,SPARC,THBS2,TP53,VCAM1,VEGFA,ZFP36 |
| hyaluronic acid | | chemical - endogenous mammalian | | 1,110 | 1,134 | 0,004 | 7,01E-06 | 9,70E-05 | ACKR3,CCL5,CCR7,CD40,CD44,CD83,CD86,CDKN1B,CSF1R,CTSK,CXCL10,CXCR4,FN1,FOS,ICAM1,LY96,MET,MMP1,MMP2,MMP9,MYD88,NCF2,NFKB1,NOS2,NR4A1,PCNA,PDCD4,PTEN,PTGES,PTPRC,RHOA,SPP1,TIMP1,TIMP2,TLR2,TLR4,VCAM1 |
| PTK2 | 0,012 | kinase | | 1,048 | 1,066 | 0,004 | 7,02E-06 | 9,70E-05 | BIRC2,BIRC3,CCND1,CDKN1B,COL1A1,COL1A2,FN1,FOS,MMP2,MMP7,MMP9,NPPA,P4HA1,PALLD,PCOLCE2,PRRX1,SERPINB1,SPP1,TIMP1,TJP1,TNC,TP53,VCAM1 |
| BMP2 | 0,022 | growth factor | | 1,004 | 1,045 | 0,006 | 7,48E-06 | 1,03E-04 | ACP5,ALPL,AOC1,ATP2C2,BHLHE40,CALB1,CCND1,CDKN1B,CLDN3,CLIC4,COL1A1,COL1A2,CRYM,CTSK,CXCL12,CYP17A1,EGF,EPHX2,ESRRG,F3,FN1,FOS,FOSB,FOXO1,GADD45B,ID1,ID2,ID3,IFI16,IHH,KLF9,LRP5,MEF2C,MMP9,MRAS,NFYB,NKX2-5,NPPA,POSTN,PTH1R,RGS6,RUNX3,SLC26A2,SMAD1,SMAD7,SPARC,SPP1,STK25,TAC1,TBX2,TG,TIMP1,TNFRSF11B,TOB1,VCAN,VDR,VEGFA |

| | | | | | | | | | |
|---|---|---|---|---|---|---|---|---|---|
| MED1 | | transcription regulator | | 0,312 | 0,354 | 0,008 | 7,96E-06 | 1,10E-04 | ACAA1,ACACA,ACOX1,AOC1,AURKA,BAX,BCL11A,BCL6,BCL7A,BHLHE40,CCNB1,CCND1,CCND2,CDK1,CFD,CYP17A1,CYP2B6,CYP4A11,DIO1,DTYMK,EHHADH,FAM89B,FOXO1,GADD45A,GHR,GSTP1,HPRT1,ID2,IL2RB,IRF8,LBR,MAD2L1,MET,NCL,NR1I3,PDK4,PRDM14,PRLR,SCP2,SMARCA4,STC2,TERT,TNNC1,TNNI1,TP53,VDR,XBP1 |
| VDR | -0,535 | transcription regulator | | 0,331 | 0,318 | -0,002 | 8,03E-06 | 1,10E-04 | ABCC3,ACACA,ACE,ACOX1,CALB1,CCND1,CD14,COL1A1,CPT2,CSHL1,CXCL10,CYP27B1,CYP2B6,CYP3A4,DOK1,F3,GADD45A,GPD1,ICAM1,IFI44L,IL18,KL,NGF,NOD2,NPHS1,NPPA,NR0B2,PDLIM2,PLD1,PLIN2,PNPLA2,PTPRF,SERPINC1,SLC10A2,SLC34A1,SLC34A2,SPP1,STAT4,TERT,TMPRSS2,TRPV5,UCP2,VDR |
| TGFA | | growth factor | | 1,639 | 1,675 | 0,007 | 8,04E-06 | 1,10E-04 | ALB,BIRC3,CASP1,CCL20,CCL5,CCND1,CXCL10,ESR1,FGF7,FOS,GJB1,GSTP1,ICAM1,IGFBP4,ITGAM,LCN2,MMP9,PECAM1,PLA2G4A,PRNP,PTGS1,PTTG1,RB1,S100A10,SLPI,TLR5,TP53,VEGFA,VIM |
| dinoprost | | chemical - endogenous mammalian | | 0,601 | 0,620 | 0,004 | 8,15E-06 | 1,11E-04 | ADM,ANGPTL3,ANXA4,CCND1,CCND2,CD36,ESR1,FOS,GSTP1,HMGCS2,HSD11B2,ID1,ID2,ID3,LIPA,MMP1,MMP9,NFIL3,NR4A1,PDIA4,PLA2G4A,POR,PRLR,PTGS1,SCP2,SPP1,VEGFA,YWHAE |
| PLG | -2,024 | peptidase | | 1,945 | 1,993 | 0,009 | 8,15E-06 | 1,11E-04 | APOH,BAX,CAV1,CD40,CLEC7A,CLIC4,CNN2,COL1A1,COLEC12,ELMO1,F3,FCGR2B,FYN,MIF,MMP1,MMP9,MRC1,MXI1,NOS2,PECAM1,PIK3CB,PLG,RAC1,RAC2,RHOA,SYK,TNFRSF11B,VAV1 |
| heparin | | chemical - endogenous mammalian | | -1,132 | -1,119 | 0,003 | 8,16E-06 | 1,11E-04 | ACADL,ACADM,ACADVL,APOE,CCND1,CD40,CD83,CD86,CDKN1B,COL1A1,DUSP1,F3,FN1,FOS,HPSE,ICAM1,IL1R1,MMP1,MMP2,MMP9,NOS2,PAPPA,PROCR,SELP,TNFAIP6,VEGFA,VIM |
| TFAM | | transcription regulator | Inhibited | -3,481 | -3,464 | 0,005 | 8,28E-06 | 1,12E-04 | ACADL,ACADM,ACADS,ACOX1,AUH,CPT2,ECH1,ECHS1,HK1,PFKP,PPARA,SLC25A20 |
| PSEN2 | 0,045 | peptidase | | -1,389 | -1,394 | -0,001 | 8,51E-06 | 1,15E-04 | ABCC6,ACSL1,ACTB,ANXA5,ATP6V0D1,BAX,C1QA,C1QB,C3AR1,CAP1,CASP3,CCS,CD74,CD9,COX8A,CTSS,CTSZ,CYTIP,ENPP2,FZD4,GJA1,GNB1,GPX5,HLA-E,INSR,KCNAB2,NDUFS8,PDGFRA,PRC1,PSENEN,PTPRF,STAB1,THY1,TP53 |
| MAPK9 | | kinase | | 0,472 | 0,500 | 0,004 | 8,62E-06 | 1,16E-04 | ACACA,AQP1,BAX,BIRC3,CACNB3,CAV1,CCL5,CDKN1B,CIDEC,EGF,FAS,FGL2,FOS,FOSL1,GADD45A,GADD45B,HIF1A,IFI16,IL15RA,IL18,IRF9,ISG20,JUND,LGALS3BP,LUC7L3,MARCKSL1,MIF,MMP9,NCL,NMI,NOS2,NR1H3,PDIA3,PLA2G4A,PLIN2,PPARA,PPP1R15A,PTEN,PTGES,RAC1,SCD,SEM1,SOD2,TBP,TLR3,TP53,VDR,ZFP36 |
| STAT5a/b | | group | | 0,696 | 0,735 | 0,008 | 9,00E-06 | 1,21E-04 | AHR,BCL2A1,BCL6,CCND1,CCND2,CD36,CD69,CDKN1B,CDKN2C,CISH,DUSP1,FCER1G,FCMR,FOS,GYPA,ID2,IDI1,IL13,IL4R,IRF1,LCP2,LEPR,MAF,MMD,MYD88,NETO2,NOX5,PCK1,RBP1,RFTN1,STK17B,TLR2,WDR3 |
| ITGAV | 0,782 | transmembrane receptor | | 1,768 | 1,789 | 0,006 | 9,05E-06 | 1,22E-04 | BAX,BIRC2,CD36,CDK1,CDKN1B,COL1A1,COL1A2,FN1,FOS,ITGAV,ITGB3,MMP1,MMP2,MMP9,PCNA,TIMP1,TP53 |
| ZEB2 | 0,287 | transcription regulator | | 1,502 | 1,485 | -0,004 | 9,20E-06 | 1,24E-04 | ALPL,AXL,CCND1,CX3CR1,ESRP2,FCER1A,FCN1,GAB2,GATM,GZMA,ID2,IL7R,INPP5D,KRT19,MYB,NKG7,PTEN,SELL,SFRP1,TERT,VCAM1,VIM |
| miR-125b-5p (and other miRNAs w/seed CCCUGAG) | | mature microRNA | Inhibited | -2,140 | -2,195 | -0,010 | 9,46E-06 | 1,27E-04 | ADAMTS1,ALOX5,CASP6,CBFB,CCNA2,CCR5,CD44,CDH5,DICER1,DUS1L,ENTPD4,ERBB3,GSS,ID1,ID2,ID3,IL6R,KRT19,LIPA,MYD88,NOS2,PCTP,PERP,PIGR,PPT2,PROM1,SCD,SGPL1,TP53,TSPAN8,UBE2I,VPS4B |
| IL33 | 0,472 | cytokine | Activated | 2,597 | 2,648 | 0,007 | 9,53E-06 | 1,28E-04 | ABCG1,ACAT1,ACTB,ADAMTS1,ALDH1A3,ALDH3A1,APOE,AXL,BCL2A1,BIRC3,CCL18,CCL5,CCND1,CCR2,CD36,CD40,CD69,CD86,CISH,COL3A1,CXCL10,EHD1,ELMO1,EPHX2,FAS,G0S2,GADD45B,GCHFR,GOSR2,GRK2,HCK,HIF1A,ICAM1,IL13,IL15,IL2RG,ITGAM,KLC1,LAMA5,LEPR,MDK,NCF4,NFKB1,NFKBIE,NOS2,NPHS2,PEA15,PILRA,PLIN2,PTPRK,RASGRP1,S100A8,SOD2,ST3GAL5,TFEC,TIMP1,TLR2,TNFAIP2,TNFRSF1B,TNFSF10,TPBG,TUBB6,VASH1,VEGFC |

| | | | | | | | | | |
|---|---|---|---|---|---|---|---|---|---|
| EZH2 | 0,364 | transcription regulator | | 1,157 | 1,158 | 0,000 | 9,60E-06 | 1,28E-04 | ACKR3,ACTB,ADAM10,ADRB2,ALOX5AP,ALPL,ATF3,BIRC3,BMP7,C3,CCL5,CCND1,CCND2,CCND3,CCR5,CD63,CDKN1B,CDX2,COL4A1,CPD,CSNK1A1,CUL1,CXCL1,CXCL10,CXCL11,CYP1B1,DNMT1,EVX1,EZH2,FBN1,FKBP5,FRZB,GSTM4,HCP5,HOXA1,HOXA10,HOXB5,HOXB6,HSD11B2,ICAM1,IL10RA,IL13,IL36A,JAM2,KRAS,KRT81,LCN2,LTB,MMP2,MMP7,MMP9,MNDA,MPZL2,MYL6,NDRG1,NME1,NOTCH2,PARP12,PDGFRA,PER1,PIK3IP1,PLA2G4A,PLAAT4,PLEKHA6,PROM1,PSMD1,PTEN,PTGES,RAC3,RNF19B,S100A4,S100A8,S100A9,SCP2,SFN,SGPL1,SLC1A3,SLC22A18AS,STT3A,TIMP2,TMPRSS2,TP53,TRIM38,TSPAN7,UBD,VEGFA,WIF1,WNT10B,WTAP |
| H1-6 | | other | Activated | 2,523 | 2,556 | 0,006 | 1,03E-05 | 1,37E-04 | ARPC5,ATP6V0E2,B2M,B4GALT5,BHMT,CANT1,CD36,CD81,DAO,DENND2D,FARP1,FKBP4,FOS,GLUL,GPHN,HIRIP3,LYRM9,MAN2B1,MCM6,PCCB,PPT2,PRKCQ,RRM1,SERINC3,SMARCE1,TEX2,THAP1,TMEM246,TMEM53,ZC3H4 |
| NPC1 | 0,294 | transporter | Inhibited | -3,424 | -3,443 | -0,005 | 1,03E-05 | 1,37E-04 | ABCG1,ALPL,CAV1,CAV2,CCND1,CCR2,CCR5,CD74,CD8A,CTSB,CTSD,CTSK,CYP27A1,CYP27B1,DDIT3,HLA-DQB1,IDI1,LIPA,LYZ,MCM2,MYLIP,PNPLA3,S100A8,S100A9,SCD,STAT1,STAT3,STAT4,TLR4,XBP1 |
| MAFB | 0,772 | transcription regulator | | 0,604 | 0,636 | 0,006 | 1,03E-05 | 1,37E-04 | ACTB,ACTN1,ADA,CACNA2D3,CAP1,CD163,CD2AP,CD38,CNN2,CXCL12,CXCL9,FPR1,GATM,HS3ST1,HTR2B,IL7R,MAFB,MARCKS,MMP9,NPHS1,NPHS2,NUAK1,OLFML2B,PCK1,RNASE1,SLC2A2,SLC7A8,STAB1,THG1L,TP53 |
| H1f1 | | other | Activated | 2,523 | 2,556 | 0,006 | 1,03E-05 | 1,37E-04 | ARPC5,ATP6V0E2,B2M,B4GALT5,BHMT,CANT1,CD36,CD81,DAO,DENND2D,FARP1,FKBP4,FOS,GLUL,GPHN,HIRIP3,LYRM9,MAN2B1,MCM6,PCCB,PPT2,PRKCQ,RRM1,SERINC3,SMARCE1,TEX2,THAP1,TMEM246,TMEM53,ZC3H4 |
| SIN3A | | transcription regulator | | 0,024 | 0,000 | -0,008 | 1,04E-05 | 1,38E-04 | ALDH6A1,ARID4B,BCL6,CASP3,CCND2,COL1A1,COL1A2,CTSK,DDIT3,ESR1,GADD45B,GSTP1,HLA-DRA,MMP9,MXI1,NCL,PMM1,S100A9,SET,SMAD7,TERT,TGFBR2,TP53BP1,TXNIP |
| KLF11 | 0,124 | transcription regulator | | -0,563 | -0,550 | 0,003 | 1,06E-05 | 1,40E-04 | ACSL5,ADAMTS1,BMP7,CAT,CAV1,CCR2,CHRND,COL1A2,COL3A1,CPT2,CXCR4,DCN,EGF,ENG,IL13,ITGA2,ITGA3,ITGAV,ITGB3,ITGB6,ITGB8,MAOB,MMP1,MMP2,MMP9,NFKB1,PDK4,PLA2G4A,PLG,PPARGC1A,SLC25A20,SMAD2,SMAD7,SOD2,STAT1,TGFBR2,THBS2,TIMP1,TIMP2,VEGFA |
| NR0B2 | -0,568 | ligand-dependent nuclear receptor | Activated | 4,153 | 4,118 | -0,008 | 1,08E-05 | 1,43E-04 | ACACA,ACADM,ACOX1,APOA1,APOM,BAAT,CD36,CEL,ESRRG,FBP1,G6PC,GPT,HNF1A,HNF4A,LPIN1,MTTP,NR0B2,P4HB,PCK1,PCK2,PDIA3,PDIA4,PDK4,PPARGC1A,SCD,SLC10A2,SPP1,TIMP1 |
| GH1 | | growth factor | | -1,391 | -1,381 | 0,002 | 1,10E-05 | 1,44E-04 | ABCC1,ATF3,CCNB1,CCND3,CISH,CLU,CSHL1,CXCR4,CYP3A4,DDIT3,DECR1,ERBB3,FN1,FOS,GADD45A,GHR,HDAC1,HOXA1,ID2,ID3,IL4R,MYB,NOS2,PRLR,PTEN,RHOA,RYK,SHMT2,SMARCA4,SOCS2,SOD1,SPP1,TKT,UBA7,VEZF1,VIM,ZFP36 |
| IGF2 | | growth factor | | 0,650 | 0,666 | 0,004 | 1,11E-05 | 1,45E-04 | ALAD,CASP6,CCND1,CD36,CDK1,CKM,CTSV,CXCR4,CYP1B1,DDIT3,DHCR7,DUSP1,ELOVL6,FN1,GADD45A,GCH1,GHR,GNAI2,GNB1,GRN,HIF1A,IDH1,IFITM2,IGFBP4,IGFBP6,KLF9,MCM2,MCM5,MCM7,MMP1,MXI1,NDRG1,NGF,NOS2,PDCD4,PMS1,PRIM1,PRKCQ,PTEN,RHOB,RRM1,RRM2,SEZ6L2,SKAP2,SPP1,TGFBI,THBS2,TP53,VEGFA |
| Ap1 | | complex | | -0,473 | -0,430 | 0,009 | 1,15E-05 | 1,50E-04 | ACE,ATF3,BCL2A1,C3AR1,CALM1 (includes others),CCL5,CCNA2,CCND1,CD40,CD44,CDX2,CLU,CXCL12,DDIT3,DNMT1,F2RL1,F3,FN1,FOS,FOSL1,GSS,GSTP1,GZMB,HBB,HPGD,HSP90B1,IFRD1,IL13,ITGA2,MET,MMP1,MMP2,MMP7,MMP9,NDRG1,NGF,NOS2,PCK1,S100A4,SNCG,SPARC,TIMP1,TLR4,UTRN,VCAM1,VDR,VEGFA |
| ABL1 | 0,131 | kinase | | -0,597 | -0,568 | 0,007 | 1,15E-05 | 1,50E-04 | C3,CAV1,CCNA2,CCND1,CCND2,CDKN1B,CGA,CTSB,DOK1,FAS,GADD45A,MMP1,NME1,NOX5,PAK2,SST,TP53,VEGFA |
| RORA | -0,589 | ligand-dependent nuclear receptor | Inhibited | -2,175 | -2,151 | 0,006 | 1,24E-05 | 1,61E-04 | ACACB,ACTB,ADIPOR1,APOA4,APOC3,APOE,ASNS,BHMT,CCL5,CCNB1,CD36,CISH,CXCL10,CYP27A1,CYP2A6 (includes others),CYP2B6,CYP2C8,CYP46A1,CYP4A11,DDC,ELOVL6,G6PC,GADD45B,GOT1,GSTM4,GSTP1,HPGD,HSD17B10,IL18,KCNK5,LCN2,NAT8,NNMT,NOCT,NR1I3,NSDHL,NTRK2,PCK1,PLIN2,POR,PPARGC1A,PPP1R3C,RORA,RORC,S100A8,S100A9,SCD,SELENBP1,SLC13A2,SLC16A7,SLC25A24,TLR3,UCP2,VLDLR |
| IGBP1 | -0,209 | phosphatase | | 1,594 | 1,600 | 0,002 | 1,25E-05 | 1,62E-04 | CASP1,CHUK,CLDN1,CLDN3,IRF1,NOS2,RAC1,TJP1 |
| corticosterone | | chemical - endogenous mammalian | | -0,882 | -0,880 | 0,000 | 1,25E-05 | 1,62E-04 | ACACA,ACOX1,ANXA1,ARC,AVP,BACE1,CALB1,CAV1,CCND1,CDKN1B,CGA,CSHL1,CYBA,DDIT3,FKBP5,FOS,GLUL,GZMB,HSPA5,KLF9,LHB,MAOA,MAPK8,MMP9,MTOR,NOS2,NOX4,NPPA,PDK4,PNPLA2,PPARGC1A,SFRP1,SGK1,SLC12A3,TLR2,UCP2,WNT10B,XBP1 |
| EDNRA | | transmembrane receptor | Activated | 2,970 | 3,000 | 0,010 | 1,29E-05 | 1,67E-04 | COL1A1,COL3A1,FOS,ICAM1,ITGAM,MMP2,PTGS1,SELL,TIMP1,TIMP2 |

| | | | | | | | | | |
|---|---|---|---|---|---|---|---|---|---|
| CAT | -0,632 | enzyme | | -1,022 | -1,046 | -0,005 | 1,31E-05 | 1,68E-04 | ADM,BAX,BCL2A1,CAV1,CD36,CD69,CDKN1B,COL1A1,CYP1B1,FN1,GCLC,HIF1A,HLA-DRA,MMP1,MMP9,NOS2,TIMP1,TNC,TP53,VCAM1,VIM,XAF1 |
| NFATC1 | | transcription regulator | | 0,226 | 0,261 | 0,008 | 1,36E-05 | 1,75E-04 | ACP5,AHR,BMP7,CCND1,CCND2,CD3G,CD40,CDKN1B,COL1A1,CTSK,CX3CR1,CYBB,CYP1B1,FAS,HNF1A,HNF1B,HNF4A,ICAM1,IL33,ITGAV,ITGB3,NOX4,PLS3,POU2AF1,PRLR,PTEN,RCAN1,RORC,SELL,SLC2A2,SPP1,TNFSF10,UTRN,VCAM1 |
| HDAC4 | | transcription regulator | | -0,281 | -0,293 | -0,004 | 1,42E-05 | 1,81E-04 | ARC,ATF3,CACNA2D1,CALB1,CAMK2A,CAP2,CCNB1,CHN1,CKM,CYFIP2,ETNPPL,FGF13,FOS,FOSL1,G6PC,GADD45A,GPHN,HDAC1,HIF1A,HLA-DRA,KLHL2,MAPK10,MAPK8,MEF2C,MMP9,MYOM2,NBEA,NOS2,NR4A1,PRKCA,RGS7,SH3GL2,TERT,TP53BP1,TRPV1,TSPAN13,TSPAN7,VCAM1,VEGFA |
| NFKB2 | | transcription regulator | Activated | 2,399 | 2,430 | 0,007 | 1,43E-05 | 1,82E-04 | AKR1B1,ANXA5,BIRC3,CCL19,CCL20,CCL5,CCND1,CCR7,CD83,CD86,CTSS,CXCL12,CXCR4,FOS,GPR183,HLA-DMB,HLA-DOA,HLA-DQA1,MARCKSL1,MAT2A,MMP2,MMP9,MYB,PTEN,SELL,SOD2,TNFAIP2,VCAM1 |
| TICAM1 | -0,022 | other | Activated | 4,907 | 4,965 | 0,009 | 1,44E-05 | 1,83E-04 | ACSL1,APOBEC3B,ATP6V1E1,CASP4,CCL5,CD38,CD40,CD86,CFB,CXCL10,CXCL11,CXCL6,DDX58,DUSP1,FAS,FPR1,ICAM1,IFI16,IFIT2,IFIT3,IL15,IL15RA,IL18,IRF1,IRF7,ISG15,ISG20,LPAR1,MAFF,MCM2,MET,NFKB1,PILRA,PSTPIP2,RHOB,RSAD2,SAMHD1,TFEC,TLR2,TLR3,TNFSF10,TSC22D1,VCAM1 |
| BCR (complex) | | complex | Activated | 3,273 | 3,314 | 0,007 | 1,44E-05 | 1,83E-04 | BCL2A1,BCL6,CALR,CASP3,CCNA2,CCND1,CCND2,CCND3,CCR7,CD1C,CD27,CD300A,CD44,CD69,CD86,CDKN1B,CTCF,CXCR4,FAS,FCMR,FOS,FOSB,FOXO1,GBP2,GZMB,HLA-DMA,HSP90B1,HSPA5,IL13,IL2RB,JAK1,KLF9,LAIR1,LCK,MS4A6A,NAMPT,POU2AF1,S100A10,S1PR1,SELL,XBP1 |
| Collagen type I (complex) | | complex | Activated | 2,609 | 2,633 | 0,005 | 1,46E-05 | 1,85E-04 | APOE,BAX,CCL5,CCND1,CD83,CDX2,COL1A1,F11,FN1,ITGA2,ITGA3,ITGAV,MMP1,MMP2,MMP9,PLOD2,PRRX1,PTEN,RAC1,RECK,S100A10,TIMP2,TNC,TP53,VIM |
| MED13 | | transcription regulator | | 0,092 | 0,077 | -0,003 | 1,56E-05 | 1,97E-04 | ACACA,ACSM3,CD36,CIB2,CIDEC,CYP2A6 (includes others),ELOVL6,ENO1,G6PC,GPD1,GYS1,KITLG,KNG1,NTRK2,PGAM2,PKLR,PYGL,SCD,SERPINA5,TKT |
| superoxide | | chemical - endogenous mammalian | Activated | 2,933 | 2,950 | 0,006 | 1,57E-05 | 1,97E-04 | CAV1,CLU,CYBA,FN1,ICAM1,ID3,MARCKSL1,MMP9,SOD2,TIMP1,TLR8 |
| SAFB2 | -0,232 | other | Inhibited | -2,481 | -2,496 | -0,004 | 1,60E-05 | 2,01E-04 | B2M,BCCIP,FOS,HLA-C,HLA-DPB1,HLA-E,MLLT3,PPP1R12A,PSMB9,RPL35A,SAFB2,UBE2B,UQCRC2 |
| DEPTOR | 0,163 | other | Inhibited | -2,620 | -2,646 | -0,010 | 1,65E-05 | 2,06E-04 | CCL20,CCL5,CXCL10,CXCL11,CXCL9,ICAM1,VCAM1 |
| OSCAR | | other | | -0,018 | 0,000 | 0,005 | 1,75E-05 | 2,19E-04 | CCL20,CCR2,CD40,CD83,CD86,CX3CR1,CXCL1,FZD2,ICAM1,IL1R1,ITGAM,MMP7,RRAS,TLR7,TLR8,TNFSF10 |
| CXCR4 | 1,330 | G-protein coupled receptor | | 1,890 | 1,919 | 0,007 | 1,91E-05 | 2,38E-04 | ACKR3,AXL,CCND1,CD44,CD69,CD9,CDH11,CXCL1,CXCL10,CXCL12,CXCR4,DPP4,F3,GAD1,ID1,ID2,IGFBP7,MMP2,RRAS,TNFRSF10D,TNFSF10,TNNC1,TP53 |
| MUC1 | 0,773 | other | | 0,540 | 0,568 | 0,006 | 1,91E-05 | 2,38E-04 | ABCC1,CAT,CCND1,CCND2,CD1D,CD40,CD83,CD86,CDH11,CDKN1B,GCLC,HIF1A,HLA-DRA,IFIT3,IFITM1,MRC1,MT1G,MUC1,SOD1,SOD2,STAT1,TERT,VIM |
| MAP2K6 | -0,107 | kinase | | 0,873 | 0,899 | 0,007 | 1,92E-05 | 2,38E-04 | ACP5,BAX,CCL8,CCND1,CD40,CD83,CD86,CKM,CXCL10,CXCL9,ESR1,IFIT2,IHH,IRF9,ISG15,MMP1,MMP2,MMP9,NOS2,NPPA,PPARGC1A,PTH1R,RGS7,SLC1A3,STAT1,TNFSF10,VDR |
| RXRB | | ligand-dependent nuclear receptor | | 0,932 | 0,928 | -0,002 | 1,95E-05 | 2,42E-04 | ACADM,ACOX1,ACSL1,ATP1A3,C1QA,CCL20,COLEC12,CSHL1,CTSB,CTSS,CXCL10,FABP1,FLRT3,FSTL1,IGFBP6,IL13,LCN2,LYZ,MMP2,PMP22,S100A4,S100A8,SEMA4D,SPP1,TERT,VCAN |
| TNFSF13B | | cytokine | | 1,411 | 1,435 | 0,006 | 1,96E-05 | 2,42E-04 | BAX,BCL2A1,BCL6,CCND2,CCR7,CD40,CD83,CD86,CTSS,CXCL6,FCGR2B,FOXO1,GPR183,HLA-DMB,HLA-DOA,HLA-DQA1,ICAM1,IL15,MMP9,NAMPT,POU2AF1,SELL,SPP1,TP53 |
| MRTFB | 0,221 | transcription regulator | | 0,265 | 0,243 | -0,006 | 2,32E-05 | 2,85E-04 | ACTB,CBFB,CCL5,CD151,CEBPE,CLDN15,CNN2,CPA3,CXCL6,CXCR4,CYBA,EYA2,FCN1,FLI1,FLNA,G0S2,HLA-A,LCN2,LTF,MAPK13,MGST2,MYH9,S100A8,S100A9,SELP,SERINC3,SLPI,TBXAS1,TST |
| TREM1 | 0,030 | transmembrane receptor | | -1,289 | -1,252 | 0,005 | 2,32E-05 | 2,85E-04 | ACKR3,ADRB2,ASNS,ATF3,CCL18,CCL20,CCL5,CCR7,CD14,CD83,CD86,CDK1,CENPU,CFB,CXCL1,CXCL10,CXCR4,DTL,F3,FERMT2,FOSL1,GADD45B,GCLM,IDO1,IFIT2,IL15RA,IL1R1,IL6R,IRF1,ISG15,KCNJ2,LAMP3,MAFF,MFN1,MMP1,MT1F,MYD88,NEDD4L,NOD2,OSGIN1,PLAC8,PLD1,PLEKHF2,PTTG1IP,RCAN1,RIN2,SIGIRR,SLC1A3,SPP1,SPRY2,TFPI2,TLR2,TLR4,YRDC |
| E2f | | group | Activated | 2,737 | 2,766 | 0,008 | 2,33E-05 | 2,85E-04 | ALG5,CAV1,CCNA2,CCNB1,CDC7,CDK1,CYP27B1,DHPS,DLEU1,DROSHA,EFHC1,EXOSC8,GINS2,HDAC1,HIF1A,MAD2L1,MCM2,MCM3,MCM5,MCM6,MFAP1,MTHFD1,MYB,NASP,NPPA,OIP5,PCNA,PCSK1N,PGRMC1,RECQL,RPA3,RPS16,RRM2,TP53,TPX2,TTK,UBE2C,YBX3 |

| | | | | | | | | | |
|---|---|---|---|---|---|---|---|---|---|
| PITX2 | | transcription regulator | Inhibited | -3,420 | -3,387 | 0,006 | 2,33E-05 | 2,85E-04 | ACAD8,CCND1,CCND2,CCND3,COL1A2,COQ6,FOXO1,FZD2,FZD4,GAD1,GADD45A,GCDH,GPX7,GRHPR,HMX1,HPGD,ITGAV,ME3,MGA,MSRA,NDUFB7,NDUFC1,NDUFS2,NDUFS3,NDUFS7,NDUFS8,NPPA,NXN,OGFOD2,PLOD1,PLOD2,PRDX2,SOD1,SOD2,TBX2,TXN2,VIM,ZEB2 |
| SRF | | transcription regulator | | -1,310 | -1,287 | 0,004 | 2,49E-05 | 3,03E-04 | A2M,ACTB,ACTG1,AKAP12,ANAPC15,ANKRD6,APOLD1,ARC,BRD3,CALB1,CAP1,CAPRIN1,CCL5,CDH9,CEBPE,CKM,CLDN1,CLDN15,CLDN3,CNN2,CPA3,CXCR4,DSTN,EIF6,EYA2,FCN1,FERMT2,FGF1,FHL1,FHL2,FLNA,FOS,FOSB,FOSL1,G0S2,GADD45A,GADD45G,GCH1,GLIPR1,IGHM,IL15,ITGAM,LCN2,LEPR,LTF,MAP4K4,MAPK10,MAPK13,MCF2L,MGST2,MMP9,MRPS35,MTMR6,MYB,MYH9,MYL3,MYO1B,NET1,NKX2-5,NPPA,NR4A1,PDCD10,PGM1,PLA2R1,PTMS,RAI2,RND3,S100A8,S100A9,SELP,SLC37A1,SLPI,SMTN,SND1,TBP,TBXAS1,TNC,TNNC1,TNNT2,TST,TUBB4B,VCAN,WDR1,WT1 |
| THBS4 | -0,295 | other | Activated | 2,552 | 2,567 | 0,004 | 2,49E-05 | 3,03E-04 | CALR,CD40,CHUK,COL3A1,CRELD2,CXCL10,HSPA5,ICAM1,MANF,MTHFD2,P4HB,SDF2L1,TNC,VCAM1 |
| LONP1 | -0,276 | peptidase | | -0,453 | -0,464 | -0,003 | 2,52E-05 | 3,04E-04 | ACADS,ACAT1,ANXA2,ASNS,ATP5F1D,C1QBP,CALU,CLPX,COX8A,DLD,ECI1,ECSIT,ETFB,GARS1,GLUD1,HSPA9,MDH2,MRPS22,MTHFD2,P4HB,PDIA3,PFN1,S100A11,SOD1,SOD2,TRAP1,TUFM |
| CCL5 | 1,439 | cytokine | | 0,488 | 0,532 | 0,009 | 2,52E-05 | 3,04E-04 | ADGRE5,AHR,CAPN2,CCL5,CCR5,CD163,CD40,CD44,CYP1B1,DDAH1,DUSP1,EGF,F2RL1,FOS,ITGAM,MMP9,NAMPT,NOS2,PNP,SGK1,SOD1,STAT1,STAT3,TLR4,TPMT,TUBB4B,ZFP36 |
| FGFR2 | 0,151 | kinase | | -0,758 | -0,743 | 0,003 | 2,59E-05 | 3,11E-04 | AFP,BAX,CA2,CA4,CCND1,CCND2,CDH11,COL1A1,COL3A1,COL4A1,COL4A2,CPE,CUBN,DAB2,DRAM1,ENPEP,FAS,FLRT3,FN1,GASK1B,GJA1,GLIPR1,GRB10,ICAM1,ID2,IHH,LAMB1,LGALS2,MRAS,MYO1F,PCOLCE,PDGFRA,PTH1R,SFRP1,SPP1,TFEC,TSPAN8,VCAM1,VEGFC |
| HDAC5 | -0,110 | transcription regulator | | -1,366 | -1,366 | 0,000 | 2,61E-05 | 3,14E-04 | ACADL,ACADM,ACSL1,ARC,ATP5F1D,CASP8,CD27,CKM,CPT2,ETNPPL,G6PC,HLA-DRA,HMGCL,HMGCS2,MEF2C,NOS2,NPPA,NR4A1,PFKL,PPARA,PPARGC1A,PTEN,SCD |
| mir-133 | | microRNA | | -1,898 | -1,938 | -0,008 | 2,63E-05 | 3,15E-04 | AK2,ARFIP2,BCL2L2,CCNB1,CCND2,CERS2,CMTM6,CORO1C,CTBP2,DNMT1,FN1,FSCN1,FTL,GSTP1,HLA-G,KLF15,LPGAT1,MET,MSN,PITX3,RHOA,TNFSF10,TNNT2,UBXN7,VCAN |
| RABL6 | | other | Activated | 3,783 | 3,808 | 0,005 | 2,64E-05 | 3,16E-04 | ABAT,CCNA2,CCNB1,DRAM1,ELF4,EZH2,FERMT2,MAD2L1,MCM2,MCM5,MCM7,MELK,NDC80,PBK,POLE2,PRC1,PRIM1,PTPRM,TOP2A,TP53,TPX2,TTK,UBE2C,VRK1 |
| ONECUT1 | | transcription regulator | | 0,134 | 0,139 | 0,002 | 2,66E-05 | 3,18E-04 | AASS,ABCC2,ACACB,ACSS3,ADH1B,AFP,ALDH3A1,ALDH5A1,AMT,APOH,ARHGAP32,BCKDHA,BNIP3L,C1S,C2,CD36,COA4,COPS7A,CRADD,CXCL1,DDB2,EED,EHF,F11,FAM117A,FAS,G6PC,GJB1,GSS,GTF2E1,HABP2,HLA-G,HMGB2,HNF1B,HNF4A,HNRNPR,HTR2B,ITGAV,LUC7L3,MFSD1,MYOF,NDUFAF4,NR0B2,OAT,PCK1,PFKL,PHTF1,PPFIBP1,PTPRN2,RARG,RECK,RMND1,RORA,RORC,SAMHD1,SCD,SH3BGRL,SLC1A3,SLC25A13,SLC2A2,SNRPD2,SNW1,SPAST,SUPT16H,TAOK3,TBX2,TDO2,TMBIM6,TNFSF10,UGT2A3,VWA5A |
| MET | 0,431 | kinase | Activated | 2,820 | 2,854 | 0,006 | 2,67E-05 | 3,19E-04 | AQP3,AURKA,CCND1,CCND2,CD44,CDK1,CDKN1B,CTSV,CXCL1,DAB2,FGL2,FN1,FOS,GCLC,IL15,IL15RA,IL18,ITGA3,MAD2L1,MMP2,MUC1,NOX4,NPHS1,PLK2,SLA,SLCO3A1,SOD1,SOD2,SPP1,TLR3,TRIM21,TSC22D1,VEGFA,VIM |
| PROC | -0,547 | peptidase | | -0,784 | -0,796 | -0,003 | 2,94E-05 | 3,51E-04 | ACE,ACE2,BCL2A1,BIRC2,CALR,CLU,ICAM1,MMP2,MMP9,MOK,NOS2,NR4A1,PCNA,PROCR,TJP1,TLR2,TLR4,TP53,VCAM1 |
| homocysteine | | chemical - endogenous mammalian | | 1,449 | 1,472 | 0,005 | 2,98E-05 | 3,55E-04 | ANXA2,APOA1,ATF3,BAX,BHMT,CCL5,CCND1,CLU,CXCL10,CXCL12,DDIT3,F3,FGF1,GADD45A,HSPA5,ID1,IDI1,ITGAM,MMP2,NDRG1,NPHS2,PKM,PRDX2,TIMP1,TP53,TRIB3,VCAM1,VEGFA,XBP1 |
| NUPR1 | 0,294 | transcription regulator | Inhibited | -2,177 | -2,209 | -0,003 | 3,01E-05 | 3,57E-04 | ABCC5,ADCY9,ADGRG1,ADM,ADRB2,AGRN,AKAP12,ALDH5A1,APOBEC3B,ASB9,ATF3,ATR,AURKA,AVPI1,BNIP3,CAMK2N1,CARD8,CARMIL1,CCNA2,CCNB2,CD63,CDKN1B,CHUK,COL1A2,COL3A1,CREB5,CSTF2T,CXADR,CXCR4,DDIT3,DENND5B,DHTKD1,ELL2,ERMP1,ESPL1,FAM111A,FCHSD2,FGF1,FHL2,GAB2,GADD45A,GBP2,GCH1,GCNT2,GINS1,GK,GPSM2,HNRNPM,HOXB5,IFIT2,IL13RA1,IL6R,ING2,ISCA1,ITPR3,KANK2,KAT5,KIF20A,LBR,LHFPL2,LMNB1,MAT2A,MMD,MSH6,MT1X,MX2,MYD88,NDRG1,NFIL3,NSF,P3H1,P4HA2,PARP1,PCTP,PCYOX1,PEA15,PFKFB3,PHTF1,PMP22,PMPCA,POLE2,PPP1R15A,PPWD1,PRNP,RAB11FIP5,RAB17,RAB29,RAB32,RAB38,RMI1,RNF19B,RPA1,SAMHD1,SHPK,SLC25A20,SPAG5,STK38,SUOX,TCTN2,TERT,TESK1,THG1L,TMEM268,TOB1,TOLLIP,TP53,TP53BP1,TP53BP2,TRIB3,UAP1,WDCP,WNK1,XBP1,ZDHHC13 |

| | | | | | | | | | |
|---|---|---|---|---|---|---|---|---|---|
| MRTFA | | transcription regulator | | 1,053 | 1,046 | -0,002 | 3,10E-05 | 3,67E-04 | ACTB,ARPC1B,CBFB,CCL5,CD151,CEBPE,CLDN15,CNN2,COMT,CPA3,CXCL12,CXCL6,CXCR4,CYBA,CYFIP2,EYA2,FCN1,FLI1,FLNA,FOS,G0S2,HLA-A,ICAM1,LCN2,LTF,MAPK13,MGST2,MMP9,MYH9,NPPA,PFN1,RAC1,S100A8,S100A9,SELP,SERINC3,SLPI,STAT1,TBXAS1,TNC,TST |
| Pdgf (complex) | | complex | | 0,911 | 0,941 | 0,006 | 3,12E-05 | 3,68E-04 | BAX,CAV1,CCN3,CCND1,CCND2,CD44,CDKN1B,COL4A1,CYBA,F3,FN1,FOS,FOSB,FOXO1,GJA1,HIF1A,ICAM1,IL13,ITGB3,MCM6,MCM7,MMP1,MMP2,NOX4,PCNA,PIK3R3,SPARC,TERT,TIMP1,TP53,VCAM1,VEGFA |
| AMPK | | complex | Inhibited | -2,667 | -2,662 | 0,001 | 3,13E-05 | 3,69E-04 | ACACA,ACE,ACO2,BAX,CAT,CCNB1,CCND2,CLDN1,CXCL10,CXCL12,CYBB,CYP2B6,CYP3A4,CYP4F2,FSCN1,G6PC,GJA1,GSTK1,HIF1A,LHB,LITAF,LPIN1,MMP9,NOS2,NOX4,NR0B2,OAT,PCK1,PFKFB3,PKLR,PPARGC1A,SOD1,STAT1,TP53,UCP2,VCAM1,VIM |
| PNPT1 | | enzyme | Inhibited | -3,585 | -3,619 | -0,008 | 3,20E-05 | 3,77E-04 | AGRN,CCL5,CDKN1B,CXCL10,DDX58,GBP2,IFI16,IFI44,IFIH1,IFIT3,IRF7,ISG15,LGALS3BP,OAS1,PARP12,RTP4,STAT1,TOR3A,UBE2L6,XAF1 |
| fatty acid | | chemical - endogenous mammalian | | -1,339 | -1,300 | 0,007 | 3,32E-05 | 3,90E-04 | ACACA,ACADL,ACOX1,AHR,AOC1,APOA1,APOC3,BAX,CD36,CDKN1B,DDIT3,DGAT1,G6PC,HSPA5,ITGAM,LEPR,LHB,MACROH2A1,MMP9,NOS2,NR1H3,NR4A1,NR4A3,PLIN2,PPARA,PPARGC1A,PTEN,RXRA,S100A8,SCD,SLC2A2,ST3GAL5,UCP2,VCAM1 |
| NEDD9 | -0,451 | other | | 0,858 | 0,894 | 0,009 | 3,34E-05 | 3,92E-04 | ADM,ALOX5,AOAH,BHLHE40,BNIP3,FABP1,FOS,ITGA2,MMP2,MMP9,PCK1,PFKP,PLAC8,PLIN2,PLOD2,TXNIP,VIM |
| RORC | -0,757 | ligand-dependent nuclear receptor | | 1,444 | 1,463 | 0,007 | 3,43E-05 | 4,01E-04 | ACACB,APOA4,ASNS,BHMT,CCL20,CCL5,CCNB1,CD36,CISH,CYP2A6 (includes others),CYP2B6,CYP2C8,CYP4A11,DDC,ELOVL6,G6PC,GADD45B,GSTM4,GSTP1,HIF1A,HPGD,IL1R1,KCNK5,KLRB1,LCN2,NAT8,NNMT,NOCT,NR1I3,NSDHL,PCK1,POR,PPP1R3C,PROCR,RORA,RORC,S100A8,S100A9,SELENBP1,SLC13A2,SLC16A7,SLC25A24,SPARC,VEGFA,VLDLR |
| PDGFB | | growth factor | | 1,157 | 1,178 | 0,005 | 3,54E-05 | 4,11E-04 | CCN3,COL1A1,COL1A2,FGF1,FGF7,FN1,FOS,FOXO1,FOXO4,ICAM1,MMP2,MMP9,MYL6,NOTCH2,NR4A1,PTEN,SERPINF1,SMAD1,SP1,STAT3,TIMP1,TNC,VEGFA |
| miR-21-5p (and other miRNAs w/seed AGCUUAU) | | mature microRNA | Inhibited | -3,054 | -3,101 | -0,010 | 3,54E-05 | 4,11E-04 | ARF4,CD69,CDH5,FAS,HIPK3,IL6R,IRAK1,LRRFIP1,MARCKS,MMP2,MSH2,MYD88,PDCD4,PECAM1,PTEN,RECK,SMAD7,SPRY1,SPRY2,STAT3,TGFBR2,TNFRSF11B,TPM1 |
| phosphate | | chemical - endogenous mammalian | | -0,475 | -0,459 | 0,003 | 3,58E-05 | 4,14E-04 | A2M,ACADSB,ACAT1,ALPL,ANKH,AXL,BLVRA,CHST12,COL5A1,CPT2,CYC1,DCN,DECR2,ECSIT,FOSL1,JAK1,MAP2K3,PCCA,PCNA,PLA2G12A,POSTN,SDF2L1,SLC25A5,SLC34A1,SLC34A2,SPP1,STC2,TIMP1,UQCR11,ZFP30 |
| PKD1 | | ion channel | Inhibited | -3,710 | -3,724 | -0,002 | 3,65E-05 | 4,23E-04 | ABCG2,ACOX2,ADCY7,ALDH8A1,ALG6,APOM,APOO,ATP1B2,AXL,BHMT2,CAVIN1,CPN2,DDIT3,DPEP1,FGFR3,FOSB,FOXO1,G6PC,GATM,GLS,GPRASP1,HGD,IDH1,KNG1,MAP3K12,MCUB,MID1,NEDD9,NNMT,PAH,PCK1,PRLR,PRODH2,PVALB,QPCT,RHOA,SEMA4C,SERPINF2,SFXN3,SLC13A2,SLC16A4,SLC2A2,SLC2A9,SLC4A1,SSTR2,ST5,THG1L,TNFAIP8,TNFRSF11B,TNS1,TREH,TRIM15,VWF,ZBTB10 |
| chenodeoxycholic acid | | chemical - endogenous mammalian | | -0,009 | 0,006 | 0,003 | 3,81E-05 | 4,40E-04 | ABCC2,ABCC3,ABCG1,ADH1B,APOA1,APOE,APOM,CAV1,CD36,COL1A1,CYP27A1,CYP3A4,FBP1,G6PC,HNF1A,HNF4A,HPGD,KNG1,MMP9,MTTP,NR0B2,PCK1,PCK2,PDK4,PLTP,PPARA,SDC1,SLC10A2,VDR |
| ACKR2 | -0,064 | G-protein coupled receptor | Inhibited | -4,168 | -4,206 | -0,009 | 3,83E-05 | 4,41E-04 | ADAR,CCL5,CCR2,CXCL10,DDX58,DDX60,IFI16,IFI44,IFIT2,IFIT3,IRF7,ISG15,ISG20,OAS1,OAS2,OAS3,RSAD2,STAT1 |
| TLR2/3/4/9 | | group | | 0,743 | 0,761 | 0,006 | 3,95E-05 | 4,54E-04 | CCR7,CD40,CD58,CD83,CD86,FCGR2B,ICAM1,TREM2 |
| NLRP12 | | other | | 1,079 | 1,061 | -0,004 | 4,24E-05 | 4,86E-04 | CASP1,CXCL12,CXCL6,CXCR4,HLA-A,HLA-B,HLA-C,HLA-E,HLA-F,HLA-G,HLA-J,ICAM1,LY96,NOS2,PSMB8,TLR4 |
| ATF2 | | transcription regulator | | -1,376 | -1,333 | 0,010 | 4,52E-05 | 5,14E-04 | ACTB,APOC3,ASNS,ATF3,BCKDHA,BCL2A1,CCNA2,CCND1,CGA,DDIT3,DUSP1,FN1,FOS,GADD45A,IL13,ITGB8,MAT2A,MEIS2,MMP1,NOS2,NPPA,PCK1,PDGFRA,PLA2G4A,PPARGC1A,PTEN,RHOB,SOD2,SST,VCAM1 |
| LIF | 0,019 | cytokine | | 1,577 | 1,594 | 0,003 | 4,52E-05 | 5,14E-04 | A2M,ACAA2,ACACA,AFP,AHNAK,BMP7,CCNA2,CCND1,CD9,CDH5,CISH,CSF1R,CSHL1,CXCL10,CYFIP1,CYP17A1,DCAF11,ENPP2,ERAP1,EVX1,FBP1,FN1,FOS,GPT,HIF1A,HLA-G,HOXB1,HPSE,IFI16,IL7,IRF1,JAM2,KLRB1,KNG1,MEF2C,MMP1,NKX2-5,NMI,NPPA,PECAM1,PLD1,PRDM14,SERPINA3,SOCS2,SOD2,SPP1,STAT3,TAC1,TIMP1,TNC,TNFRSF11B,TP53,TSPAN7,VEGFA,VWF,WT1,XBP1 |

| | | | | | | | | | |
|---|---|---|---|---|---|---|---|---|---|
| GNAQ | | enzyme | | -1,653 | -1,629 | 0,008 | 4,64E-05 | 5,26E-04 | ADAMTS1,ADM,ANXA3,ATF3,BNIP3,BNIP3L,CCND1,CD44,CTSV,CXCL12,DPYS,F2RL1,FOS,GADD45A,GALE,GBP2,GNA11,HSPB8,LHB,NEU1,NFIL3,NOX5,NPPA,NPR3,NR4A1,NR4A3,OAS1,PDE10A,PLSCR1,PRKCD,PYGL,RHOB,SELENBP1,SGK1,STAT1,TNFAIP6,VEGFA |
| platelet activating factor | | chemical - endogenous mammalian | | 1,082 | 1,112 | 0,007 | 4,66E-05 | 5,28E-04 | APOM,CD69,F3,FN1,FOS,ICAM1,IDO1,ITGAM,ITGB2,MMP1,MMP2,MMP9,NOX5,NR4A1,PLA2G7,SELL,SELP,TIMP1,TIMP2,VEGFA,VIM |
| glutathione | | chemical - endogenous mammalian | | 1,988 | 1,987 | -0,000 | 4,69E-05 | 5,30E-04 | ABCC1,ALB,ASS1,ATF3,BAX,CCND1,CDKN1B,DDIT3,DLAT,GCLC,GNL3,GPX3,HIF1A,MAPK8,MET,NDRG1,PCNA,SOD2,SP3,STAT3,TNC,VCAM1,VEGFA,ZBTB10 |
| IGF2BP1 | | translation regulator | Activated | 3,532 | 3,561 | 0,008 | 4,88E-05 | 5,48E-04 | ACTB,BIRC2,CD44,COL1A1,COL1A2,COL5A1,COL6A3,CYFIP2,KRAS,LGALS1,LUM,PTEN,TNC |
| IRS1 | -0,031 | enzyme | | -0,479 | -0,468 | 0,004 | 5,08E-05 | 5,68E-04 | ACACA,ACTR3,CCND1,CDKN2C,COL3A1,COL4A1,COL4A2,COL6A3,DBN1,DDIT3,EFNB2,FHL1,FMO1,FOS,G6PC,GAS2,GJA1,GLRB,ID2,IGFBP4,INSR,MAPK12,MEF2C,NDN,NRP1,PCK1,PHB,PPARGC1A,PTPRF,SIGLEC1,SPP1,TGFBI,TNNC1,TNNI1,TNNT2,TRIB3,UCP2,VEGFA |
| HMGB1 | | transcription regulator | Activated | 2,321 | 2,360 | 0,008 | 5,14E-05 | 5,75E-04 | BAX,CASP3,CCL20,CCNG1,CD163,CD40,CD58,CD83,CD86,CKM,CXCL6,GLUL,HIF1A,HLA-DRB1,HLA-G,ICAM1,IL1R1,INSR,MMP1,MMP9,MOK,MRC1,MX1,PTGES,SMAD7,TLR2,TLR4,VCAM1 |
| ADRB | | group | | -1,655 | -1,676 | -0,003 | 5,32E-05 | 5,94E-04 | ADGRL4,ANXA5,ATF3,AXL,BAX,BHLHE40,BNIP3,BNIP3L,CASP3,CASP4,CASP6,CCNA2,CCND1,CD40,CDC7,CDKN1B,CXCL10,DDIT3,FAS,FOS,GADD45A,GCLM,GSTA3,ICAM1,IDH1,IFIT3,LPIN1,MMP9,MSMO1,NCL,NOS2,NOTCH2,NPPA,NR4A1,NR4A3,NSDHL,PCNA,PCYT2,PPARGC1A,PPM1D,RSAD2 |
| let-7a-5p (and other miRNAs w/seed GAGGUAG) | | mature microRNA | Inhibited | -2,187 | -2,251 | -0,010 | 5,37E-05 | 5,99E-04 | ADGRG1,AGO4,BCL7A,CAPG,CARHSP1,CASP3,CCND1,CCND2,COL1A1,COL1A2,COL3A1,DICER1,EIF4G2,GAK,GYS1,IFIT5,IFRD1,ITGB3,KRAS,KRT19,MMP9,MRPS33,MTRR,NXN,PGRMC1,PRIM1,PXDN,RHOB,RHOG,RTCA,S100A4,SCD,SLC25A1,SLC25A13,SLC25A24,SLC25A32,SLC38A1,SMAD2,SPCS3,STAT3,TGFBR3,TLR4,TRMT1,VIM |
| HNF4α dimer | | complex | Inhibited | -2,979 | -2,950 | 0,010 | 5,53E-05 | 6,09E-04 | ALDOB,FABP1,FABP2,FBP1,HNF1A,PCK2,PKLR,SLC22A7,SLC2A2 |
| IFNA10 | 0,265 | cytokine | Activated | 2,744 | 2,771 | 0,010 | 5,53E-05 | 6,09E-04 | CCL8,CXCL10,CXCL11,CXCL9,IFIH1,ISG15,MX1,PLAAT4,STAT4 |
| IFNA21 | -0,136 | cytokine | Activated | 2,737 | 2,764 | 0,010 | 5,53E-05 | 6,09E-04 | CCL8,CXCL10,CXCL11,CXCL9,IFIH1,ISG15,MX1,PLAAT4,STAT4 |
| IFNA5 | -0,131 | cytokine | Activated | 2,744 | 2,771 | 0,010 | 5,53E-05 | 6,09E-04 | CCL8,CXCL10,CXCL11,CXCL9,IFIH1,ISG15,MX1,PLAAT4,STAT4 |
| IFNA7 | -0,104 | cytokine | Activated | 2,744 | 2,771 | 0,010 | 5,53E-05 | 6,09E-04 | CCL8,CXCL10,CXCL11,CXCL9,IFIH1,ISG15,MX1,PLAAT4,STAT4 |
| IFNA14 | | cytokine | Activated | 2,744 | 2,771 | 0,010 | 5,53E-05 | 6,09E-04 | CCL8,CXCL10,CXCL11,CXCL9,IFIH1,ISG15,MX1,PLAAT4,STAT4 |
| CSF3R | 0,072 | transmembrane receptor | | -0,574 | -0,577 | -0,002 | 5,53E-05 | 6,09E-04 | CD14,CDKN1B,CEBPE,CISH,CXCL12,FOS,ITGAM,LTF,LYZ |
| IFNA6 | | cytokine | Activated | 2,744 | 2,771 | 0,010 | 5,53E-05 | 6,09E-04 | CCL8,CXCL10,CXCL11,CXCL9,IFIH1,ISG15,MX1,PLAAT4,STAT4 |
| advanced glycation end-products | | chemical - endogenous mammalian | | 0,935 | 0,954 | 0,005 | 5,53E-05 | 6,09E-04 | ABCG1,BAX,CASP3,CASP8,CXCL12,FN1,FOXO1,GJA1,MMP1,MMP9,NOS2,PFN1,PTGES,ROBO1,SERPINF1,SLIT2,SPP1,VCAM1 |
| SEMA7A | -0,056 | transmembrane receptor | Activated | 2,857 | 2,891 | 0,010 | 5,58E-05 | 6,11E-04 | BAX,CCN3,CCNA2,COL1A1,COL1A2,COL3A1,CTSB,CTSS,FN1,IL18,IL4R,TIMP1 |
| ALDH2 | -0,584 | enzyme | Activated | 2,925 | 2,891 | -0,010 | 5,58E-05 | 6,11E-04 | CTH,GCLC,GCLM,GNMT,GSS,GSTA3,MTHFD2,PHGDH,PSAT1,SHMT1,SHMT2,SLC3A2 |
| C5 | -0,271 | cytokine | | 1,780 | 1,809 | 0,005 | 5,62E-05 | 6,14E-04 | ATF3,BCL6,CCL5,CCND1,CD55,CD59,CD63,CD86,CDH11,CXCL1,CXCL10,CYBB,CYP3A4,EFNB2,F3,FCGR2B,FN1,HSPA5,ICAM1,IFNGR2,IL13,ITGAM,ITGB2,MET,MMP1,MMP9,NFE2L2,NFKB1,NGF,PPP1R15A,SMAD7,ST6GAL1,TNFAIP2,VCAM1,VEGFA,VEGFC,ZFP36 |
| PELP1 | | other | | -0,825 | -0,814 | 0,003 | 5,62E-05 | 6,14E-04 | ADA,ADRB2,ARMCX1,ASNS,BCAS3,CCND1,CXCL1,DAB2,EGF,F12,FGFR3,FOS,FYN,G0S2,GCA,IFI44,KRT81,KYNU,LCP1,MMP2,MMP9,MRAS,MRC1,NDRG1,NIBAN1,PPARGC1A,PRKCQ,PRSS3,PSAT1,PTEN,PTGES,RBP1,S100A9,SLPI,SNCG,TCN2,ZGPAT |
| IFI16 | 1,554 | transcription regulator | Activated | 2,140 | 2,183 | 0,009 | 5,72E-05 | 6,23E-04 | BAX,CCL20,CCL5,CCND1,CD81,CDKN1B,CGA,CXCL10,DDX58,ESR1,GADD45A,ICAM1,ID2,IFI16,IL2RB,IL7R,ISG15,MIF,MTHFD2,NKX2-5,NPPA,OAS1,PCNA,RPA3,SOD1,VCAM1 |
| lysophosphatidylinositol | | chemical - endogenous mammalian | Activated | 2,480 | 2,524 | 0,010 | 5,95E-05 | 6,45E-04 | C3,CASP1,CD36,CD74,CFB,CXCL6,F3,FPR1,GLS,GLUL,HLA-DRB1,ICAM1,IL15,IL33,LTB,PLCE1,SOD2,TG,TLR3 |

| | | | | | | | | | |
|---|---|---|---|---|---|---|---|---|---|
| HDL | | complex | | -0,966 | -0,977 | -0,002 | 5,95E-05 | 6,45E-04 | ABCG1,ATF3,CCL5,CCR2,CD40,CD86,CX3CR1,DDIT3,HIF1A,HSPA5,ICAM1,ITGAM,ITGAV,ITGB2,ITGB3,MYD88,RAC1,SELP,VCAM1 |
| WWTR1 | -0,167 | transcription regulator | | 1,102 | 1,118 | 0,004 | 5,95E-05 | 6,45E-04 | CCNA2,CD44,CKM,COL1A1,CXCL1,DTYMK,FN1,GJA1,ICAM1,ID2,ITGB2,LAMA5,NPPA,RRM1,RRM2,SMAD7,SPP1,THY1,VCAM1 |
| miR-296-5p (miRNAs w/seed GGGCCCC) | | mature microRNA | | -0,880 | -0,913 | -0,010 | 6,08E-05 | 6,58E-04 | BAX,CCND1,CDKN1B,COL1A1,KLK1,MAP2K3,MMP1,PCNA,TNC,TP53,VEGFA |
| QKI | | other | | 0,923 | 0,923 | 0,000 | 6,16E-05 | 6,62E-04 | CSF1R,DACT1,ERCC2,HNRNPF,ITGB8,MACROH2A1,PEA15,TNC,VCAM1,VIM |
| F3 | -0,574 | transmembrane receptor | | 1,892 | 1,936 | 0,010 | 6,17E-05 | 6,62E-04 | DUSP9,EFEMP1,F3,FERMT2,FGF13,FOS,GJA1,ICAM1,ITGA2,ITGAM,KRAS,LAMB1,LCP1,MDK,MMP2,MMP7,MMP9,RAC1,RHOA,SERPINC1,SPRY1,TGFBR2,VCAM1,VEGFA |
| PLAU | 0,344 | peptidase | Activated | 2,059 | 2,078 | 0,004 | 6,17E-05 | 6,62E-04 | ABCG1,ANG,CCL5,CCND1,CCR5,F3,FN1,GJA1,HLA-DMA,HSPA5,ICAM1,IFNAR2,ITGAV,MMP1,MMP9,OAS1,OAS3,PLG,S100A8,S100A9,SERPINB1,SLPI,TP53,VIM |
| MAP3K14 | -0,082 | kinase | Activated | 2,758 | 2,797 | 0,008 | 6,17E-05 | 6,62E-04 | BIRC3,BST1,CCL5,CD44,CXCL1,FAS,GADD45A,ICAM1,IL13,IL15,IL7R,LTB,MMP9,NFKB1,NFKBIE,NOS2,PLA2G4A,PON2,PTEN,RCAN1,TNFAIP6,TNFSF10,VCAM1,XBP1 |
| mir-1 | | microRNA | Inhibited | -3,658 | -3,703 | -0,008 | 6,18E-05 | 6,63E-04 | ADAR,ANP32B,ANXA2,CACNB2,CAP1,CAPRIN1,CASP3,CCND2,CDK14,CXCL1,DHX15,ESR1,FN1,GJA1,HNRNPA3,HSP90B1,KIF2A,KRAS,MET,MMP9,NELFCD,PDCD4,PPIB,SFRP1,SH3BGRL3,SLC25A1,SMARCA4,TAGLN2,TMX1,TNNT2,UTRN,VEGFA,VEGFC,WBP1L |
| MMP12 | 0,243 | peptidase | | 1,052 | 1,067 | 0,008 | 6,32E-05 | 6,76E-04 | ACTN1,ACTR2,ACTR3,ARPC1B,CAPZA2,CCL5,CCT5,CORO1C,DNM1L,FSCN1,HNF4A,MMP2,MMP9,MYH9,PGM1,PKM,PLS3,PSMA6,PSMB7,PSMD1,RDX,TLN2,YWHAH |
| S-adenosylmethionine | | chemical - endogenous mammalian | | -0,521 | -0,543 | -0,006 | 6,39E-05 | 6,81E-04 | ACACA,ADIPOR1,BACE1,COL1A1,COL1A2,FOS,GCLC,GCLM,ITGAL,MAF,MAT2A,NOS2,PHB,PPARGC1A,PRMT1,RAC1 |
| ISGF3 | | complex | | 1,945 | 1,964 | 0,010 | 6,40E-05 | 6,81E-04 | HIF1A,IFIH1,IFIT2,IRF7,ISG15,RSAD2,TNFSF10 |
| EIF3A | 0,153 | other | | -0,286 | -0,301 | -0,006 | 6,40E-05 | 6,81E-04 | CDKN1B,ISG15,NDRG1,RPA1,RPA2,RPA3,XPC |
| MLXIPL | -0,427 | transcription regulator | | -0,632 | -0,601 | 0,008 | 6,61E-05 | 7,02E-04 | ACACA,ACACB,ELOVL6,G6PC,GPD1,HIF1A,KHK,PKLR,PPARA,PYGL,SCD,SLC2A2,TKFC,TXNIP |
| Mapk | | group | | -1,020 | -0,979 | 0,009 | 6,78E-05 | 7,18E-04 | ADA,APOA1,APOC3,ARC,CAT,CCND1,CDKN1B,COL1A1,CXCL1,CYBB,ESR1,F3,FOS,GJA1,GZMB,HPGD,MMP7,MMP9,NOS2,NR1I3,NR4A3,PER1,PIGR,PRF1,PRKCZ,PTEN,S100A8,SNCG,SOD1,SPP1,STAT1,VCAM1,VEGFA |
| GAST | -0,113 | other | | 1,695 | 1,737 | 0,008 | 6,95E-05 | 7,34E-04 | ABCC2,CCND1,COX5B,DLAT,FOS,HMGB2,IMPDH2,MCM3,MDH2,MMP7,MMP9,PHB,PLA2G7,POR,PRDX3,RAC1,REG1A,RHOA,RPL35A,RPS19,RUNX3,S100A6,SELP,SLC9A3,STAT3,TNC,VCAM1 |
| NFYB | 0,179 | transcription regulator | | -0,143 | -0,124 | 0,010 | 7,11E-05 | 7,50E-04 | ABCG2,ACTB,APOBEC3B,ARID4B,CACNA2D3,CAPN3,CAV1,CCNB1,CCNB2,CCT6B,CDK1,CHPT1,CLDN15,COL1A1,COL1A2,CYTIP,DPP4,DTL,ENTPD7,FAM214B,FAM49B,FAS,FRYL,G6PC,GADD45A,HLA-DQB1,HLA-DRA,HMGB2,HMGN3,ITM2C,KIAA0930,KLHL3,L2HGDH,LPIN1,METTL9,MSRA,OAT,OGG1,PAQR5,PCK1,PCLAF,PCNA,PDIA4,PGRMC1,PRRG2,PSMB7,PTPN6,PTTG1,PXMP4,RHCE/RHD,RHOB,RNPS1,RRM2,S100A4,SLC25A28,ST8SIA4,TGFBR2,TJP1,TM7SF3,TMBIM6,TMEM87A,TOP2A,TPM1,VIM,VWF,YBX3,ZNF468 |
| NFATC2 | | transcription regulator | | 1,144 | 1,172 | 0,004 | 7,36E-05 | 7,72E-04 | ACP5,CASP3,CASP4,CCL5,CCNA2,CCNB1,CCR5,CCR7,CD3G,CD40,CD83,CD86,CISH,CX3CR1,CXCL10,DAB2,ENPP2,FAS,FGL2,GPNMB,HDAC1,IFIT2,IFIT3,IL13,IL15,IL18,IRF1,IRF7,ISG15,ISG20,MAF,NFKB1,NMI,NOX4,NR4A1,PDCD5,PDZD2,PLD1,PLK2,PPARGC1A,PTPRK,RCAN1,RORC,RSAD2,SLC3A2,SLCO3A1,SOCS2,STAT1,TCF4,TERT,TLR2,TLR3,TSC22D1 |
| CTSS | 1,628 | peptidase | | 1,946 | 1,961 | 0,005 | 8,05E-05 | 8,40E-04 | BID,CASP3,CASP8,COL4A1,COL4A2,CTSK,CTSV,FAS,ITGA7,MMP9,PRKCD,TNFSF10,UTRN |
| CCR1 | 0,293 | G-protein coupled receptor | | 1,489 | 1,505 | 0,004 | 8,05E-05 | 8,40E-04 | CCL5,CTSK,CTSS,CTSV,FOS,ITGAM,ITGAV,ITGB3,MMP2,MMP9,SPARC,SPP1,VIM |
| ceramide | | chemical - endogenous mammalian | | 0,743 | 0,754 | 0,003 | 8,05E-05 | 8,40E-04 | BAX,CASP1,CASP3,CD1D,COL1A1,FOS,GSTA1,ICAM1,MMP1,NAT8,NAT8B,SFRP1,TP53 |
| BMI1 | | transcription regulator | | 0,951 | 0,943 | -0,003 | 8,33E-05 | 8,66E-04 | AURKA,BNIP3,CASP8,CD63,CDKN1B,CUL1,CYFIP2,DNMT1,FAS,GLIPR1,IL36A,MCM3,MET,NFKB1,P4HA1,RRM1,SFN,TERT,TP53,TUSC3,VEGFC,VIM,WNT10B |

| | | | | | | | | | |
|---|---|---|---|---|---|---|---|---|---|
| FLI1 | 0,396 | transcription regulator | | 0,346 | 0,373 | 0,006 | 8,52E-05 | 8,84E-04 | ADGRG1,CCL5,CCND1,CCND3,COL1A1,COL1A2,CSF1R,CXCL10,DNMT1,FLI1,HBB,HSPA8,ID2,INPP5D,MMP1,NEU1,RACK1,SNRPB,SRGN,TGFBR2,TNC,VEGFA |
| SERPINF1 | 0,833 | other | | 0,308 | 0,305 | -0,001 | 8,59E-05 | 8,89E-04 | BAX,BCL2A1,BID,CCND1,CDH5,CYBA,CYBB,G0S2,LOXL1,NFKB1,NGF,NOS2,PDGFRA,PPARGC1A,SELP,SERPINF1,SOD2,TJP1,TNFRSF11B,TNFSF10,VIM |
| SIM1 | 0,127 | transcription regulator | | -0,026 | 0,044 | 0,009 | 8,73E-05 | 9,03E-04 | ABCG1,AKR1B1,ANG,AOX1,APOE,ARNT2,AVP,BST1,CBFB,CCS,CD86,CDH3,CELF2,CLCN1,COPZ2,CRISP2,CYBA,EGF,ELL2,FHIT,FKBP3,FNBP1,GLUL,GPX3,GSTA3,GZMH,HLA-A,ICAM1,IFI16,IGFBP4,IL6R,ITGB3,LCP1,LRRFIP1,LTF,MCM7,OXT,PDLIM4,PEA15,PFN1,POSTN,RGS10,SELENBP1,SERPINA5,SLC12A3,SLC17A1,SNN,SNX5,SST,TACR2,THRB,TYK2,TYROBP,USP2,ZDHHC14 |
| GATA3 | 0,178 | transcription regulator | | -1,665 | -1,659 | 0,001 | 9,12E-05 | 9,42E-04 | ACBD4,ADM,AURKAIP1,BCAS3,BCL11A,BHLHE41,BLVRB,CCL5,CCND1,CCR2,CCR5,CD3D,CDKN2C,CDX2,CPA3,CSF1R,CST6,CXCR4,ELOVL6,ESR1,EVPL,FN1,FOS,FSCN1,GLUL,GZMA,ICAM1,IL13,IL2RB,IL7R,ITGAL,ITK,LAIR1,LOC102724788/PRODH,LTB,MAF,NFIL3,NFKB1,NKG7,NOTCH2,NPPA,OVOL1,PCNA,PPARA,PPL,PTGS1,RCAN1,RUNX3,S100A8,S100A9,SGPP1,SLC27A2,SLPI,SP110,STAT4,TJP1,VCAM1,VIM,XBP1,ZBTB7B,ZEB2 |
| NR2F2 | 0,354 | ligand-dependent nuclear receptor | | -0,228 | -0,228 | 0,000 | 9,43E-05 | 9,71E-04 | ALDH2,APOA1,APOA4,APOC3,EFNB2,HNF1A,HNF1B,HNF4A,LTF,NPPA,NR2F2,NRP1,PCK1,PPARA,SHBG,VEGFA |
| NR1D1 | 0,128 | ligand-dependent nuclear receptor | | 1,609 | 1,609 | 0,000 | 9,43E-05 | 9,71E-04 | ACACA,APOC3,BNIP3,CTSV,EHHADH,FOS,G6PC,ME1,MEF2C,NAMPT,PDE8A,RGR,RORC,STAT1,TLR4,ULK1 |
| Ikb | | group | Activated | 2,354 | 2,377 | 0,008 | 9,56E-05 | 9,81E-04 | BLVRA,CCNB1,CCND1,CFB,HHEX,HSPA5,ICAM1,MAT2A,MMP1,NFKB1,NR4A1,PTEN |
| TCF | | group | | -0,292 | -0,273 | 0,010 | 9,56E-05 | 9,81E-04 | ALDH3A2,CCND1,CD44,CNN2,CRAT,DPEP1,FOS,GAD1,GJA1,HTRA1,ID3,IDO1,IFITM1,IHH,IL1R1,MME,MMP2,MMP7,PCCA,PLS3,QPCT,SERPINA3,SERPINA5,SFN,SLC1A5,SLC26A2,TSC22D1,TSPAN8,UGCG |
| Integrinα | | group | Inhibited | -2,214 | -2,236 | -0,010 | 9,66E-05 | 9,84E-04 | ITGAL,ITGAM,ITGB2,TLR2,TLR4 |
| TOX | 0,284 | transcription regulator | | -1,140 | -1,135 | 0,002 | 9,66E-05 | 9,84E-04 | CDKN1B,CDKN1C,FOXO1,ID2,ZBTB7B |
| SEMA3F | | other | | -0,829 | -0,849 | -0,010 | 9,66E-05 | 9,84E-04 | HIF1A,MMP2,MMP9,TP53,VEGFA |
| VCAM1 | 1,222 | transmembrane receptor | | 0,980 | 1,000 | 0,010 | 9,66E-05 | 9,84E-04 | ABCG2,CCND1,CD44,MMP9,VCAM1 |
| myristic acid | | chemical - endogenous mammalian | | -1,430 | -1,408 | 0,010 | 9,66E-05 | 9,84E-04 | F2RL1,NR1H2,NR1H3,PLIN2,RXRA |

### Table S3. Olive

| Upstream Regulator | Expr Log Ratio | Molecule Type | Predicted Activation State | Bias-corrected z-score | Activation z-score | Bias Term | p-value of overlap | B-H corrected p-value | Target Molecules in Dataset |
|---|---|---|---|---|---|---|---|---|---|
| PDGF BB | | complex | Inhibited | -7,802 | -6,166 | 0,249 | 2,07E-21 | 9,92E-18 | ADD3,ATF3,BDKRB2,CCN2,CCNL1,CDKN1A,CDKN1B,CEBPB,CEBPD,CXCL2,DAB2,DUSP1,DUSP5,DUSP6,EDN1,EGR1,EGR3,ETS2,F3,FOS,FOSB,GADD45A,GDF15,GEM,GPR183,HBEGF,HES1,IER2,IER3,JUN,JUNB,KLF6,LDLR,MCL1,NR4A1,NR4A3,PIM1,PPP1R15A,RGS2,RHOB,RND3,SERPINE1,SGK1,TNFAIP3,TRIB1,ZFP36 |
| HNF4A | -0,036 | transcription regulator | | -0,252 | 0,167 | 0,095 | 5,77E-21 | 1,38E-17 | ABCG1,ABHD6,ACP2,ACVR1,AGA,AK2,ALDH8A1,APOE,ARMC1,ARMT1,ATOX1,ATP10A,AVPI1,BET1,C2orf42,CALML4,CCDC90B,CCNA2,CCNG2,CDC123,CDC23,CDK8,CDKN1A,CDKN1B,CEBPB,CEBPD,CETN2,CETN3,CHI3L1,COPB2,CPT2,CSTF2T,CTSA,CUL5,DBT,DUSP6,DYNC1LI1,E2F4,ECD,EGR1,EPHA2,FANCF,FOSL2,FRK,FYCO1,GADD45B,GATA6,GDF15,GIN1,GLCE,GNAI3,GNS,GTF2H1,H2BC5,HBEGF,JUN,JUNB,LARS2,LDAH,LDLR,LIPT1,LPCAT3,LRRC40,LSM5,MAP7,METTL18,MFSD1,MTMR4,MTTP,NAT8,NBR1,NOC3L,NR0B2,NSF,NSL1,NUP54,OGA,PDCD10,PDK4,PDSS2,PEX12,PEX3,PHKB,PIK3R3,PMS1,POP5,PPCS,PPFIBP2,PPP1R3C,PPP2R3C,PSMD1,PTGDS,R3HDM1,RASA1,RBM7,RIOK3,RTCA,SAP18,SERPINE1,SGK1,SLC25A13,SLC35A5,SMARCA5,SMARCE1,SNAP23,SPATA5L1,SREBF2,STAM,STAMBP,SULT1C2,SYNPO,TAOK3,TDP2,TFB2M,THAP1,TIMP3,TMEM140,TNFRSF11B,TPRKB,TRMT1L,UBE2B,UTP14C,WDR12,ZBTB11,ZC3H15,ZFP36,ZNF146,ZNF277 |
| beta-estradiol | | chemical - endogenous mammalian | Inhibited | -3,916 | -2,639 | 0,138 | 2,92E-17 | 3,77E-14 | ABCA1,AIF1,APOE,ARMT1,ATF3,BACE2,BTG2,CCL4,CCN2,CCNA2,CCNG2,CDC123,CDC23,CDKN1A,CDKN1B,CDKN1C,CEBPB,CEBPD,CHGB,CHI3L1,CITED2,CNN2,CRNKL1,CSE1L,CXCL2,DAB2,DRG1,DUSP1,DUSP5,DUSP6,EDN1,EGR1,EGR3,ERBB3,F3,FABP3,FMR1,FOS,FOSB,FOSL2,FRK,GADD45A,GADD45B,GEM,GMFB,GNS,GPR137B,GPRC5A,HBB,HBEGF,HES1,IER2,IER3,IL4R,IL6R,JUN,JUNB,JUND,KLF4,KLF6,KLF9,KPNA4,LDLR,MTMR4,MTTP,NARS1,NEDD9,NFKBIA,NR0B2,NR3C1,NR4A1,NR4A3,PDK4,PIK3C2A,PIK3R3,PPP1R15A,PPP1R3C,PROS1,PTGDS,PTGER4,PTP4A1,RARS1,RBBP8,RFC4,RGS16,RGS2,RGS3,RND3,SERPINE1,SGK1,SLC20A1,SOCS2,SPOCK2,SPRY2,STX3,SULT1C2,SYNPO,TACSTD2,TBL1XR1,THSD4,TIMP3,TNFAIP3,TNFRSF11B,TNFSF8,TNNT2,TRIB1,TSC22D3,UBE2B,USP8,VAMP2,VAV3,ZFP36,ZNF638 |
| KRAS | -0,016 | enzyme | | -1,104 | -1,097 | 0,001 | 2,80E-14 | 1,34E-11 | ABCA1,AMPH,ATF3,ATG5,CAVIN2,CCN2,CCNA2,CDKN1A,CDKN1B,CEBPB,CHI3L1,CHSY1,COL4A3,CSE1L,DDAH1,DUSP1,DUSP5,DUSP6,E2F4,EGR1,EI24,EPHA2,F3,FOS,FOSB,GADD45A,HBEGF,HES1,IER3,JUN,JUNB,KLF6,LIPG,NFKBIA,NR3C1,NR4A1,NSF,PIM1,PLA1A,RAB20,RBM7,RHOB,SERPINE2,SLC20A1,SLCO4A1,SPRY2,SREBF2,TIMP3,TNFRSF11B,TRIB1 |
| TGFB1 | | growth factor | Inhibited | -5,116 | -3,997 | 0,133 | 7,73E-14 | 3,08E-11 | ABCA1,ABCG1,ACVR1,APOE,ATG14,ATG5,BDKRB2,CALML4,CAVIN2,CCL4,CCN2,CCNA2,CCNG2,CCT6A,CDKN1A,CDKN1B,CDKN1C,CEBPB,CHI3L1,CITED2,CNN2,CSRP1,CTCF,CUL5,CXCL2,DAB2,DTX4,DUSP1,EDN1,EGR1,EGR3,EPHA2,F3,FERMT2,FILIP1L,FOS,FOSB,FOSL2,GADD45A,GADD45B,GDF15,GEM,GIN1,GLCE,GLS,GNS,GPRC5A,HBEGF,HES1,IER2,IER3,IFIH1,IL4R,IL6R,ITPR2,JUN,JUNB,JUND,KLF4,KLF9,LDLR,LIPT1,LPCAT3,LSM5,MYO1C,NAB2,NEDD9,NFKBIA,NOC3L,NR4A1,NR4A3,OARD1,OGA,PCOLCE2,PIM1,PLA1A,PPFIBP2,PPID,PPP1R3C,PSMD1,PSPH,PTGDS,PTGER4,RASA1,RFC4,RHOB,SERPINE1,SERPINE2,SGK1,SLC20A1,SLC35A5,SMC4,TIMP3,TNFAIP3,TNFRSF11B,TNNT2,TSC22D3,WDFY3,ZFP36 |
| TP53 | 0,072 | transcription regulator | Inhibited | -3,925 | -3,512 | 0,048 | 4,16E-13 | 1,33E-10 | ACP2,ADD3,AKAP9,APOE,ATF3,BDKRB2,BTG2,CAND1,CASP6,CCL4,CCN2,CCNA2,CCNG2,CCNL1,CDK7,CDKN1A,CDKN1B,CEBPB,CEBPD,CITED2,CKAP2,CNN2,COX15,CPT2,CSRP1,DBT,DRAM1,DUSP1,DUSP2,DUSP5,DUSP6,E2F4,EDN1,EGR1,EGR3,EI24,EIF2B1,EPHA2,F3,FABP3,FERMT2,FOS,GADD45A,GADD45B,GAS6,GATA6,GDF15,GTF2H1,H2BC5,HBEGF,IER3,IL4R,JUN,JUNB,JUND,KLF4,KLF6,LDLR,LIAS,MAP3K4,MCL1,MIS18A,MYL9,MYO1C,MYO6,NARS1,NFKBIA,NR0B2,PDHX,PDSS2,PIK3R3,PIM1,PPP1R15A,PSMD1,PTGDS,PTP4A1,RB1CC1,RFC4,RGS16,RHOB,RND3,RPE,SERPINE1,SERPINE2,SGK1,SLC25A13,SMC4,SOCS2,STAMBP,TDP2,TFB2M,TIMP3,TLR3,TNFAIP8,TSC22D3,UBE2B,VDAC3,XPNPEP1,ZFP36,ZNF84 |
| Raf | | group | Inhibited | -2,773 | -2,395 | 0,158 | 6,13E-13 | 1,72E-10 | CDKN1A,CDKN1B,DUSP6,EDN1,EGR1,FOS,GDF15,IER2,IER3,JUN,MAFF,MCL1,RND3,SLC20A1,SPRY2,SPRY4,TNFAIP3 |
| PRKCD | -0,183 | kinase | Inhibited | -3,854 | -2,879 | 0,198 | 1,92E-12 | 4,59E-10 | ABCA1,CCNA2,CDKN1A,CEBPB,DUSP6,ETS2,FOS,FOSL2,GDF15,GEM,GPRC5A,JUN,JUNB,KLF6,LDLR,LIPG,MCL1,NEDD4,NES,NFKBIA,NR0B2,NR4A1,PPP1R15A,SERPINE1,TNFRSF11B,ZFP36 |
| NUPR1 | 0,161 | transcription regulator | Inhibited | -4,523 | -5,337 | -0,124 | 2,63E-12 | 5,99E-10 | ANK3,ATF3,ATR,AVPI1,BRCC3,CCNA2,CDC23,CDKN1B,CEBPB,CITED2,CSTF2T,DUSP5,ELL2,GADD45A,GDF15,HBEGF,IL6R,JOSD1,KLF4,KLF6,LARS2,MRS2,MRTFB,NSF,NSL1,OSGEPL1,PCTP,PEX12,PIM1,PLK3,PPFIBP2,PPP1R15A,PPWD1,RAB20,RAB29,RPA1,SERPINE1,STK38,STX3,SUOX,TRIB1,UEVLD,VPS45 |

| | | | | | | | | | |
|---|---|---|---|---|---|---|---|---|---|
| NR3C1 | 0,306 | ligand-dependent nuclear receptor | | -1,159 | -0,982 | 0,046 | 3,18E-12 | 6,92E-10 | ABCA1,APOE,CASP6,CAVIN2,CCL4,CDKN1A,CDKN1C,CEBPB,CKAP2,CXCL2,DBT,DUSP1,EDN1,EGR1,F3,FNTA,FOS,GADD45A,GADD45B,GEM,IER2,IER3,IFIH1,JADE1,JUN,JUND,KLF9,MAEA,MCL1,NFKBIA,NME5,NR0B2,NR3C1,PER1,PIK3R3,PPP1R15A,PRKACB,RGS2,RHOB,RRAGC,SERPINE1,SGK1,THAP1,TIMP3,TIPARP,TNFAIP3,TNFAIP8,TNFRSF11B,TSC22D3,WDR12,WDR37 |
| IGF1 | -0,252 | growth factor | Inhibited | -5,145 | -3,935 | 0,199 | 7,84E-12 | 1,56E-09 | ABCA1,BTG2,CAVIN2,CCL4,CCN2,CCNA2,CCNG2,CDKN1A,CDKN1B,CEBPB,CEBPD,CISH,CITED2,DUSP1,DUSP6,EDN1,EGR1,F3,FOS,FOSB,GADD45A,HBB,IER2,IER3,IL4R,JUN,JUNB,KLF6,MCL1,MGA,NFKBIA,NR3C1,NR4A1,PIM1,PTGER4,SERPINE1,SGK1,SLC20A1,TNFRSF11B,UBE2B,ZFP36 |
| TNF | -0,149 | cytokine | Inhibited | -5,876 | -4,614 | 0,144 | 1,16E-11 | 2,23E-09 | ABCA1,AK2,ANK3,APOE,ARHGEF17,ATF3,BDKRB2,BTG2,BTG3,CASP6,CCL4,CCN2,CDKN1A,CEBPB,CEBPD,CHI3L1,CISH,CITED2,COL4A3,CPT2,CRISPLD2,CXCL2,DBT,DUSP1,DUSP2,DUSP5,DUSP6,EDN1,EGR1,EGR3,EPHA2,ETS2,F3,FOS,FOSB,FOSL2,GADD45A,GADD45B,GDF15,GEM,GLS,GNAI3,HBEGF,HES1,IER2,IER3,IFIH1,IL4R,ITPR2,JUN,JUNB,JUND,KLF4,KLF6,KMO,LDLR,MAEA,MAFF,MCL1,MTTP,NEDD9,NFKBIA,NR0B2,NR3C1,NR4A1,NR4A3,PIM1,PLA1A,PLK3,PPP1R15A,PPP1R3C,RGS16,RGS2,RGS3,RHOB,RND3,SERPINE1,SERPINE2,SGK1,SLC20A1,SOCS2,SPSB1,SYNPO,TIMP3,TLR3,TNFAIP3,TNFAIP8,TNFRSF11B,TNFSF8,TSC22D3,UBR2,VAV3,ZBTB11,ZFP36,ZNF330 |
| LDL | | complex | | -2,274 | -1,314 | 0,228 | 1,64E-11 | 2,98E-09 | ABCA1,ABCG1,APOE,ATF3,CASP6,CCL4,CCN2,CDKN1A,CXCL2,DUSP1,DUSP2,EGR1,EGR3,F3,FOS,FOSB,GAS6,GPR183,ITPR2,JUN,LDLR,MCL1,NR0B2,NR4A1,NR4A3,RND3,SERPINE1,SREBF2,TNFAIP3 |
| MAPK1 | 0,125 | kinase | Inhibited | -3,139 | -3,110 | 0,005 | 2,15E-11 | 3,38E-09 | APOE,ATF3,CCL4,CCN2,CDKN1A,CDKN1B,CISH,CPPED1,DAB2,DUSP1,DUSP2,EGR1,FOS,FOSB,FOSL2,GLS,HBB,IFIH1,ITPR2,JUN,JUNB,JUND,MCL1,NR4A1,PROS1,RGS16,RGS2,SOCS2,SPRY2,SPRY4,SPSB1,SUOX,TLR3,TRIM14 |
| MAPK9 | | kinase | | -0,898 | -0,144 | 0,177 | 2,20E-11 | 3,38E-09 | CCN2,CDKN1A,CDKN1B,CEBPB,EDN1,EGR1,FOS,FOSL2,GADD45A,GADD45B,IER2,JUN,JUNB,JUND,KLF6,LSM5,PLA1A,PPP1R15A,RAD23B,TLR3,TNFAIP3,TNFSF8,ZFP36 |
| Mek | | group | Inhibited | -3,744 | -3,135 | 0,143 | 2,26E-11 | 3,38E-09 | APOE,CCN2,CDKN1A,CDKN1B,DUSP5,DUSP6,EGR1,EGR3,EPHA2,ERBB3,FOS,GDF15,IER2,IER3,JUN,JUNB,MAFF,MCL1,NES,NR4A1,RND3,SERPINE1,SLC20A1,SPRY2,SPRY4,TFB2M,TNFAIP3 |
| NRG1 | | growth factor | | -2,231 | -1,718 | 0,174 | 2,26E-11 | 3,38E-09 | ABCA1,ATF1,BET1,CCN2,CDKN1A,CDKN1B,DUSP1,DUSP6,EDN1,EGR1,EGR3,EPHA2,ERBB3,FOS,FOSB,HES1,IER2,IER3,JUN,JUNB,JUND,LPCAT3,MCL1,NR4A1,NR4A3,ZFP36,ZNF33B |
| ESR1 | -0,038 | ligand-dependent nuclear receptor | Activated | 2,302 | 2,759 | 0,060 | 2,52E-11 | 3,65E-09 | ADD3,ANK3,APOE,ATF3,CALML4,CASP6,CCL4,CCNA2,CCNC,CCNG2,CDKN1A,CDKN1B,CDKN1C,CEBPB,CEBPD,DAB2,DUSP5,EDN1,EGR1,ERBB3,ERGIC2,F3,FGFR1OP,FMR1,FOS,FOSB,FOSL2,GADD45B,GDF15,GEM,GGA2,HBEGF,IER3,JUN,JUNB,JUND,LDLR,MAP3K4,MAP7,MYO6,NEDD4,NEDD9,NFKBIA,NLGN1,NR0B2,PDK4,PLCE1,PPP1R3C,PRKACB,PROS1,RBM22,RGS16,RGS3,RIT1,RND3,SERPINE1,SGK1,SMC4,SNAP23,SOCS2,SPRY4,SSX2IP,STEAP1,STX3,SULT1C2,SYNPO,TACC1,TNFAIP3,TNFRSF11B,TRIB1,TSC22D3,TSPYL2,USP8,VAV3,ZNF544 |
| FSH | | complex | Inhibited | -2,747 | -2,282 | 0,127 | 4,69E-11 | 6,24E-09 | ACP2,BTG2,CCN2,CDKN1C,DAB2,DUSP1,ELL2,FILIP1L,FOS,FOSB,FOSL2,GATA6,GEM,GPRC5A,IL6R,JUN,JUNB,KIDINS220,LDLR,MCL1,NR4A1,PLA1A,POP5,PSMD1,PTP4A1,RGS16,RHOB,SERPINE1,SGK1,SNAP23,TNFAIP3,TNFRSF11B,TRIB1,TYRO3,UBE2L3,ZNF331 |
| FAS | 0,053 | transmembrane receptor | | 0,908 | 0,815 | -0,026 | 1,03E-10 | 1,27E-08 | AIFM1,AKAP9,BTG3,CCL4,CCN2,CDKN1A,CDKN1B,CEBPD,CXCL2,EGR1,F3,FOS,FOSB,IER3,JUN,JUND,LDLR,NFKBIA,NR3C1,NR4A1,PLCL2,PPFIBP2,PPP1R15A,PTGER4,PTP4A1,RASA1,RGS3,SERPINE1,STAMBP,STK38,TNFAIP3,TNFRSF11B,TRIB1,ZFP36 |
| Ca2+ | | chemical - endogenous mammalian | Inhibited | -4,021 | -3,013 | 0,226 | 1,73E-10 | 1,92E-08 | ATF3,CDKN1A,CDKN1B,CEBPB,CEBPD,CHGB,CSRP1,DUSP1,EDN1,EGR1,FOS,FOSL2,GLS,HBEGF,IL6R,ITPR2,JUN,JUNB,JUND,KLF4,KLK7,NFKBIA,NR4A1,NR4A3,PER1,SERPINE1,SGK1,SREBF2 |
| TREM1 | -0,136 | transmembrane receptor | Inhibited | -5,550 | -4,491 | 0,216 | 1,82E-10 | 1,97E-08 | ATF3,CEBPB,CXCL2,EDN1,EGR1,EGR3,ETS2,F3,FABP3,FERMT2,GADD45B,GEM,GPRC5A,HBEGF,HES1,IL6R,MAFF,NFKBIA,RGS16,RHOBTB3,RIN2,SLCO4A1,SPRY2,YRDC |
| MAPK8 | -0,093 | kinase | | -1,649 | -0,940 | 0,161 | 3,30E-10 | 3,43E-08 | ABCA1,APOE,ATG5,B4GALT5,CCN2,CCNA2,CDKN1A,CDKN1B,CEBPB,DUSP1,DUSP6,EGR1,FOS,FOSL2,GADD45A,JUN,JUND,KLF6,MTTP,TNFAIP3,TSC22D3,ZFP36 |
| CREB1 | | transcription regulator | Inhibited | -5,654 | -4,841 | 0,143 | 4,51E-10 | 4,47E-08 | ABCA1,APOE,APOLD1,ATF1,ATF3,BTG2,CCL4,CCNA2,CDKN1A,CEBPB,CEBPD,CHGB,CXCL2,DUSP1,EDN1,EGR1,FOS,FOSB,GADD45A,GADD45B,GEM,GLS,IER2,IER3,IMPACT,JUN,JUNB,KLF4,LDLR,MCL1,NAB2,NR4A1,NR4A3,PER1,PPP1R15A,RGS2,SERPINI1,SGK1,SHTN1,SOCS2,TIPARP,ZFP36 |
| FADD | 0,110 | other | Activated | 1,854 | 2,689 | 0,210 | 6,00E-10 | 5,74E-08 | CDKN1A,CDKN1B,CXCL2,EGR1,FOS,GADD45B,IFIH1,JUN,JUNB,KLF6,NAT1,PIM1,PPP1R15A,TIPARP,TLR3,TNFAIP3 |

| | | | | | | | | | |
|---|---|---|---|---|---|---|---|---|---|
| CREM | | transcription regulator | Inhibited | -2,652 | -2,337 | 0,083 | 1,79E-09 | 1,55E-07 | ABCA1,APOE,APOLD1,ATF3,BTG2,CEBPB,DUSP1,EGR1,FOS,GADD45B,JUNB,LDLR,MCL1,NAB2,NR4A1,PER1,PPP1R15A,RHOB,SMC4,TIPARP |
| INS | -0,069 | other | Inhibited | -2,922 | -2,699 | 0,061 | 2,00E-09 | 1,65E-07 | CCN2,CDKN1A,CEBPB,CITED2,CUL5,DUSP1,EDN1,EGR1,ERBB3,F3,FOS,FOSB,FOSL2,IER2,IER3,JUN,JUNB,JUND,NR4A1,SREBF2 |
| IL1B | -0,244 | cytokine | Inhibited | -6,679 | -5,565 | 0,161 | 2,00E-09 | 1,65E-07 | AIF1,APOE,ATF3,BTG2,CCL4,CCN2,CDKN1A,CEBPB,CEBPD,CHI3L1,CISH,CXCL2,DAB2,DUSP1,DUSP5,EDN1,EGR1,ETS2,F3,FMR1,FOS,FOSB,GADD45A,GADD45B,GAS6,GDF15,GEM,HBEGF,HES1,IER2,IER3,IL6R,JUN,JUNB,JUND,LDLR,MCL1,NFKBIA,NR0B2,NR4A1,NR4A3,PIM1,PLK3,PTGDS,PTP4A1,RGS16,RHOB,SERPINE1,SLC20A1,SOCS2,TIMP3,TLR3,TNFAIP3,TNFRSF11B,TSC22D3,ZFP36 |
| SRF | | transcription regulator | | -0,910 | -0,400 | 0,152 | 4,05E-09 | 3,08E-07 | APOLD1,CCN2,CDKN1A,CEBPD,CNN2,CSRP1,DUSP5,EGR1,EGR3,FERMT2,FILIP1L,FOS,FOSB,GADD45A,IER2,JUN,JUNB,KRIT1,LDLR,LPCAT3,MCL1,MYL9,NR4A1,PDCD10,RFC4,RND3,SNX2,SREBF2,TMEM184C,TNNT2 |
| GH1 | | growth factor | Inhibited | -3,345 | -3,058 | 0,074 | 4,61E-09 | 3,39E-07 | ACVR1,ATF3,CEBPB,CEBPD,CISH,EGR1,ERBB3,FOS,GADD45A,GTF2H1,IL4R,JUN,JUNB,LDLR,RAD23B,SOCS2,ZFP36 |
| EGFR | | kinase | Inhibited | -3,573 | -2,600 | 0,186 | 1,38E-08 | 9,58E-07 | CCN2,CCNA2,CCNG2,CDKN1A,CDKN1B,CEBPB,CHI3L1,DUSP1,EGR1,EGR3,EPHA2,ERBB3,F3,FOS,FOSL2,GADD45A,HBEGF,IER2,IER3,JUN,JUNB,JUND,MCL1,NFKBIA,NSF,PRORP,PTGER4,RHOB,SERPINE1,SERPINE2,SPRY2,TNFRSF11B |
| Pka | | complex | Inhibited | -3,284 | -2,519 | 0,204 | 1,71E-08 | 1,13E-06 | APOE,CCN2,CDKN1A,CDKN1B,CEBPB,DUSP1,EDN1,EGR1,FOS,JUN,JUNB,KLF6,LDLR,NR4A1,NR4A3,RGS2,SGK1,TNFRSF11B |
| ADRB | | group | | 1,129 | 0,558 | -0,137 | 1,71E-08 | 1,13E-06 | ATF3,CASP6,CAVIN2,CCNA2,CDKN1A,CDKN1B,CXCL2,E2F4,FOS,GADD45A,GDF15,JUN,JUNB,KLF4,NFKBIA,NR4A1,NR4A3,SLCO4A1 |
| IgG | | complex | | 2,579 | 1,721 | -0,191 | 3,01E-08 | 1,84E-06 | APOE,CCL4,CDKN1A,CEBPB,CEBPD,DUSP1,EDN1,ETS2,FOSB,GPRC5A,IER2,JUND,KLK7,LDLR,PIM1,PPP1R15A,RND3,SERPINE1,TACSTD2,TNFAIP3,ZFP36 |
| Creb | | group | Inhibited | -3,601 | -2,854 | 0,214 | 3,56E-08 | 2,16E-06 | ANK3,ATF3,CDKN1A,CEBPB,CEBPD,CSRP1,DUSP1,EGR1,EGR3,FOS,FOSB,HES1,IER3,JUN,JUNB,LDLR,MCL1,NEDD9,NR3C1,NR4A1,NR4A3,PER1,RGS2,SYNPO,THAP1,TMEM184C,TNFRSF11B |
| Akt | | group | | -0,934 | -0,297 | 0,138 | 6,51E-08 | 3,54E-06 | AMOT,B4GALT5,BTG2,CCN2,CDKN1A,CDKN1B,CDKN1C,CEBPB,E2F4,ERBB3,F3,FOS,HES1,IER3,JUN,MCL1,NES,NFKBIA,NR0B2,PDK4,PTGER4,RHOB,SERPINE1,SGK1,TIPARP |
| EPO | | cytokine | Inhibited | -3,724 | -2,835 | 0,217 | 8,53E-08 | 4,43E-06 | CCNG2,CDKN1A,CDKN1B,CISH,EDN1,EGR1,FOS,FOSB,GDF15,HBB,JUN,JUNB,LDLR,MCL1,NAB2,NFKBIA,NR3C1,PIM1,PLCL2,PRDM14,RNF128,SNAP23,SOCS2,TNFRSF11B,TSC22D3 |
| IL4 | | cytokine | | -2,932 | -1,539 | 0,221 | 9,97E-08 | 5,02E-06 | ABCA1,ACP2,AK2,APOE,APOLD1,CAND1,CASP6,CCL4,CCN2,CD58,CDKN1A,CDKN1B,CISH,CITED2,CXCL2,DDAH1,F3,FGFR1OP,FOS,FOSB,FYCO1,HBEGF,IER2,IER3,IFIH1,IL4R,JUN,JUNB,JUND,KLF6,KLK7,KMO,LIPG,MCL1,NAB2,NEDD9,NFKBIA,NR3C1,NR4A3,PIM1,PMS1,PREPL,PROS1,SERPINE1,SOCS2,TFEC,TIMP3,TNFRSF11B,TNFSF8,TYRO3,XPOT,ZFP36,ZZZ3 |
| ESR2 | | ligand-dependent nuclear receptor | | -0,765 | -0,521 | 0,067 | 1,07E-07 | 5,32E-06 | ABCA1,APOE,ATF3,CCN2,CCNA2,CDKN1A,CDKN1B,CDKN1C,CEBPD,CXCL2,DAB2,DDAH1,EDN1,EGR1,EGR3,FMR1,FOS,GADD45B,GGCT,JUNB,KLF4,NEDD9,NFKBIA,PDK4,PPP1R3C,PTGDS,SERPINE1,SGK1,SOCS2,SYNPO,TNFRSF11B,TRIB1,USP8,VAV3 |
| Pkc(s) | | group | Inhibited | -3,269 | -2,251 | 0,239 | 1,57E-07 | 7,58E-06 | ABCA1,ATF3,CCN2,CDKN1A,CDKN1B,DUSP1,EDN1,EGR1,F3,FOS,GADD45A,GADD45B,JUN,JUNB,KLF6,NR4A1,NR4A3,PER1,PPP1R15A,RGS2,SERPINE1 |
| IL6 | -0,373 | cytokine | Inhibited | -4,406 | -3,040 | 0,221 | 1,88E-07 | 9,01E-06 | ABCA1,ACVR1,APOE,ATF3,BTG2,CCL4,CCN2,CCNA2,CDKN1A,CDKN1B,CEBPB,CEBPD,CISH,CTR9,CXCL2,DUSP1,DUSP6,E2F4,EGR1,ERBIN,ETS2,F3,FOS,GADD45A,GADD45B,IL4R,IL6R,JUN,JUNB,JUND,LDLR,MCL1,MTTP,NFKBIA,NR3C1,PIM1,PPP1R15A,PTGER4,RASA1,SERPINE1,SGK1,SOCS2,TLR3,TNFRSF11B,UBR2 |
| NFKBIA | -0,921 | transcription regulator | Inhibited | -3,304 | -2,692 | 0,117 | 2,59E-07 | 1,17E-05 | AK2,BTG2,CCNA2,CCNC,CDKN1A,CEBPB,CEBPD,CHI3L1,CXCL2,DUSP6,F3,FOS,GADD45A,GADD45B,HES1,IER3,JUN,JUNB,NBR1,NFKBIA,NR4A1,PIM1,RGS16,RGS3,SERPINE2,SGK1,TIMP3,TNFAIP3,TNFAIP8,TNFRSF11B,ZNF638 |
| OSM | | cytokine | | -2,594 | -1,643 | 0,164 | 2,80E-07 | 1,25E-05 | ABCA1,ABCG1,ATF3,BAIAP2,CCNG2,CDKN1A,CDKN1B,CEBPD,CHI3L1,CISH,CXCL2,EGR1,ERBB3,ETS2,F3,FOS,FOSB,GNS,HBEGF,IER3,IL4R,IL6R,JUN,JUNB,KLK7,LDLR,LPCAT3,NEDD4,PTP4A1,SERPINE1,TDP2,TIMP3,TLR3,TNFRSF11B,TYRO3,ZFP36,ZNF330 |
| prostaglandin E2 | | chemical - endogenous mammalian | | -2,563 | -1,913 | 0,151 | 3,88E-07 | 1,66E-05 | ABCG1,ATF3,CASP6,CCL4,CCN2,CDKN1A,CEBPB,CEBPD,DUSP1,EGR1,FOS,FOSB,FOSL2,IER3,JUN,KLF4,MCL1,NEDD9,NFKBIA,NR4A1,PTGER4,RGS2,SOCS2,TNFRSF11B,TSC22D3 |
| nitric oxide | | chemical - endogenous mammalian | | -2,168 | -1,652 | 0,153 | 5,64E-07 | 2,23E-05 | ATF3,CCN2,CDKN1A,CDKN1B,DUSP1,DUSP6,EDN1,EGR1,FOS,GADD45A,IER3,JUN,JUNB,NR3C1,RAD23B,SGK1,TIMP3 |
| NGFR | | transmembrane receptor | | -0,173 | 0,219 | 0,163 | 6,81E-07 | 2,65E-05 | CCNA2,CDKN1B,FOS,JUN,LDLR,NR0B2,RHOB,SERPINE1 |

| | | | | | | | | | |
|---|---|---|---|---|---|---|---|---|---|
| ACVR1C | | kinase | | -1,767 | -1,195 | 0,218 | 7,15E-07 | 2,74E-05 | CCNG2,CDKN1A,CDKN1B,FOS,JUNB,KLF4,SERPINE1 |
| Pdgf (complex) | | complex | Inhibited | -3,778 | -2,953 | 0,237 | 1,10E-06 | 4,09E-05 | CDKN1A,CDKN1B,EDN1,EGR1,F3,FOS,FOSB,JUN,JUNB,LDLR,PIK3R3,SERPINE1,TIMP3 |
| Insulin | | group | Inhibited | -3,336 | -2,762 | 0,120 | 1,27E-06 | 4,61E-05 | ABCA1,APOE,ATF3,CCL4,CCN2,CCNC,CCNG2,CCT6A,CDK8,CDKN1A,CDKN1B,CEBPB,DUSP1,EDN1,EGR1,FOS,FOSB,HES1,JUN,KLF4,MTTP,NFKBIA,NR3C1,NR4A1,NR4A3,OGA,PDK4,PPP1R15A,SERPINE1,SGK1,SREBF2 |
| oleic acid | | chemical - endogenous mammalian | | -1,267 | -0,880 | 0,109 | 1,35E-06 | 4,73E-05 | ABCA1,APOE,DUSP1,EDN1,FOS,GADD45A,GADD45B,GDF15,LDLR,MTTP,NFKBIA,NR4A1,PDK4,SERPINE1 |
| ZMPSTE24 | | peptidase | Activated | 3,065 | 2,451 | -0,208 | 1,40E-06 | 4,89E-05 | ATF3,BTG2,CDKN1A,CISH,CXCL2,GADD45A,GADD45B,RGS16,SOCS2 |
| HRAS | 0,032 | enzyme | | -0,799 | -0,261 | 0,126 | 1,42E-06 | 4,94E-05 | BTG2,CCN2,CCNA2,CDK7,CDKN1A,CDKN1B,CEBPB,CEBPD,CISH,CNN2,COPB2,CSRP1,DUSP1,DUSP6,EGR1,FABP3,FERMT2,FOS,FOSB,FOSL2,GAS6,HES1,JUN,JUNB,KLF6,LIPG,MYL9,MYO1C,NFKBIA,RB1CC1,RHOB,SERPINE1,SERPINE2,SOCS2,SPRY2,SPRY4,TIMP3,TYRO3 |
| Growth hormone | | group | Inhibited | -2,803 | -2,097 | 0,172 | 1,46E-06 | 5,02E-05 | APOE,CEBPB,CEBPD,CISH,CITED2,EGR1,FOS,FOSL2,GDF15,HBB,IER3,JUN,JUNB,LDLR,NEDD9,NFKBIA,PDK4,PPP1R15A,SGK1,SOCS2 |
| hydrogen peroxide | | chemical - endogenous mammalian | Inhibited | -6,022 | -4,804 | 0,228 | 1,99E-06 | 6,39E-05 | ATF3,BTG2,CCL4,CCN2,CCNA2,CDKN1A,CDKN1B,DUSP1,DUSP2,E2F4,EDN1,EGR1,EPHA2,ETS2,FOS,GADD45A,GADD45B,GDF15,HBEGF,JUN,KLF4,LDLR,NR4A1,PDCD10,PIM1,PTGER4,RIOK3,SERPINE1,SGK1,SLC20A1,SREBF2,TNFAIP3 |
| MYC | -0,718 | transcription regulator | | 0,196 | 1,110 | 0,147 | 2,29E-06 | 7,20E-05 | ABCA1,ACVR1,AK2,ANKRD17,CCNA2,CCNG2,CDK7,CDKN1A,CDKN1B,CEBPD,CPT2,CSE1L,DROSHA,DUSP1,DUSP2,DUSP5,DUSP6,EDN1,EGR1,EIF2B1,EPHA2,F3,FOS,GADD45A,GADD45B,GATA6,GDI1,GLS,HES1,IER3,IFIH1,IMPACT,JUN,KLF4,KLF6,MCL1,MFAP1,MYL9,MYO1C,NARS1,NFKBIA,PFAS,PLA1A,PPID,PRKACB,RAB40B,RARS1,RHOB,SEPHS2,SERPINE1,SERPINE2,SGK1,TRIP12,YRDC,ZFP36 |
| RASSF1 | | other | | -1,698 | -1,465 | 0,068 | 2,72E-06 | 8,35E-05 | APOE,CCNA2,CDKN1A,DAB2,EGR1,FOS,GDF15,HBEGF,KLF4,MTUS1,MYL9,PROS1 |
| tretinoin | | chemical - endogenous mammalian | | -2,503 | -1,469 | 0,139 | 3,29E-06 | 9,82E-05 | ABCA1,ABCG1,ADD3,APOE,ATR,BTG2,CAPN7,CASP6,CCL4,CCN2,CCNA2,CCNC,CDKN1A,CDKN1B,CEBPB,CHGB,CITED2,COL4A3,CSE1L,CXCL2,DAB2,DUSP1,E2F4,EDN1,EGR1,F3,FOS,FOSL2,GATA6,GDF15,GPRC5A,HBEGF,HES1,IER3,IFIH1,IL6R,ITPR2,JUN,KLF4,KLF9,KRBOX4,MAP7,MCL1,MSH3,MYO1C,NES,NR0B2,NR3C1,PIK3C2A,PIM1,PLCE1,PLK3,PTGDS,RAD23B,RCBTB2,RFC4,RPA1,SERPINE1,SERPINE2,SKAP2,SYNPO,TERF1,THSD4,TLR3,TNFAIP3,TNFRSF11B,TSC22D3,UBXN8,ZFAND5,ZNF146 |
| TP63 | | transcription regulator | | -2,108 | -1,358 | 0,176 | 3,30E-06 | 9,82E-05 | ATG5,CCNA2,CDKN1A,CDKN1B,CDKN1C,CITED2,COL4A3,CPT2,DUSP6,EI24,EPHA2,F3,FOS,GADD45A,HBEGF,HES1,IER3,JUNB,KHK,KLF6,MAFF,PIK3R3,RBBP8,RPA1,SERPINE1,TIMP3,TNFAIP8 |
| PPARG | 0,174 | ligand-dependent nuclear receptor | | 0,476 | 1,054 | 0,121 | 5,08E-06 | 1,45E-04 | ABCA1,ABCG1,APOE,CAVIN2,CCN2,CDKN1A,CEBPB,CPT2,CXCL2,EDN1,FABP3,GDF15,HES1,JUN,KLF4,KLF6,LDLR,MTTP,NR4A1,NSF,PCTP,PDK4,PHKB,RAB20,RHOB,SAP18,SERPINE1,SGK1,TSC22D3,VAMP2 |
| DACH1 | -0,073 | transcription regulator | Activated | 3,321 | 2,789 | -0,191 | 5,45E-06 | 1,54E-04 | CDKN1A,EGR1,FOS,IER2,JUN,KLF4,SERPINE1,TNFAIP3 |
| Lh | | complex | Inhibited | -2,713 | -2,433 | 0,115 | 5,55E-06 | 1,54E-04 | ACP2,CDKN1C,DAB2,DUSP1,FOS,GEM,KIDINS220,MCL1,PER1,POP5,PSMD1,PTP4A1,RGS16,RHOB,SGK1,SNAP23,TNFAIP3,TNFRSF11B,TRIB1,TYRO3 |
| FOXO1 | -0,079 | transcription regulator | | -2,658 | -1,729 | 0,186 | 5,63E-06 | 1,56E-04 | ATG14,CASP6,CCL4,CCN2,CCNA2,CCNG2,CDKN1A,CDKN1B,CDKN1C,CITED2,CPT2,EDN1,EGR1,FOS,FOSB,GADD45A,GADD45B,IER3,JUN,JUNB,KLF4,MTTP,PDK4,PPP1R15A,SERPINE1,SGK1,SPRY2,SREBF2 |
| ethanol | | chemical - endogenous mammalian | | -1,704 | -1,163 | 0,117 | 5,71E-06 | 1,57E-04 | ABCA1,ABCG1,ATF3,CCN2,CDKN1A,CDKN1B,CEBPB,CEBPD,DUSP1,DUSP6,EDN1,EGR1,FOS,FOSB,FOSL2,GADD45A,JUN,KLF6,NR3C1,PLK3,PPP1R3C,RHOB,SERPINE1,SGK1,TSC22D3,VAMP2 |
| norepinephrine | | chemical - endogenous mammalian | Inhibited | -4,079 | -3,182 | 0,231 | 5,95E-06 | 1,61E-04 | ABCA1,CCN2,CEBPB,DUSP1,EDN1,EGR1,FOS,FOSL2,GDF15,GEM,NFKBIA,NR4A1,NR4A3,PER1,SERPINE1,SGK1 |
| JUN | -1,288 | transcription regulator | | -2,245 | -1,330 | 0,218 | 5,96E-06 | 1,61E-04 | APOE,ATF3,CCN2,CCNA2,CDKN1A,CDKN1B,DROSHA,DUSP1,DUSP5,DUSP6,EDN1,F3,FOS,GADD45A,GAS6,HES1,JUN,JUNB,NFKBIA,NR0B2,NR4A1,PTGDS,RASA1,RHOB,SERPINE1,SGK1,TIMP3,VAV3,ZFP36 |
| NELFB | 0,039 | other | Activated | 2,391 | 2,236 | -0,069 | 6,52E-06 | 1,74E-04 | ATF3,CCN2,CDKN1A,GADD45A,GADD45B |

| | | | | | | | | |
|---|---|---|---|---|---|---|---|---|
| SP3 | 0,227 | transcription regulator | | -1,464 | -0,863 | 0,215 | 8,78E-06 | 2,27E-04 | ABCA1,ADD3,ATF3,CDKN1A,CDKN1B,CHI3L1,CITED2,DUSP1,EGR1,F3,FOS,GDF15,JUN,KLF4,LDLR,NES,SERPINE1,SGK1 |
| CSF3 | -0,072 | cytokine | Inhibited | -3,012 | -2,557 | 0,116 | 9,26E-06 | 2,34E-04 | CDKN1B,CEBPB,CISH,CXCL2,DUSP6,EDN1,EGR1,EGR3,FOS,GADD45A,GADD45B,GPR183,JUN,MCL1,PIM1,TNFAIP3 |
| LIF | -0,303 | cytokine | Inhibited | -2,587 | -2,174 | 0,110 | 9,36E-06 | 2,35E-04 | CCNA2,CDKN1A,CEBPB,CEBPD,CISH,CXCL2,EGR1,FOS,GATA6,HBEGF,HES1,JUN,JUNB,PIM1,PRDM14,SNAP23,SOCS2,TNFRSF11B |
| ERBB2 | | kinase | | -1,079 | -0,050 | 0,212 | 9,37E-06 | 2,35E-04 | BTG2,CAVIN2,CCL4,CCN2,CCNA2,CDC23,CDKN1A,CDKN1B,CEBPB,CPT2,CSE1L,DUSP6,EDN1,EGR1,ELL2,ERBB3,FOS,GAS6,GLS,HBEGF,HES1,IL6R,ITPR2,JUN,JUNB,KLF4,MCL1,MYL9,NEDD9,PDK4,POLR3B,RETREG1,RFC4,RHOB,RPA1,SERPINE1,SERPINI1,SLC66A2,SOCS2,TIMP3 |
| glutamine | | chemical - endogenous mammalian | | 1,611 | 1,201 | -0,131 | 1,11E-05 | 2,66E-04 | ATG5,CEBPB,DUSP1,FOS,GADD45A,JUN,JUNB,KLF4,MCL1,NES,NFKBIA,NR4A1 |
| NFATC2 | | transcription regulator | | -0,115 | 0,466 | 0,178 | 1,28E-05 | 2,99E-04 | ABCA1,CCNA2,CDKN1A,CISH,CITED2,CXCL2,DAB2,EDN1,EGR3,IER3,JUN,NFKBIA,NR4A1,RGS2,SOCS2,TLR3,TNFSF8 |
| C5 | 0,231 | cytokine | Inhibited | -3,238 | -2,510 | 0,206 | 1,39E-05 | 3,19E-04 | ATF3,CCL4,CXCL2,EGR1,F3,GDF15,NFKBIA,PLK3,PPP1R15A,RGS3,SERPINE1,TNFAIP3,ZFP36 |
| HLX | 0,032 | transcription regulator | | 0,251 | -0,222 | -0,182 | 1,41E-05 | 3,23E-04 | CDKN1C,EGR1,GDF15,HES1,IL4R,JUN,SEMA3G |
| STAT3 | -0,113 | transcription regulator | Inhibited | -4,608 | -3,656 | 0,202 | 1,65E-05 | 3,57E-04 | CCL4,CCN2,CCNC,CDKN1A,CDKN1B,CEBPB,CEBPD,CHI3L1,CXCL2,EDN1,EGR1,EGR3,F3,FERMT2,FLRT3,FOS,GADD45A,HBB,IFIH1,IL4R,IL6R,JUNB,KLF4,MCL1,NES,PIM1,SERPINE1,SERPINE2,SGK1,SYNPO,TLR3,TRIM14,ZFP36 |
| thyroid hormone | | chemical - endogenous mammalian | | -0,762 | -0,785 | -0,007 | 1,73E-05 | 3,70E-04 | ABCG1,APOE,BTG2,CCNA2,CDKN1A,CDKN1B,CHGB,DDAH1,EGR1,FOS,GDI1,KLF9,PER1,PTGDS,SERPINE2,SREBF2 |
| Hdac | | group | | 2,027 | 1,923 | -0,029 | 2,02E-05 | 4,24E-04 | ATF3,CCNG2,CDKN1A,E2F4,EDN1,EGR1,EGR3,FOS,GADD45B,IL6R,JUN,KLF6,KLF9,NR4A1,SPRY2 |
| FOXO4 | -0,015 | transcription regulator | | -1,026 | -1,228 | -0,065 | 2,25E-05 | 4,60E-04 | CCN2,CCNG2,CDKN1A,CDKN1B,GADD45A,GADD45B,IER3,SERPINE1,SGK1,SREBF2,ZFAND5 |
| MAPK14 | | kinase | | -1,511 | -0,609 | 0,238 | 2,26E-05 | 4,60E-04 | ABCA1,CCNA2,CEBPD,DAB2,DUSP1,DUSP6,FOS,FOSL2,GADD45A,JUN,JUNB,JUND,LDLR,SGK1,SREBF2,ZFP36 |
| 24-hydroxycholesterol | | chemical - endogenous mammalian | Activated | 2,032 | 2,216 | 0,083 | 2,32E-05 | 4,70E-04 | ABCA1,ABCG1,APOE,LDLR,SREBF2 |
| ADIPOQ | -0,044 | other | | 0,716 | 0,832 | 0,031 | 2,33E-05 | 4,70E-04 | CASP6,CCN2,CDKN1A,CDKN1B,CXCL2,DUSP1,EGR1,F3,FABP3,FOS,HBEGF,IL4R,LDLR,NFKBIA,TNFAIP3 |
| butyric acid | | chemical - endogenous mammalian | | -1,430 | -1,268 | 0,040 | 2,56E-05 | 5,14E-04 | ACP2,CAPN7,CCNA2,CCNC,CDKN1A,CDKN1B,CDKN1C,CETN2,DPP6,DUSP1,E2F4,FOS,GADD45A,GATA6,GDI1,GPRC5A,HBB,ITPR2,KLF4,MCL1,MPP3,NFKBIA,NR4A1,PTGDS,SERPINE1,SGK1,TIMP3 |
| SCARB1 | | transporter | | -0,876 | -1,065 | -0,068 | 2,62E-05 | 5,18E-04 | ABCA1,ABCG1,APOE,CXCL2,F3,FABP3,LDLR,SREBF2 |
| MAP3K8 | -0,098 | kinase | | -2,028 | -1,824 | 0,060 | 2,67E-05 | 5,24E-04 | APOE,CCL4,CDKN1B,CEBPD,CXCL2,DUSP6,FOS,IER3,JUN,JUNB,PTGER4,RGS3,SLCO4A1,UBE2B |
| MAP2K1/2 | | group | Inhibited | -3,661 | -2,947 | 0,242 | 2,97E-05 | 5,73E-04 | ATF3,CCL4,CCN2,CDKN1B,CHI3L1,DUSP1,EGR1,FOS,JUN,MCL1 |
| XDH | -0,168 | enzyme | | -0,357 | -0,478 | -0,046 | 3,12E-05 | 6,00E-04 | ABCA1,ABCG1,CCN2,CEBPB,DUSP1,F3,SERPINE1 |
| FOXL2 | | transcription regulator | Inhibited | -3,680 | -2,990 | 0,231 | 3,25E-05 | 6,19E-04 | ATF3,CXCL2,FOS,IER3,MAFF,NR4A3,PPP1R15A,RGS2,TNFAIP3 |
| MACROH2A1 | 0,041 | other | | 0,655 | 0,333 | -0,107 | 3,25E-05 | 6,19E-04 | ABCA1,CDKN1A,CDKN1C,CITED2,EDN1,EGR1,GADD45A,SERPINE1,SOCS2 |
| THRB | -0,119 | ligand-dependent nuclear receptor | | -1,186 | -0,971 | 0,065 | 3,44E-05 | 6,45E-04 | ANGPTL3,BTG2,CDKN1B,CXCL2,ERBB3,FOS,GDI1,JUN,KLF9,LDLR,NR4A1,PDK4,PER1,PTGDS,SERPINE1,SREBF2,ZNF185 |
| VHL | | transcription regulator | | 0,286 | -0,075 | -0,151 | 3,45E-05 | 6,45E-04 | AIF1,CCN2,CDKN1A,CDKN1B,CDKN1C,CPT2,DAB2,F3,FILIP1L,GLCE,IER3,JADE1,NEDD9 |
| ZBTB17 | | transcription regulator | | -1,286 | -1,912 | -0,238 | 3,69E-05 | 6,79E-04 | BTG3,CCNA2,CDKN1A,CDKN1C,CKAP2,EGR1,IER3,JUN,LDLR,ZFP36 |

| | | | | | | | | | |
|---|---|---|---|---|---|---|---|---|---|
| TP73 | -0,059 | transcription regulator | | -1,603 | -1,011 | 0,121 | 3,71E-05 | 6,81E-04 | ATF3,ATG5,BDKRB2,BTG2,CCNA2,CCNG2,CDKN1A,CDKN1B,CDKN1C,CXCL2,DRAM1,EDN1,EGR1,EPHA2,GADD45A,GPR137B,HBEGF,HES1,IER3,IL4R,PIK3R3,RB1CC1,SERPINE1,SERPINE2,TIMP3 |
| 22(R)-hydroxycholesterol | | chemical - endogenous mammalian | Activated | 1,578 | 2,054 | 0,185 | 3,74E-05 | 6,81E-04 | ABCA1,ABCG1,ANGPTL3,APOE,LDLR,LIPG,NR0B2 |
| cholecalciferol | | chemical - endogenous mammalian | | -2,754 | -1,977 | 0,228 | 4,01E-05 | 7,18E-04 | CDKN1B,CEBPB,CEBPD,CSTF2T,EDN1,FOS,JUN,MYO6,NR0B2,SERPINE1,SLC20A1,TNFRSF11B |
| arachidonic acid | | chemical - endogenous mammalian | Inhibited | -2,519 | -2,291 | 0,071 | 4,04E-05 | 7,21E-04 | ABCA1,ABCG1,EDN1,EGR1,F3,FOS,JUN,KLF6,LDLR,NFKBIA,RGS2 |
| STAT5A | -0,106 | transcription regulator | Inhibited | -3,014 | -2,729 | 0,087 | 4,10E-05 | 7,29E-04 | APOE,CASP6,CDKN1A,CDKN1B,CEBPB,CISH,DUSP5,FOS,FOSB,FOSL2,GADD45A,IL6R,MCL1,PIM1,PPFIBP2,RIN2,RPA1,SOCS2,TACSTD2,ZFP36 |
| AGT | 0,470 | growth factor | Inhibited | -3,845 | -2,837 | 0,207 | 4,19E-05 | 7,39E-04 | ADD3,ANK3,ATF3,BDKRB2,CCL4,CCN2,CDKN1A,CDKN1B,CXCL2,DAB2,DUSP1,EDN1,EGR1,F3,FOS,HBEGF,HES1,ITPR2,JUN,LDLR,NR4A1,PPP1R3C,RGS2,SERPINE1,SGK1,ZFP36,ZNF268 |
| STAT6 | | transcription regulator | | -1,889 | -1,601 | 0,069 | 4,37E-05 | 7,64E-04 | APOE,APOLD1,CASP6,CDKN1A,CDKN1B,CISH,CXCL2,DDAH1,EGR1,FGFR1OP,GADD45B,IER2,IFIH1,IL4R,KMO,MYO6,NEDD9,NFKBIA,PDK4,PROS1,SERPINE1,SOCS2,TFEC,ZFP36 |
| TFAP4 | -0,043 | transcription regulator | | 0,348 | 0,378 | 0,011 | 4,47E-05 | 7,71E-04 | CCNG2,CDKN1A,EGR1,FOS,GDF15,GMFB,RHOBTB3 |
| NR1H3 | 0,078 | ligand-dependent nuclear receptor | | 1,346 | 1,516 | 0,056 | 5,13E-05 | 8,73E-04 | ABCA1,ABCG1,ANGPTL3,APOE,ATF3,CCL4,CCNA2,CDKN1A,CDKN1B,CXCL2,GAS6,LDLR,MYLIP,NFKBIA,NR0B2,SREBF2,TRIB1 |
| KLF2 | -0,312 | transcription regulator | | 1,853 | 1,615 | -0,070 | 5,52E-05 | 9,29E-04 | CCL4,CCN2,CDKN1A,CXCL2,EDN1,F3,GADD45A,HBB,IL6R,JUNB,NFKBIA,PROS1,PTGDS,RGS3,SERPINE1 |
| PRKCA | 0,016 | kinase | | -2,447 | -1,814 | 0,210 | 5,57E-05 | 9,35E-04 | CCN2,CDKN1A,CXCL2,EGR1,FOS,GDF15,HES1,JUN,NFKBIA,RHOBTB3,SERPINE1,USP8 |

### Table S3. Plum

| Upstream Regulator | Expr Log Ratio | Molecule Type | Predicted Activation State | Bias-corrected z-score | Activation z-score | Bias Term | p-value of overlap | B-H corrected p-value | Target Molecules in Dataset |
|---|---|---|---|---|---|---|---|---|---|
| beta-estradiol | | chemical - endogenous mammalian | | -1,779 | -1,249 | 0,026 | 1,12E-50 | 9,24E-47 | A2M,ABCA1,ABCG2,ABLIM1,ACAA1,ACKR2,ACKR3,ACOX2,ACTL6A,ADAMTS1,ADCY1,ADGRF5,ADIPOQ,ADIRF,ADM,ADM2,AFP,AGRN,AHR,ALB,ALDH18A1,ALDH7A1,ALDOA,ALPG,ANKH,ANPEP,ANXA1,ANXA3,ANXA4,AP1G1,APBA1,APBB1,APOA1,APOD,APP,AQP5,ARC,ARMT1,ASB9,ATF3,AURKA,AVP,BAG1,BAX,BAZ1A,BCAS3,BHLHE40,BID,BLNK,BMP5,BMPR1B,BNIP3L,BTG2,BUB3,C3,CA2,CACNB4,CALM1 (includes others),CAMK2A,CAP2,CASP3,CASP4,CAST,CAV2,CBX6,CCL20,CCL25,CCN2,CCN5,CCNA2,CCNB1,CCNB2,CCR4,CCT2,CD24,CD44,CD74,CDC123,CDC20,CDC23,CDC73,CDH15,CDK1,CDK18,CDKN1B,CDKN1C,CENPM,CFB,CGA,CHI3L1,CITED2,CLCA1,CLCN3,CLDN1,CLDN11,CLU,CNTFR,COL1A1,COL1A2,COL3A1,COMT,COX6A2,CRABP2,CRK,CRNKL1,CSE1L,CSHL1,CST8,CSTB,CTLA4,CTSD,CUBN,CXADR,CYB5A,CYBB,CYP24A1,CYP2A6 (includes others),CYP2B6,CYP3A4,DBN1,DCAF6,DDB2,DDIT3,DDOST,DDR1,DEFB1,DERL1,DIO1,DLG3,DMTN,DNAJC3,DNPH1,DRD2,DRD5,DSCAM,DUSP1,DYNLL1,DYRK2,ECHDC2,EGF,EGR1,EGR3,EIF3A,EIF3D,EIF3J,ELK1,ELOC,ENO1,ERF,ESR1,ESRRA,EXT2,F12,F2RL1,F3,FAAH,FADD,FADS3,FAM111A,FARP1,FAS,FBN1,FCGRT,FEZ2,FGF1,FGFBP1,FHL1,FHL2,FLOT1,FN1,FOS,FOSB,FOSL1,FOXC2,FOXO1,FOXP3,FRK,FSTL1,G6PC,GADD45A,GADD45B,GADD45G,GALNT1,GALNT7,GARS1,GBP2,GCH1,GHR,GHRH,GK,GLRB,GM2A,GMFB,GNB1,GNL3,GNRH2,GP2,GPR12,GRB10,GSTA1,GSTP1,GTF2IRD1,GUCY1A1,GUCY1B1,GYPA,HADH,HAND1,HAPLN2,HBB,HBEGF,HDAC1,HIF1A,HLA-A,HLA-DRB1,HLA-E,HMOX1,HNF1B,HOXA10,HPGD,HRAS,HSD11B2,HSD17B4,HSP90B1,HSPA4,HSPA5,HSPB8,HTR5A,HTR6,HTRA1,IER2,IFI30,IFNGR2,IFRD1,IFT122,IGFBP4,IGSF6,IL13,IL15,IL18,IL6R,IL7,IMPDH2,INHA,INPP5F,INPP5J,IPO4,IQGAP1,IRF7,ISG15,ITGAE,ITGAV,ITGB2,ITGB3,ITIH4,IVL,JAK1,JUNB,JUND,KCNN2,KCNN3,KDELR3,KIF1A,KITLG,KLF9,KLK1,KLK11,KLK3,KLK5,KLK6,KPNA2,KPNA4,KRAS,KRT19,KRT4,KRT81,KRT83,KYNU,LAMB1,LAMB2,LAPTM5,LCN2,LDLR,LEFTY2,LEP,LEPR,LEPROT,LGALS3BP,LHB,LIPC,LPL,LPO,LTB,LTF,LUM,LYZ,MAD2L1,MAP2K3,MAPK1,MAPT,MB,MBD4,MCAM,MCM2,MCM3,MCM5,MET,MGLL,MMD,MME,MMP1,MMP7,MPC1,MPZ,MT1X,MUC1,MUC5B,MX1,MYBL2,MYL12A,MYOD1,MYOF,MYOG,NAMPT,NARS1,NCF2,NDRG1,NEDD4L,NELL2,NET1,NFE2L2,NFKB1,NFKBIA,NME1,NOS1,NOS2,NPPA,NPTX1,NR0B2,NR2F1,NR4A1,NR4A3,NRP1,NTR[illegible] |

| | | | | | | | | |
|---|---|---|---|---|---|---|---|---|
| TGFB1 | growth factor | Activated | 1,926 | 2,387 | 0,026 | 9,98E-43 | 4,12E-39 | AASS,ABCA1,ABCE1,ABCG1,ABCG2,ABL1,ACKR2,ACTN1,ADAM10,ADGRB1,ADI1,ADIPOQ,ADM,AFM,AFP,AGR2,AHR,AKR1C1/AKR1C2,ALB,ALDH18A1,ALDH2,ALDH3A2,ALDOA,ALDOB,ALOX12,ALOX5,ALOX5AP,ANKH,ANPEP,ANXA11,ANXA2,APLNR,APOB,APP,AQP8,ARF1,ARF4,ARPC2,ASNS,ASS1,ATG5,ATP13A3,AVP,B2M,BAX,BECN1,BHLHE40,BMP7,BNIP3L,BUD31,C1QA,C1QB,C1R,C1S,C2,C3,CALM1 (includes others),CALML3,CAMK2G,CAMK2N1,CAP1,CAPNS1,CAPRIN1,CASP1,CASP3,CASP4,CASP8,CAT,CAV2,CAVIN2,CBFB,CCL20,CCN2,CCN5,CCNA2,CCNB1,CCNB2,CCR4,CCT2,CCT5,CCT6A,CD163,CD44,CD59,CD83,CDC20,CDC7,CDK1,CDK2AP1,CDK4,CDKN1B,CDKN1C,CDKN3,CFB,CFD,CFI,CHI3L1,CHRNB4,CHST8,CIDEC,CITED2,CKS2,CLDN1,CLU,COL18A1,COL1A1,COL1A2,COL3A1,COL4A1,COL4A2,COL6A3,CORO1C,CRIP2,CRMP1,CSRP2,CTCF,CTLA4,CTSD,CTSS,CX3CR1,CXADR,CYBA,CYBB,CYP11B2,CYP27B1,CYP3A7,DAD1,DDB2,DLX5,DMTN,DNAJB6,DNMT1,DSG2,DTX4,DUSP1,DYNLL1,DYRK2,EGF,EGR1,EGR3,EIF4A3,ELAVL1,ELK3,ELN,ENO1,ENO3,EPCAM,ERF,ESM1,ESPL1,EXT1,EXT2,F12,F2RL1,F3,FAM110B,FAS,FBN1,FCER1G,FDXR,FERMT2,FGF5,FGF6,FGFBP1,FHL1,FILIP1L,FLRT1,FN1,FNDC3B,FOS,FOSB,FOXC2,FOXO1,FOXP3,FUBP1,FURIN,FUS,FUT7,FZD1,G6PC,GAD1,GADD45A,GADD45B,GARS1,GATM,GBP1,GBP2,GDPD5,GIN1,GLI1,GLRX2,GNAS,GNG7,GNL1,GPI,GPR12,GPR21,GZMA,HADH,HAT1,HBEGF,HEXA,HEXB,HIF1A,HK1,HLA-DMA,HLA-DQB1,HMOX1,HNF1A,HNF4A,HNMT,HNRNPAB,HPGD,HSD3B2,HSPA5,HTRA1,IDI1,IER2,IFI16,IFI30,IFIH1,IFIT3,IFRD1,IGFBP4,IL10RA,IL13,IL15,IL17RB,IL18,IL32,IL36A,IL6R,IMPDH1,INHA,INTS13,ITGA6,ITGA7,ITGAE,ITGAV,ITGB2,ITGB3,ITGB6,IVL,JUNB,JUND,KDELR2,KDELR3,KDM5B,KITLG,KLF15,KLF9,KLK11,KLK3,KRAS,KRT10,KRT19,KRT81,LAMC1,LASP1,LBR,LCN2,LDB1,LDLR,LEP,LIN37,LIPF,LOXL1,LPCAT3,LPL,LSR,LTA,LTBP2,MAP2K3,MAP3K1,MAPK1,MAPK13,MCM2,MET,MGAT3,MIA2,MID1,MITF,MLXIPL,MMP1,MMP7,MPRIP,MPZ,MSH6,MSN,MST1R,MTHFD2,MXI1,MYBL2,MYL12A,MYL6,MYOD1,MYOF,MYOG,NAA80,NAB2,NAMPT,NBEA,NCF2,NCR3,NDC80,NET1,NEU3,NFKBIA,NGF,NME1,NMNAT2,NNMT,NOC3L,NOS2,NPPA,NR4A1,NR4A3,NRP1,NUCB2,NUP153,NUP62,OGA,OPN3,OVOL1,P4HA1,PAK3,PAPPA,PCNA,PCOLCE2,PDCD1,PDCD4,PDK2,PECAM1,PFKP,PIGF,PIM1,PKM,PLA1A,PLAGL2,PLK2,PLOD1,PLOD2,PLS3,PLSCR1,PNP,PODXL,POLE2,POSTN,PPARG,PPI[illegible] |

| | | | | | | | |
|---|---|---|---|---|---|---|---|
| tretinoin | chemical - endogenous mammalian | | -0,718 | -0,233 | 0,027 | 3,63E-40 | 9,98E-37 | A2M,AATK,ABCA1,ABCE1,ABCG1,ABI1,ABLIM1,ACACB,ACKR2,ADAM10,ADAM11,ADAR,ADM,AFP,AGO1,AHR,AIMP1,AKAP12,ALAS1,ALDH1A1,ALDH1A3,ALDH3A1,ALOX5,ALPL,ANPEP,ANXA1,ANXA2,ANXA5,APLNR,APOC3,APOD,APP,ARHGAP29,ARHGEF1,ASNS,ATR,AURKA,BHLHE40,BHMT,BID,BIRC2,BMPR1B,BNIP3L,BPIFA1,BTG2,C1QA,C3,CA2,CA8,CACNA1S,CALM1 (includes others),CALML3,CAPN2,CASP1,CASP3,CASP6,CASP7,CAVIN3,CBFB,CCL20,CCL22,CCN2,CCNA2,CCNB1,CCNC,CCT5,CD44,CD47,CD52,CD53,CD63,CD70,CD74,CDC20,CDC73,CDH6,CDK4,CDKN1B,CDKN3,CDX1,CDX2,CEBPE,CGA,CHN2,CHRM2,CITED2,CKS2,CLDN1,CLDN11,CLPS,COL1A1,COL1A2,COL3A1,COL4A1,COL4A2,COL6A3,CRABP1,CRABP2,CRAT,CSE1L,CSF3,CSHL1,CSNK1A1,CTLA4,CTNNA3,CTSD,CTSS,CUBN,CX3CR1,CXCL6,CYBB,CYP11B2,CYP26A1,CYP2B6,CYP4A11,DAAM2,DAD1,DAP3,DAPK2,DBI,DCSTAMP,DDB2,DDIT3,DDX58,DDX60,DHRS3,DHX15,DIO3,DLEU1,DLGAP5,DNMT1,DPP4,DRD2,DTL,DUSP1,DYNLL1,E2F4,ECI2,EFNB1,EGR1,EIF4G1,EIF5,EIF5B,ELAVL3,ELN,ELOC,EMP3,EPCAM,EPHX1,F3,FCER1G,FGF5,FLRT1,FN1,FOS,FOSL1,FOXP3,FRAT2,FUT4,G0S2,GARS1,GATA6,GCA,GCK,GDE1,GDF2,GDF9,GHR,GIMAP6,GJB4,GJB5,GLI1,GM2A,GNAS,GPR21,GRB10,GSPT2,GSTP1,GZMA,HBEGF,HDAC1,HK1,HLA-A,HLA-B,HLA-C,HLA-DMB,HMOX1,HNF1A,HNF1B,HNF4A,HNRNPF,HOXA10,HOXB1,HOXB6,HOXB7,HPRT1,HS3ST1,HSF4,IFI16,IFI35,IFI44,IFIH1,IFIT2,IFIT3,IFIT5,IFITM1,IFNGR1,IGFBP4,IL10RA,IL13,IL15,IL1RAPL1,IL6R,IMPDH2,IRF9,ISG15,ITGA6,ITGAE,ITGB2,ITGB3,IVL,JAK1,KITLG,KLF15,KLF9,KLK3,KRT19,KRT31,LAMB1,LCN2,LEFTY2,LEPR,LGALS1,LGALS3BP,LIPA,LTF,LY6G6D,LYN,LYPD1,M6PR,MAP2K3,MAP3K1,MAP3K10,MAPK10,MAPK8,MARCKSL1,MC1R,MDK,MGST3,MID1,MLF2,MMP1,MMP7,MRAS,MTHFD2,MUC5B,MYBL2,MYOD1,MYOG,NAE1,NCF2,NDRG1,NES,NEUROD6,NEUROG1,NFE2L2,NFKB1,NKX2-5,NMI,NOS2,NPHS1,NPHS2,NPPA,NR0B2,NR2F1,NR2F2,NRGN,NRP1,NTRK3,OAS1,OAT,ODC1,OLFM4,PAM,PCK1,PDCD4,PDE1B,PDIA2,PDYN,PFN1,PGD,PIM1,PKP2,PLAAT4,PLCB2,PLCE1,PLK2,PLK3,PLOD3,PLS3,PLSCR1,PNP,POSTN,PPARA,PPARG,PPP1R16B,PRG2,PRKCA,PRKCD,PRL,PROM1,PRPH,PSMB10,PSMB8,PSMB9,PSMC4,PTEN,PTGDS,PTPRC,PTPRU,RABGGTB,RAD23B,RAE1,RAI14,RALB,RAN,RARG,RARRES1,RB1,RBP1,RFC2,RFC4,RHOA,RLBP1,RORC,RPA1,RPS19,RTP4,RUVBL1,RXRA,S100A10,S[illegible] |

| | | | | | | | | |
|---|---|---|---|---|---|---|---|---|
| HNF4A | -0,298 | transcription regulator | Inhibited | -4,743 | -4,562 | 0,018 | 1,62E-34 | 2,68E-31 | AASS,ABCC3,ABCC6,ABCE1,ABCG1,ACAT1,ACO1,ACOT13,ACOX2,ACTN1,ACTR1A,ACTR3,ADGRB1,ADGRB2,ADH6,ADIPOR1,AFP,AGA,AGTPBP1,AIMP1,AKAP13,AKR1B1,AKR1C1/AKR1C2,ALB,ALDH1A1,ALDH1L1,ALDH2,ALDH3A1,ALDOB,AMT,ANKRD49,ANPEP,ANXA5,APOA1,APOB,APOC1,APOC3,APOH,APOM,APP,AQP8,ARFIP2,ARL4C,ARMC1,ARMC6,ARMT1,ARPC5,ASNS,ATAD5,ATAT1,AVPI1,BAAT,BAZ1A,BCKDHA,BCLAF1,BET1,BHMT,BLOC1S1,BMP5,BMP7,BPHL,BST1,BUD31,C11orf71,C12orf49,C1S,C2,C3,CAP1,CAPZA2,CAT,CBX3,CBX5,CCDC47,CCDC90B,CCL16,CCNA2,CCNB1,CCT8,CDC123,CDC23,CDIPT,CDK1,CDK2AP2,CDK8,CDKN1B,CENPU,CEP83,CETN3,CFAP20,CHCHD3,CHI3L1,CHMP4A,CHRNB2,CIDEC,CKS2,CLCN6,CLDN1,CLDN3,CLPB,CLTCL1,COMMD9,COPB1,COPB2,CRABP2,CRAT,CSK,CSNK2A1,CSTF2T,CTSA,CTSZ,CUTC,CWF19L1,CXADR,CYP26A1,CYP2A6 (includes others),CYP2B6,CYP2C8,CYP3A4,CYP3A5,CYP3A7,CYP7B1,DAD1,DCK,DCTN4,DCTN5,DDOST,DDX25,DDX41,DDX47,DERL1,DHRS7B,DNAJB9,DNPH1,DPAGT1,DTL,E2F4,ECI2,EED,EGR1,EIF4ENIF1,EIF4G1,EIF5,ELP3,EPCAM,EPHX1,ESYT1,ETNK2,F11,F11R,F12,FABP1,FABP2,FAM117A,FANCF,FAS,FBP1,FDX1,FDXR,FHL1,FKBP3,FLOT1,FOXO1,FRK,FURIN,FUZ,FYCO1,FZD1,FZD6,G0S2,G6PC,GADD45B,GADD45G,GATA6,GDAP2,GIN1,GLA,GLYAT,GMDS,GNAI3,GNL3,GOLIM4,GPX7,GRHPR,GRIK3,GSTA1,GTF2E1,GTF2H1,GTF2I,HAAO,HAO1,HBEGF,HBQ1,HDAC6,HEXA,HIF1A,HIKESHI,HLA-A,HLA-B,HLA-C,HLA-F,HLA-G,HMGB2,HMGN4,HNF1A,HNF1B,HNF4A,HNRNPR,HOXC8,HPD,HPF1,HPX,HSD17B4,HSP90B1,HSPA5,IFI30,IFITM2,IFNGR1,IFT122,IL10RA,IL15,INTS13,INTS9,IPO13,ISOC2,ITGA6,ITGA9,ITIH4,JRKL,JUNB,KCNC3,KCNN2,KDM5C,KIF20A,KLF15,KLHL20,KLK14,KPNB1,KRT10,L2HGDH,LARS2,LCN2,LDB1,LDLR,LEPR,LINC00574,LIPA,LIPC,LPCAT3,LPGAT1,LSR,LTF,LYRM4,MAPK8IP2,MBD4,MCM3,MED7,METAP2,METTL18,MFSD1,MGST3,MICU1,MID1,MIS12,MLXIPL,MPZL2,MRPL15,MRPL18,MRPL3,MRPL33,MRPL49,MSN,MT1X,MTMR2,MTNR1A,MUTYH,MYL6,MYO1A,NAMPT,NAT10,NBEA,NCBP2,NCK1,NDC1,NDC80,NDRG1,NFE2L2,NME1,NMNAT2,NOC3L,NPC1L1,NR0B2,NR2F1,NR2F2,NRAS,NSF,NUP54,NUP62,OGA,ORMDL2,OSMR,OVOL1,PAPOLA,PCK1,PCK2,PCNA,PDCD10,PDCD4,PDIA5,PDK2,PDK4,PELO,PEPD,PERP,PHKB,PHTF1,PIGN,PIK3R3,PKLR,PKM,PKP2,PLA2G2D,PLAGL2,PLD2,PLG,PLSCR1,PMEL,PMS1,PNLIPRP1,PNMA1,PNP,PNPLA3,PODXL,POGLUT2,POLL,POP5,PP[illegible] |

| | | | | | | | | | |
|---|---|---|---|---|---|---|---|---|---|
| TP53 | 0,330 | transcription regulator | Activated | 2,214 | 2,383 | 0,009 | 1,09E-32 | 1,50E-29 | A2M,ABCC1,ABCG2,ACACB,ACAT1,ACO1,ACSF2,ACTL6A,ACTN1,ADGRB1,AFP,AHCY,AKAP12,AKR1B1,ALB,ALDH18A1,ALDOA,ALOX5,ANKH,ANXA1,ANXA2,ANXA3,ANXA4,APOA1,APOBEC2,APP,ARAP2,ARL6IP1,ARPC1B,ASNS,ASS1,ATF3,AURKA,BAG1,BAX,BCAP31,BCAS3,BHLHE40,BHLHE41,BID,BIRC2,BMX,BTG2,C2,CA9,CALU,CAMK2N1,CAND1,CAP1,CAPNS1,CARHSP1,CASP1,CASP3,CASP4,CASP6,CASP8,CAT,CAV2,CCN2,CCN5,CCNA2,CCNB1,CCNB1IP1,CCNB2,CCNL1,CD44,CD47,CD59,CD70,CD81,CDC20,CDC25B,CDC7,CDH3,CDK1,CDK4,CDK7,CDKN1B,CDKN3,CEL,CITED2,CKAP2,CKS2,CLU,COL18A1,COL1A1,COL1A2,COL3A1,COL4A1,COL4A2,COL5A2,COMT,COPB1,COQ2,COX6A2,CRYAB,CSHL1,CSK,CSNK1D,CSTB,CTF1,CTSD,CTSF,CYB5A,CYFIP2,CYP24A1,DBI,DCK,DDB2,DDIT3,DDR1,DDX39A,DEK,DHCR24,DLGAP5,DNAJB5,DNAJC15,DNM1L,DNMT1,DPEP1,DPP4,DRAM1,DSTN,DTYMK,DUSP1,DUSP2,E2F4,EGF,EGR1,EGR3,EI24,EIF2B1,EIF4E2,EIF4G2,EIF4G3,EIF5,ENO3,EPHX1,ERF,ESPL1,ESR1,ESRRG,F11R,F3,FAS,FDXR,FERMT2,FGFBP1,FHL1,FHL2,FKBP1A,FKBP3,FKBP6,FN1,FOS,FOSL1,FOXO1,FOXP3,FSTL1,FUBP1,FUT1,FXYD3,FZD6,G0S2,G6PC,GADD45A,GADD45B,GADD45G,GAS6,GATA6,GATM,GBP1,GDPD5,GLB1,GLRX,GLUL,GML,GNAI1,GNL3,GPD1,GPI,GPR12,GPR162,GSK3A,GSTM5,GSTP1,GTF2H1,HADH,HBEGF,HIF1A,HLA-B,HMGB2,HMOX1,HRAS,HS3ST1,HYAL1,IFI16,IFI30,IFI35,IGFBP4,IL10RA,IL7,INKA2,INSR,IRF7,IRF8,IRF9,ISG15,ITGAV,JUNB,JUND,KCNN4,KITLG,KLK3,KPNA2,KPNB1,LAMB2,LAMC1,LASP1,LBR,LDLR,LIPF,LMAN2,LPP,LRRC17,LRRC3,LYZ,MACROH2A1,MAD2L1,MAN2A1,MAP2K3,MAP4K2,MAPK1,MB,MCAM,MCM2,MCM3,MCM5,ME1,MELK,MET,MLF2,MMP1,MPZL2,MRPL15,MRPL18,MSH2,MSH6,MVP,MX1,MYBL2,MYH10,MYO6,MYOF,NAMPT,NAP1L4,NARS1,NCK2,NDC80,NDRG1,NDUFS6,NFKB1,NFKBIA,NKX2-5,NME1,NOS1,NOS2,NPTX1,NR0B2,NR2F1,NRP1,NTSR2,NUP153,OAS1,OAT,P2RX4,P4HA1,P4HB,PAK3,PARVA,PBK,PCK1,PCLAF,PCNA,PDE6A,PDHA1,PDHX,PDIA5,PDIA6,PDK2,PDLIM1,PDYN,PEA15,PECAM1,PERP,PFKFB1,PFKP,PFN1,PGD,PIGF,PIK3R3,PIM1,PKM,PLAAT4,PLK2,PLOD2,PLTP,PODXL,POLE2,POLR2E,POSTN,PPARA,PPARG,PPFIBP1,PPIC,PPM1D,PPP1CA,PPP1R15A,PPP1R17,PPT1,PRC1,PRKCA,PRKD1,PRL,PROM1,PRRX2,PSEN1,PSMA1,PSMB9,PSMD1,PSME3,PSTPIP2,PTEN,PTGDS,PTK2,PTPN12,PTPRO,PTPRU,PTTG1,PYCARD,RAB8A,RAC2,RAD50,RAN,RB1,RBBP7,RFC4,RGS16,RHOA,RHOB,RPN2,RPS19,RR[illegible] |
| OSM | | cytokine | Activated | 4,529 | 4,945 | 0,031 | 4,42E-29 | 4,52E-26 | A2M,ABCA1,ABCC1,ABCC3,ABCC4,ABCC8,ABCG1,ACAA1,ACKR2,ADAMTS1,AFP,AHR,AKR1C1/AKR1C2,ALB,ALDH3B1,ANXA1,ANXA11,ANXA2,ANXA3,ARL4C,ASNS,ATF3,ATP13A3,ATP9A,B2M,BAIAP2,BHLHE40,BST1,C1R,C1S,CAP1,CASK,CASP4,CAT,CBX3,CCL20,CCT2,CDA,CDH3,CDKN1B,CDSN,CGA,CHI3L1,CISH,COL1A1,COL1A2,COL3A1,CSF3,CST6,CTSV,CXADR,CYP3A5,DENND5A,DHCR24,DHRS3,DHX9,DNAJC3,DNM1L,DSG3,ECM2,EGR1,EMP3,EPCAM,EXOSC10,F3,FLOT1,FN1,FOS,FOSB,FOSL1,GADD45G,GBP1,GBP2,GCA,GFPT1,GHRH,GJB5,GLUL,GRHPR,HBEGF,HIF1A,HLA-A,HLA-B,HLA-C,HLA-F,HMGN4,HMOX1,HSD11B2,HSF4,HSPA5,IFI35,IFNGR1,IL13,IL13RA1,IL15,IL18,IL32,IL6R,IL7,IRAK1,IRF7,IRF9,ITGA6,ITGA7,ITPKB,JUNB,KLK1,KLK7,KPNB1,KRT19,LCN2,LDLR,LPCAT3,LRRFIP1,LY6G6C,MARCKS,MGLL,MLLT11,MMP1,MPP2,MPZL2,MT1X,MX1,MYH10,NAMPT,NEDD4L,NELL2,NMT2,NOS2,NRAS,NTSR2,OAS1,OLFML2A,OSMR,PAK2,PCNA,PDCD4,PEPD,PIGR,PLCB4,POLR2E,POLR2L,PPARG,PSMB8,PSMB9,PTEN,RAB4A,RARG,RBFOX2,RORA,RYK,SCNN1B,SERPINA3,SERPINB1,SERPING1,SLPI,SPP1,STAT1,STAT3,SULT2B1,TAP1,TAPBP,TERT,TIMP1,TLR2,TLR3,TNC,TNFRSF11B,TNNC1,TP53,TPBG,TPM1,TRIM22,TSTA3,TUBA1A,TYRO3,UBE2L6,UBQLN2,UPK1A,USP46,VCAM1,[illegible] |

| | | | | | | | | |
|---|---|---|---|---|---|---|---|---|
| IFNG | cytokine | Activated | 5,744 | 6,292 | 0,032 | 4,92E-29 | 4,52E-26 | A2M,ABCA1,ABCC1,ABI1,ACO1,ADA2,ADGRG1,ADIPOQ,ADM,AGRN,AHCY,AHR,ALDH1A3,ALDH1L1,ALDH7A1,ALOX12,ALOX5AP,ALPL,APOBEC3G,APP,ARAP2,ARC,ARFGAP3,ARL6IP5,ASNS,ASS1,ATF3,ATG5,AZGP1,B2M,BAX,BECN1,BID,BIRC2,BLNK,BST1,BTN3A2,C1QA,C1QB,C1R,C2,C3,CASP1,CASP10,CASP3,CASP4,CASP6,CASP7,CASP8,CAT,CCL1,CCL17,CCL20,CCL22,CCL25,CCN2,CCNA2,CCR4,CD163,CD44,CD63,CD74,CD83,CDK4,CDKN1B,CDX1,CDX2,CFB,CHAC1,CHRND,CHST8,CISH,CLIC5,COL1A1,COL1A2,COL5A2,CRABP1,CSE1L,CSF3,CSK,CTSD,CTSS,CTSV,CTSZ,CX3CR1,CXADR,CXCL6,CYB5A,CYBA,CYBB,CYP24A1,CYP3A4,DCT,DDB2,DDIT3,DDX58,DDX60,DEFB1,DHX9,DIO1,DMWD,DPP4,DUSP1,E2F4,EGF,EGR1,EGR3,ELAVL1,ELK1,ELN,EMID1,ENO1,ERAP1,ESM1,ETV7,F11R,F2RL1,F3,FAAH,FANCF,FAS,FBP1,FCER1G,FGF1,FGL2,FHL2,FKBP1A,FKBP6,FN1,FOS,FOSB,FOXO1,FOXP3,FURIN,FZD1,GAD1,GAS6,GBP1,GBP2,GCH1,GCK,GLA,GLUL,GNAS,GNG7,GNL1,GPR12,GSTP1,HAAO,HBEGF,HCP5,HERC6,HIF1A,HLA-A,HLA-B,HLA-C,HLA-DMA,HLA-DMB,HLA-DQB1,HLA-DRA,HLA-DRB1,HLA-E,HLA-F,HLA-G,HMOX1,HSD17B4,HSPA5,HTRA1,IDH3B,IDI1,IER2,IFI16,IFI30,IFI35,IFI44,IFIH1,IFIT2,IFIT3,IFIT5,IFITM1,IFITM2,IFNGR1,IFNGR2,IGFBP4,IL10RA,IL13,IL15,IL17RB,IL18,IL25,IL32,IL6R,IL7,INS,INSR,IRAK1,IRF7,IRF8,IRF9,ISG15,ITGA6,ITGAV,ITGB2,ITGB3,ITPKB,JUNB,JUND,KITLG,KRAS,KRT19,KYAT1,KYNU,LAMB2,LAMC1,LCN2,LEP,LGALS3BP,LPL,LTA,LTB,LY96,LYN,MAFF,MAN2A1,MARCKSL1,METAP2,MITF,MMP1,MRAS,MSH2,MST1R,MUC1,MX1,MYH10,MYOD1,MYOG,NAMPT,NBN,NCALD,NCF2,NCR3,NFKB1,NFKBIA,NGF,NMI,NOS1,NOS2,OAS1,ODC1,OPN3,PAM,PANX1,PAPPA,PBK,PCK1,PDCD1,PEA15,PECAM1,PIGR,PIM1,PKM,PLAAT4,PLSCR1,PMEL,PPARA,PPARG,PRKCA,PRKCD,PRL,PSMA4,PSMA6,PSMB10,PSMB8,PSMB9,PSME1,PSME3,PTGES2,PTPRB,RAB27A,RAB38,RAC2,RACK1,RARRES1,RB1,RBBP7,RFXANK,RHOB,RORC,RTP4,S100A10,SCNN1B,SEC11A,SEC61A1,SEMA4A,SEMA7A,SERPING1,SGPL1,SHARPIN,SLC12A2,SLC1A2,SLC28A1,SLC2A4,SLC37A4,SLPI,SMAD1,SMAGP,SOAT1,SOCS2,SOD2,SP110,SPP1,SQSTM1,SRP54,SSBP1,ST13,STAT1,STAT3,TACR2,TAP1,TAPBP,TBX21,TCIRG1,TERT,TGFBR3,TGM1,TIMP1,TLR1,TLR2,TLR3,TLR6,TMEM50B,TNFRSF11B,TNFSF10,TP53,TP73,TRIM21,TRIM22,TSC22D3,TYROBP,UBD,UBE2D1,UBE2E1,UBE2L6,VCAM1,VDR,VEGFA,VLDLR,WARS1,ZFP36,ZNF638 |

| | | | | | | | | |
|---|---|---|---|---|---|---|---|---|
| MYC | 0,404 | transcription regulator | -0,428 | -0,004 | 0,028 | 1,08E-28 | 8,93E-26 | ABCA1,ABCC1,ABCC3,ABCC4,ABCE1,ABCG4,ACACA,ACAT1,ACTN1,ADAMTS1,ADIPOR1,ADM,AFP,AHCY,AIMP2,AKAP12,ALB,ALDH18A1,ALDOA,ALOX5,ANKRD17,ANXA4,ANXA5,APOC3,APP,ARL6IP1,ASNS,ASS1,ATOH1,AXIN1,BAG1,BAX,BIRC2,BZW2,C12orf49,CAMK2N1,CANX,CAPN2,CAPNS1,CASP1,CASP10,CASP3,CASP8,CAST,CCNA2,CCNB1,CCNB2,CCT3,CD151,CD44,CD47,CD48,CDC20,CDC25B,CDK1,CDK4,CDK7,CDKN1B,CDX2,CEBPE,CIDEC,CKS2,CLPB,CLU,COL15A1,COL1A1,COL1A2,COL3A1,COL4A1,COL4A2,COL5A2,COL6A3,CPD,CRABP2,CRYAB,CSE1L,CSRP2,CST6,CSTB,CTNNA1,CTSD,CTSV,CUL1,CYFIP2,CYP26A1,DBI,DBN1,DCTPP1,DDB2,DDIT3,DEK,DLEU1,DNMT1,DNPH1,DROSHA,DUSP1,DUSP2,EED,EGR1,EIF2B1,EIF3D,EIF4G1,ELAVL1,ENO1,EPCAM,EPHB3,ERAP1,EVX1,EXOSC8,EXT2,F3,FABP1,FABP2,FARP1,FAS,FBN1,FCGRT,FGF5,FN1,FOS,FOSL1,FOXF1,FSTL1,G6PC,GADD45A,GADD45B,GADD45G,GATA6,GBP2,GCK,GFPT1,GLUD1,GLUL,GM2A,GPC3,GPI,GRPEL1,HAND1,HIF1A,HLA-A,HLA-B,HLA-E,HMOX1,HNRNPAB,HRAS,HSD11B2,HSPA14,IFI16,IFI35,IFI44,IFIH1,IFIT2,IFIT3,IFIT5,IL17RB,INS,IRF7,IRF9,ITGA6,KAT2A,KLK1,KLK14,KLK6,KRAS,LAMB2,LAMP1,LEP,LGALS1,LUM,LXN,LYZ,MAD2L1,MAN2A1,MCM5,METAP2,MFAP1,MGST3,MITF,MMP7,MSH2,MSN,MX1,MYBPH,NARS1,NBN,NDRG1,NFE2L2,NFKBIA,NGF,NIBAN1,NME1,NMI,NOCT,NPPA,NRP1,OAS1,OAT,ODC1,OVOL1,PAM,PARP1,PCK1,PCNA,PDCD4,PDHA1,PECAM1,PERP,PEX19,PFKFB1,PFKP,PHF21A,PKLR,PKM,PLA1A,PLS3,PLSCR1,POLR2G,PPARA,PPID,PPL,PRDX4,PRKACB,PROM1,PRPH,PSMB8,PTEN,PTPRC,PYCR1,RARG,RARRES1,RARS1,RB1,RBBP7,RBP1,RHOA,RHOB,RIOX2,ROCK1,RPS19,RRM2,RUVBL1,S100A10,S100A6,SAE1,SARDH,SDCBP,SEMA4A,SERINC3,SERPINE2,SFXN1,SGK1,SIRT2,SLC16A7,SLC22A4,SMAD1,SNRPD1,SOD2,SPP1,SPRR3,SQOR,ST3GAL4,STAT1,STAT3,STMN1,TERT,TES,TGM1,THBS2,TIMP1,TKT,TLE1,TMSB10/TMSB4X,TNC,TNFSF10,TOB1,TP53,TP53I3,TP73,TPP2,TRIM5,TRIP12,TSPAN7,TSPO,TXNRD1,TYMS,UBE2C,UBE2I,VCAM1,VEGFA,VIM,VLDLR,WNT7B,WNT8B,XPO1,YAP1,YRDC,YTHDC1,ZFP36 |

| | | | | | | | | | |
|---|---|---|---|---|---|---|---|---|---|
| TNF | -0,089 | cytokine | Activated | 3,672 | 4,204 | 0,028 | 7,04E-28 | 5,28E-25 | A2M,AATK,ABCA1,ABCC1,ABCC3,ABCG2,ABTB2,ACACA,ACKR2,ACKR3,ADIPOQ,ADM,AIMP1,AKAP12,AKR1B1,ALAD,ALB,ALDH1A3,ALDH2,ALDH3A1,ALOX15B,ALOX5,ALOX5AP,ANKRD55,ANPEP,ANXA1,ANXA11,APOA1,APOBEC3B,APP,AQP5,ARC,ARF4,ARHGDIB,ARHGEF17,ARL6IP5,ASS1,ATF3,AURKC,AVP,AXIN1,B2M,BAX,BCKDHA,BHLHE40,BHLHE41,BID,BIRC2,BLVRA,BTG2,BTN3A3,C3,CA2,CASP1,CASP10,CASP3,CASP4,CASP6,CASP7,CASP8,CAT,CAVIN3,CCL1,CCL17,CCL20,CCL22,CCN2,CCN5,CCR4,CD163,CD1A,CD1B,CD44,CD47,CD5,CD59,CD70,CD83,CDH3,CDK4,CDX1,CERT1,CFB,CFD,CHI3L1,CHRND,CIDEC,CISH,CITED2,CLDN11,CLTCL1,CLU,CNR2,COL15A1,COL1A1,COL1A2,COL3A1,CRYAB,CSF3,CTLA4,CTSF,CTSS,CTSZ,CX3CR1,CXCL6,CYBA,CYBB,CYP27B1,CYP2C8,CYP7B1,DBI,DCSTAMP,DDIT3,DDX58,DENND2D,DHRS3,DIO1,DLEU1,DMBT1,DNMT1,DPP4,DPYS,DUSP1,DUSP2,EGR1,EGR3,ELAVL1,ELK1,ELK3,EPCAM,ESM1,ESR1,EXT1,F11R,F2RL1,F3,FABP1,FADD,FAS,FCER1G,FCGRT,FGF5,FN1,FOS,FOSB,FOSL1,FOXC2,FOXF1,FOXO1,FOXO4,FOXP3,FUT4,FUT7,FZD5,G0S2,GABRA2,GADD45A,GADD45B,GADD45G,GAST,GBP1,GBP2,GCH1,GHR,GM2A,GNAI1,GNAI3,GNL1,GPD1,GRN,GSTA1,GSTP1,HAND1,HBEGF,HDAC1,HERC1,HEXA,HEXB,HIF1A,HLA-A,HLA-B,HLA-DRA,HLA-E,HLA-F,HLA-J,HMGN3,HMOX1,HNF4A,HPGD,HPRT1,HSD3B2,HSP90B1,HSPA4,HSPA5,IER2,IFI16,IFIH1,IFIT3,IFIT5,IFITM1,IFNAR2,IFNGR1,IFNGR2,IGFBP4,IL10RA,IL13,IL15,IL17RB,IL18,IL25,IL32,IL36A,IL7,INHA,INS,INSR,IRF7,IRF8,ISG15,ITGA10,ITGA6,ITGAV,ITGB2,ITGB3,ITGB6,ITGB8,JAK1,JUNB,JUND,KIF20A,KITLG,KLK3,KYNU,LAMC1,LCN2,LDLR,LEP,LPL,LTB,LTBP2,LXN,LY96,LYN,MAEA,MAFF,MAP2K3,MAP4K4,MAPK1,MARCKSL1,MC1R,MCAM,MCM3,MET,MFHAS1,MITF,MMD,MMP1,MMP7,MPC1,MSH2,MST1R,MUC1,MVP,MX1,MYH10,MYL6,MYOD1,MYOG,NAMPT,NCF2,NCK1,NET1,NFE2L2,NFKB1,NFKBIA,NFKBIE,NGF,NME1,NNMT,NOCT,NOS1,NOS2,NOTCH4,NPHS1,NPPA,NR0B2,NR4A1,NR4A3,NRP1,NUCB2,OAS1,ODC1,OLFML2A,OSBPL10,OSMR,P4HB,PAK2,PAPPA,PARP1,PCBP2,PCK1,PCK2,PCNA,PDCD1,PDIA4,PDZD2,PECAM1,PER2,PIGR,PIM1,PKM,PLA1A,PLA2G3,PLA2G4C,PLAAT4,PLAGL2,PLK2,PLK3,PLOD2,PLSCR1,POSTN,PPARA,PPARG,PPIF,PPP1R15A,PPP1R3C,PRKCA,PRKCD,PRL,PSEN1,PSMB10,PSMB8,PSMB9,PSME1,PTEN,PTPN12,PTPRC,PYCARD,RAB32,RAC1,RB1,RBP1,RBPMS,RGS16,RGS2,RGS7,RHOA,RHOB,RORA,RRM1,RRM2,RXRA,SCNN1B,SEC61A1,SERPINA3,SERPINB1,SERPINE [illegible] |

| | | | | | | | | | |
|---|---|---|---|---|---|---|---|---|---|
| ESR1 | -0,196 | ligand-dependent nuclear receptor | Activated | 2,724 | 2,907 | 0,012 | 3,70E-25 | 2,35E-22 | AASS,ABCC3,ABCG2,ABLIM1,ACKR3,ACTR2,ADCY1,AKAP13,ALOX5,ANGPTL2,ANXA3,ANXA4,AP2B1,AP2S1,APOA1,ARFIP2,ASS1,ATF3,AURKA,AVP,BAG1,BAX,BCAS3,BCLAF1,BECN1,BUB3,C3,CA2,CAP2,CAPN2,CASP10,CASP6,CASP8AP2,CASQ2,CAV2,CCL20,CCN5,CCNA2,CCNB1,CCNC,CD59,CD83,CDK1,CDK4,CDKN1B,CDKN1C,CDKN3,CENPM,CENPU,CGA,CHMP4A,CHRM2,CIT,CLCN3,CLDN1,CLN3,CLTCL1,COBL,COL5A2,COMT,CRABP2,CRK,CSAD,CSNK1A1,CTSD,CTSS,CXADR,CYFIP1,CYFIP2,CYP4A11,CYP7B1,DDB2,DLG5,DMTN,DR1,DSG2,DYNC1LI2,DYRK2,EGF,EGR1,ENAH,ENO1,ERGIC2,ESR1,ESRRA,F12,F2RL1,F3,FABP1,FADD,FAM102A,FAS,FHL1,FIBP,FOS,FOSB,FOSL1,FOXO1,FOXO4,FOXP3,FRAT2,GADD45B,GALNT7,GHR,GLB1,GLRX,GM2A,GNAS,GNB1,GNG12,GUCY1A1,GUCY1B1,GULP1,HACD1,HAX1,HBEGF,HCP5,HIF1A,HIP1R,HLA-B,HLA-C,HLA-E,HLA-G,HLA-J,HOMER3,HPX,HSD17B4,HSD3B2,HSP90B1,HTRA1,HYAL1,IFI44,IFITM1,IFITM2,IFRD1,IGFBP4,IMPDH2,INSR,IQGAP1,ITGA10,ITGA6,ITGB1BP1,IVL,JAK1,JUNB,JUND,KCNE2,KIF20A,KLHL20,KLK3,KLK5,KPNB1,KRAS,KRT19,LAMB1,LAMB2,LAPTM5,LCN2,LDLR,LEP,LEPR,LGALS1,LGALS3BP,LHB,LIPC,LPL,LTB,LTBP4,LTF,LYPD1,M6PR,MAD2L1,MAP3K1,MAP3K12,MAPK1,MAPK8,MAPT,MARCKS,MAST2,MC1R,MMD,MME,MMP1,MPC1,MST1R,MX1,MYH10,MYO6,NCF2,NDC1,NELL2,NFKB1,NFKBIA,NFKBIE,NLGN1,NME1,NOS2,NR0B2,NRCAM,NUP153,NUP210,PAK2,PBK,PCNA,PCNT,PDCD2,PDCD4,PDCD5,PDK4,PGLS,PIBF1,PLA2G4C,PLCE1,PLOD2,PLS1,PLXNA2,PPP1R12A,PPP1R3C,PPP6R2,PRKACB,PRKCD,PRKX,PRL,PROM1,PROS1,PSIP1,PTCH2,PTEN,PTGER3,PTK2,PTP4A2,PTPRT,PTTG1,PTTG1IP,PYCR1,RAB27A,RAI2,RB1,RBM22,RDH16,RGS16,RHOQ,RIT1,ROCK1,RORC,RRBP1,RTP4,SCARB2,SCNN1B,SEMA3B,SGK1,SLC10A1,SLC13A4,SLC25A36,SLC26A2,SLC2A4,SLC6A4,SLC9A5,SMAD2,SMC4,SNAI1,SNAP23,SNN,SOCS2,SOX4,SP110,SPINT1,SPP1,SPRY4,SPTLC2,SQSTM1,SRPK2,STAT1,STAT3,STAT5A,STC2,STEAP1,STX3,SYNPO,TACC1,TAPBP,TCIRG1,TEAD4,TERT,TESK1,TFF2,THBS4,THRB,TIMM17B,TMED10,TMPO,TNFRSF11B,TNFSF10,TNPO1,TOB1,TP53,TP53BP1,TP53I3,TPD52L1,TPM1,TPP2,TRAM1,TRPM2,TSC22D3,TSKU,TSPO,TSPYL2,TYK2,UBE2I,USP8,VEGFA,VEZF1,VIM,VPS35,VRK2,WARS1,WEE1,WNT11,WRAP73,YARS1,YIPF3,ZBTB7B,ZNF544 |
| KRAS | 0,279 | enzyme | | -0,394 | -0,392 | 0,000 | 5,82E-25 | 3,43E-22 | ABCA1,ABCC4,ACTR3,AGRN,AHR,AIMP2,ALDOA,ALDOC,AMPH,ANPEP,AQP5,ATF3,ATG5,B2M,BAG1,BZW2,CASP1,CASP3,CAVIN2,CBX5,CCN2,CCNA2,CCNB1,CCT5,CD44,CD74,CDH3,CDKN1B,CHI3L1,CLIC5,CLU,COL1A1,COL3A1,COL4A1,CRYAB,CSE1L,CSF3,CXADR,CXCL6,DBN1,DDIT3,DNMT1,DPYSL2,DUSP1,E2F4,EGR1,EI24,EIF5,ELK1,ELN,ENO1,EPCAM,EPM2A,F2RL1,F3,FAS,FBN1,FLII,FN1,FOS,FOSB,FOSL1,FOXP3,FSTL1,FXYD3,GADD45A,GCNT3,GFPT1,GLI1,GLUL,GPC3,GRN,GSTA3,GTF2I,HBEGF,HEXA,HIF1A,HLA-DRB1,HMOX1,HOPX,HSPA5,IFITM1,IFITM2,IRF9,ISG15,ITGB3,JUNB,KCNN4,KPNB1,KRAS,LAMB1,LAMP1,LCN2,LTBP4,LYZ,MAN2A1,MAPK8,MCAM,MET,MMD,MMP7,MSN,MUC6,MX1,MYBL2,NETO2,NFE2L2,NFKB1,NFKBIA,NOS2,NR4A1,NRAS,NSF,NXN,OAS1,ODC1,P3H3,PARVA,PDIA2,PDIA3,PECAM1,PIM1,PKM,PLA1A,PPARG,PPFIBP1,PPP1CA,PRC1,PRDX4,PRKCD,PRPH,PSEN1,PTEN,PTK2,PTTG1,PYGM,QTRT2,RAB27A,RAI14,RBBP7,RBM7,RCL1,RHOA,RHOB,RHOC,ROCK1,S100A4,S100A6,SEMA7A,SERPINA3,SERPINE2,SLC34A2,SLC7A10,SNAI1,SOAT1,SPP1,SPRY2,SQSTM1,ST5,STAT1,STAT3,STAT5A,TAP1,TBX21,TESC,TNC,TNFRSF11B,TP53,TRMT2A,UBE2I,VCAN,VEGFA,VIM,VPS35,W[illegible] |

| | | | | | | | | |
|---|---|---|---|---|---|---|---|---|
| IL6 | -0,273 | cytokine | 1,198 | 1,722 | 0,042 | 4,81E-23 | 2,48E-20 | A2M,ABCA1,ABCC1,ABCC3,ABCG2,ACKR2,ADAMTS1,ADGRB1,ADM,AFP,AHR,ALAS1,ALB,ANPEP,ANXA1,APOA1,APOB,APP,ARL4C,ATF3,AVP,AXIN1,BAX,BTG2,BUB3,C1QL1,C3,CAPRIN1,CASP1,CASP3,CASP4,CASP7,CCL20,CCN2,CCNA2,CCNB1,CCNB2,CCR4,CD163,CD44,CD48,CD53,CD74,CD83,CDC20,CDK1,CDKN1B,CDKN3,CDX2,CFD,CHRNB4,CISH,CLU,COL1A1,COL3A1,CRYAB,CX3CR1,CXCL6,CYBB,CYP2B6,CYP2C8,CYP3A4,CYP3A5,DIO1,DLX5,DNMT1,DUSP1,E2F4,EGR1,F12,F3,FAS,FCGRT,FN1,FOS,FOSL1,FOXP3,G6PC,GADD45A,GADD45B,GADD45G,GBP2,GCH1,GHRH,GINS1,GLI1,GLRX,GSTA1,HIF1A,HLA-A,HLA-DMA,HMOX1,HOMER3,HPGD,HPX,HSPA5,IFI16,IFIT2,IGFBP4,IK,IL13,IL15,IL25,IL6R,IL7,IRF9,ITGAV,ITGB3,JAK1,JUNB,JUND,KLK3,LCN2,LDLR,LEFTY2,LEP,LGALS1,LPL,LTF,LY96,LYZ,MACROD1,MAD2L1,MCM3,MET,MIA2,MMD,MMP1,MMP7,MRAS,MUC1,MYBL2,MYOD1,NAMPT,NCF2,NEU3,NFKB1,NFKBIA,NGF,NOS2,NRAS,NUCB2,PCLAF,PCNA,PDCD1,PECAM1,PIM1,PLA2G2D,PLG,PLK2,PPARG,PPP1R15A,PROM1,PRPH,PSMB10,PSMB8,PSMB9,PSME1,PTPRC,PTTG1,RAB27A,RASA1,RB1,REG1A,RNASE6,RORA,RORC,RRM2,SEMA4A,SEMA6D,SENP2,SERPINA3,SGK1,SH3GLB1,SLC10A1,SLC12A2,SLC2A4,SLC7A7,SNAI1,SNX10,SOCS2,SOD2,SP110,SPP1,SST,STAP2,STAT1,STAT3,TAF9,TAP1,TBX21,TERT,TIMP1,TLR1,TLR2,TLR3,TLR6,TNC,TNFRSF11B,TNFSF10,TNS4,TP53,TYK2,UBE2C,VCAM1,VEGFA,VIM,VLDLR,VTCN1,WARS1,WEE1,XPNPEP2,XRCC5 |
| NR3C1 | 0,075 | ligand-dependent nuclear receptor | -1,911 | -1,839 | 0,009 | 3,55E-22 | 1,73E-19 | A2M,AATK,ABCA1,ABI1,ABL1,ACAT1,ACTN1,ADM,ALOX5AP,ANXA1,APP,AQP5,ARPC2,AVP,B2M,BAG1,BAG2,BAX,BCKDHA,BHLHE40,BIRC2,BNIP3L,C1R,CAPN2,CASP10,CASP4,CASP6,CASP7,CAT,CAVIN2,CD47,CD70,CD83,CDKN1C,CGA,CIDEC,CKAP2,COMT,CORO1C,CRABP2,CRYAB,CUL1,CYP11B2,CYP2A6 (includes others),CYP2C8,CYP3A4,CYP3A5,CYP3A7,DAD1,DAPK2,DBI,DNAJC15,DPF2,DUSP1,DUSP22,DYNLL1,EGR1,ETNK2,F2RL1,F3,FADD,FASTKD1,FCER1G,FN1,FNTA,FOS,FOXO1,G6PC,GADD45A,GADD45B,GBP2,GLUL,GNMT,GPR65,GRIA3,GSTA1,GSTP1,GULP1,GZMA,HDAC1,HSD11B2,HSPA6,IER2,IFIH1,IFIT2,IFT122,IGFALS,IL13,IL15,IL18,INSR,IRF8,ISG15,ITGAV,ITGB2,ITGB6,JUND,KLF9,KLK3,LEP,LHB,LIPC,LTB,MAEA,MALT1,MAPK1,MAT1A,MEST,MMP1,MYOC,NAMPT,NFIL3,NFKB1,NFKBIA,NHLH1,NME5,NOS2,NOTCH4,NPPA,NR0B2,NSMAF,NTF3,OAT,PAK2,PCK1,PDCD4,PEA15,PER1,PIGR,PIK3R3,PLK2,PPARA,PPARG,PPP1R15A,PRKACB,PRKAR1B,PRKCD,PRL,PRPH,PSEN1,PSMA6,PSMG2,PTEN,PTP4A2,PTRH2,RAB11A,RABGGTB,RB1,RGS2,RHOB,ROCK1,RRAGC,SERINC3,SERPINA6,SGK1,SGPL1,SH3GLB1,SLC10A2,SLC19A2,SLC1A6,SLC1A7,SLC2A5,SLC43A1,SMAD1,SOX4,SPP1,SRD5A2,STAT3,STAT5A,STK39,STUB1,SYT2,THAP1,TIPARP,TLR1,TLR2,TM2D3,TNFAIP8,TNFRSF11B,TNS4,TP53,TP53BP2,TP53I3,TSC22D3,UBE2K,UGCG,VCAM1,VDR,WDR3,YWHAE,YWHAH |

| | | | | | | | | | |
|---|---|---|---|---|---|---|---|---|---|
| IL1B | -0,096 | cytokine | | 1,385 | 1,820 | 0,031 | 5,14E-22 | 2,36E-19 | A2M,ABCC3,ABCG2,ACKR2,ADAMTS1,ADM,AKR1B1,ALDH7A1,ALPL,ANKH,ANXA1,APOB,APP,ARC,ARL6IP5,ASIC3,ASS1,ATF3,AVP,B2M,BAX,BIRC2,BTG2,C1R,C3,CASP10,CASP3,CASP4,CAT,CCL1,CCL17,CCL20,CCL22,CCN2,CD44,CD74,CD83,CDX1,CFB,CHI3L1,CISH,CNR2,COL1A1,CRYAB,CSF3,CTSF,CTSS,CTSZ,CX3CR1,CXCL6,CYBA,CYBB,CYP11B2,CYP27B1,CYP2B6,CYP3A4,CYP3A5,DBI,DDIT3,DIO1,DNMT1,DPP4,DPY19L4,DR1,DUSP1,EGR1,ELAVL1,ELN,ESR1,ESRRA,F2RL1,F3,FABP1,FAS,FN1,FOS,FOSB,FOSL1,FOXO1,FURIN,FUT7,G0S2,GAD1,GADD45A,GADD45B,GAS6,GAST,GBP1,GBP2,GCH1,GCK,GHR,GLA,GM2A,GNAS,GNL1,GRN,GSTA1,HBEGF,HDAC1,HEXA,HEXB,HIF1A,HLA-A,HLA-DRA,HLA-E,HMOX1,HNF1B,HNF4A,HPRT1,HSPA5,IER2,IFI16,IFIT3,IFNGR1,IFRD1,IGFALS,IGFBP4,IL10RA,IL15,IL18,IL25,IL32,IL36A,IL6R,IL7,INS,INSR,IRAK1,IRF7,ISG15,ITGAV,ITGB3,ITGB8,ITPKB,JUNB,JUND,KRT19,LCN2,LDLR,LEP,LHB,LTA,LY96,MAPT,MARCKSL1,MMP1,MMP7,MUC1,MUC5B,MX1,MYOD1,MYOG,NAMPT,NFE2L2,NFIL3,NFKB1,NFKBIA,NGF,NIBAN1,NMI,NOCT,NOS1,NOS2,NPPA,NR0B2,NR4A1,NR4A3,NRP1,ODC1,OSMR,P4HB,PAPPA,PCNA,PDCD1,PFKP,PIGR,PIM1,PLA2G3,PLK3,PLSCR1,POSTN,PPARA,PPARG,PRKCD,PSEN1,PSMB10,PSMB8,PSMB9,PTGDS,RAC2,RAN,RGS16,RHOB,RORA,RORC,RXRA,S100A10,S100A6,SCNN1B,SEC61A1,SERPINA3,SGPL1,SIRT3,SLC10A1,SLC10A2,SLC1A2,SLC22A4,SLC2A4,SLC37A4,SLC6A4,SOCS2,SOD2,SPP1,SSTR2,STAT1,STAT3,STAT5A,TBP,TBX21,TFPI2,TGM1,TIMP1,TLR2,TLR3,TNFRSF11B,TNFSF10,TNMD,TOB1,TPD52L2,TSC22D3,UBD,UBE2L6,UBE2V2,UGCG,VCAM1,V |
| MAPK1 | 0,300 | kinase | Inhibited | -6,373 | -6,362 | 0,001 | 1,53E-21 | 6,63E-19 | ACO1,ADAR,ANPEP,APBB3,ARC,ATF3,AURKA,BTN3A3,C1S,CAPN2,CAV3,CCL1,CCN2,CCNB1,CDKN1B,CFB,CGA,CISH,CRABP2,CRYL1,CSTA,DDX58,DNMT1,DSE,DUSP1,DUSP2,EGR1,ENDOD1,ENO3,ERAP1,ESR1,EXOC2,FHL2,FN1,FOS,FOSB,FOXP3,GBP1,GBP2,GRAMD1B,HBB,HLA-B,HLA-C,HLA-DMA,HLA-E,HMOX1,IFI16,IFI35,IFI44,IFIH1,IFIT2,IFIT3,IFIT5,IFITM1,IL13,IRF7,IRF9,ISG15,ITGAV,ITGB3,JUNB,JUND,LGALS1,LGALS3BP,LHB,MAPK13,MAST3,MITF,MMP1,MNS1,MVP,MYBL2,NFE2L2,NMI,NOS2,NOX5,NPPA,NR4A1,NRP1,OAS1,OLFML2A,PARP12,PHF11,PLSCR1,PPT2,PRL,PROS1,PSMB8,PSMB9,PTPRK,RFPL1/RFPL3,RGS16,RGS2,SNAI1,SOCS2,SP110,SPP1,SPRY2,SPRY4,STAT1,SUN2,TAP1,TBX21,TDRD7,TLR3,TM4SF5,TMEM87A,TNFSF10,TP53,TRIM21,TRIM22,TRIM38,TRIM5,TSKU,UBE2C,UBE2I,UBE2L6,VDR,VIM,ZC3HAV1 |
| HRAS | 0,270 | enzyme | | 0,093 | 0,306 | 0,024 | 3,05E-21 | 1,26E-18 | A2M,ACTG1,ADM,AFP,ALDH1A3,ANXA2,ANXA3,APP,ARF1,ASNS,ATP1A3,AURKA,B2M,BAX,BMP7,BTG2,CACNG4,CALU,CAPG,CAPN2,CAV3,CCN2,CCNA2,CCNB1,CD24,CD44,CD81,CDH3,CDK1,CDK4,CDK7,CDKN1B,CDX2,CFB,CGA,CISH,CLU,COL18A1,COL1A1,COL1A2,COL3A1,COL4A1,COL5A2,COPB2,CRABP2,CSRP2,CSTA,CTSS,CTSV,CTSZ,CYP24A1,DDIT3,DNMT1,DPF1,DPP4,DTYMK,DUSP1,EGR1,EIF4E2,EIF4G3,ELK1,ELK3,ELN,EPHX1,FAS,FERMT2,FGFBP1,FHL1,FKBP6,FN1,FOS,FOSB,FOSL1,FOXO1,FSTL1,FURIN,FUT7,FZD1,GADD45G,GAS6,GNB1,GRN,HIF1A,HLA-B,HMOX1,HOXB6,HOXC8,HRAS,HTRA1,IFI30,IFITM2,IGFBP4,IL13RA1,ITGA6,ITGAE,ITGAV,IVL,JUNB,KLF3,KRAS,KRT19,LASP1,LOXL1,LPP,LRRC17,LXN,LYZ,MAPK1,MCAM,MET,MMP1,MYH10,MYL12A,MYOD1,MYOG,NFKBIA,NNAT,NOS2,NPPA,NRP1,OSBP2,P4HB,PAFAH1B3,PCDHGA1,PCK1,PCNA,PDCD4,PDIA5,PDLIM1,PER2,PFN1,PKP1,PLOD1,PLOD2,PLS3,PLTP,POLR2G,POSTN,PPIC,PRKCA,PRKCD,PRL,PSEN1,PSMB9,PTEN,RBBP7,RBPMS,RHOA,RHOB,RRM1,RRM2,RSU1,S100A4,SEM1,SERPINA3,SERPINE2,SLC1A7,SLC5A5,SLN,SNAI1,SOAT1,SOCS2,SOX4,SPP1,SPRY2,SPRY4,STAU1,SYN1,TAP1,TBP,TERT,TLN2,TMSB10/TMSB4X,TP53,TP73,TRPV1,TSPAN6,TYMS,TYRO3,VCAN,VEGFA,VIM,YAP1 |

| | | | | | | | | |
|---|---|---|---|---|---|---|---|---|
| dihydrotestosterone | chemical - endogenous mammalian | | -0,719 | -0,320 | 0,039 | 4,65E-21 | 1,83E-18 | ABCA1,ABCC4,ABCG1,ACACA,ACKR3,ADAMTS1,ADIPOQ,ADM,ALAS1,ALDH1A3,ALDOA,APOD,APOM,APP,ARFGAP3,AVP,AZGP1,B2M,BAX,BCAP31,BTG2,C1orf116,C1orf21,CANX,CASP1,CASP3,CASP8,CCL16,CCN2,CCNA2,CCNB1,CCT3,CD63,CD74,CDC20,CDKN1B,CGA,CHI3L1,CLU,COL3A1,COL4A1,COL5A2,COL5A3,CSNK2A1,CST6,CST8,CYBB,CYP4A11,DBI,DCTPP1,DHCR24,DPP4,DTL,DYNC1LI2,DYNLL1,EGF,EGR1,ELAVL1,ELN,ENO1,ESR1,FN1,FOS,FOXO1,FOXP3,FRK,FZD6,G0S2,GAS6,GHR,GLUL,GNMT,GPD1,GPI,GPX5,GRB10,GRN,GSTP1,GTF2I,HLA-A,HPGD,HSD11B2,HSD3B2,IGFBP4,IL17RB,INSR,KLK1,KLK3,KLK7,KRT19,LDLR,LEP,LGALS1,LHB,LPL,LYZ,MAFF,MCAM,MCM5,ME1,MGLL,MME,MMP1,MMP7,MUC1,MYOD1,NDRG1,NEDD4L,NFKBIA,NNAT,NOS1,NOS2,NR4A1,ODC1,PAM,PCK1,PDCD4,PDYN,PEA15,PER1,PER2,PFN1,PHF21A,PIP,POSTN,PPARA,PPARG,PROS1,RAB27A,RAD50,RARG,RB1,RHOB,RPLP2,SELENBP1,SERPINE2,SGK1,SHC1,SLC16A7,SLC19A2,SLC2A4,SLC2A5,SLC7A7,SNAI1,SPP1,SRD5A2,STARD8,STAT5A,TALDO1,TERT,TFPI,TGFBR3,TKT,TMPRSS2,TNFSF10,TP53,VCAM1,VCAN,VEGFA,VIM,VLDLR,WIF1,WNT7B,YTHDC1 |
| ERBB2 | kinase | Activated | 2,149 | 2,596 | 0,041 | 7,74E-20 | 2,66E-17 | ABCG2,ABL1,AGR2,AIMP2,ALAD,ALAS1,ALDOC,ANGPTL2,ANXA2,AURKA,BAG2,BCAT2,BHLHE40,BMP7,BTG2,CAPZA2,CAVIN2,CAVIN3,CCL20,CCN2,CCN5,CCNA2,CCNB1,CCNB2,CD47,CD59,CDC20,CDC23,CDC25B,CDC7,CDH3,CDK1,CDKN1B,CDKN3,CIDEC,CKS2,CLDN3,CLU,CMAS,COL18A1,COL1A1,COL3A1,COL4A1,COL5A2,COL6A3,CSE1L,CST8,CUL1,DDIT3,DERL1,DNAJB6,DPEP1,DPT,DSG2,EGR1,ELK1,ELK3,ELL2,EMP3,ESPL1,ESR1,FEZ2,FN1,FOS,FOXO1,FSTL1,FXYD3,GALNT3,GAS6,GHR,GINS1,GNAI1,GSPT2,HBEGF,HEYL,HIF1A,HMGB2,HSPB8,HTRA1,IDI1,IGFBP4,IL25,IL6R,ITGA6,ITGB3,JAK1,JUNB,KCNN4,KDM5B,KLK3,KRT81,LAMB1,LAMP1,LCN2,LGALS1,LPCAT1,LRRFIP1,LSR,LTBP2,LUM,LXN,MACF1,MAP2K3,MAPK13,MCM2,MCM3,MCM5,MITF,MME,MMP1,MMP7,MUC1,MXI1,MYBL2,NDC80,NDRG1,NET1,NFKB1,NNMT,NOS2,NOTCH4,NRP1,NUCB2,ORC2,PCK2,PCNA,PDCD4,PDHA1,PDIA4,PDK4,PHYH,POLD3,POLE2,POLR2E,POLR2G,POLR2L,PPARG,PRC1,PRL,PROM1,PRSS3,PSEN1,PSMB9,PTEN,PTPRB,PTPRK,RALB,RARG,RFC2,RFC4,RFC5,RHOB,RPA1,RPA2,RRM1,RRM2,S100A4,S100A6,SEC61A1,SERPINA3,SERPINI1,SLC1A6,SLPI,SMAD2,SNAI1,SOCS2,SOWAHC,SOX4,SPDEF,SPINK4,SPINT1,SPINT2,SQOR,ST3GAL4,ST3GAL6,STAT3,TAP1,TAPBP,TCEAL9,TERT,TGFBI,TGM1,THBS4,TMEM43,TNC,TNS4,TP53,TPD52L1,TPM1,TUBA1A,TUBB4A,TYMS,UBE2C,USP14,VCAN,VDR,VEGFA,VIM,WT1,WWC1,ZWINT |
| butyric acid | chemical - endogenous mammalian | | -0,831 | -0,760 | 0,008 | 8,34E-20 | 2,75E-17 | A2M,ADCY1,ADGRG1,ADH6,ALAD,ALB,ALDH1A1,ALOX15B,ALOX5,ALPG,ANXA1,ANXA2,ANXA5,BAG1,BAX,CALM1 (includes others),CAP1,CASP3,CCL20,CCNA2,CCNB1,CCNB2,CCNC,CD44,CD81,CD83,CDC20,CDK1,CDK4,CDKN1B,CDKN1C,CGA,CLCN3,CLDN1,CLU,COL4A1,COL4A2,COL5A2,COL6A3,CUL1,CYP2B6,CYP4A11,DDR1,DEK,DHCR24,DPP6,DTYMK,DUSP1,E2F4,EIF4G2,ELK3,EMP3,ERCC2,ESR1,FOS,FSTL1,FURIN,G0S2,G6PC,GADD45A,GATA6,GBP2,GLI3,GRN,GSTA1,GSTP1,H1-2,HBB,HK1,HLA-DRA,HLA-DRB1,HMGB2,IFI16,IFI30,IFIT3,IFITM1,IFITM2,IFNAR2,IFNGR2,IL32,INPP5J,IQGAP1,KLK3,KRAS,LEP,MAPK13,MMP7,MPP3,MUC1,MVP,MYCL,NAMPT,NEU1,NEU3,NFATC4,NFKB1,NFKBIA,NKX2-5,NME1,NOS2,NR4A1,NRAS,NRP1,PCK1,PCNA,PDIA3,PDK2,PDYN,PLCB2,PLSCR1,POLR2L,PPARG,PRKCD,PTEN,PTGDS,RB1,RHOA,RXRA,SEC61B,SGK1,SLC1A2,SLC26A2,SLC5A5,SPP1,STAT3,TGFBI,TGM1,THBS4,TIMP1,TP53,TP53BP2,TP53I3,TRPV1,TYMS,VEGFA,WEE1,WT1 |

| | | | | | | | | |
|---|---|---|---|---|---|---|---|---|
| progesterone | | chemical - endogenous mammalian | -1,134 | -0,904 | 0,020 | 1,31E-19 | 4,00E-17 | ABCG2,ACAT1,ADAMTS1,ADIPOQ,ADM,ALDH1A1,ALPL,ANKH,AP1G1,APOD,ATF3,AURKA,B3GALNT1,BAX,BHLHE40,C3,CA2,CASP3,CCL25,CCN5,CCNA2,CCNB1,CCNB2,CDKN1B,CFB,CFD,CIDEC,CITED2,CLCA1,CLDN3,COL15A1,COMT,CRYAB,CTSF,CTSV,CYP26A1,CYP2A6 (includes others),CYP2B6,CYP2C8,CYP3A4,CYP3A5,DDIT3,DPP4,DRD2,EGF,EGR1,EIF5B,ELL2,ESPL1,ESR1,F2RL1,F3,FAAH,FKBP3,FN1,FOS,FOXO1,FOXP3,FRAT2,GABARAPL1,GAD1,GARS1,GAS6,GCH1,GHR,GNMT,GRB10,GRIA3,GRINA,GSTA3,HBEGF,HGFAC,HIF1A,HLA-DQB1,HLA-G,HOPX,HOXA10,HSD11B2,HSD17B4,IHH,IL13,IL15,IL6R,INHA,INSR,KLF9,KLK1,KLK3,LCN2,LEFTY2,LEPR,LEPROT,LGALS1,LHB,LPL,LTF,LXN,MAD2L1,MAP2K3,MCM2,MCM3,MCM5,ME1,MME,MMP1,MMP26,MMP7,MPZ,MUC1,MYBL2,NDRG1,NET1,NFIL3,NFKBIA,NOS2,NR4A1,NRP1,NSUN5,NT5C2,NTF3,P2RX4,PCDH1,PCK1,PCNA,PDHA1,PDLIM1,PER2,PHF21A,PIGR,PIP5K1B,PKM,PLK2,POSTN,PPARG,PRL,PROS1,PTGDS,PTGER3,RAP2C,RBP1,RRM2,S100A6,SCNN1B,SFN,SGK1,SMAD2,SOD2,SPP1,SSTR3,ST3GAL4,STAT1,STAT3,STAT5A,STEAP1,TBX2,TERT,TGFBI,TGFBR3,TGM1,TIMP1,TIPARP,TNFSF10,TNS2,TP53,UGCG,VDR,VEGFA,VIM,WFDC2,WNT6,WNT7B,XDH |
| EGF | -1,005 | growth factor | 0,341 | 0,877 | 0,049 | 2,43E-19 | 7,18E-17 | ABCG2,ABI1,ACTN1,AFP,ALOX12,ANPEP,APOA1,ARC,ATF3,AURKA,B4GALT5,BET1,BIRC2,BTG2,CASP1,CASP3,CAVIN2,CCL17,CCL20,CCN2,CCNB1,CD44,CDK1,CDK4,CDKN1B,CDX2,CEBPE,CGA,CISH,CKS2,CLCN3,CLDN3,CLU,COL1A1,COL1A2,COL3A1,CTCF,CTSD,CXCL6,CYP2B6,DAD1,DDIT3,DIO3,DNAI1,DPP4,DUSP1,EGR1,EGR3,ELK1,ENAH,ESR1,F3,FN1,FOS,FOSB,FOSL1,FSTL1,GADD45A,GAST,GDF2,GFPT1,GSTP1,HBEGF,HIF1A,HMOX1,HNF4A,HPGD,HRAS,HSD3B2,IDI1,IER2,IGFALS,IGFBP4,ITGB3,JUNB,JUND,KLK3,KRAS,KRT19,LCN2,LPCAT3,LSM1,LTF,MAD2L1,MAPK1,MB,MET,MMP1,MUC1,MUC5B,MYOG,NES,NGF,NOS2,NPPA,NR4A1,NR4A3,NRP1,ODC1,PBK,PCNA,PDCD4,PDK4,PER1,PFKFB1,PLSCR1,PPARG,PPIB,PRL,PROM1,PTPN12,PTTG1,RAB5A,RAC1,RHOA,RHOB,RRM1,RRM2,RXRA,S100A10,S100A11,S100A4,SERPINA3,SH2B3,SHC1,SLC1A2,SLC2A4,SMAD1,SNAI1,SOX4,SPP1,SPRY2,SPRY4,ST3GAL4,STAT1,TBP,TERT,TFPI2,TGM1,TIMP1,TM4SF5,TNC,TP53,ULK1,VAMP2,VCAM1,VCAN,VDR,VEGFA,VIM,ZFP36 |
| IL4 | | cytokine | 0,502 | 1,072 | 0,042 | 4,39E-19 | 1,25E-16 | ABCA1,ABLIM1,ACTG1,AHR,ALAS1,ALDH2,ALDOC,ALOX12,ALOX15B,ALOX5,ALOX5AP,ANKRD55,ANXA1,ANXA2,ANXA4,APOLD1,ARMC6,ASCC3,ASS1,ATP1A3,BANF1,BAX,BCAT2,BHLHE40,BIRC2,BNIP3L,BPIFA1,C3,CA2,CAND1,CAPG,CASP3,CASP6,CAT,CBFB,CBX3,CCL1,CCL17,CCL20,CCL22,CCL25,CCN2,CCR4,CCT3,CD163,CD1A,CD1B,CD33,CD44,CD5,CD58,CD74,CD83,CDK1,CDKN1B,CISH,CITED2,CKAP4,CLCN3,CNR2,COL18A1,COL1A1,COL1A2,COL6A3,CRLF2,CSF3,CTNNA1,CXCL6,CYB5A,CYBB,DCSTAMP,DCT,DPP4,EBNA1BP2,EFL1,EPHX1,EXT1,F2RL1,F3,FAAH,FAS,FCER1G,FN1,FOS,FOSB,FOXP3,FUT7,FYCO1,GADD45G,GATM,GBP2,GZMA,HBEGF,HCLS1,HDAC1,HLA-DMA,HLA-DMB,HPGD,HSD3B2,IER2,IFI16,IFI30,IFI44,IFIH1,IFIT3,IFNAR2,IFNGR2,IGFBP4,IL10RA,IL13,IL13RA1,IL15,IL17RB,IL18,IPO4,IRF7,IRF8,IRF9,ISG15,ITGAE,ITGAV,ITGB2,ITGB3,ITPA,JAK1,JUNB,JUND,KIF3A,KITLG,KLK3,KLK7,KRT10,LGALS1,LGALS3BP,LMAN2,LPL,LTA,LTB,LY75,MAPK1,MCM5,MMP1,MSH2,MUC1,MYOD1,MYOF,MYOG,NAB2,NDRG1,NFIL3,NFKB1,NFKBIA,NOCT,NOS2,NR4A3,NRP1,NUP155,PDCD1,PDE1B,PFKP,PIGR,PIM1,PLA2G4C,PLBD1,PLOD2,PLSCR1,PMS1,PNP,POLE2,POSTN,PPARG,PREPL,PRKCA,PRKCD,PROS1,PSMA6,PSMB10,PSMB8,PSME1,RAB5A,RAD50,RHOA,RIDA,RIN1,S100A10,SAMSN1,SDF2L1,SEMA4A,SEMA4D,SOCS2,SOX13,SPHK2,SPINK5,SPINT2,SPP1,SQSTM1,ST14,STAT1,STAT5A,STK39,STUB1,SWAP70,SYNGR2,SYT11,TAPBP,TBX21,TES,TFAP4,TGFBI,TIMP1,TLE1,TLN2,TLR2,TMED5,TMEM131,TNC,TNFRSF11B,TP53,TPD52L2,TXLNA,TYRO3,UBR4,UCHL3,USO1,USP2,VCAM1,VDR,VEGFA,VIM,VPS13C,VPS35,VTCN1,XPO7,XPOT,XRCC5,ZBP1,ZFP36,ZNF195,ZZZ3 |

| | | | | | | | | |
|---|---|---|---|---|---|---|---|---|
| IGF1 | -0,305 | growth factor | -0,899 | -0,479 | 0,038 | 4,81E-19 | 1,29E-16 | A2M,ABCA1,ACACA,ACTR1B,ADIPOQ,ADM,AKR1B1,AKR1C1/AKR1C2,ANPEP,APP,BAX,BHLHE40,BTG2,C1QA,CACNA1S,CASP3,CAVIN2,CCN2,CCNA2,CCNB1,CD44,CD70,CDK1,CDK4,CDKN1B,CISH,CITED2,CLU,COL1A1,COL3A1,CSHL1,CTSD,CXCL6,CYBA,CYBB,CYFIP2,CYP27B1,DDIT3,DDX50,DUSP1,EGF,EGR1,ELN,ESR1,F3,FJX1,FN1,FOS,FOSB,FOXC2,FOXO1,GADD45A,GADD45G,GCK,GHR,GHRH,GPD1,HBB,HIF1A,HMOX1,HSD11B2,HSPA4,HSPA5,IER2,IFNGR2,IGFBP4,IHH,IL18,INHA,INSR,ITGAV,JUNB,KAT2A,KIF1A,KLK6,LAMP1,LASP1,LCN2,LEP,LEPR,LPL,MAPK1,MAPT,MMP1,MPZ,MUC5B,MXI1,MYOD1,MYOG,NCF2,NFE2L2,NFKBIA,NOS2,NPPA,NR4A1,ODC1,OSMR,PARP1,PBK,PCK2,PCNA,PDYN,PIM1,PITX3,PKM,PLK2,PPARG,PRL,PSMB8,PSMB9,PTEN,PTPRC,PYY,SEC23A,SFN,SGK1,SHBG,SIRT2,SLC12A6,SLC1A6,SLC2A4,SLC5A5,SLPI,SNCG,SPP1,SST,SYN1,TAP1,TBX2,TGFBI,TNFRSF11B,TP53,TRPV1,TTF2,UBE2D2,UGCG,VCAM1,VEGFA,VIM,WEE1,WT1,ZFP36 |
| CD3 | | complex | 1,239 | 1,902 | 0,058 | 1,63E-17 | 3,95E-15 | ACTR3,AHR,AIMP1,ALAS1,ANXA1,APBA1,ARHGDIB,BECN1,C1R,C2,CALM1 (includes others),CAMK2A,CASP3,CASP6,CASP8,CCL1,CCL17,CCL20,CCL22,CCR4,CD44,CD5,CD53,CD74,CD83,CDK1,CDK2AP1,CDK4,CDK7,CDKN1B,CISH,CKS2,CLCN1,CNIH4,COPB2,CRLF2,CTLA4,CYBA,DAD1,DPP4,DUSP2,DUSP22,E2F4,EGR3,EIF4A3,ELAVL3,ELK3,EXOSC8,FAS,FCER1G,FKBP1A,FKBP3,FOS,FOXP3,FUT7,GBP1,GLI1,GNAS,GNL2,GTF2IRD1,GTPBP4,GUCY1B1,GZMA,HAT1,HAX1,HCCS,HIF1A,HLA-DQB1,HNRNPF,HNRNPR,HPX,HSP90B1,HSPA5,IFI35,IFITM1,IFNGR1,IL10RB,IL13,IL32,ILF2,IRF8,IRF9,JAK1,JUNB,KATNA1,KCNA5,KCNN4,KDELR2,KLK7,LAMP1,LTA,LYN,MACROH2A1,MAP3K10,MAPK1,MCM5,MTHFD2,NAMPT,NFE2L2,NFKB1,NFKBIA,NME1,NOS1,NOS2,NR4A1,ODC1,P4HB,PAFAH1B3,PCBP2,PCNA,PDCD1,PDCD5,PDIA4,PFN1,PKLR,PLA2R1,PNP,PPA1,PPARG,PRDX4,PSMA5,PSMA6,PTEN,PTP4A2,PTPRC,RAB8A,RAC2,RAD23B,RARS1,RB1,RHOC,RORA,RORC,RXRA,RYK,S100A11,SEC61B,SH2D2A,SHC1,SLPI,SMPDL3B,SNRPB,SOCS2,SPINK4,SSB,STAT1,STMN1,SYNPO,TBX21,TFPI2,TLR2,TMPO,TNFSF10,TP73,TPST1,TRA2A,TRIM24,TRIM5,TSPAN7,TSTA3,TXNRD1,VBP1,VEGFA,VPS4B,WEE1,YARS1,ZBTB32,ZFP36,ZNF140,ZN[illegible] |
| RXRA | -0,316 | ligand-dependent nuclear receptor | -1,619 | -1,503 | 0,016 | 6,06E-17 | 1,38E-14 | ABCA1,ABCC3,ABCG1,ABCG2,ACACA,ACACB,AHCY,AKR1C1/AKR1C2,ALDH1A1,APOA1,APOC3,APOM,APP,ARL4C,ASNS,BAAT,C1QA,C3,CAT,CCL17,CCL20,CCL22,CCNA2,CCNB1,CEBPE,COL1A1,CRABP2,CRYAB,CSHL1,CTSS,CYP24A1,CYP26A1,CYP27B1,CYP2B6,CYP3A4,CYP3A5,CYP4A11,DR1,DRD2,EPHX1,FABP1,FABP2,FABP6,FLRT3,FOS,FSTL1,GAS6,GCK,GPD1,HBEGF,HIF1A,HMOX1,HNF1B,HNF4A,HOXB1,HSD17B4,IL13,KLF9,LCN2,LPL,LTF,LYZ,MLXIPL,MYL7,MYOG,NME1,NPC1L1,NPPA,NR0B2,NRGN,PCK1,PCK2,PKLR,PLTP,PPARA,PPARG,PRL,PTGDS,RARG,RBP1,RGR,RLBP1,RORA,RORC,RXRA,S100A4,SEMA4D,SLC10A1,SLC10A2,SLC2A4,SLC34A1,SPP1,STRA6,TBXA2R,VCAN,VDR,VEGFA,WEE1 |

| | | | | | | | | |
|---|---|---|---|---|---|---|---|---|
| NFkB (complex) | | complex | 0,164 | 0,896 | 0,068 | 6,20E-17 | 1,38E-14 | A2M,ABCG1,ACKR3,ADM,AHR,ALDH1A3,ALDH7A1,AMPH,APP,AQP5,ATF3,BAG2,BAX,BECN1,BID,BIRC2,BLVRA,BPIFA1,C1R,C3,CAPNS1,CASP4,CASP7,CASP8,CAV3,CCL1,CCL17,CCL20,CCL22,CCN2,CCNB1,CCNB2,CD44,CD74,CD83,CD99,CDC25B,CDX1,CDX2,CFB,CLU,COL18A1,COL1A2,CSF3,CSNK2A1,CTCF,CXCL6,CYBB,CYP3A4,CYP3A5,DAD1,DDIT3,DIO1,DMBT1,DRD2,DUSP2,EGR1,ELK1,ELL2,EPCAM,ERAP1,F2RL1,F3,FABP6,FAS,FGF1,FGF5,FN1,FOS,FOXF1,FOXP3,G0S2,GAD1,GADD45A,GADD45B,GAS6,GAST,GBP2,GCH1,GLA,GNAS,HDAC6,HGFAC,HIF1A,HLA-A,HLA-DMB,HLA-F,HMOX1,HNF4A,IFNAR2,IL10RA,IL13,IL15,IL18,IL32,IRF7,ISG15,ITGAV,ITGB8,ITPKB,JUNB,KLF3,KRT19,KYNU,LCN2,LTA,LTB,MFHAS1,MMP1,MSH2,MUC5B,NAMPT,NCF2,NFKB1,NFKBIA,NFKBIE,NGF,NOS1,NOS2,NPPA,NUP62,ODC1,PEA15,PECAM1,PKP1,PLK2,PLK3,PPARA,PPARG,PPIF,PPM1D,PRKCD,PSMB9,RGS16,RRAS,SCFD1,SENP2,SERPINA3,SLC1A2,SLC22A4,SLC2A4,SLC2A5,SLC37A4,SLIT2,SLPI,SOAT1,SOD2,SPP1,STAT5A,STX4,TAP1,TCF12,TERF2IP,TERT,TFPI2,TIMP1,TLR2,TNFAIP8,TNFRSF11B,TNFSF10,TP53,TRIM38,UBD,VCAM1,VEGFA,VIM,XRCC5,ZFP36 |
| CTNNB1 | -0,034 | transcription regulator | -1,715 | -1,193 | 0,058 | 6,93E-17 | 1,51E-14 | ABCC1,ACACA,ACAP1,ACAT1,ACTR3,ACTR8,ADIPOQ,AFP,AGRN,AHR,AKAP13,ALDH1A1,ALDH1A3,ALDH3A2,ANXA1,APOD,APP,AQP5,ARFGAP3,ATOH1,AURKA,BCAP31,BECN1,BHLHE40,BMP7,BTF3,CALM1 (includes others),CAPN1,CAPNS1,CASP7,CCN2,CCNA2,CD44,CD47,CD99,CDK7,CDKN1B,CDX1,CDX2,CENPM,CER1,CFD,CIDEC,CLU,CNTFR,COL1A1,COL4A1,COL4A2,CRABP2,CRAT,CSF3,CTNNA1,CTSF,CTSZ,CXCL6,CXXC4,CYB5A,CYFIP2,CYP24A1,CYP2A6 (includes others),CYP4F12,CYP7B1,DCT,DDIT3,DDX50,DIO1,DPEP1,EGR1,ENO3,EPCAM,EPHB3,EVX1,FAS,FBRS,FCER1G,FGF16,FGF5,FGFBP1,FN1,FOS,FOSL1,FOXC2,GAD1,GADD45B,GADD45G,GAST,GCH1,GDF9,GHR,GLI1,GLI3,GLUL,GRAMD4,HNF1A,HNF4A,HOXB7,HTATIP2,HTRA1,IFITM1,IHH,IL13,IL7,IMPDH2,INHA,IRF8,ITGA6,ITGB2,KITLG,KLK3,LAMB1,LHB,LIMD2,LIPA,LMX1B,LPL,MAN2B1,ME1,MITF,MME,MMP1,MMP7,MPZ,MPZL2,MUC6,MYOD1,MYOG,NES,NEUROG1,NKX2-5,NOS2,NPHS2,NPTX1,NR4A1,NRCAM,OAT,PARD3,PCNA,PDYN,PECAM1,PFN1,PIP5K1B,PLS3,POLR2E,PPARG,PRDM14,PRRC2A,PRX,PTCH2,PTCRA,PTEN,QPCT,RAB3A,RAI14,S100A4,SCRN1,SEC61A1,SERPINA3,SERPINE2,SFN,SGK1,SLC12A2,SLC1A2,SLC26A2,SNAI1,SNRNP70,SORBS3,SOX4,SPP1,SPRY4,SQSTM1,STAT3,STAT5A,TBX21,TCF12,TERT,TGM1,TIA1,TIMP1,TNC,TNFRSF11B,TNFSF10,TNNC1,TP53,TP73,TSPAN8,UGCG,UPB1,USO1,USP46,VCAM1,VCAN,VEGFA,VIM,WIF1,WNT11,WNT2,WNT6,WNT7B,[illegible] |
| STAT3 | 0,283 | transcription regulator | -0,340 | 0,048 | 0,039 | 4,56E-16 | 9,41E-14 | A2M,ADIPOQ,ADM,AHR,ALDH1A1,ALDH1A3,ATOH1,BECN1,BNIP3L,CA9,CASP1,CASP3,CASP7,CAT,CCL17,CCL20,CCN2,CCNB1,CCNC,CD74,CD83,CDK1,CDKN1B,CFB,CHI3L1,CLIC5,COL1A1,COL1A2,COL3A1,CSF3,CTF1,CTLA4,CXCL6,CYP26A1,DDIT3,DLEC1,DMBT1,DNMT1,DPP4,ECT2,EED,EGR1,EGR3,ESR1,F3,FAS,FCER1G,FERMT2,FGL2,FLRT3,FN1,FOS,FOXP3,FUT4,G6PC,GADD45A,GADD45G,GBP2,GPR65,HBB,HERC6,HIF1A,HLA-A,HLA-DMA,HMOX1,HNF4A,HOXA10,IFI16,IFI30,IFI35,IFI44,IFIH1,IFIT2,IFIT3,IFIT5,IFITM1,IFITM2,IHH,IL13,IL6R,IL7,IRF7,IRF9,ISG15,ITGAV,ITGB2,ITGB6,JUNB,KLK3,LEP,LEPR,LTA,LTF,LYZ,MITF,MMP7,MRAS,MUC1,MX1,MXI1,MYOD1,MYOG,NAMPT,NDRG1,NES,NFKB1,NOS2,NOX5,NPPA,NRP1,OAS1,PCK1,PDCD1,PDIA4,PECAM1,PIM1,PLSCR1,PROM1,PSIP1,PSMB8,PSMB9,PTK2,PTTG1,RAB27A,REG1A,RORA,RORC,SERPINA3,SERPINB1,SERPINE2,SGK1,SNAI1,SOD2,SP110,STAT1,STAT3,STC2,SYNPO,TAP1,TBX21,TERT,TFPI2,TIMP1,TLR3,TNFSF10,TNS1,TNS4,TP53,TRIM22,TRIM5,TRPM3,UBD,VCAN,VEGFA,VIM,WARS1,ZFP36 |

| | | | | | | | | | |
|---|---|---|---|---|---|---|---|---|---|
| FOS | -1,524 | transcription regulator | | 0,201 | 0,380 | 0,021 | 6,05E-16 | 1,22E-13 | A2M,ABCC1,ACACA,ACTG1,ADD2,ADNP,AGPS,ALAS1,ALDH1A3,ANKRD2,ANXA4,ARF1,ARPC5,ATF3,ATF7IP,CA2,CALU,CAPN2,CASP3,CAST,CAT,CCL20,CCN2,CCNA2,CD44,CGA,CHI3L1,CLU,COL18A1,COL1A1,CRABP1,CRABP2,CSF3,CSTA,CTH,CTSV,DAPK2,DNMT1,DSG3,EGR1,EIF4G2,ELOVL6,ENAH,ENO1,ESM1,EVX1,F3,FBN1,FEZ2,FN1,FOS,FOSB,FOSL1,FSTL1,FZD5,FZD6,G6PC,GM2A,GSTP1,HDAC6,HLA-B,HMOX1,HPGD,HSD3B2,HSP90B1,HSPA5,HSPB8,HTATIP2,ITGA6,IVL,JUNB,KCNN4,KIF2A,KITLG,LGALS3BP,LHB,LPCAT3,LPL,LTBP2,MAP4K4,MCM5,MET,MMP1,MMP7,MPZL2,NAA80,NAE1,NCF2,NET1,NFE2L2,NFIL3,NGF,NOTCH4,NPPA,NPTX1,NR4A1,NUBP1,ODC1,PAOX,PFN1,PIP5K1B,PLK2,PRL,PTGDS,PTP4A2,PTPN12,PTPRO,PTS,PTTG1,RALB,RARG,RBBP7,RBP1,RXRA,S100A10,SEC23B,SELENBP1,SEMA3B,SEMA4D,SEMA6D,SERPINE2,SLC10A2,SLPI,SLURP1,SNAI1,SNAP23,SNCG,SNN,SPP1,STMN1,SULT2B1,SWAP70,TGFBI,TIMP1,TIPARP,TNFRSF11B,TNMD,TP53,TUBB,UGCG,VCAM1,VEGFA,VIM,WEE1,WNT10B,WNT11,WNT6,WNT7B,XDH |
| PDGF BB | | complex | Inhibited | -2,617 | -2,186 | 0,048 | 2,02E-15 | 3,78E-13 | ACKR3,ADM,AKAP12,ANXA11,ATF3,BHLHE40,BMP3,C3,CALM1 (includes others),CASP4,CAV2,CCL20,CCN2,CCNL1,CCR4,CD44,CDK1,CDK4,CDKN1B,COL18A1,COL3A1,CRYAB,CSF3,CSRP2,CTH,DDIT3,DUSP1,EGR1,EGR3,EIF4E2,EIF4G1,ELAVL1,F3,FHL1,FHL2,FN1,FOS,FOSB,FOSL1,FOXC2,FZD1,GADD45A,GBP2,GHR,GLUL,GUCY1B1,HBEGF,HLA-E,HMOX1,HRAS,IER2,IGFBP4,JUNB,KRAS,LAMB2,LDLR,MMP1,MYBL2,NAMPT,NFIL3,NOS1,NOS2,NR4A1,NR4A3,ODC1,PCNA,PDLIM1,PIM1,PLK2,POSTN,PPARG,PPP1R15A,PRRX2,QARS1,RACK1,RBP1,RFC5,RGS2,RHOB,RPN2,S1PR2,SERPINA3,SGK1,SLC6A4,SLC7A7,SNAI1,SOD2,STAT3,TAF9,TEAD4,TIMM17A,TIMP1,TNC,TOB1,TP53,TRA2B,TRIOBP,VCAM1,VCAN,VEGFA,XDH,ZFP36 |
| RELA | | transcription regulator | | 1,148 | 1,626 | 0,053 | 2,79E-15 | 5,11E-13 | A2M,ABCG2,AFP,AHR,ALOX5AP,ANKRD2,APP,ARFGAP3,ARHGDIB,B2M,BAX,BECN1,BHLHE40,BIRC2,BLVRA,BNIP3L,BTG2,C3,CASP3,CASP8,CCL20,CCL22,CCN2,CD44,CD59,CDKN1B,CERT1,CFB,CHI3L1,CITED2,COL1A1,COL1A2,CRYAB,CTCF,CXCL6,CYBB,CYP2B6,CYP3A4,DDIT3,DUSP1,EGR1,ELN,EPCAM,ERAP1,F3,FABP6,FAS,FCGRT,FN1,FOS,FOSB,FOXP3,GAST,GBP1,GCH1,GSTA1,HIF1A,HLA-B,HMOX1,HNF4A,HSP90B1,IER2,IFNGR2,IL13,IL32,IRF7,ISG15,JUNB,LTA,LTB,LUM,LYN,MMP1,MST1R,MUC1,MYOD1,NAMPT,NFE2L2,NFKB1,NFKBIA,NFKBIE,NMI,NOS2,NOX5,NR4A1,NR4A3,PCK1,PECAM1,PKM,PLK3,PPARG,PPM1D,PRKCD,PSMB10,PSMB9,PTEN,PTGDS,RORC,S100A6,SERPINE2,SHBG,SLC1A2,SLC2A4,SLC2A5,SNAI1,SOD2,STAT5A,TAP1,TAPBP,TBX21,TERF2IP,TERT,TFPI2,THOC1,TLR2,TNFRSF11B,TP53,TP73,UBD,VCAM1,VEGFA,VIM,WT1 |
| CDKN1A | -0,252 | kinase | | -1,814 | -1,953 | -0,024 | 1,59E-14 | 2,73E-12 | AIMP1,APP,ARL6IP1,AURKA,BAX,BIRC2,BUB3,CASP3,CCN2,CCNA2,CCNB1,CDC20,CDC25B,CDK1,CDK4,CDKN1B,CDKN3,CIT,COL1A2,CRYAB,DLGAP5,DSE,DTL,DUSP1,FAS,FN1,GLB1,GRN,HMGB2,HSPA5,IL32,ITGAV,ITGB3,KIF20A,LBR,LGALS3BP,LUC7L3,MAD2L1,MCM2,MCM3,MMP1,MYBL2,MYOD1,P4HA1,PBK,PCLAF,PCNA,PELO,PLK2,PRC1,PTPRC,RB1,RFC4,RPA3,RRM1,SHC1,SLC30A5,SMC4,SOD2,STMN1,SUV39H1,TERT,TGM1,TP53,TP53I3,TRIM44,TUBB4A,TUBG2,TYMS,UBE2C,VEGFA,VGLL4,VIM,YTHDC1,ZBED5,ZNF84 |

| | | | | | | | | | |
|---|---|---|---|---|---|---|---|---|---|
| SP1 | 0,064 | transcription regulator | | -0,035 | 0,541 | 0,069 | 1,83E-14 | 2,97E-12 | ABCA1,ABCC8,ADAMTS1,AKAP12,ALDH3A2,ALOX12,ALOX15B,ALOX5,ALPL,ANKH,APOA1,APOC3,ASNS,ASS1,ATF3,B4GALT5,BAG2,BAX,BMP7,BNIP3L,CASP3,CAT,CCL20,CCL22,CCN2,CCNA2,CD59,CD99,CDC25B,CDK1,CDK4,CDKN1B,CHI3L1,CHRNB4,CITED2,COL1A1,COL1A2,CRABP2,CRYAB,CRYBB1,CRYBB2,CRYBB3,CSNK2A1,CTH,CTSD,CYP26A1,CYP2B6,CYP7B1,DHCR24,DRD2,EGR1,ESR1,ESRRA,F3,FABP6,FAS,FDX1,FN1,FOS,FOSL1,FOXP3,GHR,GJD2,GNAS,HBB,HBEGF,HDAC1,HIF1A,HMOX1,HNF4A,HSD11B2,HSPA5,IFNGR1,IGFBP4,IL15,INSR,ITGAV,ITGB3,KRT19,KRT4,KRT81,LDLR,LHB,LIPA,LPL,LTB,MET,MMP1,MSH6,MST1R,MUC5B,MYBL2,NAB2,NCF2,NDRG1,NES,NFKB1,NFKBIA,NOS1,NOX5,NPPA,PARP1,PCK1,PDHX,PIM1,PKM,PRKCA,PRKCD,PROM1,PSIP1,PTCRA,PTTG1,PYY,RB1,ROBO4,RRAS,SERPINE2,SGK1,SLC22A4,SNCG,SOAT1,SOD2,SPP1,SPTB,STAT1,STAT3,TBXA2R,TERT,TFPI2,TGFBR3,TGM1,THRB,TIMP1,TLR2,TNC,TNFSF10,TNNC1,TP53,TP73,TSPO,TUG1,TXNRD1,TYMS,UFD1,UGCG,VDR,VEGFA,VIM |
| D-glucose | | chemical - endogenous mammalian | | -0,005 | 0,272 | 0,024 | 1,95E-14 | 3,10E-12 | ABCA1,ABCG1,ACAA1,ACACA,ACTN1,ADIPOQ,ADIPOR1,AKR1B1,ALDOA,ALDOB,ALDOC,ANXA1,ANXA3,APOB,APOC3,ARF4,ASNS,ASS1,ATF3,BAX,C3,CANX,CAP1,CAPZA2,CASP1,CASP3,CASQ2,CAT,CCL20,CCN2,CCNB1,CD59,CDK1,CDK4,CDKN1B,CLU,COL1A1,COL3A1,COL4A1,COX6A2,CPD,CTSV,CYBA,CYBB,DBI,DDIT3,DNAJC3,DUSP1,EGR1,ELOVL6,ENO1,FABP2,FAS,FBP1,FN1,FOS,FUT1,G6PC,GABARAPL1,GCGR,GCH1,GCK,GFPT1,GLUL,GPD1,GPI,GRN,HIF1A,HK1,HMOX1,HNF4A,HPRT1,HSPA5,IER2,INS,INSR,ITGA8,JUNB,KDELR2,LAMB1,LAMC1,LDLR,LEP,LPL,LY96,MCAM,MLXIPL,MMP1,MMP17,NAMPT,NEU1,NFE2L2,NOS2,NPHS1,NR4A1,NR4A3,OAT,ODC1,PARP1,PBK,PCK1,PCK2,PCNA,PCSK1N,PDCD4,PDHX,PDIA3,PDIA4,PDIA6,PDYN,PFKP,PKLR,PKM,PLA2G6,PLIN3,PLK3,PNPLA3,PODXL,PON2,PPARA,PPARG,PPIB,PTGDS,PYCR1,RDH16,RGS2,RRM2,RTN3,S100A4,SEC61A1,SGK1,SHC1,SIRT2,SIRT3,SLC25A10,SLC2A4,SLC37A4,SLC39A6,SOD2,SPHK2,SPP1,SQSTM1,SSR1,SST,STUB1,TALDO1,TIMM17A,TIMP1,TKT,TLR2,TP53,TRPV4,TRPV5,TUBB4A,TXNRD1,UCP2,USO1,VAMP2,VAMP8,VCAM1,VEGFA,WARS1,WWP1,YARS1 |
| FOXO3 | -0,065 | transcription regulator | Inhibited | -2,592 | -2,496 | 0,012 | 2,79E-14 | 4,35E-12 | AGFG1,AHR,ALDH3A1,ANXA1,ANXA3,AURKA,BAX,BECN1,BNIP3L,C1QA,CASP4,CASP8,CAT,CCN2,CCNB1,CCNB2,CDC20,CDK4,CDKN1B,CDKN1C,CDKN3,CLDN1,COL4A1,COL4A2,CTSV,DDIT3,DLGAP5,DNM1L,EGR1,ESR1,FN1,FOS,FOSB,FOXO1,FOXO4,FOXP3,GABARAPL1,GABRR2,GADD45A,GADD45B,GLUL,GSTM5,GTF2I,GYPA,HIF1A,HTATIP2,IFIH1,IL10RB,IL13,IMPDH2,ITGB2,JAK1,JUNB,LCN2,MCAM,MCM2,MET,MTHFD2,MXI1,MYO6,MYOD1,NAMPT,NCAPG2,NDRG1,NFKBIA,NOS2,OVOL1,PBK,PCNA,PECAM1,PIK3IP1,PLA2G2D,PPP1R15A,PRKCD,PTEN,PTPRB,PTPRC,PTTG1,RASA1,RTN3,RUVBL1,SAC3D1,SGK1,SH2B3,SH3GLB1,SLIT2,SNAI1,SOD2,SPTLC2,TIPIN,TNFSF10,TP53,UBE2C,UCP2,ULK1,VCAM1,VEGFA,VIM,ZWINT |
| TP73 | -0,200 | transcription regulator | | -0,005 | 0,231 | 0,023 | 4,01E-14 | 6,02E-12 | ABCC1,ADAM10,AFP,AQP5,ATF3,ATG5,AURKA,B2M,BAX,BHLHE40,BID,BIRC2,BMP7,BTG2,C1R,C3,CASP1,CASP3,CASP4,CCN5,CCNA2,CCNB1,CD83,CDC20,CDC25B,CDK1,CDKN1B,CDKN1C,CDKN3,CKS2,CLIP3,CNTFR,COL18A1,COL1A1,COL5A2,CRYAB,CTH,CTSD,DDIT3,DHRS3,DIMT1,DRAM1,DTYMK,EGR1,ELOC,ERCC2,FAS,FDXR,FN1,FOXF1,G0S2,GADD45A,GRN,HAGH,HBEGF,HIF1A,HSPB8,HTATIP2,IGFBP4,IGSF3,IL7,IVL,KRT10,LYZ,MDK,MIR22HG,MPZL2,MUTYH,NFKB1,NRGN,NRP1,P2RX4,PERP,PIGF,PIK3R3,PLK2,PPL,PRL,PTEN,PTPN3,RAN,RB1,RBX1,ROBO4,S100A2,SERPINA3,SERPINE2,SERPING1,SFN,SNAI1,SPHK2,SPINT1,SPP1,STMN1,SYNE2,TAP1,TERT,THBS2,TIMP1,TNNI1,TP53,TP53I3,TP73,TRIM22,TRIM32,TSHZ2,TSPAN1,UBE2C,UBE2D1,ULK1,VDR,VEGFA,WARS2,YWHAB,ZMAT3 |

| | | | | | | | | | |
|---|---|---|---|---|---|---|---|---|---|
| SMARCA4 | 0,238 | transcription regulator | | 0,621 | 1,307 | 0,066 | 5,84E-14 | 8,61E-12 | A2M,ABCA1,ADAM15,ADGRG1,ADIPOQ,AFP,AGMAT,AGR2,AHR,ALB,ALDH1A3,ALDH2,ANGPTL2,APOA1,APOBEC3G,ARHGDIB,ARL4C,ASCC1,ASNS,AZGP1,BNIP3L,C1orf105,CA14,CASP1,CASP3,CCN2,CCNA2,CD44,CD47,CD52,CD63,CD74,CDKN1B,COL1A1,CTSS,CYP26A1,CYP3A4,DCT,ECM2,EFNB1,EGR1,ELOVL6,EMP3,ENO3,F3,FABP6,FADS3,FAS,FBP1,FHL2,FN1,FOS,FUT1,GADD45A,GADD45B,GADD45G,GBP1,GJB5,GLI1,GNAI3,GSTP1,GULP1,HBB,HCLS1,HCP5,HEXB,HKDC1,HLA-B,HLA-C,HLA-DRA,HLA-E,HLA-F,HLA-J,HNF4A,HNMT,HPGD,HS3ST1,HYI,IFI16,IFI30,IFITM1,IFITM2,IGFBP4,IL13,IL1RAPL1,ITGA7,ITGAV,KHDRBS3,KIFBP,KLK3,LDB1,LGALS1,LUM,MAFF,MAPK1,MAPK13,MC4R,MC5R,MMP1,MMP7,MUC1,MYBPH,MYOG,NCALD,NR2F2,NRP1,PARVA,PCDH1,PERP,PLS1,PMEL,PPARG,PSG4,PSMB9,PTP4A2,RAB27A,RAB38,RAC2,RAD50,RBM4B,RBP1,RGS2,ROCK1,RRM2,S100A2,S100A6,SEL1L3,SEMA3B,SEMA7A,SERPINE2,SKAP2,SLC2A4,SMARCA4,SOCS2,SPP1,SPRY2,ST6GALNAC4,TAOK3,TAP1,TBX2,TBX21,TLR2,TNFAIP8,TNFSF10,TNNC2,TNNT2,TP53,TPK1,TPM1,TRIM15,TSPAN13,TSPAN8,TUBB,TXNRD1,TYMS,UBD,VIM,WT1 |
| HGF | | growth factor | | -0,093 | 0,460 | 0,055 | 6,54E-14 | 9,48E-12 | A2M,ABCC6,ABCG2,ACKR3,ADAMTS1,ADGRF5,ADGRG1,AFP,AHR,AIMP2,AKAP12,AKAP13,ANGPTL2,ARPC5,ASNS,ATF3,AURKA,BAG2,BIRC2,CA9,CALM1 (includes others),CASP1,CASP10,CASP8,CCN2,CD44,CDC20,CDC25B,CDK1,CDK4,CDKN1B,CDKN3,CHRNB2,CITED2,CNTFR,COL1A1,COL1A2,COL3A1,COL4A1,COPB1,CRYAB,CSE1L,CYP2B6,DDIT3,DTYMK,DYNLL1,EGR1,EPCAM,ESM1,F3,FGF16,FN1,FOS,FOSB,FOSL1,G6PC,GRB10,GUCY1B1,HBEGF,HK1,HMOX1,IGFBP4,IL13,IRF8,ISG15,ITGA6,ITGB8,KITLG,KRT19,LCN2,LDLR,LSM1,LTA,LY75,MAD2L1,MAP2K3,MCM2,MCM5,MELK,MET,MID1,MMP1,MPC1,NAB2,NAMPT,NDC80,NES,NFKB1,NOS2,NR0B2,NR4A1,NR4A3,NRCAM,NRP1,OR7C2,ORC2,OSMR,PCK1,PCNA,PIM1,PLA2R1,PLOD1,PLXNA2,PRC1,PTPRF,PTTG1,RAB5A,RAC1,RAD50,RFC2,RGS2,RHOB,RIN1,SGK1,SLC19A2,SLC1A2,SNAI1,SNRNP70,SOCS2,SOD2,SPP1,SPRY2,ST14,TBXA2R,TIMP1,TP53,UBE2C,VCAM1,VDR,VEGFA,VIM,WT1,YWHAH |
| E2F1 | | transcription regulator | | 0,531 | 0,934 | 0,047 | 7,83E-14 | 1,08E-11 | ABCG2,ACTR1A,ACTR3,ADIPOQ,ALG5,ALPL,AURKA,BAX,BECN1,BID,BUB3,CA2,CALM1 (includes others),CASP3,CASP6,CASP7,CASP8,CBX5,CBX7,CCNA2,CCNB1,CCNB2,CCT2,CD2AP,CD44,CDC20,CDC25B,CDK1,CDK4,CDKN1B,CDKN1C,CITED2,CLIC1,CRABP2,CRIP2,CRYAB,CSE1L,CSHL1,CTNNBIP1,CYB5A,CYP27B1,DDB2,DDIT3,DLEU1,DNMT1,DUSP1,E2F4,ECT2,EED,EGR1,EI24,FAS,FHL1,FKBP3,FOS,FOXO1,GINS1,GM2A,GRAMD4,H2AC8,HMGB2,HNRNPR,HSD11B2,HSP90B1,JPT1,KRAS,KRT10,LEP,LHX3,MAD2L1,MCM2,MCM3,MCM5,MFAP1,MMP1,MSH2,MYBL2,NDC80,NFKBIA,NMI,NRP1,OGA,PCLAF,PCNA,PDCD5,PDK4,PLK2,PPARG,PPP1R26,PPP1R8,PPT2,PSMB9,RAN,RB1,RBFOX2,REV3L,RFC2,RFC4,RFC5,RHOQ,RPA2,RPA3,RRM1,RRM2,RUVBL1,SMARCA5,SMC4,SOD2,STAM,STMN1,TAF9,TAP1,TAPBP,TERT,TGM1,TOP2B,TP53,TP53BP2,TP73,TXNRD1,TYMS,UCP2,ULK1,VCAM1,VEGFA,VIM,WEE1,WWOX,YARS1,YWHAE,ZFP36,ZNF22 |
| PML | | transcription regulator | Activated | 2,164 | 2,205 | 0,005 | 8,82E-14 | 1,19E-11 | ACACA,ACACB,ACTG1,ADIPOQ,ANXA4,APOA1,BAX,CAT,CEBPE,CIAPIN1,CIT,DDX60,DNAJB6,DNAJB9,FABP1,FAS,GADD45A,HBB,HERC6,HLA-DMB,HLA-DRA,HMOX1,IFI35,IFI44,IFIH1,IFIT3,IFITM1,IRF7,ISG15,LDLR,LEP,LIPC,LPL,LYZ,MAFF,MAPK1,MX1,NDRG1,NFE2L2,OAS1,PCBP2,PDK4,PLSCR1,PRKCA,PSMB8,PSMB9,RSU1,RXRA,SNAI1,SQSTM1,STAT1,STMN1,TAP1,TBX2,TP53,TPD52L2,TXNRD1,UBE2M,UCP2,VIM |
| PAX3 | | transcription regulator | | -0,055 | 0,045 | 0,036 | 9,23E-14 | 1,23E-11 | ALDH3A2,ANXA1,APP,ARC,ARHGDIB,ASNS,ASS1,CAPN1,CD151,CDH3,CLU,COL1A2,CTSA,DBN1,DCT,DPEP1,DUSP2,F2RL1,F3,FHL1,FILIP1L,FKBP1A,FUBP1,G0S2,GLB1,GSTM5,GTF2H1,GUCY1A1,HARS1,HLA-DRB1,HPX,HTR6,IFITM2,IL6R,LRRC17,M6PR,MCAM,MEST,MET,MITF,MYOD1,MYOG,NNMT,P2RY4,PAK2,PCNA,PEA15,PPIB,PSME1,PTGER3,PTPN12,PTPN3,PXDN,RHOB,RLBP1,SFN,SOX10,SOX4,STX3,TFPI2,TIA1,TNC,TNNI1,TP53,TSPAN7,TYRO3,VCAN,YARS1,YWHAE |

| | | | | | | | | | |
|---|---|---|---|---|---|---|---|---|---|
| PGR | | ligand-dependent nuclear receptor | | -0,799 | -0,365 | 0,050 | 1,33E-13 | 1,72E-11 | ABCG2,ADAMTS1,AKAP13,ALDH1A3,AP1G1,BIRC2,CAPN2,CCN5,CCNB1,CCNB2,CD44,CD59,CDC20,CDK1,CDKN1B,CDKN1C,CGA,CKAP4,COMT,CTSD,DPYSL2,DSCAM,DYNLL1,EFNA3,EIF5B,ELL2,ESR1,ESRRA,F2RL1,F3,FN1,FOS,FOXO1,G6PC,GAS6,GLUL,GRB10,GRIA3,HBEGF,HIF1A,HPGD,HSD11B2,ITGA6,KLF9,LAMB2,LHB,MAD2L1,MAP2K3,MUC1,MVP,NDRG1,NET1,NFKBIA,NSUN5,NT5C2,OCEL1,P2RX4,PCDH1,PCNA,PDLIM1,PFKP,PHF21A,PIP5K1B,PLK2,POSTN,PPARG,PPIF,PRL,RAP2C,RHOC,S100A6,SGK1,SMPDL3B,SNPH,SPRY2,ST3GAL4,STAT1,STAT3,STAT5A,STEAP1,TGFBI,TIPARP,TNC,TSC22D3,UGCG,VCAN,VDR |
| TRIM24 | 0,412 | transcription regulator | Inhibited | -4,504 | -4,964 | -0,074 | 3,42E-13 | 4,21E-11 | AGRN,BLNK,CA2,CNOT6,CYP24A1,DDX58,DDX60,FAM110B,FBN1,FLRT3,GBP2,GLUL,HERC6,HNF1B,IFI35,IFI44,IFIH1,IFIT2,IFIT3,IFNGR1,IFNGR2,IRF7,IRF9,ISG15,LGALS3BP,MYH10,NMI,OAS1,PARP12,PCLAF,PHF11,PSMB10,PSMB8,PSMB9,RTP4,SOCS2,SPP1,STAT1,TAP1,TGFBI,TLR2,TP53,TRPV5,VEGFA |
| PPARA | -0,177 | ligand-dependent nuclear receptor | | -0,646 | -0,441 | 0,021 | 3,95E-13 | 4,79E-11 | ABCA1,ACAA1,ACACA,ACAT1,ADAM10,ADIPOQ,AGXT,AHCY,ALDH2,ALDH3A2,ALDOB,ANXA2,APOA1,APOC3,APOM,APP,AQP8,ASNS,ASS1,AURKA,BAX,C1QA,C1QB,C1R,C1S,C2,C3,CA9,CAT,CCNA2,CCNB1,CD63,CDK1,CDK4,CFI,CIDEC,CSAD,CTH,CYBA,CYBB,CYP2B6,CYP2C8,CYP4A11,CYP7B1,DBI,DECR2,DIO1,DR1,DYNLL1,ECI2,ELOVL6,F11R,FABP1,FABP2,FBXO21,FOS,FTCD,G0S2,G6PC,GCK,GLUD1,GLUL,GNMT,GPD1,GSTP1,H1-2,HADH,HIF1A,HLA-E,HMOX1,HNF4A,HPD,HPX,HSD17B4,IDI1,KHK,KIF20A,KYAT1,LPCAT3,LPL,MAD2L1,MASP2,MGLL,MGST3,MMD,MTNR1A,NCF2,NFE2L2,NFKB1,NFKBIA,NOS2,NPC1L1,NPPA,PAH,PBLD,PCK1,PCNA,PDK4,PKLR,PLTP,PNPLA2,PPA1,PPARA,PPARG,PPM1D,PRC1,PRL,QPCT,SARDH,SELENBP1,SLC10A2,SLC2A4,SMC4,SOCS2,TALDO1,TBX21,TERT,TLR2,TSPO,UCP2,VCAM1,VEGFA |
| SREBF1 | 0,122 | transcription regulator | | -0,119 | 0,303 | 0,056 | 4,45E-13 | 5,32E-11 | AACS,ABCA1,ACACA,ACACB,ADGRG1,ADIPOQ,AGMAT,ALDOA,ALDOC,ALOX12,ANKRD2,APOBEC3F,APOC3,ARF4,BAX,BEST1,BHLHE40,BHLHE41,CAP2,CCL17,CDK4,CFD,CFI,CSAD,CXADR,CYB5A,CYP27B1,CYP7B1,DARS1,DBI,ELOVL6,FABP6,FAS,FN1,FOXO1,G6PC,GCK,GLB1,HMOX1,HNF4A,HSPA5,IDI1,IFI30,INS,KLK3,LDLR,LEP,LPL,MAT1A,MDK,MYOD1,MYOG,NCF2,NFIL3,NOS2,NR0B2,OAT,PCK1,PCK2,PIK3R3,PKLR,PLA2G3,PLA2G6,PLS3,PLTP,PNPLA3,PPARG,PTGDS,RBP1,RNASET2,S100A13,SERPINA3,SLC22A4,SORBS3,SUCLG1,SV2A,TNNI1,TP53,UCP2,VEGFA |
| IL3 | | cytokine | | -1,527 | -1,180 | 0,056 | 4,84E-13 | 5,70E-11 | ALDH2,ALOX5AP,ARHGAP29,ARHGDIB,ARHGEF1,BNIP3L,BTG2,C1QA,CASP3,CAT,CBFB,CCL17,CCT5,CD1A,CD48,CD53,CD63,CD83,CDKN1B,CIAPIN1,CISH,CITED2,CRAT,CSF3,CSNK1A1,CTLA4,DAD1,DDIT3,EGR1,EGR3,ELAVL3,EMP3,FAS,FKBP3,FOS,FUS,GADD45A,GADD45B,GADD45G,GNAS,GPR21,GZMA,HBEGF,HK1,HLA-A,HNF1A,HNF4A,HSP90B1,HSPA5,IK,IL13,IL17RB,IMPDH2,ITGAV,ITGB3,JUNB,KLF9,KPNB1,LCN2,LTA,LY75,LYN,MAN2B1,MCM5,MID1,MLF2,MMP1,NARS1,NRP1,ODC1,PDE1B,PIM1,PIWIL1,PLCB2,PRG2,PSMA5,PSMC4,PSME1,RAN,RARS1,RB1,RIDA,RPS19,SEMA7A,SERPINA3,SH3BP1,SOX4,SSBP1,STT3A,TARS1,TERT,TLR2,TNFSF10,TNK1,TYK2,UCP2,VCAM1,VEGFA,ZFAND5 |
| IFNA2 | | cytokine | Activated | 5,646 | 6,144 | 0,066 | 5,22E-13 | 5,98E-11 | ALOX5,ANXA1,B2M,BAX,C1R,C1S,CBFB,CCDC92,CD70,CDKN1B,CISH,CYB5A,DAD1,DDX58,DDX60,DPP4,FAS,GBP1,GBP2,GNAS,HBEGF,HERC6,HIF1A,HLA-A,HLA-B,HLA-C,HLA-E,HSPA6,IFI16,IFI35,IFI44,IFIH1,IFIT2,IFIT3,IFIT5,IFITM1,IFITM2,IFNAR2,IL10RB,IRF7,IRF9,ISG15,KCNJ6,LAPTM4B,LGALS3BP,LYN,MET,MT1X,MX1,OAS1,PARP12,PHF11,PLAAT4,PLSCR1,PPP1CA,PSME1,PTTG1,RB1,SAMSN1,SOCS2,SP110,STAT1,TAP1,TBX21,TDRD7,TLR3,TNFRSF10D,TNFSF10,TP53,TRIM21,TRIM22,UBD,UBE2L6,VCAM1,VEGFA,ZBP1,ZC3HAV1 |

| | | | | | | | | | |
|---|---|---|---|---|---|---|---|---|---|
| EGR1 | -1,174 | transcription regulator | | -0,270 | 0,155 | 0,058 | 7,13E-13 | 7,95E-11 | ALOX5AP,APOA1,APP,ARC,ATF3,BAIAP2,BAX,CACNA1H,CASP3,CASP6,CASP7,CASP8,CD44,CDC20,CLU,COL1A1,COL1A2,CSHL1,CXCL6,EGR1,ELK1,F3,FAS,FN1,FOSL1,FOXP3,GAD1,GADD45A,GADD45B,GLI1,GNAS,HBEGF,HIF1A,HMOX1,HPGD,IL13,IL6R,JUNB,JUND,LDLR,LHB,LYZ,ME1,MTNR1A,NAB2,NDRG1,NFKB1,NR4A1,PNPLA2,PPARG,PRL,PTEN,RB1,SGK1,SLC12A2,SNAI1,SOD2,SPP1,SYN1,TIMP1,TNFSF10,TP53,TP73,VCAM1,VEGFA |
| NFE2L2 | 0,262 | transcription regulator | Activated | 3,741 | 4,291 | 0,053 | 8,17E-13 | 8,99E-11 | ABCC1,ABCC3,ABCC4,ABCG2,ACTG1,AHR,ALAS1,ALDH3A1,ALDOA,ARF1,ATF3,BHMT,CA11,CAT,CCN2,CCT3,CFD,COL1A1,COL3A1,COL4A2,CTSD,CUL1,CXCL6,CYBB,DAD1,DCSTAMP,DDIT3,DDX50,DNAJB5,DNAJC3,DSTN,DYNLL1,EIF4G2,ELN,EPHX1,FABP1,FN1,GABRA2,GADD45G,GHR,GNG3,GSTA1,GSTA3,GSTM5,GSTP1,HAX1,HMOX1,HNF1A,HPRT1,HSP90B1,HTATIP2,IFNGR2,IFRD1,IL13,IMPDH1,KLK3,LPL,M6PR,MAFF,MAPK10,ME1,MGLL,MGST3,MST1R,NARS1,NCF2,NFE2L2,NOCT,NOS2,NR0B2,NSMF,NUCB2,OAT,P2RX4,PDIA3,PDIA4,PDIA6,PGD,PPARG,PPIB,PSMA1,PSMA4,PSMA5,PSMA6,PSMD1,PSMD14,PSMD7,PTPRB,PTPRO,RACK1,RAN,RARS1,RBBP7,RUVBL1,S100A13,SEC23A,SEC61A1,SERINC3,SERPINA3,SERPINA6,SHC1,SLC10A1,SLC38A3,SOD2,SQSTM1,TALDO1,TKT,TP53,TUB,TXNRD1,UBE2K,UBQLN2,USO1,USP14,VCAM1,VEGFA |
| FSH | | complex | | -1,335 | -1,177 | 0,024 | 8,32E-13 | 9,03E-11 | ACTN1,ACTR2,ADAMTS1,ADGRG1,ADM,AHR,AKAP12,ALDH3A2,ALPL,ARL4C,ARL6IP5,ATP9A,BAG1,BECN1,BMPR1B,BNIP3L,BTG2,CAMK2G,CAP2,CASP4,CCN2,CDK14,CDKN1C,COL15A1,COL18A1,CSNK1A1,CTSV,CYFIP2,DHRS3,DHX15,DLX5,DNAJB9,DUSP1,ELL2,ESR1,FDX1,FDXR,FILIP1L,FOS,FOSB,FOSL1,FOXO1,GATA6,GBP1,GDE1,GDF9,GK,GNAS,GNB1,GNRH2,HSD3B2,IGFBP4,IL6R,INHA,JUNB,KIDINS220,KITLG,LDLR,LGALS1,LYPD3,MAP2K3,MAP4K4,MT1X,NR4A1,PCK1,PCNA,PDYN,PHKA2,PKM,PLA1A,PLIN3,PLK2,POP5,PPARA,PPIH,PPP1CB,PRKD1,PRKX,PSIP1,PSMD1,PTEN,PTPRF,RAB27A,RAB4A,RAB5A,RGS16,RGS7,RHOB,SGK1,SHBG,SMAD1,SMAD2,SMARCA4,SNAP23,SSTR2,STAT1,STK24,STX4,TFPI2,TGFBR3,TIMP1,TNFRSF11B,TOB1,TP53,TPM1,TUBA1A,TYRO3,UBE2L3,VEGFA,WT1,ZNF331 |
| FOXO1 | -0,278 | transcription regulator | | -0,211 | 0,117 | 0,036 | 1,13E-12 | 1,21E-10 | A2M,ACACA,ADIPOQ,ADIPOR1,AGXT,ALAS1,ALB,ALDOB,ANXA3,APOC3,APOM,BAX,BID,BLNK,C1QA,CA2,CASP6,CASP8,CAT,CCL20,CCN2,CCNA2,CCNB1,CCNB2,CD70,CDK1,CDKN1B,CDKN1C,CITED2,COL4A1,COL4A2,CTSV,DCSTAMP,DLGAP5,EGR1,ELOVL6,FABP2,FAS,FBXO21,FN1,FOS,FOSB,FOXO1,FOXP3,G6PC,GABARAPL1,GADD45A,GADD45B,GCK,GPD1,GPI,GRHPR,GZMA,HIF1A,HMOX1,HPD,HSPA5,IL10RB,IL15,IL18,INS,INSR,ITGAV,ITGB2,ITGB6,JUNB,KLK3,LEFTY2,LEP,LHB,LPL,MAVS,MB,MCM5,ME1,MFN1,MLXIPL,MMP1,MXI1,MYOG,NAMPT,OVOL1,PBK,PCK1,PCK2,PDCD1,PDK4,PIK3IP1,PKLR,PLA2G2D,PLSCR1,PNPLA2,PPARG,PPP1R15A,PRC1,PRKCD,PRL,PSMB8,PTEN,PTPRB,PTPRC,RPIA,SEMA6D,SFN,SGK1,SLC25A24,SLC2A4,SOD2,SPRY2,SPTLC2,TBX21,TKT,TNFSF10,TNNC1,TP53,TSPO,VCAM1 |
| Akt | | group | | 0,812 | 1,041 | 0,027 | 1,72E-12 | 1,82E-10 | A2M,ADIPOQ,ADM,AIMP2,ANKRD2,B4GALT5,BAX,BHLHE40,BID,BIRC2,BTG2,CAV3,CCN2,CD44,CDK4,CDKN1B,CDKN1C,COL3A1,CTSD,E2F4,ELK1,ESR1,F3,FAS,FN1,FOS,FOSL1,FOXO1,FOXP3,GCK,GLI1,HIF1A,HMOX1,HPGD,IL13,IL32,INSR,KCNA5,LCN2,LEP,LPL,MCAM,MT1X,MUC1,NDRG1,NES,NET1,NFE2L2,NFIL3,NFKB1,NFKBIA,NOS1,NOS2,NOTCH2,NPPA,NR0B2,PCK1,PCNA,PDK4,PEA15,PLG,PPARG,PSMD10,RB1,RHOB,RRM2,SEMA6D,SGK1,SLC1A2,SLC2A4,SNAI1,SOD2,SOX4,SPP1,TERT,THBS2,TIPARP,TLR2,TMED10,TNC,TNFSF10,TP53,VCAM1,VCAN,VEGFA,WT1 |

| | | | | | | | | | |
|---|---|---|---|---|---|---|---|---|---|
| Insulin | | group | | 1,364 | 1,580 | 0,023 | 1,90E-12 | 1,96E-10 | ABCA1,ACACA,ACAT1,ADAM10,ADIPOQ,ADIPOR1,ALB,ALDOA,ALDOB,APLNR,APOA1,APOM,APP,ATF3,AVP,BECN1,CANX,CCN2,CCNC,CCT2,CCT5,CCT6A,CCT8,CDK8,CDKN1B,CDX2,CIDEC,COL1A1,COL1A2,COL3A1,COL4A1,COX6A2,CRABP2,CSHL1,CTH,CYBB,DBI,DEFB1,DUSP1,EGR1,ELOVL6,FKBP8,FN1,FOS,FOSB,FOXC2,FOXO1,G6PC,GCK,GHRH,GRB10,GSTP1,HAP1,HIF1A,HMOX1,HNF1A,HSD3B2,HSP90B1,HSPA4,HSPA5,IFIT2,IFIT3,IGFALS,IGFBP4,INSR,IRF7,ITGB3,KLK3,LAMB1,LEP,LEPR,LGALS3BP,LPL,MAPT,MAT1A,ME1,MLXIPL,NFIL3,NFKBIA,NOS2,NR4A1,NR4A3,NUCB2,ODC1,OGA,P4HB,PCK1,PCNA,PDK2,PDK4,PER2,PKM,PLIN3,PNPLA2,PPARA,PPARG,PPIB,PPIC,PPP1R15A,PRKCA,PRKCD,PRL,PTEN,PTK2,RAC1,SGK1,SLC10A2,SLC2A4,SLC2A5,SLC37A4,SLC38A3,SLC5A5,SLC6A4,STAT1,STAT3,TBX21,TGFBI,THBS2,TLR2,TNC,TRPV1,UCP2,VEGFA,VLDLR |
| PRL | -0,273 | cytokine | Activated | 4,715 | 5,185 | 0,054 | 2,47E-12 | 2,49E-10 | A2M,ABCG2,ACTR3,ADAR,ANXA2,ANXA3,ANXA5,APP,B2M,BAG1,CAMK2N1,CAPN2,CCL20,CCNB1,CDK4,CISH,CLDN3,CLU,COL1A1,COL1A2,COL3A1,CPD,CTSA,CTSD,CTSS,CTSV,DDX58,EGF,EGR1,ELOC,ENO1,ESR1,FN1,FOS,HERC6,HSD11B2,IFI35,IFI44,IFIH1,IFIT3,IFIT5,IFITM1,IGFALS,IRF7,IRF9,ISG15,KRT19,KRT31,LAMC1,MAGED1,MARS1,MME,MSN,NFKB1,NOS2,OAS1,ODC1,P4HB,PARP12,PCNA,PDIA4,PDIA5,PDK2,PDK4,PIM1,PLSCR1,PRL,PSME1,RALB,SERPINA3,SLC10A1,SND1,SOCS2,SP110,SPP1,STAT1,STAT5A,TDRD7,TIMP1,TMEM140,TP53,TUBA1B,VDR,VIM,XDH |
| Ins1 | | other | | -2,193 | -1,920 | 0,031 | 2,65E-12 | 2,63E-10 | ACACA,ADIPOQ,ADIPOR1,ALAD,ALAS1,ALB,AP2M1,APOA1,APOC3,ARF1,BCKDHA,CA14,CDKN1B,CFD,COL18A1,COL1A1,CRAT,CRYAB,CSHL1,CYB5A,DDIT3,DLX5,DYNLL1,E2F4,ECI2,EGR1,ELOVL6,FOS,FOXC2,G6PC,GAST,GCK,GPD1,GSK3A,GSTP1,HSD11B2,HSD17B4,IGFBP4,IMPDH2,INS,INSR,ITGA6,JUNB,LDLR,LEP,LGALS3BP,LPL,MAPK1,MAPT,MLXIPL,MMP1,MPZ,MYOD1,MYOG,NGF,NOS1,NOS2,NPPA,NTF3,ODC1,PCK1,PCNA,PDIA2,PER1,PKLR,PPARA,PPARG,PPIB,PRKCA,PRL,PTEN,RHOA,RRM1,RRM2,RXRA,SLC28A1,SLC2A4,SLC37A4,SOCS2,SOD2,STAT3,STAT5A,STX4,TBX21,TIMP1,TTF2,UBE2D2,UGT8,VAMP2,VEGFA,VIM |
| ethanol | | chemical - endogenous mammalian | | 0,968 | 1,185 | 0,022 | 3,98E-12 | 3,92E-10 | ABCA1,ABCG1,ACACA,ACKR3,ADIPOQ,ADIPOR1,ADM,ALDH1A3,ALOX15B,ALOX5,ANPEP,APOA1,ATF3,AVP,BAX,BHMT,CAPN1,CASP3,CAT,CCN2,CDKN1B,CHRNA6,CLU,COL1A2,COL3A1,CYP2A6 (includes others),CYP2B6,CYP2C8,CYP3A4,CYP3A5,CYP3A7,CYP4A11,DCT,DDIT3,DRD2,DUSP1,EGR1,EPHX1,ESR1,FAAH,FABP1,FABP2,FAS,FOS,FOSB,G6PC,GADD45A,GCK,GDE1,GHRH,GNAI1,GNAS,HIF1A,HLA-A,HMOX1,HSBP1,HSP90B1,HSPA5,HTATIP2,IL15,IRF7,LEP,LGALS1,MAPK8,MAT1A,MGST3,MLXIPL,MME,MYCL,NCF2,NMI,NOS2,NPPA,PARP1,PCK1,PCNA,PDCD1,PDYN,PLK3,PNPLA2,PPARA,PPARG,PPP1R3C,PRL,PTEN,RBP1,RHOB,RORA,SEC23A,SGK1,SLC1A2,SNAI1,SOD2,STAT1,STAT3,TBP,TFPI2,TIMP1,TLR1,TLR2,TLR6,TP53,TSC22D3,VAMP2,VCAM1,WEE1 |
| TCF4 | 0,083 | transcription regulator | | -0,900 | -0,699 | 0,023 | 4,68E-12 | 4,51E-10 | AMHR2,APOBEC3G,ARHGEF18,BAG2,BHLHE40,BMP7,CASP1,CASP3,CASP4,CASP8,CCN5,CCNA2,CCNB2,CCT6A,CD24,CDC20,CDC25B,CDH17,CDK1,CDKN1C,CDKN3,CKAP4,CKS2,CRELD2,CSF3,DCK,DCT,DDX58,DIMT1,DPF2,DPT,DSTN,EAF2,EPCAM,FAM111A,FAS,FGF1,FKBP8,FN1,FOS,FOXO1,GLI1,GP9,GRPEL1,HIF1A,HMGB2,HSP90B1,IFI16,IGFBP4,IL15,KCNN4,LEPROTL1,MAD2L1,MANF,MITF,MMP1,MVP,NDC80,NEIL1,NELL1,NOD1,NOS2,NOTCH2,OCA2,OTOR,PARP8,PDIA4,PDIA6,PIWIL1,PLK2,POP7,PPARG,PPID,PPIF,PPIH,PRC1,PRDX4,PTCRA,QTRT2,RHOC,RPS6KC1,RRM2,SACS,SDF2L1,SEC24D,SERPINB1,SGK1,SH3BP5,SH3GLB1,SMAD1,SMAD2,SMAGP,SORBS2,SP140L,SPP1,SQSTM1,STAT3,SUB1,TERT,TIA1,TMEM140,TMEM184B,TNFSF10,TRIM38,VCAN,WARS1,ZBP1,ZP2 |
| IRF1 | 0,232 | transcription regulator | Activated | 5,083 | 5,496 | 0,060 | 8,50E-12 | 7,89E-10 | B2M,BAX,C1R,CASP1,CASP3,CASP7,CASP8,CCL20,CCNB1,CDK4,CFB,CTSS,CYBB,DDX58,EIF4A3,ERAP1,FGL2,FOXP3,GBP2,HLA-G,IFI35,IFIH1,IFIT2,IFIT3,IFIT5,IFITM1,IL13,IL15,IL18,IL7,IRF7,IRF9,ISG15,LTB,MAP4K4,MX1,NOS2,OAS1,ODC1,PCNA,PIGR,PLAAT4,PSMB10,PSMB8,PSMB9,PSME1,SLPI,SOCS7,SP110,STAT1,STAT3,TAP1,TAPBP,TERT,TLR3,TLR6,TNFSF10,TP53,TRIM21,TRIM22,UBD,VCAM1 |

| | | | | | | | | | |
|---|---|---|---|---|---|---|---|---|---|
| EGFR | | kinase | | 1,132 | 1,479 | 0,036 | 1,06E-11 | 9,66E-10 | ABCG2,ABI1,ANXA1,BAX,BZW2,C12orf49,CAPZA2,CASP3,CASP4,CBX5,CCN2,CCNA2,CCT5,CD47,CDK1,CDK4,CDKN1B,CHI3L1,COL1A1,COL3A1,COL5A2,CRMP1,CRYAB,CUL1,DDIT3,DSG2,DUSP1,EGF,EGR1,EGR3,ELK1,ESR1,F2RL1,F3,FBN1,FOS,FOSL1,FOXP3,GADD45A,GAST,GBP1,GHR,GRN,HBEGF,HIF1A,HMGB3,HSD11B2,HSP90B1,HSPA5,IER2,IGFBP4,IL36A,ITGA6,JUNB,JUND,KCNN4,KLK3,KPNB1,LCN2,MACF1,MARCKS,MET,MME,MMP1,MUC1,MXI1,MYBL2,NFKBIA,NOS2,NPPA,NSF,ODC1,OSMR,PCNA,PDIA2,PKM,POSTN,PROM1,PSEN1,PTEN,QTRT2,RAI14,RB1,RBBP7,RCL1,RHOB,RRAS,S100A2,SDF2L1,SEMA7A,SERPINA3,SERPINE2,SLPI,SPRY2,STAT3,TM4SF5,TNC,TNFRSF11B,TPST1,TRMT2A,TUBB4A,VCAN,VDR,VEGFA,VIM,WEE1,WNT10B,WNT6,WNT7B |
| AGT | 0,279 | growth factor | Activated | 1,865 | 2,275 | 0,040 | 1,18E-11 | 1,05E-09 | ADIPOQ,ADIPOR1,ADM,ALOX12,ANXA1,ANXA3,ATF3,AVP,BAX,BECN1,CASP3,CASP8,CAT,CAV3,CCN2,CD44,CDK1,CDK4,CDKN1B,CIT,COL11A2,COL1A1,COL1A2,COL3A1,COL4A1,CRABP2,CTF1,CTSS,CYBA,CYBB,CYP11B2,CYP4A11,DRD5,DUSP1,EGR1,F3,FBXO21,FDX1,FDXR,FN1,FOS,FRAT2,GCH1,GFPT1,GSTA3,GSTM5,GSTP1,HBEGF,HIF1A,HMGB2,HMOX1,HSD3B2,IDI1,IGFBP4,IL13,IL15,IL18,INSR,ITGA7,ITGAV,ITGB3,ITPKB,KITLG,KRAS,LAMB1,LDLR,LEP,LEPR,LIMD2,LPP,LTA,MAD2L1,MAPT,MMP1,MYH10,NAMPT,NCALD,NCF2,NDRG1,NGF,NMNAT2,NOS1,NOS2,NOTCH4,NOX5,NPHS1,NPPA,NR4A1,PARP1,PECAM1,PFN1,POSTN,PPARG,PPFIBP1,PPP1R3C,PSMB10,PSMB8,PTEN,RAC1,RGS2,RHOA,RRAS,SGK1,SHC1,SOD2,SOX4,SPP1,TBX2,TFPI,TIMP1,TLN2,TP53,UCP2,UGCG,VCAM1,VEGFA,VIM,ZFP36,ZNF268 |
| NFKBIA | -0,487 | transcription regulator | | 0,182 | 0,403 | 0,022 | 1,31E-11 | 1,15E-09 | A2M,ACKR3,ALOX15B,ANKRD49,APOA1,AZGP1,BAX,BID,BIRC2,BPIFA1,BTG2,C3,CAPZA2,CASP4,CASP8,CCL20,CCNA2,CCNB1,CCNC,CDC25B,CDH3,CHI3L1,CLU,COL1A2,COL3A1,COL5A2,CRYAB,CSF3,CSK,CTNNA1,CTSF,CTSZ,CXCL6,CYBA,CYP7B1,DDIT3,EIF4G2,ERAP1,ESM1,F3,FAS,FCGRT,FHL2,FN1,FOS,FOXC2,GADD45A,GADD45B,GADD45G,GBP2,GCH1,GM2A,GRN,GSTM5,HIF1A,HLA-A,HMOX1,HOXA10,HSPA5,IFI16,IL13,IL15,IRAK1,ISG15,ITGAV,JUNB,KCNN4,LCN2,MMP1,MSH2,MYOD1,NFKB1,NFKBIA,NFKBIE,NGF,NOCT,NOS2,NPPA,NR4A1,PCNA,PDHA1,PDLIM1,PEA15,PICALM,PIM1,PRL,PTEN,PTP4A2,RAC1,RBP1,RGS16,ROCK1,RRM1,SCFD1,SERPINE2,SGK1,SLC12A2,SLC1A2,SOAT1,SOD2,TCEAL9,TCF12,TIMP1,TLR2,TMPO,TNC,TNFAIP8,TNFRSF11B,TNFSF10,TP53,VCAM1,VEGFA,XRCC5,YAP1,ZNF638 |
| ANGPT2 | | growth factor | Activated | 3,200 | 3,628 | 0,057 | 1,69E-11 | 1,41E-09 | ABCC1,ARL4C,BAG1,C1R,CASP3,CAT,CFB,COL1A1,COL1A2,COL3A1,COMT,CRYAB,CRYBB3,CYBA,CYBB,CYP11B2,DDIT3,EXT2,FAS,FN1,GATA6,HIF1A,HMOX1,HSPA4,HSPA5,INPP5F,KPNA2,LEP,LEPR,LYN,MAPK10,MMP1,MMP7,MYOD1,MYOG,NME1,NPHS1,NPPA,P4HB,PDCD4,PDIA3,PDIA5,PDIA6,PLG,POSTN,PPARA,PROS1,RFC2,RFC4,RHOB,RPA2,RPA3,SERPING1,SMYD5,SNAI1,SOCS7,SOD2,ST13,THBS2,TLR3,TP53,UBE2A,VCAM1,VIM,YIPF5 |
| ERK1/2 | | group | | -1,513 | -1,098 | 0,049 | 1,69E-11 | 1,41E-09 | ABCA1,ABCC6,ADIPOQ,ALOX5,ANKRD2,ATP1A3,AVP,B2M,BAX,BHMT,BIRC2,C3,CAT,CAV3,CCL20,CCL22,CCN2,CD44,CDK1,CDK4,CDKN1B,COL1A1,COL3A1,CTSV,CXCL6,CYBB,DDIT3,DUSP1,EGR1,ELK1,ENO1,ESR1,F3,FN1,FOS,FOSB,FOSL1,FOXO1,FZD1,GAD1,HBEGF,HIF1A,HLA-A,HPGD,HSPA5,INS,ITGB3,JUNB,JUND,KIF2A,LDLR,LEPR,MAP3K10,MAPK1,MAPT,MMP1,MUC5B,MYOG,NAMPT,NKX2-5,NOS2,NPPA,NR4A3,PCNA,PKM,POSTN,PRL,PSMB10,PSMB8,PSMB9,RAC1,RB1,RHOC,S100A6,SGK1,SLC12A6,SNAI1,SPP1,SPRY4,SSTR3,TAP1,TAPBP,TBP,TERT,TIMP1,VCAM1,VCAN,VEGFA |

| | | | | | | | | | |
|---|---|---|---|---|---|---|---|---|---|
| CD40LG | -0,097 | cytokine | | 1,264 | 1,786 | 0,066 | 2,51E-11 | 2,05E-09 | AHR,AKAP13,APOBEC3G,ARMC6,ASCC3,ATF3,B2M,BAX,BCAT2,BIRC2,CA2,CASP1,CAST,CBX3,CCL17,CCL20,CCL22,CCN2,CD44,CD63,CD70,CD83,CDC25B,CDKN1B,CHRM5,CLU,DUSP1,DUSP2,EGR1,F3,FAS,FOS,FOSB,FURIN,G0S2,GADD45A,GCH1,HIF1A,HLA-DQB1,IFI44,IFIT2,IFIT3,IFITM1,IL10RA,IL13RA1,IL15,ISG15,ITPA,JUNB,JUND,KIF2A,LMAN2,LTA,LTB,MAP2K3,MARCKS,MARCKSL1,MSH2,MX1,NAMPT,NET1,NFKB1,NFKBIA,NFKBIE,NOS2,NR4A3,PIM1,PLA2G4C,PREPL,PSMB10,PSMB8,PSMB9,PSME1,PTPN12,RAD50,RASA1,RB1,RGS2,RIDA,RNASE3,SAMSN1,SEMA4D,SERPINA10,SOD2,SPHK2,STAT1,STAT3,STAT5A,SWAP70,TAP1,TFAP4,TNFRSF11B,TNFRSF13B,TNFSF10,TNPO1,TP53,TPD52L2,TUBB,UBD,UCHL3,VCAM1,VEGFA,XPO7,XRCC5,ZNF195,ZZZ3 |
| Pkc(s) | | group | | -1,460 | -1,133 | 0,046 | 2,61E-11 | 2,11E-09 | ABCA1,ACAT1,ACO1,ADM,AHR,APOA1,APP,ATF3,CCN2,CDKN1B,CDX2,CGA,CLDN1,COL3A1,CYBB,CYP2A6 (includes others),DDIT3,DNAJC3,DUSP1,EGR1,F3,FAS,FGFBP1,FN1,FOS,FOSL1,GADD45A,GADD45B,HIF1A,HMOX1,HPGD,HSP90B1,HSPA5,IVL,JUNB,LHB,LIPA,MAPK1,MMP1,MYOG,NFKB1,NOS1,NOS2,NOTCH4,NR4A1,NR4A3,OVOL1,P4HB,PDIA4,PER1,PER2,PLA2G6,PPARA,PPARG,PPP1R15A,PRKCA,PRL,RB1,RGS2,S100A1,SLC1A2,SLC6A4,SPP1,STC2,TGM1,TP73,VCAM1,VDR,VEGFA |
| STAT1 | 0,761 | transcription regulator | Activated | 6,287 | 6,766 | 0,055 | 2,69E-11 | 2,12E-09 | A2M,ABCA1,ALDH1A3,APOBEC3B,APOBEC3G,B2M,BAX,C1R,C1S,C3,CAPN1,CASP1,CASP3,CASP4,CASP6,CASP8,CCL20,CDKN1B,CFB,CLIC5,CTSS,CX3CR1,CYP2C8,CYP3A5,CYP4A11,DCSTAMP,DDX60,DPP4,EGR1,FAS,FCER1G,FGL2,FOS,FOXO1,FOXP3,FURIN,GBP1,GBP2,GCK,HERC6,HIF1A,HLA-E,HTRA1,IFI16,IFI30,IFI35,IFI44,IFIH1,IFIT2,IFIT3,IFITM1,IFITM2,IL15,IL18,INS,IRF7,IRF8,IRF9,ISG15,LCN2,LY96,MUC1,MX1,NOS2,NOX5,NPPA,OAS1,PECAM1,PIM1,PLSCR1,PPARG,PRL,PSMB10,PSMB8,PSMB9,PSME1,RORC,RTP4,S100A10,SERPINA3,SERPING1,SMAD2,SMAGP,SOAT1,SP110,STAT1,STAT3,TAP1,TBX21,TLR3,TNFSF10,TP53,TP53BP2,TRIM21,TRIM22,UBD,ULK1,WARS1,ZBP1 |
| RARA | | ligand-dependent nuclear receptor | | -0,083 | -0,085 | -0,000 | 2,70E-11 | 2,12E-09 | AASS,ABCA1,ABLIM1,ACACA,ACACB,ADAM10,ALDH1A1,ALOX5,APOA1,APOC3,APOM,CCNA2,CCNC,CD1A,CD24,CD44,CD53,CDK1,CDKN1B,CDKN3,CDX1,CEBPE,CENPM,CENPU,COBL,CRABP2,CSF3,CSHL1,CTNNA3,CTSS,CYP26A1,DHRS3,DRD2,DUSP1,EGR1,EGR3,ELL2,ENAH,F12,FOS,FOXP3,FUT4,GCK,GDF2,GHR,GJB4,GLI1,GSK3A,HCP5,HLA-B,HLA-C,HLA-G,HLA-J,HNF1B,HOXA10,HOXB1,HOXB7,IFI44,IL13,IL6R,KLK3,LAMB1,LGALS1,LGALS3BP,LTF,LY6G6D,LYPD1,MAD2L1,MAML1,MAP3K10,MAPK1,MED7,NR2F1,NRP1,OAS1,PBK,PCK1,PGLS,PLXNA2,PTTG1,RARG,RARRES1,RBP1,RORC,RTP4,SLC10A1,SLC25A36,SMC4,SP110,SPP1,STAT1,STRA6,STX3,TAPBP,TERT,TNFRSF11B,WNT8B,ZNF544 |
| CEBPA | 0,005 | transcription regulator | | -1,568 | -1,317 | 0,027 | 2,74E-11 | 2,13E-09 | A2M,ACACA,ADH6,ADIPOQ,AKAP12,AKR1B1,AKR1C1/AKR1C2,ALB,ALOX5AP,ANPEP,ANXA1,APLNR,APOB,APOC3,AQP5,ARL4C,ARL6IP5,ASNS,ASS1,BHMT,BTG2,C3,CA2,CCL20,CCNA2,CCNB2,CDK4,CEBPE,CFD,CHI3L1,CKAP4,COL1A1,COL1A2,CRABP2,CSF3,CTNNA1,CYP2A6 (includes others),CYP2B6,CYP3A4,CYP3A5,CYP3A7,EDF1,EPHA7,EPHX1,FHL1,FOS,FOXO1,FZD1,G0S2,G6PC,GADD45A,GATA6,GBP1,GCH1,GLI1,GLRX,GP9,GSTP1,HDAC1,HLA-B,HMOX1,HNF1A,HNF1B,HPGD,HSPA5,IGFBP4,IL6R,INSR,ISG15,IVL,JUNB,KLK14,LCN2,LEP,LEPR,LGALS1,LPL,LTF,MALT1,MC1R,MYBL2,MYOD1,MYOG,NFIL3,NRP1,OVOL1,PCK1,PCNA,PGD,PLOD2,PPARA,PPARG,PPL,PPP1R3C,PRL,PTPN3,PTPRC,RGS2,RIN1,RORA,SERPINB1,SERPINI1,SLC10A1,SLC2A4,SOD2,SPINT2,SPP1,SPRR3,THRB,TNFSF10,TSC22D3,UBE2I,VCAN,VDR,VLDLR,ZMIZ1 |

| | | | | | | | | | |
|---|---|---|---|---|---|---|---|---|---|
| TP63 | | transcription regulator | | -0,357 | -0,067 | 0,034 | 2,95E-11 | 2,26E-09 | ACTL6A,ADA2,ADAMTS1,ADM,AGR2,AHR,ALDH1A3,ALOX12,ATG5,ATOH1,BAX,BECN1,BHLHE40,BMP7,CASP3,CAST,CCNA2,CCNB1,CDK1,CDK18,CDKN1B,CDKN1C,CITED2,CKS2,CLDN3,COL4A1,CSTA,CTSD,CYP2A6 (includes others),DAP3,DDB2,DNMT1,EFNA4,EI24,F3,FAS,FBN1,FDXR,FN1,FOS,FOSL1,FUBP1,FUT1,GADD45A,GCNT3,HAGH,HBEGF,HSPA4,IHH,INSR,ITGA7,ITGB8,IVL,JUNB,KHK,KLK3,KRT10,KRT31,LLGL1,LYN,MAD2L1,MAFF,MAP2K3,MAP4K4,MAPK8,MPZL2,PCNA,PERP,PIK3R3,PKLR,PKP2,POSTN,PPARA,PTEN,PTPN12,RAC2,RIN1,RPA1,S100A2,S100A4,SFN,SH2B3,SMAD2,SNAI1,SPINK4,SPRR3,TERT,TFF2,TIPIN,TNC,TNFAIP8,TNFSF10,TNS4,TP53,TP53I3,TP73,TPM1,TRIM29,ULK1,UPK1A,UPK2,VDR,VEGFA,VIM,WEE1,WFDC2 |
| IL13 | -0,261 | cytokine | | 0,067 | 0,389 | 0,034 | 4,14E-11 | 3,11E-09 | ABCA1,ACKR3,ADAM15,ALOX5AP,ATF3,AVPI1,BAX,BID,BIRC2,BLVRA,BPIFA1,BZW2,C3,CA2,CAPN2,CASP1,CASP3,CASP6,CASP7,CASP8,CCL17,CCL20,CCL22,CCL25,CCN2,CD163,CD1A,CD1B,CD44,CD48,CD52,CFI,CHI3L1,CHN2,CISH,CLCA1,COL18A1,COL1A1,COL1A2,COL3A1,CTSS,CTSV,CXCL6,CYBB,DHCR24,DNASE2,EGR1,F3,FAS,FKBP1A,FLOT1,FXYD3,G0S2,GAS6,GP2,HK1,HMOX1,HTRA1,IFNGR1,IFNGR2,IL13,IL13RA1,IL17RB,IL18,IVL,JAK1,KITLG,LILRB3,LIPA,MCUR1,MMP1,MS4A4A,MT1X,MUC5B,MYOD1,MYOG,NHLH1,NOS2,PAPSS1,PFKP,PLA2G2D,POSTN,PPARG,RHOA,RORC,RYK,SAMSN1,SLC26A6,SLC7A7,SLPI,SPDEF,SPINT1,SPINT2,SPP1,ST14,ST3GAL6,SWAP70,SYNGR2,TIMP1,TLR1,TNFRSF11B,TNS1,TRPV1,TYRO3,VCAM1 |
| IL2 | | cytokine | | 0,875 | 1,381 | 0,048 | 4,18E-11 | 3,11E-09 | ABCA1,ABCC1,AHR,ANXA1,APOBEC3G,ARHGAP12,ARHGAP32,ARHGEF18,AVP,BAG1,BAX,BHLHE40,CACNA1E,CACNG3,CAMK2N1,CAPG,CASP1,CASP3,CASP4,CCL1,CCL17,CCL22,CCR4,CD44,CD47,CD5,CD58,CD59,CD74,CD83,CDK4,CDKN1B,CDKN1C,CHN2,CISH,CPD,CRMP1,CSF3,CTCF,CTLA4,CX3CR1,CYP2B6,DAPK2,DENND2D,DKKL1,DPP4,DRD5,DSG2,DUSP2,E2F4,EEF1E1,FAM13B,FAS,FCER1G,FGL2,FOS,FOXO1,FOXP3,GADD45B,GADD45G,GARS1,GNL3,GRN,GZMA,H1-2,HDAC1,HSP90B1,HSPA5,IDI1,IFNGR1,IFNGR2,IGFBP4,IL10RA,IL13,IL17RB,IL18,ITGA6,ITGAE,ITGB8,JAK1,JUND,LAPTM5,LDLR,LTA,LTB,MLLT3,MMD,MME,MMP25,MX1,NAB2,NCR3,NDRG1,NET1,NETO2,NFKB1,NME1,NMI,NOS2,NRGN,NRP1,PCNA,PDCD1,PDCD4,PDIA3,PEA15,PECAM1,PIK3R3,PIM1,PLA2G2D,PNP,POLE2,PPARG,PRKX,PROS1,PTPN3,PTPRC,PTRH2,RGS16,RGS2,RHOB,RHOC,RORC,RUFY1,RXRA,S100A4,SNAP23,SOCS2,SPP1,STAM,STAT3,STC2,SWAP70,TBX21,TGFBR3,TIMP1,TLR2,TNFAIP8,TNFRSF10D,TNFRSF11B,TNFRSF13B,TNFSF10,TP53,VEGFA,VIM,WDR3,WT1,YARS1 |
| LDL | | complex | Activated | 2,512 | 2,822 | 0,044 | 4,55E-11 | 3,35E-09 | ABCA1,ABCG1,AP1G1,APOBEC3B,ATF3,C3,CAPN1,CAPN2,CASP1,CASP10,CASP3,CASP6,CASP7,CASP8,CCL20,CCN2,CDK1,CX3CR1,CXCL6,CYBB,CYP11B2,CYP2C8,DDIT3,DNMT1,DUSP1,DUSP2,EGR1,EGR3,F3,FAS,FOS,FOSB,G0S2,GAS6,GJD2,GNAI1,GTF2I,HIF1A,HMOX1,HSP90B1,HSPA5,HSPA6,IFIT2,IFITM1,IGFBP4,IL10RA,IRF7,ITGAV,ITGB3,LASP1,LDLR,LTA,MMP1,MX1,NAMPT,NFKB1,NME1,NOS2,NR0B2,NR4A1,NR4A3,PANX1,PARP1,PECAM1,PPARG,PTEN,PTPRF,RNASEH1,SHC1,SOD2,STAT3,TGFBR3,TNFSF10,TP53,UBE2K,VCAM1,VEGFA |
| IL15 | 0,210 | cytokine | | 0,531 | 0,907 | 0,047 | 4,70E-11 | 3,43E-09 | AHR,ALDOA,ALDOC,ANXA1,APBA1,ARHGDIB,CALM1 (includes others),CAMK2G,CASP3,CCL17,CCN2,CD33,CD44,CD5,CD53,CD59,CD74,CD83,CDK2AP1,CDK4,CDKN1B,CISH,CKS2,COPB2,CSF3,CX3CR1,DPP4,ELK3,ELOC,ENO1,ENO3,EXOSC8,FAS,FBP1,FKBP1A,FOXP3,GARS1,GNAS,GNL2,GPI,HAT1,HK1,HMOX1,HNRNPF,IFI35,IFITM1,IL13,IL15,IL32,JUNB,LEP,LTA,LYN,MACROH2A1,MAPK1,MCM5,MLLT3,MX1,NCR3,NFKB1,NFKBIA,NME1,NOS2,PAFAH1B3,PCBP2,PCNA,PDCD1,PDCD4,PDCD5,PDIA3,PDIA4,PFKP,PFN1,PGD,PIK3R3,PIM1,PKM,PNP,PRDX4,PSMA5,PSMB10,PSMB8,PSMB9,PTP4A2,PTPRC,RAB8A,RAC2,RBX1,RGS16,RPIA,S100A11,SET,SMAD2,SOCS2,SYNPO,TALDO1,TBX21,TIGAR,TIMP1,TKT,TLR2,TNFSF10,TNNI1,TRIM5,TSPAN7,TXNRD1,TYROBP,VBP1,VCAM1,VDR,VEGFA,YARS1,ZFP36 |

| | | | | | | | | | |
|---|---|---|---|---|---|---|---|---|---|
| THRB | -0,175 | ligand-dependent nuclear receptor | | 0,113 | 0,191 | 0,012 | 4,75E-11 | 3,44E-09 | ACACA,APOA1,APOC3,APOM,ATP1A3,BTG2,CASP1,CCL17,CCNB1,CDK4,CDKN1B,CFD,CGA,CSHL1,CSNK1A1,CTSD,CYP27B1,DAD1,DIO1,DR1,EGF,F12,FOS,FOSL1,FOXN1,FSTL1,FZD1,FZD6,G6PC,GCH1,HIF1A,IL7,KLF9,LDLR,LPL,LYZ,MAPK8,MC4R,ME1,MET,MLXIPL,MMP1,MMP7,NME1,NPPA,NR4A1,NRAS,NRGN,PAM,PCK1,PDK4,PER1,PGD,PHYH,PPARG,PRL,PROM1,PTGDS,PTTG1,SLC2A4,TGFBI,THRB,TP53,WIF1,WNT11,WNT2,WNT8B,YWHAE |
| PTP4A1 | -0,116 | phosphatase | | 0,758 | 0,889 | 0,020 | 9,85E-11 | 6,95E-09 | ACTN1,ACTR2,ACTR3,AHCY,ALDOA,ANXA5,BAX,C1orf21,COL15A1,CRK,CSNK2A1,ELAVL1,FKBP1A,FLRT1,GSTP1,HIF1A,HNF4A,HNRNPF,HRAS,HSPA6,IFI16,IMPDH2,ITGB8,MAP3K1,MAPK10,MAPK8,NCK1,NME1,NRAS,PCNA,PECAM1,PKM,PPA1,PPP1R12A,RAN,ROCK1,RUVBL1,SHC3,SPINT2,STMN1,TKT,TMEM267,TP53,VCAN,VIM |
| P38 MAPK | | group | | -0,652 | -0,063 | 0,065 | 1,20E-10 | 8,34E-09 | ABCA1,ADIPOQ,ANXA5,AQP5,ASS1,ATF3,BAX,BNIP3L,CAV3,CCN2,CD44,CD70,CD83,CDX2,COL3A1,CSF3,CXCL6,CYBB,DDB2,DDIT3,DIO3,DUSP1,EGR1,ELK1,F3,FADD,FAS,FDXR,FN1,FOS,GBP1,HIF1A,HMOX1,HSPA5,HTRA1,IL13,IL32,IRF7,ITGAV,ITGB2,ITGB3,ITGB8,IVL,JUNB,KDELR3,KLF15,LEP,MAP4K4,MITF,MMP1,MSH2,MUC5B,MYBPH,MYOG,NFATC4,NFE2L2,NFKB1,NGF,NKX2-5,NOS2,NPPA,NR4A1,NR4A3,PCK1,PCNA,POSTN,PPARG,PPM1D,PTEN,PTPN12,RAB5A,RASA1,RB1,RBP1,RHOB,SATB2,SGK1,SLC5A5,SMAD2,SNAI1,STAT1,TBP,TBX21,TERT,TIMP1,TLR2,TNFSF10,TNNC1,TNNC2,TNNI1,TP53,TP53I3,ULK1,VCAM1,VDR,VEGFA,VLDLR,ZFP36,ZMAT3 |
| BRCA1 | | transcription regulator | | 1,789 | 1,956 | 0,024 | 1,22E-10 | 8,40E-09 | ALDH1A3,AQP5,BAX,CCNB1,CCNB2,CDH3,CDKN1B,COL18A1,CTSD,CTSF,CYP3A4,DDB2,DDIT3,DDX58,EGR1,ELAVL1,ESR1,FAS,FHL2,FOXO1,FXYD3,GADD45A,GADD45B,GTF2H1,HBB,HMOX1,HSD11B2,IFIT2,IFIT3,IFITM1,IRF7,JAK1,KLK3,MAD2L1,MX1,NFE2L2,PCBP2,PCNA,PERP,PLSCR1,PLTP,PTEN,RB1,S100A2,SERPINE2,SFN,SNAI1,STAT1,STAT5A,TAP1,TBP,TERT,TP53,TP53I3,UBE2I,VCAN,VEGFA,VIM,WEE1 |
| XBP1 | 0,270 | transcription regulator | Activated | 3,550 | 4,014 | 0,059 | 1,25E-10 | 8,55E-09 | ACACA,ACACB,ADAM10,AFP,APOA1,APP,ARFGAP3,ATG5,BECN1,BET1,CAT,COPB1,COPB2,CREB3L1,CRK,DAD1,DDIT3,DDOST,DERL1,DNAJB9,DNAJC10,DNAJC3,ESR1,FAS,FN1,GOLGA3,GOLPH3L,GORASP2,HLA-DRA,HMOX1,HSP90B1,HSPA5,INS,KDELR2,KDELR3,LMAN2,MGAT2,NOS2,NUCB2,P3H1,PDIA2,PDIA3,PDIA4,PDIA6,PGC,PPIB,RAB3A,RPN2,S100A6,SDF2L1,SEC11A,SEC23A,SEC23B,SEC24D,SEC61A1,SEC61B,SEC61G,SPCS3,SRP54,SSR1,STT3A,TFF2,TRAM1,USO1,VAMP2,VCAM1,WNT10B,YIF1A,YIPF5 |
| IFNL1 | | cytokine | Activated | 4,741 | 5,166 | 0,074 | 1,77E-10 | 1,18E-08 | ATF3,CD83,CHAC1,DDX58,DDX60,GBP1,HERC6,HLA-B,HLA-C,IFI35,IFI44,IFIH1,IFIT2,IFIT3,IFIT5,IFITM1,IFITM2,IL13,IRF9,ISG15,LGALS3BP,MX1,OAS1,PHF11,PLAAT4,PLSCR1,PSMB9,RTP4,SP110,STAT1,TDRD7,TLR2,TMEM140,TRIM22,UBE2L6,ZC3HAV1 |
| PTEN | 0,319 | phosphatase | | 0,702 | 0,680 | -0,002 | 1,77E-10 | 1,18E-08 | ABCC4,ACAA1,ACACA,ADIPOQ,ADM,AGFG1,ALOX5AP,ANXA1,ARF4,ATF3,AURKA,BAX,BTG2,C3,CA2,CA8,CADM3,CASP7,CCNB1,CCNB2,CCNC,CD44,CDC20,CDK4,CDKN1B,CFD,CITED2,CKS2,CST6,CTSV,CTSZ,CX3CR1,DENR,DHRS11,DNPH1,DUSP1,DYNC1LI2,ECI2,EED,EGF,ESR1,F3,FAS,FGF1,FGF5,FN3K,FOS,FOXO1,FZD6,G0S2,GALK1,GMDS,GP9,GPR12,GPR162,GTF2I,HADH,HCLS1,HEXB,HGFAC,HIF1A,HLA-DMA,HOXA10,IDH3B,IDI1,IFI30,IFNGR2,IFRD1,ISG15,JUND,KAT2A,KIAA0930,KIF3A,KRT19,LEPROT,LHPP,LMX1B,LRRFIP1,MACROH2A1,MAD2L1,MAPK8,MCAM,MET,MFAP3L,MME,MYBL2,MYO6,NDRG1,NEU1,NFIL3,NFKB1,NKX2-5,NME1,NOD1,NOS2,NPPA,NR2F2,NTSR2,PAPOLA,PARP1,PCBP2,PDCD4,PDHA1,PECAM1,PIM1,PITX3,PKLR,PLOD1,PPARG,PPP1R17,PRL,PTEN,PTP4A2,PTPRC,PTPRF,PTPRK,RAB38,RB1,RMI1,RNASEH1,RRM1,SEC23A,SERINC3,SMAD1,SMAD2,SMARCA4,SNAI1,SNRPF,SOAT1,SOX4,SPP1,SPRY2,SQOR,SRSF10,STARD8,STAT3,STAT5A,STEAP1,SUCLG1,SYNCRIP,TNFRSF10D,TNFRSF11B,TNFSF10,TP53,TP53BP2,TRAM1,TSC22D3,TSPO2,VCAM1,VEGFA,XRCC5 |

| | | | | | | | | | |
|---|---|---|---|---|---|---|---|---|---|
| NR4A1 | -1,162 | ligand-dependent nuclear receptor | | 0,916 | 0,910 | -0,001 | 1,85E-10 | 1,22E-08 | ACACA,ACO1,ADIPOQ,ADM,AKR1C1/AKR1C2,ALDOA,ALDOB,APOB,APOD,ATF3,CAV3,CDKN1B,COL1A1,COL1A2,CRABP2,DDIT3,ELK3,ENO1,ENO3,EPCAM,FBP1,FOXP3,G0S2,G6PC,GCK,GPD1,GPI,GSK3A,GSTM5,HDAC1,HDAC6,HIF1A,HMOX1,HSD3B2,IL13,INS,ITGAE,KITLG,LDLR,LEP,LIPC,LPL,MAPK8,MARCKSL1,MMP7,MPZ,NDUFS6,NFKBIA,NR4A3,NUCB2,PCK1,PCK2,PCNA,PDCD1,PDHA1,PDIA3,PDK4,PHKB,PKLR,PPA1,PPARG,PPP1R3C,PRL,PTGDS,PTK2,PYGM,RPIA,RXRA,SLC2A4,SOAT1,SPINT2,SUCLG1,TALDO1,TNFSF10,UBE2C,UCP2,VCAM1 |
| EDN1 | 0,125 | cytokine | | -0,083 | 0,350 | 0,063 | 1,90E-10 | 1,24E-08 | ADIPOQ,ANXA1,ANXA4,ANXA5,AVP,BAG2,CCN2,CDK1,CDKN1B,COL1A2,COL4A1,CYBB,CYP11B2,DCT,EGR1,EIF3J,ENAH,FN1,FOS,FOSB,FOSL1,GAST,GRN,HAND1,HBEGF,HIF1A,HSPA5,IL18,INSR,ITGAV,ITGB3,IVL,JUNB,LEP,LEPR,MARCKSL1,MCAM,MITF,MMP1,MMP7,MPRIP,MSN,MYL6,NOX5,NPPA,ODC1,PDIA6,PLCB4,PLOD1,PPARA,PRKCA,PRKCD,PRL,RAB3A,SLC1A2,TIMP1,TNC,TP53,VCAM1,VCAN,VEGFA,VIM,YWHAE |
| CEBPB | -0,173 | transcription regulator | | 0,689 | 1,182 | 0,052 | 2,10E-10 | 1,37E-08 | ADIPOQ,ADM,ALB,ALDH1A1,ALOX5AP,APLNR,APOB,APOC3,APP,ASNS,ATR,BHMT,BLNK,C3,CCNA2,CCNB2,CDC25B,CDH3,CDKN1B,CFD,COL1A1,COL1A2,COL5A2,CSF3,CYB5A,CYP24A1,CYP27B1,CYP2A6 (includes others),CYP3A4,DDB2,DDIT3,DHX9,DMWD,DYNLT1,EPHA7,FAS,FHL1,FHL2,FKBP6,FOS,FZD1,G6PC,GADD45A,GNAS,GSTA1,H4C4,HBB,HLA-A,HPX,HSD17B4,HSD3B2,IDH3B,IGFBP4,IL7,INS,INSR,IRF9,IVL,LAMC1,LCN2,LDLR,LEP,LEPR,LY96,LYN,MAPT,MCM3,MMP1,MSH2,MYBL2,NOS2,NRP1,ODC1,PCK1,PCNA,PDK4,PEA15,PLG,PPARA,PPARG,PPP1R15A,PRG2,PRKCD,PRL,PSMA3,PSMA6,RAC2,RBBP7,RORA,SEC11A,SGK1,SLC10A1,SLC12A2,SMAD2,SPP1,SQOR,SRP54,SST,STAT3,TMEM50B,TP53,USP33,VCAN,VDR,VIM,VLDLR,XDH,ZNF638 |
| TCF3 | | transcription regulator | | -1,039 | -0,803 | 0,036 | 2,15E-10 | 1,39E-08 | ABCC8,ACACB,ADIPOQ,ARHGEF18,ATF3,ATOH1,AURKA,AZGP1,BID,BLNK,BTNL2,CA2,CASP3,CASP6,CCNA2,CCNB1,CCNB2,CD5,CDC20,CDC25B,CDK1,CDKN3,CKAP4,CKS2,COL1A1,CRELD2,CTSV,DCK,DDX58,DSTN,E2F4,EAF2,ECT2,EPB42,EPCAM,FOXO1,FOXP3,GADD45A,GADD45B,GM2A,GYPA,HAND1,HMGB2,HS3ST1,HSD17B14,HSP90B1,IFI16,INS,INTS13,JUND,KCNN4,LEPROTL1,MAD2L1,MANF,MYOD1,MYOG,NDC80,NEIL1,NELL1,NFIL3,NPPA,NR0B2,NR4A3,PAFAH1B3,PARP8,PDIA4,PDIA6,PFKP,PRC1,PRDX4,PRSS3,PTCRA,PTPRU,RGS16,RORA,RORC,RRM2,SCN4A,SDF2L1,SEC24D,SEMA3G,SERPINB1,SLC12A2,SMAGP,SUB1,SULT2B1,TCEAL9,TMEM184B,TYMS,ZBP1,ZP2 |
| Rxr | | group | | -0,678 | -0,616 | 0,018 | 2,80E-10 | 1,78E-08 | ABCA1,ADAM10,ALAS1,BMP7,BTG2,C1QA,CEBPE,CRABP1,CSF3,CSHL1,CTSS,CYP24A1,CYP26A1,CYP2B6,CYP2C8,CYP3A4,CYP3A5,DIO1,EMX1,FABP6,FURIN,GADD45A,GSTA1,HBEGF,HNF1B,HR,INSR,ITGB2,LEP,LTF,MMP1,NOS2,NR0B2,PCK1,PDK4,PLTP,PPARG,PTPRO,RXRA,SEMA4D,SLC10A1,SPP1,TERT |
| bisphenol A | | chemical - endogenous mammalian | | 1,408 | 1,715 | 0,046 | 2,82E-10 | 1,78E-08 | ADIPOQ,ALAS1,APOB,AQP5,BAX,BECN1,C3,CASP3,CCN2,CDK4,CDKN1B,CFD,CITED2,CLU,CTSD,DNM1L,EGR1,ELAC2,ENO1,ESR1,F11R,FABP2,FOS,GHRH,HOXA10,HSP90B1,LCN2,LEP,NMD3,NR4A1,NR4A3,PCNA,PICALM,PKM,POSTN,PPARG,PPP1R10,PRL,RBM22,RBM4B,RBPMS,RPH3A,RPN2,SLC22A6,SLC2A4,SLC6A4,SLFN12,SRD5A2,STAT3,SYN1,SYNCRIP,TP53,TRA2A,TWF2 |
| Ifnar | | group | Activated | 4,755 | 5,206 | 0,075 | 2,88E-10 | 1,80E-08 | APOBEC3B,B2M,C3,CASP1,CCL22,CD74,DDIT3,DDX58,FCER1G,GBP2,HLA-A,HLA-E,HLA-G,IFI16,IFI35,IFIH1,IFIT2,IFIT3,IFNGR1,IL15,IRF7,IRF8,IRF9,ISG15,OAS1,PSMB8,PSMB9,SPP1,STAT1,TAP1,TAPBP,TLR2,TLR3,TNFSF10,TRIM21,UBE2L6,VCAM1,ZBP1 |
| IL5 | | cytokine | | 0,626 | 1,299 | 0,076 | 2,95E-10 | 1,83E-08 | A2M,ALDOA,ALDOC,ANXA2,ASNS,ASS1,BIRC2,BNIP3L,CASP4,CCL22,CD24,CD5,CD63,CDC123,CISH,CITED2,CKAP4,CRELD2,CX3CR1,EAF2,EGR1,EGR3,ELL2,ENO1,FAS,FGL2,GADD45A,GADD45G,GALK1,GBP2,GK,HBEGF,HIF1A,HSP90B1,HSPA5,HSPA6,IDI1,IER2,IFI30,IL13,IL13RA1,IL6R,IRF7,ITGAV,ITGB6,LTA,LTB,LUM,MANF,MMD,NDRG1,NFIL3,NOS2,P4HA1,PDCD2,PDIA6,PDLIM1,PFKP,PIM1,PKM,PPIF,PRDX4,PRRC2A,PTPRC,PYCR1,RBPMS,S100A4,S100A6,SDF2L1,SEMA4A,SGK1,SNAP23,SOCS2,SRI,STK39,TLR1,TRA2A,UCHL3,UPB1,VIM,ZBTB32 |

| | | | | | | | | | |
|---|---|---|---|---|---|---|---|---|---|
| NRAS | 0,370 | enzyme | Inhibited | -2,180 | -2,367 | -0,030 | 3,15E-10 | 1,94E-08 | AGRN,AKAP12,ANXA4,B2M,BAX,BUB3,CCNB1,CCNB2,CD48,CDC20,CDKN3,COL1A2,CRABP2,CX3CR1,CYBB,DLX5,DUSP2,EGR1,ELK1,EPHX1,FHL1,FN1,GBP2,HIF1A,HLA-A,HOMER3,HTRA1,IFI16,IFI35,IFIH1,IL13RA1,ISG15,KRAS,LCN2,LGALS1,MCM3,MEST,MITF,MYBL2,NCF2,NPPA,NRAS,P3H4,PERP,PLTP,PSMB8,RBPMS,RHOB,RNASE6,SARDH,SERPINE2,SLC2A4,STAT1,STAT3,TAP1,TMEM43,UBE2C,VEGFA,WEE1,XPNPEP2,YIF1A |
| FAS | 0,557 | transmembrane receptor | | -1,068 | -1,096 | -0,005 | 3,73E-10 | 2,20E-08 | ANXA2,ARAP2,ARHGEF1,BAX,BID,BIRC2,BTN3A3,CAMK2G,CASP3,CASP8,CAST,CCL20,CCN2,CD44,CD48,CD59,CD83,CDIPT,CDKN1B,CEACAM4,CLTB,COL15A1,COL18A1,COL1A1,COL1A2,COL3A1,COL4A1,COL4A2,COL6A3,CXCL6,DGKZ,EGR1,F2RL1,F3,FADD,FAS,FBN1,FGL2,FOS,FOSB,FOXO1,GADD45G,GBP1,GBP2,GDE1,GNG7,GRIA3,HLA-DQB1,HLA-F,IL10RA,IL13RA1,IL18,ITGA7,ITGAV,ITGB8,JUND,LDLR,LTB,LUM,LY75,MAP4K4,MET,MME,MMP1,MPZL1,NFKBIA,NFKBIE,NOS2,NR4A1,P2RX4,PAK2,PECAM1,PLD2,PPP1R15A,PTP4A2,PVALB,RASA1,SHC1,SIT1,SLPI,SOD2,SPP1,STAMBP,STK38,TLN2,TLR2,TNC,TNFRSF11B,TNFSF10,TP53BP1,TPM1,TWF2,TYK2,UGCG,VCAM1,VEGFA,VRK2,ZFP36 |
| ERK | | group | | 0,791 | 1,168 | 0,050 | 4,89E-10 | 2,84E-08 | ABCA1,ACTN1,ADAM10,APOBEC3G,ARPC2,BAX,BECN1,BTG2,CCN2,CDK4,CDKN1B,CGA,CKS2,CLU,COL1A1,COL3A1,CXCL6,DIO3,DPYSL2,DUSP1,E2F4,EGR1,ESR1,ESRRA,FAS,FN1,FOS,FOSL1,FOXO1,FOXP3,GDF9,GK,HBEGF,HMOX1,IFNAR2,ITGAV,ITGB2,ITGB3,JUNB,KYNU,LAMB1,LCN2,LDLR,MAFF,MAP4K4,MCAM,MMP1,MUC1,NDRG1,NFE2L2,ODC1,OSBPL10,PDK4,PECAM1,PIWIL1,ROCK1,RORC,SFN,SOX4,STAT1,STMN4,TBX21,TERT,TIMP1,TNFAIP8,TP53,TUBA1A,UGCG,VCAN,VEGFA,ZFP36 |
| Interferon alpha | | group | Activated | 4,948 | 5,427 | 0,051 | 5,19E-10 | 3,00E-08 | ADA2,ADAR,AFP,ANXA5,APOBEC3B,APOBEC3F,APOBEC3G,ATF3,B2M,BTG2,C3,CASP1,CASP3,CASP8,CCL22,CD70,CD81,CD83,CDKN1B,CISH,CLTCL1,CTH,CYP3A4,DDX58,DEFB1,DYNLT1,E2F4,EMID1,F3,FAS,FOS,GBP1,GBP2,GLI1,HERC6,HLA-A,HLA-B,HLA-C,HLA-F,HLA-G,IFI16,IFI35,IFI44,IFIH1,IFIT2,IFIT3,IFITM1,IFITM2,IFNAR2,IGFBP4,IL10RA,IL13,IL15,IL32,IL6R,IL7,INSR,IRF7,IRF8,IRF9,ISG15,JAK1,MX1,MYLIP,NCR3,NFIL3,NFKBIA,NMI,NOS2,OAS1,PARP12,PHF11,PIM1,PLAAT4,PLSCR1,PRL,PSMB8,PSMB9,RB1,RBX1,RTP4,SERPINB1,SLC6A4,SOCS2,SP110,SPRY1,SPRY2,SPRY4,STAT1,STAT3,TAP1,TAPBP,TBX21,TDRD7,TERT,TGM1,TLR1,TLR2,TLR3,TMEM140,TNFSF10,TRIM21,TRIM22,TRIM5,TRIOBP,TYMS,UBE2L6,VCAM1,WARS1,ZBP1,ZC3HAV1 |
| HIF1A | 0,522 | transcription regulator | | 0,234 | 0,742 | 0,056 | 6,04E-10 | 3,46E-08 | ABCC8,ACO1,ADAM10,ADAM15,ADAMTS1,ADIPOQ,ADM,AFP,AKAP12,ALDOA,ALDOC,ALOX5AP,ARPC2,ASNS,AURKA,BAX,BHLHE40,BHLHE41,BID,BIRC2,BNIP3L,CA9,CCN2,CD24,CD44,CD70,CDKN1B,CITED2,CLDN1,CTSD,CYB5A,EED,EIF4G1,ENO1,ESRRG,FHL1,FHL2,FHL3,FN1,FOS,FURIN,FUT7,GADD45B,GATA6,GCK,GHR,GPI,H1-2,H2BC12,HIF1A,HMOX1,HSPA4,HSPA5,IL15,ITGAV,ITGB2,ITGB3,KITLG,KLHL20,KRT19,LEP,LGALS1,MAFF,MANF,MB,MET,MITF,MMP1,MMP7,MST1R,MUC1,NAE1,NBN,NDRG1,NOS2,NPPA,NR4A1,NR4A3,P4HA1,PDHA1,PKM,PLOD2,PPARA,PRKCA,PROM1,PTEN,RORC,SIRT2,SLC2A4,SNAI1,SOD2,SSBP1,STAT3,STC2,TERT,TLR2,TLR6,TMPRSS6,TNFRSF11B,TP53,TRIM21,VCAM1,VEGFA,VIM,WNT10B,WT1,ZNF267 |

| | | | | | | | | | |
|---|---|---|---|---|---|---|---|---|---|
| APP | 0,227 | other | Activated | 1,961 | 2,233 | 0,028 | 6,18E-10 | 3,52E-08 | ABCA1,ABCG1,ABCG2,ABL1,ACTG1,ALB,ALDOA,ALDOC,AMPH,ANXA5,APP,ARC,ARF4,ARL6IP5,ASCC1,ATF1,ATP1A3,ATP4A,AXIN1,BAX,C1R,C1S,C3,CALML3,CANX,CAP1,CAPN1,CASP3,CAST,CAT,CCL20,CCN2,CD151,CD44,CD59,CD74,CDH3,CDK1,CDK4,CITED2,CLDN1,CLIC1,CLTB,CLU,COL18A1,CPNE6,CRABP1,CRABP2,CRK,CRYL1,CSF3,CSNK2A1,CTSD,CTSV,CXCL6,CYBB,CYFIP1,CYFIP2,DBI,DBN1,DDX58,DNM1L,DPYSL2,DRD2,DSTN,EGR1,ELN,ENO1,ERMP1,ESR1,EXT1,F2RL1,FKBP1A,FN1,FOS,FOSB,FUZ,FZD1,GALK1,GBP2,GHR,GNB1,GPI,GSTP1,HAP1,HARS1,HDAC1,HECW1,HERC6,HK1,HLA-E,HMOX1,HSPA5,IFI16,IFI35,IFIH1,IFIT2,IL13,IL15,IL18,IL7,INA,INSR,IRF7,ITGA6,ITGB3,JUNB,JUND,KAT2A,KIF2A,KLK3,KPNB1,KYNU,LDLR,MAFF,MAPK1,MAPT,MBD4,MDK,MGAT3,MME,MMP1,MXI1,NAMPT,NDUFS6,NES,NEUROG1,NFKB1,NME1,NNAT,NOS1,NOS2,NPTX1,NRP1,NTF3,PAK3,PDCD4,PDK2,PEA15,PFKP,PITX3,PJA1,PKM,PPARG,PRKCA,PSEN1,PSME1,PTCH2,PTGDS,PTPRC,PVALB,RAN,RFC4,RHOA,ROCK1,RTP4,SET,SH3GL2,SHC1,SLC13A3,SLC1A2,SLIT2,SMARCA4,SOD2,SQSTM1,STAT3,STMN1,STUB1,SUCLG1,SYN1,TAF9,TERT,TLR2,TLR3,TNFSF10,TOP2B,TOR3A,TP53,TP73,TPD52L1,TPM1,TRIM21,TUBA1A,TUBA1B,TUBB,TUBB6,UBE2K,UCHL1,VAMP2,VCAM1,VCAN,VEG[illegible] |
| AR | -0,026 | ligand-dependent nuclear receptor | | 0,044 | 0,489 | 0,050 | 6,86E-10 | 3,88E-08 | ABCA1,ABCC4,ABCG1,ACTR3,ADGRA3,ADM,ALDH1A1,ALOX15B,AMHR2,AQP8,ATP1A3,ATP4A,AZGP1,C3,CA5B,CA7,CADM3,CALU,CAMK2A,CASQ2,CAST,CAV2,CCDC70,CCNA2,CCT3,CDC25B,CDK14,CEND1,CGA,CIT,CKAP4,CLCN1,CLDN11,CLDN3,COL3A1,CTSO,DBI,DCTPP1,DHCR24,DNAJB9,DNM1L,DR1,DSG2,EGR1,EPHX1,EPPIN,ESPL1,FXYD3,GAS6,GDF9,GPD1,GPX5,GSTP1,GUCY1A1,HOPX,HSD3B2,HSPA5,IMPDH2,INHA,ITGA6,KDELR2,KITLG,KLK3,KPNA2,MAD2L1,MCM2,MME,MRAS,MT1X,MTHFD2,MUC1,MYOD1,MYOG,NDRG1,ODC1,P4HA1,PBK,PCSK1N,PDIA5,PDYN,PER1,PGC,PIGR,PIP,PLOD2,PLS3,PRKCA,PRKCD,PROS1,PTEN,PTGDS,RARRES1,RHBG,RHOB,SCN4A,SEC61B,SERPINA3,SGK1,SHBG,SLC22A4,SLC43A1,SNAI1,SPP1,SRD5A2,STAT1,STK39,TERT,TMPRSS2,TNFSF10,TNNT2,TPD52L1,TSPAN8,UBE2C,VCAN,VEGFA,VIM,WEE1,WT1,ZBP1,ZWINT |
| OGA | 0,233 | enzyme | | 0,775 | 0,699 | -0,007 | 7,23E-10 | 4,06E-08 | ABLIM1,ACTG1,ADIPOQ,ALDOC,ALOX5AP,APOBEC3G,ARHGAP29,ASS1,AZGP1,BAX,C1QA,C1QB,C1RL,C6orf15,CAPN1,CASP1,CASQ2,CAVIN2,CCL20,CD151,CD24,CD44,CD47,CD48,CD53,CDH15,CIAPIN1,CLDN1,COLEC11,CRABP1,CX3CR1,CXCL6,CYFIP2,CYP2B6,DPEP1,EFNB1,ELK1,EPHB3,FAS,FBP1,FERMT2,FGF5,FGFBP1,FN1,FSCN3,FZD6,GABRA2,GADD45A,GADD45G,GALNT7,GAS6,GLI3,GLUL,GP9,GPR21,GTF2IRD1,HPGD,IFIT3,IFNGR2,IGFBP4,IL18,IL6R,ITPKB,JAK1,KRT81,LAMB1,LEP,LIPC,MCAM,MGLL,NDUFA2,NOTCH2,NPHS1,PDCD2,PDHA1,PHKA2,PHKB,PLCE1,PLIN3,PRDX4,PRKAR1B,PROM1,PTGDS,RAB27A,RB1,RHOB,S100A2,S100A6,SERPINE2,SOCS2,SRPK3,ST14,STAT1,TBP,TGFBI,TGFBR3,TIMP1,TNC,TNFRSF11B,TP53,TP53BP1,TP73,TPD52L1,TUBA1A,VIM,WNT11 |
| LEP | -0,132 | growth factor | | -1,740 | -1,437 | 0,032 | 7,55E-10 | 4,19E-08 | ABCC3,ACACA,ACAT1,ACO1,ADIPOQ,AKR1C1/AKR1C2,ALDOB,APOA1,APOH,APOM,APP,ASS1,BAX,BID,CASP10,CASP3,CASP6,CASP7,CCNA2,CD63,CDKN1B,CEL,CFD,COL1A1,COL3A1,COL4A1,CSF3,CSHL1,CTSV,CYB5A,CYBA,CYBB,CYP24A1,CYP27B1,CYP2C8,CYP3A5,CYP4A11,DGKZ,DRD2,EGR1,ELOVL6,ESR1,F3,FAAH,FADD,FAS,FDX1,FOS,FOXP3,G6PC,GADD45A,GCK,GNAS,GSTA3,HPX,HSPA5,IL13,INS,JUNB,JUND,KIF3A,LDLR,LEP,LEPR,LIPA,LIPC,LPL,MLXIPL,MMP1,MMP7,MUC5B,MYOD1,MYOG,NAMPT,NCF2,NEUROG1,NOS1,NOS2,NPPA,P2RX4,PCK1,PCNA,PECAM1,PER2,PNPLA2,PPARA,PPARG,PPP1CA,PRL,RAC1,RORC,SLC13A2,SLC2A4,SOD2,SPP1,SST,STAT3,SYN1,TERT,TIMP1,TMED9,TNFRSF11B,TNFSF10,TP53,TRPV1,TSPO,TTPA,UCP2,VCAM1,VEGFA,VIM,ZFP36 |

| | | | | | | | | | |
|---|---|---|---|---|---|---|---|---|---|
| Growth hormone | | group | Inhibited | -2,565 | -2,344 | 0,033 | 7,88E-10 | 4,34E-08 | ACAA1,ACACA,ADIPOQ,AKR1B1,ALB,APLP1,BMP7,BPHL,C3,CAT,CDH3,CISH,CITED2,CLDN15,CLU,COL18A1,COL1A1,COL3A1,COMT,CSHL1,CTH,CYP27B1,CYP2C8,CYP3A4,DDIT3,DDR1,DYRK2,EGR1,FN1,FOS,FZD5,G6PC,GHR,GLUD1,GSTA1,GSTM5,GSTP1,HADH,HBB,IGFALS,INSR,JUNB,LDLR,LEPR,LPL,MYOD1,NAMPT,NFKBIA,NPPA,PCK1,PCNA,PDK4,PLK2,PPARA,PPARG,PPP1R15A,PRL,RAB3A,RPH3A,SGK1,SOCS2,SPP1,SST,STAT5A,TKT,TNFSF10,TUBA1A,XDH |
| MKNK1 | -0,007 | kinase | | 0,056 | 0,552 | 0,080 | 8,40E-10 | 4,59E-08 | AGRN,ANXA2,ANXA5,ARVCF,ATAT1,ATP1A3,BMP8B,CAMK2A,CRMP1,EPB41L2,FOS,FOSB,GNAI1,GPC4,GPS1,HSD17B4,IFIT2,IFRD1,ISG15,KIF21B,LAMB2,LAMC1,LCN2,MAN2A1,MAPK1,MAPT,MYO6,NELL2,NR4A3,PLTP,PRKAR1B,RAB3A,SMAD2,TM9SF2,TP53BP1,TRIM32,USO1,VAMP2,VIM |
| hydrogen peroxide | | chemical - endogenous mammalian | | 0,562 | 1,008 | 0,044 | 1,18E-09 | 6,30E-08 | ADAM9,ADIPOQ,AKR1C1/AKR1C2,ALOX5,ANXA2,APP,ATF3,AURKA,BAX,BTG2,CASP1,CASP3,CAT,CBX5,CCN2,CCNA2,CCNB1,CD59,CD63,CD81,CDK1,CDKN1B,CFD,CLU,COL1A1,COL1A2,CRYAB,CSF3,CTSD,CTSS,CXCL6,DDIT3,DNMT1,DPAGT1,DUSP1,DUSP2,E2F4,EGR1,ELN,F2RL1,FAS,FDXR,FN1,FOS,FOSL1,FOXO4,GADD45A,GADD45B,GLI1,GLRX,GSTA1,HBEGF,HIF1A,HLA-DRA,HMOX1,HRAS,HSPA5,IFIT2,IL10RB,IL13,IL32,ITGA6,ITGB3,KITLG,KRAS,LCN2,LDLR,MAPK8,MB,MCAM,MCM2,MDK,MITF,MMP1,MUC1,NAT10,NDC80,NFE2L2,NFIL3,NGF,NOS1,NOS2,NOX5,NR4A1,ODC1,PAH,PARP1,PCBP2,PDCD10,PDCD4,PDIA3,PDIA4,PIM1,PPARG,PPM1D,PPP1CA,PSMC4,PSTPIP2,PTEN,PTP4A2,RAB27A,RAC1,RAN,RBMX2,RTN3,SGK1,SHC1,SNAI1,SOD2,SPP1,STAT3,STMN1,SUV39H1,TAP1,TGFBR3,TP53,TP73,UCP2,VCAM1,VDR,VEGFA,VIM |
| HTT | | transcription regulator | Activated | 2,639 | 2,741 | 0,016 | 1,30E-09 | 6,82E-08 | ABCA1,ABCC10,ADRA2B,AGRN,AKAP12,ALDOA,ALDOC,APOA1,APP,ATF3,ATP1A3,AVP,AZIN1,B2M,BAIAP2,BCAP31,BHLHE40,BID,BIRC2,BMP3,C1R,CAMK2A,CANX,CASP1,CASP3,CAST,CAVIN3,CBX5,CCN2,CD74,CDHR5,CDK2AP1,CHRNB2,CIT,CITED2,CLCN1,CLDN1,CNR2,COL18A1,COL4A1,COL4A2,COL6A3,COX6A2,CRYAB,CTSD,CYFIP2,DDIT3,DDX50,DLG3,DNAI1,DNAJC3,DRD2,DRD5,EGR1,EMC8,EMP3,ENO3,FGF1,FHL1,FN1,FOS,FOSL1,GAD1,GADD45G,GHRH,GJD2,GLUL,GNG7,GPI,HAP1,HBEGF,HIF1A,HMGB2,HOXB1,HRAS,HSPA5,IFI16,IL15,JUNB,KAT2A,KCNC3,KLF9,LDLR,LTBP2,LTF,MAST3,MCAM,MICU1,MLF2,MPRIP,MTNR1A,MYOD1,NAA15,NAB2,NDUFB2,NFE2L2,NFIL3,NOS1,NOS2,NPTX1,NR2F1,NR4A1,NSF,PCNA,PDE10A,PDE1B,PDK2,PEA15,PER1,PER2,PFN1,PIGF,PLOD3,PPARA,PPARG,PPP1R16B,PPP1R1A,PRELP,PRKACB,PRKCD,PROM1,PSMA5,PSMB8,PSMB9,PSME1,PVALB,RGS16,RNF34,RPH3A,SERPINA3,SERPING1,SGK1,SIRT3,SLC12A2,SLC1A2,SNN,SOCS2,SOD2,SPP1,SPTBN2,SQSTM1,SST,STAU1,STMN1,STX4,SYN1,TBX6,TESC,TGFBI,THBS2,THRB,TP53,TUBB,TYRO3,UCHL1,UCHL3,USP2,VCAM1 |
| VEGFA | -0,459 | growth factor | | -0,611 | -0,099 | 0,060 | 1,73E-09 | 8,97E-08 | ABCC1,ADAM15,ADAMTS1,ALAS1,ALDH2,ANPEP,BAX,BCAT2,CASP3,CBFB,CCN2,CD1A,CD83,CDKN1B,CITED2,COL1A1,CSF3,CSTB,CTLA4,CTSS,CTSV,CXCL6,EGR1,ERAP1,ESM1,F3,FN1,FOS,FOSB,GBP1,GRB10,HBEGF,HIF1A,HK1,HLA-DQB1,HMOX1,HSPA5,IGFBP4,ITGAV,ITGB3,JUNB,LDB1,MMP1,MRPL3,MTIF2,MVP,NME1,NOS2,NOTCH4,NPHS1,NR2F2,NR4A1,NRGN,NRP1,PARP1,PCNA,PDCD1,PECAM1,PIM1,PNPLA2,PPA1,PRKCA,PSEN1,PTK2,RAB5A,SET,SLC12A6,SNCG,SOD2,ST5,STARD8,TERT,TFPI2,TIMP1,TLR1,TMSB10/TMSB4X,TNC,TOMM70,TP53,TSPO,VCAM1,VEGFA |
| APOE | -0,217 | transporter | | -0,818 | -1,099 | -0,046 | 1,92E-09 | 9,92E-08 | ABCA1,ACAT1,ADAM10,ADGRB1,ADGRG1,ADIPOQ,ADNP,ALOX12,ANXA11,APOA1,APOB,APOD,APP,ATF3,BAX,BEGAIN,C1QA,C1QB,C2,CAPNS1,CASP1,CASP3,CAT,CCL22,CCN2,CD44,CLU,COL18A1,COL1A1,CREB3L1,CTSD,CTSS,CTSV,CYBB,CYP3A5,DCSTAMP,DDIT3,EGR1,ESR1,F2RL1,F3,FOS,GJD2,GLUL,GRIA3,HMOX1,HSP90B1,HSPA5,IL10RA,IL10RB,INSR,JAK1,LCN2,LDLR,LEP,LIMS2,LIPA,LPL,LTBP2,M6PR,MAP2K3,MAP3K1,MAPT,MYLIP,NCF2,NELL2,NMNAT2,NOS2,OGA,P4HB,PEA15,PHF21A,PPARG,PPP1R15A,PSMC4,SEMA6B,SERPINA3,SLC25A44,SMAD1,SOD2,SPP1,TCIRG1,TIMP1,UCP2,VCAM1,WBP2,WNT2 |

| | | | | | | | | | |
|---|---|---|---|---|---|---|---|---|---|
| RNA polymerase II | | complex | | -0,005 | 0,152 | 0,080 | 2,05E-09 | 1,05E-07 | ADIPOQ,ALAS1,ALPL,APOC3,APOM,ASNS,B2M,BLNK,CAP2,CASP7,CBX5,CCNB1,CCNB2,CD83,CLDN3,COL1A2,CSDC2,CSHL1,CYP24A1,DCTPP1,DDIT3,DIO1,DLGAP5,DUSP1,ENO1,ESR1,ESRRA,EXOSC8,FOS,FOSL1,FOXC2,GADD45A,GADD45B,GAST,GBP2,GHR,HBB,HNF1A,HNF4A,HPRT1,HSD11B2,IFNGR1,INS,JUNB,KITLG,KPNA2,LDLR,MAD2L1,MAPK1,MAPK8,MCM3,MYBL2,MYOG,NFKBIA,NMD3,NOS2,PCK1,PCNA,PKLR,PPARG,PSMB9,PTK2,RAD50,RORC,RPL13A,SFN,SOD2,SPP1,TAP1,TAPBP,TERT,THBS2,TNNT2,WNT7B,ZNF268,ZNF83 |
| PI3K (complex) | | complex | | 0,717 | 1,116 | 0,055 | 2,07E-09 | 1,05E-07 | ABCA1,ABCC4,ADIPOR1,ALOX5AP,ATF3,ATG5,ATOH1,CASP3,CCN2,CCNB1,CCT6A,CD44,CDK4,CDKN1B,CNR2,COL1A1,CTCF,CTSD,CYBB,DDIT3,DUSP1,E2F4,FN1,FOS,FOSL1,FOXO1,FRY,FUT7,GLI1,GSTA1,GSTM5,GSTP1,HBEGF,HIF1A,HK1,HMOX1,INHA,INS,ITGA6,ITGB2,KRAS,LASP1,LCN2,LEFTY2,LEP,LTBP2,MCAM,MITF,MMP1,MTHFD2,MYOD1,NFE2L2,NFKBIA,NME1,NOS2,NPPA,NRAS,PDCD4,PIM1,PMEL,POSTN,PTPN12,RGS2,RHOB,SEMA6D,SGK1,SLC12A6,SLC5A5,SNAI1,SNCG,SOAT1,SPP1,SST,STMN1,TNFSF10,TP53,UGCG,VCAM1,VEGFA,WT1 |
| BDNF | -0,043 | growth factor | Inhibited | -3,838 | -3,585 | 0,032 | 2,36E-09 | 1,19E-07 | AGRN,ALDH7A1,ANXA2,ANXA5,APP,ARC,ARVCF,ATAT1,ATP1A3,BHLHE40,BID,BMP7,CAMK2A,CASP3,CAV2,CRMP1,DNAJB5,DRD2,DUSP1,EGR1,ELOVL6,EPB41L2,FOS,FOSB,GLUL,GNAI1,GPC4,GPS1,HSD17B4,HSPA5,IFRD1,KIF1A,KIF21B,LAMB2,LAMC1,LCN2,LEP,LGALS1,LYPD1,MACROD1,MAN2A1,MAPK1,MAPT,MMP1,MYO6,MYOD1,MYOG,NAB2,NECAB3,NELL2,NOS1,NOS2,NR4A1,NR4A3,NTF3,P4HA1,PCNA,PER2,PLG,PLTP,PRKAR1B,PSEN1,PTEN,RAB3A,REEP5,RHOA,S100A10,SLC16A7,SLC17A7,SOD2,SPP1,SPRY2,SST,SYN1,TIPARP,TM9SF2,TNFRSF11B,TP53BP1,TRIM32,TUBA1A,USO1,VAMP2,VIM,WNT2 |
| CREBBP | | transcription regulator | | -1,447 | -1,135 | 0,053 | 2,55E-09 | 1,27E-07 | AKAP12,ALAS1,AMHR2,B2M,BMP7,CA2,CAMK2G,CAPG,CD44,CD53,CD59,CDKN1B,CGA,COL1A2,CRYBB2,CTSG,CYBB,CYP24A1,DHRS3,DUSP1,DUSP22,EGR1,EGR3,EPN1,EVI2A,FDX1,FNDC3B,FOS,FOSB,FUT7,G6PC,GHRH,GLI1,GM2A,GNAS,GRIA3,HBB,HLA-B,HLA-DRA,HLA-G,HSD3B2,IL10RA,INHA,ISG15,ITGA6,ITGA9,JUND,KLK3,LDLR,LGALS3BP,LPL,LTA,MCM3,MFHAS1,MPZ,MSH2,MYOD1,NDRG1,NFE2L2,NFKBIA,NOS2,NPPA,NR4A1,NR4A3,NTRK3,PCK1,PCNA,PDE10A,PHIP,PIP5K1B,PKLR,PLEKHA1,PRKCD,PRL,PSMB9,PTPRF,RB1,RGS2,RHOA,RNF39,RTP4,SERPINA3,SERPINE2,SLC1A2,SLC43A1,SST,TAPBP,TBX4,TESC,TIPARP,TLE6,TLR2,TMPRSS2,TMPRSS4,TNC,TNFSF10,VCAM1,VEGFA,WNT8B |
| RAF1 | -0,028 | kinase | | -1,120 | -0,619 | 0,070 | 2,73E-09 | 1,36E-07 | AKAP12,ANKRD2,ANXA1,AP2B1,BECN1,CA2,CCN5,CD44,CD74,CDKN1B,CLDN1,CRYAB,CSTA,DHRS3,DUSP2,EGR1,EGR3,ELK1,EMP3,ESR1,FAS,FGFBP1,FOS,GRN,HBEGF,HIF1A,HMGB3,HNRNPAB,HSD11B2,IER2,INS,ITGB3,LHB,LPGAT1,MET,MGLL,MMP1,MYBL2,NFKB1,NGF,NPPA,OSMR,PCK1,PNP,PPP1R10,PRL,PRSS3,RAC1,RAN,RRM2,SELENBP1,SLC16A7,SLC26A2,SMAGP,SNRPD1,SPRY2,ST3GAL4,TBP,TERT,TFPI2,TIMP1,TNNI1,TSPOAP1,VEGFA |
| NOS2 | -0,309 | enzyme | | 0,664 | 0,733 | 0,009 | 2,95E-09 | 1,46E-07 | ACO1,ADIPOQ,ALDH3A1,APP,AVP,AZGP1,BAX,CASP3,CAV3,CD44,CD53,CFD,CHI3L1,COL4A1,COX6A2,FAS,FOXO1,FOXP3,G6PC,GPD1,GSPT2,HIF1A,HMGB2,HMOX1,IFIT3,IL32,IRAK1,ISG15,ITIH4,KRT4,LCN2,LEP,LGALS3BP,MAPK8,MB,MCM5,NFKBIA,NKX2-5,NOS2,NPPA,PCNA,PERP,PIGR,PLSCR1,PRL,SERPINA3,SFXN1,SLC2A4,SPRR3,TIMP1,TKT,TNFSF10,TNNC1,TNNT2,TOR3A,TSPYL2,TYROBP,VEGFA |
| MAPK8 | -0,221 | kinase | | -1,129 | -0,915 | 0,031 | 3,32E-09 | 1,63E-07 | ABCA1,APOA1,APOD,ATG5,B4GALT5,CASP3,CAT,CCN2,CCNA2,CDKN1B,COL1A1,CSTA,CYBB,DDIT3,DNMT1,DUSP1,EGF,EGR1,ELK1,FOS,FOSL1,FOXO1,FOXO4,FUS,G6PC,GADD45A,GCK,HDAC6,HMOX1,IL13,JUND,LEP,LHB,MMP1,MMP7,MUC5B,MVP,MYOG,NOS2,NPPA,NRP1,PTEN,RBP1,SH2D2A,SLC10A1,SOD2,TBP,TBX21,TLR2,TP53,TSC22D3,VDR,WNT6,ZFP36 |
| ACOX1 | -0,146 | enzyme | Inhibited | -2,150 | -2,252 | -0,015 | 3,40E-09 | 1,66E-07 | ACAA1,ADAMTS1,ANXA2,BAAT,CAT,CD63,CHRNB2,COL4A2,CRAT,CSTB,CTSV,CYP26A1,CYP2C8,CYP4A11,CYP4F12,CYP7B1,DDIT3,DIO1,DPP4,FABP2,G6PC,GADD45A,GNMT,GSTP1,HDAC1,HPD,HSPA5,LCN2,LGALS3BP,LHPP,LPCAT3,LPL,MGLL,NFE2L2,OAT,PAH,PDK4,PLSCR1,QPCT,RGS2,SELENBP1,SGK1,SLC37A4,SQSTM1,TP53,UCP2,UGCG,UGT8 |

| | | | | | | | | | |
|---|---|---|---|---|---|---|---|---|---|
| FGF19 | | growth factor | | -0,443 | -0,473 | -0,010 | 3,41E-09 | 1,66E-07 | ACACA,ACACB,AFP,ALDH1A1,AQP8,CYP7B1,EGR1,ENO3,FBP1,FOS,G0S2,GADD45A,GCK,HLA-DQB1,HSD3B2,LCN2,LEP,LEPR,MCAM,MME,NNAT,NR0B2,PKLR,PRL,PROM1,PTGDS,RGS16,RGS2,SERPINA3,SGK1,SLC10A2,SLC2A5,SLPI,TNNI1 |
| testosterone | | chemical - endogenous mammalian | | 1,543 | 1,873 | 0,037 | 3,71E-09 | 1,79E-07 | A2M,ADIPOQ,ADRA2B,ALDOA,ANPEP,ANXA1,ANXA3,AVP,BAX,BMP7,CASP3,CASP8,CAST,CAT,CCNC,CDK4,CDKN1B,CGA,CLCN3,CLU,COX6A2,CRK,CST8,CTSV,CYB5A,CYP7B1,DHCR24,EGF,EGR1,EIF4G3,ELOC,ESR1,FADD,FAS,FN1,FOS,FOSB,FOSL1,FOXO1,GADD45A,GDF9,GHRH,GMFB,GPX5,HIF1A,HSD3B2,JUND,KLK3,KRAS,LEP,LGALS1,LHB,MMP7,MX1,MYOD1,MYOG,NDRG1,NFKBIA,NGF,NOS1,NOS2,OAT,ODC1,PARP1,PCNA,PDIA4,PDYN,PPARA,PPARG,PSEN1,PSMA1,RAI14,RARG,SHBG,SLC25A10,SLC5A5,SMAD2,SOD2,SRD5A2,SST,SSTR2,SSTR3,STAT3,STX3,SYT11,TAC3,TNFRSF11B,TXNRD1,UGT8,VCAM1,VCAN,YWHAE |
| FOXA1 | 0,002 | transcription regulator | Inhibited | -3,082 | -3,121 | -0,007 | 4,43E-09 | 2,13E-07 | AFP,AGR2,ALDH6A1,ANXA1,BEST2,CD58,CDX2,COL18A1,CSHL1,DCT,DSCAM,DSE,ELK3,EPHX1,ESR1,FABP2,FNDC3B,FSTL1,G6PC,GBP2,GFRA4,HADH,HK1,HNF1A,HNF1B,HNF4A,KLK3,LHFPL6,LYN,MALT1,MLXIPL,MSH2,MUC5B,MUC6,NES,PAM,PCK1,PFKFB1,PPARA,PYY,RORA,RORC,S100A2,SLC12A2,SLPI,SPDEF,SST,TMPRSS2,TNNI1,WNT7B |
| PPARG | 0,186 | ligand-dependent nuclear receptor | | -1,003 | -0,791 | 0,023 | 4,69E-09 | 2,23E-07 | ABCA1,ABCG1,ABCG2,ACAA1,ACACA,ADIPOQ,ALOX12,ALOX15B,APOA1,APP,ATP4A,BAX,BECN1,C3,CA2,CAT,CAV2,CAVIN2,CCL1,CCL17,CCL22,CCN2,CFD,CIDEC,COL1A1,COL1A2,CRABP2,CRAT,CSF3,CXCL6,CYBA,CYP26A1,CYP4A11,DHRS3,DR1,ELOVL6,EPHX1,ESRRA,F11R,FABP1,FABP2,FGF1,FN1,FOSL1,FZD1,G6PC,GCK,GPD1,GSTA1,HMOX1,HR,IHH,IL13,IL25,INS,INSR,KLK11,KLK6,KRT19,LDLR,LEP,LHPP,LPL,ME1,MGLL,MUC1,MYOD1,MYOG,NDRG1,NFE2L2,NOS2,NPPA,NR2F2,NR4A1,NSF,ODC1,PCK1,PCK2,PCLO,PCNA,PDK4,PEA15,PECAM1,PEPD,PHKB,PLA2G3,PPARA,PPARG,PPIC,PTEN,PTPRF,RHOB,RORC,SGK1,SHBG,SLC27A3,SLC2A4,SNCG,SPP1,TBCC,TBXA2R,TKT,TNFSF10,TP53,TSC22D3,TUBB,UCP2,VAMP2,VAMP8,VCAM1,VDR,VEGFA,VIM,VLDLR,YWHAE |
| VCAN | 0,866 | other | | 0,698 | 0,744 | 0,007 | 4,97E-09 | 2,33E-07 | ADM,AGRN,ASS1,C1R,C1S,C3,CASP1,CASP3,CASQ2,CBX6,CCL20,CD59,CEACAM4,CLU,CXCL6,CYBA,ELN,F3,FAS,FN1,GLB1,GUCY1B1,IFI44,IFIT2,IFITM1,IGFBP4,IRF8,LAMB1,LCN2,MX1,MYH10,NFKB1,NFKBIA,PAM,PCSK1N,PRELP,PTK2,RBP1,RGS2,ROCK1,RXRA,SERPING1,SMPDL3B,SOD2,STAT1,TIMP1,TP53,TRIM22,VCAM1,VCAN,VEGFA |
| MED1 | | transcription regulator | | 1,032 | 1,387 | 0,064 | 4,97E-09 | 2,33E-07 | ACAA1,ACACA,ADIPOQ,APOB,AURKA,BAX,BCL7A,BHLHE40,CCN5,CCNB1,CDC20,CDK1,CDK4,CFD,CYP24A1,CYP2B6,CYP3A5,CYP4A11,DIO1,DTYMK,EDF1,FOXO1,GADD45A,GCK,GHR,GSTP1,HPRT1,HSD17B4,IRF8,KLK3,LBR,MAD2L1,MB,MET,MYOD1,NFKBIA,ODC1,PDK4,PIM1,PODXL,PPARG,PRDM14,SLC2A4,SMARCA4,STC2,TBX21,TERT,TNNC1,TNNI1,TP53,VDR |
| INS | -0,150 | other | | -0,635 | -0,561 | 0,012 | 5,65E-09 | 2,64E-07 | ACACA,ADAM10,ADIPOQ,ADIPOR1,ADM,ALDH3A2,APOA1,AVP,CAT,CCN2,CIDEC,CITED2,CRABP2,CSHL1,DDIT3,DUSP1,EGR1,ELK1,F3,FADS3,FAS,FOS,FOSB,FOSL1,FOXO1,G6PC,GABARAPL1,GCK,GRB10,HIF1A,HSPA5,IER2,IGFBP4,INSR,JUNB,JUND,LEP,LPL,NOS1,NR4A1,PCK1,PLK2,PPARG,RDH16,SLC2A4,SNAI1,TP53,UGCG,VEGFA,VLDLR |
| AKT1 | 0,070 | kinase | | 0,063 | 0,351 | 0,037 | 5,76E-09 | 2,67E-07 | ADIPOQ,AKR1C1/AKR1C2,ANKH,APOC3,ARL6IP1,ASS1,BAX,CASP3,CD74,CDK4,CDKN1B,CISH,CKS2,CLU,COL4A1,CTSV,CXCL6,EGR1,ENO1,ESR1,FAS,FN1,FOS,FOSB,FOXO1,FOXP3,FSTL1,G6PC,GATM,GCK,GSTM5,HIF1A,HMGB2,HMOX1,HNRNPM,JUND,KLF15,LAMB1,LCN2,LEP,LIPA,ME1,MEPE,MYH10,MYOG,NDC80,NFE2L2,NFKB1,NOS2,NPPA,PCK1,PDCD1,PKM,PLG,PPARA,PPARG,PRL,RAB11A,RFC4,RHOB,SEMA6D,SLC5A5,SNAI1,SPP1,STUB1,TGFBR3,THBS2,TIMP1,TNFSF10,TP53,TPD52L1,TYMS,UBE2C,UBE2L3,VCAN,VEGFA,VIM,ZWINT |
| linoleic acid | | chemical - endogenous mammalian | | -0,432 | -0,284 | 0,028 | 6,13E-09 | 2,81E-07 | ABCA1,APOA1,APOB,APP,CD47,CLU,CYP4F12,DDIT3,DGKZ,DUSP1,FABP1,G6PC,HSP90B1,HSPA5,LDLR,LHB,LPL,MLXIPL,MMP1,NEDD4L,NFKBIA,NOS2,PECAM1,PKLR,PSEN1,RXRA,SLC2A4,UCP2,VCAM1,VEGFA |

| | | | | | | | | | |
|---|---|---|---|---|---|---|---|---|---|
| Lh | | complex | | -1,720 | -1,617 | 0,022 | 6,85E-09 | 3,11E-07 | ACTN1,ACTR2,ADAMTS1,ADGRG1,AKAP12,ALPL,ARL4C,ARL6IP5,ATP9A,BAG1,BECN1,BNIP3L,CAMK2G,CAP2,CASP4,CDK14,CDKN1C,COL15A1,COL18A1,CSNK1A1,CYFIP2,CYP11B2,DHRS3,DUSP1,FDX1,FDXR,FOS,GBP1,GK,GNAS,GNB1,GNRH2,HSD3B2,INHA,KIDINS220,MAP2K3,MAP4K4,NTF3,PER1,PHKA2,PLIN3,POP5,PPIH,PRKD1,PRKX,PSIP1,PSMD1,PTPRF,RAB27A,RAB4A,RAB5A,RGS16,RGS7,RHOB,SGK1,SLIT2,SMARCA4,SNAP23,SOD2,SRD5A2,STAT1,STK24,TFPI2,THBS2,TNFRSF11B,TP53,TPM1,TUBA1A,TYRO3,VEGFA |
| RB1 | 0,238 | transcription regulator | Inhibited | -2,493 | -2,354 | 0,017 | 7,80E-09 | 3,52E-07 | ALG5,ALPL,ATOH1,BAX,BCKDHA,BECN1,CACNA1S,CASP3,CASP4,CASP6,CASP7,CASP8,CBX5,CCN2,CCNA2,CCNB1,CDK1,CDKN1B,CDX1,CDX2,CERT1,CHCHD3,CITED2,CLDN3,CMAS,CYP27B1,DCK,DCTN4,DDB2,DDIT3,DEK,DNMT1,EED,EGR1,EI24,F3,FABP2,FAM111A,FANCL,FAS,FN1,FOS,FOSL1,FOXC2,FOXN1,FZD6,GPR12,GPR162,GRPEL1,H2AC8,HMGB2,HRAS,HSBP1,IDH3B,IVL,KPNA4,KRT10,KRT4,MAP2K3,MAPK1,MCM2,MCM3,MCM5,MELK,MET,MFAP1,MYBL2,MYOM2,NDUFA2,NKX2-5,NME1,NTSR2,OGA,PARP1,PCLAF,PCNA,PGD,PIM1,POLD3,PPARG,PPFIBP1,PPP1R17,PPT2,PYY,RAB27A,RB1,RFC4,RMI1,RRM1,RRM2,SMARCA5,SOD2,TERT,TMPO,TNNC1,TNNC2,TNNT2,TP53,TP73,TUBA1A,TYMS,ULK1,VEGFA,YWHAE,ZNF638 |
| oleic acid | | chemical - endogenous mammalian | | -1,506 | -1,380 | 0,021 | 9,05E-09 | 4,04E-07 | ABCA1,ACACA,ALOX5,APOB,APP,BAX,CLU,CYP4F12,DDIT3,DUSP1,FABP1,FABP2,FOS,G6PC,GADD45A,GADD45B,HMOX1,HSPA5,LCN2,LDLR,LHB,LPL,MLXIPL,NFKBIA,NOS2,NPC1L1,NR4A1,PCK1,PDK4,PIK3IP1,PLIN3,PNPLA3,PPARA,PSEN1,PTEN,RXRA,SLC2A4,SOAT1,TLR2,UCP2,VCAM1 |
| FGF2 | | growth factor | | -0,327 | 0,026 | 0,040 | 9,84E-09 | 4,34E-07 | ADM,AFP,AKR1B1,ALB,ANPEP,ARC,BAX,BTG2,C1QL1,CAPZA2,CBFB,CCL20,CDKN1B,CDX2,CIDEC,COL1A1,COL1A2,COL3A1,CRYAB,CSF3,CXCL6,DAD1,DDIT3,DIO3,EGR1,ELK1,ELN,ENO1,EPCAM,F3,FAS,FBN1,FDX1,FN1,FOS,FOSL1,FOXO1,FOXO4,GADD45A,GBP1,GCH1,GLI1,GRIA3,HBEGF,HIF1A,IGFBP4,JUNB,KATNA1,KRAS,MACROD1,MAPT,MCAM,MET,MIP,MITF,MMD,MMP1,MPZ,MYOD1,MYOG,NES,NEUROD6,NKX2-5,NOS2,NOTCH4,NR4A1,NTF3,ODC1,PCNA,PLK2,PPARG,PPP1R12A,PRKCD,PRL,PTTG1,RB1,S100A10,S100A4,SEMA6D,SHBG,SLC34A1,SNAI1,SPP1,SPRY1,SPRY2,SPRY4,SQSTM1,ST3GAL4,TEAD4,TFPI,TGFBR3,TIMP1,TNFSF10,TP53,USP47,VCAM1,VEGFA,VIM,WNT10B,XRCC5 |
| JUN | -0,496 | transcription regulator | | -0,574 | -0,226 | 0,042 | 1,01E-08 | 4,44E-07 | A2M,AFP,AKAP12,ALAS1,ALOX12,ANGPTL2,ANKRD2,ANXA1,ANXA2,APOC3,APOM,APP,ASNS,ATF3,BAX,BIRC2,CALM1 (includes others),CAPN2,CAPNS1,CCL20,CCN2,CCNA2,CD44,CDC20,CDK1,CDKN1B,CGA,CLU,COL1A1,COL1A2,COMT,CSTA,DARS1,DNMT1,DROSHA,DUSP1,EVX1,F3,FAS,FN1,FOS,FOSL1,FUS,GADD45A,GAS6,GSTP1,HLA-B,HLA-G,HMOX1,HPGD,IFI16,IGFBP4,ITGAV,ITGB8,IVL,JUNB,LHB,MET,MMP1,MMP7,MPZ,MSH2,MXI1,MYOD1,NCF2,NFKBIA,NGF,NKX2-5,NOS2,NOX5,NPPA,NR0B2,NR4A1,NRCAM,PADI3,PAK3,PARVA,PDCD4,PEA15,PPM1D,PRL,PTEN,PTGDS,PTPRO,PXDN,RARG,RASA1,RHOB,RNF7,RXRA,S100A10,SGK1,SLC10A2,SNCG,SNRNP70,SOD2,SPP1,SST,STAT1,STMN1,TIMP1,TNC,TNNC1,TP53,TP73,VCAM1,VDR,VEGFA,VIM,ZFP36 |
| NR1H3 | -0,126 | ligand-dependent nuclear receptor | | 1,431 | 1,489 | 0,011 | 1,08E-08 | 4,72E-07 | ABCA1,ABCG1,ACACA,APOA1,APOC3,APOD,APOM,ARL4C,ATF3,AURKA,C1QA,CAT,CCL17,CCNA2,CCNB1,CCNB2,CD47,CDC25B,CDK1,CDKN1B,CFB,CSF3,CX3CR1,DDX60,FBP1,FGL2,G6PC,GAS6,GCK,GLI1,GRAP2,GSTM5,GSTP1,HK1,IFIT2,IRF7,ITGB3,KIF20A,LDLR,LPL,LYZ,MELK,MLXIPL,MYLIP,NFKBIA,NOD1,NOS2,NR0B2,PFKFB1,PKLR,PLTP,PPARG,PRC1,RB1,SLC2A4,SNAI1,SYN1,TAP1,THRB,TLR1,UBE2C,VCAN,VEGFA,ZBP1 |
| FOXA2 | | transcription regulator | | -0,288 | -0,348 | -0,010 | 1,27E-08 | 5,54E-07 | A2M,ABCC8,AFP,AGR2,ALAS1,ALB,ALDOB,ALOX5,APOA1,APOB,APOC3,APOM,BIRC2,C3,CCL17,CCL22,CDX2,CER1,CHI3L1,CLCA1,CYP26A1,CYP2A6 (includes others),DCT,EPCAM,EPHX1,EPX,FOXO1,G6PC,GCK,GFRA4,HADH,HNF1A,HNF1B,HNF4A,HPGD,IL13,LPL,LYZ,MLXIPL,MUC5B,MUC6,NFKBIE,PCK1,PDHX,PIM1,PKM,PNLIPRP1,PPARA,PPARG,PROS1,PTGDS,PYY,RARG,RORA,RORC,SLC10A1,SLC2A4,SNAI1,SST,TMPRSS2,TNNI1,UCP2,WNT7B |

| | | | | | | | | | |
|---|---|---|---|---|---|---|---|---|---|
| ELAVL1 | 0,140 | other | | -0,424 | -0,171 | 0,054 | 1,42E-08 | 6,12E-07 | ABCA1,ANXA11,ATF3,CALM1 (includes others),CD83,CLDN1,CLDN3,DUSP1,EIF4E2,EIF4ENIF1,EIF4G1,ELAVL1,FOS,HMOX1,IL13,MYBL2,MYOD1,MYOG,NOS2,PBK,PIGR,PRC1,PROM1,QARS1,RACK1,RFC5,RPN2,SLC7A7,SQSTM1,STAT3,TAF9,TIA1,TIMM17A,TP53,TRIOBP,TSC22D3,VEGFA,WEE1,ZFP36 |
| SP3 | 0,223 | transcription regulator | | 0,560 | 0,739 | 0,041 | 1,50E-08 | 6,42E-07 | ABCA1,AKAP12,ALOX12,ALOX15B,APBA1,ASNS,ATF3,CCL20,CD2AP,CDC25B,CDKN1B,CHI3L1,CITED2,COL1A1,COL1A2,CYP26A1,CYP7B1,DRD2,DUSP1,EGR1,ESR1,F3,FAS,FDX1,FOS,FOSL1,FOXP3,GHR,HSD11B2,HSPA5,ITGB8,KRT81,LDLR,LPL,MET,MSH6,NCF2,NES,NFKB1,NOS1,PKM,PRKCD,PTCRA,PTTG1,PYY,RRAS,SGK1,SOD2,STAT1,STAT3,TERT,TGFBR3,TGM1,TIMP1,TLR2,TNC,TNNC1,TP53,TSPO,TXNRD1,VEGFA,VIM |
| CREB1 | | transcription regulator | | -1,347 | -1,077 | 0,027 | 1,51E-08 | 6,42E-07 | ABCA1,ADM,AKAP13,ALAS1,ALOXE3,ANGPTL2,APOLD1,ARC,ARHGAP33,ATF1,ATF3,AURKA,AVP,BCAP29,BHLHE40,BTG2,CA10,CAMKV,CCNA2,CCNB1,CCNB2,CDA,CDK14,CGA,CPD,CRMP1,CSRP2,CYFIP2,DMBT1,DUSP1,EGR1,ESRRG,FABP2,FGL2,FN1,FOS,FOSB,FOXP3,FUT7,G6PC,GAD1,GADD45A,GADD45B,GADD45G,GALNT1,GLA,GPR12,GPR3,GRIA3,HLA-A,HLA-DQB1,HLA-DRA,HLA-G,HMOX1,HPGD,HSPA4,HSPA5,IDI1,IER2,INA,INHA,IRF7,JPT1,JUNB,KDELR3,KYAT1,LAPTM4B,LCN2,LDLR,LEP,LPL,MARCKSL1,MCAM,MCM5,MEST,MMP1,MNS1,MRPL49,MUC5B,MYBL2,MYH10,NAB2,NEUROD6,NFIL3,NNAT,NOS1,NOS2,NOX5,NR4A1,NR4A3,NRP1,PAM,PCK1,PCNA,PDYN,PER1,PER2,PLXNA2,PPARG,PPP1R15A,PRL,PSEN1,RALYL,RB1,RGS2,SEMA4A,SEMA7A,SERPINI1,SGK1,SHTN1,SLC2A4,SLC8A2,SOCS2,SOD2,SQSTM1,SRPK2,SRSF10,SST,STAT3,STMN1,SULT2B1,TCEAL9,TIPARP,TMBIM1,TSKU,VEGFA,VIM,WARS2,ZFP36 |
| IGF2 | | growth factor | | 0,018 | 0,141 | 0,031 | 2,14E-08 | 9,03E-07 | ALAD,CASP6,CAV3,CCN2,CDK1,CTSV,DDIT3,DUSP1,EGR1,ELOVL6,FN1,GADD45A,GATA6,GCH1,GHR,GNB1,GRN,HBEGF,HIF1A,IER2,IFITM2,IGFBP4,JUNB,KLF9,LEP,MCM2,MCM5,MMP1,MXI1,MYBL2,MYOD1,MYOG,NDRG1,NFKBIA,NGF,NOS2,PDCD4,PIM1,PMS1,PTEN,RGS16,RHOB,RRM1,RRM2,SKAP2,SLC2A4,SPP1,TGFBI,THBS2,TP53,VEGFA,ZFAND5 |
| NRG1 | | growth factor | | 0,285 | 0,470 | 0,033 | 2,28E-08 | 9,54E-07 | ABCA1,ACAT1,ACTN1,ATF1,BDH2,BET1,CAPN1,CASP3,CCN2,CDKN1B,CDX2,CHCHD3,CHRND,CKS2,COL1A1,COL6A3,CXCL6,DAD1,DDIT3,DUSP1,EGR1,EGR3,ENO3,FGF1,FN1,FOS,FOSB,FSTL1,GAD1,GPI,HIF1A,HMOX1,IDH3B,IDI1,IER2,ITGAV,ITGB3,JUNB,JUND,KLK3,LPCAT3,LSM1,MGLL,MYBPC3,MYOG,NKX2-5,NPPA,NR4A1,NR4A3,PDCD4,PFKFB1,PKLR,PLG,PRL,PYGM,RAB3A,SHBG,SLC2A4,SNAI1,SOX4,VEGFA,VIM,ZFP36,ZNF33B |
| MAP2K1 | 0,132 | kinase | | -1,081 | -0,692 | 0,063 | 2,29E-08 | 9,54E-07 | ABCC1,ADGRG1,AHR,ALDH1L1,APOBEC3G,ATF3,BIRC2,CCN2,CCNA2,CD44,CDKN1B,COL1A1,COL1A2,COL3A1,CRYAB,CSTA,DNMT1,DPP4,DUSP1,ELK1,ENO1,F3,FOS,FOSL1,FURIN,HIF1A,HSPA5,INS,ITGB3,IVL,JUND,LDLR,LEP,LHB,MITF,MMP1,MPZ,MUC5B,NET1,NFKBIA,NOS1,NOS2,NOX5,OLFM4,PCNA,PECAM1,PIM1,RAB38,RRM2,SERPINE2,SMAD2,SPDEF,TFPI2,TNC,TNFSF10,VEGFA,ZWINT |
| inosine | | chemical - endogenous mammalian | Activated | 3,980 | 4,276 | 0,061 | 2,49E-08 | 1,03E-06 | A2M,ANXA1,ANXA2,ANXA5,ARPC1B,C1QA,C1QB,C1S,C2,C3,CD63,CFB,CFD,DPP4,EMP3,LGALS3BP,RHOC,RNASET2,SERPINB1,SERPING1,TIMP1,TSPO,TUBA1A,VIM |
| TWIST1 | -0,002 | transcription regulator | | 0,636 | 0,818 | 0,028 | 2,88E-08 | 1,17E-06 | ADAMTS1,ALDH1A1,APOD,BAX,C3,CASP3,CD44,CHI3L1,CLU,COL1A1,EMP3,ESR1,FAS,FN1,FOS,FOSL1,FOXC2,FOXO1,GLI1,IGFBP4,ITGA8,ITGB3,KCNN4,LEFTY2,MET,MGST3,MME,MMP1,MXRA8,PAM,PAMR1,PDZD2,PRL,PROM1,PRRX2,PTPRU,PXDN,RHOA,RHOC,SLC28A3,SNAI1,SPOCK2,SPP1,TBX21,TGFBI,THBS2,TP53,VCAN,VEGFA,VIM |
| KDM5B | 0,293 | transcription regulator | Inhibited | -4,118 | -4,366 | -0,039 | 4,12E-08 | 1,64E-06 | ARL6IP5,ASF1A,AURKA,BUB3,CCN2,CCNB1,CDIPT,CDK1,CDKN1B,CLDN1,CYB5A,DDIT3,DHCR24,DLGAP5,DPY19L1,ECT2,EGR1,EIF5B,ESR1,FHL1,FJX1,GADD45A,GCA,HACD1,HSD17B8,ISG15,ITGB1BP1,LGALS3BP,MCAM,MCM2,MCM3,MPC1,MT1X,NDC80,NKX2-5,PBK,PLS3,PSIP1,PSME3,RNASET2,SNRPD1,STAT5A,SWAP70,WNT7B |

| | | | | | | | | | |
|---|---|---|---|---|---|---|---|---|---|
| Vegf | | group | | -0,269 | 0,333 | 0,058 | 4,28E-08 | 1,70E-06 | A2M,ACKR3,ADAM10,ADAM15,ADAMTS1,ADGRF5,ADGRG1,AHR,AKAP12,AKAP13,ALOX5AP,ANGPTL2,ANPEP,APLNR,APOM,ARHGEF17,ASNS,ATF3,AURKA,BAG2,BAX,BMP7,CA2,CALM1 (includes others),CASP1,CASP3,CASP8,CD44,CDC20,CDC25B,CDK1,CDKN3,CHI3L1,CHRNB2,CLDN1,CNTFR,CRABP1,CRYAB,CSE1L,CYP4A11,DDIT3,DENND2D,DNAJB9,DTYMK,EGR1,EGR3,ELN,ESM1,F3,FAS,FGF16,FN1,FOSB,FOSL1,GRB10,GUCY1B1,HBEGF,HIF1A,HMOX1,HOPX,IGFBP4,IHH,IL18,ITGA10,ITGA6,ITGB8,KITLG,LCN2,LDLR,LY75,LYN,MAD2L1,MAP2K3,MCM2,MCM5,MELK,MET,MID1,MPC1,NAB2,NAMPT,NDC80,NFKB1,NGB,NOTCH4,NR2F2,NR4A1,NR4A3,NRCAM,NRP1,OR7C2,OSBPL10,OSMR,PLOD2,PLXNA2,PRC1,PTK2,RAC1,RGS2,SFN,SGK1,SLC19A2,SOCS2,STAT3,TBXA2R,TIMP1,TLR3,TNC,TNFRSF11B,TPST1,UBE2C,UGCG,VCAM1,VEGFA,VIM,XDH |
| ADIPOQ | -0,184 | other | | 0,466 | 0,503 | 0,006 | 4,76E-08 | 1,87E-06 | ACACA,ACACB,ADIPOQ,ADIPOR1,BECN1,CASP6,CCL20,CCN2,CD163,CDKN1B,CGA,COL3A1,CYBA,CYBB,DUSP1,EGR1,F3,FN1,FOS,FOXP3,G6PC,GCK,GPD1,HBEGF,HMOX1,LDLR,LEP,LEPR,LPL,MMP1,NCF2,NFKBIA,NOS2,NPPA,PCK1,PCNA,PDCD4,PFKFB1,PPARA,PPARG,PTEN,ROCK1,SHC1,SLC2A4,SOAT1,SOD2,STAT3,TIMP1,TP53,UCP2,VCAM1 |
| F2 | -0,082 | peptidase | | 1,217 | 1,800 | 0,072 | 5,29E-08 | 2,07E-06 | ADIPOQ,AKR1B1,BAX,BIRC2,CASP1,CASP3,CCL20,CCN2,CCNA2,CD44,CD63,CDKN1B,CKAP4,CLIC1,CLU,COL1A1,COL4A1,CORO1C,CYBB,CYP2B6,DIO3,DUSP1,EGR1,ELL2,ESM1,F2RL1,F3,FHL1,FN1,FOS,FOSB,FOSL1,HBEGF,HIF1A,HMOX1,IER2,IGFBP4,IL13,IL32,JUNB,LAMP1,MFHAS1,MGLL,MMP1,MYBL2,MYOG,NCF2,NFKBIA,NOS2,NR4A3,NUP62,PDIA4,PPIF,PROS1,RAB3A,RAC1,RAC2,RHOA,RHOB,SLC16A5,SLIT2,SOD2,SOX15,TFPI,TFPI2,TMED9,TNFSF10,TNNT2,VCAM1,VEGFA,YIF1A |
| CLOCK | 0,022 | transcription regulator | | 0,137 | 0,095 | -0,008 | 5,43E-08 | 2,12E-06 | APP,AVP,BHLHE40,BPHL,CA1,CBFB,CCN2,CPD,CYP26A1,DDIT3,DNAJB9,DR1,DTX4,EFHC1,EIF4G2,ELOVL6,ESYT1,FGF1,FZD6,GADD45A,GADD45B,GCC2,GCK,GNPTAB,GPR3,GTF2E1,HLA-DQB1,HSD17B8,IHH,ITM2C,JUNB,KCNN3,KCNS1,KIF20A,MAP2K3,MAPK8,MEST,MGAT2,MMP7,MOXD1,NAA15,NFIL3,NOCT,NR0B2,NTSR2,OPN3,PER1,PER2,PICALM,PPARA,PPIF,RIOX2,SEC23A,SLC12A3,SLC1A2,SLC30A5,ST6GALNAC4,TEF,TMED3,TMX1,TOR1B,TP53BP1,TPST1,UBE3C,USP2,VSTM4,WEE1,WT1,ZZZ3 |
| ESR2 | | ligand-dependent nuclear receptor | | -0,558 | -0,474 | 0,013 | 5,77E-08 | 2,23E-06 | ABCA1,ABLIM1,ADAMTS1,AKAP12,ANGPTL2,ANPEP,ANXA4,ATF3,AVP,BAG1,BID,C3,CA2,CADM3,CAPN2,CCN2,CCN5,CCNA2,CDKN1B,CDKN1C,CGA,CLDN1,CLDN11,CLIC5,COL1A1,COL1A2,CTSD,CYP2A6 (includes others),DDB2,DMTN,EGR1,EGR3,ELK1,ESR1,F2RL1,FADD,FHL1,FN1,FOS,FOXO1,GADD45B,GALNT7,GLB1,GLRX,GM2A,GRPR,GSTP1,HAND1,HPGD,HRAS,HSD17B4,IGFBP4,IMPDH2,IVL,JAK1,JUNB,KLK3,KRT19,LAPTM5,LEP,LHB,LPL,MAP2K3,MAPK1,MGAT3,MME,MMP1,MPC1,MUC1,MYCL,MYOD1,NDRG1,NELL2,NFKB1,NFKBIA,NOS2,PDK4,PERP,PIBF1,PPP1R12A,PPP1R3C,PRL,PTEN,PTGDS,PYCR1,RAI2,RHOQ,ROCK1,RORC,SEMA3B,SGK1,SHC1,SLC5A7,SMAD2,SNN,SOCS2,SOX4,SPP1,SYNPO,TEAD4,TERT,TESK1,THBS4,TIMP1,TNFRSF11B,TP53I3,TPM1,TSPAN13,TSPO,USP8,VEGFA,VIM,WRAP73 |
| CD38 | 0,036 | enzyme | | 0,237 | 0,731 | 0,072 | 6,44E-08 | 2,46E-06 | ALDOA,ALDOC,ANXA2,ASNS,ASS1,BNIP3L,CCL22,CD44,CISH,CITED2,CKAP4,CLCN3,COL1A1,CRELD2,EAF2,ELL2,FGL2,FN1,GADD45G,GALK1,GBP2,IFI30,IL6R,LTA,MANF,NDRG1,NFIL3,P4HA1,PDCD2,PDIA6,PDLIM1,PFKP,PIM1,PKM,PRDX4,PYCR1,RBPMS,S100A4,S100A6,SDF2L1,SEMA4A,SOCS2,STK39,TMED5,UCHL3,UPB1,VCAM1,VIM,ZBTB32 |
| thyroid hormone | | chemical - endogenous mammalian | Inhibited | -3,079 | -3,087 | -0,001 | 6,75E-08 | 2,54E-06 | ABCG1,AFP,ALDOB,BTG2,CCNA2,CDKN1B,COMT,CRAT,CSNK2A1,DAD1,DBI,DNAJC3,EGF,EGR1,FOS,FZD6,G6PC,GCH1,GHR,HR,IGFBP4,INHA,KCNA5,KLF3,KLF9,LEP,LPL,ME1,NHLH1,NNAT,NPPA,NRAS,OAT,PCK1,PER1,PGD,PHYH,PKLR,PPARA,PRL,PTEN,PTGDS,RYK,S100A10,SEMA6B,SERPINE2,SLC6A4,SST,ST14,STAT1,THRB,UCP2,VIM,YWHAE |

| | | | | | | | | | |
|---|---|---|---|---|---|---|---|---|---|
| HDAC1 | 0,351 | transcription regulator | | -1,147 | -1,229 | -0,013 | 7,41E-08 | 2,77E-06 | ADIPOQ,AKAP12,APOA1,ARC,ATF3,ATG5,BAX,CAVIN2,CCNA2,CCNB1,CCNB2,CD44,CDK1,CDKN1B,COL1A1,COL1A2,CSHL1,DUSP1,EGR1,ESR1,FABP1,FABP2,FAS,FOS,FOXP3,GABARAPL1,GAST,GLI1,GLUL,GNAS,GSTP1,IL13,INA,KLK3,LCN2,MCM3,MCM5,MME,MPZ,MYBL2,NFKBIA,NKX2-5,NOS2,OLFM4,PCK1,POLL,PPARG,PTEN,RBBP7,RGS10,RHOB,RORC,RRM2,SGK1,SLC1A2,SNAI1,SOX10,SPP1,SQSTM1,ST5,STAT3,TBX2,TBX6,TERT,TNS1,TP53,TP73,TSC22D3,TYMS,VDR |
| Jnk | | group | | 0,807 | 1,163 | 0,052 | 7,75E-08 | 2,88E-06 | A2M,ABCA1,ANXA5,APP,ATG5,BAX,CASP3,CCL22,CCN2,CD44,CD83,CDK4,COL1A1,COL3A1,CRIP2,CXCL6,DDIT3,DNMT1,DUSP1,E2F4,EGR1,F12,F2RL1,F3,FAS,FN1,FOS,FOSB,GADD45A,HMOX1,HNF4A,HSPA5,IL13,INS,JUND,LHB,MMP1,MSH2,NFE2L2,NOS2,NR4A1,PLG,PLSCR1,PON2,POSTN,PPM1D,PRL,PTEN,RARG,RBP1,RXRA,SOD2,STRA6,TBP,TERT,TIMP1,TP53,VCAM1,VCAN,ZFP36 |
| Ifn | | group | Activated | 3,685 | 4,001 | 0,055 | 8,65E-08 | 3,20E-06 | ADAMTS1,B2M,BAG1,CASP8,CD44,CD58,CD83,CYP4A11,DDX58,FAS,FOS,HIF1A,HLA-A,IFI16,IFIH1,IFITM1,IL10RA,IL15,IL32,IL7,IRF7,IRF8,ISG15,LCN2,MAVS,MX1,NOS2,PECAM1,PLAAT4,SPP1,STAT1,TAP1,TLR1,TLR2,TLR3,TP53,TRIM21,TRIM22,XDH,ZBP1 |
| EP300 | | transcription regulator | | -1,643 | -1,198 | 0,067 | 1,11E-07 | 4,09E-06 | ACACB,ADIPOQ,AKAP12,AMHR2,AQP5,B2M,BAX,BCLAF1,CA2,CAPG,CAV3,CD53,CD59,CDKN1B,CHPT1,COL1A2,COMT,CRABP2,CRYBB2,CSF3,DHRS3,DIO1,DRD2,DUSP1,DUSP22,EGR1,EPN1,EVI2A,FDX1,FN1,FNDC3B,FOS,FOXO1,G6PC,GAST,GM2A,HBB,HLA-B,HLA-G,HMOX1,HSD11B2,IL10RA,INS,ITGA6,ITGA9,ITGB2,KLK3,LEFTY2,LEP,LGALS3BP,LHB,LTA,MAPT,MCM3,MFHAS1,MMD,MME,MMP1,MMP7,MSH2,MYOG,NDRG1,NFKBIA,NOS2,NPPA,NR4A1,NTRK3,PARP1,PCK1,PDE10A,PDK4,PERP,PHIP,PIP5K1B,PKLR,PLEKHA1,PPARA,PRKCD,PRL,PSEN1,PTEN,PTPRF,RB1,RBP1,RHOA,RNF39,RORC,RTP4,SLC43A1,SOD2,TERT,TESC,TIPARP,TLE6,TLR2,TMPRSS2,TMPRSS4,TNC,TP53,UBE2C,VEGFA,WNT8B |
| CNOT7 | 0,116 | transcription regulator | | -1,553 | -1,698 | -0,060 | 1,15E-07 | 4,18E-06 | B2M,CLDN1,CSTA,HERC6,IFI35,IFIT5,IFITM1,ISG15,LGALS3BP,OAS1,PARP12,PLSCR1,PSMB8,SP110,STAT1,TAP1,UBE2L6 |
| SRF | | transcription regulator | | -0,508 | -0,330 | 0,029 | 1,27E-07 | 4,61E-06 | A2M,ACTG1,AKAP12,ANKRD6,APOLD1,ARC,BICC1,BRD3,CAP1,CAPRIN1,CCN2,CD177,CDK4,CEBPE,CLDN1,CLDN15,CLDN3,CSRP2,CTSG,DSTN,EGR1,EGR3,ELK1,EPX,FERMT2,FGF1,FHL1,FHL2,FILIP1L,FOS,FOSB,FOSL1,G0S2,GADD45A,GADD45G,GCH1,GP9,GSTM5,HMOX1,IER2,IL15,ITGA6,ITGA9,JUNB,LCN2,LDLR,LEPR,LPCAT3,LPL,LTF,MAP4K4,MAPK10,MAPK13,MAT1A,MED6,MTMR6,MYOD1,MYOG,NET1,NEUROD6,NKX2-5,NPPA,NR4A1,OPN3,PDCD10,PLA2R1,PMFBP1,PRG2,PTMS,RAI2,RFC4,SEMA6D,SLC6A4,SLPI,SND1,SNX2,SPTBN2,TBP,TMEM184C,TNC,TNNC1,TNNT2,VCAN,WDR1,WT1 |
| SP110 | 0,251 | transcription regulator | Inhibited | -2,605 | -2,714 | -0,016 | 1,34E-07 | 4,84E-06 | ANXA5,ATF3,CD74,CLU,EGR3,F3,FLII,FUS,GRN,IFIH1,IFIT3,IFITM1,IL18,IL6R,IRF9,ITGAV,LY96,MRTFB,MX1,NCK1,NFIL3,OAS1,PANX1,PERP,PLSCR1,PPARG,PRKCA,PTPN12,RAC1,RPA1,RXRA,SMAD2,SMARCA4,SMARCD3,SOX4,SQSTM1,STAT1,STAT5A,TGFBR3,TIA1,TIMP1,TRIM5,WWOX,YAP1 |
| MAPK14 | | kinase | | -0,171 | 0,139 | 0,046 | 1,47E-07 | 5,28E-06 | ABCA1,ALOX5AP,BAX,CAT,CCL25,CCNA2,CCNB1,CD83,COL1A1,CRYAB,DUSP1,ELAVL1,FAS,FGFBP1,FN1,FOS,G6PC,GADD45A,GNAS,HMOX1,IL13,IL18,ITGB8,IVL,JUNB,JUND,KITLG,LDLR,MAPT,MMP1,MUC5B,MYOG,NOS2,NPPA,NRP1,PCK1,PECAM1,PRL,PSEN1,PSMD10,PTEN,SGK1,SH2D2A,SHC1,SOD2,TBX21,TIMP1,TP53,TRPV1,VCAM1,VDR,VEGFA,WIF1,ZFP36 |
| IgG | | complex | | 1,053 | 0,787 | -0,037 | 1,53E-07 | 5,46E-06 | ADGRG1,ADM,AFM,APP,B2M,C3,CD1B,COL3A1,CRABP2,DDIT3,DSG3,DUSP1,EFNA3,ELK1,FGFBP1,FN1,FOSB,FOXP3,GNAS,HLA-E,HSPA5,IER2,IFITM1,IL13,IL25,ISG15,ITGB8,IVL,JUND,KLK11,KLK6,KLK7,KRT10,KRT31,LCN2,LDLR,LGALS1,NGF,PEA15,PIM1,PLS3,PPIF,PPL,PPP1R15A,SPINT1,ST14,STK24,TAPBP,TNC,TNFRSF13B,TOB1,TRIM29,UGCG,VCAM1,VEGFA,ZFP36 |

| | | | | | | | | | |
|---|---|---|---|---|---|---|---|---|---|
| STK11 | | kinase | | 0,027 | 0,209 | 0,022 | 1,54E-07 | 5,49E-06 | A2M,ADSL,AHCY,ALB,ALDOA,ASS1,ATF3,ATIC,BMP5,BTF3,CCNA2,CCT6A,CDKN1B,COL3A1,CSF3,CTSD,CXCL6,CYP2B6,DCTPP1,DLGAP2,DNMT1,ECM2,EIF3A,ENO1,FBXO9,FN1,FOSL1,GADD45A,GADD45B,GPI,GRIA3,HIF1A,IMPDH2,JUNB,LTB,LUM,LYPD3,MCAM,MMP17,NDUFA2,NET1,NOD1,NOS2,NRCAM,PAPSS1,PCDHA10,PCK2,PDYN,PKM,PNP,POLR2E,PPA1,PTEN,PTPN3,PYCR1,RB1,RPIA,RRM1,RRM2,SARDH,SERPINI1,SNAI1,SPP1,TALDO1,TARS1,TKT,TNFSF10,TWF2,TXNRD1,TYMS,VEGFA,WNT2 |
| EIF2AK3 | -0,120 | kinase | | -1,804 | -1,537 | 0,064 | 1,76E-07 | 6,23E-06 | ALDH18A1,ASNS,ATF3,ATG5,BECN1,BIRC2,BTG2,CA9,CCL20,CLCN3,DDIT3,DNAJB9,DNAJC3,EGR1,ELP3,FOSL1,GADD45A,GCH1,HSPA5,NARS1,NDRG1,NFIL3,NOS2,PCK2,PNLIP,PON2,PPP1R15A,SLC1A7,SNW1,SQSTM1,TARS1,TP53,VEGFA,WARS1,YARS1,ZFP36 |
| CSF3 | -0,210 | cytokine | | -0,894 | -0,743 | 0,022 | 1,99E-07 | 6,96E-06 | BAX,CAPG,CD177,CD44,CDK4,CDKN1B,CEBPE,CISH,CTLA4,CTSD,CX3CR1,CYBB,EGR1,EGR3,FOS,FUS,GADD45A,GADD45B,GADD45G,HLA-DPA1,HLA-DRA,HLA-DRB1,HMOX1,HOXB7,IK,IL7,ITGB2,KITLG,LCN2,LTA,LTB,LTF,LY96,LYZ,MET,NFIL3,ODC1,PIM1,PIWIL1,PLCB2,PLSCR1,PPARG,PRKCA,RAB27A,RORC,STAT3,TLR2,TNFSF10,TP53,VCAM1,VEGFA |
| SMARCB1 | 0,044 | transcription regulator | | -1,013 | -0,723 | 0,045 | 2,34E-07 | 8,09E-06 | A2M,ACAT1,ACTR2,AMPH,AURKA,BNIP3L,CA8,CCNA2,CD44,CDC7,CDK1,COL18A1,COL1A1,COL1A2,COL5A2,CYP2C8,CYP3A7,CYP4A11,DSG2,F11,F11R,FAS,FBP1,FOS,G6PC,GADD45A,HBEGF,HOXC8,IFI16,IFITM1,IFNAR2,MCM2,MCM3,MCM5,MMP1,MX1,MYBL2,OAS1,PCK1,PFKFB1,POSTN,PPARG,PPP1R3C,PTMS,RFC5,S100A1,SLC37A4,SMC4,TBX21,TP53 |
| mir-21 | | microRNA | Inhibited | -2,702 | -3,148 | -0,055 | 2,49E-07 | 8,49E-06 | ARF4,ARL6IP1,BTG2,C1R,CANX,CASP4,CCL1,CCL17,CCNA2,CCNB1,CCT8,CKS2,CLPB,CLU,COL1A1,COL3A1,CRY2,CSE1L,DLGAP5,ECT2,ELN,FAS,FGF1,FGL2,FN1,FOXP3,FUBP1,GBP2,IFI16,IL18,IL6R,IRAK1,LEPR,LTB,MSH2,NME1,NMT2,NOS2,PBK,PDCD4,PECAM1,PER2,PPA1,PPARA,PRC1,PTEN,RHOB,SLPI,SMAD2,SMARCA4,SOD2,SPRY1,SPRY2,SSR1,STAT1,STAT3,STMN1,TAP1,TBX21,TERT,TGM1,TLR1,TLR2,TP53,TPM1,TRIM37,UBE2A,UBE2L6,VCAM1,YWHAB,ZWILCH |
| IRF4 | | transcription regulator | Inhibited | -3,134 | -3,337 | -0,032 | 2,52E-07 | 8,58E-06 | ALPL,B2M,CANX,CASP10,CCL1,CCL17,CCL20,CCNB1,CCNC,CDK1,CTLA4,CTSS,CX3CR1,DBI,DDIT3,DNASE2,ELL2,FAIM,FOXP3,GALNT2,GBP1,IL13,IRF7,IRF9,ISG15,MANF,OAS1,PCNA,PDCD6,PHF11,PLSCR1,PPIB,PRKCA,PSMA1,RAC1,RHOA,RHOB,RORA,RORC,SMARCA4,SPP1,STAT1,SUB1,TBX21,TNFSF10,TRIM21,YWHAH |
| MST1 | -0,121 | growth factor | | -1,715 | -1,820 | -0,029 | 2,63E-07 | 8,80E-06 | CCN2,FAS,FOS,FOXP3,G6PC,GPC3,IL15,IL18,IRF8,MST1R,NR0B2,PCK1,PTK2,SOX4,SPP1 |
| SIRT1 | -0,193 | transcription regulator | Inhibited | -2,136 | -2,410 | -0,031 | 2,84E-07 | 9,44E-06 | ABCA1,ABCG1,ACAP1,ADAR,ADIPOQ,AFP,ANXA4,ATOH1,BAX,BCLAF1,CAT,CCNB2,CD74,CD83,CDKN1B,CLCA1,CYP2B6,DDX58,DDX60,ERMP1,ESR1,FAS,FGF5,FN1,FOXO1,FOXP3,G6PC,GABARAPL1,GADD45A,GCK,GFPT1,GLI1,GSTA3,HIF1A,HLA-A,HLA-DQB1,HMOX1,IFI44,IFIT3,IL13,IRF7,KDM5B,KIF1A,LAMB1,LAPTM5,LDLR,LEP,LGALS3BP,LTBP2,MAPT,MGAT2,MMP1,MYL6,MYOG,NEDD4L,NELL2,NFKBIE,NLGN1,OAS1,P3H3,PARP1,PARP12,PCK1,PCK2,PDK4,PER1,PHKB,PLAGL2,PNPLA3,PSMB9,PYGM,RAC1,RGS16,RNF39,ROCK1,RTP4,SEC61A1,SIRT3,SLC7A10,SNRNP70,SOD2,SP110,SSBP3,STAT1,STAT3,SYNPO,TAP1,TERT,TFPI,TMPRSS4,TP53,TP73,UCP2,WNT2 |
| S100A8 | 1,146 | other | | 1,016 | 1,154 | 0,020 | 2,93E-07 | 9,67E-06 | A2M,ABCG1,ADAMTS1,AKAP12,ALDH1A1,ALPL,ATF3,BECN1,BHLHE40,BIRC2,C1QB,C1S,CASP1,CCL20,CCN5,CDH3,COX6A2,CTLA4,ELN,EPCAM,FAS,FOSL1,FURIN,HMOX1,ISG15,KLF15,MMP1,MMP17,NFKB1,NFKBIA,NOS2,NR4A3,NTRK3,PCDHGB5,PER1,PIK3IP1,PLK3,PLXNB3,POSTN,PTPRO,RBFOX1,RNASE6,SEMA3B,SGK1,SHC3,SLC26A1,SOCS7,TNFAIP8,VCAM1,VCAN,WT1 |
| L-methionine | | chemical - endogenous mammalian | | 0,326 | 0,286 | -0,012 | 2,99E-07 | 9,82E-06 | ALB,CCNB1,CCNB2,CDK1,CLU,DDIT3,F3,FANCL,G6PC,GADD45B,HSPA5,IFI16,IGFBP4,MAT1A,NFKBIA,PCK1,PCK2,PRC1,RHOB,SOD2,SPHK2,SPP1,TYMS |
| cholesterol | | chemical - endogenous mammalian | | 0,197 | 0,370 | 0,025 | 3,09E-07 | 1,01E-05 | ABCA1,ABCG1,ACAT1,ADAM11,APOB,APP,AQP8,ATF3,CASP1,CAV2,CCL1,COL1A1,COL3A1,CYBB,CYP3A5,CYP7B1,DBI,DCSTAMP,DDIT3,EPHX1,F3,FABP6,FN1,GLI3,GNAI1,GNMT,HBEGF,HSP90B1,HSPA5,IDI1,IHH,IL10RA,IL10RB,JAK1,KCNA5,KITLG,LAPTM5,LDLR,LEPR,LPL,LYZ,MANF,MAPT,ME1,MITF,MME,MMP1,NCF2,NPC1L1,NR0B2,NR4A1,NR4A3,PLTP,RGS16,RHOA,SLC28A3,SLC2A4,STX3,TIMP1,TMEM123,VCAM1 |

| | | | | | | | | | |
|---|---|---|---|---|---|---|---|---|---|
| HNF1A | -0,233 | transcription regulator | Inhibited | -3,719 | -3,171 | 0,074 | 3,21E-07 | 1,04E-05 | ADH6,AFP,AIMP1,AKR1C1/AKR1C2,ALB,ALDOB,ANPEP,ANXA4,APOB,APOC3,APOH,APOM,ATP4A,BMP3,C1S,C2,C8B,CDKN1B,CLIC5,COL3A1,COX6A2,CSHL1,CTSZ,CYB5A,DCT,DPP4,DTL,F11,FABP1,FAS,FBP1,FRK,FUT7,FXYD3,G0S2,G6PC,GARS1,GATM,GCK,GCNT3,GLA,GLUL,GMDS,GNG12,GRHPR,GSTA1,GTF2E1,HAO1,HGFAC,HNF1A,HNF1B,HNF4A,HNMT,HOPX,HPX,HSPA5,IHH,IL36A,INS,INSR,ITIH4,KIF20A,LCT,LDLR,LIPA,MIA2,MLXIPL,MRPL15,MSH6,MT1X,NAT10,NOD1,NPC1L1,NR0B2,PAH,PAMR1,PCK1,PDHX,PFKP,PIGR,PKLR,PLA1A,PLG,PPP1R1A,PSMA5,RACK1,RO60,SEMA4A,SERPINA10,SERPING1,SLC10A1,SLC10A2,SLC26A1,SLC37A4,SLPI,SRI,SSTR4,SUPV3L1,TARS1,TEF,TMED10,TMEM140,TMPRSS2,TMPRSS4,TRPM2,UCP2,VLDLR |
| IKBKB | | kinase | | 1,578 | 1,957 | 0,049 | 3,23E-07 | 1,04E-05 | ABCA1,AFP,ATF3,BAX,BIRC2,C3,CASP8,CCL1,CCL17,CCL20,CCL25,CCN2,CCNA2,CD44,CDC25B,CLU,CTSF,CTSZ,CXCL6,CYBA,CYP7B1,ECT2,EGR1,F3,FAS,FOS,FOXO1,GADD45A,GBP2,GCH1,GLI1,GM2A,GRN,HIF1A,HLA-A,HMOX1,IFI16,IL13,ISG15,ITGB3,JUNB,KIF20A,LCN2,LTB,MMP1,MX1,NFKB1,NFKBIA,NGF,NOCT,NOS2,NPPA,POSTN,PPM1D,PTEN,RBP1,RGS16,S100A4,SERPINE2,SGK1,SNAI1,SOCS2,SOD2,STAT3,TERF2IP,TLR3,TNC,TNFRSF11B,TP53,VCAM1,VCAN,VEGFA |
| IRF2 | 0,131 | transcription regulator | Activated | 2,339 | 2,577 | 0,044 | 3,72E-07 | 1,18E-05 | B2M,BNIP3L,C1S,CASP1,CDKN1B,CFB,CIDEC,CLIC1,CTSS,DNAJB6,DPY19L1,ERAP1,FAM111A,GBP1,IFI35,IL7,IRF7,IRF8,ISG15,LCN2,MMP7,NAMPT,NOS2,OAS1,OPN3,PSMB10,PSMB8,PSMB9,PSME1,RARRES1,RRBP1,TAP1,TAPBP,TLR3,TNFSF10,TPM1,TRIM21,TXNRD1,UBE2L6,VCAM1 |
| HNF1B | 0,282 | transcription regulator | | -1,639 | -1,604 | 0,006 | 3,84E-07 | 1,22E-05 | ACTN1,AFP,ALB,ALDOB,ANXA4,BHLHE40,CA2,CD44,CD99,CDK1,CITED2,CLDN1,COL5A2,DEFB1,DUSP1,EXT1,F11R,FN1,FZD1,GATA6,GSTA1,HGFAC,HNF1A,HNF4A,IHH,ITGB6,LAMB1,LUM,MET,MMP7,MPZL2,PDE4C,PODXL,PSTPIP2,S100A2,S100A4,S100A6,SNAI1 |
| CG | | complex | | 0,031 | 0,482 | 0,052 | 4,41E-07 | 1,38E-05 | ADAMTS1,ADM,APLNR,B2M,BAX,BHLHE40,BHMT,BTG2,CDK4,CDKN1B,CLDN11,CLU,CTSV,CYP26A1,DBI,DDX25,DPT,DTL,DUSP1,EGR1,ELK3,EPHA7,ESM1,F2RL1,FAS,FGL2,FHL1,FJX1,FOS,FZD1,G0S2,G6PC,GATA6,GM2A,GTF2E1,HIF1A,HLA-A,HPGD,HSD11B2,HSD3B2,IFIT3,IGFBP4,INHA,ITGAV,ITGB3,KHK,LDLR,LEPR,LGALS3BP,MCAM,MMP1,MYCL,NOS2,NR2F1,NR4A1,NRP1,NUCB2,PAPPA,PCNA,PDYN,PECAM1,PODXL,PPARA,PPARG,RGS2,RORA,S100A10,S100A6,SACS,SLC5A5,SLC5A7,SMAD2,SMARCA1,SMARCA5,SPP1,SPRY2,SRD5A2,STEAP1,TFPI2,TGFBR3,TIMP1,TIPIN,TP53,TSPAN13,VCAN,VEGFA |
| cholic acid | | chemical - endogenous mammalian | | 1,441 | 1,523 | 0,016 | 4,45E-07 | 1,39E-05 | ABCA1,ABCC3,ABCC4,APOA1,APOM,CDX2,CYP2B6,CYP3A4,CYP3A5,CYP4A11,CYP7B1,EGF,EPHX1,FABP1,FABP6,FBP1,G6PC,LPL,MDK,MVP,NR0B2,PCK1,PLTP,RAD23B,RBP1,SLC10A1,SLC10A2,TM4SF5 |
| RBL2 | -0,071 | other | | 1,094 | 0,749 | -0,070 | 4,84E-07 | 1,50E-05 | ATOH1,AURKA,CASP4,CASP6,CCNA2,CCNB1,CCNB2,CDK1,CDKN1B,CDX1,CDX2,DNMT1,E2F4,ELK1,FAS,FOXN1,GRPR,HADH,IVL,KPNA2,MAP2K3,MYBL2,MYOD1,PCNA,PIM1,PPP1R8,RB1,RRM2,STMN1,TEAD4,TERT,TP73,TYMS,VEGFA |
| IRF7 | 0,451 | transcription regulator | Activated | 5,494 | 6,039 | 0,080 | 4,94E-07 | 1,52E-05 | ADAR,APOBEC3G,CASP4,CTLA4,DDX58,FOXP3,FZD1,GBP1,IFI16,IFI35,IFI44,IFIH1,IFIT2,IFIT3,IFITM1,IFITM2,IL15,IRF7,IRF8,IRF9,ISG15,MX1,NAMPT,NMI,OAS1,PARP12,PDCD2,PHF11,PLSCR1,PSMB10,PSMB8,PSMB9,PSME1,RTP4,STAT1,TAP1,TDRD7,TMPO,TNFAIP8,TNFSF10,TOR1B,TPST1,TRIM21,TRIM22,TRIM5,UBE2L6,USP8,ZBP1,ZC3HAV1 |
| TCR | | complex | | -1,600 | -1,235 | 0,050 | 5,07E-07 | 1,55E-05 | ABLIM1,ADM,ALDOA,ALOX12,APOC3,APOD,ARHGEF1,CCL1,CCL17,CCL22,CCR4,CD44,CD5,CD70,CDKN1B,CISH,CTLA4,DDX58,EBNA1BP2,EGR1,ELL2,ENO1,FAS,FOS,FOXP3,GADD45B,GADD45G,GLUD1,GRIA3,GZMA,HADH,HIF1A,HSP90B1,HSPA5,IFI44,IFIH1,IFIT2,IFIT3,IFNGR1,IL13,IL7,IRF7,ISG15,JUNB,KCNK5,LAMP1,LBR,LIPA,LTA,MAPK1,MAPK8,MX1,NAB2,NFKB1,NFKBIA,NME1,NR4A1,NRP1,OAS1,ODC1,P4HB,PDCD1,PDCD4,PDIA4,PIK3R3,PIM1,PRKACB,PTEN,PTPRC,RPL13A,SCARB2,SDCBP,SERPINA3,SOCS2,SPP1,STAT1,STAT3,STAT5A,SYNCRIP,TALDO1,TBX21,TLR3,TOB1,TSC22D3,VDR,ZBTB32,ZBTB7B,ZC3HAV1 |

| | | | | | | | | | |
|---|---|---|---|---|---|---|---|---|---|
| FOXM1 | 0,147 | transcription regulator | Activated | 1,714 | 2,133 | 0,069 | 5,08E-07 | 1,55E-05 | ANXA1,CAT,CCNA2,CCNB1,CCNB2,CDC20,CDC25B,CDK1,CDKN1B,CDKN3,CKS2,ESR1,FOS,FOXF1,FUT4,FZD1,GLB1,HSD11B2,KIF20A,MAPK8,MET,MMP7,NBN,NES,NFKB1,PCNA,PECAM1,PRC1,PROM1,PTCH2,PTTG1,RMI1,SNAI1,SOD2,SQSTM1,STAT3,STMN1,TP53,VCAN,VEGFA,VIM |
| NGF | -0,241 | growth factor | | -2,041 | -1,709 | 0,049 | 5,11E-07 | 1,55E-05 | A2M,ADM,AGRN,ANXA2,APOBEC3G,APP,ASIC2,ASIC3,ATF3,BAX,BTG2,C1QL1,CD83,CDK1,CDKN1B,CHRNB4,CRYAB,DHX9,DRD2,DUSP1,EGR1,ELK1,FAS,FOS,FOSL1,GADD45A,GCH1,GUCY1A1,GUCY1B1,HMOX1,JUNB,KIF2A,KLK3,LDLR,LRRFIP1,MACROD1,MAP3K12,MMD,NAB2,NOS1,NR4A1,PLG,PLK2,PPARG,PTEN,RAC1,RACK1,S1PR2,SEMA6D,SHC1,SHTN1,SQSTM1,STAT3,STMN4,TP73,TRPV1,TRPV4,VEGFA,WT1,ZWINT |
| levodopa | | chemical - endogenous mammalian | | -2,043 | -1,938 | 0,009 | 5,50E-07 | 1,66E-05 | ABI1,ADAR,ADGRB2,ALPL,APLP1,ARC,AREL1,BANF1,BAX,BICC1,C12orf43,C1GALT1C1,C1QA,C8B,CACNA1H,CAP1,CAPG,CAPN10,CASK,CASP3,CD63,CHAC1,CHRM2,CHST8,CINP,COL4A1,COMT,CRY2,CTNNBIP1,CTSZ,CYP2A6 (includes others),DDOST,DENND1C,DMTN,DNAJC2,DOC2B,DUSP1,EIF4G2,F11R,FAAH,FADD,FAM3A,FCGRT,FOS,FOSB,FOSL1,FOXO1,FRAT2,FSTL4,GABARAPL1,GADD45B,GLB1,GNAS,GNG7,GOLIM4,GPC3,GPD1,GPS1,GRB10,GUCY1A1,HBEGF,HCLS1,HECW1,HEXA,HEXB,HLA-A,HRAS,HTATIP2,HTR1B,IFNAR2,INPP5F,INTS13,IQGAP1,ITGB1BP1,JUNB,KCNA5,KCNA6,KCNE5,KDM5B,KLF9,KYAT1,LAPTM5,LASP1,LPP,LRRC1,LYPD1,MAP3K1,MITF,MRPL33,NAB2,NEDD4L,NOCT,NTRK3,PALM,PAOX,PARD6A,PDCD4,PDYN,PDZD2,PER1,PER2,PFKP,PKP2,PLK2,PRL,PRPF18,PTPRT,PVALB,RAB8A,RC3H2,RESF1,RFC2,RTL8C,RUNDC3B,S100A10,S1PR2,SCRN1,SH3BGRL3,SH3GL2,SHC1,SLC1A2,SLC25A44,SRPK3,SUB1,SYN1,TBP,TCIRG1,TLE1,TMED10,TMEM106C,TNR,TP53,TRIOBP,UBE3C,UBFD1,UPP1,VLDLR,WDR1,WNT7B,ZNF281,ZNF76 |
| ERN1 | -0,079 | kinase | Activated | 2,281 | 2,504 | 0,041 | 6,33E-07 | 1,89E-05 | ACACA,ARF4,ATF3,BECN1,BLOC1S1,CDH6,DDIT3,DERL2,DNAJB9,DNAJC10,DNAJC3,EPCAM,FAS,G6PC,GOLGA3,HSP90B1,HSPA5,IFITM1,ITGB6,MLXIPL,MPZL2,NARS1,NOS2,NPPA,P3H1,PPARG,PROS1,SDF2L1,SEC11A,SEC24D,SEC61A1,SLC25A28,ST14,SYT11,TBCB,TMED3,TMPRSS2,UBE2J1,USO1,VEGFA,WARS1 |
| Ifn gamma | | complex | Activated | 2,243 | 2,472 | 0,043 | 7,04E-07 | 2,07E-05 | ADAR,APOBEC3G,BAG1,CASP1,CASP3,CDKN1B,CHI3L1,COL1A1,CSE1L,CYBB,ESR1,FAS,FN1,GBP1,HLA-DRA,IGFBP4,IL32,IRF8,LY96,MAP4K4,MAPK1,NOS2,NR4A3,PCK1,PPARG,PSME1,SOCS7,STAT1,TACR2,TERT,TLR3,TNFSF10,VSIG4 |
| EPAS1 | -0,138 | transcription regulator | | -0,698 | -0,325 | 0,060 | 7,27E-07 | 2,13E-05 | ABCG2,ABI1,ACACA,ADIPOQ,ADM,AKAP12,ALDOC,ATG5,BHLHE40,C1QA,CA9,CAT,CCN2,CCN5,CITED2,CLDN1,DDIT3,DECR2,ENO1,FABP2,FHL1,FN1,FOS,GADD45B,H1-2,HIF1A,HSPA4,HSPA5,ITGAV,ITGB3,LDLR,LEP,LPL,MAFF,MANF,MB,MYO7A,MYOM2,NAMPT,NDRG1,NFIL3,PLOD2,PPARG,PRKCA,SLC2A4,SOD2,SOX15,STC2,TERT,TMPRSS6,TNFRSF11B,TNNI1,TP53,TPP2,TRIM21,VEGFA,WNT10B,YAP1 |
| HMGA1 | -0,067 | transcription regulator | | -1,217 | -1,069 | 0,024 | 7,51E-07 | 2,19E-05 | ALPL,B2M,CAV2,CD44,CLCN6,COL1A1,COL1A2,COL4A1,COL4A2,COX6A2,CRYAB,CSK,DDIT3,EGR1,ELK1,ESR1,FOS,FSTL1,GHR,HAND1,HS3ST1,IDI1,IER2,IGFALS,IGFBP4,IL15,INSR,JUNB,KITLG,KRT10,LAMB1,LTA,MAPT,NKX2-5,NOS2,PPARG,PTPRU,RAN,RHOB,SHC3,SOX4,STAT3,TFEB,WNT6 |
| EHF | 0,303 | transcription regulator | | 0,625 | 0,786 | 0,030 | 7,84E-07 | 2,28E-05 | ALOX5,ANPEP,BLNK,CASP3,CCL20,CDH3,CDK1,CDKN1B,CRABP2,FOXN1,GLB1,HMOX1,HOPX,IL13,KLK3,KLK5,KLK6,KLK7,LYN,MET,MMP1,MUC1,NOTCH2,PGLYRP4,RHCG,SERPINA3,SPDEF,SPRR3,TFPI,TIMP1,TNC,VEGFA |
| Creb | | group | | -2,065 | -1,783 | 0,041 | 7,91E-07 | 2,29E-05 | AASS,ADIPOQ,ADM,ALOX5,ARPC5,ATF3,CACNA1S,CASP1,CD59,CGA,CIDEC,COMT,COX6A2,CTH,CTSV,CYP11B2,DUSP1,DYNC1LI2,EGR1,EGR3,FAS,FCER1G,FOS,FOSB,FOSL1,G6PC,GABRR2,GNAS,HLA-G,HPGD,HSPB8,IFI16,INHA,ITGA10,IVL,JUNB,LASP1,LDLR,LEP,MAPK8,MCAM,MCM3,MITF,MMP1,MUC5B,NMNAT2,NOX5,NR4A1,NR4A3,NRL,PCK1,PCNA,PDYN,PER1,PER2,PHTF1,PLIN3,PPARG,PPFIBP1,PRKCA,PRL,PSENEN,PYCR1,RBFOX1,RGS2,SLC25A44,SLIT2,SOX4,SST,SSTR2,SYNPO,THAP1,TIA1,TMEM184C,TNFRSF11B,VEGFA,ZBP1 |

| | | | | | | | | | |
|---|---|---|---|---|---|---|---|---|---|
| MAPK9 | | kinase | | -0,686 | -0,469 | 0,034 | 8,34E-07 | 2,41E-05 | ACACA,BAX,CCN2,CD70,CDKN1B,CIDEC,EGF,EGR1,FAS,FGL2,FOS,FOSL1,FOXP3,GADD45A,GADD45B,HIF1A,IER2,IFI16,IL18,IRF9,JUNB,JUND,LGALS3BP,LPL,LTA,LUC7L3,MARCKSL1,METAP2,MUC5B,NMI,NOS2,PDIA3,PLA1A,PPARA,PPARG,PPP1R15A,PTEN,RAC1,RAD23B,SEM1,SOD2,TBP,TBX21,TLR3,TP53,VDR,ZFP36 |
| HDAC4 | | transcription regulator | | 0,554 | 0,447 | -0,029 | 8,51E-07 | 2,45E-05 | ARC,ATF3,CAMK2A,CAP2,CCNB1,CYFIP2,FOS,FOSL1,G6PC,GADD45A,HDAC1,HIF1A,HLA-DRA,KLHL2,MAPK1,MAPK10,MAPK8,MYH10,MYOG,MYOM2,MYOZ1,NBEA,NOS2,NR4A1,NRCAM,PRKCA,RGS2,RGS7,SH3GL2,SLC17A7,SLC2A4,SYN1,TERT,TP53BP1,TRPV1,TSPAN13,TSPAN7,VCAM1,VEGFA |
| NR2F2 | 0,200 | ligand-dependent nuclear receptor | | -0,557 | -0,557 | 0,000 | 8,64E-07 | 2,47E-05 | ALDH2,APOA1,APOC3,CYP11B2,GATA6,HNF1A,HNF1B,HNF4A,LIPC,LPL,LTF,NPPA,NR2F2,NRP1,PCK1,PPARA,SHBG,VEGFA |
| IL1A | | cytokine | | 0,668 | 1,077 | 0,054 | 9,44E-07 | 2,66E-05 | ADAMTS1,ALDH1A1,ALDH1A3,ALDH3A2,APOD,APP,ASIC2,ASIC3,ATF3,BAX,BIRC2,BTN3A3,CA2,CCL20,CD44,CD83,CSF3,CXCL6,CYP2B6,CYP3A4,CYP3A5,DPP4,F2RL1,FAS,FOS,FOSB,FOSL1,GBP1,GCH1,HMOX1,HSD11B2,IFNGR1,IFNGR2,IK,IL13,IL18,IL32,IRAK1,ITGAV,ITGB3,JUNB,KITLG,LCN2,LEP,MCAM,MMP1,NFKB1,NFKBIA,NGF,NOS2,NR2F2,PPARG,RHCG,SERPINA3,SOD2,SPP1,SSTR2,TERF2IP,TLR2,TNFRSF11B,VCAM1 |
| WT1 | -0,429 | transcription regulator | | -0,203 | -0,136 | 0,011 | 9,44E-07 | 2,66E-05 | ACTN1,AHCY,AMHR2,ANPEP,ANXA11,ARL2,ASNS,BTG2,CAPRIN1,CCN2,CD2AP,CDC73,CLIC1,CMPK1,COL4A1,CTSV,EGR1,ESR1,EXT2,FGF1,FIBP,FOSB,GRB10,HBEGF,HCFC1R1,HSP90B1,IDI1,INHA,INSR,IRF8,ITGB1BP1,JUNB,JUND,KRAS,MAPK8,MT1X,MTIF2,MYL6,NDRG1,NME1,NPHS1,ODC1,PCK2,PDIA4,PECAM1,PODXL,PPIB,ROCK1,SH3BP5,SPRY1,SQSTM1,TARS1,TERT,TRIM21,TYROBP,VDR,VEGFA,WARS1,WT1,YARS1,ZMIZ1 |
| PPARGC1A | -0,177 | transcription regulator | Inhibited | -3,605 | -3,455 | 0,018 | 9,48E-07 | 2,66E-05 | ACACA,ACACB,ACAT1,ADAMTS1,ADGRG1,ADIPOQ,ALAS1,APOC3,APOD,BAX,BCAT2,C2,C3,CALM1 (includes others),CAT,CAV3,CCN2,COL1A2,COL6A3,CYP2A6 (includes others),CYP2B6,CYP3A4,CYP3A5,DIO1,DPT,DR1,ESRRA,G6PC,GAS6,GK,HSD3B2,IL18,INHA,INPP5J,ITGB1BP1,KYAT1,LAMB1,LDLR,LIPA,LIPC,LPL,LUM,MB,MCM3,MCM5,ME1,MITF,NPC1L1,NPPA,PCK1,PCK2,PDHA1,PDIA5,PDK2,PDK4,PHYH,PLA2R1,PNPLA2,POSTN,PPARA,PVALB,RXRA,SEMA3G,SERPINA3,SERPINE2,SIRT3,SLC2A4,SOD2,SYT2,TGFBR3,TIGAR,TIMP1,TNFRSF11B,TNFSF10,TNNI1,TP53,UCP2,VAMP2,VCAN,VEGFA,XDH |
| glutamine | | chemical - endogenous mammalian | | 1,007 | 0,876 | -0,025 | 1,05E-06 | 2,92E-05 | AKR1C1/AKR1C2,ASNS,ASS1,ATG5,COL1A1,DDIT3,DUSP1,FN1,FOS,GADD45A,HIF1A,HK1,HSBP1,HSPA5,IL15,JUNB,LTA,NES,NFKBIA,NR4A1,PCK1,PCK2,PCNA,PROM1,PYCR1,SOX10,THBS2,TIMM17A,TP53,TUBB4A,VEGFA,VIM,WARS1,WT1 |
| MTOR | -0,088 | kinase | | 1,405 | 1,566 | 0,023 | 1,15E-06 | 3,18E-05 | ACACA,ACACB,ADM,AKAP13,AKR1B1,ALDH1A3,ARL4C,CASP1,CASP3,CASP8,CAV3,CITED2,CTCF,DDIT3,DEFB1,EIF3A,ENO1,FADD,FKBP1A,FN1,FOXP3,GLUD1,GSTM5,HIF1A,HK1,HSP90B1,IFI16,IL13,INHA,IQSEC2,IRAK1,IRF8,KLK3,LEP,MAPT,MB,MLXIPL,MUC1,MYOG,NAB2,NOS2,OAS1,PCNA,PDHA1,PDHX,PDK2,PDK4,PECAM1,PFKP,PFN1,PKM,PLD2,PNLIP,PPARA,PPARG,PPIB,PRX,PTEN,RAB4A,RAP1GDS1,RHOA,RORC,SERPINB1,SGCA,SLC12A2,SLC2A4,SLC8A2,SNCG,SND1,SOAT1,SOX4,SQSTM1,STAT1,STAT3,TBX21,TMPRSS2,TNFRSF11B,TNFSF10,TP53,UBE2C,VCAM1,VEGFA |
| TLR4 | 0,059 | transmembrane receptor | Activated | 1,632 | 2,069 | 0,059 | 1,17E-06 | 3,21E-05 | ADM,APOBEC3B,APP,ATF3,BAX,C3,CASP7,CASP8,CAST,CCL1,CCL20,CD163,CD44,CD70,CERT1,CFB,CISH,CITED2,CLN3,CSF3,CXCL6,CYP27B1,CYP3A5,DDIT3,DUSP2,EGF,F3,FAS,FGL2,GADD45G,GBP2,HBEGF,HDAC1,HMOX1,HSPA5,HTRA1,IFI16,IFIT2,IFIT3,IL13,IL15,IL18,IRF7,IRF8,ISG15,LCN2,LHB,LTA,MET,METTL1,MMP1,MX1,NAA80,NFKB1,NFKBIA,NMI,NOS1,NOS2,NR4A1,NR4A3,NSMAF,PLA1A,PLK2,PPARG,RAC1,RGS16,RXRA,SGPL1,SH3BP5,SIRT3,SMPDL3B,SOD2,SPP1,SPTLC1,SPTLC2,STAT1,STAT3,STAT5A,TLR2,TLR3,TNFRSF13B,TNFSF10,TPM1,TRIM21,TRIM38,TSPO,UGCG,VCAM1 |
| ATF4 | | transcription regulator | | -0,849 | -0,445 | 0,067 | 1,19E-06 | 3,27E-05 | ABCA1,ABCG2,ASNS,ATF3,CA9,CANX,CDKN1B,CHAC1,CTH,DDIT3,FN1,FUT7,GADD45A,GARS1,GCH1,GRB10,HNF4A,HSP90B1,HSPA5,IHH,KLF9,MARS1,MITF,MTHFD2,NARS1,NDRG1,OSMR,PCK1,PCK2,PPARG,PPP1R15A,PRRX2,PSEN1,PYCR1,SLC38A3,SOD2,SST,STAT3,STC2,TKT,TNFRSF11B,VEGFA,WARS1,WNT11,XDH,XPOT |

| | | | | | | | | | |
|---|---|---|---|---|---|---|---|---|---|
| SMAD3 | | transcription regulator | | -0,269 | 0,051 | 0,045 | 1,21E-06 | 3,30E-05 | ALB,APOA1,APOB,APOC3,C3,CCN2,CD59,CDK4,COL1A1,COL1A2,COL3A1,COL6A3,DDIT3,EGR1,ELN,ESR1,F12,FN1,FOS,FOXP3,G6PC,GADD45B,GAST,GLI1,GZMA,HBEGF,HMOX1,HNF4A,HSP90B1,HSPA5,IL13,IL15,INHA,JUNB,JUND,KLK3,LEFTY2,LHB,LUM,MMP1,MYOD1,NET1,NOS2,PCNA,PDCD1,PTPRK,RAC1,RHOB,S1PR4,SLC5A5,SMAD1,SNAI1,SPP1,SSTR2,STAT3,TBX21,TERT,TIMP1,TNC,TPM1,VEGFA,VIM,ZFP36 |
| HOXA10 | 0,235 | transcription regulator | | -0,270 | -0,444 | -0,026 | 1,26E-06 | 3,42E-05 | ADIPOQ,ADM,AIMP2,ALDH1A1,ALOX5AP,ALPL,ARL2,B3GALNT1,CFD,COL15A1,COL3A1,CSTA,CTSG,CTSV,CYBB,DPP4,EIF3A,ELOC,FCER1G,FKBP3,FN1,FZD1,FZD5,GAS6,GSTA3,HLA-DQB1,ITGB3,KLF9,KRT4,LCN2,LEPR,LTF,LYZ,ME1,MMP26,MSN,NCF2,NR4A1,P4HB,PIGR,PRKCD,PROM1,PROS1,PTGER3,RBP1,TGFBR3,TP53,WNT10B,XDH,ZNF22 |
| SRC | | kinase | | -0,611 | -0,361 | 0,046 | 1,30E-06 | 3,53E-05 | ACAT1,CA2,CDK1,CGA,COL1A2,CSF3,CTSV,DUSP1,ENO1,F3,FN1,FOS,FOSL1,GBP1,HIF1A,INS,KLK3,LAMP1,LDLR,LHB,MMP1,MYOD1,MYOG,NFKB1,NPPA,POSTN,PRKCA,PRKCD,PRL,SPP1,STAT3,SYNPO,TFPI2,VCAM1,VEGFA,VIM |
| AHR | 0,308 | ligand-dependent nuclear receptor | | -1,189 | -1,119 | 0,009 | 1,31E-06 | 3,54E-05 | A2M,ABCC4,ABCG2,ACACA,ACTL6A,ACTN1,ADM,ALB,ALDH2,ALDH3A1,ALOX5,APP,ATP4A,BAX,BTG2,BZW2,CAMK2N1,CASP8,CCL1,CCNA2,CCNB1,CDC25B,CDK1,CDK4,CDKN1B,COL1A1,COL1A2,COL3A1,COL4A1,COL4A2,COL5A2,COL6A3,CTSD,CYP2A6 (includes others),CYP2B6,CYP2C8,CYP3A4,DIO1,ENO1,ESR1,ESRRG,FAS,FN1,FOS,FOSL1,FOXP3,GADD45A,GAS6,GSTA1,GSTM5,HIF1A,HSPA5,ITGA6,ITGA7,JUNB,LTBP2,NFE2L2,NMT2,NNMT,NOS2,NPTX1,PCK1,PJA1,PKLR,PLXNA2,PPARG,PPP1R15A,PRL,RBP1,RDH16,RORC,SLC16A5,SNAI1,SNTA1,SOD2,SPP1,STAT3,TGFBI,TIPARP,TP53,TP73,TSPYL2,VEGFA,VIM |
| KITLG | 0,309 | growth factor | | 0,809 | 1,176 | 0,064 | 1,32E-06 | 3,54E-05 | ALOX5,ALOX5AP,B2M,BZW2,CA2,CASP1,CASP3,CCL22,CCN2,CCNA2,CDKN1B,CIAPIN1,CISH,CLIC1,DHCR24,EGR1,FOS,FRK,GAS6,GCC2,HMOX1,HNRNPF,IK,IL13,IL18,ITGAV,ITGB3,KPNA2,KPNB1,LAPTM5,LDLR,LTF,MME,NFKBIA,NPPA,PDCD4,PGLS,PIK3IP1,PIM1,PIWIL1,PKM,PLSCR1,PRKCA,PTGES3,RAB4A,RBBP7,SAMSN1,SHC1,SLC4A1,SNAP23,ST3GAL4,STAT3,TCF12,TERT,TSC22D3,UCP2,UGCG,WDR1,WEE1,ZNF207 |
| IRS1 | -0,035 | enzyme | | -0,979 | -0,861 | 0,031 | 1,32E-06 | 3,55E-05 | ACACA,ACTR3,ADIPOQ,COL3A1,COL4A1,COL4A2,COL6A3,CRABP2,DBN1,DDIT3,EGR1,FHL1,FOS,G6PC,GCK,GLRB,HMOX1,IGFBP4,INS,INSR,LDLR,LEP,LPL,MYBL2,NR2F1,NRP1,PCK1,PPARG,PTPRF,SLC2A4,SNAI1,SPP1,TGFBI,TNNC1,TNNI1,TNNT2,UCP2,VEGFA,WNT6 |
| RHO | | G-protein coupled receptor | | -1,149 | -1,091 | 0,013 | 1,36E-06 | 3,64E-05 | APP,B2M,C1QA,CASP3,CASP7,CD44,CD74,COL1A2,CRABP1,CTSS,DDIT3,FOS,GNAT1,GNB1,H1-2,HCLS1,HMGB2,IMPDH1,LAMB2,PDE6A,PRPH,S100A6,SERPING1,SGK1 |
| CHUK | 0,018 | kinase | Activated | 1,726 | 2,172 | 0,065 | 1,43E-06 | 3,80E-05 | ACKR3,ARL6IP5,ATF3,C3,CASP4,CCNB1,CDKN1B,CLU,COL18A1,CTSF,CTSZ,CXCL6,CYBA,CYP7B1,DNMT1,EGF,ENAH,F3,FAS,FOS,FOXC2,GADD45B,GBP2,GCH1,GM2A,GRN,HLA-A,HMOX1,IFI16,IFI35,IFIT2,IL13,ISG15,ITGB3,JUNB,LCN2,NFKBIA,NGF,NOCT,OVOL1,PCNA,PLSCR1,RB1,RBP1,RGS16,SERPINE2,SGK1,SNX10,SPP1,SQSTM1,TERF2IP,TLR3,TNFRSF11B,TP53,VCAM1,VEGFA |
| cyclic AMP | | chemical - endogenous mammalian | Inhibited | -4,280 | -3,882 | 0,051 | 1,46E-06 | 3,86E-05 | ABCA1,APP,ARC,ASS1,CA2,CACNA1H,CCN2,CDK4,CDKN1B,CGA,CRYAB,CTLA4,CYP11B2,CYP2A6 (includes others),DCT,DDIT3,EGR1,EPCAM,ESRRA,EVX1,F3,FN1,FOS,G6PC,GATA6,GCH1,GCK,HBEGF,HIF1A,IGFBP4,IL15,INHA,INSR,JUNB,LEP,LGALS3BP,LHB,MEST,MITF,MME,MMP1,NGF,NOS1,NOS2,NR4A1,NTF3,OAT,ODC1,PAH,PCK1,PER1,PFKFB1,PRL,RORC,RXRA,SLC37A4,SLC5A5,SOD2,SST,STAT1,STAT3,TIMP1,TNFSF10,TP53,TRPV4,WNT10B |
| palmitic acid | | chemical - endogenous mammalian | | 0,776 | 1,009 | 0,032 | 1,61E-06 | 4,24E-05 | ACAA1,ACACA,ADIPOQ,AHR,ALDOA,APOM,ATF3,ATG5,BAX,BECN1,CASP3,CCN2,CYBA,CYBB,CYP4F12,DDIT3,ELOVL6,F2RL1,FABP1,FABP2,FN1,FOS,FOXO1,G6PC,GCK,HMOX1,HNF1A,HNF4A,HSPA5,IL6R,INS,INSR,LCN2,LEP,MET,MLXIPL,NCF2,NFE2L2,NFKBIA,NOS2,NR4A1,NR4A3,PCK1,PDK4,PKLR,PNPLA2,PNPLA3,PPARA,PPARG,PTEN,SLC2A4,SQSTM1,TLR2,TNFSF10,TP53,UCHL1,UCP2,VCAM1 |

| | | | | | | | | | |
|---|---|---|---|---|---|---|---|---|---|
| CSF2 | | cytokine | | 1,009 | 1,577 | 0,062 | 1,65E-06 | 4,29E-05 | ABCA1,ABCG1,ALOX5,ALOX5AP,ANXA1,AURKA,BAX,BID,C3,CASP1,CASP3,CCL17,CCL25,CCNA2,CCNB1,CD163,CD1A,CD1B,CD24,CD33,CD5,CD63,CD74,CD83,CDC123,CDC20,CDK1,CDKN1B,CISH,CSF3,CTLA4,CYBB,DDIT3,EGR1,EGR3,ELN,EPCAM,F2RL1,F3,FANCL,FAS,FOS,FOSL1,FOXP3,GCH1,GK,HBEGF,HIF1A,HLA-DQB1,IK,IL13,IL15,IL32,IRAK1,ITGAE,ITGB3,JUNB,LEP,LTB,LY75,LY96,MAEA,MCM3,MCM5,MET,MMD,MME,MMP1,MNS1,NET1,NFKB1,NFKBIA,NOS2,NR4A1,NRP1,ODC1,OSMR,PDE1B,PIM1,PLAGL2,PPARG,PPIF,PPP1R15A,PRC1,PSEN1,PTK2,RB1,RRM2,SGK1,SLC2A4,SNAP23,SOCS2,SOD2,SPP1,SQSTM1,STMN1,TERT,TLR1,TLR2,TNFRSF10D,TP53,UBD,UBE2C,VDR,VTCN1,ZFP36 |
| NKX2-3 | | transcription regulator | Inhibited | -2,211 | -2,200 | 0,001 | 1,65E-06 | 4,29E-05 | ACO1,ADM,ALPL,ANKRD55,APOBEC3G,ARHGDIB,ASNS,CASP1,CAVIN2,CCNB2,CDKN3,CLEC1A,CRYAB,CXADR,DDX58,DDX60,F2RL1,GBP1,GBP2,GCA,GCH1,GHR,GNG12,HCP5,HLA-B,HLA-C,HLA-F,KRT81,LYPD1,MMP7,NETO2,NR2F1,NT5DC2,PARP12,PLA1A,PLSCR1,POSTN,PSMB8,PSMB9,RTP4,SERPINE2,SLC25A28,SNCAIP,SP110,STAT1,SYT11,TAP1,TFPI2,TIPARP,TNFRSF10D,TRIM21,TRIM22,TRIM5,UBE2L6,VCAM1,ZC3HAV1,ZMAT3 |
| 15-deoxy-delta-12,14 -PGJ 2 | | chemical - endogenous mammalian | | 0,627 | 0,449 | -0,026 | 1,79E-06 | 4,63E-05 | BAX,BECN1,BIRC2,CASP3,CAV2,CCN2,CCNB1,CDK4,COL1A1,COL1A2,CTSD,CYBB,DDIT3,DDX58,E2F4,EGR1,ESR1,FABP2,FN1,FOS,GADD45A,GSTP1,HLA-DRA,HMOX1,KLK3,LAMP1,MMP1,MUC1,MYOD1,MYOG,NFE2L2,NFKB1,NFKBIA,NOS2,P4HA1,PARP1,PLAAT4,PPARG,PTGES2,RORA,SLC2A4,SOAT1,SPP1,STAT1,TBXA2R,TMPRSS2,TP53,TXNRD1,VCAM1,VEGFA |
| IGF1R | 0,010 | transmembrane receptor | | -0,780 | -0,455 | 0,052 | 1,85E-06 | 4,78E-05 | ABCC4,ALDOA,ANXA3,APP,ATOH1,BAX,CALU,CAPG,CAVIN3,CBFB,CD48,CD53,CDK1,CDKN1B,CKS2,CLU,COL18A1,COL1A1,COL3A1,COL4A1,COL4A2,COX6A2,CSHL1,CTSD,DDIT3,DIO3,EGR1,ESR1,FOS,GCK,GHRH,GLI1,GRB10,HIF1A,HSPA5,HTRA1,IFNGR2,IGFBP4,KIF20A,KRT19,LASP1,MYOD1,MYOG,MYOM2,NME1,NR4A1,PDCD10,PKM,PROM1,RAC1,RBBP7,RFC2,RHOA,RRAGC,S100A4,SNAI1,SNCG,SST,STAT3,TGFBI,TMEM87A,TNFSF10,TNNI1,TP53,TYMS,UBE2V2,VEGFA |
| IDH1 | -0,129 | enzyme | | 0,905 | 1,033 | 0,036 | 1,90E-06 | 4,89E-05 | CD47,CFD,CLDN3,CREB3L1,DDIT3,EGR1,FBP1,FN1,G6PC,GLI1,GLUD1,LEP,LPL,MAPK10,PCK1,PKM,PPARG,PPP1R15A,SLC2A4,TP53,VIM |
| CDKN2A | | transcription regulator | | -1,863 | -1,922 | -0,007 | 1,93E-06 | 4,96E-05 | A2M,BAX,BTG2,C3,CAPG,CASP3,CCL1,CCL22,CCN2,CCNA2,CCNB1,CCR4,CDC25B,CDK1,CDKN1B,CITED2,COL1A2,CUL1,CXCL6,DCK,DCTN4,DDR1,DEK,DUSP1,EED,EGR1,FAM111A,FANCL,FHL2,FOS,GADD45G,GLB1,GNAS,GNL3,HMGB2,IFI35,IL15,IL36A,ITGAV,ITGB3,KLK3,KRAS,LAMP1,LTA,MAD2L1,MCM5,MELK,MMP1,MYBL2,ODC1,PCNA,POLD3,PPFIBP1,PSMA5,PSMB9,PTEN,RAB27A,RB1,RBBP7,RFC4,RMI1,RRM1,RRM2,S100A6,SAE1,TMPO,TOB1,TP53,TPM1,VEGFA,VIM,ZMAT3 |
| CREM | | transcription regulator | Inhibited | -2,412 | -2,324 | 0,016 | 2,01E-06 | 5,15E-05 | ABCA1,ANXA4,APOLD1,ARC,ATF3,BHLHE40,BTG2,CCNB1,CGA,COL3A1,CRABP2,CYP11B2,DUSP1,EGR1,FOS,FUT7,GADD45B,GJD2,HLA-DRA,HSPA4,IDI1,INA,INHA,INS,IRAK1,JUNB,LDLR,MEST,MNS1,NAB2,NFIL3,NOS1,NOS2,NR4A1,PDYN,PER1,PPP1R15A,PRL,RHOB,SMC4,SST,TIPARP,VEGFA,ZNRF4 |
| GH1 | | growth factor | Inhibited | -2,843 | -2,762 | 0,014 | 2,06E-06 | 5,26E-05 | ABCC1,ATF3,CCNB1,CISH,CLU,CSHL1,CYP3A4,DBI,DDIT3,EGR1,FN1,FOS,GADD45A,GAST,GHR,GHRH,GTF2H1,HDAC1,IGFALS,JUNB,LDLR,NOS2,NRAS,PTEN,RAD23B,RHOA,RYK,SMARCA4,SNAI1,SOCS2,SPP1,TAF9,TKT,VEZF1,VIM,ZFP36 |
| S100A9 | 0,438 | other | | 1,553 | 1,728 | 0,026 | 2,11E-06 | 5,36E-05 | A2M,ABCG1,ADAMTS1,AKAP12,ALDH1A1,ALPL,ATF3,BECN1,BHLHE40,C1QB,C1S,C3,CASP1,CCN5,CDH3,COX6A2,CTLA4,ELN,EPCAM,FAS,FOSL1,FURIN,ISG15,KLF15,MMP1,MMP17,NOS2,NR4A3,NTRK3,PCDHGB5,PER1,PIK3IP1,PLK3,PLXNB3,POSTN,PTPRO,RBFOX1,RNASE6,SEMA3B,SGK1,SHC3,SLC26A1,SOCS7,TNFAIP8,VCAM1,VCAN,WT1 |
| PDX1 | | transcription regulator | | -1,269 | -1,153 | 0,020 | 2,11E-06 | 5,36E-05 | ACTG1,AKR1B1,ALB,ALDOB,ANXA2,ASNS,ATF3,ATG5,CAMK2N1,CAT,CD44,COL1A1,COL3A1,CRELD2,DNAJC10,EGR1,F3,FGF1,FGF5,FUT1,G6PC,GCH1,GCK,GNAS,GNG12,GRN,HAP1,HK1,HNF4A,HSPA5,INS,JUNB,LRRC49,LXN,MAPT,MITF,NFKB1,NNAT,PCNA,PDIA4,PDIA6,PPP1R1A,PTGER3,PTPRF,RHOBTB1,SPP1,SSBP3,SST,TGFBR3,TIPRL,TSPAN8 |

| | | | | | | | | | |
|---|---|---|---|---|---|---|---|---|---|
| RBL1 | | transcription regulator | | -0,852 | -1,083 | -0,056 | 2,19E-06 | 5,50E-05 | CASP4,CASP6,CCNA2,CDK1,CDKN1B,E2F4,F3,FAS,FOS,FOXN1,H2AC8,IVL,KRT10,MCM2,MCM3,MCM5,MET,MYBL2,PCNA,PPP1R8,RB1,RRM1,RRM2,TERT,TP53,TP73,TYMS |
| SEMA7A | -0,192 | transmembrane receptor | | 1,151 | 1,437 | 0,080 | 2,32E-06 | 5,80E-05 | BAX,CCN2,CCN5,CCNA2,COL1A1,COL1A2,COL3A1,CTSS,EGR1,ELN,FN1,IL18,TIMP1 |
| POU5F1 | | transcription regulator | | 0,304 | 0,530 | 0,028 | 2,50E-06 | 6,19E-05 | AFP,AHR,ALB,ANKRD17,AQR,AXIN1,BAG1,BAX,BID,BIRC2,BNIP3L,CASP1,CASP10,CASP3,CASP6,CASP7,CD70,CDX2,CGA,CRABP2,CST6,CYP26A1,DLX5,DNMT1,DUSP1,EED,EPCAM,EVX1,EXT2,FABP1,FABP2,FADD,FARP1,FAS,FGF5,FOXF1,FUT4,GADD45A,GADD45G,GATA6,GCK,HAND1,HIF1A,HOPX,HOXB1,KLK14,LEFTY2,LOXL1,LTA,MYBL2,NFATC4,NR2F2,NRP1,OVOL1,PARP1,PECAM1,PODXL,PPARA,PPL,PRDM14,PROM1,PYCARD,SH3GLB1,SMAD1,SNAI1,SPP1,SPRR3,TDRD7,TGM1,TNFRSF11B,TNFSF10,TP53,TP53BP2,TP73,TXNRD1,VEGFA,VIM,WNT8B,YAP1 |
| SYVN1 | | transporter | | 1,533 | 1,998 | 0,074 | 2,62E-06 | 6,46E-05 | ABCC3,ABCC4,ADAM9,AHR,APP,BHLHE40,CALM1 (includes others),CD151,CD44,CDA,DUSP1,EMC7,EXT2,F3,FAS,HLA-A,HLA-C,HNRNPM,IFI44,IFITM2,ITGA6,ITGAV,KCNN4,LGALS3BP,MCAM,MYOF,NFE2L2,PFN1,PLD2,PNPLA2,PODXL,RPN2,SETX,SLC12A3,SLC27A3,SNAP23,TARS1,TNS4,ZC3HAV1,ZNF281 |
| folic acid | | chemical - endogenous mammalian | | 0,329 | 0,342 | 0,003 | 2,65E-06 | 6,52E-05 | ABCC1,ABCG2,APP,CLU,COL1A1,COL3A1,CSNK1A1,CYBB,DDIT3,DNMT1,FABP1,FN1,FZD1,FZD6,HDAC6,NOS2,PCNA,PECAM1,PLG,PSEN1,TP53,TYMS,VCAM1,WNT6 |
| IRF5 | | transcription regulator | Activated | 3,771 | 4,082 | 0,061 | 2,67E-06 | 6,54E-05 | BAX,BID,CASP3,COL1A1,COL3A1,COL4A1,DAPK2,DDX58,FOXO1,IFI44,IFIH1,IFIT2,IFIT3,IRF7,ISG15,NAMPT,OAS1,PARP12,PDCD2,PLSCR1,SP110,STAT1,TMPO,TNFSF10,UBE2L6,USP8,VEGFA,YWHAE |
| CHD4 | | enzyme | Activated | 2,317 | 2,065 | -0,058 | 2,75E-06 | 6,68E-05 | ARC,CASQ1,CDA,CDKN1B,CPLX3,CPNE6,CPNE7,FAS,GNB3,HBB,HMOX1,IL13,NHLH1,NT5DC2,PDE1B,PRPH,RAD50,RORC,SPRY1,SYDE1,TBX21,TNNC2,TP53 |
| ARNT | | transcription regulator | | -0,217 | 0,051 | 0,064 | 2,76E-06 | 6,68E-05 | ADM,AHR,ALDH3A1,ALDH7A1,ALDOA,ALDOC,BHLHE40,BHLHE41,CA9,CD81,CDK4,CITED2,CLDN1,CSF3,CTSD,CYP2A6 (includes others),DNAJC2,ENO1,ESRRG,FURIN,G6PC,GPI,GSTA1,HIF1A,HNF1A,HNF4A,INSR,ITGAV,ITGB3,KIF20A,LEP,MYL6,PCK2,TP53,UBE2C,VEGFA,VIM |
| EPO | | cytokine | Inhibited | -2,385 | -2,152 | 0,042 | 2,77E-06 | 6,68E-05 | B2M,BAX,BNIP3L,BZW2,CA1,CA2,CASP3,CD52,CDKN1B,CISH,CLIC1,CTSZ,CUL1,DCSTAMP,DDIT3,DHCR24,EGR1,FABP1,FAS,FOS,FOSB,G6PC,GCC2,GLUL,GYPA,HBB,HLA-DRB1,HNRNPF,IL18,ILF2,JUNB,KCNH6,KPNA2,KPNB1,LAPTM5,LDLR,MACROH2A1,MAN2A1,MIA2,MYOD1,MYOG,NAB2,NFKBIA,PCNA,PDCD4,PIK3IP1,PIM1,PLCB2,PRDM14,PRKCA,PRKCD,PTPRB,RAB4A,RHOA,ROCK1,SAMSN1,SLC4A1,SNAP23,SOCS2,SPP1,ST3GAL4,STAT3,TNFRSF11B,TNFSF10,TP53,TSC22D3,UCP2,UGCG,VEGFA,WDR1 |
| MITF | 0,269 | transcription regulator | | 0,158 | 0,683 | 0,071 | 2,78E-06 | 6,70E-05 | APOLD1,ATP6V1C1,BEST1,CA14,CCNB1,CD151,CD44,CDC25B,CENPM,CLCN7,COL1A1,CTSG,DCSTAMP,DCT,ECT2,ESPL1,FN1,FOS,GAPDHS,GLI1,GM2A,GNPTAB,HIF1A,HPGD,IL6R,ITPKB,KCNN2,KIF20A,MC1R,MCM2,MCM5,MET,MGAT4B,MITF,MMS19,PLA1A,PMEL,POLE2,POLM,RAB27A,RFC5,RHOQ,RMI1,RPA3,SHTN1,SLC19A2,SNAI1,SNW1,SOX10,SOX13,ST3GAL6,STX7,TBC1D16,TBX2,TDRD7,TERT,TMEM251,TP53,UBE2C,YAP1 |
| L-triiodothyronine | | chemical - endogenous mammalian | Inhibited | -2,261 | -2,054 | 0,024 | 2,89E-06 | 6,94E-05 | A2M,ABCA1,ADIPOQ,ADM,AFP,ALB,ALDH1A1,APOA1,APOB,APOC1,APOC3,APOM,APP,ATIC,ATP1A3,BNIP3L,C1S,CA1,CCN2,CD44,CDKN1B,CGA,CLU,COL4A1,CSHL1,CTH,CYP27B1,CYP4A11,DIO1,DIO3,DUSP1,EGF,EGR1,ESR1,FABP1,FAS,FOS,G6PC,GCK,HIF1A,HR,IGFBP4,INHA,ITGA7,ITGAV,ITGB3,KLF9,LCT,LDLR,LEP,LYZ,MC4R,ME1,MGST3,MLXIPL,MYBPH,MYOD1,NKX2-5,NME1,NOS2,NR4A1,NRGN,NTF3,OAT,ODC1,PARP1,PCK1,PCK2,PDK2,PDK4,PER1,PITX3,PPARA,PPARG,PRL,PTGDS,PTTG1,RGS2,ROCK1,RYK,SEMA7A,SHBG,SLC2A4,SSB,ST3GAL4,THRB,TP53,UCP2 |

| | | | | | | | | | |
|---|---|---|---|---|---|---|---|---|---|
| MYOD1 | -0,148 | transcription regulator | | -2,135 | -1,840 | 0,045 | 2,95E-06 | 7,06E-05 | ACACA,ACACB,ALDOA,ANKRD2,ASS1,CAV3,CBX5,CDH15,CDKN1C,CDX2,CHRND,COL4A1,CRYAB,DAO,DUSP1,ENO3,ESR1,FOSL1,FSTL1,GADD45A,GADD45G,GUCA2A,HADH,HSPB8,IFI16,ITGA7,JUNB,MYBPH,MYCL,MYOD1,MYOG,NPTX1,PNP,POSTN,PTK2,RB1,RRAS,SCN4A,SGCA,SH3GLB1,SLC12A2,SLC2A4,SST,STAM,TGFBR3,TKT,TNNC1,TNNC2,TNNI1,TNNT2,TNS1 |
| EIF4E | 0,070 | translation regulator | Activated | 2,182 | 2,680 | 0,080 | 2,97E-06 | 7,08E-05 | ATR,BCLAF1,BIRC2,CAPG,CD70,CDK4,CKS2,CSHL1,FGF5,FN1,FOSL1,GPS1,HIF1A,HMOX1,HRAS,IGFBP4,IL18,ILF2,LSM1,LSM7,NCK2,NFKBIA,NOP10,NOS2,NRAS,NRP1,ODC1,PCNA,PIM1,POP7,PRG2,PRL,PTGES3,RB1,SIRT2,SLC6A4,SNAI1,TGFBI,TNC,TP53,TP53BP1,TP53BP2,TRIP6,UBE2A,UBE2C |
| CD28 | | transmembrane receptor | | -0,124 | 0,311 | 0,050 | 2,98E-06 | 7,08E-05 | ACTR3,AHR,ANXA1,ARHGDIB,BHLHE40,C2,CAMK2A,CASP3,CASP6,CASP8,CCR4,CDK4,CDK7,CDKN1B,CISH,CNIH4,CRLF2,CTLA4,DAD1,DDX58,DUSP22,E2F4,FCER1G,FOS,FOXP3,GBP1,GBP2,GLI1,GNL2,GTF2IRD1,GTPBP4,GUCY1B1,GZMA,HAT1,HAX1,HCCS,HLA-DQB1,IFITM1,IFNGR1,IL10RA,IL10RB,IL13,IL7,ILF2,IRF8,KDELR2,LTA,MACROH2A1,MAP3K10,NAMPT,NFKB1,NFKBIA,NOS1,NR4A1,ODC1,PDCD1,PKLR,PLA2R1,PNP,PPA1,PRDX4,PSMA5,PSMA6,PTEN,RAD23B,RHOC,RORA,RORC,RYK,SEC61B,SH2D2A,SNRPB,SOCS2,SSB,STAT3,TBX21,TFPI2,TNFSF10,TOB1,TRIM24,TSTA3,VBP1 |
| POR | -0,004 | enzyme | | -1,362 | -1,479 | -0,029 | 3,00E-06 | 7,10E-05 | ABCA1,ABCC3,ACTG1,AHR,ALDH1A1,ANXA5,AQP8,ASNS,CD74,CLDN3,CSAD,CYP26A1,CYP2A6 (includes others),CYP2B6,CYP2C8,CYP4A11,CYP7B1,DHCR24,ELOVL1,ELOVL6,FABP2,FGF1,GADD45B,GBP2,GCK,GSTA3,GSTM5,HIF1A,HLA-A,HLA-DMA,HLA-DQB1,HMOX1,IDI1,IHH,LDLR,LGALS1,LPL,NAA80,NOCT,NR0B2,PDK4,PGD,PPARG,SERPINA3,STAT1,TMEM184C,TMEM267,UBD,VEGFA |
| TSC2 | -0,100 | other | | -1,504 | -1,726 | -0,039 | 3,08E-06 | 7,26E-05 | A2M,ANXA1,ANXA2,ARC,ATF3,BAX,CASP1,CDKN1B,CEACAM4,CRYAB,CST6,DDIT3,EGR1,EMP3,FOS,GLA,HIF1A,HMOX1,HSPA5,NEU1,NIBAN1,NTRK3,OLFM4,PDK4,PLP2,PRELP,PRKCA,PSMA1,PSMD1,PSMD14,PTEN,PVALB,S100A11,S100A6,SOD2,STAT1,UCP2,VAMP8,ZFP36 |
| SOD1 | -0,061 | enzyme | | -1,097 | -1,219 | -0,047 | 3,31E-06 | 7,76E-05 | ACO1,ALDOC,APP,ASS1,ATF1,ATP1A3,B2M,BAX,BID,C1QA,CA7,CACNB4,CASP1,CASP3,CASP4,CAST,CCNC,CDK4,CGA,CLU,COL1A1,COMT,CRYAB,DAO,DCXR,DSG3,FGF6,FOS,GNAS,GNB1,GNG7,GSTM5,GSTP1,HMOX1,HSBP1,HSPA4,HSPB8,IL15,JPT1,JUNB,LPL,MET,NETO2,NFE2L2,NFIL3,NOS1,NOS2,P2RX4,PLCB4,PRPH,PSMB8,RAC1,RPGRIP1,SH3BGRL3,SLC1A2,SNCG,SOD2,SPP1,SQSTM1,STMN1,TP53,TUBB4A,VCAM1 |
| Mek | | group | | -1,133 | -0,947 | 0,027 | 3,44E-06 | 8,04E-05 | ABCE1,ABCG2,ACAT1,ANPEP,BECN1,BID,CCN2,CDKN1B,CDX2,CLU,COL1A2,CRYAB,DCT,DDR1,DDX58,DNMT1,EGR1,EGR3,ESR1,FGFBP1,FOS,FOSL1,FOXP3,GBP2,GNL3,HAND1,HIF1A,IER2,IL15,ITGA6,ITGB3,IVL,JUNB,LEP,LHB,MAFF,MANSC1,MST1R,NES,NGF,NR4A1,PIWIL1,PMEL,PRL,PSEN1,PTEN,PUS7,RABGGTB,ROCK1,SACS,SH2B3,SLC5A5,SNCG,SPRY1,SPRY2,SPRY4,TP53,VIM |
| 26s Proteasome | | complex | | -0,023 | -0,253 | -0,036 | 3,47E-06 | 8,10E-05 | AHR,ATG5,BAX,BIRC2,CCNA2,CDK1,CDKN1B,CLCN7,CTSA,CTSD,CTSF,DDIT3,DDX58,EGF,ESR1,FOSL1,FOXO1,GCH1,HBEGF,HMGB2,HSBP1,HSPB8,IFI35,JUNB,LAMP1,MAD2L1,NFKB1,PRKCA,PSMC4,PTEN,RB1,SMAD2,SNAI1,SQSTM1,STAT1,STAT5A,TNFSF10,TP53,UBD,UBE2I,UBFD1,VIM,WBP2 |
| MTPN | | transcription regulator | Activated | 2,854 | 3,270 | 0,080 | 3,51E-06 | 8,17E-05 | ANXA1,APOD,BAX,CASP3,CASP7,CASP8,CASQ1,CCNA2,CCNB1,CCNB2,CCNC,CDK1,COL1A1,COL1A2,FADD,FAS,FN1,FOS,GAS6,LTB,NFKB1,NPPA,S100A10,S100A6,SPP1,TNFSF10,TUBA1A,VCAM1 |
| PTH | | other | | 0,382 | 0,655 | 0,048 | 3,56E-06 | 8,27E-05 | ADAMTS1,ALPL,BHLHE40,CCN2,CCN5,CDKN1B,COL1A1,COL1A2,CYBA,CYP24A1,CYP27B1,CYP3A4,DDIT3,DUSP1,FN1,FOS,FOSB,GALNT3,GNG12,HSPA5,IL6R,ITGAV,MEPE,MME,MMP1,NFIL3,NR4A1,ODC1,PALM,POSTN,PPP1R1A,RGS2,SEC61B,SLC2A4,SLC34A1,SPP1,ST14,TFPI2,THBS4,TNFRSF11B,TNS1,TRPV5,VDR,WIF1,WNT10B |
| arachidonic acid | | chemical - endogenous mammalian | | -0,583 | -0,511 | 0,014 | 3,61E-06 | 8,35E-05 | ABCA1,ABCG1,ACACA,CD83,CLU,COL1A2,DDIT3,DNAJB9,EGR1,ELOVL6,F3,FABP1,FAS,FOS,FOSL1,GLI1,HMOX1,LDLR,NFKBIA,PECAM1,PKLR,PLTP,PNPLA2,PPARG,PRKD1,RGS2,RXRA,SLC2A4,SOAT1,UCP2,VCAM1,VIM |
| NCOR1 | | transcription regulator | Activated | 2,267 | 2,121 | -0,052 | 3,64E-06 | 8,38E-05 | ADIPOQ,AFP,AZGP1,CCNA2,CDKN1B,CEBPE,CGA,DBI,DHCR24,DIO1,DUSP1,FXYD3,G6PC,HK1,KLK3,LYZ,MB,ME1,MME,MRPL49,MT1X,MYOD1,MYOG,NDRG1,PDK4,PFKFB1,PPARG,PRKCD,STK39,WARS2 |

| | | | | | | | | | |
|---|---|---|---|---|---|---|---|---|---|
| TCF | | group | | -1,248 | -1,091 | 0,080 | 3,64E-06 | 8,38E-05 | AKAP13,ALDH1A1,ALDH3A2,APOD,CCT8,CD44,CRAT,CYP24A1,DPEP1,EGR1,EPCAM,EPHB3,FOS,GAD1,HTRA1,IFITM1,IHH,JUNB,KLK3,MME,MMP7,MUC6,NRCAM,PLS3,QPCT,SERPINA3,SFN,SLC26A2,TSPAN8,UGCG |
| NR0B2 | -0,721 | ligand-dependent nuclear receptor | Activated | 4,189 | 3,903 | -0,063 | 3,88E-06 | 8,88E-05 | ABCA1,ACACA,APOA1,APOM,BAAT,CEL,CYP11B2,CYP7B1,EGR1,ESRRG,FBP1,G6PC,GCK,HNF1A,HNF4A,NR0B2,P4HB,PCK1,PCK2,PDIA3,PDIA4,PDK4,PPARG,SLC10A1,SLC10A2,SPP1,TIMP1 |
| CYP27B1 | -1,287 | enzyme | | -1,198 | -1,270 | -0,021 | 3,92E-06 | 8,94E-05 | ALPL,CCL1,CCL17,CCL20,CCR4,CTLA4,CYP27B1,CYP3A4,CYP3A5,IL15,IL18,SLC34A1,SPP1,TLR2,TRPV5,VDR |
| EZH2 | 0,264 | transcription regulator | | 0,475 | 0,477 | 0,000 | 3,95E-06 | 8,97E-05 | ACKR3,ADAM10,ALOX5AP,ALPL,ATF3,ATOH1,BMP7,BMPR1B,C3,CACNA1E,CACNG4,CD63,CD70,CDK4,CDKN1B,CDX2,CER1,COL4A1,CPD,CSF3,CSNK1A1,CTNNA1,CUL1,DNMT1,EGR1,EVX1,FBN1,FGF5,FHOD3,FOXP3,FZD1,GABRA2,HCP5,HOXA10,HOXB6,HOXC13,HSD11B2,IL10RA,IL13,IL36A,KLK3,KRAS,KRT81,LCN2,LTB,MMP7,MPZL2,MYL6,MYOD1,NDRG1,NES,NFKBIA,NME1,NME5,NOTCH2,PARP12,PER1,PER2,PIK3IP1,PLAAT4,PPARG,PRG2,PRL,PROM1,PSMD1,PTEN,RNASEH1,S100A4,SFN,SGPL1,SNAI1,SOX10,SPOCK2,STT3A,TBX21,TMPRSS2,TP53,TRIM38,TSPAN7,UBD,VEGFA,WIF1,WNT10B |
| chenodeoxycholic acid | | chemical - endogenous mammalian | | 0,210 | 0,333 | 0,023 | 4,10E-06 | 9,28E-05 | ABCA1,ABCC3,ABCG1,APOA1,APOB,APOM,COL1A1,CYP3A4,CYP3A5,EGR1,FABP6,FBP1,G6PC,HNF1A,HNF4A,HPGD,LDLR,LIPC,NR0B2,PCK1,PCK2,PDK4,PLTP,PPARA,SLC10A1,SLC10A2,SNAI1,TXNRD1,VDR |
| IL27 | | cytokine | Activated | 3,502 | 3,765 | 0,044 | 4,11E-06 | 9,28E-05 | AHR,B2M,CCL16,CCL20,CD163,CD63,CD74,CDK4,CKLF,CTLA4,EGR1,FOS,FOXP3,GADD45G,GBP2,HLA-A,HLA-B,HLA-C,HLA-DMA,HLA-DMB,HLA-DPA1,HLA-DQB1,HLA-DRA,HLA-E,HPGD,IL10RB,IL13,IL18,MX1,NOS2,OAS1,PIM1,PLAAT4,RORC,STAT1,TAP1,TBX21,TLR3,TNFSF10,UBD,VCAM1,VIM |
| E2F3 | 0,111 | transcription regulator | | 0,744 | 0,962 | 0,038 | 4,24E-06 | 9,54E-05 | AKR1C1/AKR1C2,BECN1,CASP7,CAV2,CCL20,CCNA2,CCNB1,CCNB2,CDK1,COL18A1,CSHL1,DEK,ECHDC2,EED,FAM102A,FOXN1,H1-2,HMGB2,KRAS,LBR,LEP,LPL,MAD2L1,MCM2,MCM3,MCM5,MT1X,MYBL2,NCAPG2,PCLAF,PCNA,PLCB4,PPP1R8,PTGER3,PTTG1,RB1,RBPMS,RRM1,RRM2,TERT,TM9SF2,TMPO,TOP2B,TP53BP2,TP73,UBE2C,WFDC2,ZNF76 |
| STAT5A | -0,279 | transcription regulator | Inhibited | -2,909 | -2,818 | 0,017 | 4,28E-06 | 9,55E-05 | A2M,ADIPOQ,AGR2,ALDH3A1,BAX,CASP3,CASP4,CASP6,CASP8,CD24,CD70,CDC7,CDH17,CDK4,CDKN1B,CISH,CSF3,CYP27B1,ESR1,FAS,FCER1G,FGFBP1,FOS,FOSB,GADD45A,GADD45G,GCNT3,GZMA,H1-2,HOXB6,HSD3B2,IFITM2,IGFALS,IL6R,KLK3,LCN2,LTA,LTB,LY6G6D,MMP7,OAS1,PIM1,PIP,PLAAT4,PPARG,PRL,PSMB8,QPCT,RNF39,RO60,RORA,RPA1,SELENBP1,SERPINA3,SH3GL2,SLC10A1,SLC34A2,SLC7A7,SOCS2,STAT5A,TBX21,TGFBI,TNNC1,TNNI1,TP53,TSPAN13,TSPAN8,WARS1,ZFP36 |
| PSEN1 | 0,214 | peptidase | | 0,077 | 0,029 | -0,009 | 4,47E-06 | 9,92E-05 | ABCC6,ABCG1,ABCG2,ACTG1,ALB,ALDOA,AMPH,APP,ARC,ARF4,ATP1A3,ATP4A,BAX,C1QA,C1QB,C1R,C1S,C3,CALML3,CANX,CAP1,CASP3,CD74,CDX1,CLTB,CTSD,CTSS,CTSZ,CYFIP2,DBI,DNM1L,DPYSL2,DSTN,DUSP1,EGR1,ENO1,EPM2A,ERMP1,FKBP1A,FOS,GADD45B,GNB1,GPX5,GSTP1,HAND1,HK1,HLA-E,HSPA5,INSR,IRF7,MAPK1,MAPT,MME,NDUFS6,NME1,NOS1,NOS2,NTF3,OCEL1,PAK3,PEA15,PKM,PPARG,PRC1,PSEN1,PSENEN,PTPRF,PVALB,RHOQ,SCN10A,SH3GL2,SLC1A2,SQSTM1,STMN1,SUCLG1,SYN1,TIPARP,TOP2B,TP53,TP73,TUBA1A,TUBA1B,TUBB,TUBB6,UCHL1,VAMP2,WARS1,YWHAE |
| TFRC | -0,144 | transporter | | -0,732 | -1,043 | -0,065 | 4,59E-06 | 1,01E-04 | ADGRG1,ANXA2,ATF3,CASP3,CCNA2,CCNB1,CDC20,CDK1,CHAC1,CSTF2T,DDIT3,FOS,GADD45A,GPX7,HMOX1,HSPA5,ITGB2,LY75,ME1,MS4A6A,PIP5K1B,PPARG,PPP1R15A,STC2,TNFSF10,TP53,TREML2,UBE2E1 |
| RAS | | group | | -1,676 | -1,404 | 0,052 | 4,60E-06 | 1,01E-04 | ANPEP,AURKA,B4GALT5,CCN2,CDKN1B,CDX1,CDX2,CYP24A1,EGR1,FN1,FOS,FOSL1,GBP2,GSTP1,HMOX1,IVL,LDLR,LHB,MET,MITF,MMP1,MUC5B,NFKB1,NOS2,ODC1,PDCD4,PRL,PSMD10,PTEN,RAE1,SERPINE2,SNAI1,SOD2,STAT1,TBP,TFPI2,TP53,VEGFA,VIM |
| mir-8 | | microRNA | | -0,911 | -1,201 | -0,050 | 4,60E-06 | 1,01E-04 | ADAMTS1,ATF3,ATG5,BAX,BST1,CD47,CDC25B,CITED2,CLDN3,DLX5,DNAJC3,DUSP26,EPCAM,ESM1,HIF1A,HMOX1,KITLG,LEPR,LSR,MAP3K1,MMP1,MPRIP,MYH10,MYOF,PERP,PTEN,SEC23A,SMAD2,SNAI1,SPRY2,SQSTM1,STAT5A,TOB1,TSPAN1,UBE2D1,VIM,YAP1,YWHAB,ZFP36 |

| | | | | | | | | | |
|---|---|---|---|---|---|---|---|---|---|
| Immunoglobulin | | complex | | -0,714 | -0,708 | 0,001 | 4,77E-06 | 1,04E-04 | ACACA,ACKR2,ACO1,ADIPOR1,ADM,ANPEP,APP,CAPG,CASP4,CCL1,CCL17,CCL22,CD44,CD74,CD83,CDK4,CDKN1B,CDKN1C,CHN2,CISH,CLCA1,CTH,CTSZ,CXCL6,CYP27B1,DPYSL2,DUSP1,EGR1,EGR3,ENO1,ENO3,EXT1,F3,FAIM,FAS,FBP1,FCER1G,FOS,FOSB,FOXP3,G6PC,GCK,GPI,GSK3A,HLA-DQB1,HMOX1,HSD11B2,HSPA6,IFI30,IFNGR2,IGFBP4,IL10RA,IL10RB,IL13,IL17RB,IL18,IL25,IL32,IMPDH2,IRAK1,IRF7,ITGAE,ITGB8,LCN2,LILRB3,LTA,MAFF,MAPT,MMP1,MTHFD2,NAMPT,NCF2,NDUFS6,NFIL3,NFKBIA,NOS1,NOS2,NR4A1,NRP1,PCK1,PCK2,PDCD1,PDHA1,PDIA6,PDK4,PHKB,PKLR,PLSCR1,PPA1,PYGM,RB1,RGS16,RGS2,RORC,RPIA,RRBP1,SOD2,SPP1,STAT3,SUCLG1,TALDO1,TBX21,TGFBR3,TIMP1,TLR2,TXNRD1,UBD,VCAM1,VCAN,VEGFA |
| E2F4 | -0,197 | transcription regulator | | -0,793 | -0,733 | 0,022 | 4,77E-06 | 1,04E-04 | ALG5,BECN1,BUB3,CALM1 (includes others),CASP4,CASP6,CBX5,CCNA2,CDK1,CEP57,CFAP20,CKS2,DROSHA,ECT2,EFHC1,EXOSC8,FAS,FOXN1,FPGT,GINS1,HMGB3,HSP90B1,MAD2L1,MCM3,MCM5,MSH2,MYBL2,NAP1L4,NDC80,PCLAF,PCNA,PCSK1N,PLSCR1,PPARG,PPP1R8,PRC1,PTTG1,RB1,RFC2,RFC4,RPA3,RRM1,RRM2,SMARCA5,SMC4,STAM,STMN1,TEAD4,TERT,TMPO,TOP2B,TP53,TP53BP2,TP73,UBE2C,ZNF267,ZNF83 |
| PIK3R1 | 0,126 | kinase | | 1,227 | 1,511 | 0,057 | 5,11E-06 | 1,11E-04 | CDKN1B,COL1A2,DDIT3,DNAJB9,DNAJC3,F3,FOS,FOXO1,GCH1,HIF1A,HMOX1,HSP90B1,HSPA5,IRF7,ISG15,MITF,NFE2L2,NOS2,PCK1,PDHA1,PKM,PPARG,PTEN,SLC5A5,SOD2,VCAM1,VEGFA |
| LGALS3 | -0,080 | other | | 0,138 | 0,213 | 0,017 | 5,25E-06 | 1,14E-04 | ABCC1,ADIPOQ,BAX,CASP3,CCNB2,CDKN1B,COL3A1,COL4A1,COL5A2,CTSF,CTSV,DDOST,FN1,IL13,ITGA10,ITGA6,ITGAV,KRAS,KRT19,LAMP1,MAP3K10,MUC1,NOS2,NRAS,PNPLA2,PPARG,PRSS3,PTEN,RAC1,RB1,VCAM1 |
| PRKCD | -0,382 | kinase | | -0,249 | 0,010 | 0,038 | 5,73E-06 | 1,23E-04 | ABCA1,ALOXE3,ARHGAP32,BIRC2,CCL20,CCNA2,CD70,COL1A1,COL1A2,DDIT3,FBN1,FOS,FOSL1,FZD1,GLI1,IL13,IL32,IVL,JUNB,LAMP1,LDLR,MPZ,MUC5B,NES,NFKBIA,NOCT,NOS2,NOTCH2,NR0B2,NR4A1,OAS1,ODC1,PDCD4,PLAGL2,PLSCR1,PPP1R15A,PRL,PROM1,SLITRK3,SPRY1,STAT3,TLR2,TNFRSF11B,TP53,TPBG,TRIM48,UGCG,VCAM1,VEGFA,ZFP36 |
| SMAD7 | -0,235 | transcription regulator | | -0,197 | -0,339 | -0,022 | 5,96E-06 | 1,28E-04 | ALDOB,APOC3,BIRC2,BMP5,BMP7,BMPR1B,BZW2,CCN2,CCNA2,CCNB1,CDK1,CDKN1B,CITED2,COL1A1,COL1A2,COL3A1,COL6A3,FAS,FGF5,FN1,FOXO1,FURIN,GADD45A,GAS6,HBEGF,HMOX1,ITGA6,ITGA7,LTBP2,MET,MMP1,NET1,NFKBIA,NOS2,PCNA,PROM1,RAC1,SLC5A5,TERT,TGFBI,TIMP1,VEGFA |
| Mapk | | group | Inhibited | -2,834 | -2,505 | 0,072 | 6,27E-06 | 1,34E-04 | APOA1,APOC3,APP,ARC,CAT,CDKN1B,CHRNB4,COL1A1,CYBB,EGR1,ESR1,F3,FOS,FURIN,HMOX1,HPGD,JUNB,LEP,MMP7,NOS1,NOS2,NR4A3,PER1,PER2,PIGR,PRL,PTEN,SNAI1,SNCG,SPP1,STAT1,VCAM1,VEGFA |
| GATA4 | | transcription regulator | | -1,195 | -1,070 | 0,020 | 6,42E-06 | 1,37E-04 | ACACA,ATOH1,BECN1,CASQ2,CCN2,CDX2,CLDN1,COL1A1,COL1A2,COL3A1,CRABP2,CXCL6,ECM2,EPHX1,ESPL1,FABP1,FABP6,FGF16,FOS,FSTL4,HSD3B2,HSPA5,IFI16,INHA,IRF8,KCNA5,KLK14,LCT,LRRFIP1,LTBP2,MYL7,MYOG,NKX2-5,NPPA,OVOL1,POSTN,PPL,PTPRB,PYY,S100A4,SGPL1,SLC10A2,SPP1,SPRR3,TGFBI,THBS4,TIMP1,TNNC1,TNNT2,TPM1,VCAM1,VIM |
| MAPT | -0,688 | other | | -0,875 | -0,836 | 0,013 | 6,45E-06 | 1,37E-04 | ABCG1,ABCG2,ACAT1,ACTG1,ACTN1,ACTR1A,ADD2,ALB,ALDOA,AMPH,ANXA5,APP,ARF1,ARF4,ATP1A3,ATP4A,BSN,C1R,C1S,C3,CALML3,CAMK2A,CAMK2G,CANX,CAP1,CAPN1,CAPN2,CASP3,CAST,CCNB1,CCT8,CDKN1B,CLTB,CTSD,CYFIP2,DBI,DNM1L,DPYSL2,DSTN,ENO1,ERMP1,FKBP1A,FOS,FUZ,GAD1,GSTM5,GSTP1,GUCY1B1,HK1,HSPA5,MAP1A,MAPK1,MAPK8,MAPT,NDUFS6,NFKB1,NME1,NMNAT2,NOS1,NOS2,PAK3,PCLO,PEA15,PKM,PPA1,PSEN1,PVALB,SH3GL2,SLC1A2,SQSTM1,STMN1,STUB1,SUCLG1,SYN1,TLN2,TOMM70,TOP2B,TUBA1A,TUBA1B,TUBB,TUBB6,UCHL1,VAMP2,YWHAE |

| | | | | | | | | | |
|---|---|---|---|---|---|---|---|---|---|
| STAT5B | | transcription regulator | | -1,793 | -1,727 | 0,009 | 6,59E-06 | 1,39E-04 | A2M,AKR1C1/AKR1C2,ALDH3A2,BAIAP2,BAX,C8B,CA2,CASP3,CASP4,CASP6,CASP8,CD70,CISH,COX6A2,CSHL1,CYP2A6 (includes others),CYP2B6,CYP2C8,CYP3A5,CYP4A11,DDIT3,ESR1,FAS,FOS,FOXP3,G0S2,GADD45A,GADD45B,GCK,GPD1,GPX7,GSTP1,HEXA,HSD3B2,IGFALS,KRT19,LEP,LMAN1L,LTA,LTB,MB,ME1,MOXD1,MYL7,NNMT,NPPA,PDK4,PIM1,PIP,PLTP,PPP1R3C,PROM1,RO60,RPA1,SLC16A5,SLC34A2,SLN,SOCS2,SOX15,STAT5A,TLR2,TNNC1,TNNI1,TRIM24,TULP2,WARS1,YBX2,ZFP36 |
| EIF2AK2 | 0,159 | kinase | Activated | 2,348 | 2,727 | 0,072 | 6,65E-06 | 1,40E-04 | ATF3,BAX,BHLHE40,BIRC2,DDIT3,DDX58,EGR1,FAS,FOS,IFI35,IFIT5,IFITM1,IFITM2,ISG15,LGALS3BP,NFKBIA,NMI,NNMT,NOS2,OAS1,PARP12,PLSCR1,PPP1R15A,PSMB10,SOD2,STAT1,TP53,TUBA1A,UBE2L6,ZC3HAV1,ZNF292 |
| miR-29b-3p (and other miRNAs w/seed AGCACCA) | | mature microRNA | Inhibited | -3,031 | -3,412 | -0,080 | 6,72E-06 | 1,41E-04 | CAV2,CNOT8,COL15A1,COL1A1,COL1A2,COL3A1,COL4A1,COL4A2,COL5A2,COL5A3,DCP2,DUSP2,FBN1,FRAT2,GMFB,HMGN3,LAMC1,MYBL2,PPIC,PPM1D,PTEN,SRSF10,TP53,VEGFA |
| Ap1 | | complex | | -1,472 | -1,139 | 0,073 | 7,20E-06 | 1,51E-04 | ATF3,CALM1 (includes others),CCN2,CCNA2,CD44,CDX2,CLU,DDIT3,DLX5,DNMT1,F2RL1,F3,FN1,FOS,FOSL1,GSTP1,HBB,HMOX1,HPGD,HSP90B1,IFRD1,IL13,KCNN4,KLK3,LPL,MET,MMP1,MMP7,MYOD1,NDRG1,NGF,NOS2,NOTCH4,PADI3,PCK1,PRL,RNF7,S100A4,SNCG,TIMP1,UFD1,VCAM1,VDR,VEGFA |
| ERG | | transcription regulator | | 1,028 | 1,402 | 0,064 | 7,27E-06 | 1,52E-04 | ABCC4,ACTN1,ADGRG1,ALOX12,APOC1,ARHGAP28,CCNA2,CD59,CHRNB2,CHRNB4,CHTOP,COL11A2,CYFIP1,DBN1,DSTN,EXT1,FHL1,FOSB,GDF2,GMFB,GUCY1A1,GUCY1B1,HDAC6,HPGD,ITGA6,KLK3,MET,MMP1,NDRG1,NFIL3,NLGN1,NRCAM,PARVA,PECAM1,PIM1,PLA1A,PSTPIP2,RHOA,RHOBTB1,RPA1,RRAS,RSU1,SLIT2,SNAI1,SOX4,SPP1,TACC1,TMPRSS2,TRIOBP,TSC22D3,VIM,WNT11,WNT2 |
| mir-30 | | other | | -0,897 | -1,185 | -0,072 | 7,76E-06 | 1,61E-04 | BAX,BECN1,CCN2,GADD45A,ITGB3,KLF9,MYBL2,NEDD4L,NPPA,RAB38,SMAD1,SMAD2,SNAI1,STK39,TP53,UBE2I,UGT8,VIM |
| RXRB | | ligand-dependent nuclear receptor | | 1,561 | 1,522 | -0,013 | 8,23E-06 | 1,70E-04 | ABCA1,ALDH1A1,ATP1A3,C1QA,CCL17,CCL20,CCL22,CRYAB,CSHL1,CTSS,CYP24A1,DRD2,FABP1,FLRT3,FSTL1,GAS6,HBEGF,IL13,LCN2,LYZ,S100A4,SEMA4D,SPP1,TERT,VCAN |
| TERT | -0,231 | enzyme | | -0,398 | -0,346 | 0,016 | 8,23E-06 | 1,70E-04 | ALDH1A3,ASNS,ATF3,B2M,BAX,CALU,CCN2,CD24,COL5A2,ELK3,FUT4,GATA6,GLB1,HLA-B,HOXB6,HTRA1,IFI16,IFIT2,IGFBP4,IL13RA1,ISG15,ITGAV,KDM5C,KLK3,KRT19,MMP1,MX1,NFKBIA,PCDHGA1,PLOD2,PLTP,PROM1,RB1,RRM2,SOD2,SPP1,TERT,TP53,VIM |
| CST5 | | other | Inhibited | -3,417 | -3,731 | -0,039 | 8,30E-06 | 1,71E-04 | ABCC1,ABLIM1,ACYP2,ADSL,AGO1,AKAP12,AKR1C1/AKR1C2,ANXA2,ANXA3,ANXA5,ARHGAP12,ARHGAP29,ATR,BCAP31,BLOC1S1,CAMK2G,CD44,CNOT8,DHX9,DR1,EBNA1BP2,EIF2B1,EIF5B,ELK3,EMP3,ESRRA,FHL1,FRAS1,GAR1,GNL3,KDM5B,LAMC1,LIMD2,MALT1,MAPK13,MSN,MVP,NCBP2,NOC3L,NOP10,NR2F1,NRP1,NUP107,NUP155,PCLAF,PLS3,PPIC,PPL,PRKACB,PUF60,RBM28,RBP1,REEP5,RTN3,S100A11,SGPL1,SMAD2,SNF8,SOX4,STEAP1,SUPV3L1,TAF1,TESC,VCAN,VIM,WDR3 |
| ALDH1A2 | 0,060 | enzyme | | -0,181 | -0,276 | -0,025 | 8,36E-06 | 1,71E-04 | ABCG2,ALDH1A1,ALDH1A3,ATF3,CCN2,CD44,COL1A2,COL3A1,HOXC8,IGFBP4,NKX2-5,NPPA,SPP1,TGFBI,TIMP1 |
| ADRB | | group | Inhibited | -2,156 | -2,320 | -0,026 | 8,39E-06 | 1,71E-04 | AACS,ABL1,ANXA5,ATF3,BAX,BHLHE40,BNIP3L,CASP3,CASP4,CASP6,CAVIN2,CCNA2,CDC7,CDK4,CDKN1B,CRYAB,DDIT3,E2F4,ENDOD1,FAS,FOS,GADD45A,GSTA3,HMOX1,IFIT3,JUNB,NBN,NFKBIA,NOD1,NOS2,NOTCH2,NPPA,NR4A1,NR4A3,PCNA,PPM1D,RAN,SQSTM1,WEE1,WNT7B |
| TCF7L2 | | transcription regulator | Activated | 1,535 | 2,114 | 0,070 | 8,54E-06 | 1,74E-04 | AATK,ACAA1,ADGRG1,ADO,ALDH3A2,ALOX5,ANKRD49,ANXA1,APLP1,APOD,ARAP2,ASPA,CAMK2N1,CAP1,CARHSP1,CCN2,CCN5,CD24,CDKN1B,CLDN11,CPD,CTNNA1,CYP2C8,CYP3A7,DHCR24,ELOVL1,EPCAM,ERF,EVI2A,EVI2B,FBP1,FKBP1A,G6PC,GAST,GJC2,GLUL,HAPLN2,INS,IPO13,KLHL2,LAMP1,MAP4K4,MCAM,ME1,MMP1,MMP7,MTMR2,MTMR6,MYO6,NFASC,NKAIN1,PAPSS1,PCK1,PICALM,PLEKHA1,PPP1R16B,PROP1,PRRG1,RCBTB1,RHOA,ROCK1,SEMA4D,SIRT2,SLC12A2,SLC1A2,SNAP23,SORBS3,SOX10,SPP1,STK39,TMEM123,TNS1,UGT8,UTP14C,VEGFA,YAP1 |
| JUND | -0,375 | transcription regulator | | -0,246 | -0,109 | 0,039 | 9,01E-06 | 1,83E-04 | ALAS1,APOC3,CCR4,CLU,CSTA,CTLA4,CYBB,F3,FOSL1,HMOX1,HPGD,IVL,JUND,MMP1,NCF2,NOS2,NR4A1,PRL,SOX4,SPP1,SST,TBX21,TIMP1,VEGFA |

| | | | | | | | | | |
|---|---|---|---|---|---|---|---|---|---|
| GNAQ | | enzyme | | -1,522 | -1,344 | 0,064 | 9,16E-06 | 1,86E-04 | ADAMTS1,ADM,ANXA3,ATF3,BNIP3L,BTG2,CCN2,CD44,CITED2,CTSV,DPYS,EGR1,F2RL1,FOS,GADD45A,GAS6,GBP2,HSPB8,LHB,NEU1,NFIL3,NOX5,NPPA,NR4A1,NR4A3,OAS1,PDE10A,PLSCR1,PRKCD,RHOB,SELENBP1,SGK1,SH3BP5,STAT1,VEGFA,YAP1 |
| SPARC | 0,382 | other | Inhibited | -2,539 | -2,609 | -0,012 | 9,72E-06 | 1,96E-04 | ABCC4,BHLHE40,CDH3,CFD,CIDEC,CLDN1,COL1A1,COL1A2,CTSF,FN1,GPD1,HAT1,HSF4,MBD4,MPZL1,MSH2,NDRG1,NES,NIPSNAP2,NOTCH2,PER2,PIK3C2G,POLE2,PPIH,RFC4,RIC8A,RRM1,SLC34A2,SNAI1,TRIM5,TSKU,USP2,VIM |
| TAZ | | enzyme | | 1,009 | 0,980 | -0,008 | 1,05E-05 | 2,11E-04 | ADAMTS1,ALDOC,AMPH,ANXA1,ARHGAP29,CAMK2N1,CAV2,CCN2,CD44,COL1A1,COL5A2,DUSP1,ESM1,F2RL1,FN1,HIF1A,HMOX1,IFI16,LDLR,LUM,MET,MYOF,NOS2,PCLO,PKM,PLK2,SORBS2,TFPI2,VEGFA,VLDLR,WNT11 |
| RORA | -0,330 | ligand-dependent nuclear receptor | Inhibited | -2,899 | -2,730 | 0,046 | 1,07E-05 | 2,13E-04 | ABCA1,ACACB,ADIPOR1,APOC3,AQP8,ASNS,BHMT,CAV3,CCNB1,CISH,CLCA1,CYP2A6 (includes others),CYP2B6,CYP2C8,CYP3A5,CYP4A11,CYP7B1,DHCR24,ELOVL6,G6PC,GADD45B,GSTP1,HPGD,IL18,KCNK5,LCN2,MC1R,MGST3,MUC5B,MYOG,NFKBIA,NLGN1,NNMT,NOCT,PCK1,PPP1R3C,RBFOX1,RDH16,RHBG,RORA,RORC,SELENBP1,SLC13A2,SLC16A7,SLC1A2,SLC25A24,SLC2A4,TLR3,UCP2,VLDLR |
| EBI3 | -0,049 | cytokine | Activated | 1,950 | 2,142 | 0,043 | 1,10E-05 | 2,18E-04 | B2M,CDK4,FOS,GADD45G,GBP2,HLA-A,HLA-B,HLA-C,HLA-DMA,HLA-DMB,HLA-DPA1,HLA-DRA,IL13,MX1,PDCD1,PIM1,RORC,STAT1,TAP1,TBX21,TNFSF10 |
| GDF2 | -0,224 | growth factor | | -0,026 | 0,350 | 0,072 | 1,11E-05 | 2,19E-04 | ABCG2,ANKH,ANXA3,BID,CDH6,COL1A1,CRABP1,CRYAB,DLX5,ELOVL1,FOS,FOXC2,GADD45B,HIF1A,HSPB8,HTRA1,NFKBIA,NGF,NRCAM,NRP1,NTF3,POSTN,SEMA3B,SERPINE2,SMAD1,SOCS2,SPP1,VCAM1,YWHAH |
| RAC1 | 0,223 | enzyme | | 0,113 | 0,366 | 0,054 | 1,20E-05 | 2,35E-04 | APBB1,ATF3,BID,CCN2,COL1A2,DPYSL2,DUSP1,ESR1,FAS,FN1,FOS,FOSL1,HIF1A,HMOX1,HSD11B2,IL13,IL6R,ITGA6,KIR3DL2,MAPK8,MMP1,MYOG,NPPA,PRL,RAC1,RHOA,SHC1,SLC5A5,SOD2,STAT1,STAT3,VCAM1,VEGFA |
| TNFSF11 | | cytokine | | -1,390 | -0,918 | 0,063 | 1,25E-05 | 2,43E-04 | AHR,AKAP13,AURKA,BST1,CA2,CAPN2,CCR4,CD1A,CD44,CD83,CDK14,CDKN1B,CSF3,DCSTAMP,DUSP1,ECT2,ESR1,F3,FAS,FHL2,FOS,FOSL1,FOXO1,GADD45B,GCH1,GLRX,GRB10,GRN,GTF2H1,HCLS1,HMOX1,IFRD1,IL13,IL13RA1,IL15,IRF8,ITGAV,ITGB3,ITPKB,JUNB,LSR,MARCKSL1,MMP1,NAB2,NFIL3,NFKB1,NFKBIA,NOS2,PIM1,RCL1,RGS16,RHOB,RRAS,SENP2,SLC7A7,SOD2,SPP1,STAT1,STMN1,TLR2,TNFRSF11B,TSPAN13,VCAM1 |
| STAT6 | | transcription regulator | | -0,677 | -0,570 | 0,013 | 1,26E-05 | 2,45E-04 | ABLIM1,ADAM11,ALDH2,ANKRD55,ANXA1,APOLD1,ASS1,BAX,CASP6,CCL17,CCL20,CCL22,CDKN1B,CEL,CIDEC,CISH,CLPS,CNR2,COL1A1,COL1A2,CTSS,CYBB,CYP4A11,DCSTAMP,EFL1,EGR1,EPHX1,EXT1,FABP1,FCER1G,FOXP3,GADD45B,GADD45G,GATM,GBP2,HLA-DMA,HLA-DMB,IER2,IFI16,IFI44,IFIH1,IFIT3,IFNAR2,IL10RA,IL13,IL15,IRF7,IRF9,ISG15,LGALS3BP,LTA,LTB,LY75,MYO6,NFIL3,NFKBIA,PDK4,PLBD1,PLSCR1,PNPLA2,POLE2,PPARA,PRKCA,PROS1,RAD50,S100A10,SAMSN1,SELENBP1,SOCS2,SOX13,SPDEF,SQSTM1,STAT5A,STK39,SYNGR2,TBX21,TES,TIMP1,TLE1,TMEM131,USP2,VCAM1,VIM,ZBP1,ZFP36 |
| NFYA | | transcription regulator | | -1,106 | -0,887 | 0,060 | 1,27E-05 | 2,46E-04 | ALB,CAT,CCNA2,CCNB1,CCNB2,CDC25B,CDK1,COL1A2,COL5A3,EGR1,EPHX1,F12,G6PC,GADD45A,GADD45B,GUCY1B1,HLA-DQB1,HLA-DRA,HSPA5,JUNB,JUND,PCK1,PKM,PPP1R15A,PTTG1,RHOB,RRM2,UBE2I |
| BMI1 | | transcription regulator | | 0,647 | 0,579 | -0,025 | 1,31E-05 | 2,54E-04 | ATOH1,AURKA,BTG2,CASP8,CD63,CDKN1B,CUL1,CYFIP2,DNMT1,FAS,IL36A,MCM3,MET,NFKB1,P4HA1,RRM1,SFN,TERT,TP53,TP53I3,TUSC3,VIM,WNT10B |
| RAB1B | -0,052 | other | | 1,098 | 1,273 | 0,046 | 1,33E-05 | 2,57E-04 | ARF4,BET1,COPZ2,DERL1,GOLGA3,GORASP2,KDELR3,RAB32,RRAGC,SEC24D,SLC5A5,SRP54,STX3,USO1,YIF1A |
| NFATC2 | | transcription regulator | | 0,771 | 0,994 | 0,034 | 1,40E-05 | 2,70E-04 | ABCA1,AQP5,CASP3,CASP4,CCL17,CCL22,CCNA2,CCNB1,CD83,CDK4,CISH,CITED2,CRYAB,CTLA4,CX3CR1,EGR3,FAS,FGL2,FOXP3,HDAC1,IFIT2,IFIT3,IL13,IL15,IL18,IRF7,ISG15,MITF,NAA80,NFKB1,NFKBIA,NMI,NR4A1,PDCD1,PDCD5,PDZD2,PLK2,PMEL,PPARG,PTPRK,RGS2,RORC,SH3BP5,SOCS2,STAT1,STAT5A,TBX21,TERT,TLR2,TLR3,ZBTB32 |
| ATF2 | | transcription regulator | | -1,954 | -1,600 | 0,077 | 1,40E-05 | 2,70E-04 | ANGPTL2,APOC3,ASNS,ATF3,BCKDHA,CCNA2,CGA,CYP11B2,DDIT3,DMBT1,DUSP1,EGR1,FN1,FOS,GADD45A,IL13,ITGB8,MMP1,NOS2,NPPA,PCK1,PPARG,PSEN1,PTEN,RHOB,SOD2,SST,VCAM1,ZNF268 |

| | | | | | | | | | |
|---|---|---|---|---|---|---|---|---|---|
| SIM1 | 0,094 | transcription regulator | | -2,131 | -1,580 | 0,076 | 1,51E-05 | 2,89E-04 | ABCG1,ABI1,AKR1B1,AVP,BST1,CBFB,CDH3,CLCN1,COPZ2,CRISP2,CYBA,EGF,EGR1,ELL2,FKBP3,FOXH1,GLI1,GLUL,GSTA3,HGFAC,HLA-A,IFI16,IGFBP4,IL6R,ITGB3,KCNE5,KLK3,LEP,LRRFIP1,LTF,MYCL,NAP1L4,PADI3,PEA15,PFN1,PMFBP1,POSTN,RGS10,SELENBP1,SLC12A3,SNN,SNX5,SOX10,SST,TACR2,TCF7L1,THRB,TSC22D3,TSSK2,TYK2,TYROBP,USP2,WNT7B |
| ELK1 | -0,204 | transcription regulator | Inhibited | -2,269 | -2,022 | 0,062 | 1,52E-05 | 2,90E-04 | ANKH,AVP,CCT8,CDKN1B,EGR1,ELK1,FOS,FOSB,FOSL1,FUT4,ITGAV,ITGB6,JUNB,MTHFD2,PRKCA,PRL,SNAI1,SP P1,TIPARP,TPD52L1,ZFP36 |
| Ige | | complex | Activated | 1,746 | 2,239 | 0,060 | 1,59E-05 | 3,03E-04 | ACTN1,ALAS1,ALOX5,ANXA1,BAIAP2,C3,CAMK2G,CAPN2,CARHSP1,CCL1,CCL17,CCL22,CD53,CD74,CHST8,CISH,COL18A1,CSF3,CXADR,DUSP2,EGR1,EIF4G2,FAS,FCER1G,FHL2,GADD45B,HBEGF,HIP1R,HLA-DMA,HMOX1,IL13,IL15,ITGAV,JUNB,KRAS,LTA,MALT1,MARCKSL1,MMD,NFKB1,NFKBIA,NFKBIE,NPHS2,NRP1,NXPE4,PEA15,PECAM1,PFN1,PIK3IP1,PLK3,PPARG,PPIF,PRKCA,PTPRF,RAI14,RGS16,RNASE3,SEMA7A,SERPINB1,SNRPD1,SPP1,STAT5A,TCIRG1,TLR2,TNC,TNFRSF11B,TNFRSF13B,TNFSF10,TPPP3,TUBB6,UGCG,VCAM1,VDR,VIM,WNT10B |
| estrogen receptor | | group | | -1,462 | -1,473 | -0,002 | 1,60E-05 | 3,04E-04 | ABCA1,ABCG1,ABCG2,ADIPOQ,AGO1,ANXA1,BAX,BMP7,C3,CAPG,CAV2,CCN2,CD24,CD44,CD5,CD59,CD99,CDH15,CDH3,CLDN3,COL4A1,COL4A2,CYP2B6,DNMT1,EGR1,F11R,F12,FGF1,FGF5,FN1,FOS,GP9,HBEGF,HRAS,KRT19,LDLR,LTF,LYN,MAPT,MMP1,MMP17,MSN,MUC1,PCDH1,PDYN,RGS2,SMAD1,SNAI1,SYNPO,TERT,TGFBR3,TIMP1,TNC,TP53,TSC22D3,UGT8,VEGFA,VIM |
| RETNLB | | other | | 1,236 | 1,610 | 0,074 | 1,64E-05 | 3,10E-04 | CASP3,CCN2,COL1A1,COL1A2,ELN,FABP1,FN1,FZD1,FZD6,IL13,IL13RA1,JAK1,KLF15,MID1,MSN,NR4A1,PLA1A,PTEN,SMAD1,SOCS2,TERT,THBS4,TIMP1,TLR2,TNNC1,UBE2C |
| SGPL1 | 0,216 | enzyme | | 0,649 | 0,637 | -0,003 | 1,73E-05 | 3,24E-04 | ABCA1,ABCC1,APP,BAX,DDIT3,DDX58,HSPA5,ISG15,MX1,OAS1,PPARG,SEC61A1 |
| CETP | -0,100 | enzyme | | 1,282 | 1,257 | -0,007 | 1,73E-05 | 3,24E-04 | APOA1,APOB,DDIT3,HSPA5,LDLR,LPL,NR0B2,P4HB,PDIA3,PDIA4,PLIN3,PPARG |
| EFNA1 | -0,242 | other | | -0,436 | -0,512 | -0,017 | 1,75E-05 | 3,27E-04 | APOBEC3G,CCN2,CRABP2,FN1,FOSL1,GATA6,HERC6,HRAS,IRF7,ITGB6,IVL,KLK7,KRAS,KRT10,KRT4,KRT75,NEDD4L,NRAS,PKP1,TGM1,TRIM5,UBE2C,WWP1 |
| IL12 (complex) | | complex | Activated | 1,928 | 2,252 | 0,052 | 1,81E-05 | 3,38E-04 | AHR,CASP1,CASP3,CCL1,CCL17,CCL20,CCL22,CCR4,CD44,CD63,CISH,CXCL6,DDB2,FAAH,FAS,FOS,FOXP3,FURIN,FUT7,GADD45B,GADD45G,GZMA,IFIH1,IFIT2,IFNGR1,IFNGR2,IL13,IL15,IL18,IL32,IRF8,ISG15,ITGA6,ITGAE,ITGB2,LTA,LTB,NOS2,PDCD1,PDCD4,PIM1,RAD23B,RPA1,RPA2,RPA3,SET,SMAD2,SOCS2,TBX21,TIMP1,TLR2,VEGFA |
| TGFB2 | | growth factor | | 0,449 | 0,615 | 0,031 | 1,82E-05 | 3,40E-04 | ABCA1,AHR,BHLHE40,CCN2,CD59,COL1A1,COL1A2,COL3A1,COL4A1,CYP24A1,EGR1,FAS,FN1,FOS,FURIN,HBEGF,HMOX1,MLLT3,NGF,NOS2,PDK4,PECAM1,PLOD2,PTEN,SMAD2,SPP1,TGFBR3,TIMP1,TNFRSF11B,VCAN,VDR,VEGFA,WNT11 |
| KLF2 | -0,097 | transcription regulator | | 0,635 | 0,561 | -0,014 | 1,83E-05 | 3,41E-04 | ACKR3,ADAM15,ADM,APLNR,AQP5,ASS1,CCN2,CCR4,CD1A,CD44,COL4A1,CX3CR1,ELN,F2RL1,F3,FGF5,FOXP3,FUT4,GADD45A,HBB,HIF1A,IL6R,ITGB2,ITGB3,JUNB,KCNN4,KITLG,LEPR,MVP,MYH10,NDRG1,NFKBIA,NOTCH2,PECAM1,PLOD2,PPARG,PROS1,PTGDS,STAT1,STAT3,TBX21,TERT,TFPI,TFPI2,TIMP1,VCAM1,WEE1 |
| NTRK2 | 0,062 | kinase | | 1,652 | 1,931 | 0,067 | 1,85E-05 | 3,43E-04 | ABCG2,AMBN,APP,BID,CASP1,CCNA2,CDK1,CDK4,EGR1,FOS,GAD1,MCM5,NOS2,PCNA,PTEN,PVALB,PYCARD,RAB3A,RB1,SPP1,SYN1,TNFSF10,TP53BP2,VEGFA,VIM |
| MEF2D | | transcription regulator | | -1,717 | -1,547 | 0,040 | 1,91E-05 | 3,53E-04 | CCN2,COL1A2,COL3A1,FN1,FOS,FOSB,JUNB,JUND,MCM3,MCM5,MYOD1,MYOG,NPPA,NR4A1,PCNA,PPARA,PTEN,SLC2A4,TNNC1,TNNI1,VIM,ZFP36 |
| PRKCA | -0,182 | kinase | | -1,182 | -0,973 | 0,040 | 1,95E-05 | 3,60E-04 | APOBEC3G,APOM,APP,ARHGDIB,ARL4C,ARPC2,AVP,BNIP3L,CCN2,CHRND,COL1A1,CSF3,CSTA,CYBB,EGR1,FN1,FOS,IFIT3,IVL,LPL,MMP1,NFKB1,NFKBIA,NOS2,NOTCH4,NPPA,PLA2G6,PRKCD,PTEN,RB1,SDCBP,TP53,USP8,VCAM1,VEGFA |
| ADRB1 | -0,117 | G-protein coupled receptor | | 0,296 | 0,314 | 0,007 | 2,03E-05 | 3,73E-04 | ANXA4,APP,BECN1,COL3A1,DDIT3,GNAS,HSPA5,IRAK1,LEP,NGF,SMC4 |
| F2R | 0,109 | G-protein coupled receptor | | 0,093 | 0,367 | 0,052 | 2,03E-05 | 3,73E-04 | BIRC2,CASP3,CASP4,CCN2,CD24,CD44,CLDN3,DUSP1,EGR1,EGR3,ESR1,F11,F3,FBP1,FN1,FOS,HMOX1,ITGA6,KRT19,LAMB1,MCAM,MYOG,NR4A1,RHOA,S100A4,TFPI,TFPI2,TP53,VCAM1,VEGFA,VIM |

| | | | | | | | | | |
|---|---|---|---|---|---|---|---|---|---|
| PRKAA1 | 0,079 | kinase | | -0,278 | -0,464 | -0,031 | 2,05E-05 | 3,75E-04 | ACKR3,ALB,ARHGDIB,ATF3,BAX,CA2,CAT,CD24,CDH3,CDKN1B,CLU,CYP2B6,CYP3A5,DDIT3,DLGAP2,EAF2,EGF,FN1,G6PC,GHR,GRIA3,GSTA1,HSD3B2,HSPA5,IGFBP4,NCF2,NFKBIA,OAT,PDYN,PKLR,PRKACB,PSMB8,RGS2,SERPINI1,SIRT2,SIRT3,SOD2,TP53,VCAM1,YAP1 |
| MAP2K1/2 | | group | | -1,217 | -1,004 | 0,046 | 2,11E-05 | 3,83E-04 | ARC,ATF3,C3,CCN2,CDK1,CDKN1B,CHI3L1,DDIT3,DUSP1,EGR1,ELN,FOS,FOSL1,FOXP3,FURIN,HMOX1,HSPA5,IL13,JAK1,NFKB1,NOS2,PCNA,PPARA,RRM1,VEGFA,VIM |
| ID3 | 0,266 | transcription regulator | | -0,045 | -0,075 | -0,005 | 2,25E-05 | 4,06E-04 | ADGRG1,ADIPOQ,ADM,BMP7,BTG2,CCNB1,CD44,CD83,CDC25B,CDK1,CDKN1B,DTX2,DUSP1,DUSP22,ELOVL6,FCER1G,FOXO1,FOXP3,GADD45B,GADD45G,GNL3,HIF1A,IFI16,IFNGR1,IFNGR2,IGFBP4,IL10RA,IL17RB,IL6R,IRF8,ITGB3,LEPR,LGALS1,LTA,LTB,NEUROD6,NPPA,NR4A1,NR4A3,PDCD1,PIK3IP1,PTPN3,RORA,SEMA7A,SOX13,SOX4,TGFBR3,TNFSF10,TP53,VCAM1 |
| LEPR | 0,236 | transmembrane receptor | | 0,024 | 0,117 | 0,017 | 2,25E-05 | 4,06E-04 | ABCG1,ACACA,ADIPOQ,ANXA5,APOA1,APOB,CASP8,CD5,CD63,CDK8,CNR2,COL1A1,COL4A1,EIF3A,FN1,FOS,GADD45B,GADD45G,GCK,GTF2A2,HIF1A,HPX,LCN2,LDLR,LEP,LEPR,LIPC,LPL,MAP4K4,MMP7,NBN,NOS2,PPARA,PPARG,PSMB8,RNASET2,SLC2A4,SMARCD3,SOCS2,STAT3,STUB1,TIMP1,TRPV1,TSC22D3,UCP2 |
| mir-155 | | microRNA | Inhibited | -3,727 | -3,959 | -0,042 | 2,31E-05 | 4,14E-04 | ADAM10,ANXA2,CD47,CDC73,CLDN1,CSF3,DHX40,FADD,HIF1A,HMOX1,IL13,IL13RA1,IL7,IRF7,IRF8,IRF9,MME,MSH2,MSH6,MX1,NOS2,NUP153,PDCD4,PPARG,PSIP1,PTEN,RORA,SDCBP,SMAD2,STAT1,STAT3,TP53,TRIM32,WEE1 |
| CLDN7 | 0,258 | other | | -1,205 | -1,197 | 0,001 | 2,36E-05 | 4,20E-04 | ABLIM1,ALDH3A1,ANXA3,ARHGDIB,C1S,C3,CCL20,CCN2,CKAP4,CYP11B2,DENND5A,DNAJB6,F3,FADS3,FOSL1,GRINA,HLA-B,IFI44,MAGED1,MLEC,MRPS17,MT1X,MUC1,MX1,NETO2,NNMT,PLGRKT,PSG11,PSMG2,RNASET2,RPP40,SERPINB1,SPRR3,TPK1,UBE2L6 |
| Pdgf (complex) | | complex | | -0,310 | -0,072 | 0,045 | 2,38E-05 | 4,22E-04 | BAX,CD44,CDKN1B,COL4A1,CYBA,DIO3,EGR1,F3,FN1,FOS,FOSB,FOXO1,HIF1A,HMOX1,IL13,ITGB3,JUNB,LDLR,MMP1,NTF3,ODC1,PCNA,PIK3R3,PPARG,SV2A,TERT,TIMP1,TP53,VCAM1,VEGFA |
| prostaglandin E2 | | chemical - endogenous mammalian | | -2,182 | -1,971 | 0,029 | 2,56E-05 | 4,53E-04 | ABCG1,ADAMTS1,AHR,ANXA2,APP,ARC,ATF3,BAX,CASP1,CASP3,CASP4,CASP6,CASP7,CCL20,CCL22,CCN2,CCNB1,CCR4,CD44,CD83,CNNM2,COL1A1,CSF3,CTLA4,DUSP1,DYNLL1,EGR1,ESRRA,F2RL1,FAS,FN1,FOS,FOSB,FOSL1,FOXO1,FOXP3,HIF1A,HMOX1,HSD3B2,IGFBP4,LEFTY2,MITF,MMP1,MXI1,MYOZ1,NFIL3,NFKBIA,NOS2,NR4A1,NRP1,PKLR,PLK2,PPARG,PRL,PTK2,RAB5A,RGS2,RIN1,RORC,SOCS2,SPP1,ST3GAL4,TBX21,TGFBR3,TNFRSF11B,TP53,TSC22D3,UCP2,VEGFA,VIM,XRCC5 |
| INSR | -0,328 | kinase | Inhibited | -3,070 | -2,786 | 0,063 | 2,62E-05 | 4,63E-04 | ACOT13,ALAD,ALB,ALDH2,ALDH6A1,ATF3,BAG1,C1QA,C2,CASP6,CCN2,CD83,CDK1,COL6A3,COX6A2,CRAT,CSTB,CTSV,DCTN4,DEK,DIO3,DUSP1,EGR1,FBP1,FHL1,FOS,FOXC2,FURIN,GADD45A,GATA6,GCH1,GCK,GNB1,GRB10,GRN,HBEGF,HOXC8,HSD17B4,IDI1,IER2,IFITM2,IFNAR2,IL25,INS,INSR,IQGAP1,JUNB,KLF9,LEP,LEPR,LGALS1,LRP10,LYZ,MARCKSL1,MCM2,MCM5,MSH6,MXI1,MYBL2,MYOM2,NAMPT,NDRG1,NFE2L2,NGF,NPPA,PDCD4,PDHA1,PDK4,PIM1,PKLR,PMS1,PPARA,PRKD1,RGS16,RHOB,RRM1,RRM2,SIRT2,SKAP2,SLC2A4,SOCS2,THBS2,TNNI1,UCP2,VCAM1,ZFAND5 |
| NR1I3 | -0,229 | ligand-dependent nuclear receptor | | -0,573 | -0,242 | 0,067 | 2,63E-05 | 4,64E-04 | ABCC3,ABCC4,AHR,ALAS1,ALDH1A1,APOA1,CSHL1,CYP2A6 (includes others),CYP2B6,CYP2C8,CYP3A4,CYP3A5,CYP3A7,CYP7B1,DIO1,EI24,EPHX1,FBRS,G6PC,GADD45B,GSTM5,GSTP1,HK1,HNF4A,HSD17B4,MAFF,PCK1,PGD,RARRES1,RPA2,SULT2B1,TMPRSS2 |
| EGR2 | -0,609 | transcription regulator | | -1,638 | -1,245 | 0,063 | 2,80E-05 | 4,93E-04 | ABCC1,ASS1,BHLHE40,BNIP3L,CAST,CCL1,CDH6,CNTFR,CRABP1,DDN,DLG3,DMBT1,EGR1,EIF5B,FOS,GHR,GHRH,HSPA5,IL6R,ITGAE,KITLG,LHB,MME,MPZ,NAB2,NSF,NUP54,PIP,PPT2,PRKCD,PRX,RORA,RORC,S100A1,SLC12A2,SMAD1,STAT5A,TBX21,TGFBR3,TNFSF10,TOP2B,VEGFA |
| LIPE | | enzyme | | 0,113 | 0,176 | 0,019 | 2,85E-05 | 5,00E-04 | ABCA1,ADIPOQ,ALDH1A1,ANKRD2,ATF3,CA2,CASP8,CD74,CITED2,COL1A1,COL1A2,EGR1,FOS,G0S2,GRB10,ITGA6,KITLG,MAPT,NET1,NR4A1,PAM,PCK1,PDHA1,PDK4,PER1,PFKFB1,POSTN,PPARG,PVALB,RB1,RXRA,SCARB2,SLC2A4,TKT,TWF2 |

| | | | | | | | | | |
|---|---|---|---|---|---|---|---|---|---|
| ARNT2 | 0,271 | transcription regulator | | -2,143 | -1,575 | 0,080 | 2,87E-05 | 5,02E-04 | ABCG1,ABI1,AKR1B1,BST1,CBFB,CDH3,CLCN1,COPZ2,CRISP2,CYBA,EGF,EGR1,ELL2,FKBP3,FOXH1,GLI1,GLUL,GSTA3,HGFAC,HLA-A,IFI16,IGFBP4,IL6R,ITGB3,KCNE5,KLK3,LRRFIP1,LTF,MYCL,NAP1L4,PADI3,PEA15,PFN1,PMFBP1,POSTN,RGS10,SELENBP1,SLC12A3,SNN,SNX5,SOX10,TACR2,TCF7L1,THRB,TSC22D3,TSSK2,TYK2,TYROBP,USP2,VEGFA,WNT7B |
| ONECUT1 | | transcription regulator | | 0,379 | 0,417 | 0,016 | 2,95E-05 | 5,15E-04 | AASS,ACACB,AFP,AKAP13,ALDH3A1,AMT,APOH,ARHGAP32,ARMC6,ARMT1,BCKDHA,BNIP3L,C1S,C2,CCT8,CFI,COL5A3,COPB2,DDB2,EED,ELOVL1,F11,FADS3,FAM117A,FAS,G6PC,GLA,GTF2E1,HLA-G,HMGB2,HNF1B,HNF4A,HNRNPR,HSD17B4,ITGAV,ITIH1,KLF3,LPL,LRRC59,LUC7L3,MFSD1,MGST3,MYOF,NDC1,NFKBIA,NR0B2,OAT,PCK1,PFKFB1,PHTF1,PMEL,PPFIBP1,PSMA1,RARG,RORA,RORC,SH3BGRL,SNW1,SPAST,TAOK3,TBX2,TNFSF10,USP46,UTP3,VWA5A |
| EIF3A | 0,226 | other | | -1,088 | -1,203 | -0,044 | 2,98E-05 | 5,18E-04 | CDKN1B,ISG15,NDRG1,RAD23B,RPA1,RPA2,RPA3 |
| LIF | -0,074 | cytokine | | 0,915 | 1,051 | 0,021 | 3,01E-05 | 5,22E-04 | A2M,ACACA,AFP,BMP7,CCNA2,CISH,CSHL1,CYFIP1,CYP11B2,CYP26A1,EGR1,ERAP1,EVX1,FBP1,FGF5,FN1,FOS,FOXF1,FOXP3,GATA6,GHRH,HBEGF,HIF1A,HLA-G,HOXB1,IFI16,IL7,JUNB,LEP,LPL,MMP1,MYOG,NKX2-5,NMI,NPPA,PECAM1,PIM1,PPARG,PRDM14,PRPH,SERPINA3,SNAP23,SOCS2,SOD2,SPP1,STAT3,TIMP1,TNC,TNFRSF11B,TP53,TSPAN7,VEGFA,WT1 |
| Pro-inflammatory Cytokine | | group | | 0,022 | 0,230 | 0,064 | 3,03E-05 | 5,23E-04 | ADM,AKR1B1,ASS1,CD47,CLU,F3,GLUL,GSTA1,HDAC6,HMOX1,HSPA5,LCN2,LGALS1,NFE2L2,NOS2,NR4A1,S100A1,SLC6A4,SPINT1,TIMP1,TLR2,UBD,VCAM1 |
| GAPDH | | enzyme | Inhibited | -2,580 | -2,706 | -0,033 | 3,16E-05 | 5,44E-04 | C2,CCL20,DUSP1,ENO1,FCER1G,HLA-C,IFIT2,IFITM1,IFITM2,KLK3,OAS1,SPP1,STAT1,UBE2L6,WARS1 |
| IL10 | | cytokine | | 0,315 | 0,379 | 0,008 | 3,26E-05 | 5,58E-04 | ABCA1,ADM,ALOX5AP,ARAP2,BAX,CASP3,CASP8,CCL1,CCL16,CCL17,CCL20,CCL22,CCR4,CD163,CD1A,CD44,CD83,CDKN1B,CLCA1,COL1A1,CSF3,CTLA4,CTSS,CTSZ,CX3CR1,CXCL6,DUSP1,ELAVL1,F3,FAAH,FAS,FCER1G,FOS,FOXP3,GZMA,HLA-G,HMOX1,HPGD,HTATIP2,IFIT2,IGFBP4,IL10RA,IL13,IL17RB,IL18,IL7,IRAK1,ITGAE,KITLG,LGALS1,LTB,LY75,MMP1,MUC1,MYOG,NOS2,NPHS1,NR4A3,PAM,PDCD1,PSMB9,RARRES1,RGS16,SLC6A4,SNAP23,STAT1,STAT3,TAP1,TERT,TIMP1,TLR2,TLR3,TNFRSF13B,TNFSF10,TSC22D3,VCAM1,VCAN,VEGFA,VTCN1,ZFP36 |
| GATA1 | -0,126 | transcription regulator | | -2,093 | -1,875 | 0,031 | 3,29E-05 | 5,59E-04 | ABCC4,ACTN1,ALOX12,ALOX5,ALOX5AP,BTG2,BZW2,CA1,CA2,CBX5,CCR4,CDK4,CDKN1B,CPD,CYBB,CYFIP1,DMTN,EGR1,EPB42,EXOC1,FHL1,FOSB,GNAS,GP9,GUCY1B1,GYPA,GYPB,HBB,HNRNPAB,IL13,IL6R,INHA,IRF8,ITGA6,ITGB3,JUNB,LEPR,LYZ,MAGED1,MCM3,MCM5,MITF,MSH2,NAB2,NFIL3,NFKB1,NPPA,PCNA,PECAM1,PIM1,PLXNA2,PRG2,PSTPIP2,RFC4,RRM1,SLC4A1,SMARCE1,SPOCK2,SPTB,SSBP3,ST3GAL6,STAT5A,TAC3,TFEB,TRIM29,USP16,VDR |
| FN1 | 0,341 | enzyme | | 0,722 | 1,005 | 0,046 | 3,29E-05 | 5,59E-04 | ACTR2,AHCY,APP,BNIP3L,CANX,CAPN2,CASP7,CD83,CDK4,CDKN1B,CHRNB4,COL1A1,CSNK2A1,CTSD,DHCR24,DPF2,F3,FERMT2,FN1,FOS,GSK3A,HK1,HLA-B,IL7,ITGAE,ITGAV,ITGB3,JUND,KIF21B,LAMP1,MAPK8,MARCKS,MMP1,NES,NFKBIA,NOS2,PCNA,PPP6R2,RALB,SCRN1,SDCBP,SEC23B,SMAD2,SMARCA4,SNAI1,SNAP23,SPP1,SQSTM1,TLR2,VIM,VPS45 |
| ZBTB17 | | transcription regulator | Inhibited | -2,511 | -2,708 | -0,046 | 3,42E-05 | 5,76E-04 | ATOH1,BHLHE40,CAMK2N1,CCNA2,CCNB1,CCNB2,CDC20,CDK1,CDKN1C,CKAP2,DCT,DDB2,DDIT3,ECT2,EGR1,KIF20A,LDLR,NDC80,NOCT,ODC1,OSMR,PBK,PRC1,RHOA,UBE2C,ZFP36 |
| JUNB | -0,667 | transcription regulator | | 0,939 | 1,164 | 0,052 | 3,43E-05 | 5,76E-04 | APOC3,APOM,ATF3,C3,CCR4,CDC20,CDKN1B,CGA,CLU,COL1A1,COL1A2,COMT,DARS1,DUSP1,FN1,FOSL1,FOXP3,HMOX1,HPGD,ITGAV,ITGB3,JUNB,LCN2,MMP1,MYL12A,NCF2,PAK3,PNPLA2,ROCK1,SGK1,SNAI1,SST,SWAP70,TIMP1,VAMP8 |
| let-7 | | microRNA | Inhibited | -4,282 | -4,819 | -0,077 | 3,43E-05 | 5,76E-04 | AURKA,BTG2,CASP3,CBFB,CCNA2,CCNB1,CD44,CDC20,CDC23,CDK1,CDK8,COL1A1,COL1A2,COL3A1,E2F4,EIF4G2,ERF,FARP1,FN1,FOSL1,GPI,GPX7,GTF2I,IL6R,ITGB3,KRAS,MAD2L1,MCM2,MCM3,MCM5,NFKB1,NRAS,PBX2,PKM,PLAGL2,PPP1R12B,PROM1,RFC2,RFC4,RFC5,RIOX2,RRM1,RRM2,S100A4,SMAD2,SSR1,STAT3,VIM,YAP1,ZFP36 |

| | | | | | | | | | |
|---|---|---|---|---|---|---|---|---|---|
| ATF3 | -1,334 | transcription regulator | | 1,709 | 1,629 | -0,017 | 3,44E-05 | 5,76E-04 | ADIPOQ,ADIPOR1,ASNS,ATF3,AURKA,BHLHE41,CD83,CDK1,CDK4,CGA,CHAC1,DDIT3,FBP1,G6PC,GBP2,HSPA5,KLK3,LDLR,MMP1,MSH2,NOS2,PCK1,PPP1R15A,SRI,STAT3,TLR2,TP53,TP53I3,TP73 |
| Ca2+ | | chemical - endogenous mammalian | Inhibited | -3,773 | -3,460 | 0,043 | 3,53E-05 | 5,88E-04 | AHR,ANKH,APP,AQP5,ARC,ATF3,CD83,CDK4,CDKN1B,CGA,COL1A1,CUL9,CYP27B1,DDIT3,DUSP1,EGR1,ELK1,FAAH,FN1,FOS,FOSL1,GAST,GUCY1A2,HBEGF,HSPA5,IL6R,ITGA6,IVL,JUNB,JUND,KLK5,KLK7,KRT10,LHB,MCAM,NDRG1,NFKBIA,NOS1,NOX5,NPPA,NR4A1,NR4A3,ODC1,PADI3,PER1,PER2,PLS3,PPARA,PPARG,PRL,PTTG1,PYY,SGK1,SLC12A3,SPP1,SST,SSTR2,TGM1,UGCG,VCAM1,VDR,VEGFA |
| TGFB3 | -0,172 | growth factor | | 0,329 | 0,630 | 0,057 | 3,91E-05 | 6,49E-04 | BHLHE40,CCN2,CD59,CDH6,CDKN1B,CLDN11,COL1A1,COL1A2,COL3A1,ELN,F2RL1,FN1,FOS,FOSB,HTRA1,ITGAV,ITGB3,JUND,LTBP2,MMP1,PLOD2,PTEN,SERPINE2,SMAD2,SNAI1,TGFBR3,TNC,TNFRSF11B,TNS1,TPM1,TSPAN13,VEGFA |
| JAK1 | 0,545 | kinase | | 1,201 | 1,518 | 0,070 | 3,95E-05 | 6,50E-04 | BID,CAPN1,CDKN1B,CHI3L1,CSF3,FOS,HIF1A,HLA-A,HLA-C,HLA-F,IFIT2,IRF7,IRF9,JUNB,MX1,MYOD1,OSMR,PSMB9,SNAI1,STAT1,STAT3,TAP1,TNS4 |
| mir-133 | | microRNA | | -1,677 | -1,988 | -0,066 | 3,95E-05 | 6,50E-04 | ARFIP2,CCN2,CCNB1,CMTM6,CORO1C,CTBP2,DNMT1,ELAVL1,FN1,GSTP1,HLA-G,KLF15,LASP1,LPGAT1,MET,MSN,PITX3,PPARG,RHOA,SLC2A4,TNFSF10,TNNT2,VCAN |
| 5-hydroxytryptamine | | chemical - endogenous mammalian | | -1,164 | -0,881 | 0,057 | 4,01E-05 | 6,59E-04 | AP2S1,ARC,ARHGEF1,ASIC2,ASIC3,CCN2,CYBB,EGR1,F3,FADD,FOS,GAD1,HBEGF,HIF1A,LIPC,MAPK13,MYL6,NCF2,NR4A1,NUCB2,PFN1,PLCB2,PLIN3,PPP1CA,PRL,RAB8A,SLC10A2,SLC6A4,STX4,TRPV1,TUBB,UCP2,VCAM1 |
| PRDM1 | -0,108 | transcription regulator | | -0,754 | -0,880 | -0,018 | 4,05E-05 | 6,64E-04 | ACOX2,ADGRG1,AFP,ALPL,APOM,ATP9A,BMP5,CA9,CCL20,CD44,CD53,CD74,CHI3L1,CLDN1,CSF3,CXCL6,ERAP1,ESR1,FOS,FOSL1,GAD1,GALNT2,GPX5,HLA-DRA,IRF8,KHDRBS3,KRT81,LGALS1,LHX3,LIPF,ME1,MMP25,MS4A4A,MYO1A,NOS2,PDCD1,PLAAT4,PSMB10,PSMB8,REG1A,SERPINA3,SLC16A5,SLC2A5,SMPDL3B,SNAI1,ST14,SWAP70,TAPBP,TBX21,TNFRSF10D,TNFRSF13B,TNFSF10.TREH.TRIM38.XRCC5 |
| corticosterone | | chemical - endogenous mammalian | | 0,422 | 0,433 | 0,002 | 4,08E-05 | 6,66E-04 | ACACA,ANXA1,APOB,APP,ARC,AVP,CCN2,CDK4,CDKN1B,CGA,CSHL1,CYBA,DDIT3,FOS,GCK,GLUL,HSPA5,KLF9,LEP,LHB,MAPK8,NOS1,NOS2,NPPA,PDK4,PNPLA2,PRL,SGK1,SLC12A3,SLC1A2,TLR2,UCP2,WNT10B,WNT7B |
| hydrocortisone | | chemical - endogenous mammalian | | -1,864 | -1,789 | 0,013 | 4,11E-05 | 6,70E-04 | ACO1,ADM,ALDH18A1,ALDH1L1,APP,BCLAF1,CCL17,CCN2,CD163,CDK1,CFD,CSHL1,CYP3A4,FAS,FN1,GADD45B,GBP1,GLUL,GNAS,GPR153,IGFBP4,INS,IVL,KLK3,LPL,LY75,NFKBIA,NOTCH4,PLG,PPARG,PRL,STAT5A,STC2,TLR2,TNFRSF11B,TP53,TUBB,VAMP2 |
| Tgf beta | | group | | -0,134 | 0,128 | 0,036 | 4,17E-05 | 6,79E-04 | ABCA1,ACKR2,ANKH,AXIN1,BECN1,BHLHE40,CASP1,CASP8,CCL1,CCN2,CDKN1B,CLU,COL1A1,COL1A2,COL4A1,COL4A2,CSF3,CTF1,CTLA4,CX3CR1,FN1,FOS,FOXP3,FSTL1,FURIN,GADD45B,HMOX1,HNRNPAB,HTATIP2,IGFBP4,IL13,IL32,ITGA6,ITGAV,ITGB3,JAK1,JUNB,MET,MMP1,MMP7,MUC1,NCR3,NOS2,PDCD1,POSTN,PROM1,PTEN,RHOB,RNF111,RORA,RORC,SGK1,SMAD2,SNAI1,SPP1,SSBP3,TBX21,TERT,TGFBI,TIMP1,TLR2,TNFRSF11B,VCAM1,VEGFA.VIM |
| ZBTB20 | 0,088 | transcription regulator | | 0,320 | 0,333 | 0,003 | 4,37E-05 | 7,08E-04 | AFP,CISH,CYP2B6,CYP2C8,CYP3A5,G6PC,GAS6,GCGR,GCK,GHR,GPC3,GSTM5,IGFALS,LEP,LEPR,NFKBIA,PCK1,PDK4,SOCS2 |
| GPER1 | -0,229 | G-protein coupled receptor | Inhibited | -4,022 | -3,720 | 0,073 | 4,37E-05 | 7,08E-04 | ATF3,BAX,BHLHE40,CCN2,DDIT3,DUSP1,EGR1,ESR1,ESRRA,FOS,FOSB,HSPA5,IER2,LTF,MT1X,NFKBIA,PPP1R15A,RGS16,ZFP36 |
| PARP9 | | enzyme | Activated | 2,916 | 3,138 | 0,071 | 4,38E-05 | 7,08E-04 | CD74,HLA-DQB1,HLA-E,IFI44,IFIT2,IFIT3,IRF7,ISG15,SP110,STAT1 |
| L-glutamic acid | | chemical - endogenous mammalian | | 0,129 | 0,372 | 0,045 | 4,62E-05 | 7,40E-04 | APP,B2M,BAX,CASP3,CDS1,CFD,CIDEC,CYP2B6,CYP3A4,EGR1,FOS,FOSB,GADD45B,HEXA,HIF1A,IL18,JUNB,LEP,MGLL,NFIL3,NOS1,NOS2,PCNA,PER1,RHOA,ROCK1,SLC1A2,TAP1,TIMP1,TOR3A,TP53 |
| AVP | -0,233 | other | Inhibited | -2,402 | -2,204 | 0,051 | 4,63E-05 | 7,40E-04 | ADM,ATF3,AVP,COL3A1,EGR1,FOS,HSPA5,MYOG,NGF,NOS1,NOS2,NPPA,PRKCA,RRM1,RRM2,S100A6,SCNN1B,SST |
| ERBB3 | -0,009 | kinase | | -0,356 | -0,403 | -0,009 | 4,68E-05 | 7,47E-04 | ADAM9,BAX,CD47,CDKN1B,CLU,COL1A1,DUSP1,EGR1,F3,FBN1,FN1,GHR,HBEGF,HIF1A,HMOX1,IER2,MXI1,PRL,PTEN,RRAS,SERPINA3,SLPI,SNAI1,SPINK4,TNC,USP14,VEGFA |
| ITGAV | 0,474 | transmembrane receptor | | 1,309 | 1,471 | 0,046 | 4,72E-05 | 7,51E-04 | BAX,BIRC2,CDK1,CDKN1B,COL1A1,COL1A2,FN1,FOS,ITGAV,ITGB3,LDLR,MMP1,PCNA,TIMP1,TP53 |

| | | | | | | | | | |
|---|---|---|---|---|---|---|---|---|---|
| NCOA1 | | transcription regulator | | -0,830 | -0,950 | -0,022 | 4,79E-05 | 7,60E-04 | ABCA1,ADIPOQ,AGRN,AHR,ALDOA,BTG2,C3,CASP7,CASP8,CD24,CSHL1,CYP2B6,DIO1,EGR1,ELK1,FOS,FRAT2,HIF1A,HNF1A,HSD3B2,JUNB,KLK3,LEP,LHB,LYZ,MAPK8IP2,NFKBIA,NOCT,NR0B2,PCK1,PDCD2,PKLR,PMM2,PPARG,PRL,PSMG2,PTGES2,RB1,TLE1 |
| TGM2 | 0,238 | enzyme | Activated | 2,273 | 2,518 | 0,034 | 4,85E-05 | 7,67E-04 | ALPL,BTG2,C3,CA2,CCL22,CD74,CSF3,CX3CR1,DAAM2,DAPK2,DCSTAMP,DDX60,DLEU1,FCER1G,FN1,GCA,GIMAP6,GM2A,HIF1A,HLA-B,IFI35,IFIT2,IFIT3,IFIT5,IL10RA,IRF9,ITGA6,ITGB3,NCF2,OAS1,OLFM4,PIM1,PLAAT4,PLSCR1,PPP1R16B,SACS,SDCBP,SEMA6B,SEMA7A,SLPI,SP110,SPP1,STAT1,TAP1,TNFRSF11B,TOR1B,TP53,TPCN1,TRIM22,TYROBP,UBXN8,VSIG4,WDR3,ZNF146 |
| CNR1 | | G-protein coupled receptor | | 0,500 | 0,715 | 0,036 | 4,86E-05 | 7,67E-04 | ACACA,ADIPOQ,ALPL,APBB1,BMP7,CA9,CCNB2,CDK1,DDX25,DRD2,DRD5,DYRK2,EAF2,EGF,FOS,GAD1,HAGH,HEYL,HPRT1,HSPA4,KLK7,LEP,LIPC,MDK,NPHS1,NPHS2,PCNA,PDYN,PECAM1,PLSCR1,RAC1,S100A6,SLC2A4,SLIT2,STAT3,TIMP1,UPK2,WNT6 |
| MAP3K7 | | kinase | | 0,216 | 0,384 | 0,035 | 4,99E-05 | 7,83E-04 | CCL20,CFB,CXCL6,DDX58,GBP1,GBP2,GCH1,HMOX1,IFI44,IFIH1,IFIT2,IFIT3,IFITM1,IRF7,MAP2K3,MMP1,MYOD1,MYOG,NFKB1,NKX2-5,NPPA,RGS2,SLPI,TERT,TNFSF10 |
| TLR3 | 0,546 | transmembrane receptor | Activated | 1,664 | 2,155 | 0,072 | 5,02E-05 | 7,87E-04 | ABCA1,ATF3,C3,CAST,CCL1,CCL17,CCL20,CD70,CFB,CLCA1,CLU,CSF3,CTH,DDX58,DEFB1,DNMT1,DUSP1,FAS,FOS,FURIN,GADD45B,GBP2,GCH1,HMOX1,IFI16,IFI44,IFIH1,IFIT2,IFIT3,IL13,IL15,IL36A,IRF7,ISG15,JUNB,LCN2,LIPA,MMP1,MX1,NFKB1,NMI,NOS2,NR4A1,OAS1,PECAM1,PIK3R3,PLK2,PPP1CA,RHOB,SLPI,SPRY1,STAT1,TIMP1,TLR3,TNC,TNFSF10,TREML2,TRIM38,ZBTB32,ZC3HAV1 |
| PPARD | | ligand-dependent nuclear receptor | | -0,617 | -0,384 | 0,033 | 5,11E-05 | 7,98E-04 | ACAA1,ACACA,ACACB,ALDH2,ALDH3A2,APOA1,APOB,C1QA,C1QB,CDK1,CDK4,CDKN1B,CSAD,CYP2B6,ECI2,FABP1,FBP1,FN1,GAS6,GCK,GPD1,HSD11B2,KYAT1,LPCAT3,LPL,MAPK1,MYCL,MYOM2,NOS2,NPC1L1,NPPA,OCEL1,PCNA,PDE4C,PDK4,PKLR,PNPLA2,PPARG,PTEN,RBP1,S100A6,SCARB2,SLC10A2,SLC2A4,STAT1,TBX21,TNFRSF11B,TP53,UCP2,VEGFA,VLDLR,YWHAE,ZBED5 |
| miR-16-5p (and other miRNAs w/seed AGCAGCA) | | mature microRNA | Inhibited | -2,774 | -3,328 | -0,080 | 5,18E-05 | 8,08E-04 | ACTR1A,ARL2,CAPRIN1,CDK7,CSHL1,EIF4G2,FNDC3B,GALNT7,GFPT1,GOLPH3L,GRB10,GRN,GTF2H1,HARS1,HERC6,HMOX1,HPF1,HSP90B1,IFRD1,IPO4,KCNN4,KITLG,LAMC1,LAMTOR3,MLLT11,MSH2,NAA15,NOTCH2,PANX1,PDCD4,PHKB,PMS1,PNP,PPIF,RARS1,RIDA,SERPINE2,SKAP2,SLC12A2,SPTLC1,SQSTM1,TIA1,TMEM251,TMEM43,TPPP3,UCP2,VEGFA,VPS45,WEE1,WT1 |
| Histone h3 | | group | Inhibited | -2,795 | -2,621 | 0,066 | 5,38E-05 | 8,38E-04 | ACKR3,ADAM10,ADD2,ADGRB1,ADIPOQ,AFP,AKAP12,AQP5,ARHGEF17,ARR3,ATF7IP,ATOH1,B2M,BAHCC1,CA9,CCNA2,CCNB1,CDK14,CDK4,CDKN1B,CDX2,CGA,CKAP4,CLPS,COL1A2,CRIP2,CSHL1,CTNNA1,CYP3A5,DIO1,DUSP2,EGR1,ENO3,EPHB3,ESR1,EVX1,FGF5,FOS,GAD1,GATA6,GHR,GLI1,HAND1,HNF4A,HOXA10,HOXB1,HOXB6,IGFBP4,IL13,IL6R,INHA,INS,IRF7,KLK3,MAD2L1,MCM2,MCM3,MCM5,MET,MGLL,MPZ,MX1,MYBL2,NFKBIA,NOS2,PADI3,PCNA,PKLR,PPARG,PROS1,PTEN,RAB38,RB1,RGS10,SATB2,SGK1,SNAI1,SOD2,SPINK4,SPP1,SYNE2,TBX2,TBX21,TBX6,TERT,TLR2,TMPRSS2,TNFSF10,TYRO3,USP3,VAMP8,VEGFA,WDR1,WNT8B,ZNF271P |
| CFTR | 0,009 | ion channel | | -1,808 | -1,838 | -0,006 | 5,43E-05 | 8,40E-04 | ABCA1,ACAA1,ASNS,BLNK,CAPG,CFI,CUBN,CYP2B6,CYP3A5,CYP4A11,DMBT1,FAAH,FABP2,FAS,GML,HADH,HPGD,HSD17B4,IDI1,MYL7,NFKB1,NFKBIA,NOS2,SLC37A4,SPINK4,STAT1 |
| PTK2 | 0,165 | kinase | | 0,505 | 0,635 | 0,033 | 5,43E-05 | 8,40E-04 | BIRC2,CDKN1B,COL1A1,COL1A2,FN1,FOS,LDLR,LPP,MMP7,NPPA,P4HA1,PCOLCE2,PTK2,SERPINB1,SNAI1,SPP1,TIMP1,TNC,TP53,VCAM1 |
| SERPINF1 | 0,589 | other | | -1,065 | -1,084 | -0,005 | 5,43E-05 | 8,40E-04 | ADIPOQ,BAX,BID,CCN2,CRYAB,CYBA,CYBB,G0S2,LOXL1,NFKB1,NFKBIA,NGF,NOS2,PPARG,RLBP1,SOD2,SOX10,TNFRSF11B,TNFSF10,VIM |
| Atg5 | | other | | 0,483 | 0,447 | -0,016 | 5,46E-05 | 8,41E-04 | BAX,BID,CASP3,NOS2,SQSTM1 |
| RCHY1 | | enzyme | | -0,495 | -0,651 | -0,080 | 5,46E-05 | 8,41E-04 | CDKN1B,HDAC1,KLK3,TP53,TP73 |

| | | | | | | | | | |
|---|---|---|---|---|---|---|---|---|---|
| SOX2 | | transcription regulator | | 1,333 | 1,227 | -0,011 | 5,51E-05 | 8,47E-04 | ABCC3,ABCC6,ABCG2,ABTB2,ADRA2B,AHR,ALB,ALDH1A1,ALOX5,ANKRD17,ARPC1B,ATF3,AXIN1,BHLHE40,C1QL1,CCN2,CCN5,CD44,CDKN1B,CDX2,CER1,CITED2,CLDN11,COL11A2,COL1A1,CRABP2,CST6,CYP26A1,DLX5,DNMT1,DUSP1,ELOVL1,EPB41L2,EPCAM,EVX1,EXT2,FABP1,FABP2,FARP1,FGF5,FOS,FOXF1,FOXH1,FOXO4,FUT4,GADD45B,GATA6,GCK,GJC2,GLI3,HAND1,HHAT,HOPX,HOXB1,KITLG,KLK14,KRT19,LIMS2,MPZL1,NEUROG1,NFASC,NOTCH4,NRP1,OVOL1,PECAM1,PIK3R3,PLXNB3,PPARA,PPARG,PPL,PRDM14,PRKCD,PROM1,S100A1,S100A13,SIRT2,SLC12A2,SMAD1,SNAI1,SOX10,SOX13,SOX21,SPRR3,SPRY4,STRA6,TBX6,TGM1,TIA1,TNNI1,TNR,TXNRD1,UGT8,VCAN,VEGFA,VIM,WNT8B,YAP1 |
| IFN Beta | | group | Activated | 3,045 | 3,375 | 0,059 | 5,73E-05 | 8,78E-04 | ATF3,CASP4,CD74,CD83,CYP3A4,DDX58,DPP4,DUSP1,FN1,HLA-A,HLA-B,HLA-G,IFI16,IFI35,IFI44,IFIH1,IFIT2,IFIT3,IFITM1,IFNAR2,IL15,IRF7,IRF9,ISG15,MX1,NOS2,OAS1,PRKCA,SPRY1,SPRY2,SPRY4,STAT1,TAPBP,TLR3,TNFSF10,TP53,TXNRD1,ZBP1 |
| cholecalciferol | | chemical - endogenous mammalian | Activated | 2,102 | 2,348 | 0,044 | 5,82E-05 | 8,88E-04 | ADGRG1,ALOX5,ANXA2,BECN1,CD83,CDA,CDKN1B,COL1A1,CSTF2T,CYP24A1,FABP6,FOS,FOXP3,ITGB2,LY75,ME1,MS4A6A,MYO6,NR0B2,PIP5K1B,PPARG,PRKX,PTPRB,PVALB,RB1,RORC,SPP1,TNFRSF11B,TNFSF10,TREML2,UBE2E1,VDR,WT1 |
| THRA | | ligand-dependent nuclear receptor | | -0,962 | -0,949 | 0,003 | 5,85E-05 | 8,91E-04 | ACACA,APOA1,APP,CCL17,CD44,CDKN1B,CDX1,CDX2,CGA,CSHL1,CSNK1A1,CYP27B1,DIO1,DIO3,DR1,FOS,FURIN,HMOX1,IL7,KLF9,LDLR,LPL,MC4R,MMP7,NME1,NPPA,NRGN,PCK1,PPARG,PRL,PROM1,PTTG1,RB1,SLC2A4,THRB,TP53 |
| DCN | -0,085 | other | | -0,425 | -0,459 | -0,008 | 5,92E-05 | 9,00E-04 | BAX,CASP3,CASP8,CCN2,CDKN1B,CDKN1C,COL1A2,DUSP1,FBN1,HIF1A,HLA-DQB1,ITGAV,MET,MMP1,MYOG,NOS2,TP53,VCAM1,VEGFA |
| LDLR | -0,509 | transporter | | 0,955 | 0,892 | -0,020 | 6,11E-05 | 9,22E-04 | ABCA1,ABCG1,ACACA,APOB,AURKA,C1QA,CCNA2,CCNB1,CCNB2,CD47,CDC25B,CDK1,CFB,CX3CR1,CYP7B1,DDIT3,DUSP1,EGR1,ELOVL6,F3,FGL2,FOXP3,G6PC,GADD45G,GAS6,GATM,GRAP2,HSPA5,IRF7,ITGB3,KIF20A,LCN2,LDLR,LPL,LYZ,NOD1,PLTP,PPARA,PPARG,PPP1R15A,PRC1,PSMB9,SIGLEC9,SPP1,SYN1,TAP1,TLR1,UBE2C,VCAM1,VCAN,VLDLR,ZBP1 |
| FOXO4 | -0,303 | transcription regulator | | -1,691 | -1,753 | -0,012 | 6,15E-05 | 9,27E-04 | ACACA,ANXA3,C1QA,CASP8,CCN2,CDKN1B,COL4A1,COL4A2,DHCR24,ELOVL6,GADD45A,GADD45B,HIF1A,IDI1,IL10RB,ITGB2,NAMPT,OVOL1,PLA2G2D,PRKCD,PTPRB,PTPRC,SGK1,SOD2,SPTLC2,VCAM1,VEGFA,ZFAND5 |
| MYCN | | transcription regulator | | -0,864 | -0,738 | 0,017 | 6,21E-05 | 9,34E-04 | ABCA1,ABCC1,ABCC3,ABCC4,ABCE1,ABCG2,ABCG4,ACTG1,ALDH1A1,ALDOA,ARPC1B,B2M,BAX,BID,CCN2,CDK7,CDKN1B,CITED2,CKAP4,CLU,COL18A1,COL1A1,COL4A1,COL5A2,DUSP2,EED,EIF4G2,FBN1,FGF5,FN1,GATA6,HLA-A,KDM5B,KLK3,LGALS1,MXI1,MYL12A,NBN,NDRG1,NME1,ODC1,P3H1,PDIA4,POLR2L,PTK2,RBBP7,RPL13A,RPLP2,RPS19,RUSC2,S100A10,SLC16A7,SLC1A2,TERT,TMED9,TMPRSS2,TP53,TPM1,TUBA1B,TUBB,UBE2V2,UCHL1,VIM,ZFAND5 |
| ATM | | kinase | | 0,717 | 0,916 | 0,054 | 6,55E-05 | 9,80E-04 | ADIPOQ,BAX,BIRC2,CCN2,CD70,CDK4,CDKN1B,CLU,DUSP1,FAS,FDXR,FN1,GADD45A,GADD45B,GADD45G,HMOX1,LEP,PCNA,PSMC4,RRM1,RRM2,TP53,XRCC5 |
| MAP2K3 | -0,241 | kinase | | -1,122 | -0,838 | 0,070 | 6,55E-05 | 9,80E-04 | BAX,CDKN1B,COL1A1,COL1A2,COL3A1,DDIT3,GP9,GSTP1,IRF9,ISG15,IVL,MMP1,MUC5B,NOS2,NPPA,PPA1,RGS7,SGK1,SLC2A4,SLC5A5,STAT1,TNFSF10,VEGFA |
| SPI1 | | transcription regulator | | 0,782 | 0,989 | 0,031 | 6,70E-05 | 9,98E-04 | BAX,BCL7A,BLNK,CCL17,CCL22,CCNA2,CCNB1,CCNB2,CDK1,CDK4,CEBPE,CHI3L1,CTSS,CYBB,DCSTAMP,EPB41L2,EPX,FCER1G,FCGRT,FOS,FOXO1,FUT7,IFI44,IFIT2,IFIT3,IFITM1,IL13,IL18,IRF7,IRF9,ISG15,ITGB2,KLK1,KLK3,LBR,LILRB3,LTF,LYZ,MME,MMP1,MT1X,MX1,NCF2,NKX2-5,NOS2,PARP12,PCNA,PRG2,PSMB10,PSMB8,PSMB9,PTCRA,PTEN,PTPRC,RACK1,SP110,SPP1,TLR2,TNFSF10,TRIM22,VIM |
